# An Ensemble Deep Learning Algorithm for Structural Heart Disease Screening Using Electrocardiographic Images: PRESENT SHD

**DOI:** 10.1101/2024.10.06.24314939

**Authors:** Lovedeep S Dhingra, Arya Aminorroaya, Veer Sangha, Aline F Pedroso, Sumukh Vasisht Shankar, Andreas Coppi, Murilo Foppa, Luisa CC Brant, Sandhi M Barreto, Antonio Luiz P Ribeiro, Harlan M Krumholz, Evangelos K Oikonomou, Rohan Khera

## Abstract

**Background:** Identifying structural heart diseases (SHDs) early can change the course of the disease, but their diagnosis requires cardiac imaging, which is limited in accessibility.

**Objective:** To leverage images of 12-lead ECGs for automated detection and prediction of multiple SHDs using an ensemble deep learning approach.

**Methods:** We developed a series of convolutional neural network models for detecting a range of individual SHDs from images of ECGs with SHDs defined by transthoracic echocardiograms (TTEs) performed within 30 days of the ECG at the Yale New Haven Hospital (YNHH). SHDs were defined as LV ejection fraction <40%, moderate-to-severe left-sided valvular disease (aortic/mitral stenosis or regurgitation), or severe left ventricular hypertrophy (IVSd > 1.5cm and diastolic dysfunction). We developed an ensemble XGBoost model, PRESENT-SHD, as a composite screen across all SHDs. We validated PRESENT-SHD at 4 US hospitals and the prospective, population-based Brazilian Longitudinal Study of Adult Health (ELSA-Brasil), with concurrent protocolized ECGs and TTEs. We also used PRESENT-SHD for risk stratification of new-onset SHD or heart failure (HF) in clinical cohorts and the population-based UK Biobank (UKB).

**Results:** The models were developed using 261,228 ECGs from 93,693 YNHH patients and evaluated on a single ECG from 11,023 individuals at YNHH (19% with SHD), 44,591 across external hospitals (20-27% with SHD), and 3,014 in the ELSA-Brasil (3% with SHD). In the held-out test set, PRESENT-SHD demonstrated an AUROC of 0.886 (0.877-894), 90% sensitivity, and 66% specificity. At hospital-based sites, PRESENT-SHD had AUROCs ranging from 0.854-0.900, with sensitivities and specificities of 93-96% and 51-56%, respectively. The model generalized well to ELSA-Brasil (AUROC, 0.853 [0.811-0.897], 88% sensitivity, 62% specificity). PRESENT-SHD demonstrated consistent performance across demographic subgroups, novel ECG formats, and smartphone photographs of ECGs from monitors and printouts. A positive PRESENT-SHD screen portended a 2- to 4-fold higher risk of new-onset SHD/HF, independent of demographics, comorbidities, and the competing risk of death across clinical sites and UKB, with high predictive discrimination.

**Conclusion:** We developed and validated PRESENT-SHD, an AI-ECG tool identifying a range of SHD using images of 12-lead ECGs, representing a robust, scalable, and accessible modality for automated SHD screening and risk stratification.

**CONDENSED ABSTRACT:** Screening for structural heart disorders (SHDs) requires cardiac imaging, which has limited accessibility. To leverage 12-lead ECG images for automated detection and prediction of multiple SHDs, we developed PRESENT-SHD, an ensemble deep learning model. PRESENT-SHD demonstrated excellent performance in detecting SHDs across 5 US hospitals and a population-based cohort in Brazil. The model successfully predicted the risk of new-onset SHD or heart failure in both US clinical cohorts and the community-based UK Biobank. By using ubiquitous ECG images and smartphone photographs to predict a composite outcome of multiple SHDs, PRESENT-SHD establishes a scalable paradigm for cardiovascular screening and risk stratification.

## BACKGROUND

Structural heart diseases (SHDs) represent a spectrum of prevalent cardiac disorders with a long presymptomatic course and with substantially elevated risk of heart failure (HF) and premature death.^1^ The detection of these disorders has traditionally required advanced cardiac imaging, including echocardiography and cardiac magnetic resonance imaging, which are resource-intensive and, therefore, not suitable for large-scale disease screening.^2,3^ Consequently, these conditions are often diagnosed after the development of clinical symptoms, leading to poor health outcomes.^4–6^ Moreover, there are no strategies to identify individuals at risk of developing SHDs, despite the presence of evidence-based interventions that can alter the course of patients.^6–8^ Thus, there is an urgent need for the development of an automated, accessible, and scalable strategy for the screening and risk stratification of SHDs.^1,9^

Previously applications of artificial intelligence for electrocardiograms (AI-ECG) have shown potential to detect signatures of SHDs.^10–18^ A key challenge of AI-ECG models in detecting specific cardiac disorders using ECGs is the low precision driven by the low prevalence of individual disorders.^10–12^ To overcome this limitation, ensemble models for detecting a composite of multiple SHDs have been proposed.^19^ Nonetheless, these models use raw ECG voltage data as inputs, which are inaccessible to clinicians at the point of care and often require modifications to the technical infrastructure to account for vendor-specific data formats.^19^ This precludes the widespread use of AI-ECG approaches for broad cardiovascular screening, as these data integrations are not commonly available. Further, most AI-ECG approaches focus on cross-sectional detection and do not quantify the risk of new-onset disease in those without SHD, which would identify a group for continued monitoring. Thus, there is a critical unmet need for an AI-ECG-based strategy to enable cross-sectional detection and longitudinal prediction of multiple SHDs simultaneously using ubiquitous, interoperable, and accessible data input in the form of ECG images.

In this study, we report the development and multinational validation of an ensemble deep learning approach that uses an image of a 12-lead ECG, independent of the format, for the accurate detection and prediction of multiple SHDs.

## METHODS

The Yale Institutional Review Board approved the study protocol and waived the need for informed consent as the study involves secondary analysis of pre-existing data. An online version of the model is publicly available for research use at https://www.cards-lab.org/present-shd.

### Data Sources

For model development, we included data from the Yale New Haven Hospital (YNHH) during 2015-2023. YNHH is a large 1500-bed tertiary medical center that provides care to a diverse patient population across Connecticut. For external validation of our approach to detect SHDs, we included multiple clinically and geographically diverse cohorts: (i) 4 distinct community hospitals in the Yale-New Haven Health System, the Bridgeport Hospital, Greenwich Hospital, Lawrence + Memorial Hospital, and Westerly Hospital, and (ii) a community-based cohort of individuals in Brazil with protocolized concurrent ECG and TTE assessments, the ELSA-Brasil study.

To evaluate the longitudinal prediction of SHD in people without baseline disease, in addition to serial monitoring data from hospitals in the Yale-New Haven Health System, we included data from the UK Biobank (UKB). UKB is the largest population-based cohort with protocolized ECG assessments and clinical encounters derived from the integrated EHR of the National Health Service in the UK. An overview of all data sources is included in the **Supplementary Methods**.

### Study Population for SHD Detection

At YNHH, we identified all adults (≥18 years) who underwent a 12-lead ECG within 30 days before or after a transthoracic echocardiogram (TTE), excluding those with prior cardiac surgery to replicate the intended use of these models in a screening setting (**Central Illustration; Supplementary Figure 1**). In the internal validation and internal held-out test sets, and all external validation sites, one ECG was randomly selected from one or more ECGs performed within 30 days of a TTE for each individual. In ELSA-Brasil, all participants who underwent both ECG and TTE at their baseline study visit were included.

**Central Illustration.**
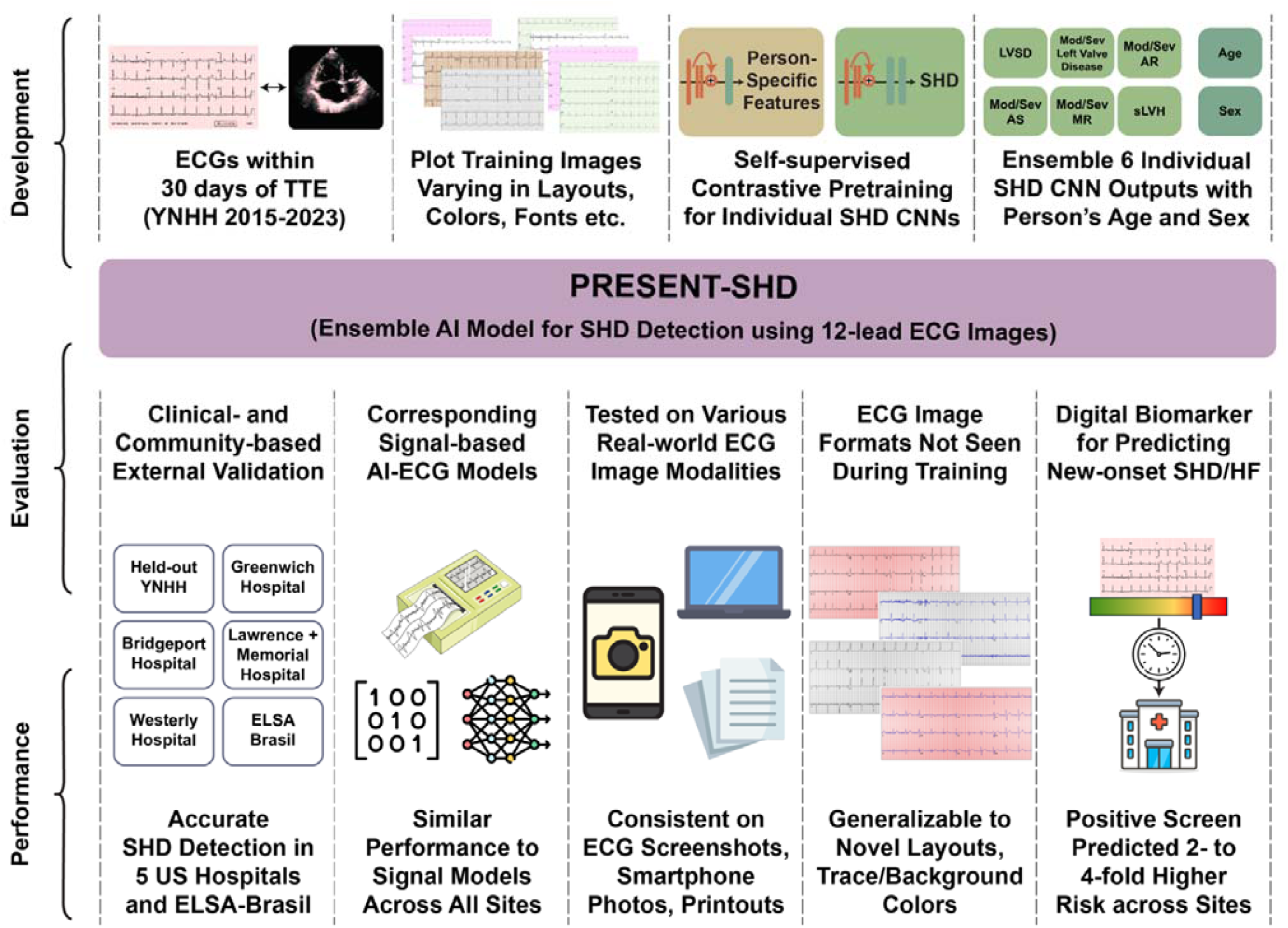
Study Design. Abbreviations: AR, aortic regurgitation; AS, aortic stenosis; CNN, convolutional neural network; ECG, electrocardiogram; FC, fully-connected layers; LVSD, left ventricular systolic dysfunction; MR, mitral regurgitation; SHD, structural heart diseases; sLVH, severe left ventricular hypertrophy; TTE, transthoracic echocardiogram.

### SHD Outcome

The study outcome of SHD was defined as any LVSD, moderate-or-severe left-sided valve disease, or sLVH. All conditions were ascertained based on the interpretation of TTE by board-certified cardiologists according to the American Society of Echocardiography guidelines.^20^ Echocardiography variables were available in tabular format, obviating manual ascertainment. The left ventricular ejection fraction (LVEF) was primarily measured as a continuous variable using the biplane method. When the LVEF measurement using the biplane method was unavailable, measurements using the three-dimensional or visual estimation methods were used. LVSD was defined as an LVEF < 40%. Left-sided valve diseases included aortic stenosis (AS), aortic regurgitation (AR), mitral regurgitation (MR), or mitral stenosis (MS), graded as mild to moderate, moderate, moderate to severe, or severe, based on established echocardiographic guidelines.^21,22^ We defined sLVH by a combination of an interventricular septal diameter at end-diastole (IVSd) of greater than 15 mm, along with moderate to severe (grade II and grade III) LV diastolic dysfunction.^23^

### Signal Processing and Image Generation

We used a strategy for developing models that can detect SHD from images from ECGs regardless of their layout. This was done using a custom waveform plotting strategy where ECG signals are processed and plotted as images in a format randomly chosen from 2880 formats, encompassing variations in lead layout, trace and background color, lead label font, size and position, and grid and signal line width (**Supplementary Figure 2**). We also included non-conventional variations in ECG lead placements, with the chest leads on the left and limb leads on the right side of the plotted ECGs. The plotted signals were processed using a standard preprocessing strategy described previously (and included in **Supplemental Methods**). For evaluation, ECG images were plotted in standard clinical layout from signal waveform data, with a voltage calibration of 10 mm/mV, with the limbs and precordial leads arranged in four columns of 2.5-second each, representing leads I, II, and III; aVR, aVL, and aVF; V1, V2, and V3; and V4, V5, and V6 (**Supplementary Figure 3**). A 10-second recording of the lead I signal was included as a rhythm strip. We further evaluated the model on 4 novel image formats that were not encountered during model training (**Supplementary Methods**; **Supplementary Figure 4**). All images were converted to greyscale and down-sampled to 300×300 pixels using Python Image Library.^24^ Examples of ECG images used for model training and evaluation are presented in **Online Supplement 2**.

### Model Development for Individual SHDs

We trained 6 independent convolutional neural network (CNN) models to detect individual components of SHD (**Central Illustration**). We randomly divided individuals at YNHH into training, validation, and test sets (85:5:10) without any patient spanning these sets (**Supplementary Figure 1**). We retained multiple ECGs per person in the training set to ensure the adequacy of training data. However, in evaluating the model in the internal validation, held-out test, and external validation sets, only one ECG was randomly chosen for every individual. Of note, none of the patients in the external validation sets were in the model development population.

We used CNN models built upon the EfficientNet-B3 architecture, which has 384 layers and over 10 million trainable parameters.^11,25^ To enable label-efficient model development, we initialized the CNNs with weights from a model pretrained to recognize individual patient-specific patterns in ECGs, independent of their interpretation, using a self-supervised, contrastive learning framework (**Figure 1**).^26^ None of the ECGs on the self-supervised pretraining task represented individuals in the SHD model development.

**Figure 1.**
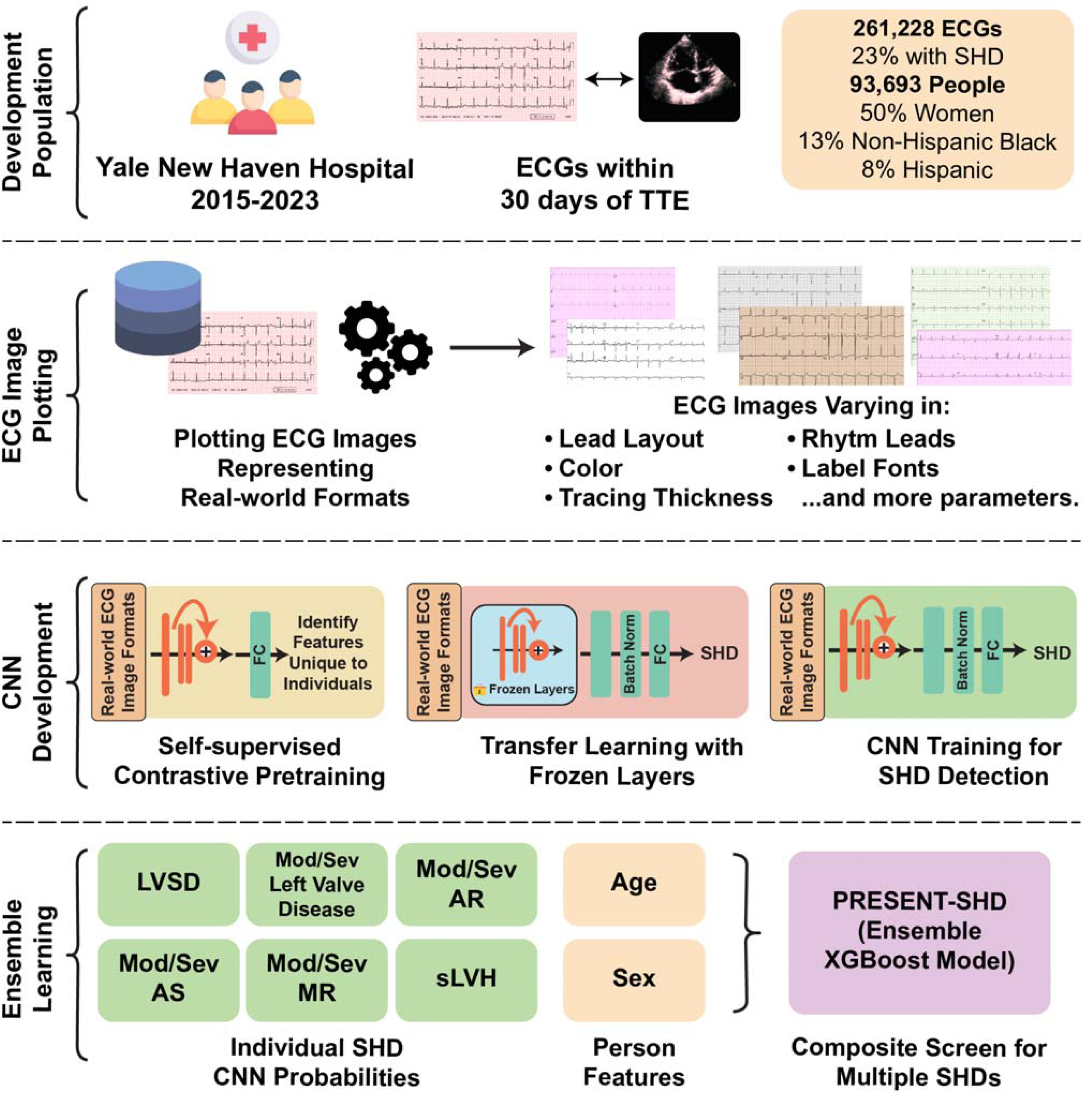
Model Development Strategy. Abbreviations: AR, aortic regurgitation; AS, aortic stenosis; CNN, convolutional neural network; ECG, electrocardiogram; FC, fully-connected layers; LVSD, left ventricular systolic dysfunction; MR, mitral regurgitation; SHD, structural heart diseases; sLVH, severe left ventricular hypertrophy; TTE, transthoracic echocardiogram; XGBoost, extreme gradient boosting

Each ECG in the training set was plotted using one of the randomly assigned plotting formats described above. We used an Adam optimizer, gradient clipping, and a minibatch size of 128 throughout training, with sequential unfreezing of the final layers (learning rate, 0.001), and all layers (learning rate, 10^-5^), with training stopped when validation loss did not improve in 5 consecutive epochs. A custom class-balanced loss function (weighted binary cross-entropy) based on the effective number of samples was used, given the case and control imbalance.

The CNNs for the individual components of SHD had the same model backbone but differed in the populations for training. Five of the 6 models, specifically for those detecting LVSD, the presence of any moderate to severe left-sided valvular heart diseases, and those for moderate-to-severe AR, AS, or MR, were trained using all ECGs in the training set, spanning those with and without each disease. However, given the low prevalence of sLVH (<1%), we age- and sex-matched cases and controls for model development. Each case, representing an ECG corresponding to an individual with sLVH, was matched to 10 control ECGs without sLVH from someone of the same sex and within 5 years of age as the case. These individual models were combined in an ensemble model to detect the presence of any SHD. As a sensitivity analysis, we used the same training strategy and model architecture to develop a classifier CNN model directly detecting the presence of SHD. For each SHD component, we also trained corresponding signal-based models within the same label and training population (**Supplementary Methods**).

### Ensemble Learning Strategy

Following CNN development, output probabilities from the 6 component SHD CNN models, along with a person’s age and sex, were used as input features for an extreme gradient boosting (XGBoost) model, PRESENT-SHD (Practical scREening using ENsemble machine learning sTrategy for SHD detection; **Figure 1**). The XGBoost model was exclusively trained using data from the same training sets as the CNN models. Before being included as features, age and the CNN model output probabilities were standardized to a mean of 0 and a variance of 1 to improve learning stability and ensure consistent feature contribution across different datasets. The standardization algorithm was derived based on the distribution of these variables in the training set and was applied for inference across all other datasets, including internal validation, testing, and external validation sets.

### Model Evaluation on Screenshots and Smartphone Photographs of ECGs

To evaluate model performance across different strategies for obtaining images in the real world, we selected a random subset of 100 ECGs from the held-out test set. From the EHR, we identified the PDFs for these ECGs and saved screenshots to reflect the exact images used during clinical care. We then photographed the ECGs from a laptop screen displaying them. We also printed them on A4 sheets and photographed using default camera settings on three different smartphones (**Supplementary Figure 5; Supplementary Methods**). Finally, we applied the 6 SHD-component CNNs and the PRESENT-SHD model to these screenshots and smartphone photographs (**Online Supplement 2**), compared the predictions with the images of the same ECGs plotted in the standard format, and evaluated performance for SHD detection.

### Prediction of New-onset SHD

To evaluate the use of the model for stratifying the risk of new-onset disease across data sources, we identified a population without evidence of SHD or HF at baseline. In YNHH, we identified the first recorded encounter for all individuals within the EHR and instituted a 1-year blanking period to define prevalent diseases (**Supplementary Figure 6**). Among 204,530 patients with ECGs following a one-year blanking period, we excluded 6,909 individuals with prevalent SHD, 1,197 with a prior valvular repair or replacement procedure, and 13,632 with prevalent HF (**Supplementary Table 1**). Those included in the model training set (n = 55,245) were also excluded from this analysis. We used a similar strategy across the hospital-based external validation sites to identify patients at risk for new-onset disease – a one-year blanking period to identify prevalent diseases and exclude those with prevalent SHD/HF, prior valvular procedures. Across sets, new-onset SHD/HF was defined as the first occurrence of any SHD detected on the TTE, any valvular replacement or repair procedure, or hospitalization with HF. Data were censored at death or loss to follow-up.

Further, we identified participants with ECGs in the UKB. We used the national EHR linkage to identify those who had not undergone any hospitalizations with HF and had not undergone valvular procedures before their baseline ECG. We followed these individuals till their first encounter with an SHD or HF diagnosis code or a left-sided valve replacement or repair procedure (**Supplementary Table 1**).

### Statistical Analysis

We reported continuous variables as median and interquartile range (IQR), and categorical variables as counts and percentages. Model performance for detecting SHD was reported as area under the receiver operating characteristic curve (AUROC) and area under the precision-recall curve (AUPRC), with 95% confidence intervals (CI) for these computed using bootstrapping with 1000 iterations. Additional performance measures included sensitivity, specificity, positive predictive value (PPV), negative predictive value (NPV), and F1 score with 95% CIs using the standard error formula for proportion. Finally, we calculated the model’s PPV in simulated screening scenarios with different prevalences of composite SHD using the model’s sensitivity and specificity corresponding to the probability threshold with sensitivity above 90% in the internal validation set.

Among those without SHD at baseline, the predictive role of PRESENT-SHD for new-onset SHD/HF was evaluated in age- and sex-adjusted Cox proportional hazard models. The time-to-first SHD/HF event was the dependent variable and the PRESENT-SHD-based screen status – presumably “false positive” or “true negative” status – was the key independent variable. Further, to account for the competing risk of death while evaluating new-onset SHD, we used age- and sex-adjusted multi-outcome Fine-Gray subdistribution hazard models.^28^ The discrimination of the model for SHD prediction was assessed using Harrell’s C-statistic.^29,30^ The statistical analyses were two-sided, and the significance level was set at 0.05. Analyses were executed using Python 3.11.2 and R version 4.2.0. Our study follows the Transparent Reporting of a multivariable prediction model for Individual Prognosis Or Diagnosis + Artificial Intelligence (TRIPOD + AI) checklist from the EQUATOR network (**Supplementary Table 2**).^31^

## RESULTS

### Study Population

There were 261,228 ECGs from 93,693 unique patients in the training set, and the validation and internal held-out test sets had a single ECG per person from 5,512 and 11,023 patients, respectively (**Supplementary Figure 1**). The development population (model training and validation sets) had a median age of 67.8 (IQR, 56.1-78.3) years, 49,947 were (50.3%) women, 13,383 (13.8%) non-Hispanic Black, and 7,754 were (8.1%) Hispanic (**Supplementary Table 3**). In the development population, 60,096 (22.5%) ECGs were paired with TTEs with an SHD, including 25,552 (9.5%) with LVSD, 42,989 (16.1%) with moderate or severe left-sided valvular disease, and 1,004 (0.4%) with sLVH.

At the external hospital sites, 18,222 patients at Bridgeport Hospital, 4,720 patients at Greenwich Hospital, 17,867 patients at Lawrence + Memorial Hospital, and 3,782 patients from Westerly Hospital were included (**Supplementary Figure 1**), with 44,591 ECGs, randomly one chosen per person, across these sites for model evaluation. Across hospital sites, the median age at ECG ranged from 66 to 74 years, with cohorts comprising 48.3% to 50.5% women, 1.5% to 19.4% Black, and 1.4% to 15.9% Hispanic individuals. The distribution of SHDs across these cohorts are described in **Supplementary Table 4**.

Of the 15,105 participants in ELSA-Brasil, 3,014 who underwent ECG and TTE during their baseline visit were included. The median age of the cohort was 62.0 (IQR, 57.0-67.0) years, 1,596 (53.0%) were women, 1,661 (55.1%) were White, 455 (15.1%) were Black, and 753 (25.0%) were Pardo (or mixed race) individuals. A total of 88 (2.9%) individuals had SHD, with 37 (1.2%) with LVSD, 55 (1.8%) with moderate or severe left-sided valvular disease, and 6 (0.2%) with sLVH (**Supplementary Table 4**).

### Detection of Structural Heart Disease

The ensemble XGBoost model, PRESENT-SHD, demonstrated an AUROC of 0.886 (95% CI, 0.877-0.894) and an AUPRC of 0.807 (95% CI, 0.791-0.823) for the detection of the composite SHD outcome in the held-out test set (**Table 1**). At the probability threshold for sensitivity above 90% in the internal validation set, the model had a sensitivity of 89.8% (95% CI, 89.0-90.5), specificity of 66.2% (95% CI, 65.0-67.4), PPV of 57.4% (95% CI, 56.1-58.6), and NPV of 92.8% (95% CI, 92.1-93.4) for detecting SHD in the held-out test set in YNHH (**Table 2; Supplementary Figure 7**). PRESENT-SHD performed consistently across subgroups of age, sex, race, and ethnicity (**Table 1**), and generalized well to novel ECG formats not encountered during training (**Supplementary Table 5**). Moreover, the model had consistent performance across subsets where TTEs were performed before, on the same day as, or after the ECG (**Supplementary Table 6**) and had high discrimination for detecting the severe SHD phenotype (LVSD, severe left-sided valve disease, or sLVH; **Supplementary Figure 8**). Notably, the performance of PRESENT-SHD was higher than the CNN models trained to directly detect SHD and other XGBoost ensemble strategies (**Supplementary Tables 7 and 8**).

**Table 1.** Performance metrics for detecting structural heart disease across demographic subgroups in the held-out test set. Abbreviations: AUPRC, area under the precision-recall curve; AUROC, area under the receiver operating characteristic curve; NPV, negative predictive value; PPV, positive predictive value.

| Subgroup | Total Number | Diagnostic OR | AUROC | AUPRC | F1 Score | Prevalence | Sensitivity | Specificity | PPV | NPV |
| --- | --- | --- | --- | --- | --- | --- | --- | --- | --- | --- |
| **Overall** | 6203 | 17.2 (14.7-20.1) | 0.886 (0.877-0.894) | 0.807 (0.791-0.823) | 0.7 | 33.60% | 89.8% (89.0-90.5) | 66.2% (65.0-67.4) | 57.4% (56.1-58.6) | 92.8% (92.1-93.4) |
| **Age ≥ 65 years** | 2897 | 8.0 (6.1-10.5) | 0.822 (0.807-0.838) | 0.839 (0.820-0.857) | 0.735 | 53.20% | 95.7% (94.9-96.4) | 26.6% (25.0-28.2) | 59.7% (57.9-61.5) | 84.3% (83.0-85.7) |
| **Age < 65 years** | 3306 | 16.2 (13.1-20.2) | 0.873 (0.856-0.889) | 0.679 (0.641-0.716) | 0.595 | 16.50% | 73.2% (71.7-74.7) | 85.6% (84.4-86.8) | 50.1% (48.4-51.8) | 94.2% (93.4-95.0) |
| **Women** | 3150 | 17.5 (14.1-21.7) | 0.884 (0.873-0.896) | 0.778 (0.752-0.804) | 0.689 | 30.80% | 88.5% (87.3-89.6) | 69.5% (67.9-71.1) | 56.4% (54.7-58.1) | 93.1% (92.2-94.0) |
| **Men** | 3052 | 16.7 (13.3-20.9) | 0.886 (0.874-0.898) | 0.830 (0.810-0.851) | 0.71 | 36.50% | 90.9% (89.9-92.0) | 62.4% (60.7-64.2) | 58.2% (56.4-59.9) | 92.3% (91.3-93.2) |
| **Non-Hispanic White** | 3966 | 16.7 (13.7-20.5) | 0.882 (0.871-0.892) | 0.824 (0.805-0.841) | 0.711 | 37.40% | 91.7% (90.9-92.6) | 60.2% (58.7-61.7) | 58.0% (56.4-59.5) | 92.4% (91.6-93.2) |
| **Non-Hispanic Black** | 834 | 17.2 (11.4-25.9) | 0.877 (0.852-0.902) | 0.774 (0.724-0.823) | 0.697 | 31.50% | 87.8% (85.6-90.1) | 70.4% (67.3-73.5) | 57.8% (54.4-61.1) | 92.6% (90.9-94.4) |
| **Hispanic** | 537 | 15.8 (9.7-25.8) | 0.882 (0.846-0.916) | 0.786 (0.720-0.844) | 0.666 | 25.50% | 81.0% (77.7-84.3) | 78.8% (75.3-82.2) | 56.6% (52.4-60.8) | 92.4% (90.1-94.6) |
| **Others** | 866 | 18.1 (11.9-27.5) | 0.893 (0.868-0.918) | 0.740 (0.668-0.803) | 0.648 | 23.10% | 84.0% (81.6-86.4) | 77.5% (74.7-80.3) | 52.8% (49.5-56.2) | 94.2% (92.6-95.7) |

**Table 2.** Model performance characteristics for PRESENT-SHD for detection of structural heart disease across the held-out test set and external validation cohorts. Abbreviations: AUPRC, area under the precision-recall curve; AUROC, area under the receiver operating characteristic curve; NPV, negative predictive value; PPV, positive predictive value.

| Cohort Type | Site Name | Total Number | Diagnostic OR | AUROC | AUPRC | F1 Score | Prevalence | Sensitivity | Specificity | PPV | NPV |
| --- | --- | --- | --- | --- | --- | --- | --- | --- | --- | --- | --- |
| **Held-out test set** | **Yale New Haven Hospital** | 6203 | 17.2 (14.7-20.1) | 0.886 (0.877-0.894) | 0.807 (0.791-0.823) | 0.700 | 33.60% | 89.8% (89.0-90.5) | 66.2% (65.0-67.4) | 57.4% (56.1-58.6) | 92.8% (92.1-93.4) |
| **External validation – Hospital sites** | **Bridgeport Hospital** | 8944 | 14.8 (12.9-16.9) | 0.854 (0.847-0.862) | 0.834 (0.823-0.845) | 0.751 | 46.60% | 93.2% (92.6-93.7) | 52.0% (51.0-53.1) | 62.9% (61.9-63.9) | 89.7% (89.1-90.3) |
|  | **Greenwich Hospital** | 2271 | 30.6 (22.2-42.1) | 0.900 (0.888-0.913) | 0.894 (0.878-0.910) | 0.798 | 49.80% | 96.0% (95.2-96.8) | 55.9% (53.9-58.0) | 68.3% (66.4-70.2) | 93.4% (92.4-94.4) |
|  | **Lawrence + Memorial Hospital** | 11447 | 16.0 (14.0-18.2) | 0.871 (0.864-0.878) | 0.771 (0.757-0.784) | 0.643 | 31.50% | 92.5% (92.0-93.0) | 56.4% (55.5-57.3) | 49.3% (48.4-50.3) | 94.3% (93.8-94.7) |
|  | **Westerly Hospital** | 1843 | 19.9 (14.5-27.3) | 0.887 (0.874-0.902) | 0.906 (0.890-0.922) | 0.810 | 55.60% | 95.1% (94.1-96.1) | 50.5% (48.3-52.8) | 70.6% (68.6-72.7) | 89.2% (87.8-90.6) |
| **External validation – Population-based cohort** | **ELSA-Brasil** | 2988 | 11.4 (6.0-21.5) | 0.853 (0.811-0.897) | 0.354 (0.253-0.460) | 0.121 | 2.90% | 87.5% (86.3-88.7) | 61.9% (60.2-63.6) | 6.5% (5.6-7.4) | 99.4% (99.1-99.7) |

PRESENT-SHD performance was similar to the corresponding signal-based model for detecting SHD (**Supplementary Table 9**). Across ECG screenshots and smartphone photographs of monitors and printouts, the model demonstrated high agreement with plotted images (Pearson correlation coefficients, 0.959–0.977; **Supplementary Figure 9**) and consistent performance across all image types (AUROCs: Plotted images, 0.939 [95% CI, 0.887-0.976]; ECG screenshots, 0.934 [95% CI, 0.885-0.973]; Smartphone photographs of computer monitors, 0.932 [95% CI, 0.878-0.970]; Smartphone photographs of printouts, 0.924 [95% CI, 0.866-0.970]; **Supplementary Table 10**).

Further, PRESENT-SHD generalized well to the external validation cohorts at Bridgeport (AUROC, 0.854 [95% CI, 0.847-0.862]), Greenwich (AUROC, 0.900 [95% CI, 0.888-0.913]), Lawrence + Memorial (AUROC, 0.871 [95% CI, 0.864-0.878]), and Westerly (AUROC, 0.887 [95% CI, 0.874-0.902]) Hospitals, with sensitivities and specificities ranging 88-96% and 51-66%, respectively. PRESENT-SHD also generalized well to the population-based ELSA-Brasil, with an AUROC of 0.853 (95% CI: 0.811-0.897) and a sensitivity and specificity of 87.5% and 61.9%, respectively (**Table 2**; **Supplementary Table 11**). Across validation sites, model performance was consistent across demographic subgroups (**Supplementary Tables 12-16**). The F1 scores, PPVs, and NPVs for a range of putative prevalences of SHDs representing different screening scenarios are presented in **Supplementary Table 17**.

### Detection of Individual Diseases

The models for LVSD, moderate or severe valvular disease, and sLVH had AUROCs of 0.914 (95% CI, 0.904-0.924), 0.805 (95% CI, 0.793-0.817), and 0.903 (95% CI, 0.850-0.946; **Figure 2**), respectively. The performance of CNN models for individual valvular heart diseases varied, with an AUROC of 0.722 (95% CI, 0.784-0.824) for moderate or severe AR, 0.804 (95% CI, 0.784-0.824) for AS, and 0.792 (95% CI, 0.776-0.807) for MR. The CNN model AUPRCs varied with individual disease prevalence (**Supplementary Tables 18-23**). The performance for individual disease CNNs was consistent across external validation cohorts (**Supplementary Figure 10; Supplementary Tables 18-23**) and real-world ECG image modalities (**Supplementary Table 24**).

**Figure 2.**
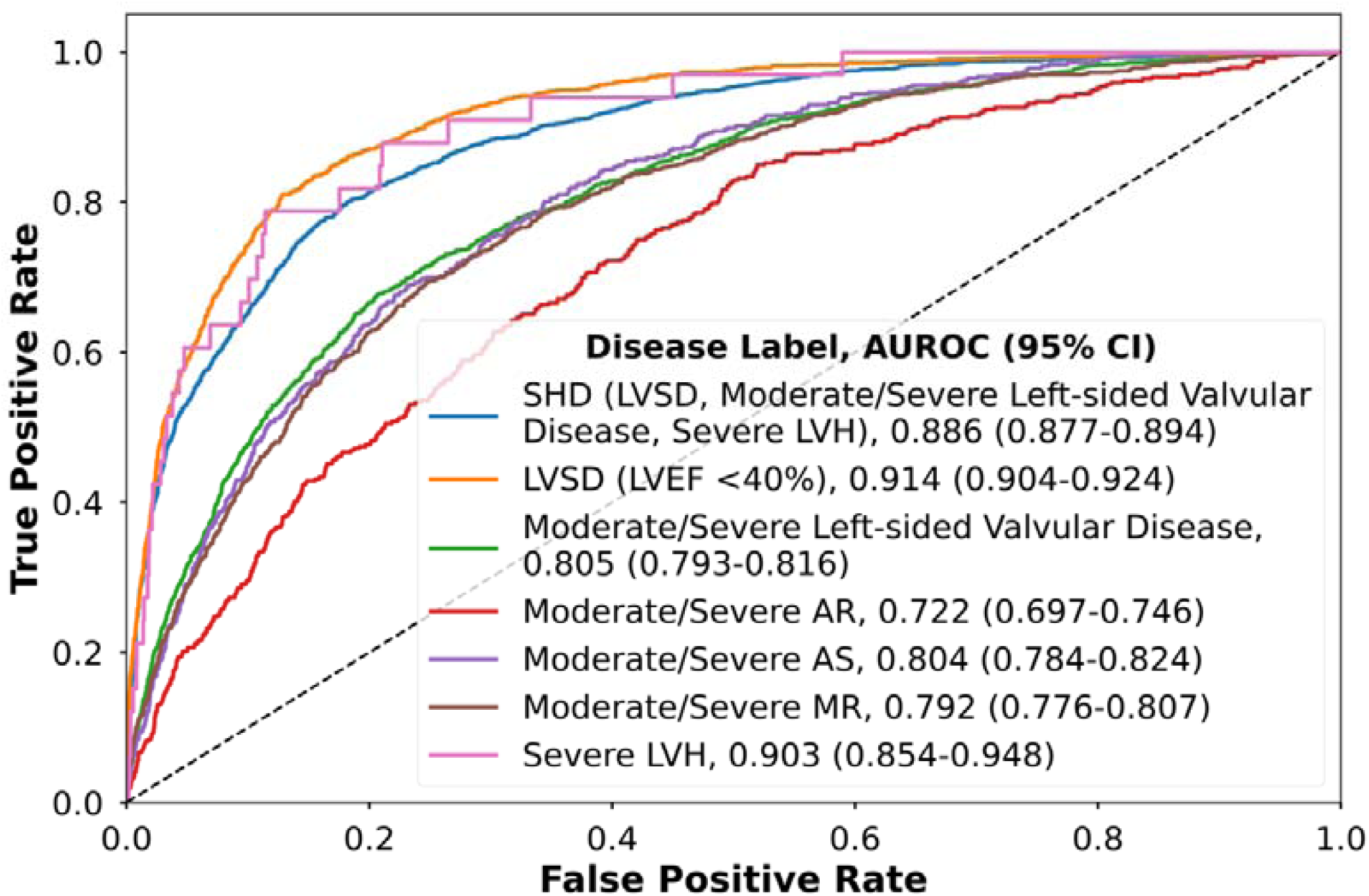
Model discrimination for the detection of composite structural heart disease and individual components in the held-out test set. Abbreviations: AR, aortic regurgitation; AS, aortic stenosis; CI, confidence intervals; LVEF, left ventricular ejection fraction; LVH, severe left ventricular hypertrophy; LVSD, left ventricular systolic dysfunction; MR, mitral regurgitation; SHD, structural heart diseases

### Prediction of SHD and Cardiovascular Risk

Of the 127,547 individuals at risk in YNHH, 5,346 (4.2%) had new-onset SHD/HF over a median of 4.0 (IQR 1.7-6.4) years of follow-up. Across the hospital-based external validation sites, there were 63,748 individuals without SHD/HF at baseline and 4,593 (7.2%) developed incident SHD/HF over a median of 3.1 years (IQR, 1.3-5.0) of follow up (**Supplementary Table 25**). In UKB, 413 (1.0%) of 41,800 individuals developed SHD/HF over 3.0 (IQR 2.1-4.5) years of follow-up.

A positive PRESENT-SHD screen portended a 4-fold higher risk of new-onset SHD/HF in YNHH (age- and sex-adjusted HR [aHR], 4.28 [95% CI, 3.95-4.64], Harrell’s C-statistic, 0.823 [95% CI, 0.817-0.828]) and every 10% increment in model probability was progressively associated with a 36% higher hazard for incident SHD/HF (aHR, 1.36 [1.35-1.38]). A similar pattern was observed across all external validation hospital sites (**Supplementary Tables 26 and 27**). This association remained consistent after adjusting for comorbidities at baseline and the competing risk of death (**Supplementary Table 26**).

In the UKB, a positive vs. negative PRESENT-SHD screen was associated with twice the hazard of developing SHD/HF (aHR, 2.39 [95% CI, 1.87-3.04], Harrell’s C-statistic, 0.754 [95% CI, 0.728-0.780]). Across all sites, higher model probabilities were associated with progressively higher risk of new-onset SHD/HF (**Supplementary Table 28; Figure 3**).

**Figure 3.**
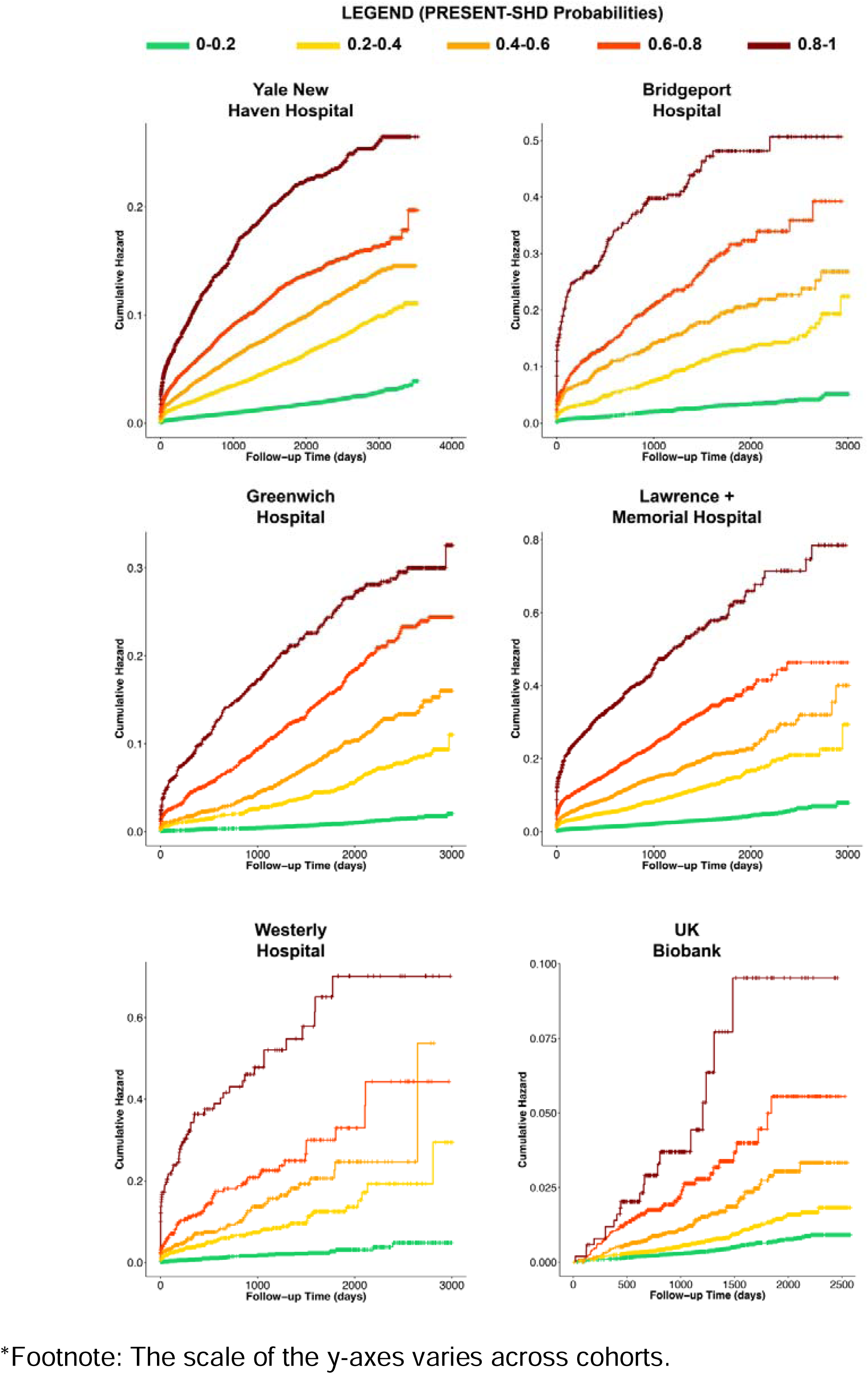
Cumulative hazard for new-onset structural heart disease or heart failure hospitalization in individuals without structural heart disease or heart failure at baseline.

## DISCUSSION

We developed and validated PRESENT-SHD, an ensemble deep learning model that uses an ECG image as the input to detect a range of SHDs. PRESENT-SHD had excellent performance in detecting SHDs across five distinct US hospitals with unique patients and in a population-based cohort study from Brazil. Model performance was consistent across demographic subgroups and similar to the corresponding signal-based models. Additionally, PRESENT-SHD maintained high performance when tested on novel ECG formats, screenshots of ECGs in the EHR, as well as smartphone photographs of ECGs captured from laptop monitors and printouts. Further, in individuals without SHD at baseline, PRESENT-SHD identified those with an up to 4-fold higher risk of developing new-onset SHD/HF, across both health system-centered cohorts in the US and in a community-based cohort in the UK. The model was well calibrated to the risk of new-onset disease, such that higher PRESENT-SHD probabilities were associated with progressively higher risk of developing SHD/HF. Thus, an image-based AI-ECG approach is a scalable and accessible strategy for screening for SHDs and identifying those at high risk for developing SHDs.

Prior studies have reported the use of deep learning on 12-lead ECGs to detect individual structural cardiovascular conditions, including LVSD,^10,11,13^ hypertrophic cardiomyopathy,^16,32,33^ cardiac amyloidosis,^34,35^ aortic stenosis,^12^ among others.^15,35–37^ While these models provide a strong foundation for the role of ECG-based detection of SHDs, the low prevalence of these individual diseases, their potential implementation for broad screening is limited by the low PPVs of the proposed models.^10–12,16,32–34^ The simultaneous detection of multiple SHDs increases the composite disease prevalence and improves model precision.^19^ Through a focus on detecting any of the clinically relevant SHDs that require TTE for confirmation, PRESENT-SHD enables efficient screening by limiting false discovery. Moreover, the use of ECG images as the input, and a flexible strategy that allows for varying formats, supports the scalability of the approach across resources settings.^38^

Our work has additional features that build upon the studies reported in the literature. A focus on developing PRESENT-SHD in diverse populations enabled its consistent performance in demographic subgroups across validation sites. Moreover, in addition to the accurate detection of cross-sectional disease, PRESENT-SHD also predicted the risk of new-onset disease in those without baseline SHD, representing a novel strategy for cardiovascular risk stratification. The model was well calibrated to predict the risk of SHD, suggesting that those with high PRESENT-SHD scores can benefit from surveillance, evaluation, and management of risk factors.^7,39–41^

The application of PRESENT-SHD has important implications for cardiovascular screening. Since early disease detection and intervention can alter the trajectory and outcomes of patients with SHDs, an AI-ECG-based approach that leverages ECG images and photographs can enable opportunistic screening through automated deployment across clinical settings where ECGs are obtained.^42,43^ The focus on a composite model that detects a broad range of SHDs simultaneously reduces the burden of false positive screens and downstream testing, which is a major concern for AI-ECG models developed for individual cardiovascular conditions. This high PPV can allow for a sensitive threshold to be selected during implementation to identify those who should be referred for further imaging. Given that the individual components of SHD share a common diagnostic test, a TTE, screening with PRESENT-SHD can help triage the use of TTE testing. Those with a positive AI-ECG screen can be prioritized for cardiac imaging, which is especially helpful in settings where access may be limited.^1,38,44^

Our study has limitations that merit consideration. First, the development population represented a selected set of patients with a clinical indication for an ECG and a TTE. The consistent validation of the model across populations with a broad range of clinical subpopulations seen in community as well as referral hospitals suggests that the model learned generalizable signatures of the SHDs. This is further supported by the consistent validation of PRESENT-SHD in the ELSA-Brasil study, where individuals underwent protocolized ECGs and echocardiograms concurrently at enrollment without any confounding by indication. Nonetheless, continued prospective validation studies are necessary before broad use in a screening population. Second, while PRESENT-SHD consistently performed well on ECG screenshots from EHR and smartphone-captured ECG photographs, it is essential to prospectively evaluate the feasibility and the performance of PRESENT-SHD in real-world settings before broad clinical adoption. Third, while we used age- and sex-matched controls for the development of the CNN model for sLVH detection, we did not evaluate alternative approaches that additionally use clinical risk factors for case-control matching.

Fourth, although the development of the model focused on plotted images, the signal preprocessing before image plotting represented standard steps used in ECG machines before ECG images are generated or printed. Thus, any processing of ECG images is not required for the real-world application of PRESENT-SHD, as also demonstrated in the publicly accessible application of the model. Fifth, model performance was lower in individuals aged 65 and older, potentially limiting reliability as a standalone tool to rule out the need for cardiac imaging. Adjusting model thresholds or developing age-specific models could be evaluated to improve performance. Sixth, we did not evaluate the cost-effectiveness of PRESENT-SHD use in clinical settings. However, the model had a high PPV for cross-sectional disease detection and identified individuals at high risk of developing SHD/HF, representing features favorable for deployment. Finally, in the predictive evaluation of the model, despite broad geographic coverage, some outcome events may have occurred outside the YNHH and the community hospitals, potentially resulting in incomplete capture of longitudinal outcomes. Nonetheless, the model risk stratification was consistent in the UKB, where the ECGs were protocolized and outcomes were ascertained using national EHR linkage.

## CONCLUSION

We developed and validated a novel approach for the detection and the prediction of a range of SHDs using images of 12-lead ECGs, representing a scalable and accessible tool for SHD screening and risk stratification.

## Data Sharing Statement

Data from the UK Biobank and the Brazilian Longitudinal Study of Adult Health are available for research to licensed users. Individual-level data for the Yale New Haven Health System cannot be made available due to HIPAA regulations enforced by the Yale IRB. The model is publicly accessible for research use on our website and programming code for generating key results is available from the authors on request.

## ABBREVIATIONS LIST

AR: aortic regurgitation
AS: aortic stenosis
AUPRC: area under the precision-recall curve
AUROC: area under the receiver operating characteristic curve
CI: confidence intervals
CNN: convolutional neural network
ECG: electrocardiogram
ELSA-Brasil: Brazilian Longitudinal Study of Adult Health
HF: heart failure
HR: hazard ratio
IVSd: interventricular septal diameter at end-diastole
IQR: interquartile range
LVEF: left ventricular ejection fraction
LVSD: left ventricular systolic dysfunction
MR: mitral regurgitation
MS: mitral stenosis
NPV: negative predictive value
PPV: positive predictive value
PRESENT-SHD: Practical scREening using ENsemble machine learning sTrategy for Structural Heart Disease
SHD: structural heart disease
sLVH: severe left ventricular hypertrophy
TRIPOD + AI: Transparent Reporting of a multivariable prediction model for Individual Prognosis Or Diagnosis + Artificial Intelligence
TTE: transthoracic echocardiogram
UKB: UK Biobank
XGBoost: extreme gradient boosting
YNHH: Yale New Haven Hospital

