## Supplementary material for "An Ensemble Deep Learning Algorithm for Structural Heart Disease Screening Using Electrocardiographic Images: PRESENT SHD": Online Supplement

**SUPPLEMENTARY METHODS** ---------- 5

**Data Sources** ---------- 5

**Signal Preprocessing** ---------- 5

**Model Evaluation on Novel ECG Formats** ---------- 5

**Signal Model Development** ---------- 6

**Model Evaluation on Screenshots and Smartphone Photographs of ECGs** ---------- 6

**SUPPLEMENTARY REFERENCES.** ---------- 7

**SUPPLEMENTARY FIGURES** ---------- 8

**Supplementary Figure 1. Flow Diagram of study population and analysis.** ---------- 8

**Supplementary Figure 2. Examples of 12 variations in the electrocardiographic images used for convolutional neural network training.** ---------- 9

**Supplementary Figure 3. Example of 4 electrocardiographic images plotted in the standard layout and used for model evaluation.** ---------- 10

**Supplementary Figure 4. Novel electrocardiogram formats used for model evaluation.** ---------- 11

**Supplementary Figure 5. Example of an electrocardiogram image from a 47-year-old female patient used for PRESENT-SHD evaluation on real-world screenshots and smartphone photographs of electrocardiograms.** ---------- 12

**Supplementary Figure 6. Overview of methodology to identify individuals at risk of new-onset disease in the hospital-based validation sites.** ---------- 13

**Supplementary Figure 7. PRESENT-SHD performance metrics across probability thresholds in the held-out test set.** ---------- 14

**Supplementary Figure 8. PRESENT-SHD performance for detection of structural heart disease, including left ventricular systolic dysfunction, severe left-sided valve diseases, and severe left ventricular hypertrophy across study cohorts.** ---------- 15

**Supplementary Figure 9. PRESENT-SHD model predictions for a random subset of 100 electrocardiograms with real-world images captured using different strategies.** ---------- 16

**Supplementary Figure 10. Receiver operating characteristic curves for detecting individual structural heart disease across study cohorts.** ---------- 17

**SUPPLEMENTARY TABLES** ---------- 18

**Supplementary Table 1. Diagnosis and procedure codes used to identify longitudinal outcomes.** - 18

**Supplementary Table 2. Transparent Reporting of a multivariable prediction model for Individual Prognosis Or Diagnosis + Artificial Intelligence (TRIPOD + AI) checklist.** ---------- 19

**Supplementary Table 3. Demographic and clinical characteristics of the model development population (including training and validation sets).** ---------- 21

**Supplementary Table 4. Demographic and clinical characteristics of the held-out test set and the external validation cohorts.** ---------- **22**

**Supplementary Table 5. Model performance on novel electrocardiogram formats not encountered during training.** ---------- **24**

**Supplementary Table 6. Model discrimination in subsets of the held-out test set where transthoracic echocardiograms were performed before, after, or on the same day as the electrocardiogram.** ---------- **25**

**Supplementary Table 7. Performance for detection of structural heart diseases for convolutional neural network model for structural heart disease in the held-out test set.** ---------- **26**

**Supplementary Table 8. Performance for detection of structural heart diseases for extreme gradient boosting model variations in the held-out test set.** ---------- **27**

**Supplementary Table 9. Model performance characteristics for signal-based ensemble model for detection of structural heart disease across the held-out test set and external validation cohorts.** **28**

**Supplementary Table 10. PRESENT-SHD performance for detecting structural heart disease in a random subset of 100 electrocardiograms with real-world images captured using different strategies.** ---------- **29**

**Supplementary Table 11. Confusion matrices for PRESENT-SHD in the held-out test set and across external validation cohorts.** ---------- **30**

**Supplementary Table 12. Performance metrics for detecting structural heart disease across key demographic subgroups in Bridgeport Hospital.** ---------- **31**

**Supplementary Table 13. Performance metrics for detecting structural heart disease across key demographic subgroups in Greenwich Hospital.** ---------- **32**

**Supplementary Table 14. Performance metrics for detecting structural heart disease across key demographic subgroups in Lawrence + Memorial Hospital.** ---------- **33**

**Supplementary Table 15. Performance metrics for detecting structural heart disease across key demographic subgroups in Westerly Hospital.** ---------- **34**

**Supplementary Table 16. Performance metrics for detecting structural heart disease across key demographic subgroups in Brazilian Longitudinal Study of Adult Health.** ---------- **35**

**Supplementary Table 17. Performance metrics for PRESENT-SHD for detecting structural heart disease in simulated screening cohorts with varying prevalence.** ---------- **36**

**Supplementary Table 18. Performance metrics for convolutional neural network for detecting left ventricular systolic dysfunction in the held-out test set and across external validation cohorts.** -- **37**

**Supplementary Table 19. Performance metrics for convolutional neural network for detecting moderate or severe left-sided valvular disease in the held-out test set and across external validation cohorts.** ---------- **38**

**Supplementary Table 20. Performance metrics for convolutional neural network for detecting moderate or severe aortic regurgitation in the held-out test set and across external validation cohorts.** ---------- **39**

**Supplementary Table 21. Performance metrics for convolutional neural network for detecting moderate or severe aortic stenosis in the held-out test set and across external validation cohorts. -------------------- 40**

**Supplementary Table 22. Performance metrics for convolutional neural network for detecting moderate or severe mitral regurgitation in the held-out test set and across external validation cohorts.-------------------- 41**

**Supplementary Table 23. Performance metrics for convolutional neural network for detecting severe left ventricular hypertrophy in the held-out test set and across external validation cohorts. -------------------- 42**

**Supplementary Table 24. Performance for respective convolutional neural networks for detecting structural heart disease components in a random subset of 100 electrocardiograms with real-world images captured using different strategies.-------------------- 43**

**Supplementary Table 25. Baseline demographic and clinical characteristics of individuals without structural heart disease or heart failure included for the assessment of PRESENT-SHD for prediction of new-onset disease. -------------------- 44**

**Supplementary Table 26. Performance metrics for risk stratification of new-onset structural heart disease or heart failure in individuals at risk in the Yale New Haven Hospital and external validation sites. -------------------- 45**

**Supplementary Table 27. Cumulative hazard across for new-onset structural heart disease or heart failure over median follow-up time across the cohort. -------------------- 46**

**Supplementary Table 28. Age- and sex-adjusted Cox proportional hazard models for the prediction of new-onset structural heart disease or heart failure across model output probabilities in individuals at risk in the Yale New Haven Hospital and external validation sites. -------------------- 47**

### SUPPLEMENTARY METHODS

#### Data Sources

The electronic health records (EHR) data was acquired during patient care at the hospital sites in the Yale New Haven Health System using Epic and was extracted from the Clarity database.[1,2] The YNHHS EHR data are linked to the CT death index to capture out-of-hospital mortality.

The Brazilian Longitudinal Study of Adult Health (ELSA-Brasil) study, a large multicenter prospective cohort study conducted in Brazil, enrolled,105 community-dwelling adults aged 35-74 years at their baseline visit during 2008-2010.[3,4] These participants represent active and retired civil servants from six higher education and research institutions in Brazilian state capitals in three geographical regions of the country: Southeast (Belo Horizonte, Rio de Janeiro, São Paulo and Vitória), South (Porto Alegre) and Northeast (Salvador).[5] The ELSA-Brasil study aimed to investigate the development and progression of chronic diseases and their determinants in the Brazilian adult population. Baseline data were collected using validated instruments, physical examinations, laboratory assessments, and imaging modalities.[3] Additionally, all participants underwent protocolized 12-lead ECG and echocardiogram.[3,4] To ascertain exposure status and to identify changes in baseline, ELSA-Brasil participants present for in-person follow-up visits every three to four years. Moreover, telephone interviews occur annually to obtain information on new diagnoses, hospitalization, and death with adjudicated clinical events based on expert medical record review.[3]

UK Biobank (UKB) is a prospective cohort of 502,468 community-dwelling adults aged 40-69 years recruited during 2006-2010.[3] A group of these participants accepted to participate in the third or fourth UKB study visit during which the participants underwent 12-lead electrocardiograms (ECGs) in 2014-2021. The UKB dataset is linked with the national EHR from the UK National Health Service predating UKB enrollment, enabling access to EHR diagnosis and procedure codes.[7,8] It is also linked to the national death index for complete capture of mortality data. We used data from UKB under research application #71033.

#### Signal Preprocessing

We used a standard preprocessing strategy to extract the signal waveform data from 12-lead ECGs, predominantly acquired using Philips PageWriter and GE MAC machines. We used linear interpolation to resample the ECGs that were obtained at 250Hz to align with a majority that were recorded at a sampling frequency of 500Hz as 10-second ECGs. Median pass filtering was done by subtracting a one-second median filter from the acquired signals to eliminate baseline drift. ECG signals were divided by a factor of 1000 and scaled to millivolts.

#### Model Evaluation on Novel ECG Formats

As a sensitivity analysis, we also evaluated the model on ECG images plotted in 4 novel formats that were not encountered by the model during training, including (a) Black-on-Red Standard: black ECG trace on red background grid plotted in standard clinical format, (b) Blue-on-Black Standard: blue ECG trace on black background grid plotted in standard clinical format, and (c) Black-on-black rhythm-on-top: black ECG trace plotted on black background with a single 10-second rhythm strip (lead I) above the 12 limb and precordial leads, and (d) Blue-on-red rhythm-on-top: blue

ECG trace plotted on red background in the rhythm-on-top layout (**Supplementary Figure 3.5**)."

**Signal Model Development**
For each image-based CNN, a corresponding signal-based CNN model was trained using the same disease labels and in the same training population as the image models. We evaluated multiple CNN architectures, experimenting with the number and size of convolutional layers as well as dropout and learning rates. The architecture with the highest AUROC for LVSD detection in the validation set was selected as the final architecture for training the individual disease detection models.[9,10] This architecture comprised an input layer with dimensions of (5000, 12, 1), representing a 10-second, 500 Hz, 12-lead ECG. The input layer was followed by 7 2-dimensional convolutional layers, progressively increasing the number of filters from 16 to 64 while incorporating varying kernel sizes (7x1, 5x1, and 3x1) to capture different levels of feature abstraction. A batch normalization layer, a ReLU activation layer, and a 2-dimensional max-pooling layer with different pool sizes (2x1 and 4x1) followed each convolutional layer. Next, the output of the 7$^{th}$ convolutional layer was used as the input for a fully connected network that included two dense layers. Each dense layer was followed by a batch normalization layer, a ReLU activation layer, and a dropout layer with a rate of 0.5. Finally, the model output was a dense layer with a single class and a sigmoid activation to generate the output probability of the label. The loss function was adjusted by calculating model weights using the effective number of samples class re-weighting approach to ensure that the learning is not impacted by the differential prevalence of positive and negative labels. The LVSD model was trained first and the weights from the optimal epoch were transferred to initialize the training for the models for sLVH and valve disease labels.

**Model Evaluation on Screenshots and Smartphone Photographs of ECGs**
To evaluate model performance across ECG screenshots, we accessed the YNHH's Epic EHR. Using the 'cardio-server' tab to view ECGs within Epic, we identified the printable PDF versions, and captured their screenshots. These screenshots included only the graphed area with the ECG waveforms, excluding any identifiable patient information. Of note, these ECGs from the EHR featured lead II as the rhythm lead, while the test set images plotted in the standard format used lead I as the rhythm lead.

For capturing smartphone photographs, each screenshot was displayed using the default 'Preview' app on a laptop monitor (MacBook Pro 2021). Photographs were taken using default camera settings on three smartphones: Apple iPhone 15 Pro (30 photographs), Apple iPhone 12 (40 photographs), and Samsung Galaxy S21 5G (30 photographs). These photographs were taken at a distance of 25–40 cm, ensuring the entire ECG image was captured. The same protocol was followed for capturing photographs of printouts where the ECGs were printed on A4 sheets using a color printer. All cropped screenshots and smartphone photographs used for this assessment have been included in **Online Supplement 2.**"

**SUPPLEMENTARY REFERENCES.**

### SUPPLEMENTARY FIGURES

**Supplementary Figure 1. Flow Diagram of study population and analysis.**
Abbreviations: ECG, electrocardiogram; TTE, transthoracic echocardiogram; YNHHS, Yale New Haven Health System.

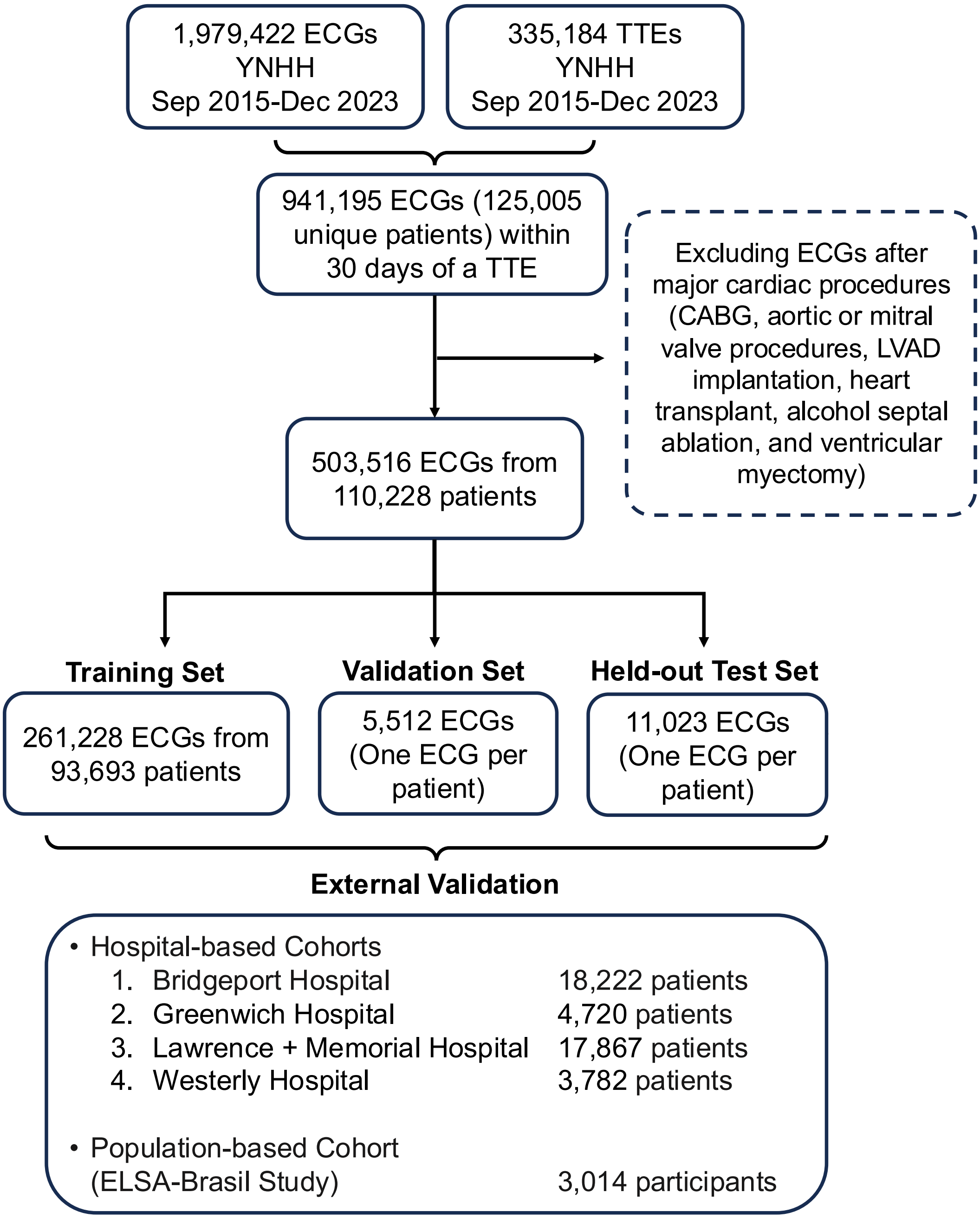

**Supplementary Figure 2. Examples of 12 variations in the electrocardiographic images used for convolutional neural network training.**

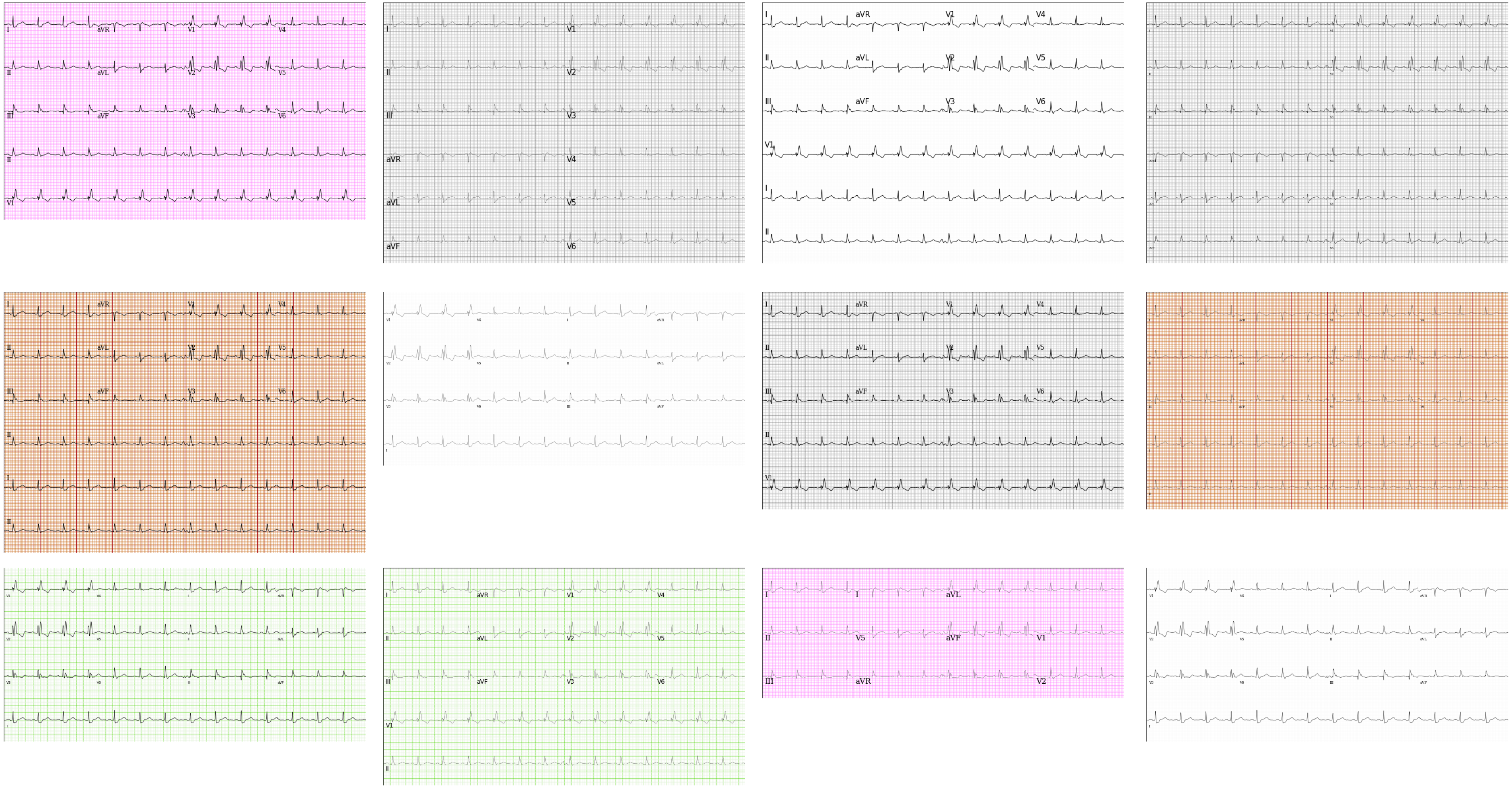

**Supplementary Figure 3. Example of 4 electrocardiographic images plotted in the standard layout and used for model evaluation.**

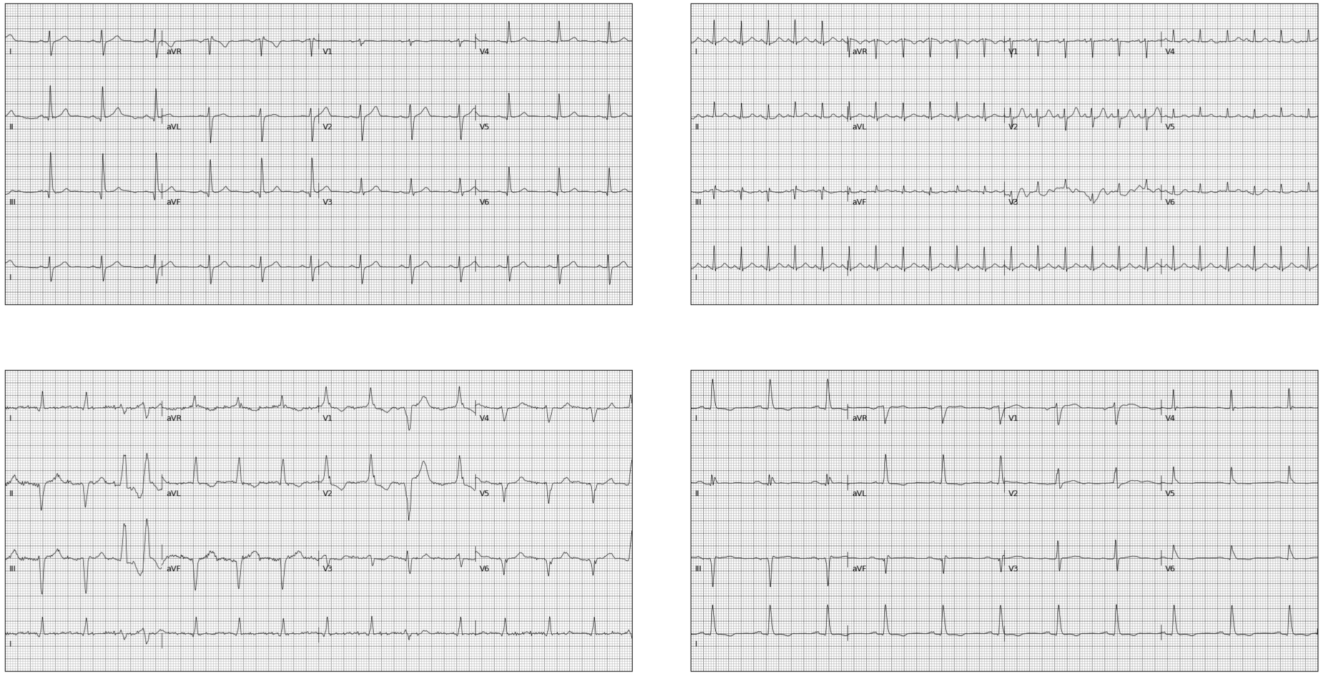

**Supplementary Figure 4. Novel electrocardiogram formats used for model evaluation.**
Abbreviations: ECG, electrocardiogram

**(A) Standard ECG format (presented here for reference)**

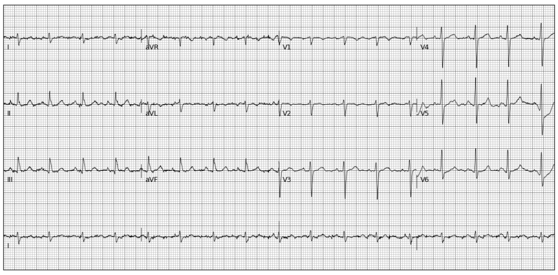

**(B) Black-on-Red colors in standard ECG layout**

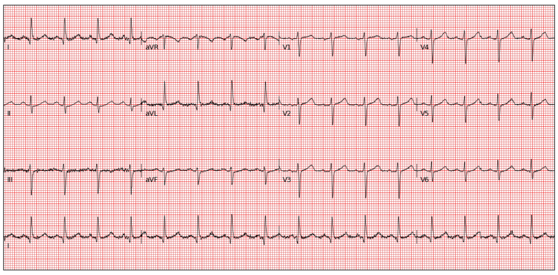

**(C) Blue-on-Black colors in standard ECG layout**

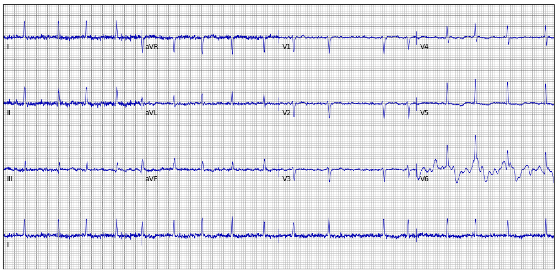

**(D) Black-on-Black colors in rhythm-on-top layout**

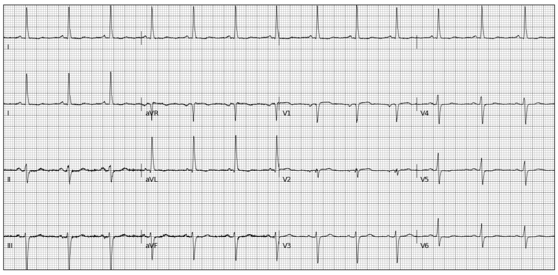

**(E) Blue-on-Red colors in rhythm-on-top layout**

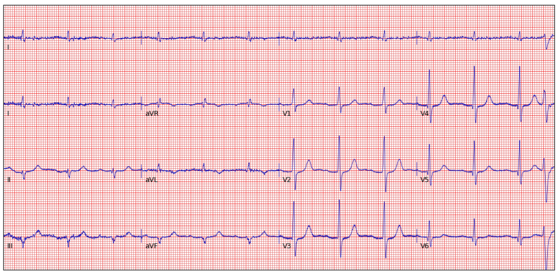

**Supplementary Figure 5. Example of an electrocardiogram image from a 47-year-old female patient used for PRESENT-SHD evaluation on real-world screenshots and smartphone photographs of electrocardiograms.**
Abbreviations: ECG, electrocardiogram; EHR, electronic health record.

**(A) ECG Image Plotted in Standard Format**

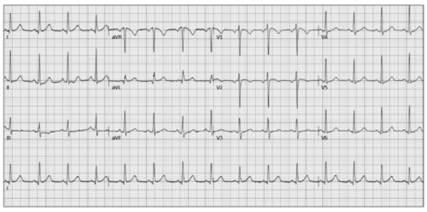

**PRESENT-SHD Probability = 4.1%**

**(B) ECG Image Screenshots from EHR**

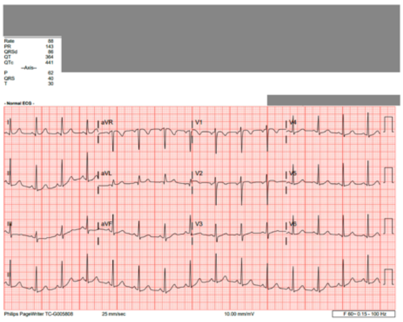

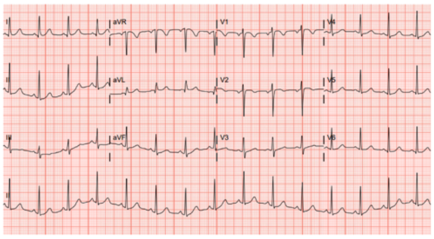

**Cropped Image**
**PRESENT-SHD Probability = 4.6%**

**(C) Smartphone Photos of ECGs on Computer Screen**

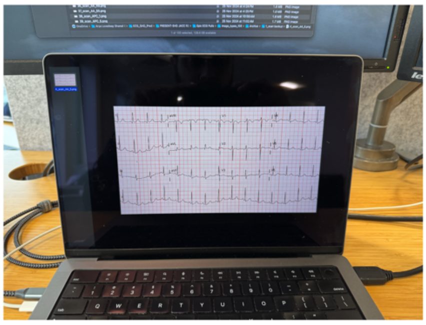

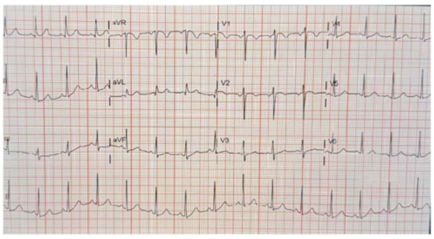

**Cropped Image**
**PRESENT-SHD Probability = 5.0%**

**(D) Smartphone Photos of ECG Printouts**

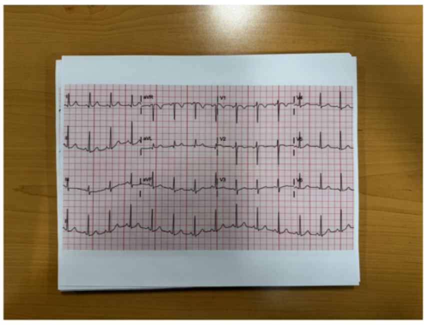

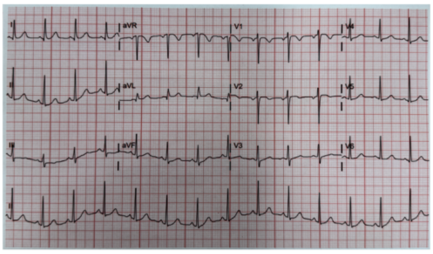

**Cropped Image**
**PRESENT-SHD Probability = 5.2%**

**Supplementary Figure 6. Overview of methodology to identify individuals at risk of new-onset disease in the hospital-based validation sites.**
Abbreviations: ECG, electrocardiograms; HF, heart failure; SHD, structural heart disease; TTE, transthoracic echocardiograms.

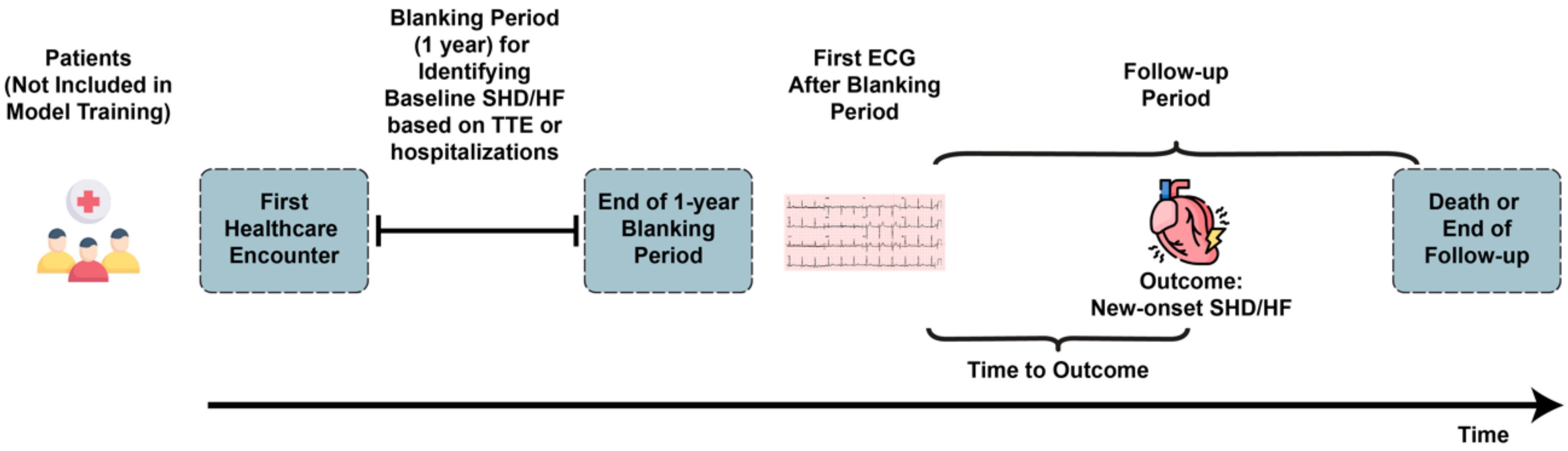

**Supplementary Figure 7. PRESENT-SHD performance metrics across probability thresholds in the held-out test set.**
Abbreviations: NPV, negative predictive value; PPV, positive predictive value.

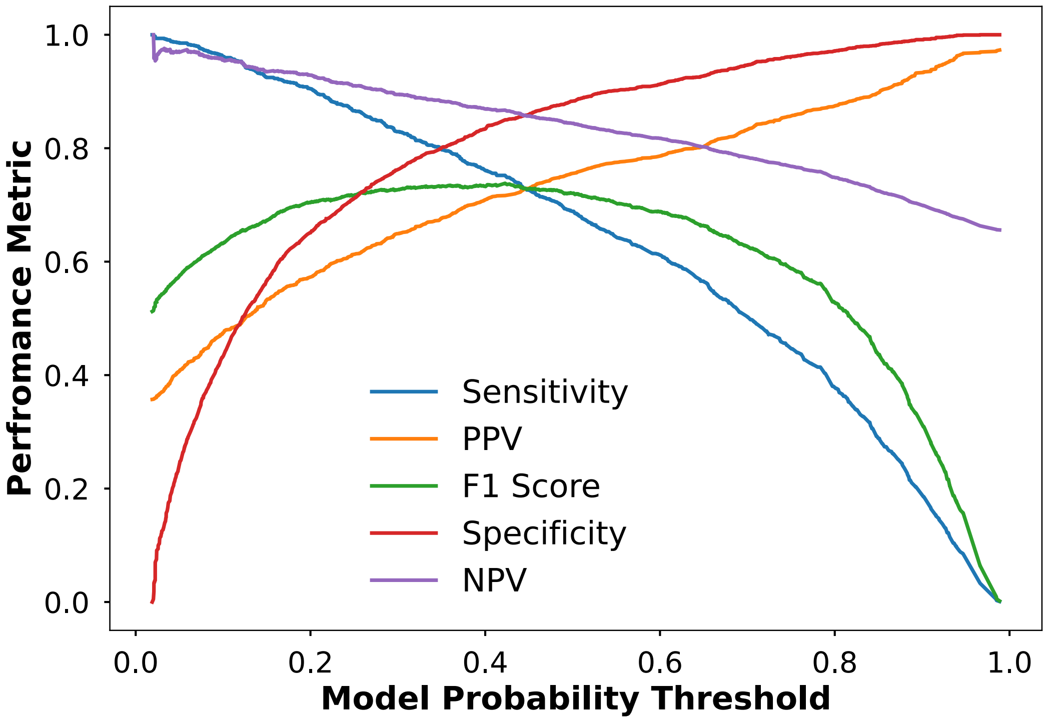

**Supplementary Figure 8. PRESENT-SHD performance for detection of structural heart disease, including left ventricular systolic dysfunction, severe left-sided valve diseases, and severe left ventricular hypertrophy across study cohorts.**

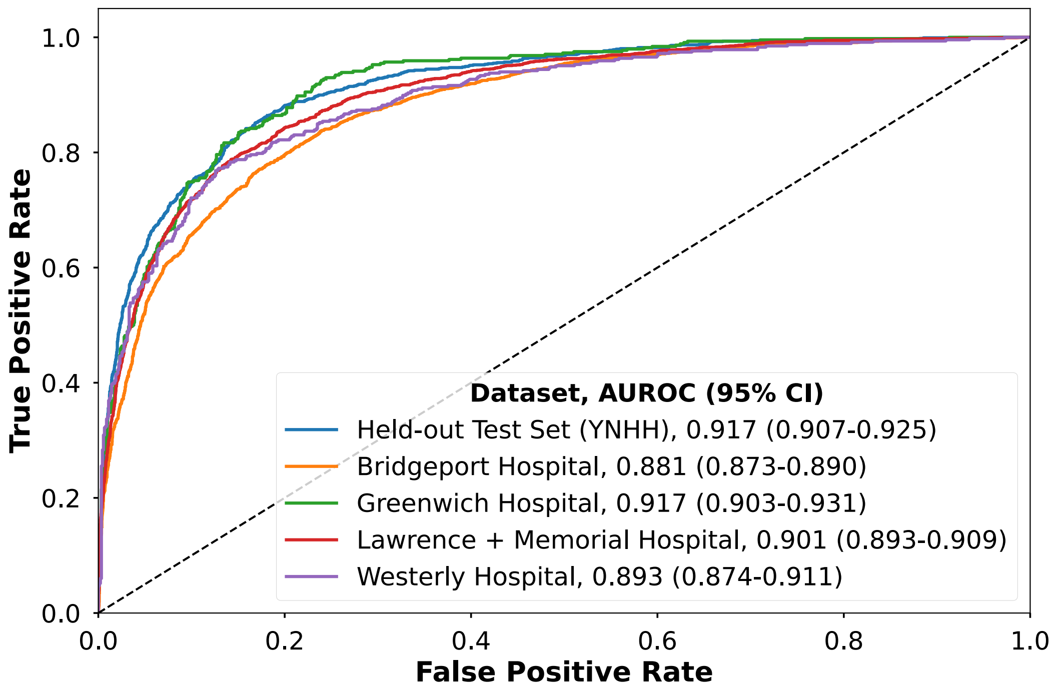

**Supplementary Figure 9. PRESENT-SHD model predictions for a random subset of 100 electrocardiograms with real-world images captured using different strategies.**
Abbreviations: AUPRC, area under the precision-recall curve; AUROC, area under the receiver operating characteristic curve; ECG, electrocardiogram; NPV, negative predictive value; PPV, positive predictive value.

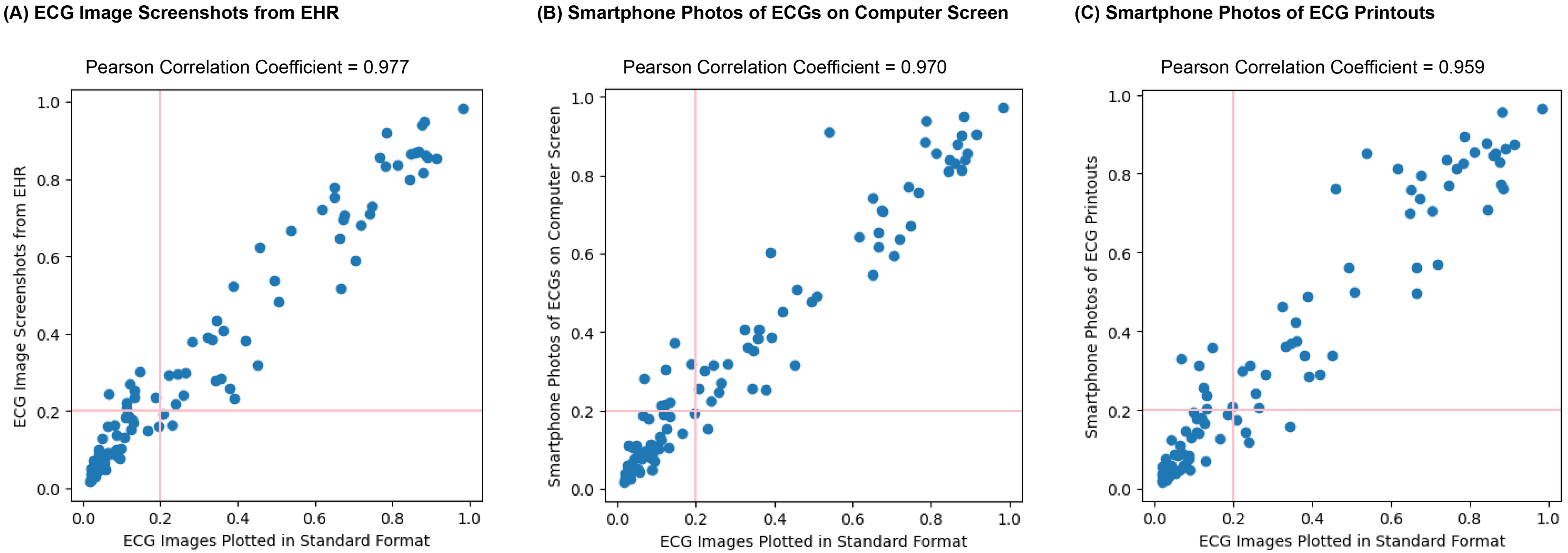

**Supplementary Figure 10. Receiver operating characteristic curves for detecting individual structural heart disease across study cohorts.**
Abbreviations: AR, aortic regurgitation; AS, aortic stenosis; AUROC, area under the receiver operating characteristic curve; sLVH, severe left ventricular hypertrophy; IVSd, interventricular septal diameter at end-diastole; LVDD, left ventricular diastolic dysfunction; LVEF, left ventricular ejection fraction; MR, mitral regurgitation; NPV, negative predictive value; PPV, positive predictive value.

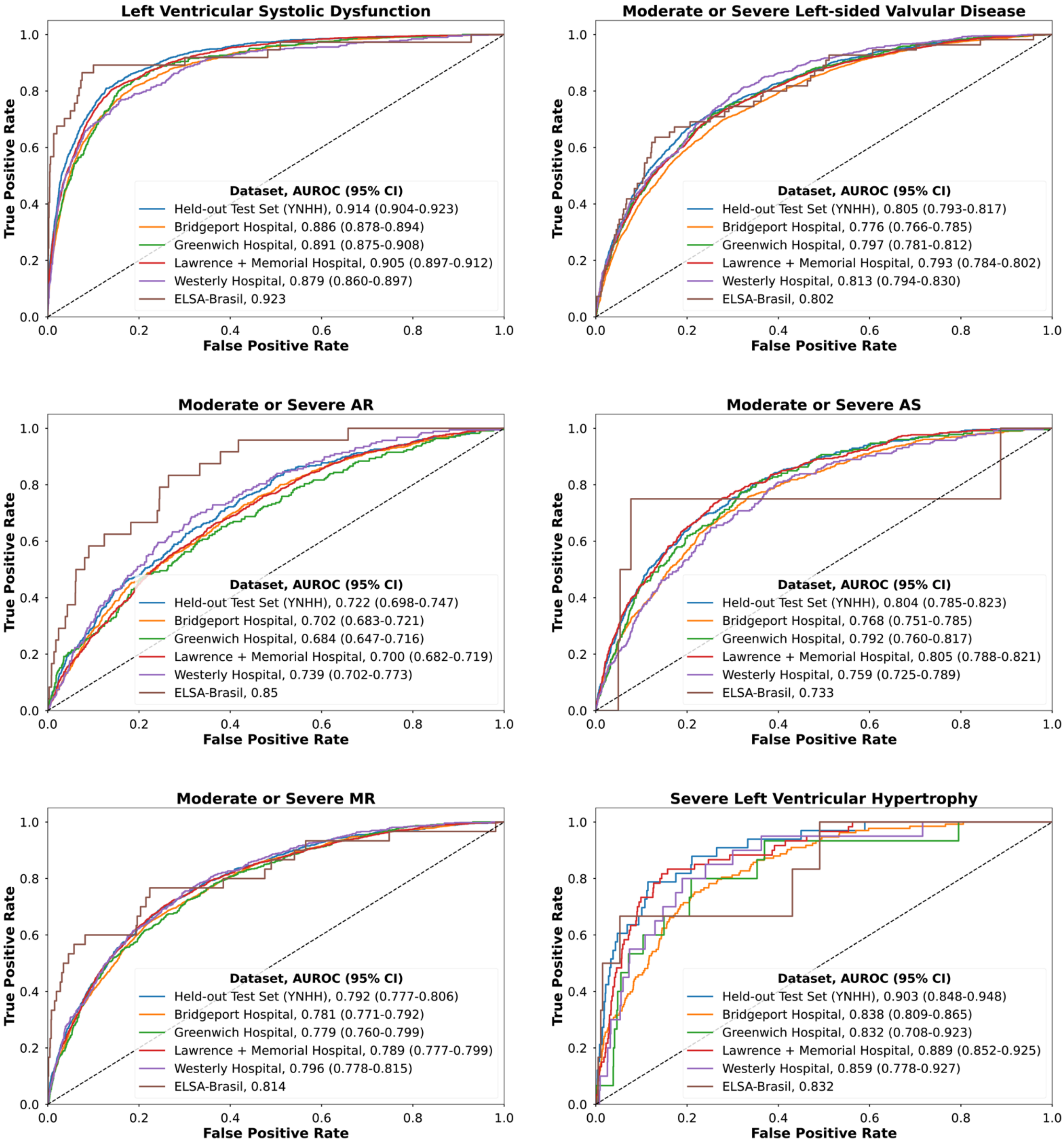

### SUPPLEMENTARY TABLES

#### Supplementary Table 1. Diagnosis and procedure codes used to identify longitudinal outcomes.

| Condition | ICD-10-CM codes |
|---|---|
| **Heart Failure** | ‘I11.0’,’I13.0’,’I13.2’,’I50’,’I50.0’,’I50.1’,’I50.9’,’Z95.81’,’I09.81’ |
| **Acute Myocardial Infarction** | ‘I21’, ‘I22’, ‘I23’, ‘I24.0’, ‘I24.8’, ‘I24.9’ |
| **Stroke** | ‘G45’,’G45.0’,’G45.1’,’G45.2’,’G45.3’,’G45.4’,’G45.8’,’G45.9’, ‘I63’,’I63.0’,’I63.1’,’I63.2’,’I63.3’,’I63.4’,’I63.5’,’I63.8’,’I63.9’,’I64’, ‘I65’,’I65.0’,’I65.1’,’I65.2’,’I65.3’,’I65.8’,’I65.9’,’I66’,’I66.0’,’I66.1’, ‘I66.2’,’I66.3’,’I66.4’,’I66.8’,’I66.9’,’I67.2’,’I69.3’,’I69.4’ |
| **Type 2 Diabetes Mellitus** | ‘E11’,’E11.0’,’E11.1’,’E11.2’,’E11.3’,’E11.4’,’E11.5’,’E11.6’, ‘E11.7’,’E11.8’,’E11.9’,’O24.1’ |
| **Hypertension** | ‘I10’,’I11’,’I11.0’,’I11.9’,’I12’,’I12.0’,’I12.9’, ‘I13’,’I13.0’,’I13.1’,’I13.2’,’I13.9’,’I67.4’, ‘O10’,’O10.0’,’O10.1’,’O10.2’,’O10.3’,’O10.9’,’O11’ |

### Supplementary Table 2. Transparent Reporting of a multivariable prediction model for Individual Prognosis Or Diagnosis + Artificial Intelligence (TRIPOD + AI) checklist.

| Section/Topic | Item | Development / evaluation[1] | Checklist item | Reported on page |
|---|---|---|---|---|
| **TITLE** | | | | |
| *Title* | 1 | D;E | Identify the study as developing or evaluating the performance of a multivariable prediction model, the target population, and the outcome to be predicted | Pg 1 |
| **ABSTRACT** | | | | |
| *Abstract* | 2 | D;E | See TRIPOD+AI for Abstracts checklist | Pg 2 |
| **INTRODUCTION** | | | | |
| *Background* | 3a | D;E | Explain the healthcare context (including whether diagnostic or prognostic) and rationale for developing or evaluating the prediction model, including references to existing models | Pg 6 |
| | 3b | D;E | Describe the target population and the intended purpose of the prediction model in the context of the care pathway, including its intended users (e.g., healthcare professionals, patients, public) | Pg 6 |
| | 3c | D;E | Describe any known health inequalities between sociodemographic groups | Pg 6,7 |
| *Objectives* | 4 | D;E | Specify the study objectives, including whether the study describes the development or validation of a prediction model (or both) | Pg 7 |
| **METHODS** | | | | |
| *Data* | 5a | D;E | Describe the sources of data separately for the development and evaluation datasets (e.g., randomised trial, cohort, routine care or registry data), the rationale for using these data, and representativeness of the data | Pg 7 and suppl. |
| | 5b | D;E | Specify the dates of the collected participant data, including start and end of participant accrual; and, if applicable, end of follow-up | Pg 7 and suppl. |
| *Participants* | 6a | D;E | Specify key elements of the study setting (e.g., primary care, secondary care, general population) including the number and location of centres | Pg 7-8 |
| | 6b | D;E | Describe the eligibility criteria for study participants | Pg 7-8 |
| | 6c | D;E | Give details of any treatments received, and how they were handled during model development or evaluation, if relevant | N/A |
| *Data preparation* | 7 | D;E | Describe any data pre-processing and quality checking, including whether this was similar across relevant sociodemographic groups | Pg 8-9 and suppl. |
| *Outcome* | 8a | D;E | Clearly define the outcome that is being predicted and the time horizon, including how and when assessed, the rationale for choosing this outcome, and whether the method of outcome assessment is consistent across sociodemographic groups | Pg 8-9, 11-12 |
| | 8b | D;E | If outcome assessment requires subjective interpretation, describe the qualifications and demographic characteristics of the outcome assessors | N/A |
| | 8c | D;E | Report any actions to blind assessment of the outcome to be predicted | N/A |
| *Predictors* | 9a | D | Describe the choice of initial predictors (e.g., literature, previous models, all available predictors) and any pre-selection of predictors before model building | Pg 8 |
| | 9b | D;E | Clearly define all predictors, including how and when they were measured (and any actions to blind assessment of predictors for the outcome and other predictors) | N/A |
| | 9c | D;E | If predictor measurement requires subjective interpretation, describe the qualifications and demographic characteristics of the predictor assessors | Pg 8 |
| *Sample size* | 10 | D;E | Explain how the study size was arrived at (separately for development and evaluation), and justify that the study size was sufficient to answer the research question. Include details of any sample size calculation | Pg 6 |
| *Missing data* | 11 | D;E | Describe how missing data were handled. Provide reasons for omitting any data | Pg 7-8 |
| *Analytical methods* | 12a | D | Describe how the data were used (e.g., for development and evaluation of model performance) in the analysis, including whether the data were partitioned, considering any sample size requirements | Pg 11-12 |
| | 12b | D | Depending on the type of model, describe how predictors were handled in the analyses (functional form, rescaling, transformation, or any standardisation). | Pg 9-11 |
| | 12c | D | Specify the type of model, rationale[2], all model-building steps, including any hyperparameter tuning, and method for internal validation | Pg 9-11 |
| | 12d | D;E | Describe if and how any heterogeneity in estimates of model parameter values and model performance was handled and quantified across clusters (e.g., hospitals, countries). See TRIPOD-Cluster for additional considerations[3] | N/A |
| | 12e | D;E | Specify all measures and plots used (and their rationale) to evaluate model performance (e.g., discrimination, calibration, clinical utility) and, if relevant, to compare multiple models | Pg 9-11 |
| | 12f | E | Describe any model updating (e.g., recalibration) arising from the model evaluation, either overall or for particular sociodemographic groups or settings | N/A |
| | 12g | E | For model evaluation, describe how the model predictions were calculated (e.g., formula, code, object, application programming interface) | Pg 9-11 |
| *Class imbalance* | 13 | D;E | If class imbalance methods were used, state why and how this was done, and any subsequent methods to recalibrate the model or the model predictions | N/A |
| *Fairness* | 14 | D;E | Describe any approaches that were used to address model fairness and their rationale | Pg 12 |
| *Model output* | 15 | D | Specify the output of the prediction model (e.g., probabilities, classification). Provide details and rationale for any classification and how the thresholds were identified | Pg 12 |

[1] D=items relevant only to the development of a prediction model; E=items relating solely to the evaluation of a prediction model; D;E=items applicable to both the development and evaluation of a prediction model
[2] Separately for all model building approaches.
[3] TRIPOD-Cluster is a checklist of reporting recommendations for studies developing or validating models that explicitly account for clustering or explore heterogeneity in model performance (eg, at different hospitals or centres). Debray et al, BMJ 2023; 380: e071018 [DOI: 10.1136/bmj-2022-071018]

| | | | | |
|---|---|---|---|---|
| *Training versus evaluation* | 16 | D;E | Identify any differences between the development and evaluation data in healthcare setting, eligibility criteria, outcome, and predictors | N/A |
| *Ethical approval* | 17 | D;E | Name the institutional research board or ethics committee that approved the study and describe the participant-informed consent or the ethics committee waiver of informed consent | Pg 7 |
| **OPEN SCIENCE** | | | | |
| *Funding* | 18a | D;E | Give the source of funding and the role of the funders for the present study | Pg 21 |
| *Conflicts of interest* | 18b | D;E | Declare any conflicts of interest and financial disclosures for all authors | Pg 19-20 |
| *Protocol* | 18c | D;E | Indicate where the study protocol can be accessed or state that a protocol was not prepared | Not prepared |
| *Registration* | 18d | D;E | Provide registration information for the study, including register name and registration number, or state that the study was not registered | N/A |
| *Data sharing* | 18e | D;E | Provide details of the availability of the study data | Pg 21 |
| *Code sharing* | 18f | D;E | Provide details of the availability of the analytical code[4] | Pg 21 |
| **PATIENT & PUBLIC INVOLVEMENT** | | | | |
| *Patient & Public Involvement* | 19 | D;E | Provide details of any patient and public involvement during the design, conduct, reporting, interpretation, or dissemination of the study or state no involvement. | N/A |
| **RESULTS** | | | | |
| *Participants* | 20a | D;E | Describe the flow of participants through the study, including the number of participants with and without the outcome and, if applicable, a summary of the follow-up time. A diagram may be helpful. | Suppl. |
| | 20b | D;E | Report the characteristics overall and, where applicable, for each data source or setting, including the key dates, key predictors (including demographics), treatments received, sample size, number of outcome events, follow-up time, and amount of missing data. A table may be helpful. Report any differences across key demographic groups. | Suppl. |
| | 20c | E | For model evaluation, show a comparison with the development data of the distribution of important predictors (demographics, predictors, and outcome). | Suppl. |
| *Model development* | 21 | D;E | Specify the number of participants and outcome events in each analysis (e.g., for model development, hyperparameter tuning, model evaluation) | Pg 13-14 |
| *Model specification* | 22 | D | Provide details of the full prediction model (e.g., formula, code, object, application programming interface) to allow predictions in new individuals and to enable third-party evaluation and implementation, including any restrictions to access or re-use (e.g., freely available, proprietary)[5] | Pg 13-15 |
| *Model performance* | 23a | D;E | Report model performance estimates with confidence intervals, including for any key subgroups (e.g., sociodemographic). Consider plots to aid presentation. | Pg 14-15 |
| | 23b | D;E | If examined, report results of any heterogeneity in model performance across clusters. See TRIPOD Cluster for additional details[3]. | N/A |
| *Model updating* | 24 | E | Report the results from any model updating, including the updated model and subsequent performance | N/A |
| **DISCUSSION** | | | | |
| *Interpretation* | 25 | D;E | Give an overall interpretation of the main results, including issues of fairness in the context of the objectives and previous studies | Pg 16 |
| *Limitations* | 26 | D;E | Discuss any limitations of the study (such as a non-representative sample, sample size, overfitting, missing data) and their effects on any biases, statistical uncertainty, and generalizability | Pg 18 |
| *Usability of the model in the context of current care* | 27a | D | Describe how poor quality or unavailable input data (e.g., predictor values) should be assessed and handled when implementing the prediction model | Pg 19 |
| | 27b | D | Specify whether users will be required to interact in the handling of the input data or use of the model, and what level of expertise is required of users | N/A |
| | 27c | D;E | Discuss any next steps for future research, with a specific view to applicability and generalizability of the model | Pg 19 |

From: Collins GS, Moons KGM, Dhiman P, et al. *BMJ* 2024;385:e078378. doi:10.1136/bmj-2023-078378

[4] This relates to the analysis code, for example, any data cleaning, feature engineering, model building, evaluation.
[5] This relates to the code to implement the model to get estimates of risk for a new individual.

**Supplementary Table 3. Demographic and clinical characteristics of the model development population (including training and validation sets).**
Abbreviations: AR, aortic regurgitation; AS, aortic stenosis; ECG, electrocardiogram; IQR, interquartile range; LVSD, left ventricular systolic dysfunction; SD, standard deviation; SHD, structural heart disease; sLVH, severe left ventricular hypertrophy; MR, mitral regurgitation

| Characteristic* | Train Set (ECG Level) | Train Set (Patient Level) | Internal Validation Set |
|---|---|---|---|
| **Number** | 261,228 | 93,693 | 5,512 |
| **Age (years; Median [IQR])** | 67.8 [56.1-78.3] | 66.4 [54.1-77.3] | 66.5 [54.1-77.4] |
| **Age (years; Mean [SD])** | 66.4 [16.2] | 64.0 [17.5] | 64.5 [17.6] |
| **Female Sex** | 125735 (48.1%) | 47153 (50.3%) | 2794 (50.7%) |
| **Race and Ethnicity** | | | |
| White | 172972 (66.2%) | 61656 (65.8%) | 3612 (65.5%) |
| Black | 38938 (14.9%) | 12630 (13.4%) | 753 (13.7%) |
| Hispanic | 20941 (8.0%) | 7346 (7.8%) | 408 (7.4%) |
| Others | 28377 (10.9%) | 12061 (12.9%) | 739 (13.4%) |
| **SHD** | 59005 (22.6%) | 17805 (19.0%) | 1091 (19.8%) |
| Indeterminate | 119837 (45.9%) | 40952 (43.7%) | 2344 (42.5%) |
| **LVSD (LVEF <40%)** | 25162 (9.6%) | 6601 (7.0%) | 390 (7.1%) |
| Indeterminate | 4705 (1.8%) | 1424 (1.5%) | 82 (1.5%) |
| **Moderate or Severe Left-sided Valvular Disease** | 42170 (16.1%) | 13397 (14.3%) | 819 (14.9%) |
| Indeterminate | 91537 (35.0%) | 30510 (32.6%) | 1745 (31.7%) |
| **Moderate or Severe AR** | 10271 (3.9%) | 3446 (3.7%) | 214 (3.9%) |
| Indeterminate | 28423 (10.9%) | 8806 (9.4%) | 539 (9.8%) |
| **Moderate or Severe AS** | 10270 (3.9%) | 3389 (3.6%) | 202 (3.7%) |
| Indeterminate | 84492 (32.3%) | 28143 (30.0%) | 1614 (29.3%) |
| **Moderate or Severe MR** | 27347 (10.5%) | 8399 (9.0%) | 503 (9.1%) |
| Indeterminate | 21087 (8.1%) | 6428 (6.9%) | 387 (7.0%) |
| **Severe Left-sided Valvular Disease** | 6193 (2.4%) | 2060 (2.2%) | 128 (2.3%) |
| Indeterminate | 108234 (41.4%) | 35658 (38.1%) | 2064 (37.4%) |
| **Severe AR** | 348 (0.1%) | 109 (0.1%) | 10 (0.2%) |
| Indeterminate | 28423 (10.9%) | 8806 (9.4%) | 539 (9.8%) |
| **Severe AS** | 3123 (1.2%) | 1132 (1.2%) | 70 (1.3%) |
| Indeterminate | 84492 (32.3%) | 28143 (30.0%) | 1614 (29.3%) |
| **Severe MR** | 2672 (1.0%) | 813 (0.9%) | 44 (0.8%) |
| Indeterminate | 21087 (8.1%) | 6428 (6.9%) | 387 (7.0%) |
| **sLVH** | 975 (0.4%) | 276 (0.3%) | 29 (0.5%) |
| Indeterminate | 118906 (45.5%) | 37952 (40.5%) | 2233 (40.5%) |

Footnote: Missing values were considered ‘indeterminate’ for individual SHD components. For composite SHD, the label was considered positive if any of the SHD components was flagged positive, negative if all SHD components were flagged negative, and ‘indeterminate’ otherwise.

**Supplementary Table 4. Demographic and clinical characteristics of the held-out test set and the external validation cohorts.**
Abbreviations: AR, aortic regurgitation; AS, aortic stenosis; ECG, electrocardiogram; IQR, interquartile range; LVSD, left ventricular systolic dysfunction; SHD, structural heart disease; sLVH, severe left ventricular hypertrophy; MR, mitral regurgitation

| | Held-out Test Set | Bridgeport Hospital | Greenwich Hospital | Lawrence + Memorial Hospital | Westerly Hospital | ELSA-Brasil |
|---|---|---|---|---|---|---|
| **Number** | 11,023 | 18,222 | 4,720 | 17,867 | 3,782 | 3,014 |
| **Age (years; Median [IQR])** | 66.3 [53.7-77.4] | 68.5 [56.0-80.0] | 74.0 [59.9-84.5] | 68.7 [57.3-79.5] | 73.3 [62.2-82.4] | 62.0 [57.0-67.0] |
| **Age (years; Mean [SD])** | 64.3 [17.7] | 66.9 [17.1] | 70.8 [17.6] | 67.1 [16.6] | 71.4 [15.1] | 60.7 [8.8] |
| **Female Sex** | 5501 (49.9%) | 9210 (50.5%) | 2316 (49.1%) | 8634 (48.3%) | 1821 (48.1%) | 1596 (53.0%) |
| **Race and Ethnicity** | | | | | | |
| White | 7264 (65.9%) | 10420 (57.2%) | 3110 (65.9%) | 12944 (72.4%) | 3181 (84.1%) | 1661 (55.1%) |
| Black | 1474 (13.4%) | 3472 (19.1%) | 182 (3.8%) | 1184 (6.6%) | 56 (1.5%) | 455 (15.1%) |
| Hispanic | 897 (8.1%) | 2849 (15.6%) | 504 (10.7%) | 1271 (7.1%) | 53 (1.4%) | - |
| Pardo | - | - | - | - | - | 753 (25.0%) |
| Others | 1388 (12.6%) | 1481 (8.1%) | 924 (19.6%) | 2468 (13.8%) | 492 (13.0%) | 145 (4.8%) |
| **SHD** | 2085 (18.9%) | 4167 (22.9%) | 1130 (23.9%) | 3601 (20.2%) | 1024 (27.1%) | 88 (2.9%) |
| Indeterminate | 4820 (43.7%) | 9278 (50.9%) | 2449 (51.9%) | 6420 (35.9%) | 1939 (51.3%) | 26 (0.9%) |
| **LVSD (LVEF <40%)** | 821 (7.4%) | 1772 (9.7%) | 368 (7.8%) | 1466 (8.2%) | 386 (10.2%) | 37 (1.2%) |
| Indeterminate | 163 (1.5%) | 307 (1.7%) | 414 (8.8%) | 137 (0.8%) | 168 (4.4%) | 2 (0.1%) |
| **Moderate or Severe Left-sided Valvular Disease** | 1569 (14.2%) | 3053 (16.8%) | 924 (19.6%) | 2640 (14.8%) | 818 (21.6%) | 55 (1.8%) |
| Indeterminate | 3597 (32.6%) | 6979 (38.3%) | 1243 (26.3%) | 4537 (25.4%) | 1611 (42.6%) | 27 (0.9%) |
| **Moderate or Severe AR** | 392 (3.6%) | 688 (3.8%) | 224 (4.7%) | 694 (3.9%) | 188 (5.0%) | 24 (0.8%) |
| Indeterminate | 997 (9.0%) | 2436 (13.4%) | 472 (10.0%) | 1204 (6.7%) | 277 (7.3%) | 12 (0.4%) |
| **Moderate or Severe AS** | 426 (3.9%) | 714 (3.9%) | 226 (4.8%) | 607 (3.4%) | 236 (6.2%) | 4 (0.1%) |
| Indeterminate | 3312 (30.0%) | 5958 (32.7%) | 1167 (24.7%) | 4343 (24.3%) | 1252 (33.1%) | 9 (0.3%) |
| **Moderate or Severe MR** | 958 (8.7%) | 2104 (11.5%) | 627 (13.3%) | 1670 (9.3%) | 537 (14.2%) | 30 (1.0%) |
| Indeterminate | 759 (6.9%) | 1222 (6.7%) | 310 (6.6%) | 823 (4.6%) | 170 (4.5%) | 15 (0.5%) |
| **Severe Left-sided Valvular Disease** | 279 (2.5%) | 400 (2.2%) | 81 (1.7%) | 281 (1.6%) | 81 (2.1%) | 0 (0%) |
| Indeterminate | 4166 (37.8%) | 8318 (45.6%) | 1560 (33.1%) | 5407 (30.3%) | 2054 (54.3%) | 0 (0%) |
| **Severe AR** | 15 (0.1%) | 27 (0.1%) | 7 (0.1%) | 12 (0.1%) | 1 (0.0%) | 0 (0%) |
| Indeterminate | 997 (9.0%) | 2436 (13.4%) | 472 (10.0%) | 1204 (6.7%) | 277 (7.3%) | 0 (0%) |
| **Severe AS** | 155 (1.4%) | 220 (1.2%) | 47 (1.0%) | 167 (0.9%) | 59 (1.6%) | 0 (0%) |
| Indeterminate | 3312 (30.0%) | 5958 (32.7%) | 1167 (24.7%) | 4343 (24.3%) | 1252 (33.1%) | 0 (0%) |

| | | | | | | |
|---|---|---|---|---|---|---|
| **Severe MR** | 104 (0.9%) | 146 (0.8%) | 28 (0.6%) | 94 (0.5%) | 19 (0.5%) | 0 (0%) |
| Indeterminate | 759 (6.9%) | 1222 (6.7%) | 310 (6.6%) | 823 (4.6%) | 170 (4.5%) | 0 (0%) |
| **sLVH** | 33 (0.3%) | 133 (0.7%) | 15 (0.3%) | 60 (0.3%) | 20 (0.5%) | 6 (0.2%) |
| Indeterminate | 4446 (40.3%) | 8814 (48.4%) | 2979 (63.1%) | 5991 (33.5%) | 2152 (56.9%) | 0 (0%) |

Footnote: Missing values were considered ‘indeterminate’ for individual SHD components. For composite SHD, the label was considered positive if any of the SHD components was flagged positive, negative if all SHD components were flagged negative, and ‘indeterminate’ otherwise.

**Supplementary Table 5. Model performance on novel electrocardiogram formats not encountered during training.**

| Novel Image Format | Key Novel Features | PRESENT-SHD AUROC (95% CI) | PRESENT-SHD AUPRC (95% CI) |
|---|---|---|---|
| **Black-on-Red Standard** | **1. Novel background grid color (red)**<br>**2. Standard ECG trace color**<br>**3. Standard ECG layout** | 0.884 (0.877-0.893) | 0.806 (0.789-0.822) |
| **Blue-on-Black Standard** | **1. Standard background grid color**<br>**2. Novel ECG trace color (blue)**<br>**3. Standard ECG lead layout** | 0.885 (0.877-0.893) | 0.807 (0.788-0.821) |
| **Black-on-Black Rhythm-on-top** | **1. Standard background grid color**<br>**2. Standard ECG trace color**<br>**3. Novel ECG lead layout (rhythm strip on top of the 12 limb and precordial leads)** | 0.883 (0.875-0.892) | 0.802 (0.785-0.818) |
| **Blue-on-Red Rhythm-on-top** | **1. Novel background grid color (red)**<br>**2. Novel ECG trace color (blue)**<br>**3. Novel ECG lead layout (rhythm strip on top of the 12 limb and precordial leads)** | 0.883 (0.874-0.892) | 0.803 (0.785-0.820) |

**Supplementary Table 6. Model discrimination in subsets of the held-out test set where transthoracic echocardiograms were performed before, after, or on the same day as the electrocardiogram.**
Abbreviations: AUPRC, area under the precision-recall curve; AUROC, area under the receiver operating characteristic curve; ECG, electrocardiogram; TTE, transthoracic echocardiogram.

| Subset of interest | Number of patients | AUROC |
|---|---|---|
| **TTE performed within 30 days before the ECG** | 2745 | 0.885 |
| **TTE performed on the same day as the ECG** | 1671 | 0.882 |
| **TTE performed within 30 days after the ECG** | 6607 | 0.885 |

**Supplementary Table 7. Performance for detection of structural heart diseases for convolutional neural network model for structural heart disease in the held-out test set.**

| Performance metric | Convolutional Neural Network for Structural Heart Disease Detection | PRESENT-SHD (Mentioned for reference) |
|---|---|---|
| **AUROC** | 0.856 (0.846-0.866) | 0.886 (0.877-0.894) |
| **AUPRC** | 0.774 (0.758-0.792) | 0.807 (0.791-0.823) |
| **Diagnostic OR** | 10.94 (9.41-12.76) | 17.2 (14.7-20.1) |
| **Sensitivity** | 89.3% (88.0-90.6) | 89.8% (89.0-90.5) |
| **Specificity** | 56.6% (55.1-58.3) | 66.2% (65.0-67.4) |
| **PPV** | 51.0% (49.4-52.4) | 57.4% (56.1-58.6) |
| **NPV** | 91.3% (90.1-92.3) | 92.8% (92.1-93.4) |
| **F1 score** | 0.649 | 0.7 |

**Supplementary Table 8. Performance for detection of structural heart diseases for extreme gradient boosting model variations in the held-out test set.**
Abbreviations: AUROC, area under the receiver operating characteristic curve; CNN, convolutional neural network; SHD, structural heart disease; XGBoost, extreme gradient boosting.

| Model Variation | XGBoost Model Input Features | AUROC (95% CI) |
|---|---|---|
| **Variation 1**<br>**(without age, sex, or CNNs for individual valve diseases)** | **1. LVSD CNN probability**<br>**2. Moderate/Severe Valve Disease CNN Probability**<br>**3. sLVH CNN Probability** | 0.869 (0.859-0.878) |
| **Variation 2**<br>**(without CNNs for individual valve diseases)** | **1. Age**<br>**2. Sex**<br>**3. LVSD CNN probability**<br>**4. Moderate/Severe Valve Disease CNN Probability**<br>**5. sLVH CNN Probability** | 0.885 (0.876-0.893) |

**Supplementary Table 9. Model performance characteristics for signal-based ensemble model for detection of structural heart disease across the held-out test set and external validation cohorts.**
Abbreviations: AUPRC, area under the precision recall curve; AUROC, area under the receiver operating characteristic curve; NPV, negative predictive value; PPV, positive predictive value.

| Cohort Type | Site Name | Total Number | Diagnostic OR | AUROC | AUPRC | F1 Score | Prevalence | Sensitivity | Specificity | PPV | NPV |
|---|---|---|---|---|---|---|---|---|---|---|---|
| **Held-out test set** | **Yale New Haven Hospital** | 6203 | 19.9 (16.9-23.5) | 0.894 (0.885-0.902) | 0.823 (0.806-0.838) | 0.701 | 33.6% | 91.6% (90.9-92.3) | 64.7% (63.6-65.9) | 56.8% (55.6-58.0) | 93.8% (93.2-94.4) |
| **External validation – Hospital sites** | **Bridgeport Hospital** | 8944 | 16.6 (14.5-19.0) | 0.868 (0.861-0.875) | 0.852 (0.842-0.862) | 0.757 | 46.6% | 93.6% (93.1-94.1) | 53.0% (52.0-54.0) | 63.5% (62.5-64.5) | 90.5% (89.9-91.1) |
| | **Greenwich Hospital** | 2271 | 36.7 (26.3-51.2) | 0.908 (0.897-0.920) | 0.903 (0.887-0.918) | 0.808 | 49.8% | 96.4% (95.6-97.1) | 58.0% (56.0-60.0) | 69.5% (67.6-71.3) | 94.2% (93.2-95.1) |
| | **Lawrence + Memorial Hospital** | 11447 | 19.2 (16.6-22.1) | 0.880 (0.874-0.887) | 0.783 (0.769-0.797) | 0.643 | 31.5% | 94.1% (93.6-94.5) | 54.8% (53.8-55.7) | 48.8% (47.9-49.7) | 95.3% (94.9-95.6) |
| | **Westerly Hospital** | 1843 | 21.6 (15.6-29.8) | 0.895 (0.882-0.910) | 0.916 (0.901-0.928) | 0.813 | 55.6% | 95.4% (94.5-96.4) | 50.9% (48.6-53.2) | 70.8% (68.8-72.9) | 89.9% (88.5-91.2) |
| **External validation – Population-based cohort** | **ELSA-Brasil** | 2988 | 9.4 (5.5-16.1) | 0.854 (0.806-0.895) | 0.335 (0.234-0.442) | 0.136 | 2.9% | 80.7% (79.3-82.1) | 69.3% (67.7-71.0) | 7.4% (6.5-8.3) | 99.2% (98.8-99.5) |

**Supplementary Table 10. PRESENT-SHD performance for detecting structural heart disease in a random subset of 100 electrocardiograms with real-world images captured using different strategies.**
Abbreviations: AUPRC, area under the precision-recall curve; AUROC, area under the receiver operating characteristic curve; ECG, electrocardiogram; NPV, negative predictive value; PPV, positive predictive value.

| ECG Image Variation | Pearson correlation coefficient[a] | AUROC | AUPRC | Diagnostic OR | F1 Score | Sensitivity | Specificity | PPV | NPV |
|---|---|---|---|---|---|---|---|---|---|
| **Images plotted in standard format** | - | 0.939 (0.887-0.976) | 0.854 (0.709-0.948) | - | 0.744 | 100.0% (100.0-100.0) | 71.8% (63.0-80.6) | 59.2% (49.6-68.8) | 100.0% (100.0-100.0) |
| **Screenshots of ECGs from EHR** | 0.977 (0.966-0.985) | 0.934 (0.885-0.973) | 0.819 (0.664-0.943) | - | 0.690 | 100.0% (100.0-100.0) | 63.4% (53.9-72.8) | 52.7% (42.9-62.5) | 100.0% (100.0-100.0) |
| **Smartphone Photographs of ECGs on Computer Monitor** | 0.970 (0.951-0.984) | 0.932 (0.878-0.970) | 0.839 (0.696-0.947) | - | 0.690 | 100.0% (100.0-100.0) | 63.4% (53.9-72.8) | 52.7% (42.9-62.5) | 100.0% (100.0-100.0) |
| **Smartphone Photographs of ECG Printouts** | 0.959 (0.939-0.975) | 0.924 (0.866-0.970) | 0.843 (0.715-0.936) | 54.8 (7.0-427.8) | 0.691 | 96.6% (93.0-100.0) | 66.2% (56.9-75.5) | 53.8% (44.1-63.6) | 97.9% (95.1-100.0) |

[a] Pearson correlation coefficient calculated for model predictions on images plotted in standard format and other image types.

**Supplementary Table 11. Confusion matrices for PRESENT-SHD in the held-out test set and across external validation cohorts.**

| Cohort Type | Site Name | Total Number | True Positive | False Negative | True Negative | False Positive |
|---|---|---|---|---|---|---|
| **Held-out test set** | **Yale New Haven Hospital** | 6203 | 1872 (30.2%) | 213 (3.4%) | 2726 (43.9%) | 1392 (22.4%) |
| **External validation – Hospital sites** | **Bridgeport Hospital** | 8944 | 3882 (43.4%) | 285 (3.2%) | 2485 (27.8%) | 2292 (25.6%) |
| | **Greenwich Hospital** | 2271 | 1085 (47.8%) | 45 (2.0%) | 638 (28.1%) | 503 (22.1%) |
| | **Lawrence + Memorial Hospital** | 11447 | 3331 (29.1%) | 270 (2.4%) | 4426 (38.7%) | 3420 (29.9%) |
| | **Westerly Hospital** | 1843 | 974 (52.8%) | 50 (2.7%) | 414 (22.5%) | 405 (22.0%) |
| **External validation – Population-based cohort** | **ELSA-Brasil** | 2988 | 77 (2.6%) | 11 (0.4%) | 1795 (60.1%) | 1105 (37.0%) |

**Supplementary Table 12. Performance metrics for detecting structural heart disease across key demographic subgroups in Bridgeport Hospital.**

Abbreviations: AUPRC, area under the precision-recall curve; AUROC, area under the receiver operating characteristic curve; NPV, negative predictive value; PPV, positive predictive value.

| Subgroup | Total Number | Diagnostic OR | AUROC | AUPRC | F1 Score | Prevalence | Sensitivity | Specificity | PPV | NPV |
|---|---|---|---|---|---|---|---|---|---|---|
| **Overall** | 8944 | 14.8 (12.9-16.9) | 0.854 (0.847-0.862) | 0.834 (0.823-0.845) | 0.751 | 46.6% | 93.2% (92.6-93.7) | 52.0% (51.0-53.1) | 62.9% (61.9-63.9) | 89.7% (89.1-90.3) |
| **Age ≥ 65 years** | 5139 | 9.4 (7.3-12.2) | 0.798 (0.786-0.810) | 0.853 (0.840-0.866) | 0.777 | 60.3% | 97.7% (97.3-98.1) | 18.3% (17.2-19.3) | 64.5% (63.2-65.8) | 83.8% (82.8-84.8) |
| **Age < 65 years** | 3805 | 13.6 (11.4-16.2) | 0.859 (0.846-0.873) | 0.756 (0.732-0.780) | 0.671 | 28.1% | 80.1% (78.8-81.3) | 77.2% (75.8-78.5) | 57.8% (56.2-59.3) | 90.8% (89.9-91.8) |
| **Female Sex** | 4528 | 11.9 (9.9-14.2) | 0.833 (0.821-0.844) | 0.794 (0.776-0.812) | 0.729 | 44.7% | 91.7% (90.9-92.6) | 51.6% (50.2-53.1) | 60.5% (59.1-62.0) | 88.6% (87.6-89.5) |
| **Male Sex** | 4416 | 18.9 (15.4-23.2) | 0.875 (0.865-0.885) | 0.867 (0.853-0.882) | 0.772 | 48.5% | 94.5% (93.8-95.2) | 52.4% (51.0-53.9) | 65.2% (63.8-66.6) | 91.0% (90.1-91.8) |
| **Non-Hispanic White** | 5010 | 13.8 (11.5-16.7) | 0.839 (0.828-0.850) | 0.846 (0.831-0.860) | 0.776 | 53.3% | 94.7% (94.1-95.3) | 43.5% (42.2-44.9) | 65.7% (64.4-67.0) | 87.8% (86.9-88.7) |
| **Non-Hispanic Black** | 1754 | 11.6 (8.8-15.2) | 0.847 (0.827-0.864) | 0.813 (0.784-0.838) | 0.713 | 41.5% | 90.4% (89.0-91.8) | 55.2% (52.8-57.5) | 58.9% (56.6-61.2) | 89.0% (87.5-90.5) |
| **Hispanic** | 1355 | 15.9 (11.5-21.8) | 0.875 (0.854-0.893) | 0.826 (0.798-0.855) | 0.725 | 38.0% | 89.7% (88.1-91.3) | 64.5% (62.0-67.1) | 60.8% (58.2-63.4) | 91.1% (89.6-92.6) |
| **Others** | 825 | 18.6 (11.6-30.1) | 0.863 (0.835-0.888) | 0.765 (0.711-0.813) | 0.665 | 30.8% | 91.7% (89.9-93.6) | 62.7% (59.4-66.0) | 52.2% (48.8-55.7) | 94.5% (92.9-96.0) |

**Supplementary Table 13. Performance metrics for detecting structural heart disease across key demographic subgroups in Greenwich Hospital.**

Abbreviations: AUPRC, area under the precision-recall curve; AUROC, area under the receiver operating characteristic curve; NPV, negative predictive value; PPV, positive predictive value.

| Subgroup | Total Number | Diagnostic OR | AUROC | AUPRC | F1 Score | Prevalence | Sensitivity | Specificity | PPV | NPV |
|---|---|---|---|---|---|---|---|---|---|---|
| **Overall** | 2271 | 30.6 (22.2-42.1) | 0.900 (0.888-0.913) | 0.894 (0.878-0.910) | 0.798 | 49.8% | 96.0% (95.2-96.8) | 55.9% (53.9-58.0) | 68.3% (66.4-70.2) | 93.4% (92.4-94.4) |
| **Age ≥ 65 years** | 1440 | 11.8 (6.8-20.4) | 0.829 (0.807-0.851) | 0.904 (0.887-0.921) | 0.822 | 67.2% | 98.3% (97.7-99.0) | 16.5% (14.6-18.4) | 70.7% (68.4-73.1) | 83.0% (81.0-84.9) |
| **Age < 65 years** | 831 | 23.6 (15.0-37.0) | 0.907 (0.881-0.934) | 0.790 (0.733-0.839) | 0.659 | 19.5% | 82.1% (79.5-84.7) | 83.7% (81.2-86.2) | 55.0% (51.6-58.3) | 95.1% (93.6-96.5) |
| **Female Sex** | 1139 | 28.5 (17.9-45.4) | 0.889 (0.870-0.907) | 0.873 (0.846-0.898) | 0.776 | 47.4% | 96.1% (95.0-97.2) | 53.6% (50.7-56.5) | 65.1% (62.4-67.9) | 93.9% (92.5-95.3) |
| **Male Sex** | 1132 | 33.2 (21.3-51.7) | 0.911 (0.894-0.927) | 0.912 (0.891-0.931) | 0.82 | 52.1% | 95.9% (94.8-97.1) | 58.5% (55.6-61.4) | 71.6% (68.9-74.2) | 93.0% (91.5-94.5) |
| **Non-Hispanic White** | 1440 | 27.4 (17.9-41.8) | 0.884 (0.866-0.901) | 0.907 (0.888-0.925) | 0.827 | 58.8% | 96.9% (96.0-97.8) | 46.5% (43.9-49.0) | 72.1% (69.7-74.4) | 91.4% (89.9-92.8) |
| **Non-Hispanic Black** | 77 | 12.2 (3.6-41.3) | 0.887 (0.799-0.951) | 0.903 (0.814-0.964) | 0.773 | 49.4% | 89.5% (82.6-96.3) | 59.0% (48.0-70.0) | 68.0% (57.6-78.4) | 85.2% (77.3-93.1) |
| **Hispanic** | 235 | 112.4 (26.4-477.8) | 0.920 (0.882-0.949) | 0.887 (0.832-0.936) | 0.816 | 41.3% | 97.9% (96.1-99.8) | 70.3% (64.4-76.1) | 69.9% (64.0-75.7) | 98.0% (96.2-99.8) |
| **Others** | 519 | 19.8 (10.8-36.3) | 0.899 (0.869-0.927) | 0.809 (0.749-0.859) | 0.659 | 28.7% | 91.3% (88.8-93.7) | 65.4% (61.3-69.5) | 51.5% (47.2-55.8) | 94.9% (93.0-96.8) |

**Supplementary Table 14. Performance metrics for detecting structural heart disease across key demographic subgroups in Lawrence + Memorial Hospital.**
Abbreviations: AUPRC, area under the precision-recall curve; AUROC, area under the receiver operating characteristic curve; NPV, negative predictive value; PPV, positive predictive value.

| Subgroup | Total Number | Diagnostic OR | AUROC | AUPRC | F1 Score | Prevalence | Sensitivity | Specificity | PPV | NPV |
|---|---|---|---|---|---|---|---|---|---|---|
| **Overall** | 11447 | 16.0 (14.0-18.2) | 0.871 (0.864-0.878) | 0.771 (0.757-0.784) | 0.643 | 31.5% | 92.5% (92.0-93.0) | 56.4% (55.5-57.3) | 49.3% (48.4-50.3) | 94.3% (93.8-94.7) |
| **Age ≥ 65 years** | 6165 | 11.7 (9.1-15.0) | 0.822 (0.812-0.832) | 0.795 (0.779-0.810) | 0.673 | 45.5% | 97.6% (97.2-98.0) | 22.5% (21.4-23.5) | 51.3% (50.0-52.5) | 91.7% (91.0-92.4) |
| **Age < 65 years** | 5282 | 13.1 (11.0-15.7) | 0.861 (0.845-0.875) | 0.653 (0.619-0.686) | 0.537 | 15.0% | 74.5% (73.4-75.7) | 81.8% (80.8-82.8) | 42.0% (40.6-43.3) | 94.8% (94.2-95.4) |
| **Female Sex** | 5634 | 13.5 (11.3-16.1) | 0.858 (0.848-0.868) | 0.737 (0.715-0.758) | 0.63 | 30.5% | 91.1% (90.3-91.8) | 56.9% (55.6-58.2) | 48.1% (46.8-49.4) | 93.6% (92.9-94.2) |
| **Male Sex** | 5813 | 19.1 (15.7-23.3) | 0.884 (0.876-0.893) | 0.800 (0.782-0.817) | 0.657 | 32.4% | 93.8% (93.2-94.4) | 55.9% (54.6-57.2) | 50.5% (49.2-51.8) | 94.9% (94.4-95.5) |
| **Non-Hispanic White** | 8085 | 15.9 (13.5-18.7) | 0.869 (0.861-0.876) | 0.787 (0.772-0.801) | 0.658 | 34.8% | 93.7% (93.2-94.3) | 51.5% (50.4-52.6) | 50.7% (49.7-51.8) | 93.9% (93.4-94.4) |
| **Non-Hispanic Black** | 776 | 14.0 (8.7-22.4) | 0.879 (0.851-0.905) | 0.766 (0.707-0.819) | 0.615 | 27.2% | 89.6% (87.4-91.7) | 61.9% (58.5-65.4) | 46.8% (43.3-50.3) | 94.1% (92.4-95.7) |
| **Hispanic** | 882 | 19.4 (11.8-31.8) | 0.885 (0.856-0.911) | 0.692 (0.616-0.762) | 0.58 | 19.5% | 88.4% (86.3-90.5) | 71.8% (68.9-74.8) | 43.2% (39.9-46.5) | 96.2% (95.0-97.5) |
| **Others** | 1704 | 13.0 (9.5-17.7) | 0.851 (0.830-0.871) | 0.686 (0.640-0.729) | 0.588 | 23.9% | 87.3% (85.7-88.8) | 65.4% (63.2-67.7) | 44.3% (41.9-46.6) | 94.2% (93.1-95.3) |

**Supplementary Table 15. Performance metrics for detecting structural heart disease across key demographic subgroups in Westerly Hospital.**

Abbreviations: AUPRC, area under the precision-recall curve; AUROC, area under the receiver operating characteristic curve; NPV, negative predictive value; PPV, positive predictive value.

| Subgroup | Total Number | Diagnostic OR | AUROC | AUPRC | F1 Score | Prevalence | Sensitivity | Specificity | PPV | NPV |
|---|---|---|---|---|---|---|---|---|---|---|
| **Overall** | 1843 | 19.9 (14.5-27.3) | 0.887 (0.874-0.902) | 0.906 (0.890-0.922) | 0.81 | 55.6% | 95.1% (94.1-96.1) | 50.5% (48.3-52.8) | 70.6% (68.6-72.7) | 89.2% (87.8-90.6) |
| **Age ≥ 65 years** | 1282 | 16.3 (9.2-29.1) | 0.847 (0.827-0.869) | 0.916 (0.899-0.933) | 0.833 | 67.6% | 98.4% (97.7-99.1) | 21.2% (18.9-23.4) | 72.2% (69.8-74.7) | 86.3% (84.4-88.2) |
| **Age < 65 years** | 561 | 14.3 (9.2-22.4) | 0.885 (0.853-0.914) | 0.810 (0.759-0.857) | 0.683 | 28.2% | 77.2% (73.7-80.7) | 80.9% (77.6-84.1) | 61.3% (57.3-65.3) | 90.1% (87.6-92.5) |
| **Female Sex** | 895 | 19.6 (12.4-31.1) | 0.879 (0.856-0.900) | 0.890 (0.863-0.913) | 0.797 | 53.6% | 95.2% (93.8-96.6) | 49.6% (46.4-52.9) | 68.6% (65.6-71.7) | 90.0% (88.0-91.9) |
| **Male Sex** | 948 | 20.3 (13.2-31.3) | 0.895 (0.875-0.914) | 0.918 (0.896-0.938) | 0.822 | 57.4% | 95.0% (93.7-96.4) | 51.5% (48.3-54.7) | 72.5% (69.7-75.4) | 88.5% (86.5-90.5) |
| **Non-Hispanic White** | 1527 | 19.7 (13.9-28.1) | 0.884 (0.867-0.902) | 0.916 (0.900-0.932) | 0.831 | 60.2% | 95.5% (94.5-96.6) | 47.9% (45.4-50.4) | 73.6% (71.3-75.8) | 87.7% (86.0-89.3) |
| **Non-Hispanic Black** | 23 | 14.0 (1.3-147.4) | 0.750 (0.526-0.938) | 0.619 (0.326-0.888) | 0.7 | 34.8% | 87.5% (74.0-101.0) | 66.7% (47.4-85.9) | 58.3% (38.2-78.5) | 90.9% (79.2-102.7) |
| **Hispanic** | 28 | 10.7 (1.0-109.8) | 0.886 (0.741-1.000) | 0.612 (0.316-1.000) | 0.556 | 21.4% | 83.3% (69.5-97.1) | 68.2% (50.9-85.4) | 41.7% (23.4-59.9) | 93.8% (84.8-102.7) |
| **Others** | 265 | 15.1 (6.6-34.5) | 0.890 (0.847-0.928) | 0.843 (0.777-0.900) | 0.664 | 34.0% | 92.2% (89.0-95.4) | 56.0% (50.0-62.0) | 51.9% (45.9-57.9) | 93.3% (90.3-96.3) |

**Supplementary Table 16. Performance metrics for detecting structural heart disease across key demographic subgroups in Brazilian Longitudinal Study of Adult Health.**
Abbreviations: AUPRC, area under the precision-recall curve; AUROC, area under the receiver operating characteristic curve; NPV, negative predictive value; PPV, positive predictive value.

| Subgroup | Total Number | Diagnostic OR | AUROC | AUPRC | F1 Score | Prevalence | Sensitivity | Specificity | PPV | NPV |
|---|---|---|---|---|---|---|---|---|---|---|
| **Overall** | 2988 | 11.4 (6.0-21.5) | 0.853 (0.811-0.897) | 0.354 (0.253-0.460) | 0.121 | 2.9% | 87.5% (86.3-88.7) | 61.9% (60.2-63.6) | 6.5% (5.6-7.4) | 99.4% (99.1-99.7) |
| **Age ≥ 65 years** | 1087 | 7.0 (2.5-19.7) | 0.813 (0.741-0.885) | 0.364 (0.223-0.501) | 0.123 | 4.5% | 91.8% (90.2-93.5) | 38.4% (35.5-41.3) | 6.6% (5.1-8.1) | 99.0% (98.4-99.6) |
| **Age < 65 years** | 1901 | 13.7 (6.0-31.2) | 0.860 (0.781-0.926) | 0.366 (0.225-0.548) | 0.119 | 2.1% | 82.1% (80.3-83.8) | 75.0% (73.0-76.9) | 6.4% (5.3-7.5) | 99.5% (99.2-99.8) |
| **Female Sex** | 1584 | 6.5 (2.8-15.1) | 0.809 (0.725-0.885) | 0.214 (0.097-0.382) | 0.083 | 2.1% | 78.8% (76.8-80.8) | 63.6% (61.3-66.0) | 4.4% (3.4-5.4) | 99.3% (98.9-99.7) |
| **Male Sex** | 1404 | 19.0 (6.8-53.0) | 0.877 (0.819-0.923) | 0.449 (0.315-0.584) | 0.157 | 3.9% | 92.7% (91.4-94.1) | 59.9% (57.3-62.5) | 8.6% (7.1-10.1) | 99.5% (99.1-99.9) |
| **Non-Hispanic White** | 1644 | 29.1 (7.0-121.1) | 0.889 (0.830-0.939) | 0.381 (0.239-0.532) | 0.106 | 2.4% | 94.9% (93.8-95.9) | 61.1% (58.8-63.5) | 5.6% (4.5-6.7) | 99.8% (99.6-100.0) |
| **Non-Hispanic Black** | 451 | 9.8 (2.9-33.4) | 0.876 (0.795-0.948) | 0.527 (0.342-0.723) | 0.192 | 5.5% | 88.0% (85.0-91.0) | 57.3% (52.7-61.8) | 10.8% (7.9-13.6) | 98.8% (97.8-99.8) |
| **Hispanic** | 748 | 5.3 (2.1-13.7) | 0.770 (0.651-0.863) | 0.220 (0.088-0.408) | 0.116 | 3.1% | 73.9% (70.8-77.1) | 65.2% (61.8-68.7) | 6.3% (4.6-8.1) | 98.7% (98.0-99.5) |
| **Others** | 145 | N/A | 0.917 | 0.077 | 0.041 | 0.7% | 100.0% (100.0-100.0) | 67.4% (59.7-75.0) | 2.1% (-0.2-4.4) | 100.0% (100.0-100.0) |

**Supplementary Table 17. Performance metrics for PRESENT-SHD for detecting structural heart disease in simulated screening cohorts with varying prevalence.**
Abbreviations: AR, aortic regurgitation; AS, aortic stenosis; MR, mitral regurgitation; LVSD, left ventricular systolic dysfunction; sLVH, severe left ventricular hypertrophy; NPV, negative predictive value; PPV, positive predictive value; SHD, structural heart disease; SVD, severe valvular disease.

| Simulated Prevalence | F1 Score | PPV | NPV |
|---|---|---|---|
| **40%** | 0.747 | 63.0% | 90.7% |
| **33.6%[a]** | 0.700 | 57.3% | 92.8% |
| **20%** | 0.553 | 39.9% | 96.3% |
| **10%** | 0.364 | 22.8% | 98.3% |
| **5%** | 0.216 | 12.3% | 99.2% |
| **2.5%** | 0.119 | 6.4% | 99.6% |
| **1%** | 0.051 | 2.6% | 99.8% |

[a] Prevalence in the held-out test set

**Supplementary Table 18. Performance metrics for convolutional neural network for detecting left ventricular systolic dysfunction in the held-out test set and across external validation cohorts.**
Abbreviations: AUPRC, area under the precision-recall curve; AUROC, area under the receiver operating characteristic curve; NPV, negative predictive value; PPV, positive predictive value.

| Cohort Type | Site Name | Total Number | Diagnostic OR | AUROC | AUPRC | F1 Score | Prevalence | Sensitivity | Specificity | PPV | NPV |
|---|---|---|---|---|---|---|---|---|---|---|---|
| **Held-out test set** | **Yale New Haven Hospital** | 10860 | 27.2 (21.7-34.0) | 0.914 (0.904-0.923) | 0.543 (0.507-0.579) | 0.377 | 7.6% | 89.2% (88.6-89.7) | 76.8% (76.0-77.6) | 23.9% (23.1-24.7) | 98.9% (98.7-99.1) |
| **External validation – Hospital sites** | **Bridgeport Hospital** | 17915 | 18.8 (16.1-22.1) | 0.886 (0.878-0.895) | 0.517 (0.494-0.543) | 0.369 | 9.9% | 90.2% (89.7-90.6) | 67.2% (66.5-67.9) | 23.2% (22.6-23.8) | 98.4% (98.2-98.6) |
| | **Greenwich Hospital** | 4306 | 22.2 (15.6-31.7) | 0.891 (0.874-0.907) | 0.508 (0.455-0.556) | 0.354 | 8.5% | 90.5% (89.6-91.4) | 70.0% (68.7-71.4) | 22.0% (20.8-23.2) | 98.7% (98.4-99.1) |
| | **Lawrence + Memorial Hospital** | 17730 | 24.5 (20.5-29.2) | 0.905 (0.897-0.912) | 0.534 (0.509-0.562) | 0.362 | 8.3% | 90.5% (90.0-90.9) | 72.1% (71.4-72.7) | 22.6% (22.0-23.2) | 98.8% (98.7-99.0) |
| | **Westerly Hospital** | 3614 | 16.6 (12.2-22.7) | 0.879 (0.860-0.898) | 0.544 (0.491-0.594) | 0.401 | 10.7% | 87.6% (86.5-88.6) | 70.2% (68.7-71.7) | 26.0% (24.6-27.5) | 97.9% (97.5-98.4) |
| **External validation – Population-based cohort** | **ELSA-Brasil** | 3012 | 75.7 (29.2-196.1) | 0.923 | 0.485 | 0.212 | 1.2% | 86.5% (85.3-87.7) | 92.2% (91.2-93.2) | 12.1% (11.0-13.3) | 99.8% (99.7-100.0) |

**Supplementary Table 19. Performance metrics for convolutional neural network for detecting moderate or severe left-sided valvular disease in the held-out test set and across external validation cohorts.**
Abbreviations: AUPRC, area under the precision-recall curve; AUROC, area under the receiver operating characteristic curve; NPV, negative predictive value; PPV, positive predictive value.

| Cohort Type | Site Name | Total Number | Diagnostic OR | AUROC | AUPRC | F1 Score | Prevalence | Sensitivity | Specificity | PPV | NPV |
|---|---|---|---|---|---|---|---|---|---|---|---|
| **Held-out test set** | **Yale New Haven Hospital** | 7426 | 8.4 (7.0-9.9) | 0.805 (0.794-0.817) | 0.536 (0.510-0.564) | 0.471 | 21.1% | 89.8% (89.1-90.5) | 48.7% (47.5-49.8) | 31.9% (30.9-33.0) | 94.7% (94.2-95.2) |
| **External validation – Hospital sites** | **Bridgeport Hospital** | 11243 | 7.2 (6.3-8.2) | 0.776 (0.766-0.786) | 0.555 (0.536-0.575) | 0.522 | 27.2% | 91.3% (90.7-91.8) | 40.8% (39.9-41.7) | 36.5% (35.6-37.4) | 92.6% (92.1-93.1) |
| | **Greenwich Hospital** | 3477 | 8.3 (6.4-10.9) | 0.797 (0.779-0.813) | 0.579 (0.545-0.615) | 0.512 | 26.6% | 93.1% (92.2-93.9) | 38.2% (36.6-39.8) | 35.3% (33.7-36.9) | 93.8% (93.0-94.6) |
| | **Lawrence + Memorial Hospital** | 13330 | 7.6 (6.6-8.8) | 0.793 (0.784-0.802) | 0.494 (0.475-0.514) | 0.424 | 19.8% | 91.7% (91.3-92.2) | 40.7% (39.9-41.5) | 27.6% (26.9-28.4) | 95.2% (94.9-95.6) |
| | **Westerly Hospital** | 2171 | 13.5 (9.4-19.5) | 0.813 (0.794-0.830) | 0.709 (0.677-0.741) | 0.637 | 37.7% | 96.0% (95.1-96.8) | 36.3% (34.3-38.3) | 47.7% (45.6-49.8) | 93.7% (92.7-94.7) |
| **External validation – Population-based cohort** | **ELSA-Brasil** | 2987 | 8.8 (2.7-28.1) | 0.802 | 0.111 | 0.051 | 1.8% | 94.5% (93.7-95.4) | 33.6% (31.9-35.3) | 2.6% (2.0-3.2) | 99.7% (99.5-99.9) |

**Supplementary Table 20. Performance metrics for convolutional neural network for detecting moderate or severe aortic regurgitation in the held-out test set and across external validation cohorts.**
Abbreviations: AUPRC, area under the precision-recall curve; AUROC, area under the receiver operating characteristic curve; NPV, negative predictive value; PPV, positive predictive value.

| Cohort Type | Site Name | Total Number | Diagnostic OR | AUROC | AUPRC | F1 Score | Prevalence | Sensitivity | Specificity | PPV | NPV |
|---|---|---|---|---|---|---|---|---|---|---|---|
| **Held-out test set** | **Yale New Haven Hospital** | 10026 | 4.8 (3.4-6.8) | 0.722 (0.695-0.749) | 0.109 (0.088-0.135) | 0.097 | 3.9% | 91.3% (90.8-91.9) | 31.3% (30.4-32.2) | 5.1% (4.7-5.6) | 98.9% (98.7-99.1) |
| **External validation – Hospital sites** | **Bridgeport Hospital** | 15786 | 4.3 (3.3-5.5) | 0.702 (0.683-0.722) | 0.097 (0.085-0.112) | 0.107 | 4.4% | 90.3% (89.8-90.7) | 31.5% (30.7-32.2) | 5.7% (5.3-6.0) | 98.6% (98.4-98.8) |
| | **Greenwich Hospital** | 4248 | 3.2 (2.1-5.0) | 0.684 (0.650-0.719) | 0.128 (0.099-0.170) | 0.119 | 5.3% | 89.7% (88.8-90.6) | 27.0% (25.7-28.3) | 6.4% (5.7-7.1) | 97.9% (97.5-98.4) |
| | **Lawrence + Memorial Hospital** | 16663 | 4.6 (3.5-6.0) | 0.700 (0.680-0.719) | 0.091 (0.080-0.104) | 0.1 | 4.2% | 91.8% (91.4-92.2) | 29.0% (28.3-29.7) | 5.3% (5.0-5.7) | 98.8% (98.6-98.9) |
| | **Westerly Hospital** | 3505 | 7.3 (3.6-14.9) | 0.739 (0.706-0.772) | 0.127 (0.104-0.161) | 0.125 | 5.4% | 95.7% (95.1-96.4) | 24.5% (23.1-25.9) | 6.7% (5.9-7.5) | 99.0% (98.7-99.3) |
| **External validation – Population-based cohort** | **ELSA-Brasil** | 3002 | inf (nan-inf) | 0.85 | 0.078 | 0.018 | 0.8% | 100.0% (100.0-100.0) | 14.4% (13.1-15.7) | 0.9% (0.6-1.3) | 100.0% (100.0-100.0) |

**Supplementary Table 21. Performance metrics for convolutional neural network for detecting moderate or severe aortic stenosis in the held-out test set and across external validation cohorts.**
Abbreviations: AUPRC, area under the precision-recall curve; AUROC, area under the receiver operating characteristic curve; NPV, negative predictive value; PPV, positive predictive value.

| Cohort Type | Site Name | Total Number | Diagnostic OR | AUROC | AUPRC | F1 Score | Prevalence | Sensitivity | Specificity | PPV | NPV |
|---|---|---|---|---|---|---|---|---|---|---|---|
| **Held-out test set** | **Yale New Haven Hospital** | 7711 | 8.0 (6.0-10.7) | 0.804 (0.782-0.822) | 0.206 (0.175-0.244) | 0.178 | 5.5% | 87.6% (86.8-88.3) | 53.3% (52.2-54.4) | 9.9% (9.2-10.5) | 98.7% (98.4-98.9) |
| **External validation – Hospital sites** | **Bridgeport Hospital** | 12264 | 5.7 (4.6-7.0) | 0.768 (0.751-0.785) | 0.175 (0.156-0.201) | 0.172 | 5.8% | 84.9% (84.2-85.5) | 50.5% (49.6-51.3) | 9.6% (9.1-10.1) | 98.2% (97.9-98.4) |
| | **Greenwich Hospital** | 3553 | 8.5 (5.4-13.5) | 0.792 (0.763-0.818) | 0.199 (0.163-0.248) | 0.187 | 6.4% | 90.7% (89.8-91.7) | 46.7% (45.0-48.3) | 10.4% (9.4-11.4) | 98.7% (98.3-99.0) |
| | **Lawrence + Memorial Hospital** | 13524 | 7.6 (5.8-9.9) | 0.805 (0.788-0.822) | 0.178 (0.154-0.205) | 0.133 | 4.5% | 90.0% (89.4-90.5) | 45.8% (45.0-46.7) | 7.2% (6.8-7.7) | 99.0% (98.8-99.1) |
| | **Westerly Hospital** | 2530 | 6.0 (3.9-9.3) | 0.759 (0.729-0.789) | 0.243 (0.205-0.302) | 0.232 | 9.3% | 90.3% (89.1-91.4) | 39.3% (37.4-41.2) | 13.3% (11.9-14.6) | 97.5% (96.9-98.1) |
| **External validation – Population-based cohort** | **ELSA-Brasil** | 3005 | 2.5 (0.3-23.9) | 0.733 | 0.008 | 0.004 | 0.1% | 75.0% (73.5-76.5) | 45.3% (43.5-47.1) | 0.2% (0.0-0.3) | 99.9% (99.8-100.0) |

**Supplementary Table 22. Performance metrics for convolutional neural network for detecting moderate or severe mitral regurgitation in the held-out test set and across external validation cohorts.**
Abbreviations: AUPRC, area under the precision-recall curve; AUROC, area under the receiver operating characteristic curve; NPV, negative predictive value; PPV, positive predictive value.

| Cohort Type | Site Name | Total Number | Diagnostic OR | AUROC | AUPRC | F1 Score | Prevalence | Sensitivity | Specificity | PPV | NPV |
|---|---|---|---|---|---|---|---|---|---|---|---|
| **Held-out test set** | **Yale New Haven Hospital** | 10264 | 7.2 (5.9-8.9) | 0.792 (0.778-0.806) | 0.312 (0.285-0.343) | 0.257 | 9.3% | 88.5% (87.9-89.1) | 48.4% (47.5-49.4) | 15.0% (14.3-15.7) | 97.6% (97.3-97.9) |
| **External validation – Hospital sites** | **Bridgeport Hospital** | 17000 | 7.1 (6.1-8.3) | 0.781 (0.771-0.790) | 0.343 (0.323-0.365) | 0.3 | 12.4% | 90.9% (90.4-91.3) | 41.6% (40.9-42.4) | 18.0% (17.5-18.6) | 97.0% (96.7-97.3) |
| | **Greenwich Hospital** | 4410 | 6.9 (5.2-9.2) | 0.779 (0.760-0.797) | 0.369 (0.333-0.409) | 0.327 | 14.2% | 91.5% (90.7-92.4) | 38.9% (37.5-40.4) | 19.9% (18.7-21.1) | 96.5% (96.0-97.1) |
| | **Lawrence + Memorial Hospital** | 17044 | 6.9 (5.9-8.1) | 0.789 (0.778-0.799) | 0.305 (0.282-0.327) | 0.254 | 9.8% | 89.8% (89.4-90.3) | 43.9% (43.1-44.6) | 14.8% (14.3-15.3) | 97.5% (97.3-97.8) |
| | **Westerly Hospital** | 3612 | 10.0 (6.8-14.7) | 0.796 (0.780-0.816) | 0.407 (0.366-0.453) | 0.338 | 14.9% | 94.6% (93.9-95.3) | 36.4% (34.8-38.0) | 20.6% (19.3-21.9) | 97.5% (97.0-98.0) |
| **External validation – Population-based cohort** | **ELSA-Brasil** | 2999 | 5.7 (2.0-16.3) | 0.814 | 0.194 | 0.031 | 1.0% | 86.7% (85.5-87.9) | 46.6% (44.9-48.4) | 1.6% (1.2-2.1) | 99.7% (99.5-99.9) |

**Supplementary Table 23. Performance metrics for convolutional neural network for detecting severe left ventricular hypertrophy in the held-out test set and across external validation cohorts.**
Abbreviations: AUPRC, area under the precision-recall curve; AUROC, area under the receiver operating characteristic curve; NPV, negative predictive value; PPV, positive predictive value.

| Cohort Type | Site Name | Total Number | Diagnostic OR | AUROC | AUPRC | F1 Score | Prevalence | Sensitivity | Specificity | PPV | NPV |
|---|---|---|---|---|---|---|---|---|---|---|---|
| **Held-out test set** | **Yale New Haven Hospital** | 6577 | 20.4 (6.2-66.8) | 0.903 (0.848-0.946) | 0.065 (0.037-0.131) | 0.028 | 0.5% | 90.9% (90.2-91.6) | 67.1% (65.9-68.2) | 1.4% (1.1-1.7) | 99.9% (99.9-100.0) |
| **External validation – Hospital sites** | **Bridgeport Hospital** | 9408 | 11.3 (6.7-19.1) | 0.838 (0.809-0.866) | 0.090 (0.063-0.136) | 0.06 | 1.4% | 88.0% (87.3-88.6) | 60.8% (59.8-61.8) | 3.1% (2.8-3.5) | 99.7% (99.6-99.8) |
| | **Greenwich Hospital** | 1741 | 9.0 (2.5-31.9) | 0.832 (0.702-0.919) | 0.072 (0.018-0.222) | 0.043 | 0.9% | 80.0% (78.1-81.9) | 69.1% (66.9-71.3) | 2.2% (1.5-2.9) | 99.7% (99.5-100.0) |
| | **Lawrence + Memorial Hospital** | 11876 | 12.1 (5.5-26.7) | 0.889 (0.851-0.924) | 0.064 (0.031-0.127) | 0.024 | 0.5% | 88.3% (87.8-88.9) | 61.6% (60.7-62.4) | 1.2% (1.0-1.3) | 99.9% (99.8-100.0) |
| | **Westerly Hospital** | 1630 | 31.5 (4.2-235.7) | 0.859 (0.772-0.926) | 0.067 (0.036-0.137) | 0.058 | 1.2% | 95.0% (93.9-96.1) | 62.4% (60.0-64.7) | 3.0% (2.2-3.9) | 99.9% (99.7-100.1) |
| **External validation – Population-based cohort** | **ELSA-Brasil** | 3014 | 4.0 (0.7-22.0) | 0.832 | 0.035 | 0.008 | 0.2% | 66.7% (65.0-68.3) | 66.8% (65.1-68.5) | 0.4% (0.2-0.6) | 99.9% (99.8-100.0) |

**Supplementary Table 24. Performance for respective convolutional neural networks for detecting structural heart disease components in a random subset of 100 electrocardiograms with real-world images captured using different strategies.**
Abbreviations: AUROC, area under the receiver operating characteristic curve; ECG, electrocardiogram.

| ECG Image Variation | Left Ventricular Systolic Dysfunction | | Moderate/Severe Left Valve Disease | | Moderate/Severe Aortic Regurgitation | | Moderate/Severe Aortic Stenosis | | Moderate/Severe Mitral Regurgitation | | Severe Left Ventricular Hypertrophy | |
|---|---|---|---|---|---|---|---|---|---|---|---|---|
| | Model AUROC | Pearson correlation coefficient[a] | Model AUROC | Pearson correlation coefficient[a] | Model AUROC | Pearson correlation coefficient[a] | Model AUROC | Pearson correlation coefficient[a] | Model AUROC | Pearson correlation coefficient[a] | Model AUROC[b] | Pearson correlation coefficient[a] |
| **Images plotted in standard format** | 0.902 (0.739-1.000) | - | 0.911 (0.845-0.960) | - | 0.757 (0.552-0.928) | - | 0.903 (0.792-0.990) | - | 0.836 (0.725-0.928) | - | - | - |
| **Screenshots of ECGs from EHR** | 0.872 (0.628-1.000) | 0.952 (0.928-0.975) | 0.923 (0.863-0.973) | 0.958 (0.943-0.971) | 0.756 (0.538-0.941) | 0.914 (0.876-0.946) | 0.909 (0.807-0.982) | 0.919 (0.875-0.950) | 0.844 (0.745-0.93) | 0.946 (0.909-0.971) | - | 0.914 (0.851-0.957) |
| **Smartphone Photographs of ECGs on Computer Monitor** | 0.872 (0.671-1.000) | 0.928 (0.879-0.965) | 0.909 (0.842-0.961) | 0.954 (0.932-0.971) | 0.780 (0.574-0.941) | 0.892 (0.847-0.931) | 0.900 (0.791-0.981) | 0.922 (0.875-0.951) | 0.847 (0.748-0.941) | 0.950 (0.923-0.971) | - | 0.847 (0.726-0.926) |
| **Smartphone Photographs of ECG Printouts** | 0.878 (0.682-1.000) | 0.913 (0.857-0.951) | 0.904 (0.831-0.962) | 0.918 (0.878-0.948) | 0.741 (0.564-0.891) | 0.864 (0.798-0.919) | 0.931 (0.815-0.997) | 0.900 (0.838-0.939) | 0.811 (0.667-0.919) | 0.920 (0.866-0.958) | - | 0.876 (0.788-0.929) |

[a] Pearson correlation coefficient calculated for model predictions on images plotted in standard format and other image types.
[b] Model AUROC could not be calculated because there were no severe left ventricular hypertrophy cases in the subset.

**Supplementary Table 25. Baseline demographic and clinical characteristics of individuals without structural heart disease or heart failure included for the assessment of PRESENT-SHD for prediction of new-onset disease.**

| Characteristic | Yale New Haven Hospital | Bridgeport Hospital | Greenwich Hospital | Lawrence + Memorial Hospital | Westerly Hospital | UK Biobank |
|---|---|---|---|---|---|---|
| **Number** | 127,547 | 46,883 | 26,835 | 28,344 | 3,930 | 41,800 |
| **Age (years)** | 53 [37-67] | 53 [38-67] | 59 [45-74] | 59 [43-72] | 63 [51-74] | 65 [59-71] |
| **Sex** | 73031 (57.3%) | 27588 (58.8%) | 15443 (57.5%) | 16314 (57.6%) | 2138 (54.4%) | 21671 (51.8%) |
| **Race/Ethnicity** | | | | | | |
| White | 75450 (60.7%) | 19597 (42.8%) | 19002 (72.8%) | 20609 (74.4%) | 3596 (92.8%) | 40359 (96.8%) |
| Black | 24481 (19.7%) | 11519 (25.1%) | 1356 (5.2%) | 2448 (8.8%) | 71 (1.8%) | 300 (0.7%) |
| Hispanic | 20217 (16.3%) | 13814 (30.1%) | 4848 (18.6%) | 3724 (13.4%) | 127 (3.3%) | - |
| Others | 4062 (3.3%) | 905 (2.0%) | 900 (3.5%) | 908 (3.3%) | 81 (3.1%) | 1028 (3.4%) |
| **Hypertension** | 56313 (44.2%) | 21259 (45.3%) | 10705 (39.9%) | 14437 (50.9%) | 2289 (58.2%) | 5941 (14.2%) |
| **Type-2 Diabetes Mellitus** | 21355 (16.7%) | 9776 (20.9%) | 3830 (14.3%) | 5485 (19.4%) | 855 (21.8%) | 1224 (2.9%) |
| **New-onset SHD/HF Outcome** | 5353 (4.2%) | 3507 (7.5%) | 1493 (5.6%) | 2290 (8.1%) | 298 (7.6%) | 413 (1.0%) |
| **TTE-defined SHD** | 4178 (3.3%) | 2880 (6.1%) | 1021 (3.8%) | 1810 (6.4%) | 221 (5.6%) | - |
| **HF Hospitalization** | 1751 (1.4%) | 1229 (2.6%) | 761 (2.8%) | 876 (3.1%) | 138 (3.5%) | 44 (0.1%) |
| **Aortic Valve Repair/Replacement** | 518 (0.4%) | 172 (0.4%) | 48 (0.2%) | 72 (0.3%) | 10 (0.3%) | 228 (0.5%) |
| **Mitral Valve Repair/Replacement** | 199 (0.2%) | 55 (0.1%) | 20 (0.1%) | 27 (0.1%) | 4 (0.1%) | 264 (0.6%) |
| **Follow-up (years)** | 4.0 [1.7-6.4] | 4.2 [2.4-6.2] | 4.7 [2.7-6.5] | 2.5 [1.1-4.1] | 2.4 [0.8-4.0] | 3.0 [2.1-4.5] |

**Supplementary Table 26. Performance metrics for risk stratification of new-onset structural heart disease or heart failure in individuals at risk in the Yale New Haven Hospital and external validation sites.**

| Model | Covariates | YNHH | Bridgeport Hospital | Greenwich Hospital | Lawrence + Memorial Hospital | Westerly Hospital | UK Biobank |
|---|---|---|---|---|---|---|---|
| **Harrell's C-statistic** | Model Probability | 0.823 (0.817-0.828) | 0.831 (0.819-0.844) | 0.851 (0.841-0.861) | 0.832 (0.824-0.840) | 0.820 (0.796-0.845) | 0.754 (0.728-0.780) |
| **Cox Proportional Hazard Model** | Per 0.1 increase | 1.46 (1.45-1.48) | 1.48 (1.45-1.51) | 1.51 (1.48-1.54) | 1.49 (1.47-1.51) | 1.47 (1.42-1.53) | 1.58 (1.51-1.64) |
| | Per 0.1 increase + Age + Sex | 1.36 (1.35-1.38) | 1.43 (1.39-1.47) | 1.42 (1.38-1.47) | 1.43 (1.4-1.45) | 1.43 (1.36-1.51) | 1.45 (1.38-1.52) |
| | Positive Screen | 8.16 (7.69-8.66) | 8.65 (7.41-10.09) | 14.92 (12.31-18.08) | 9.34 (8.4-10.39) | 9.16 (6.65-12.63) | 4.2 (3.42-5.17) |
| | Positive Screen + Age + Sex | 4.28 (3.95-4.64) | 5.11 (4.18-6.26) | 6.14 (4.8-7.85) | 4.6 (4.01-5.28) | 5.03 (3.39-7.47) | 2.39 (1.87-3.04) |
| | Positive Screen + Age + Sex + HTN + T2DM | 4.04 (3.73-4.37) | 4.73 (3.87-5.78) | 5.55 (4.34-7.1) | 4.21 (3.68-4.83) | 4.72 (3.18-7.01) | 2.34 (1.84-2.99) |
| **Fine-Gray Subdistribution Hazard Model** | Positive Screen + Age + Sex + Competing Risk of Death | 4.24 (3.88-4.64) | 5.07 (4.03-6.37) | 6.25 (4.76-8.21) | 4.56 (3.93-5.29) | 5.09 (3.39-7.63) | 2.62 (2.07-3.32) |
| | Positive Screen + Age + Sex + HTN + T2DM + Competing Risk of Death | 3.99 (3.66-4.36) | 4.69 (3.74-5.88) | 5.64 (4.29-7.41) | 4.18 (3.61-4.84) | 4.77 (3.20-7.13) | 2.56 (2.02-3.25) |

**Supplementary Table 27. Cumulative hazard across for new-onset structural heart disease or heart failure over median follow-up time across the cohort.**

| Cohort | Median Follow-up Time (years) | Cumulative Hazard for New-onset SHD/HF |
|---|---|---|
| **Yale New Haven Hospital** | 4.0 [1.7-6.4] | 0.015 |
| **Bridgeport Hospital** | 4.2 [2.4-6.2] | 0.022 |
| **Greenwich Hospital** | 4.7 [2.7-6.5] | 0.010 |
| **Lawrence + Memorial Hospital** | 2.5 [1.1-4.1] | 0.021 |
| **Westerly Hospital** | 2.4 [0.8-4.0] | 0.018 |
| **UK Biobank** | 3.0 [2.1-4.5] | 0.004 |

**Supplementary Table 28. Age- and sex-adjusted Cox proportional hazard models for the prediction of new-onset structural heart disease or heart failure across model output probabilities in individuals at risk in the Yale New Haven Hospital and external validation sites.**

| Model output probability bins | YNHH | Bridgeport Hospital | Greenwich Hospital | Lawrence + Memorial Hospital | Westerly Hospital | UK Biobank |
|---|---|---|---|---|---|---|
| **0-0.2** | Reference | Reference | Reference | Reference | Reference | Reference |
| **0.2-0.4** | 3.55 (3.25-3.88) | 3.88 (3.09-4.86) | 5.52 (4.22-7.23) | 3.86 (3.31-4.49) | 4.53 (2.92-7.01) | 1.88 (1.43-2.47) |
| **0.4-0.6** | 5.53 (5-6.12) | 6.55 (5.12-8.37) | 9.45 (7.08-12.61) | 6.25 (5.29-7.39) | 7.49 (4.63-12.09) | 3.86 (2.79-5.34) |
| **0.6-0.8** | 7.56 (6.79-8.42) | 9.85 (7.64-12.71) | 16.68 (12.31-22.6) | 10.52 (8.84-12.51) | 11.74 (7.04-19.58) | 7.49 (5.18-10.84) |
| **0.8-1.0** | 12.87 (11.47-14.44) | 18.72 (14.42-24.32) | 25.24 (18.39-34.63) | 20.15 (16.82-24.14) | 27.79 (16.76-46.07) | 13.7 (8.2-22.9) |
