## Supplementary material for "An Ensemble Deep Learning Algorithm for Structural Heart Disease Screening Using Electrocardiographic Images: PRESENT SHD": Online Supplement 2

**Examples of electrocardiogram images used for:**

- **Model development and validation**
- **Model evaluation on previously unseen ECG formats**
- **Model evaluation on real-world ECG screenshots and smartphone images**

### Contents.

**Section 1. Random examples of the same electrocardiogram plotted in 50 different formats used during model development**

**Section 2. Random examples of 25 electrocardiogram images plotted in standard clinical layout used during model evaluation**

**Section 3. Random examples of 25 electrocardiogram images plotted in novel format (Black-on-Red Standard) not encountered during model development**

**Section 4. Random examples of 25 electrocardiogram images plotted in novel format (Blue-on-Black Standard) not encountered during model development**

**Section 5. Random examples of 25 electrocardiogram images plotted in novel format (Black-on-Black Rhythm-on-top) not encountered during model development**

**Section 6. Random examples of 25 electrocardiogram images plotted in novel format (Blue-on-Red Rhythm-on-top) not encountered during model development**

**Section 7. Screenshots of 100 electrocardiograms from the electronic health record used for model evaluation on real-world images (cropped).**

**Section 8. Smartphone photographs of 100 electrocardiograms on computer screen (30 Apple iPhone 15 Pro, 40 Apple iPhone 12, 30 Samsung Galaxy S21 5G; cropped)**

**Section 9. Smartphone photographs of 100 electrocardiogram printouts (30 Apple iPhone 15 Pro, 40 Apple iPhone 12, 30 Samsung Galaxy S21 5G; cropped)**

### Section 1. Random examples of the same electrocardiogram plotted in 50 different formats used during model development

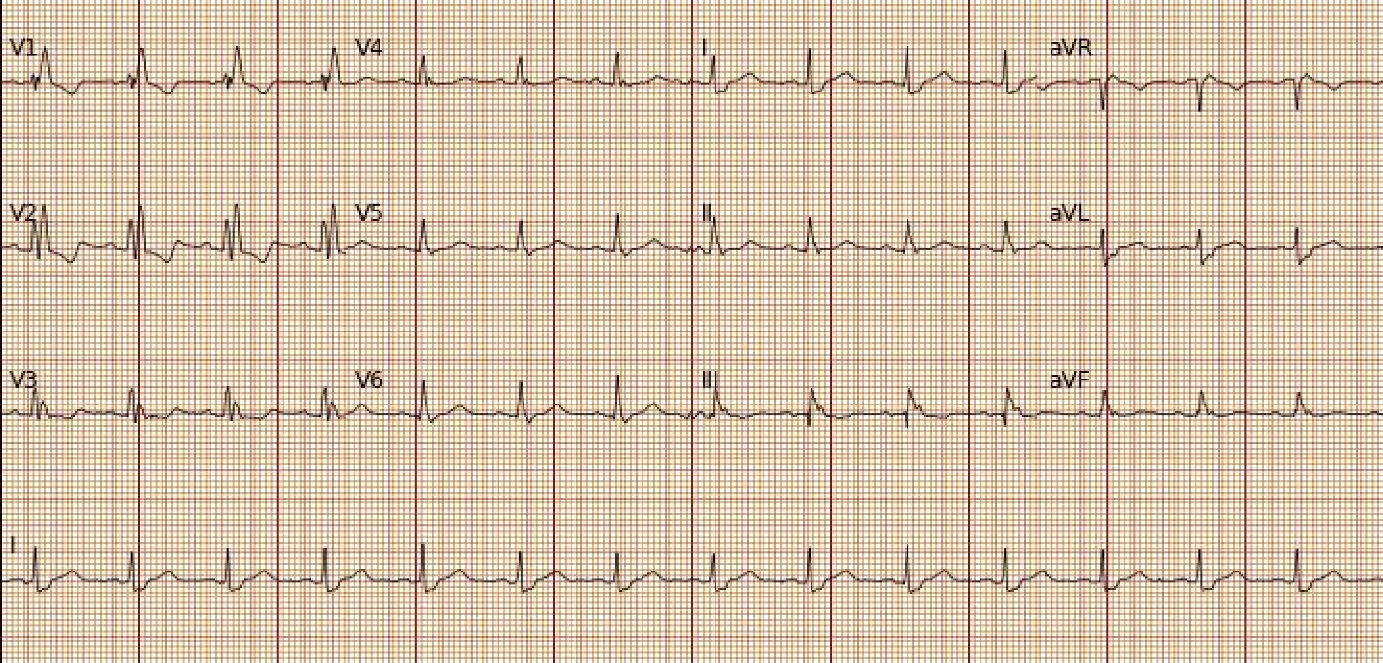
V1
V4
I
aVR
V2
V5
II
aVL
V3
V6
III
aVF
I

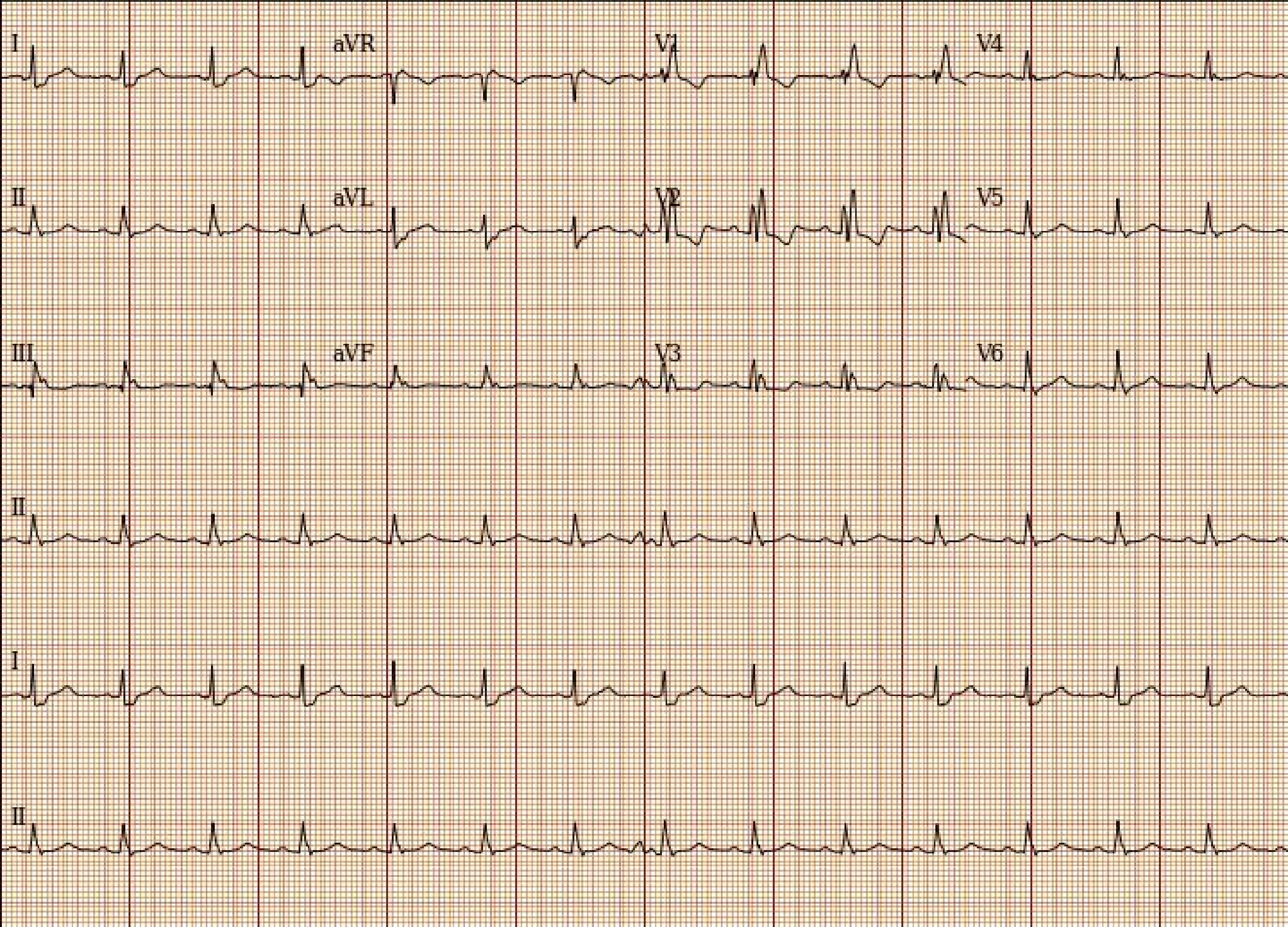
I
aVR
V1
V4
II
aVL
V2
V5
III
aVF
V3
V6
II
I
II

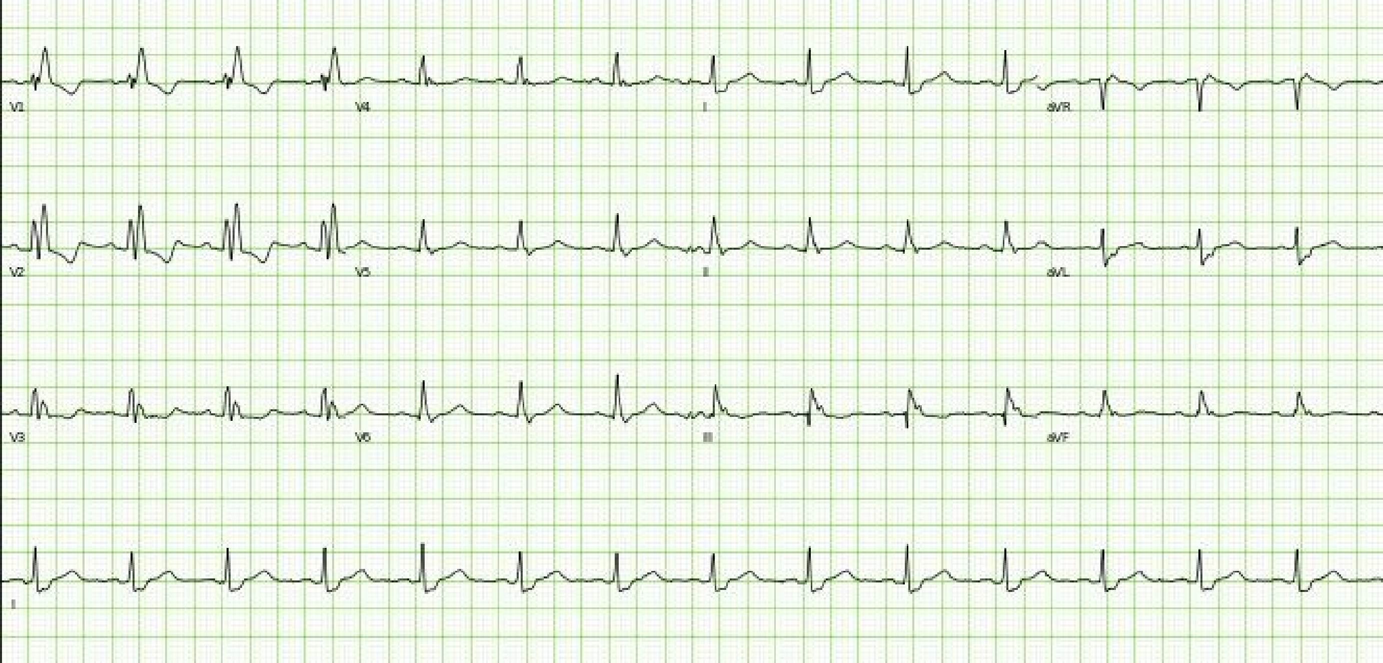
V1
V4
I
aVR
V2
V5
II
aVL
V3
V6
III
aVF
II

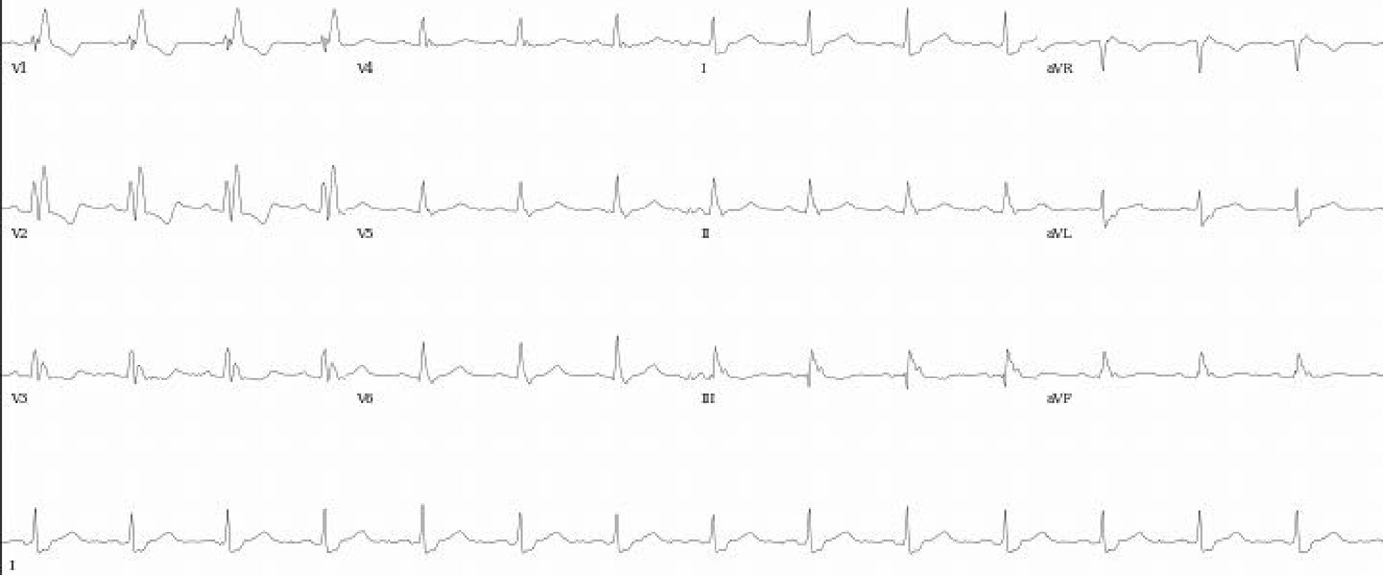
V1
V4
I
aVR
V2
V5
II
aVL
V3
V6
III
aVF
I

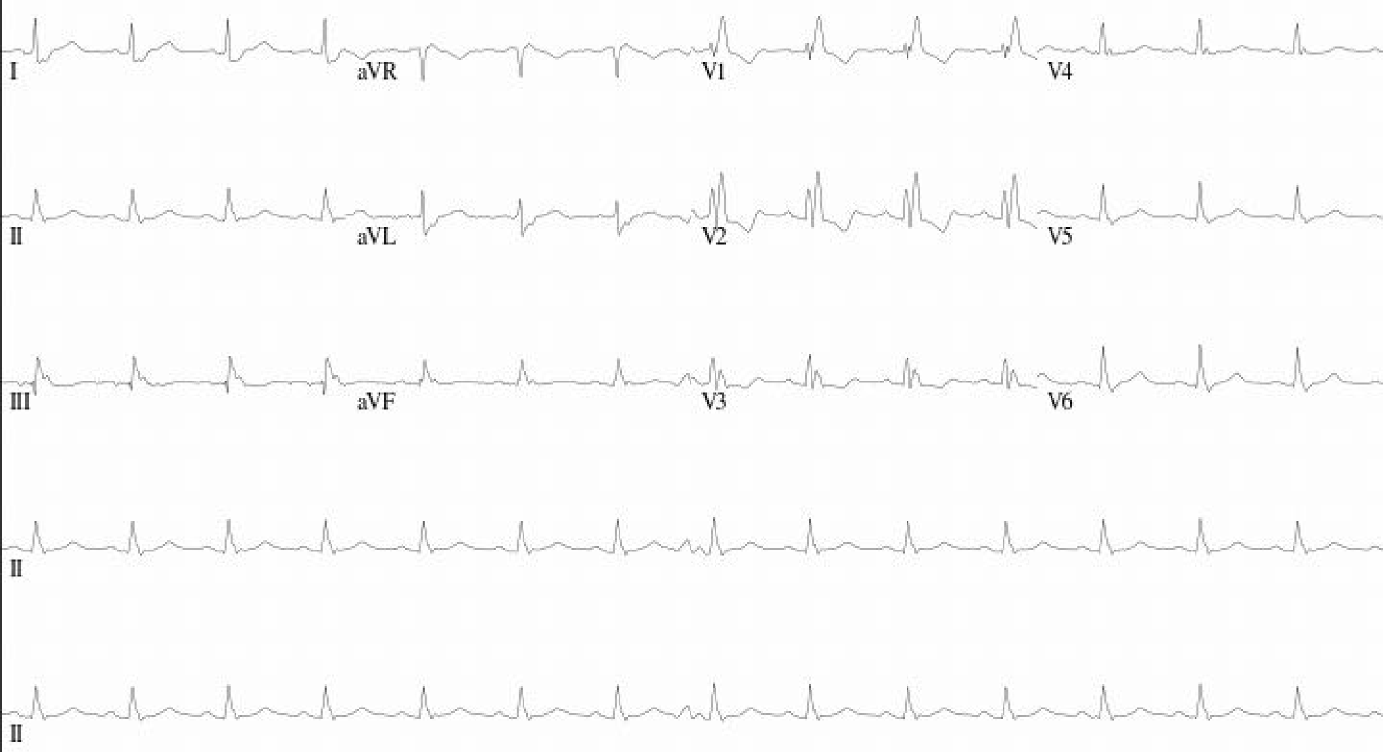
I
aVR
V1
V4
II
aVL
V2
V5
III
aVF
V3
V6
II
II

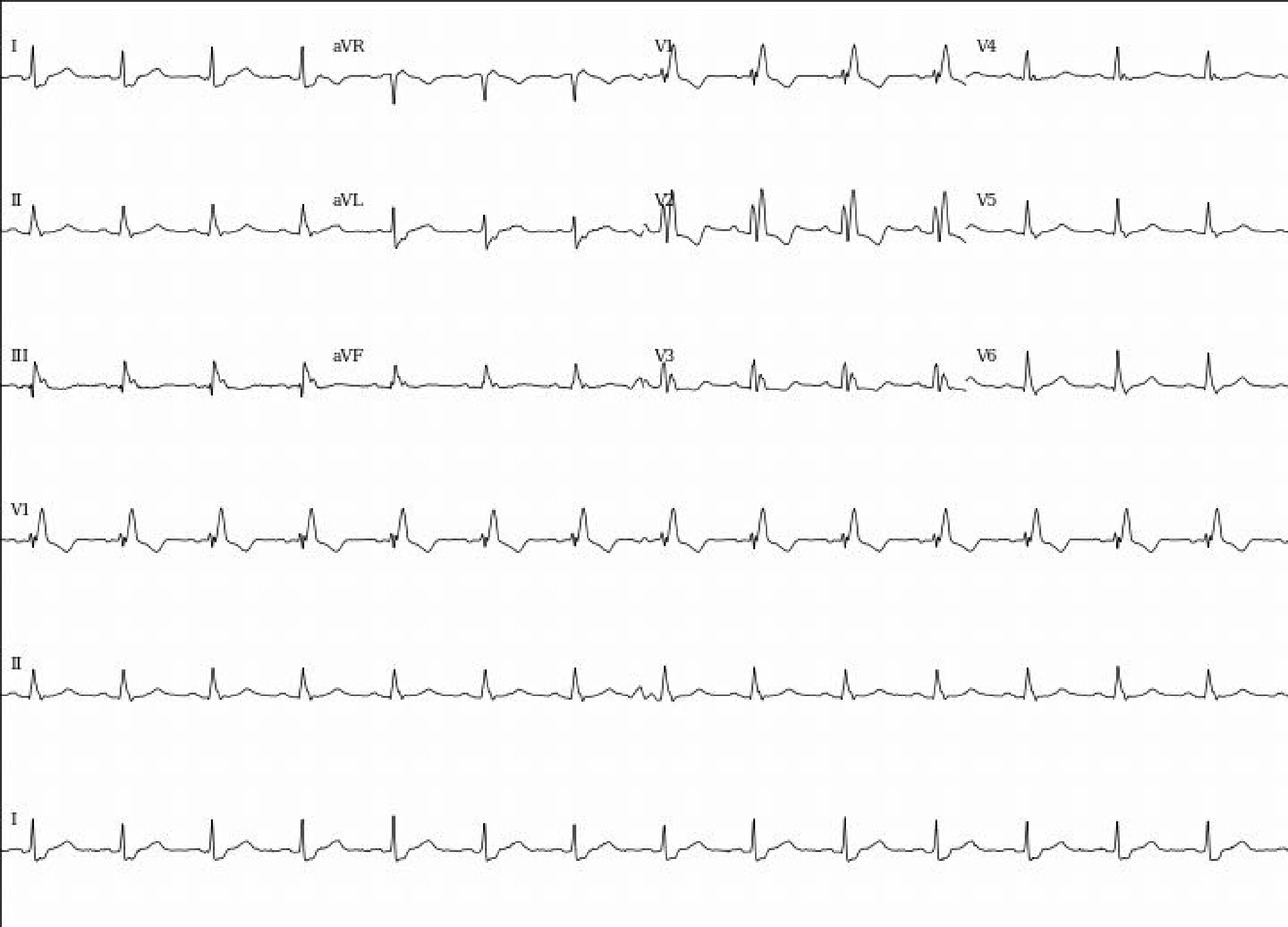
I
aVR
V1
V4
II
aVL
V2
V5
III
aVF
V3
V6
V1
II
I

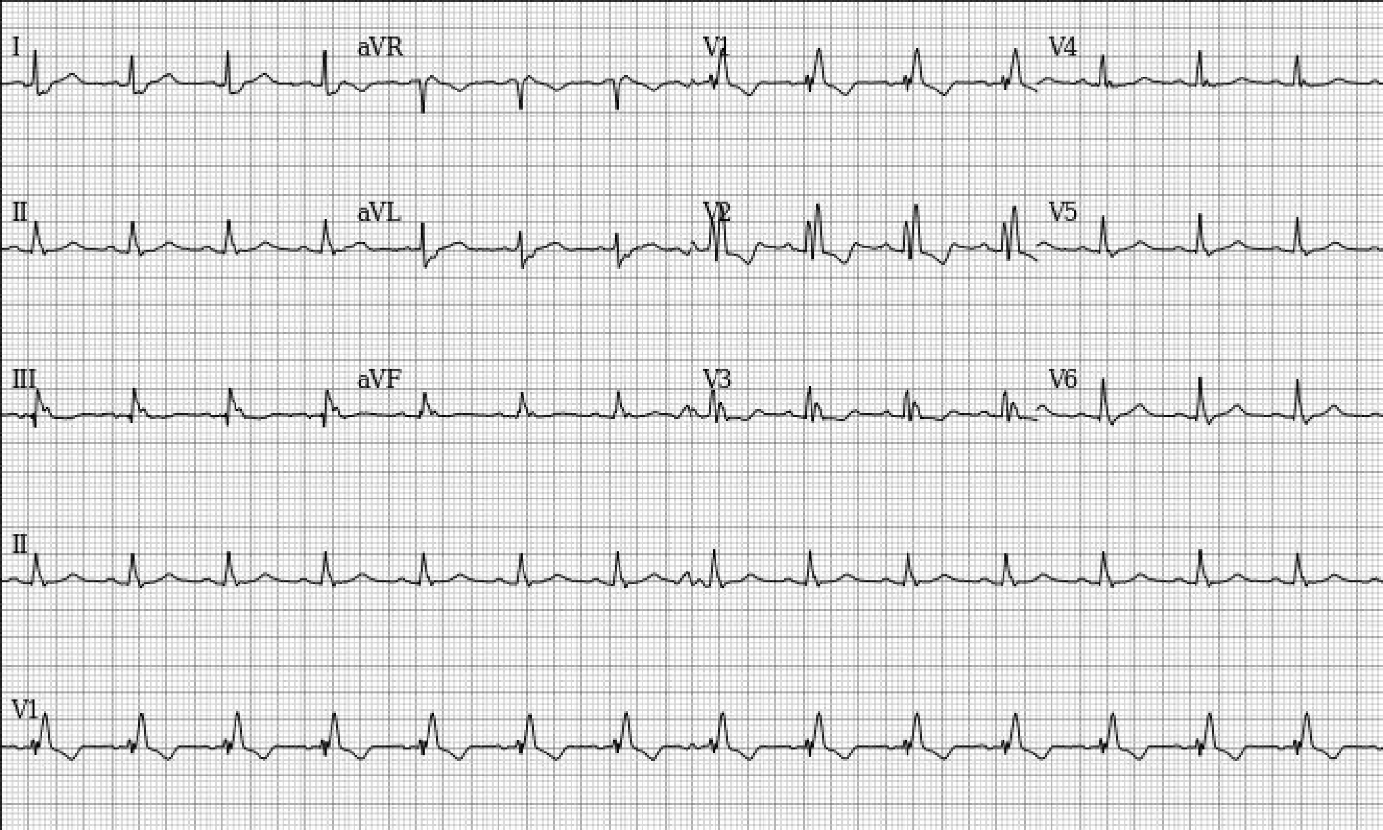
I
aVR
V1
V4
II
aVL
V2
V5
III
aVF
V3
V6
II
V1

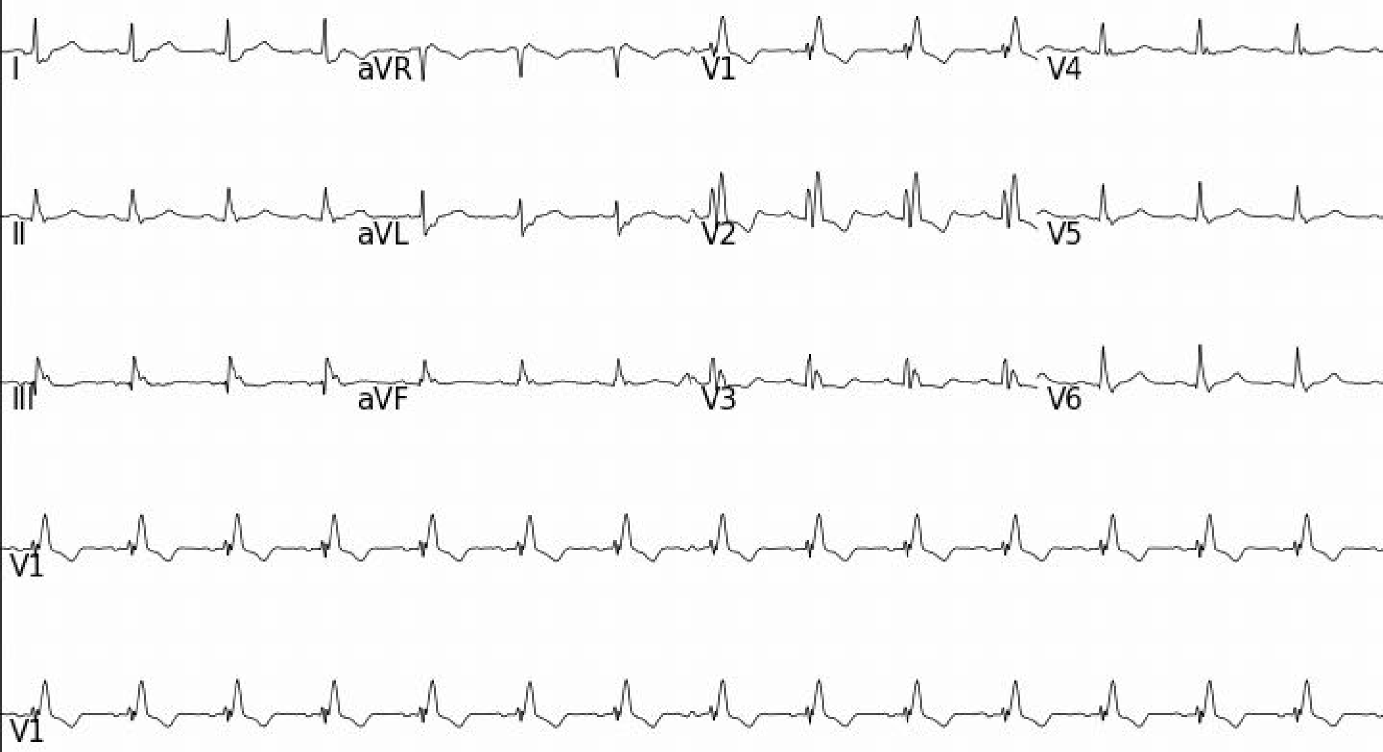
I
aVR
V1
V4
II
aVL
V2
V5
III
aVF
V3
V6
V1
V1

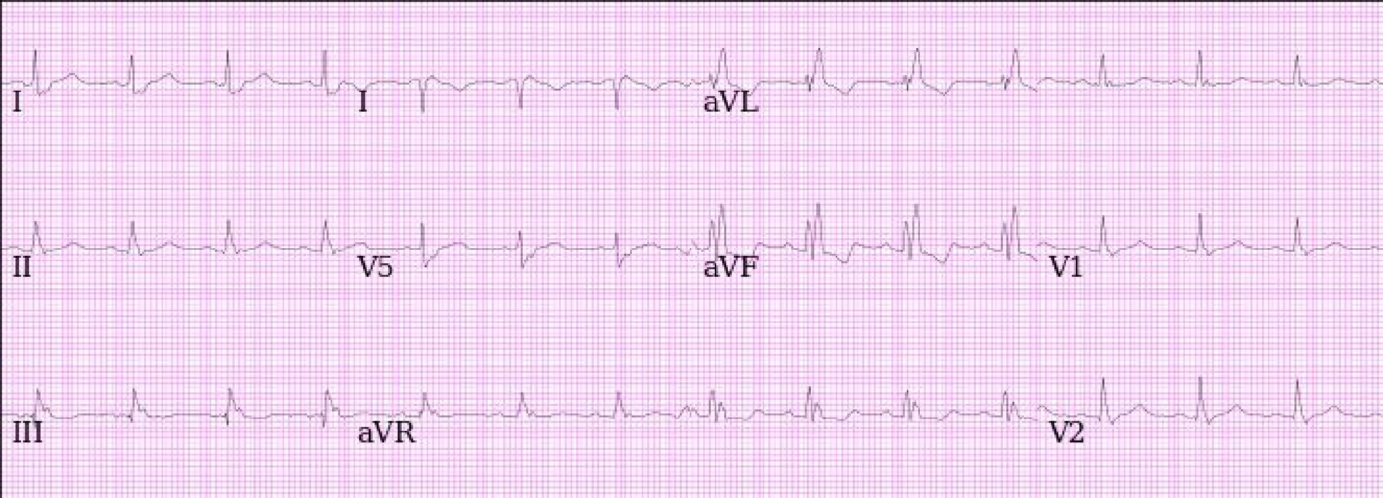
I
I
aVL
II
V5
aVF
V1
III
aVR
V2

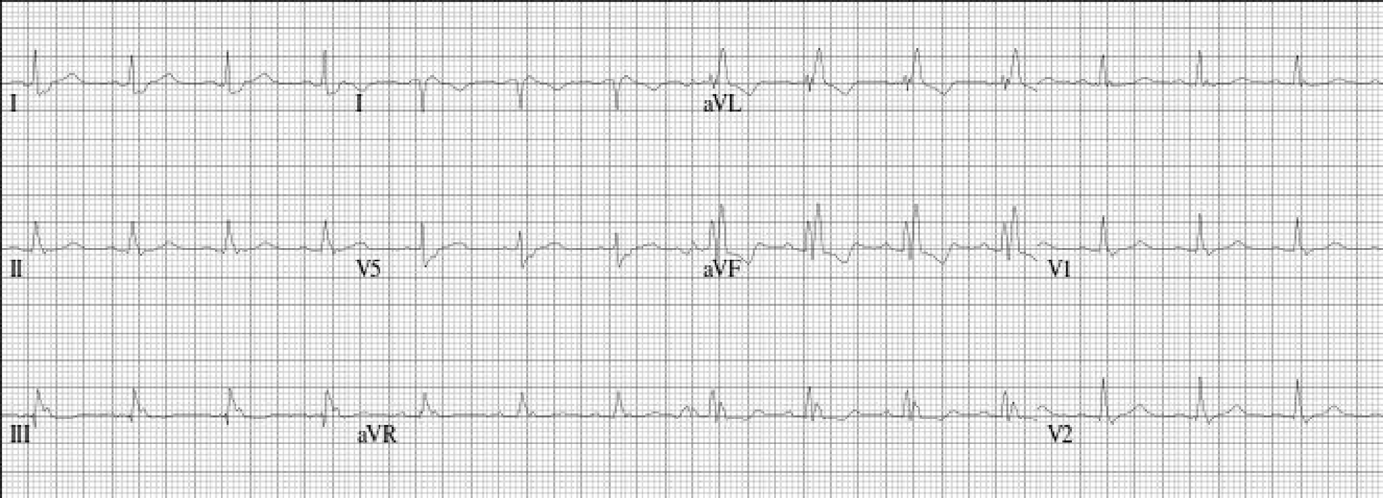
I
I
aVL
II
V5
aVF
V1
III
aVR
V2

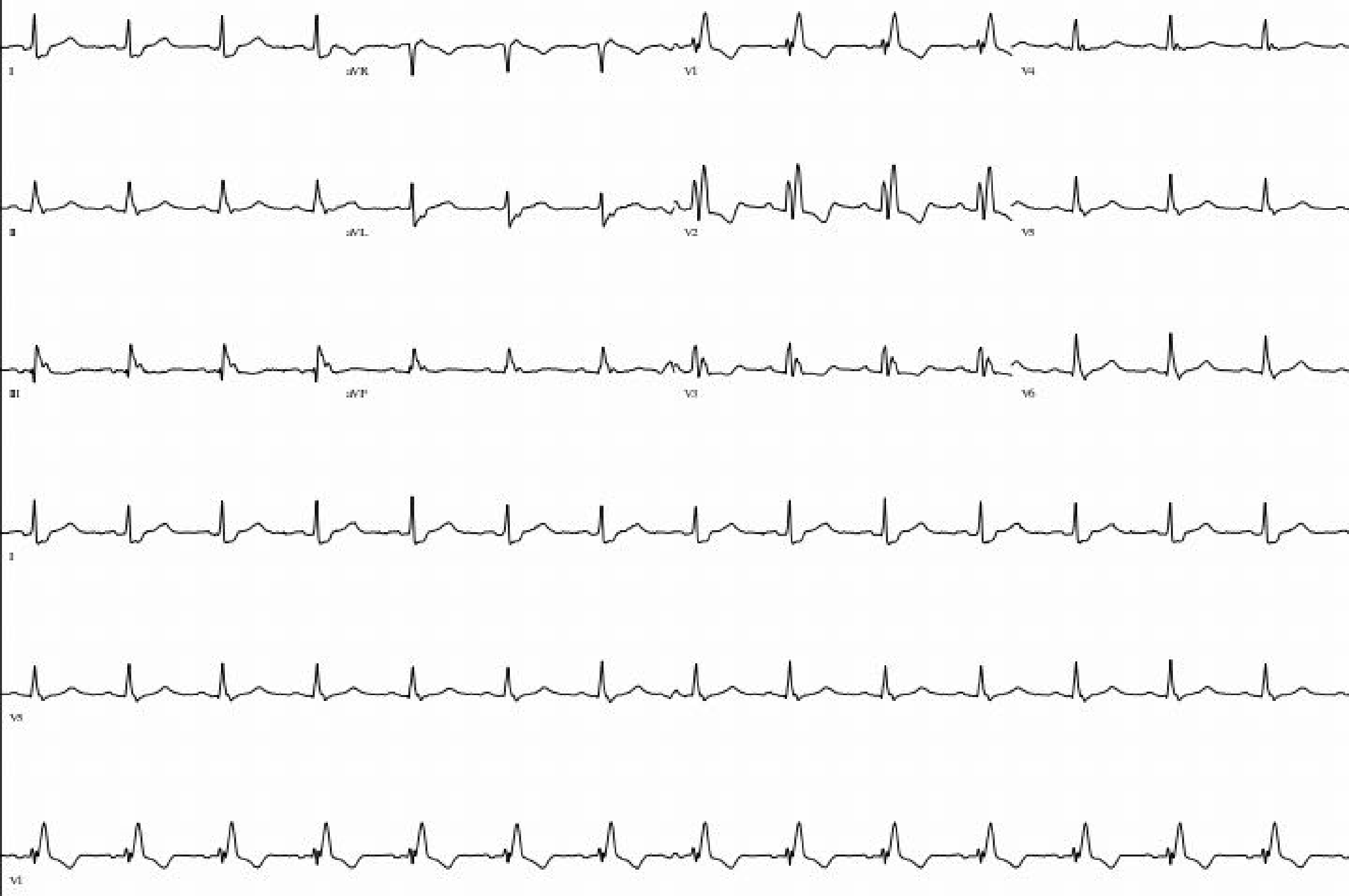
I
aVR
V1
V4
II
aVL
V2
V5
III
aVF
V3
V6
I
V5
V1

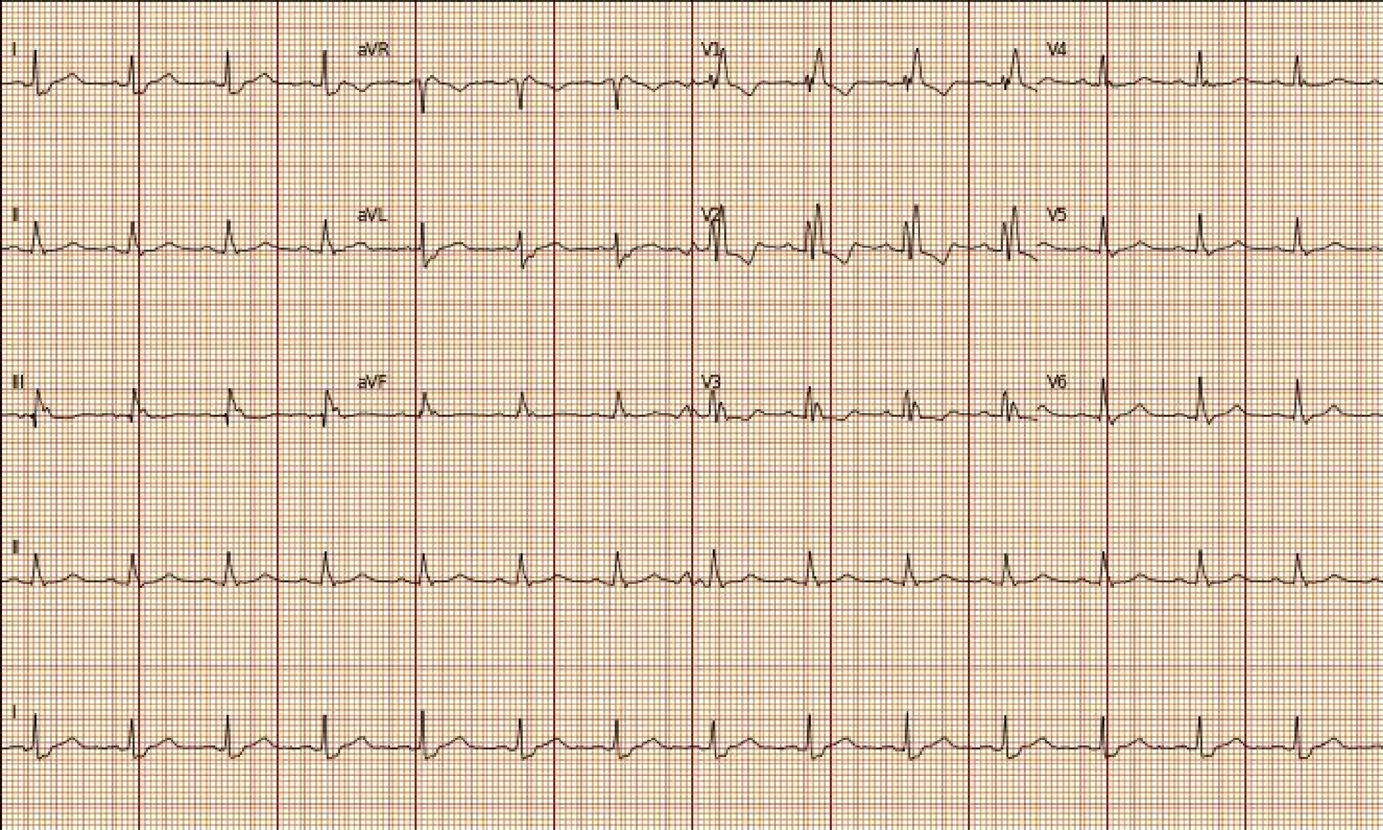

I
aVR
V1
V4
II
aVL
V2
V5
III
aVF
V3
V6
II
I

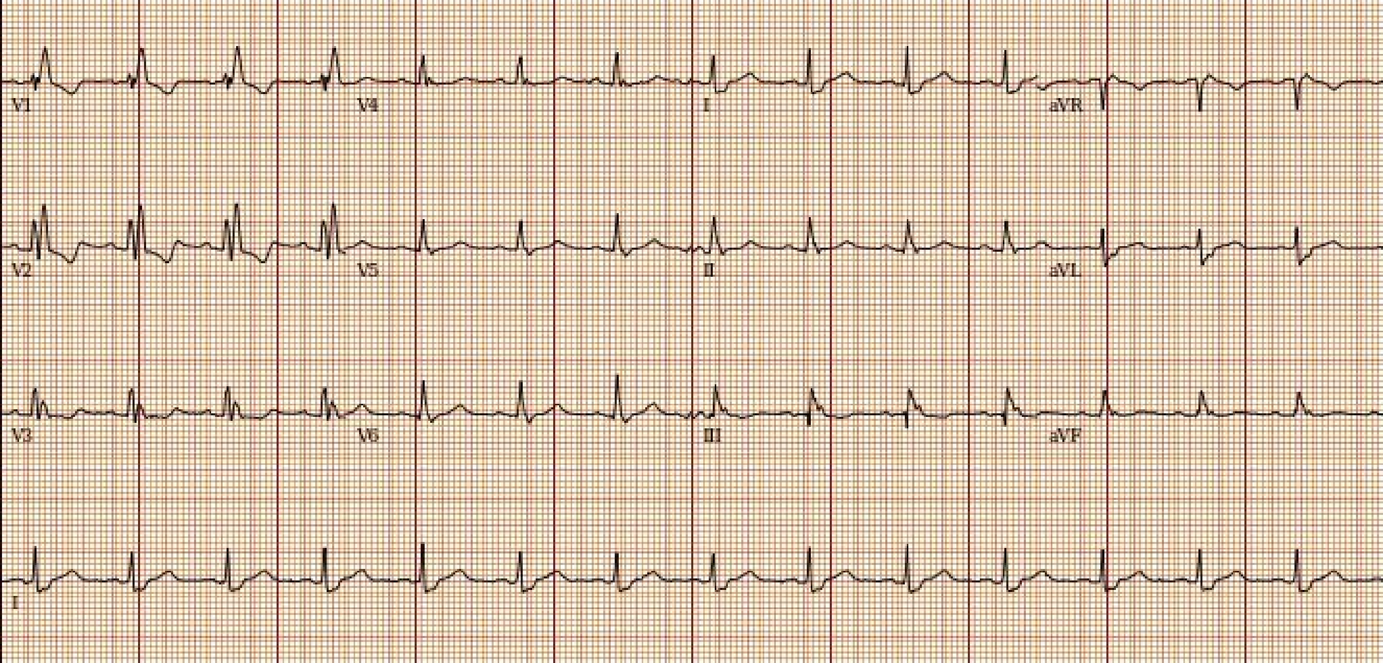
V1
V4
I
aVR
V2
V5
II
aVL
V3
V6
III
aVF
I

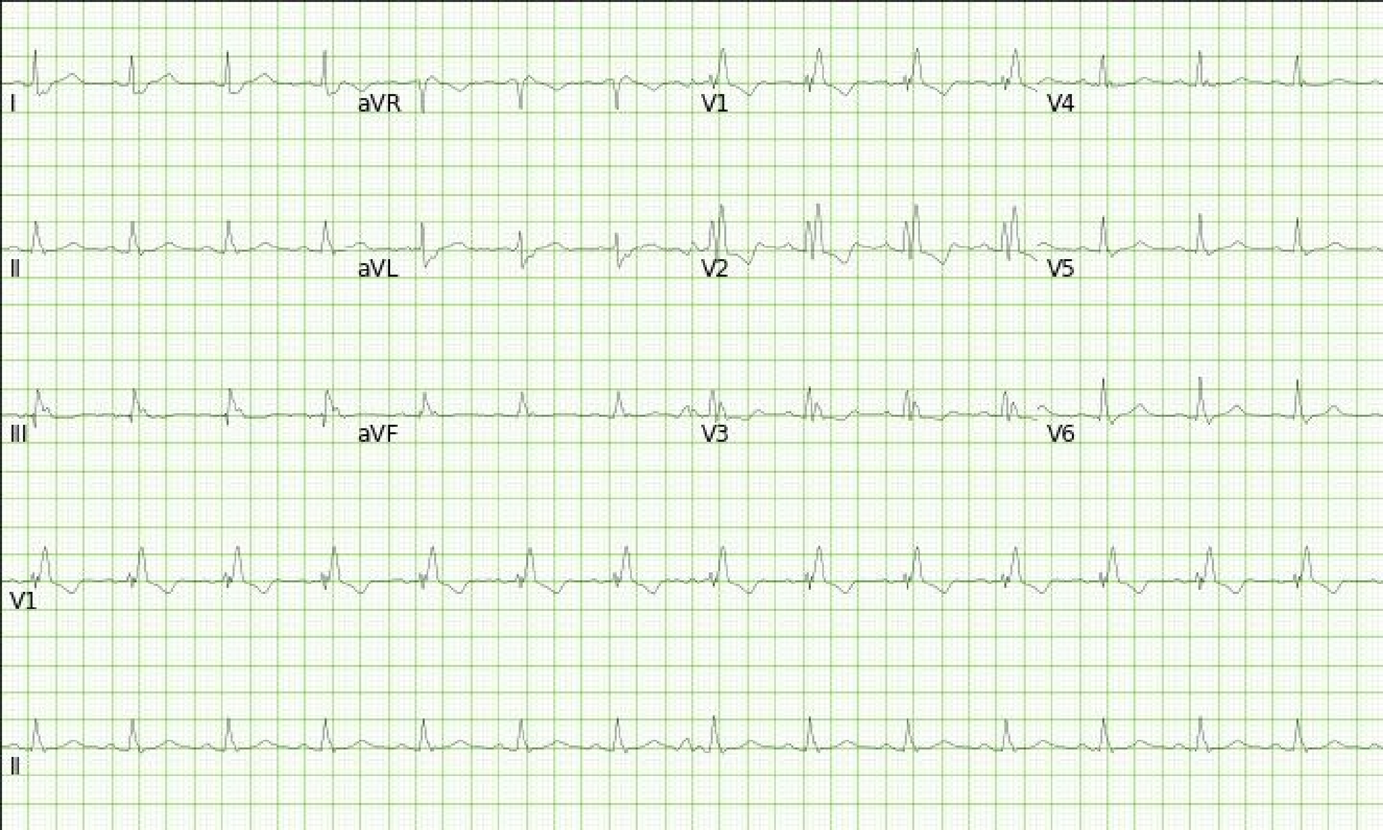
I
aVR
V1
V4
II
aVL
V2
V5
III
aVF
V3
V6
V1
II

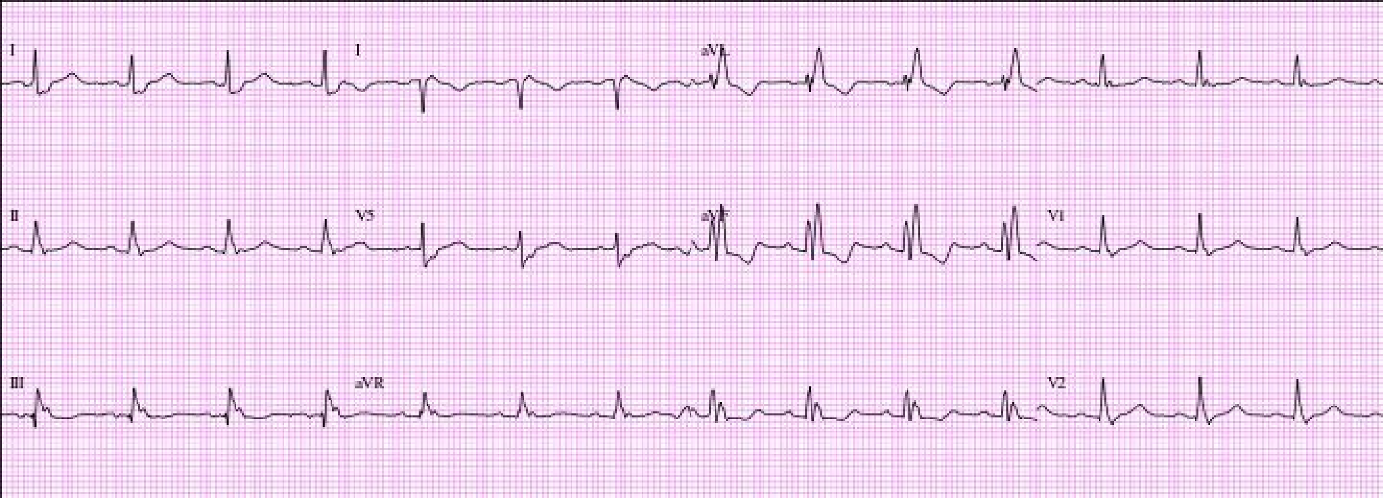
I
I
aVL
II
V5
aVF
V1
III
aVR
V2

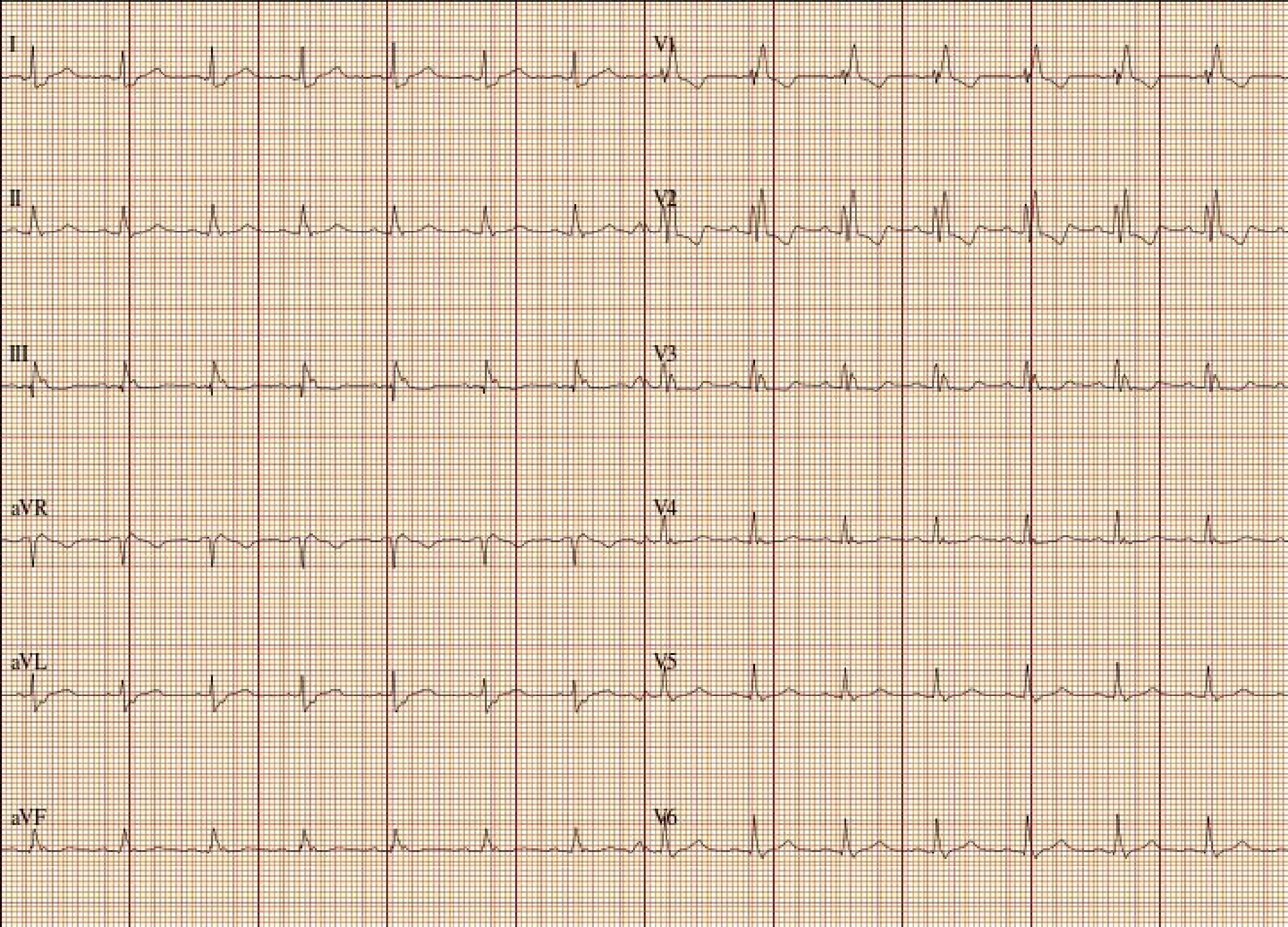
I
II
III
aVR
aVL
aVF
V1
V2
V3
V4
V5
V6

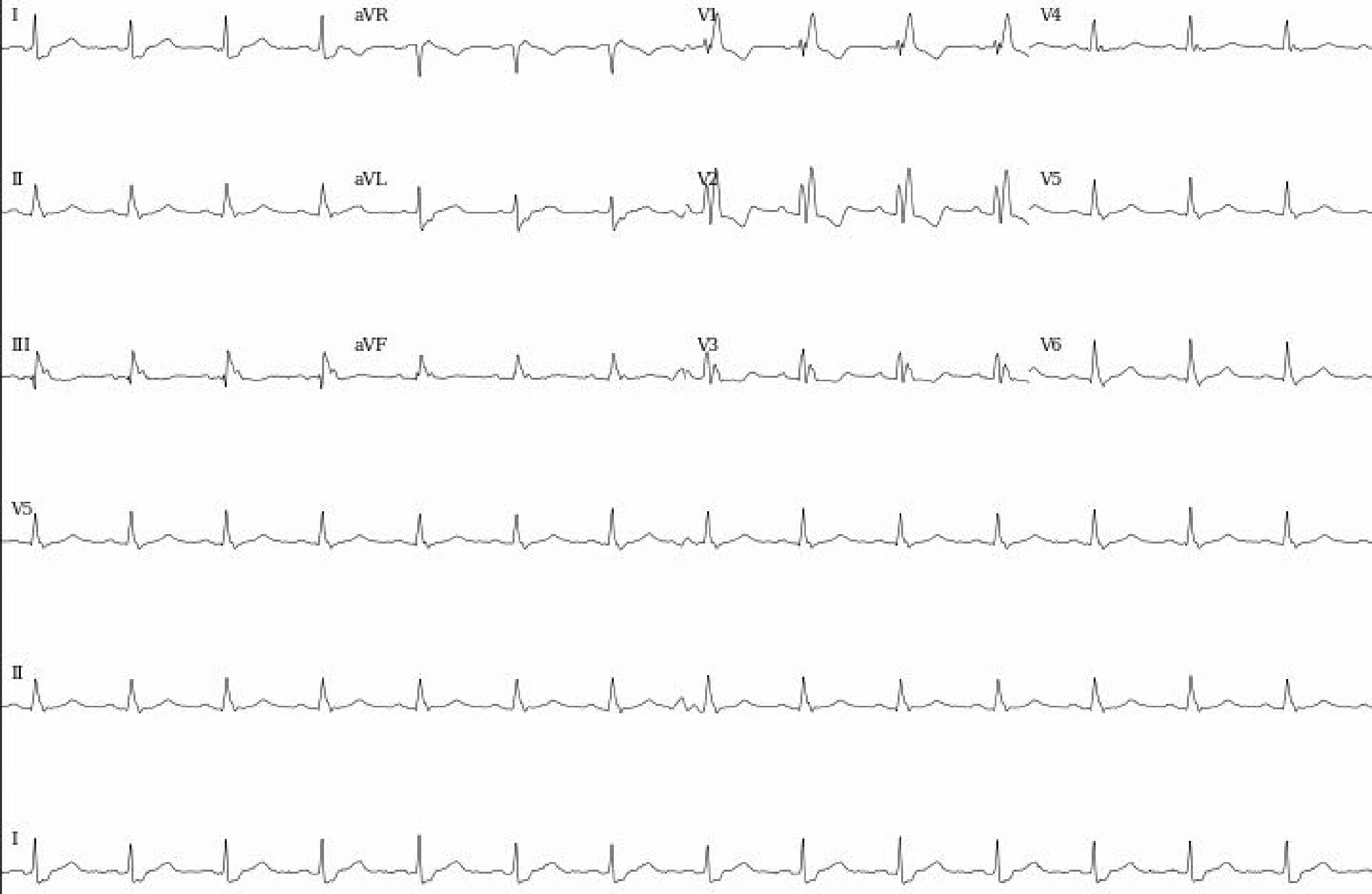
I
aVR
V1
V4
II
aVL
V2
V5
III
aVF
V3
V6
V5
II
I

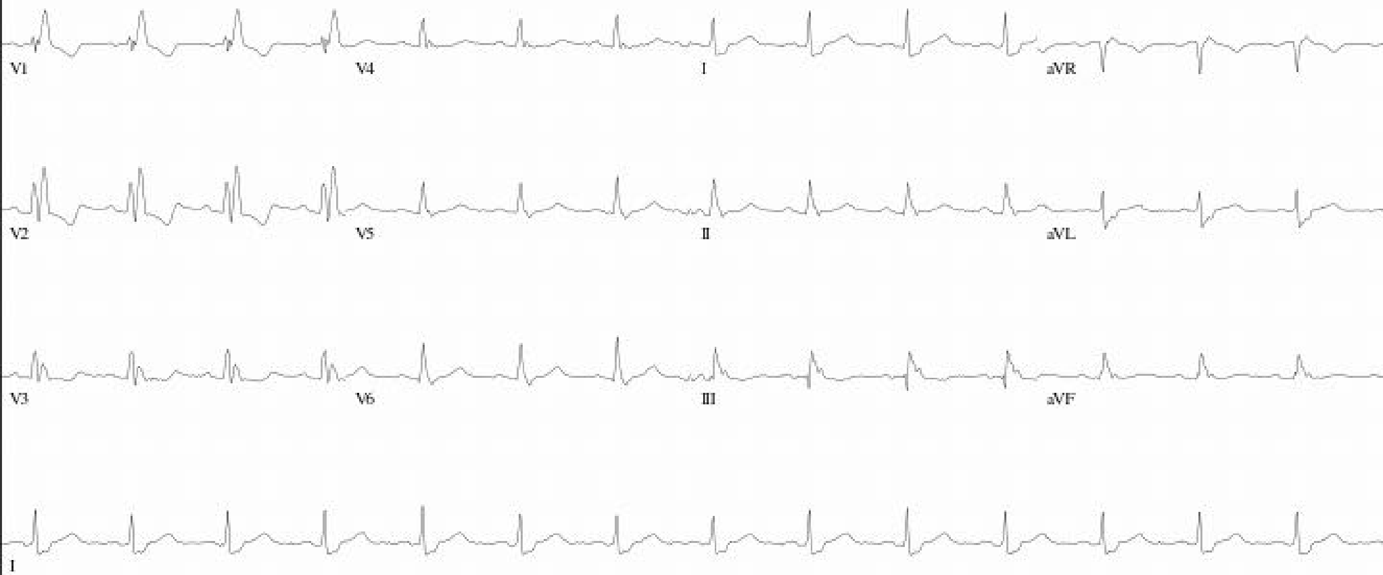
V1
V4
I
aVR
V2
V5
II
aVL
V3
V6
III
aVF
I

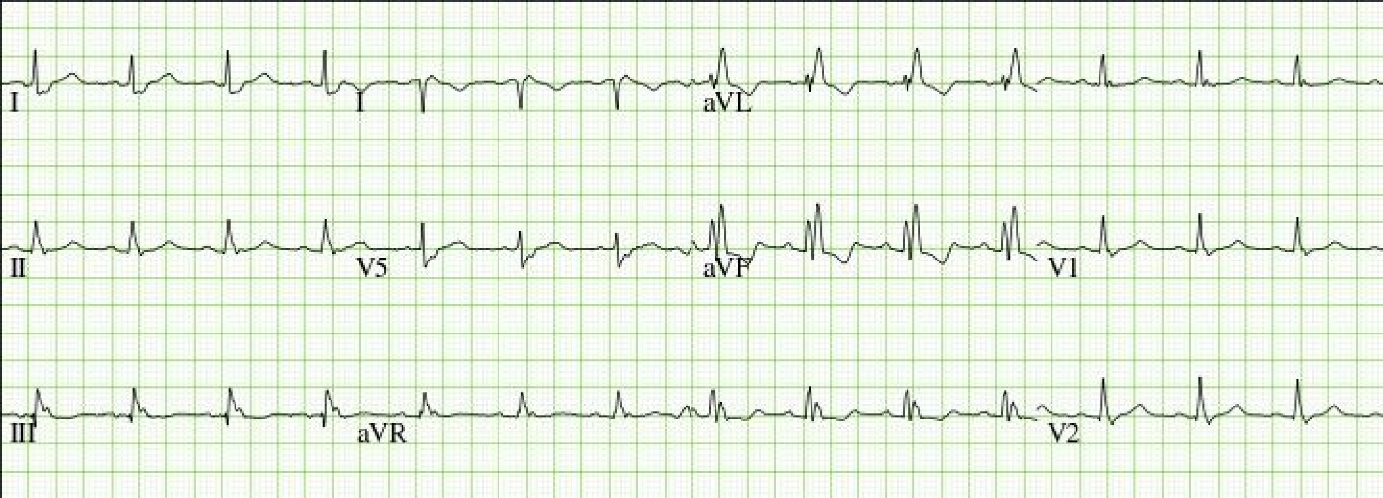
I
I
aVL
II
V5
aVF
V1
III
aVR
V2

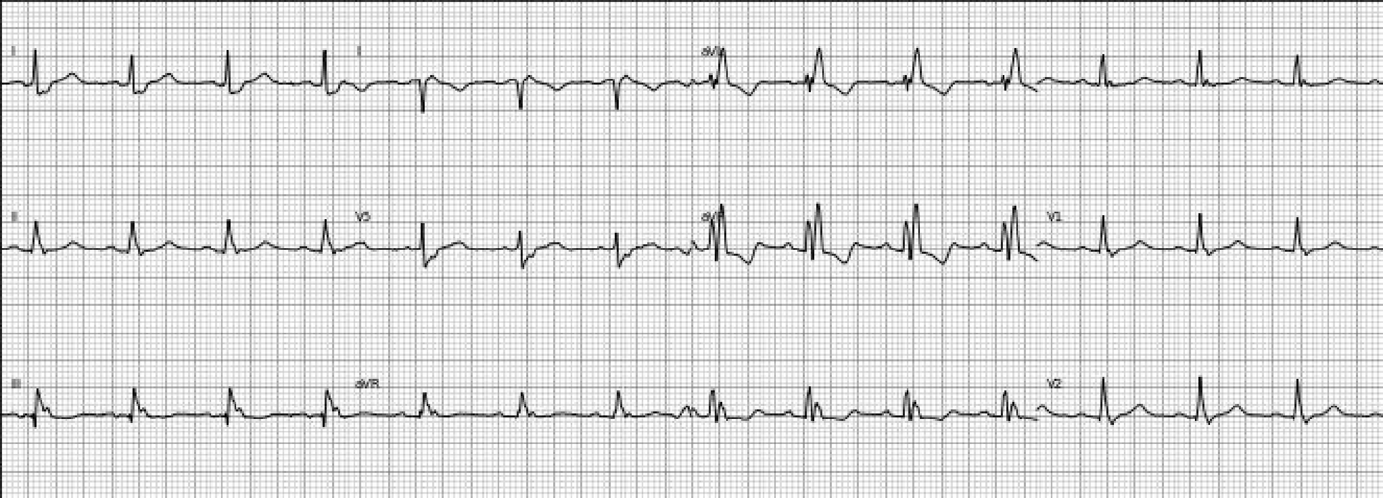
I
II
V5
III
aVR
V1
V2

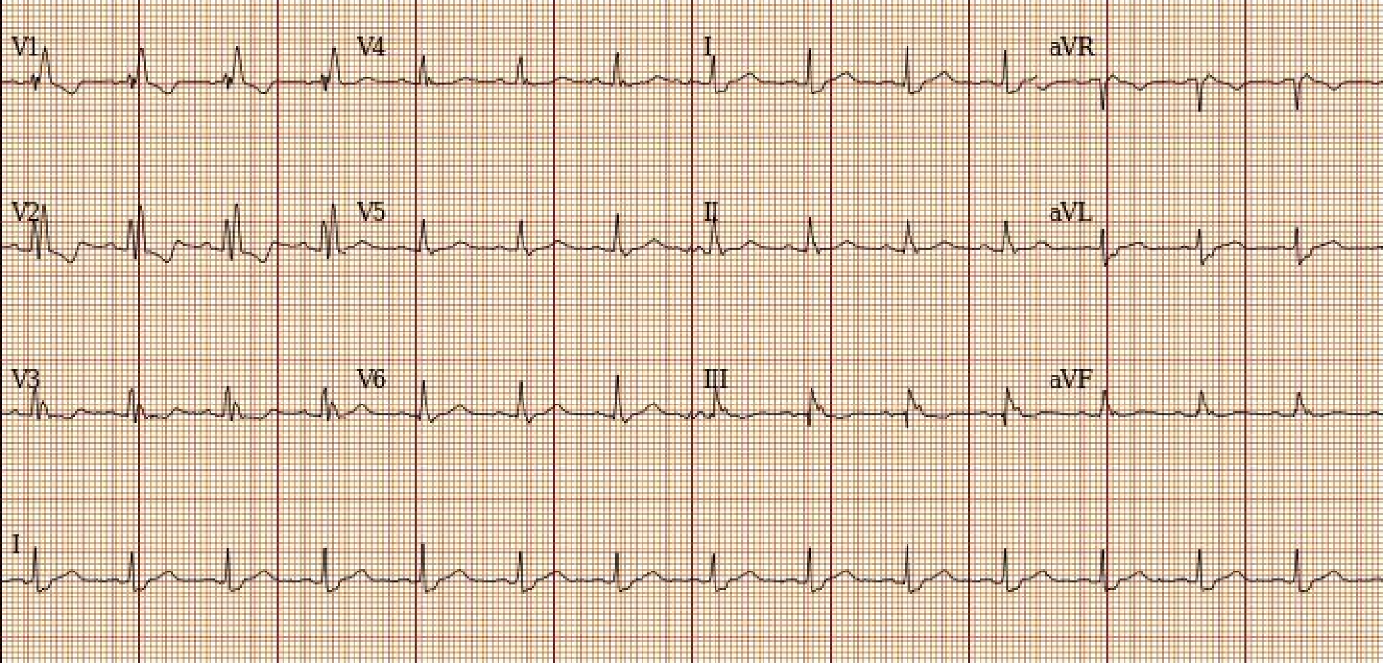
V1
V4
I
aVR
V2
V5
II
aVL
V3
V6
III
aVF
I

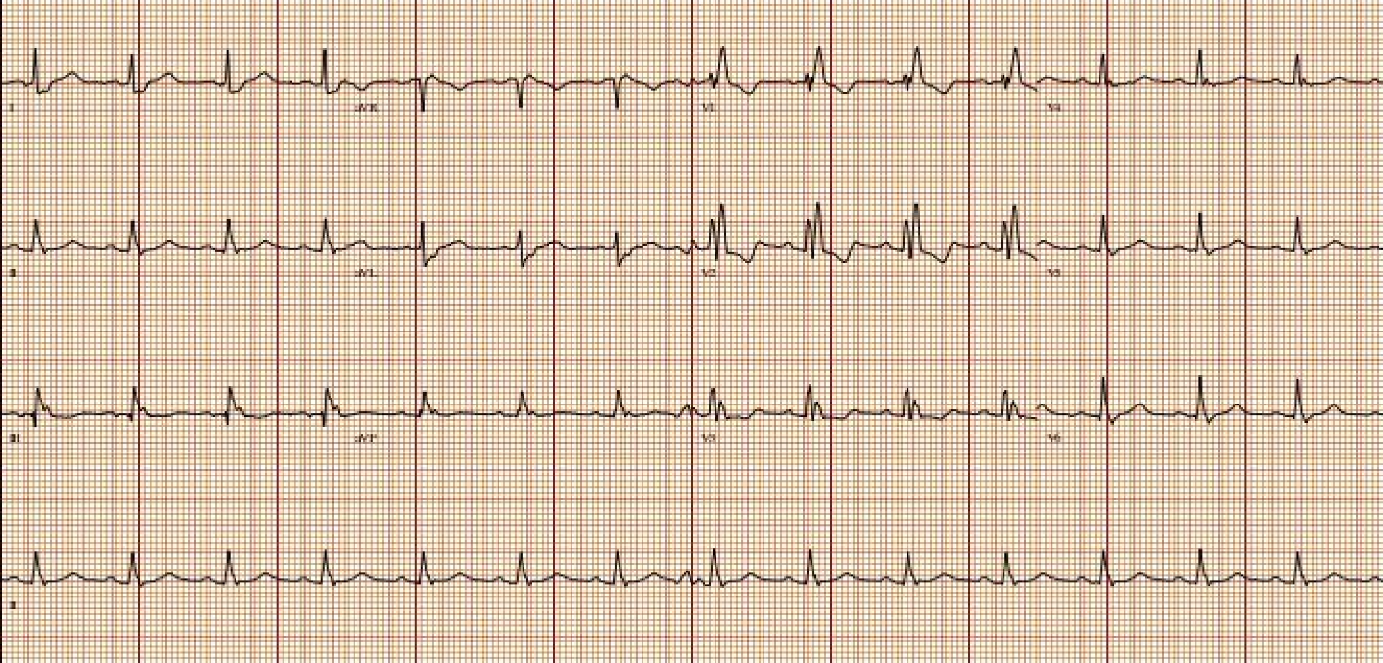

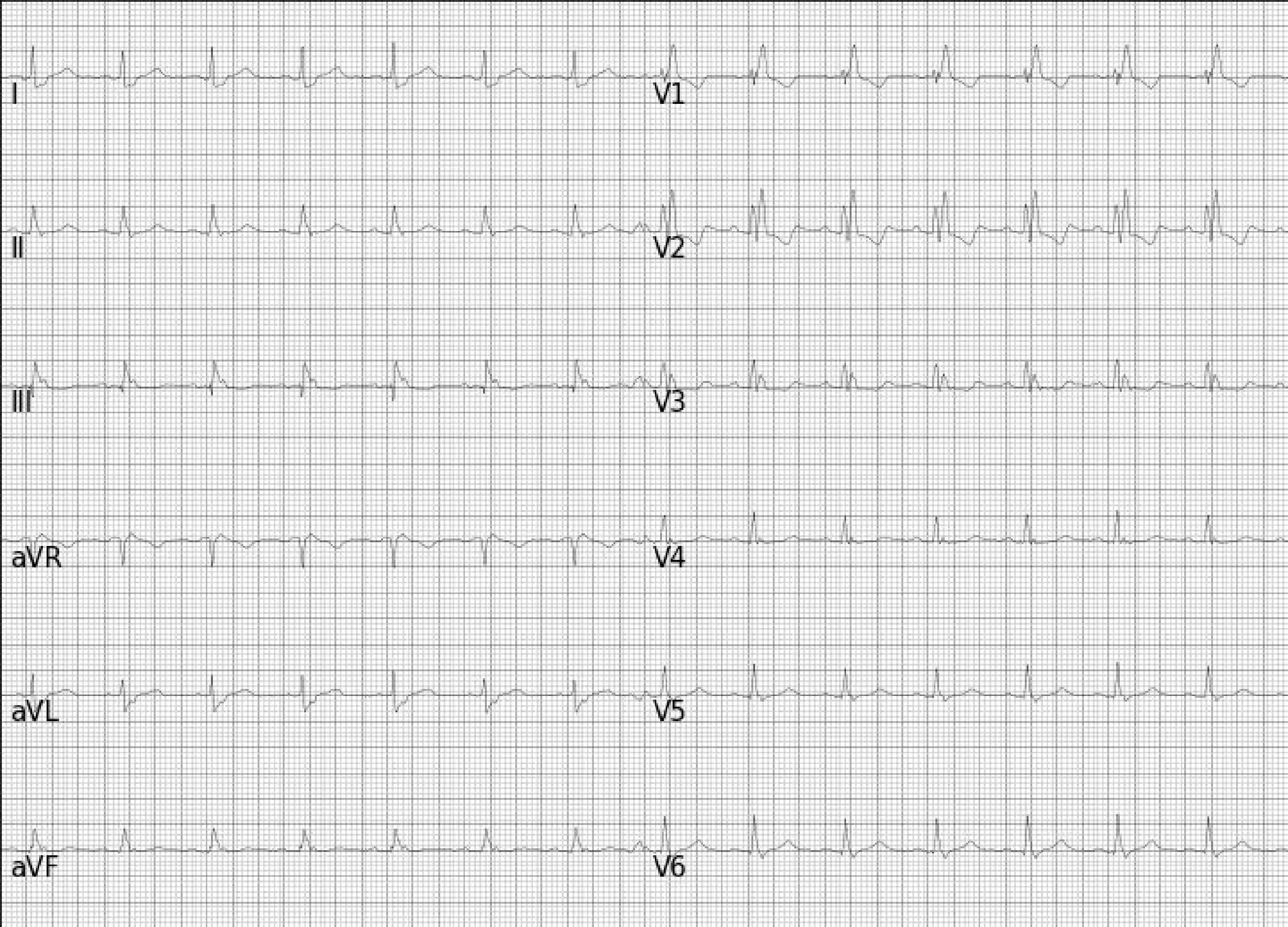
I
II
III
aVR
aVL
aVF
V1
V2
V3
V4
V5
V6

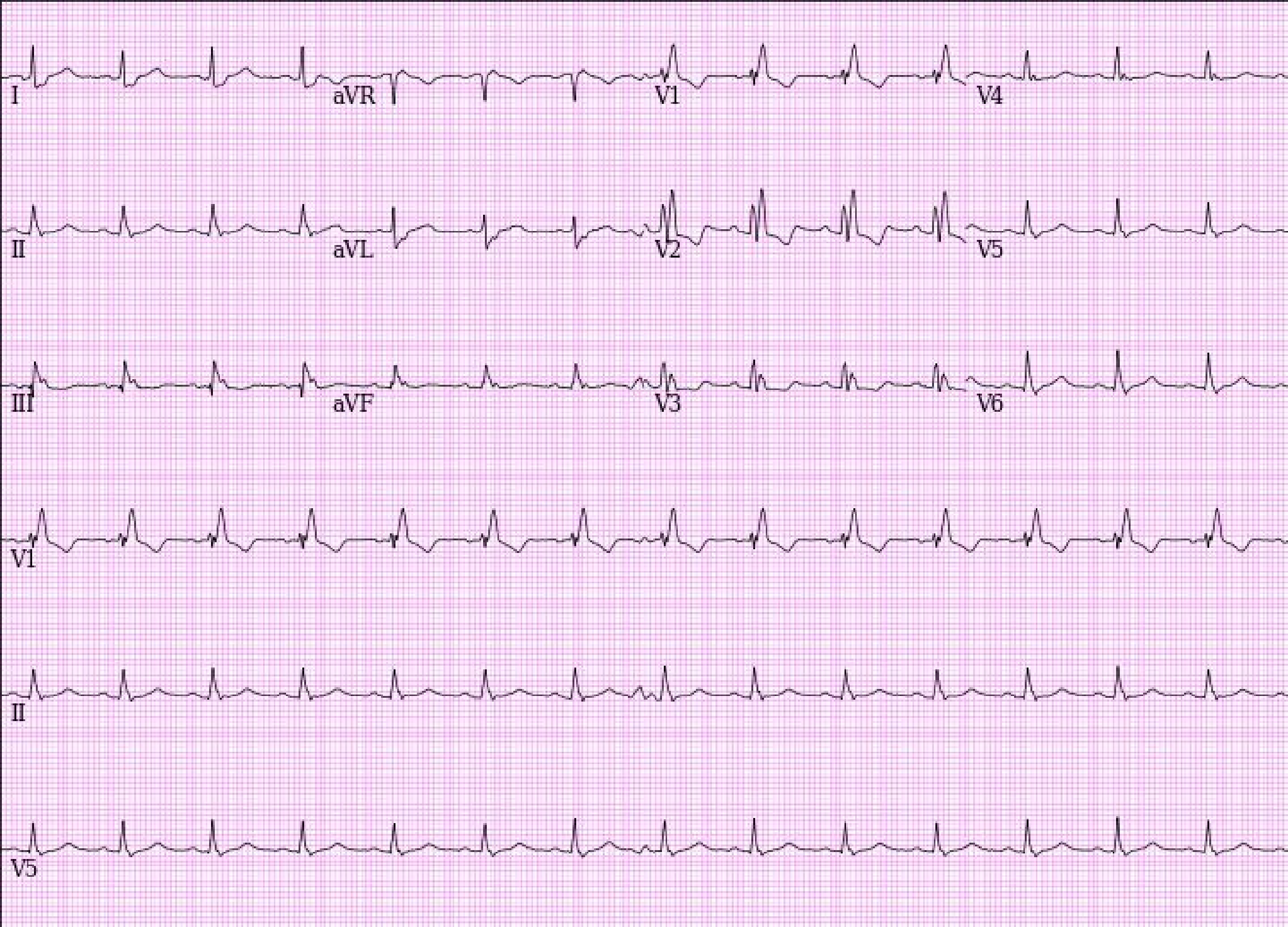
I
aVR
V1
V4
II
aVL
V2
V5
III
aVF
V3
V6
V1
II
V5

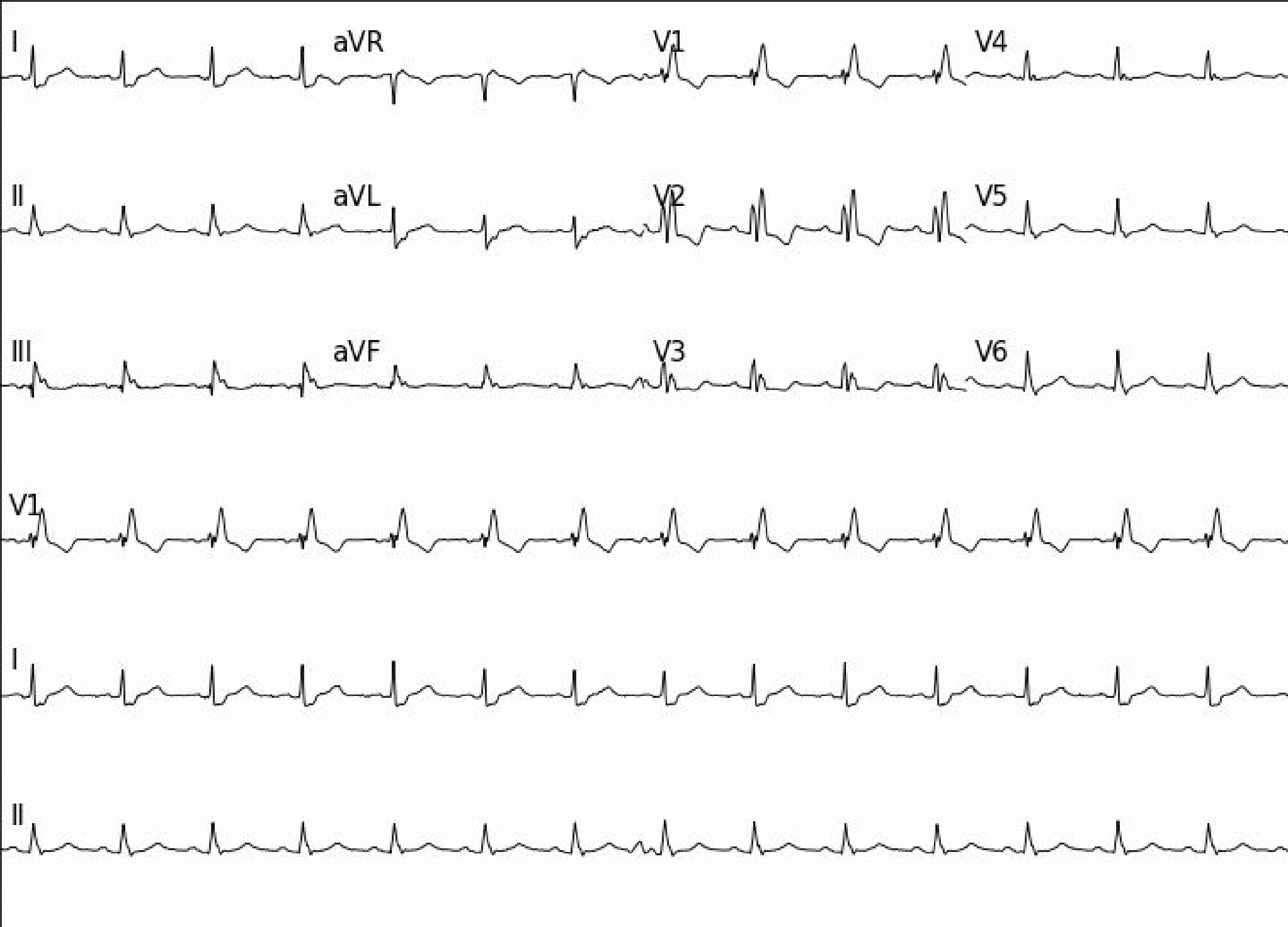
I
aVR
V1
V4
II
aVL
V2
V5
III
aVF
V3
V6
V1
I
II

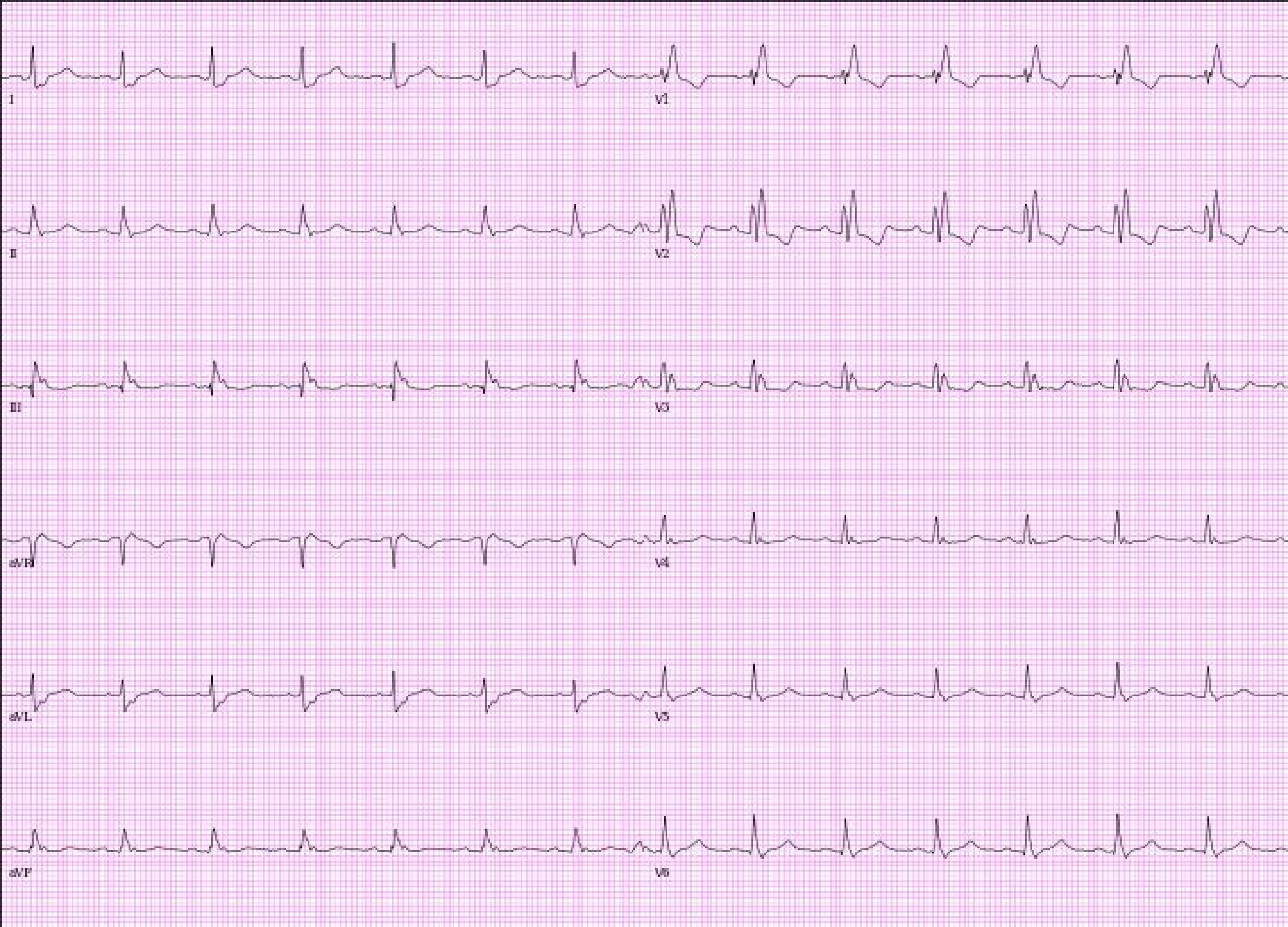

I
V1
II
V2
III
V3
aVR
V4
aVL
V5
aVF
V6

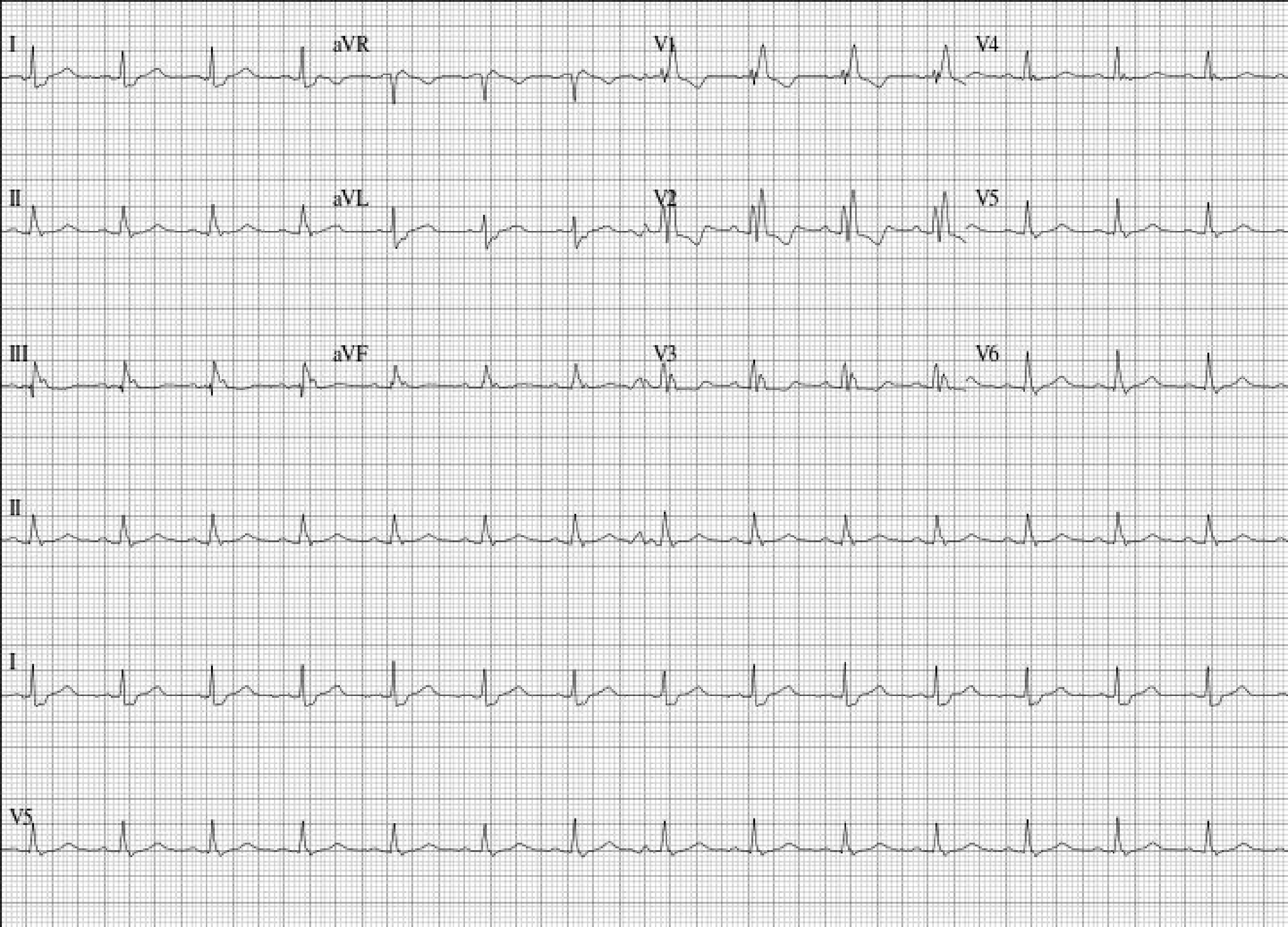

I
aVR
V1
V4
II
aVL
V2
V5
III
aVF
V3
V6
II
I
V5

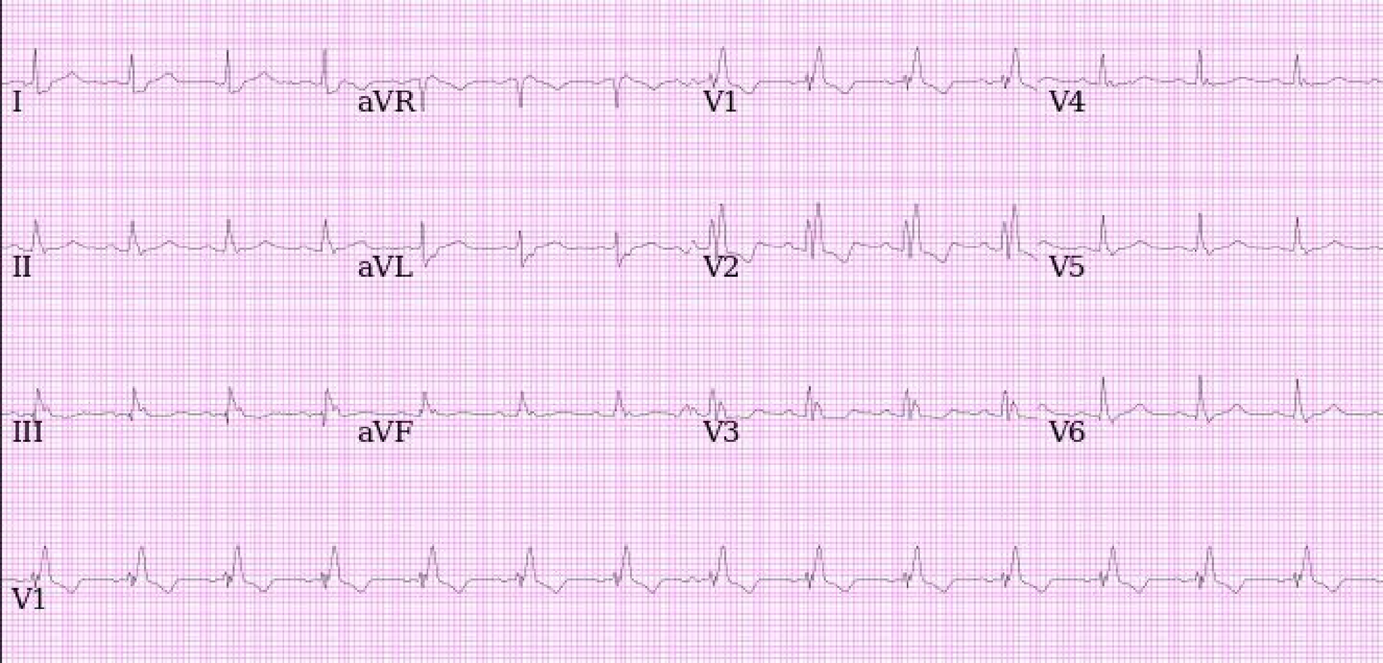
I
aVR
V1
V4
II
aVL
V2
V5
III
aVF
V3
V6
V1

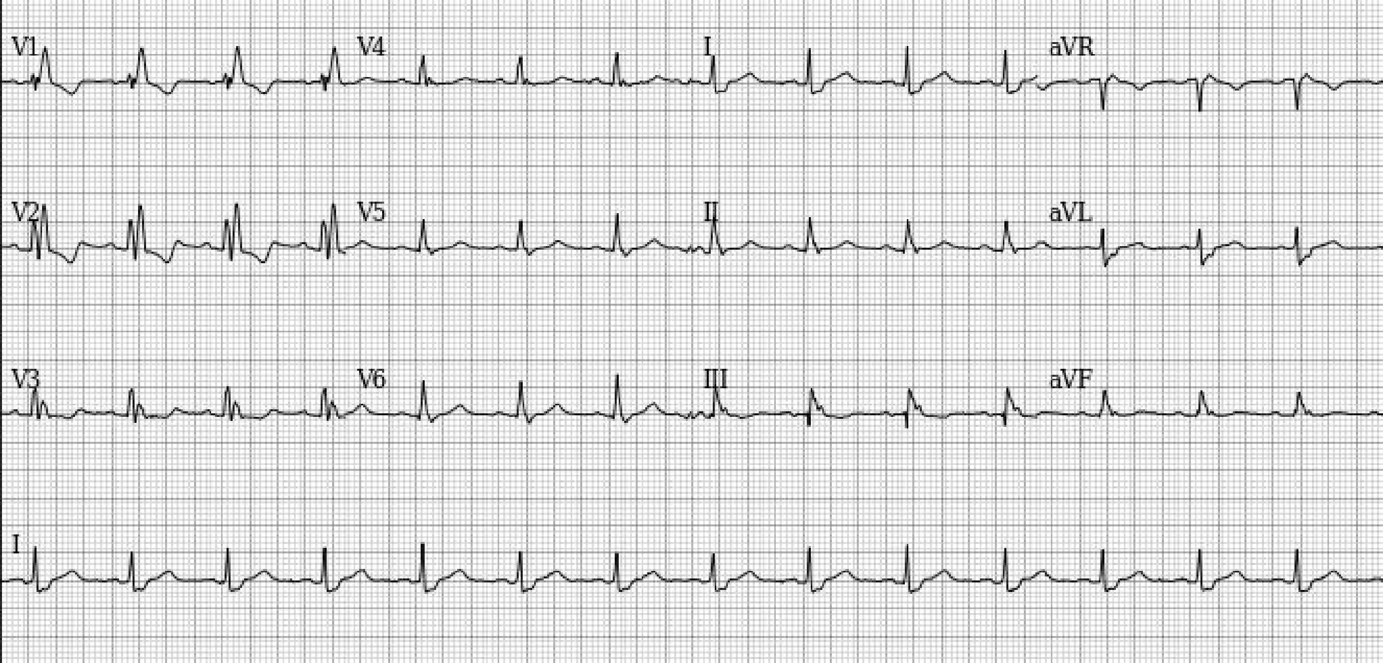
V1
V4
I
aVR
V2
V5
II
aVL
V3
V6
III
aVF
I

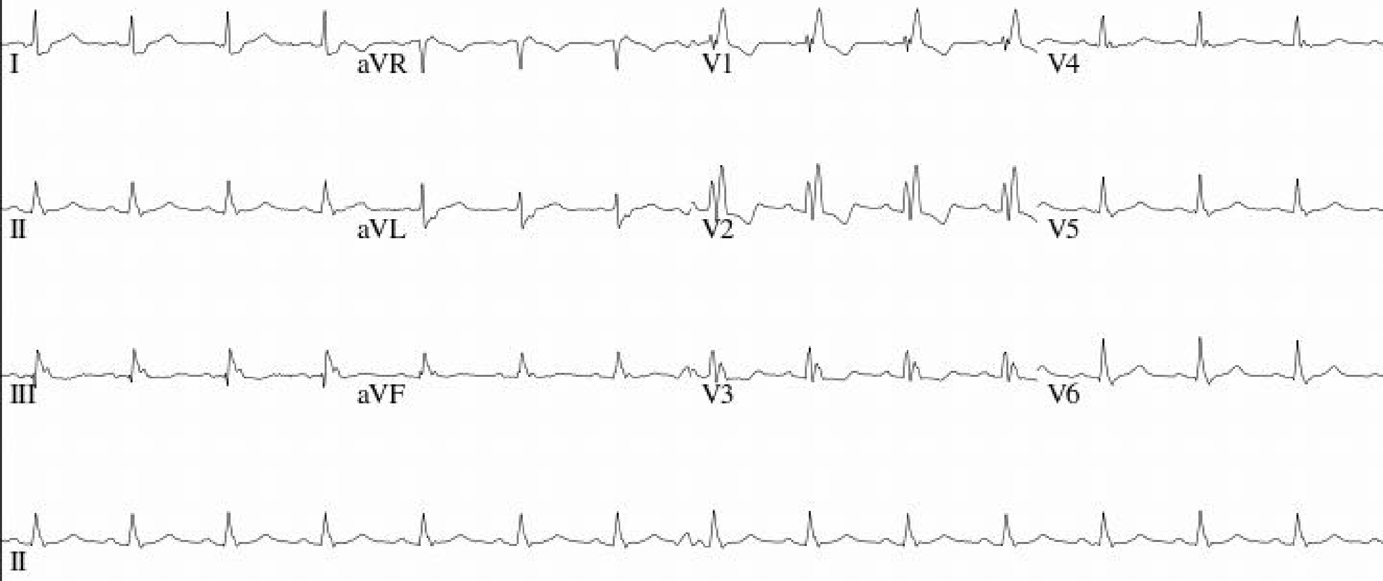
I
aVR
V1
V4
II
aVL
V2
V5
III
aVF
V3
V6
II

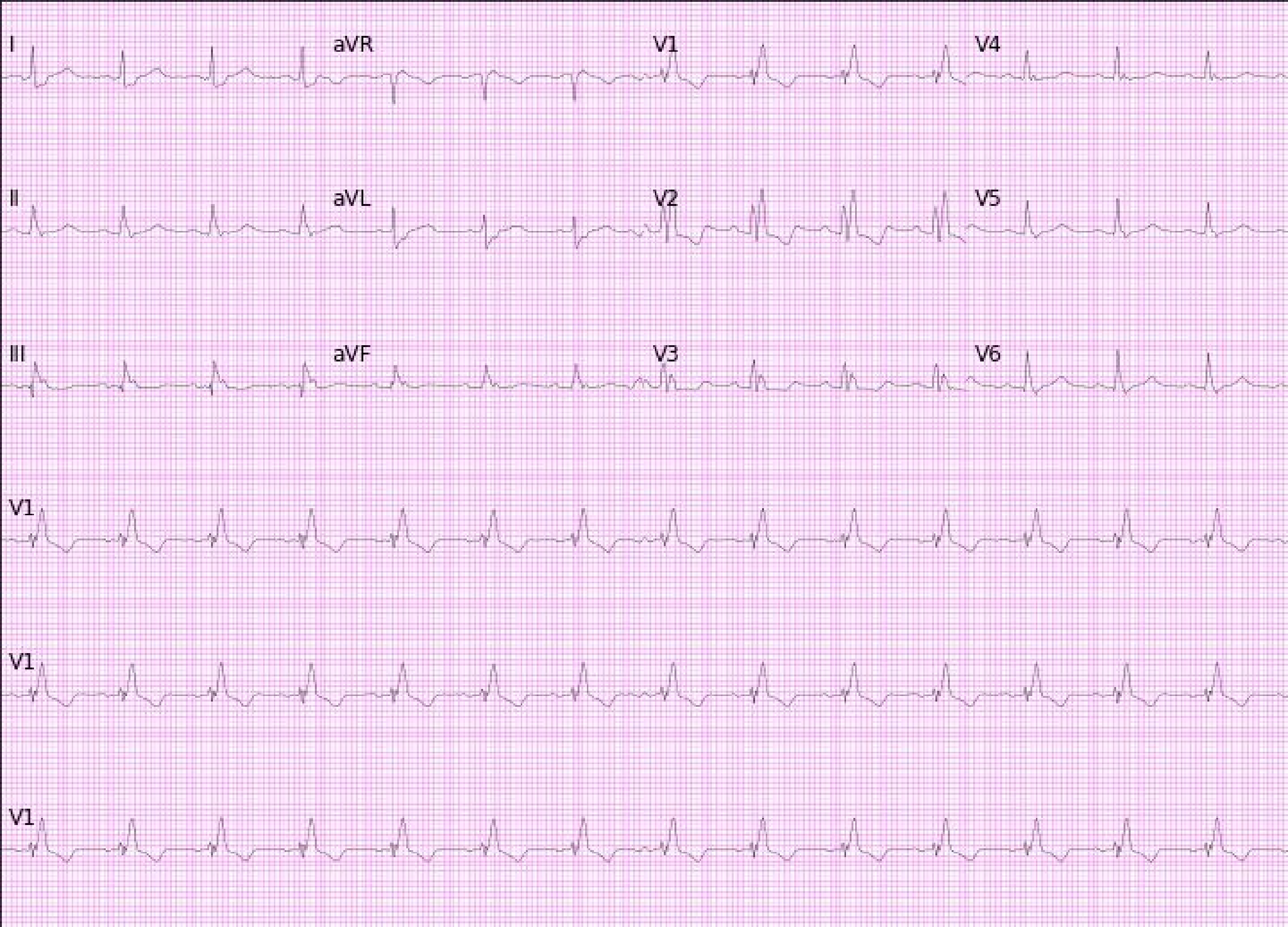
I
aVR
V1
V4
II
aVL
V2
V5
III
aVF
V3
V6
V1
V1
V1

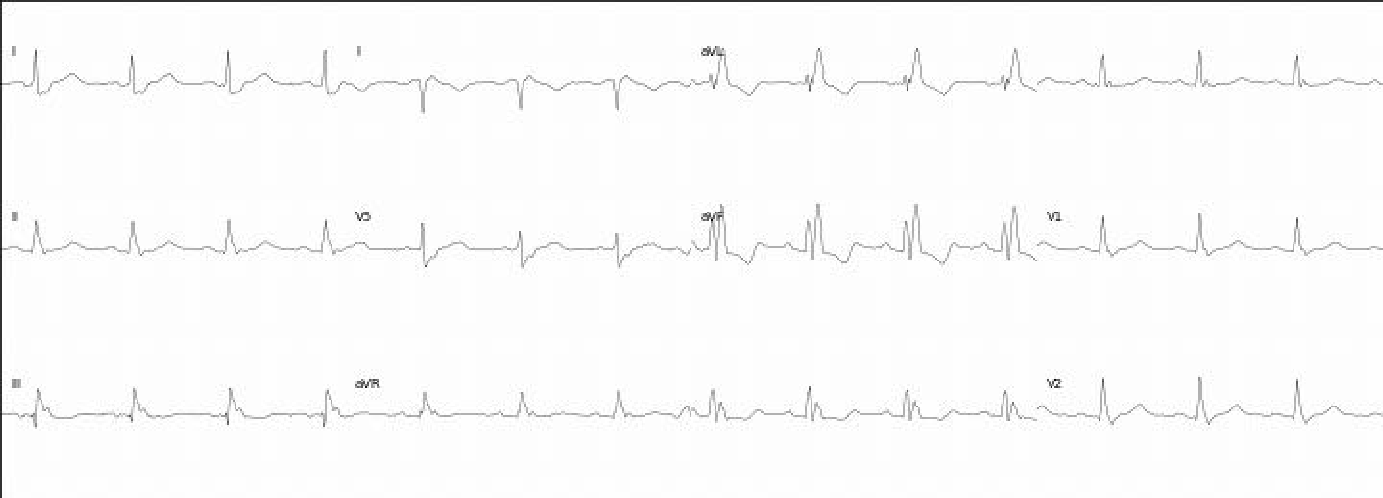
I
aVL
II
aVF
V1
III
aVR
V2

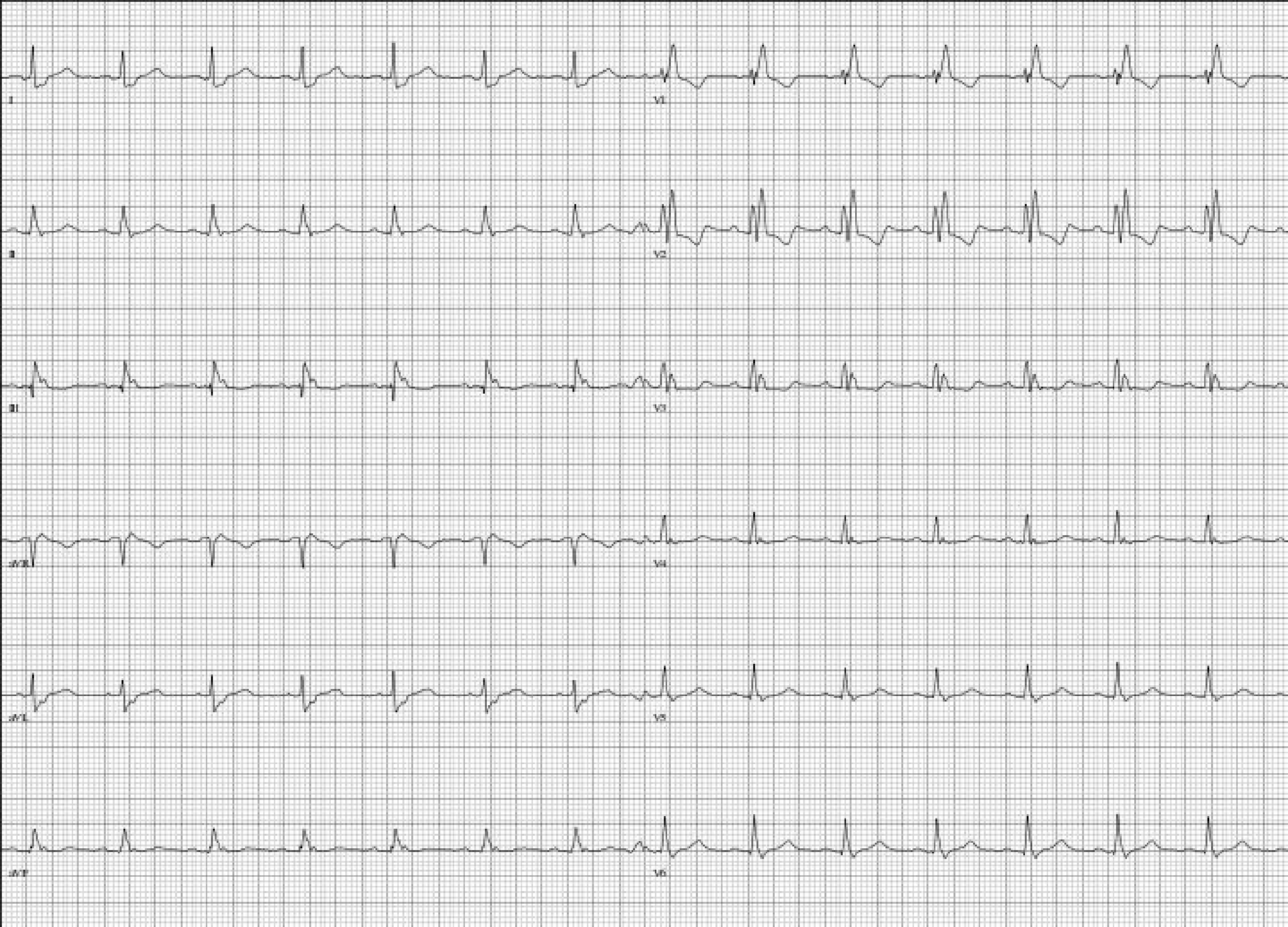
I
V1
II
V2
III
V3
aVR
V4
aVL
V5
aVF
V6

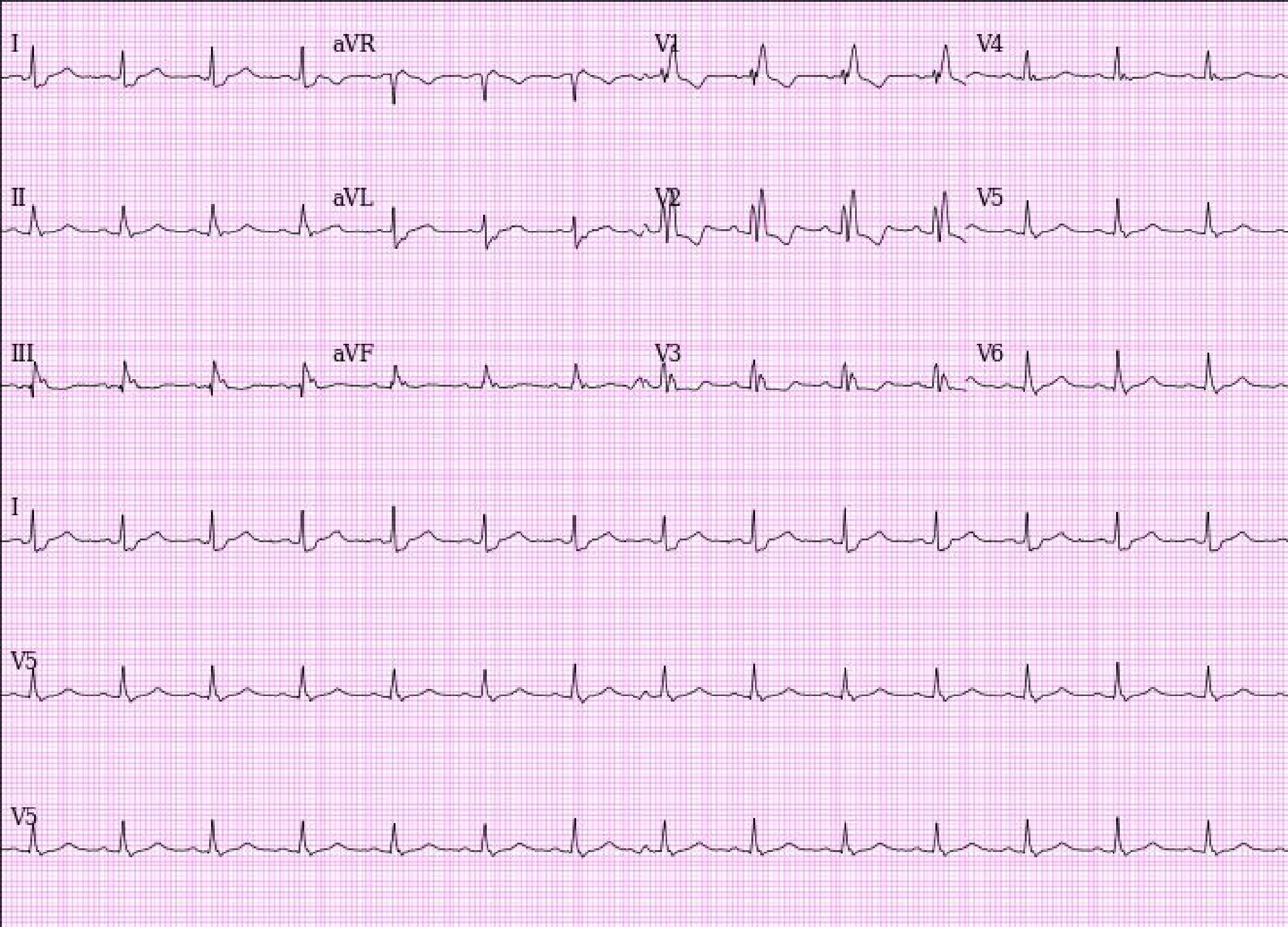

I
aVR
V1
V4
II
aVL
V2
V5
III
aVF
V3
V6
I
V5
V5

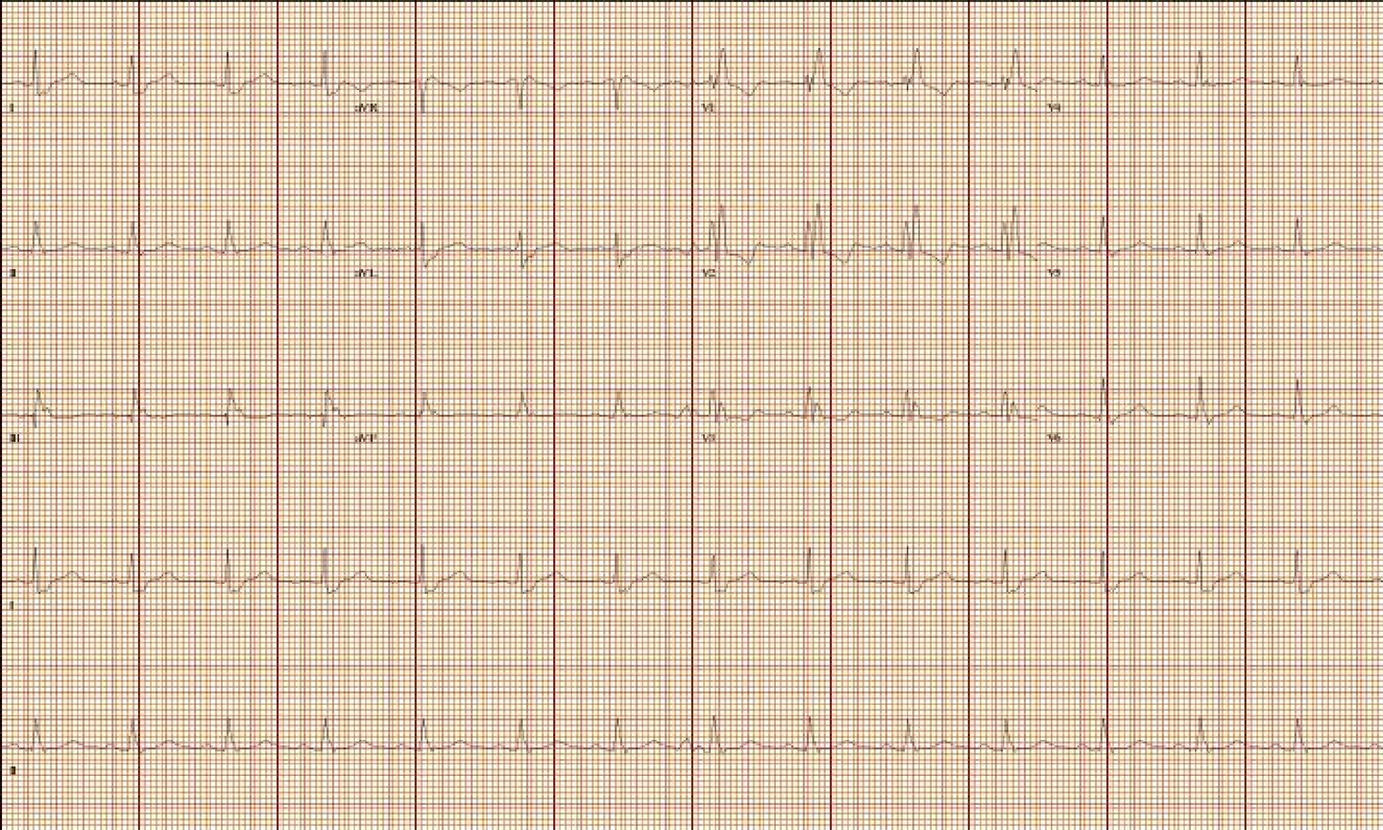
I
aVR
V1
V4
II
aVL
V2
V5
III
aVF
V3
V6

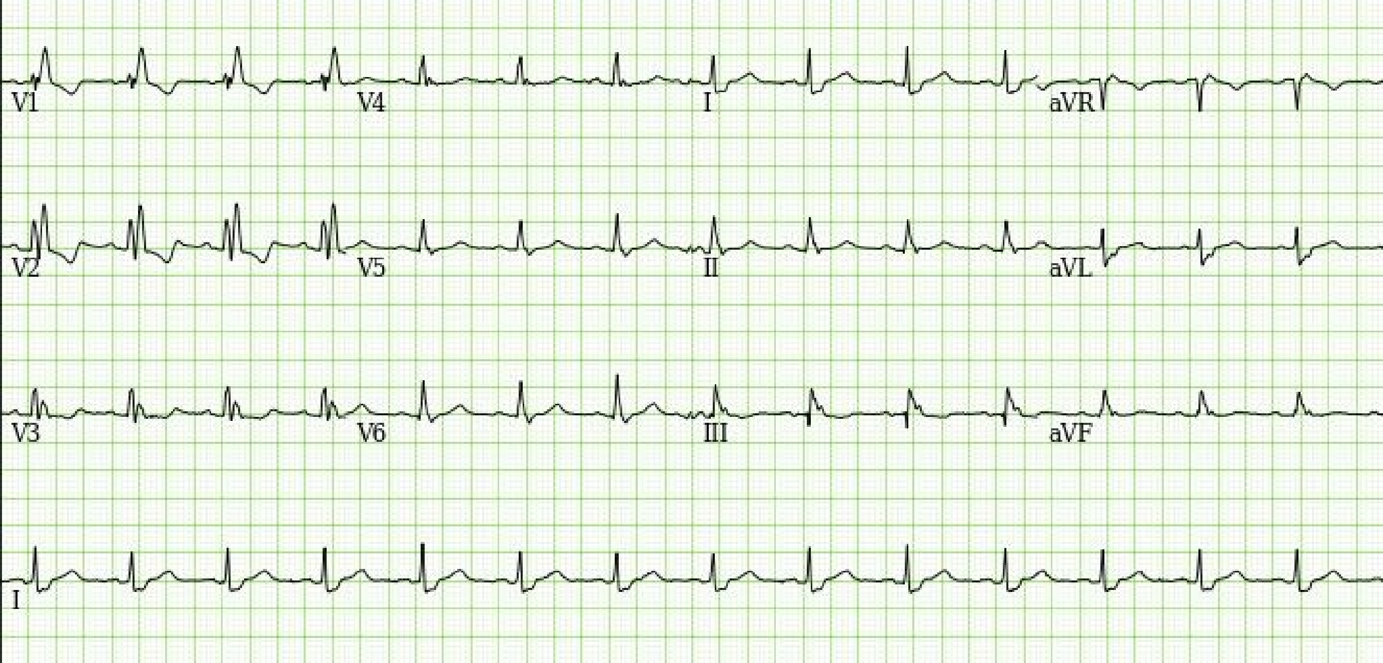
V1
V4
I
aVR
V2
V5
II
aVL
V3
V6
III
aVF
I

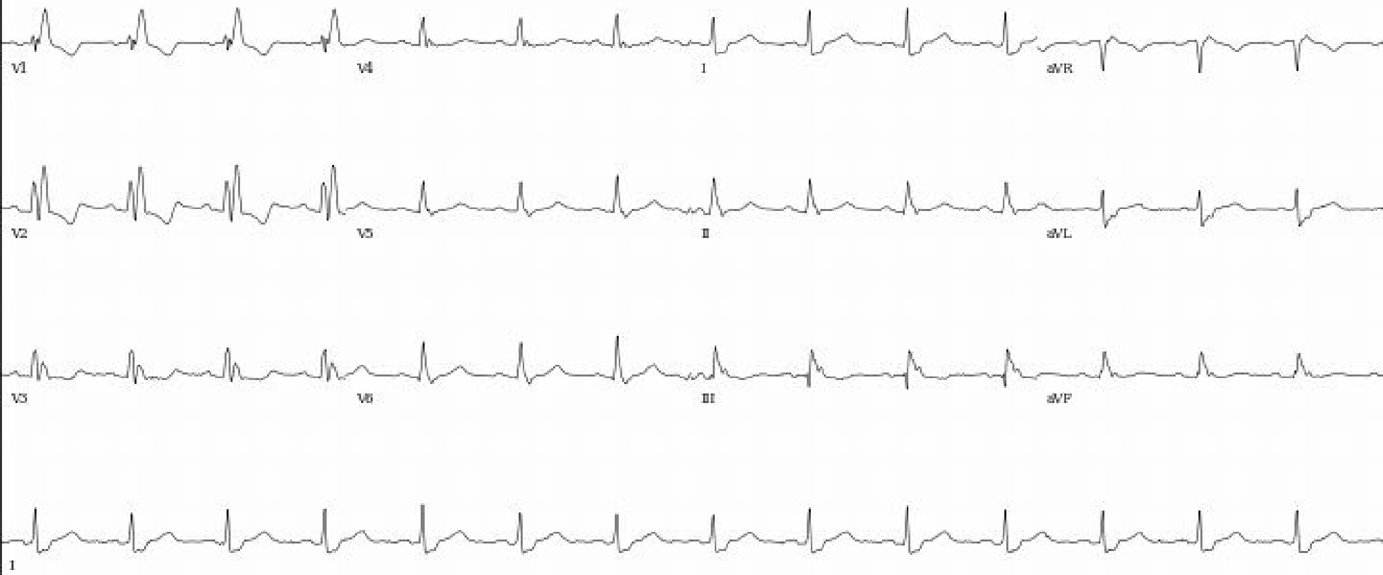
V1
V4
I
aVR
V2
V5
II
aVL
V3
V6
III
aVF
I

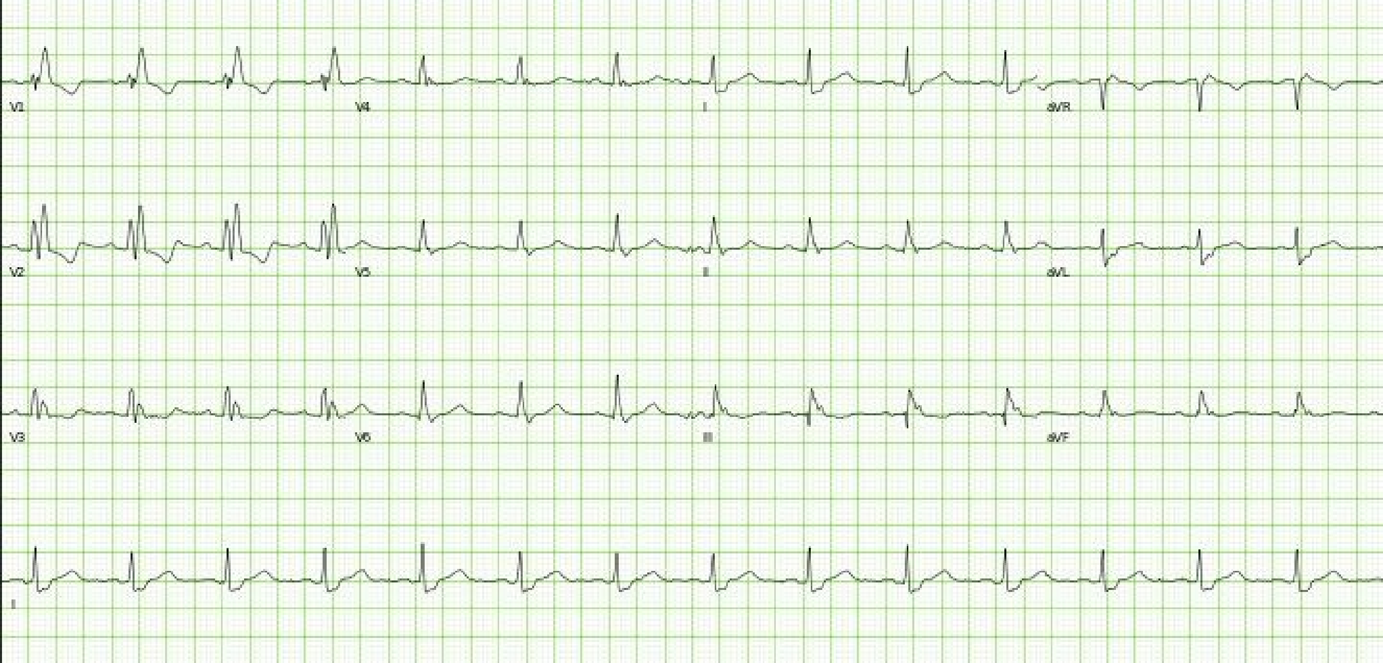
V1
V4
I
aVR
V2
V5
II
aVL
V3
V6
III
aVF
II

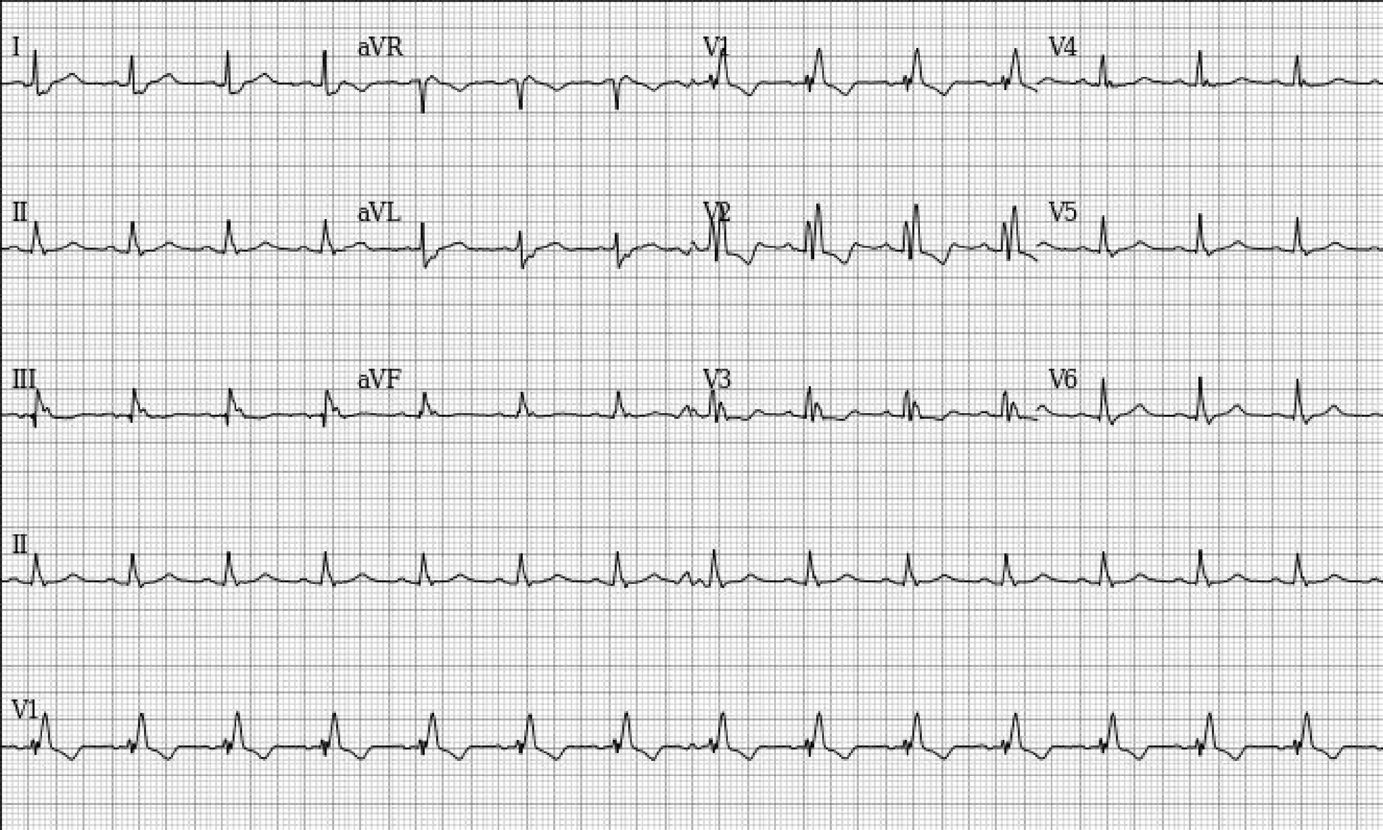
I
aVR
V1
V4
II
aVL
V2
V5
III
aVF
V3
V6
II
V1

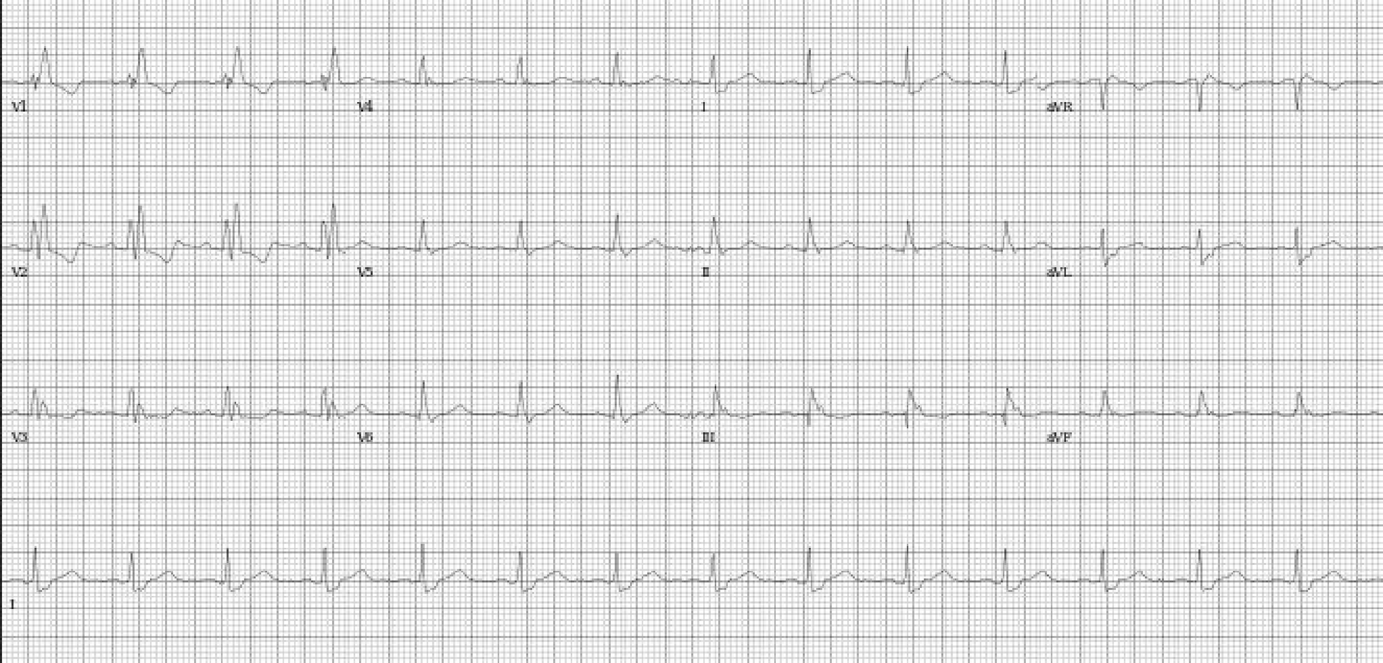
V1
V4
I
aVR
V2
V5
II
aVL
V3
V6
III
aVF
I

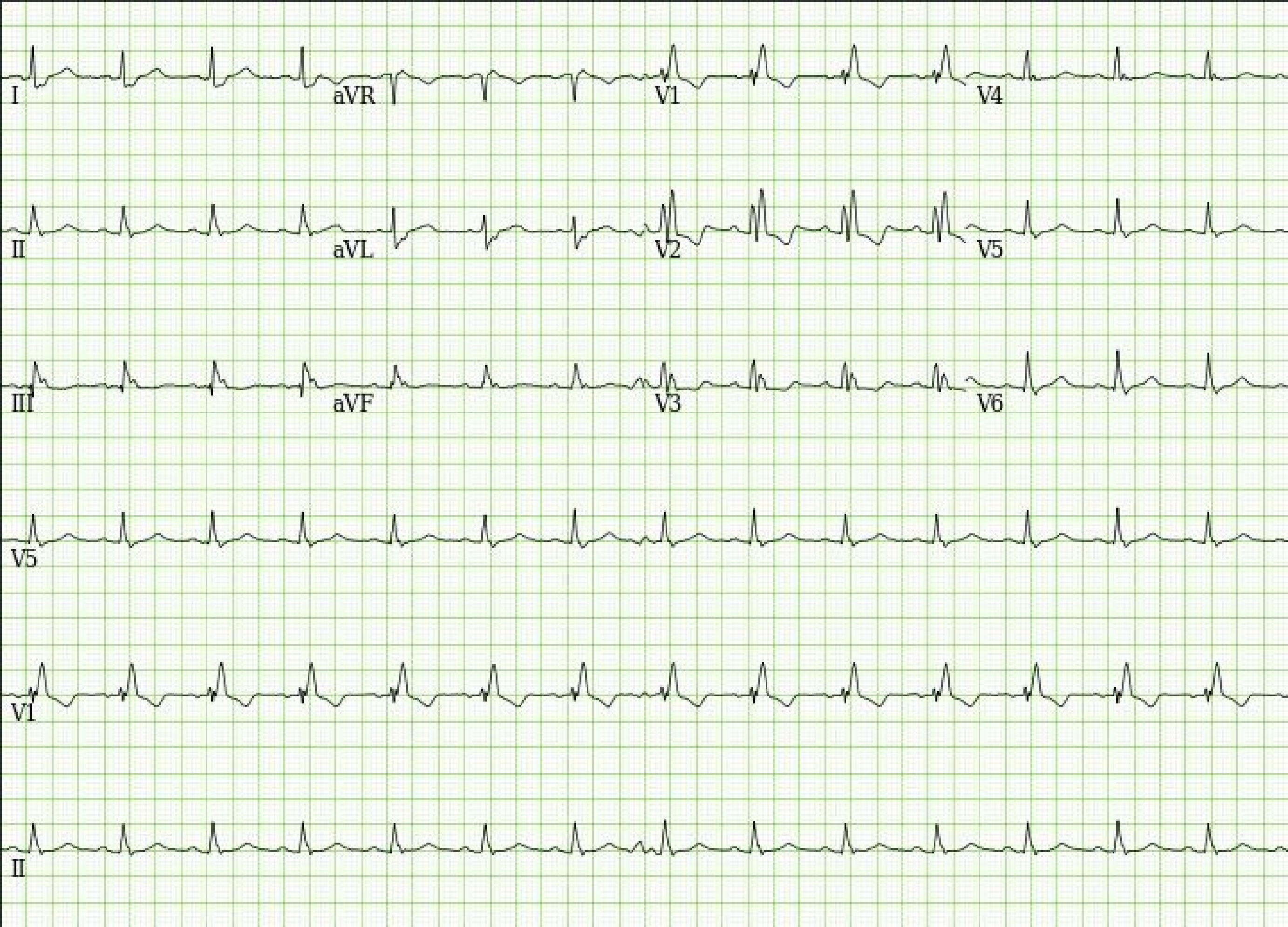
I
aVR
V1
V4
II
aVL
V2
V5
III
aVF
V3
V6
V5
V1
II

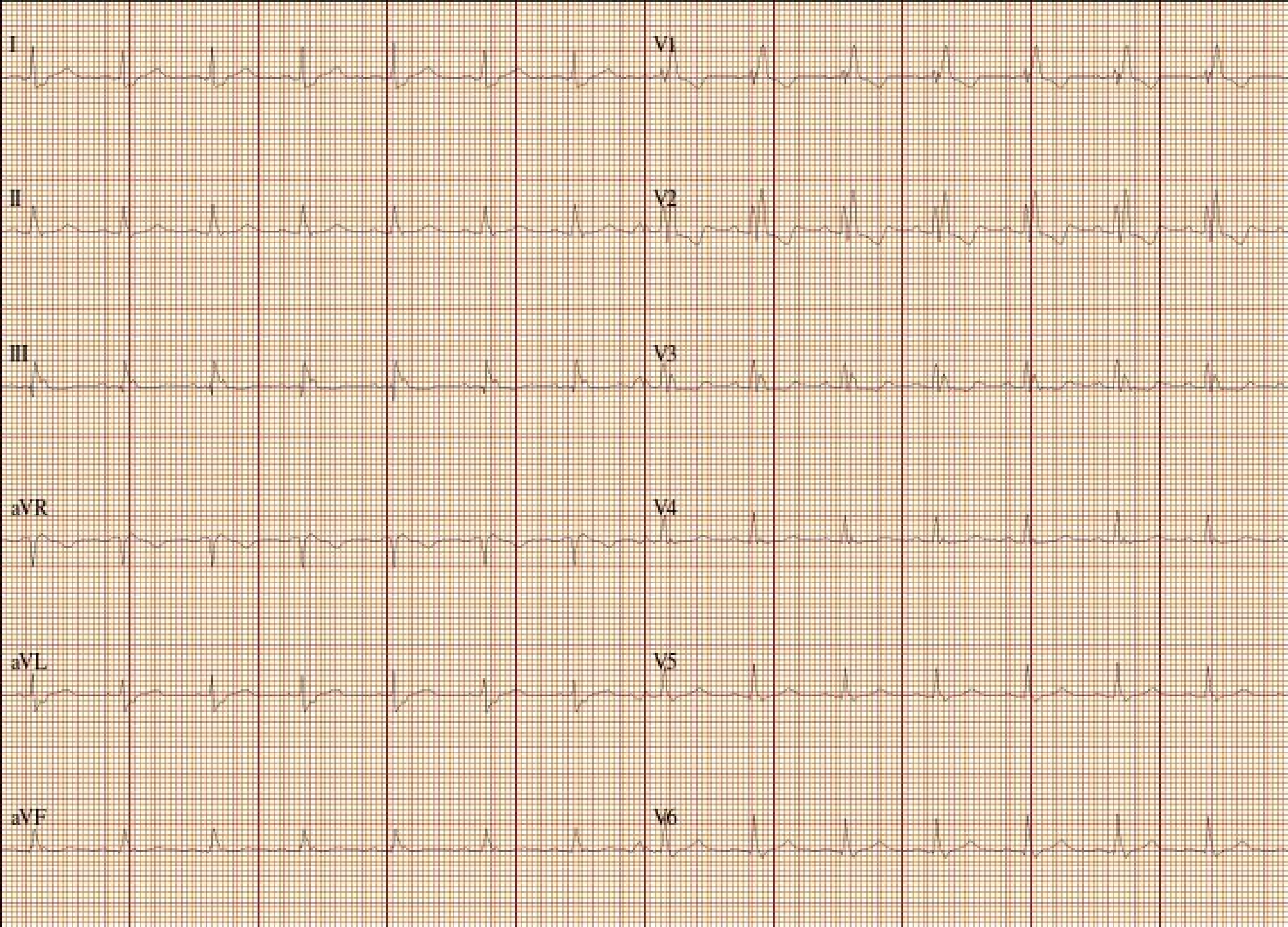
I
II
III
aVR
aVL
aVF
V1
V2
V3
V4
V5
V6

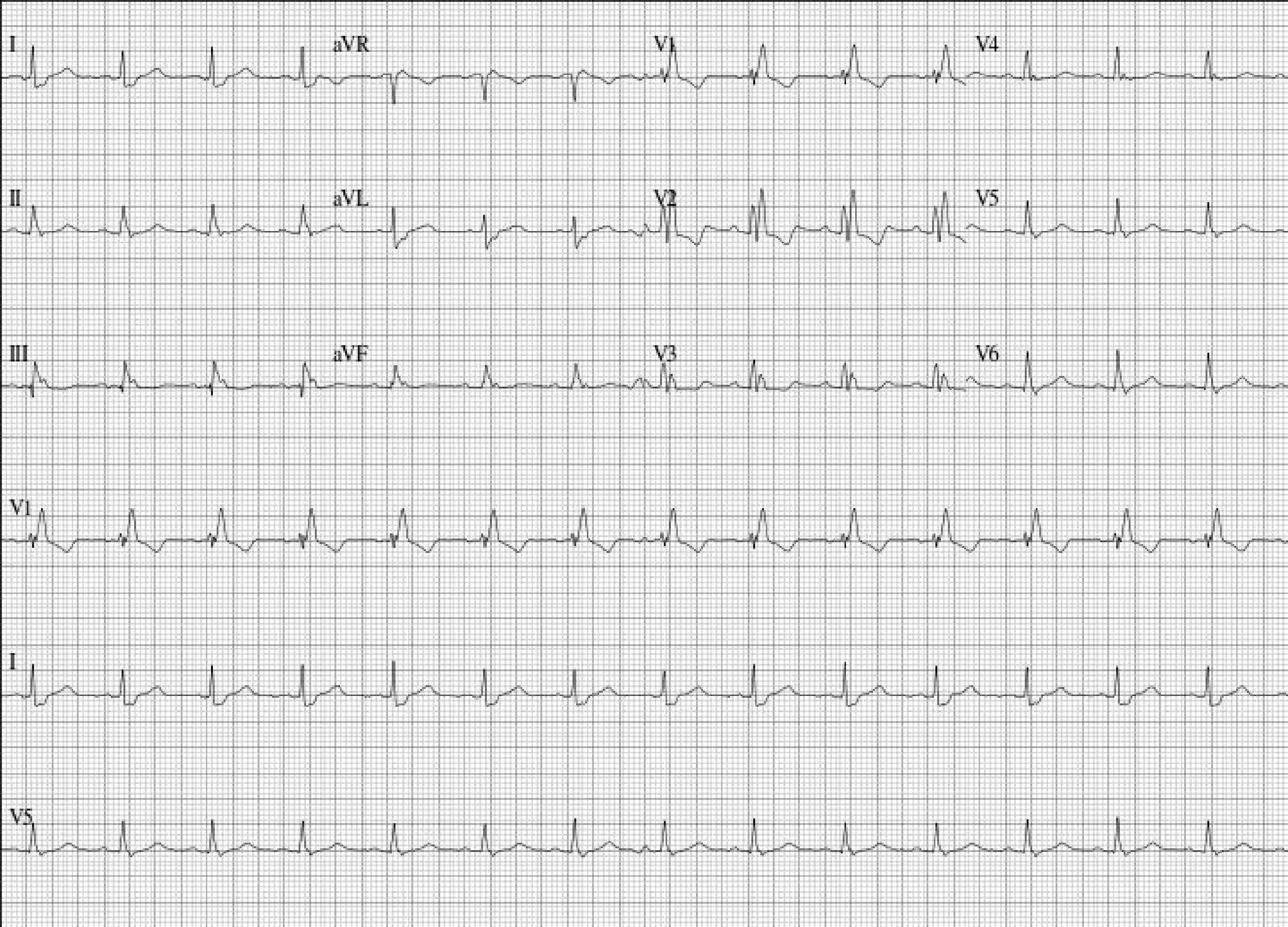
I
aVR
V1
V4
II
aVL
V2
V5
III
aVF
V3
V6
V1
I
V5

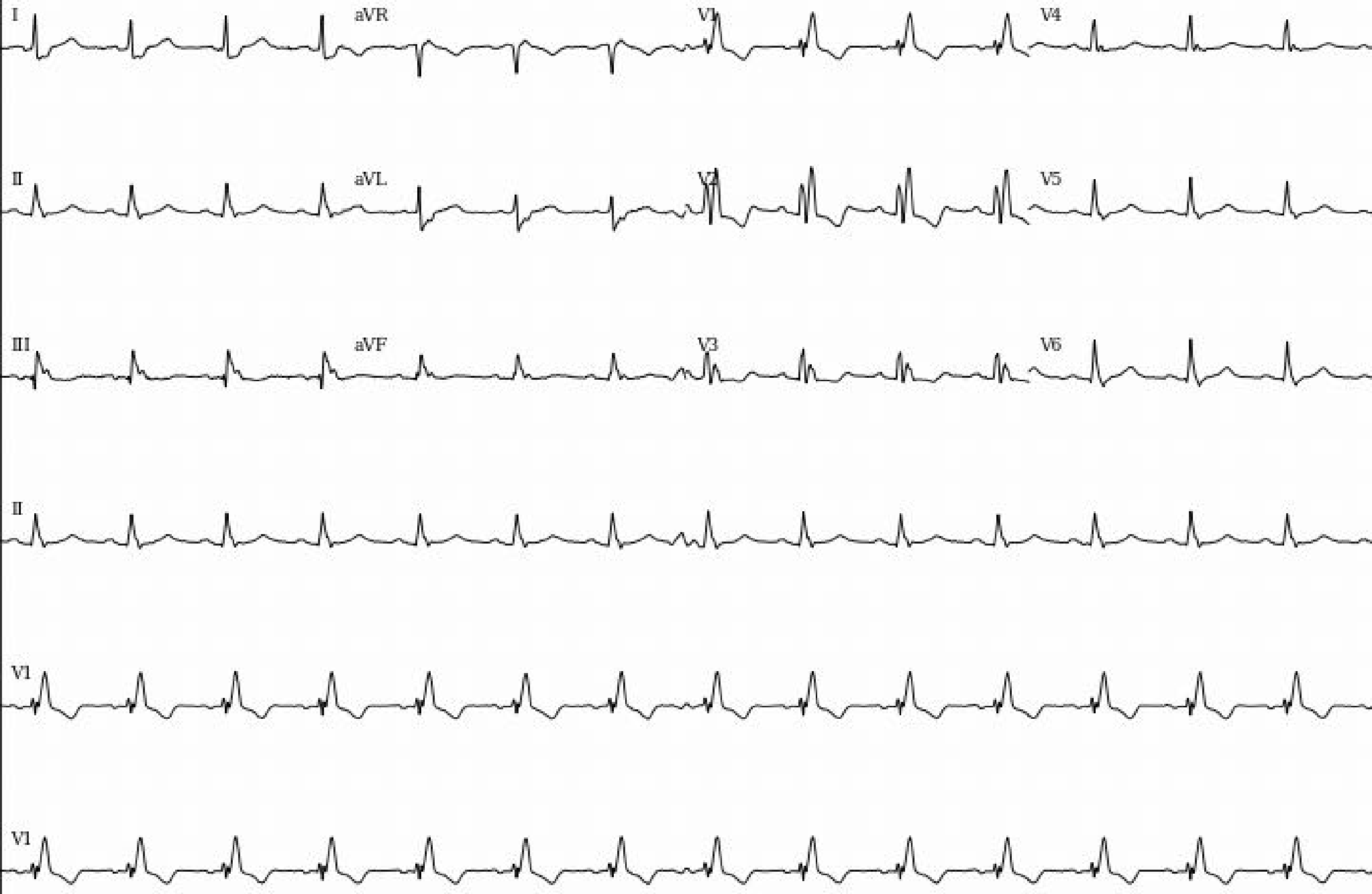
I
aVR
V1
V4
II
aVL
V2
V5
III
aVF
V3
V6
II
V1
V1

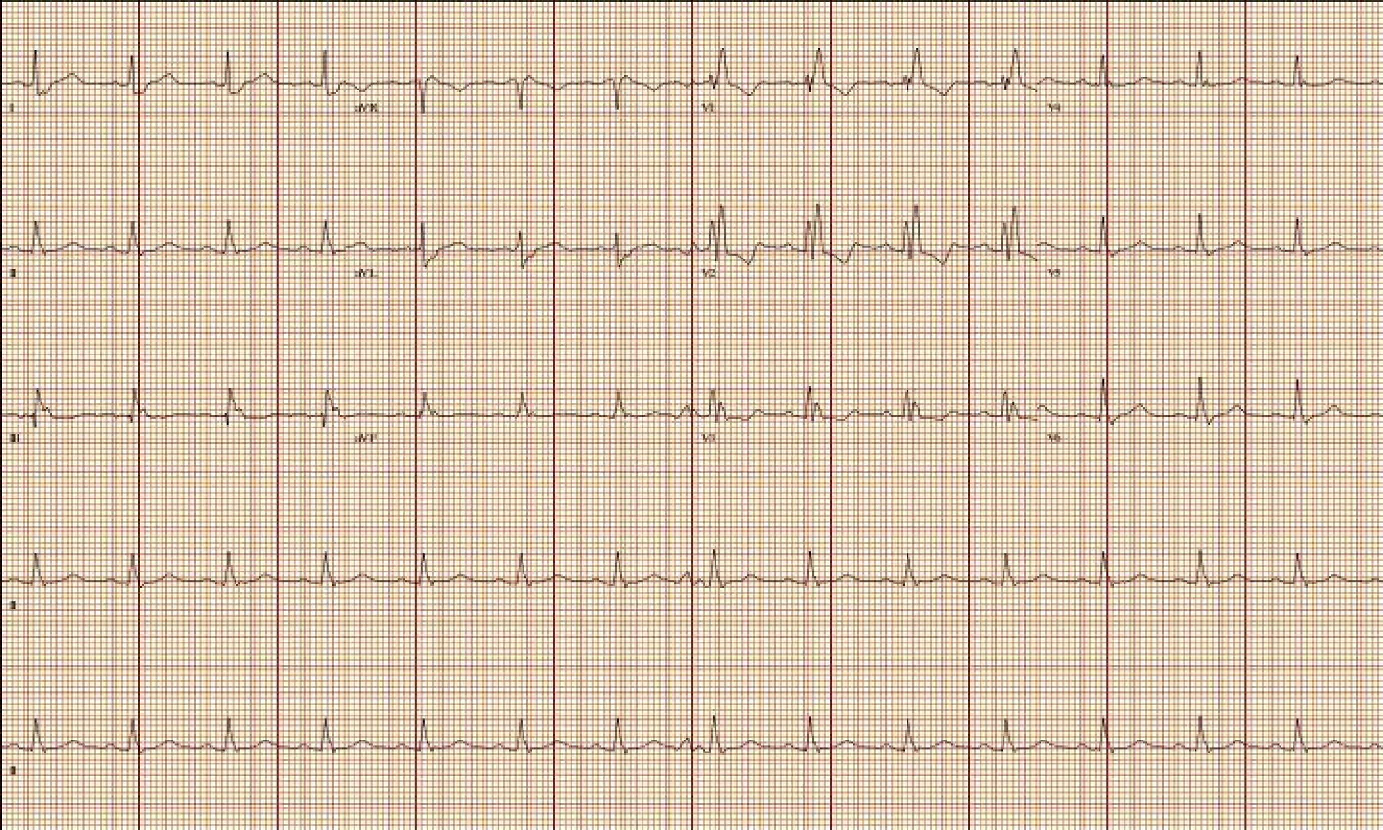
I
aVR
V1
V4
II
aVL
V2
V5
III
aVF
V3
V6
II
II

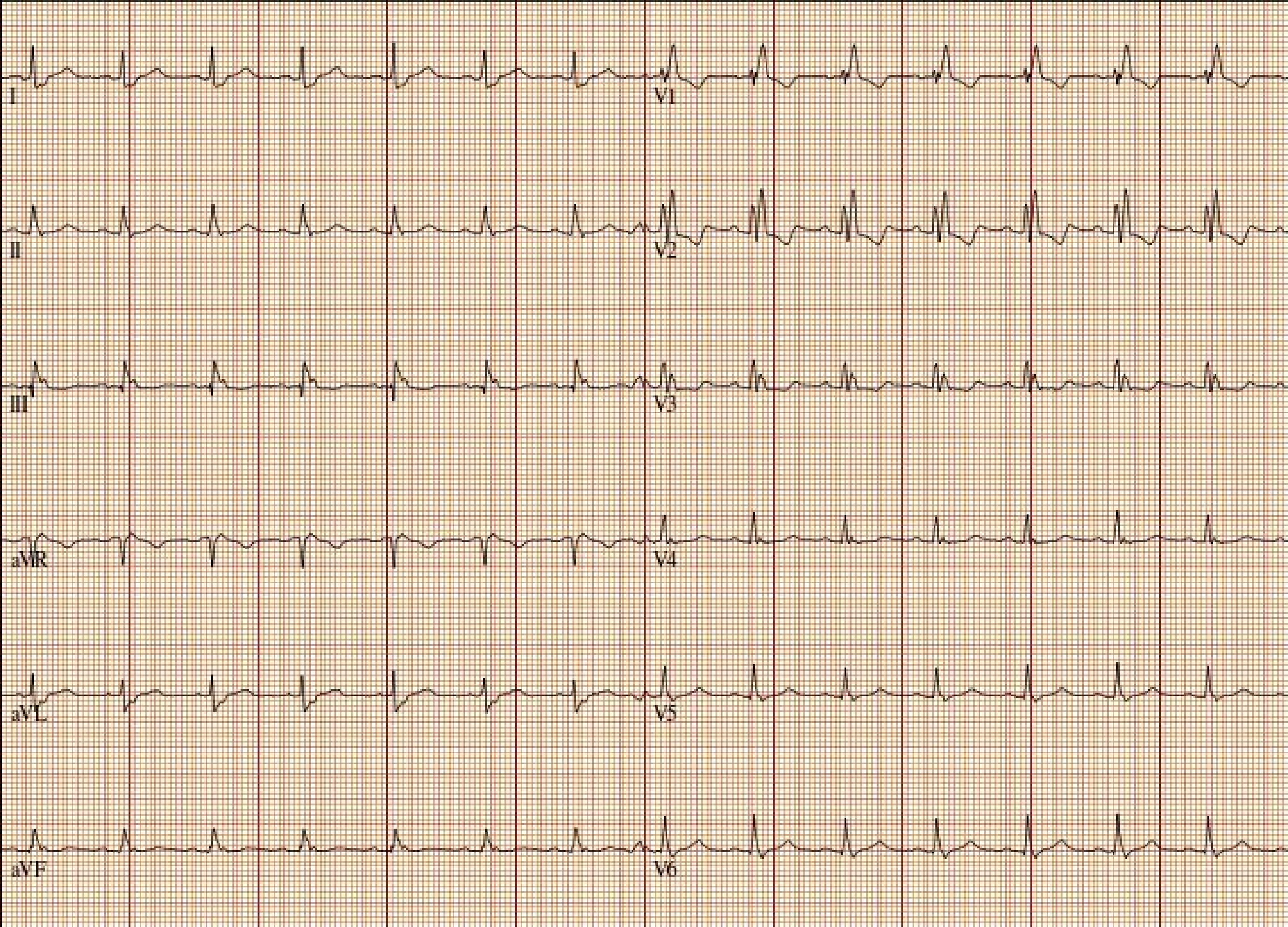
I
II
III
aVR
aVL
aVF
V1
V2
V3
V4
V5
V6

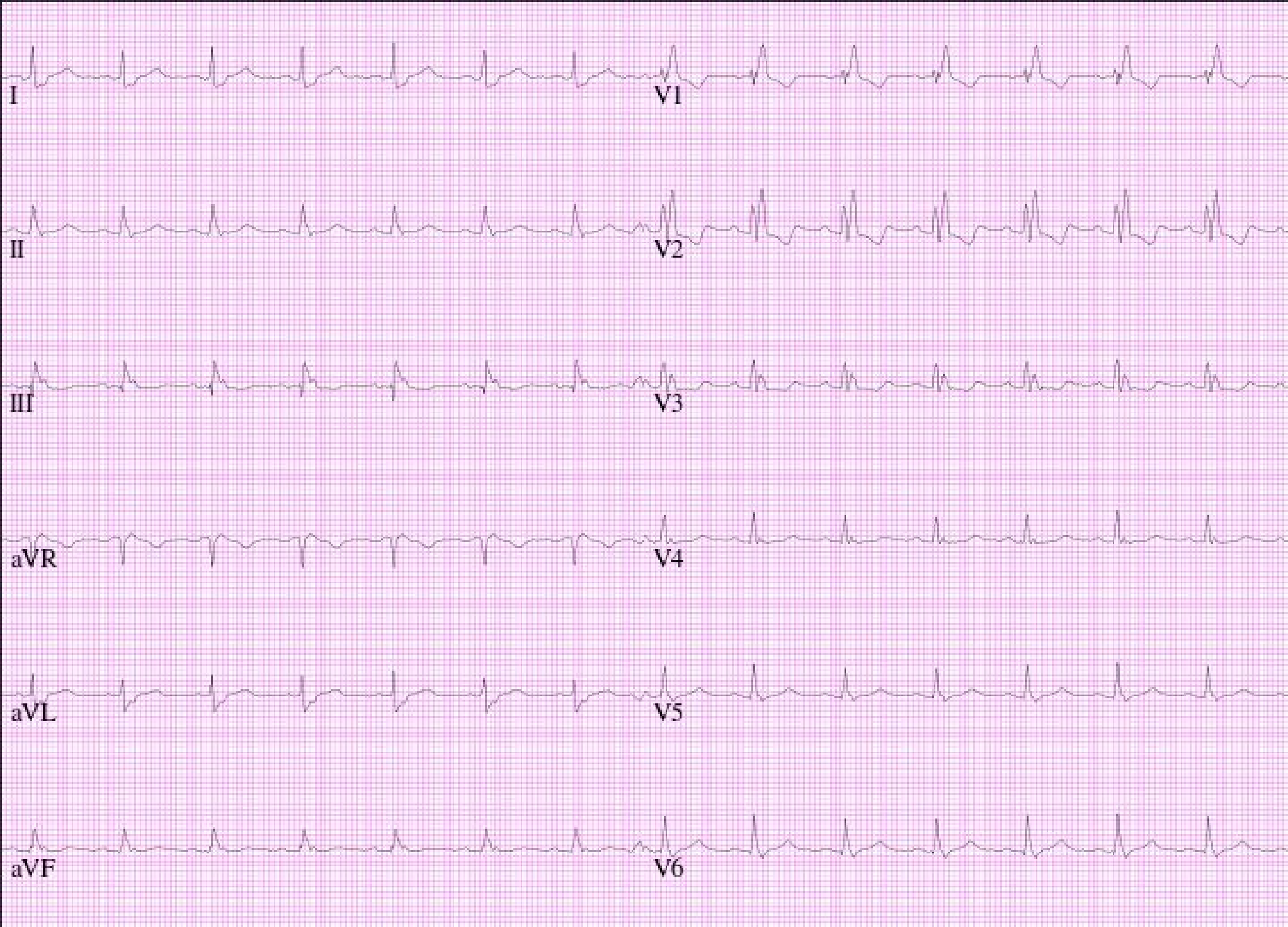
I
II
III
aVR
aVL
aVF
V1
V2
V3
V4
V5
V6

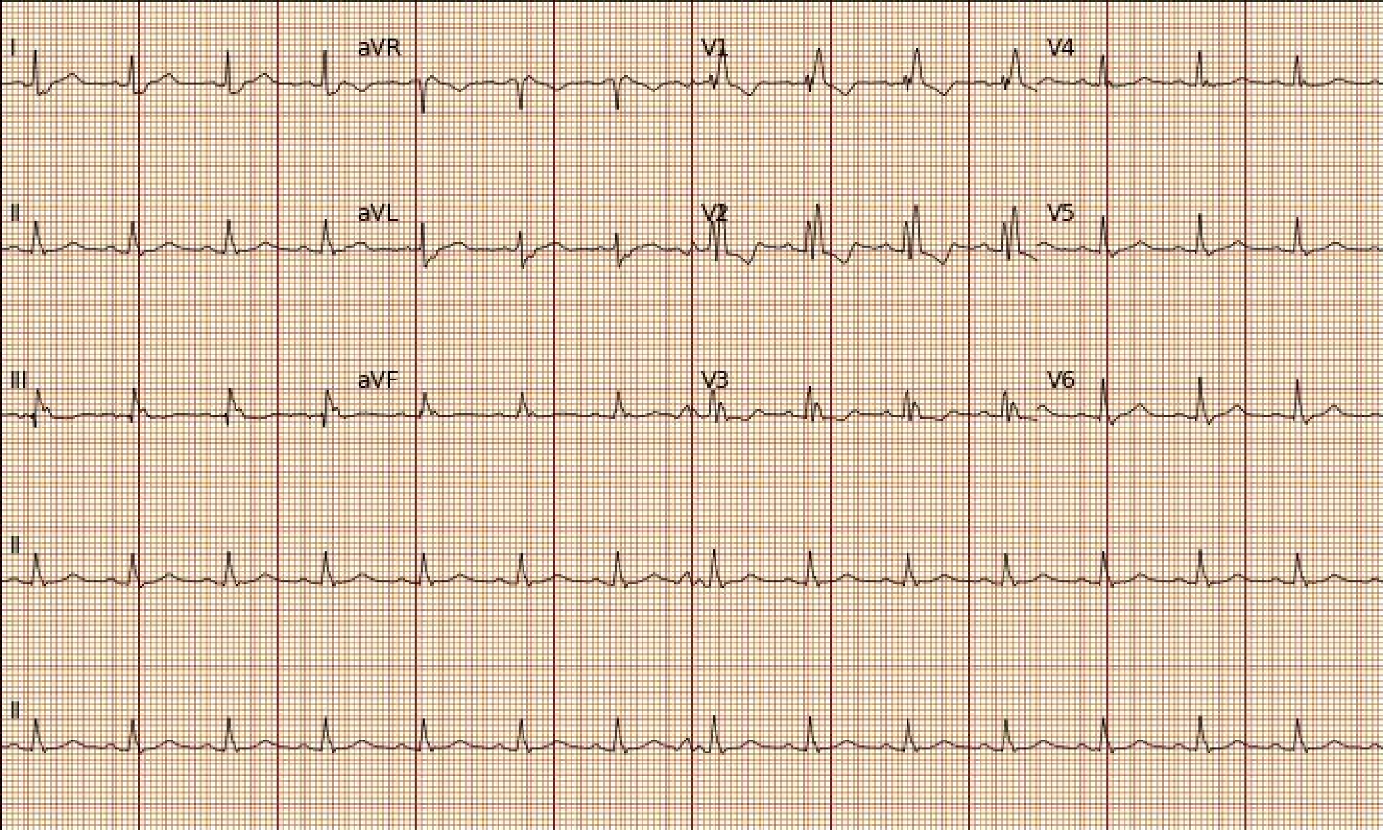
I
aVR
V1
V4
II
aVL
V2
V5
III
aVF
V3
V6
II
II

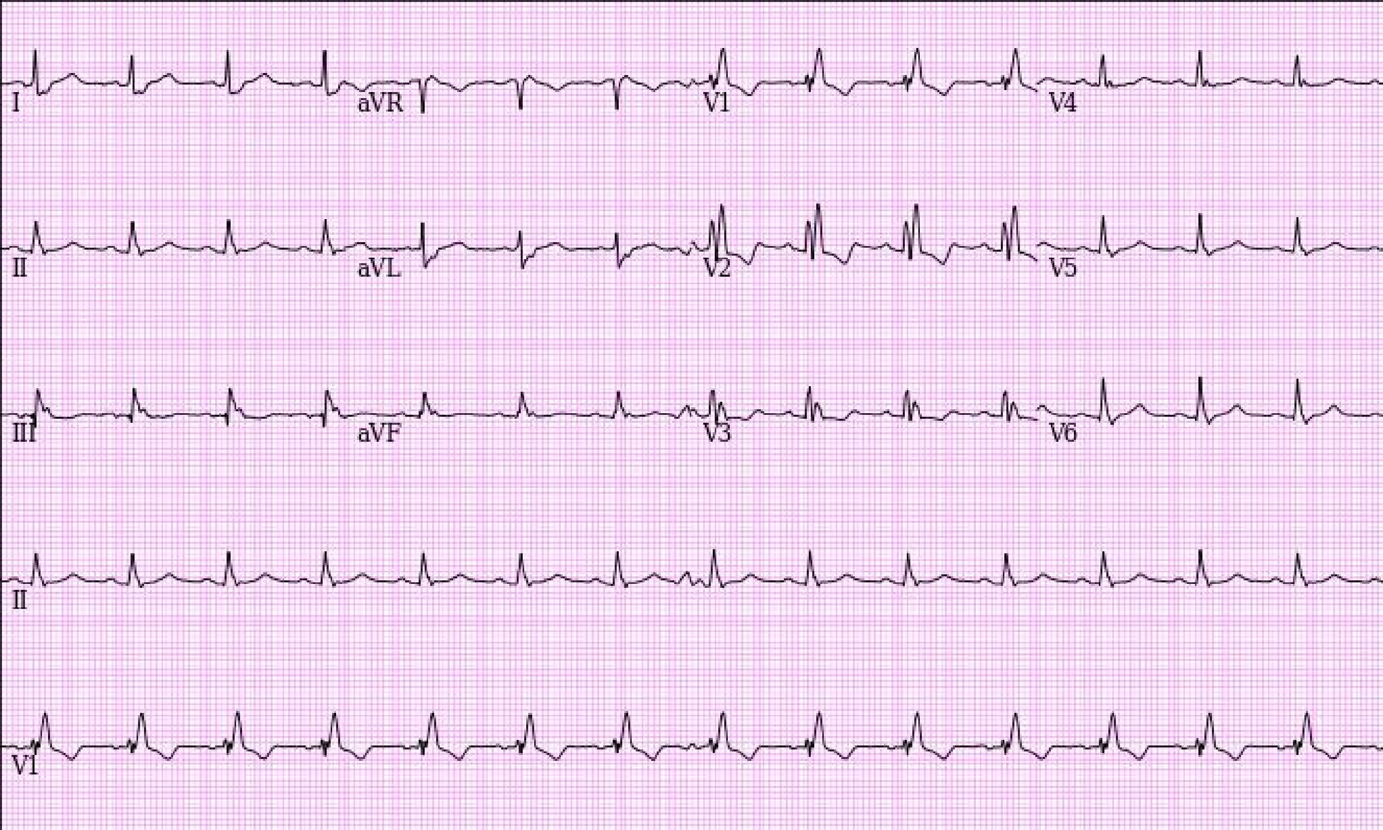
I
aVR
V1
V4
II
aVL
V2
V5
III
aVF
V3
V6
II
V1

### Section 2. Random examples of 25 electrocardiogram images plotted in standard clinical layout used during model evaluation

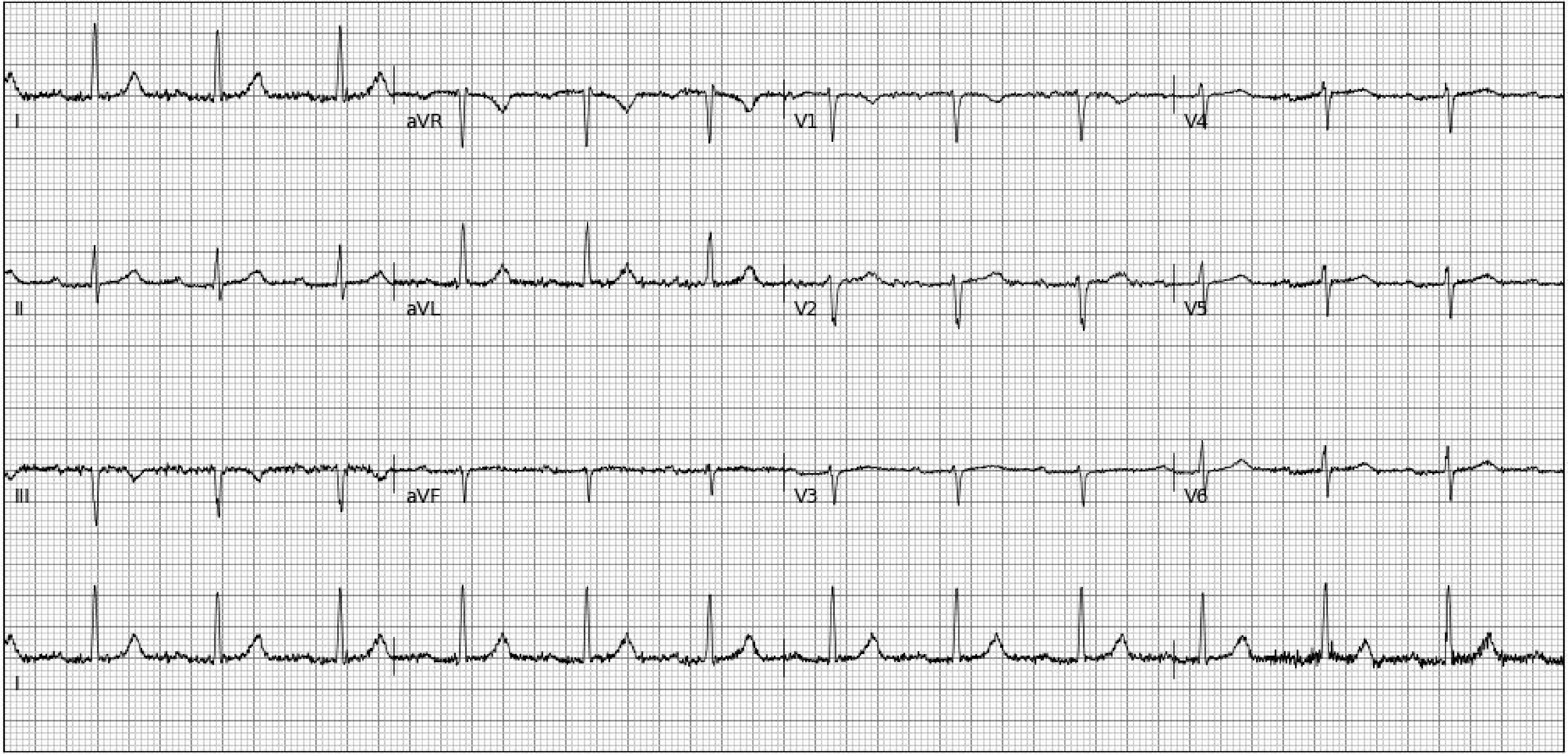
I
aVR
V1
V4
II
aVL
V2
V5
III
aVF
V3
V6
I

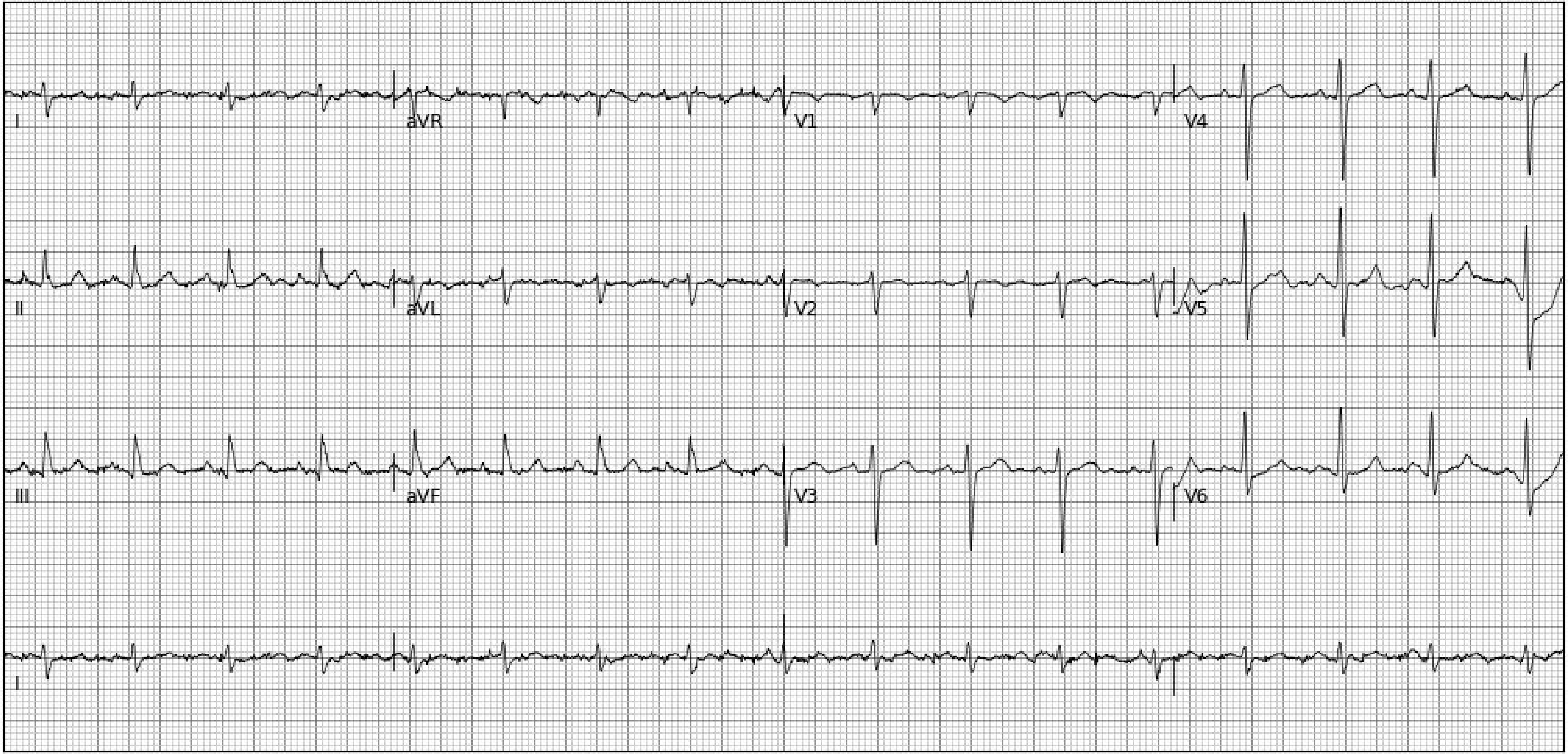
I
aVR
V1
V4
II
aVL
V2
V5
III
aVF
V3
V6
I

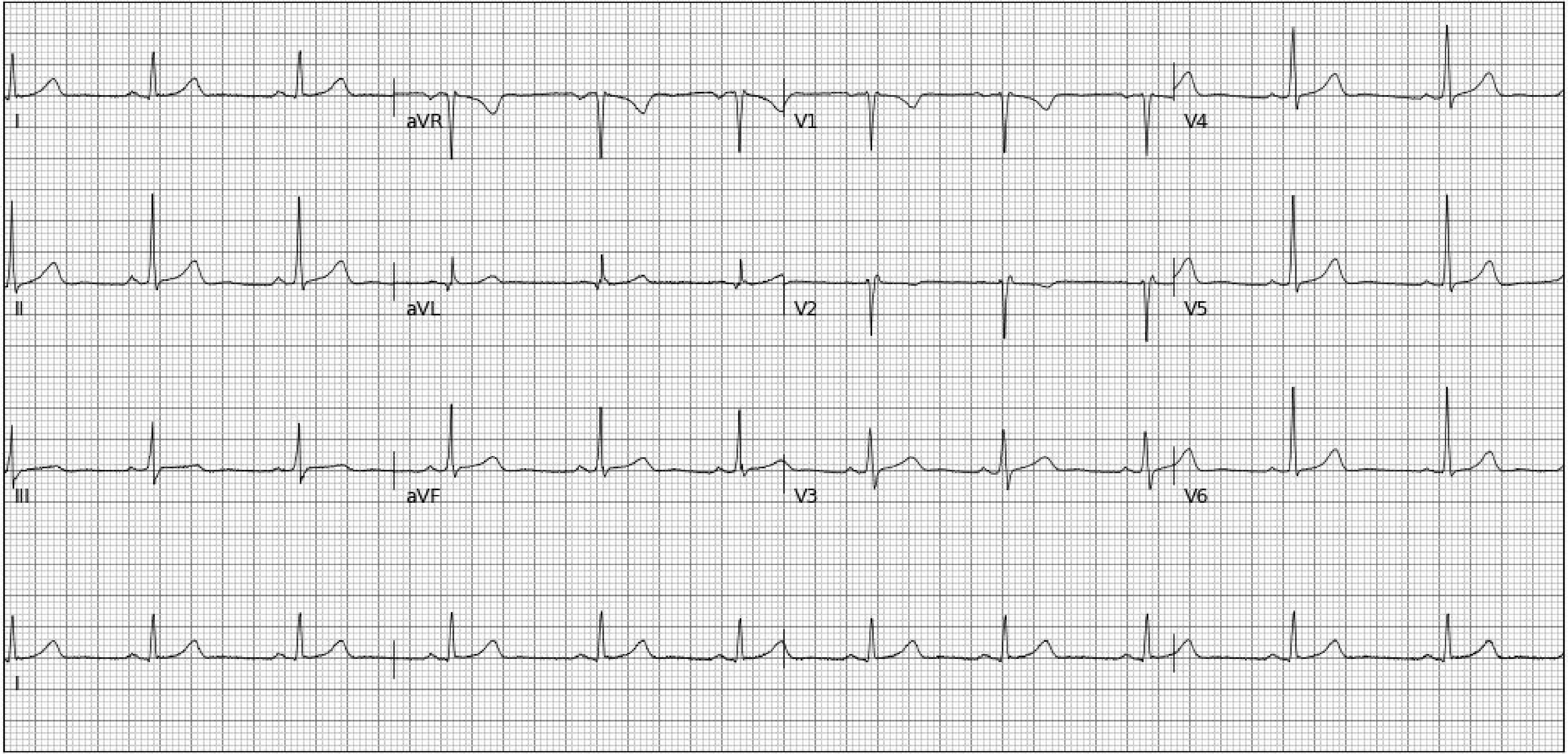

I
aVR
V1
V4
II
aVL
V2
V5
III
aVF
V3
V6
I

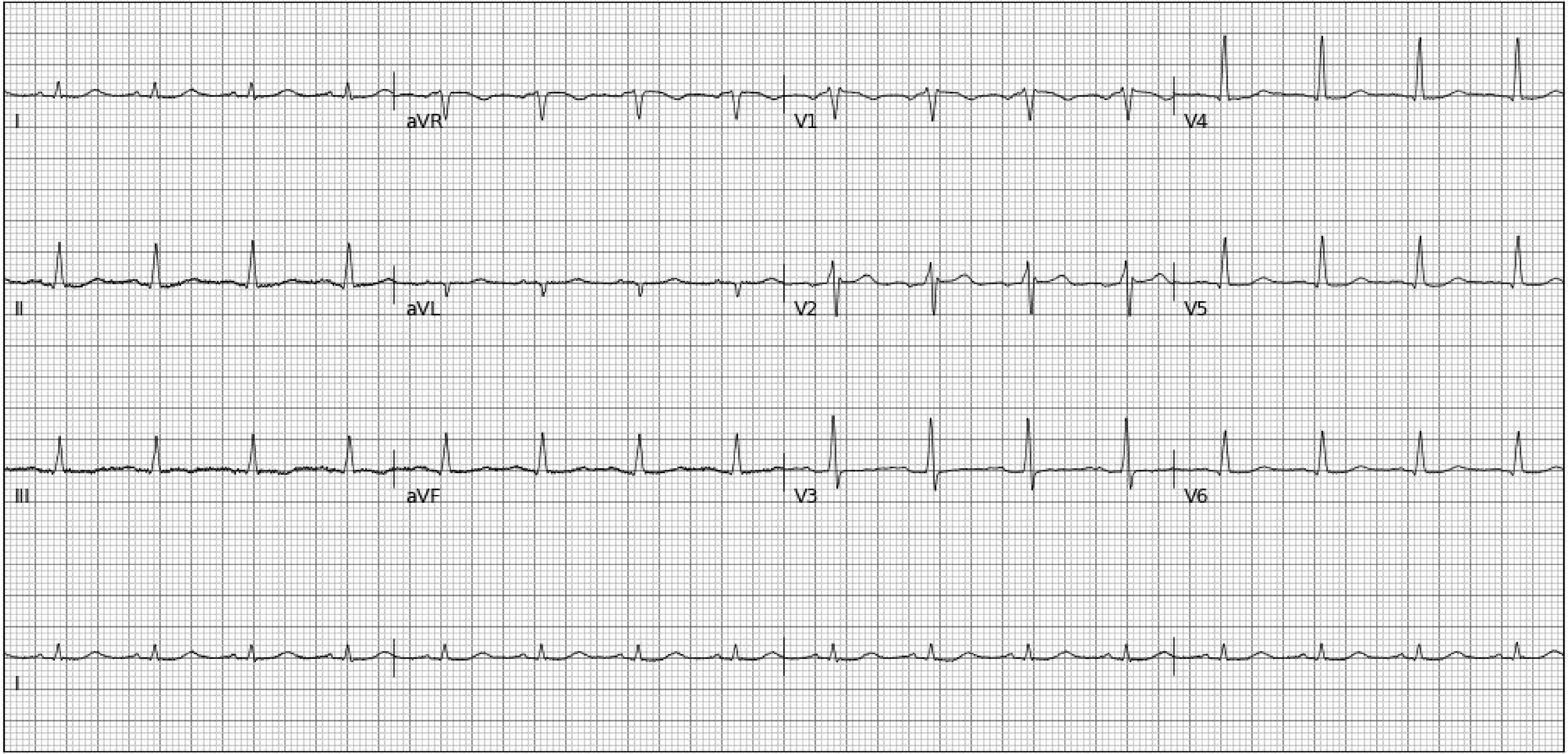
I
aVR
V1
V4
II
aVL
V2
V5
III
aVF
V3
V6
I

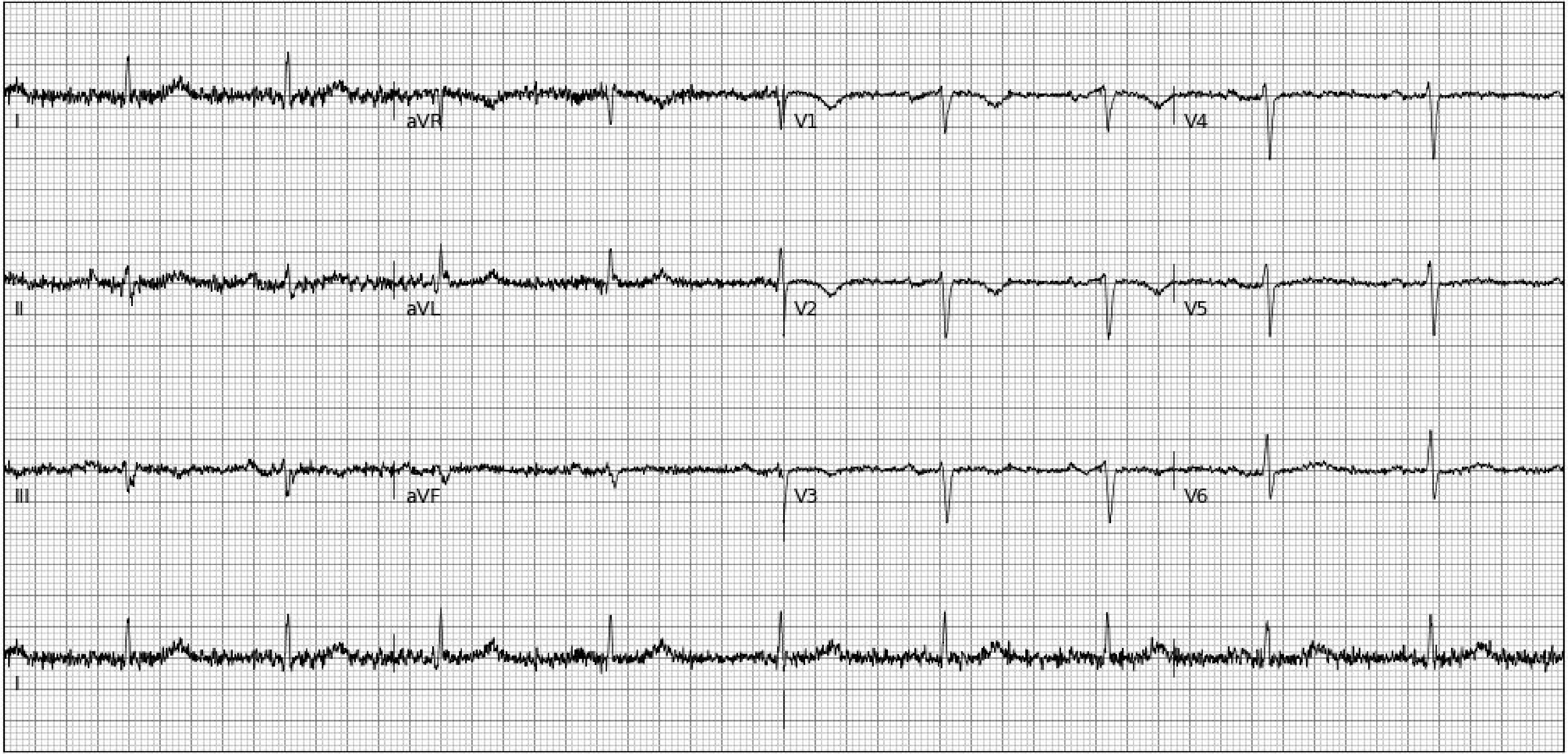
I
aVR
V1
V4
II
aVL
V2
V5
III
aVF
V3
V6
I

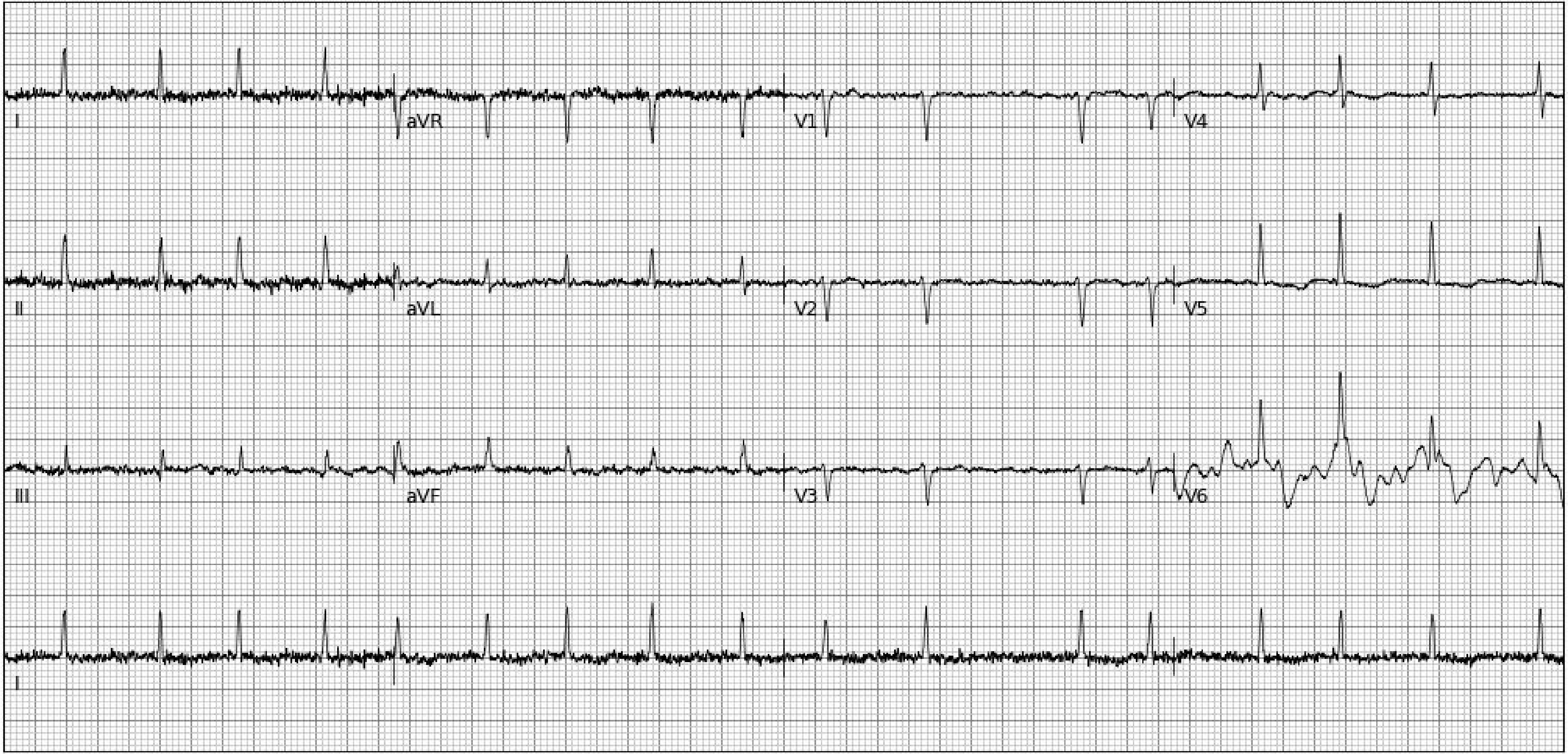
I
aVR
V1
V4
II
aVL
V2
V5
III
aVF
V3
V6
I

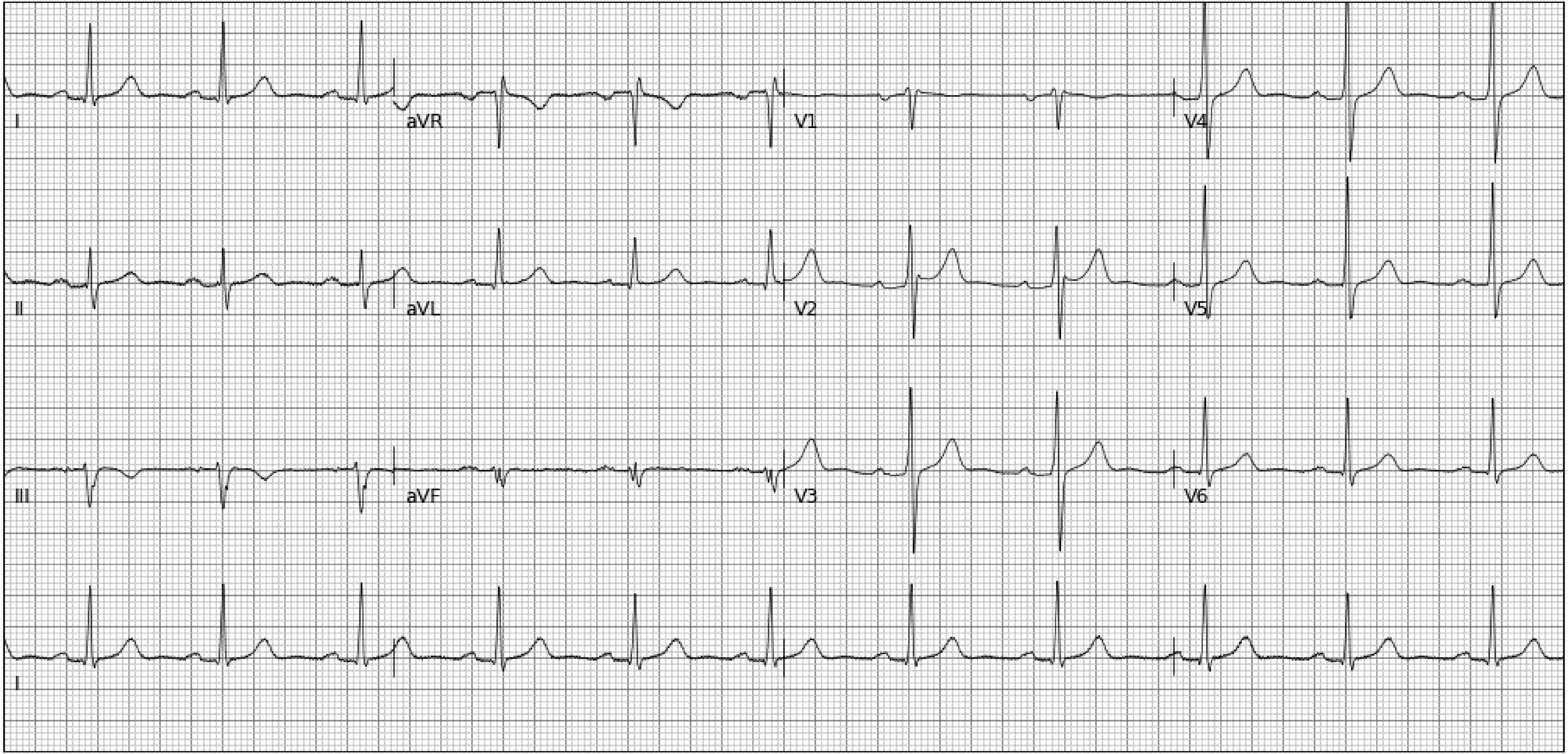

I
aVR
V1
V4
II
aVL
V2
V5
III
aVF
V3
V6
I

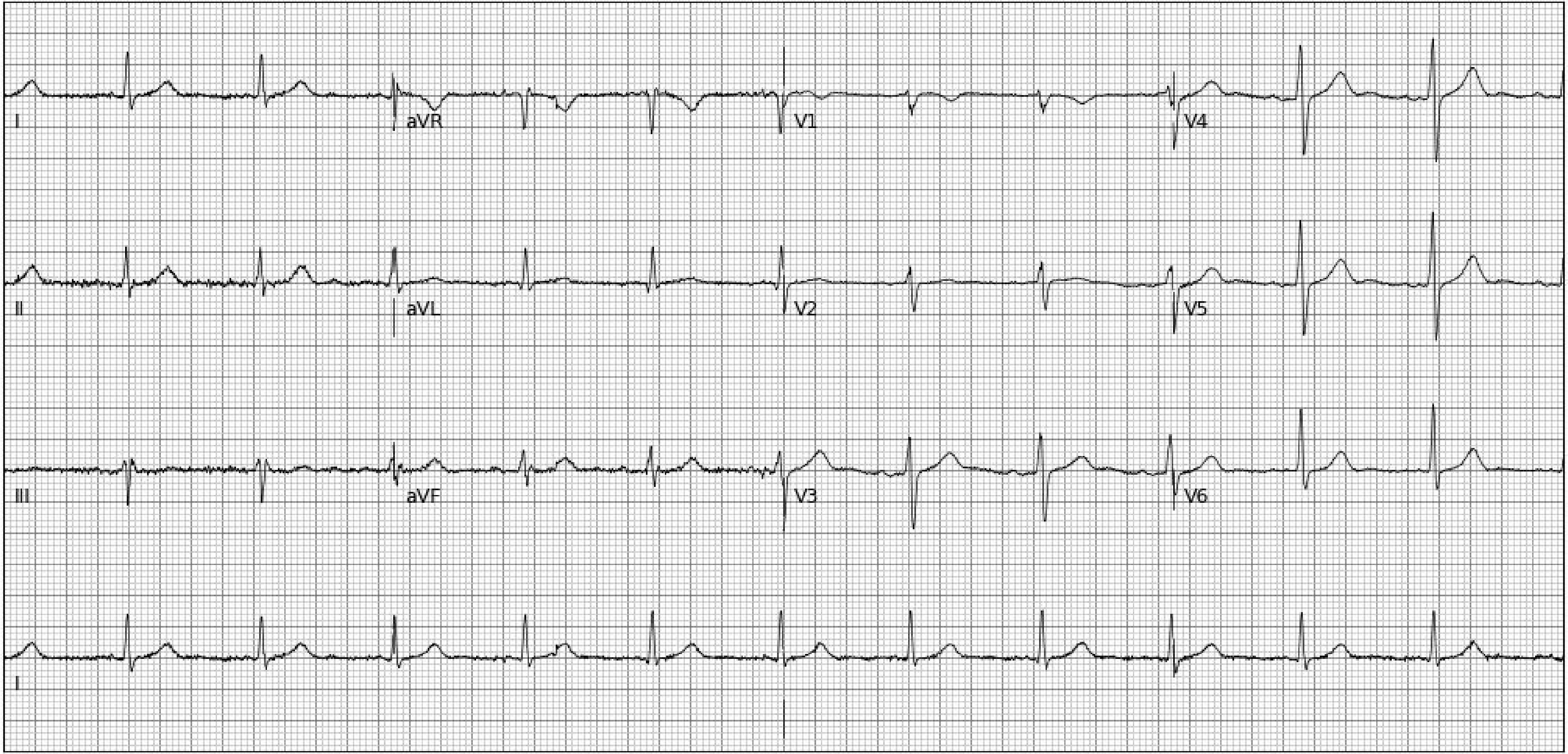
I
aVR
V1
V4
II
aVL
V2
V5
III
aVF
V3
V6
I

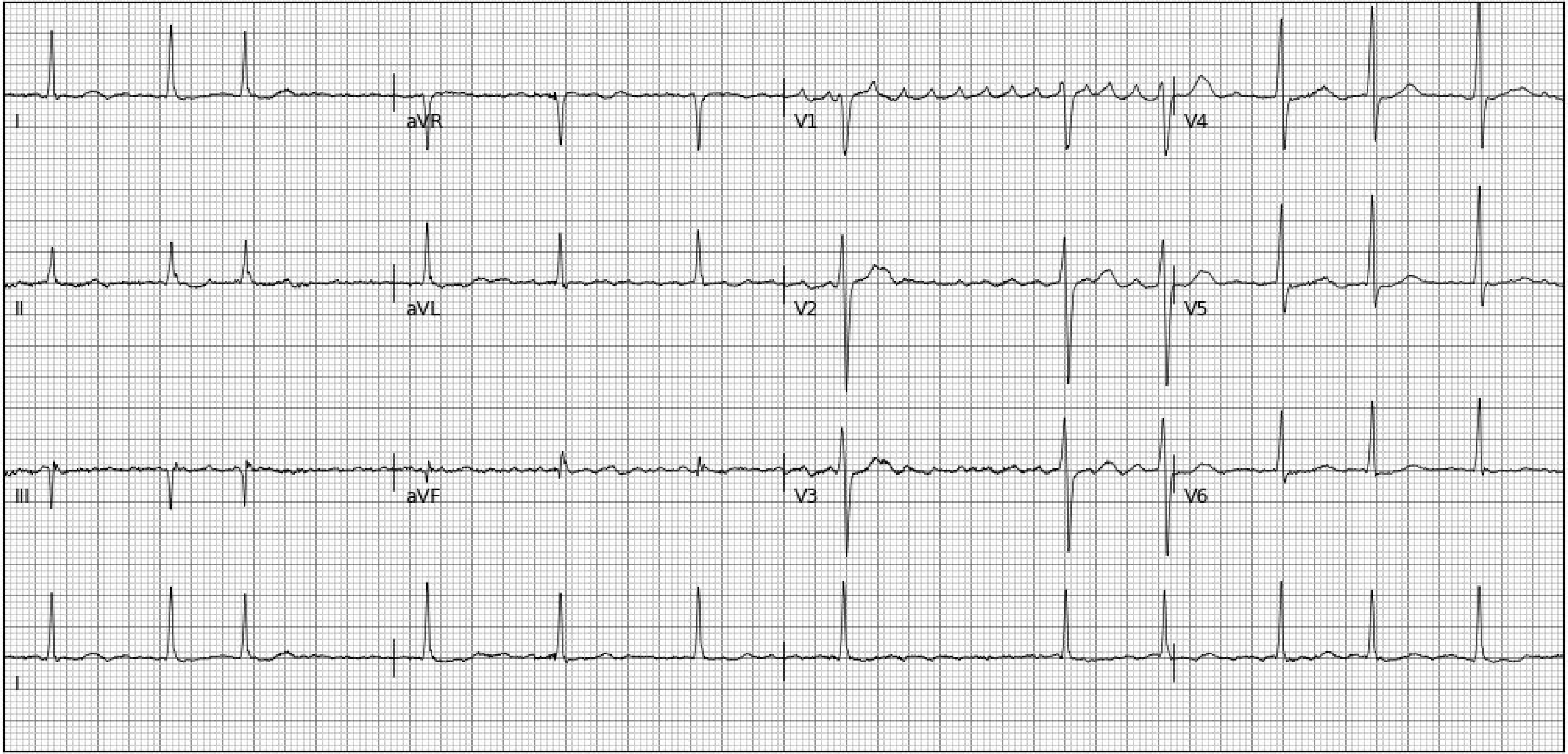
I
aVR
V1
V4
II
aVL
V2
V5
III
aVF
V3
V6
I

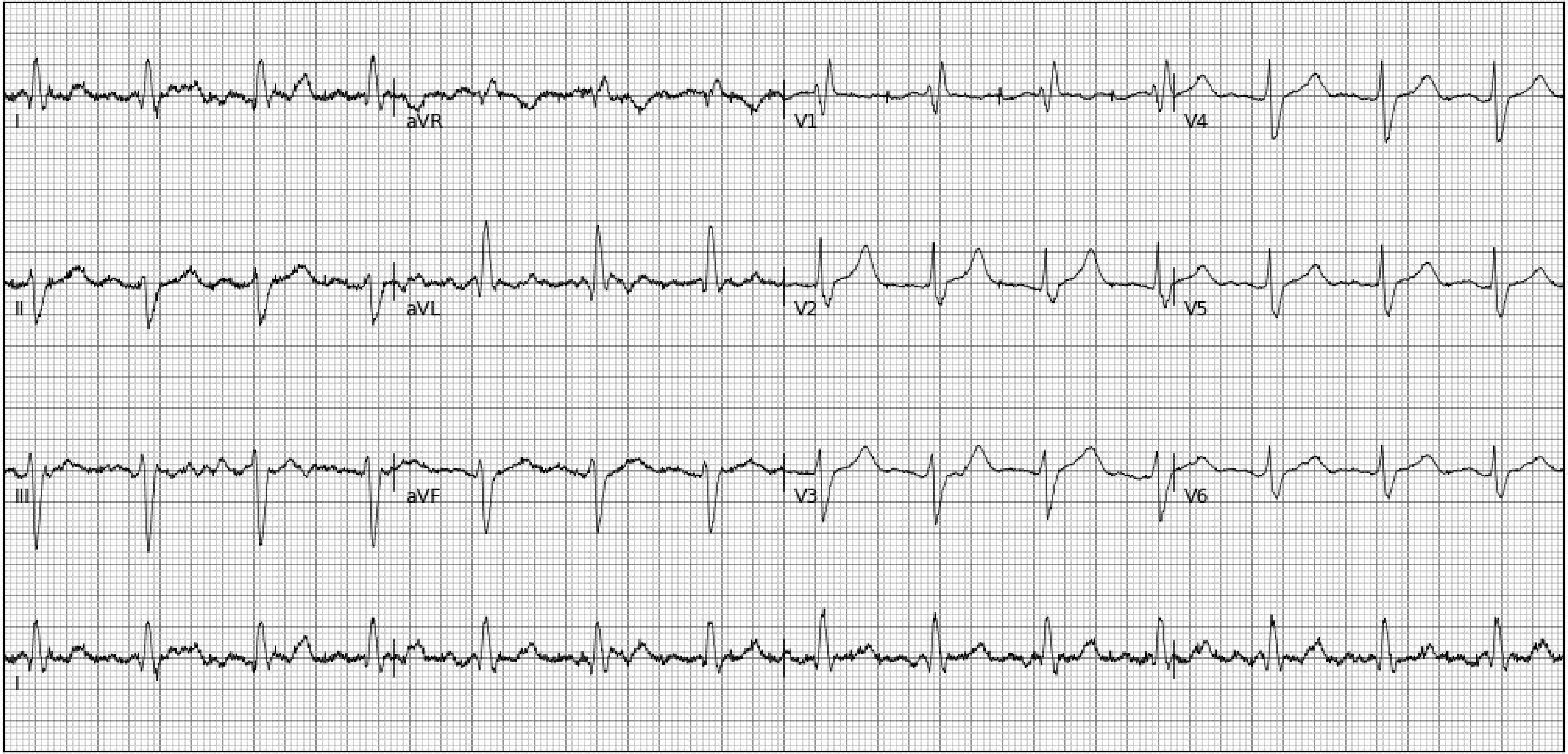
I
aVR
V1
V4
II
aVL
V2
V5
III
aVF
V3
V6
I

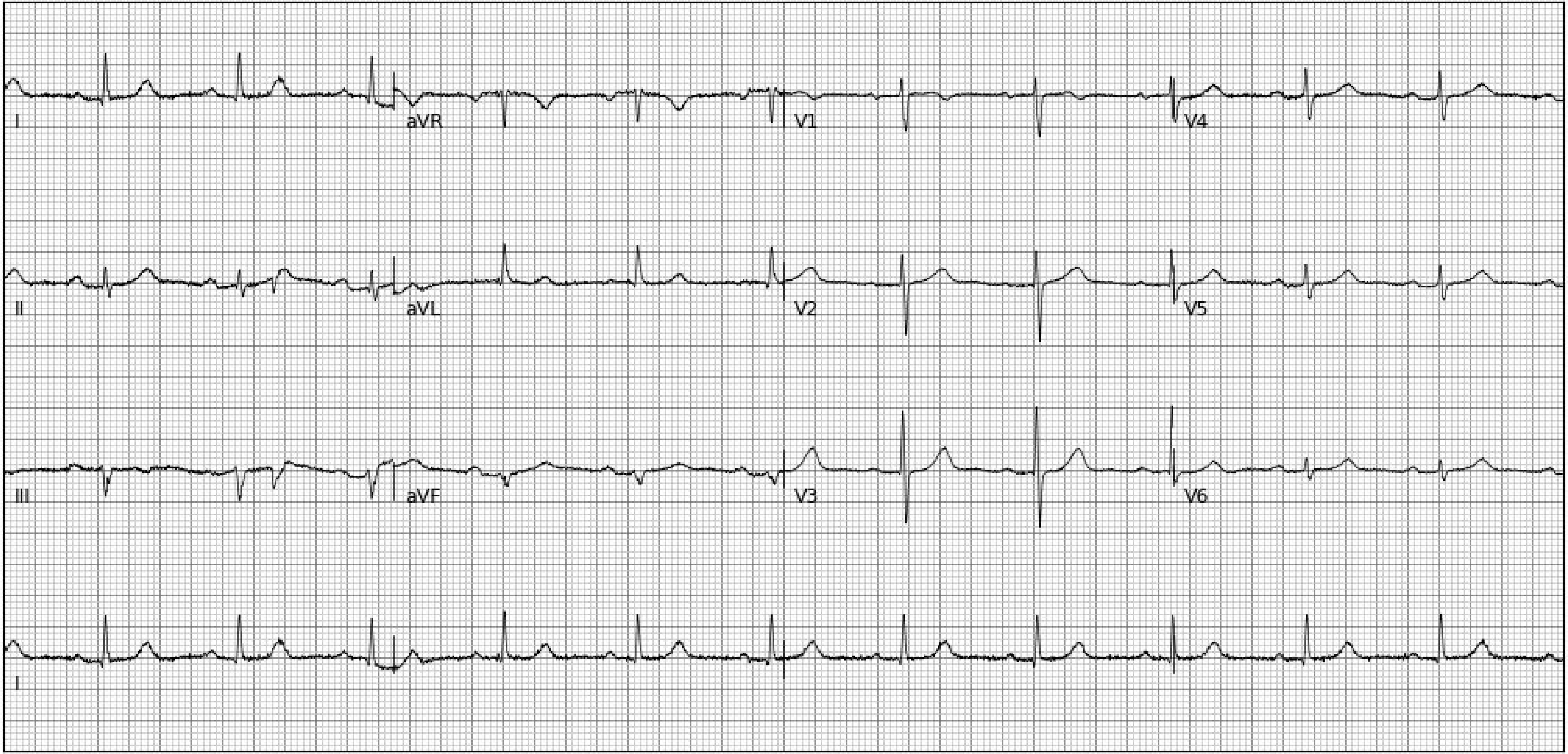
I
aVR
V1
V4
II
aVL
V2
V5
III
aVF
V3
V6
I

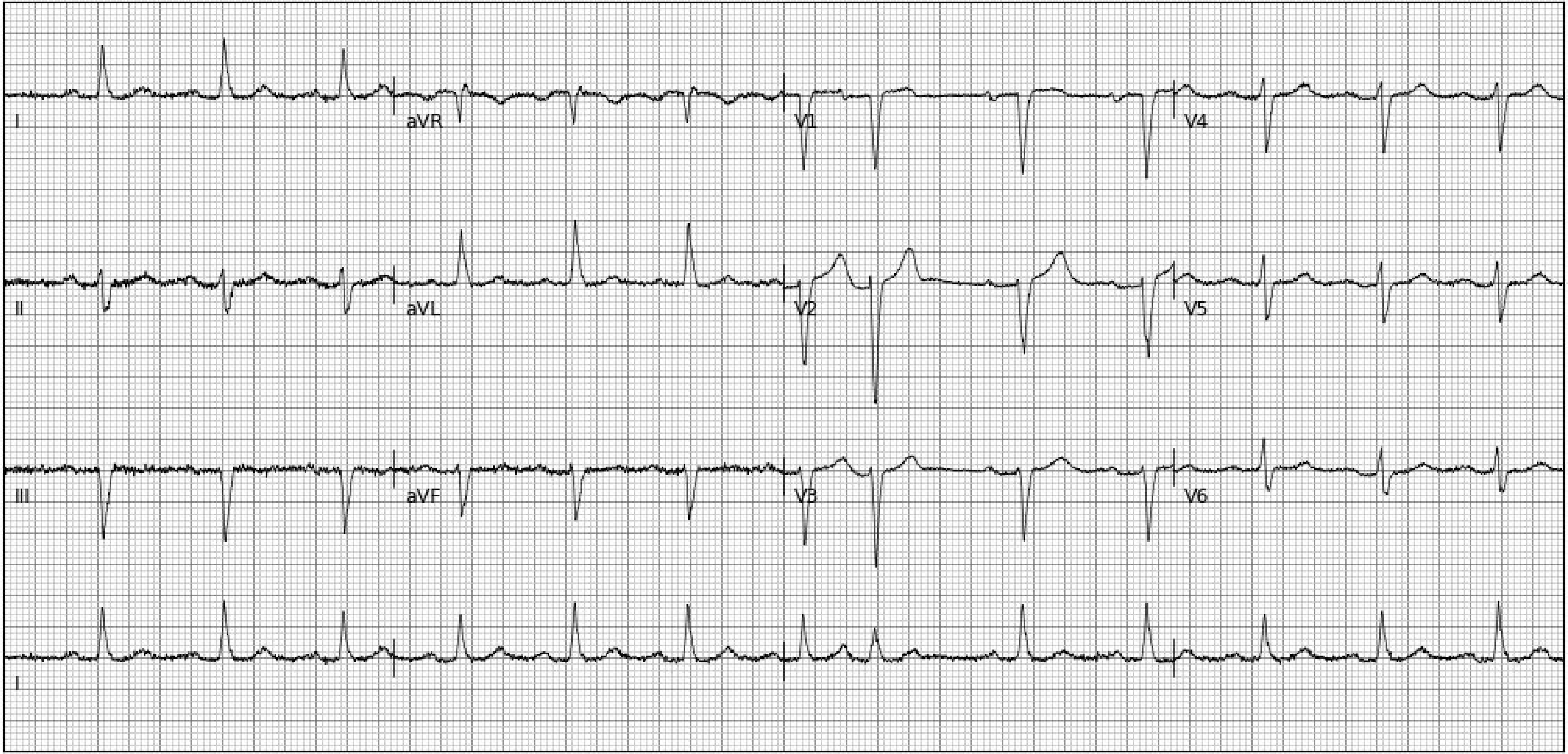
I
aVR
V1
V4
II
aVL
V2
V5
III
aVF
V3
V6
I

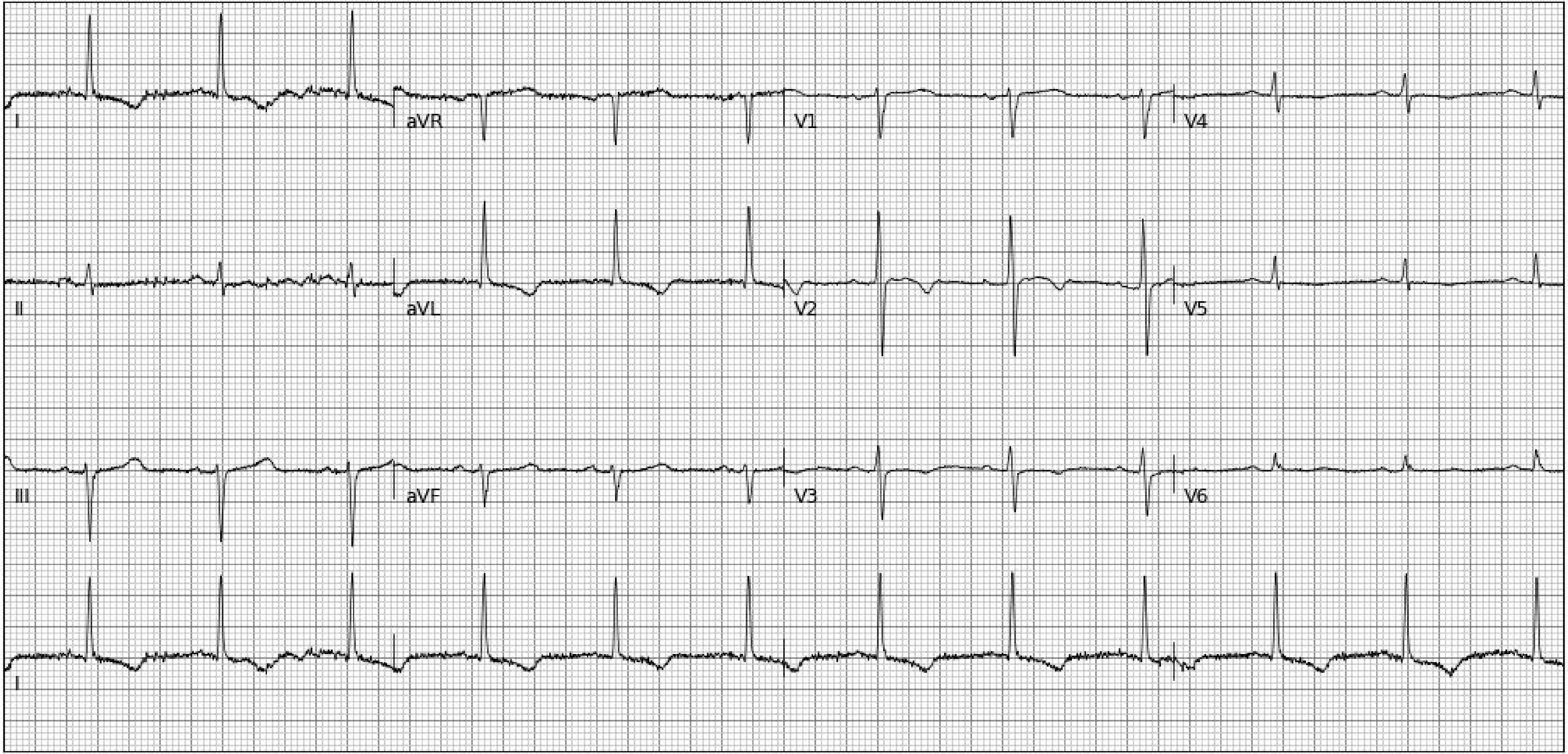
I
aVR
V1
V4
II
aVL
V2
V5
III
aVF
V3
V6
I

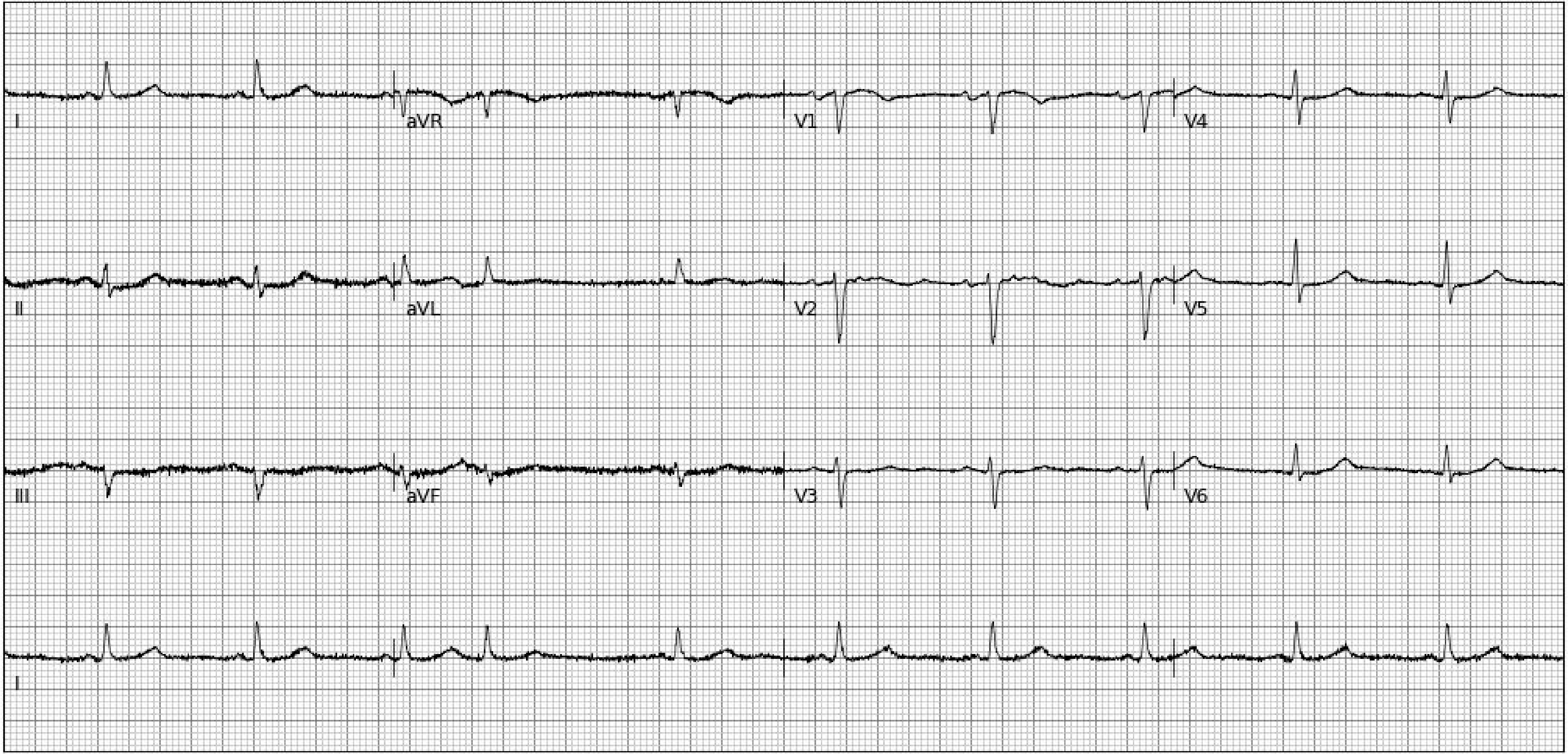
I
aVR
V1
V4
II
aVL
V2
V5
III
aVF
V3
V6
I

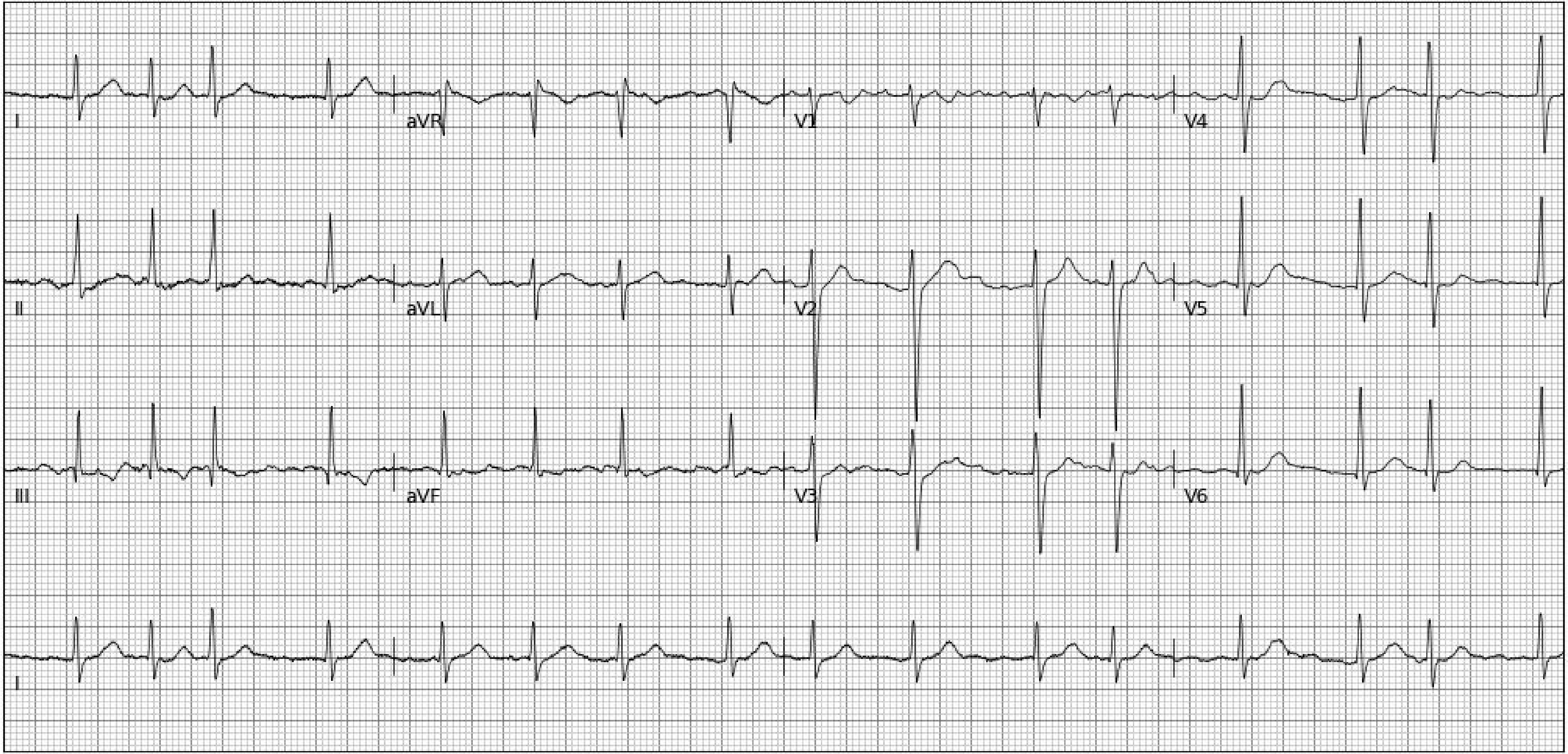
I
aVR
V1
V4
II
aVL
V2
V5
III
aVF
V3
V6
I

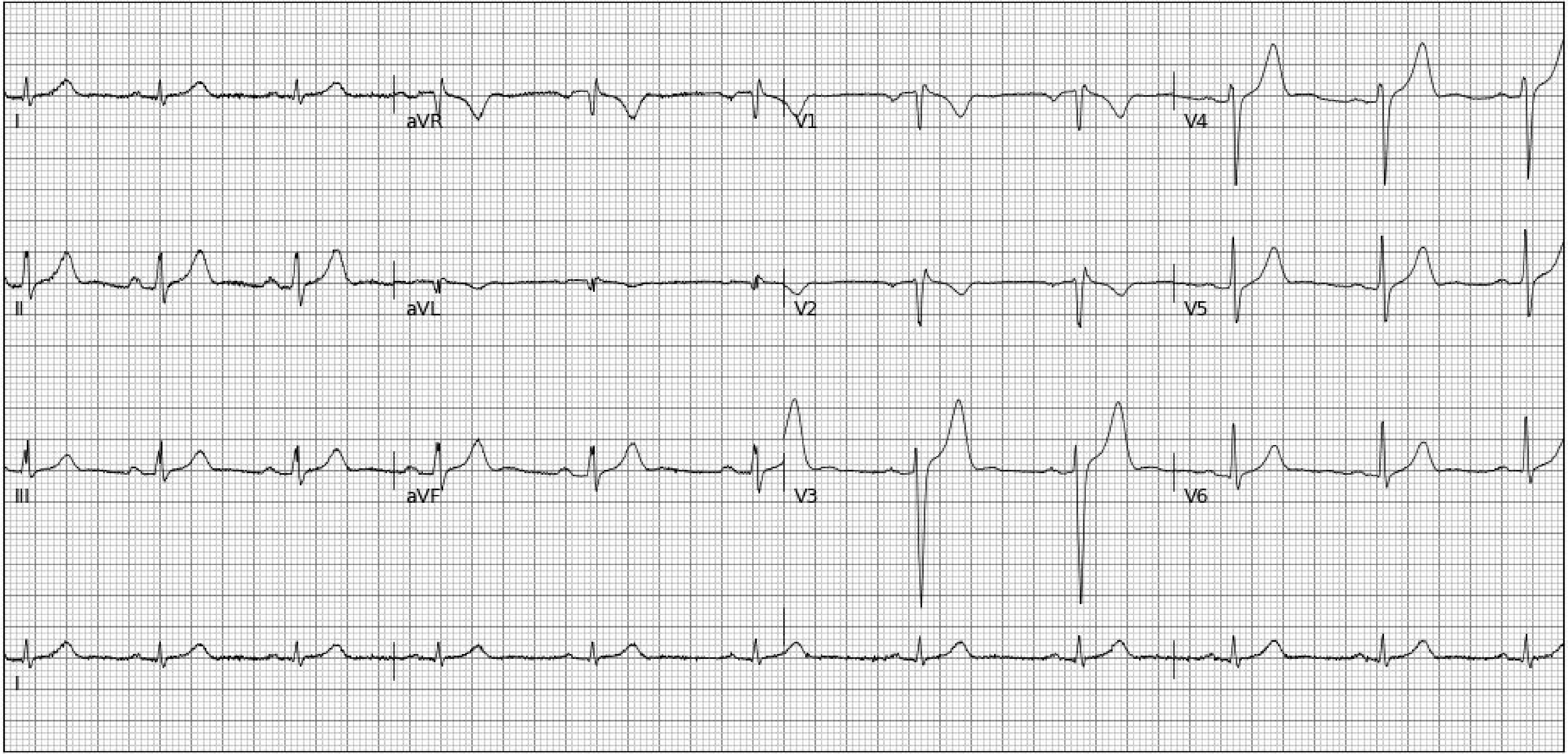
I
aVR
V1
V4
II
aVL
V2
V5
III
aVF
V3
V6
II

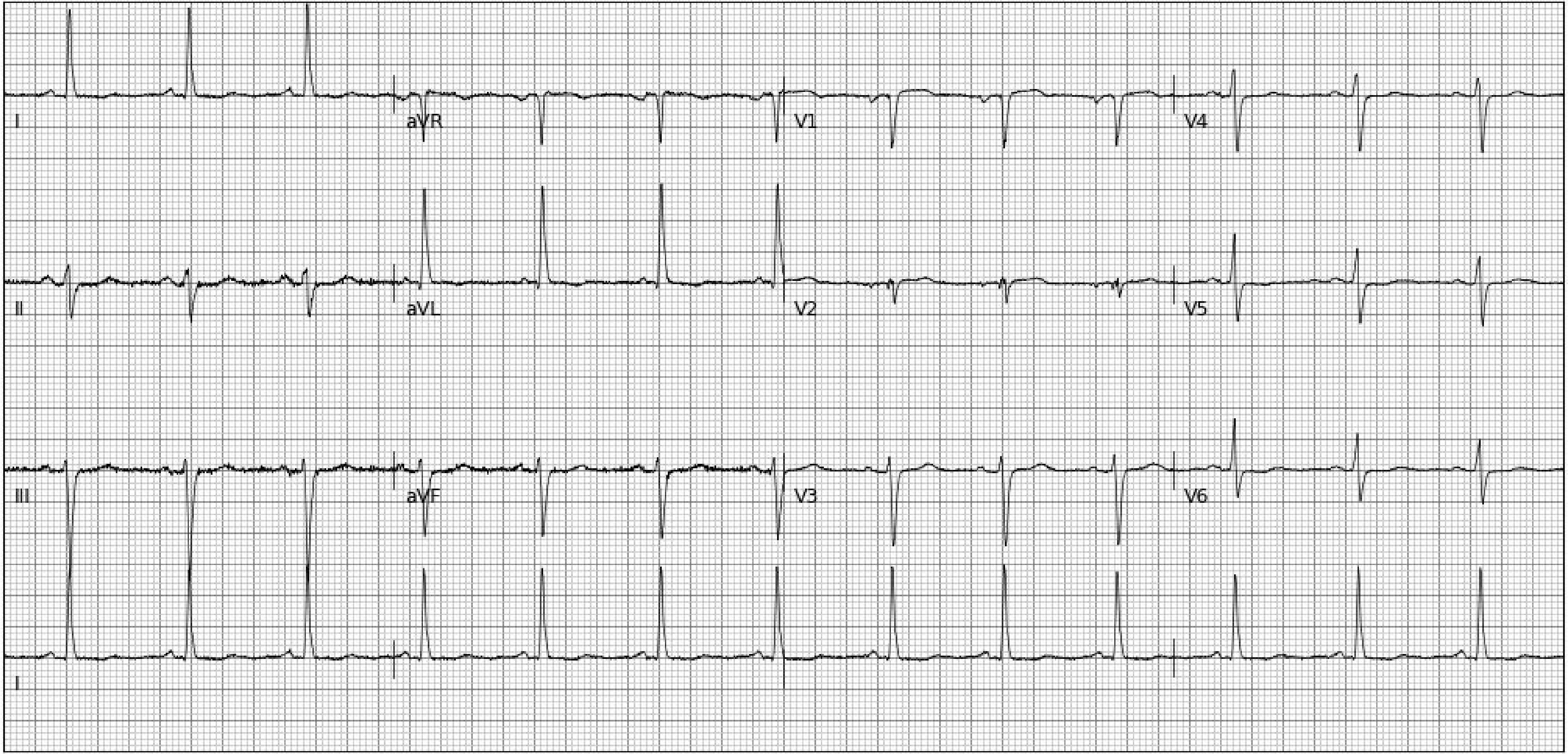
I
aVR
V1
V4
II
aVL
V2
V5
III
aVF
V3
V6
I

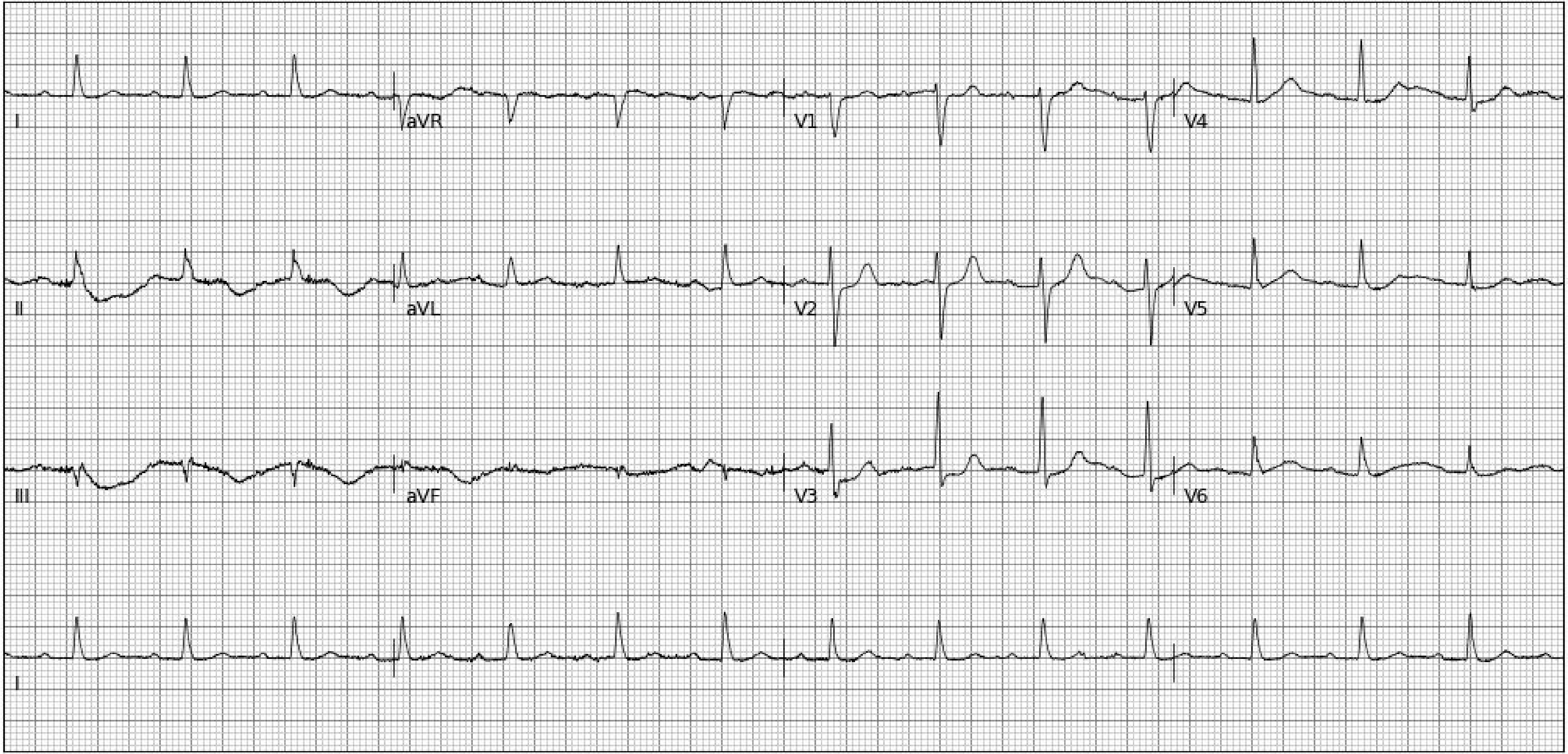
I
aVR
V1
V4
II
aVL
V2
V5
III
aVF
V3
V6
I

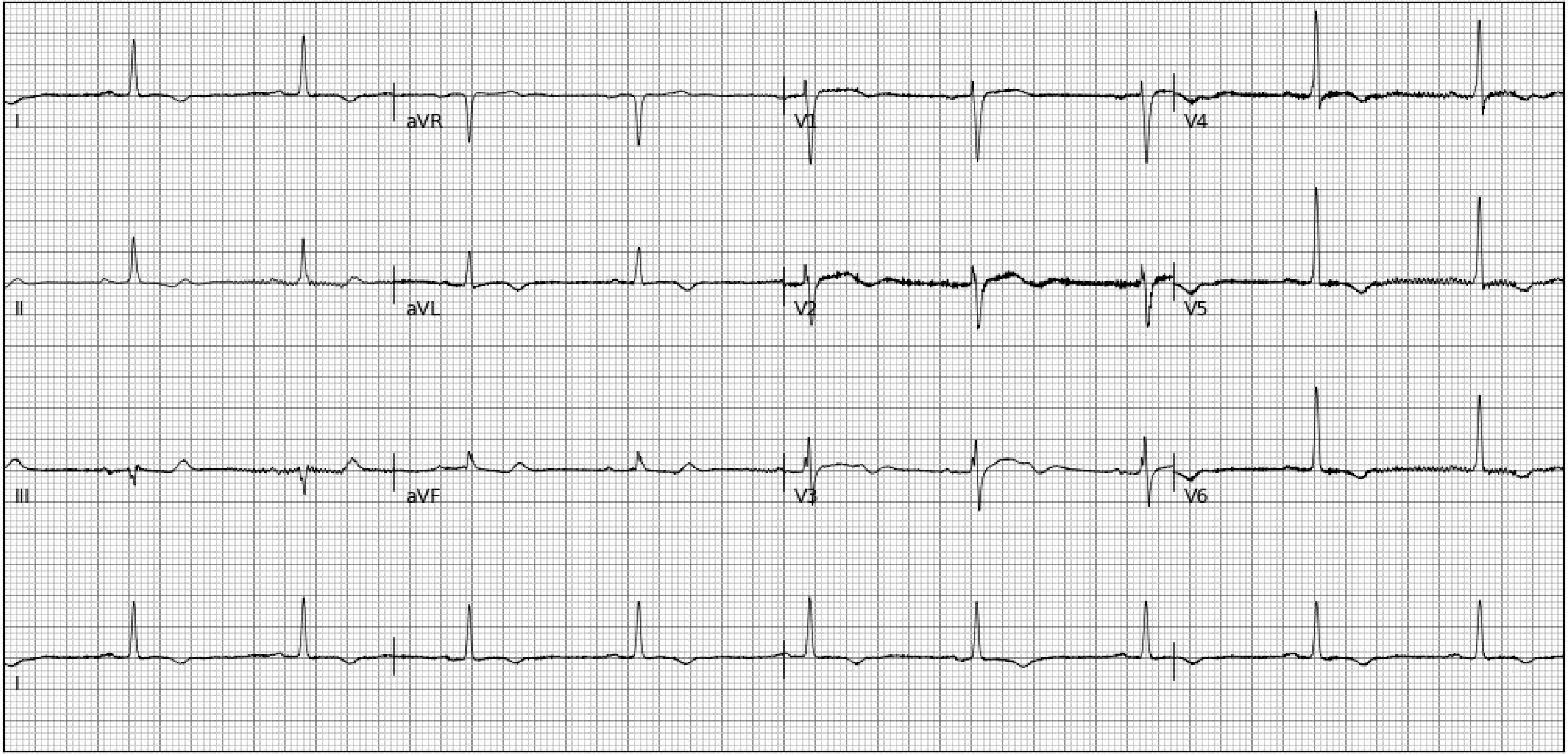
I
aVR
V1
V4
II
aVL
V2
V5
III
aVF
V3
V6
I

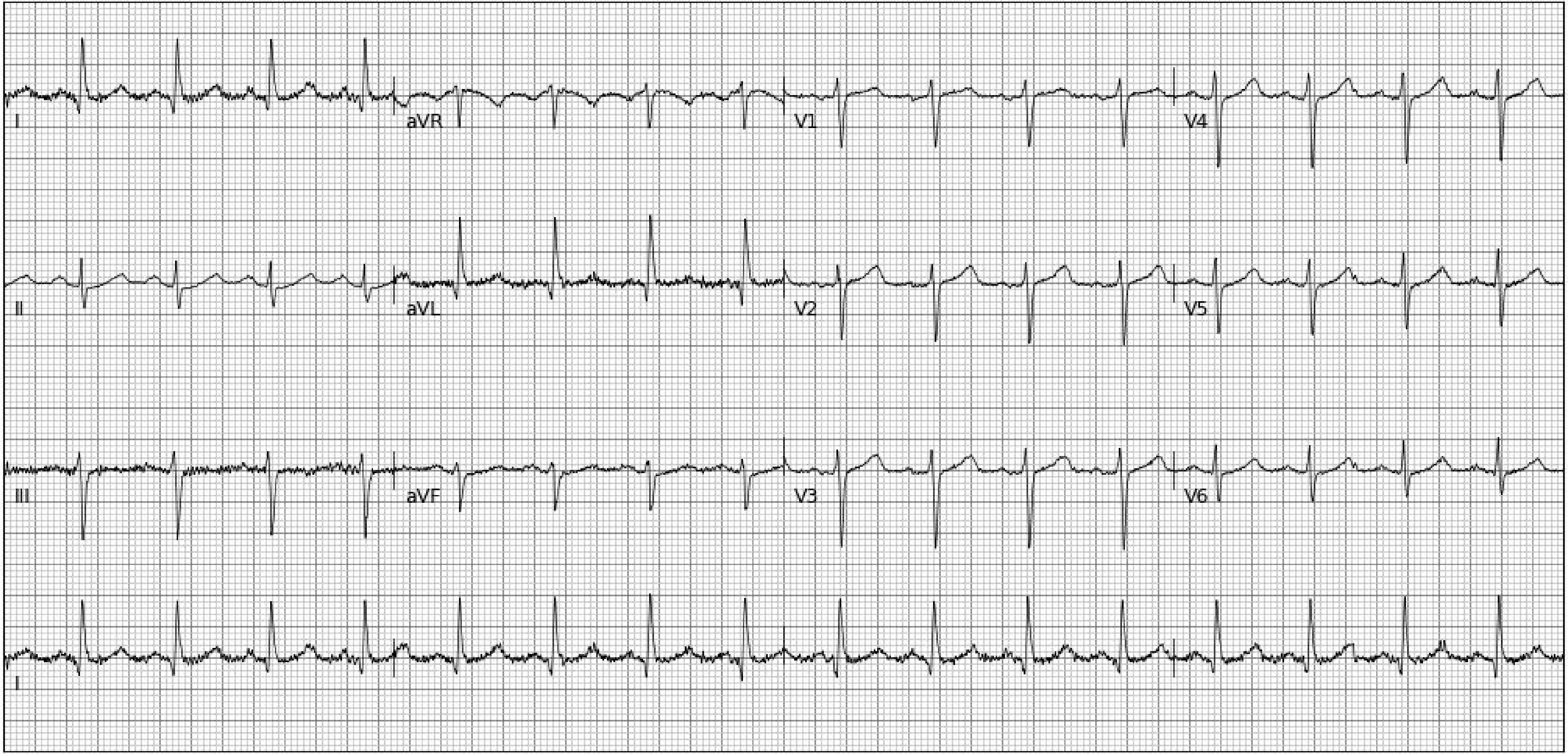
I
aVR
V1
V4
II
aVL
V2
V5
III
aVF
V3
V6
I

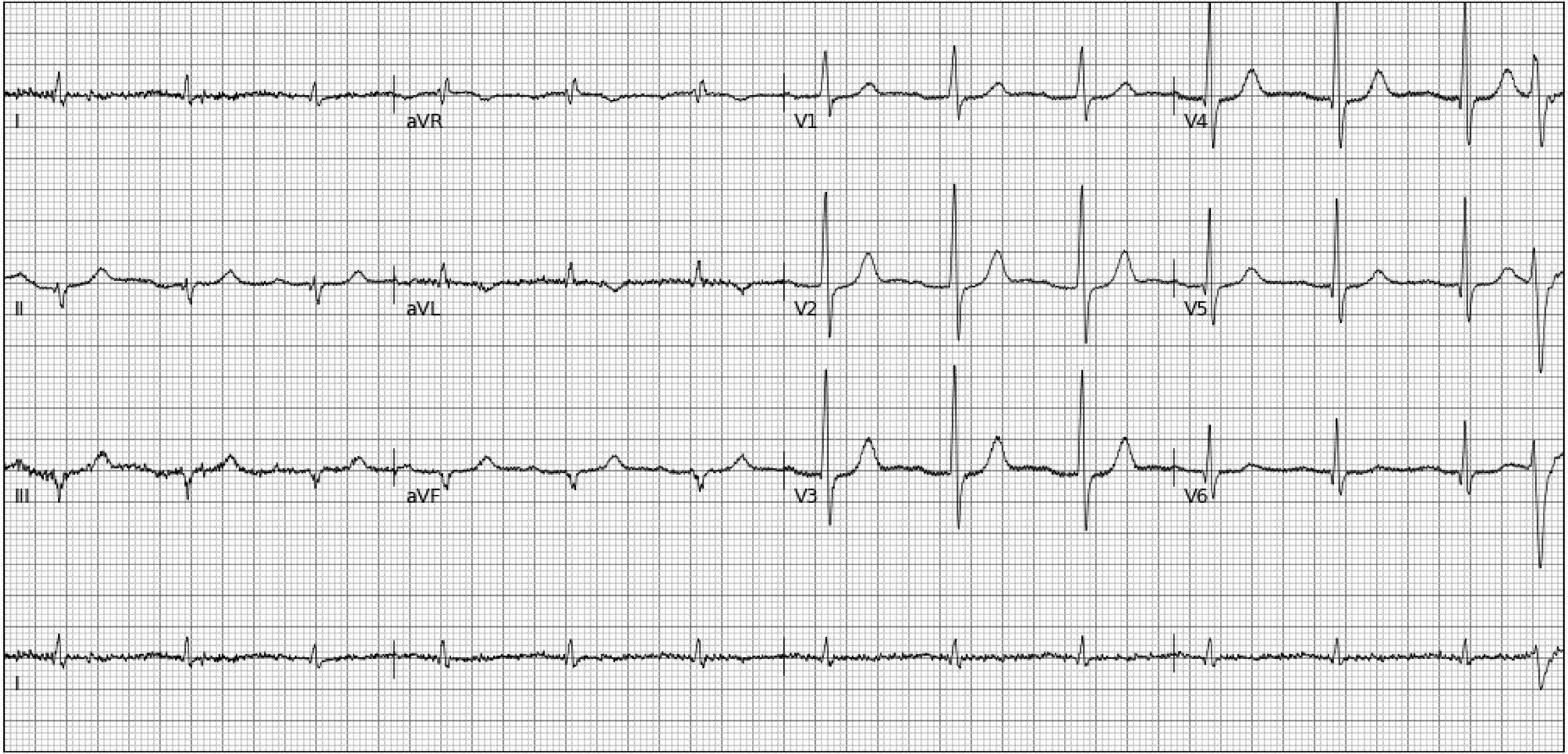
I
aVR
V1
V4
II
aVL
V2
V5
III
aVF
V3
V6
I

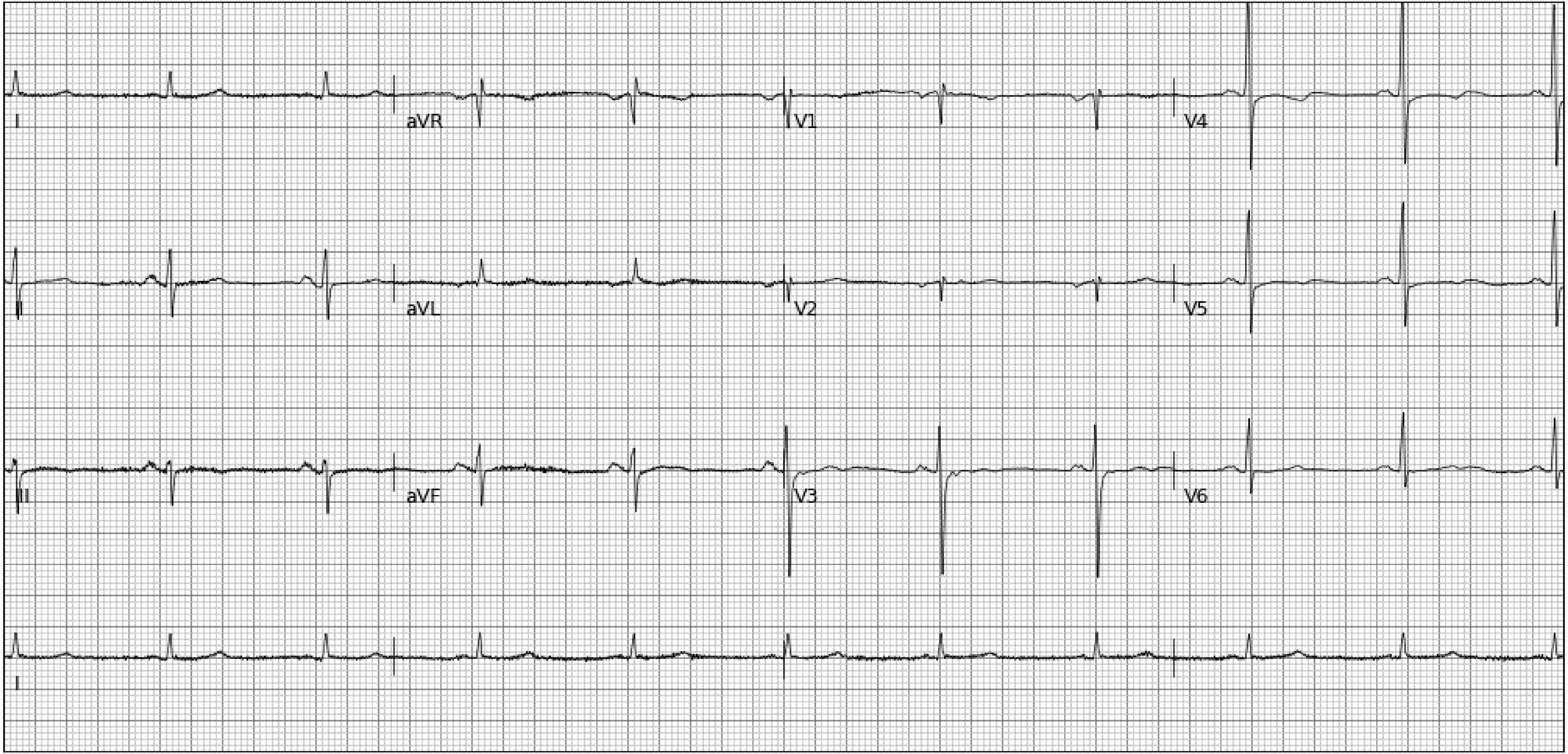
I
aVR
V1
V4
II
aVL
V2
V5
III
aVF
V3
V6
II

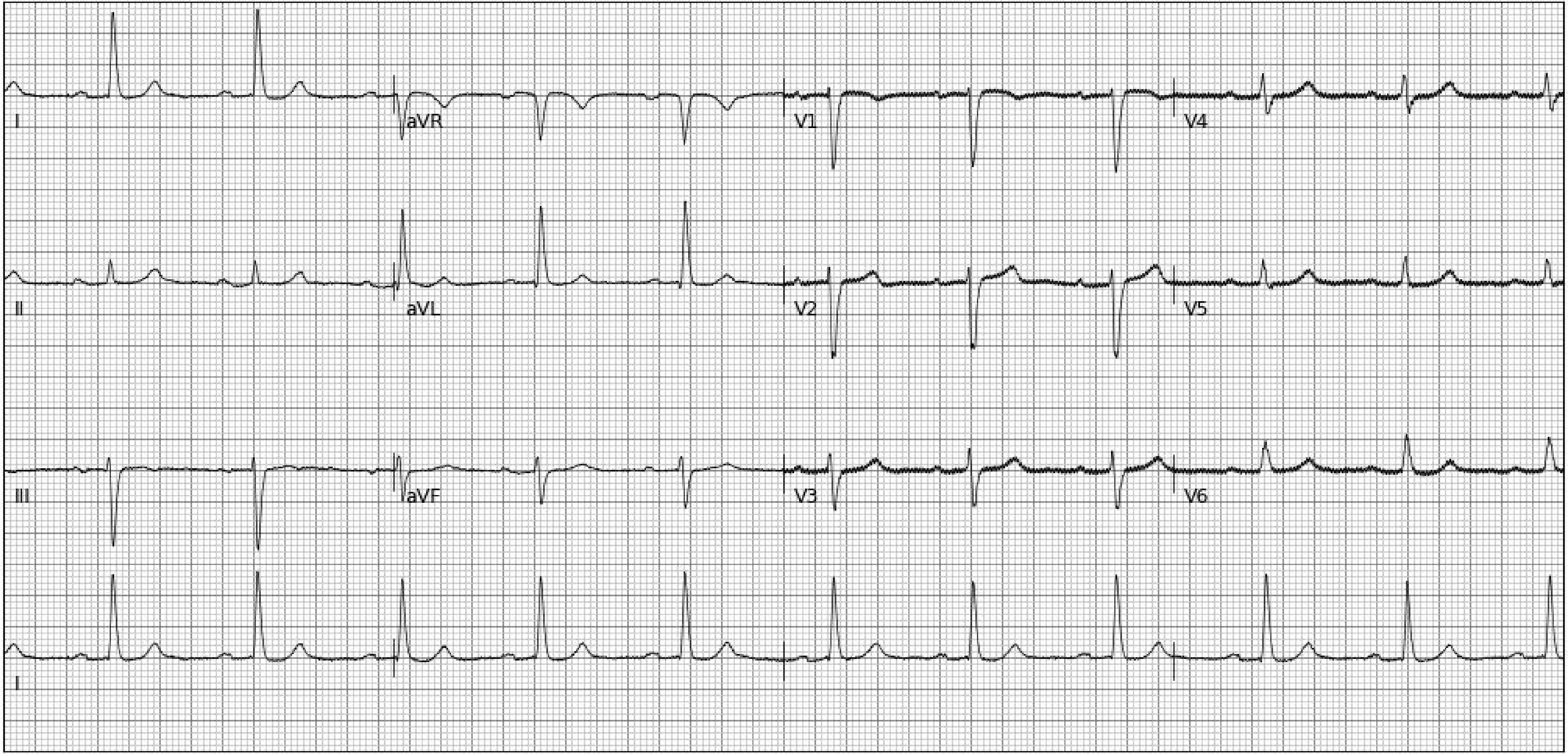
I
aVR
V1
V4
II
aVL
V2
V5
III
aVF
V3
V6
I

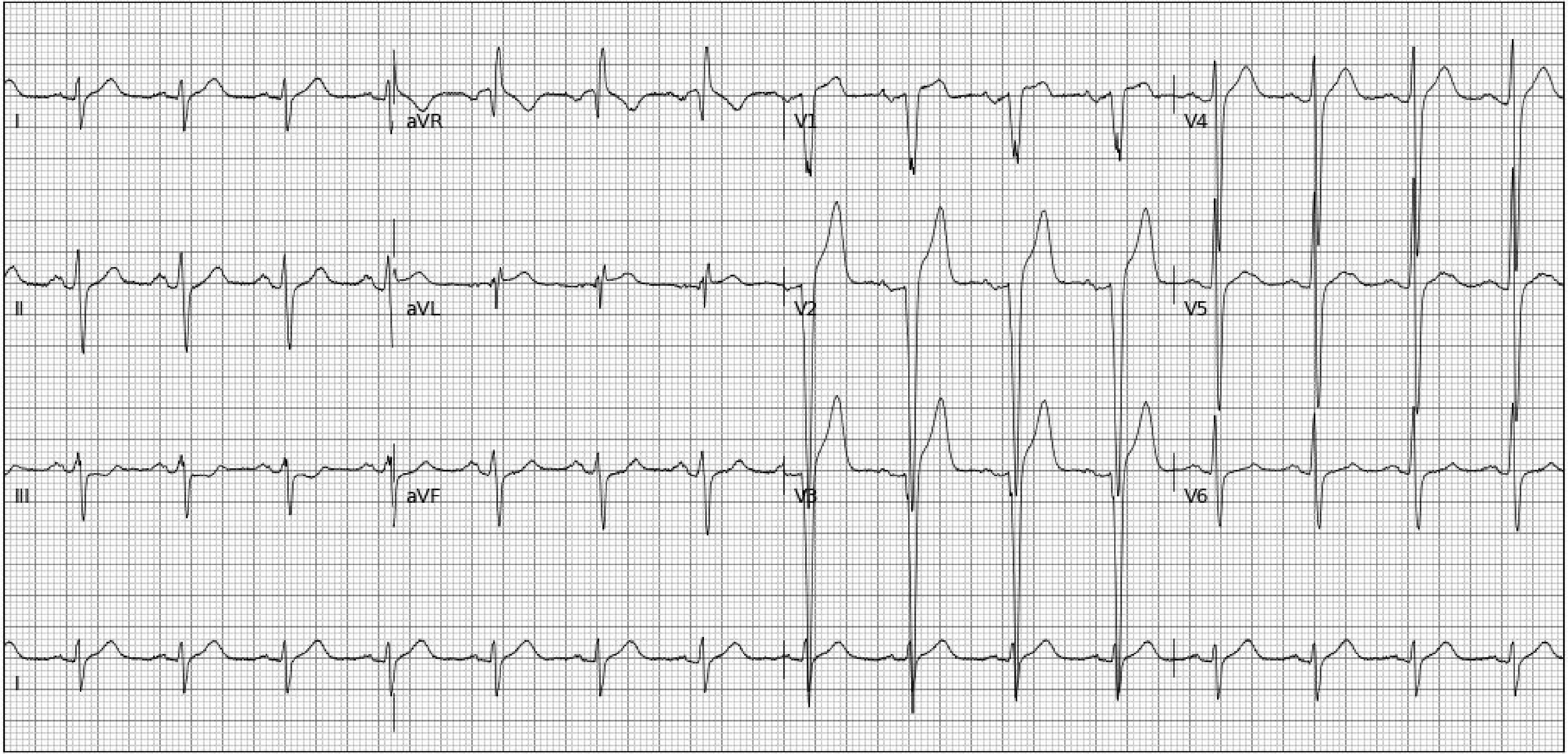
I
aVR
V1
V4
II
aVL
V2
V5
III
aVF
V3
V6
I

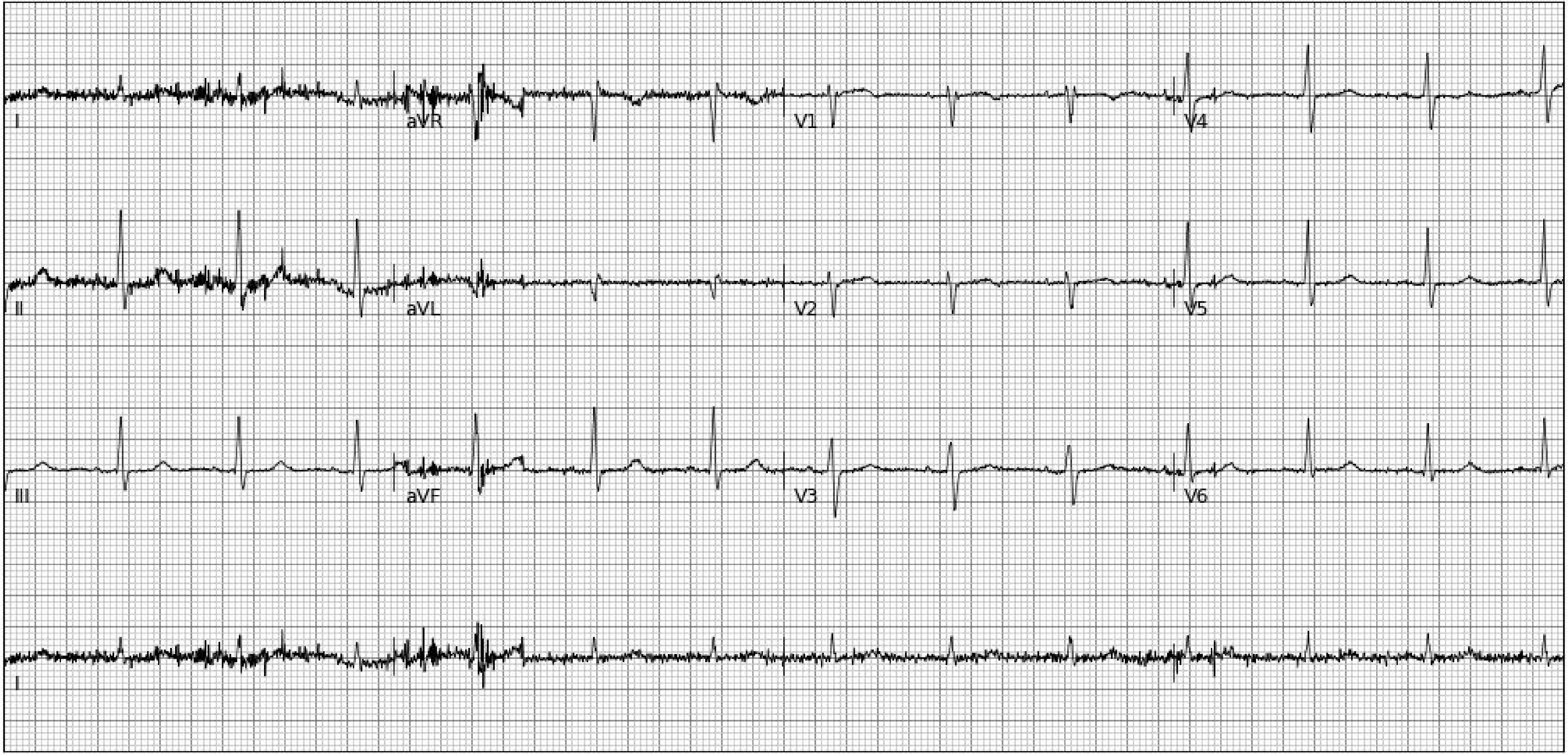
I
aVR
V1
V4
II
aVL
V2
V5
III
aVF
V3
V6
I

**Section 3. Random examples of 25 electrocardiogram images plotted in novel format (Black-on-Red Standard) not encountered during model development**

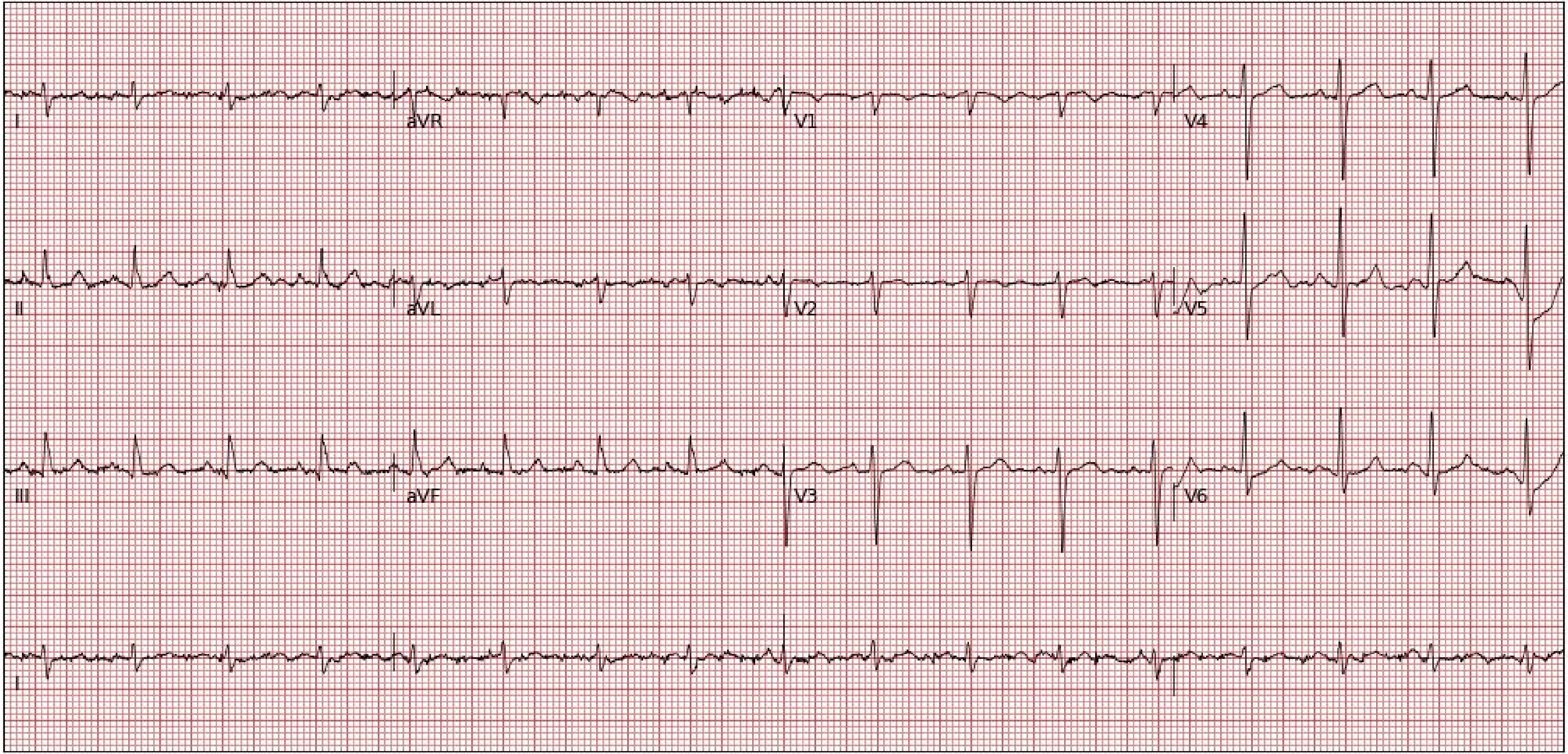
I
aVR
V1
V4
II
aVL
V2
V5
III
aVF
V3
V6
I

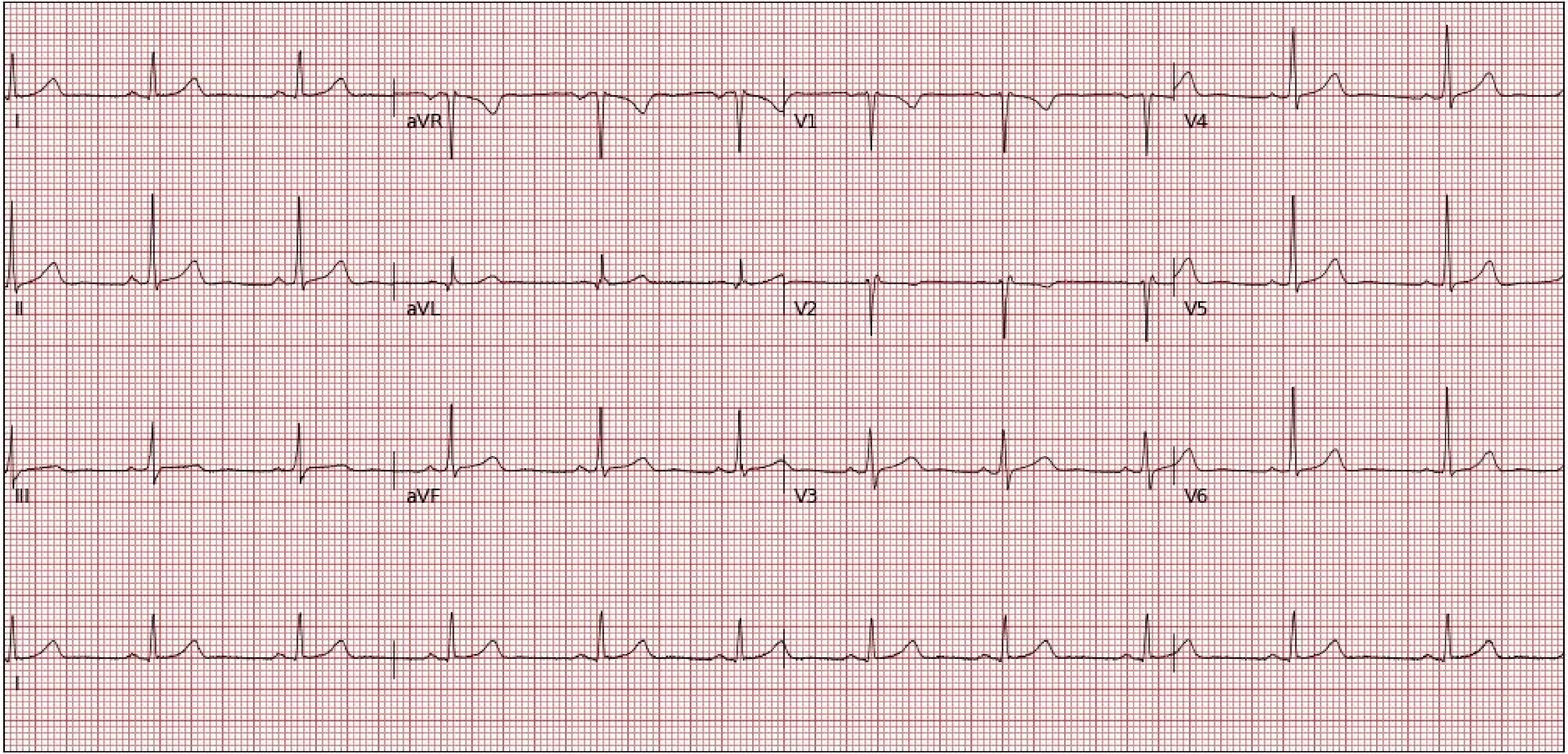
I
aVR
V1
V4
II
aVL
V2
V5
III
aVF
V3
V6
II

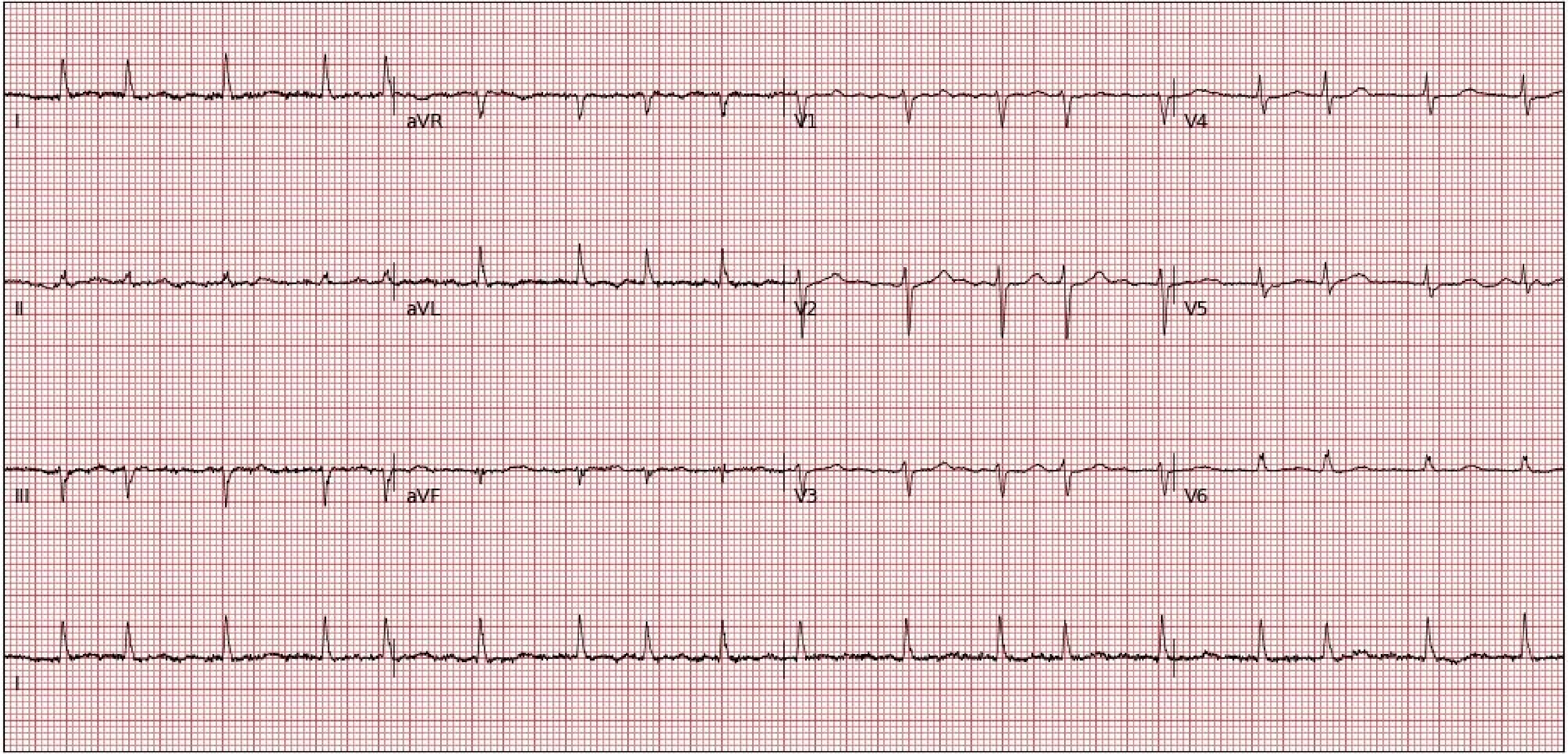
I
aVR
V1
V4
II
aVL
V2
V5
III
aVF
V3
V6
I

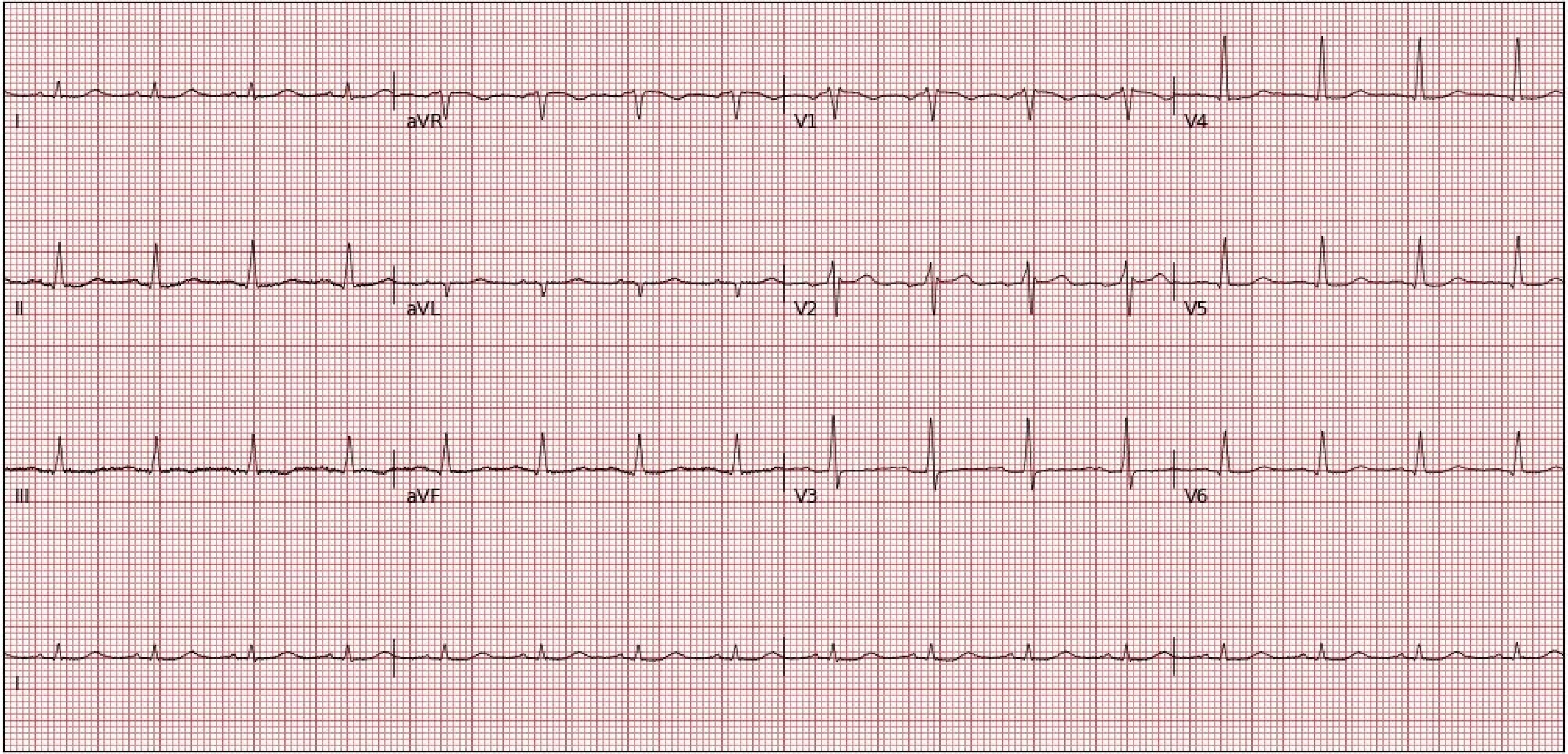
I
aVR
V1
V4
II
aVL
V2
V5
III
aVF
V3
V6
II

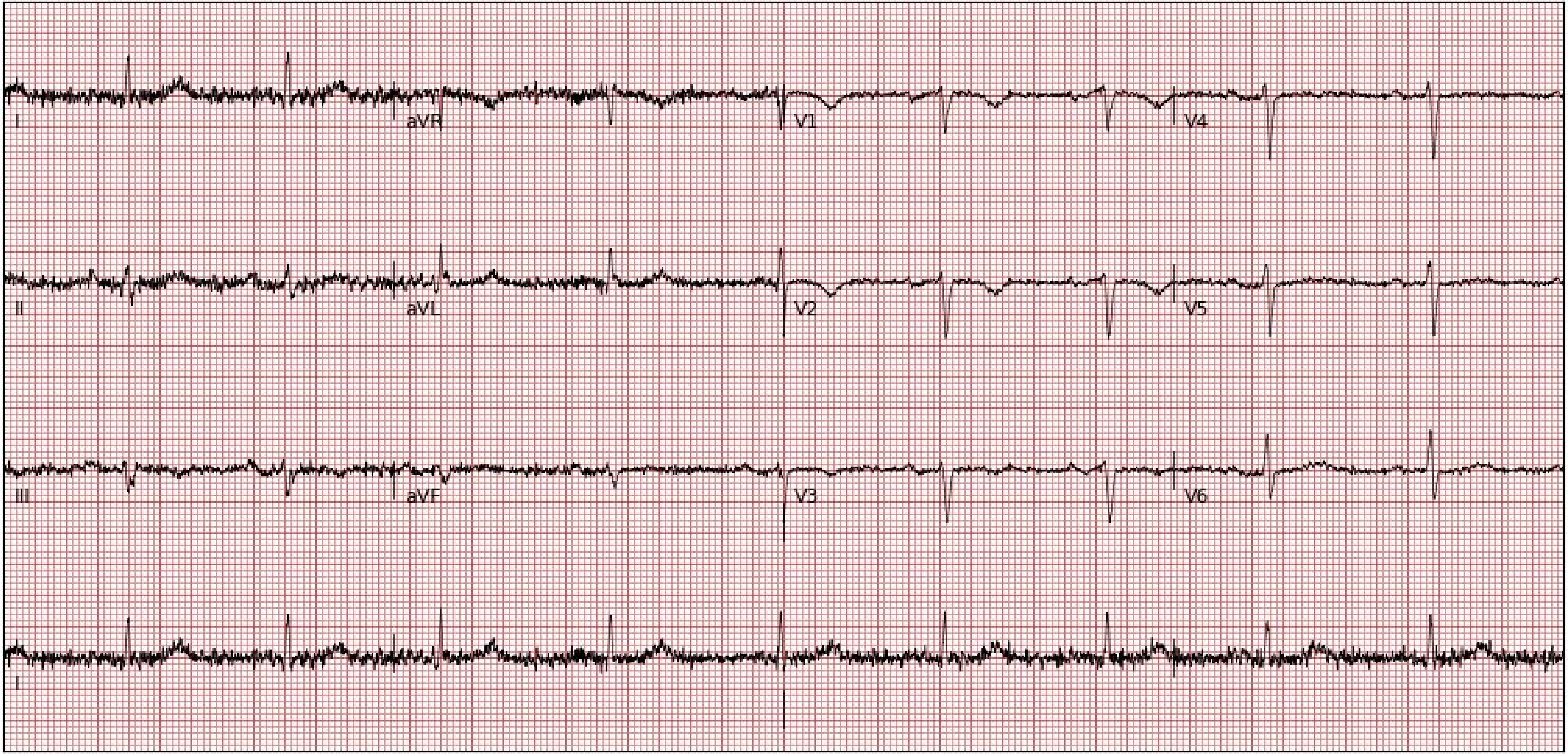
I
aVR
V1
V4
II
aVL
V2
V5
III
aVF
V3
V6
I

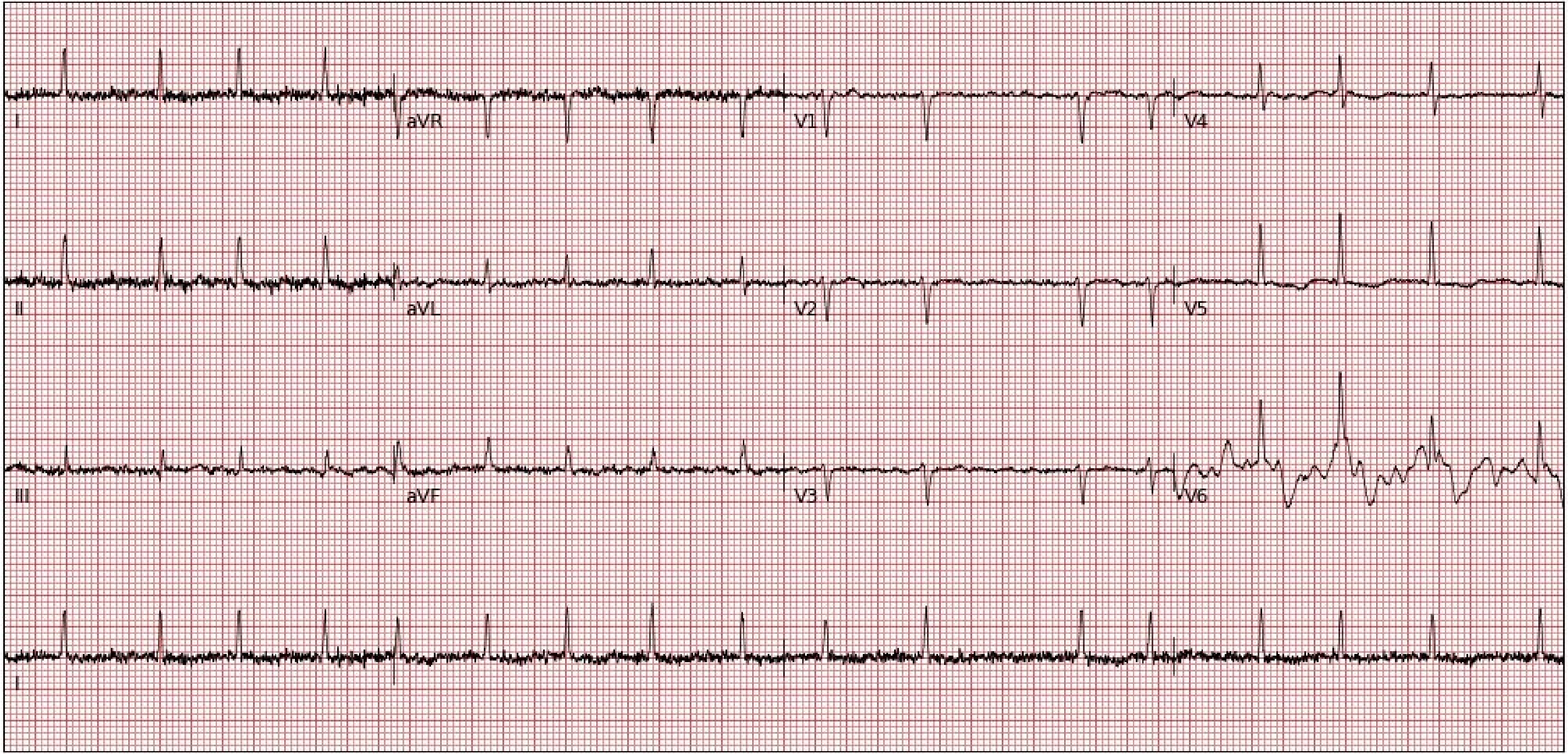
I
aVR
V1
V4
II
aVL
V2
V5
III
aVF
V3
V6
I

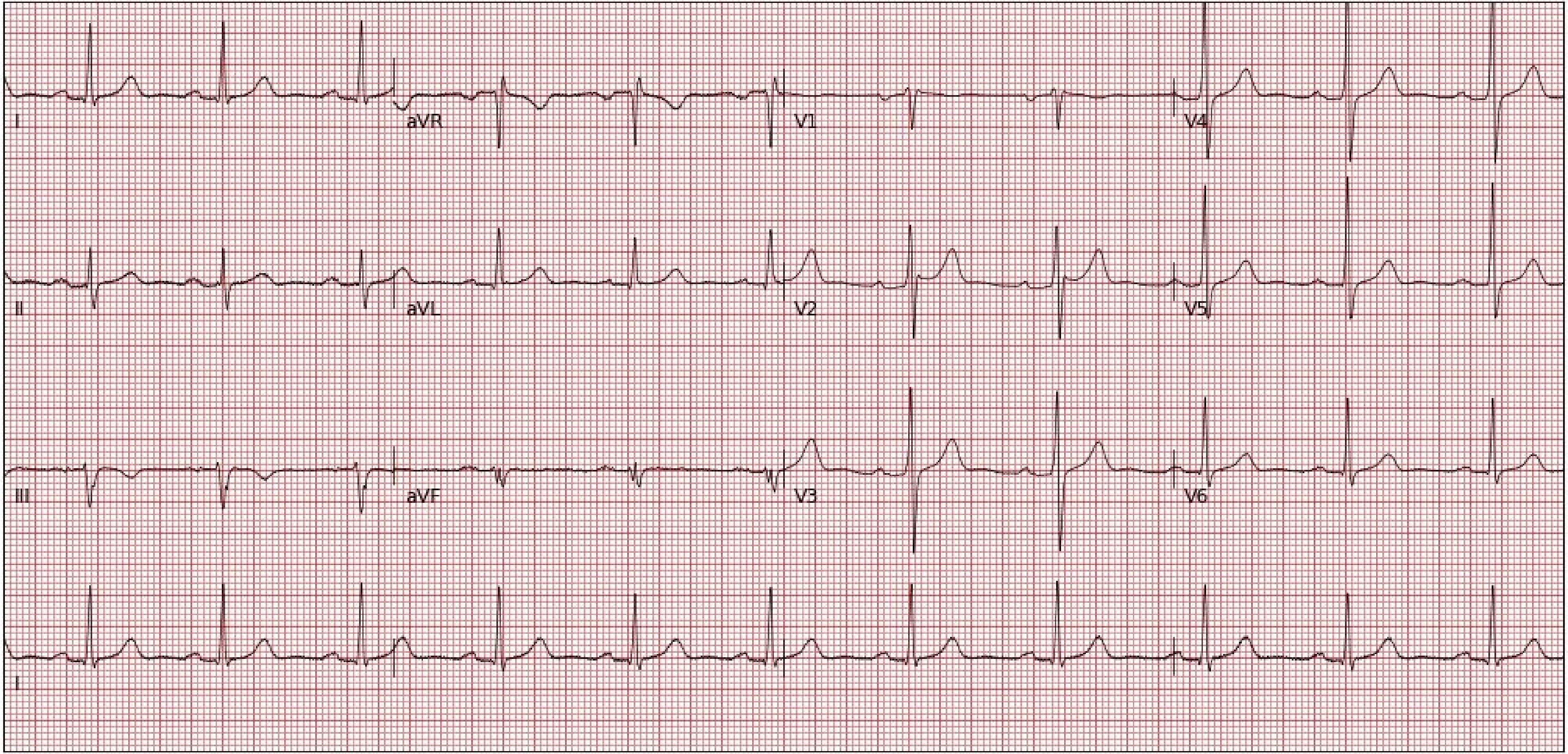
I
aVR
V1
V4
II
aVL
V2
V5
III
aVF
V3
V6
I

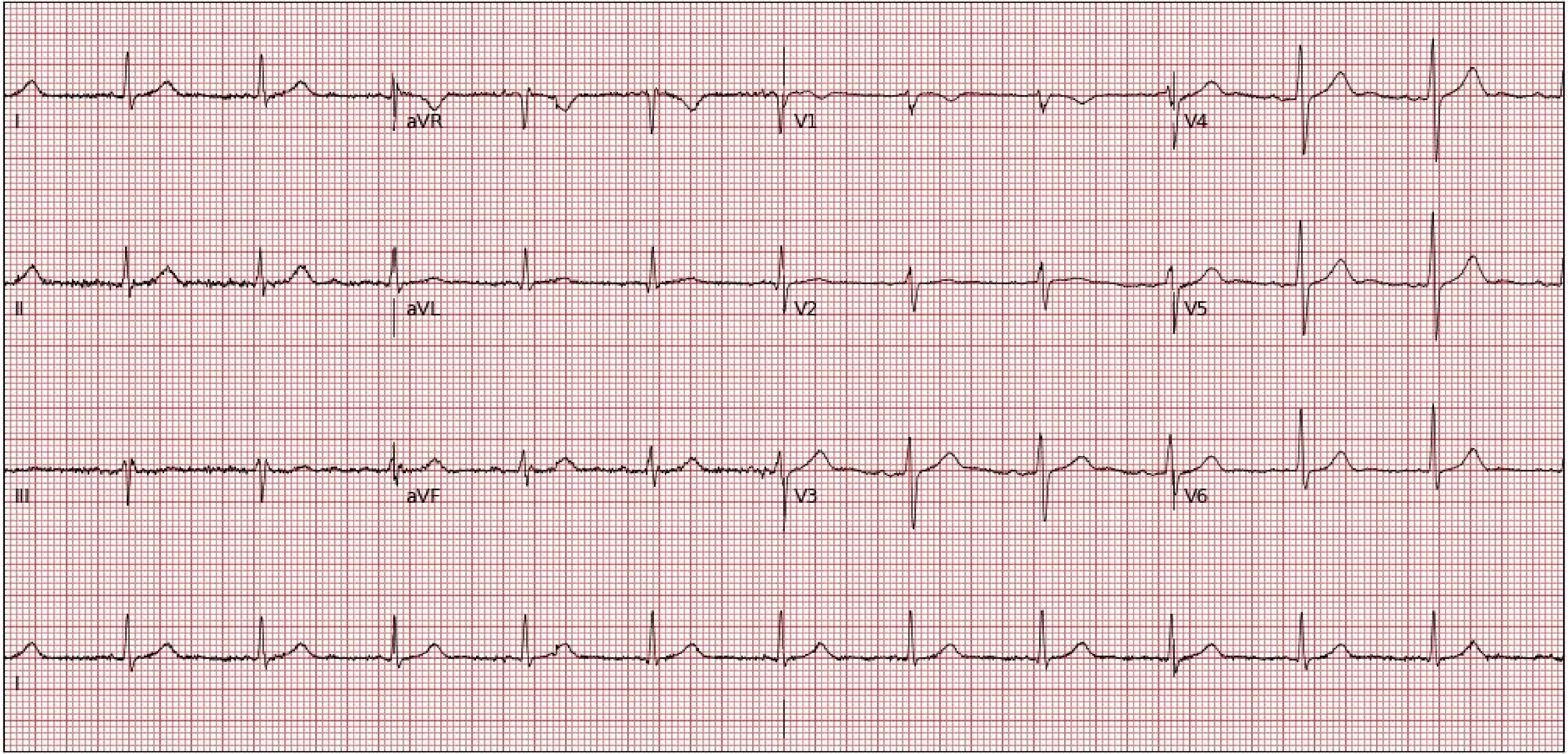
I
aVR
V1
V4
II
aVL
V2
V5
III
aVF
V3
V6
I

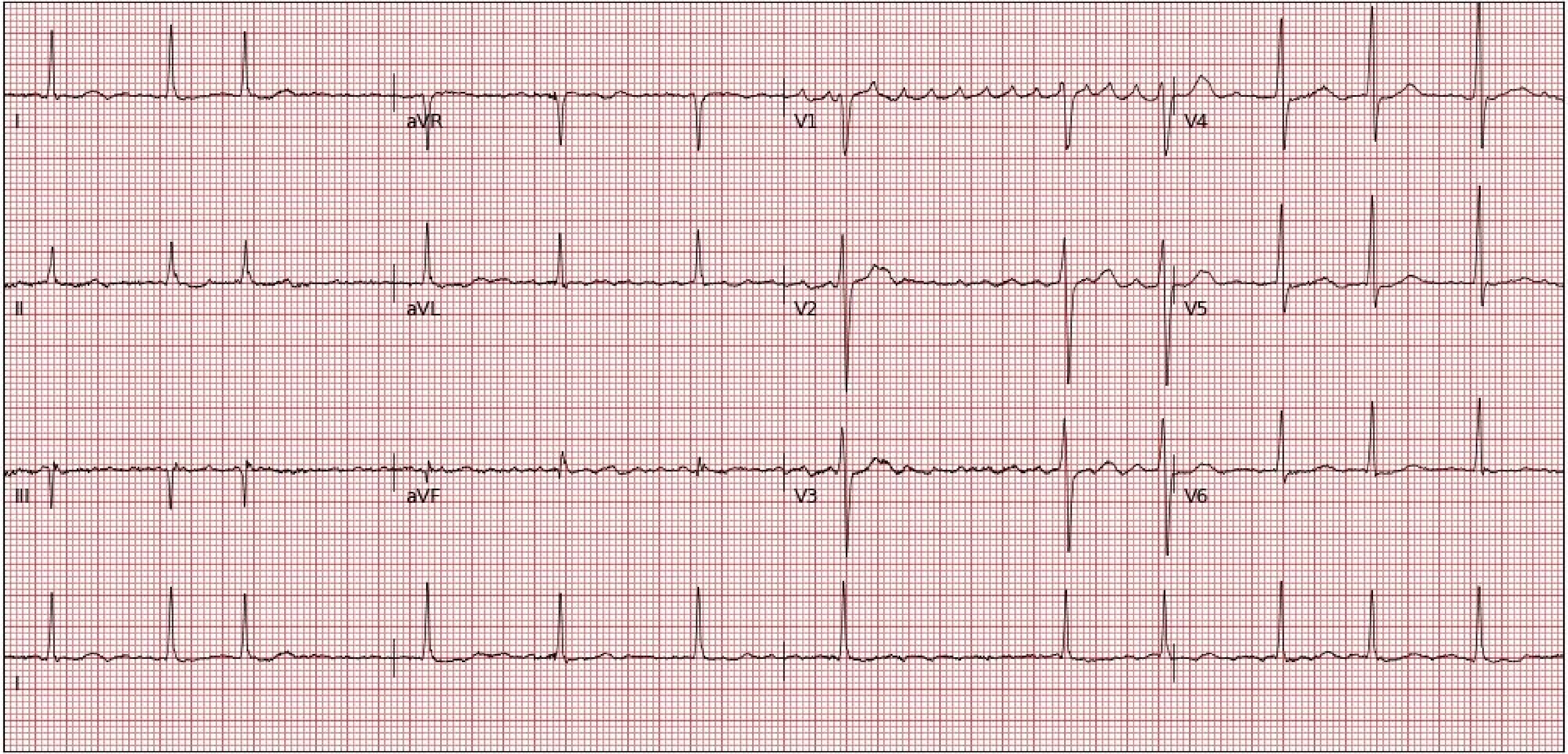

I
aVR
V1
V4
II
aVL
V2
V5
III
aVF
V3
V6
I

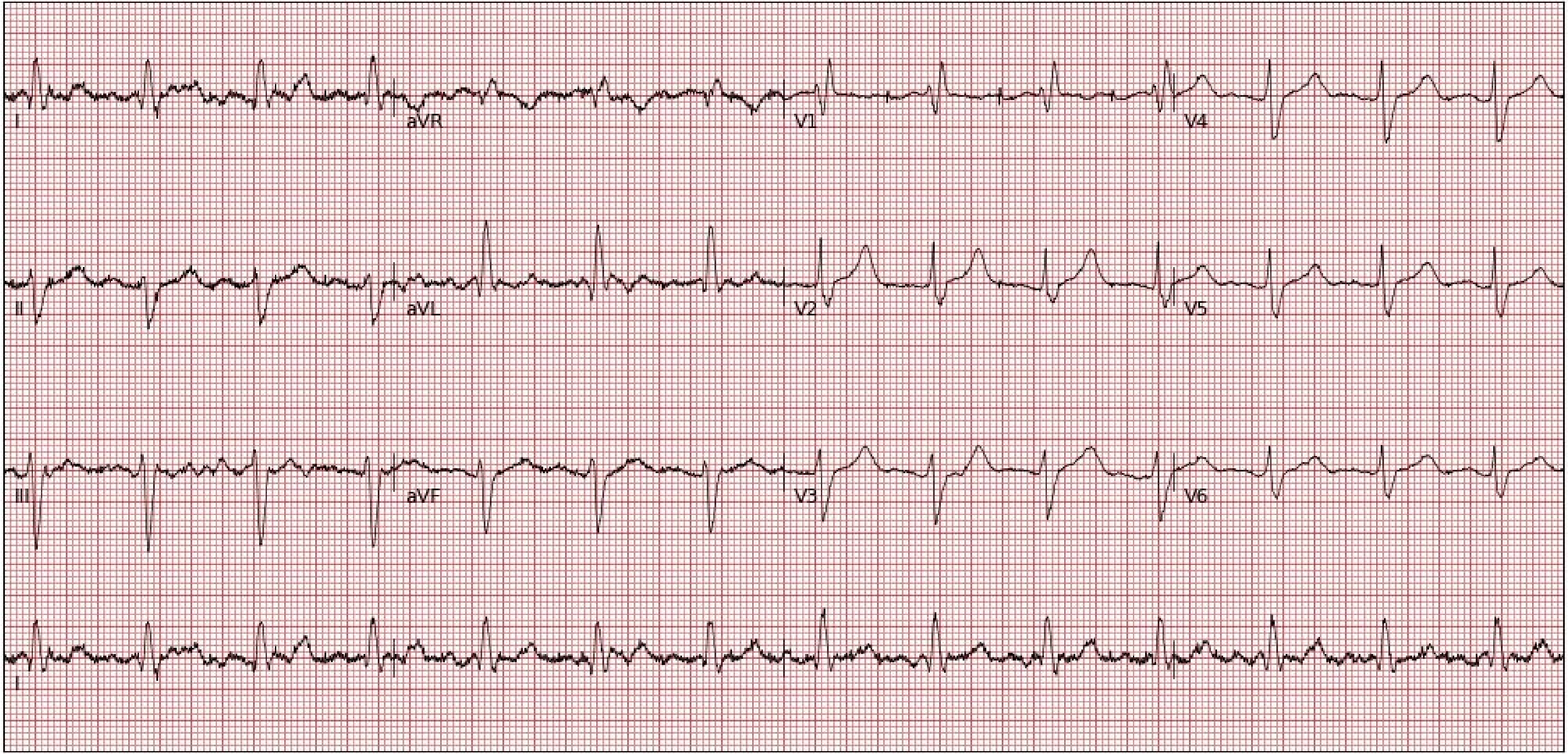
I
aVR
V1
V4
II
aVL
V2
V5
III
aVF
V3
V6
I

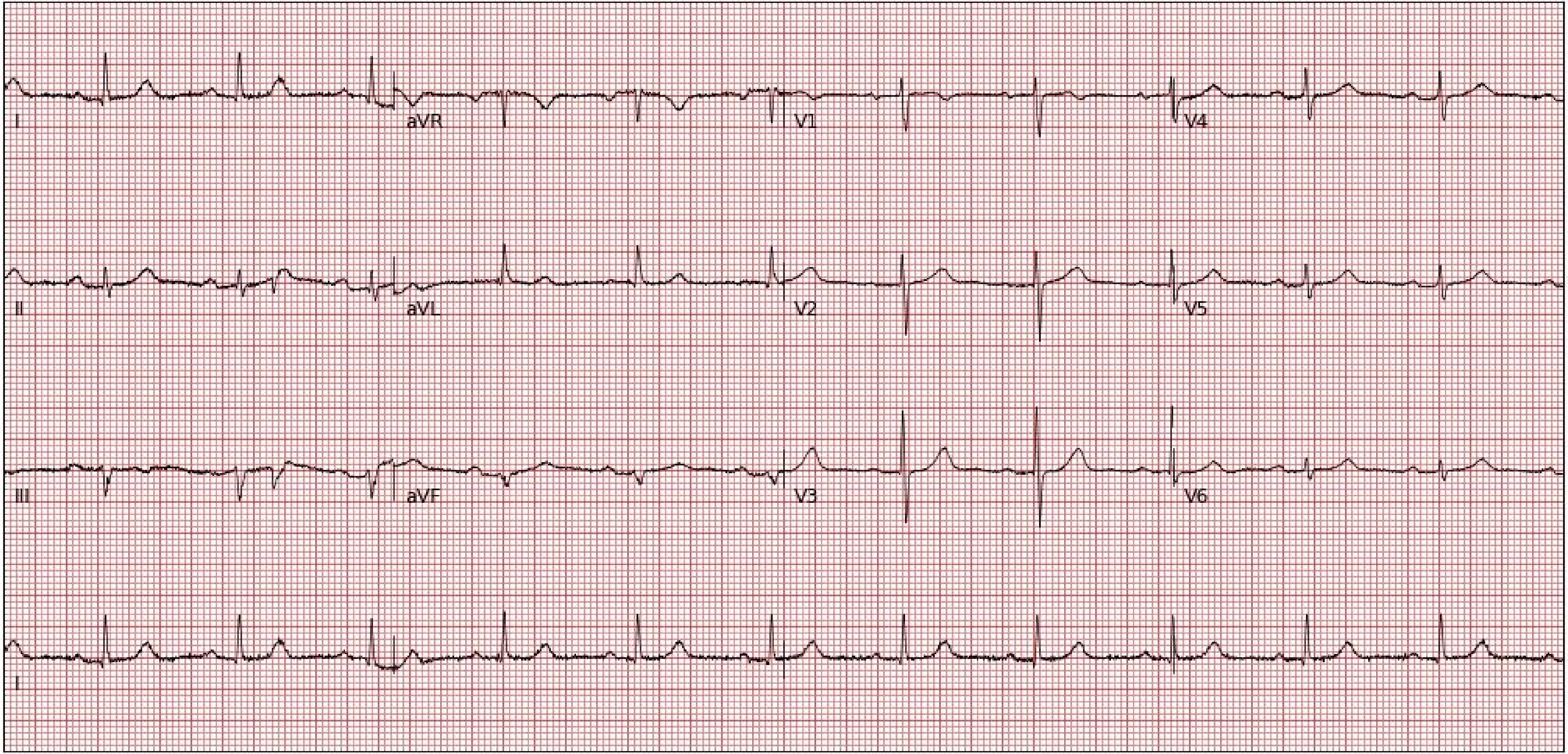
I
aVR
V1
V4
II
aVL
V2
V5
III
aVF
V3
V6
I

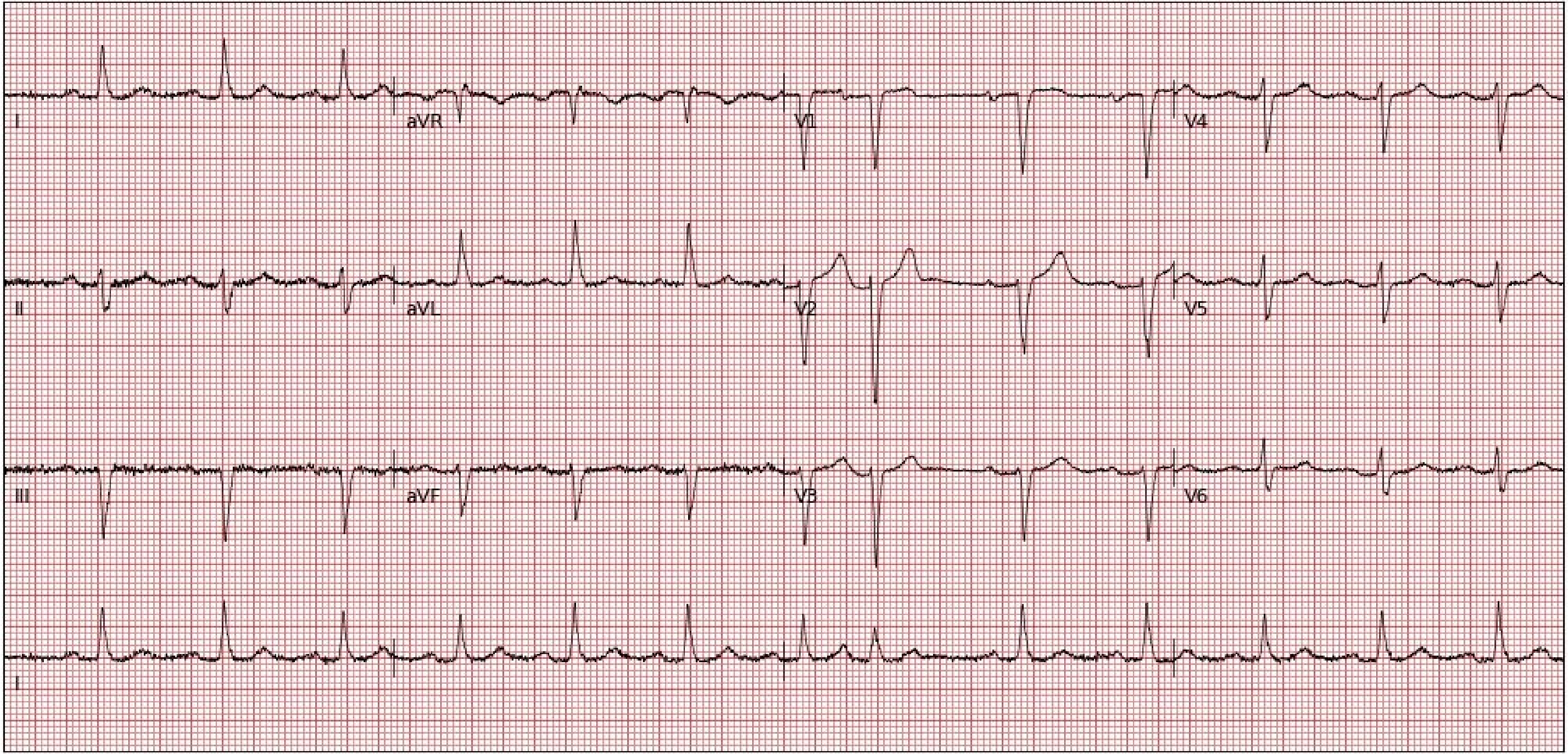
I
aVR
V1
V4
II
aVL
V2
V5
III
aVF
V3
V6
I

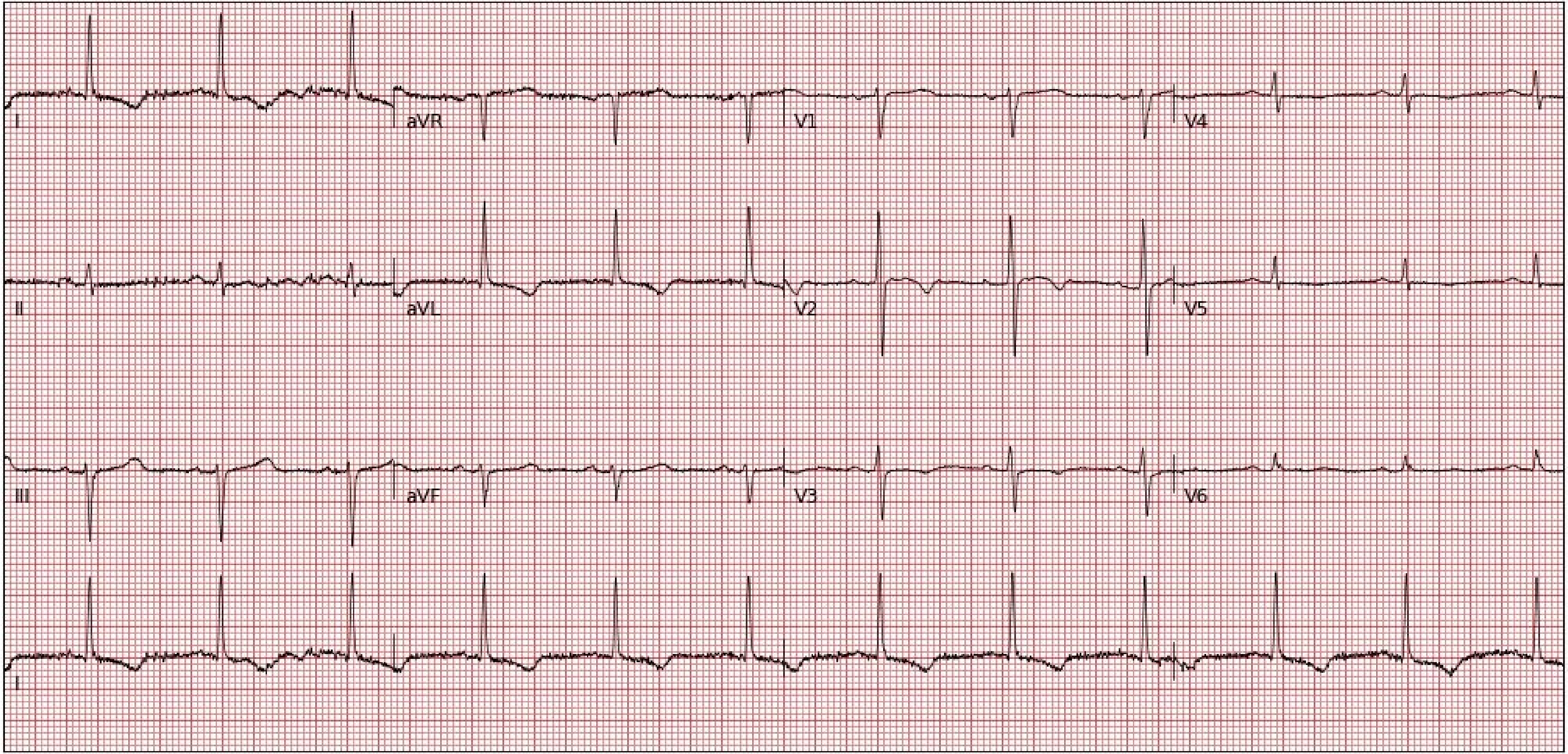
I
aVR
V1
V4
II
aVL
V2
V5
III
aVF
V3
V6
I

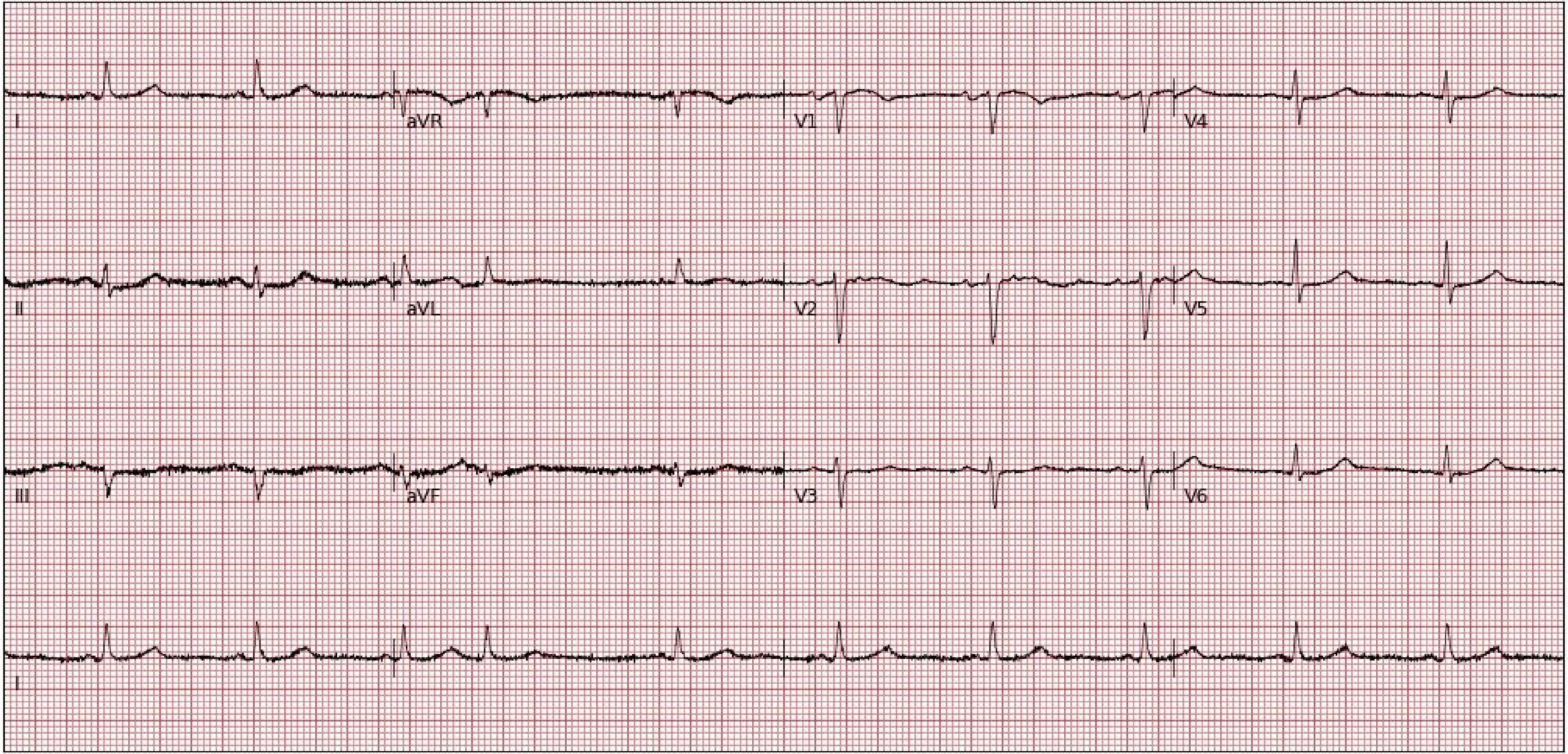
I
aVR
V1
V4
II
aVL
V2
V5
III
aVF
V3
V6
I

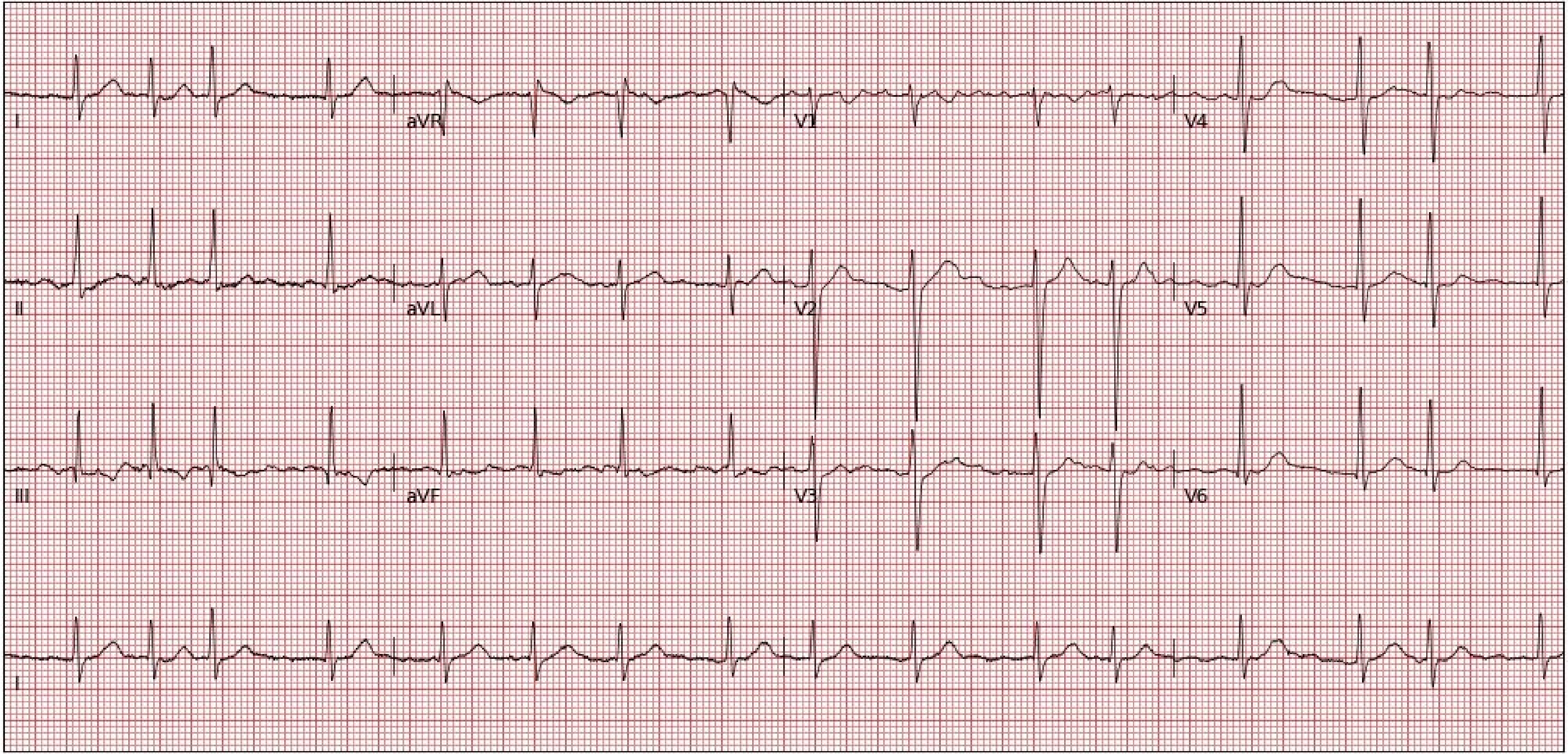
I
aVR
V1
V4
II
aVL
V2
V5
III
aVF
V3
V6
II

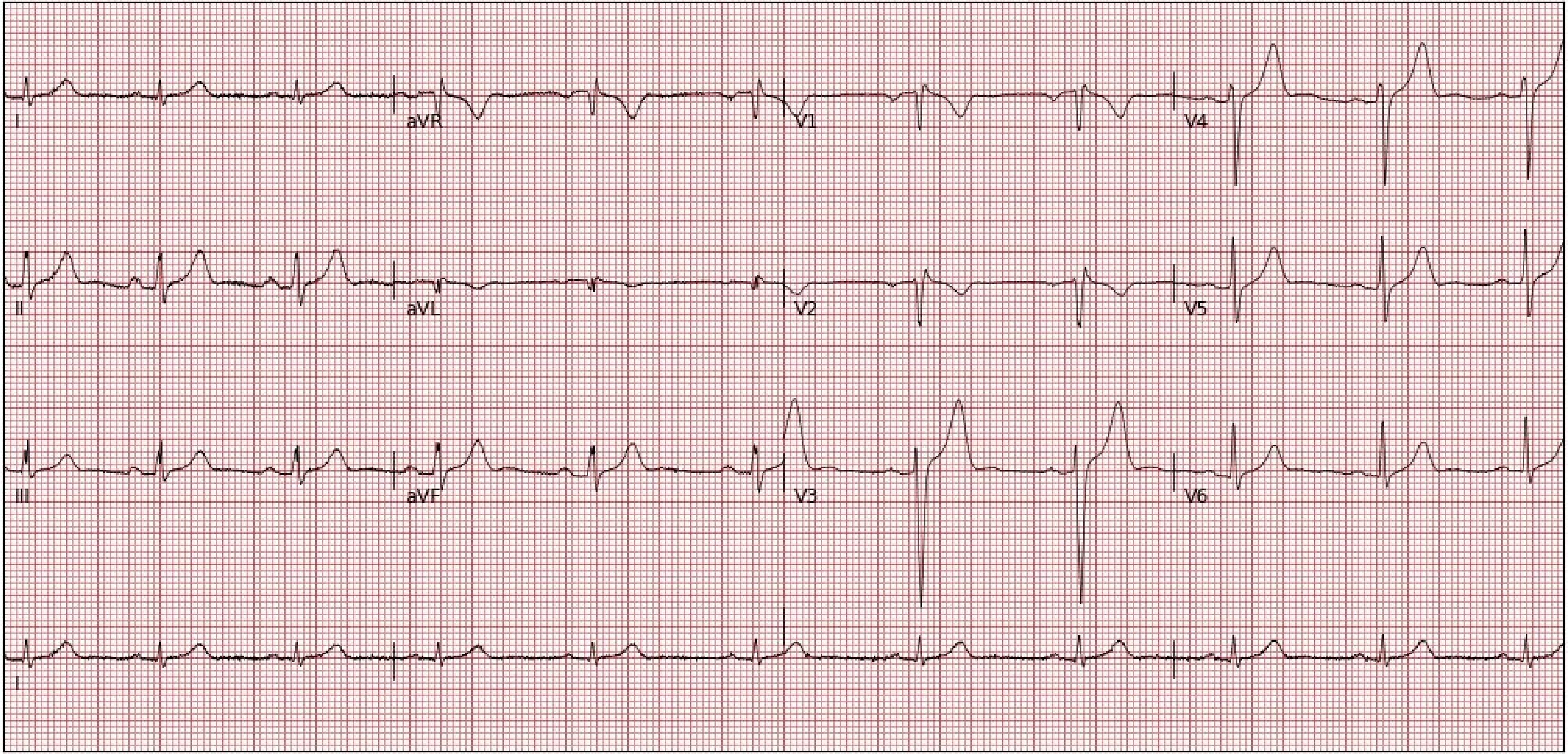
I
aVR
V1
V4
II
aVL
V2
V5
III
aVF
V3
V6
I

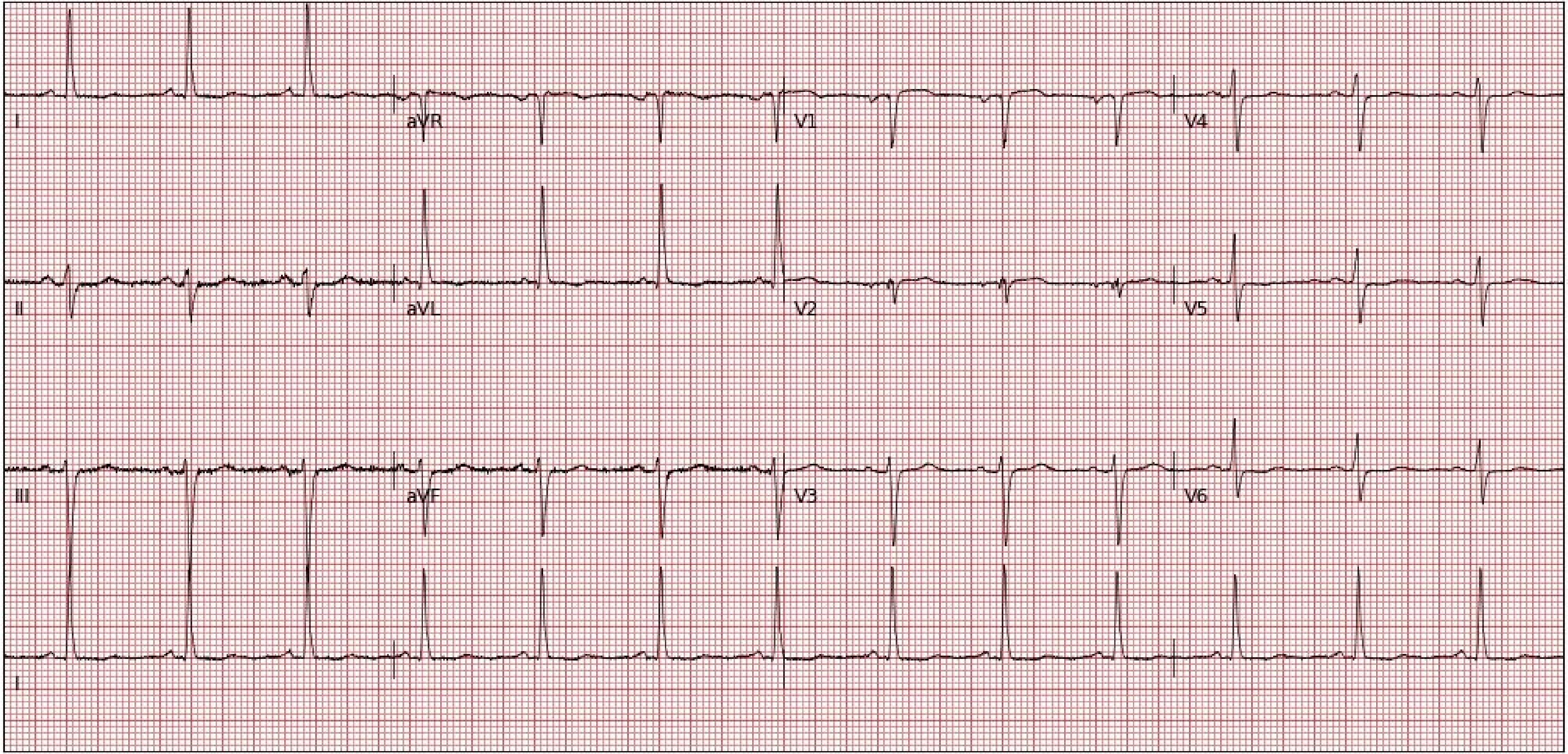
I
aVR
V1
V4
II
aVL
V2
V5
III
aVF
V3
V6
I

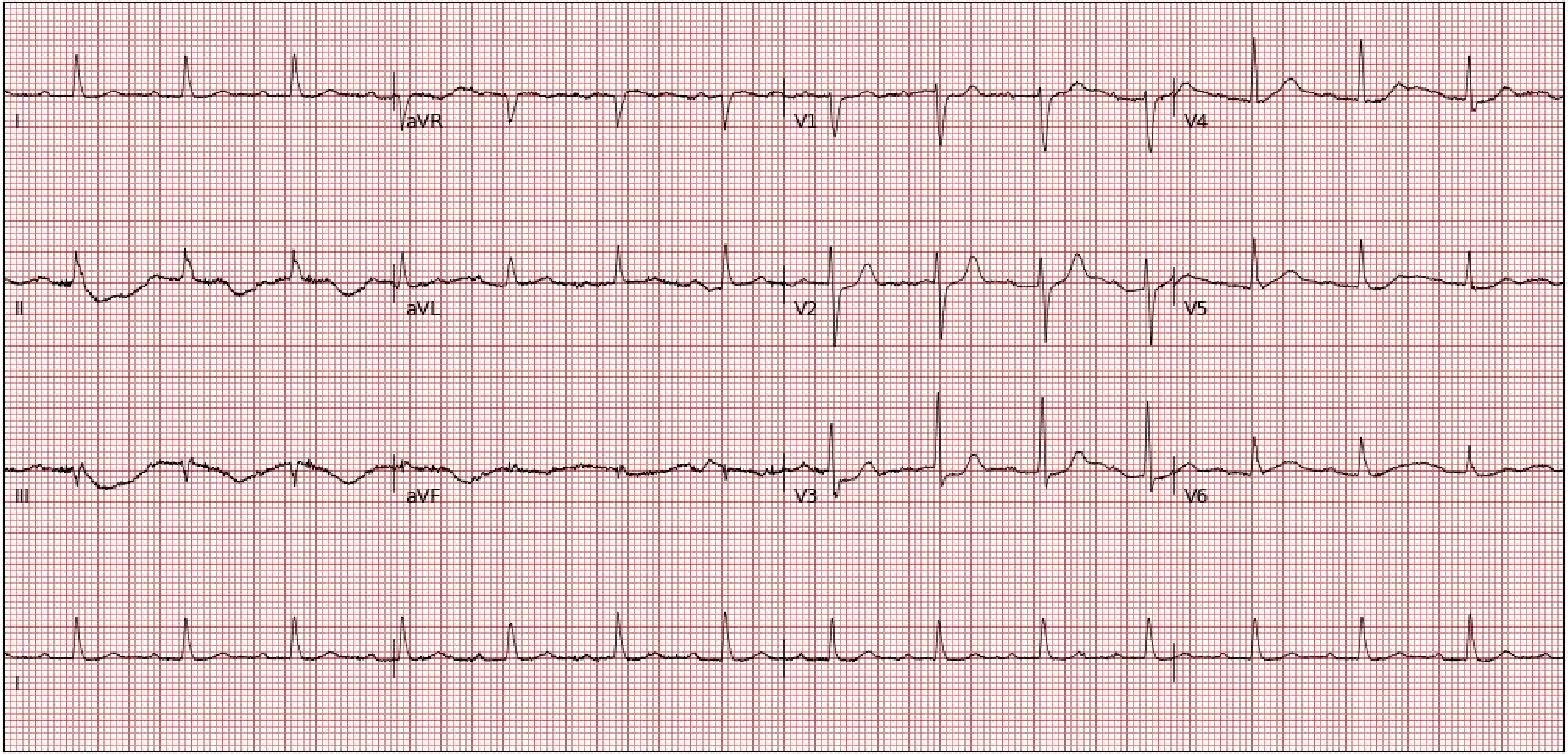
I
aVR
V1
V4
II
aVL
V2
V5
III
aVF
V3
V6
I

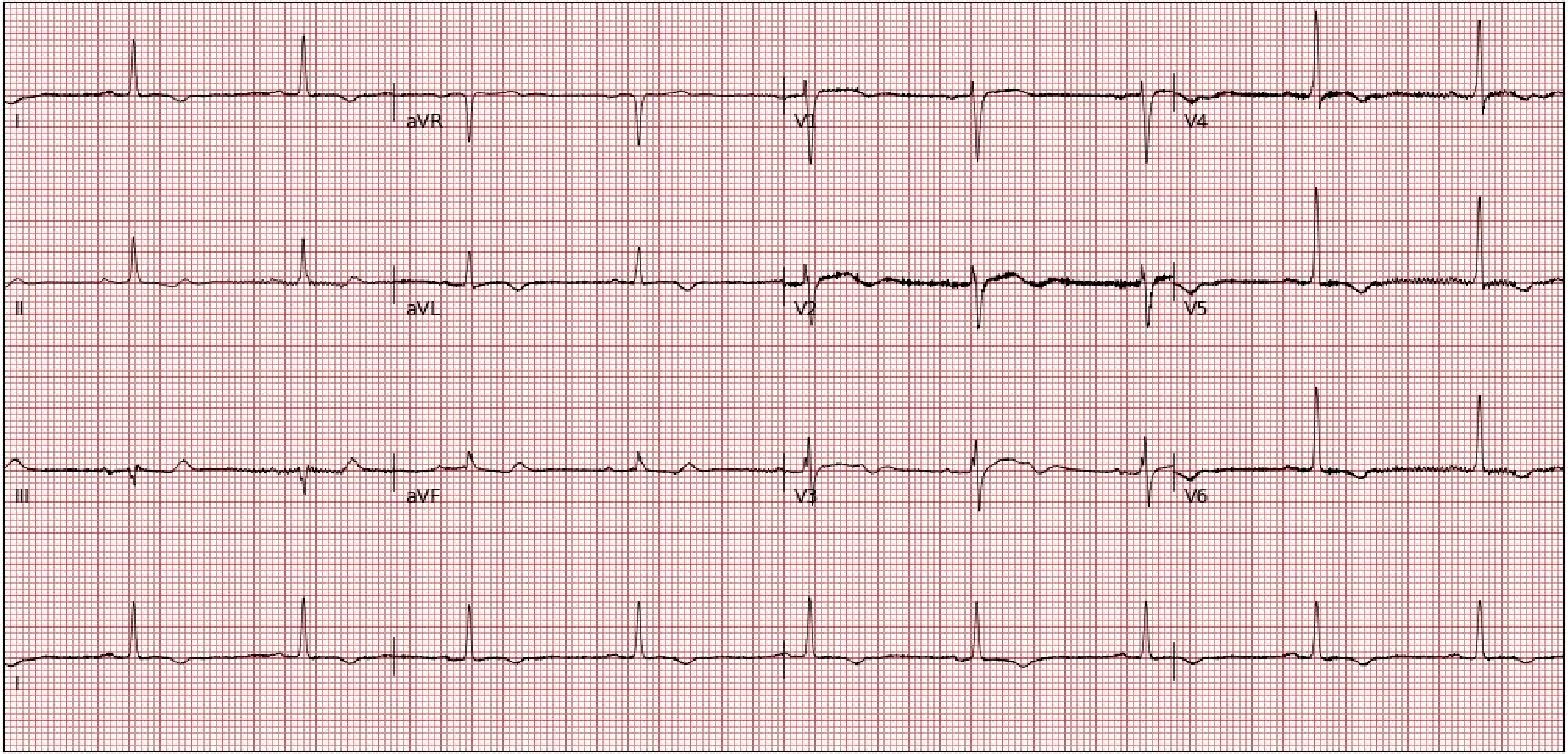
I
aVR
V1
V4
II
aVL
V2
V5
III
aVF
V3
V6
I

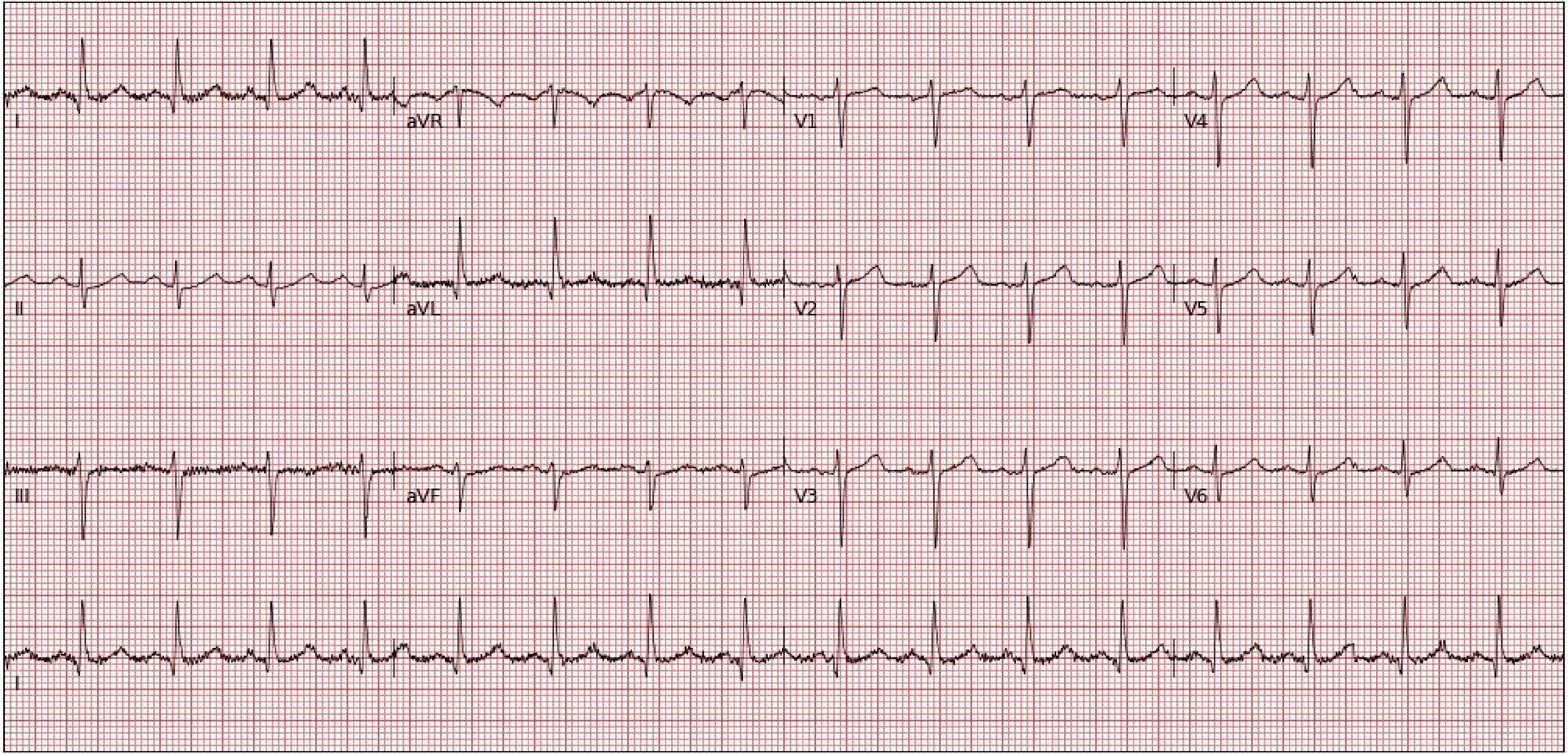
I
aVR
V1
V4
II
aVL
V2
V5
III
aVF
V3
V6
I

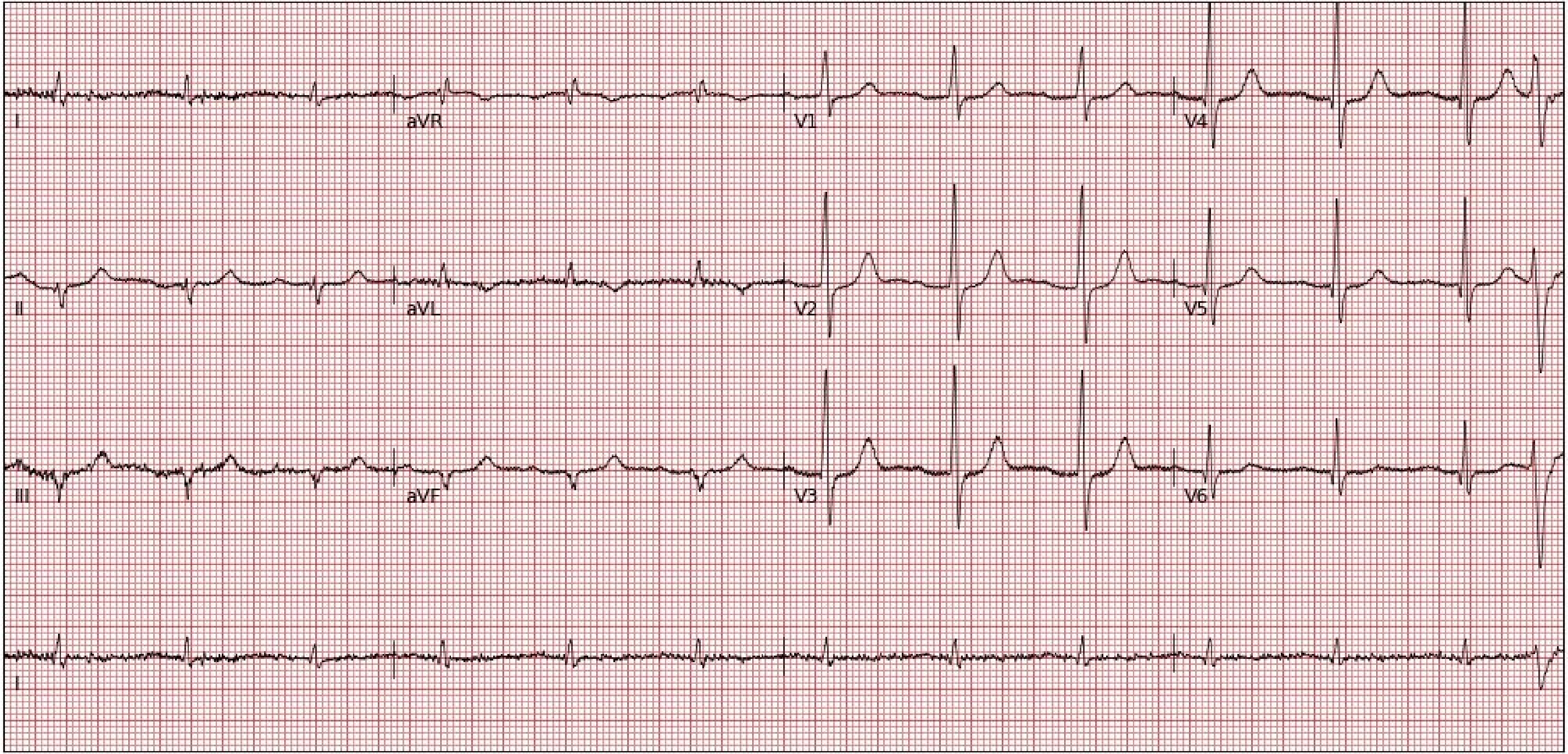
I
aVR
V1
V4
II
aVL
V2
V5
III
aVF
V3
V6
I

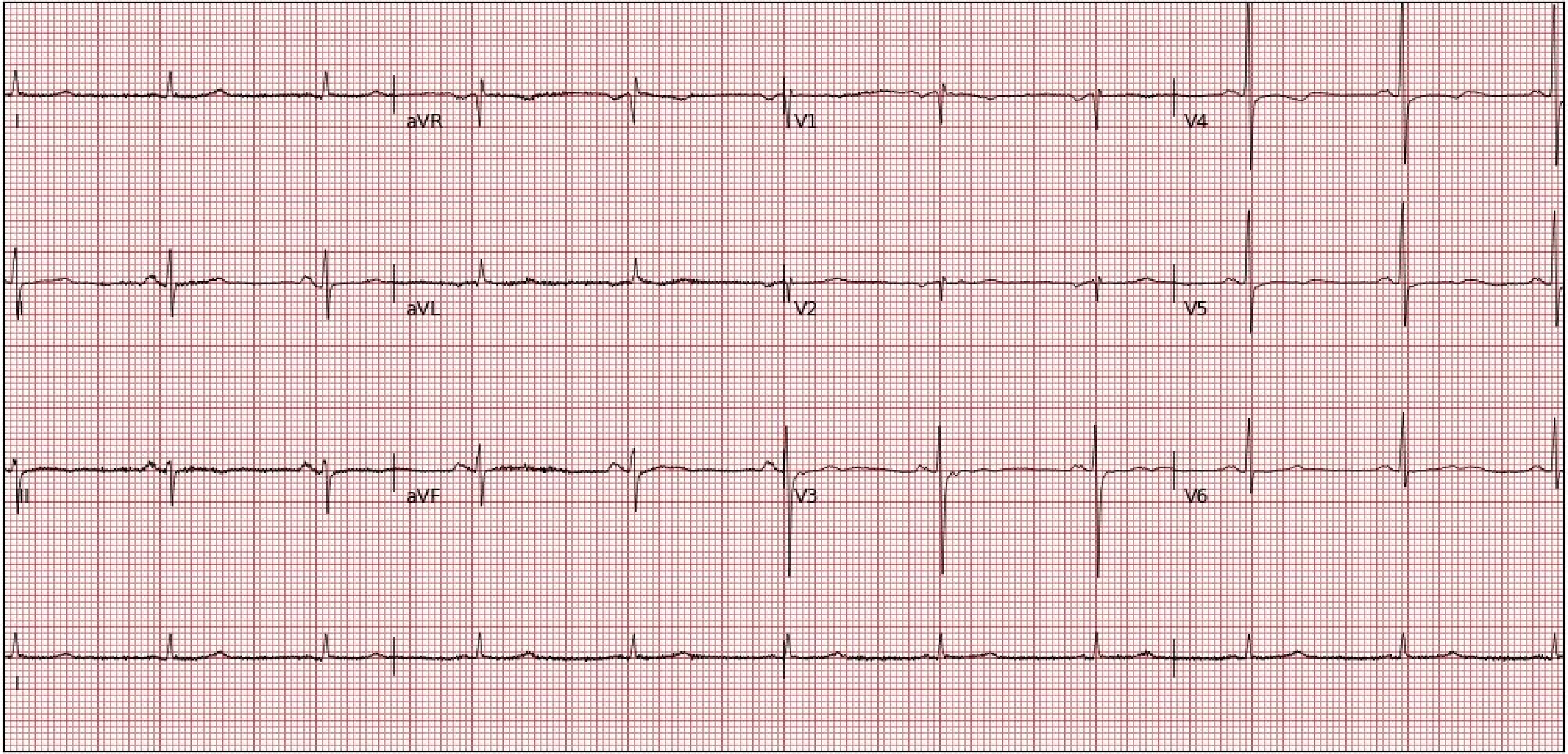
I
aVR
V1
V4
II
aVL
V2
V5
III
aVF
V3
V6
I

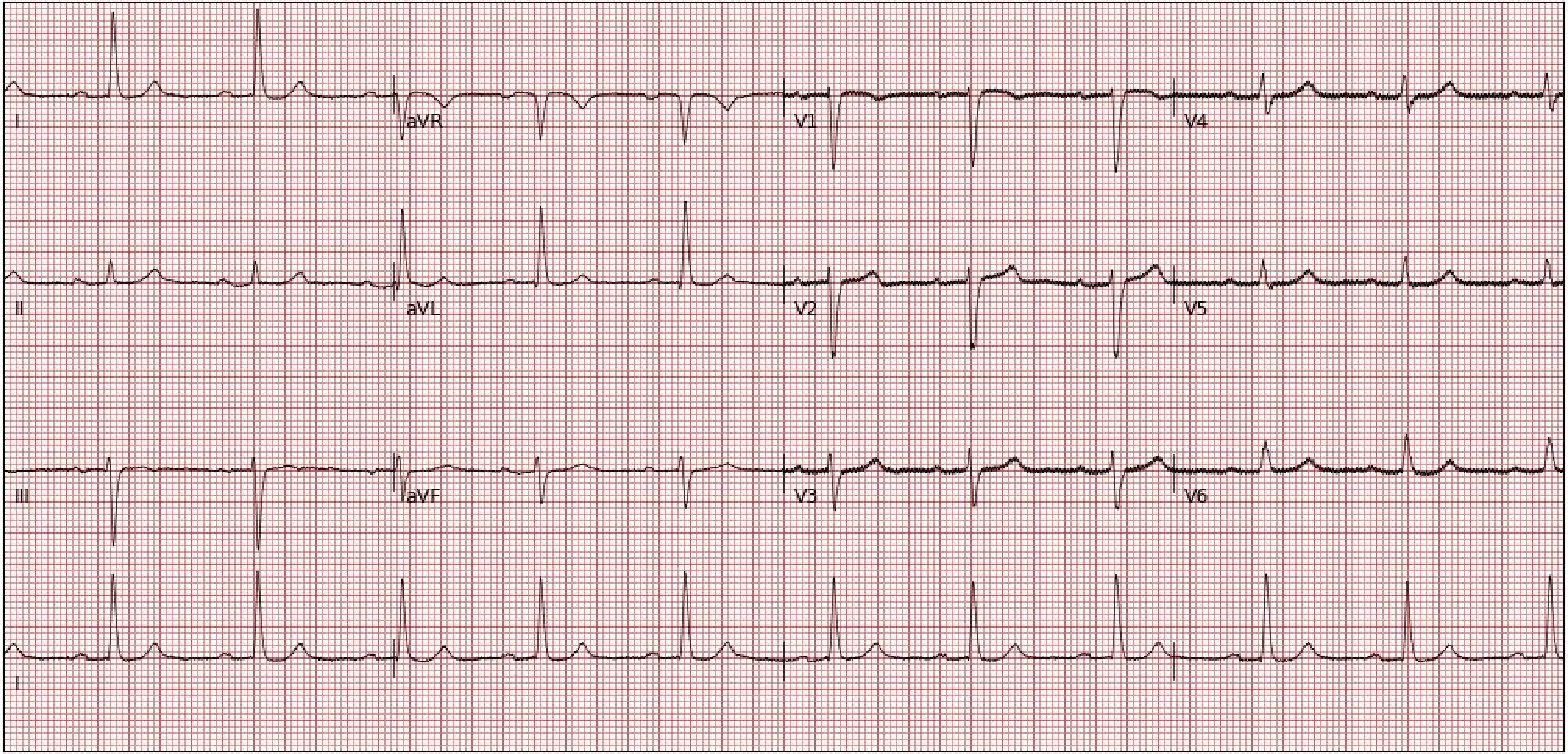
I
aVR
V1
V4
II
aVL
V2
V5
III
aVF
V3
V6
I

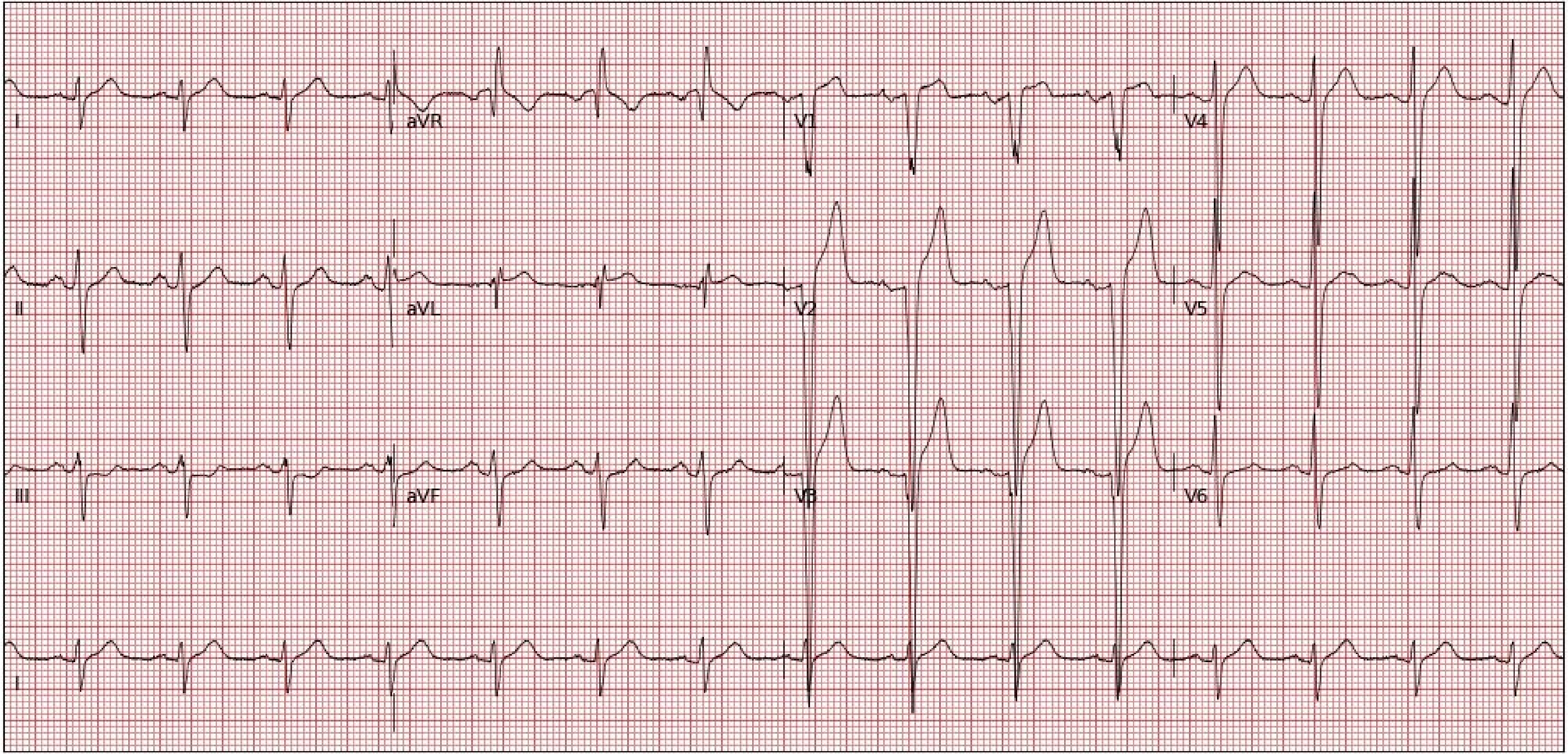
I
aVR
V1
V4
II
aVL
V2
V5
III
aVF
V3
V6
I

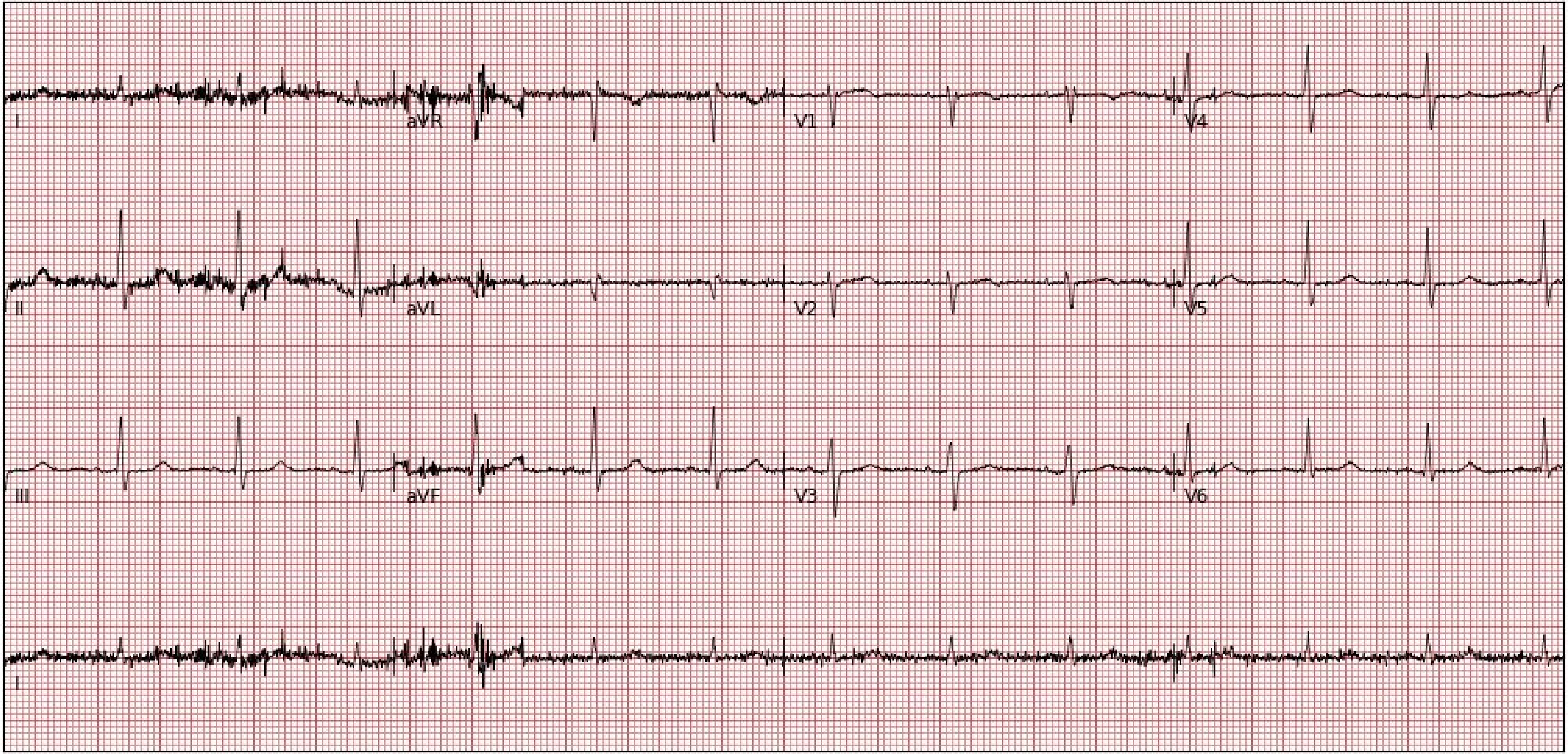
I
aVR
V1
V4
II
aVL
V2
V5
III
aVF
V3
V6
I

**Section 4. Random examples of 25 electrocardiogram images plotted in novel format (Blue-on-Black Standard) not encountered during model development**

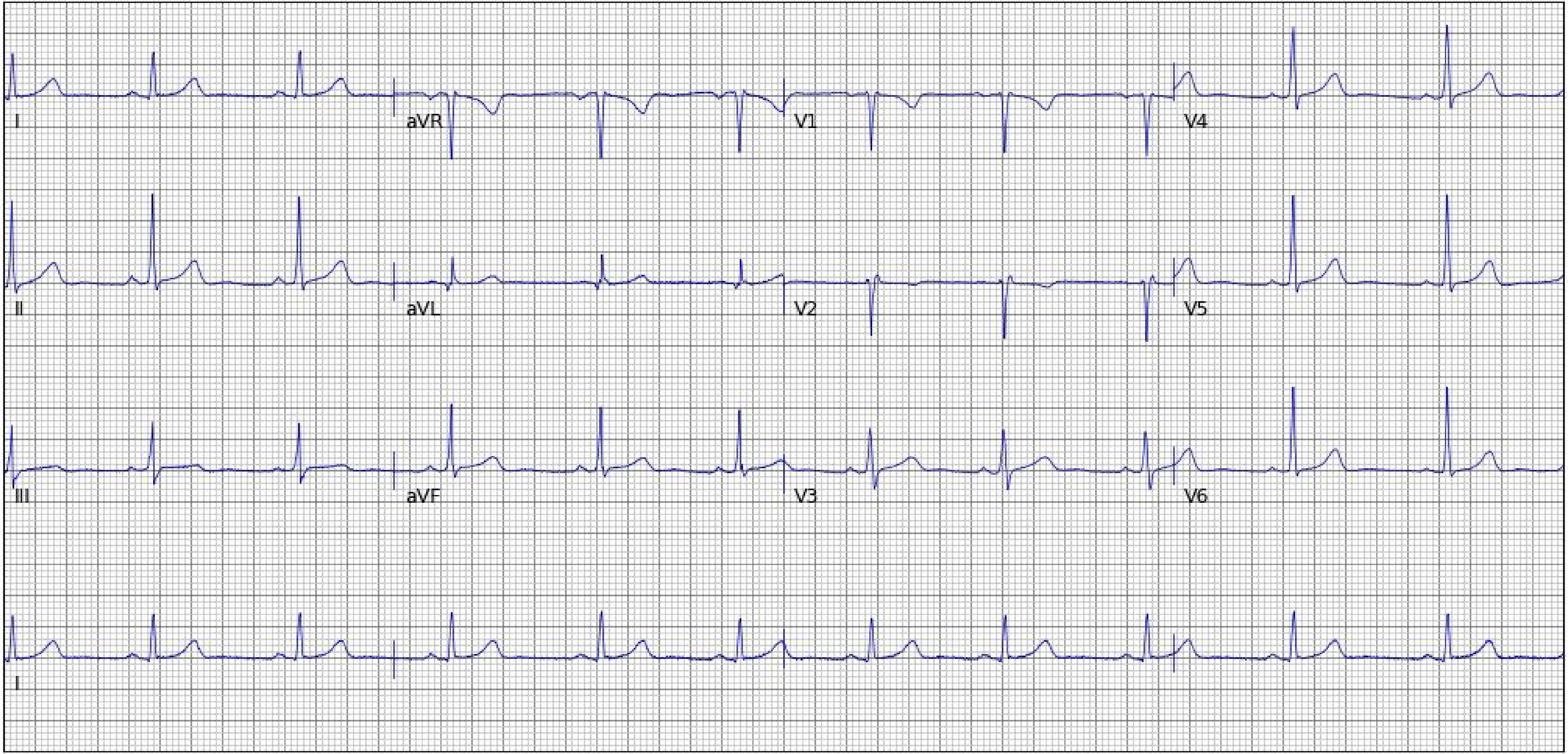
I
aVR
V1
V4
II
aVL
V2
V5
III
aVF
V3
V6
I

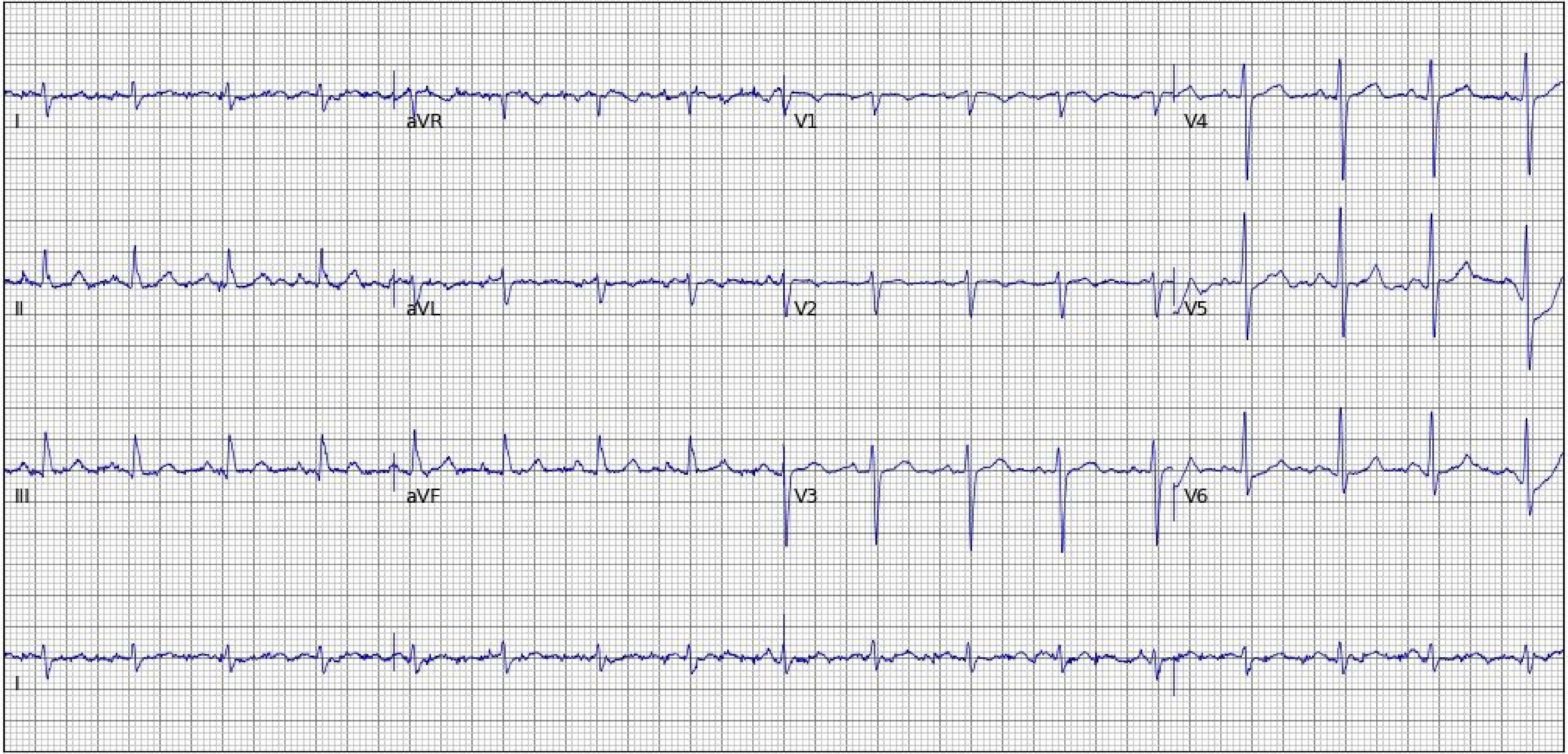
I
aVR
V1
V4
II
aVL
V2
V5
III
aVF
V3
V6
I

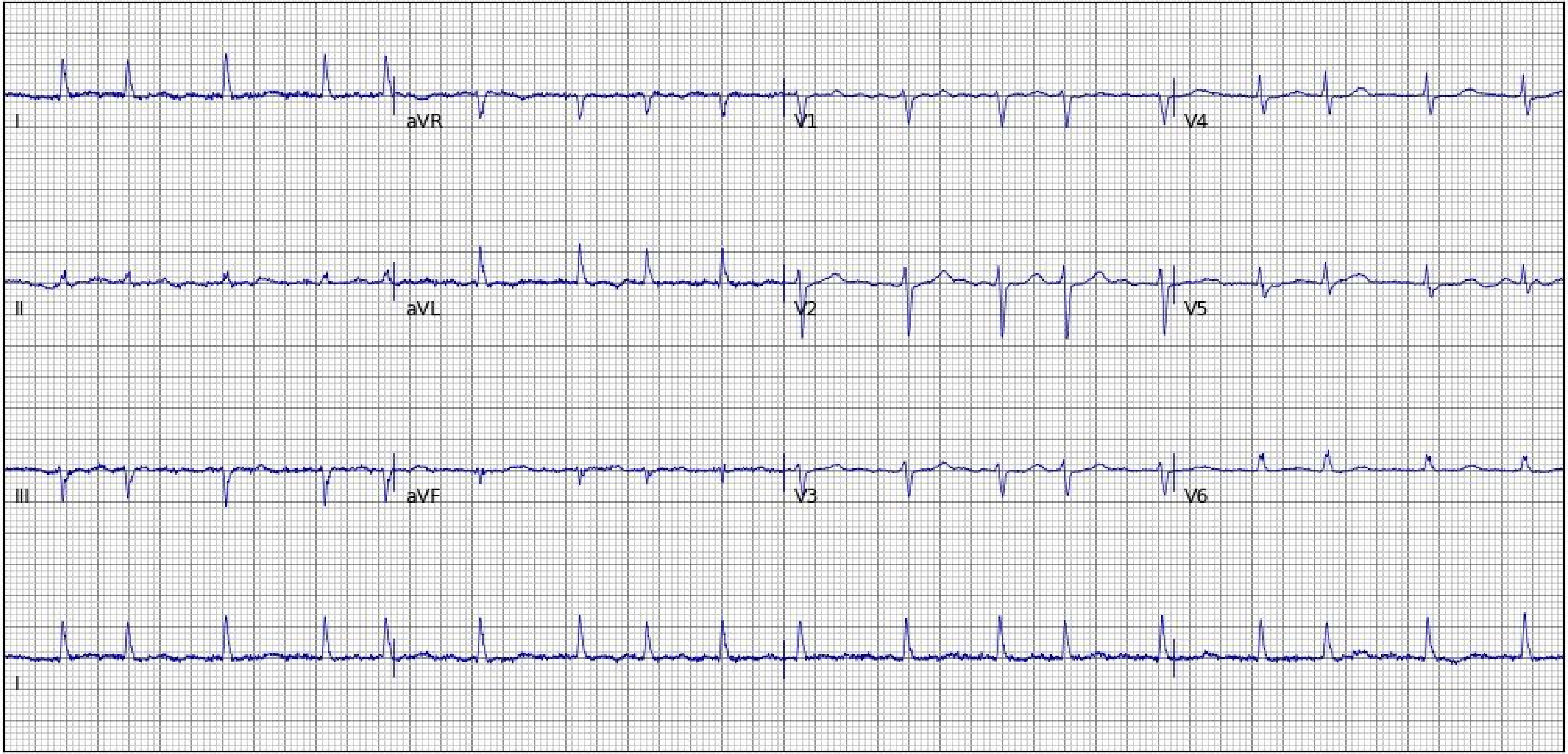
I
aVR
V1
V4
II
aVL
V2
V5
III
aVF
V3
V6
I

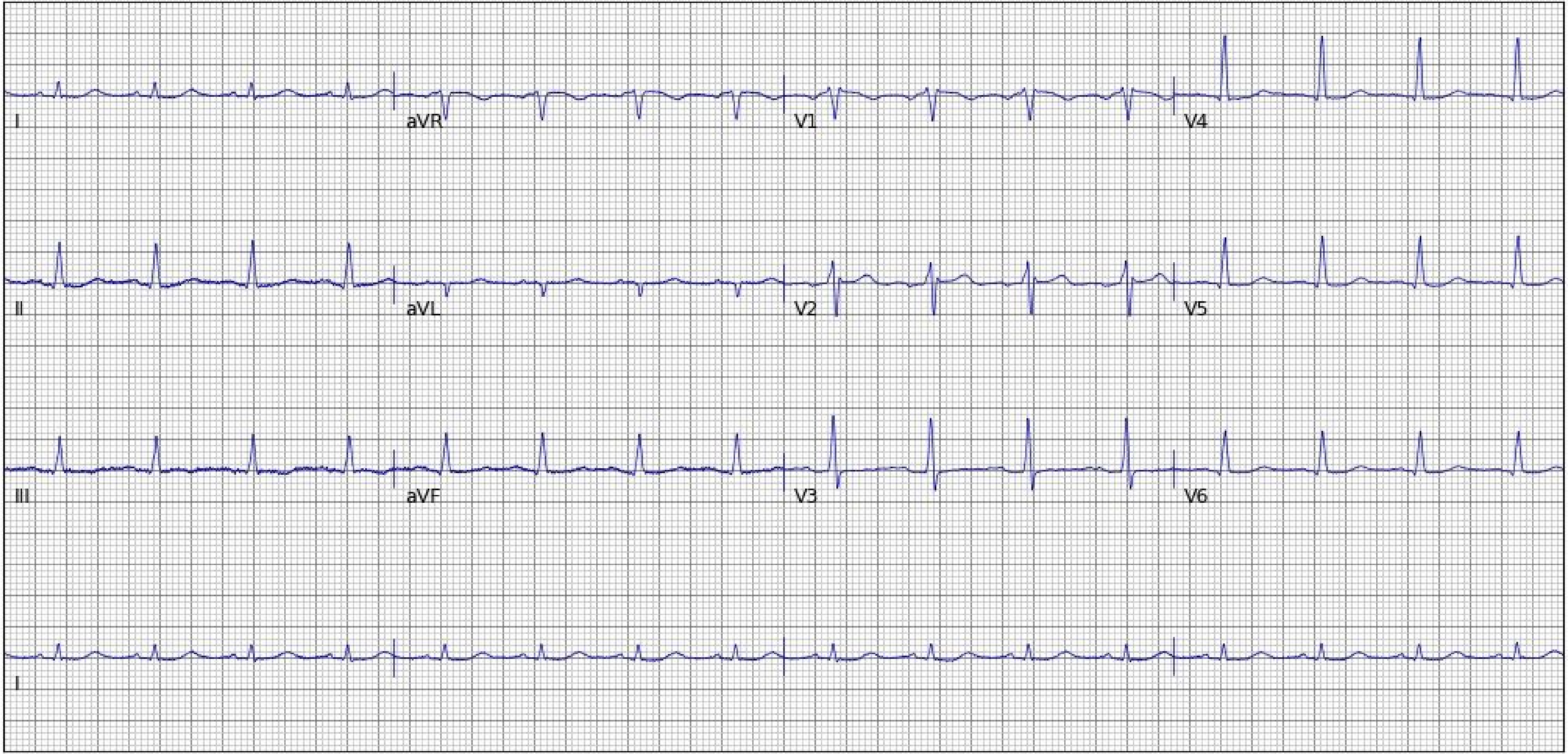
I
aVR
V1
V4
II
aVL
V2
V5
III
aVF
V3
V6
I

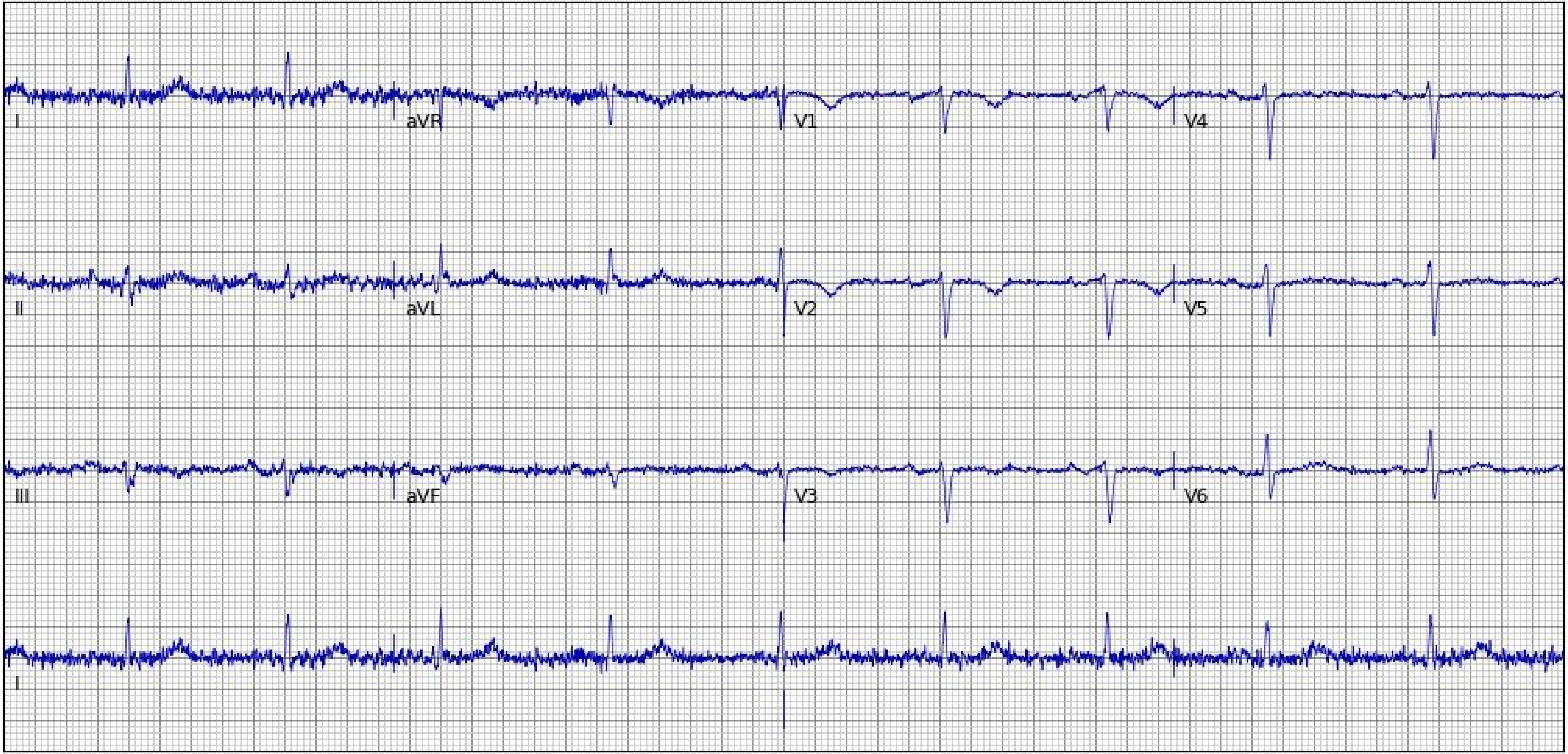
I
aVR
V1
V4
II
aVL
V2
V5
III
aVF
V3
V6
II

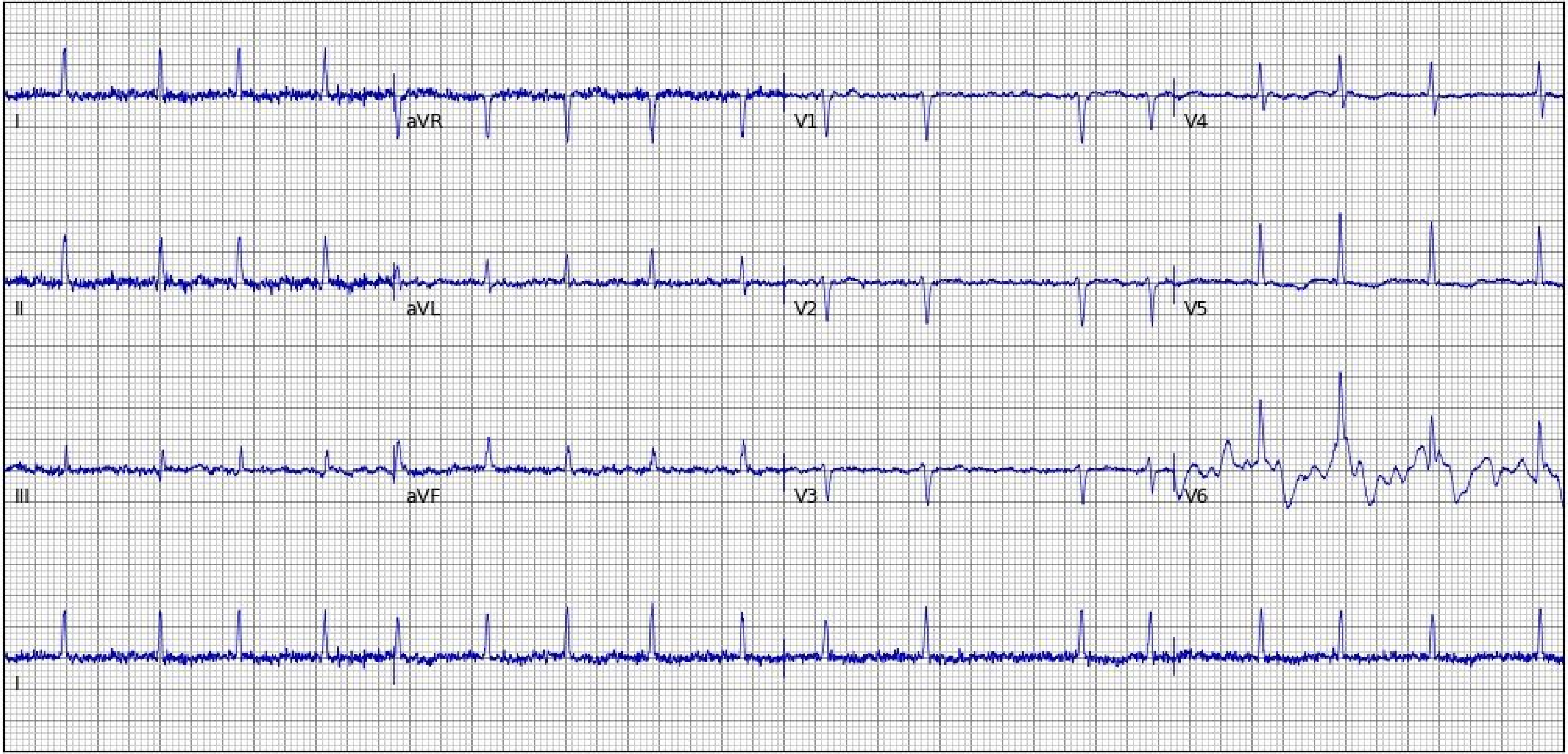
I
aVR
V1
V4
II
aVL
V2
V5
III
aVF
V3
V6
I

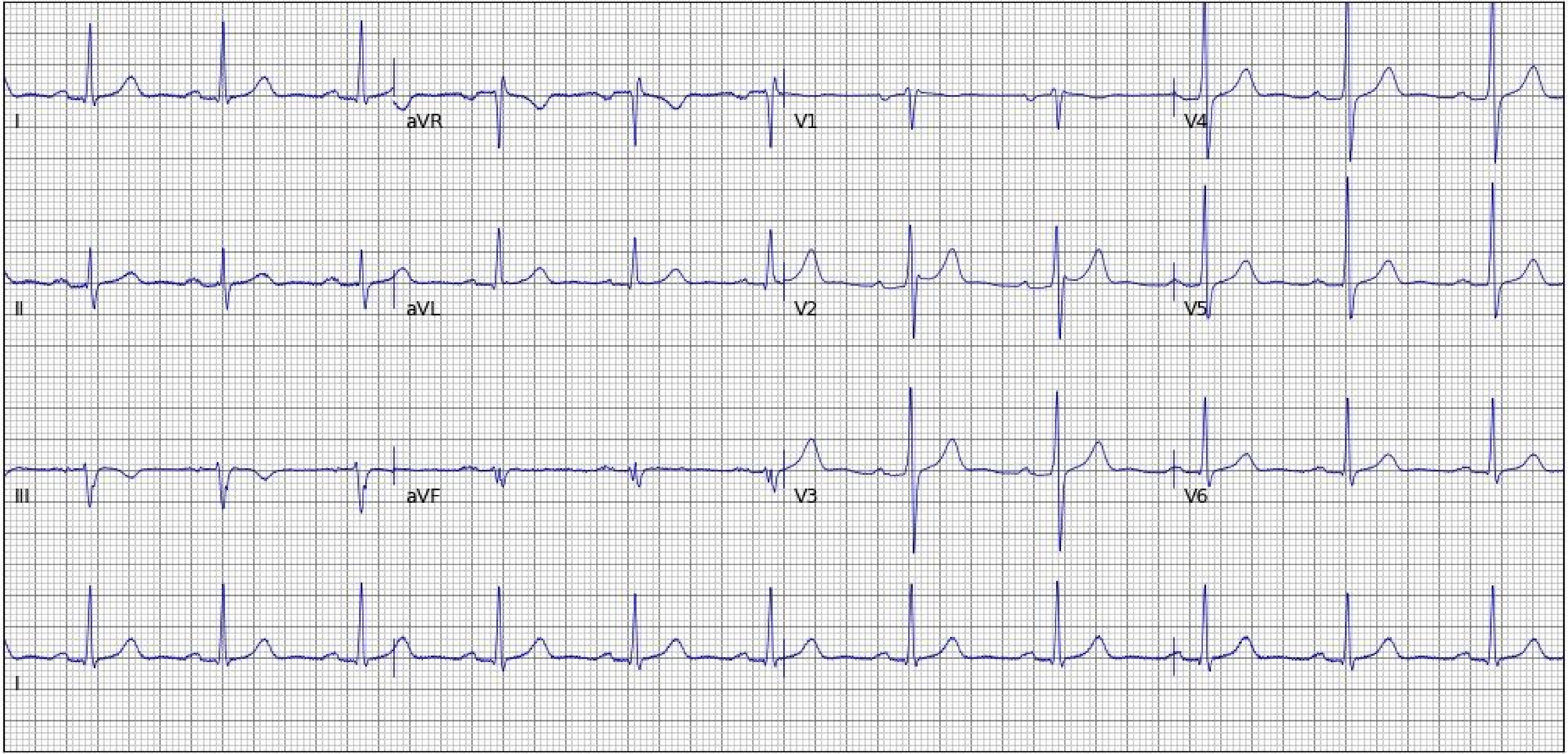
I
aVR
V1
V4
II
aVL
V2
V5
III
aVF
V3
V6
I

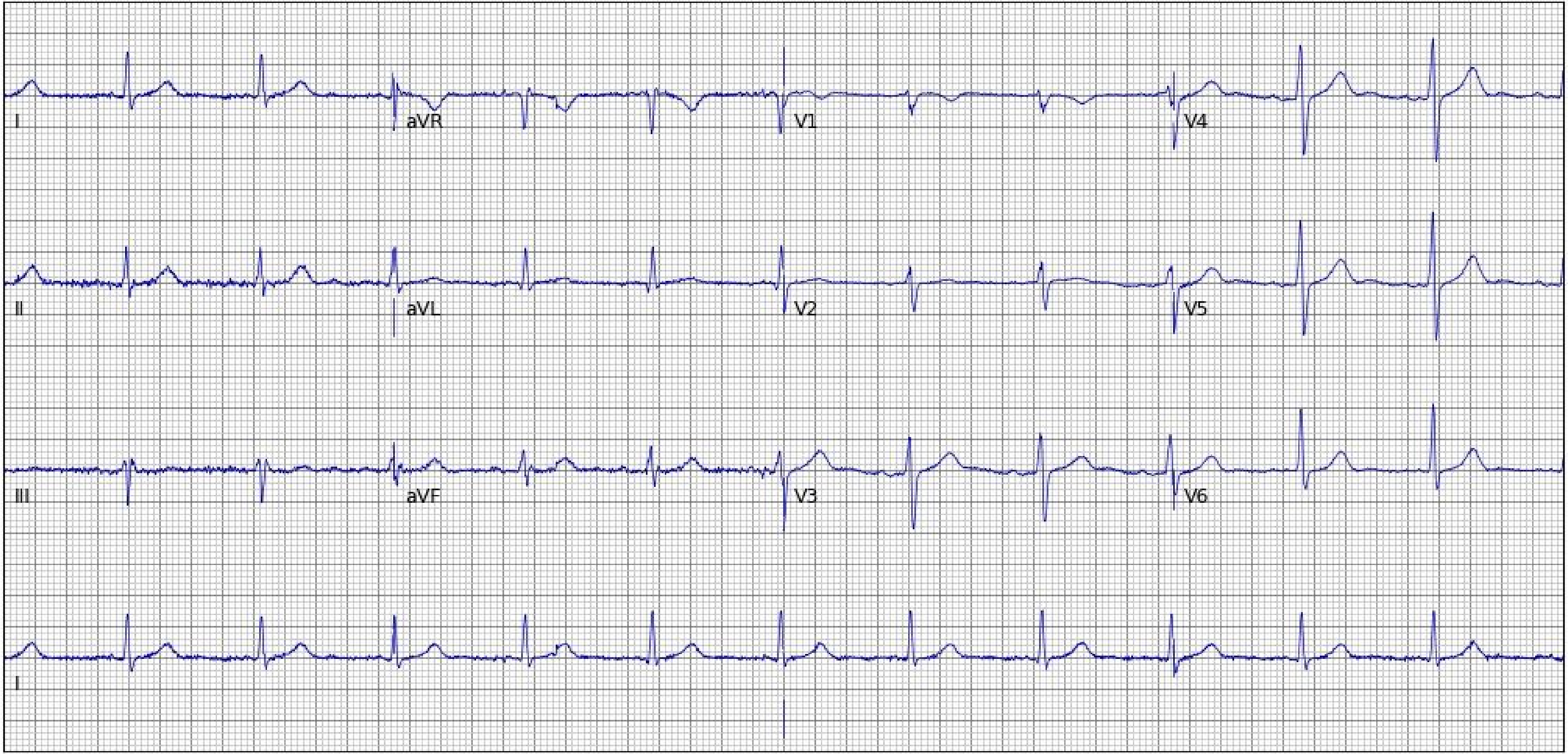
I
aVR
V1
V4
II
aVL
V2
V5
III
aVF
V3
V6
I

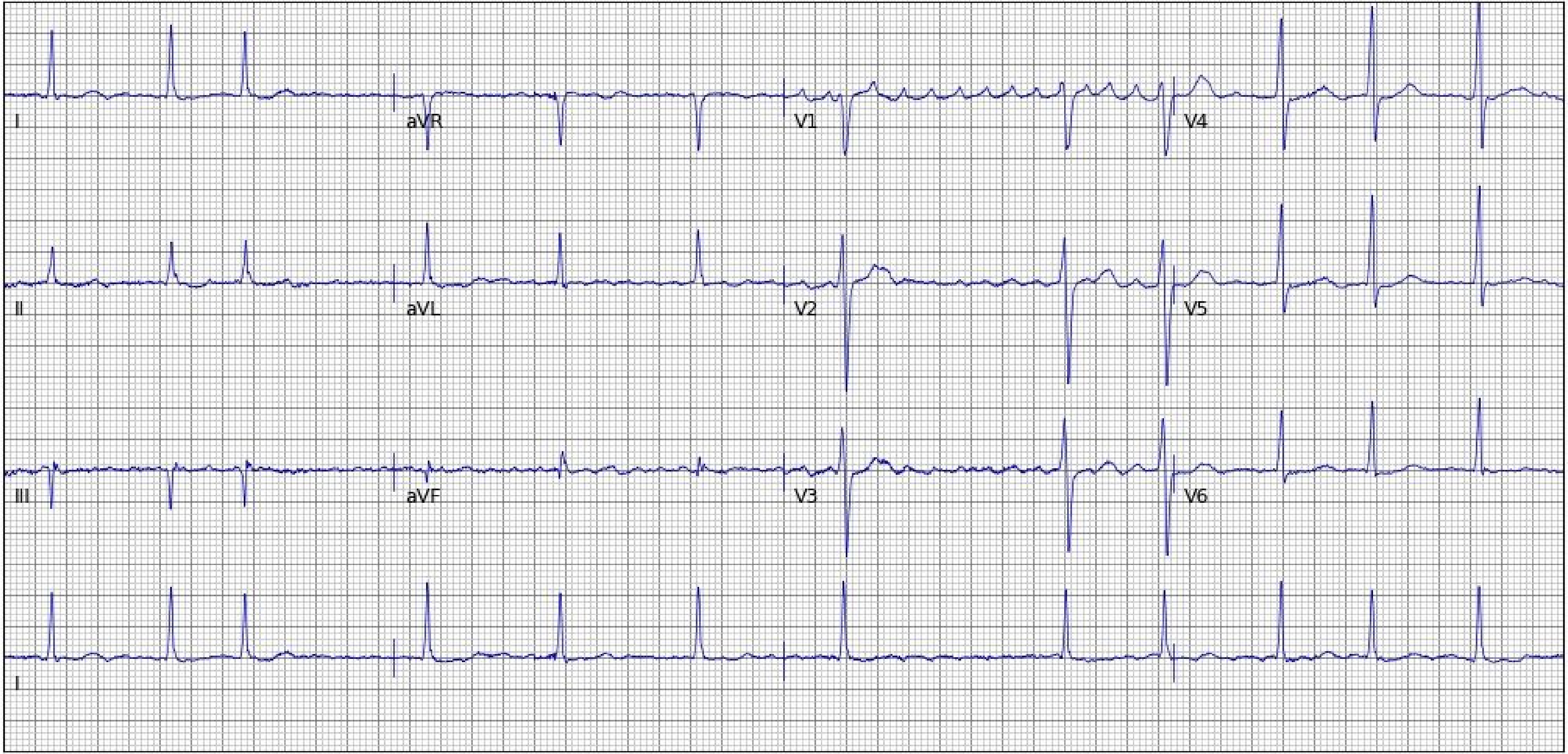
I
aVR
V1
V4
II
aVL
V2
V5
III
aVF
V3
V6
I

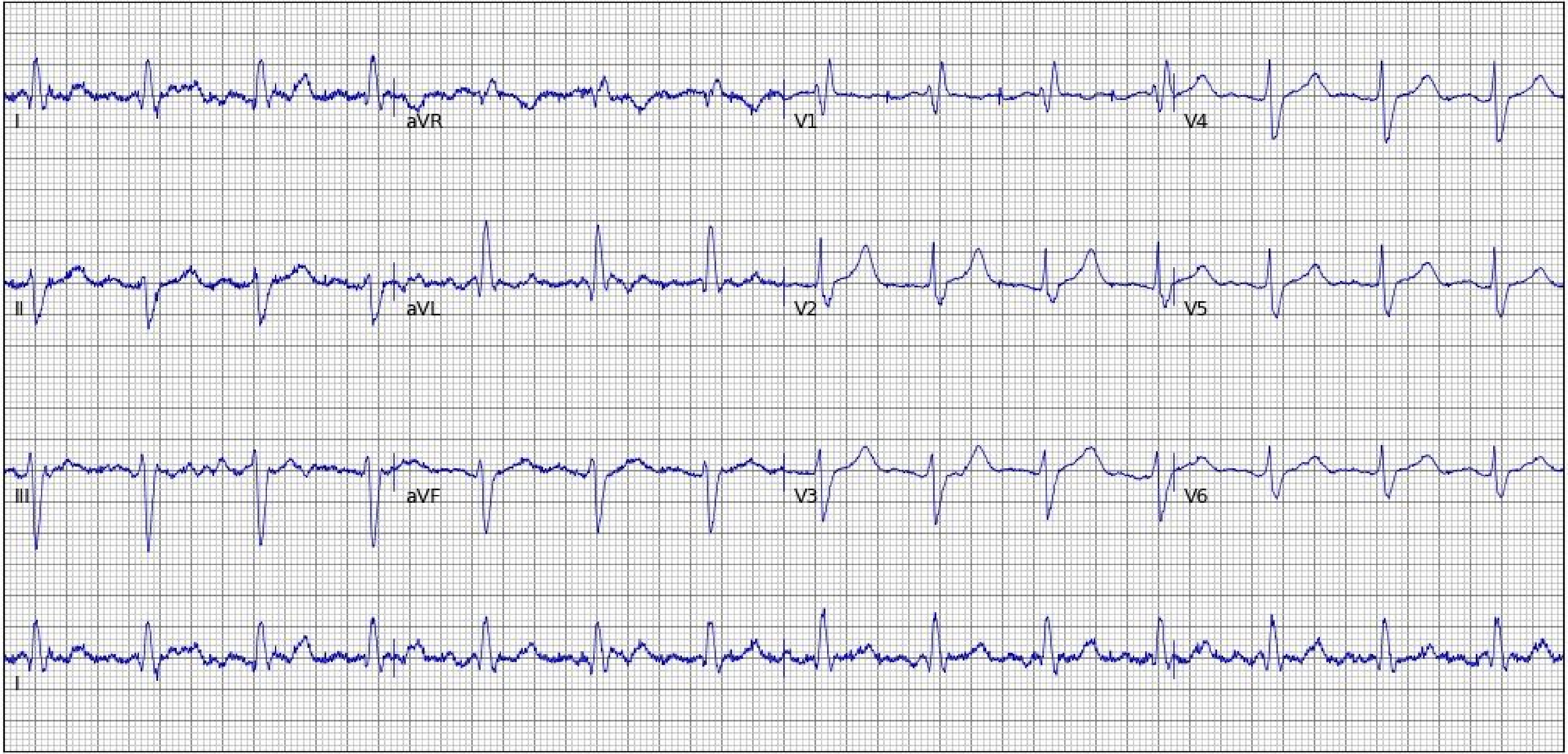
I
aVR
V1
V4
II
aVL
V2
V5
III
aVF
V3
V6
I

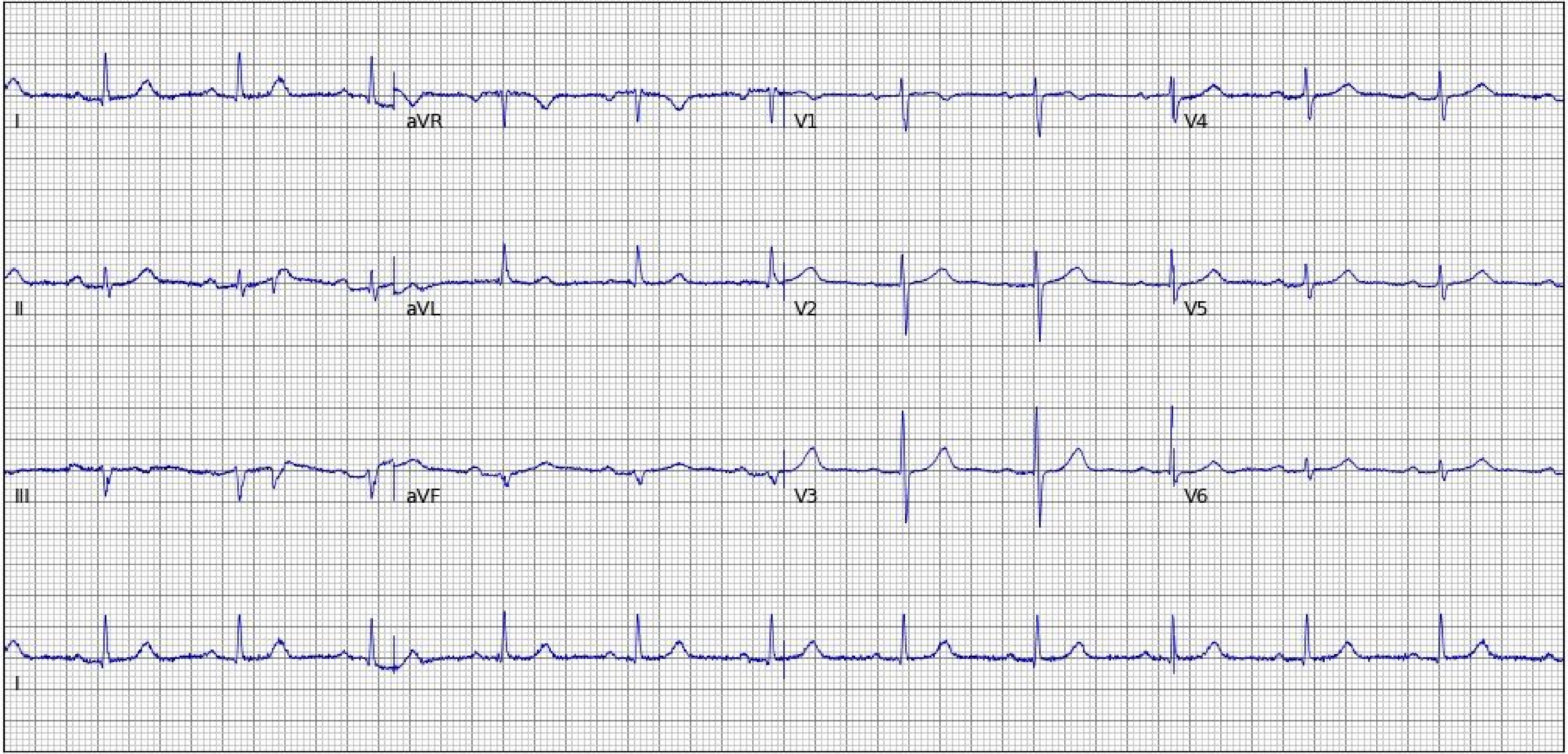
I
aVR
V1
V4
II
aVL
V2
V5
III
aVF
V3
V6
I

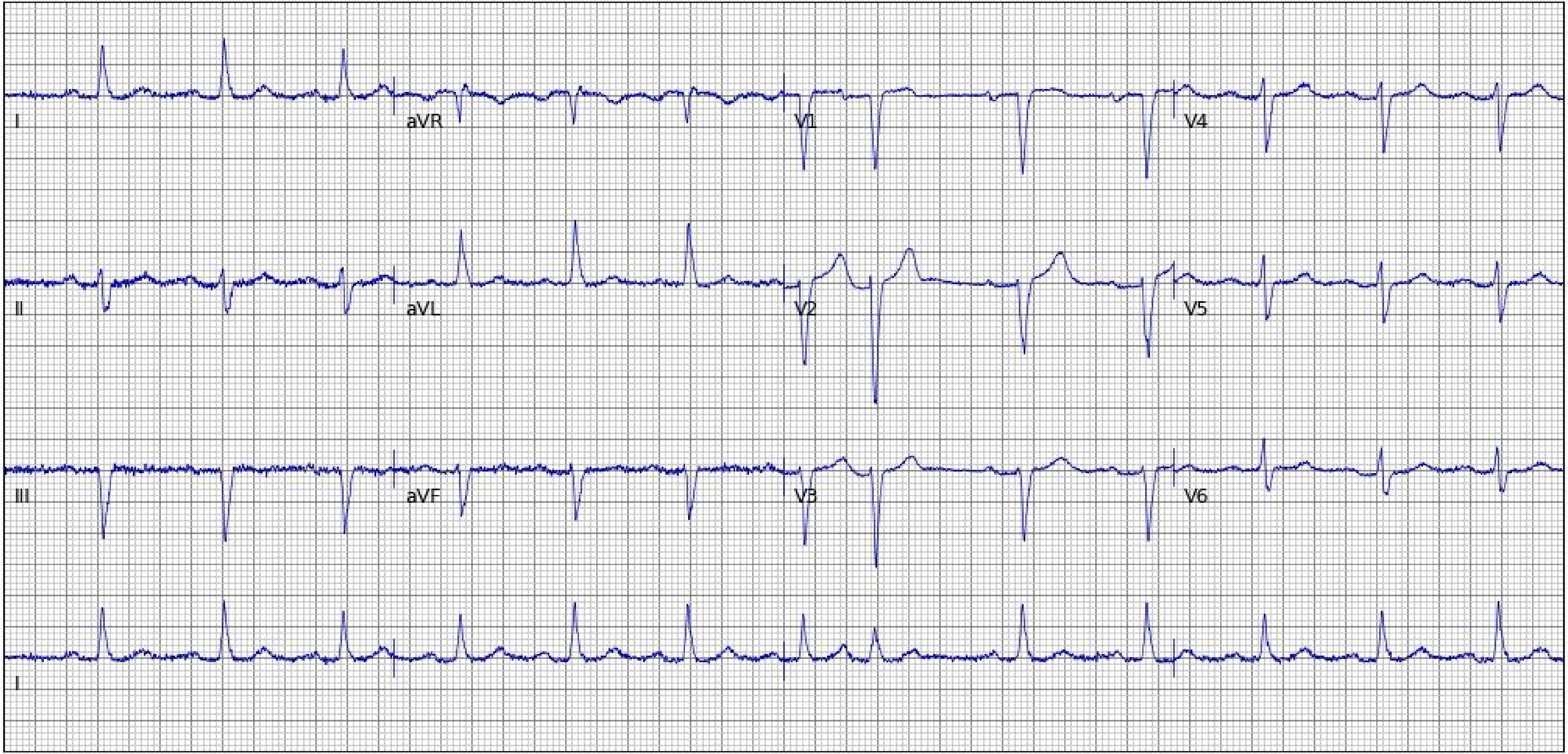
I
aVR
V1
V4
II
aVL
V2
V5
III
aVF
V3
V6
I

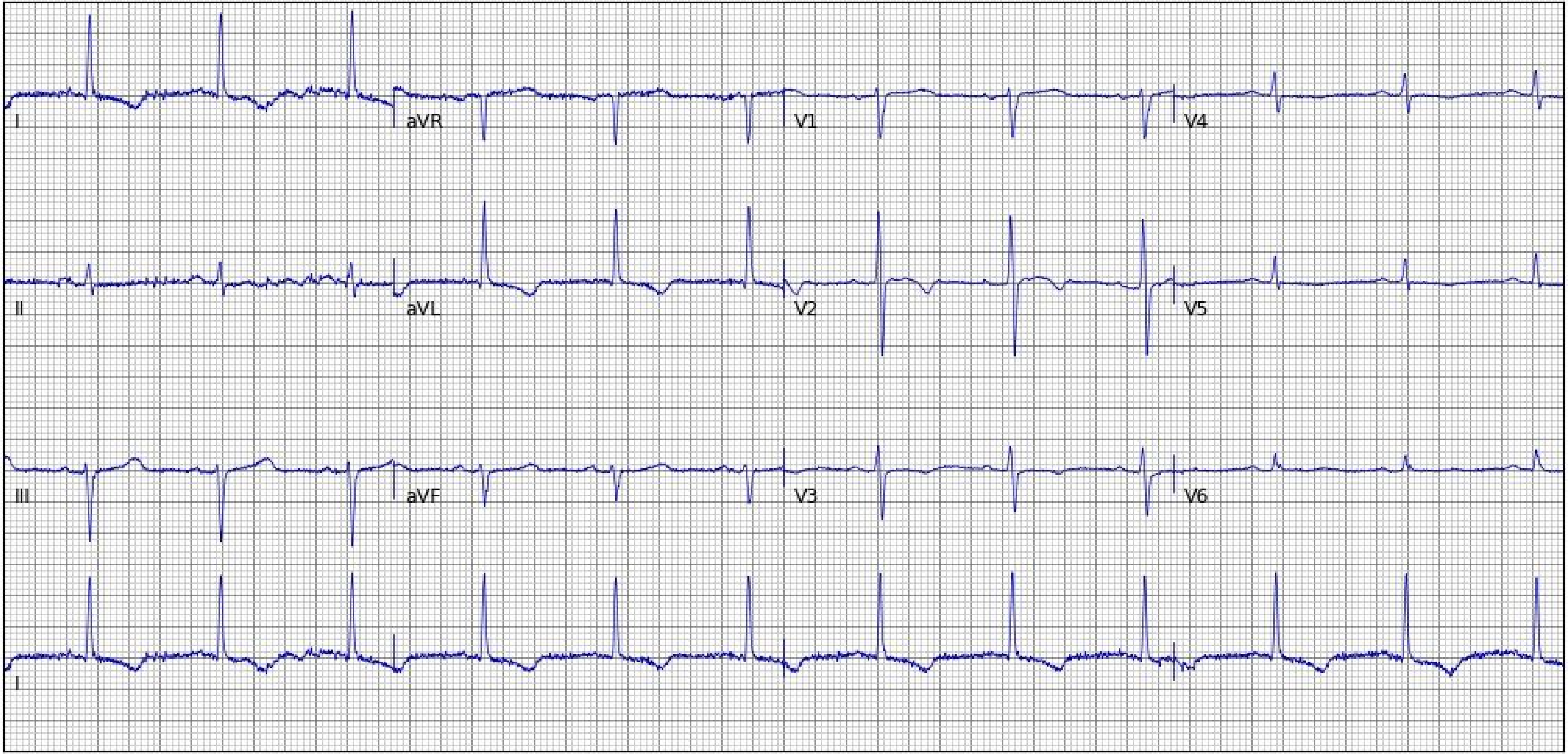
I
aVR
V1
V4
II
aVL
V2
V5
III
aVF
V3
V6
I

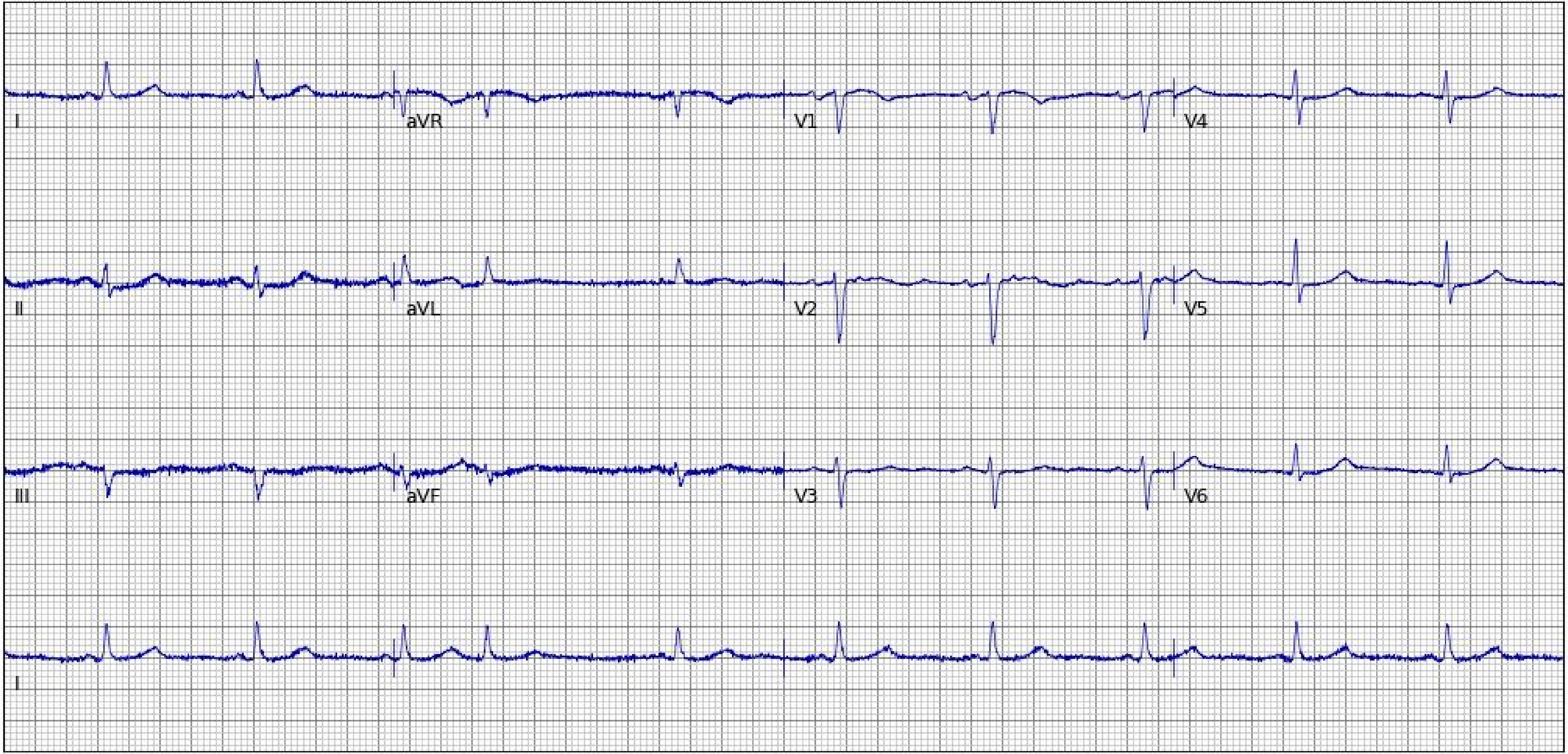
I
aVR
V1
V4
II
aVL
V2
V5
III
aVF
V3
V6
I

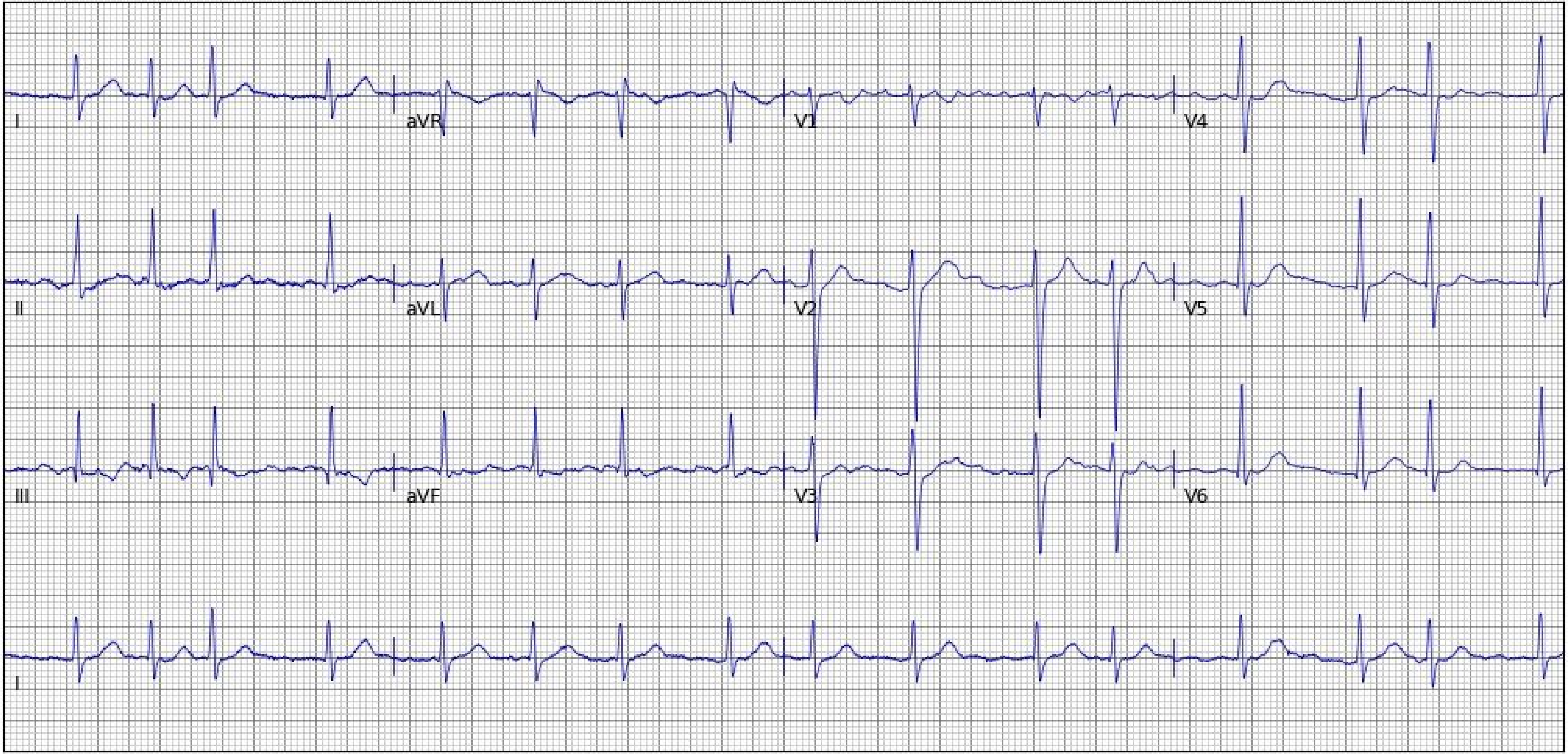

I
aVR
V1
V4
II
aVL
V2
V5
III
aVF
V3
V6
II

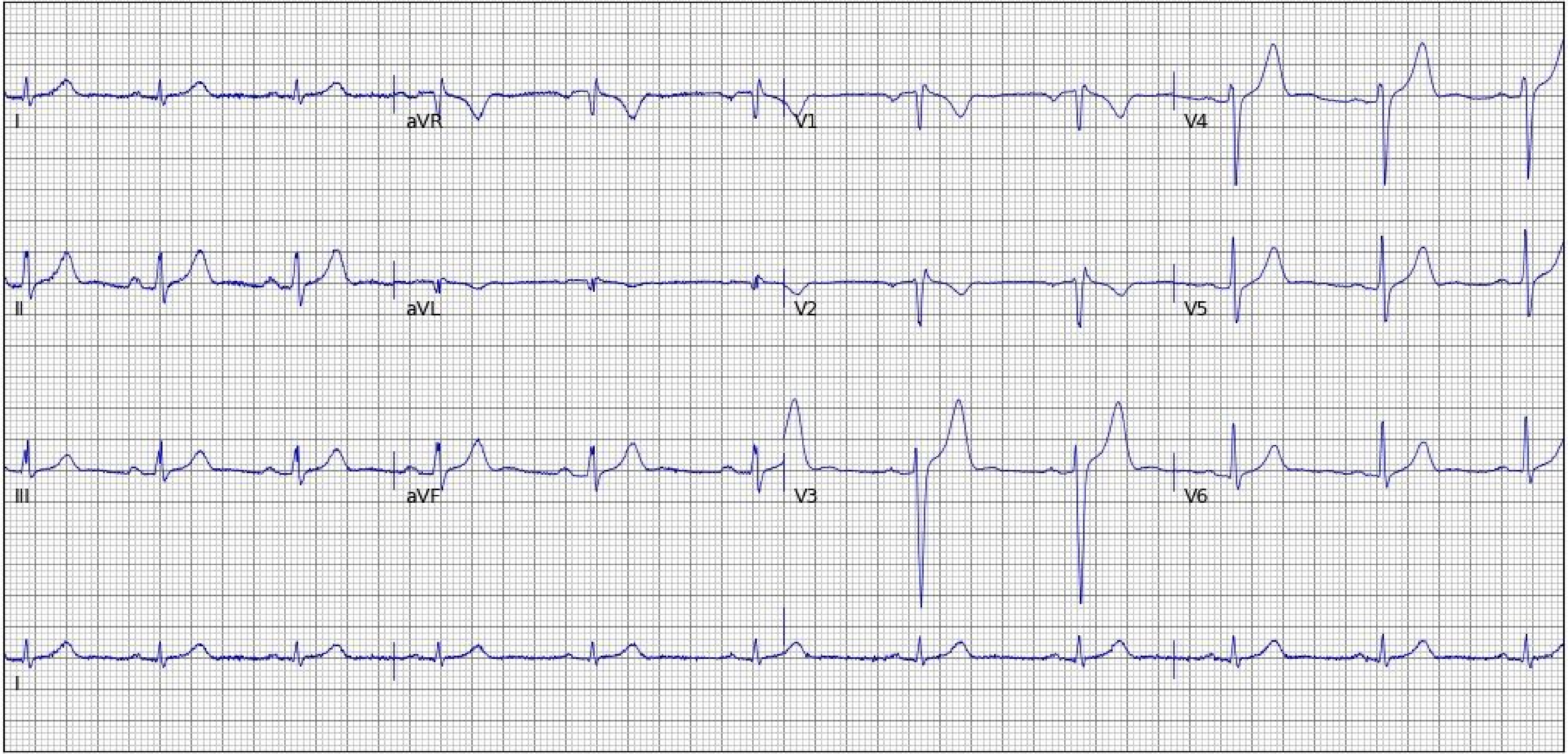
I
aVR
V1
V4
II
aVL
V2
V5
III
aVF
V3
V6
I

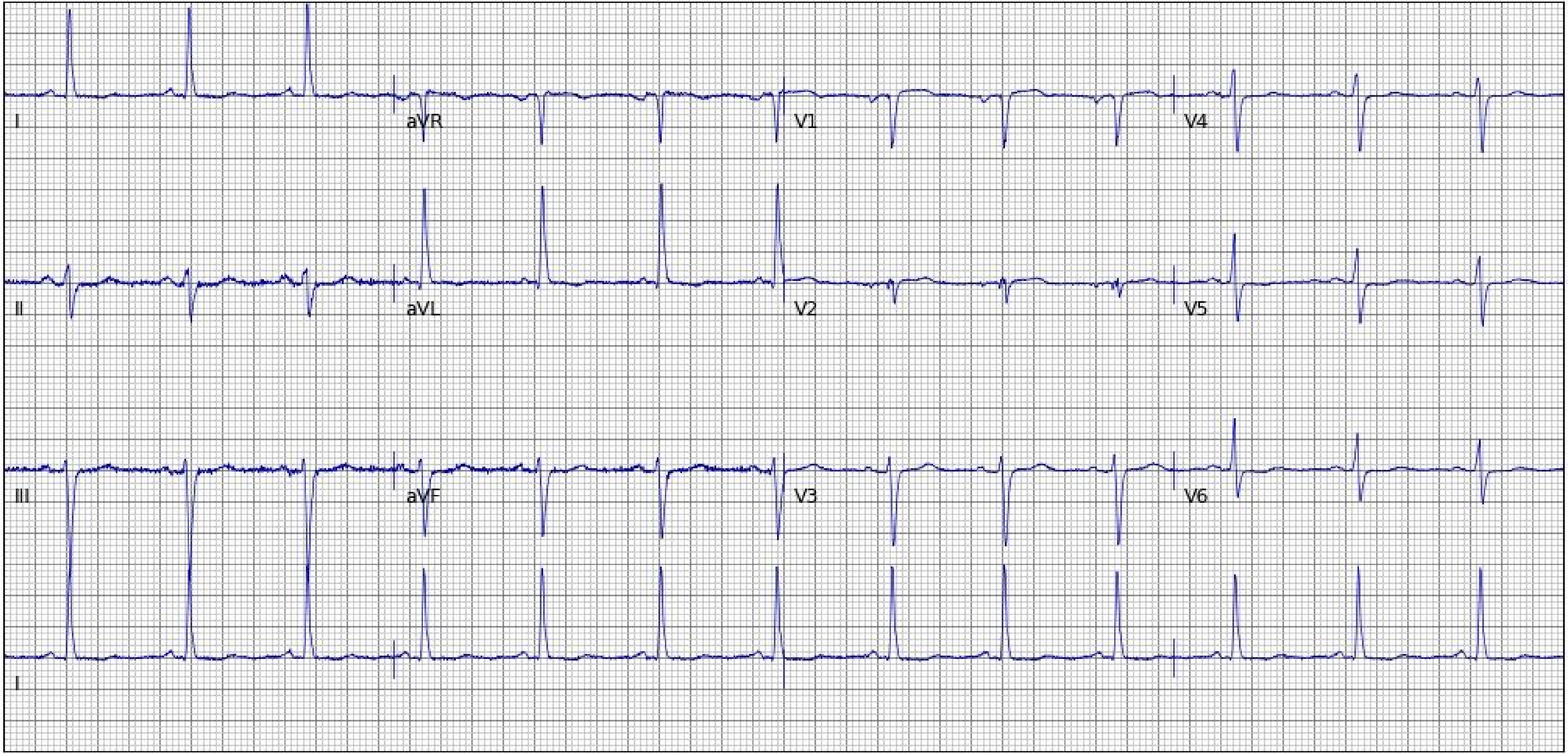

I
aVR
V1
V4
II
aVL
V2
V5
III
aVF
V3
V6
I

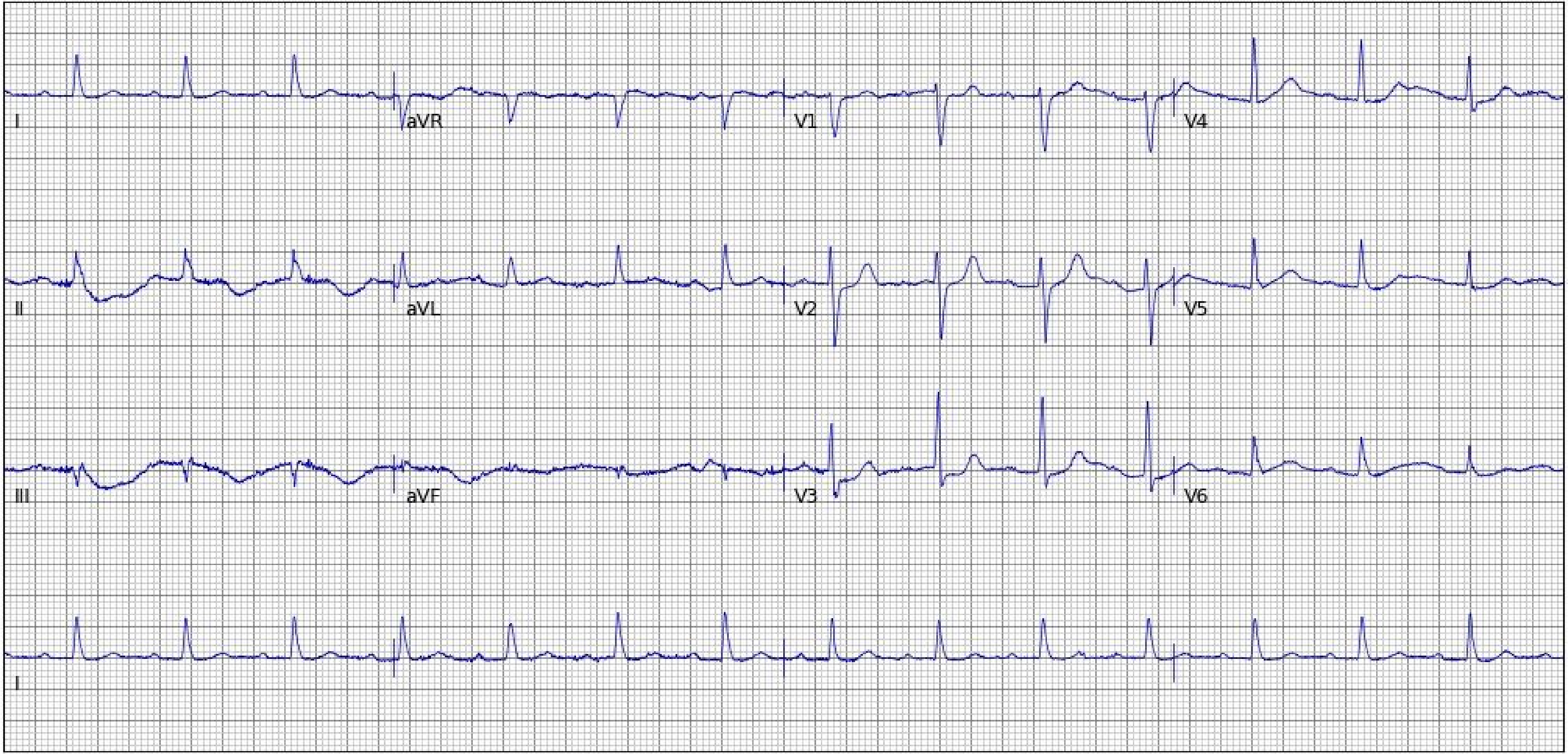
I
aVR
V1
V4
II
aVL
V2
V5
III
aVF
V3
V6
I

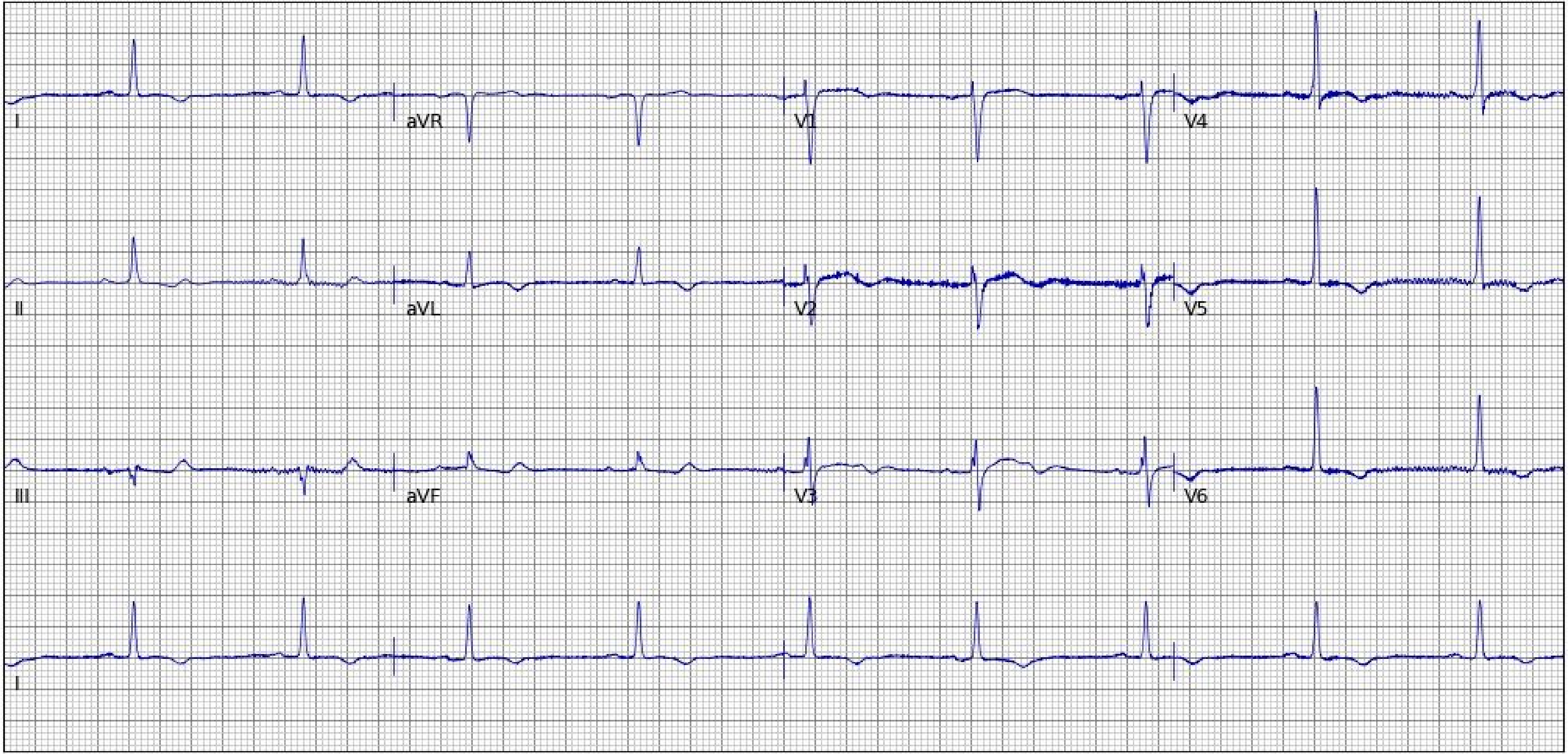
I
aVR
V1
V4
II
aVL
V2
V5
III
aVF
V3
V6
I

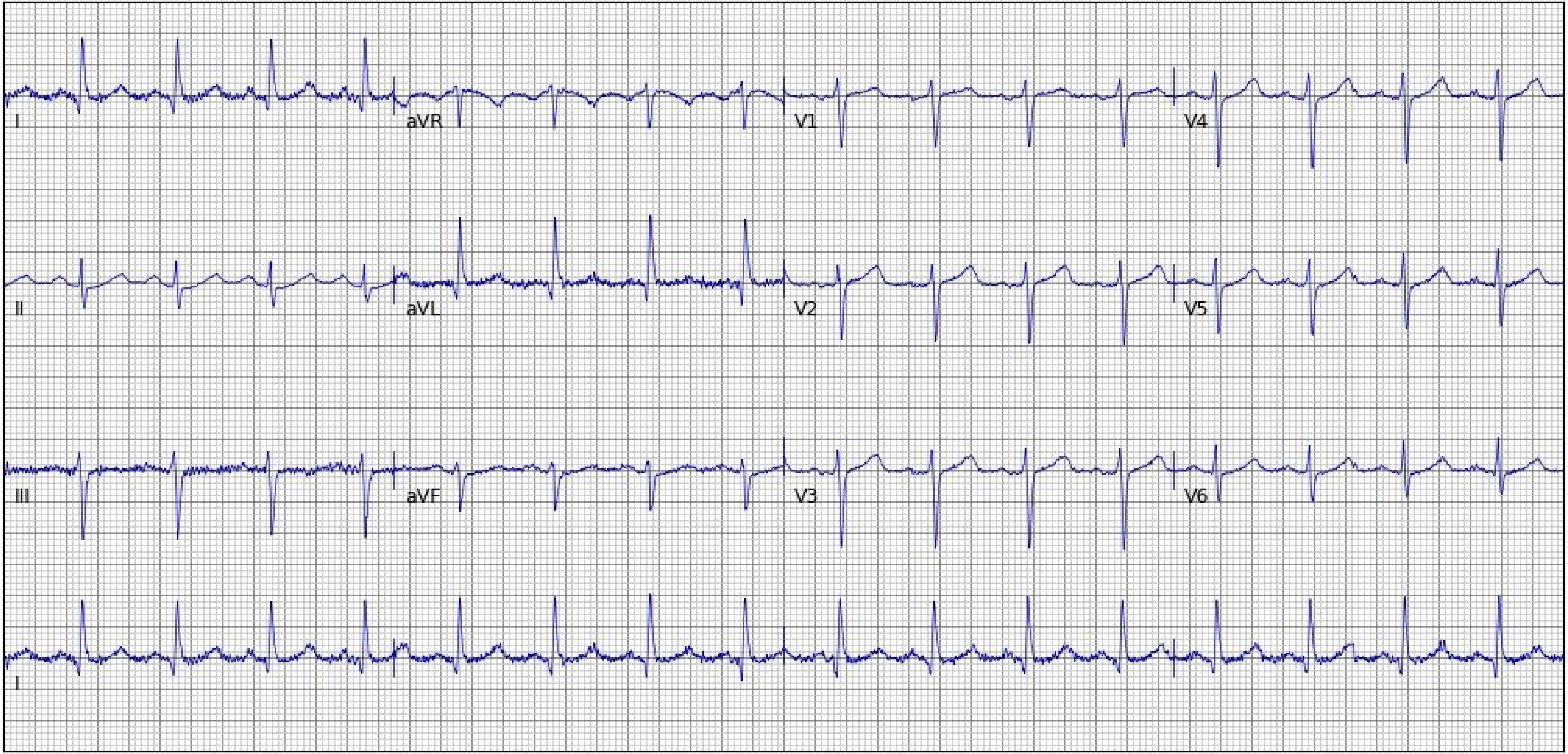
I
aVR
V1
V4
II
aVL
V2
V5
III
aVF
V3
V6
II

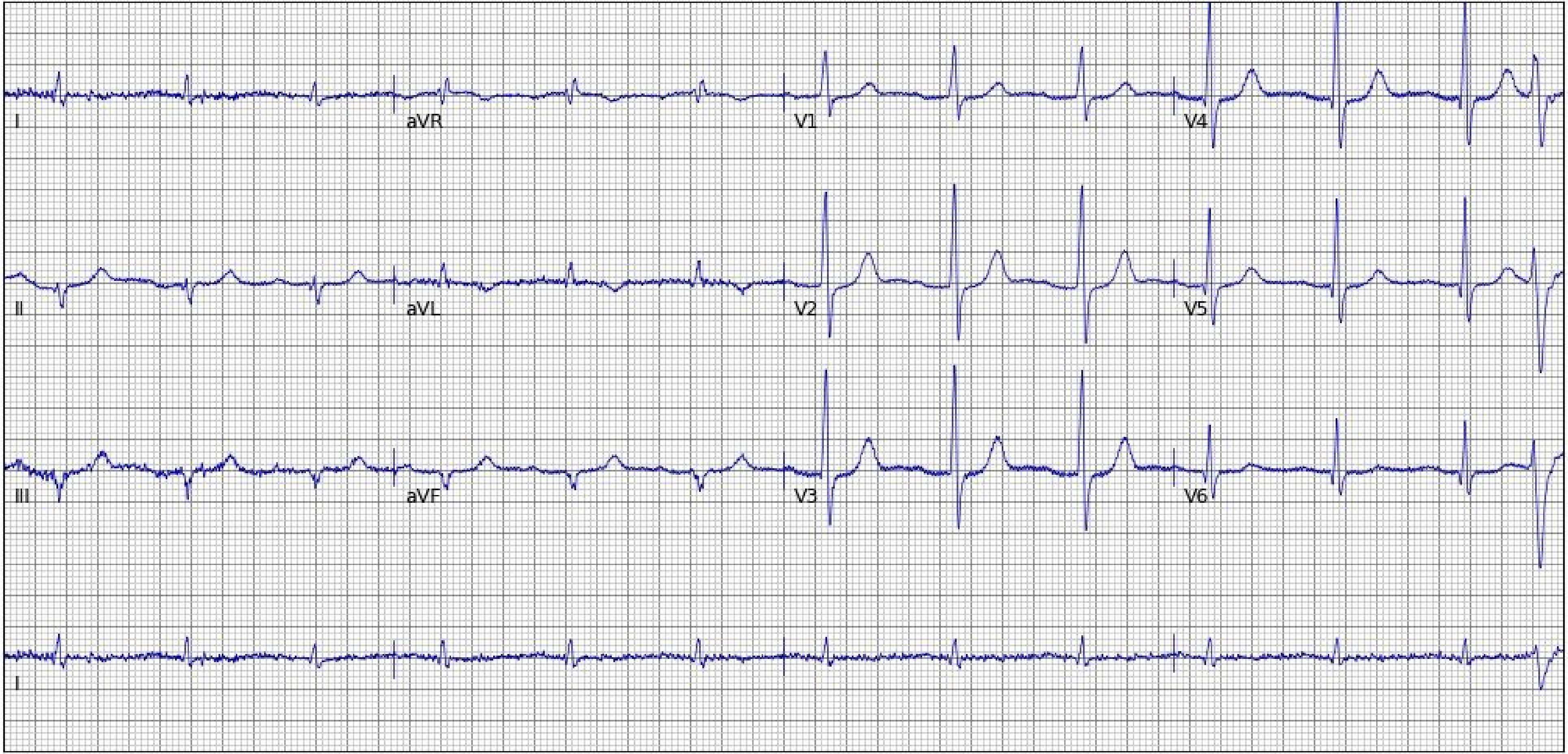
I
aVR
V1
V4
II
aVL
V2
V5
III
aVF
V3
V6
I

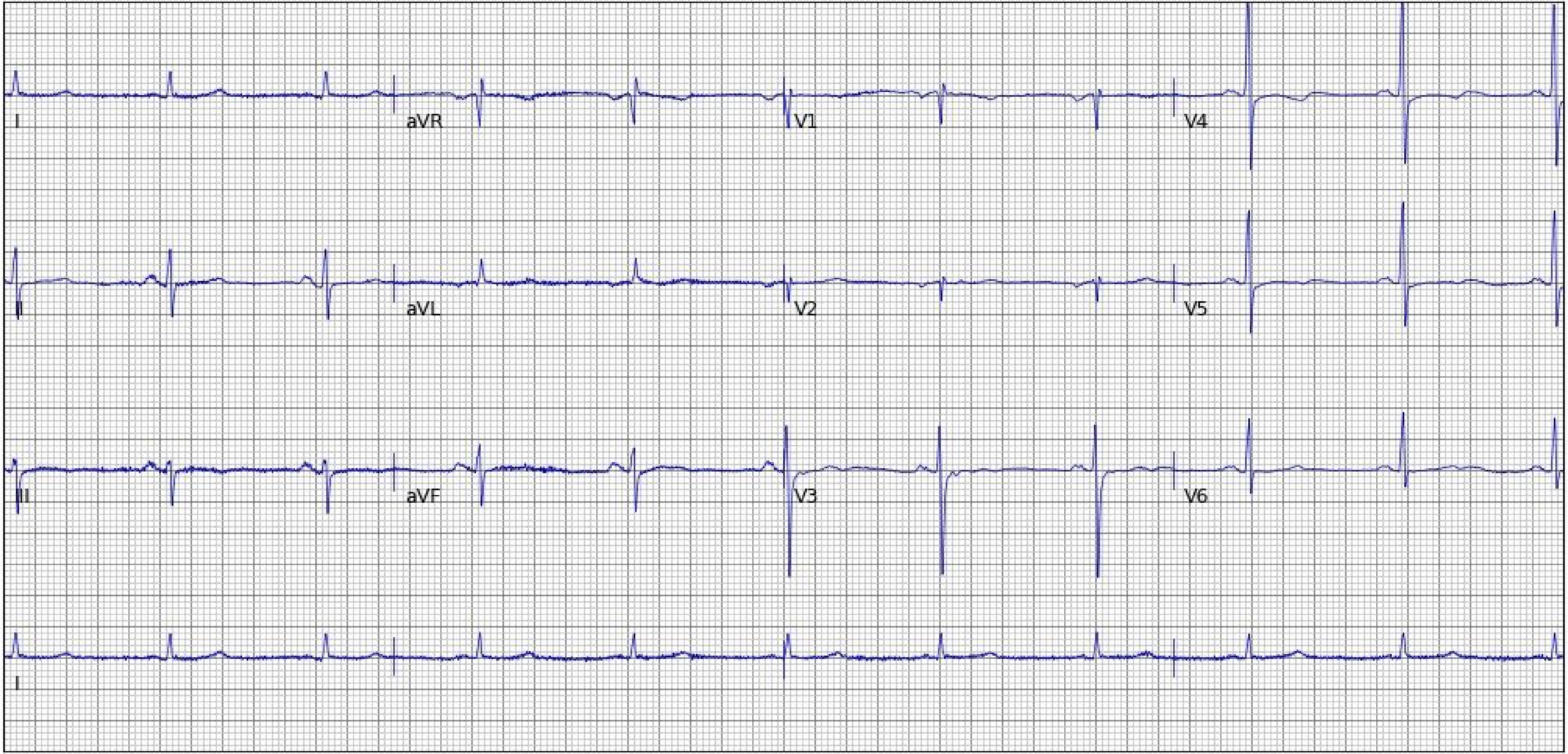
I
aVR
V1
V4
II
aVL
V2
V5
III
aVF
V3
V6
I

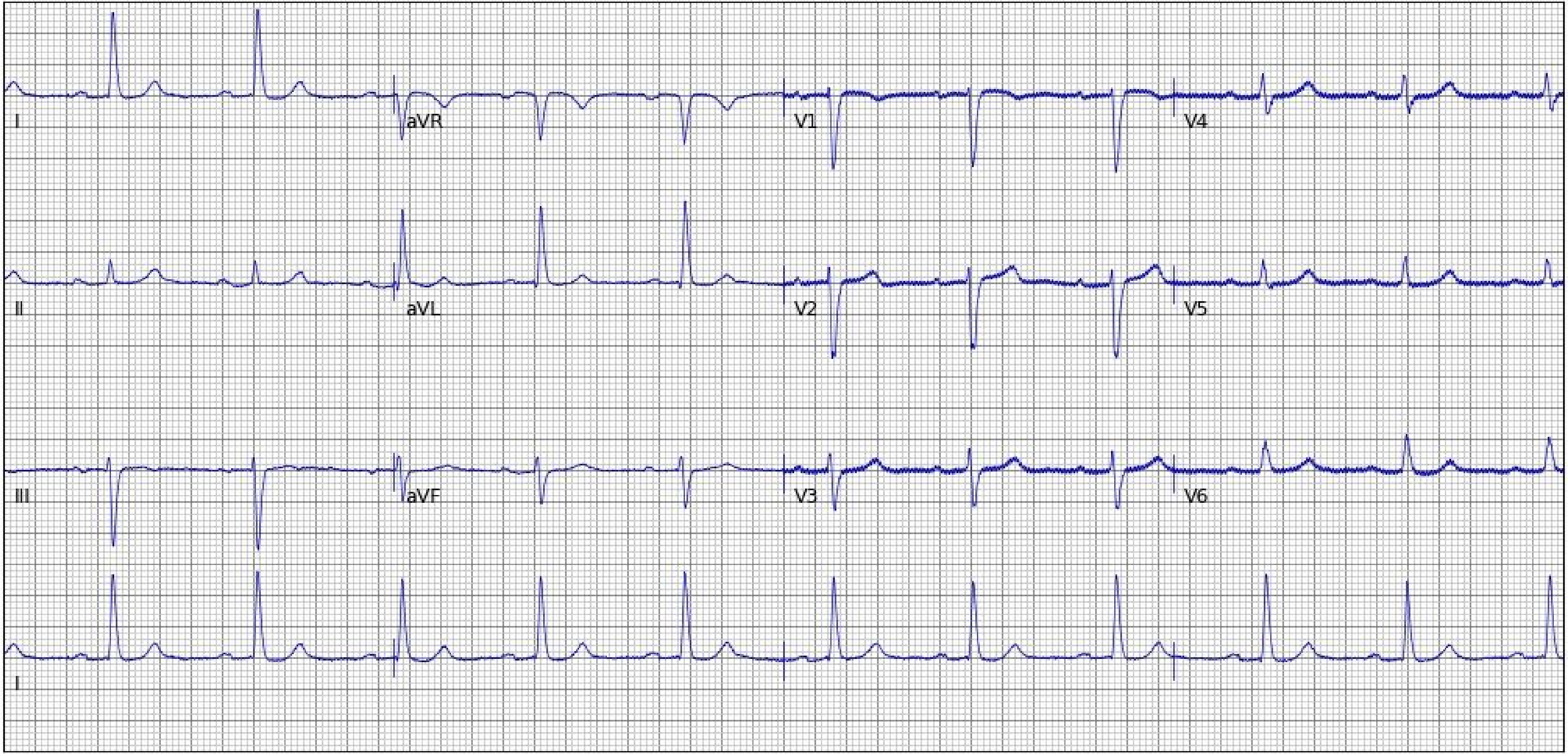
I
aVR
V1
V4
II
aVL
V2
V5
III
aVF
V3
V6
I

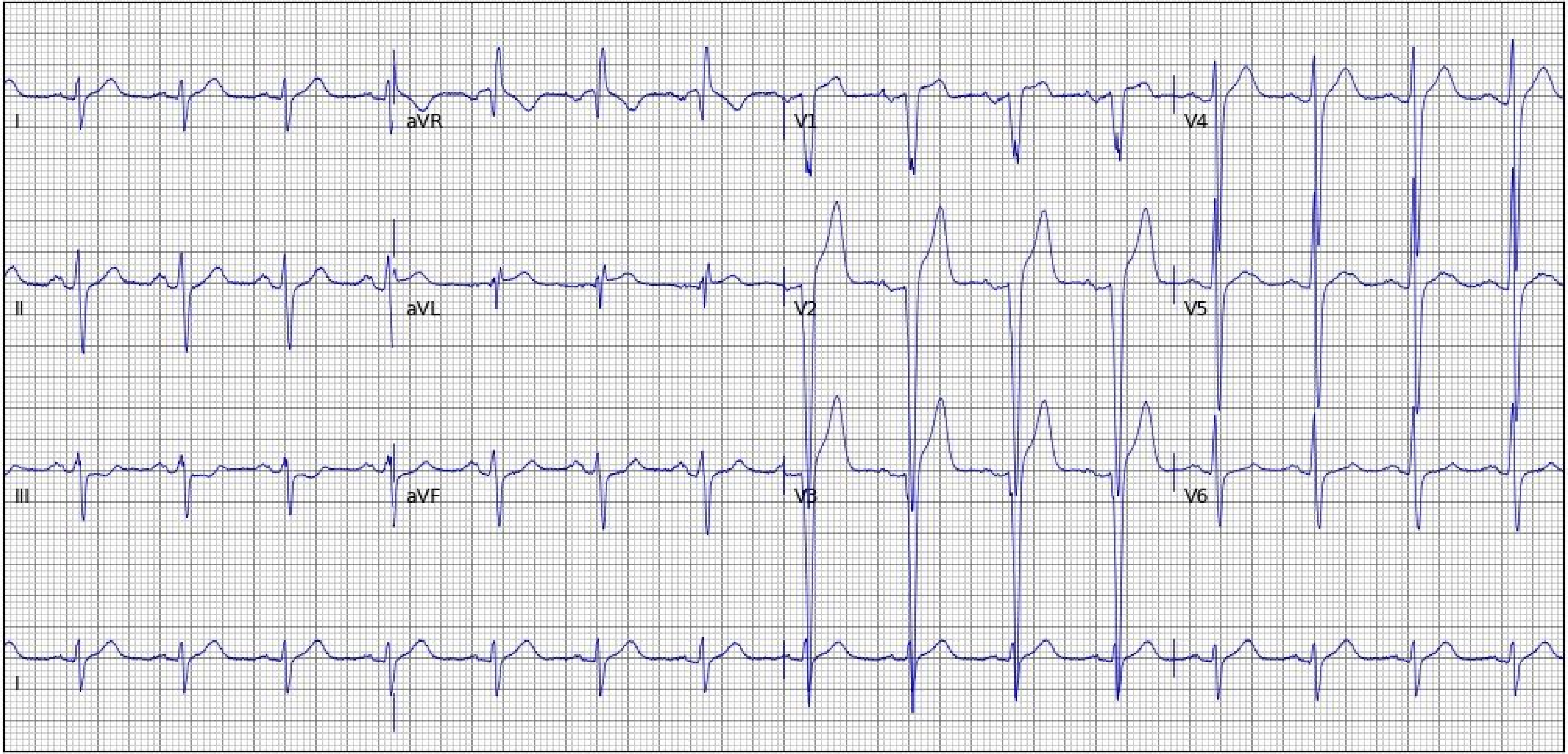
I
aVR
V1
V4
II
aVL
V2
V5
III
aVF
V3
V6
I

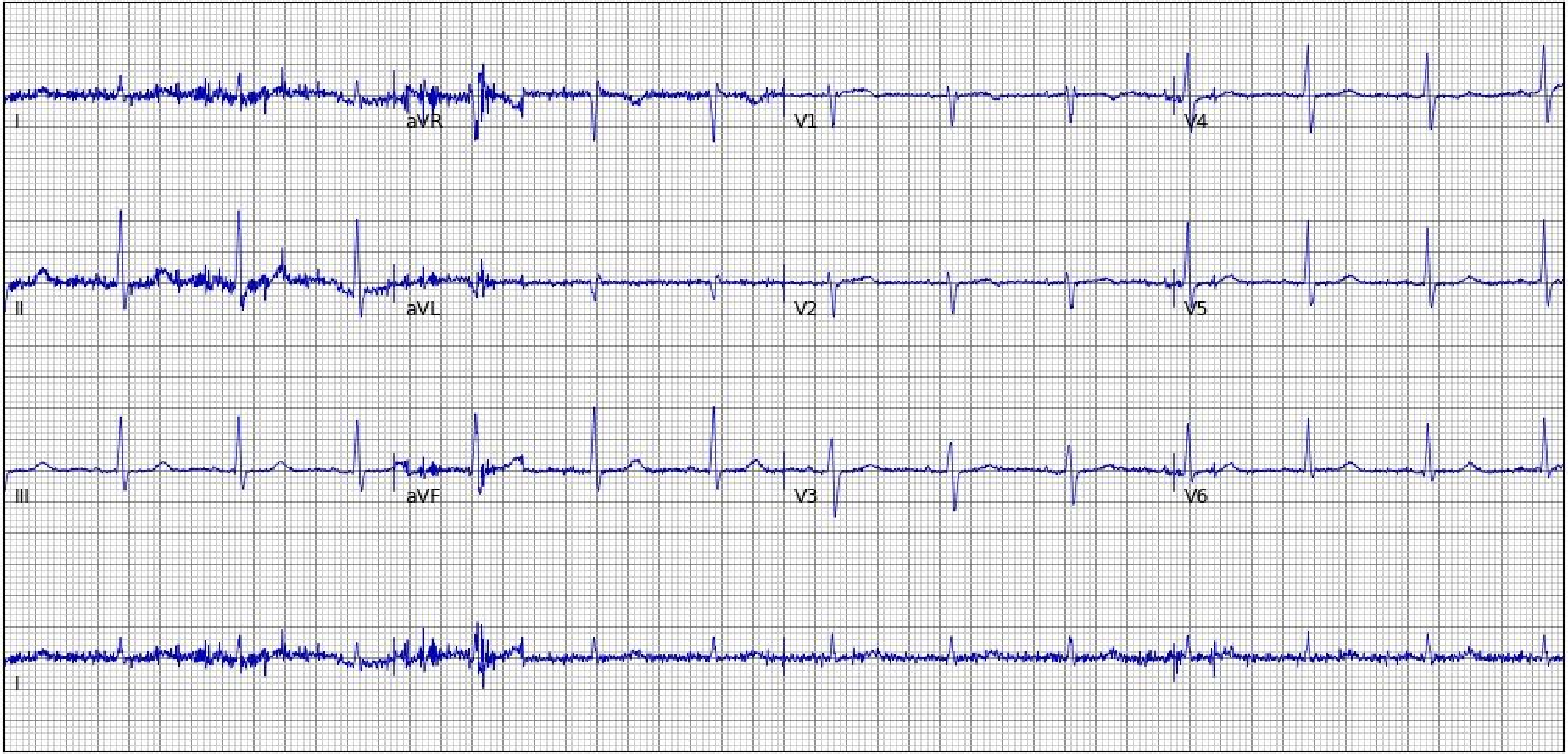
I
aVR
V1
V4
II
aVL
V2
V5
III
aVF
V3
V6
I

### Section 5. Random examples of 25 electrocardiogram images plotted in novel format (Black-on-Black Rhythm-on-top) not encountered during model development

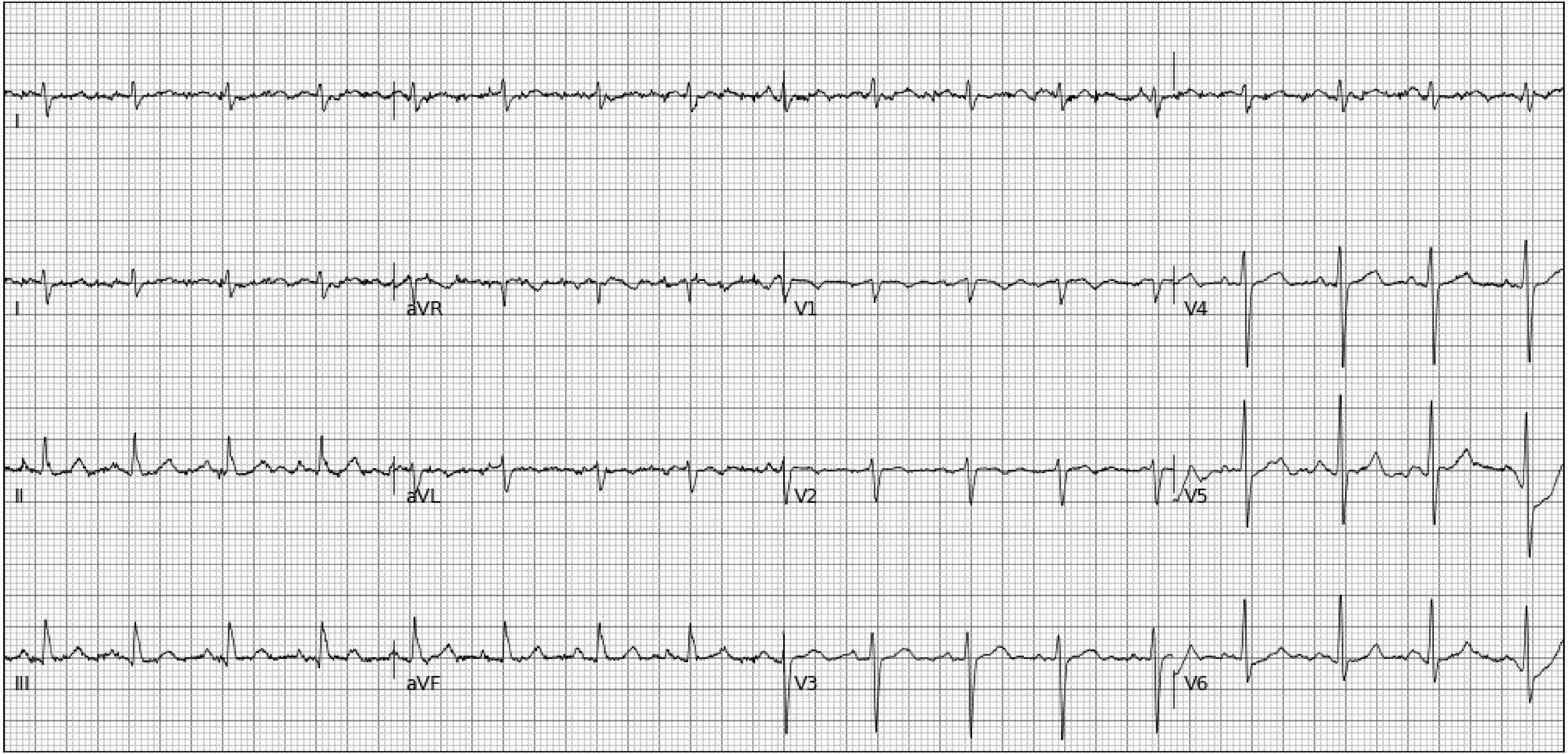
I
I
aVR
V1
V4
II
aVL
V2
V5
III
aVF
V3
V6

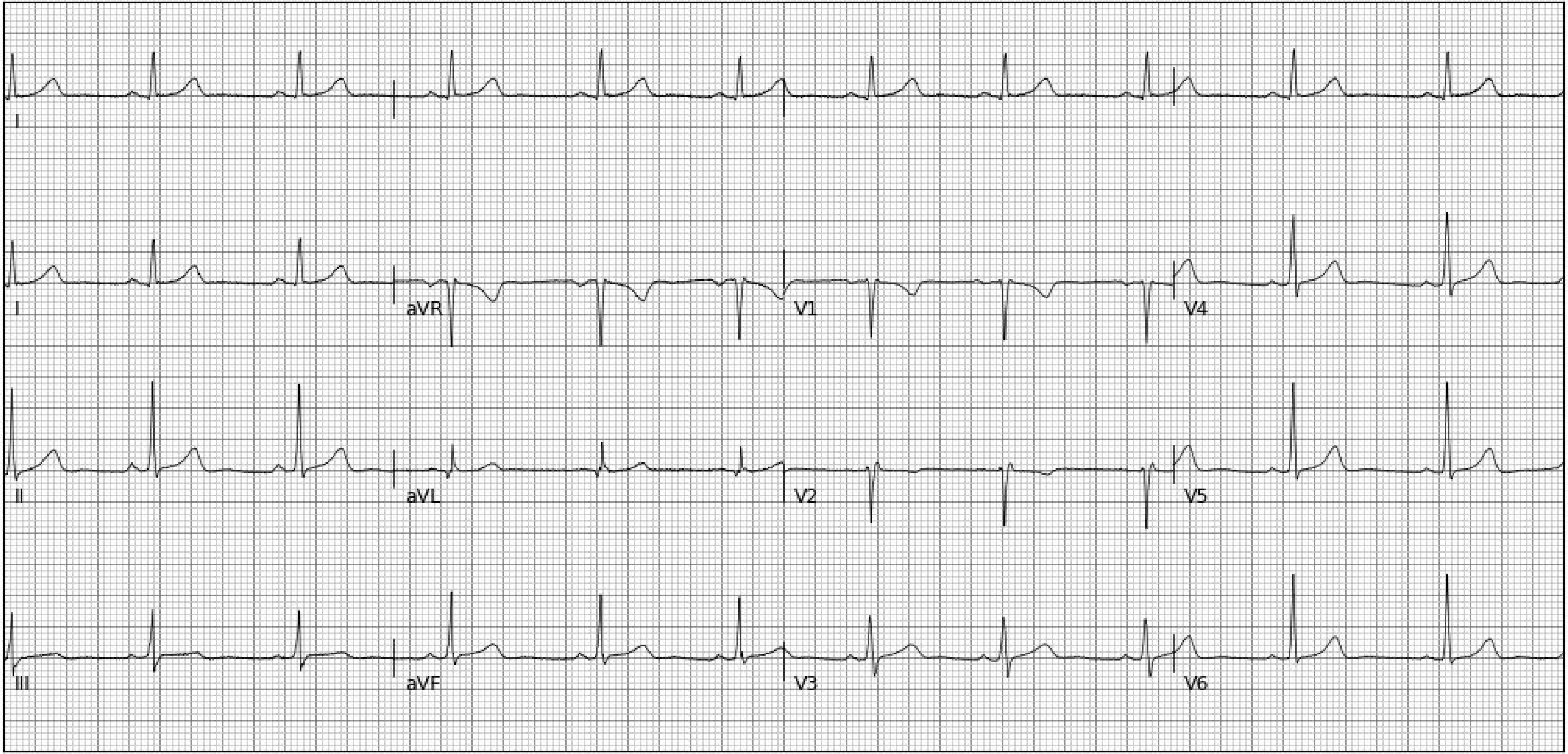
I
I
aVR
V1
V4
II
aVL
V2
V5
III
aVF
V3
V6

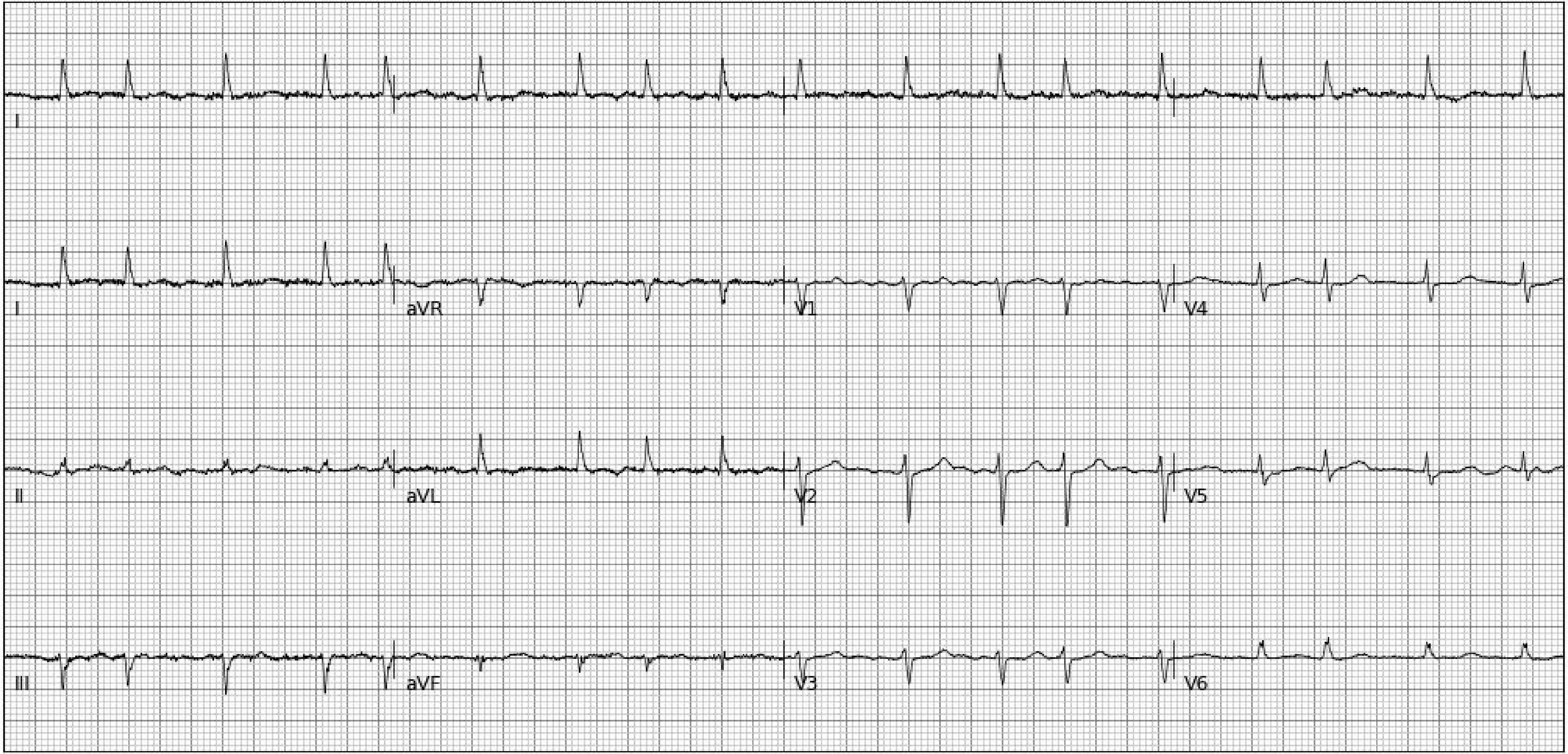
I
I
aVR
V1
V4
II
aVL
V2
V5
III
aVF
V3
V6

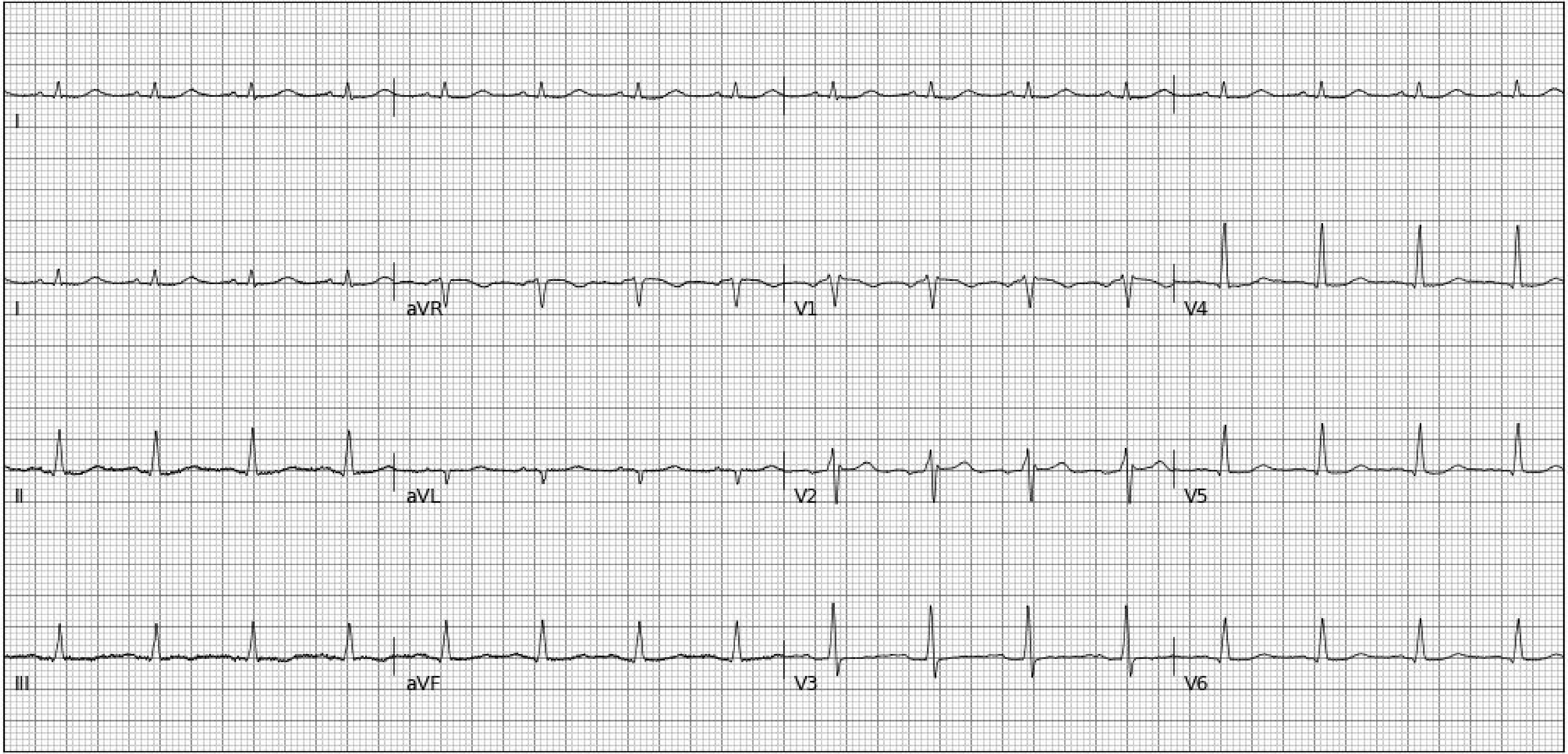
I
I
aVR
V1
V4
II
aVL
V2
V5
III
aVF
V3
V6

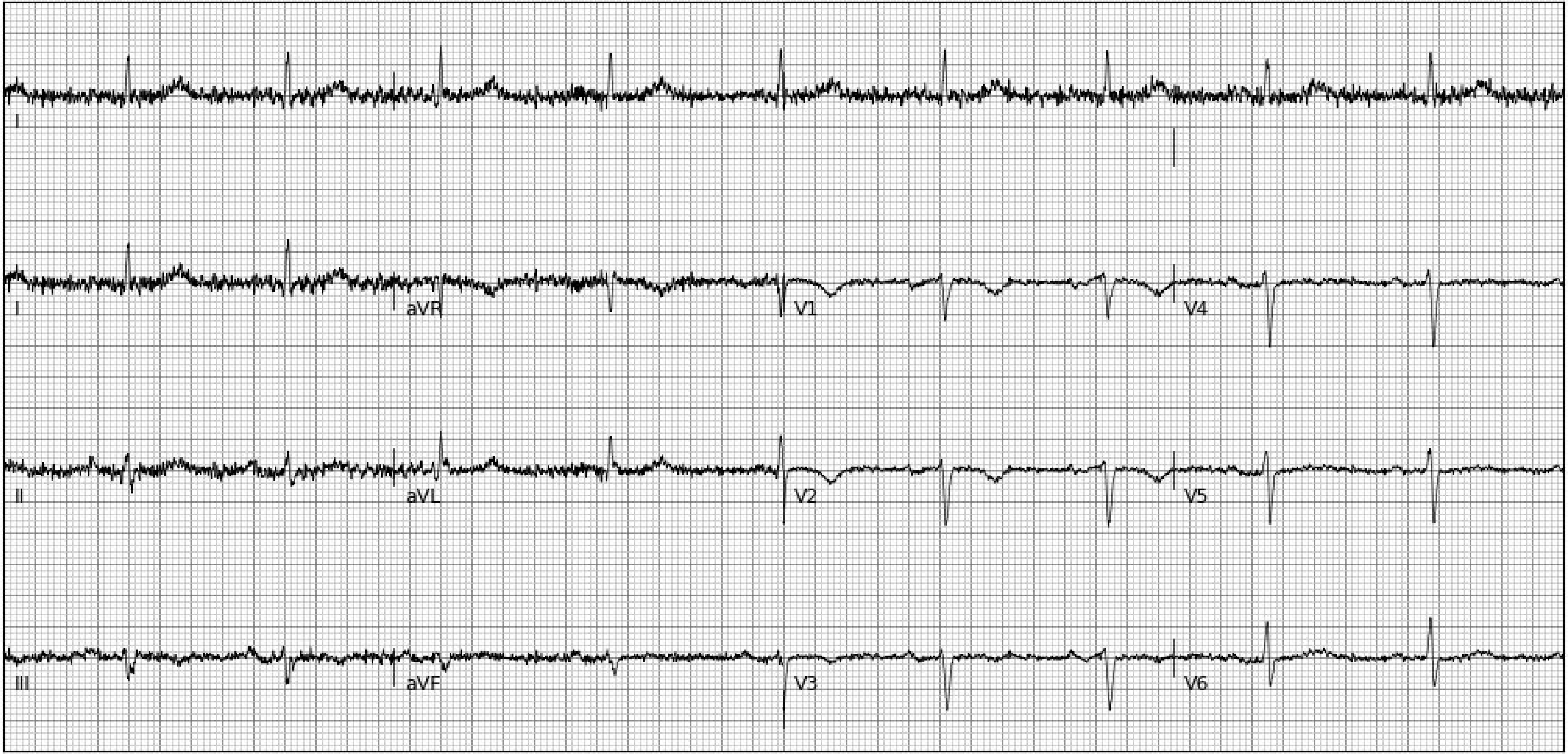
I
I
aVR
V1
V4
II
aVL
V2
V5
III
aVF
V3
V6

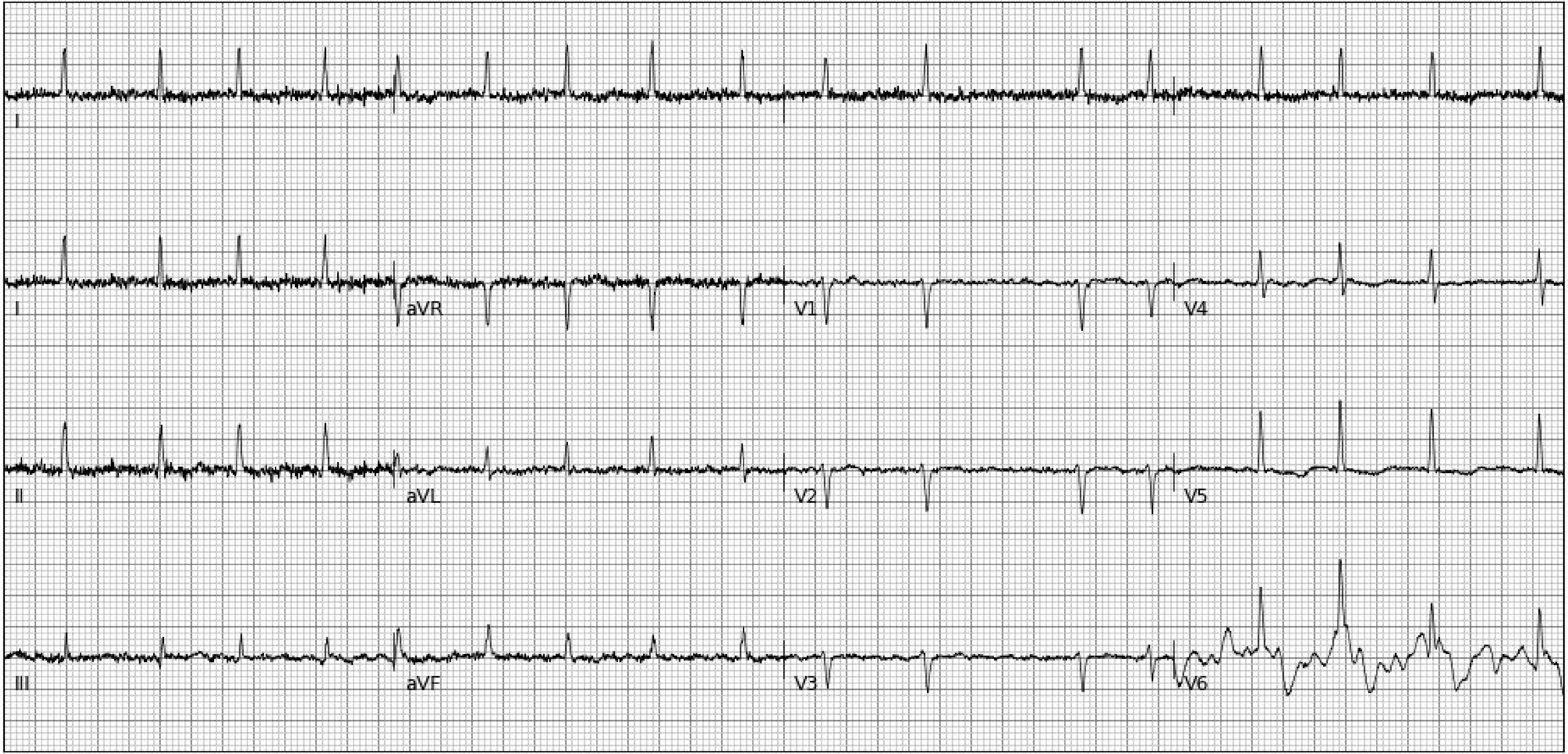
I
I
aVR
V1
V4
II
aVL
V2
V5
III
aVF
V3
V6

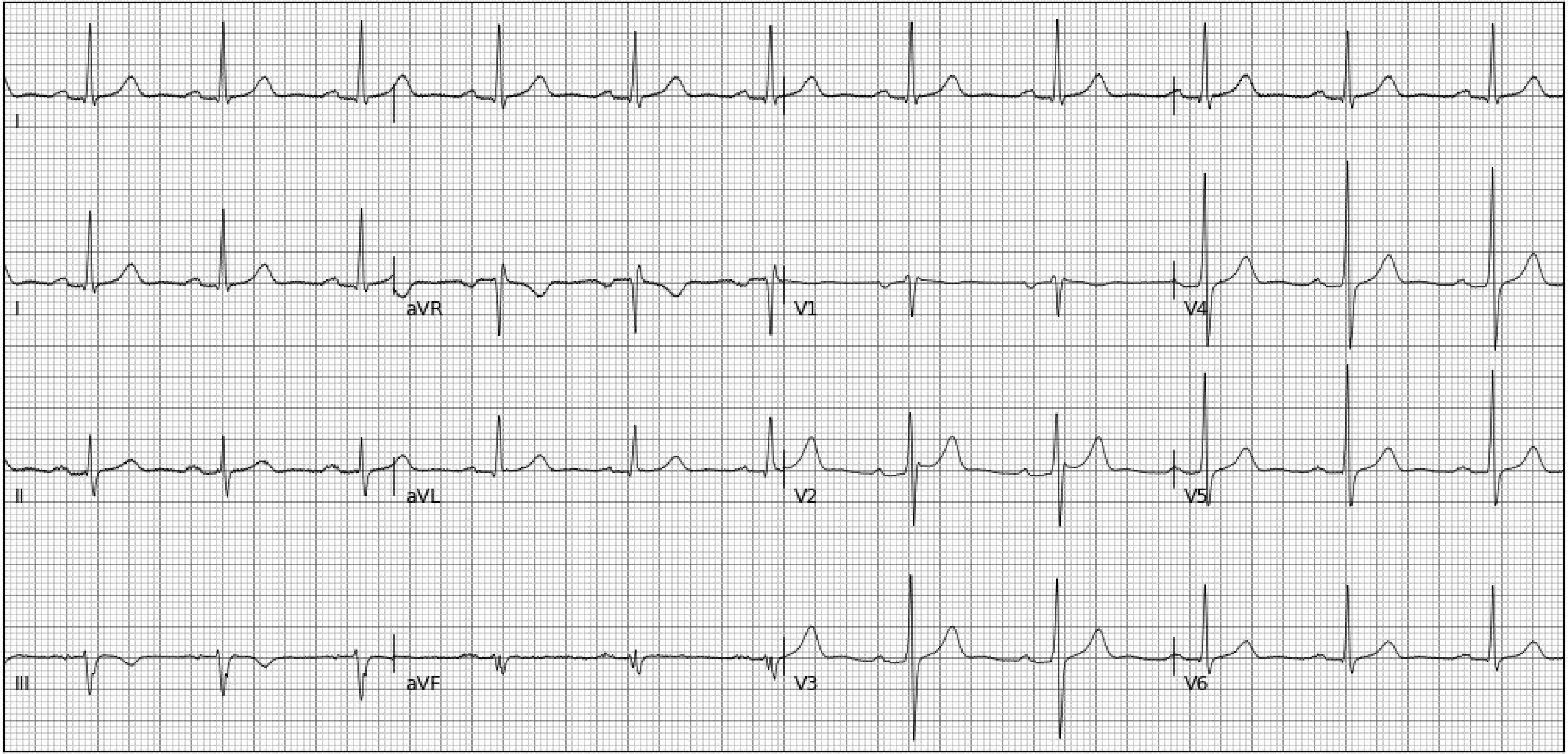
I
I
aVR
V1
V4
II
aVL
V2
V5
III
aVF
V3
V6

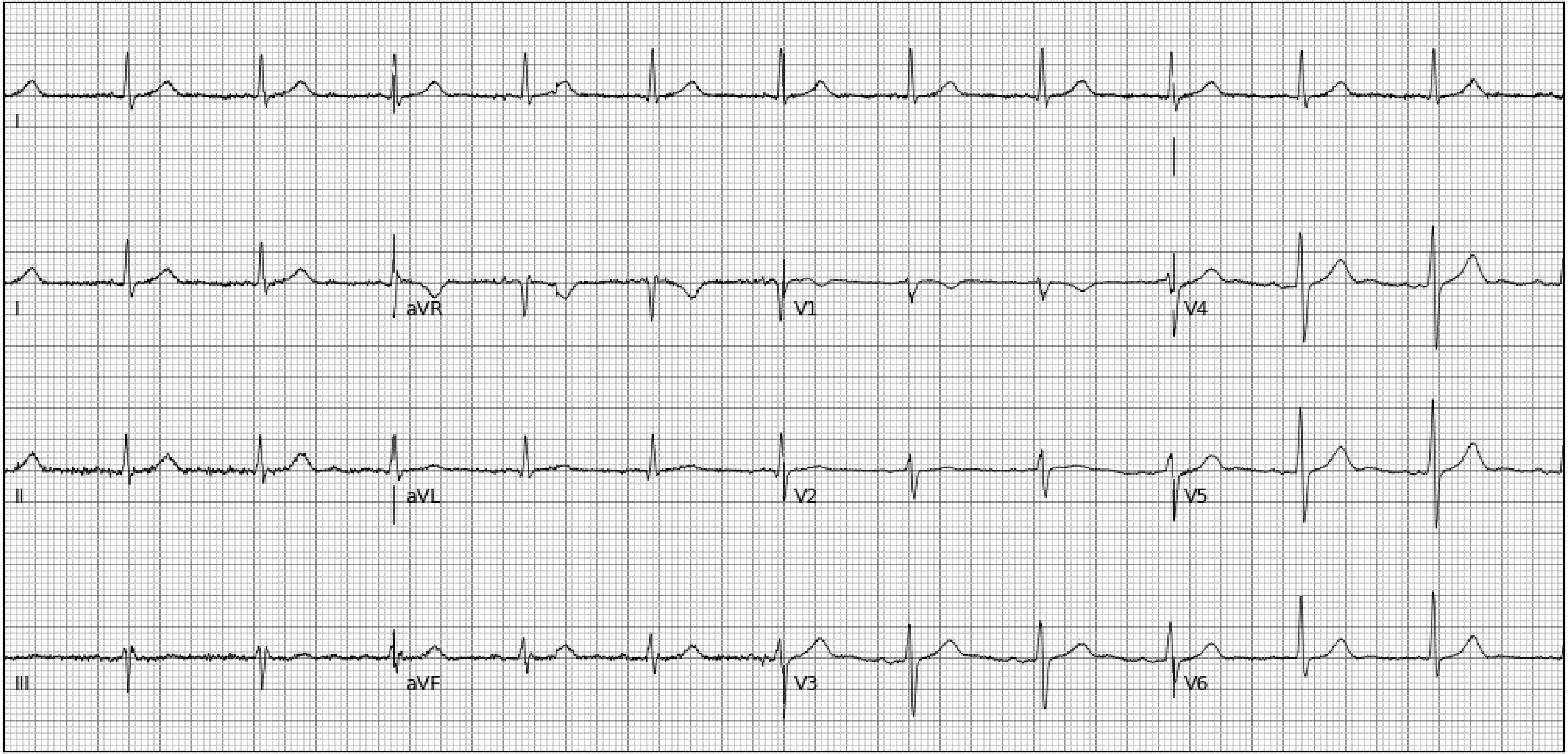
I
I
aVR
V1
V4
II
aVL
V2
V5
III
aVF
V3
V6

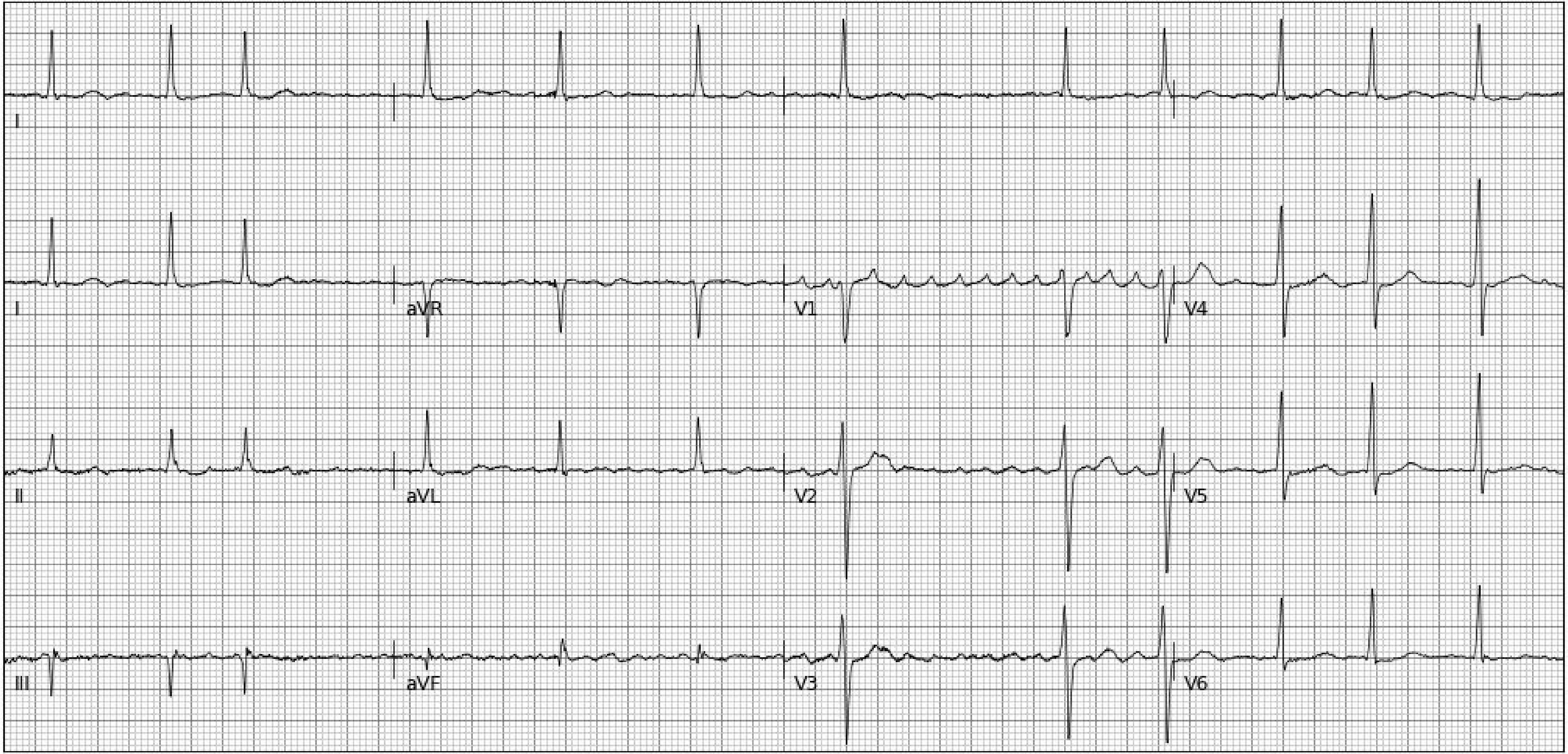
I
I
aVR
V1
V4
II
aVL
V2
V5
III
aVF
V3
V6

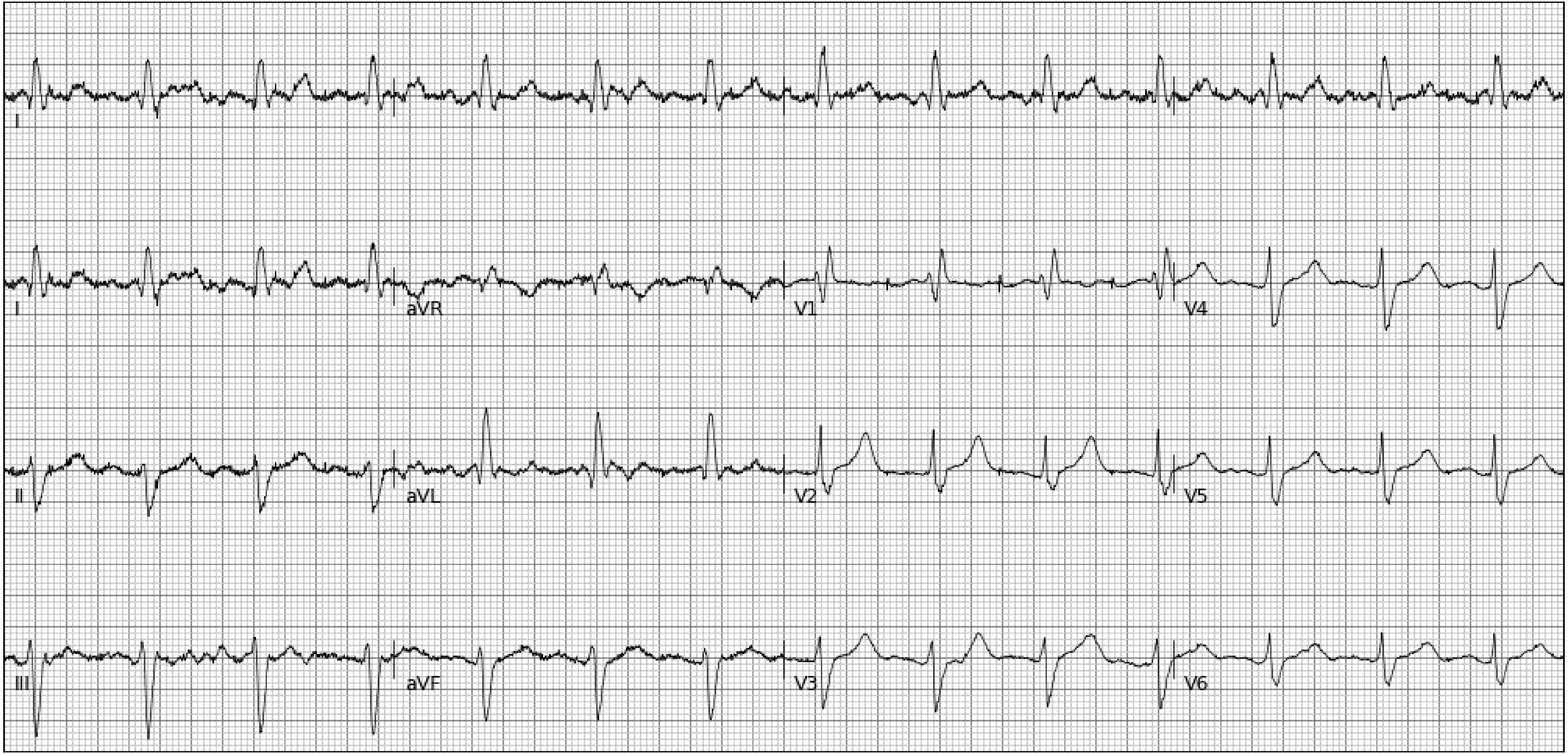
I
I
aVR
V1
V4
II
aVL
V2
V5
III
aVF
V3
V6

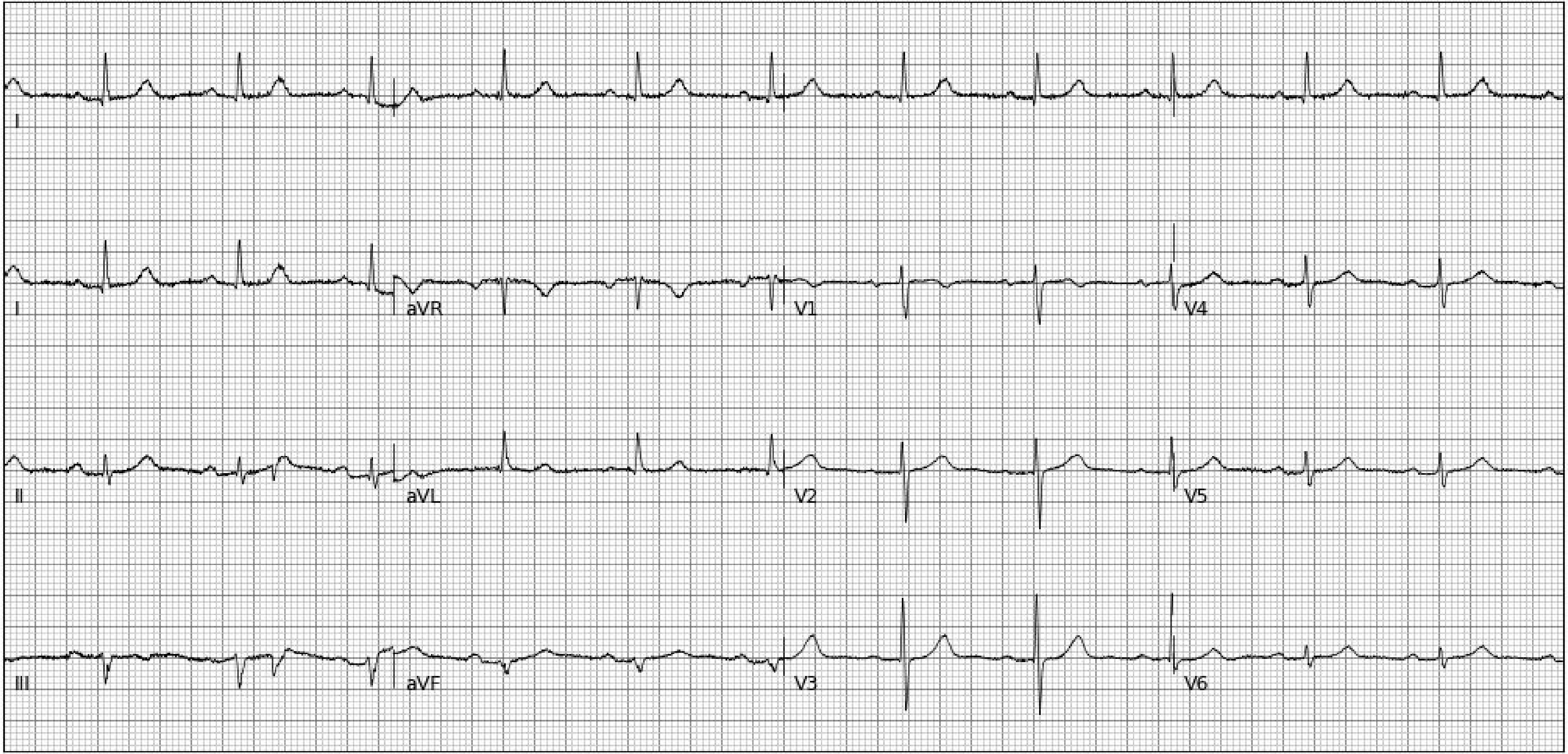
I
I
aVR
V1
V4
II
aVL
V2
V5
III
aVF
V3
V6

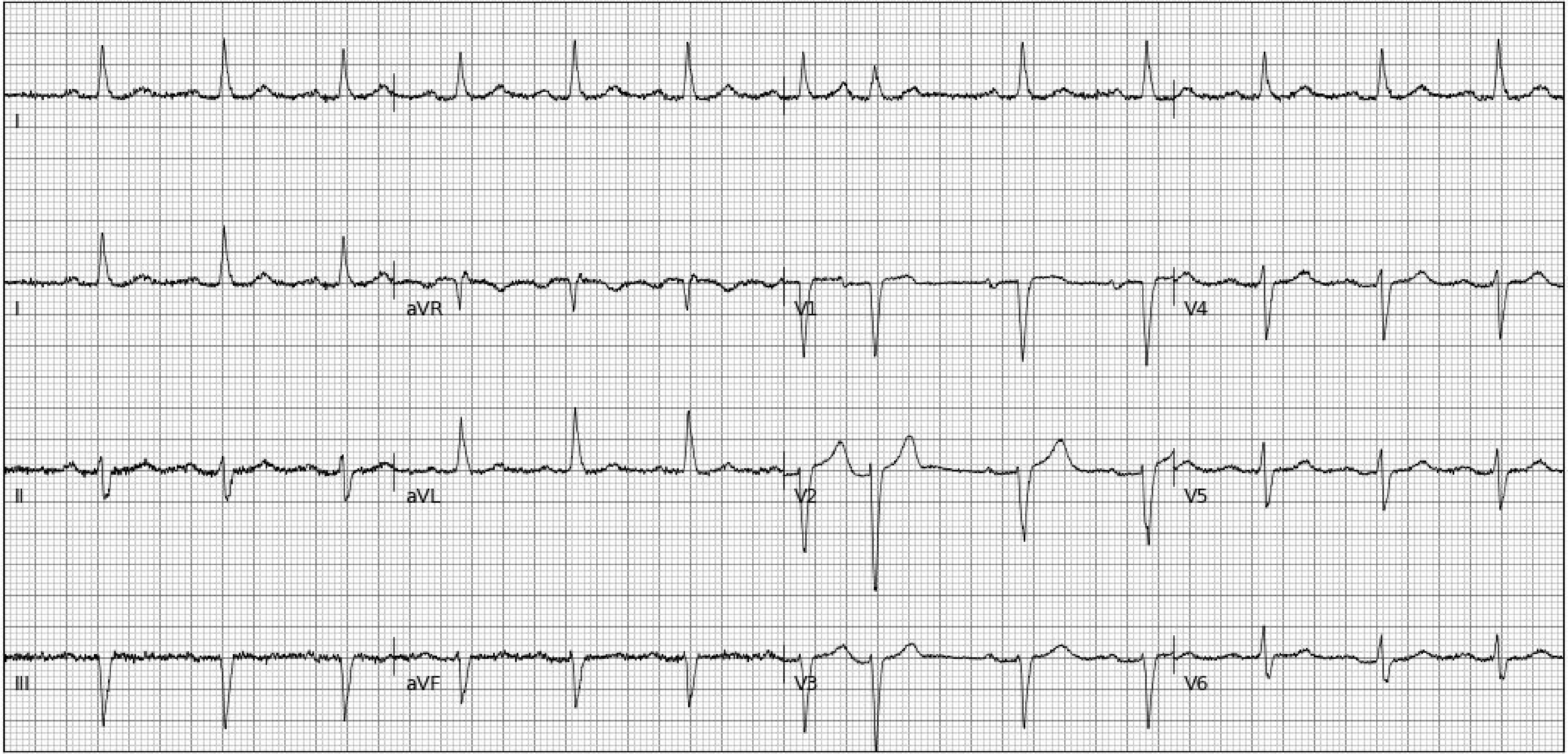
I
I
aVR
V1
V4
II
aVL
V2
V5
III
aVF
V3
V6

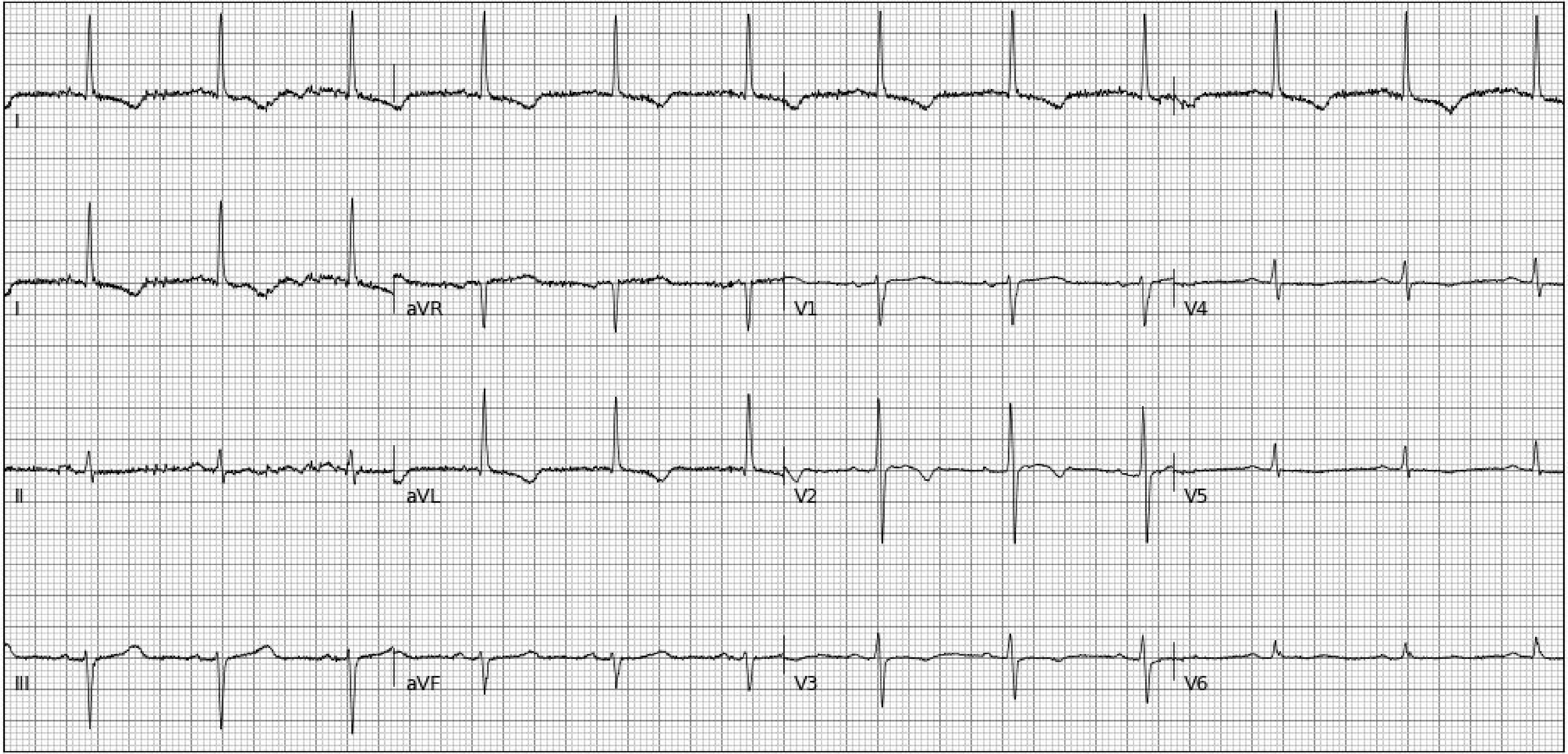
I
I
aVR
V1
V4
II
aVL
V2
V5
III
aVF
V3
V6

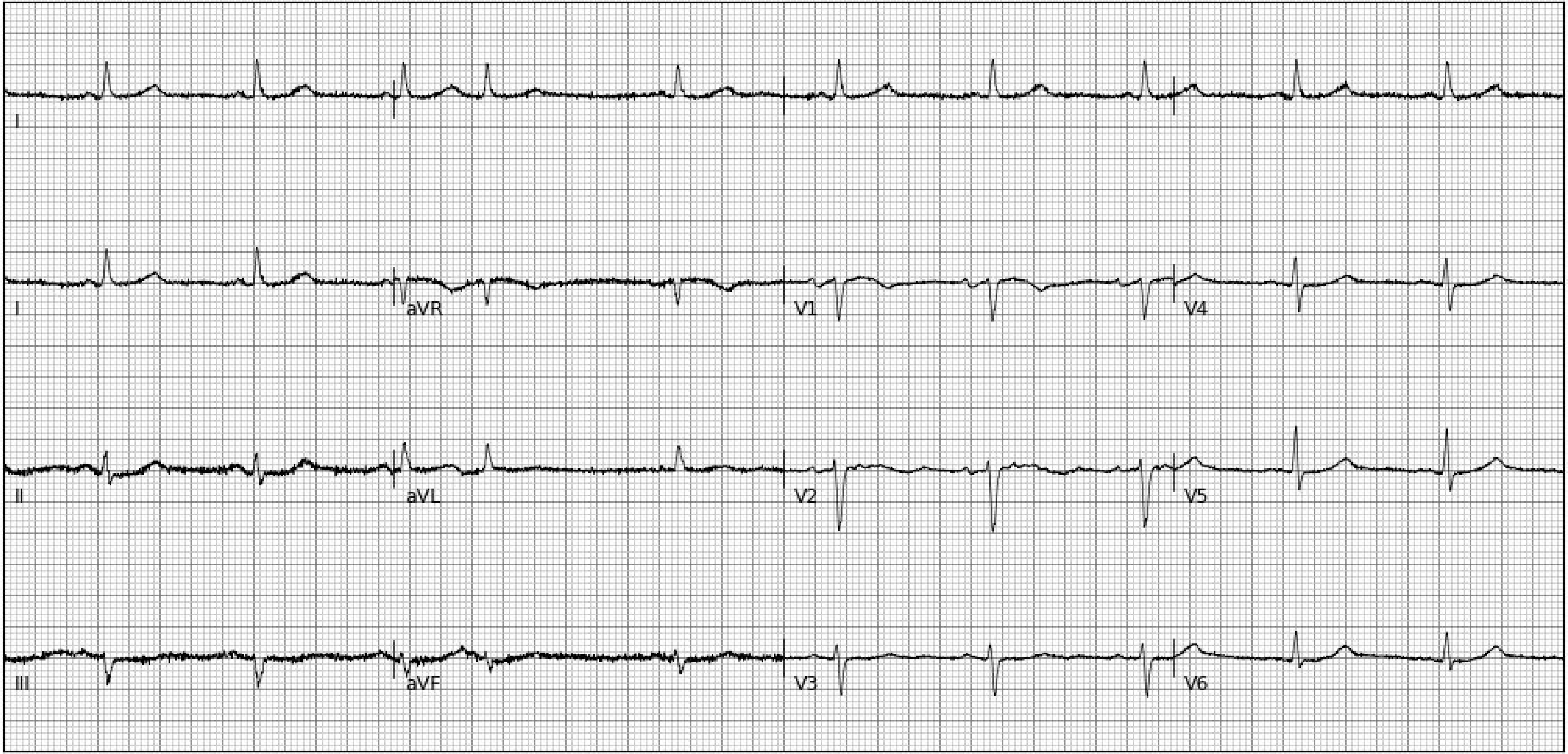
I
I
aVR
V1
V4
II
aVL
V2
V5
III
aVF
V3
V6

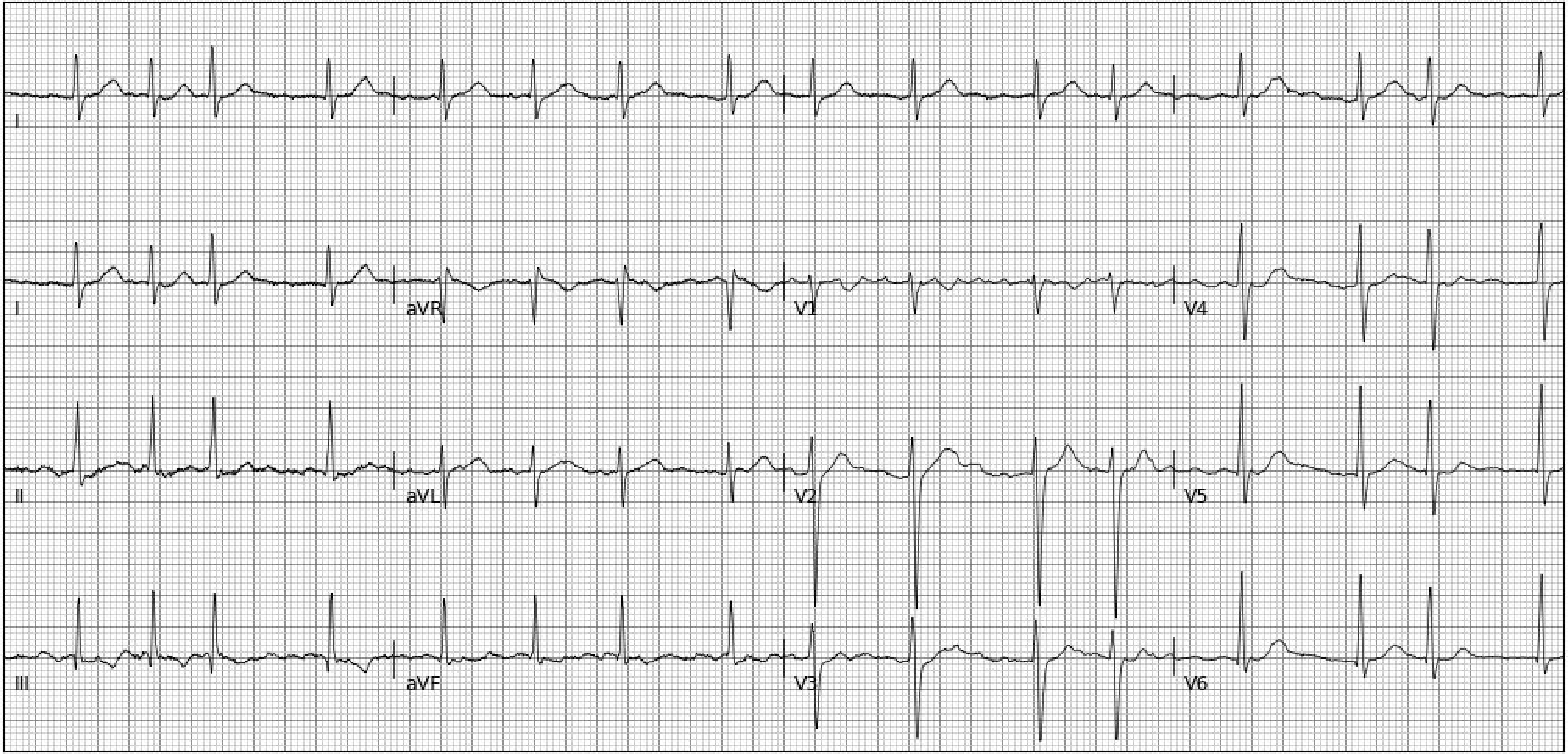
I
I
aVR
V1
V4
II
aVL
V2
V5
III
aVF
V3
V6

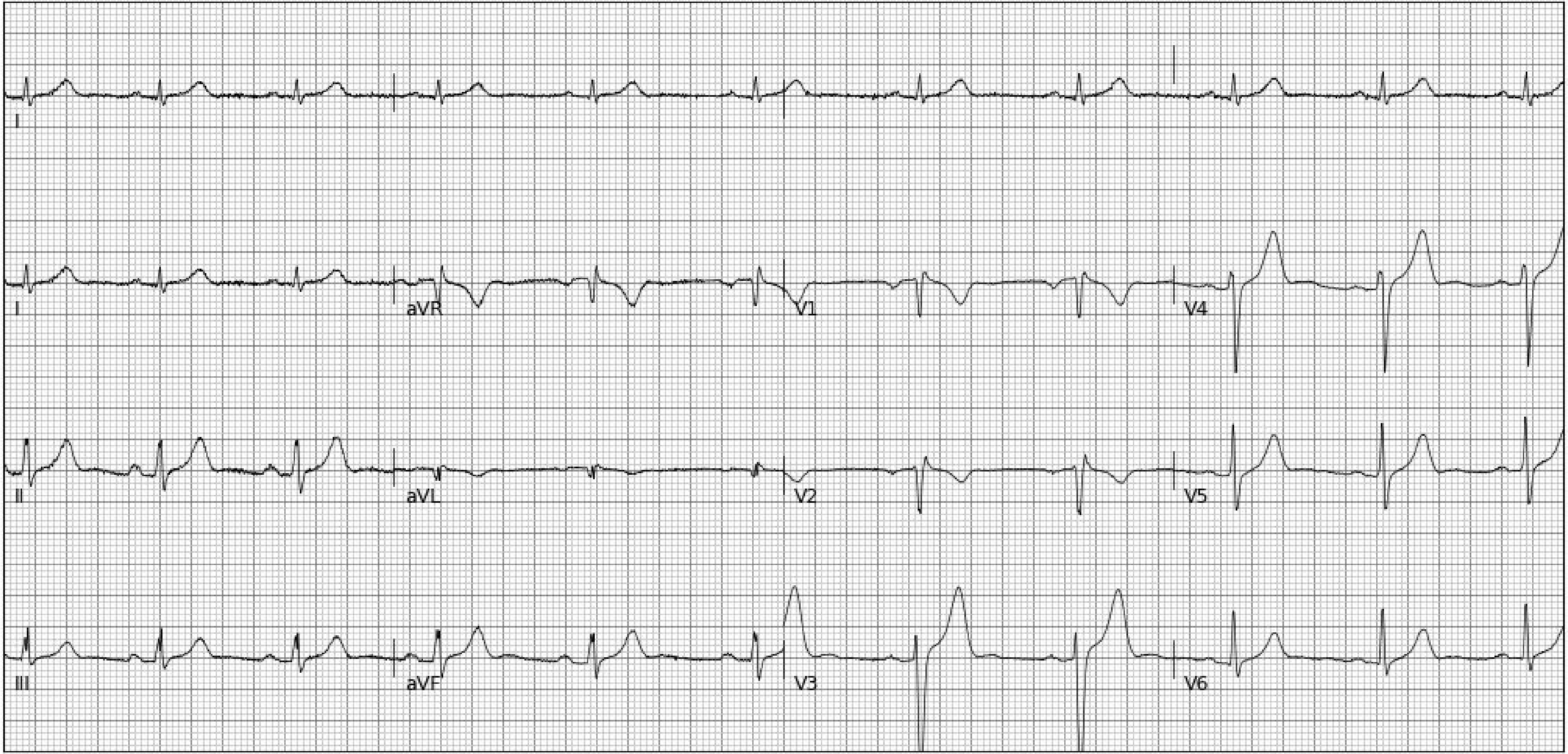
I
I
aVR
V1
V4
II
aVL
V2
V5
III
aVF
V3
V6

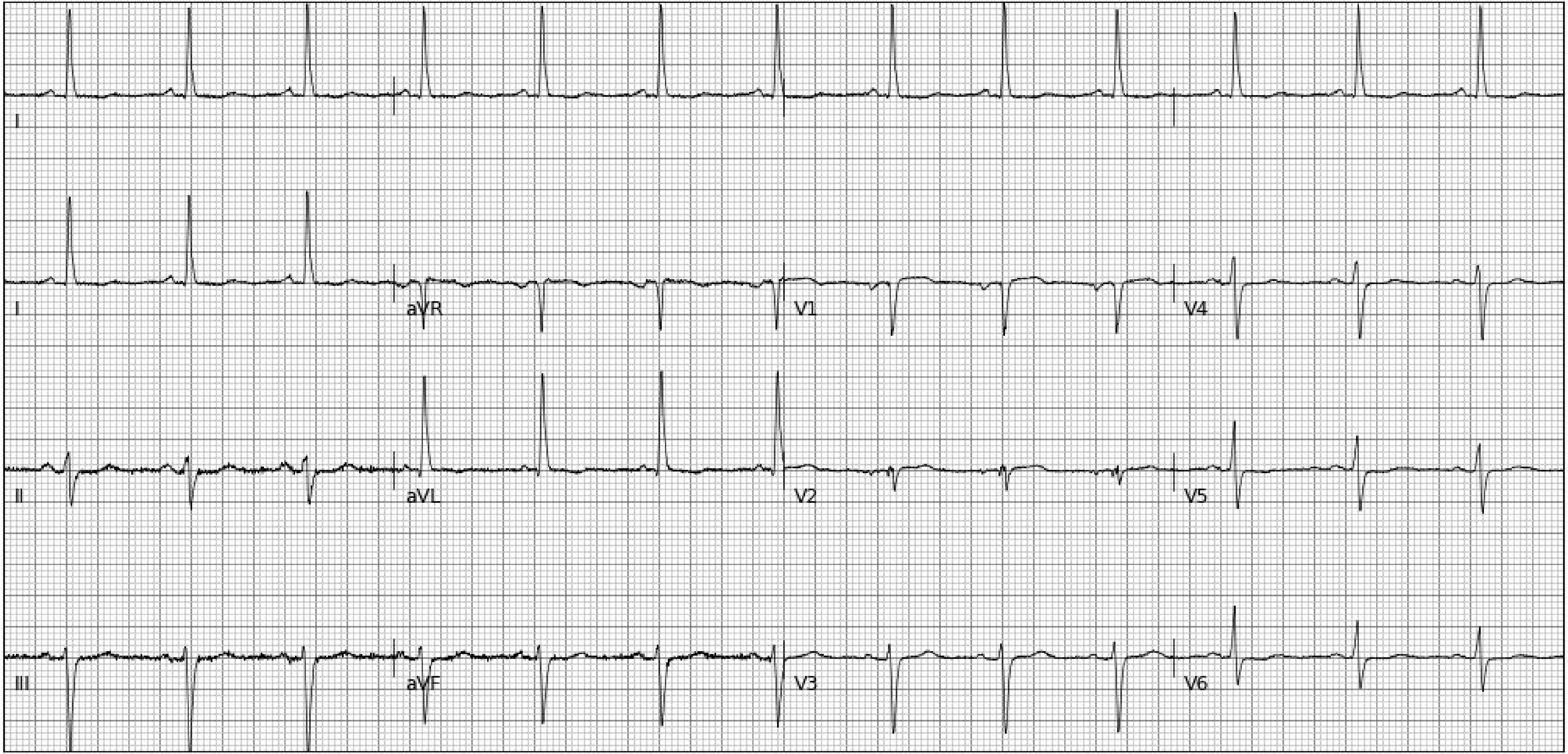
I
I
aVR
V1
V4
II
aVL
V2
V5
III
aVF
V3
V6

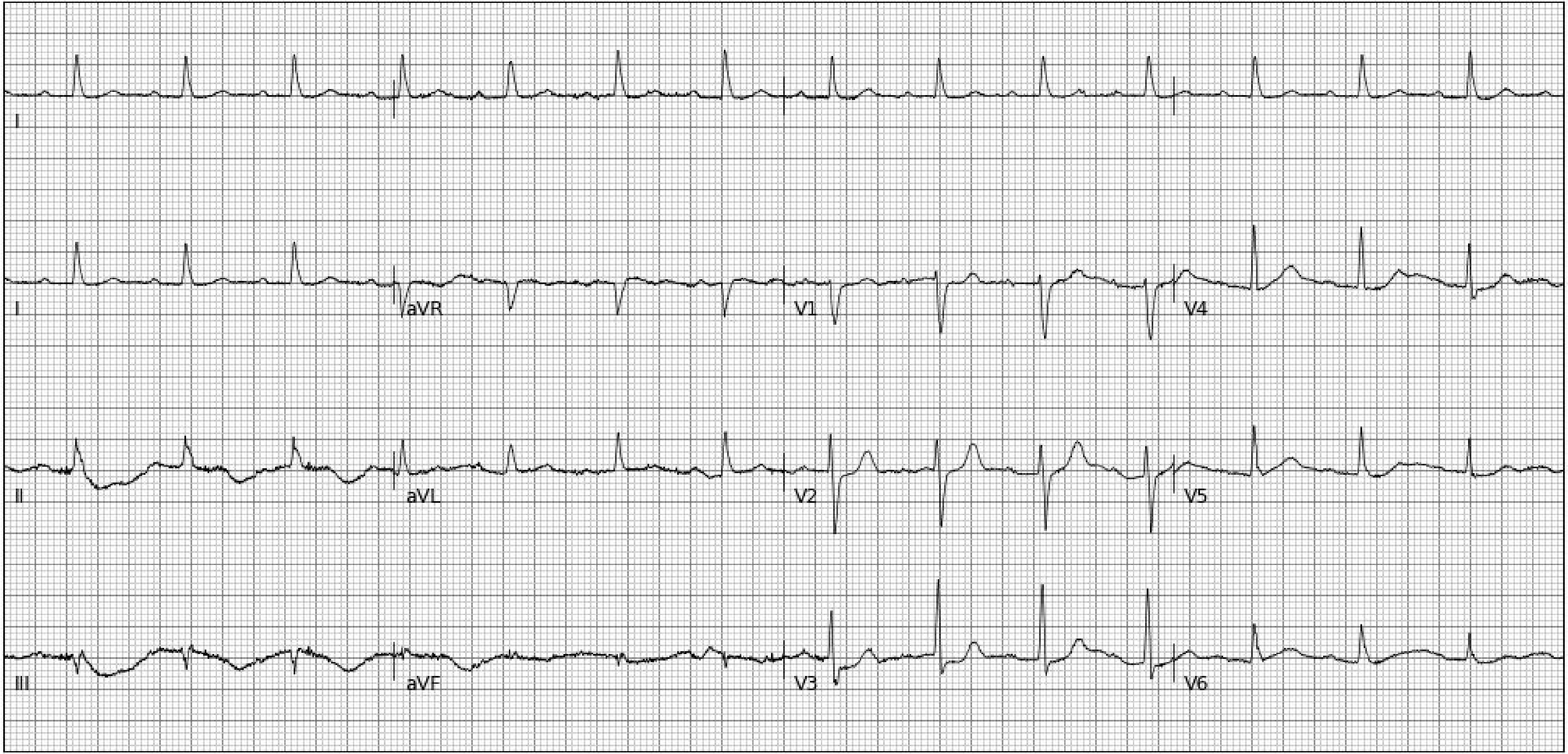
I
I
aVR
V1
V4
II
aVL
V2
V5
III
aVF
V3
V6

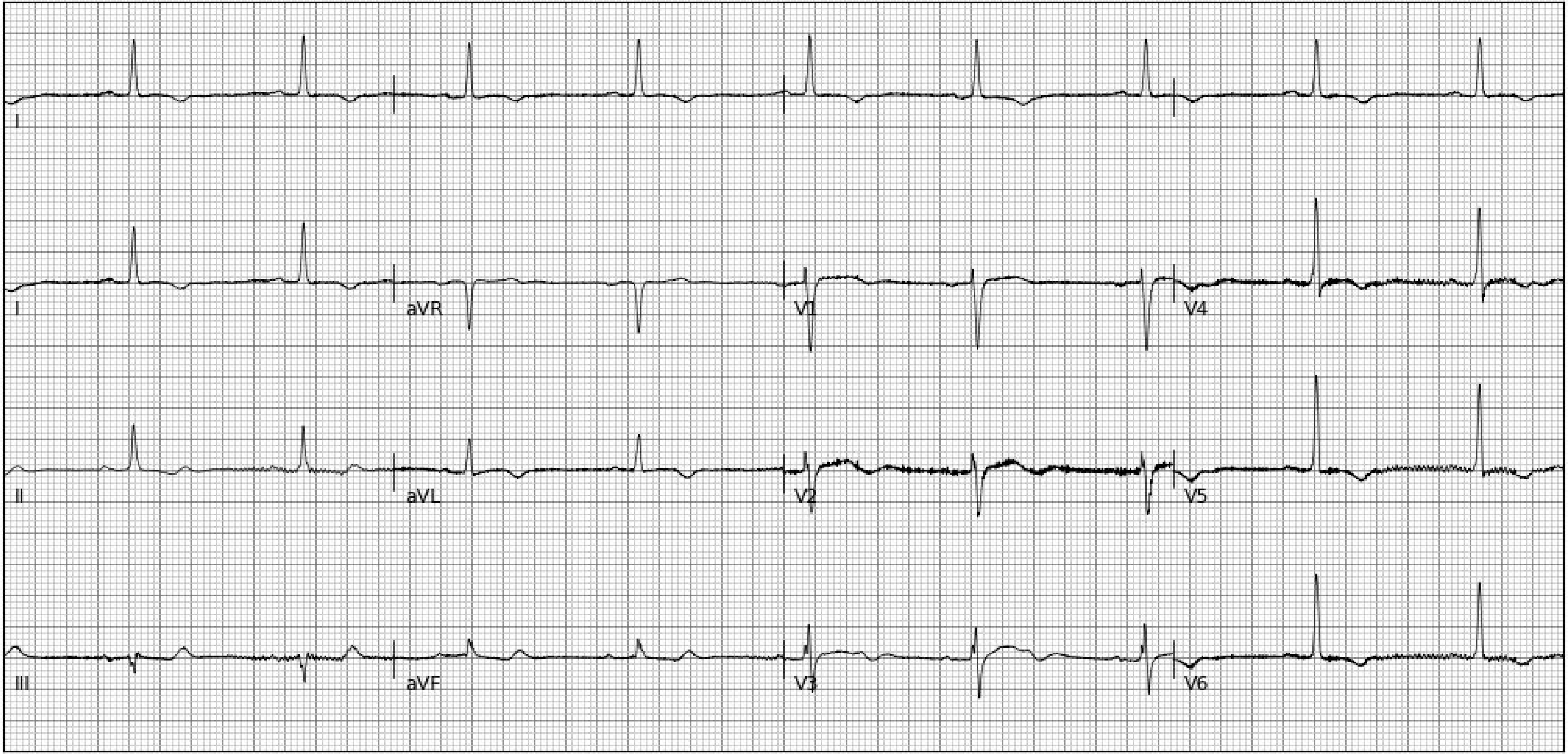
I
I
aVR
V1
V4
II
aVL
V2
V5
III
aVF
V3
V6

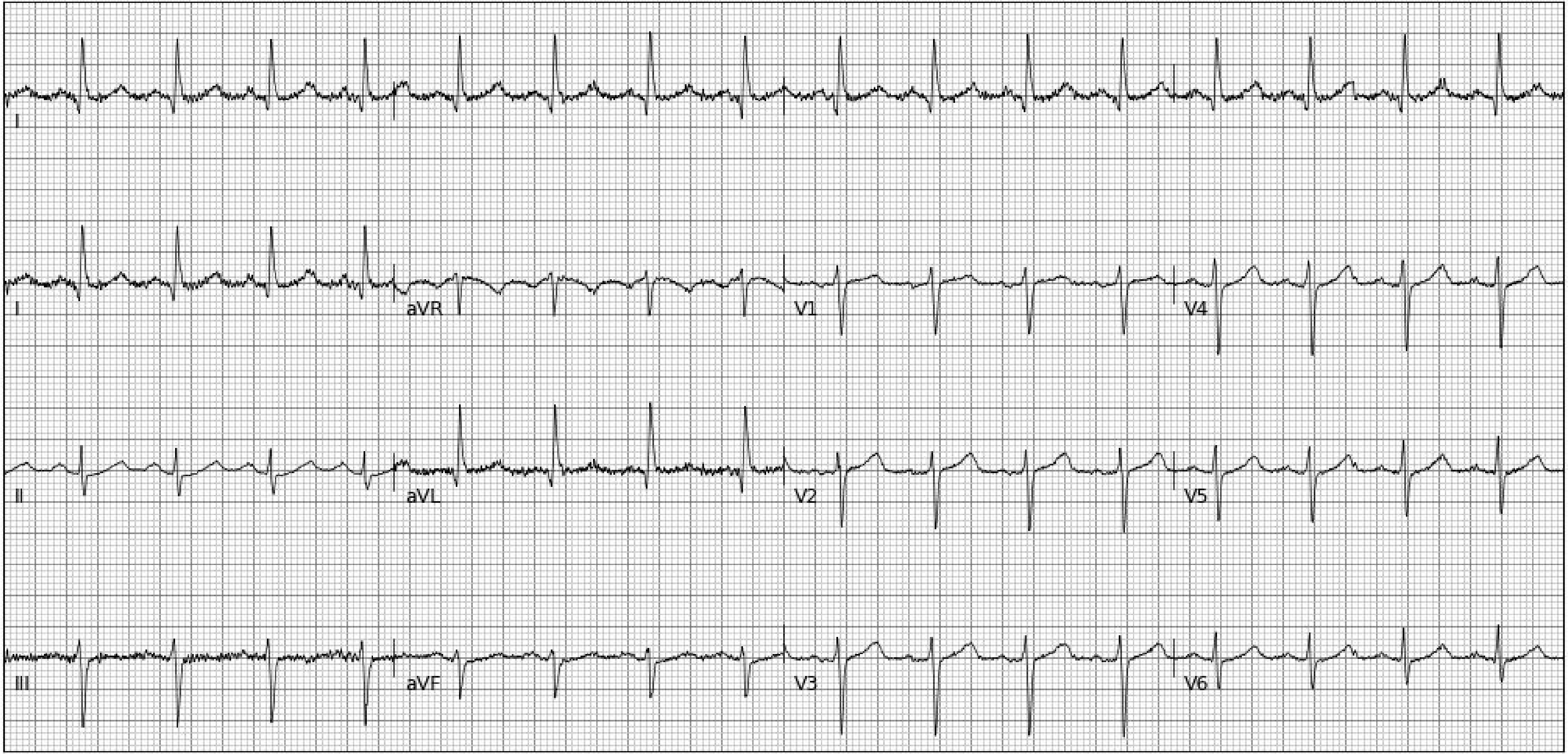
I
I
aVR
V1
V4
II
aVL
V2
V5
III
aVF
V3
V6

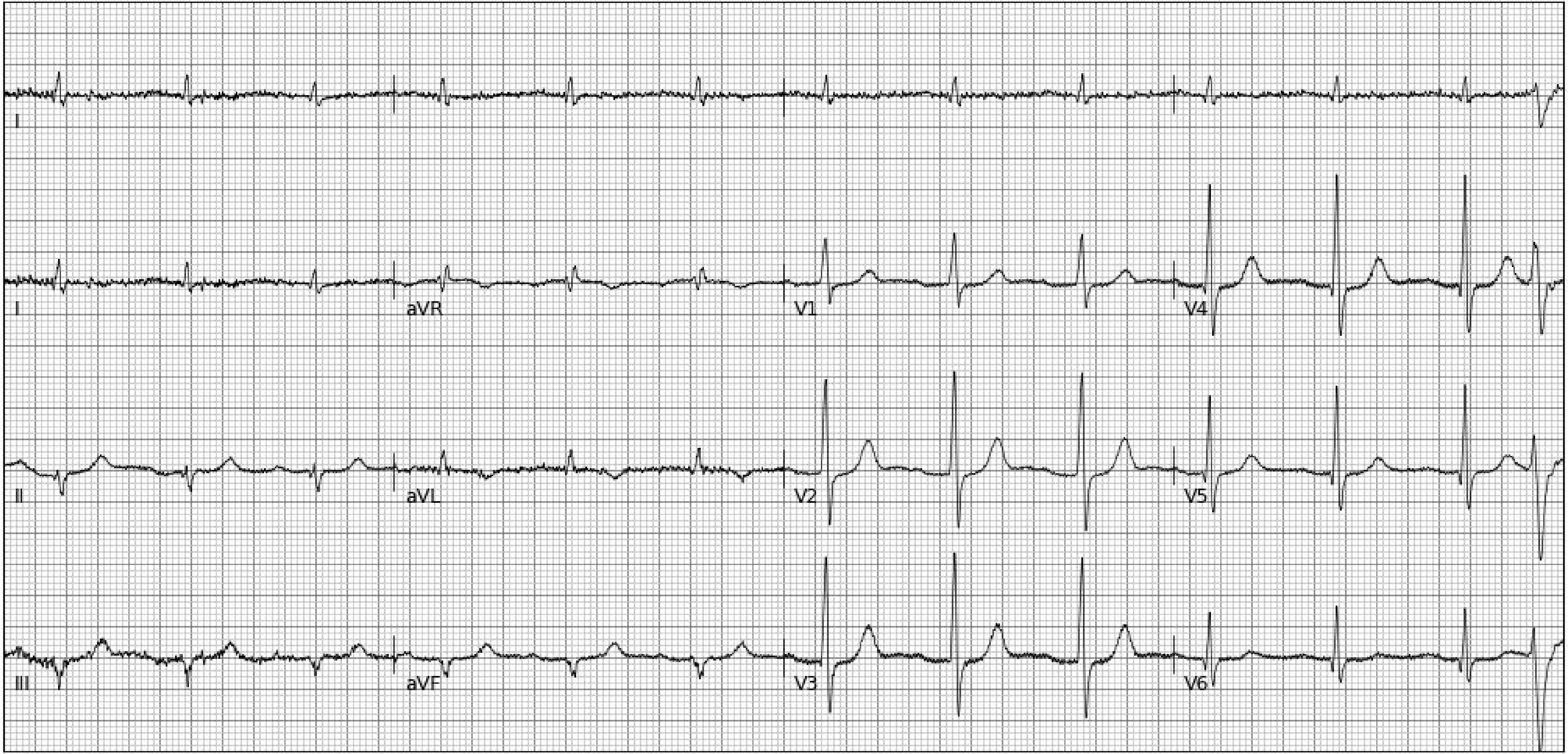
I
I
aVR
V1
V4
II
aVL
V2
V5
III
aVF
V3
V6

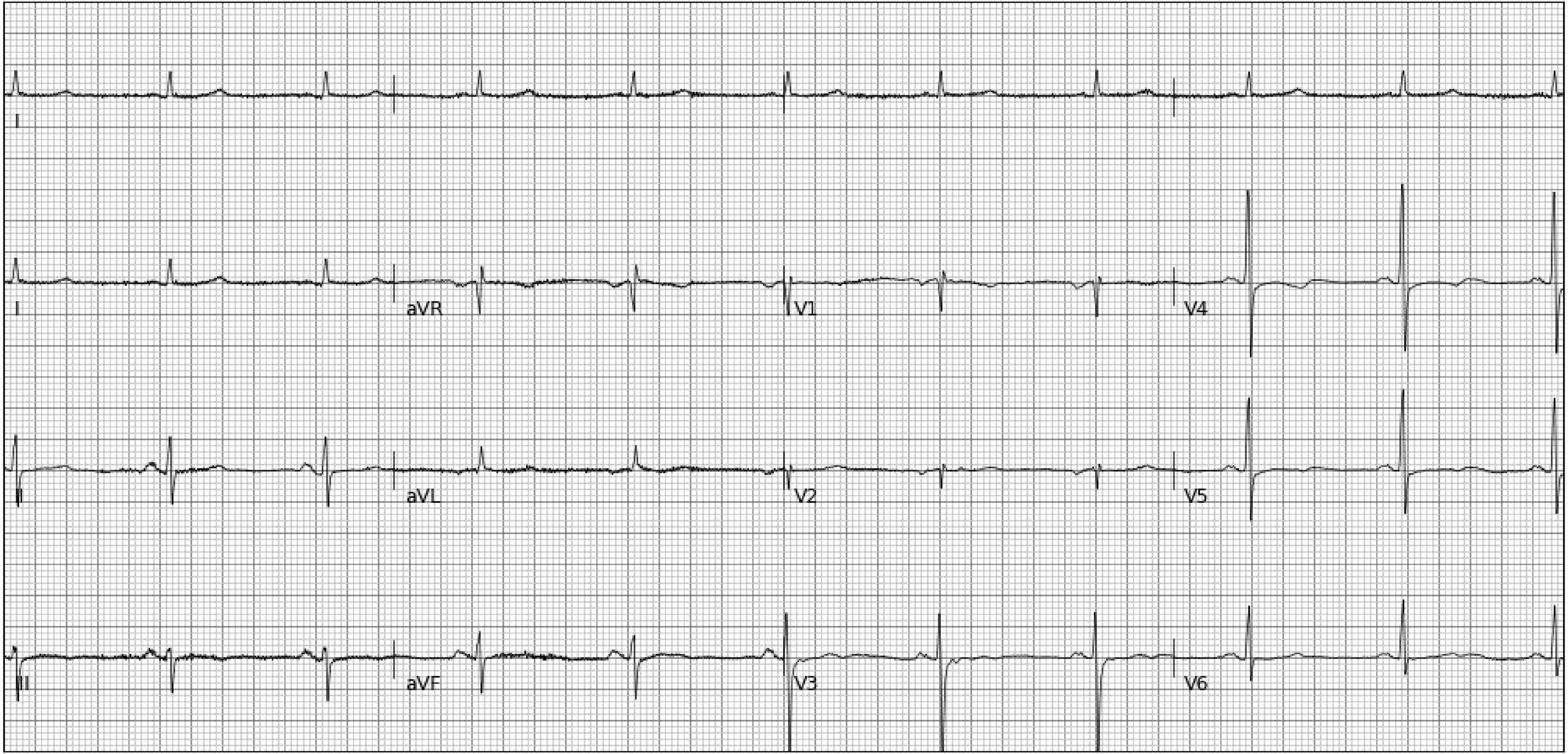
I
I
aVR
V1
V4
II
aVL
V2
V5
III
aVF
V3
V6

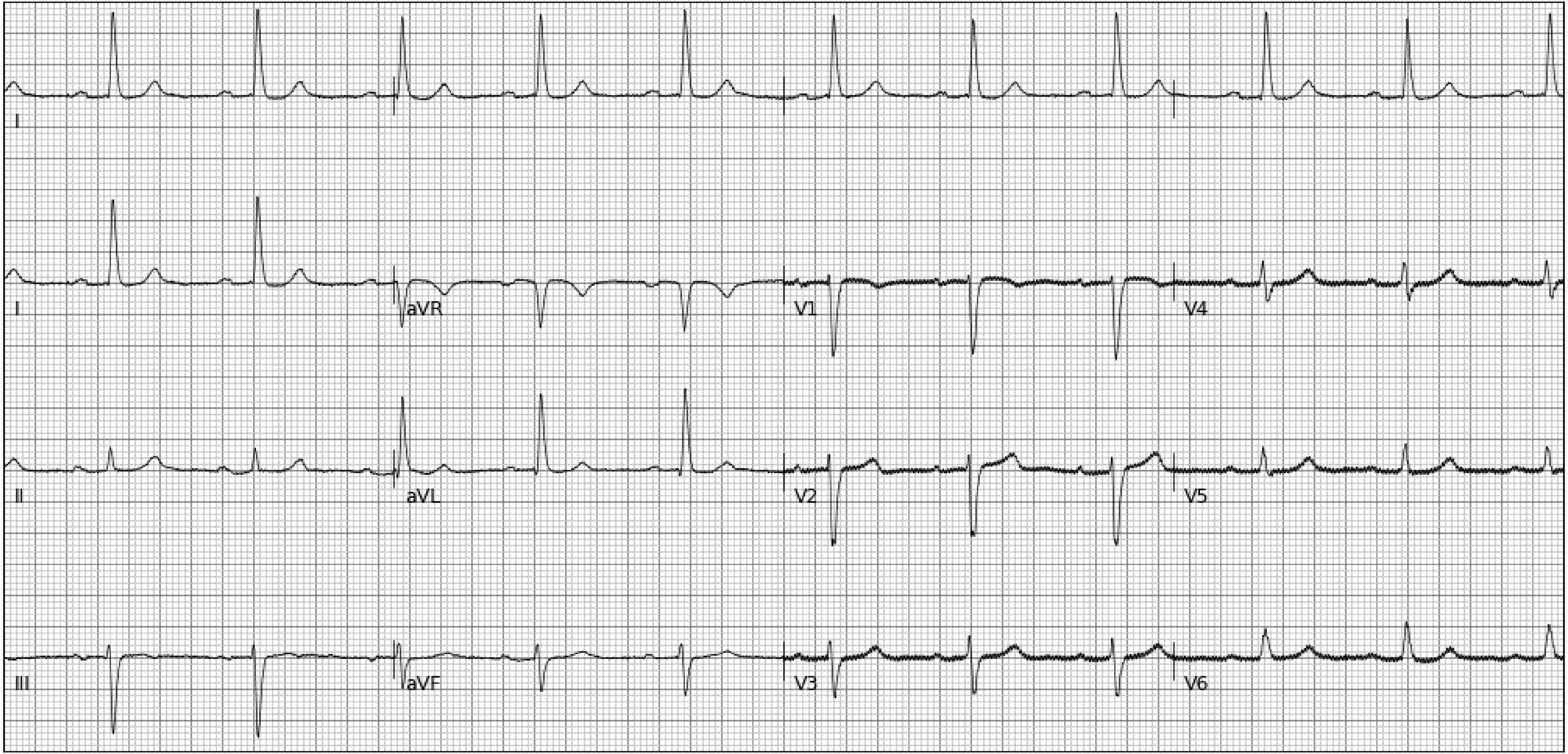
I
I
aVR
V1
V4
II
aVL
V2
V5
III
aVF
V3
V6

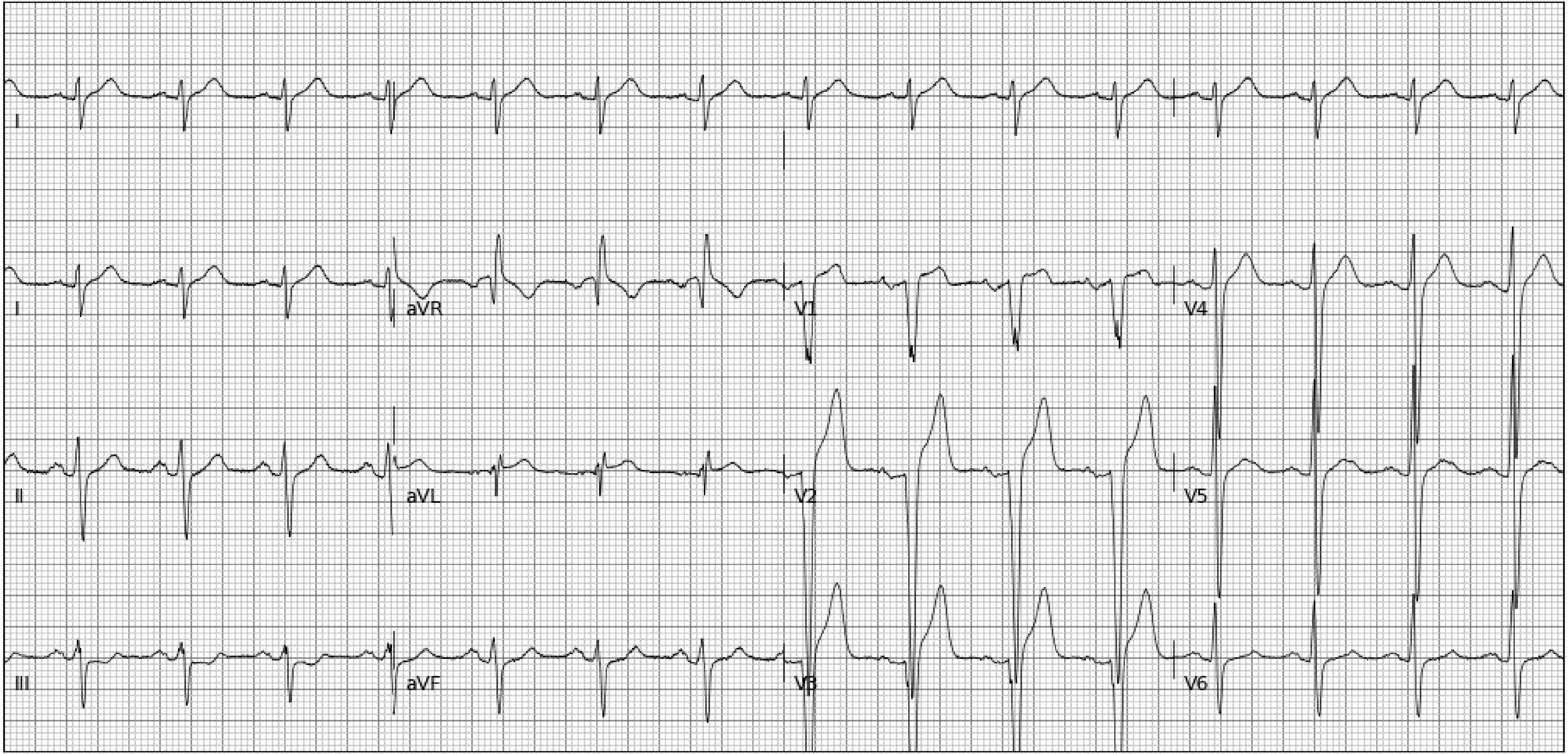
I
I
aVR
V1
V4
II
aVL
V2
V5
III
aVF
V3
V6

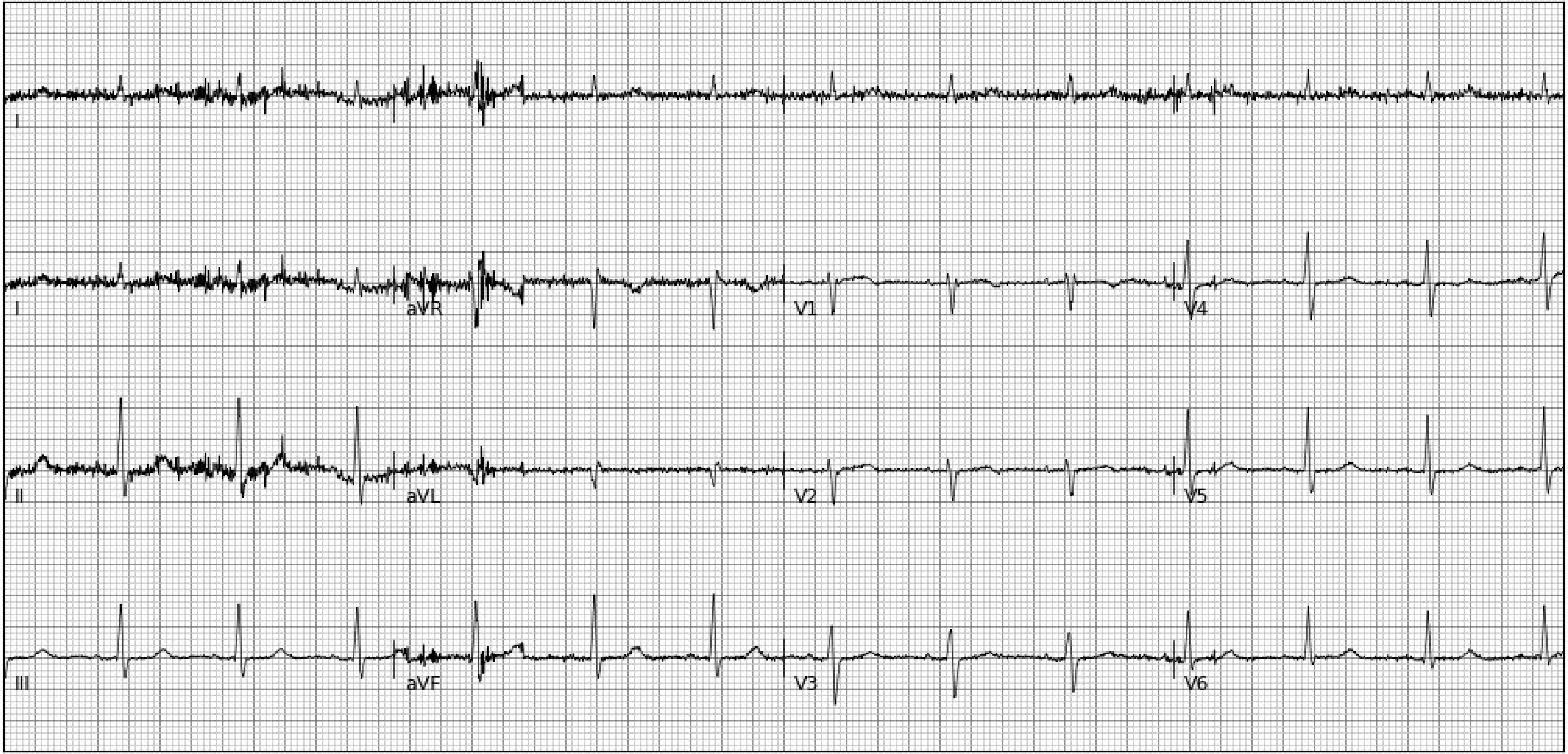
I
I
aVR
V1
V4
II
aVL
V2
V5
III
aVF
V3
V6

**Section 3. Random examples of 25 electrocardiogram images plotted in novel format (Blue-on-Red Rhythm-on-top) not encountered during model development**

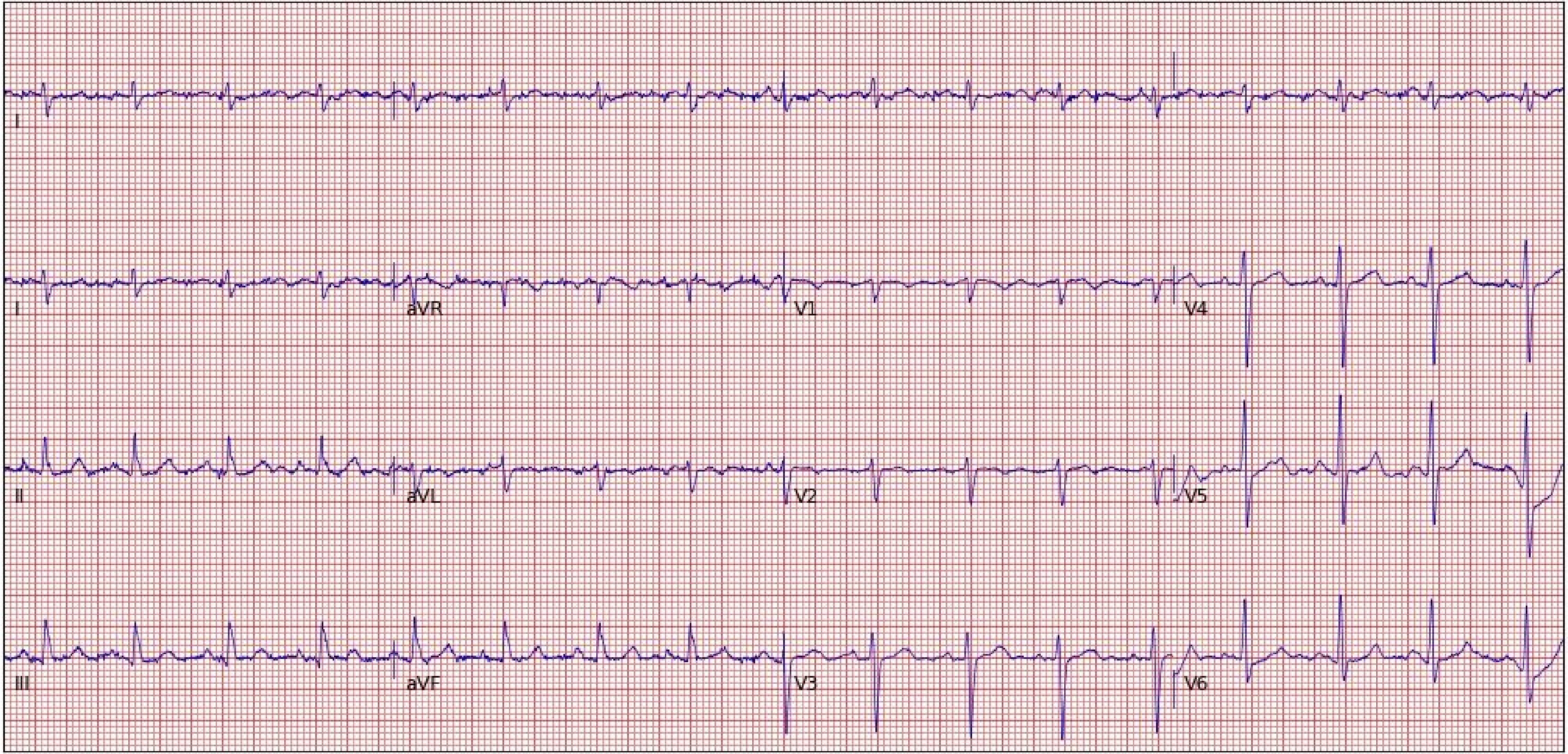
I
I
aVR
V1
V4
II
aVL
V2
V5
III
aVF
V3
V6

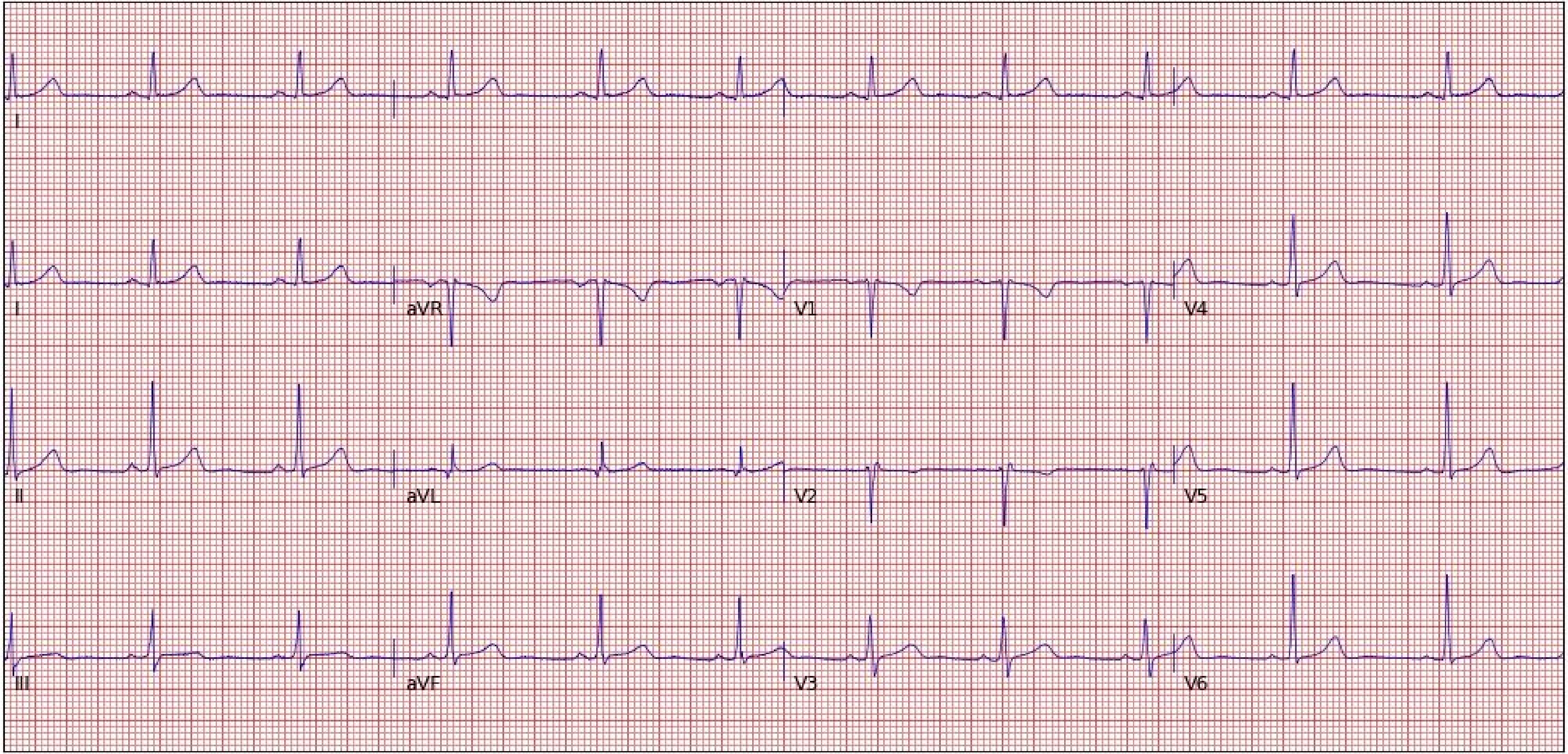
I
I
aVR
V1
V4
II
aVL
V2
V5
III
aVF
V3
V6

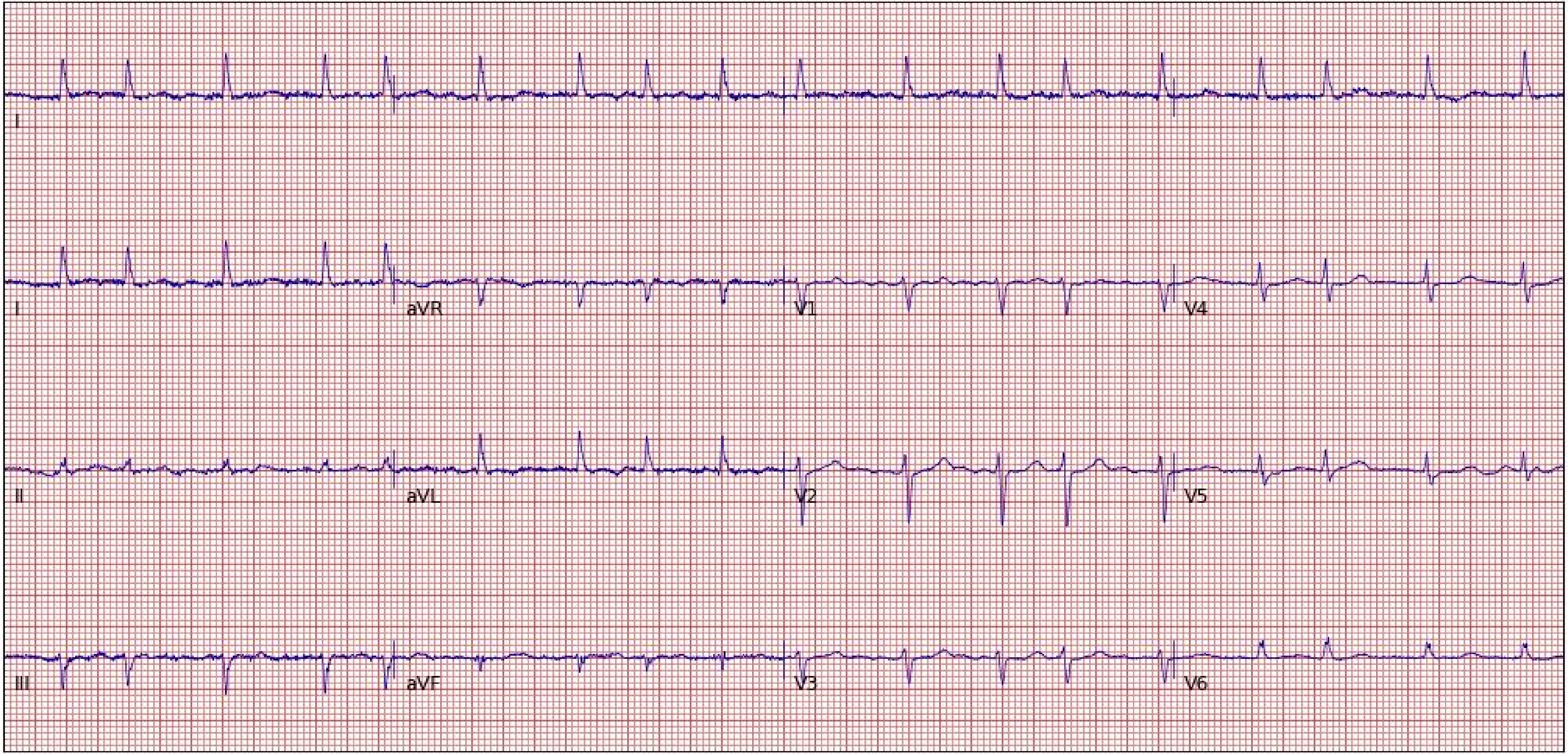
I
I
aVR
V1
V4
II
aVL
V2
V5
III
aVF
V3
V6

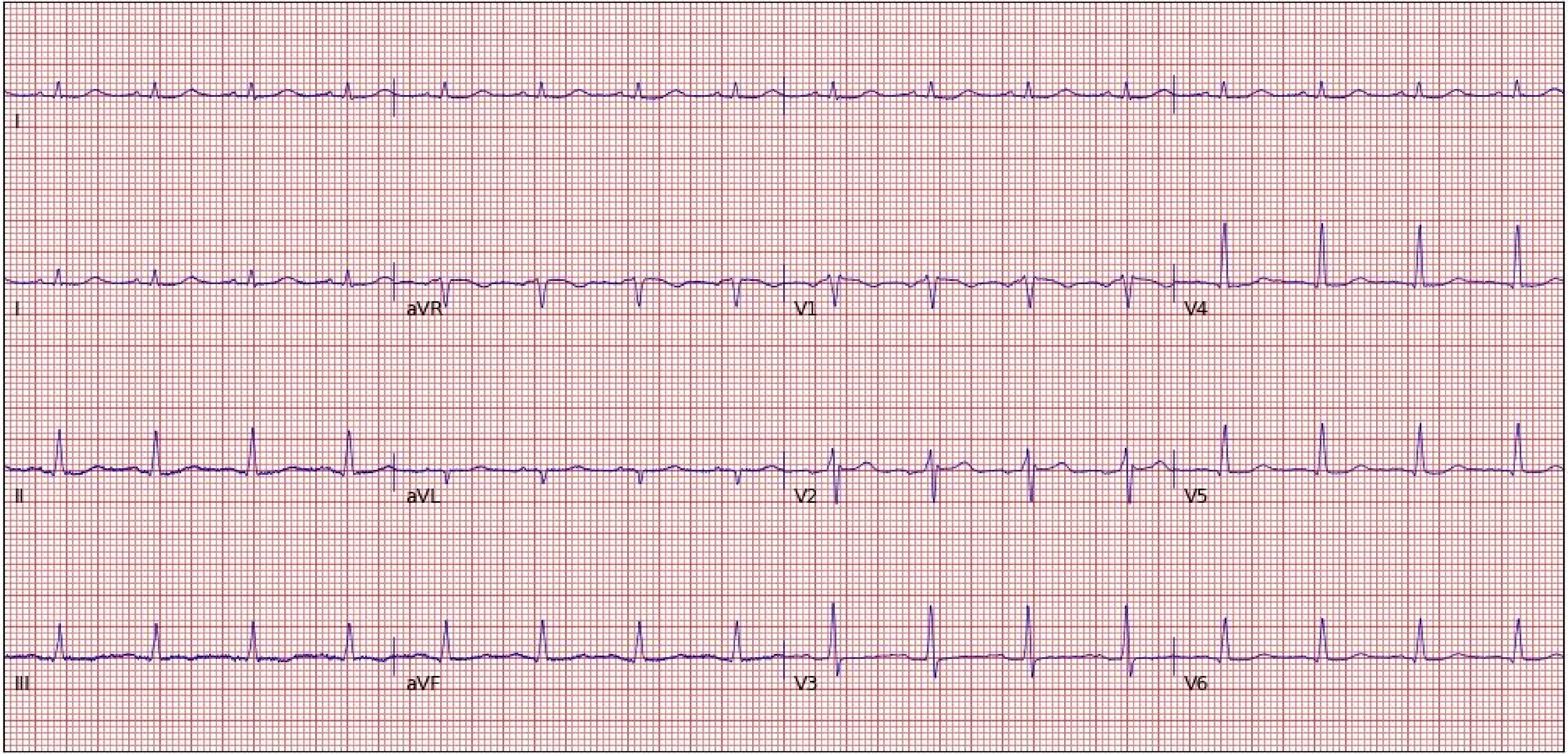

I
I
aVR
V1
V4
II
aVL
V2
V5
III
aVF
V3
V6

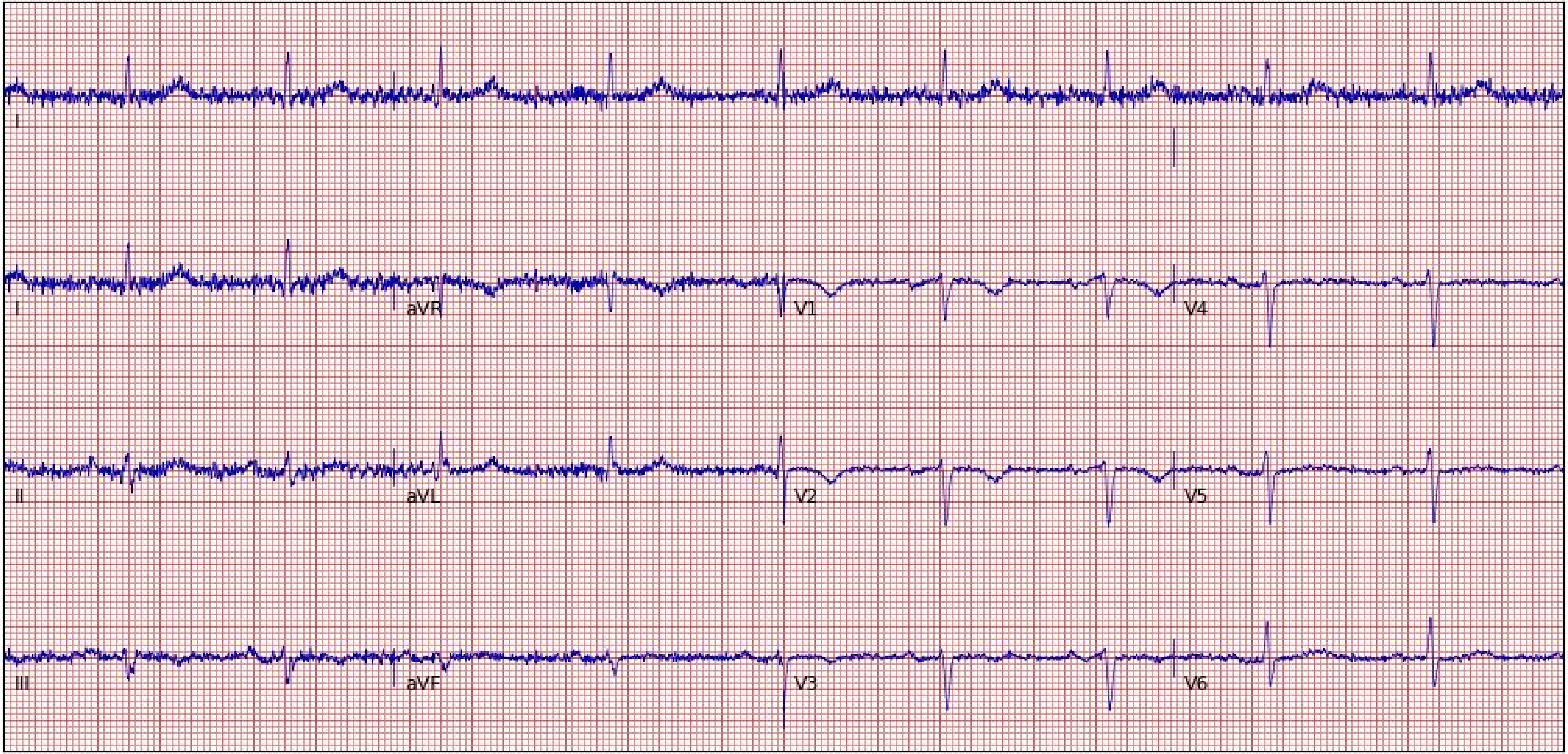
I
I
aVR
V1
V4
II
aVL
V2
V5
III
aVF
V3
V6

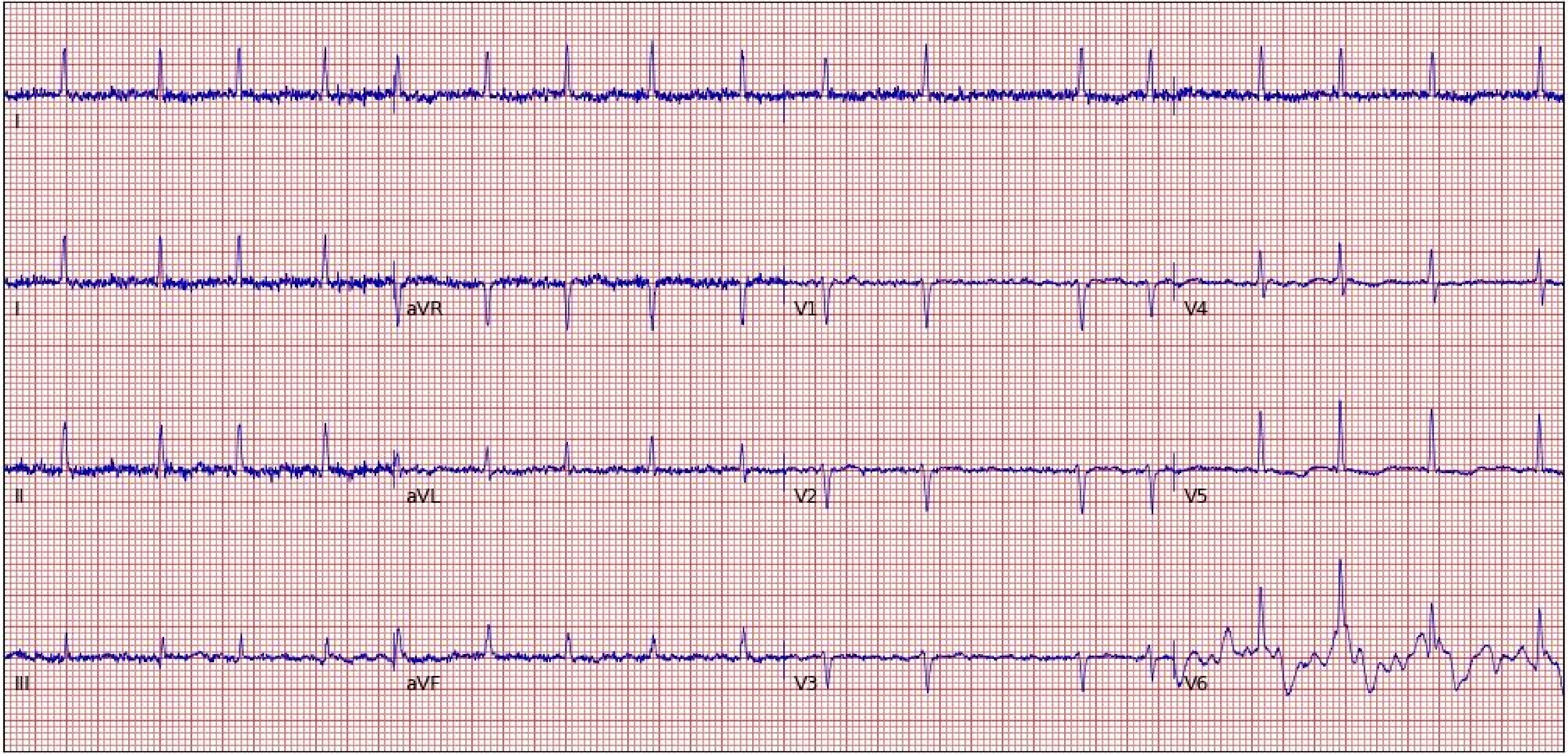
I
I
aVR
V1
V4
II
aVL
V2
V5
III
aVF
V3
V6

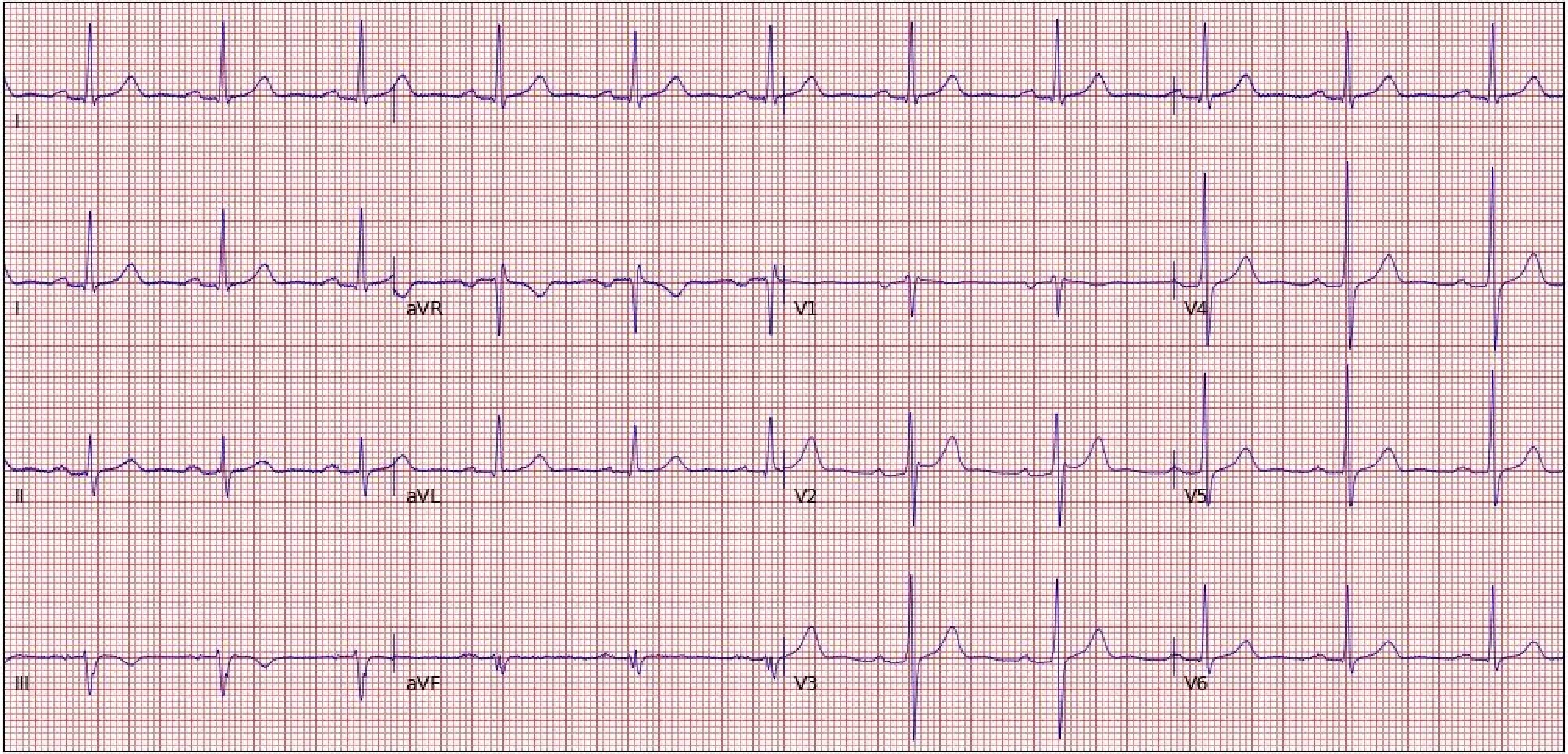

I
II
aVR
V1
V4
II
aVL
V2
V5
III
aVF
V3
V6

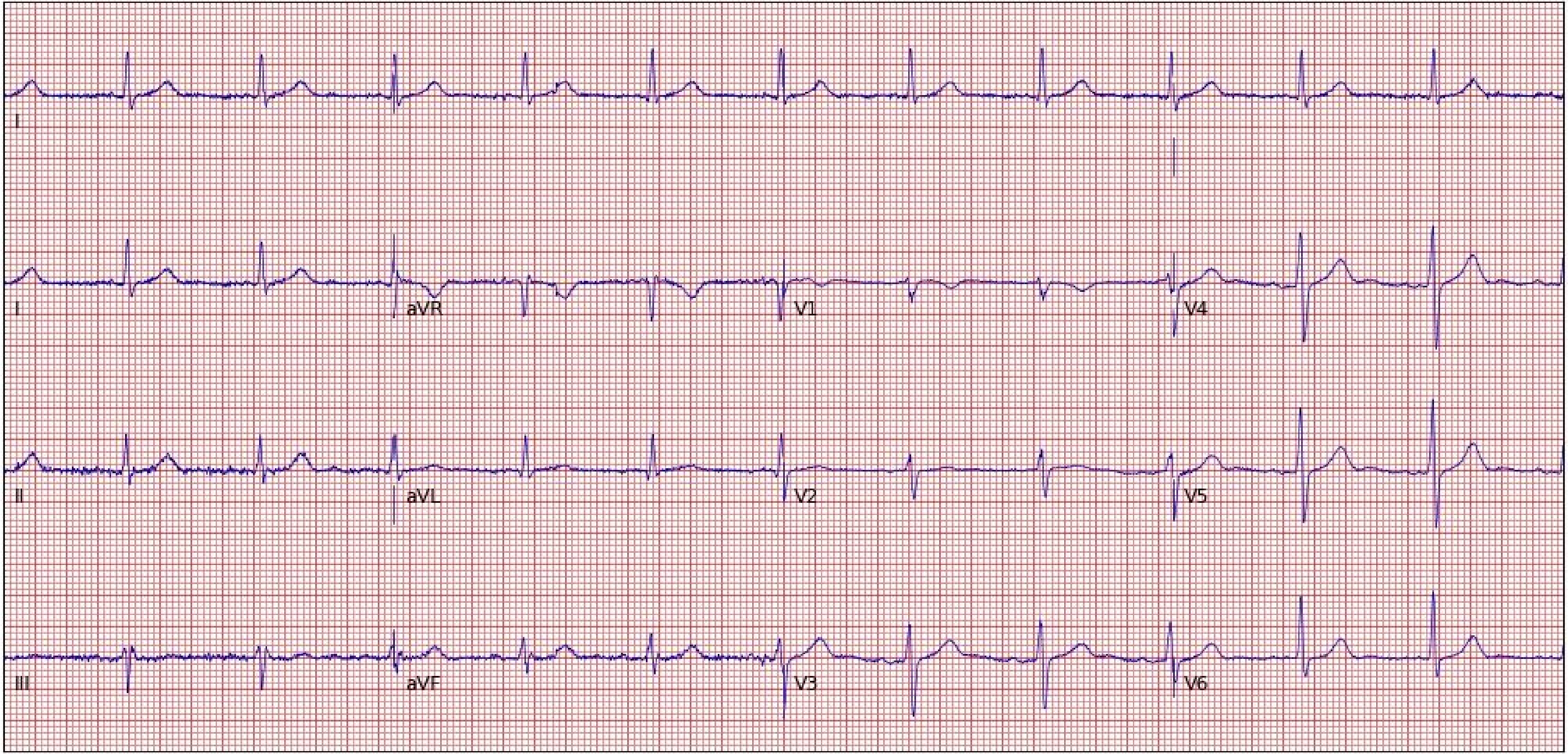
I
I
aVR
V1
V4
II
aVL
V2
V5
III
aVF
V3
V6

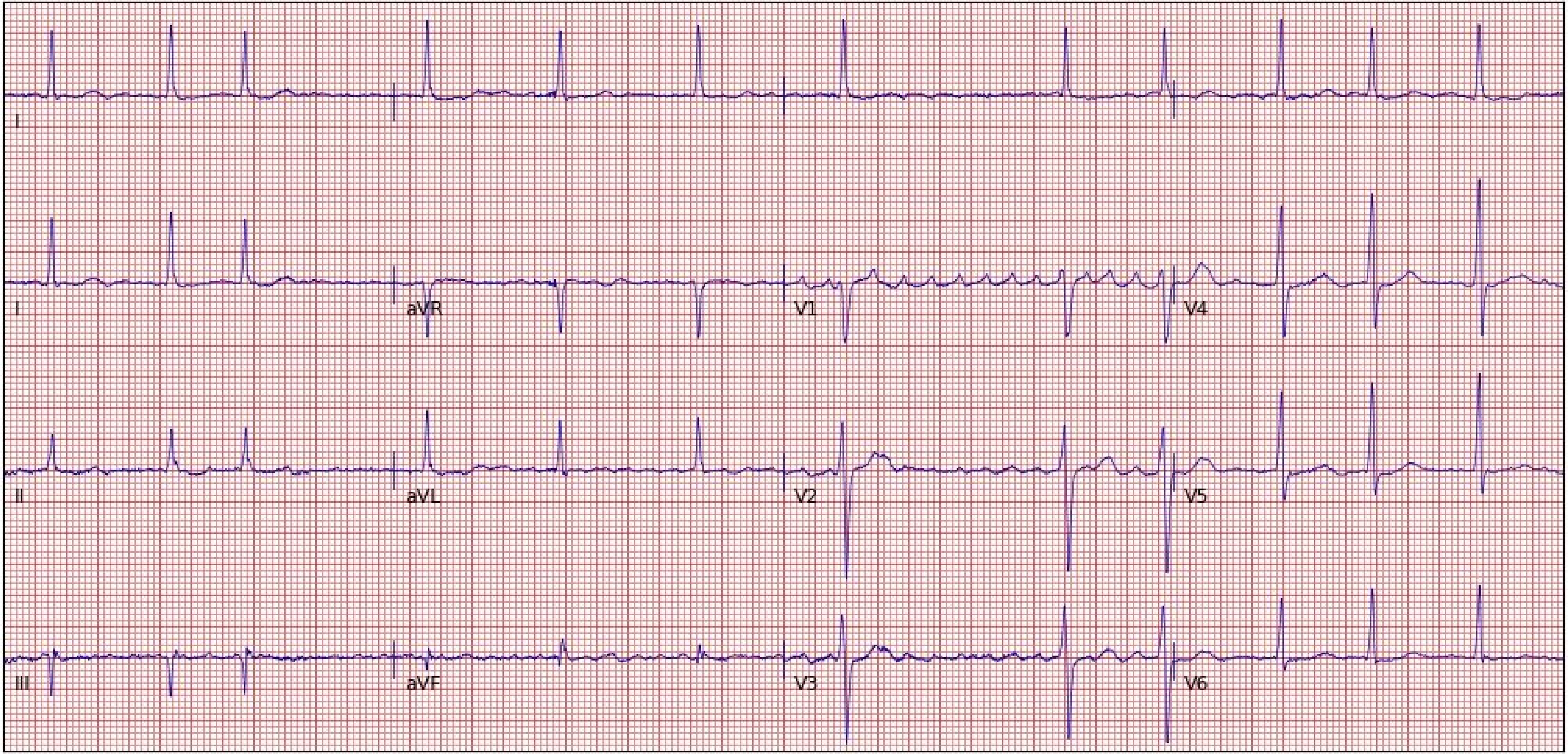
I
I
aVR
V1
V4
II
aVL
V2
V5
III
aVF
V3
V6

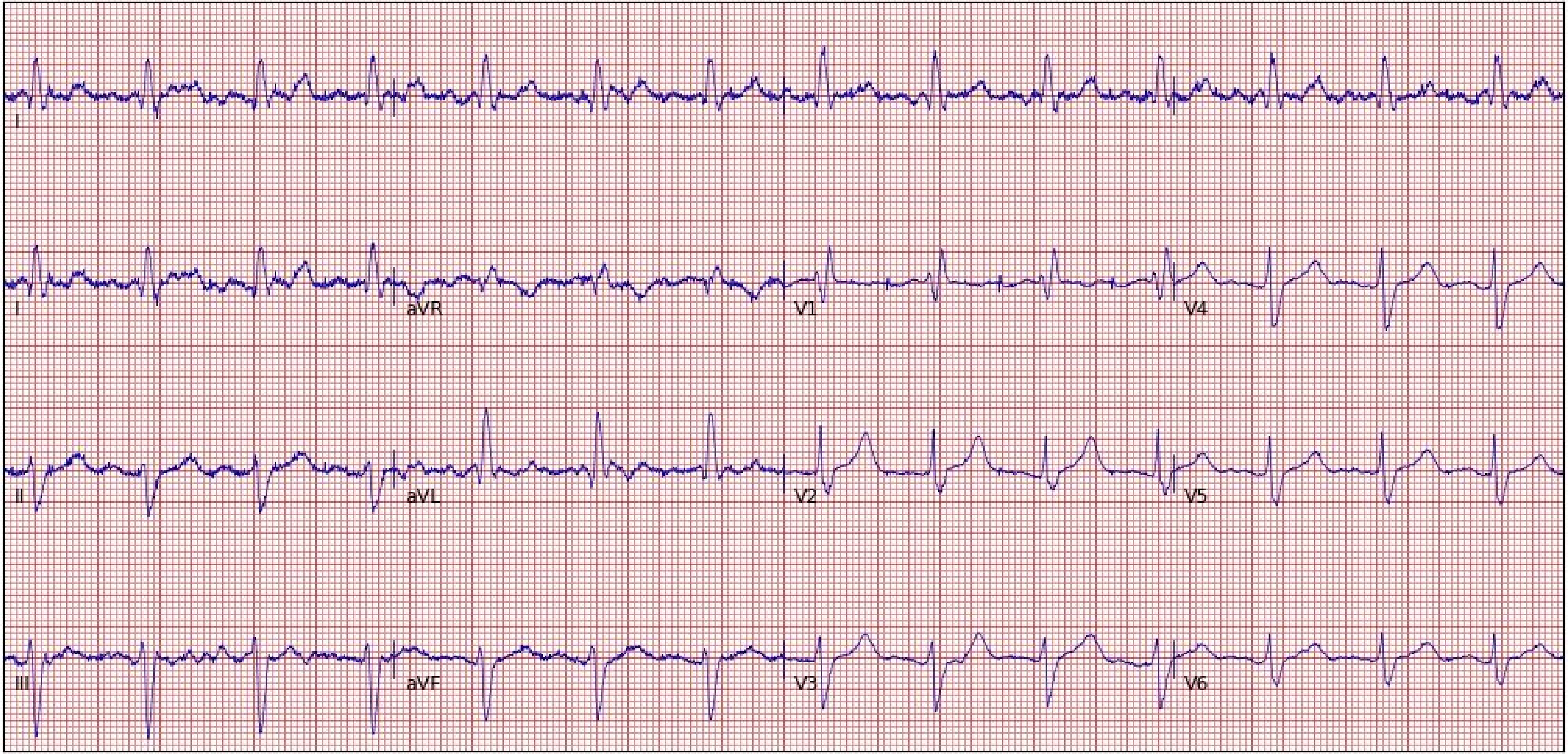

I
I
aVR
V1
V4
II
aVL
V2
V5
III
aVF
V3
V6

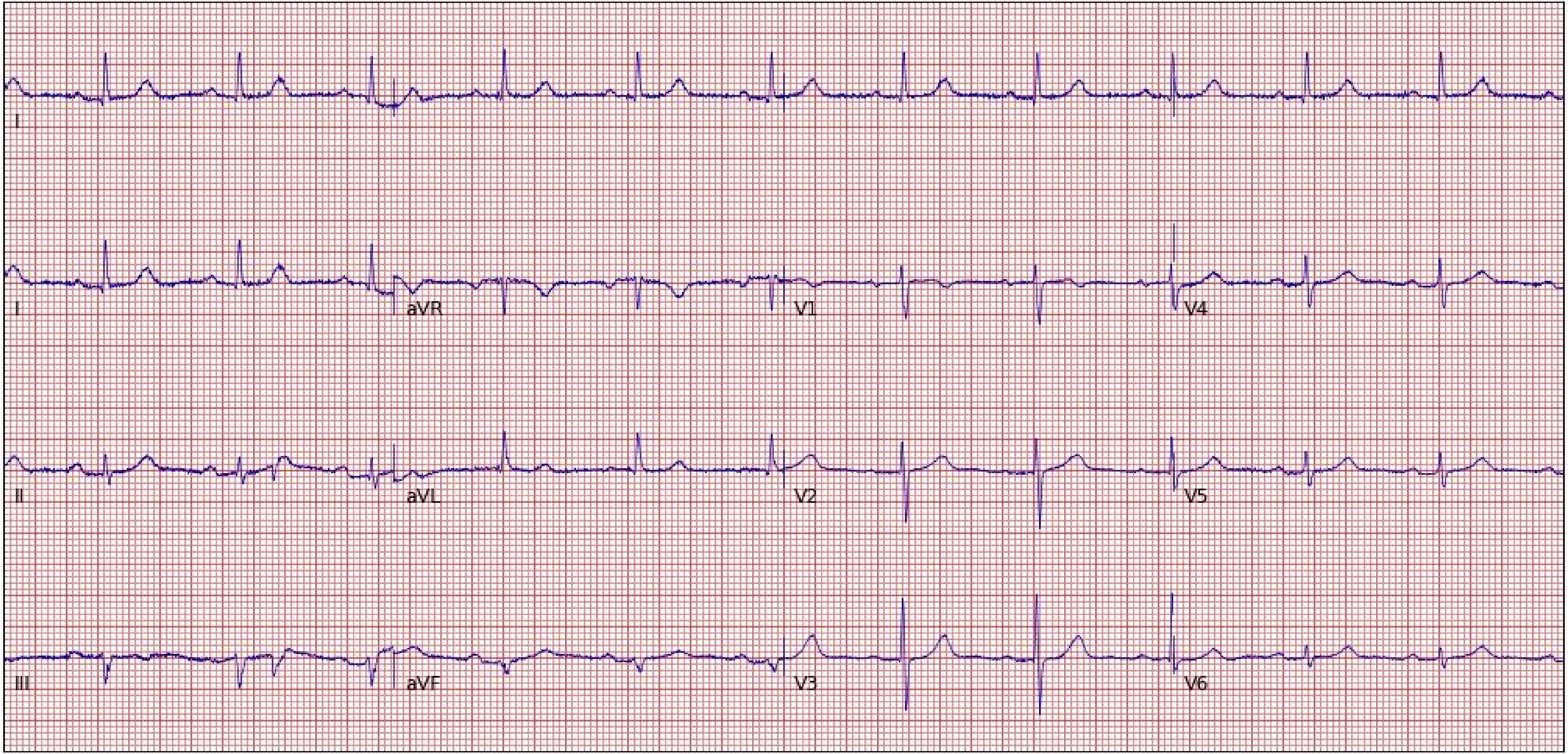

I
I
aVR
V1
V4
II
aVL
V2
V5
III
aVF
V3
V6

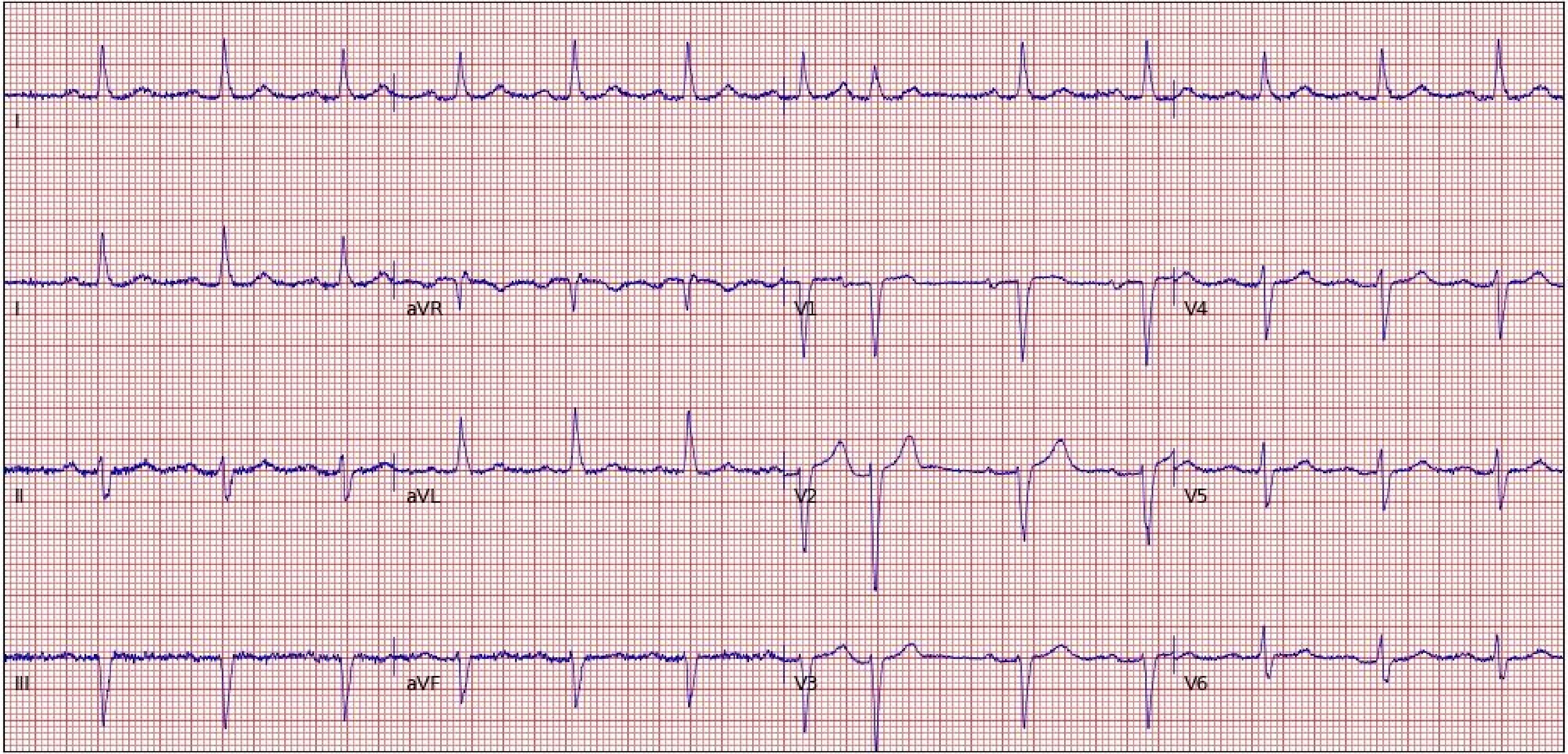
I
I
aVR
V1
V4
II
aVL
V2
V5
III
aVF
V3
V6

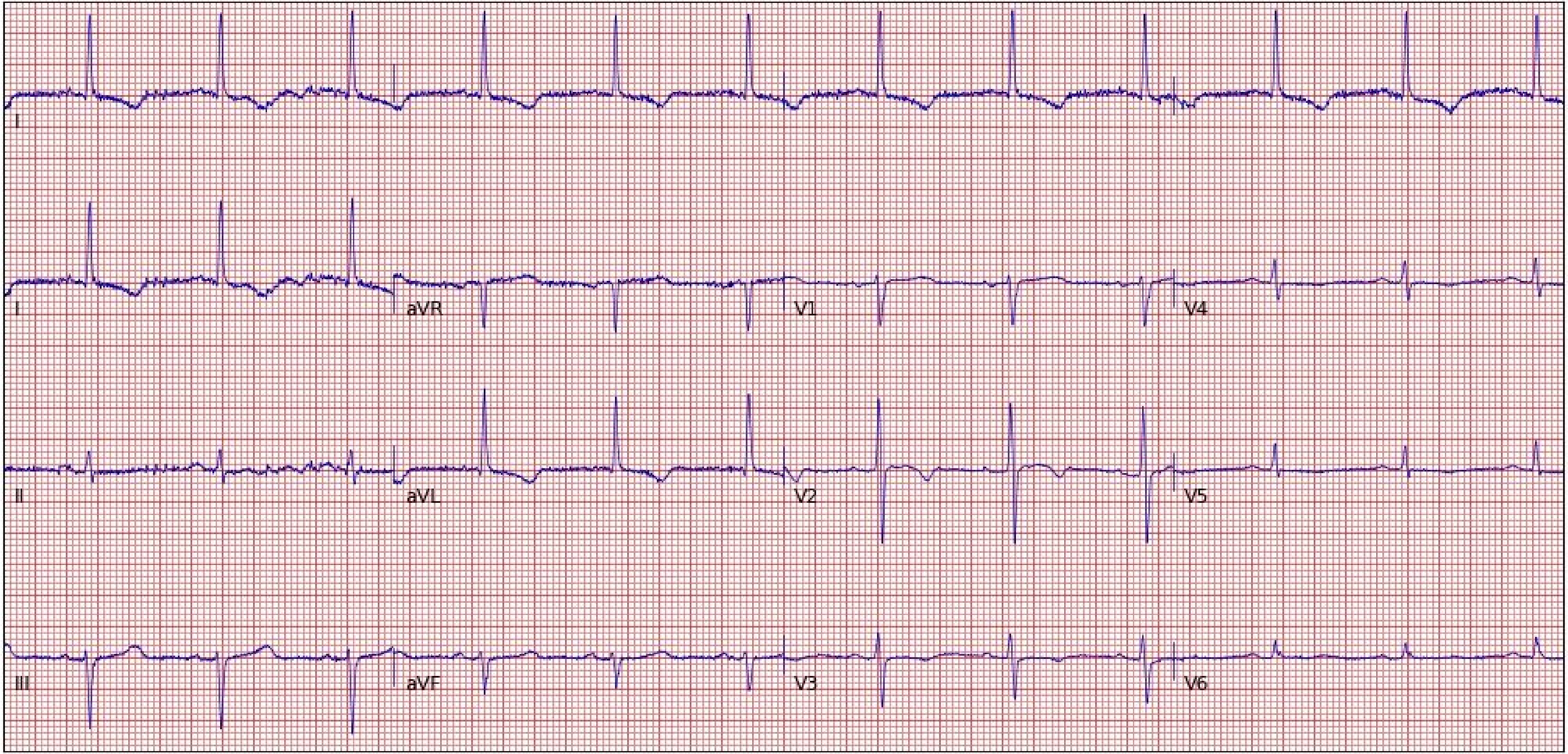
I
I
aVR
V1
V4
II
aVL
V2
V5
III
aVF
V3
V6

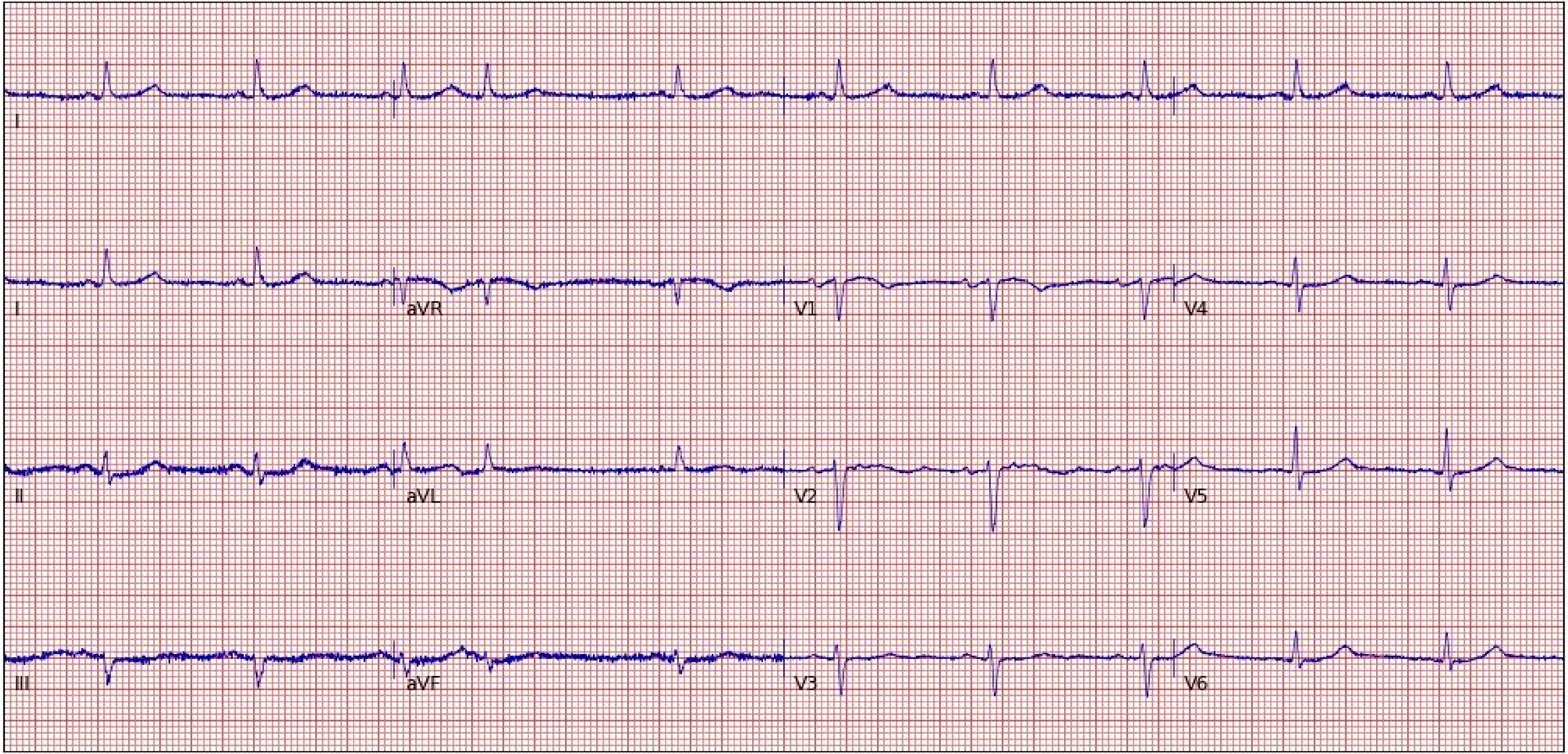
I
I
aVR
V1
V4
II
aVL
V2
V5
III
aVF
V3
V6

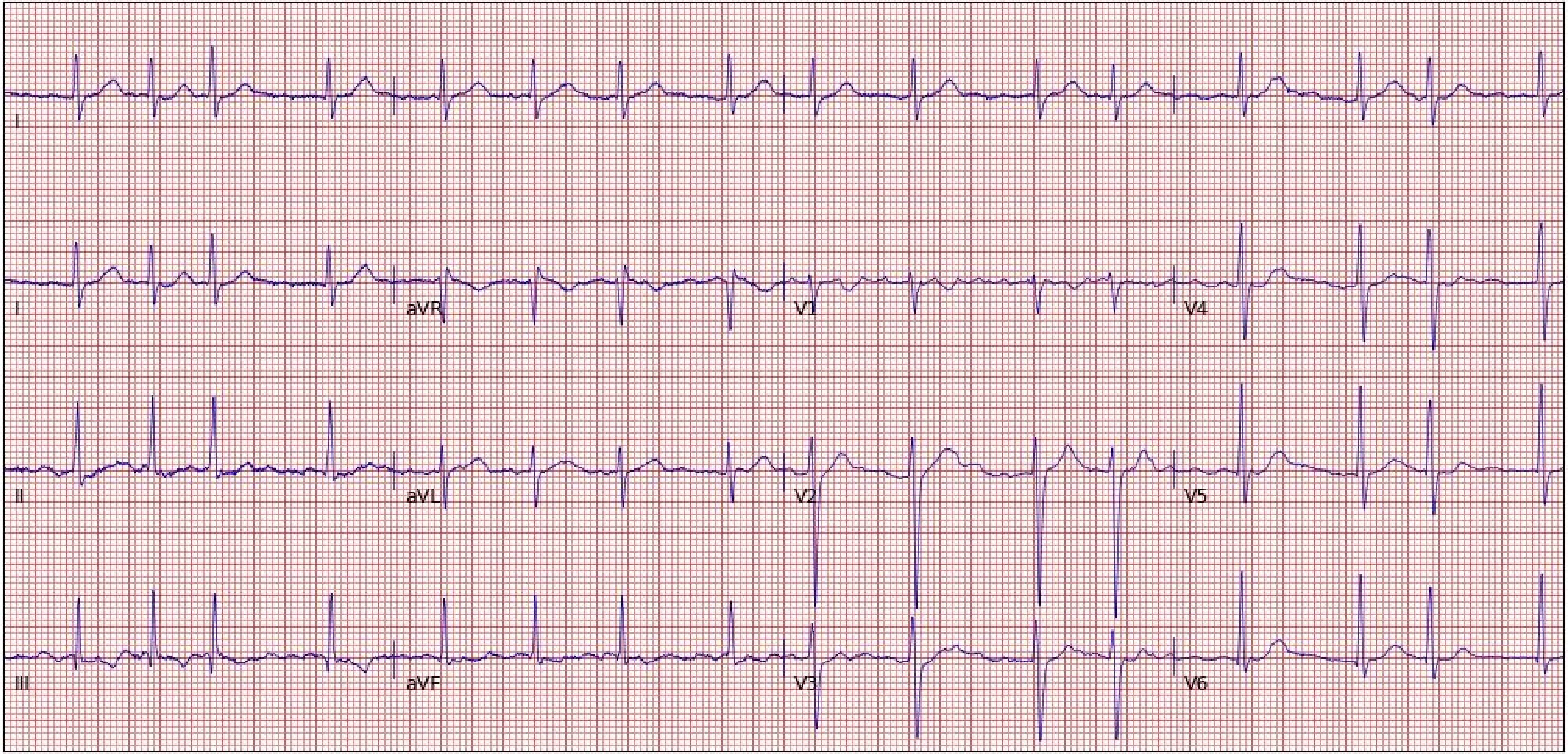
I
I
aVR
V1
V4
II
aVL
V2
V5
III
aVF
V3
V6

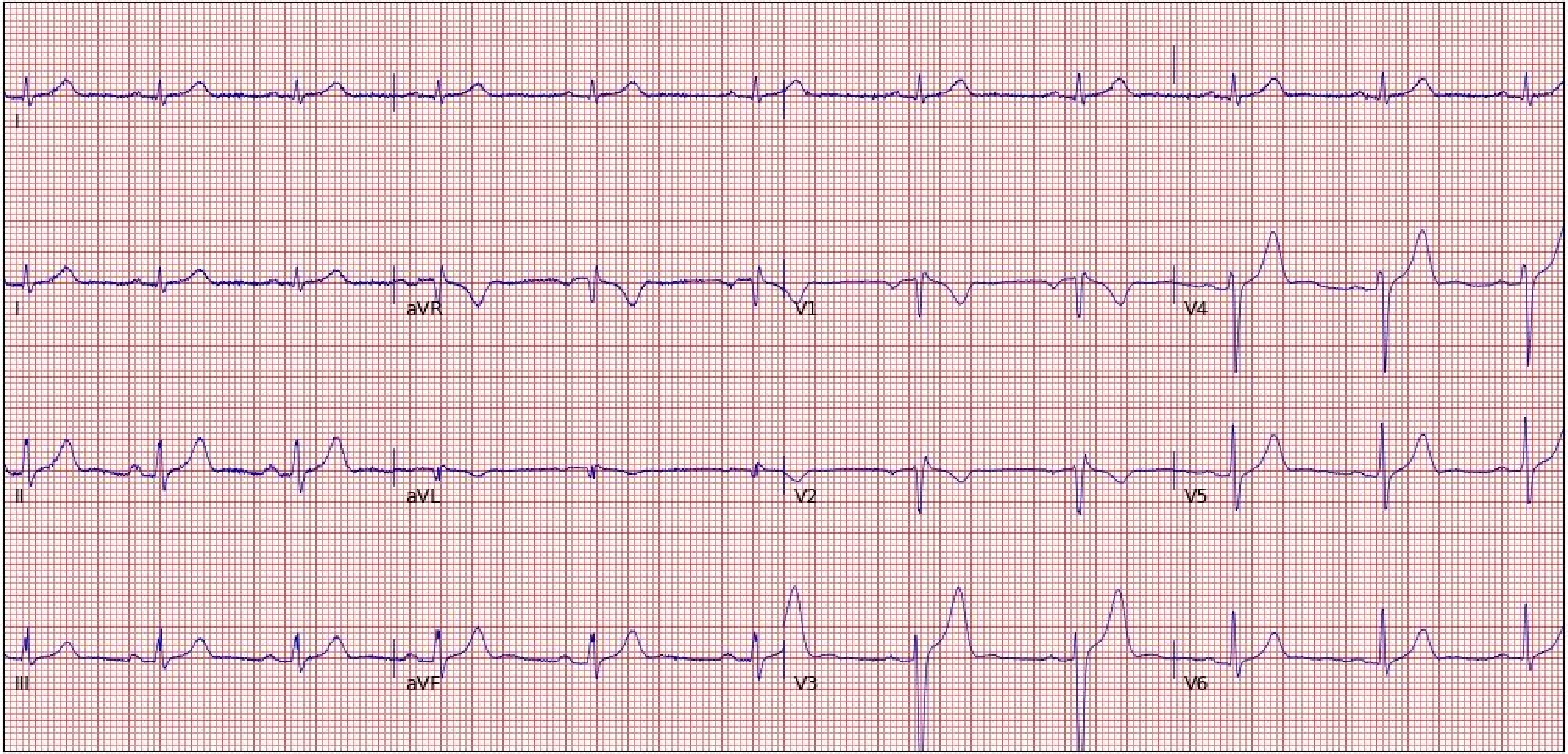
I
I
aVR
V1
V4
II
aVL
V2
V5
III
aVF
V3
V6

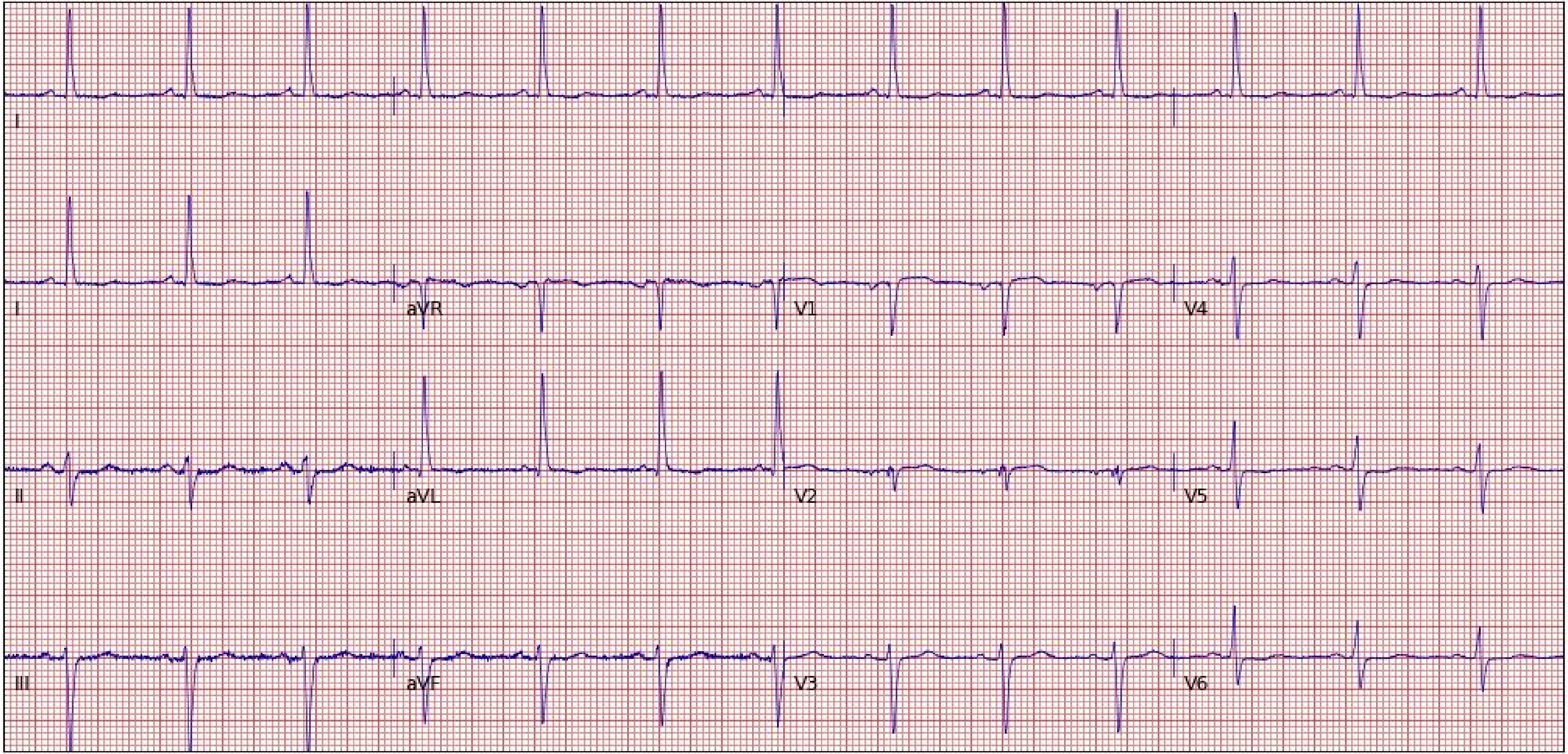
I
I
aVR
V1
V4
II
aVL
V2
V5
III
aVF
V3
V6

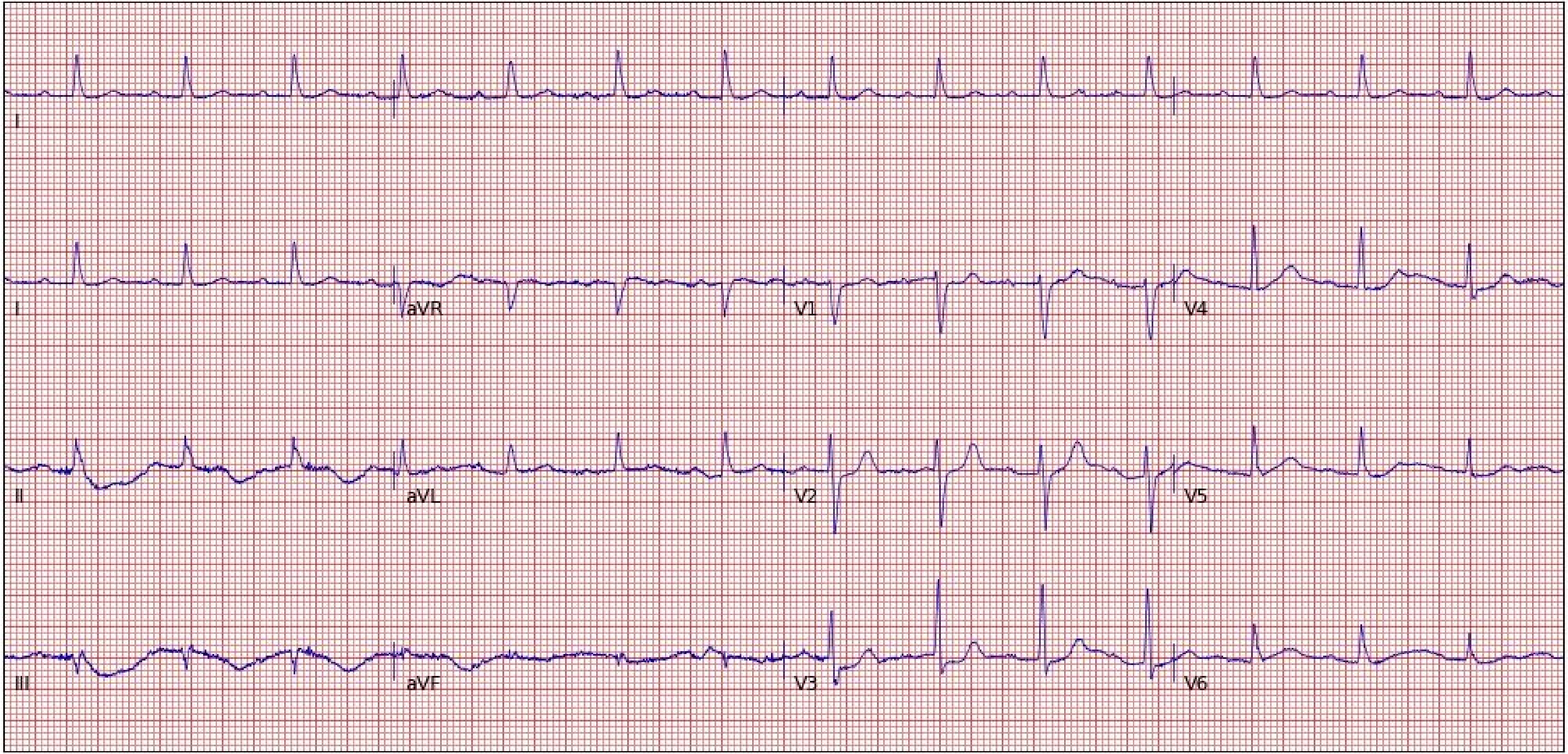
I
I
aVR
V1
V4
II
aVL
V2
V5
III
aVF
V3
V6

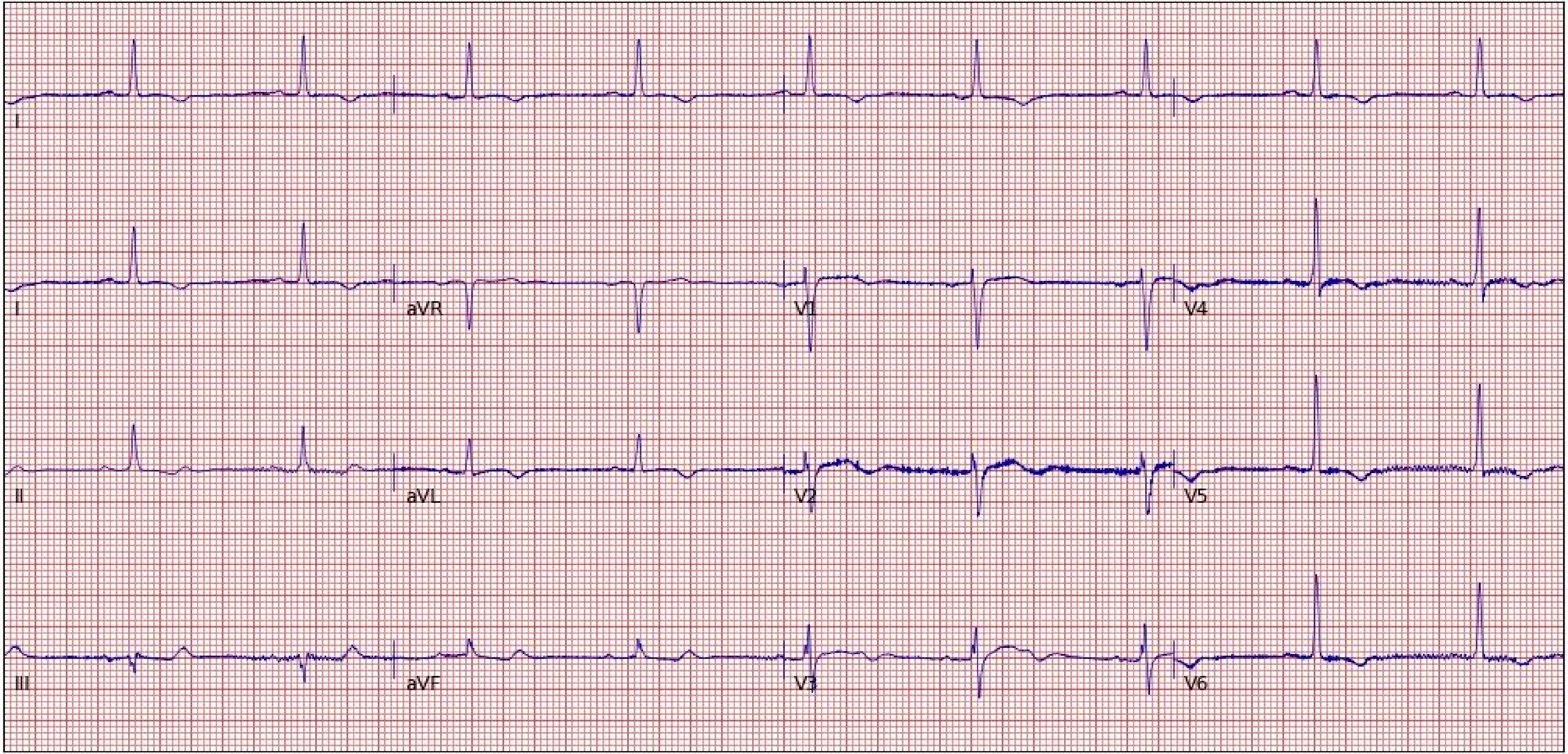
I
I
aVR
V1
V4
II
aVL
V2
V5
III
aVF
V3
V6

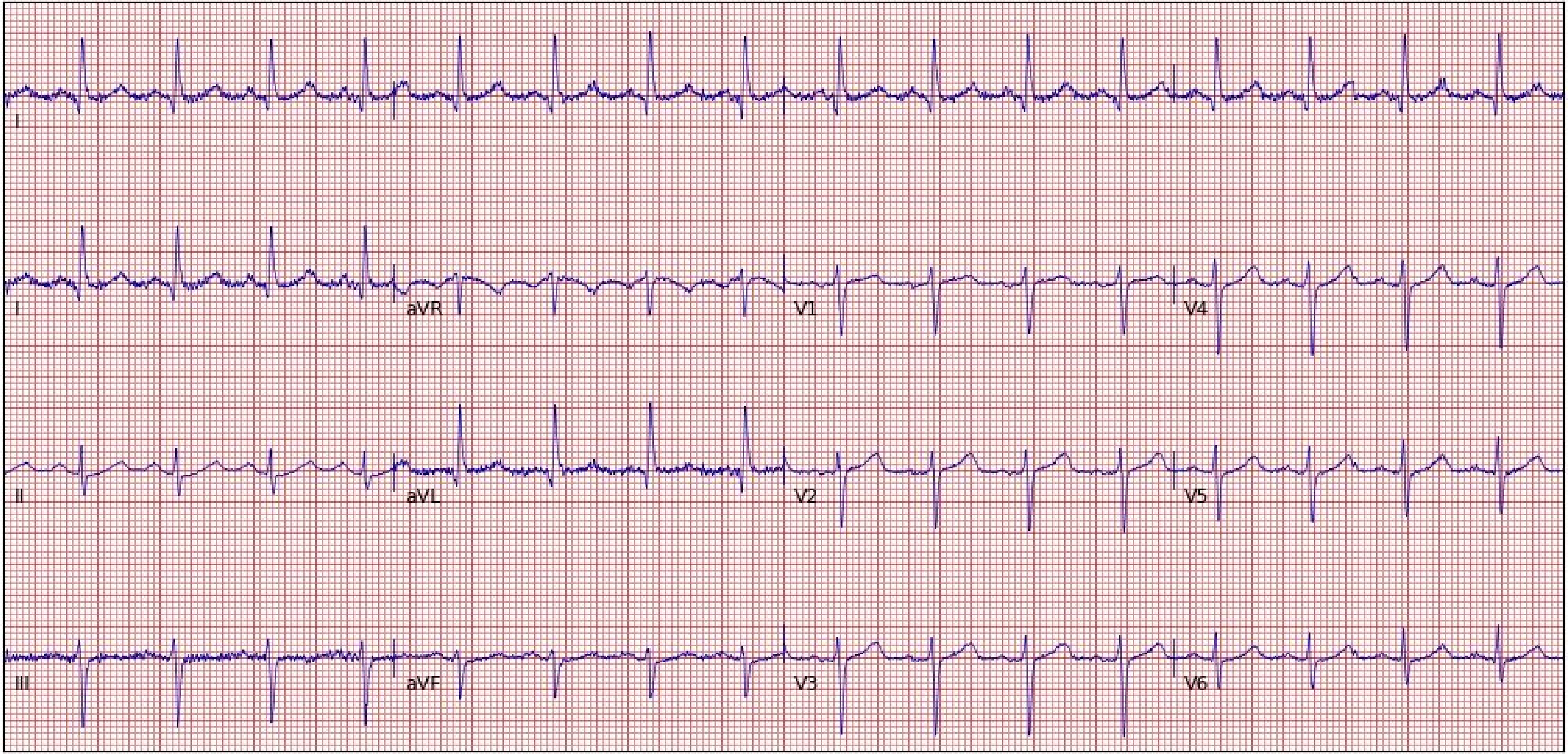
I
I
aVR
V1
V4
II
aVL
V2
V5
III
aVF
V3
V6

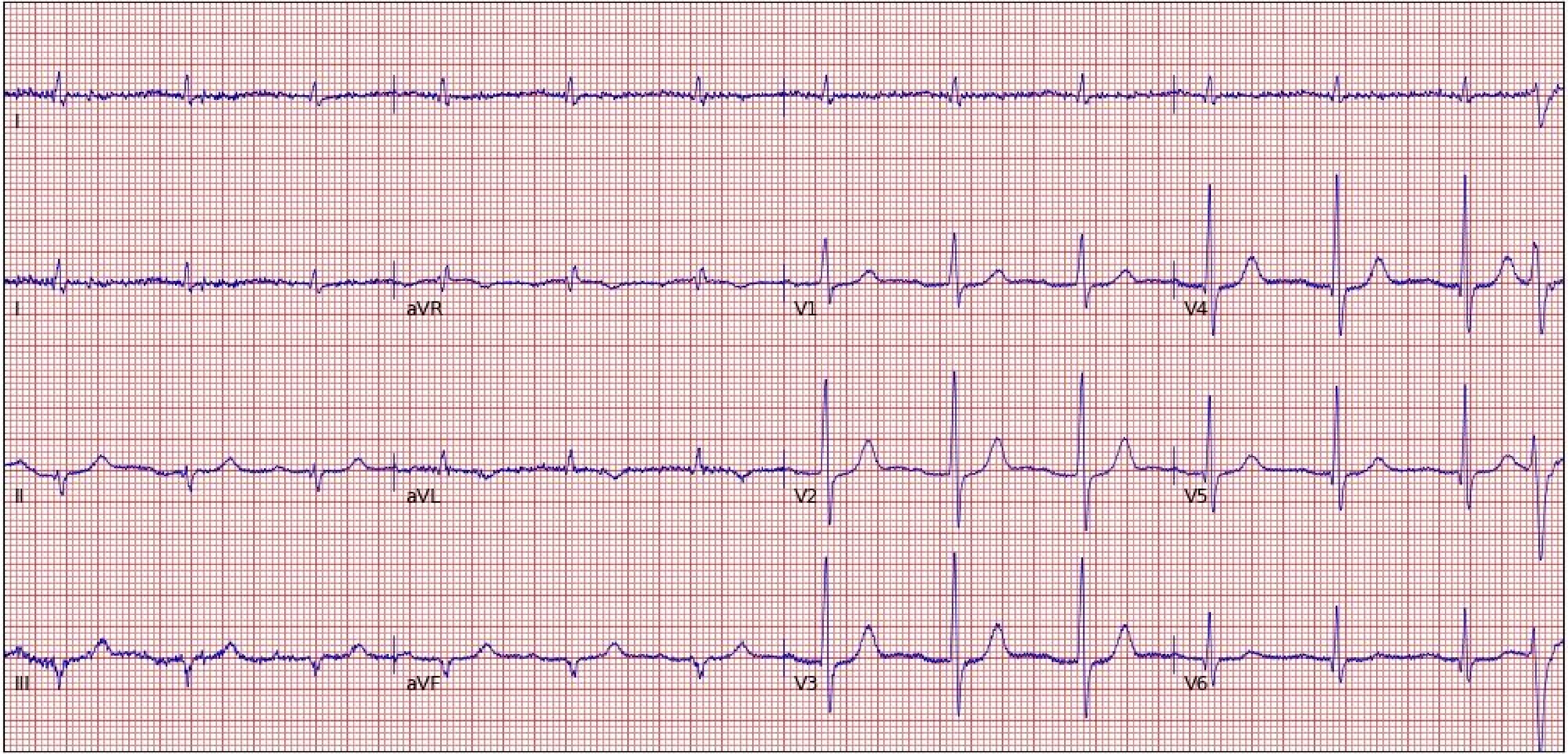

I
I
aVR
V1
V4
II
aVL
V2
V5
III
aVF
V3
V6

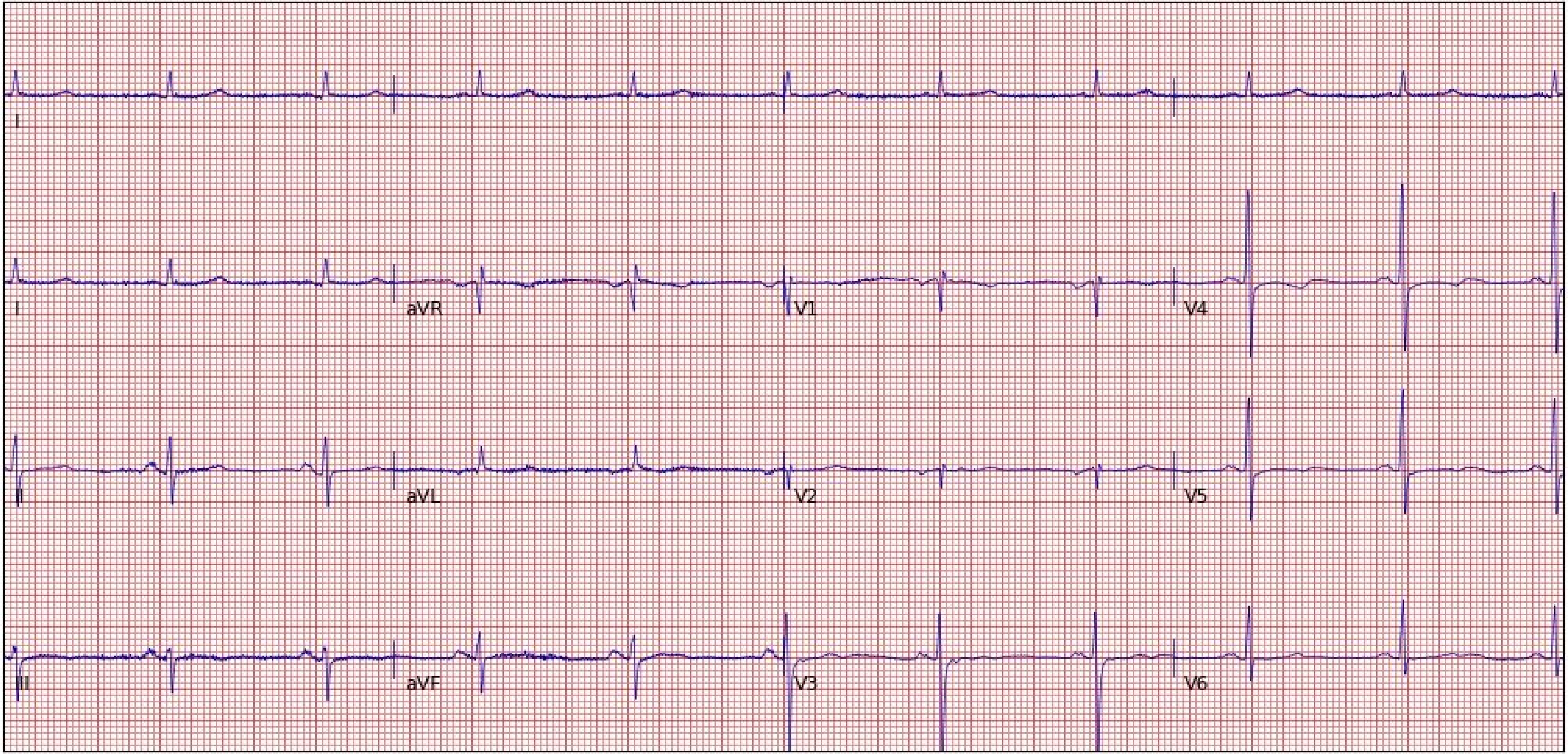

I
I
aVR
V1
V4
II
aVL
V2
V5
III
aVF
V3
V6

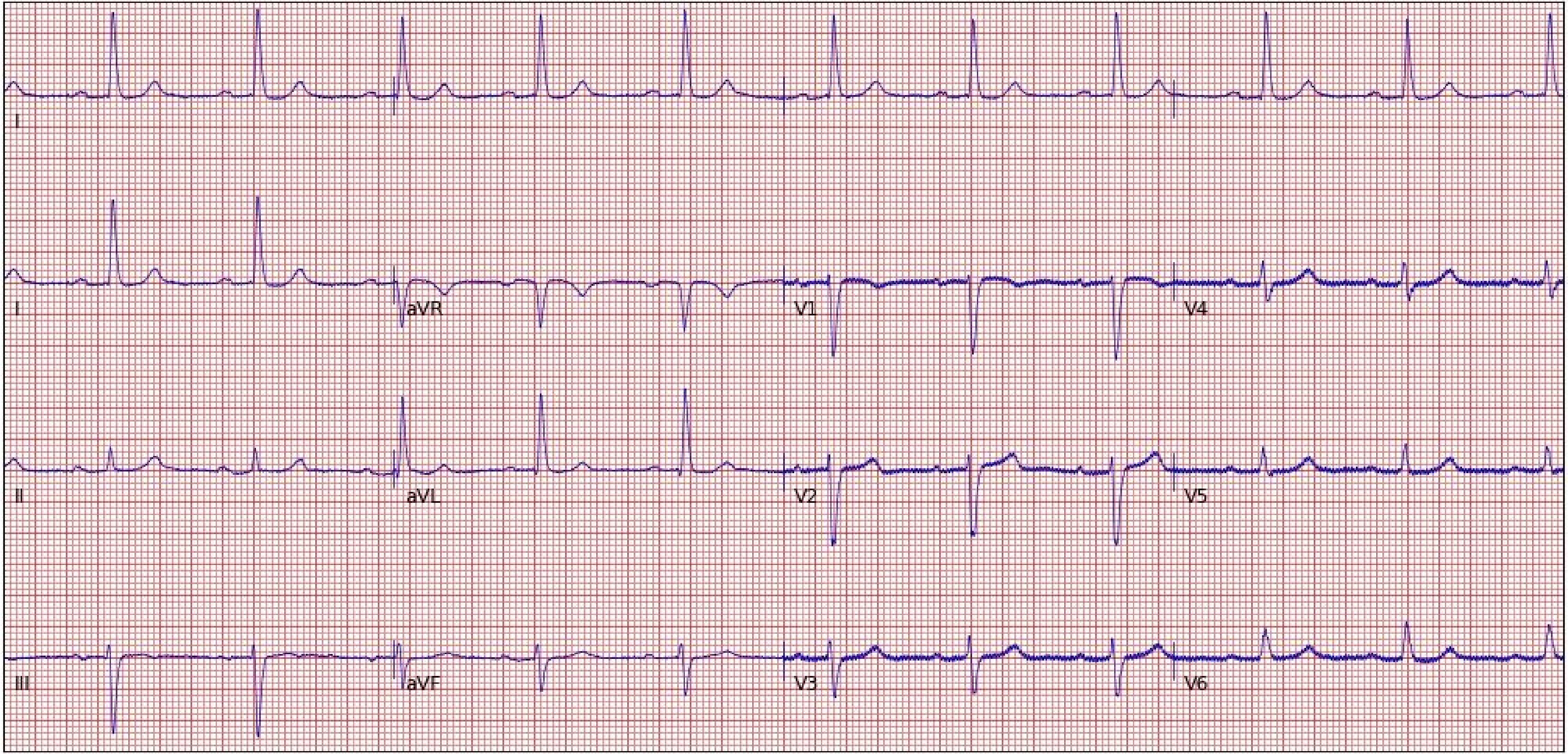
I
I
aVR
V1
V4
II
aVL
V2
V5
III
aVF
V3
V6

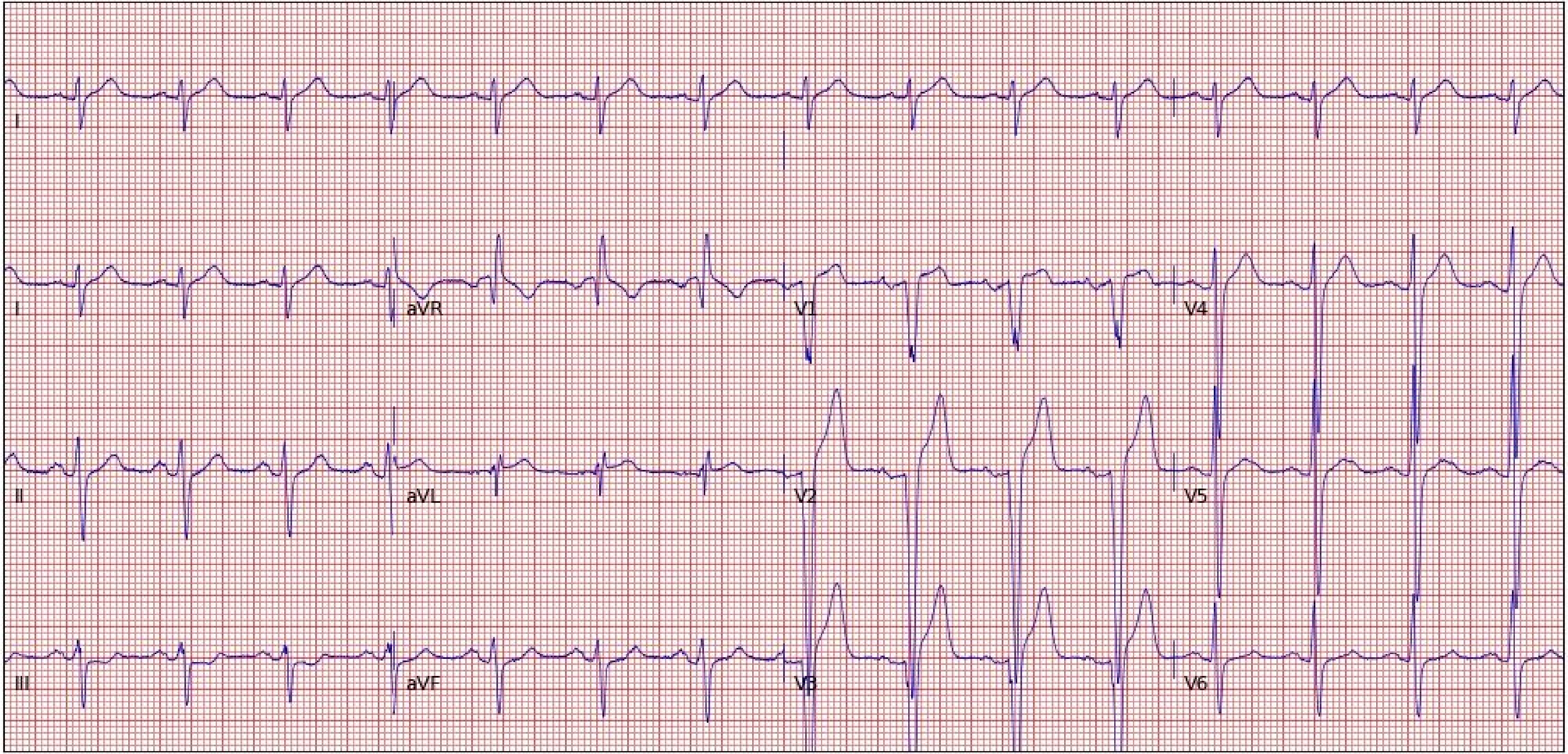
I
I
aVR
V1
V4
II
aVL
V2
V5
III
aVF
V3
V6

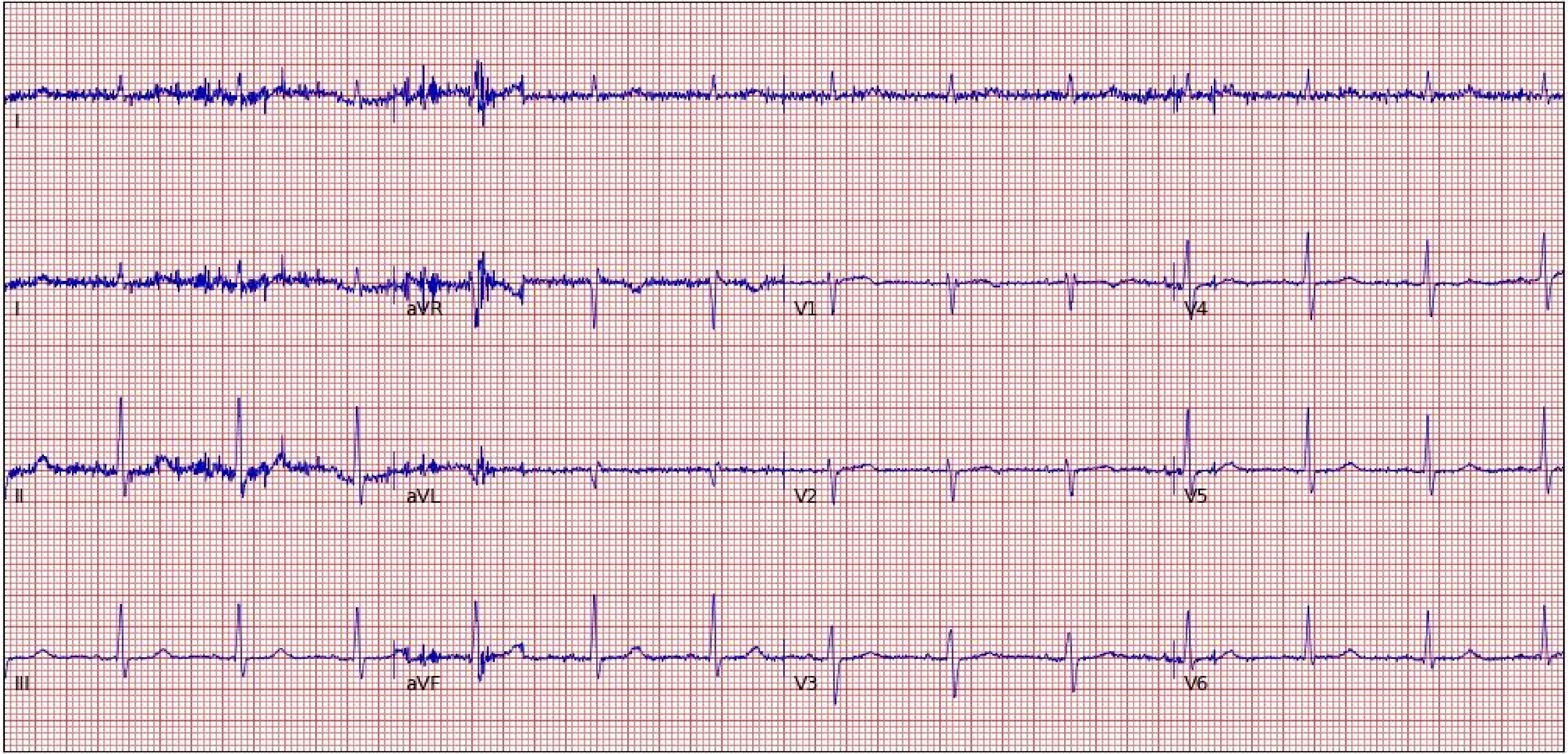
I
I
aVR
V1
V4
II
aVL
V2
V5
III
aVF
V3
V6

**Section 7. Screenshots of 100 electrocardiograms (cropped) from the electronic health record used for model evaluation on real-world images.**

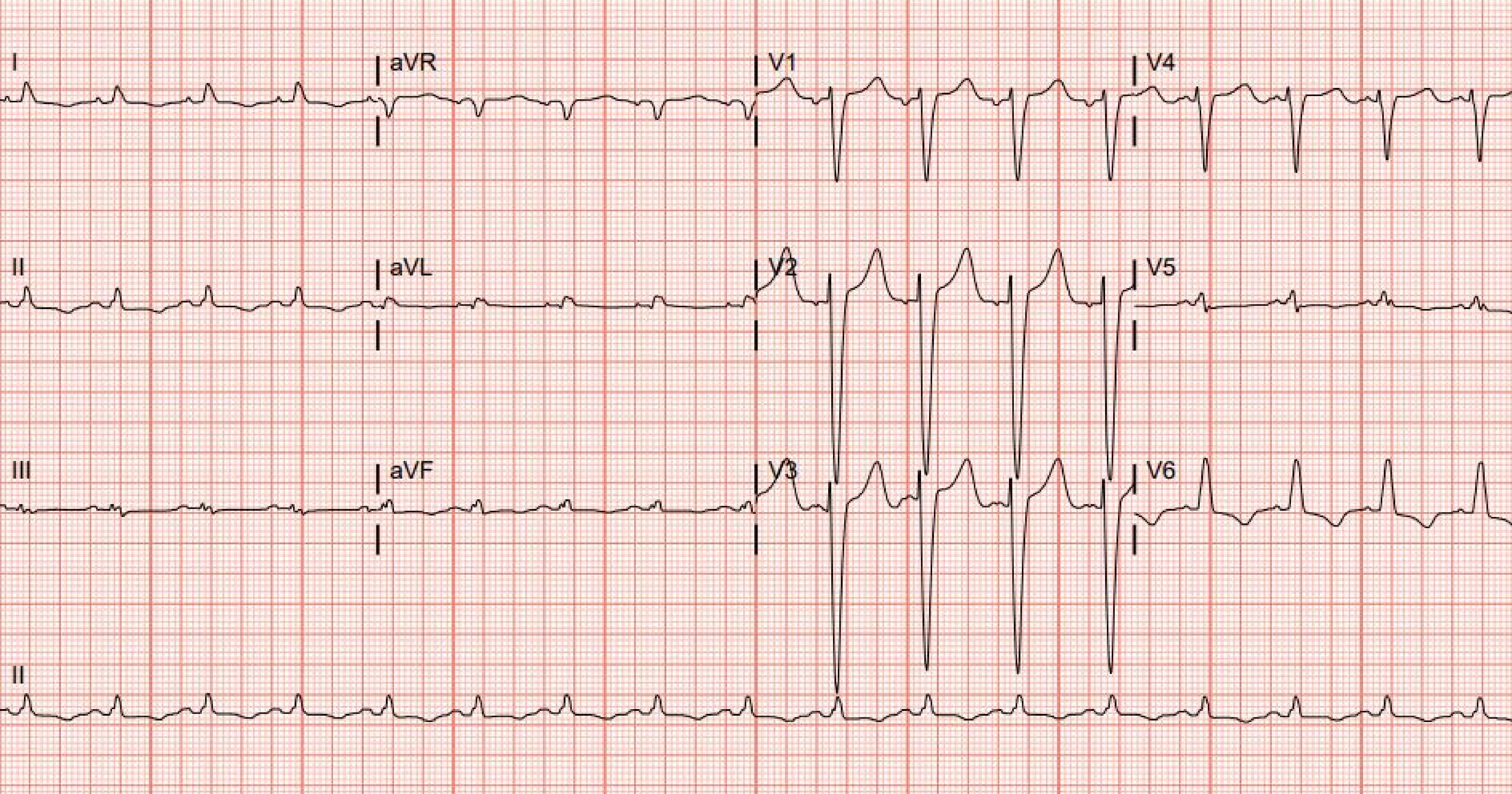
I
aVR
V1
V4
II
aVL
V2
V5
III
aVF
V3
V6
II

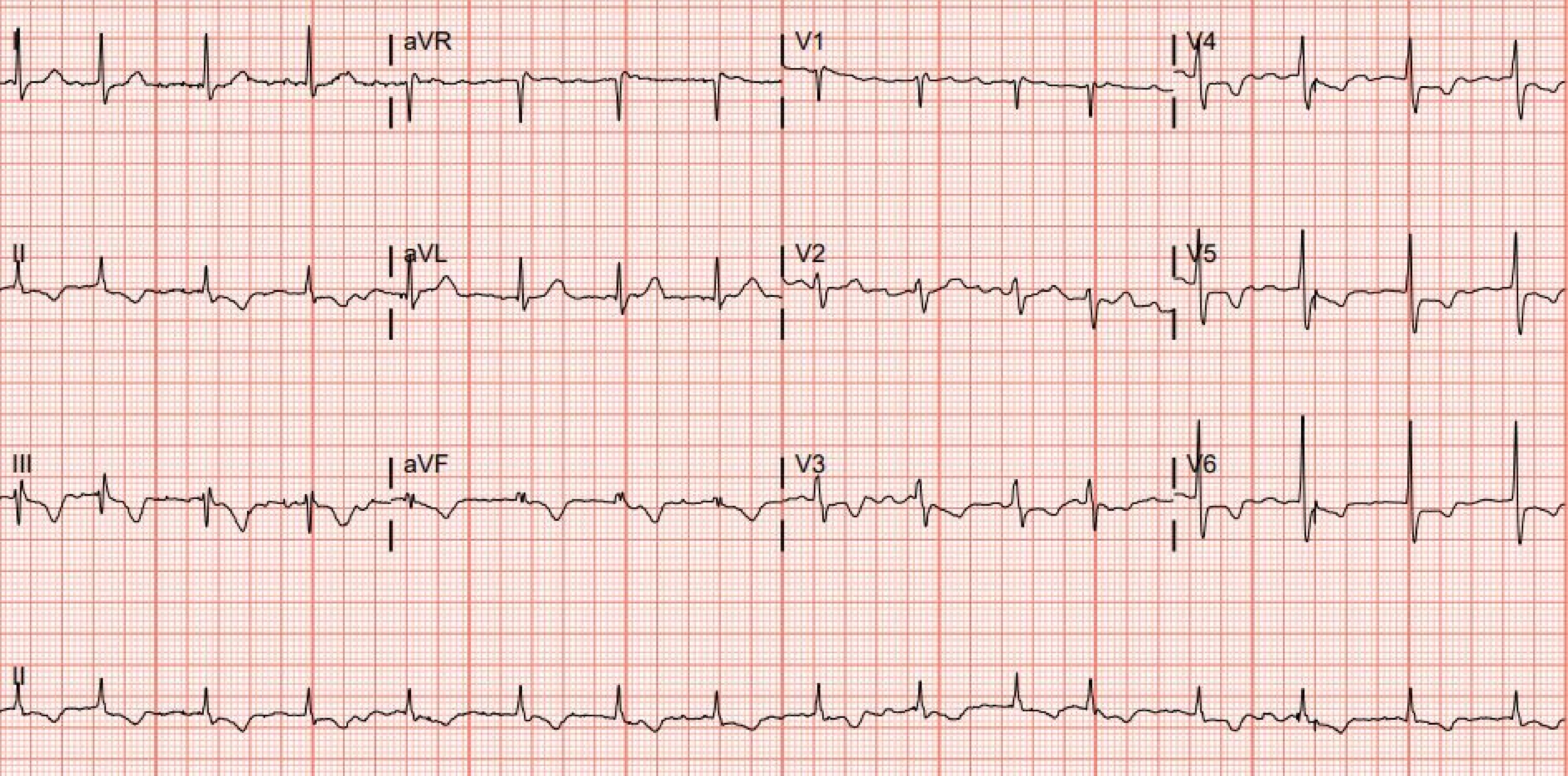
I
aVR
V1
V4
II
aVL
V2
V5
III
aVF
V3
V6
II

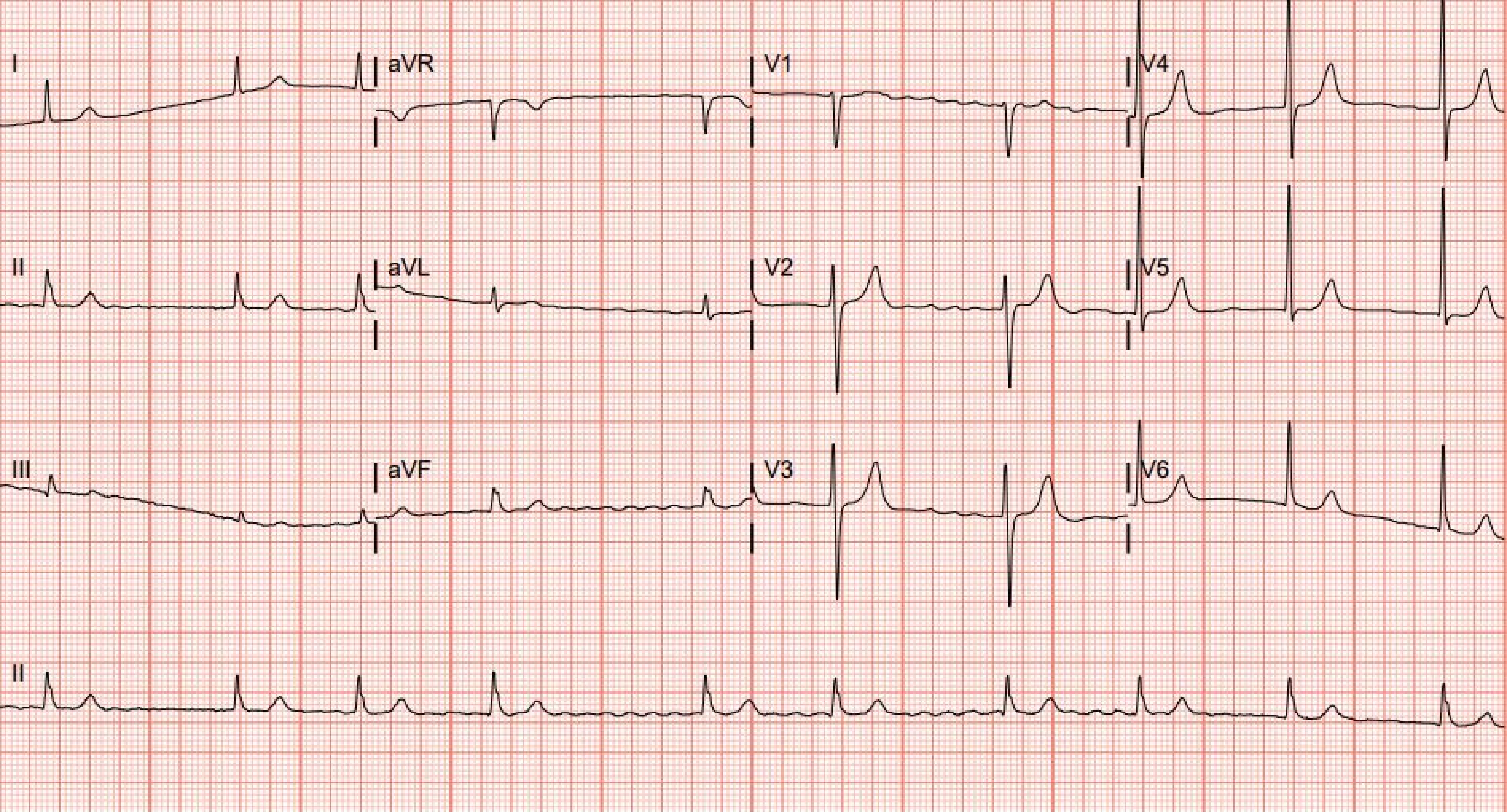
I
aVR
V1
V4
II
aVL
V2
V5
III
aVF
V3
V6
II

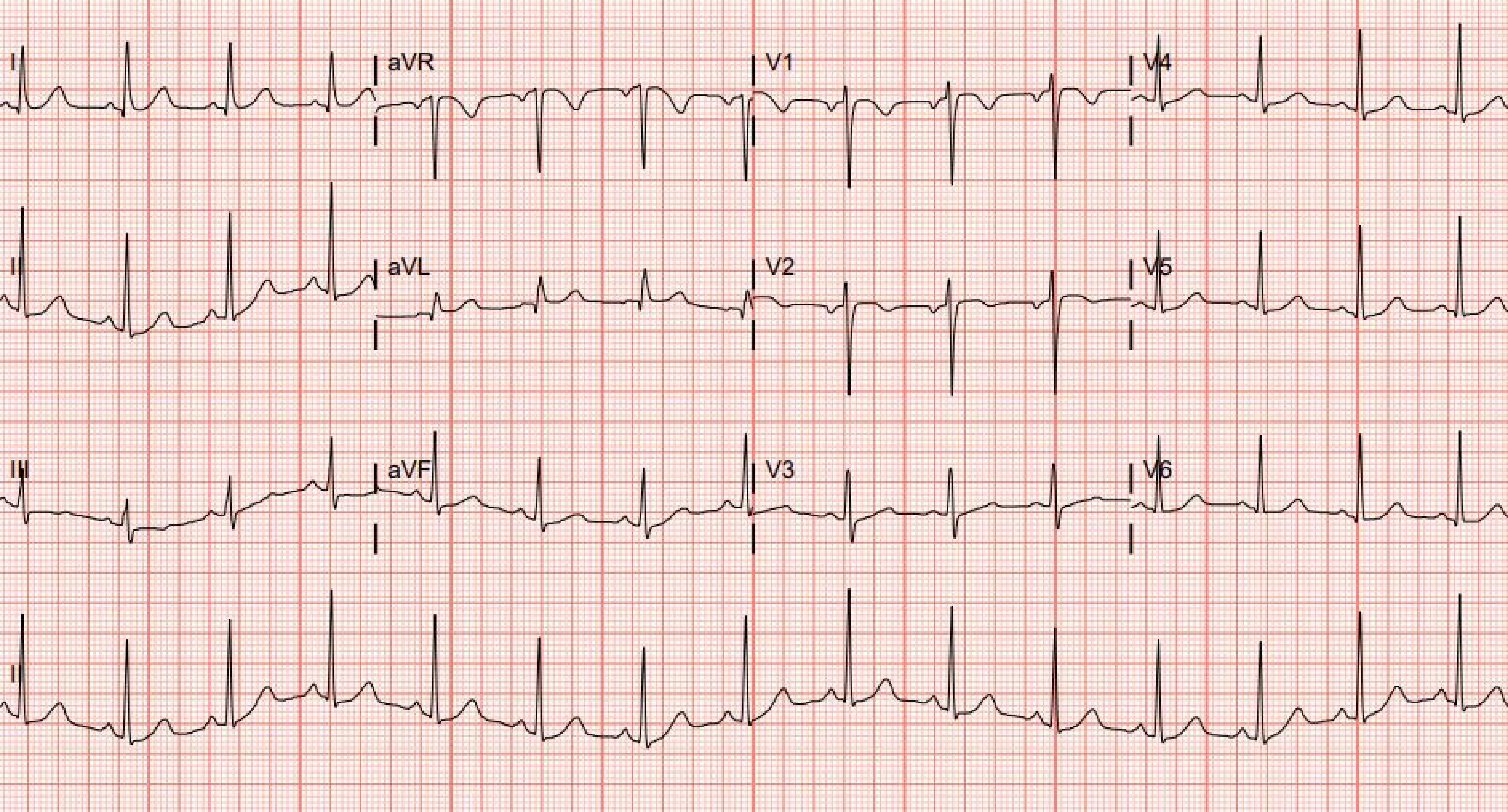
I
aVR
V1
V4
II
aVL
V2
V5
III
aVF
V3
V6
II

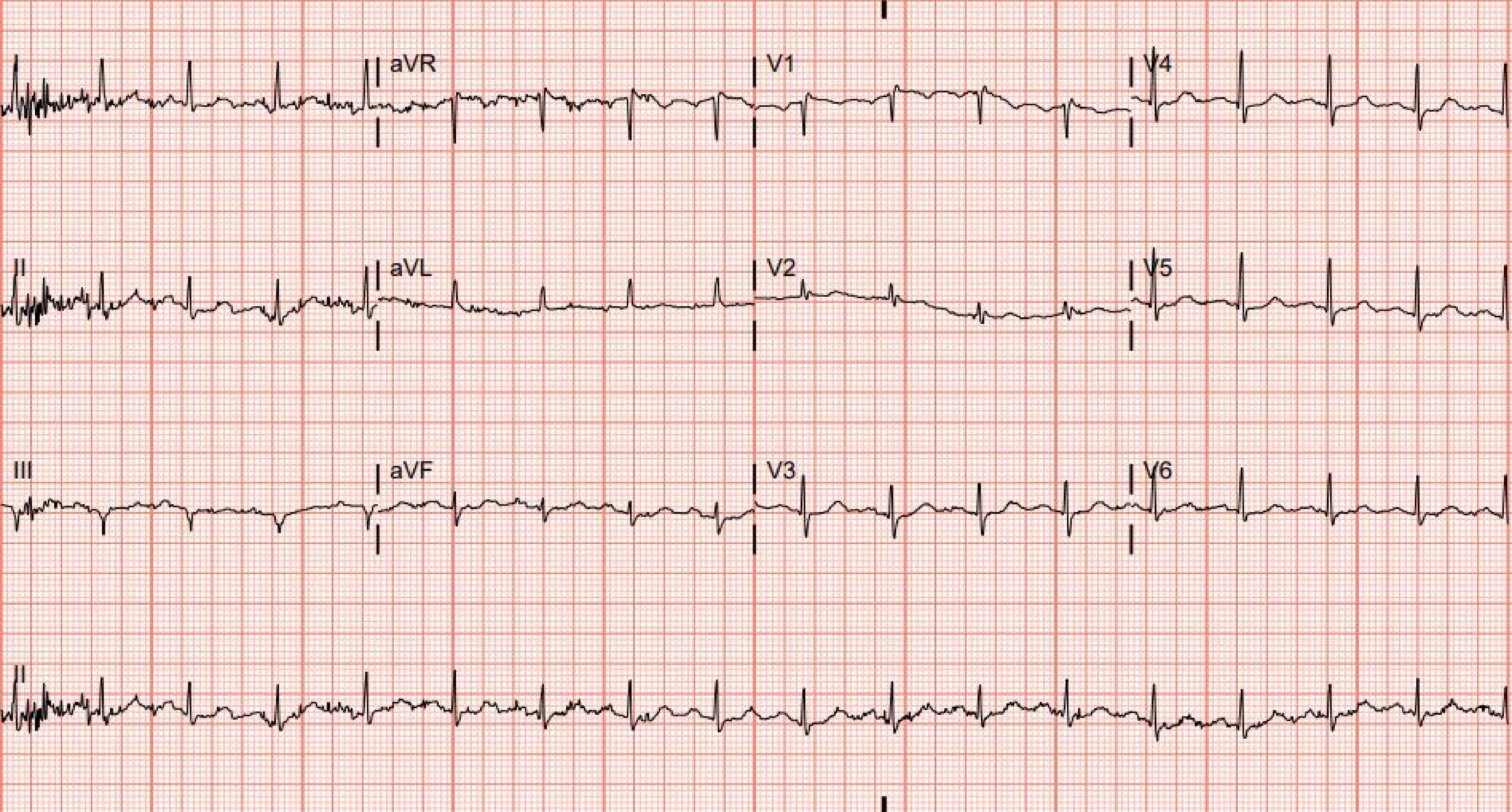

I
aVR
V1
V4
II
aVL
V2
V5
III
aVF
V3
V6
II

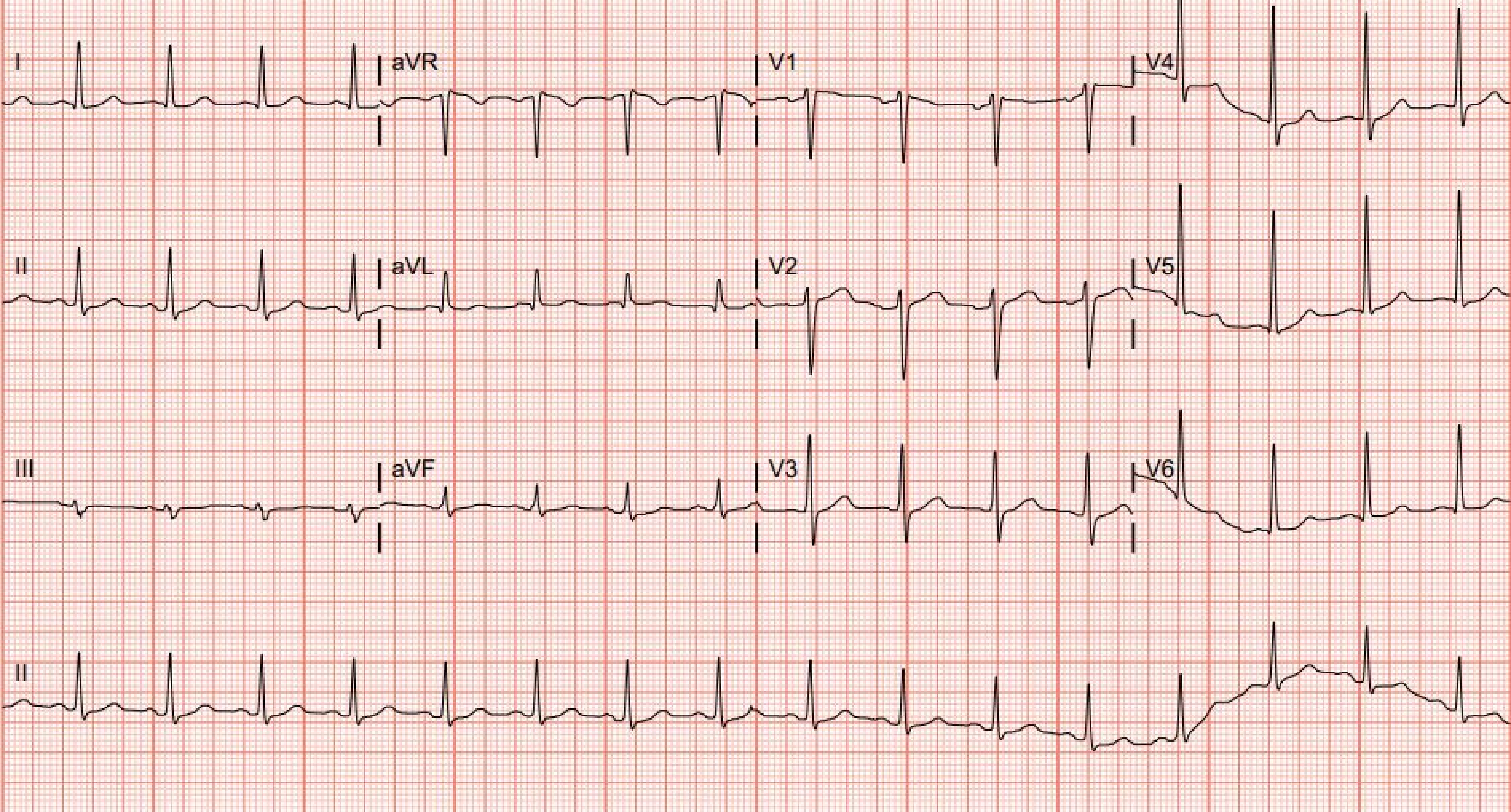
I
aVR
V1
V4
II
aVL
V2
V5
III
aVF
V3
V6
II

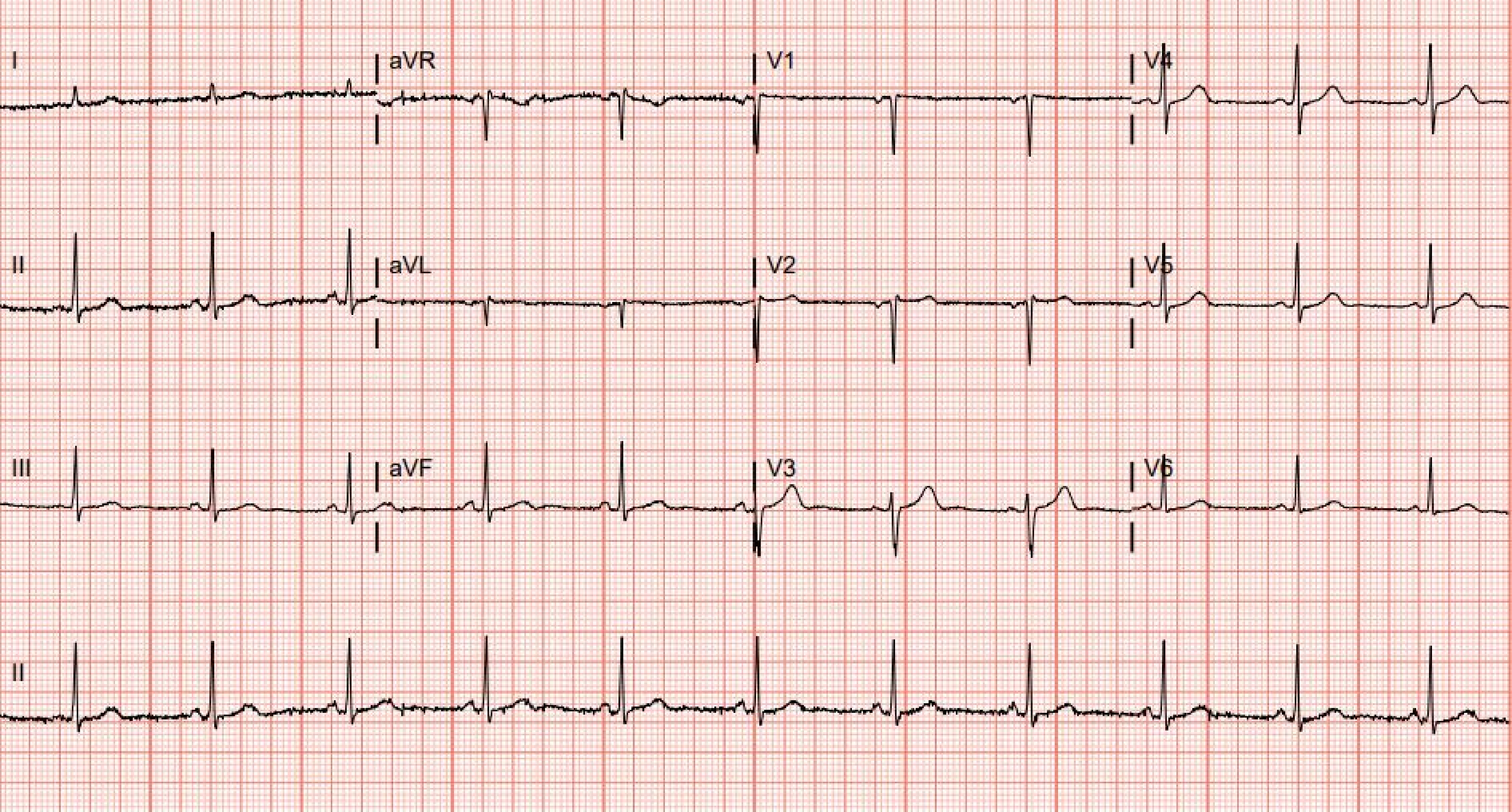
I
aVR
V1
V4
II
aVL
V2
V5
III
aVF
V3
V6
II

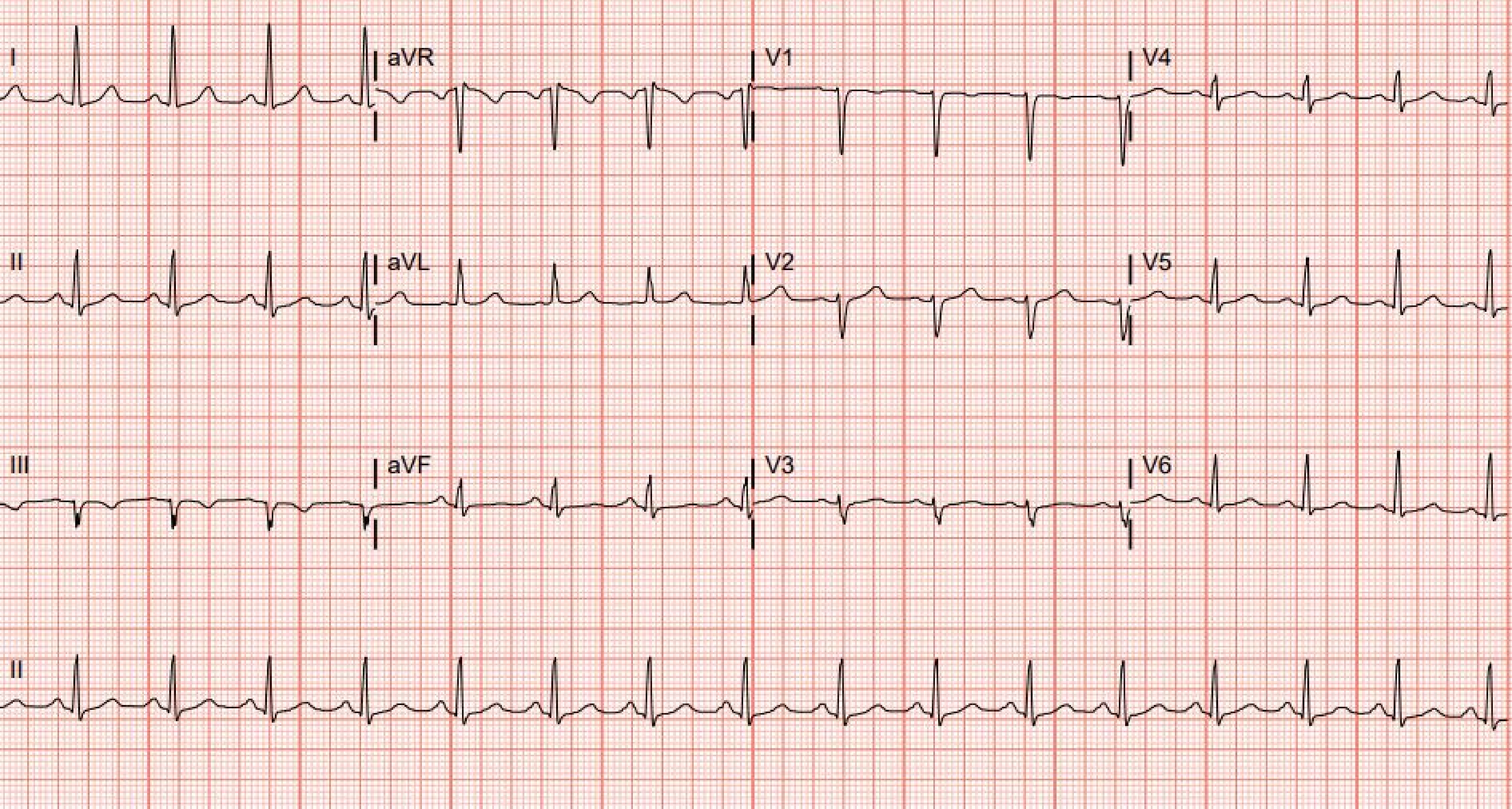
I
aVR
V1
V4
II
aVL
V2
V5
III
aVF
V3
V6
II

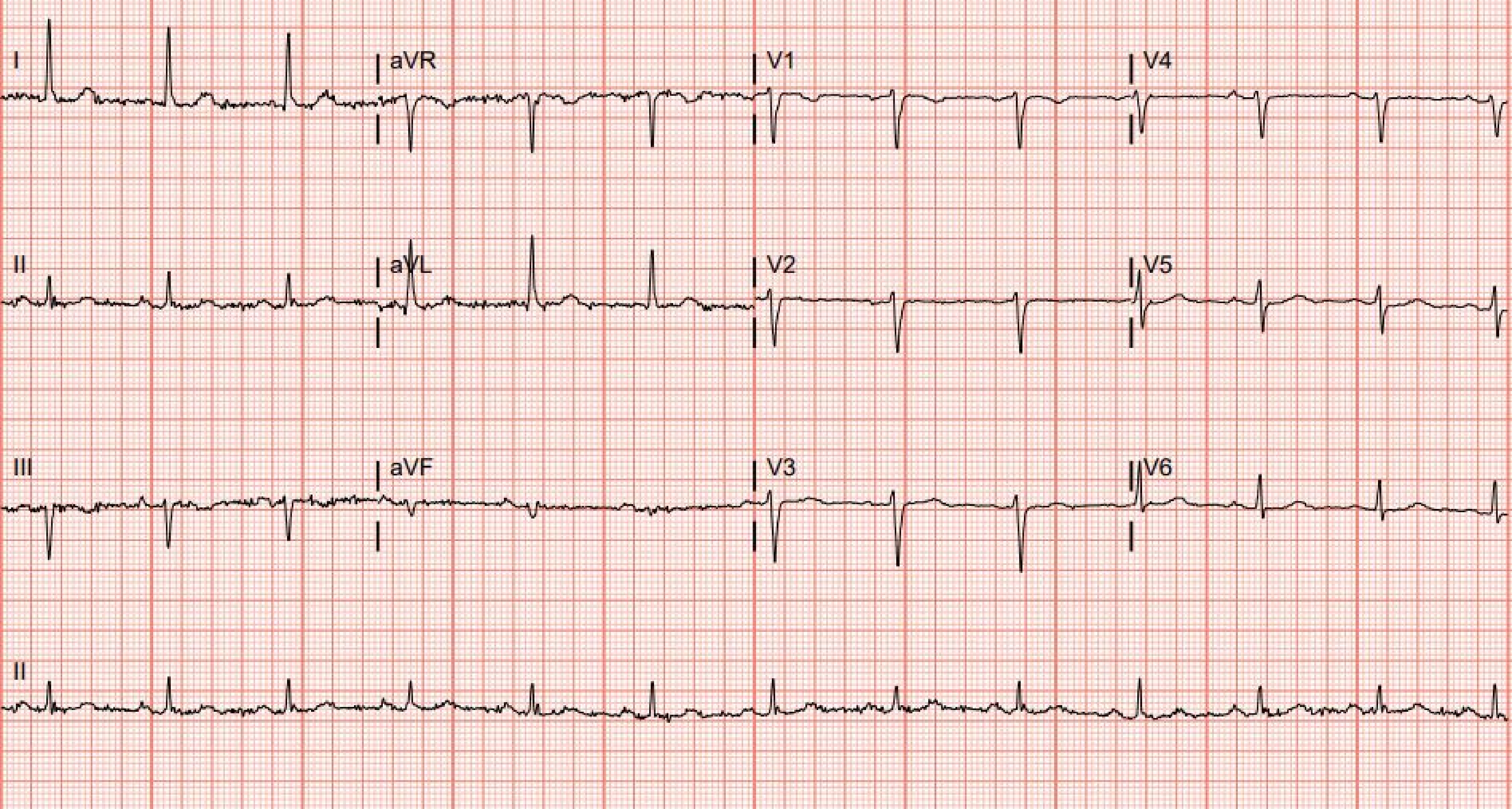
I
aVR
V1
V4
II
aVL
V2
V5
III
aVF
V3
V6
II

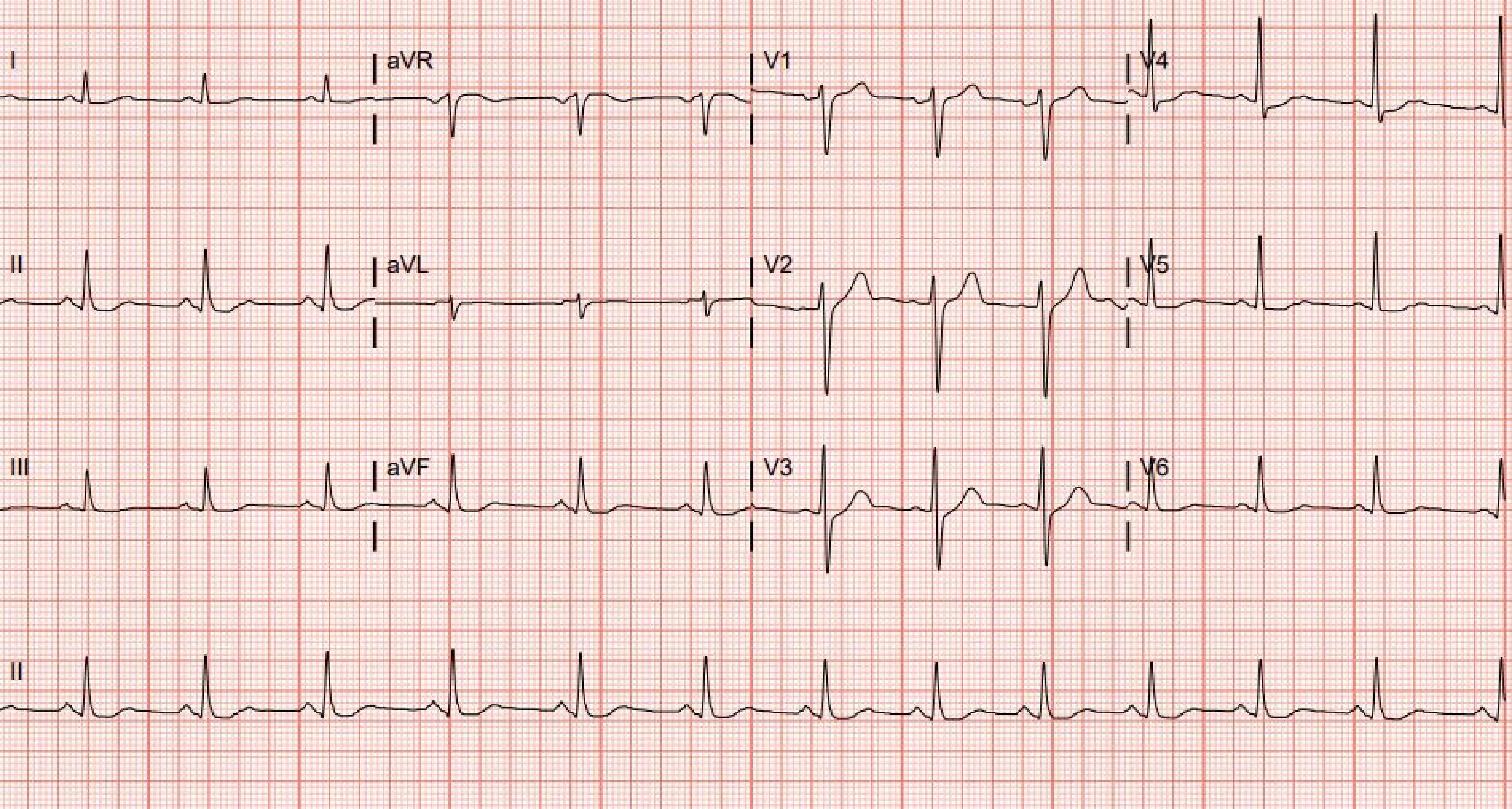
I
aVR
V1
V4
II
aVL
V2
V5
III
aVF
V3
V6
II

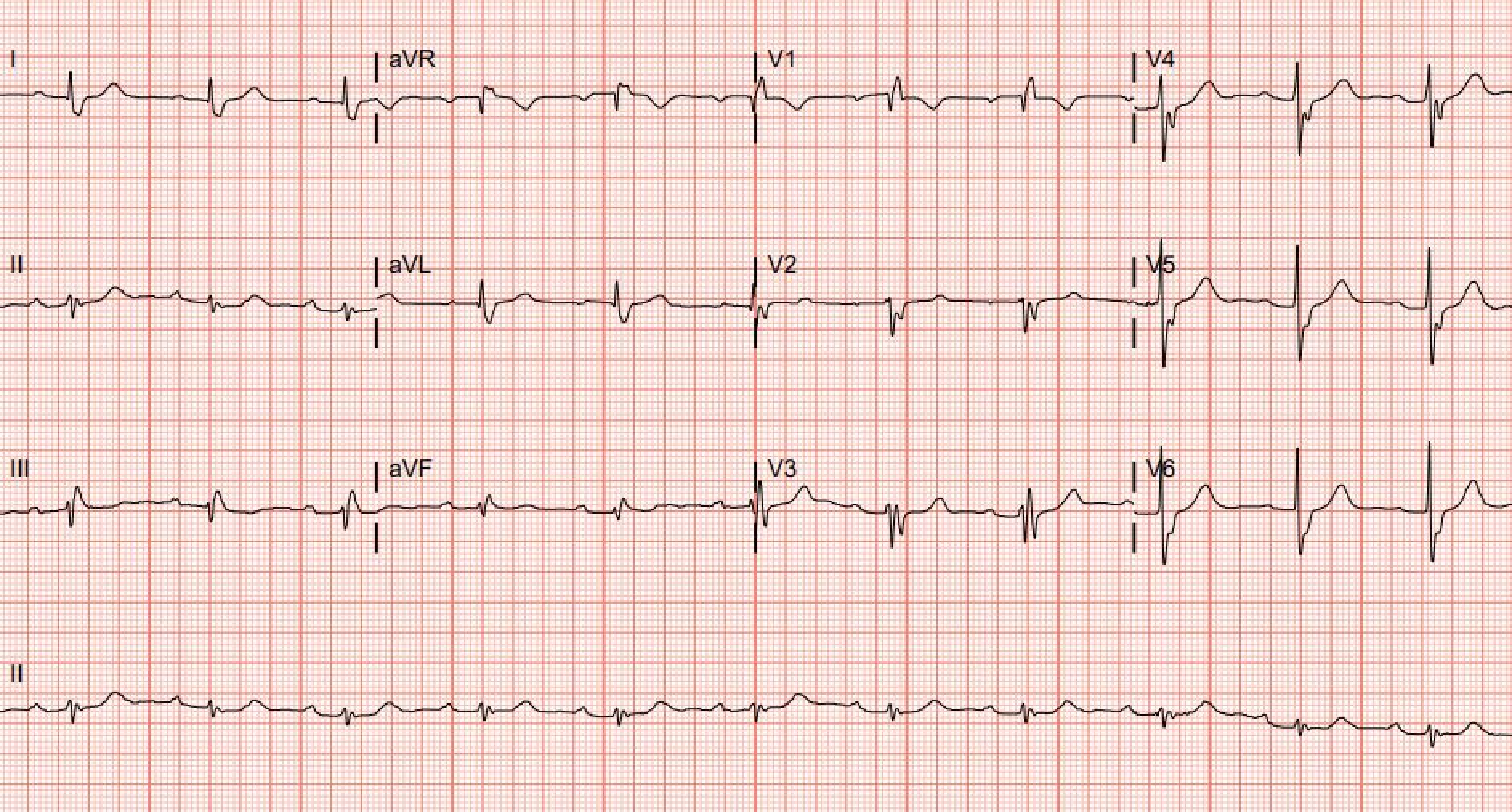
I
aVR
V1
V4
II
aVL
V2
V5
III
aVF
V3
V6
II

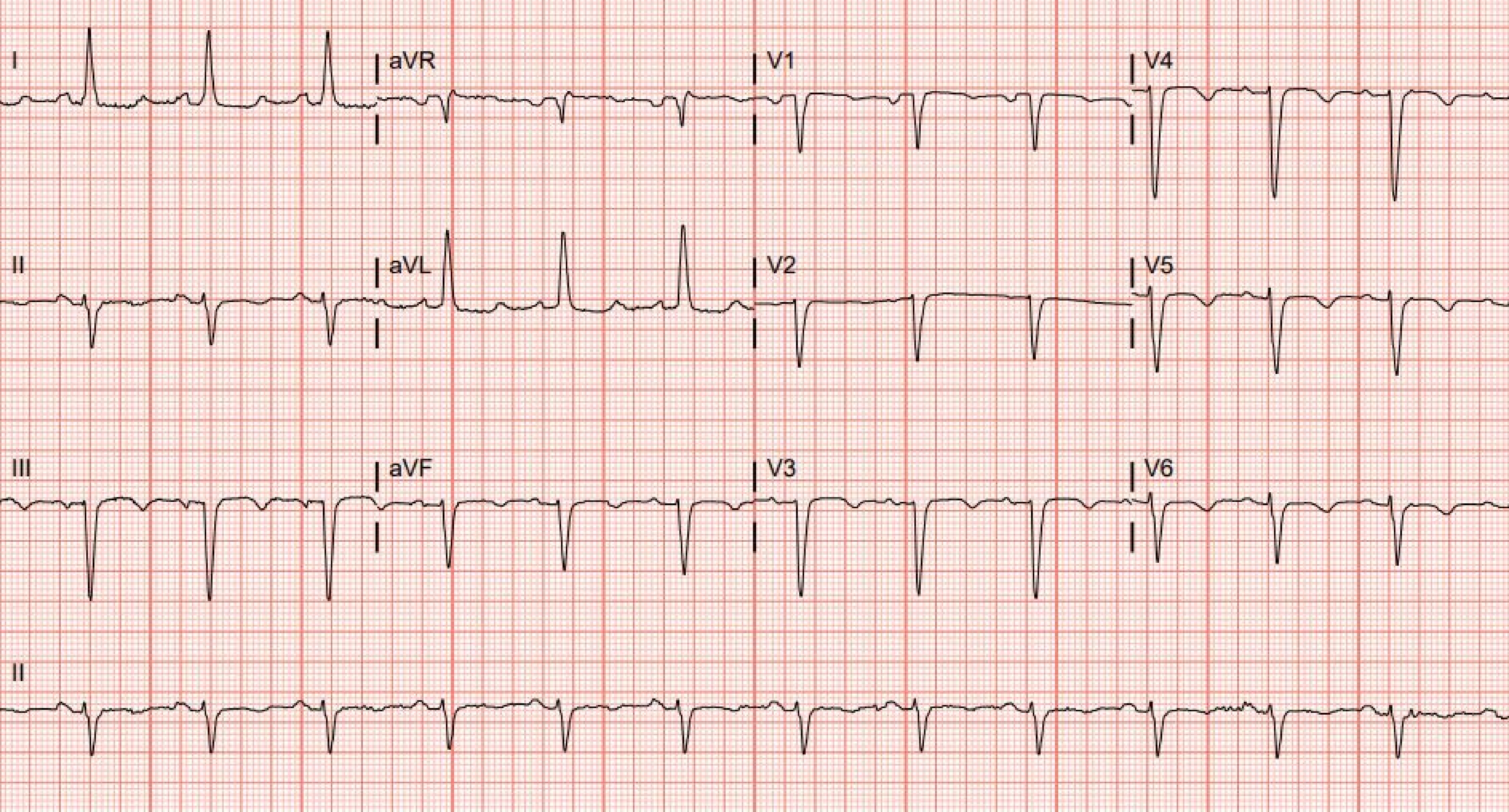
I
aVR
V1
V4
II
aVL
V2
V5
III
aVF
V3
V6
II

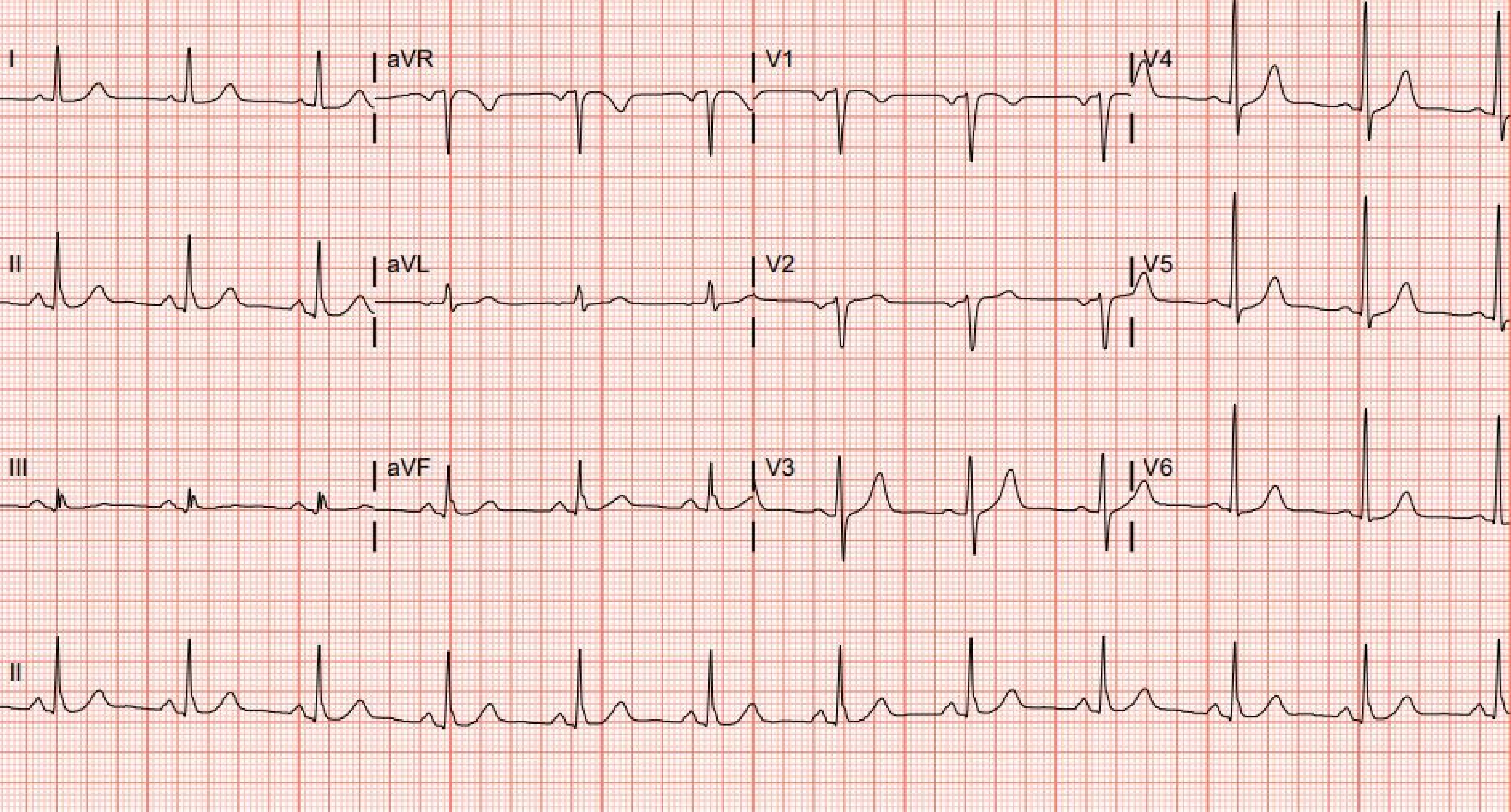
I
aVR
V1
V4
II
aVL
V2
V5
III
aVF
V3
V6
II

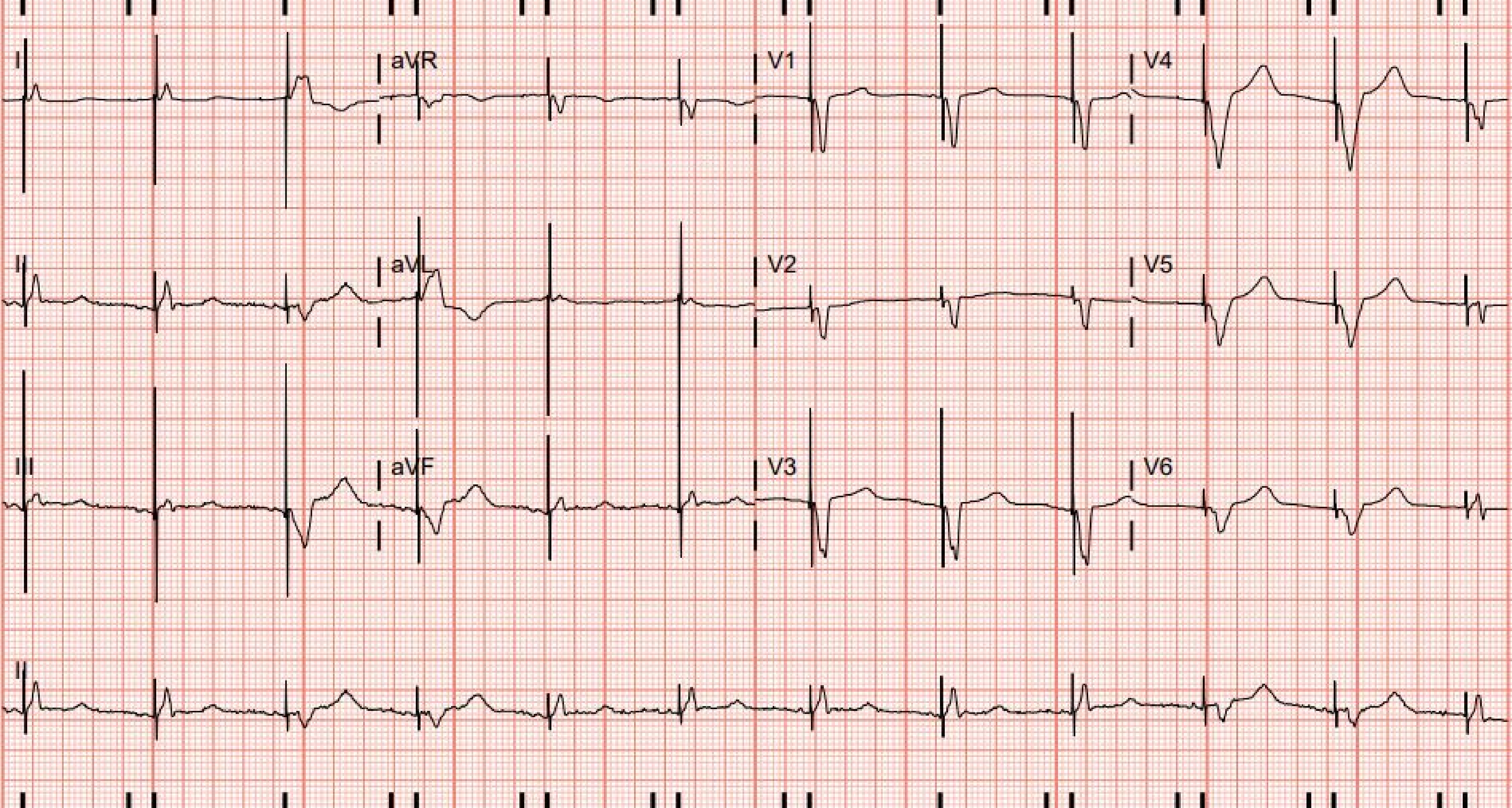
I
aVR
V1
V4
II
aVL
V2
V5
III
aVF
V3
V6
II

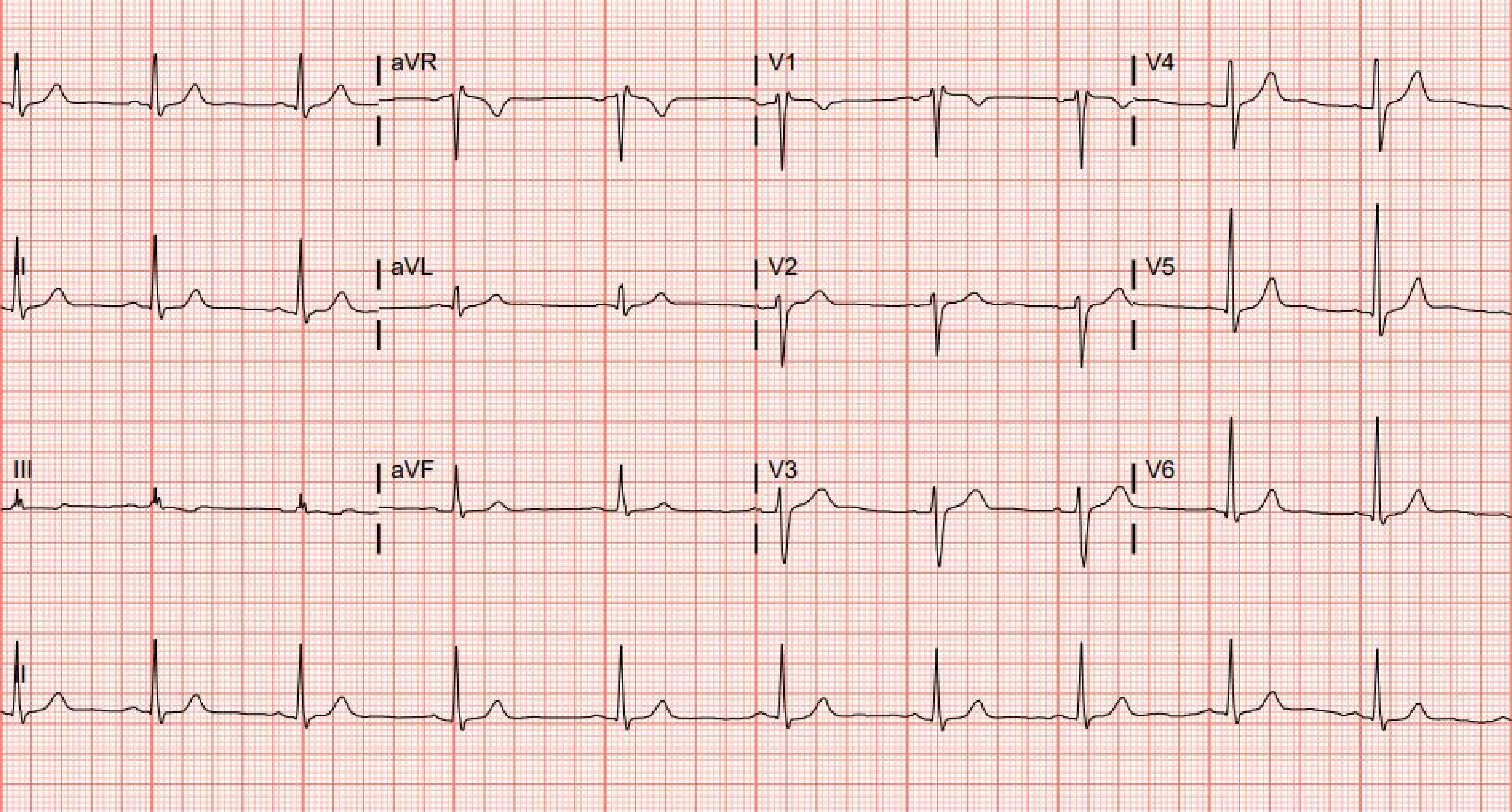
I
aVR
V1
V4
II
aVL
V2
V5
III
aVF
V3
V6
II

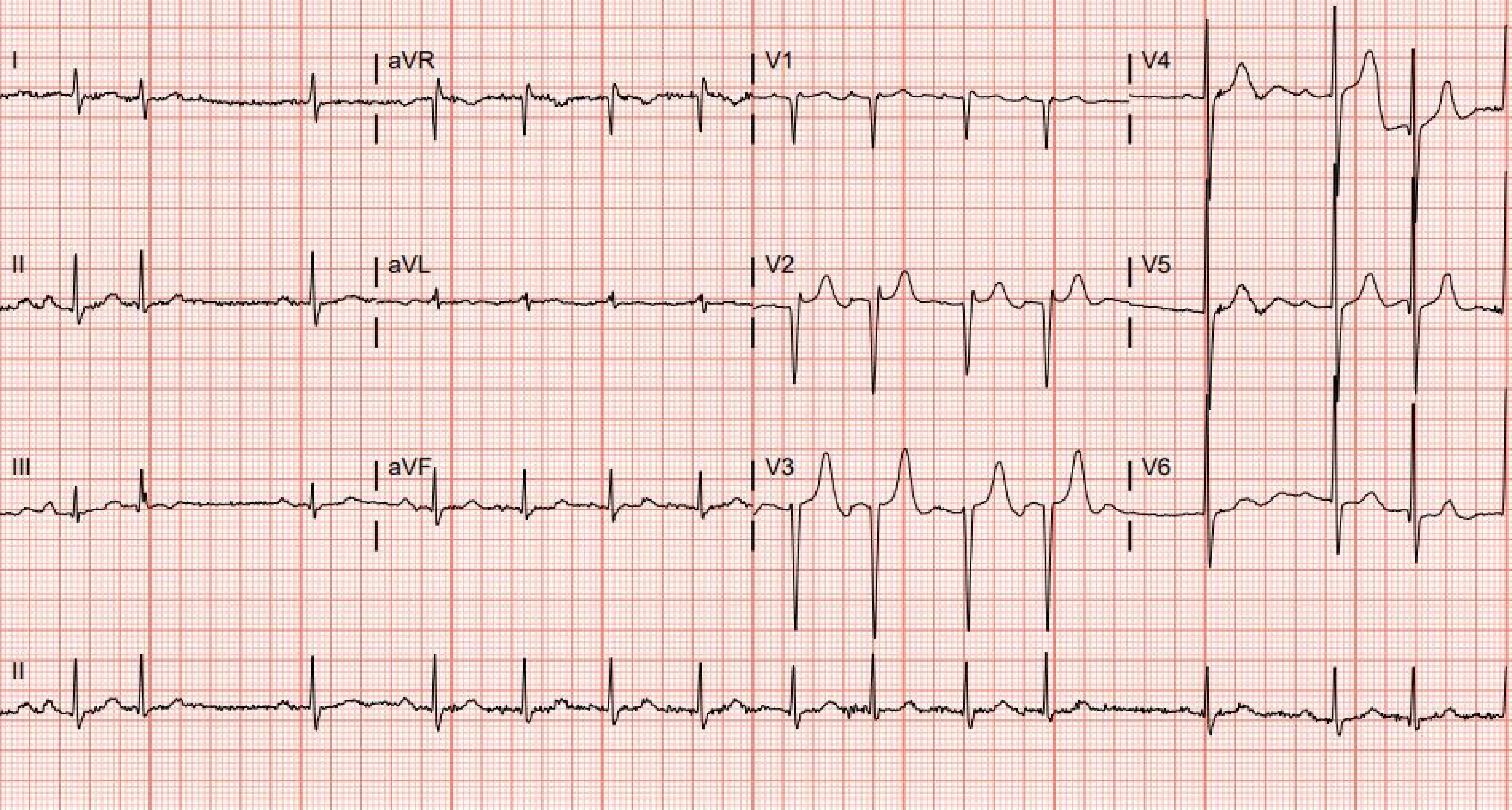
I
aVR
V1
V4
II
aVL
V2
V5
III
aVF
V3
V6
II

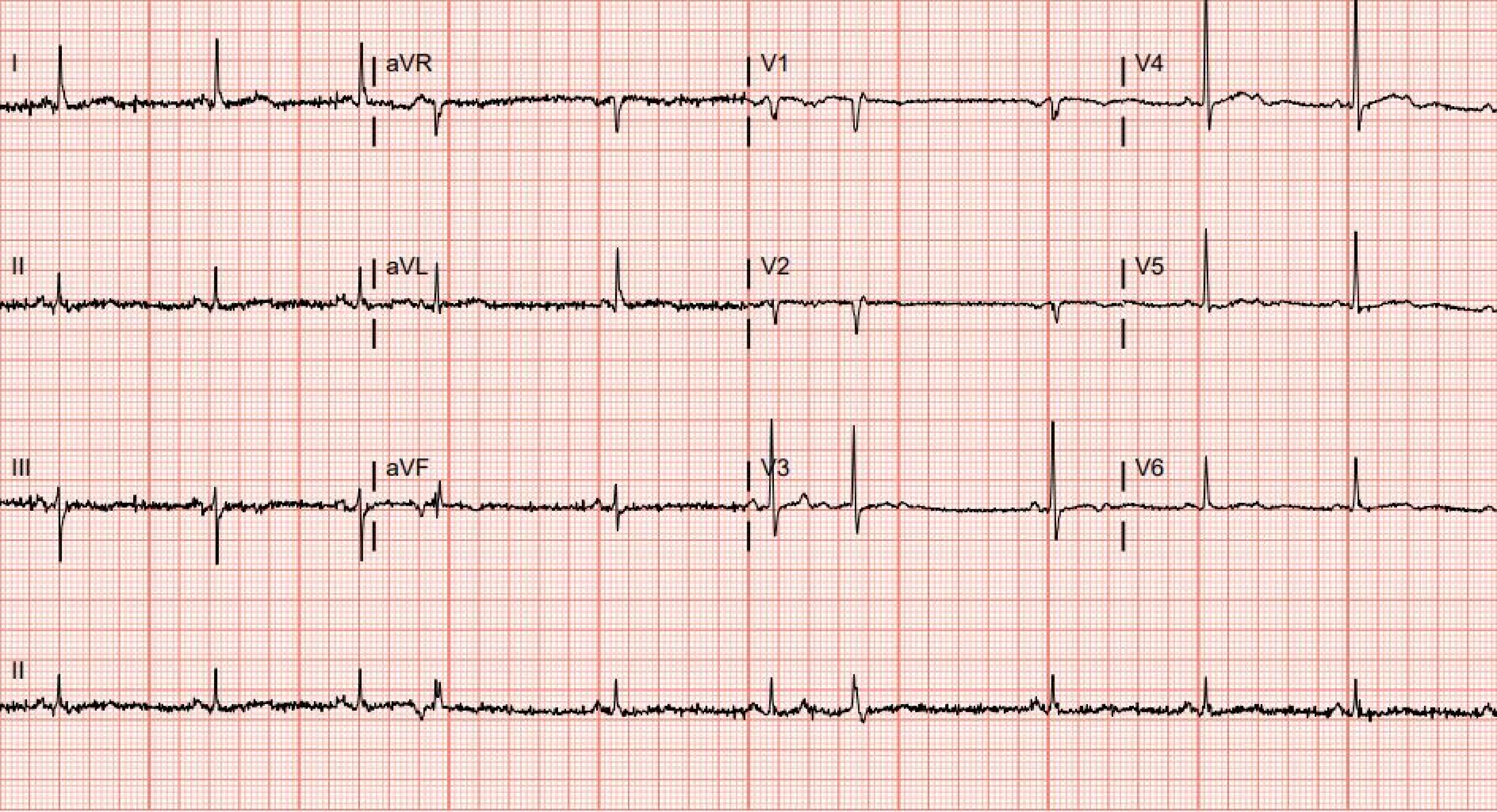
I
aVR
V1
V4
II
aVL
V2
V5
III
aVF
V3
V6
II

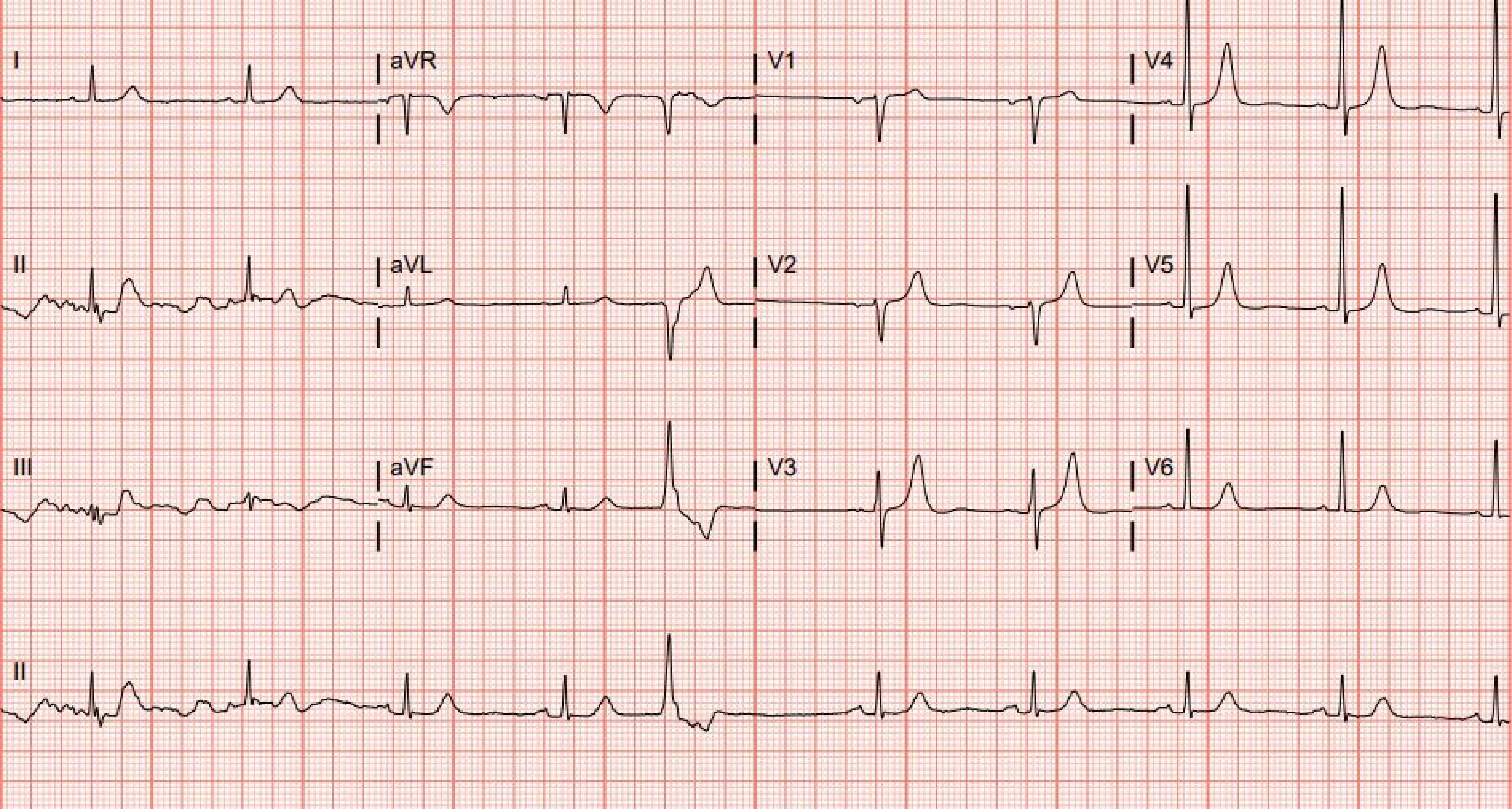
I
aVR
V1
V4
II
aVL
V2
V5
III
aVF
V3
V6
II

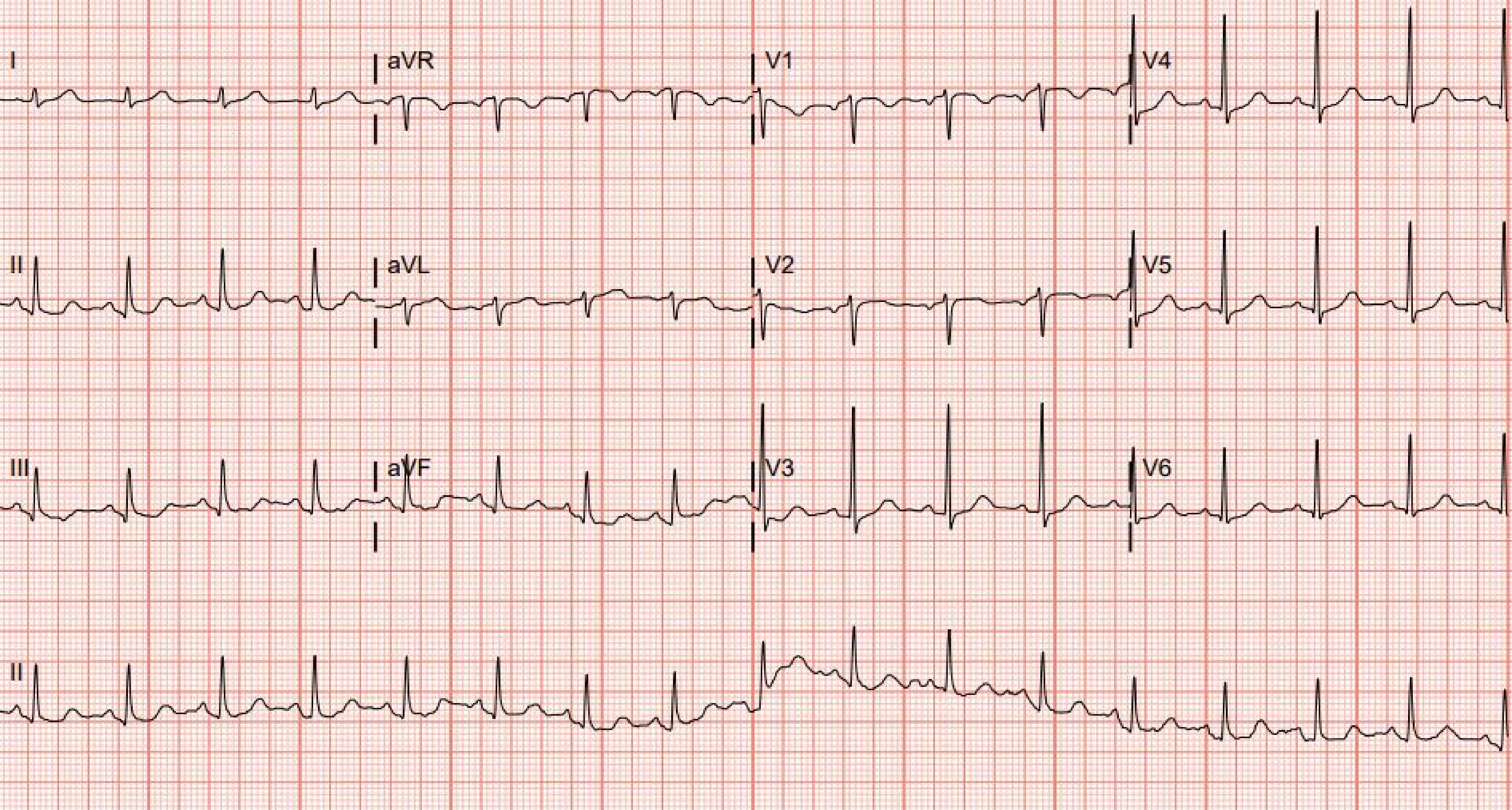
I
aVR
V1
V4
II
aVL
V2
V5
III
aVF
V3
V6
II

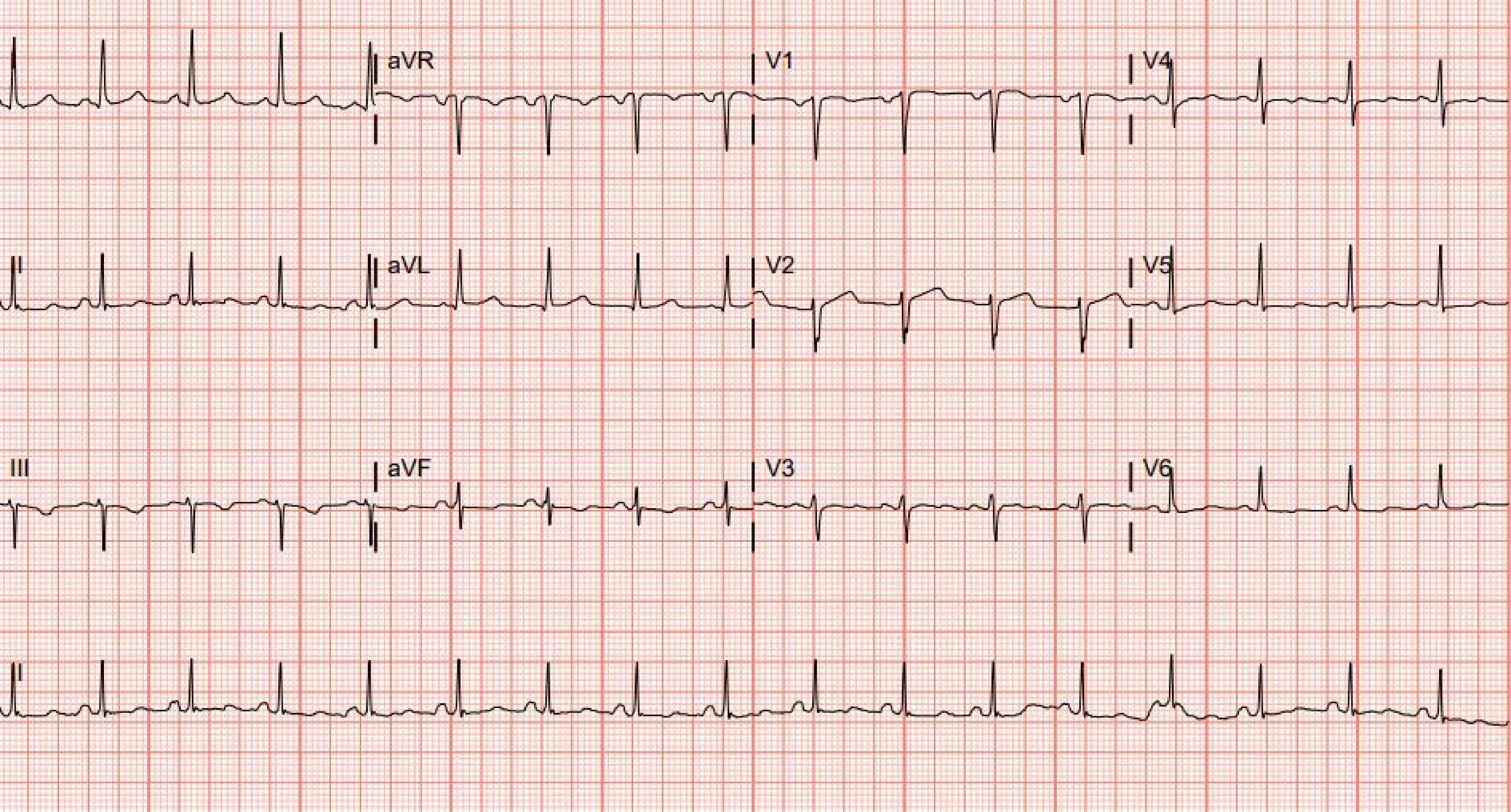

aVR
V1
V4
II
aVL
V2
V5
III
aVF
V3
V6
II

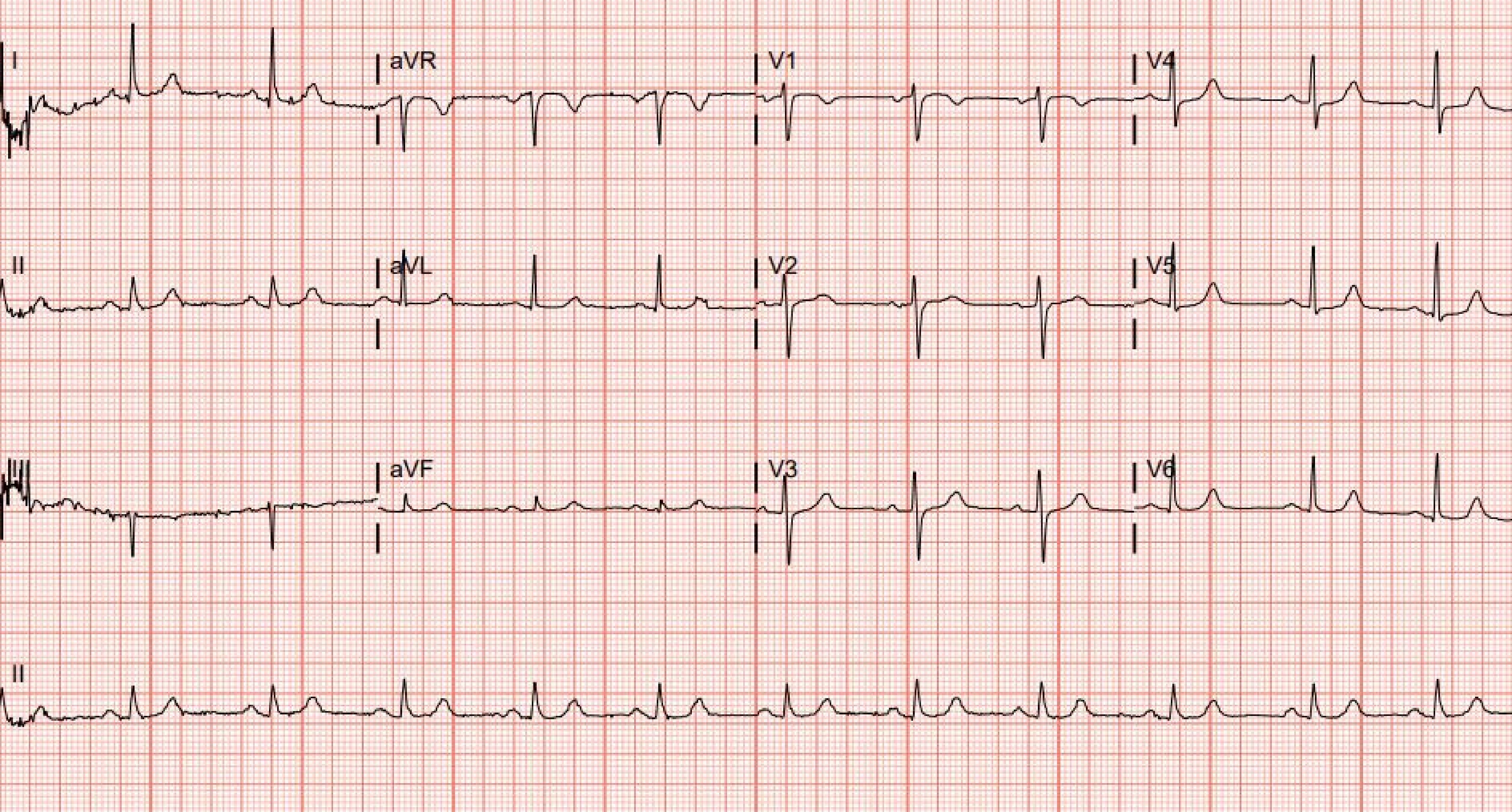
I
aVR
V1
V4
II
aVL
V2
V5
III
aVF
V3
V6
II

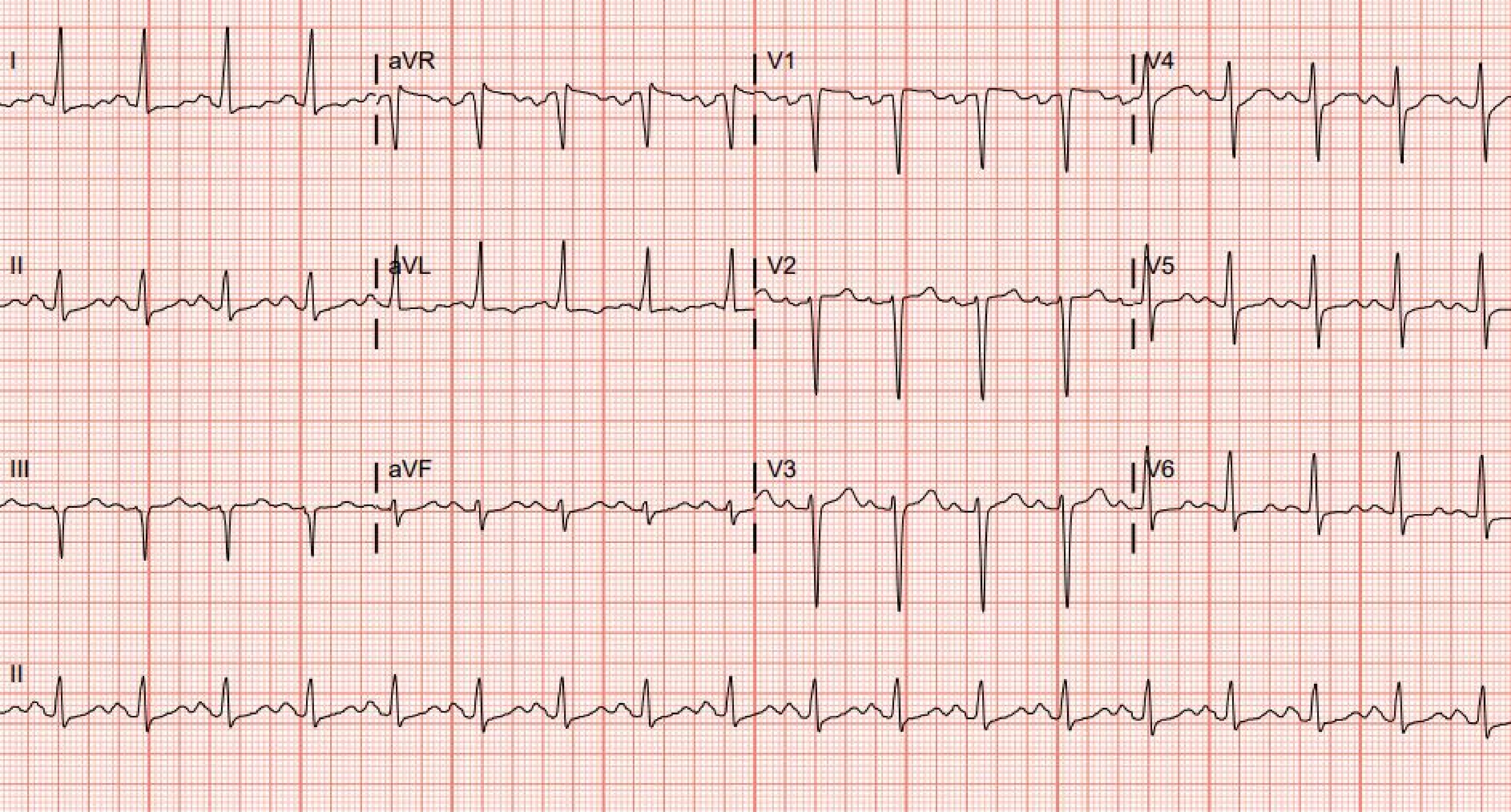
I
aVR
V1
V4
II
aVL
V2
V5
III
aVF
V3
V6
II

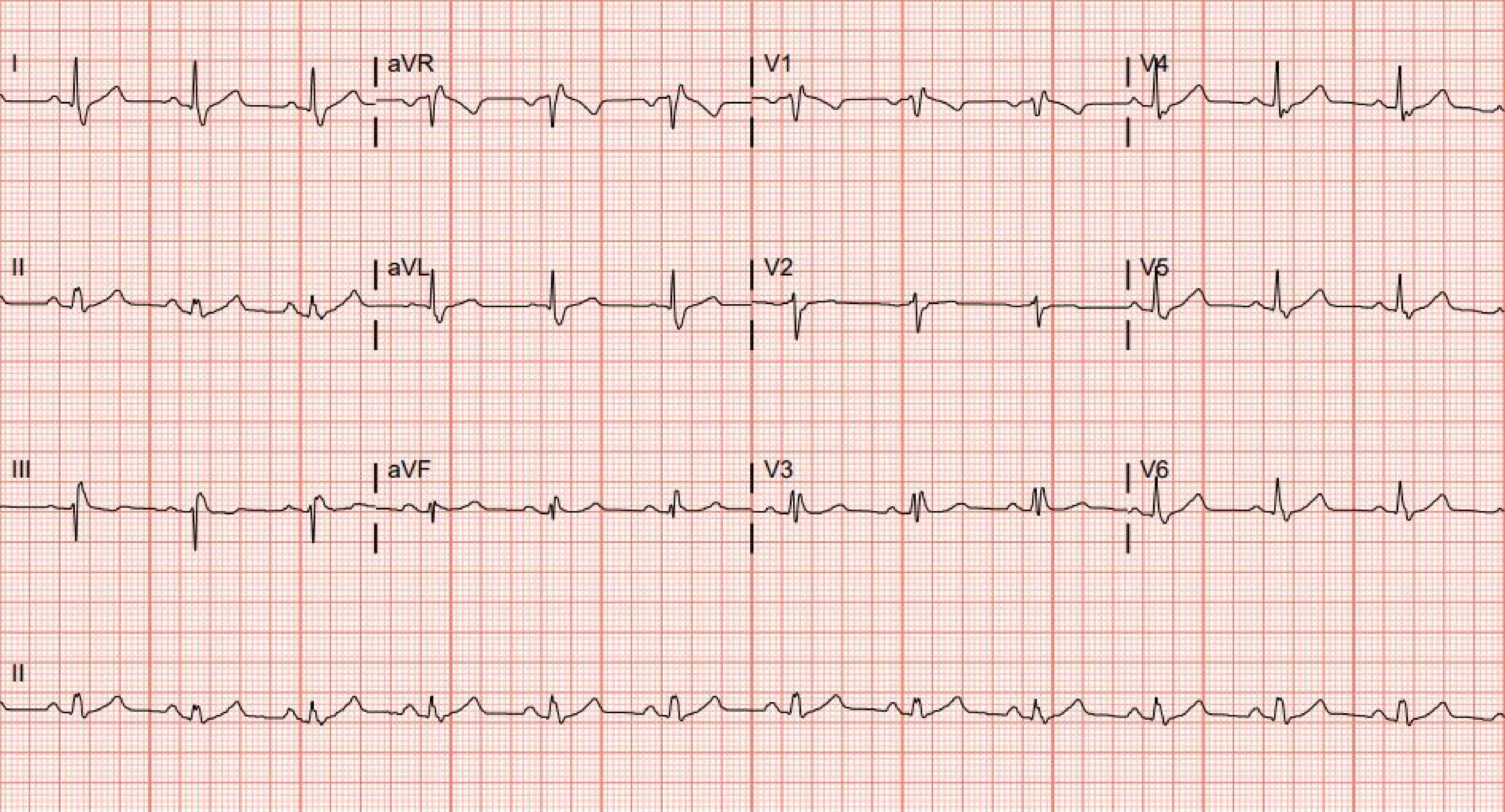
I
aVR
V1
V4
II
aVL
V2
V5
III
aVF
V3
V6
II

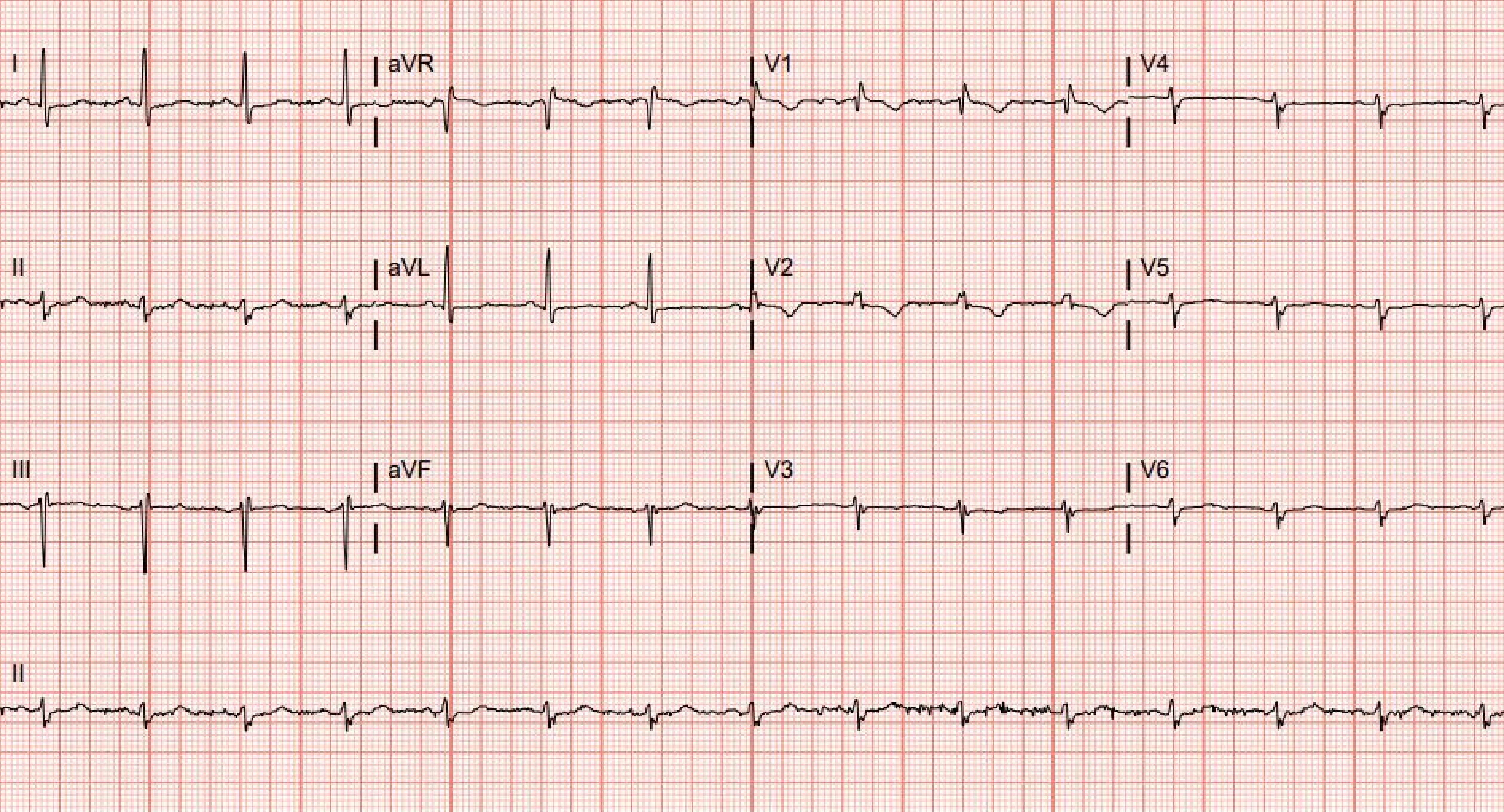
I
aVR
V1
V4
II
aVL
V2
V5
III
aVF
V3
V6
II

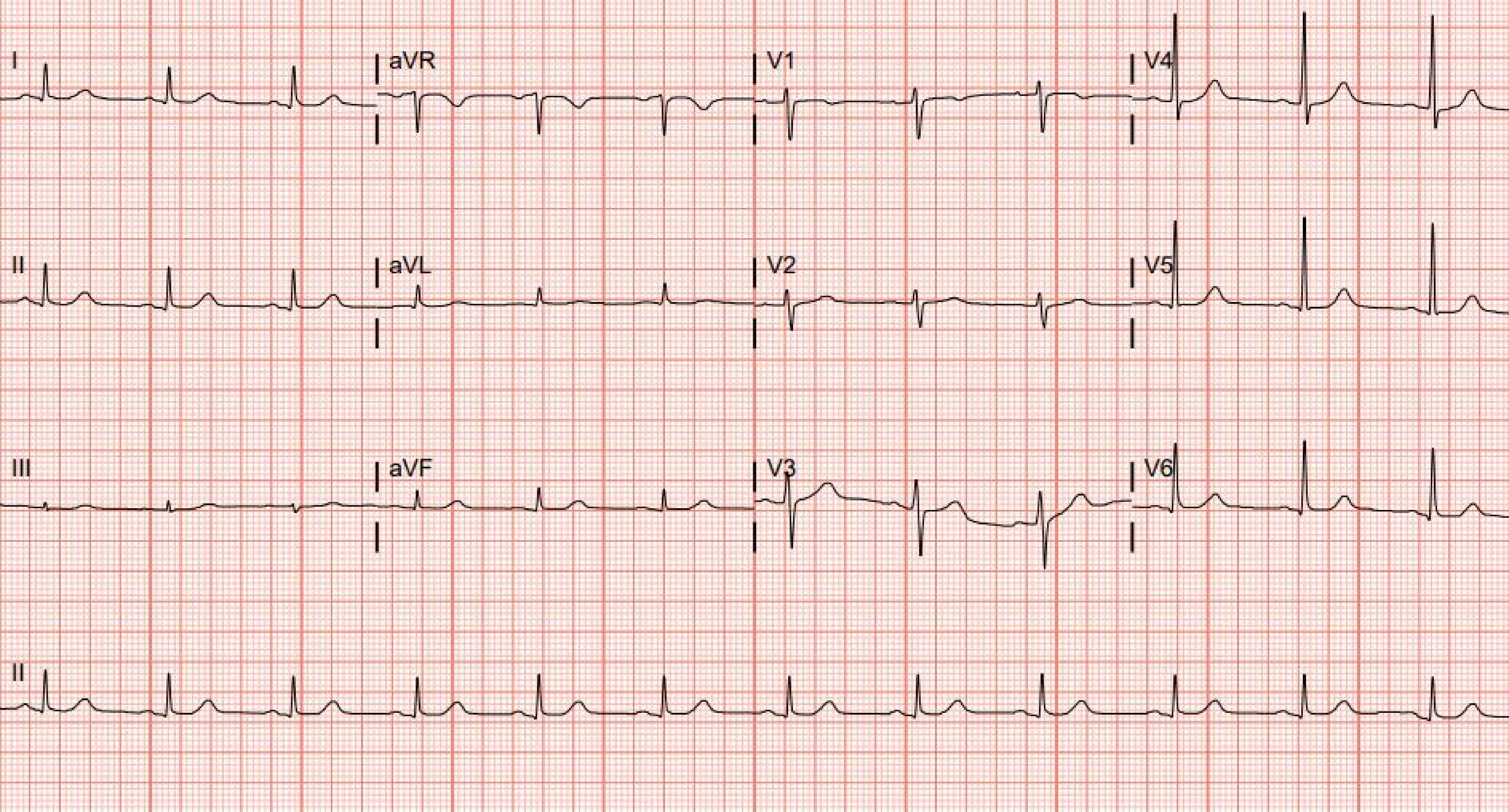
I
aVR
V1
V4
II
aVL
V2
V5
III
aVF
V3
V6
II

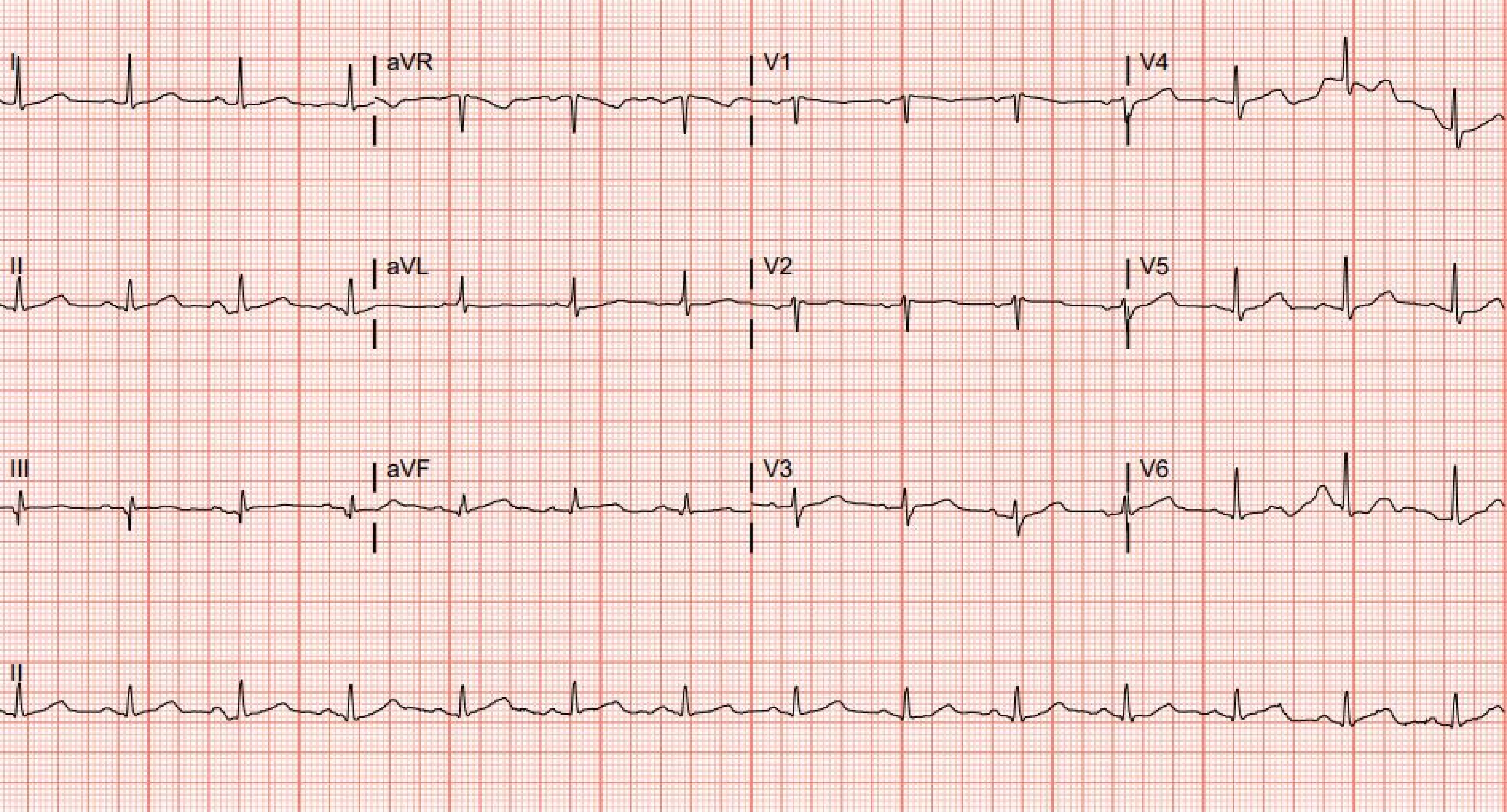
I
aVR
V1
V4
II
aVL
V2
V5
III
aVF
V3
V6
II

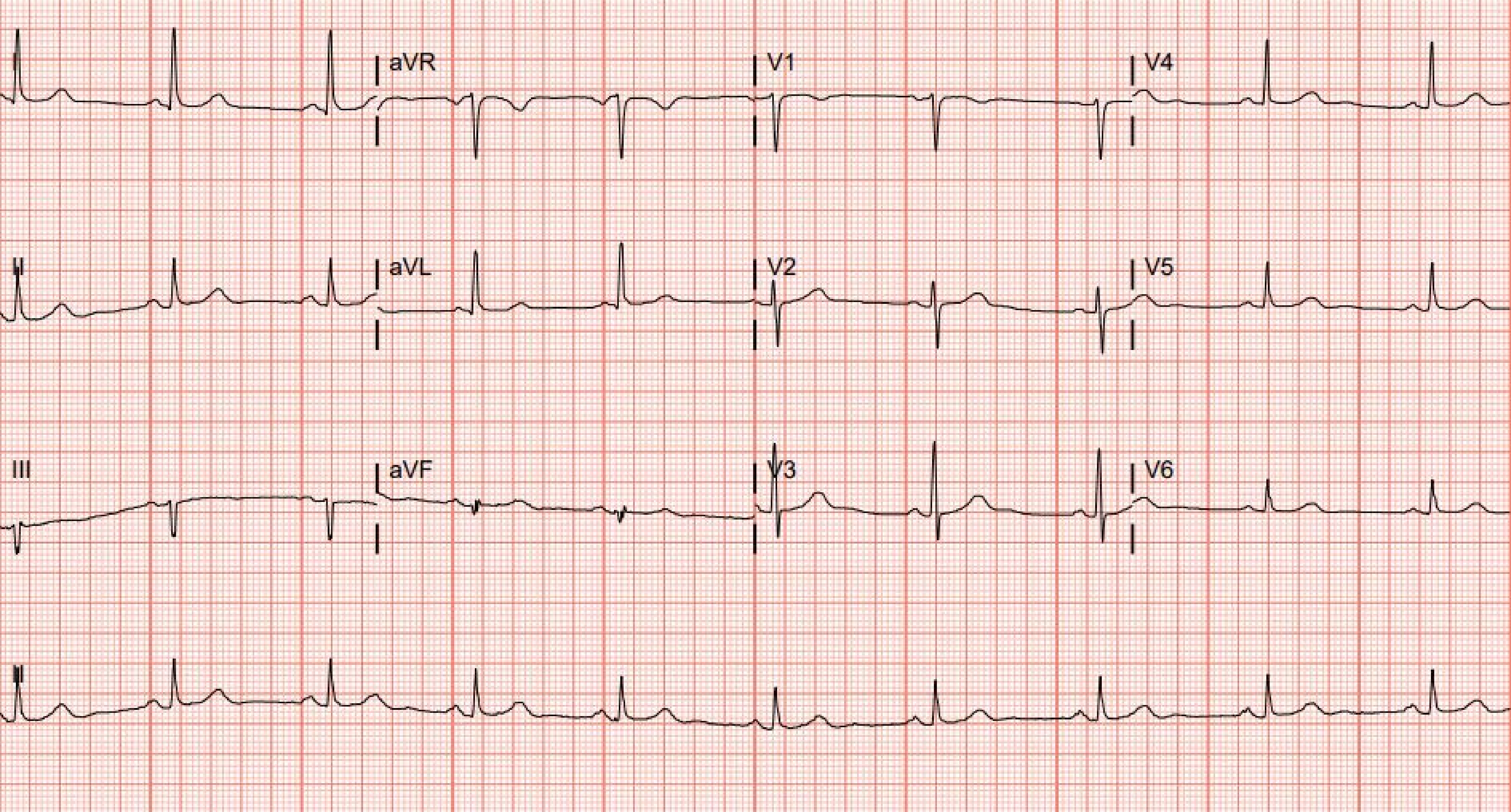
aVR
V1
V4
II
aVL
V2
V5
III
aVF
V3
V6

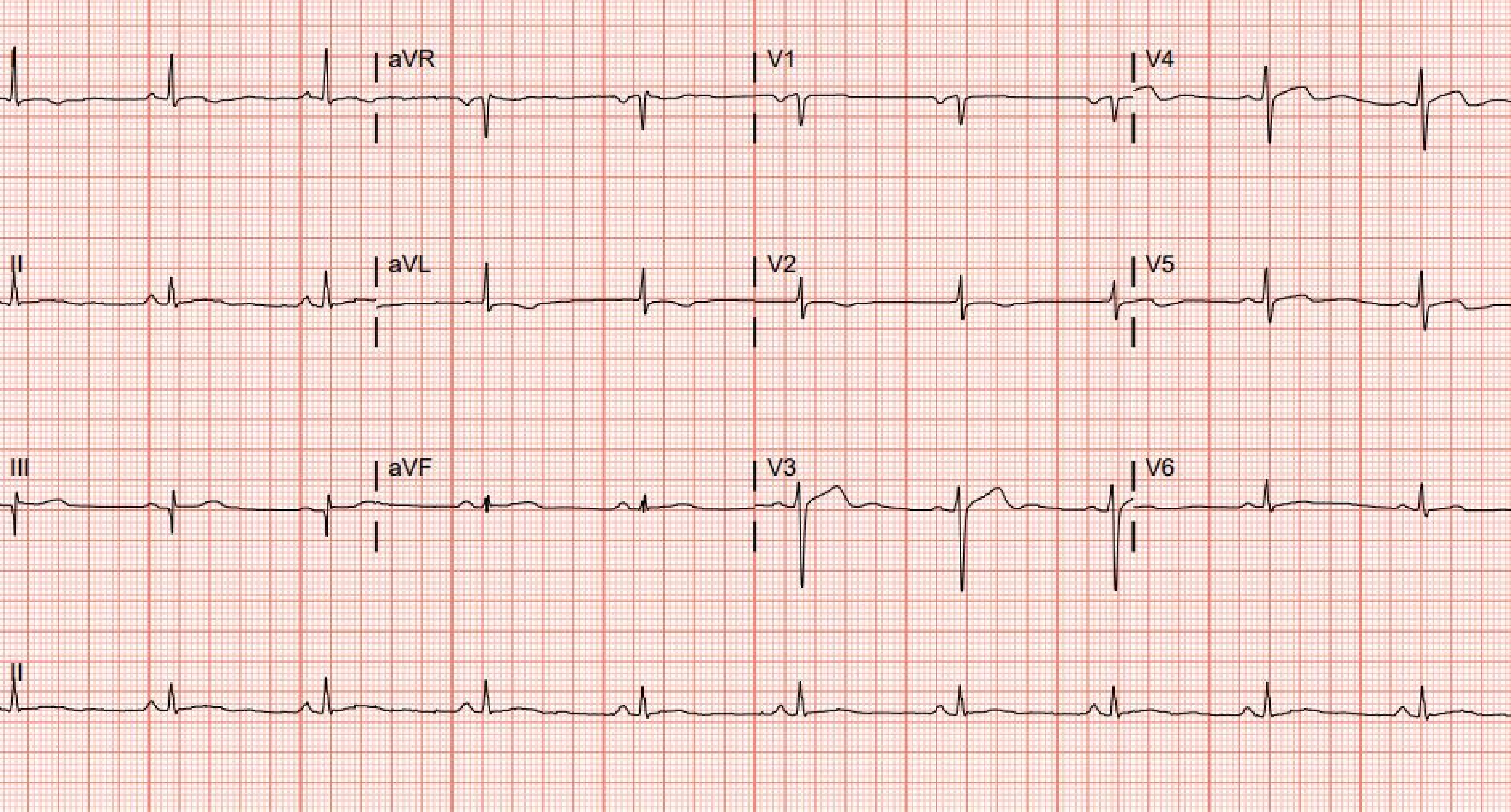
I
aVR
V1
V4
II
aVL
V2
V5
III
aVF
V3
V6
II

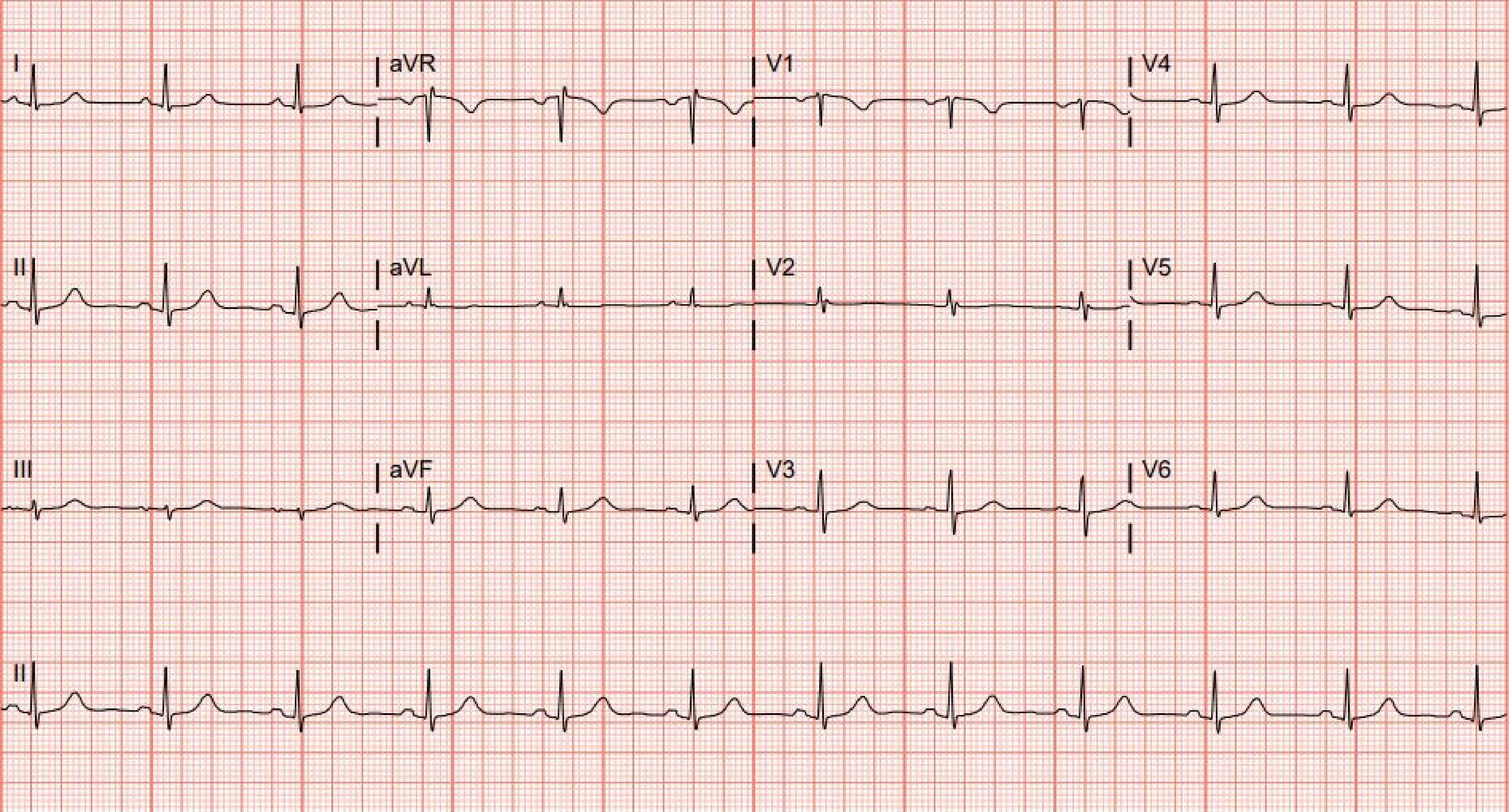
I
aVR
V1
V4
II
aVL
V2
V5
III
aVF
V3
V6
II

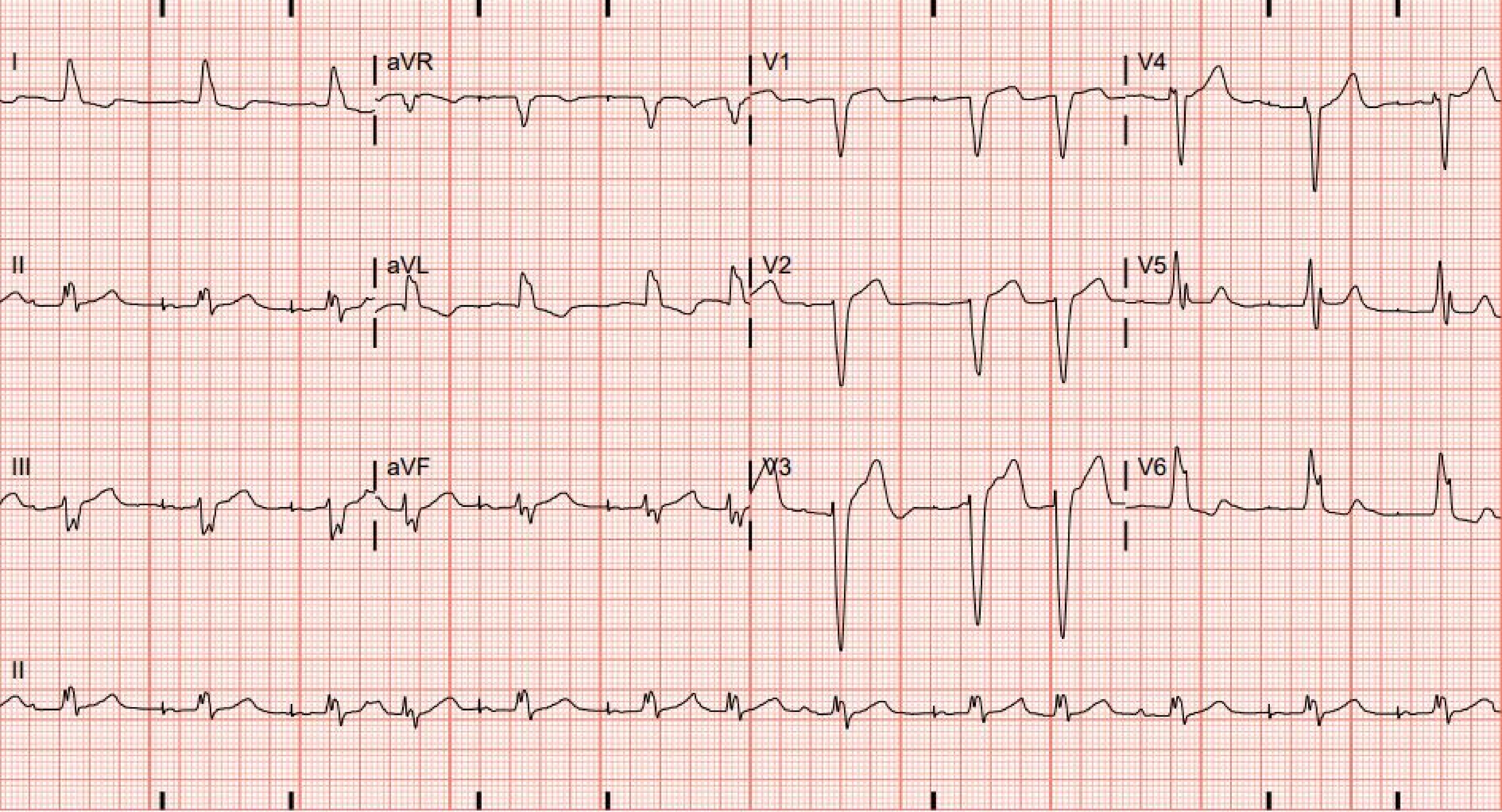

I
aVR
V1
V4
II
aVL
V2
V5
III
aVF
V3
V6
II

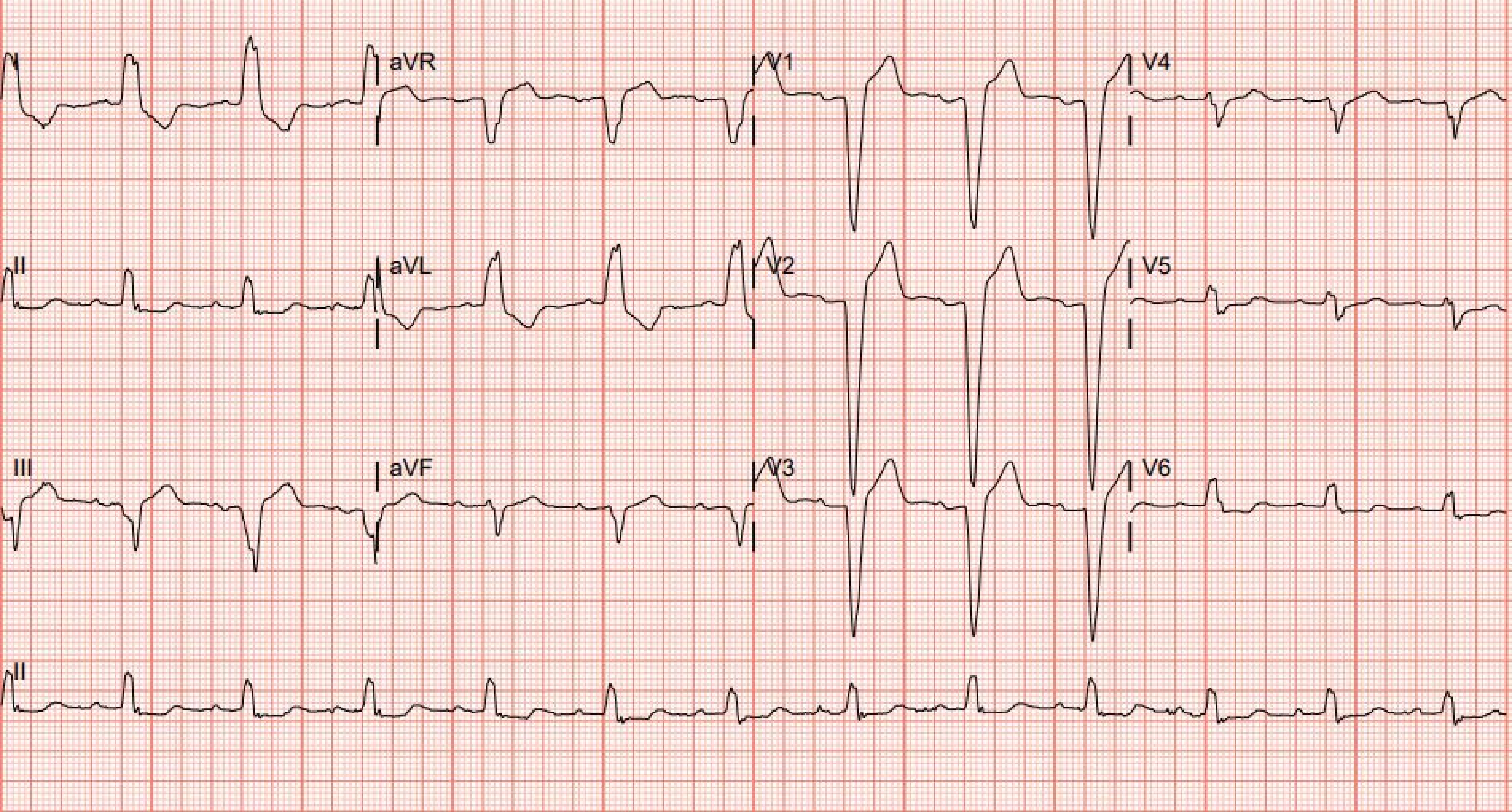

I
aVR
V1
V4
II
aVL
V2
V5
III
aVF
V3
V6
II

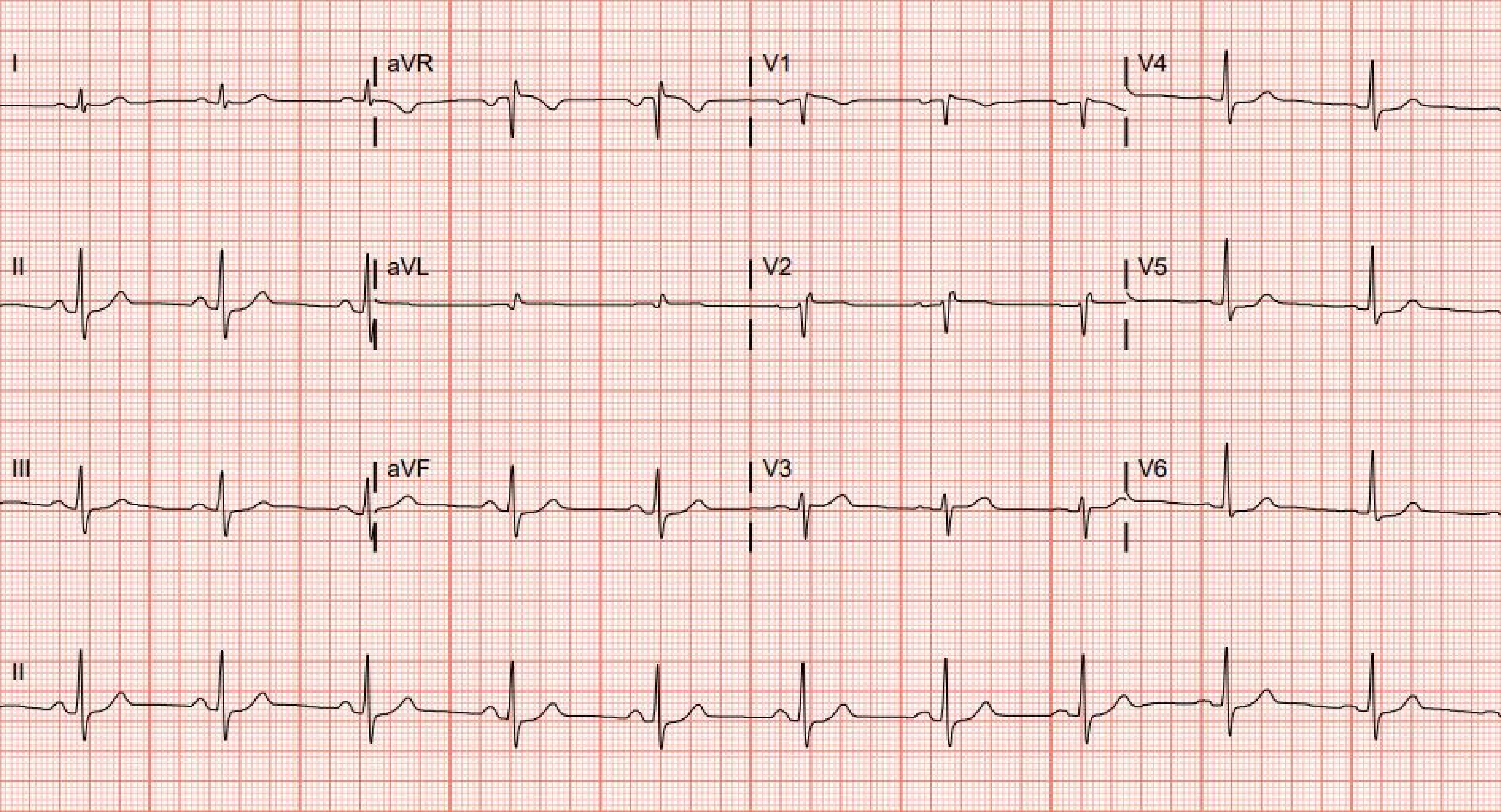
I
aVR
V1
V4
II
aVL
V2
V5
III
aVF
V3
V6
II

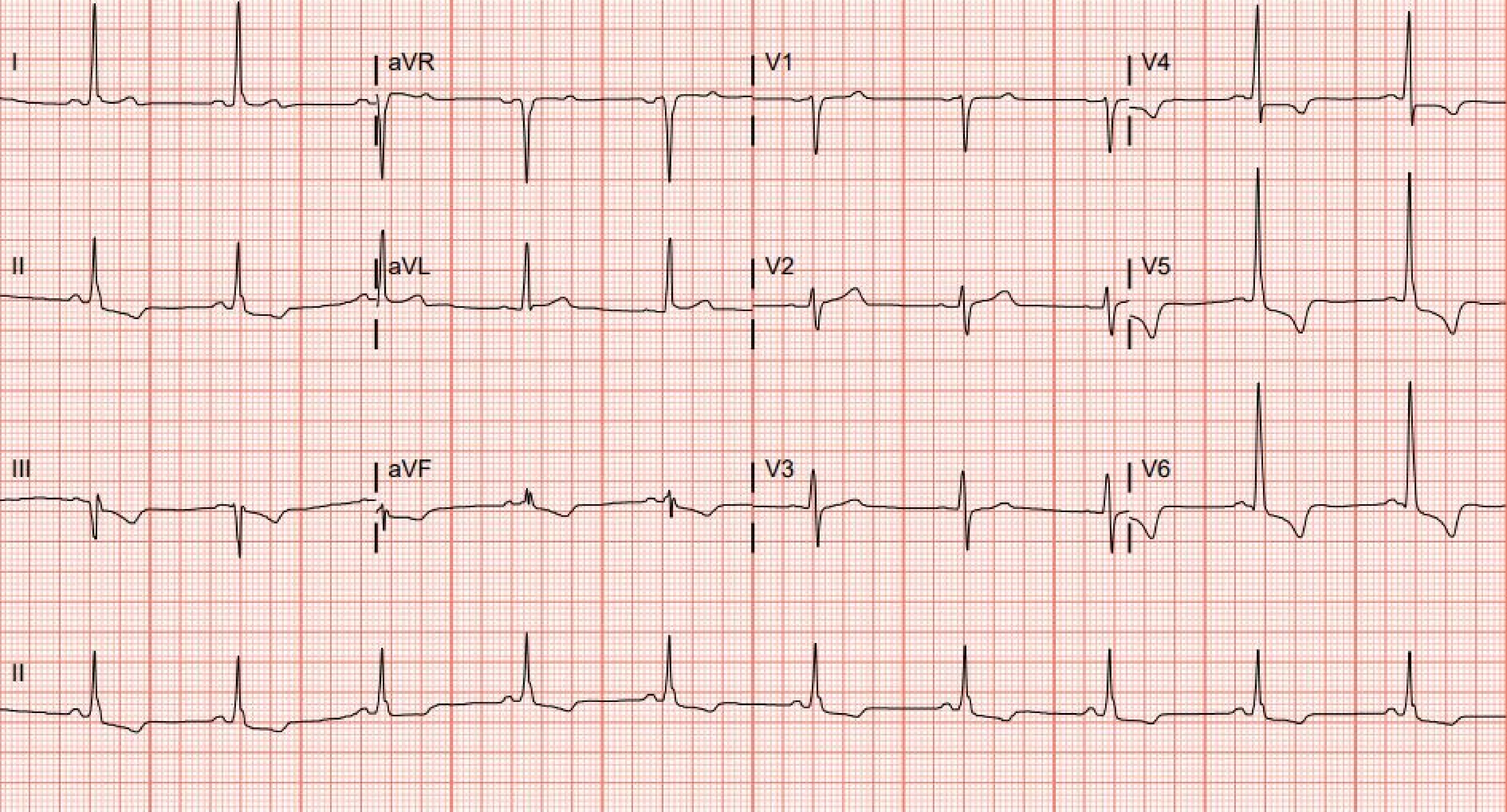
I
aVR
V1
V4
II
aVL
V2
V5
III
aVF
V3
V6
II

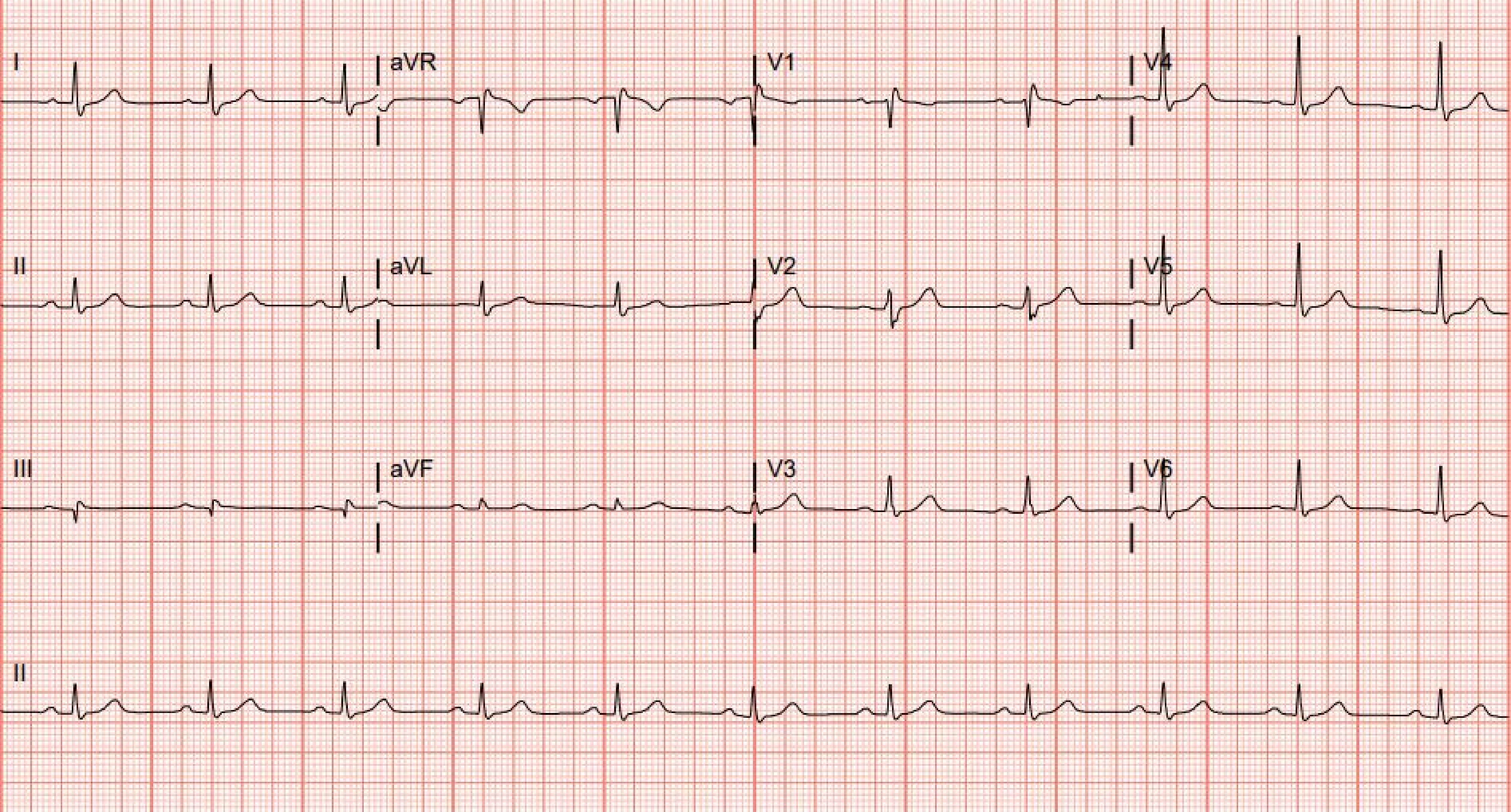
I
aVR
V1
V4
II
aVL
V2
V5
III
aVF
V3
V6
II

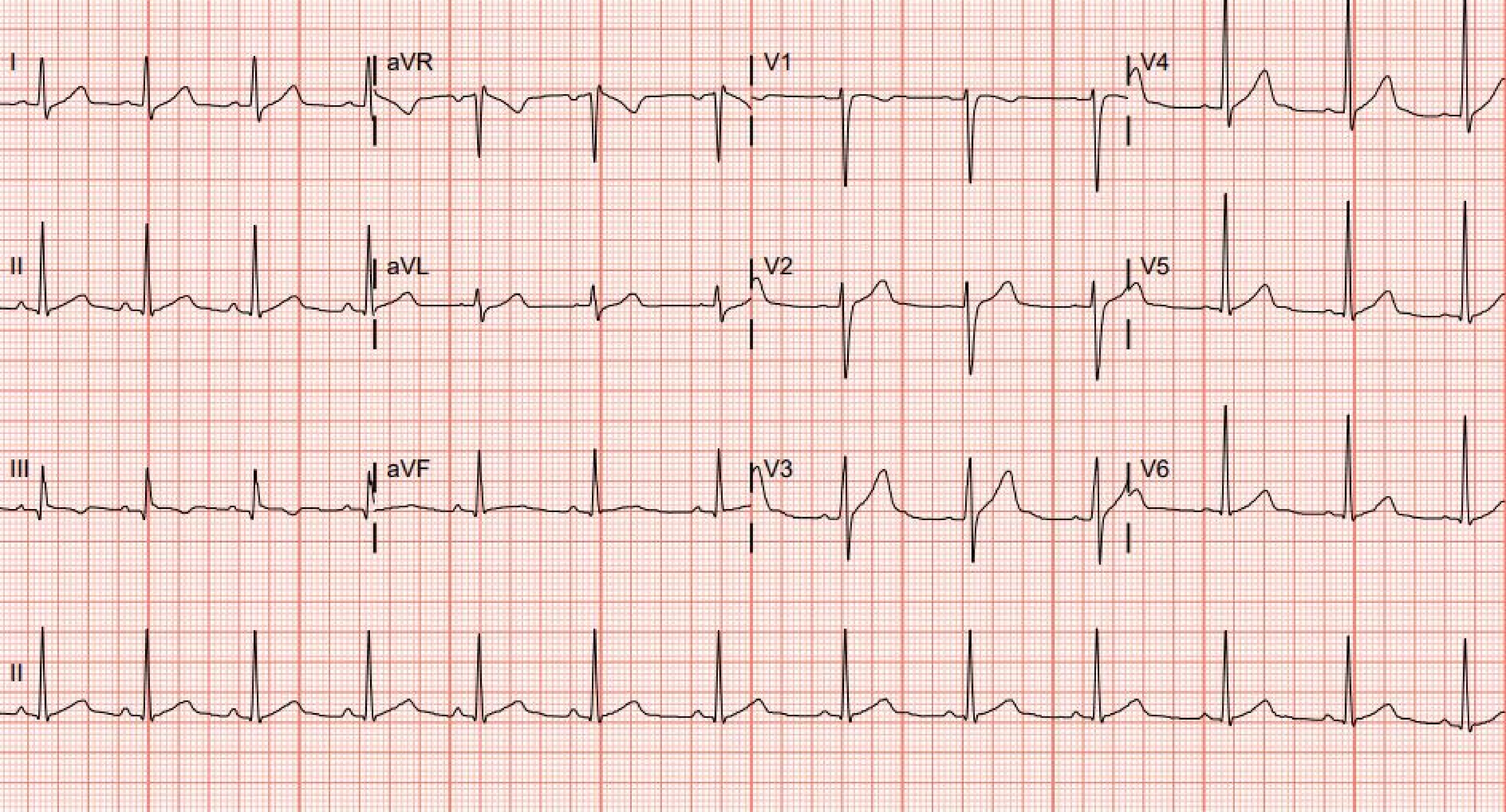
I
aVR
V1
V4
II
aVL
V2
V5
III
aVF
V3
V6
II

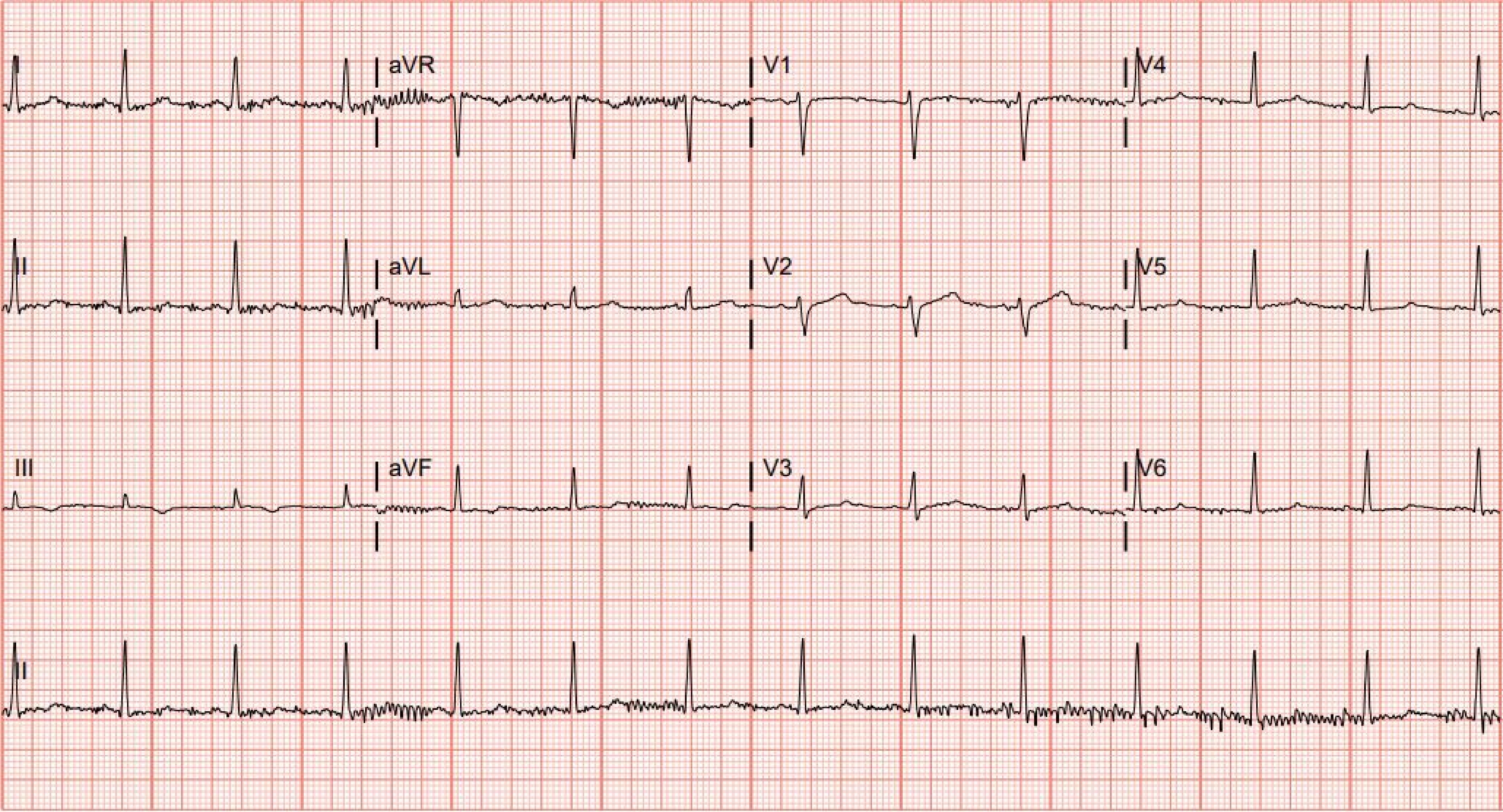
I
aVR
V1
V4
II
aVL
V2
V5
III
aVF
V3
V6
II

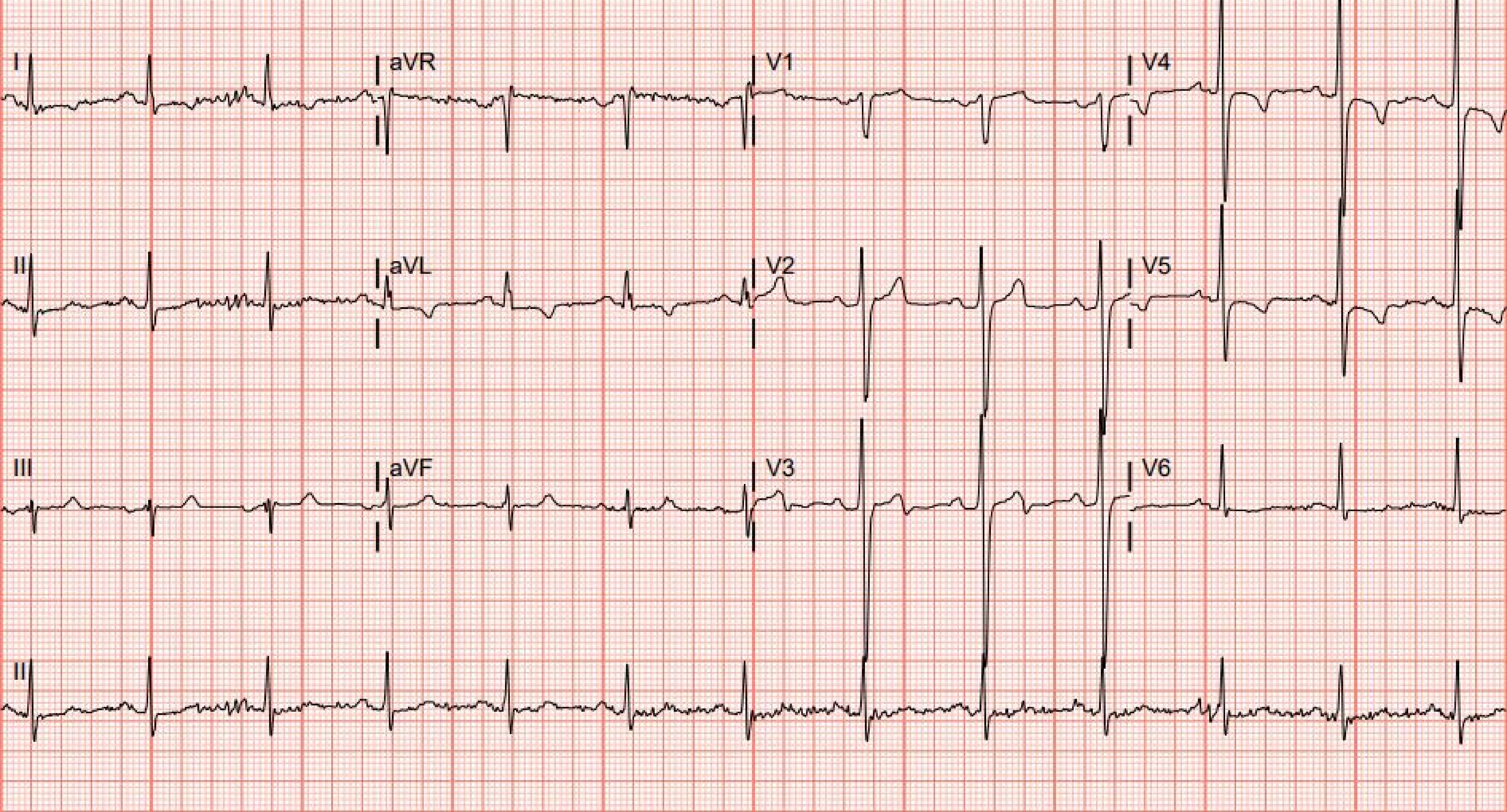
I
aVR
V1
V4
II
aVL
V2
V5
III
aVF
V3
V6
II

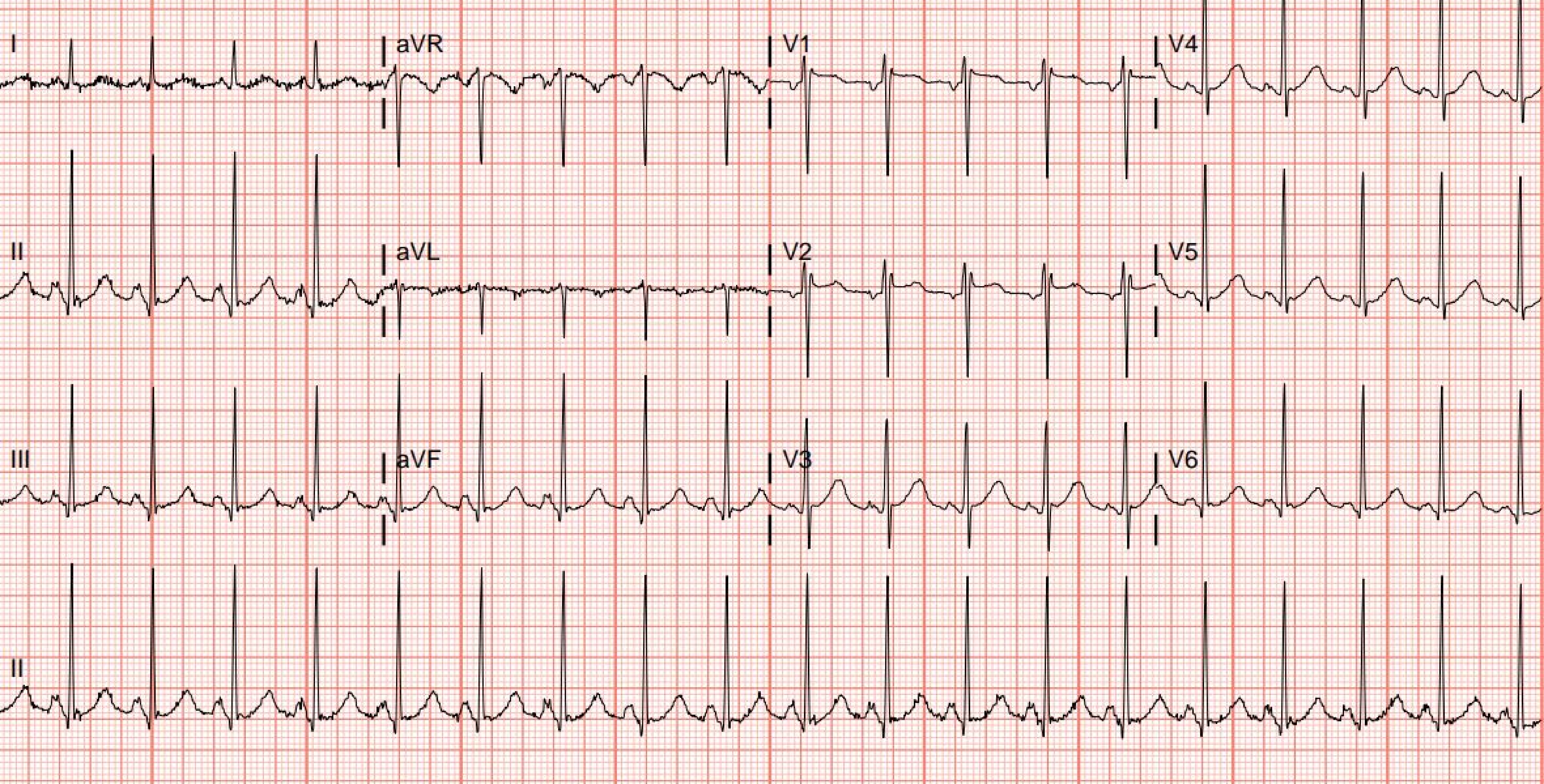
I
aVR
V1
V4
II
aVL
V2
V5
III
aVF
V3
V6
II

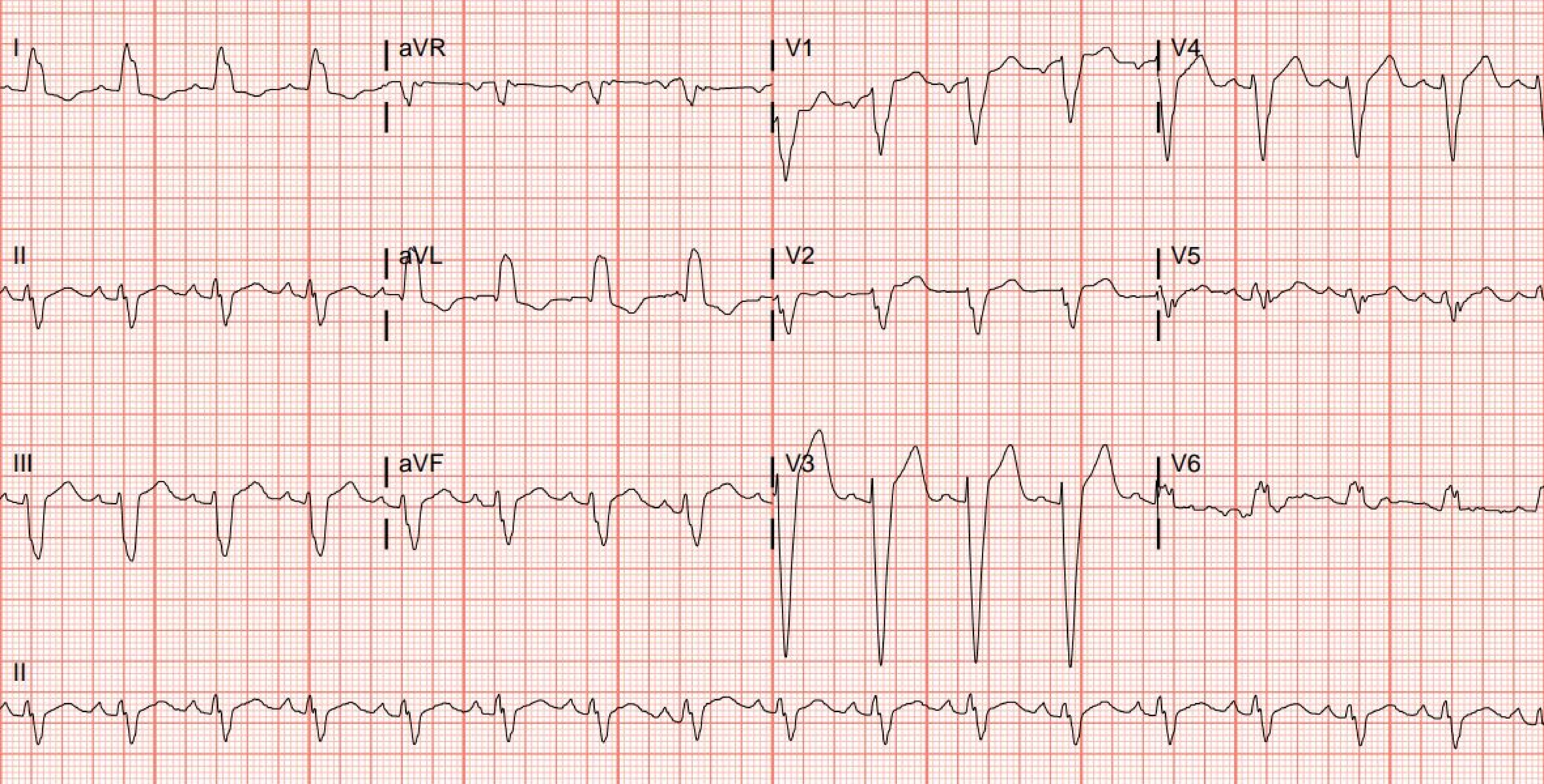
I
aVR
V1
V4
II
aVL
V2
V5
III
aVF
V3
V6
II

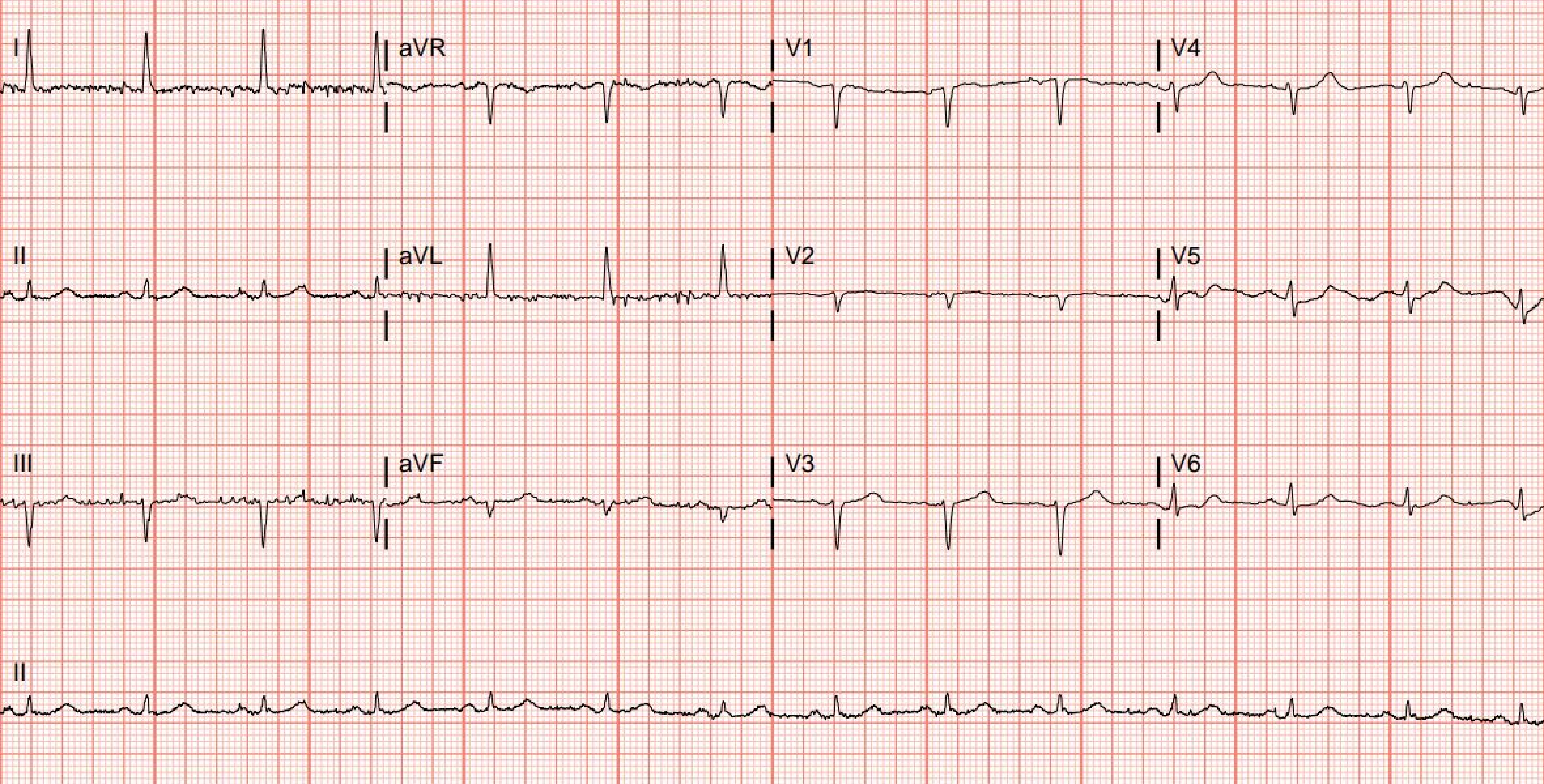
I
aVR
V1
V4
II
aVL
V2
V5
III
aVF
V3
V6
II

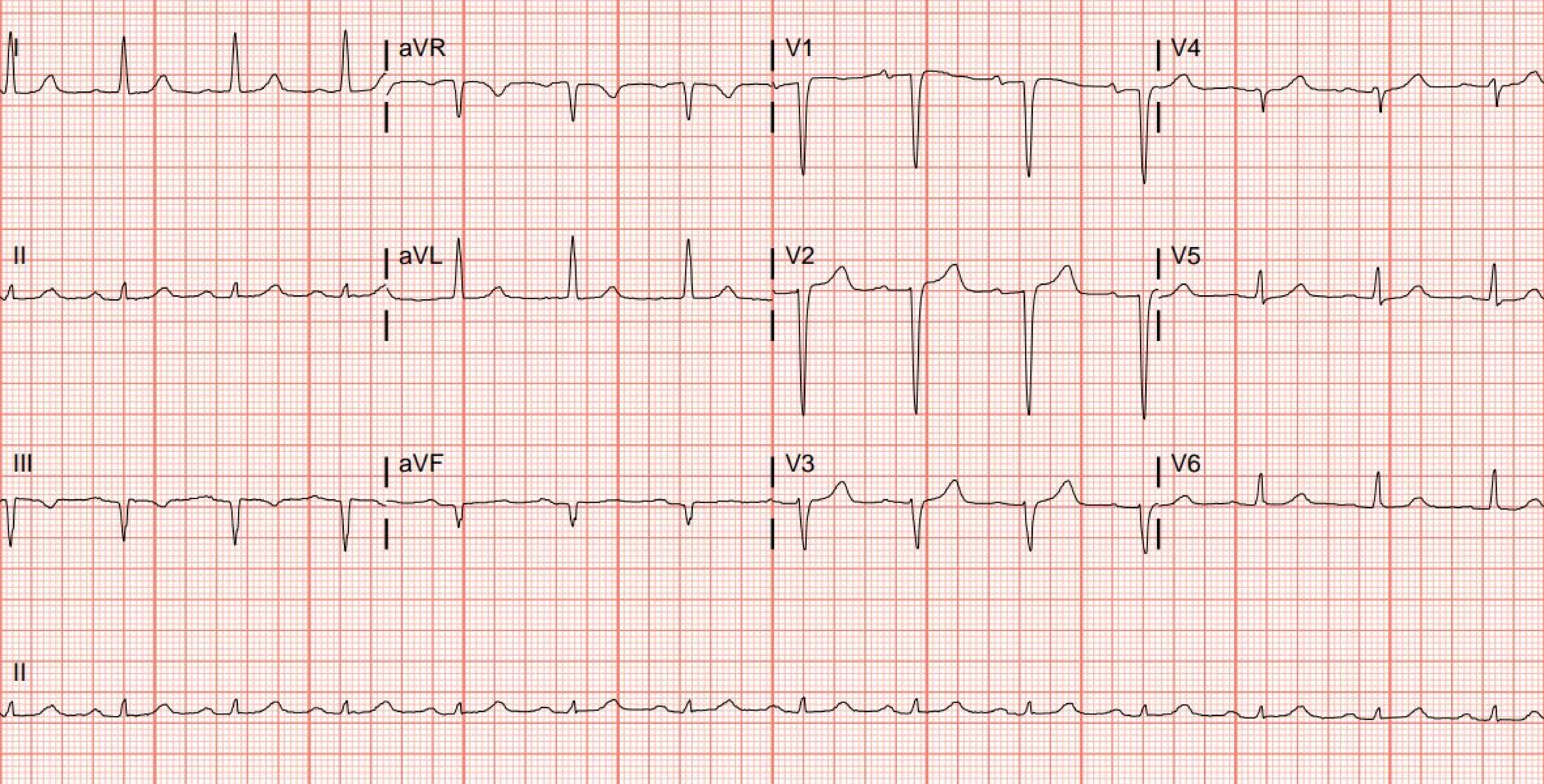
I
aVR
V1
V4
II
aVL
V2
V5
III
aVF
V3
V6
II

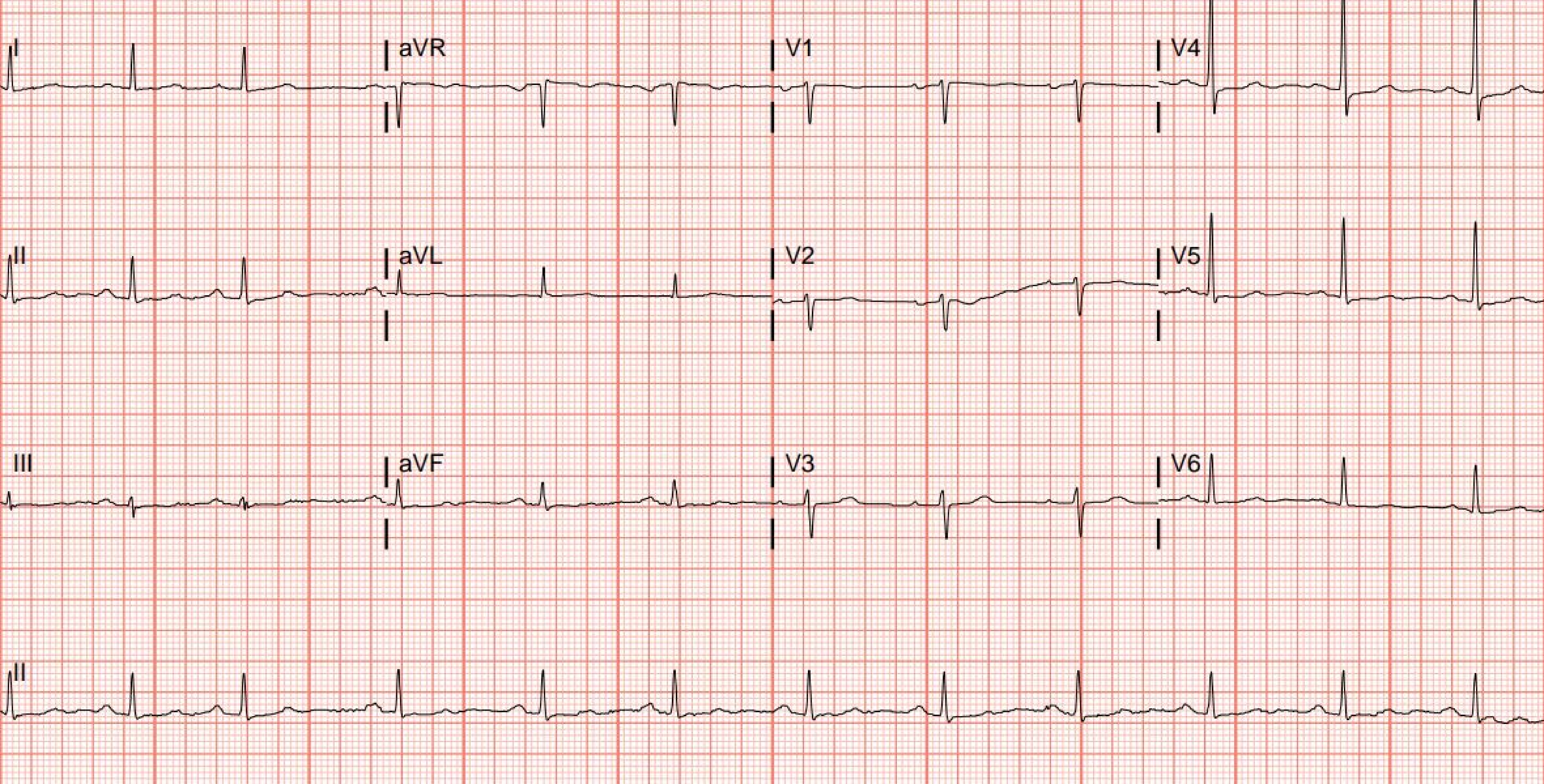
I
aVR
V1
V4
II
aVL
V2
V5
III
aVF
V3
V6
II

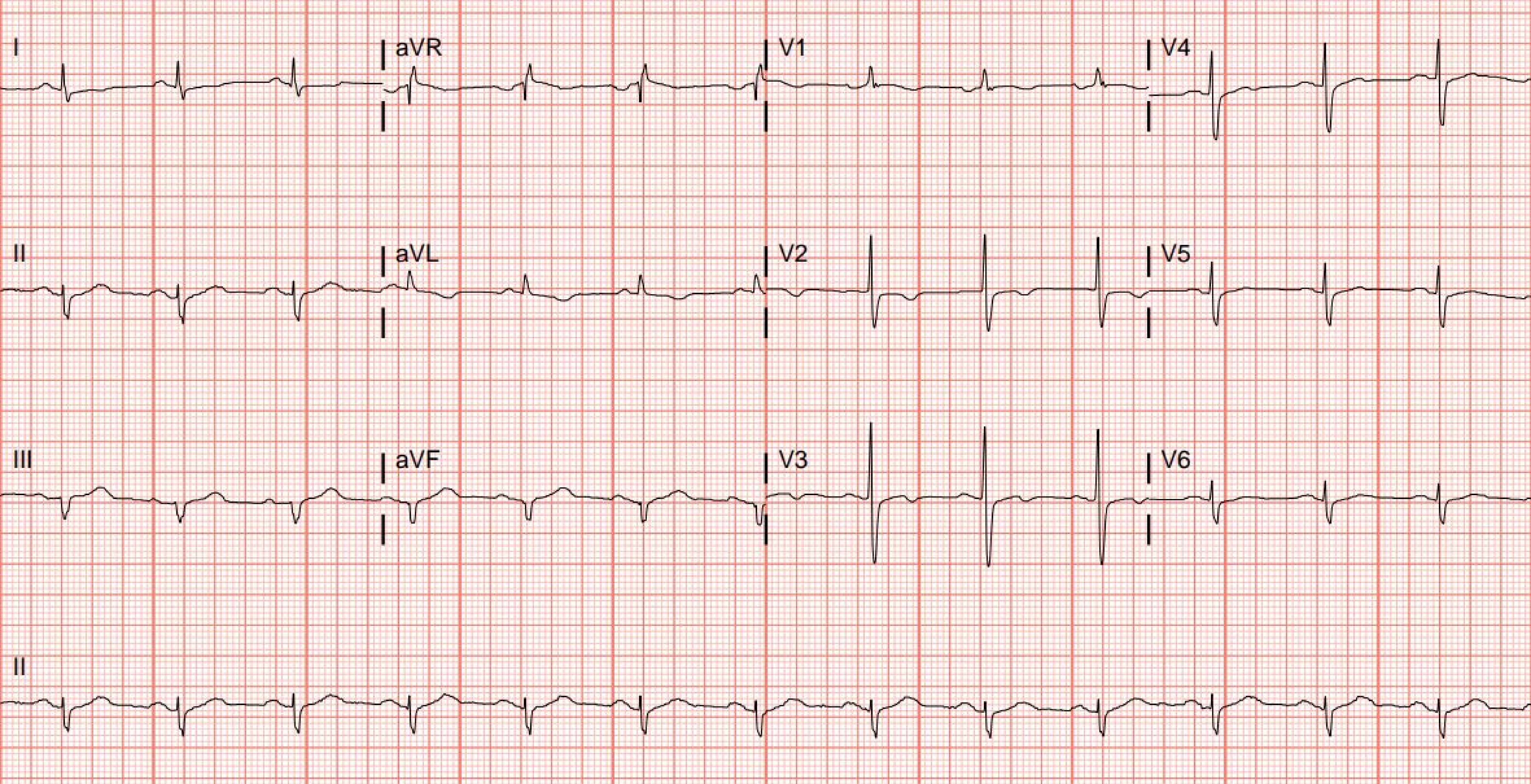
I
aVR
V1
V4
II
aVL
V2
V5
III
aVF
V3
V6
II

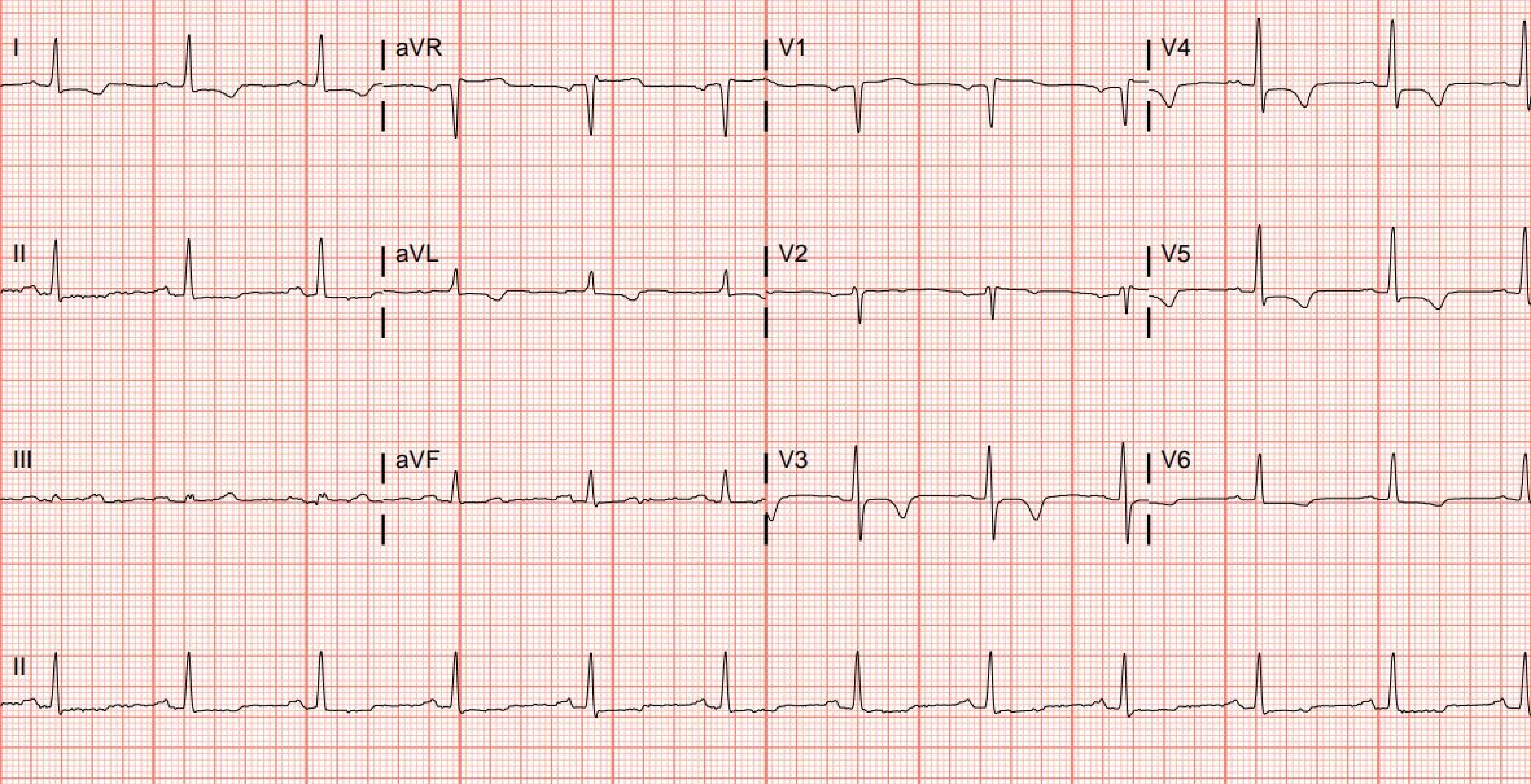
I
aVR
V1
V4
II
aVL
V2
V5
III
aVF
V3
V6
II

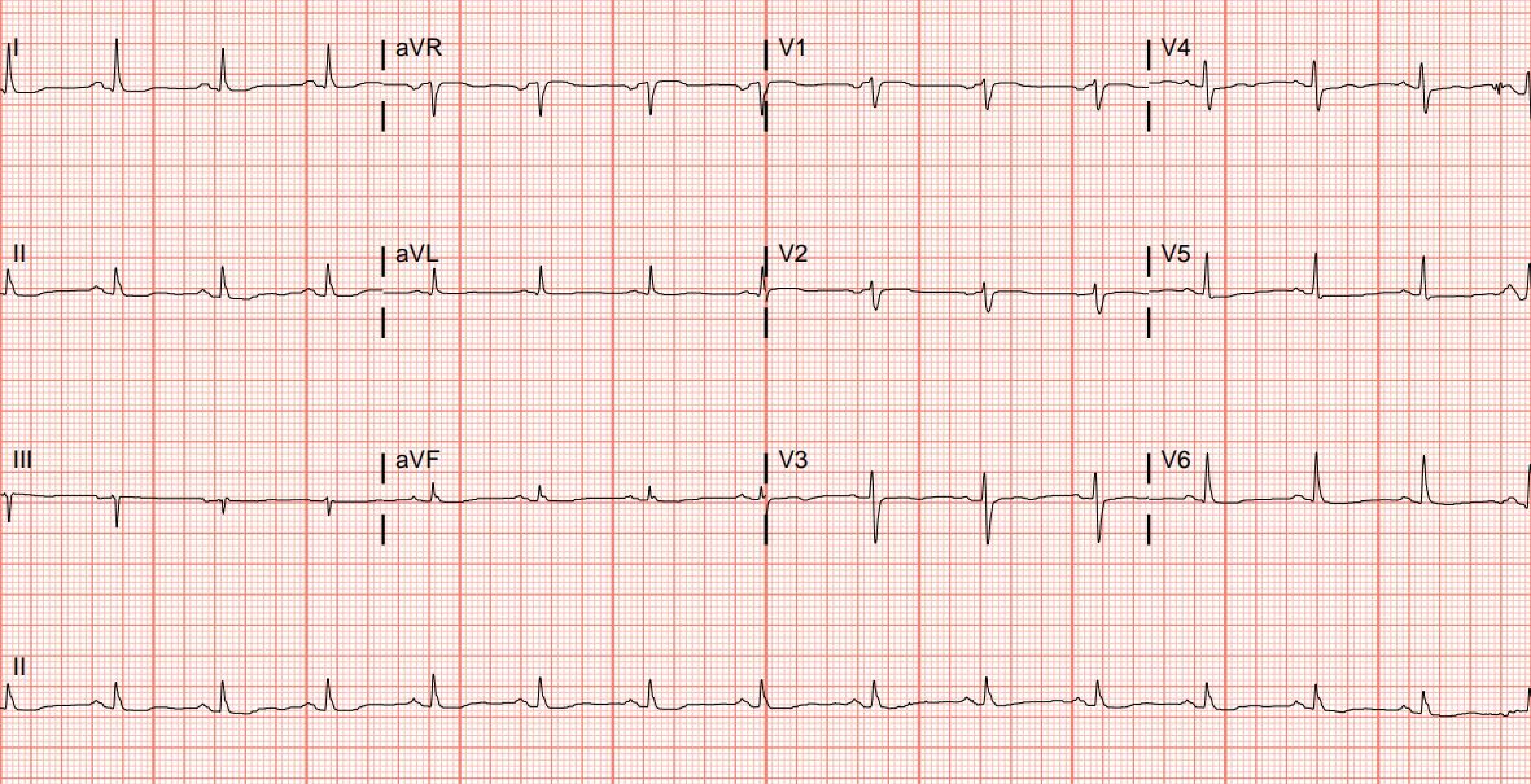
I
aVR
V1
V4
II
aVL
V2
V5
III
aVF
V3
V6
II

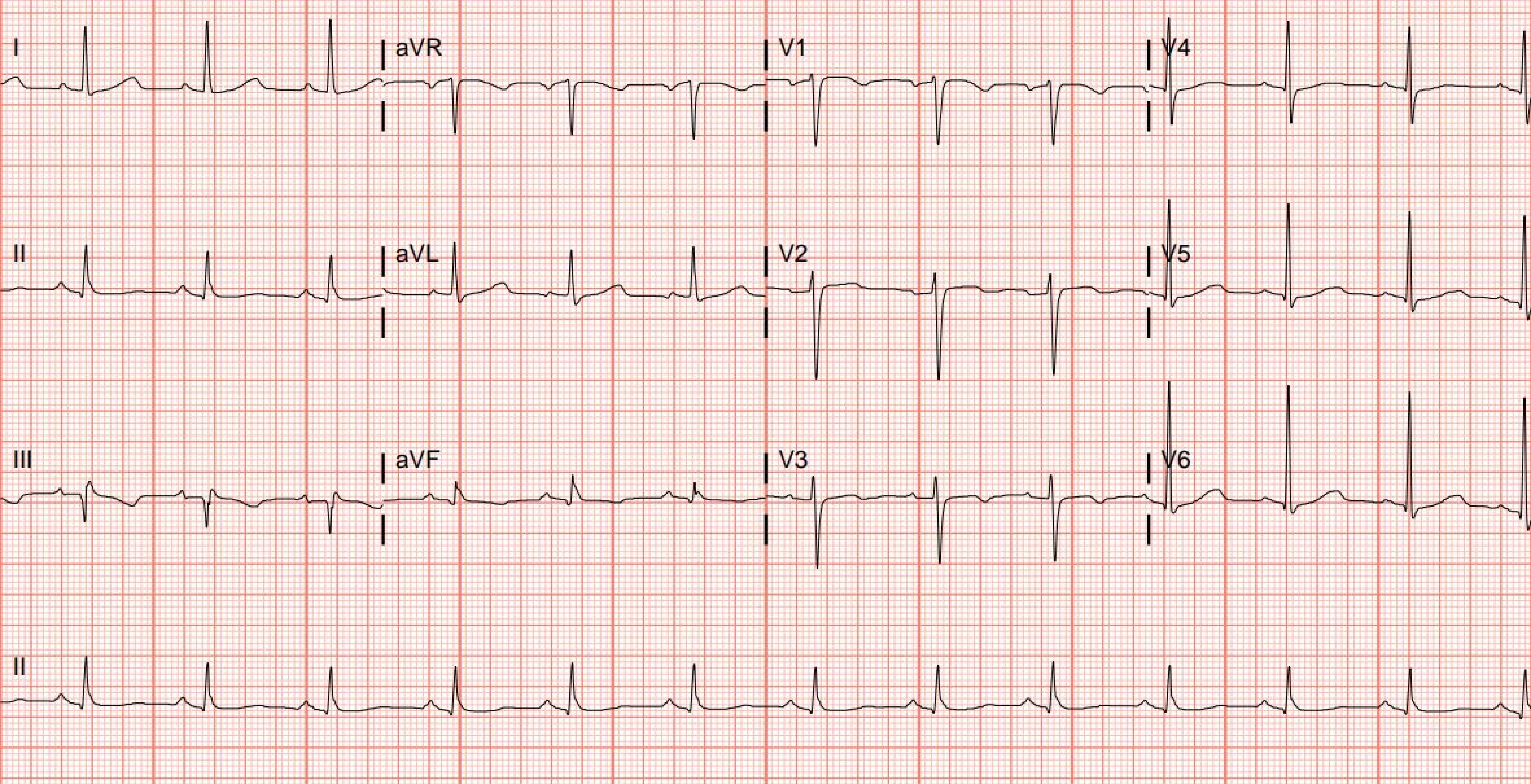
I
aVR
V1
V4
II
aVL
V2
V5
III
aVF
V3
V6
II

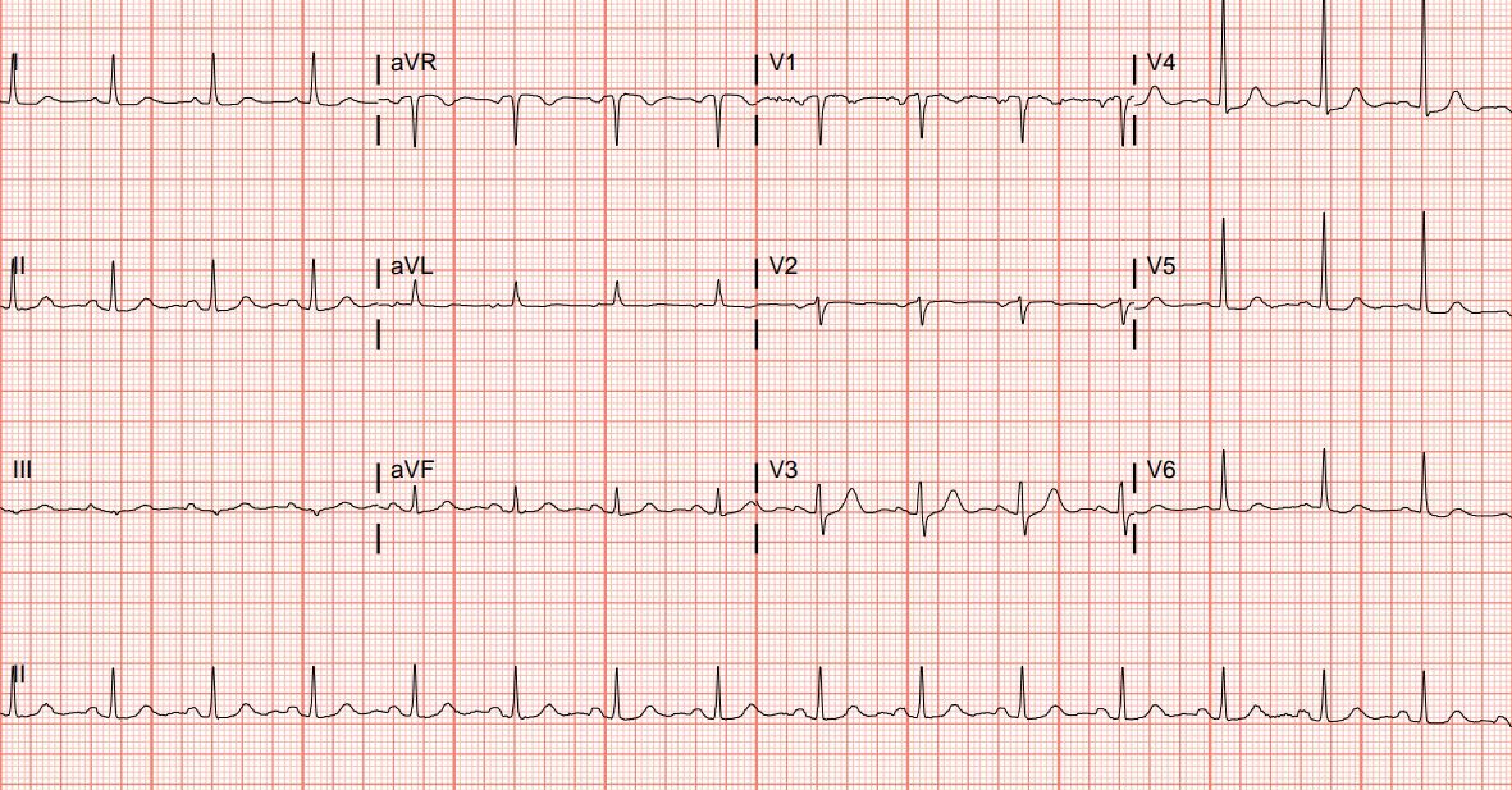
I
aVR
V1
V4
II
aVL
V2
V5
III
aVF
V3
V6
II

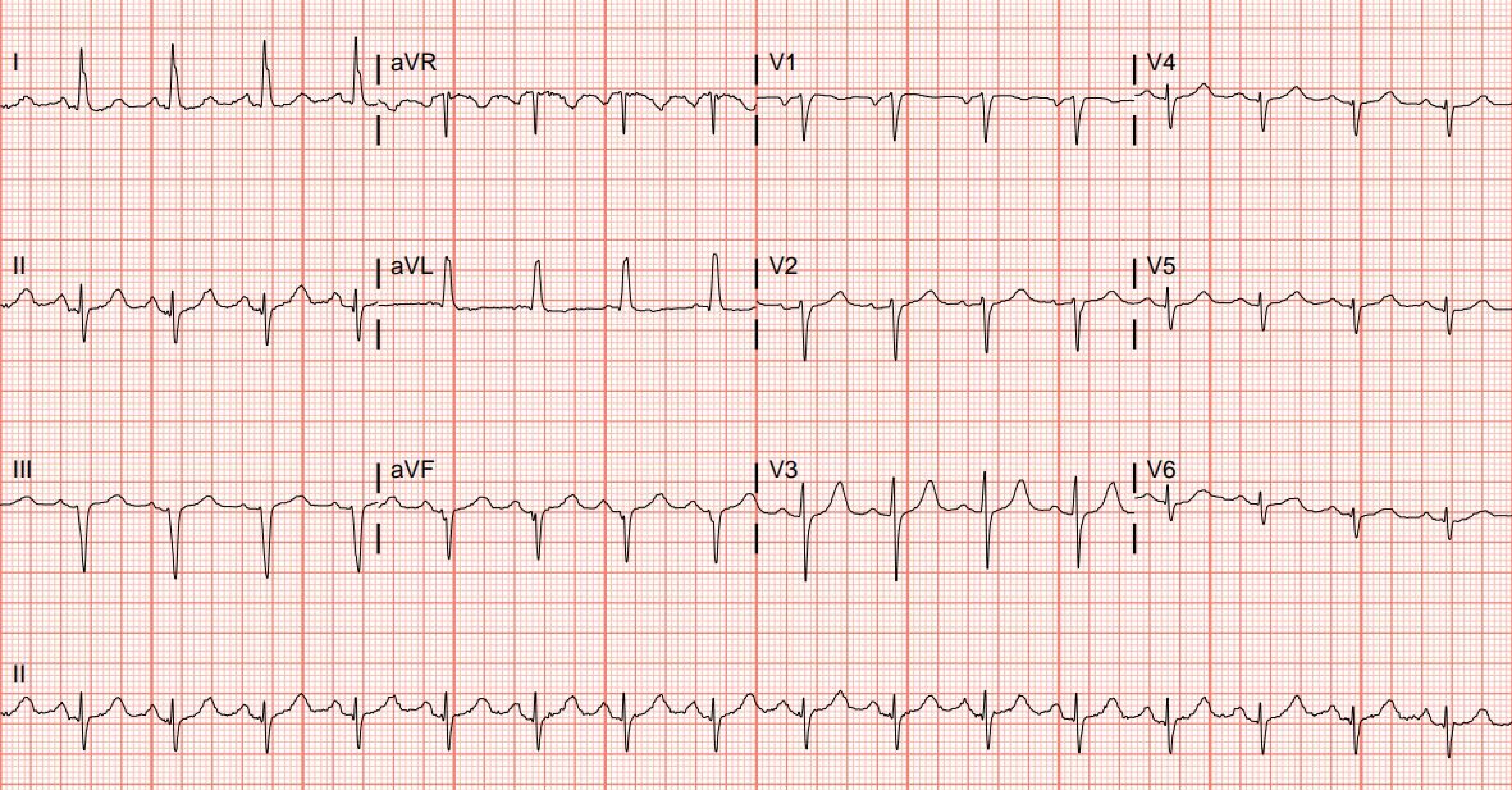
I
aVR
V1
V4
II
aVL
V2
V5
III
aVF
V3
V6
II

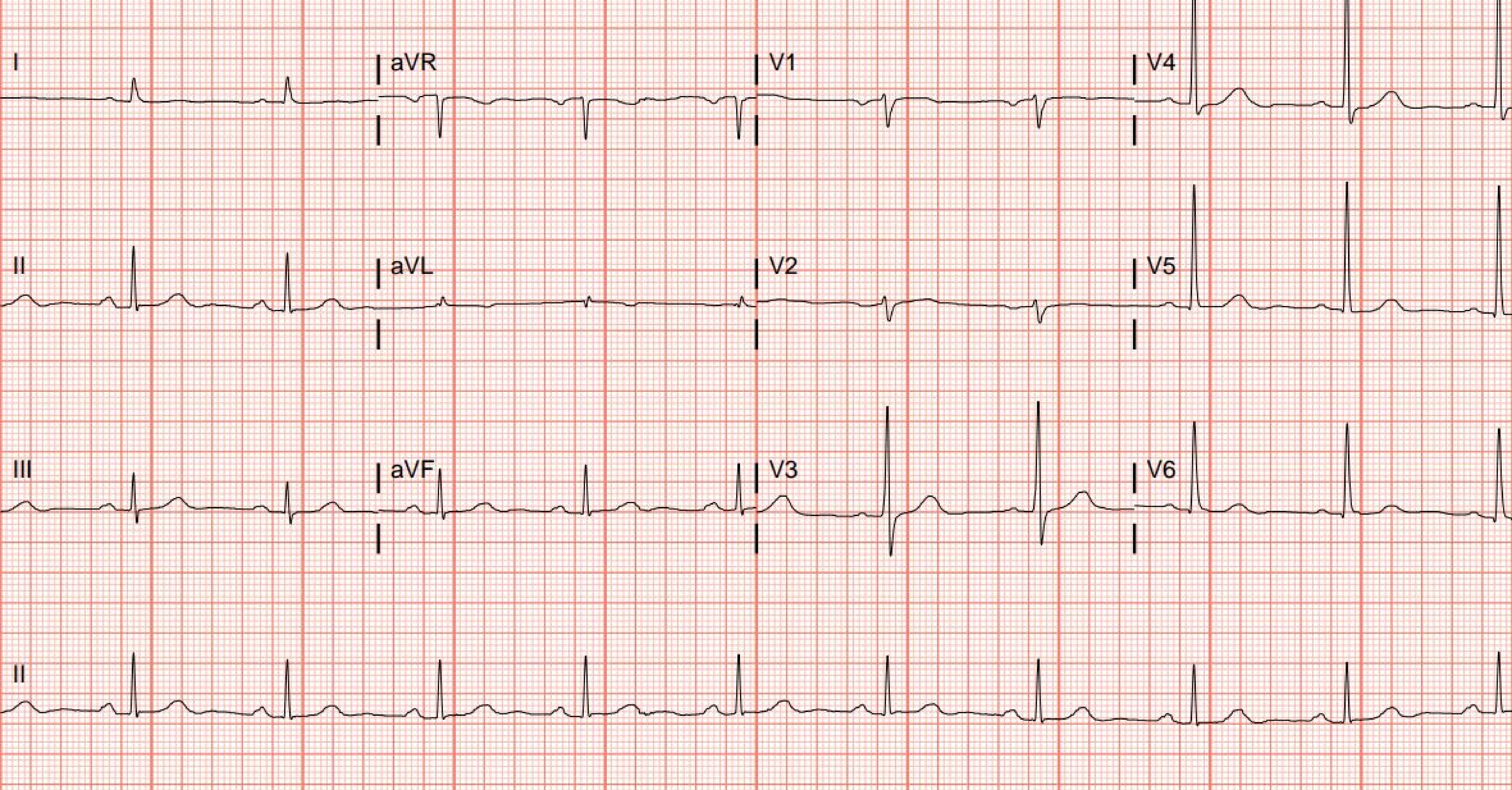
I
aVR
V1
V4
II
aVL
V2
V5
III
aVF
V3
V6
II

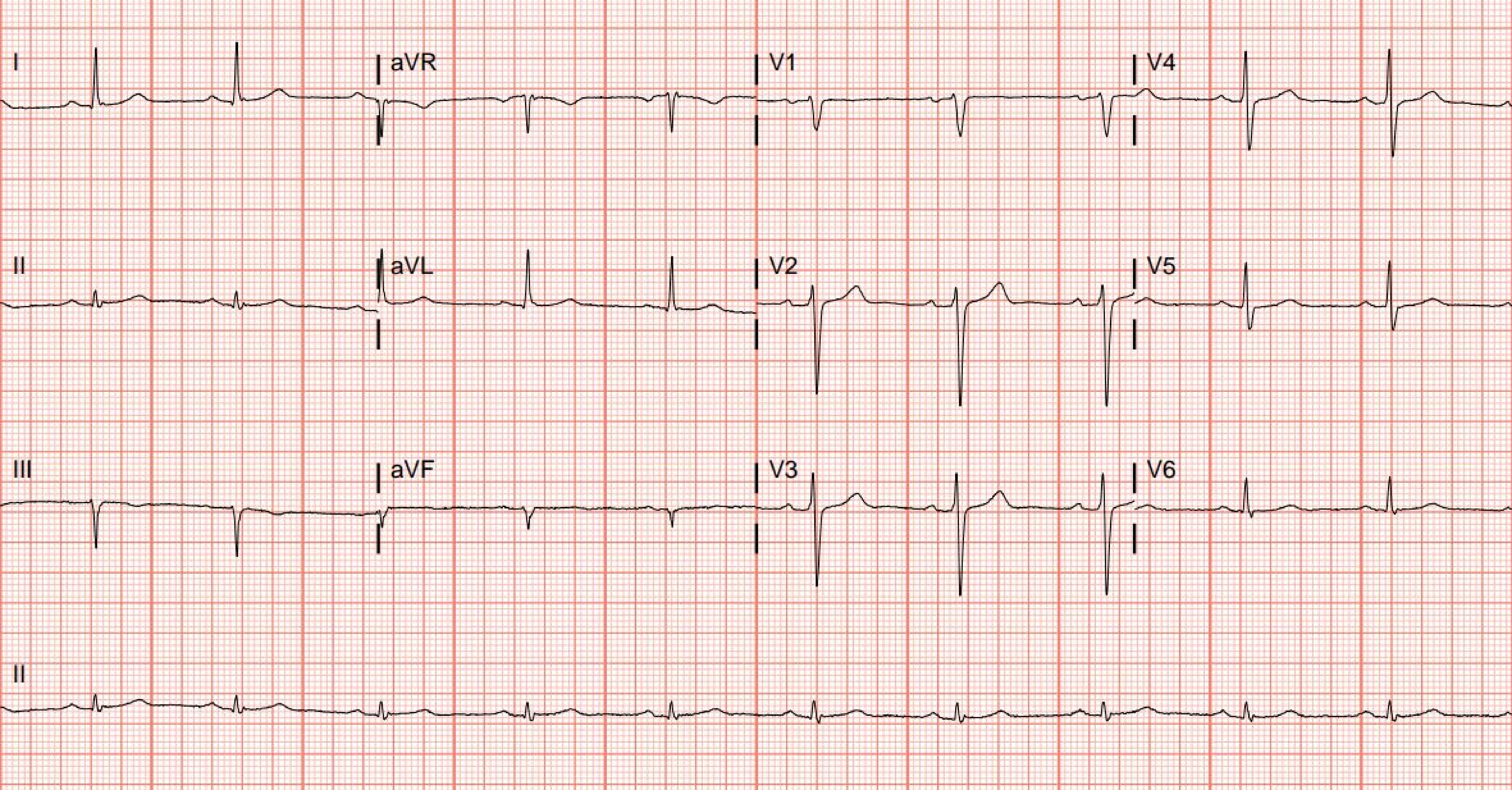
I
aVR
V1
V4
II
aVL
V2
V5
III
aVF
V3
V6
II

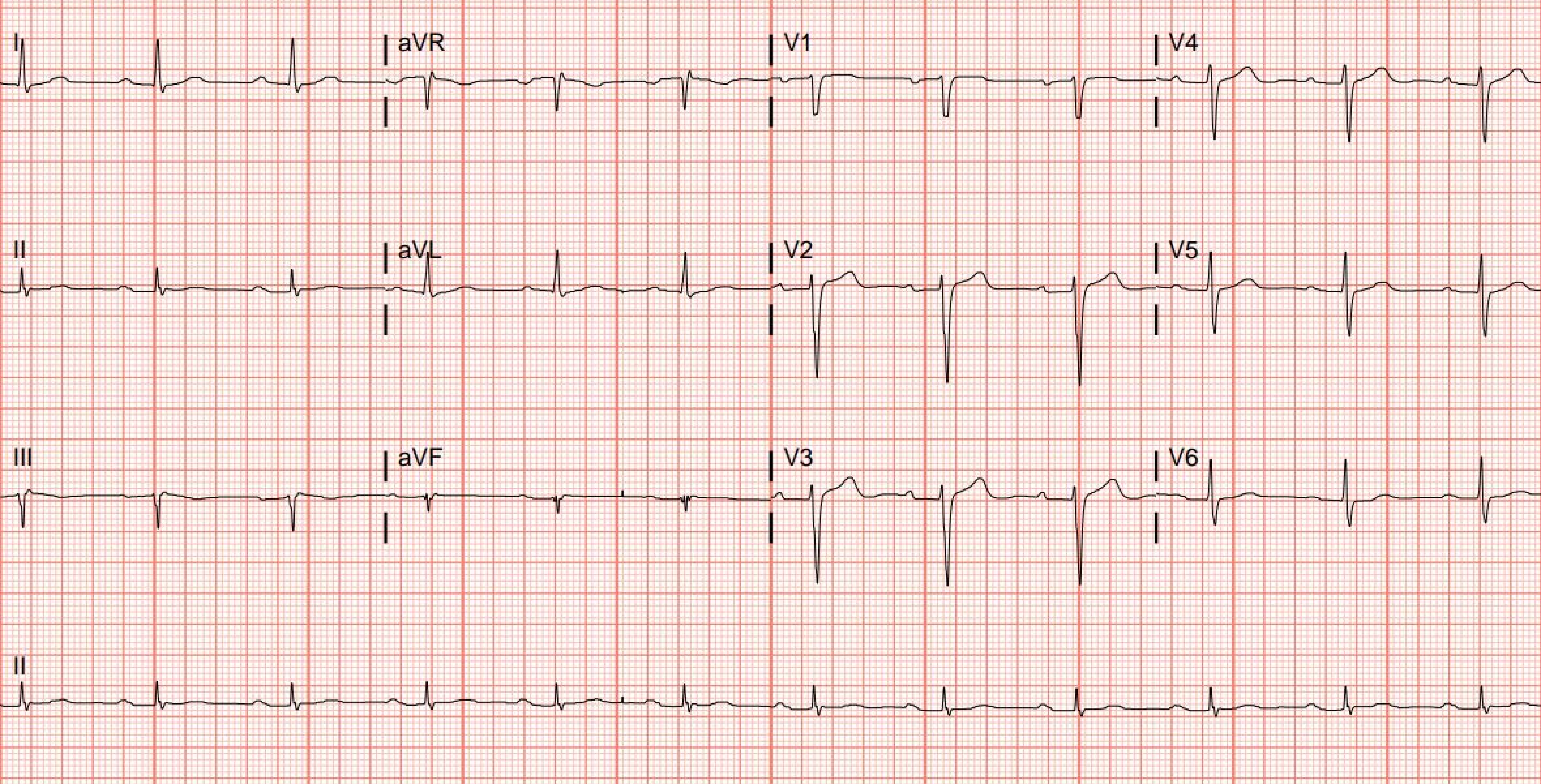
I
aVR
V1
V4
II
aVL
V2
V5
III
aVF
V3
V6
II

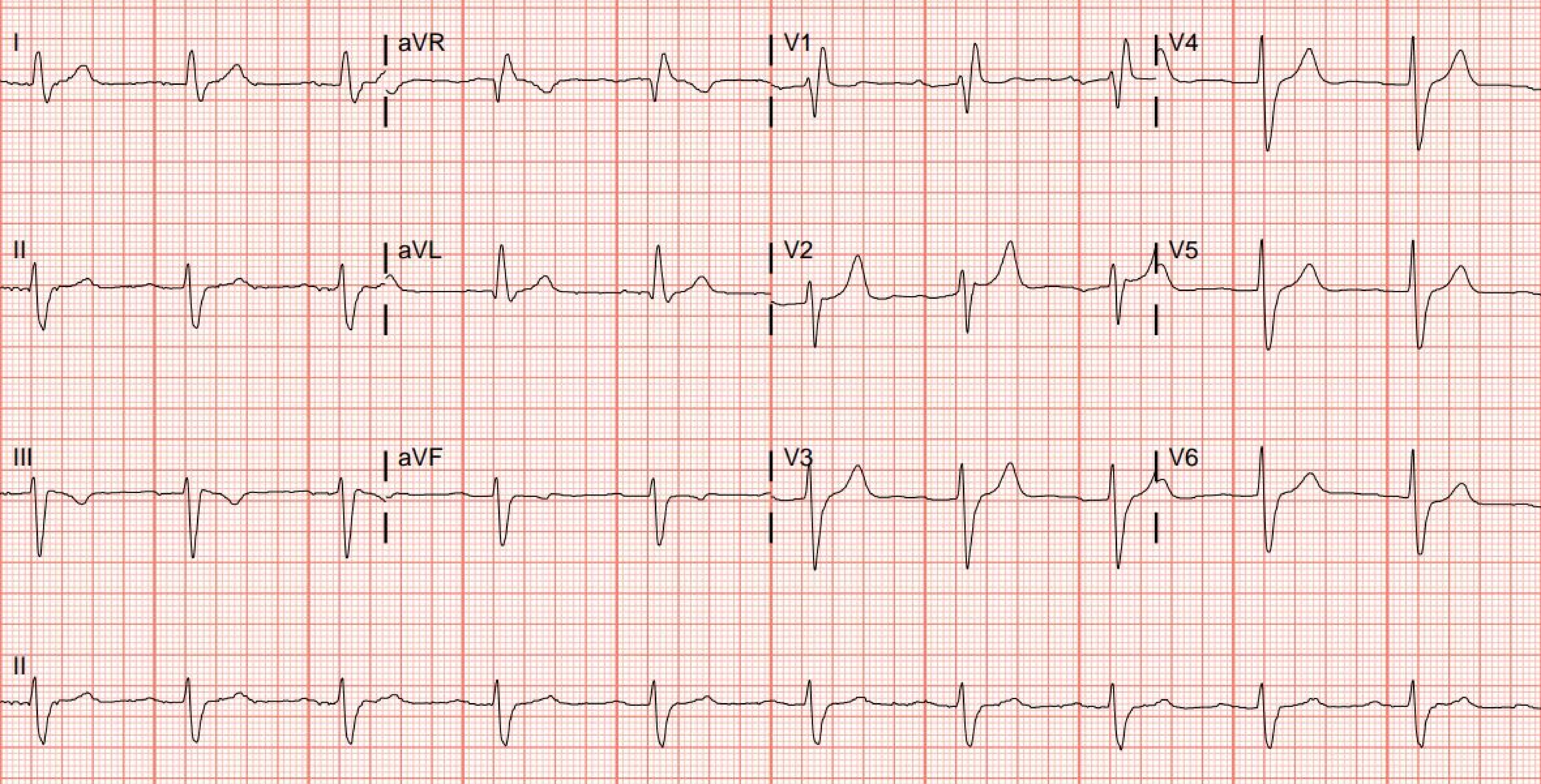
I
aVR
V1
V4
II
aVL
V2
V5
III
aVF
V3
V6
II

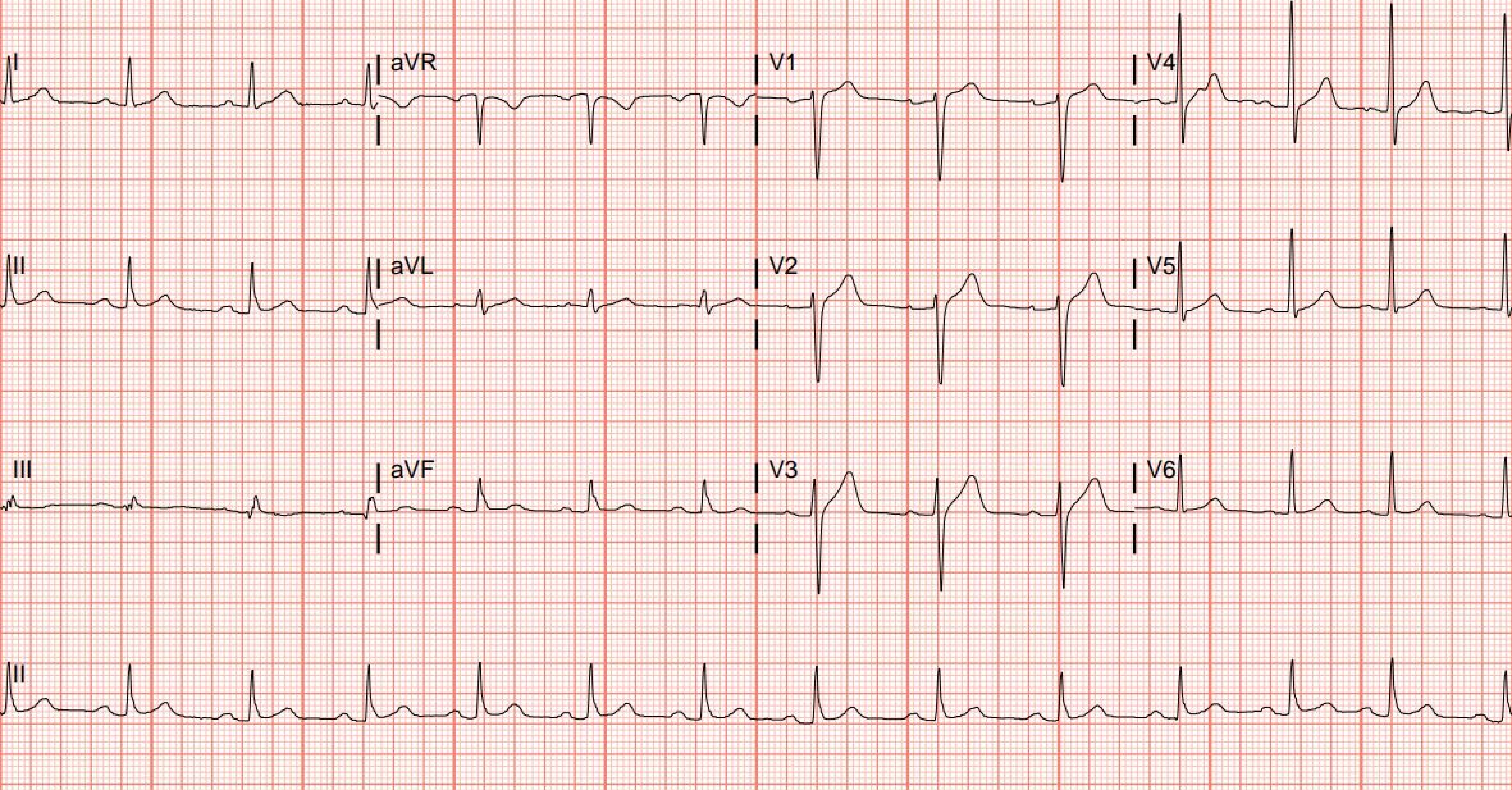
I
aVR
V1
V4
II
aVL
V2
V5
III
aVF
V3
V6
II

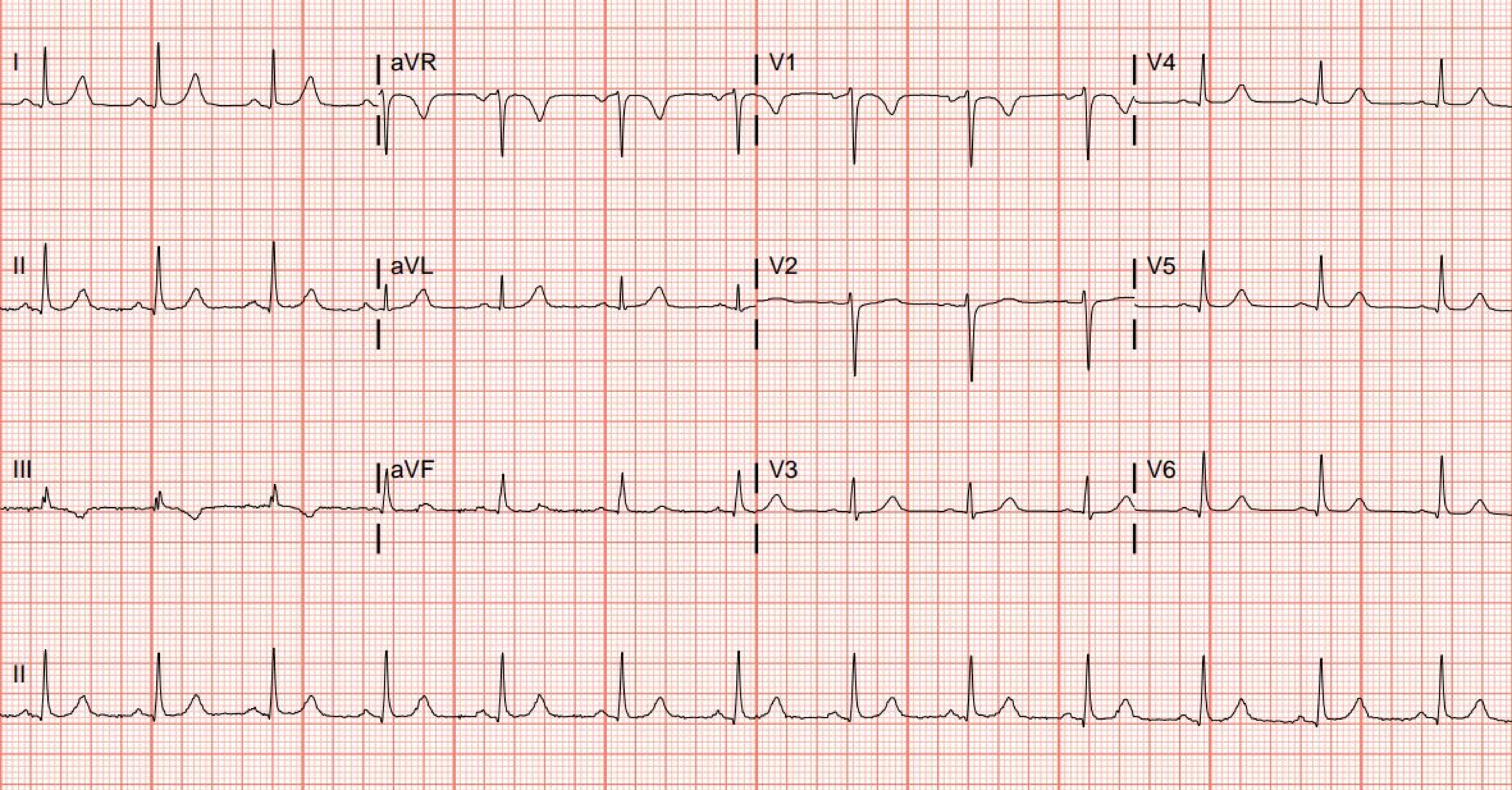

I
aVR
V1
V4
II
aVL
V2
V5
III
aVF
V3
V6
II

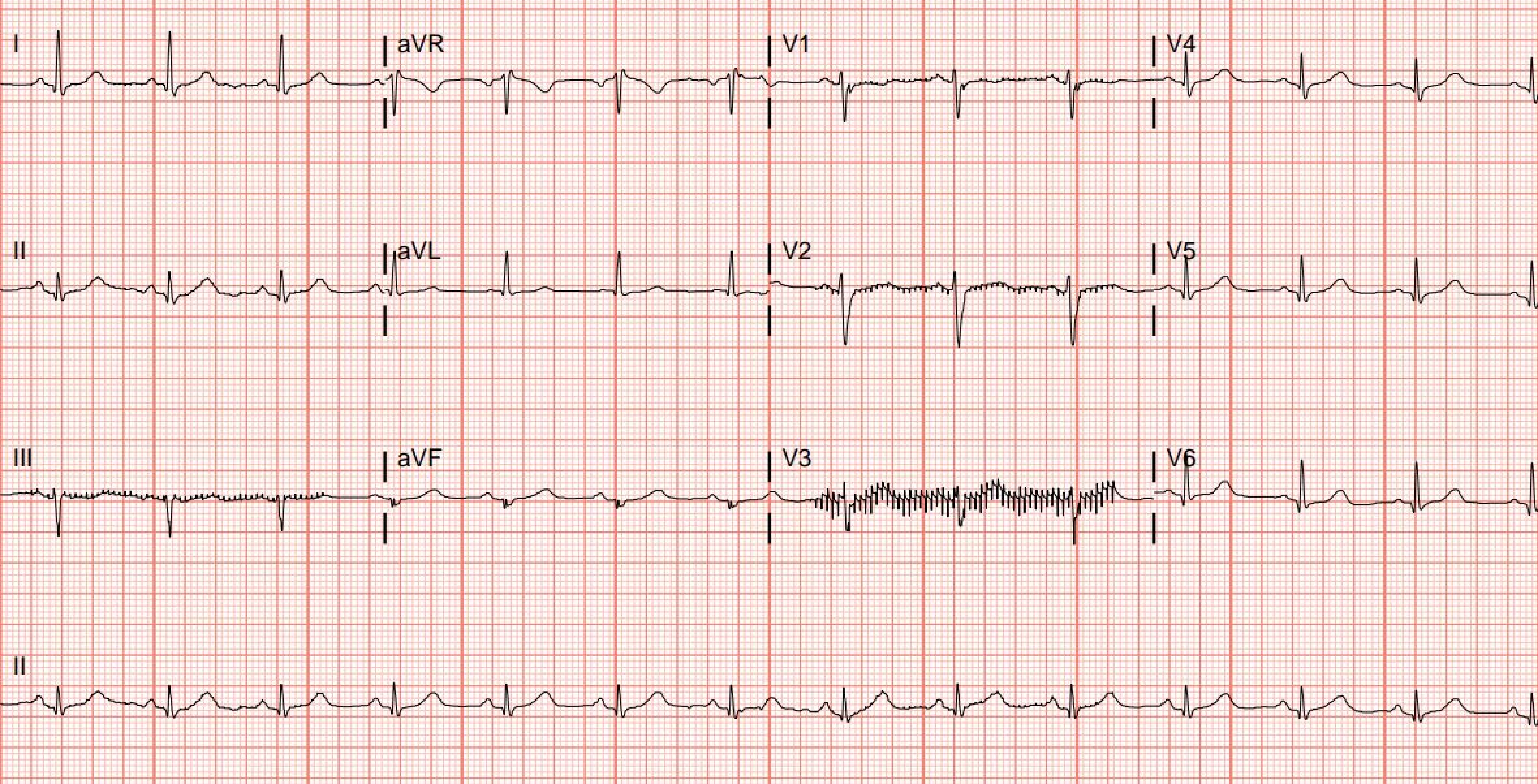
I
aVR
V1
V4
II
aVL
V2
V5
III
aVF
V3
V6
II

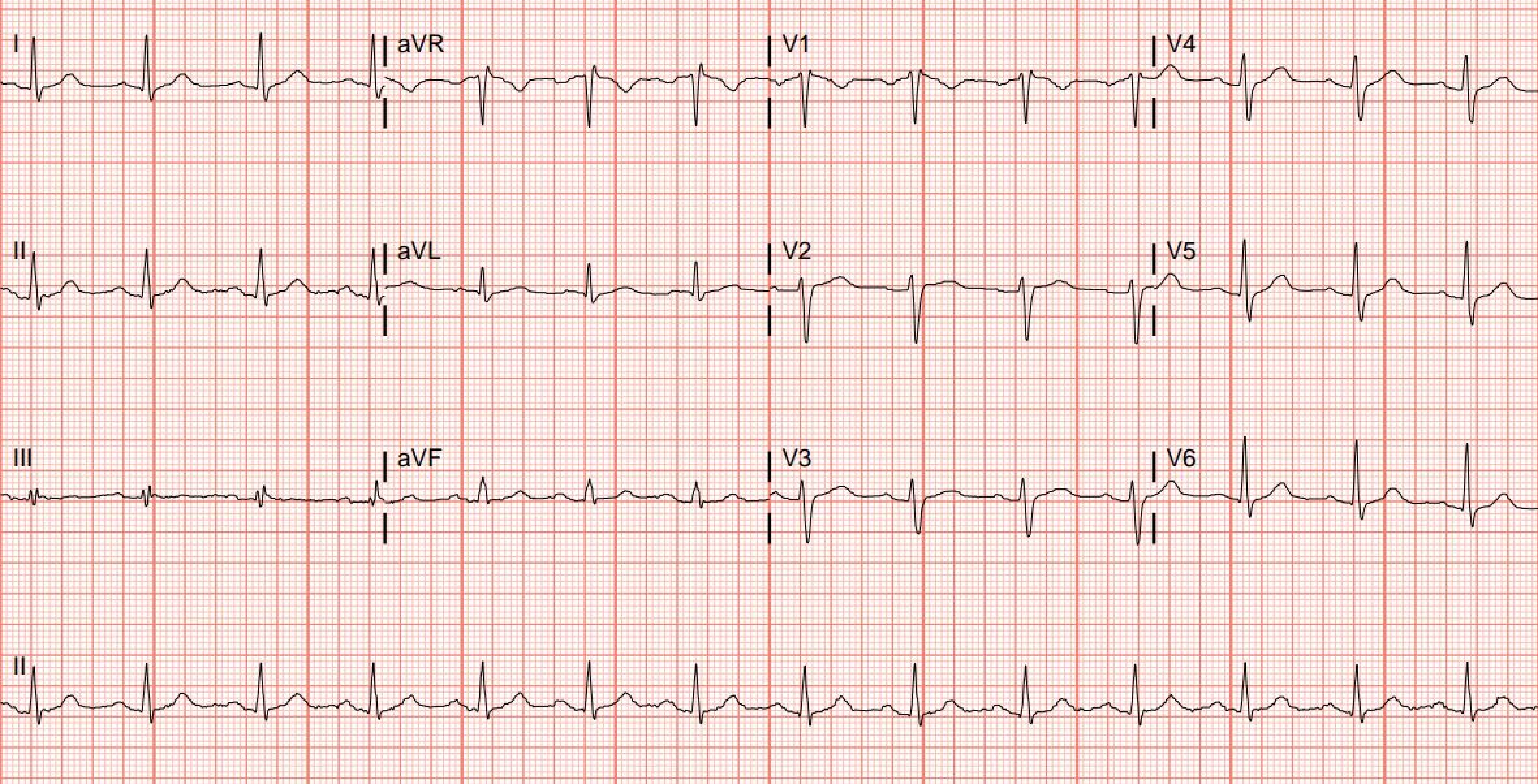
I
aVR
V1
V4
II
aVL
V2
V5
III
aVF
V3
V6
II

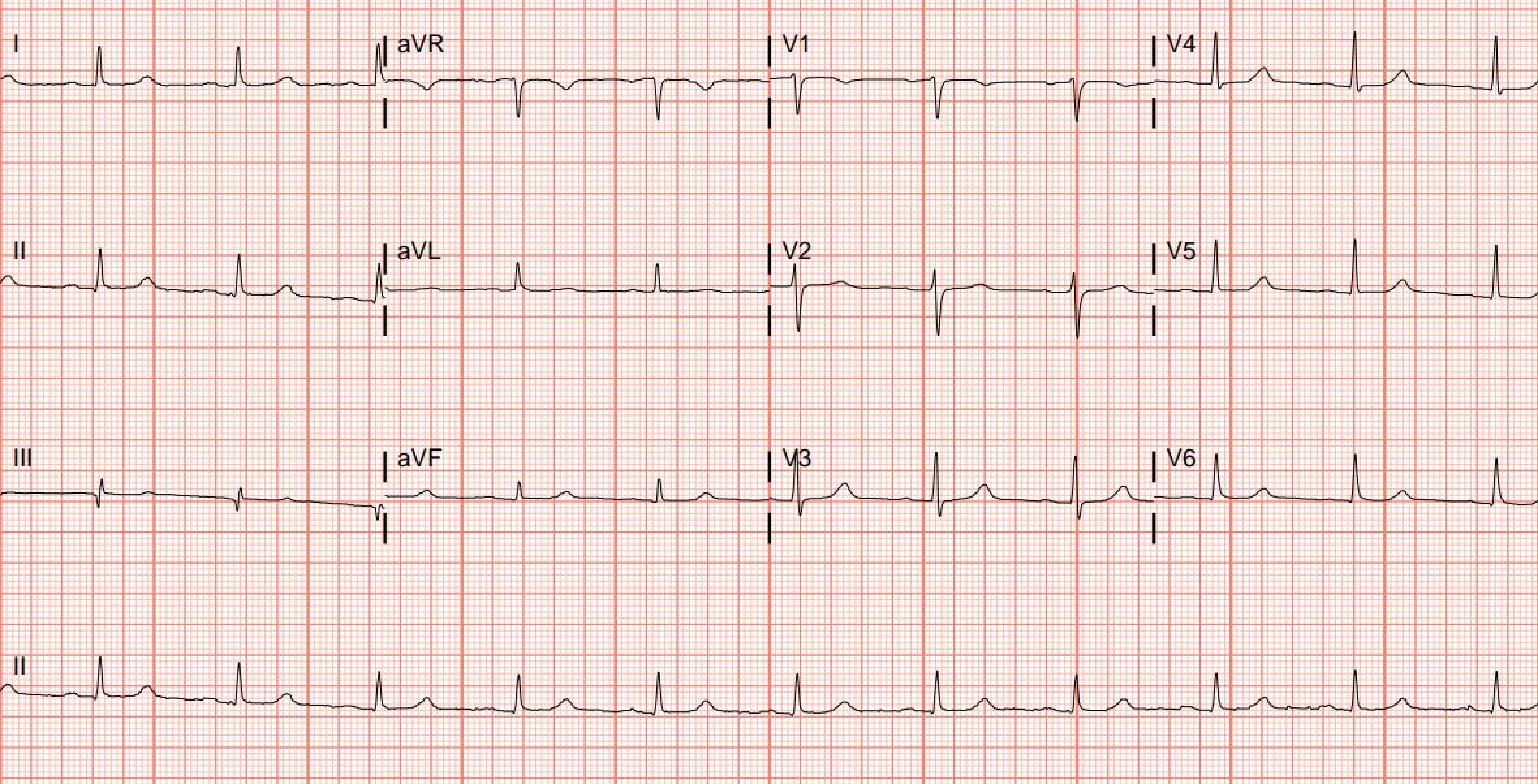
I
aVR
V1
V4
II
aVL
V2
V5
III
aVF
V3
V6
II

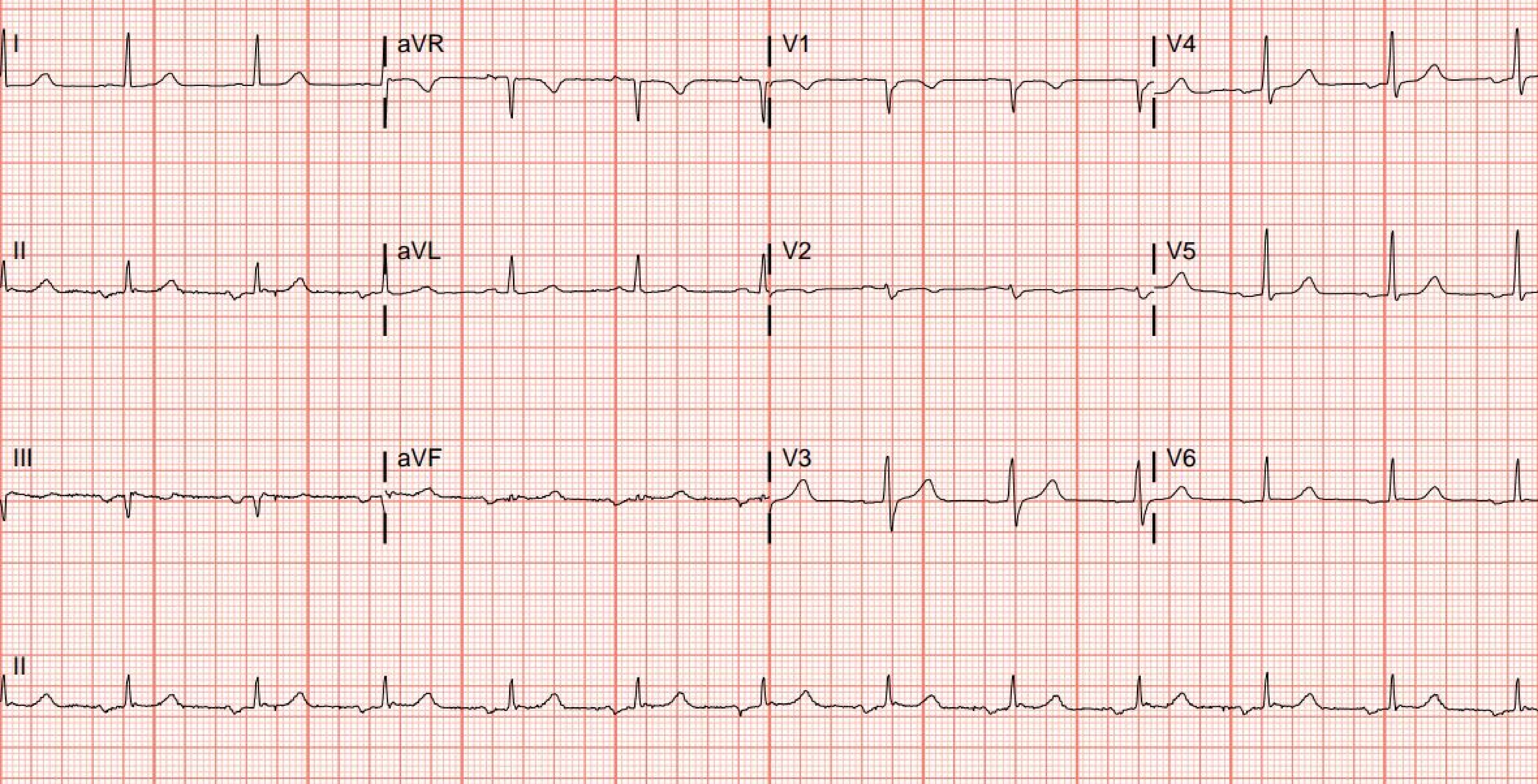
I
aVR
V1
V4
II
aVL
V2
V5
III
aVF
V3
V6
II

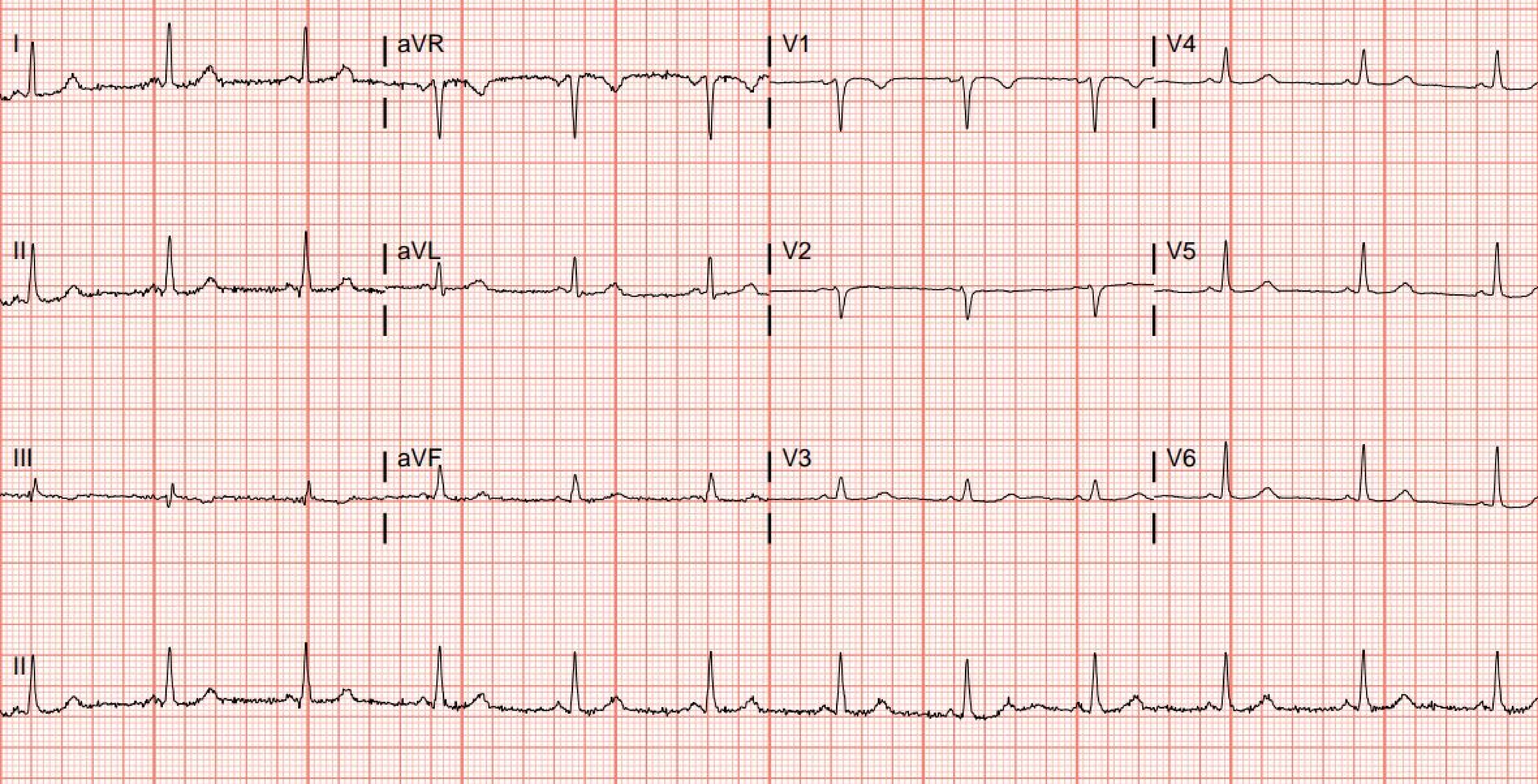
I
aVR
V1
V4
II
aVL
V2
V5
III
aVF
V3
V6
II

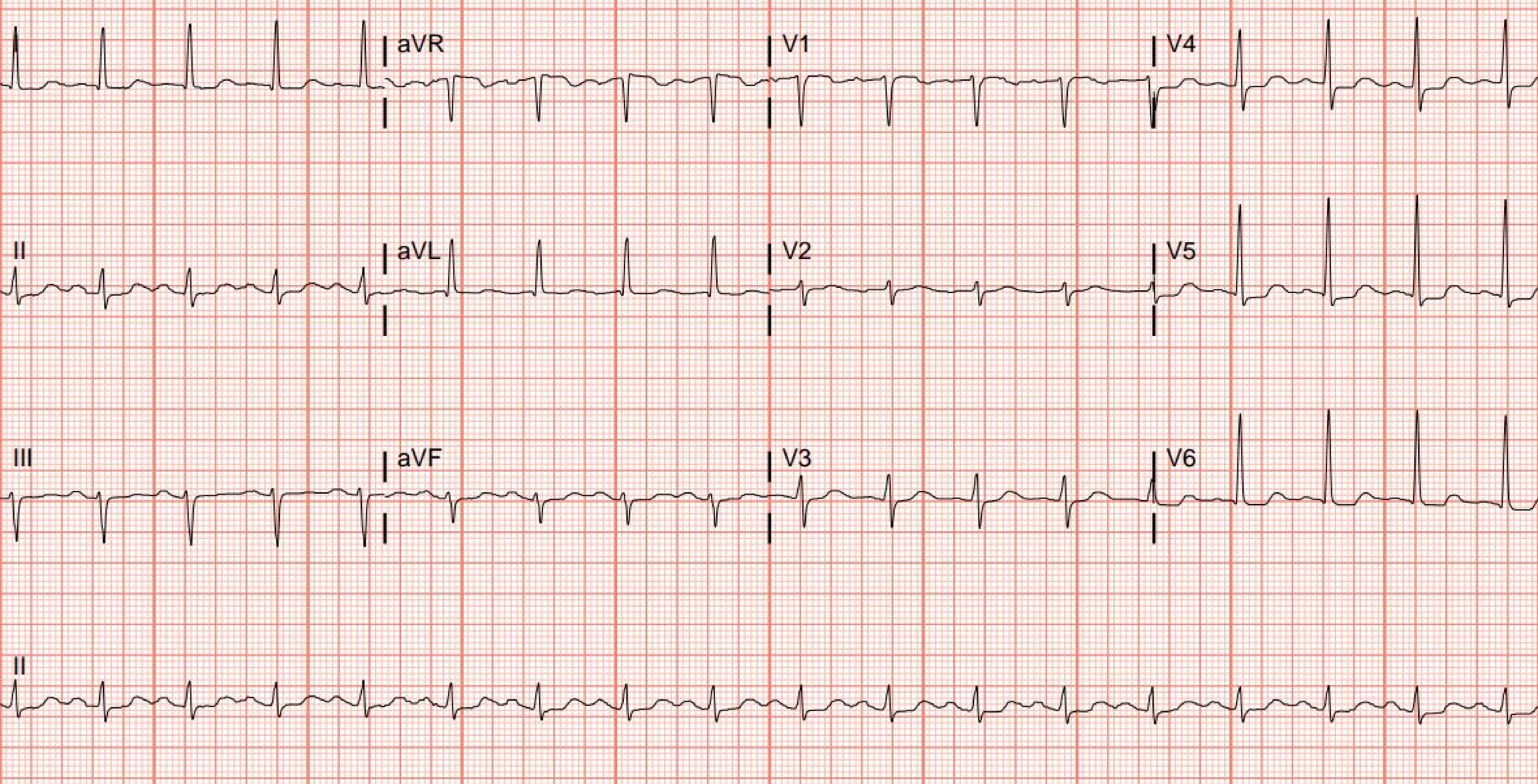
aVR
V1
V4
II
aVL
V2
V5
III
aVF
V3
V6
II

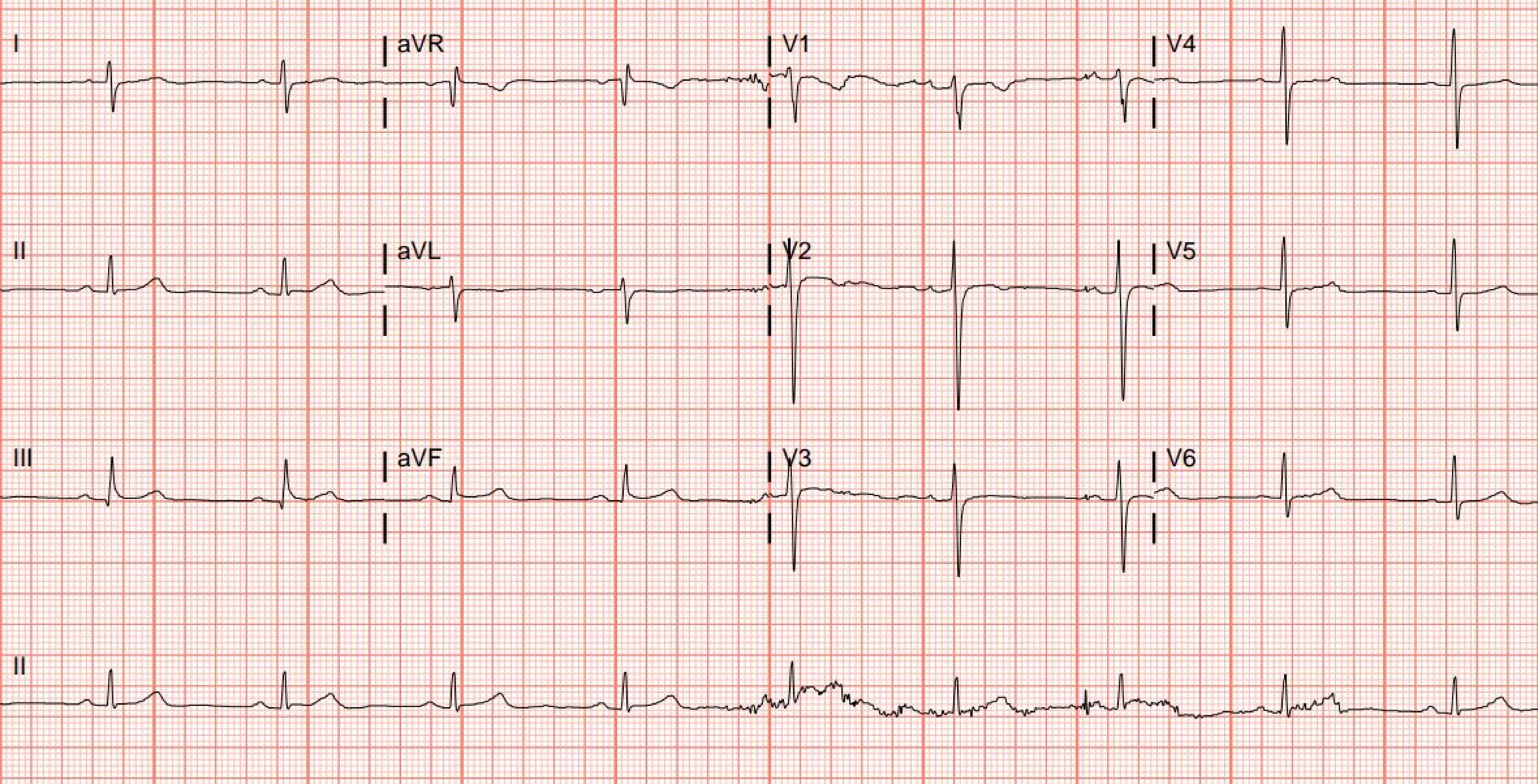
I
aVR
V1
V4
II
aVL
V2
V5
III
aVF
V3
V6
II

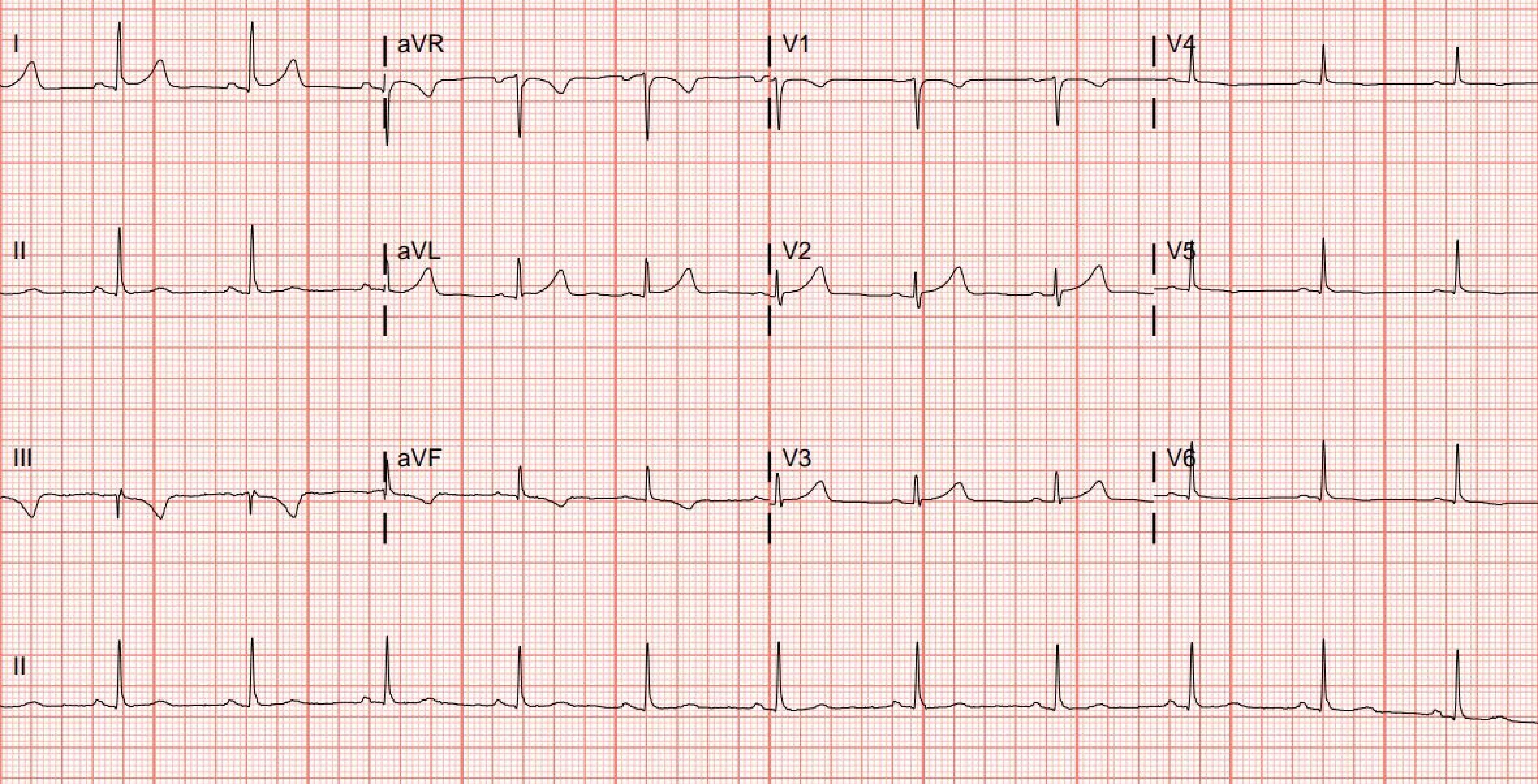
I
aVR
V1
V4
II
aVL
V2
V5
III
aVF
V3
V6
II

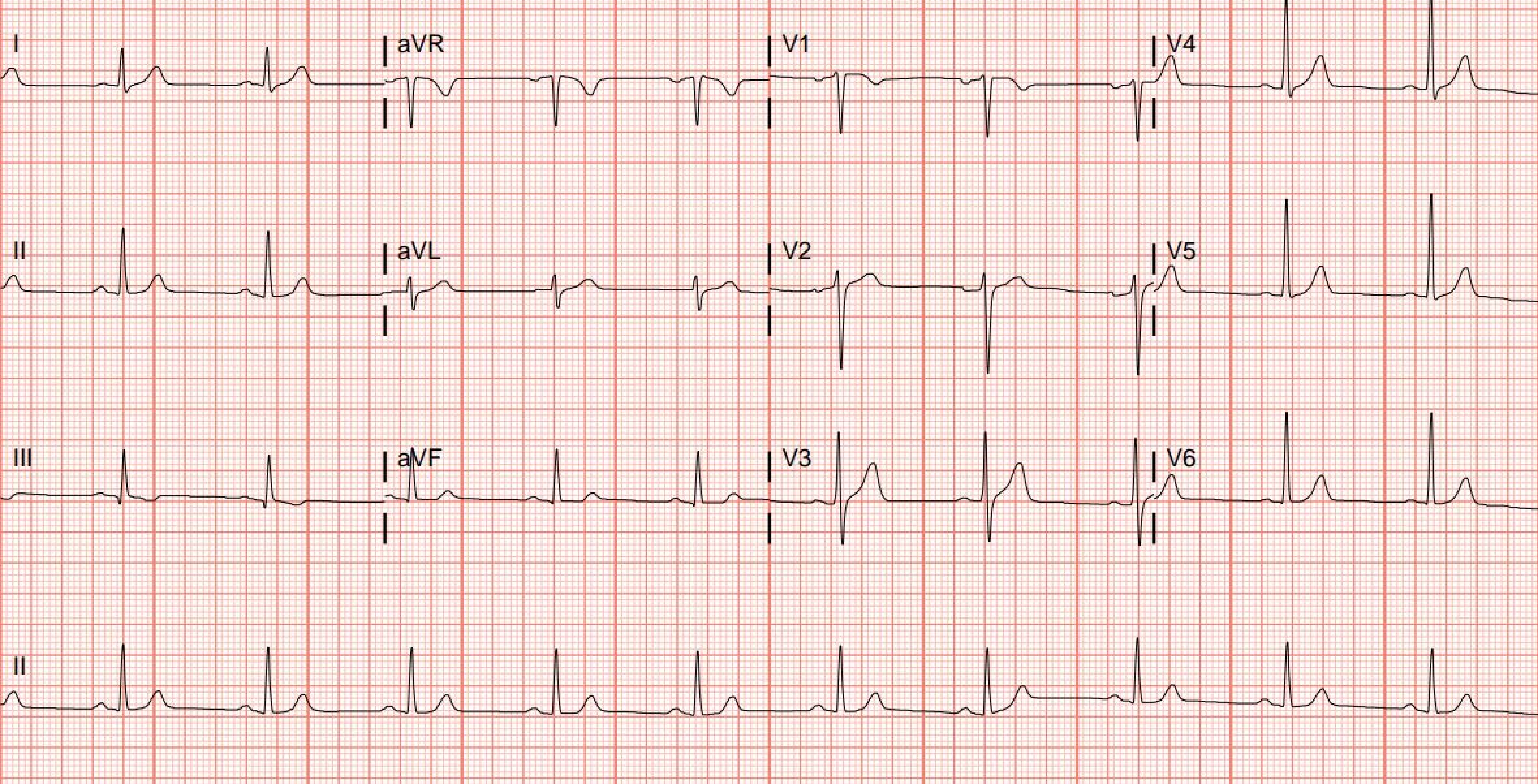
I
aVR
V1
V4
II
aVL
V2
V5
III
aVF
V3
V6
II

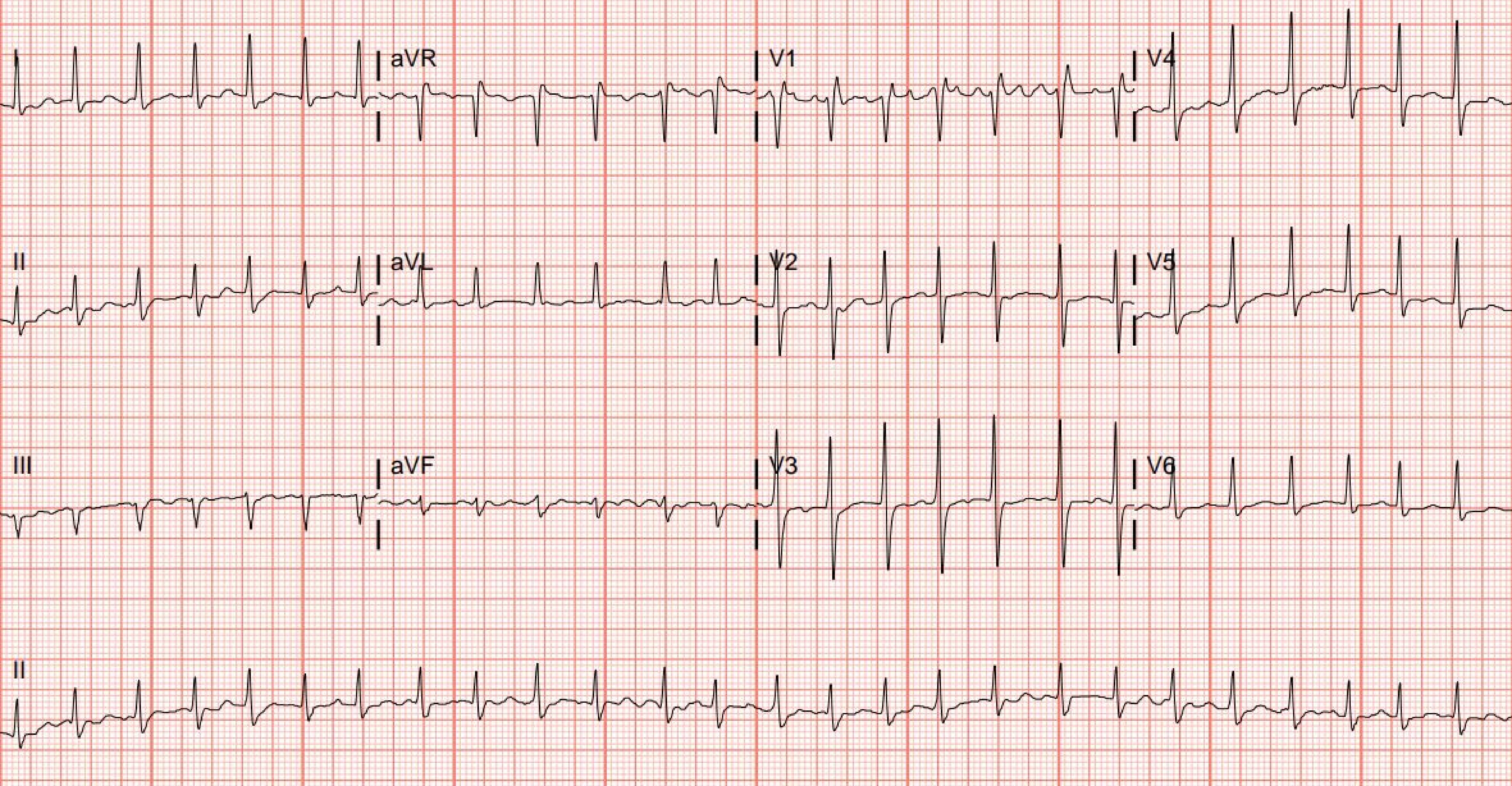

I
aVR
V1
V4
II
aVL
V2
V5
III
aVF
V3
V6
II

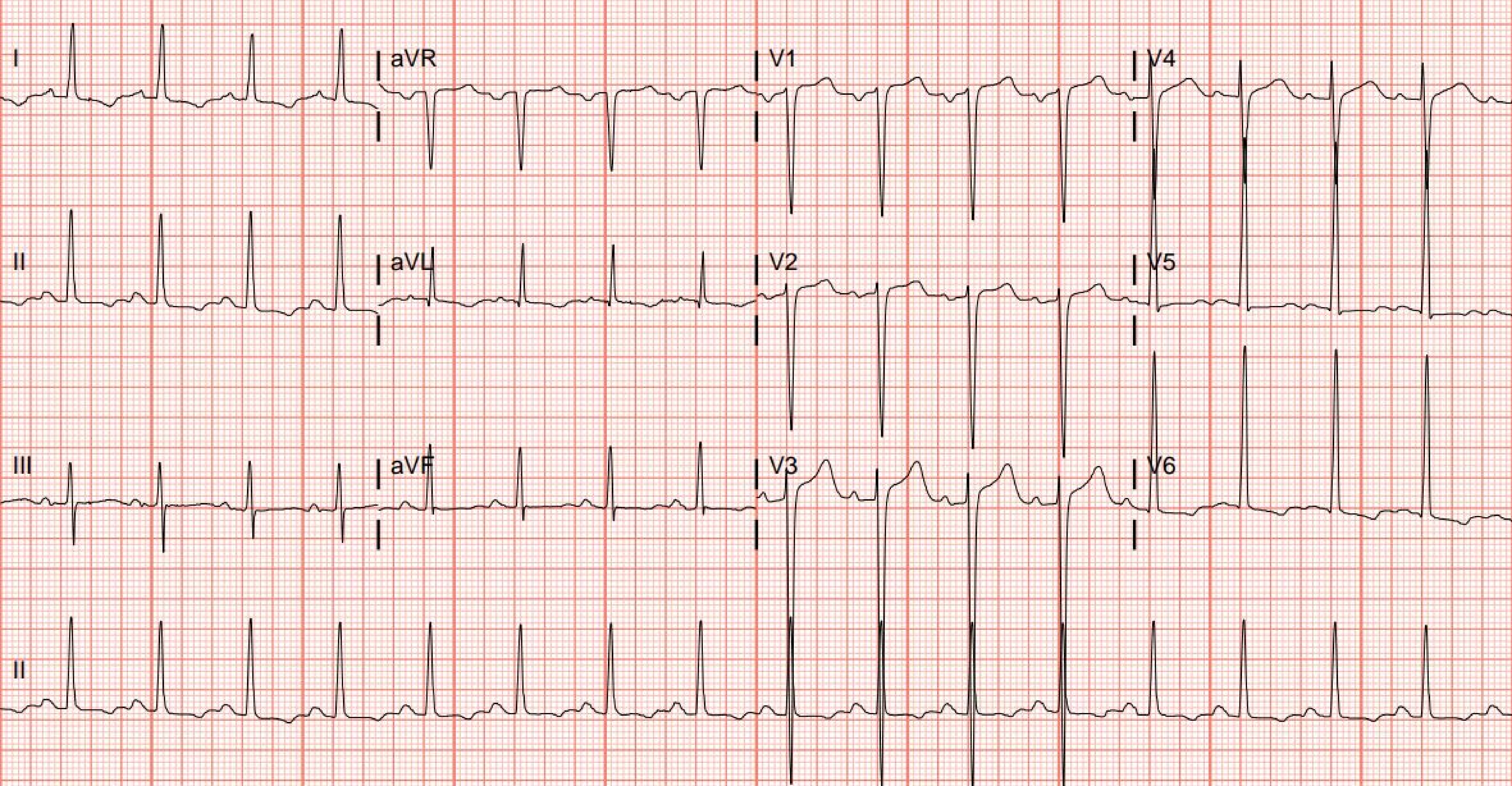

I
aVR
V1
V4
II
aVL
V2
V5
III
aVF
V3
V6
II

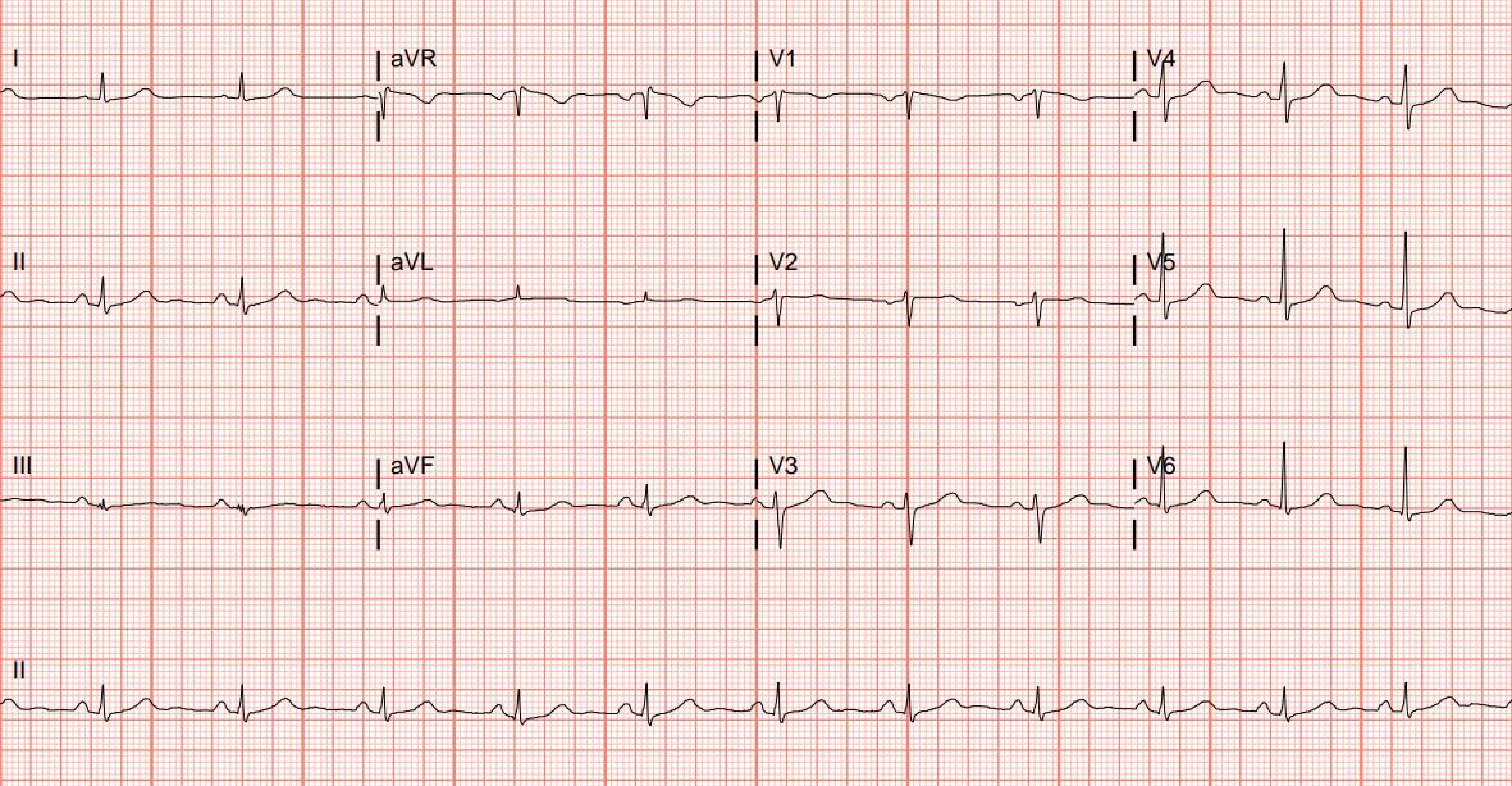
I
aVR
V1
V4
II
aVL
V2
V5
III
aVF
V3
V6
II

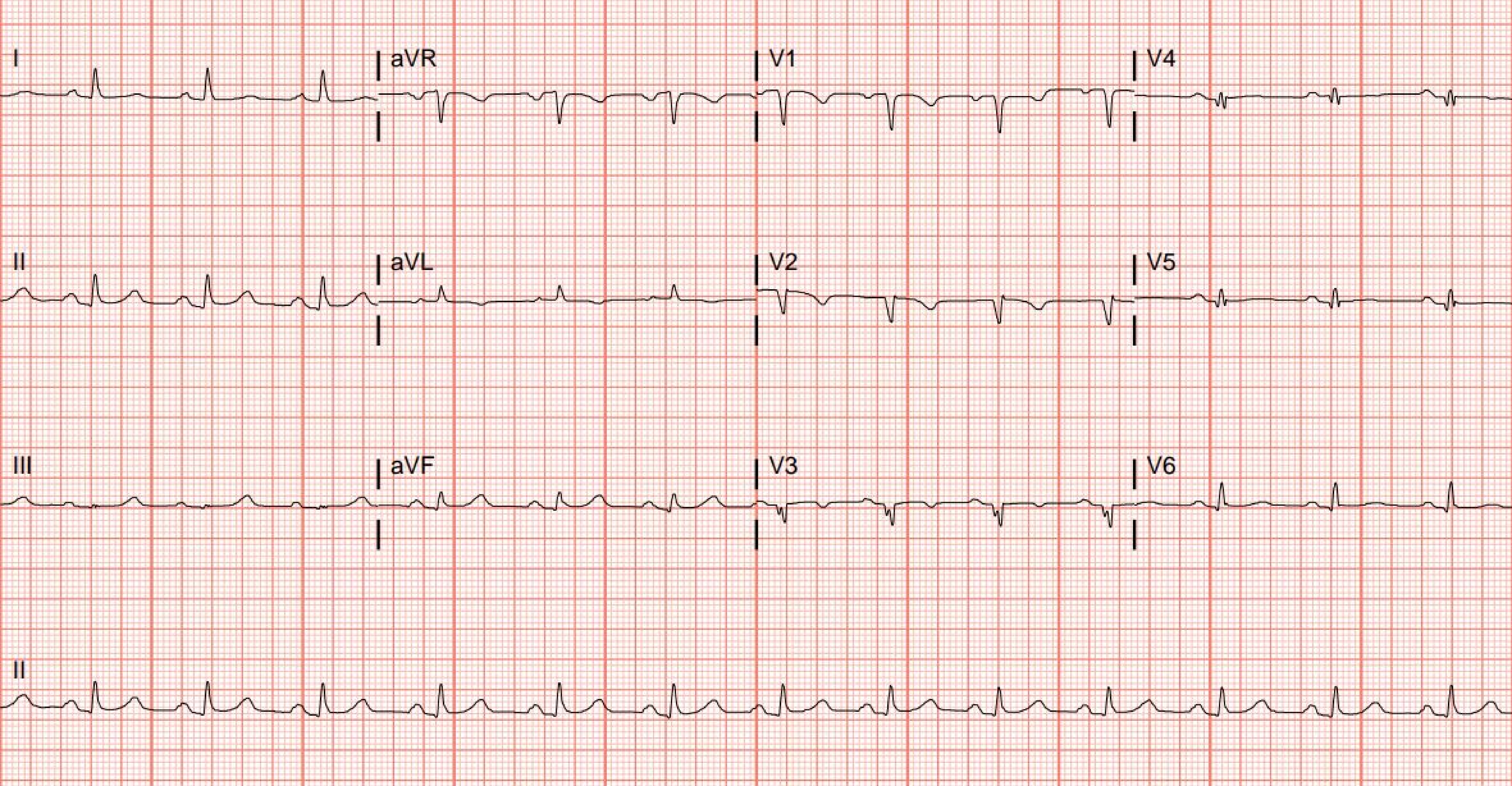
I
aVR
V1
V4
II
aVL
V2
V5
III
aVF
V3
V6
II

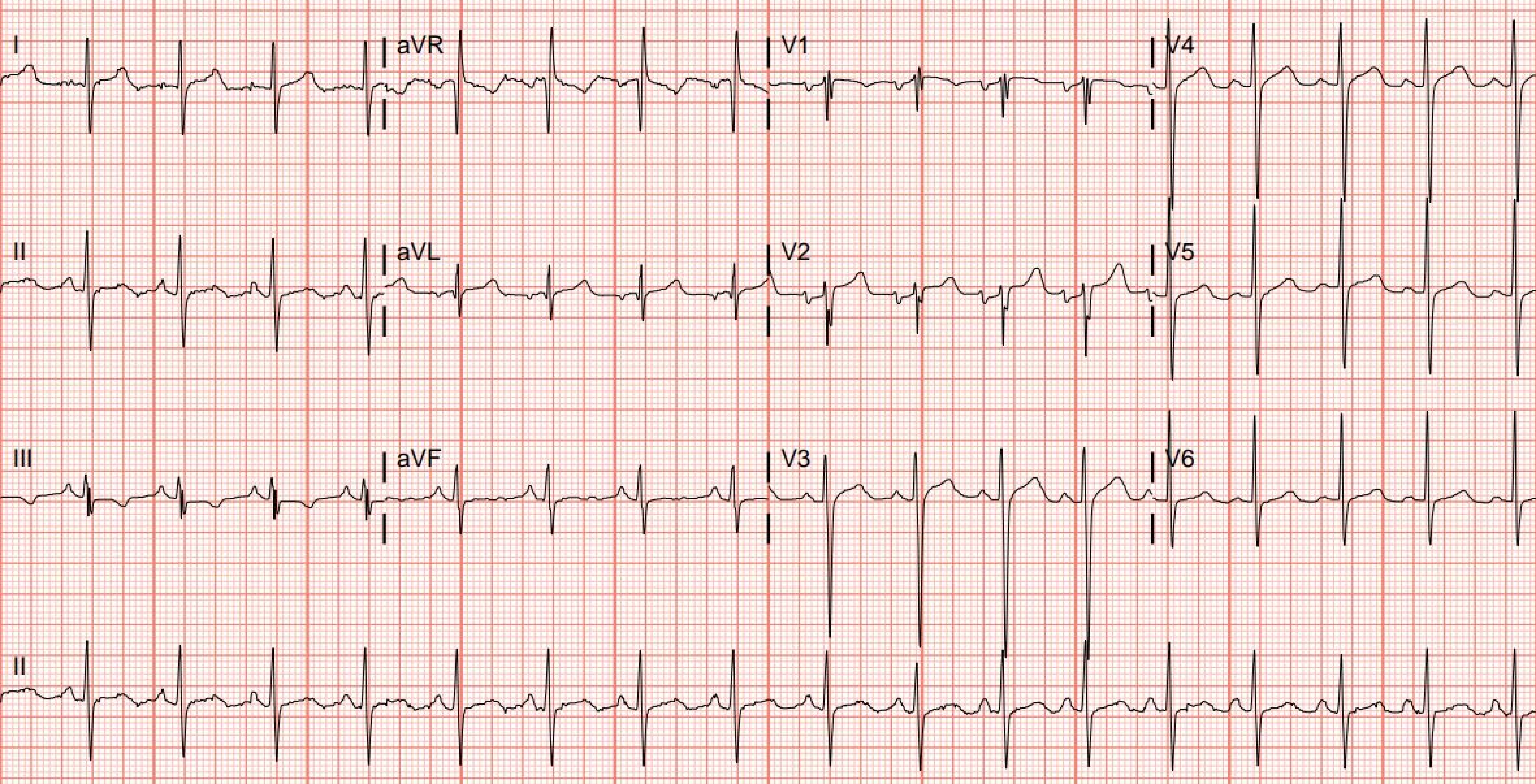
I
aVR
V1
V4
II
aVL
V2
V5
III
aVF
V3
V6
II

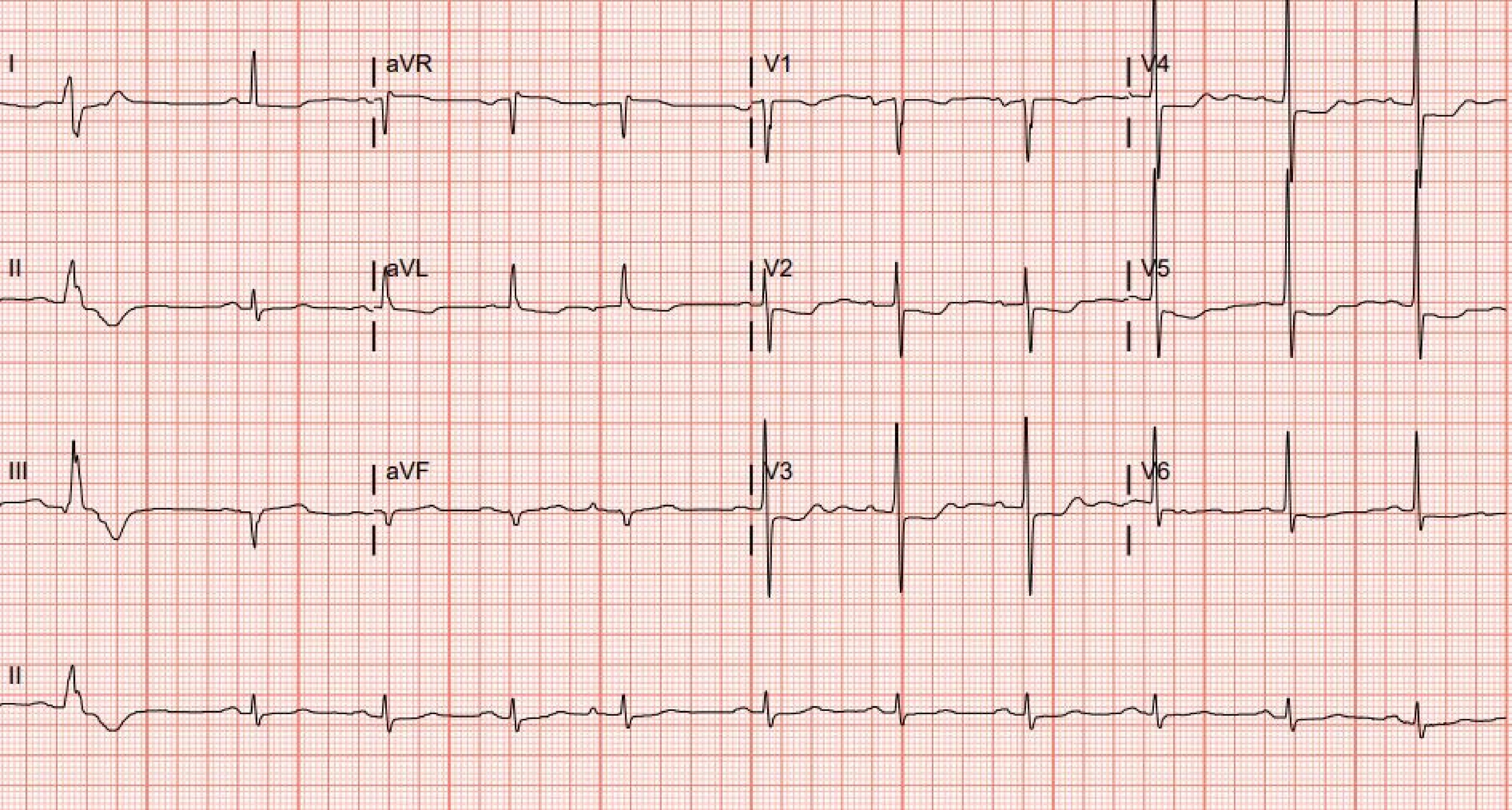
I
aVR
V1
V4
II
aVL
V2
V5
III
aVF
V3
V6
II

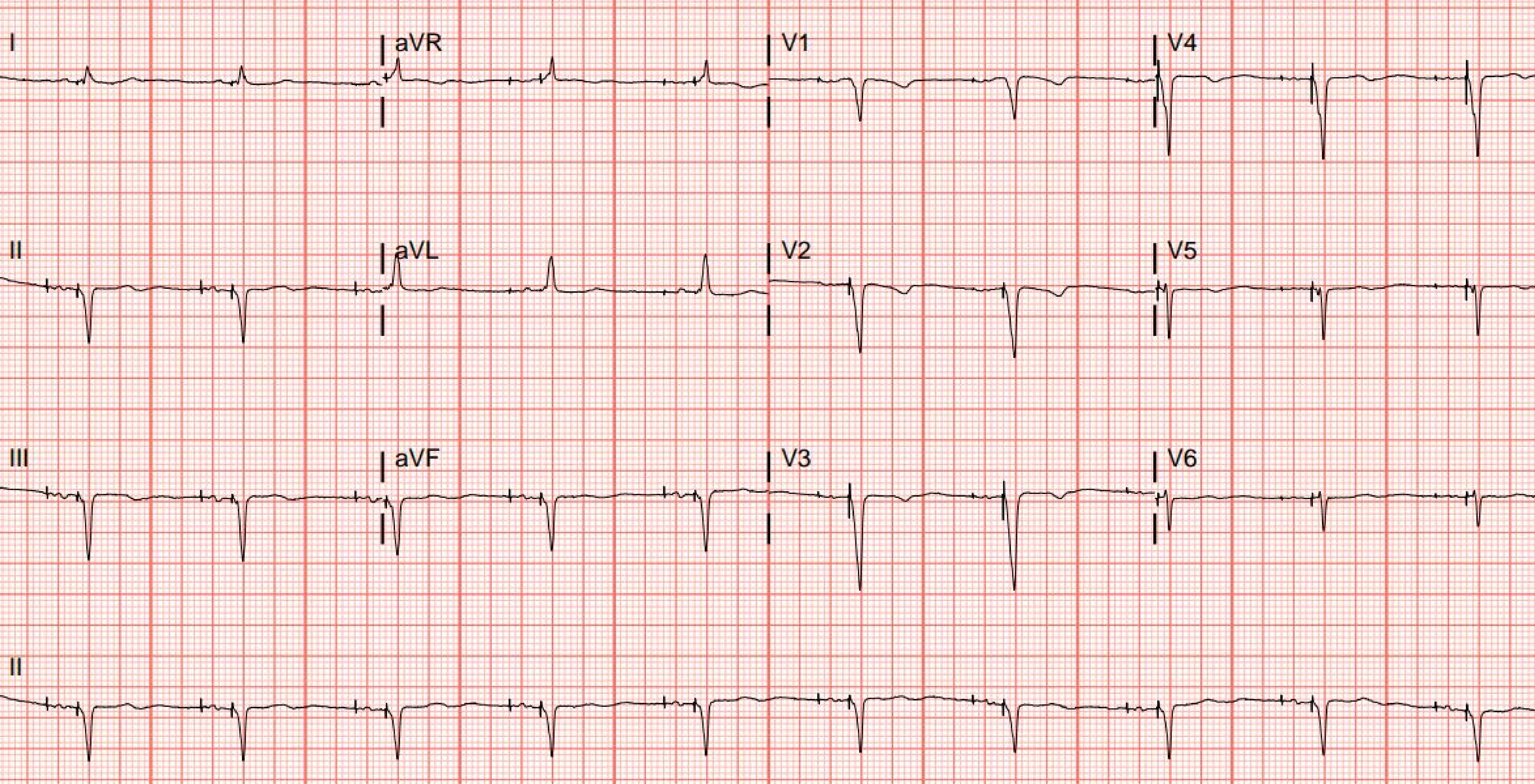
I
aVR
V1
V4
II
aVL
V2
V5
III
aVF
V3
V6
II

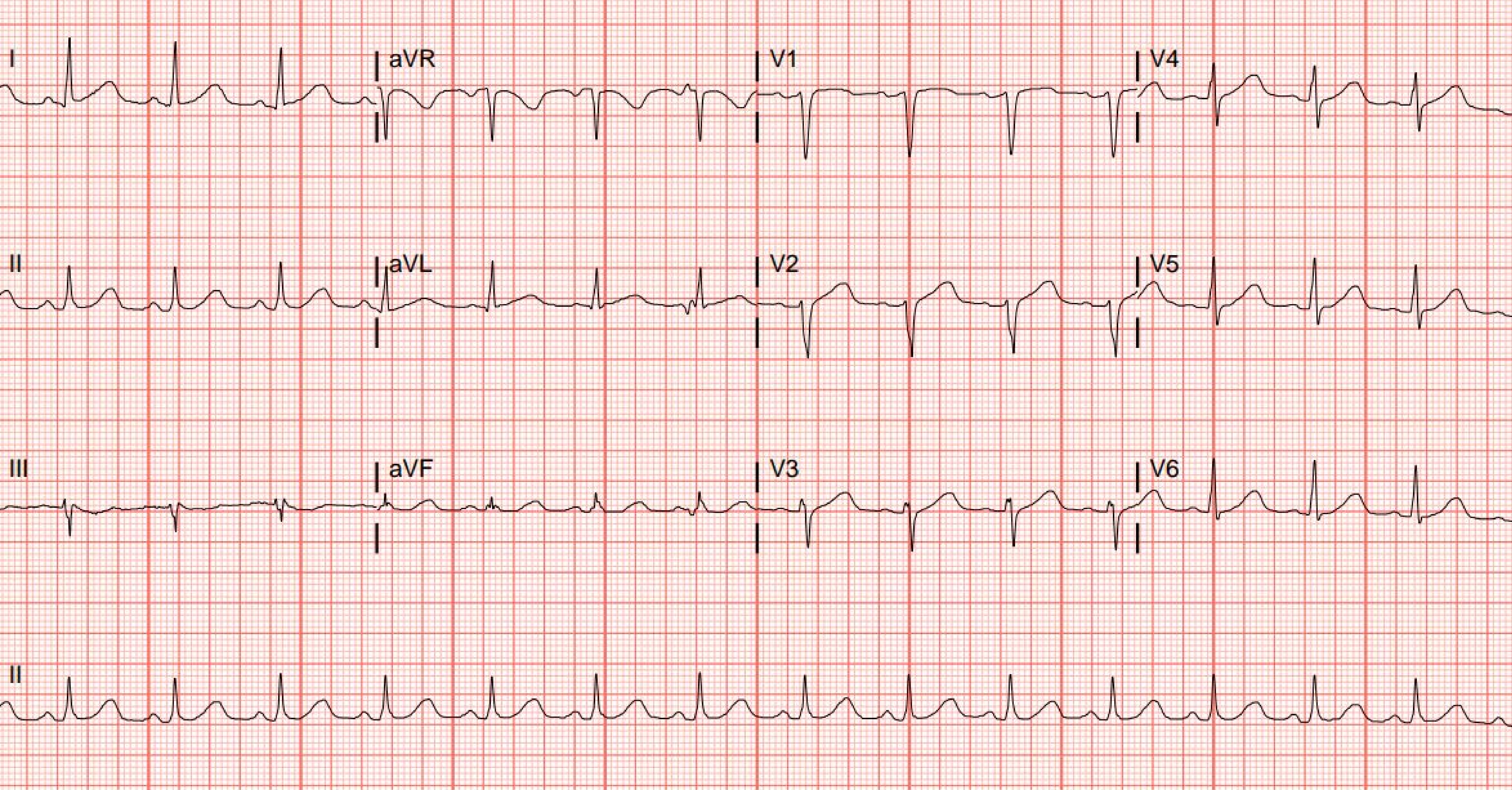
I
aVR
V1
V4
II
aVL
V2
V5
III
aVF
V3
V6
II

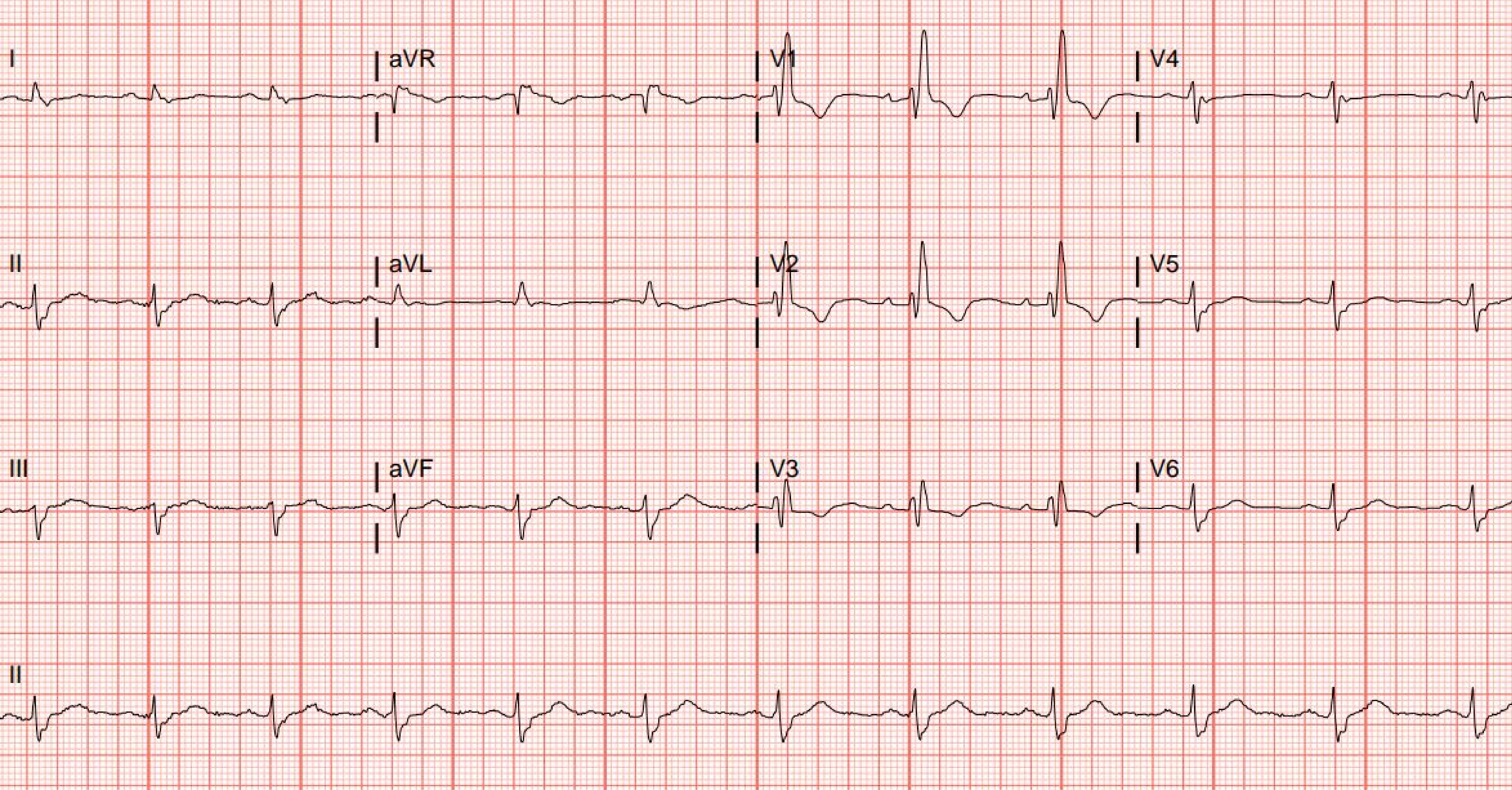
I
aVR
V1
V4
II
aVL
V2
V5
III
aVF
V3
V6
II

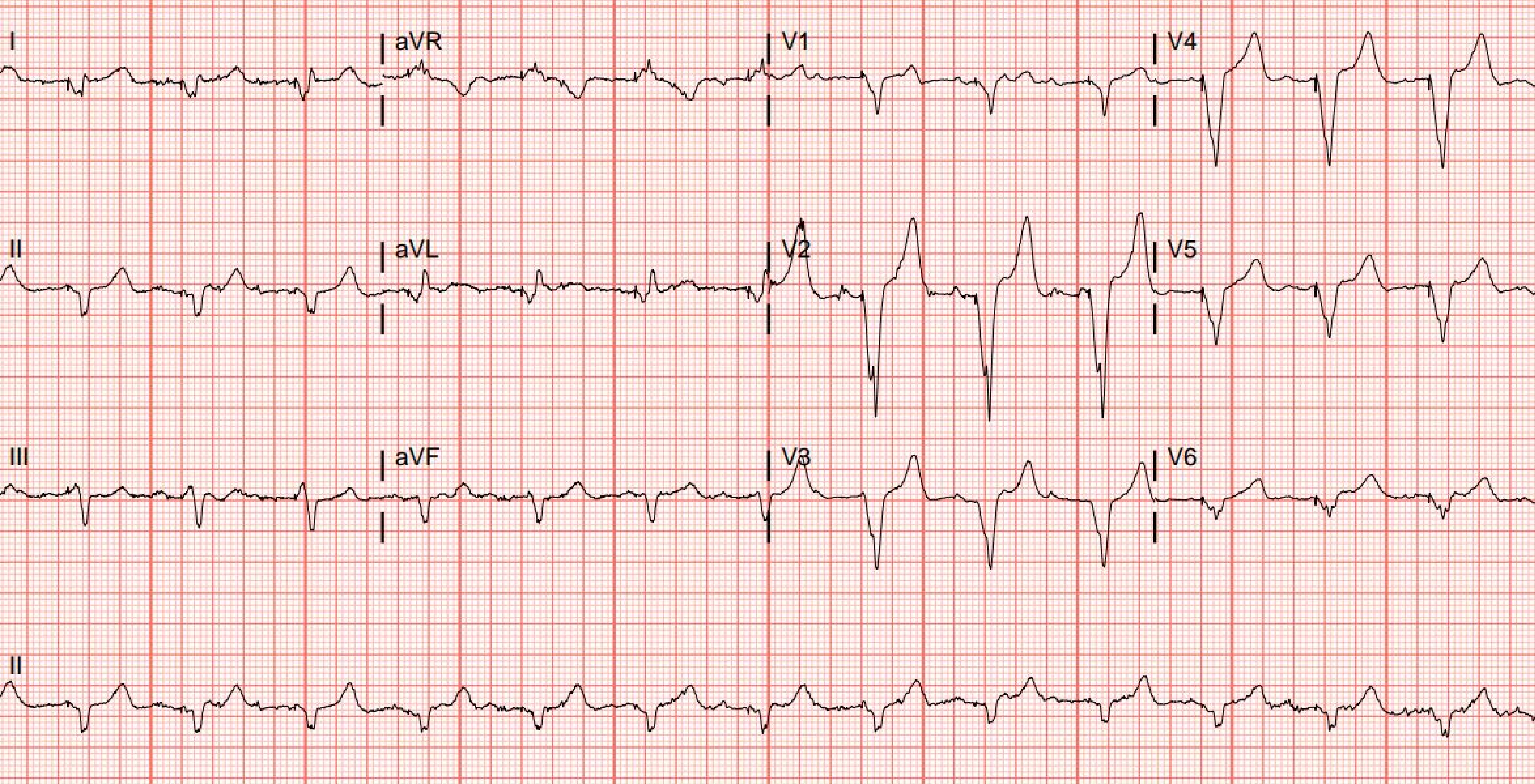
I
aVR
V1
V4
II
aVL
V2
V5
III
aVF
V3
V6
II

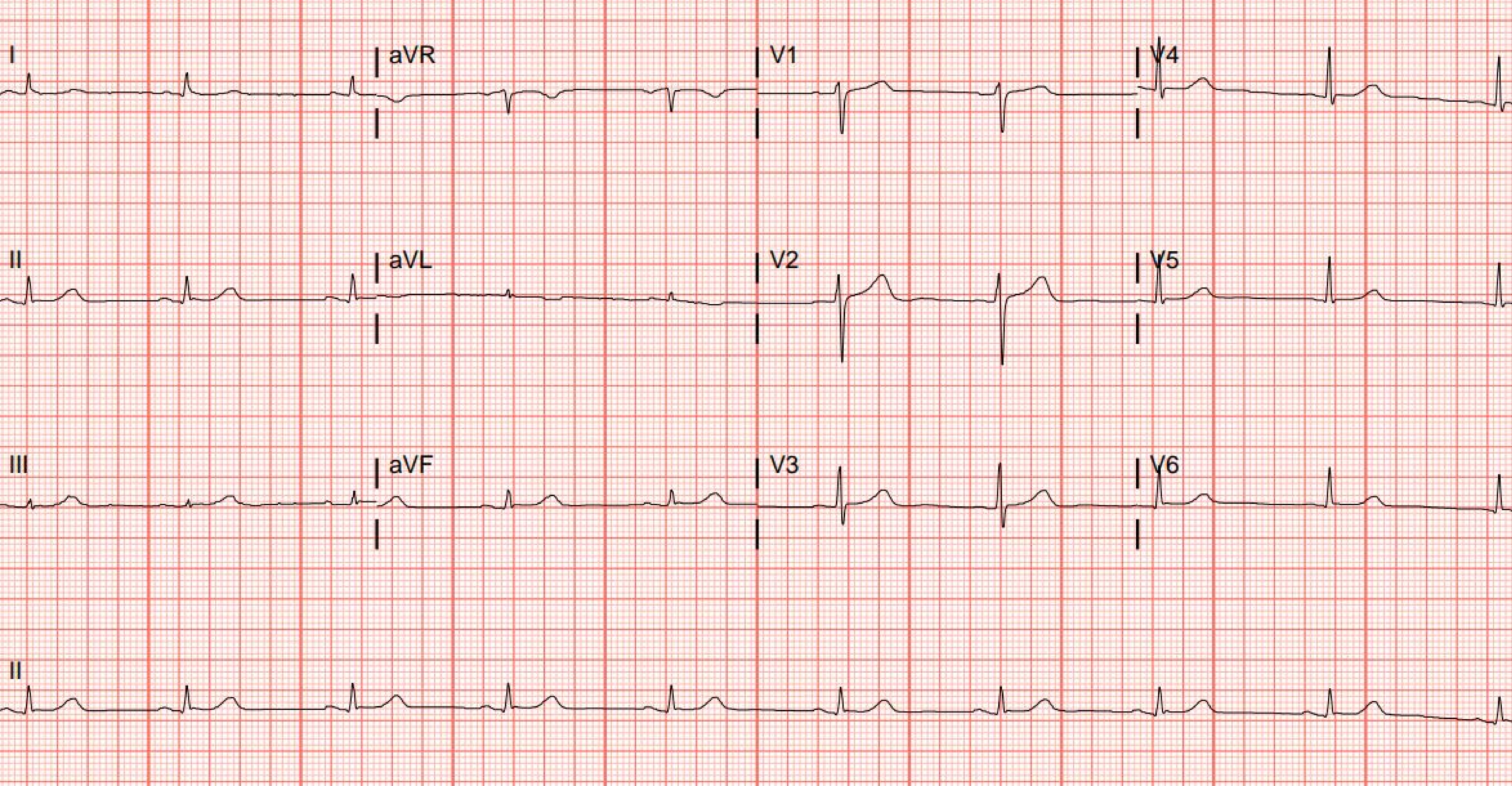
I
aVR
V1
V4
II
aVL
V2
V5
III
aVF
V3
V6
II

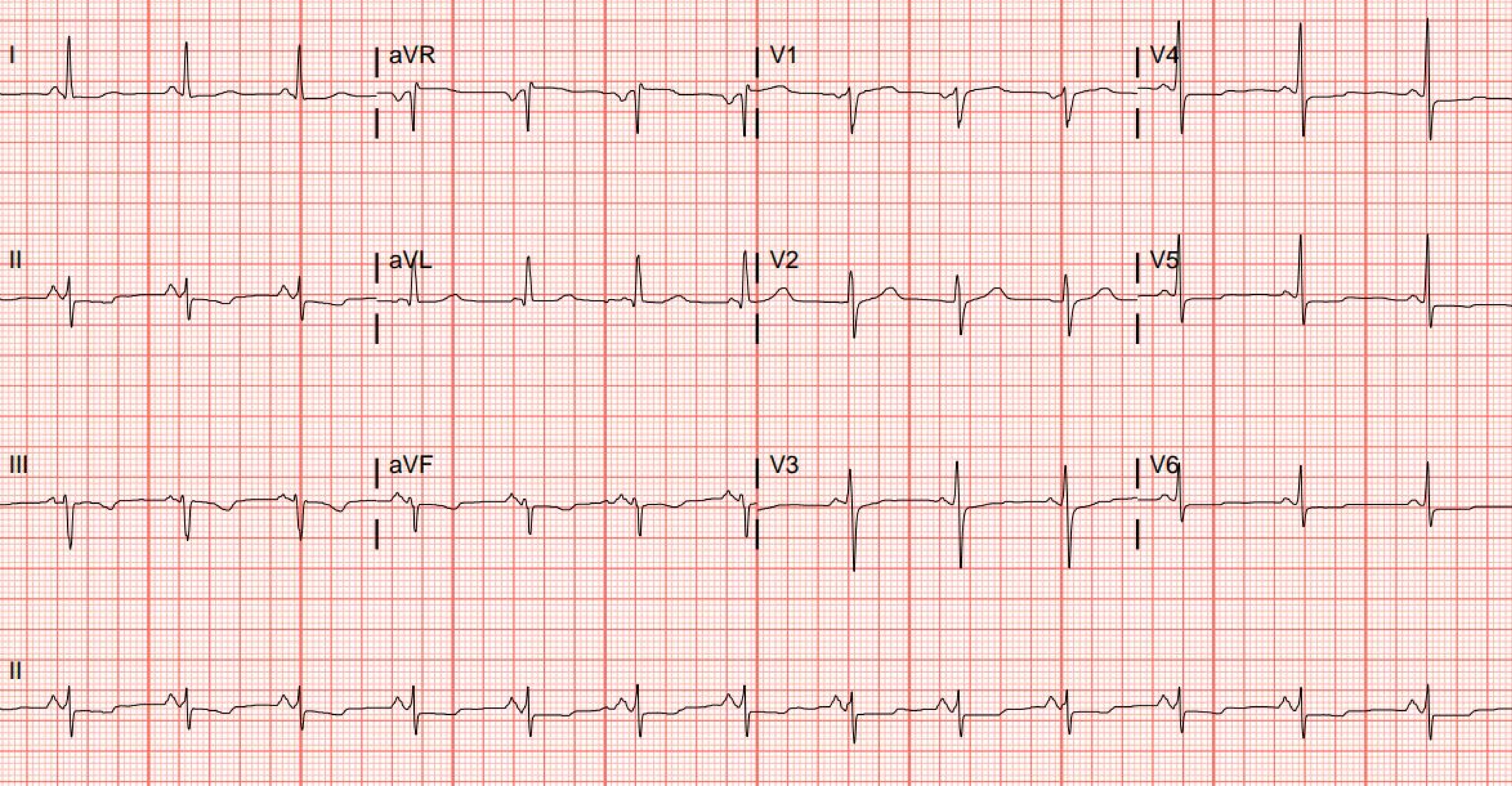
I
aVR
V1
V4
II
aVL
V2
V5
III
aVF
V3
V6
II

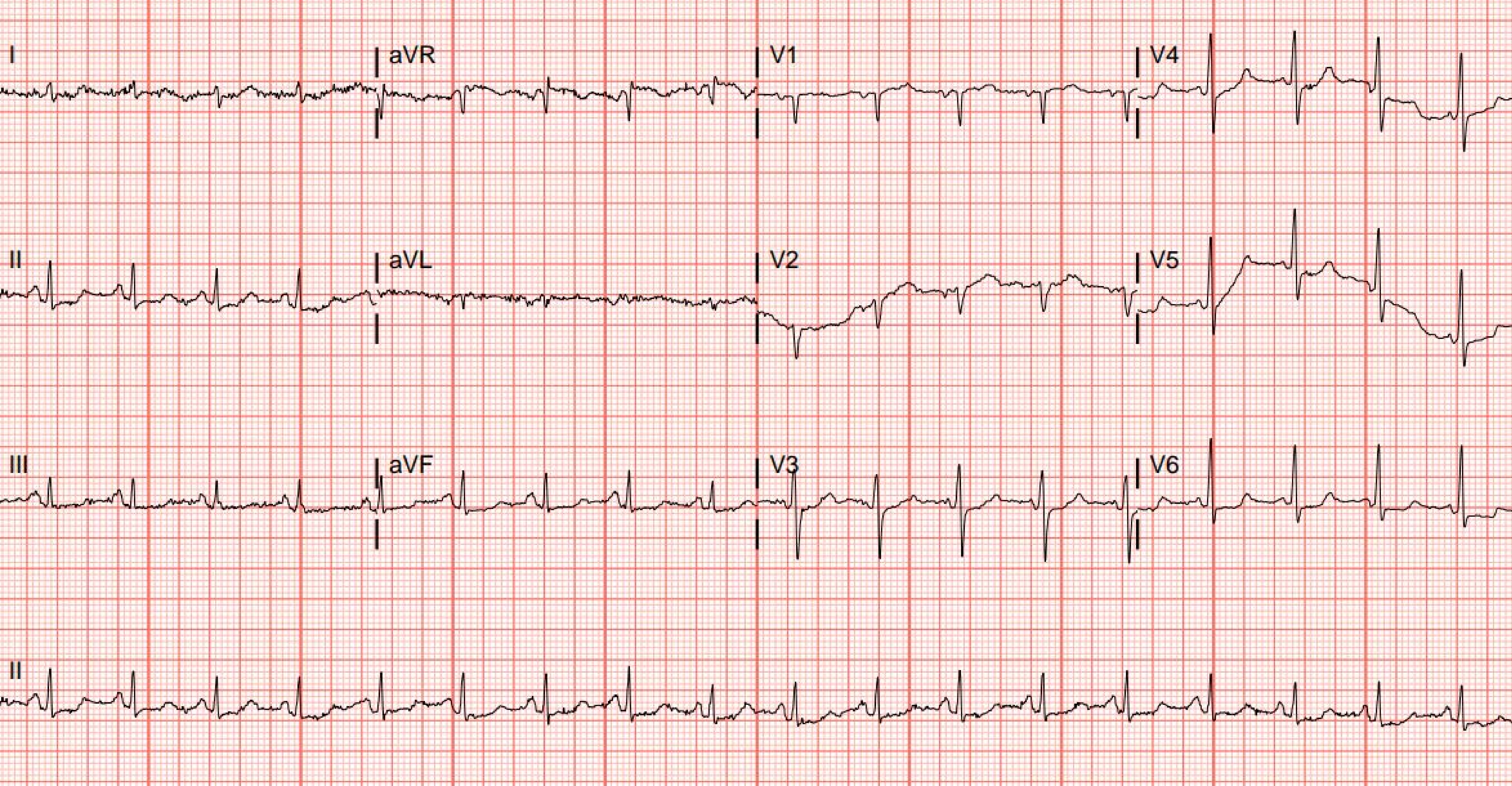
I
aVR
V1
V4
II
aVL
V2
V5
III
aVF
V3
V6
II

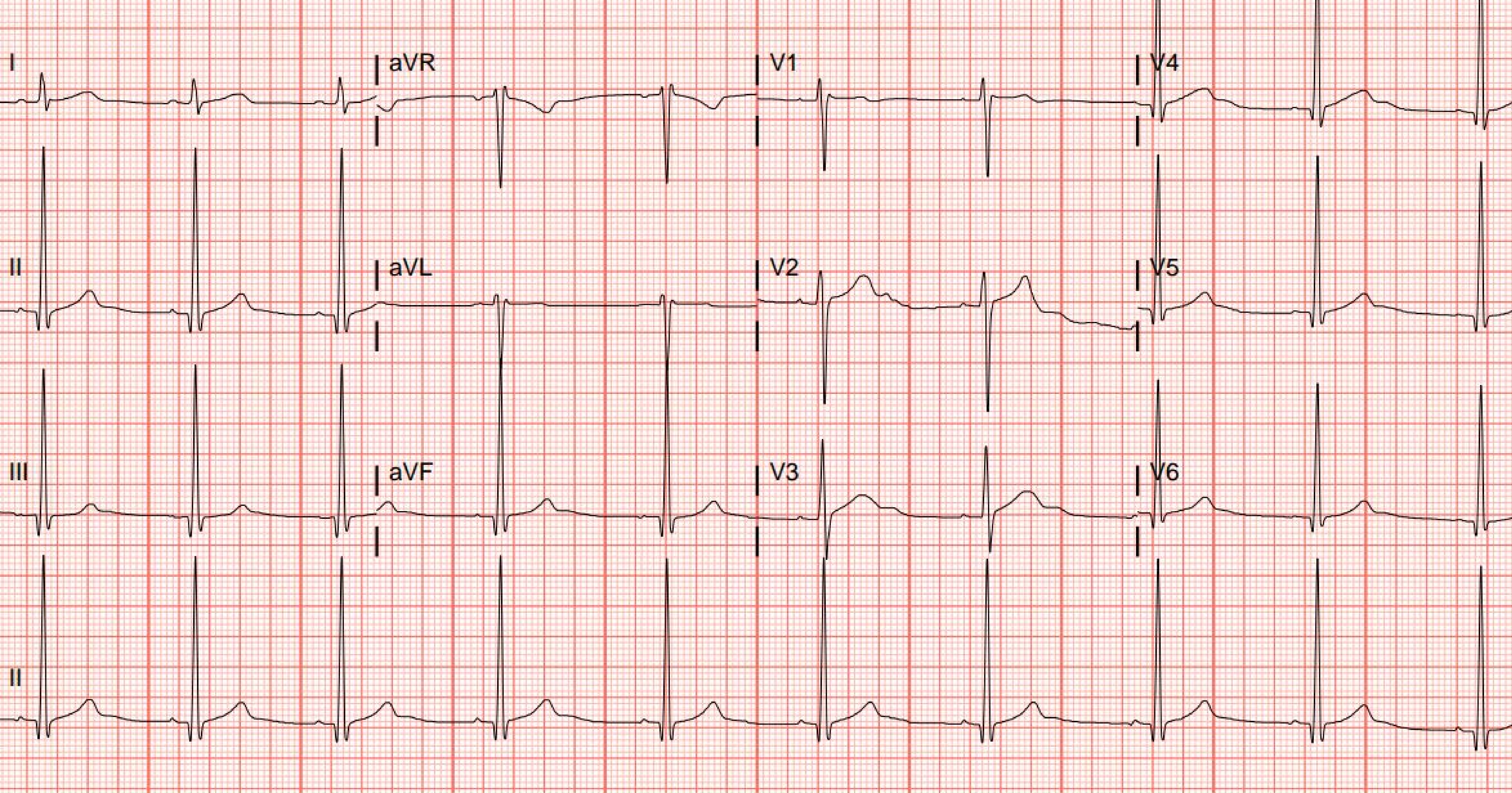
I
aVR
V1
V4
II
aVL
V2
V5
III
aVF
V3
V6
II

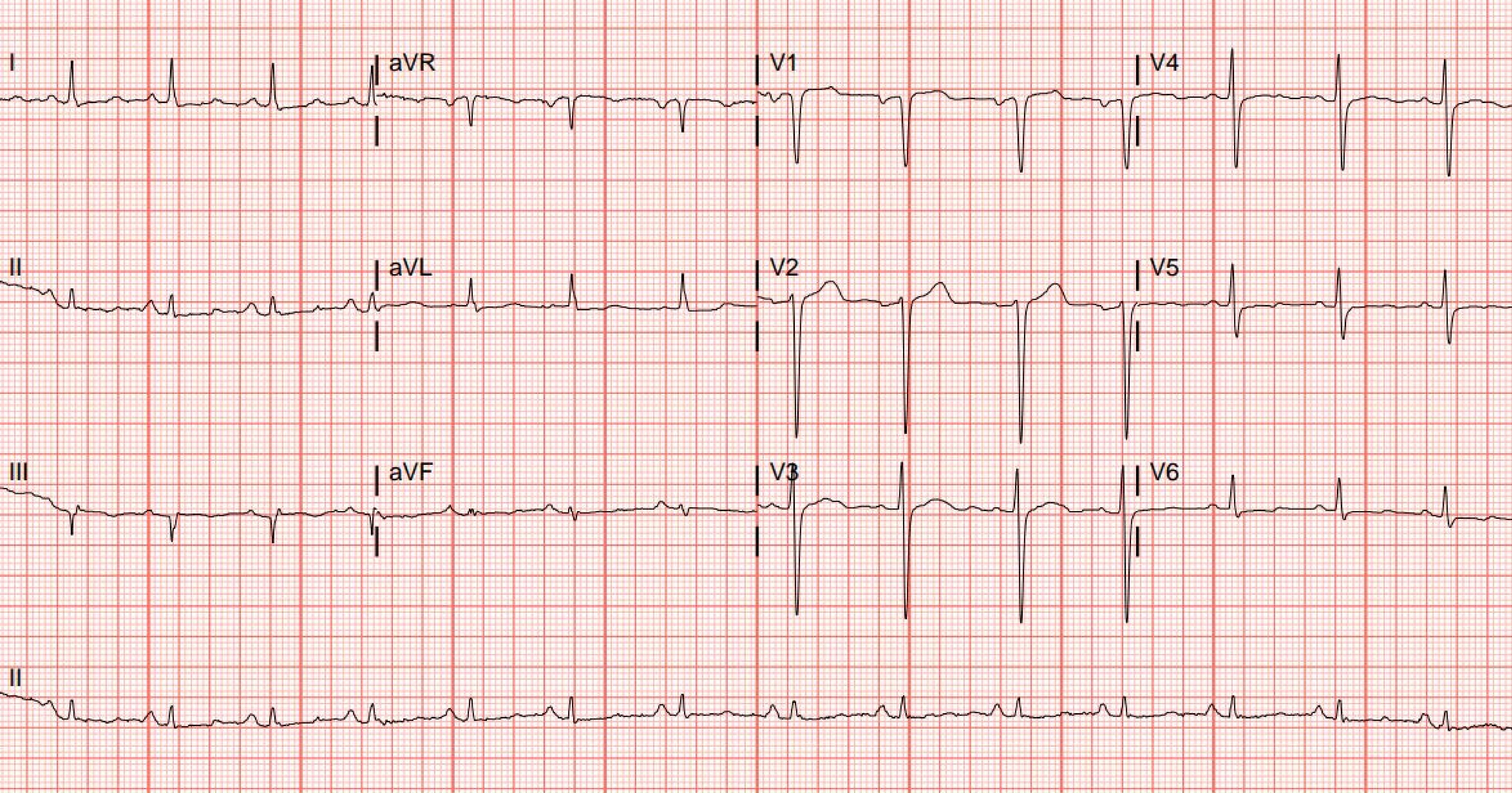
I
aVR
V1
V4
II
aVL
V2
V5
III
aVF
V3
V6
II

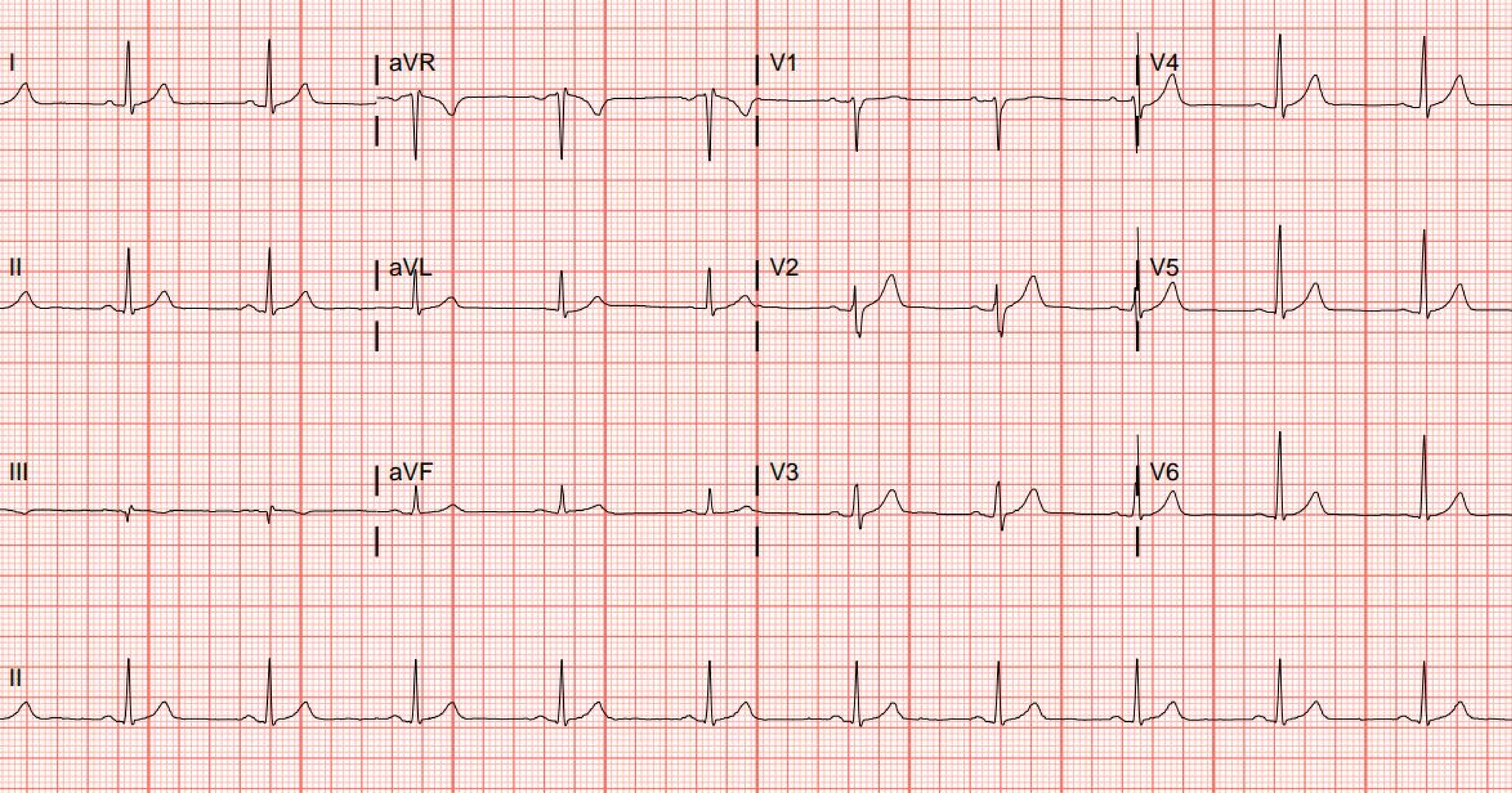

I
aVR
V1
V4
II
aVL
V2
V5
III
aVF
V3
V6
II

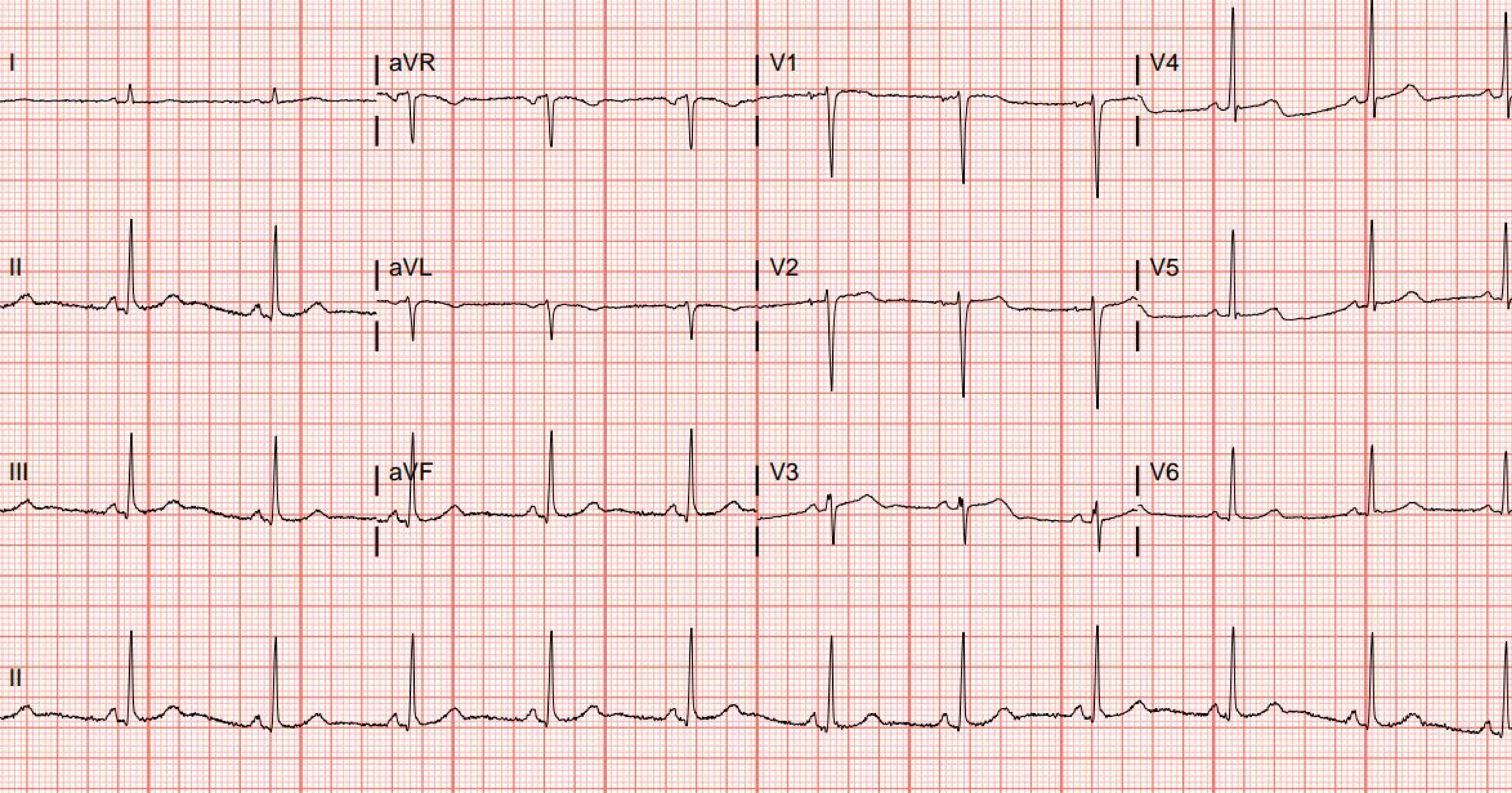
I
aVR
V1
V4
II
aVL
V2
V5
III
aVF
V3
V6
II

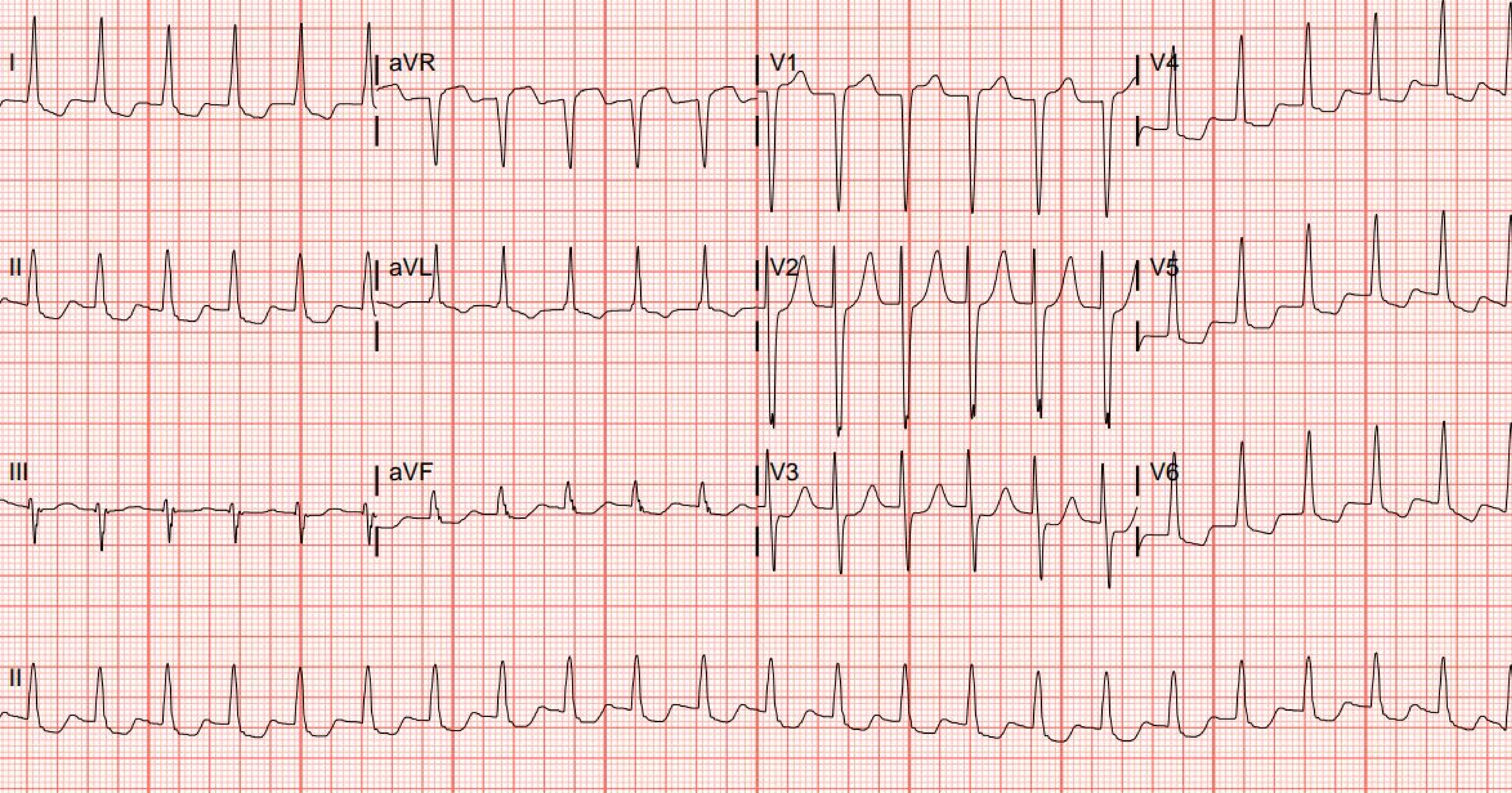
I
aVR
V1
V4
II
aVL
V2
V5
III
aVF
V3
V6
II

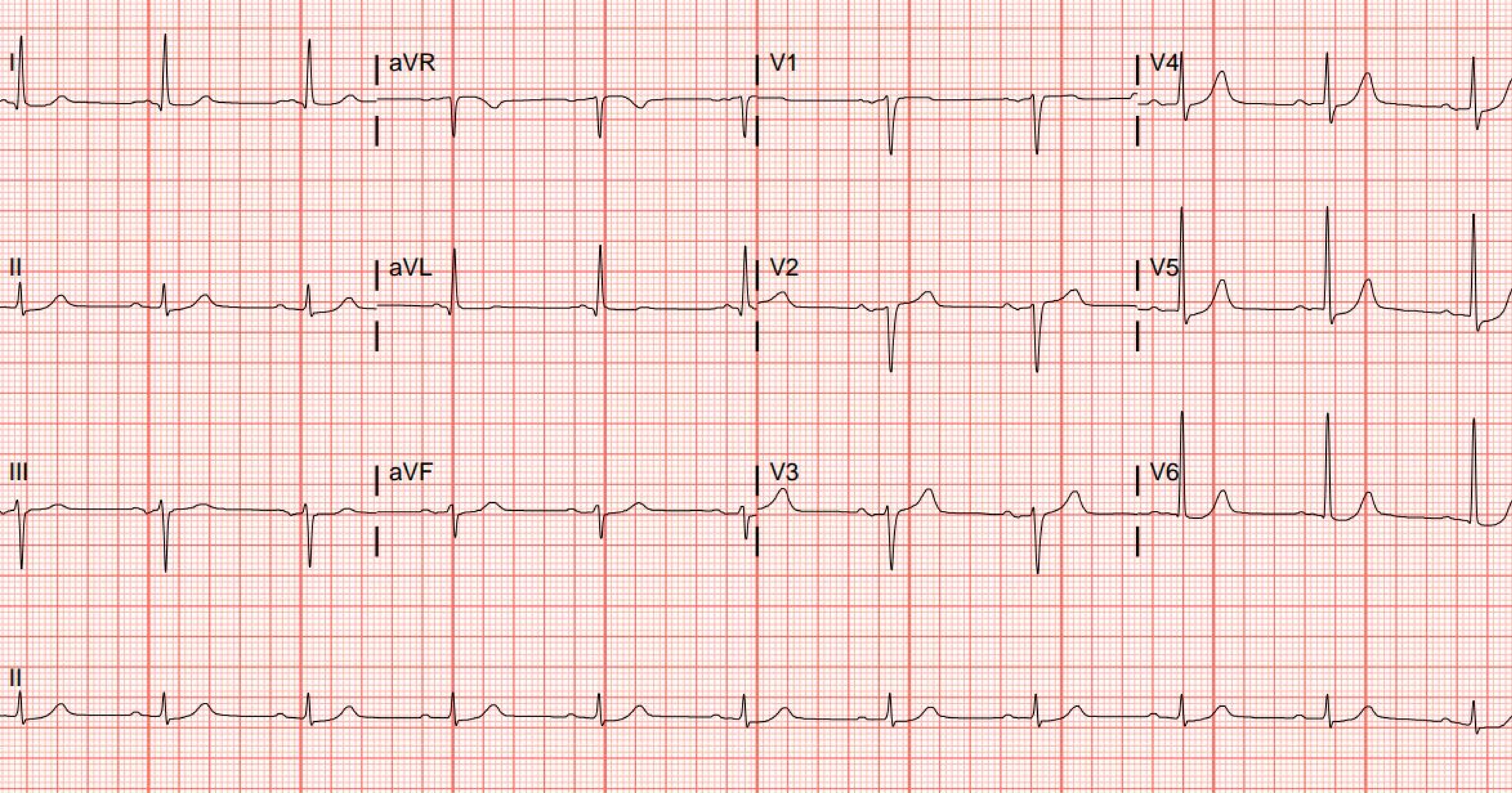
I
aVR
V1
V4
II
aVL
V2
V5
III
aVF
V3
V6
II

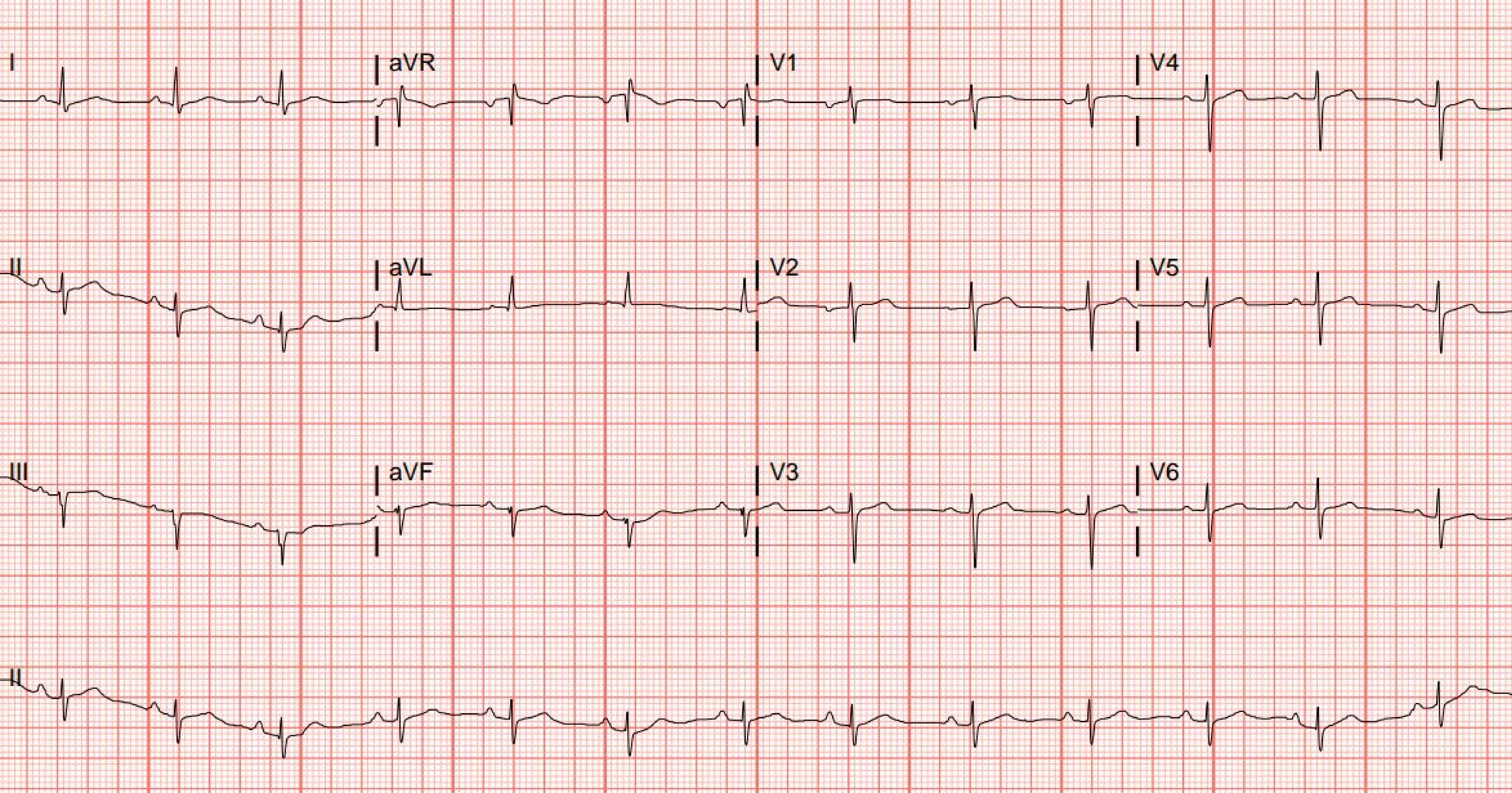
I
aVR
V1
V4
II
aVL
V2
V5
III
aVF
V3
V6
II

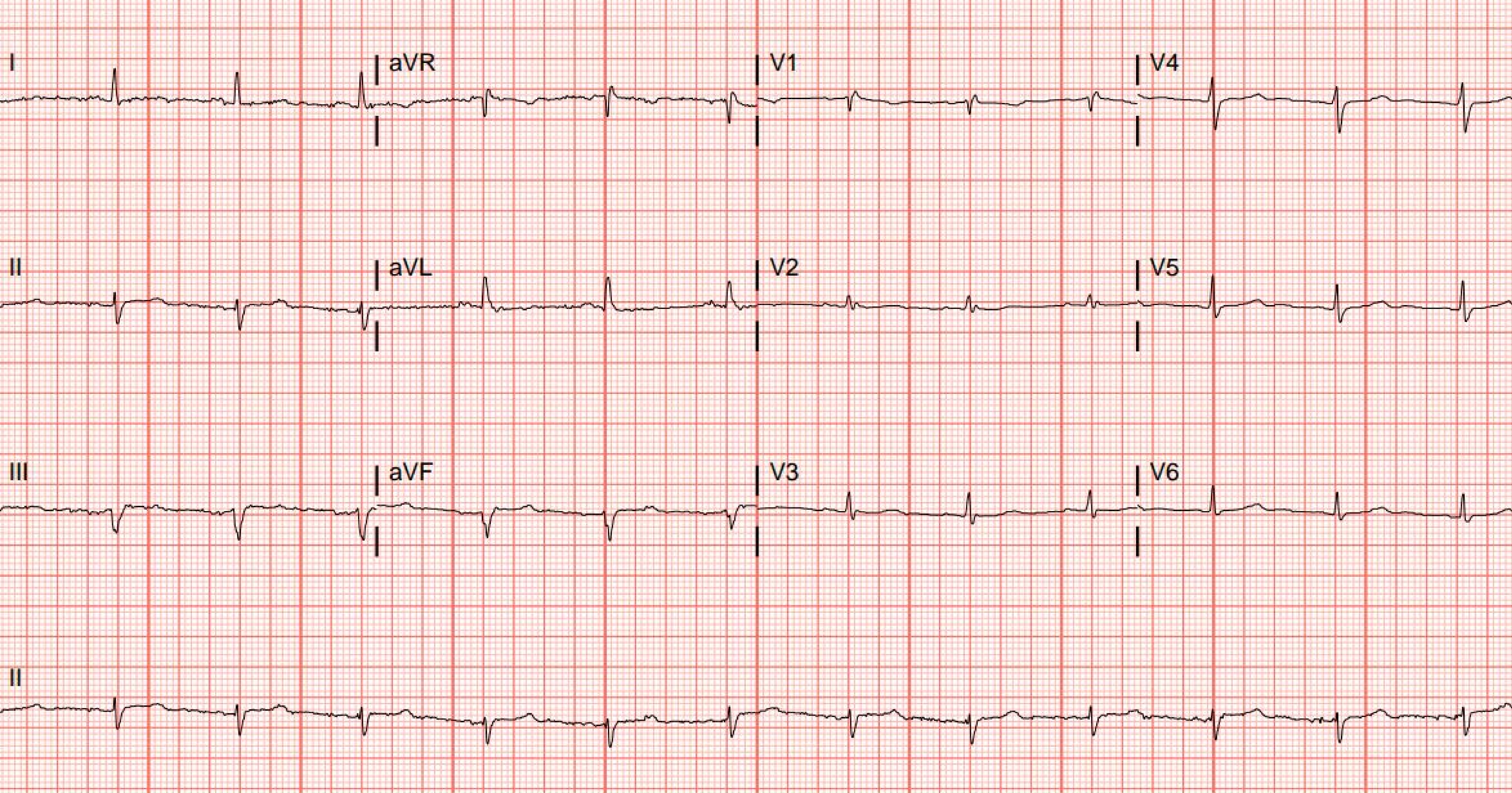
I
aVR
V1
V4
II
aVL
V2
V5
III
aVF
V3
V6
II

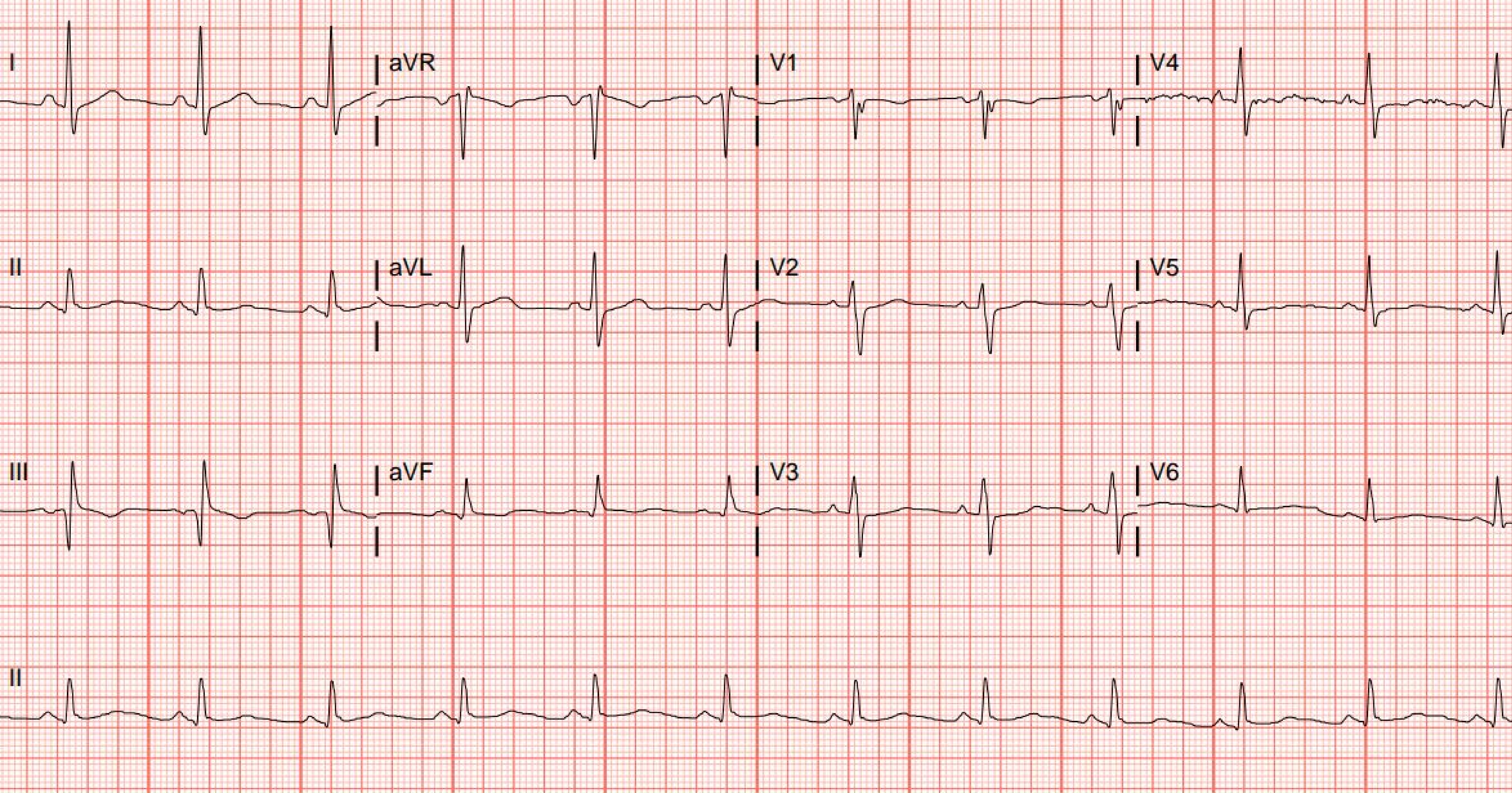
I
aVR
V1
V4
II
aVL
V2
V5
III
aVF
V3
V6
II

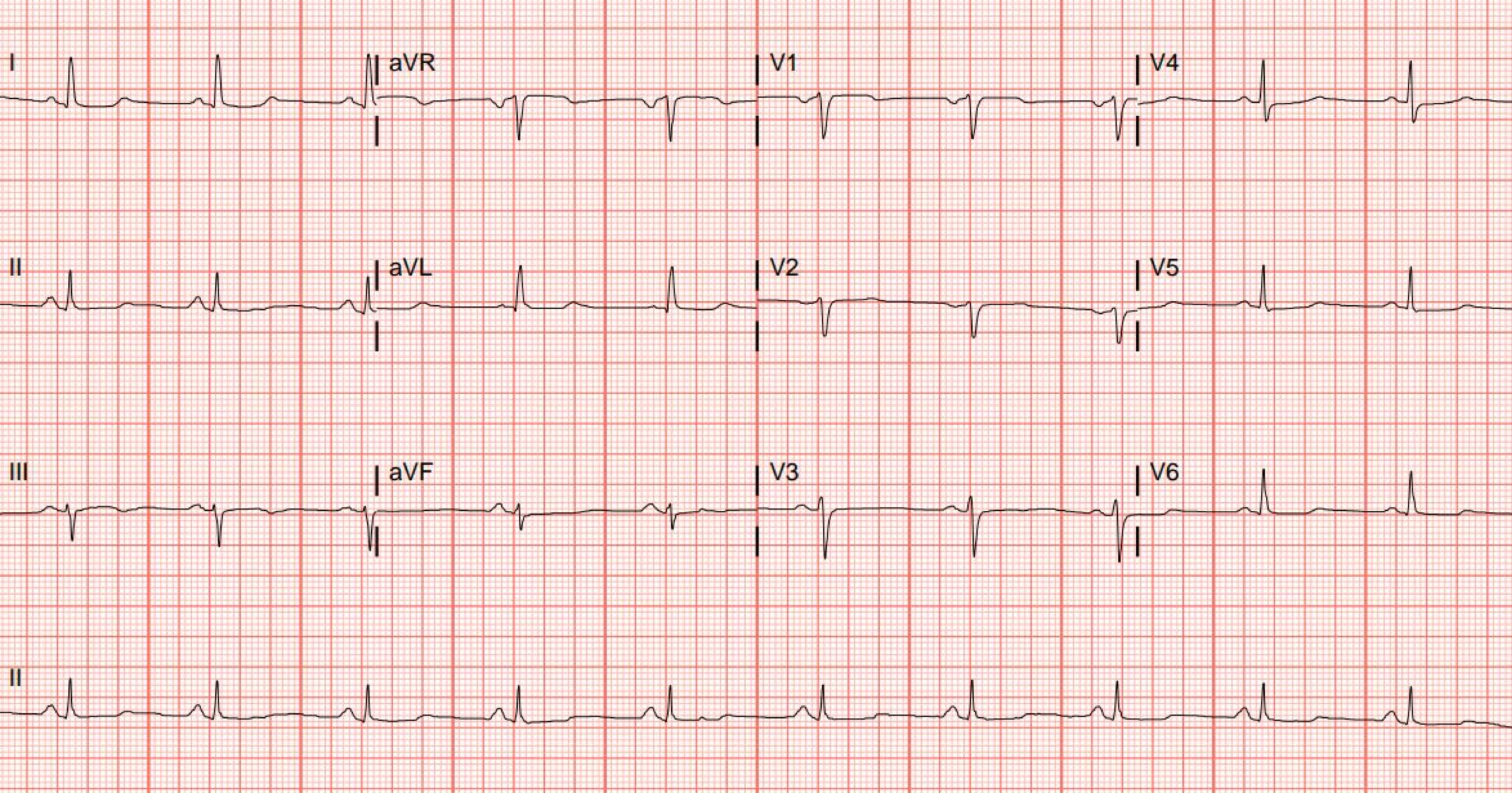
I
aVR
V1
V4
II
aVL
V2
V5
III
aVF
V3
V6
II

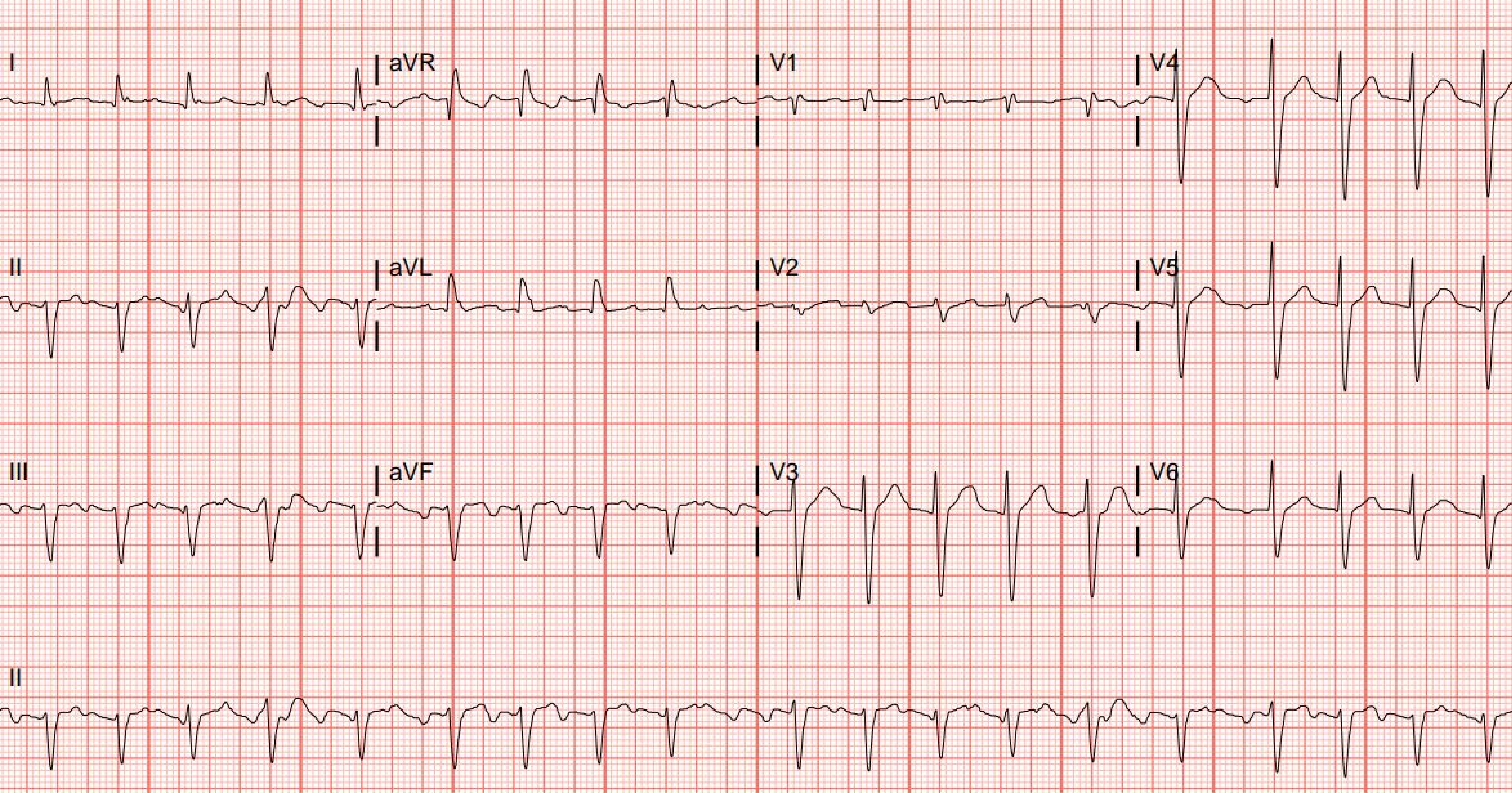
I
aVR
V1
V4
II
aVL
V2
V5
III
aVF
V3
V6
II

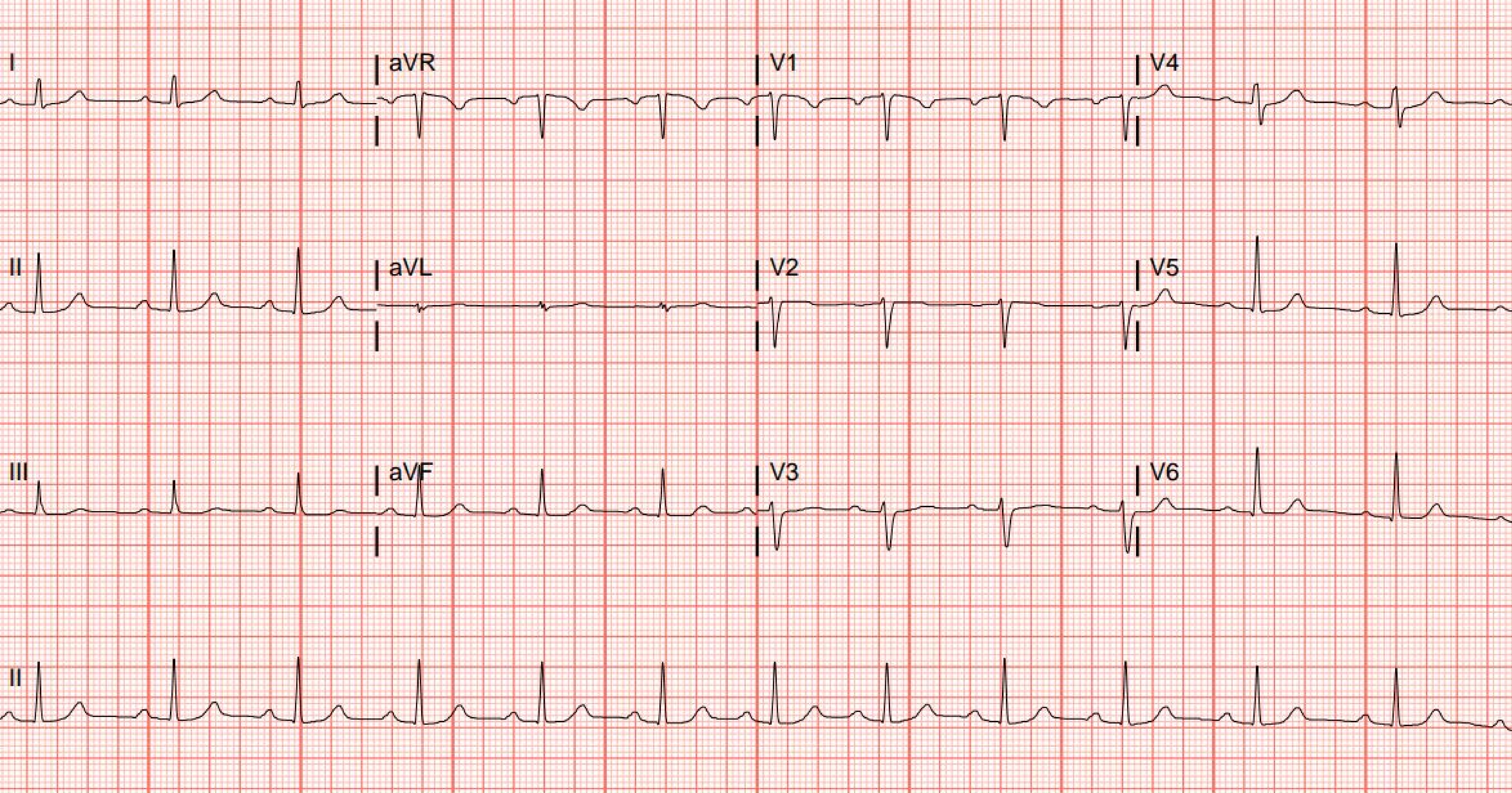
I
aVR
V1
V4
II
aVL
V2
V5
III
aVF
V3
V6
II

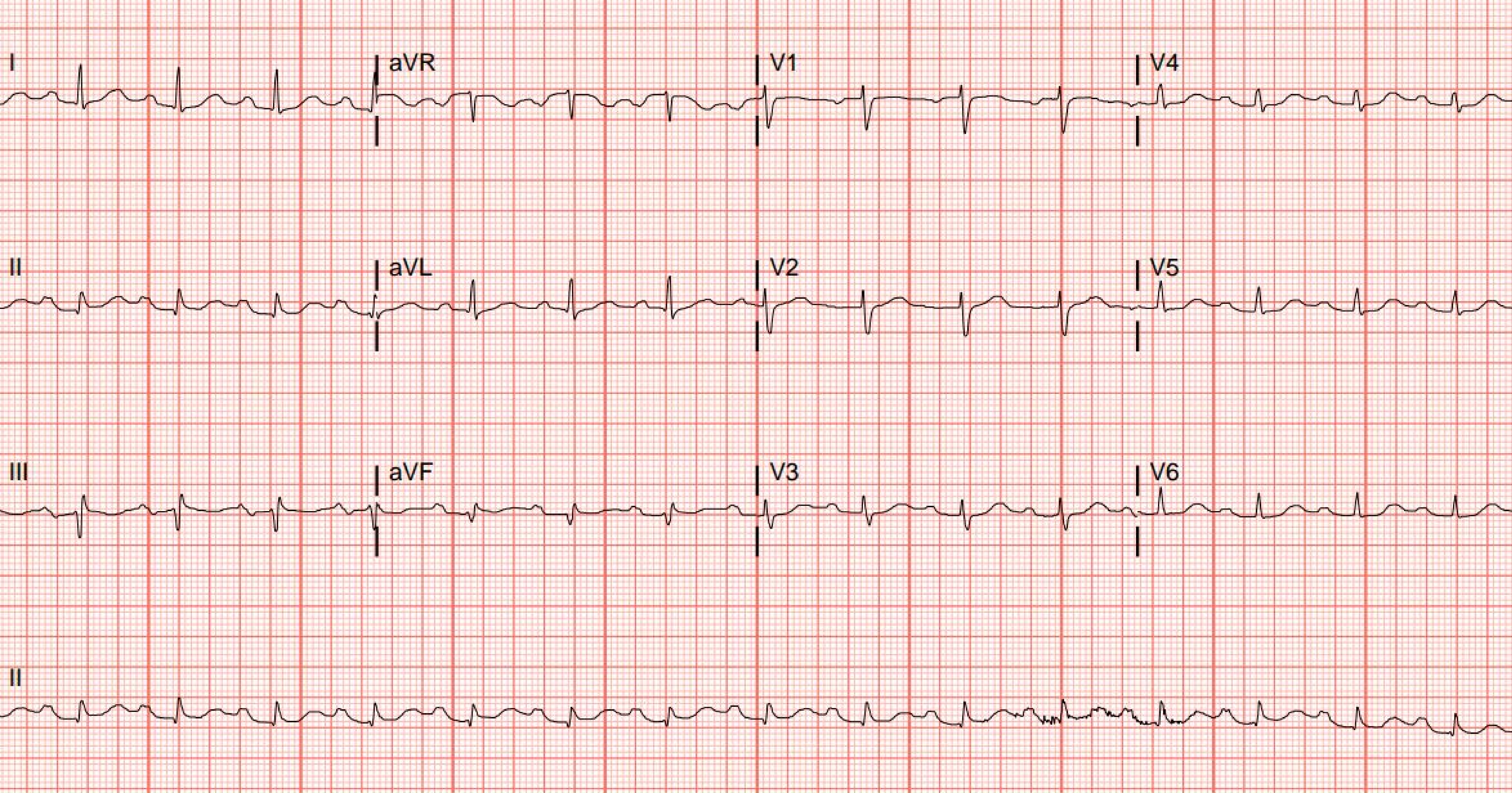
I
aVR
V1
V4
II
aVL
V2
V5
III
aVF
V3
V6
II

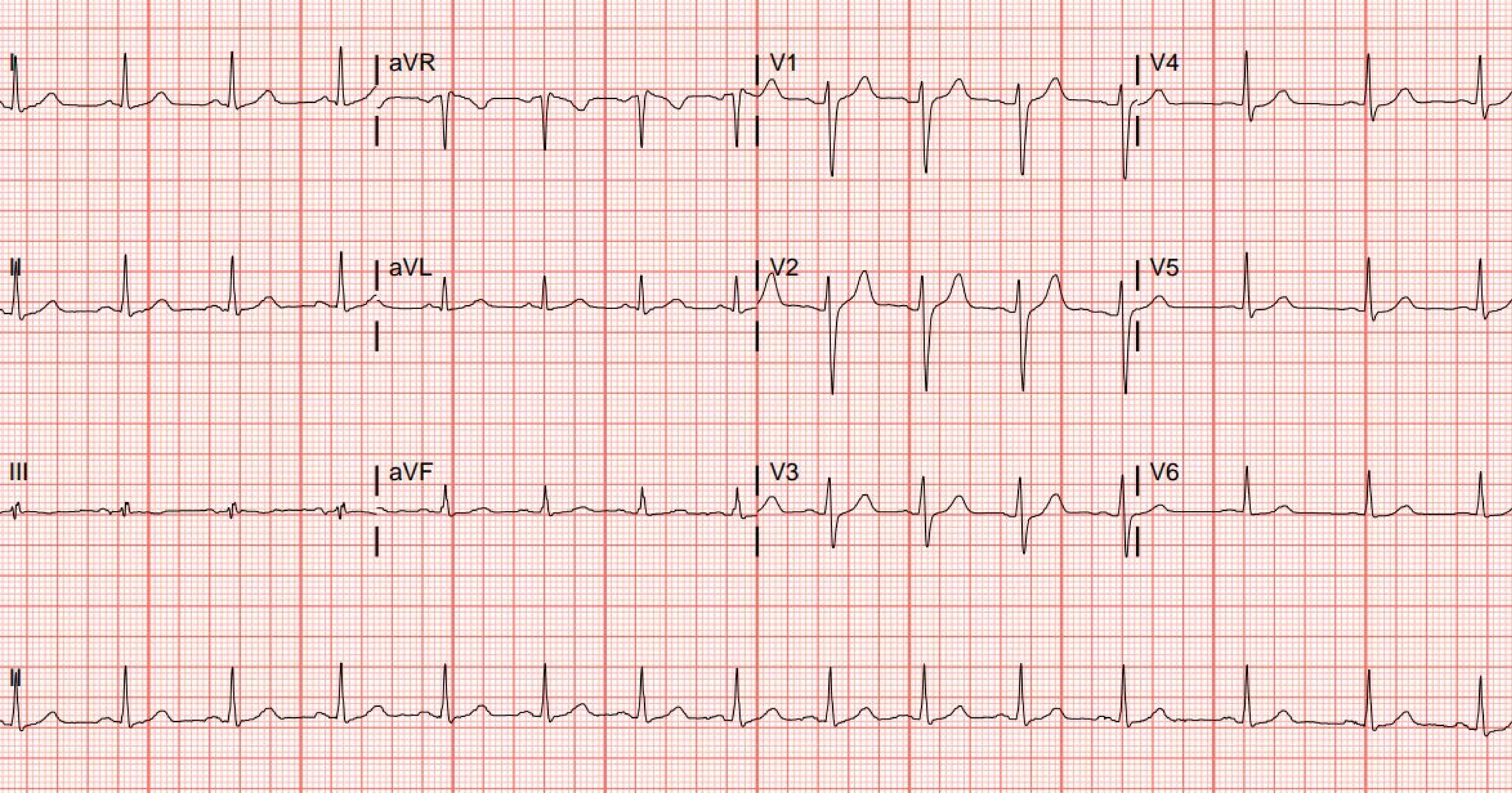

I
aVR
V1
V4
II
aVL
V2
V5
III
aVF
V3
V6
II

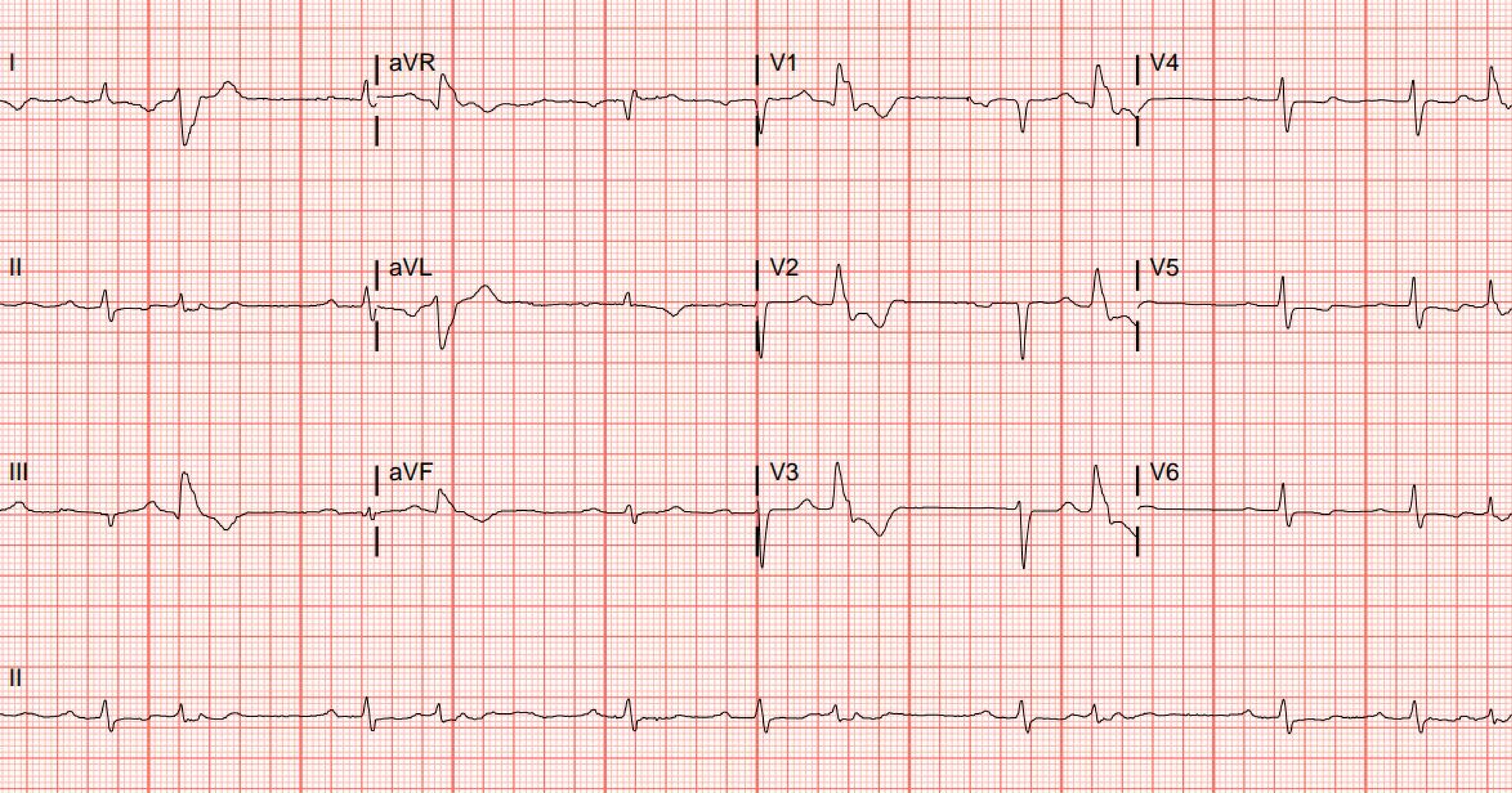

I
aVR
V1
V4
II
aVL
V2
V5
III
aVF
V3
V6
II

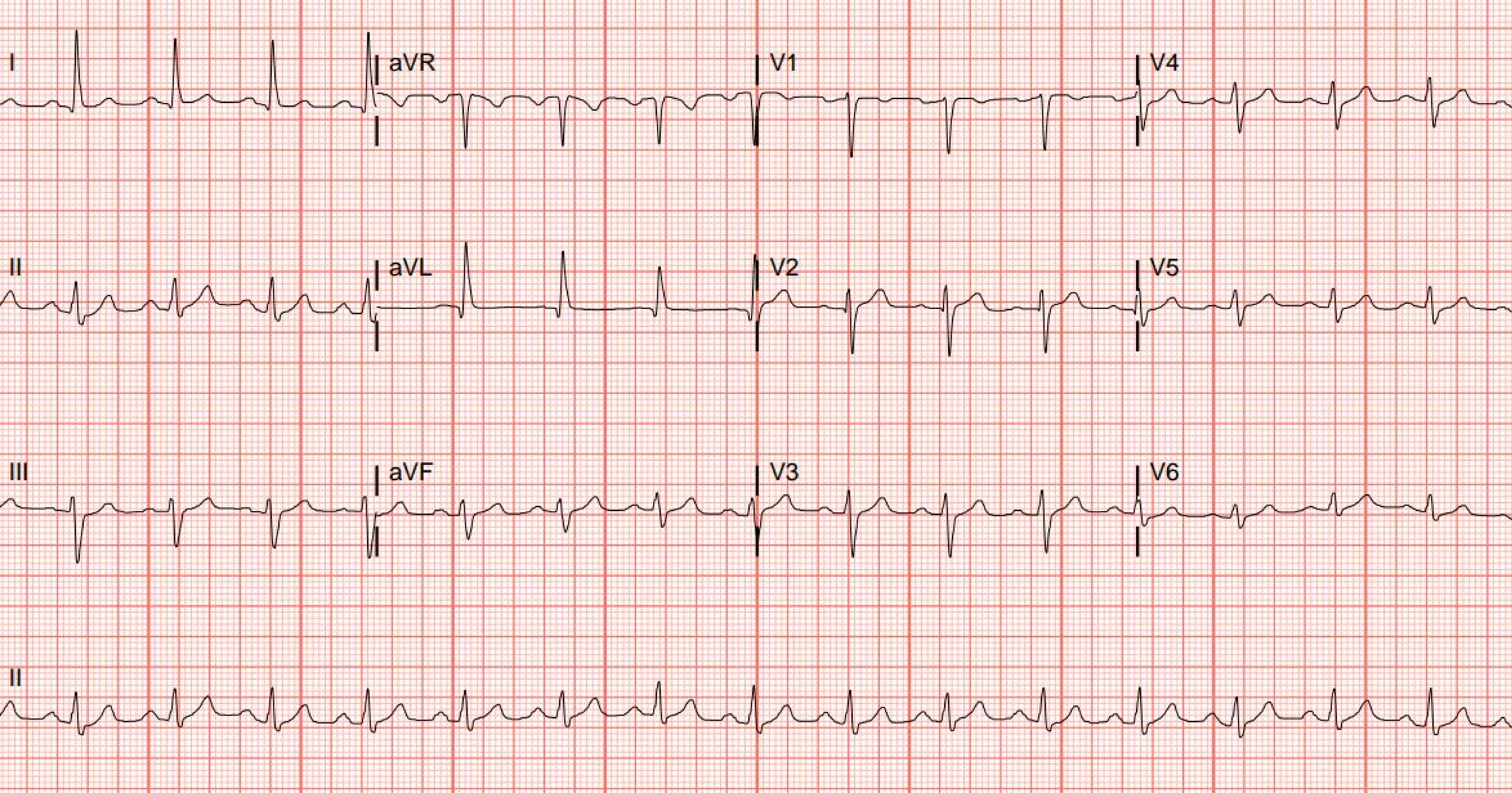
I
aVR
V1
V4
II
aVL
V2
V5
III
aVF
V3
V6
II

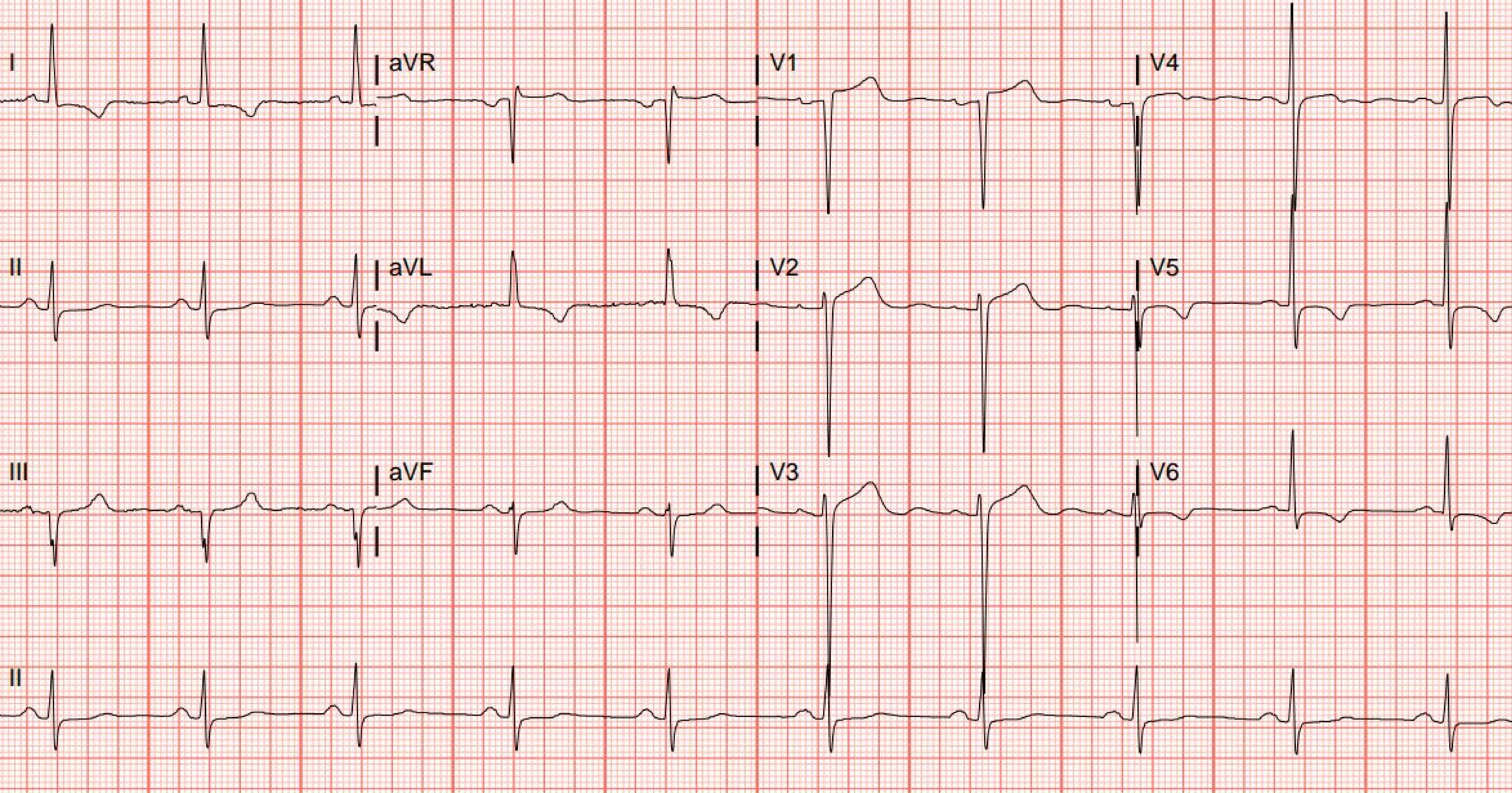
I
aVR
V1
V4
II
aVL
V2
V5
III
aVF
V3
V6
II

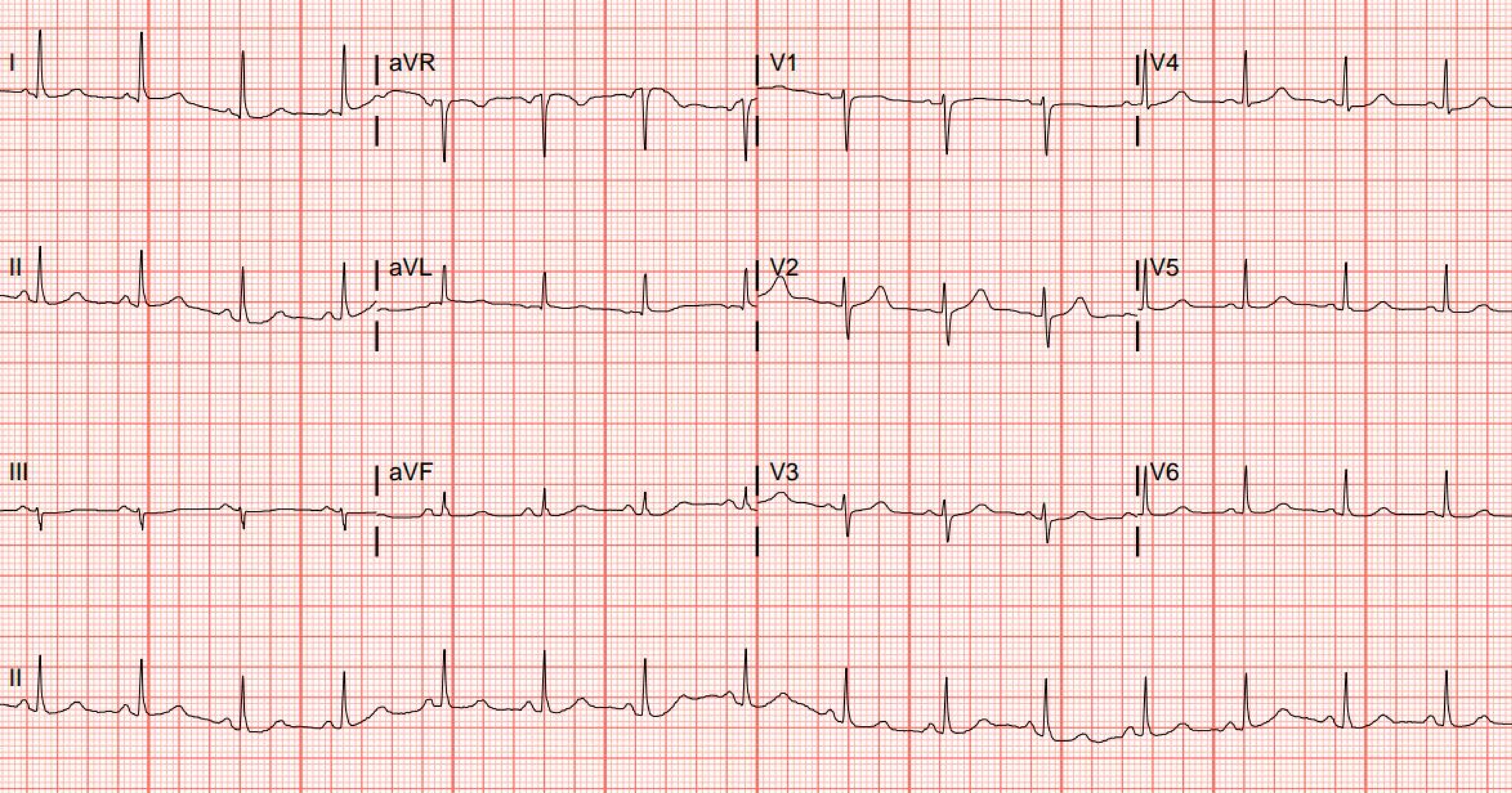
I
aVR
V1
V4
II
aVL
V2
V5
III
aVF
V3
V6
II

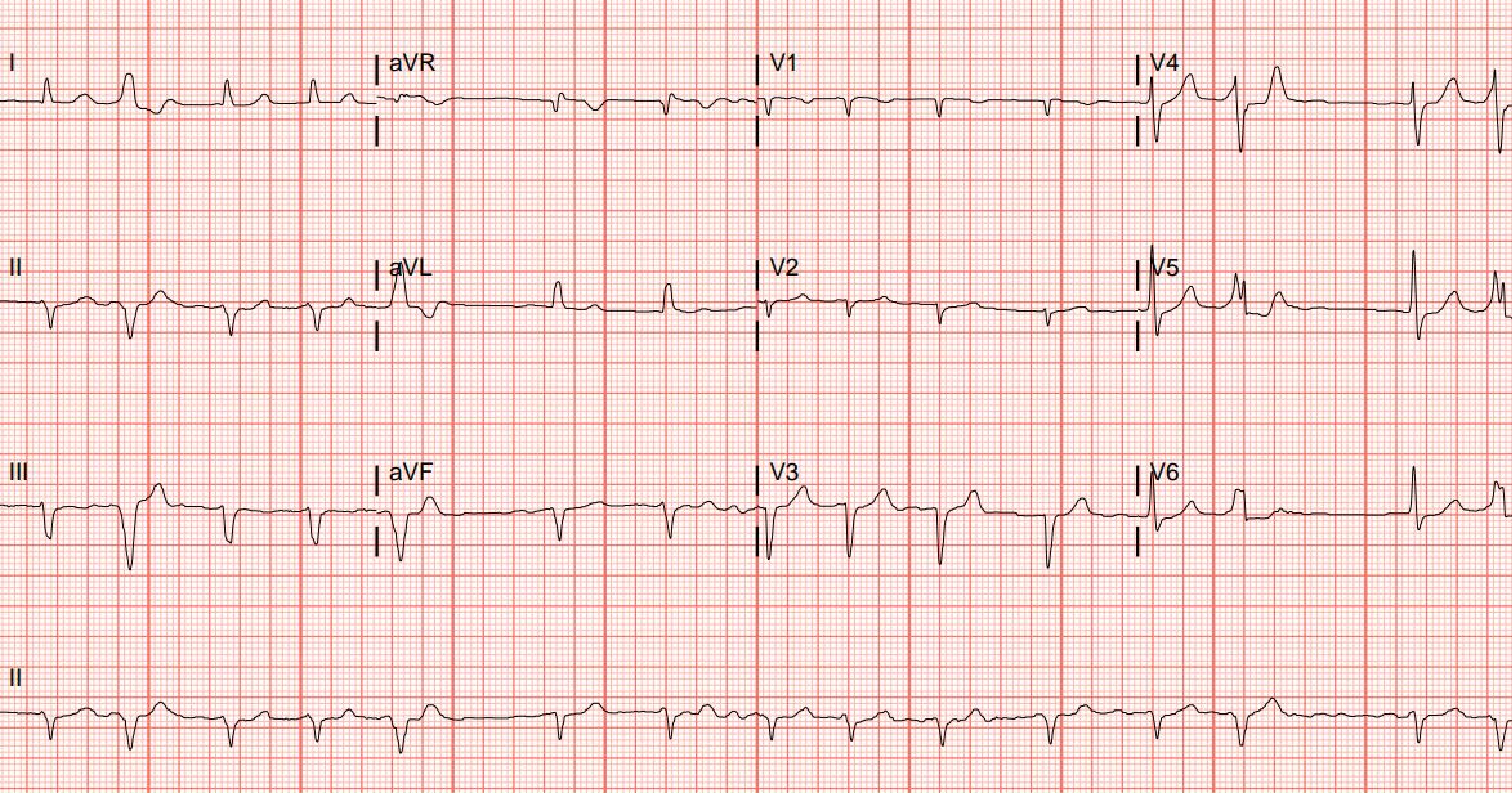
I
aVR
V1
V4
II
aVL
V2
V5
III
aVF
V3
V6
II

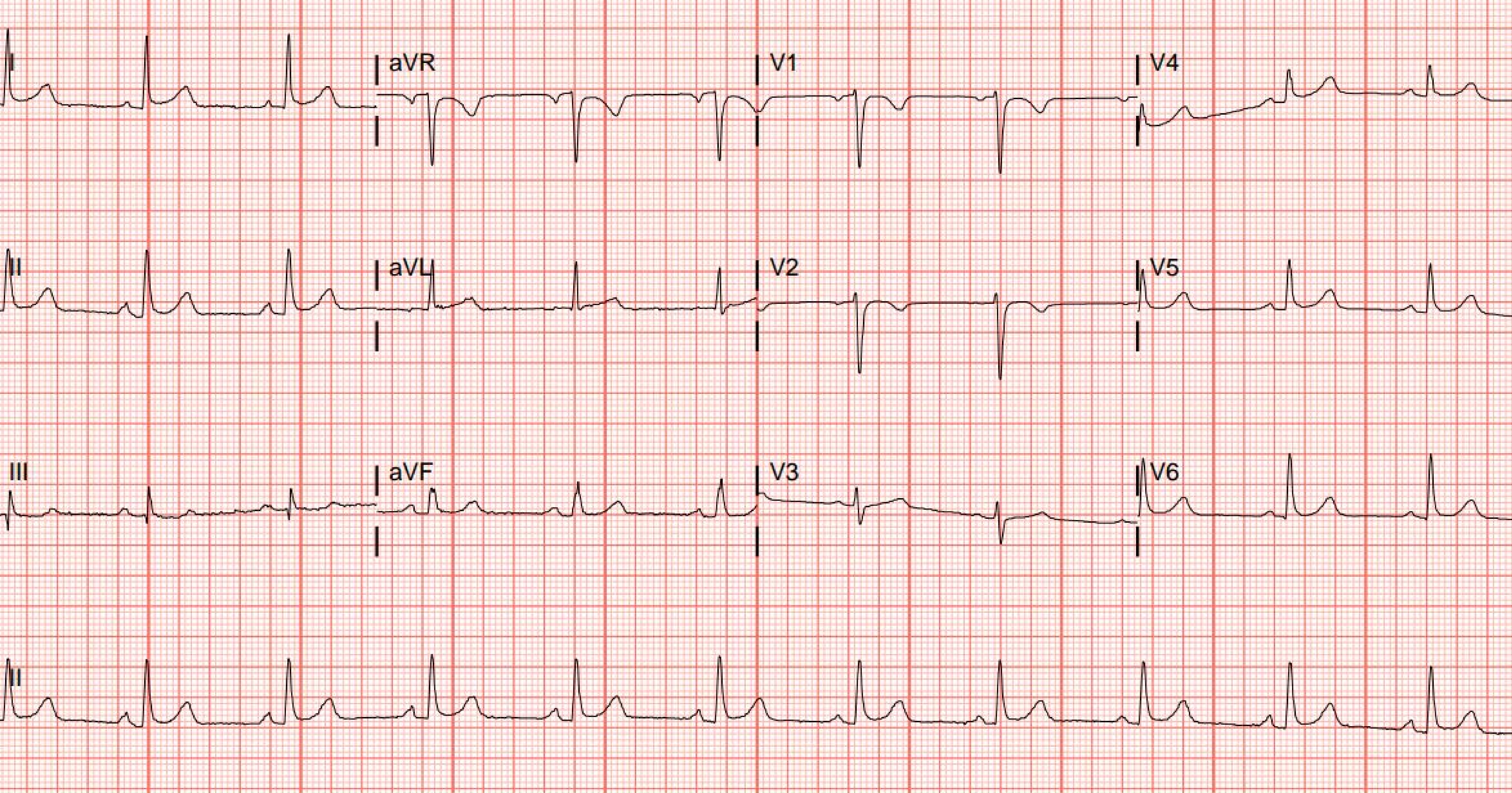
I
aVR
V1
V4
II
aVL
V2
V5
III
aVF
V3
V6
II

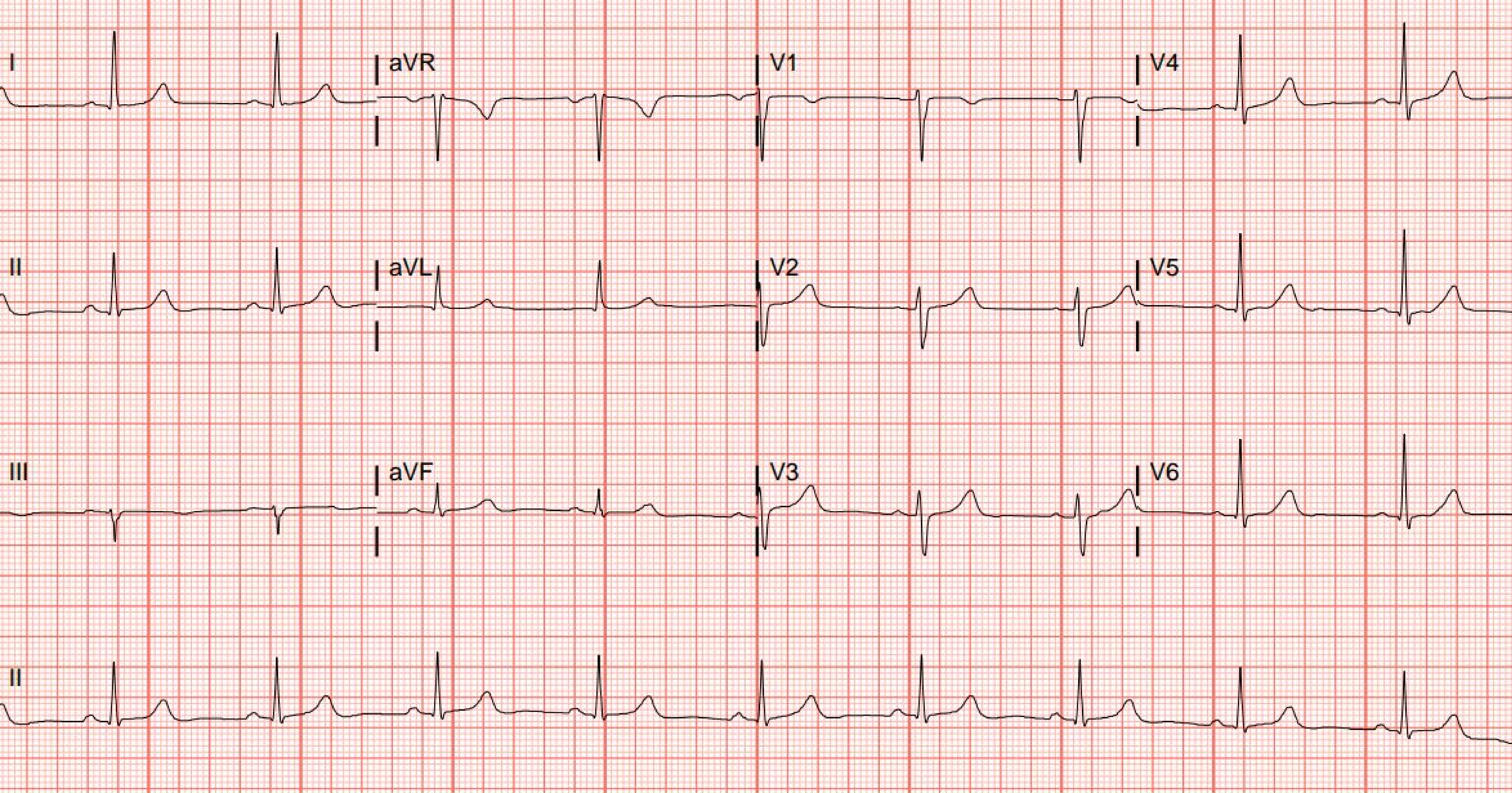
I
aVR
V1
V4
II
aVL
V2
V5
III
aVF
V3
V6
II

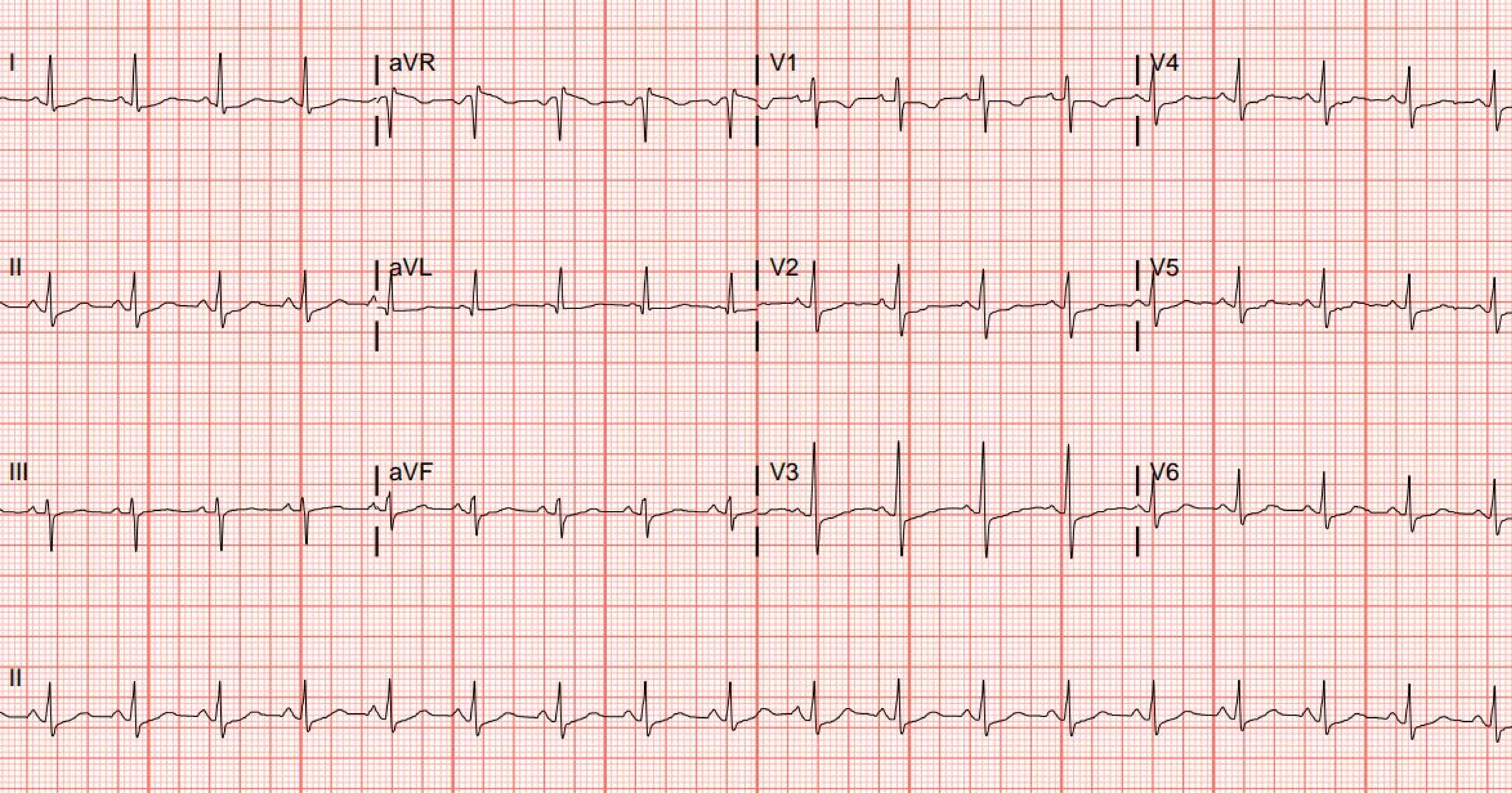
I
aVR
V1
V4
II
aVL
V2
V5
III
aVF
V3
V6
II

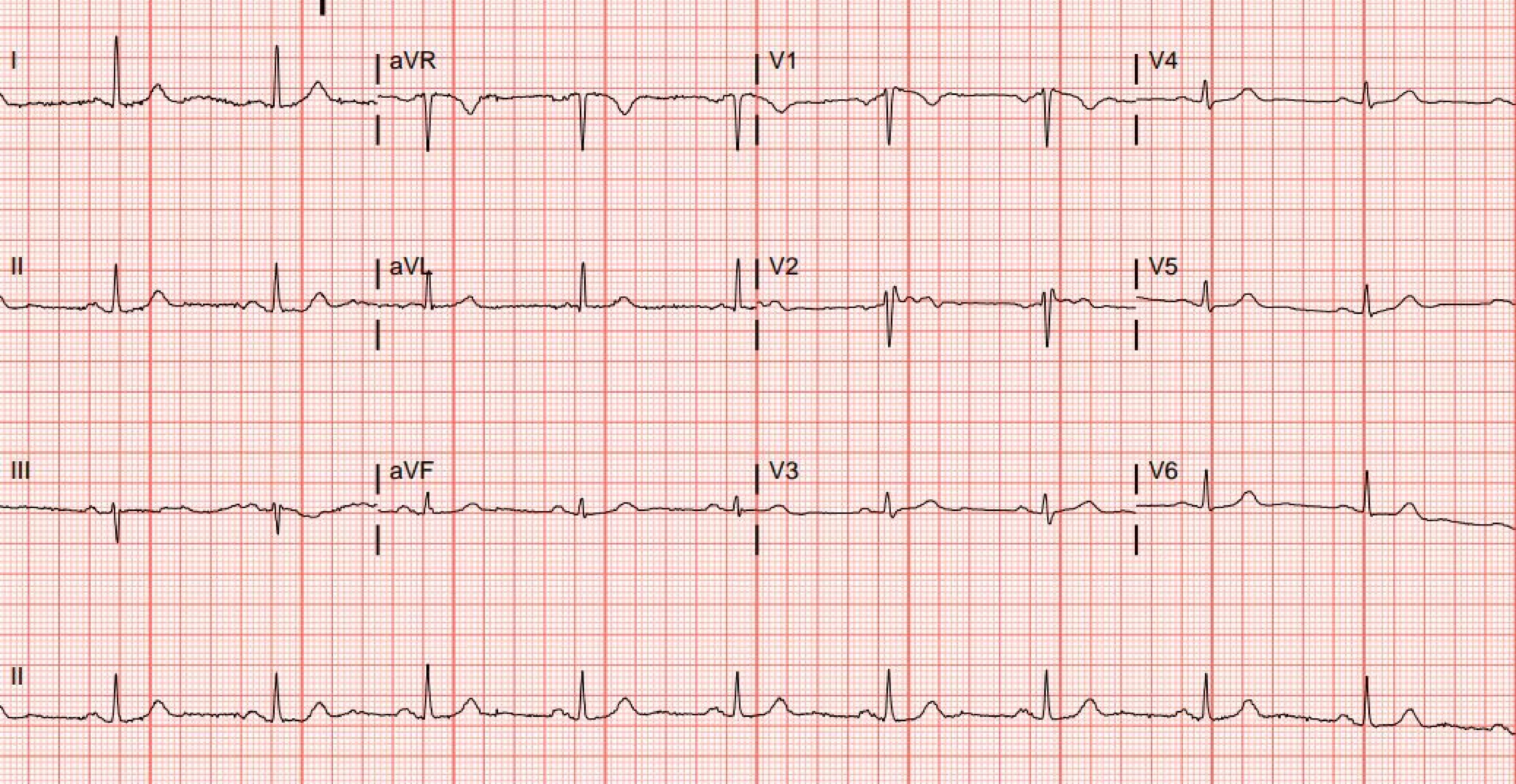
I
aVR
V1
V4
II
aVL
V2
V5
III
aVF
V3
V6
II

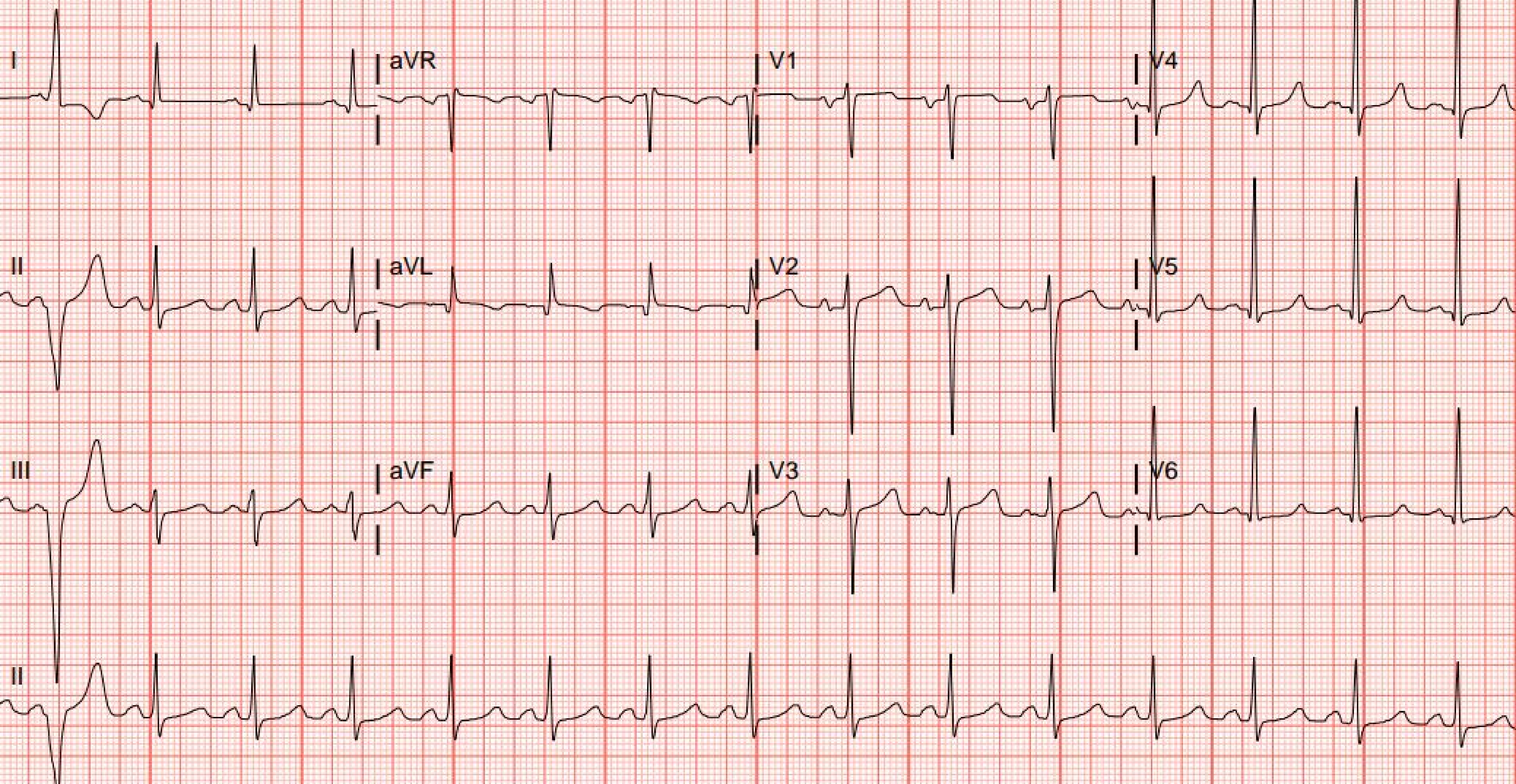
I
aVR
V1
V4
II
aVL
V2
V5
III
aVF
V3
V6
II

### Section 8. Smartphone photographs of 100 electrocardiograms on computer screen (30 Apple iPhone 15 Pro, 40 Apple iPhone 12, 30 Samsung Galaxy S21 5G; cropped)

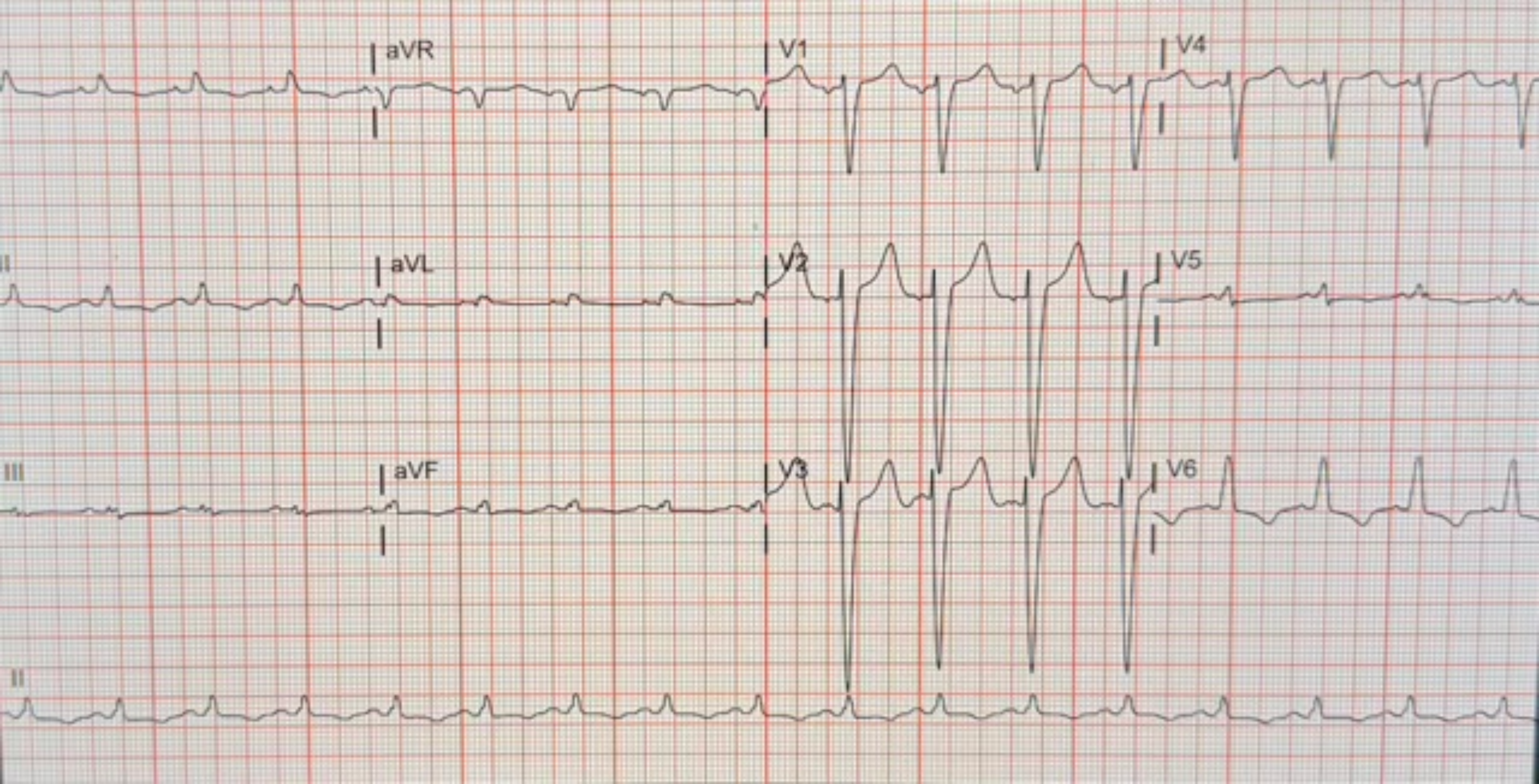
aVR
V1
V4
II
aVL
V2
V5
III
aVF
V3
V6
II

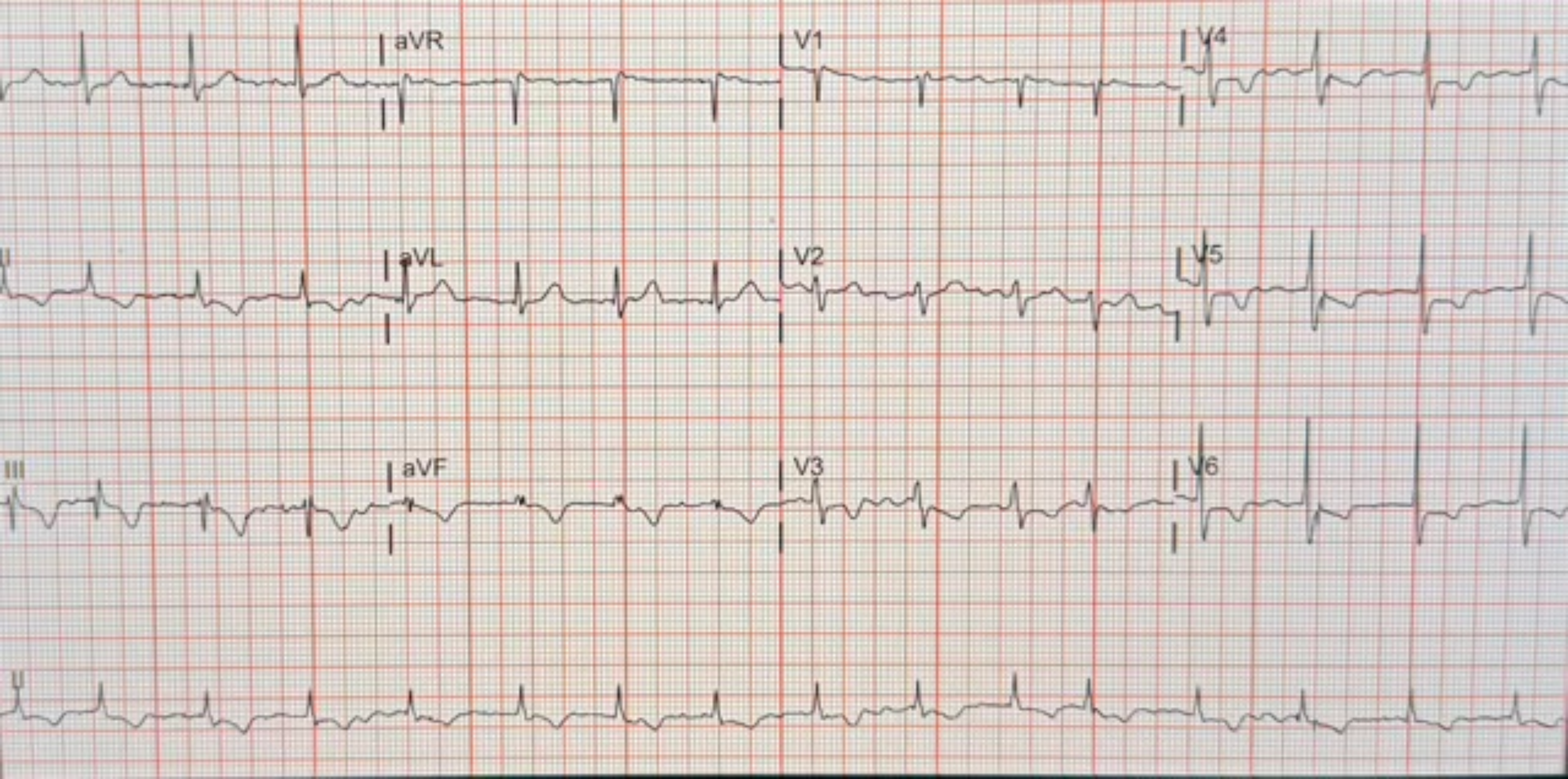
aVR
V1
V4
II
aVL
V2
V5
III
aVF
V3
V6
II

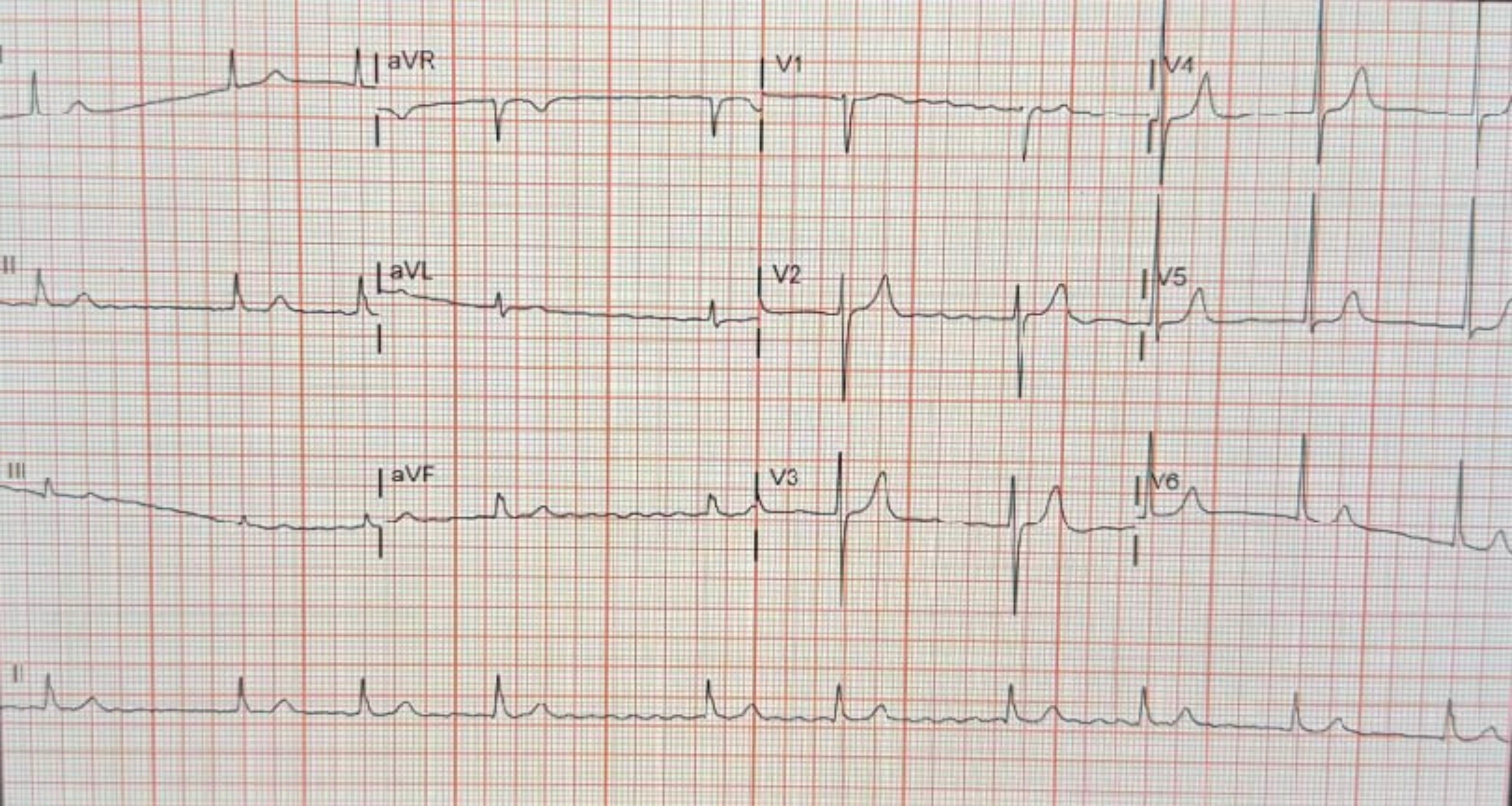
I
aVR
V1
V4
II
aVL
V2
V5
III
aVF
V3
V6
II

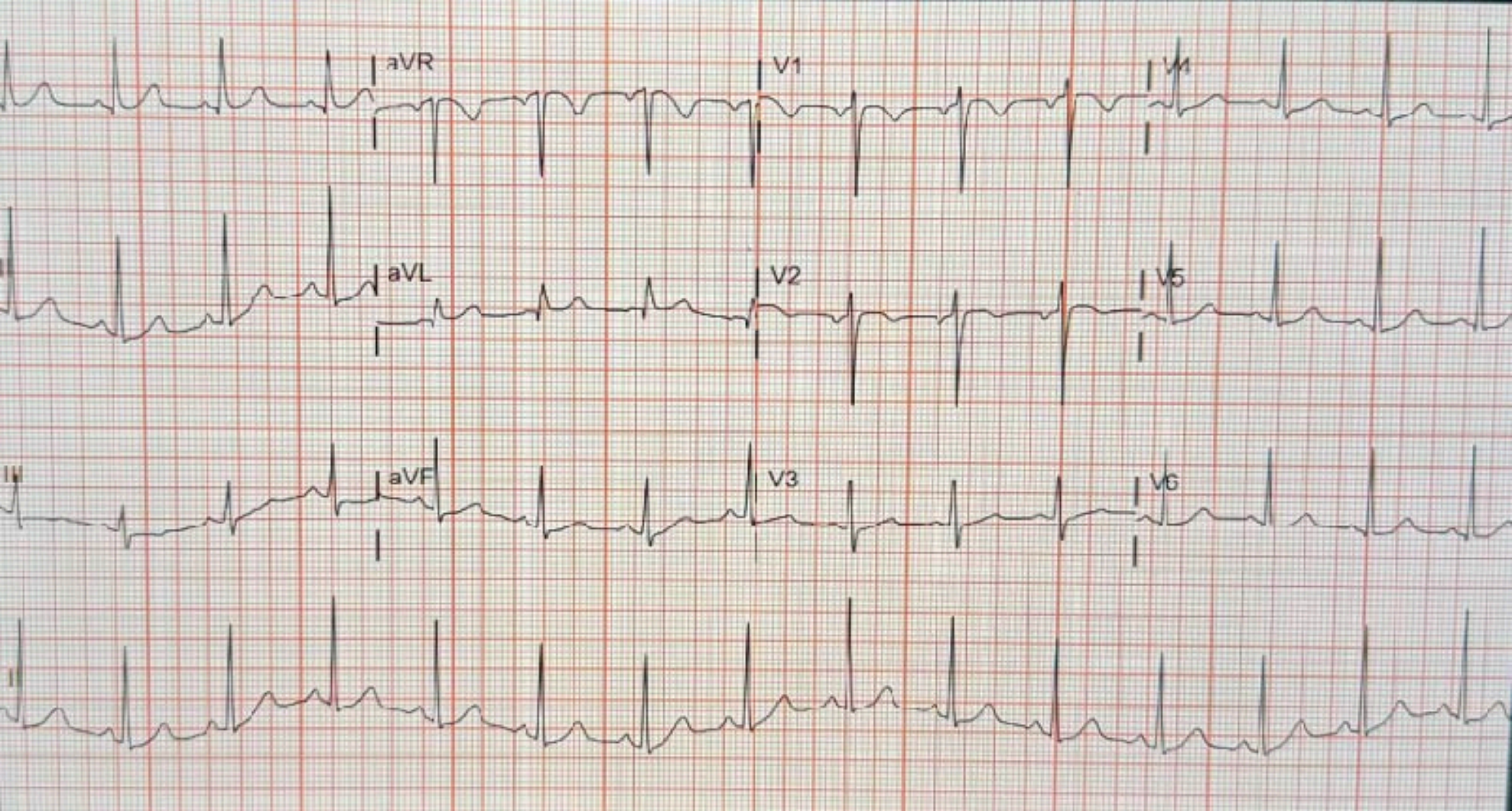
aVR
V1
V4
aVL
V2
V5
aVF
V3
V6

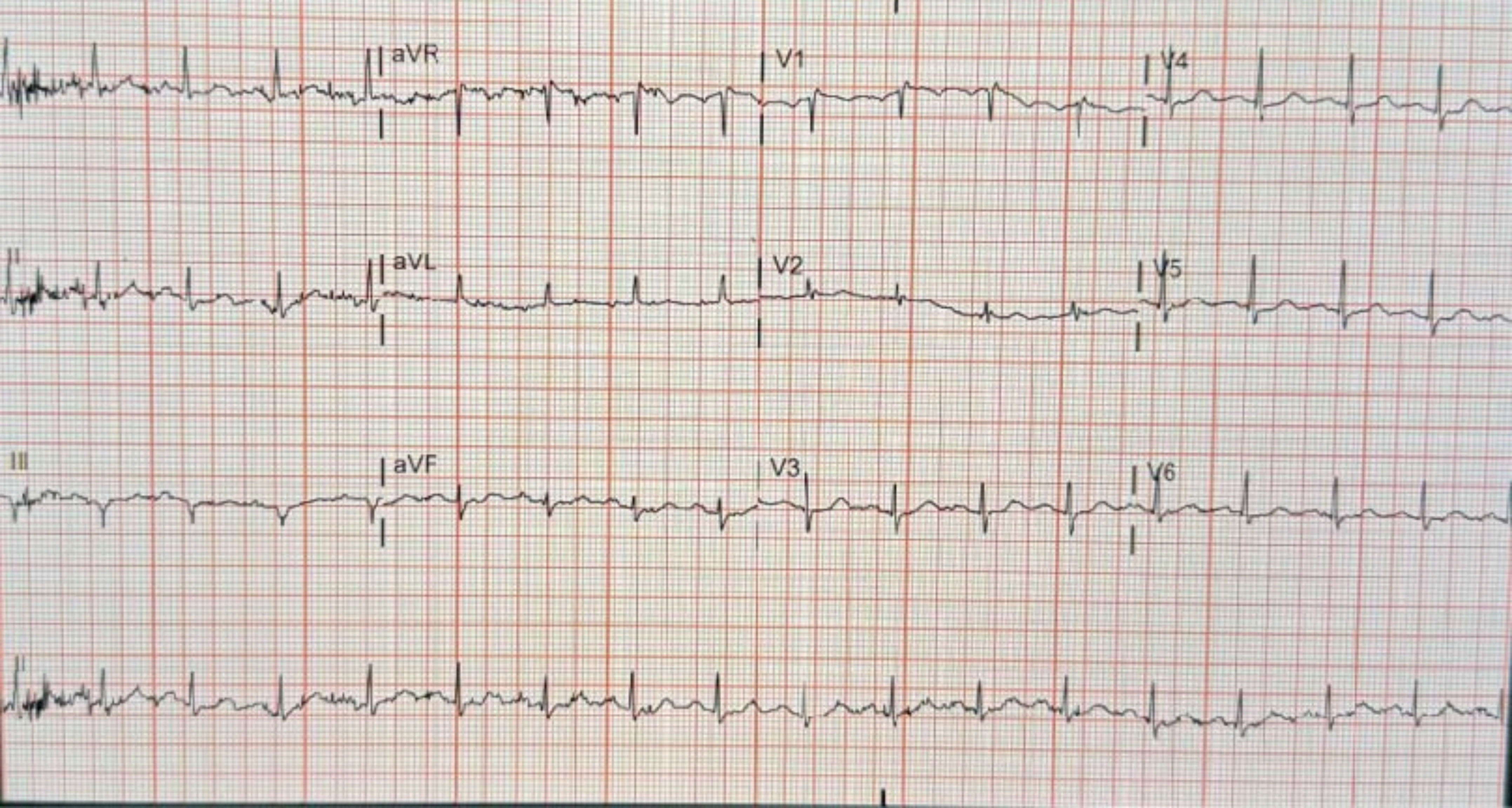
aVR
V1
V4
II
aVL
V2
V5
III
aVF
V3
V6

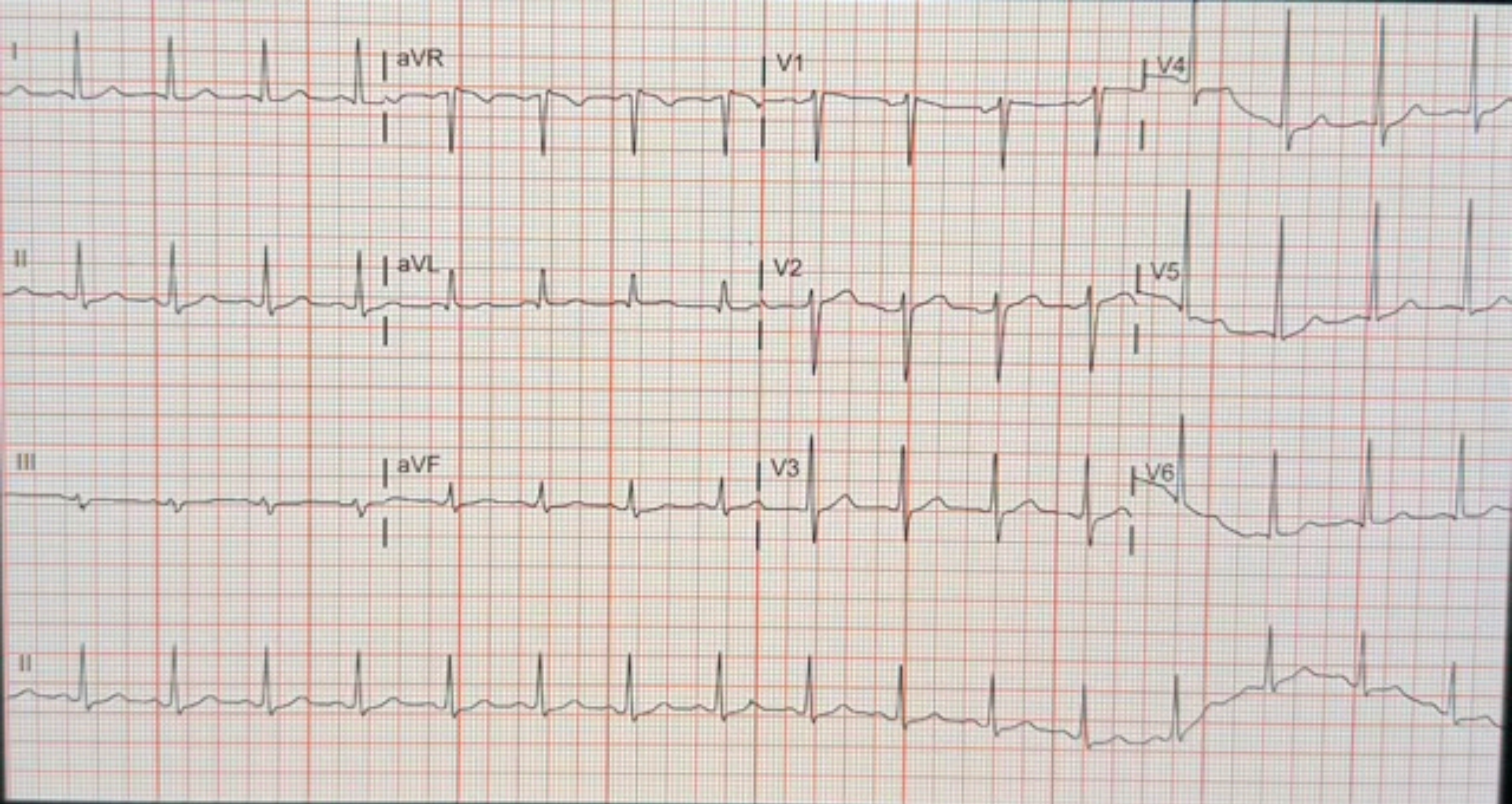
I
aVR
V1
V4
II
aVL
V2
V5
III
aVF
V3
V6
II

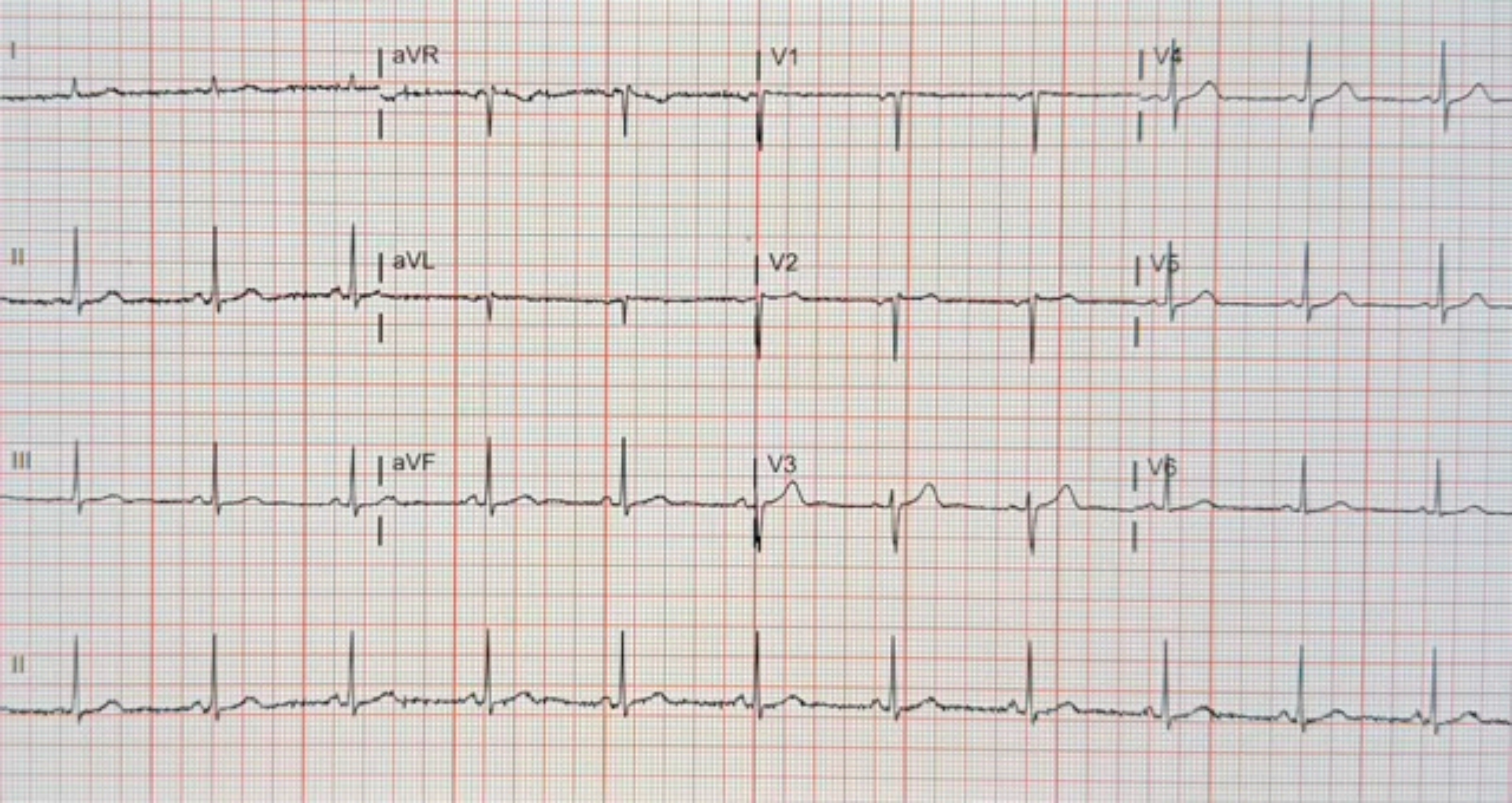
I
aVR
V1
V4
II
aVL
V2
V5
III
aVF
V3
V6
II

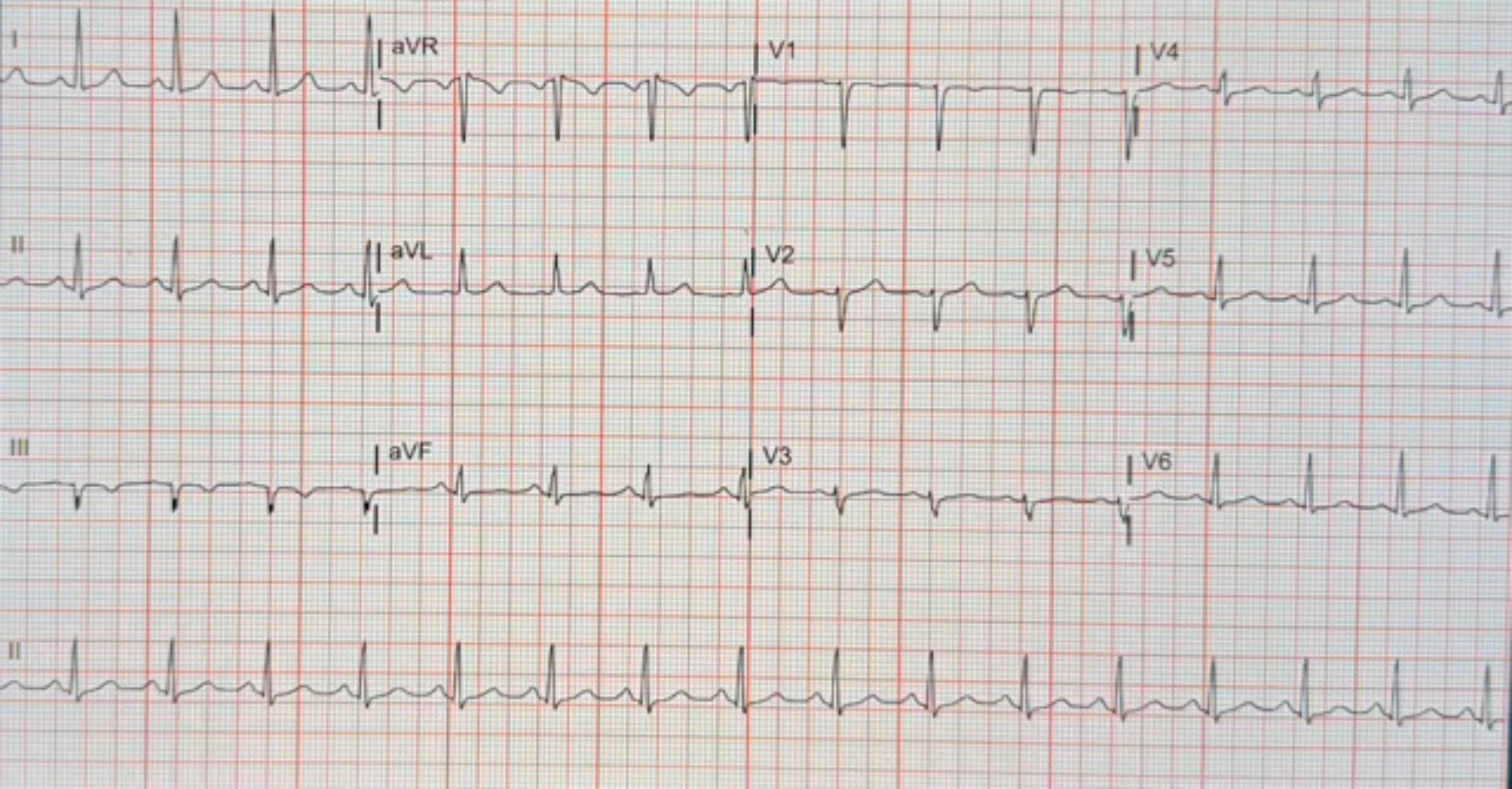
I
aVR
V1
V4
II
aVL
V2
V5
III
aVF
V3
V6
II

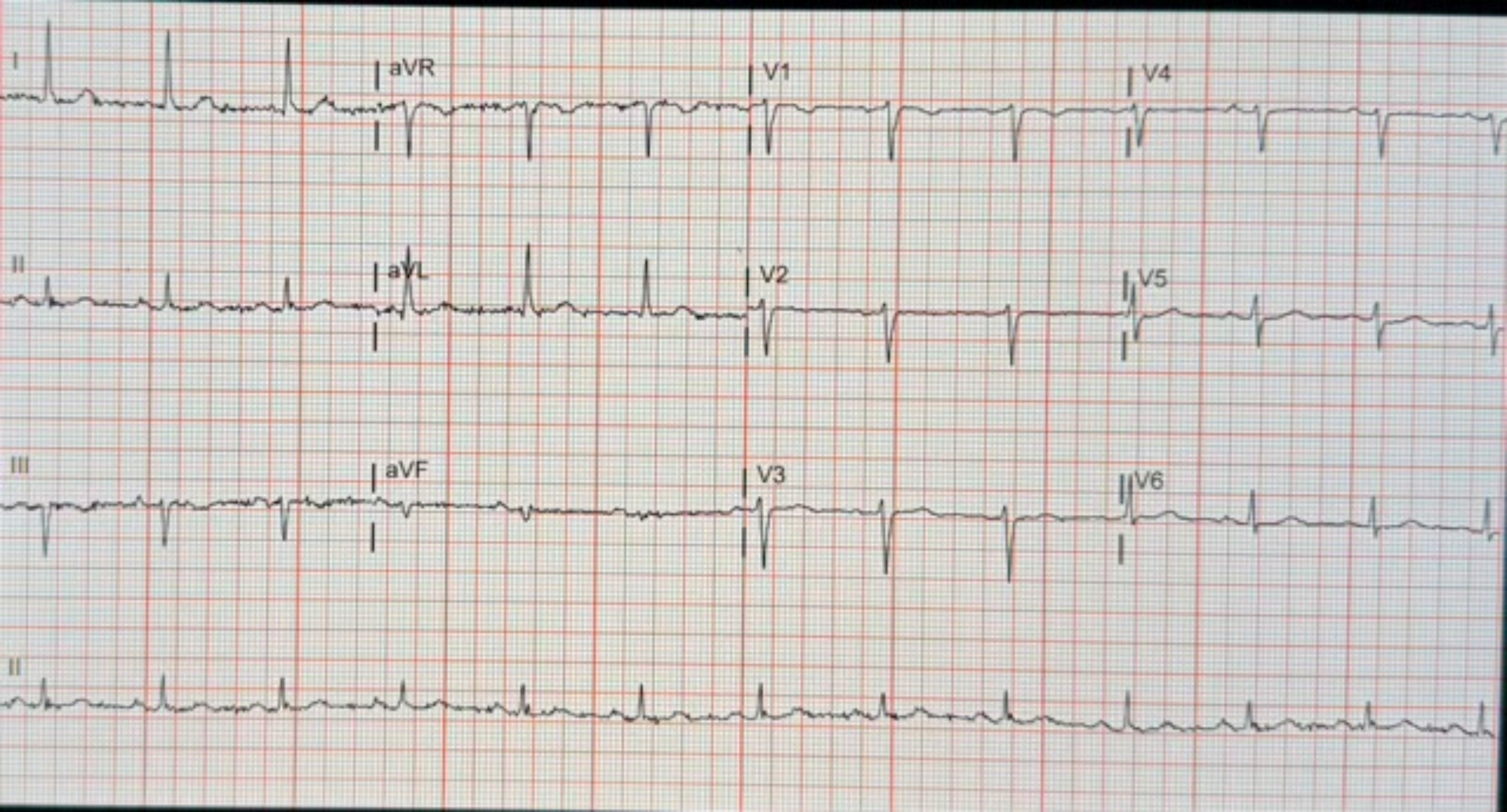
I
aVR
V1
V4
II
aVL
V2
V5
III
aVF
V3
V6
II

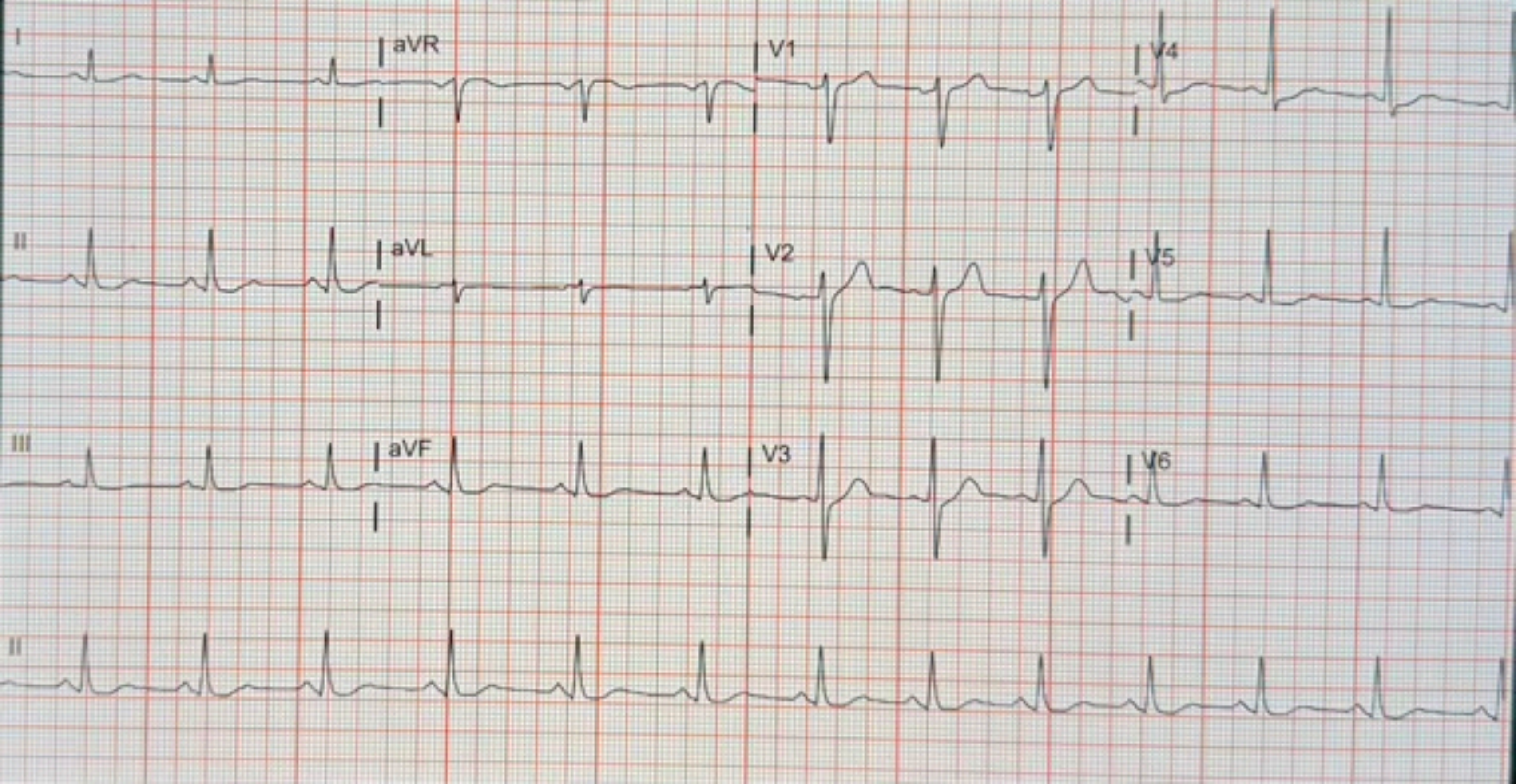
I
aVR
V1
V4
II
aVL
V2
V5
III
aVF
V3
V6
II

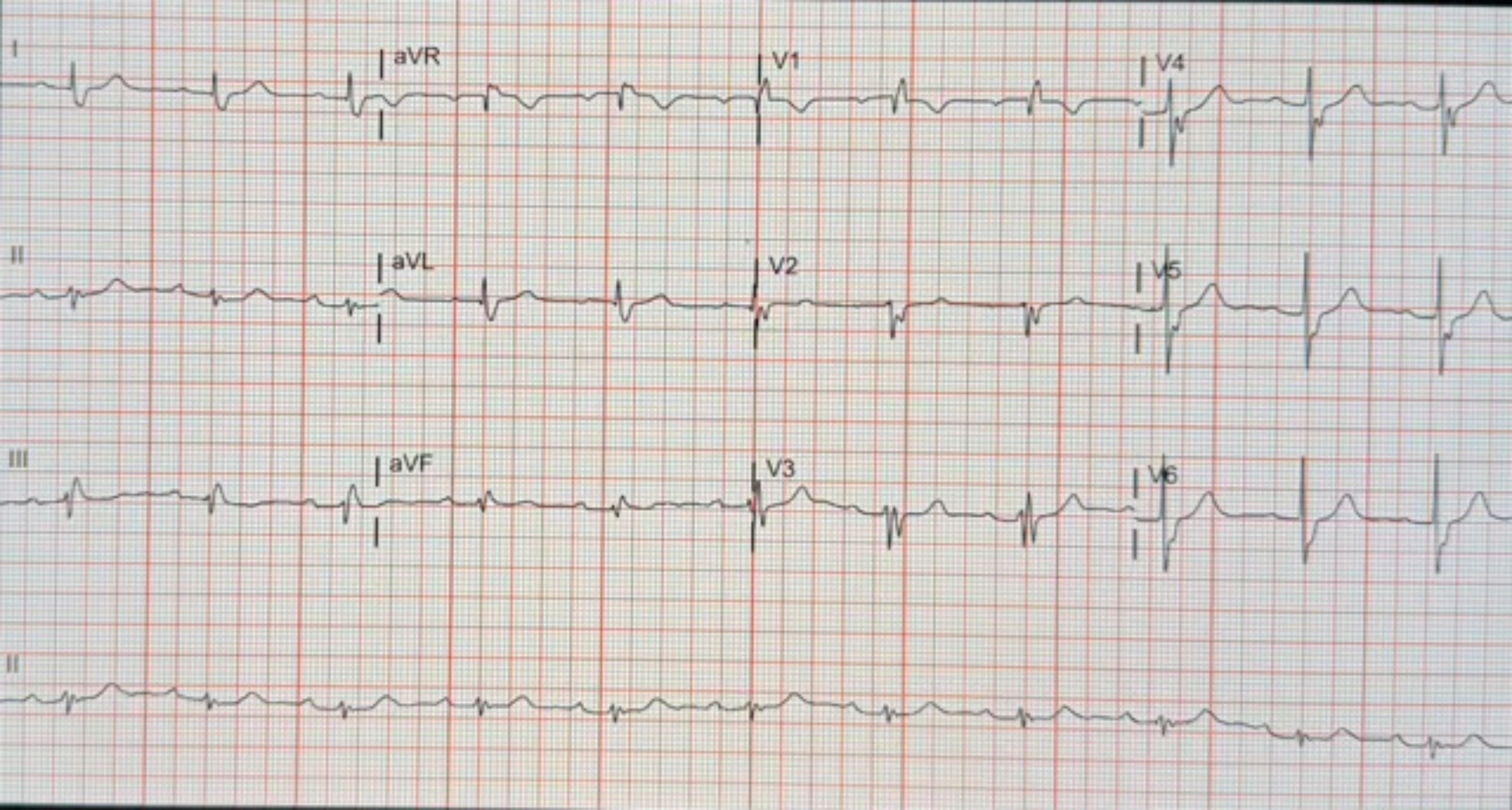
I
aVR
V1
V4
II
aVL
V2
V5
III
aVF
V3
V6
II

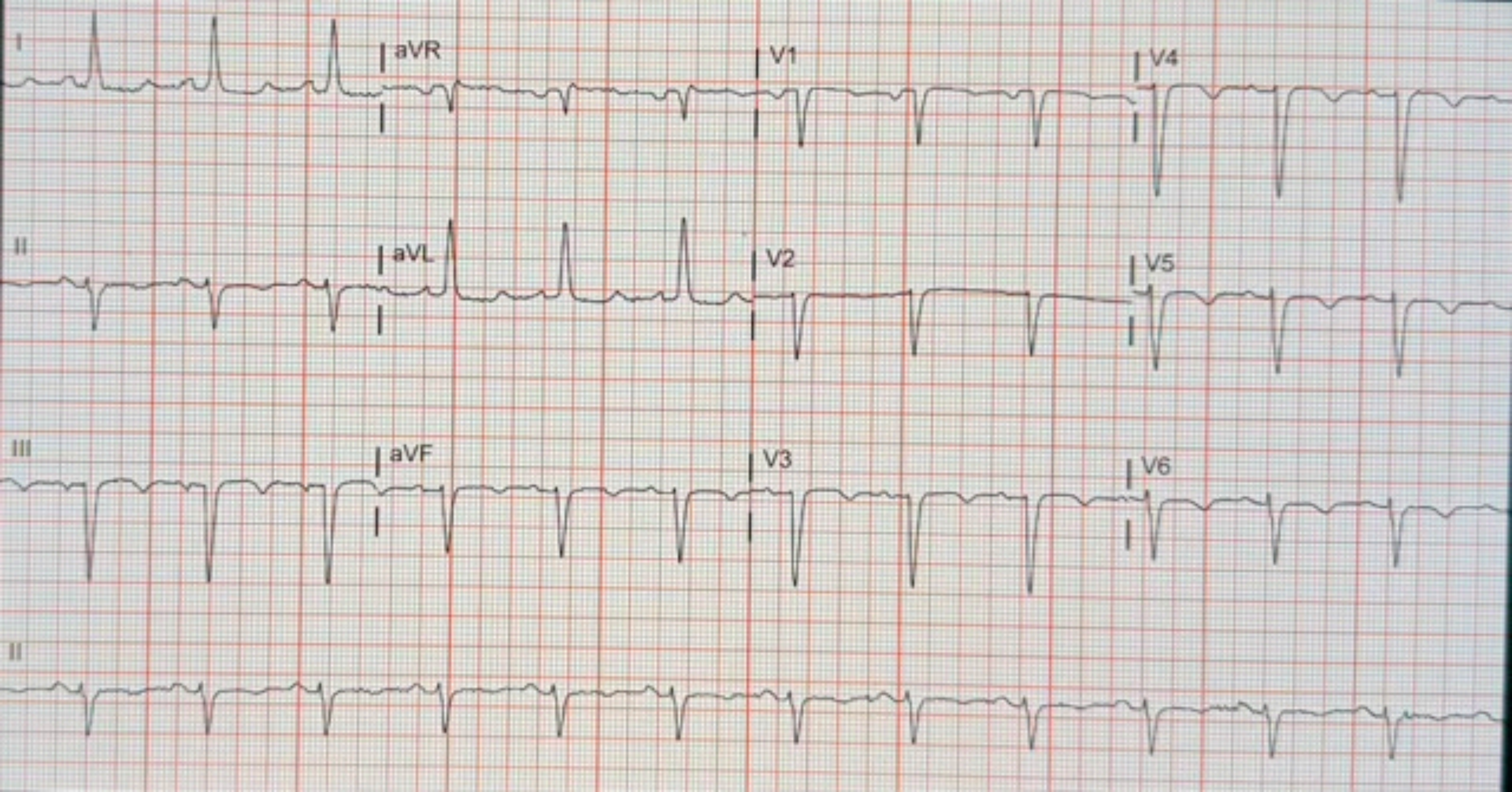
I
aVR
V1
V4
II
aVL
V2
V5
III
aVF
V3
V6
II

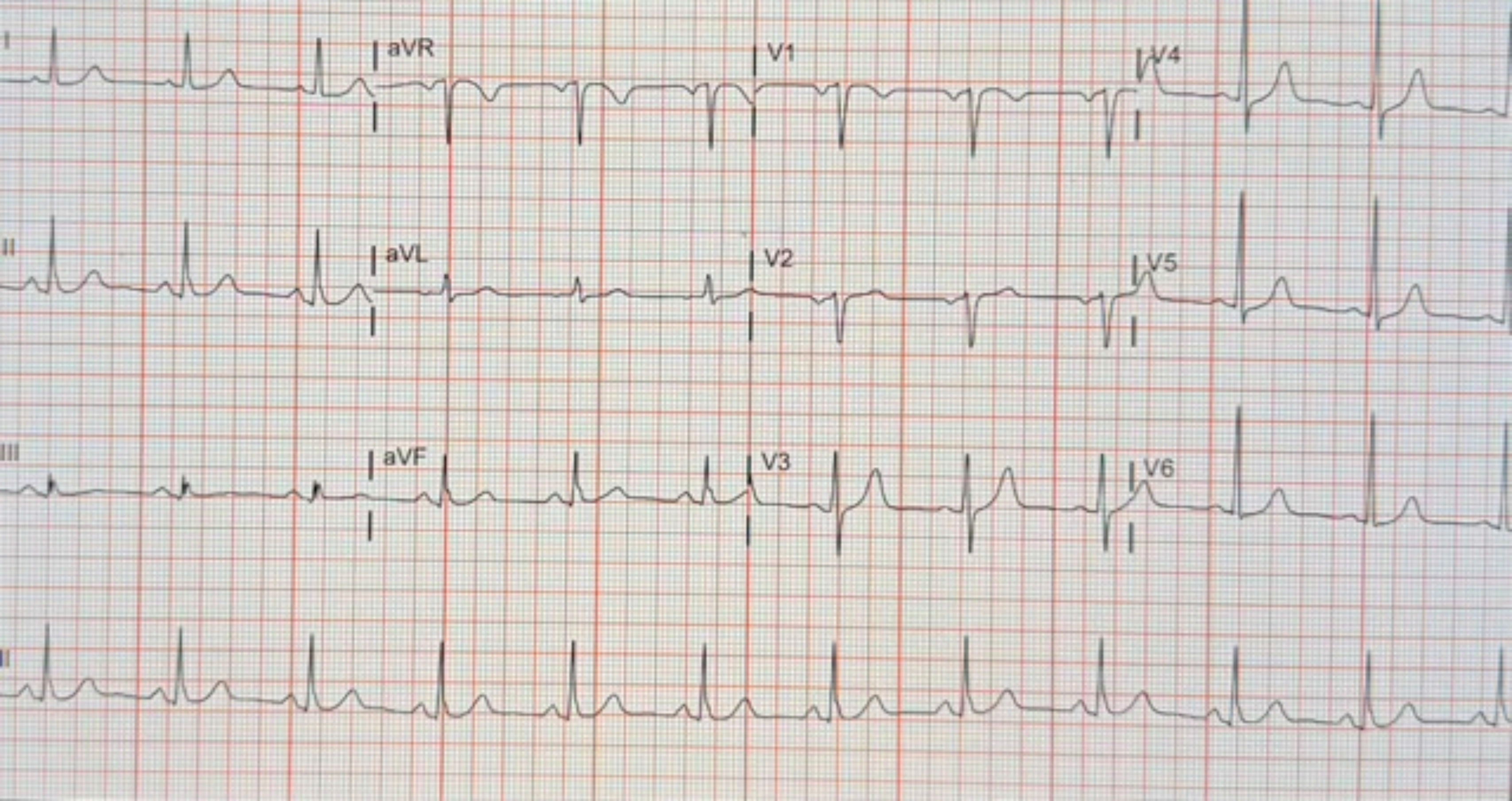
I
aVR
V1
V4
II
aVL
V2
V5
III
aVF
V3
V6
II

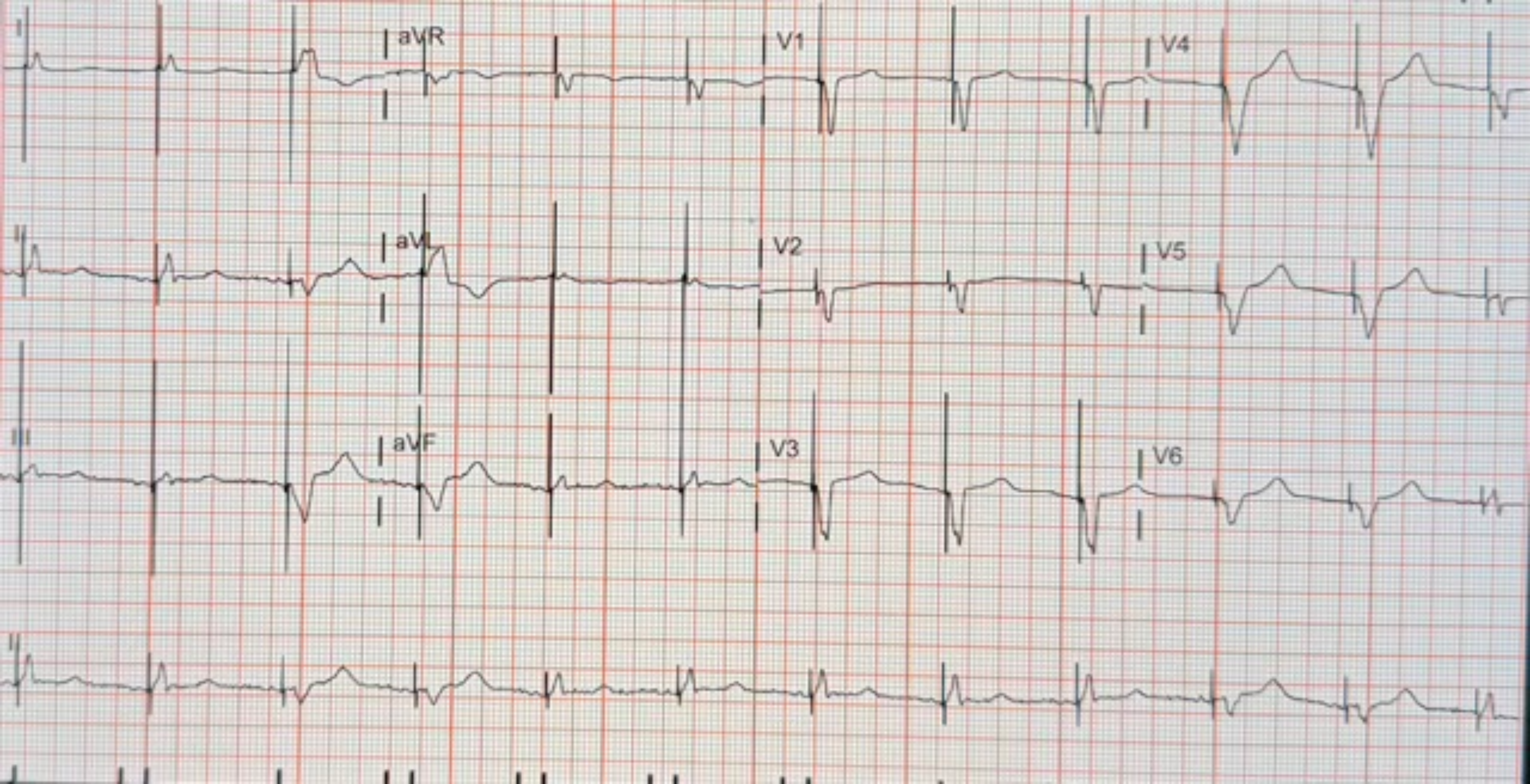

I
aVR
V1
V4
II
aVL
V2
V5
III
aVF
V3
V6
II

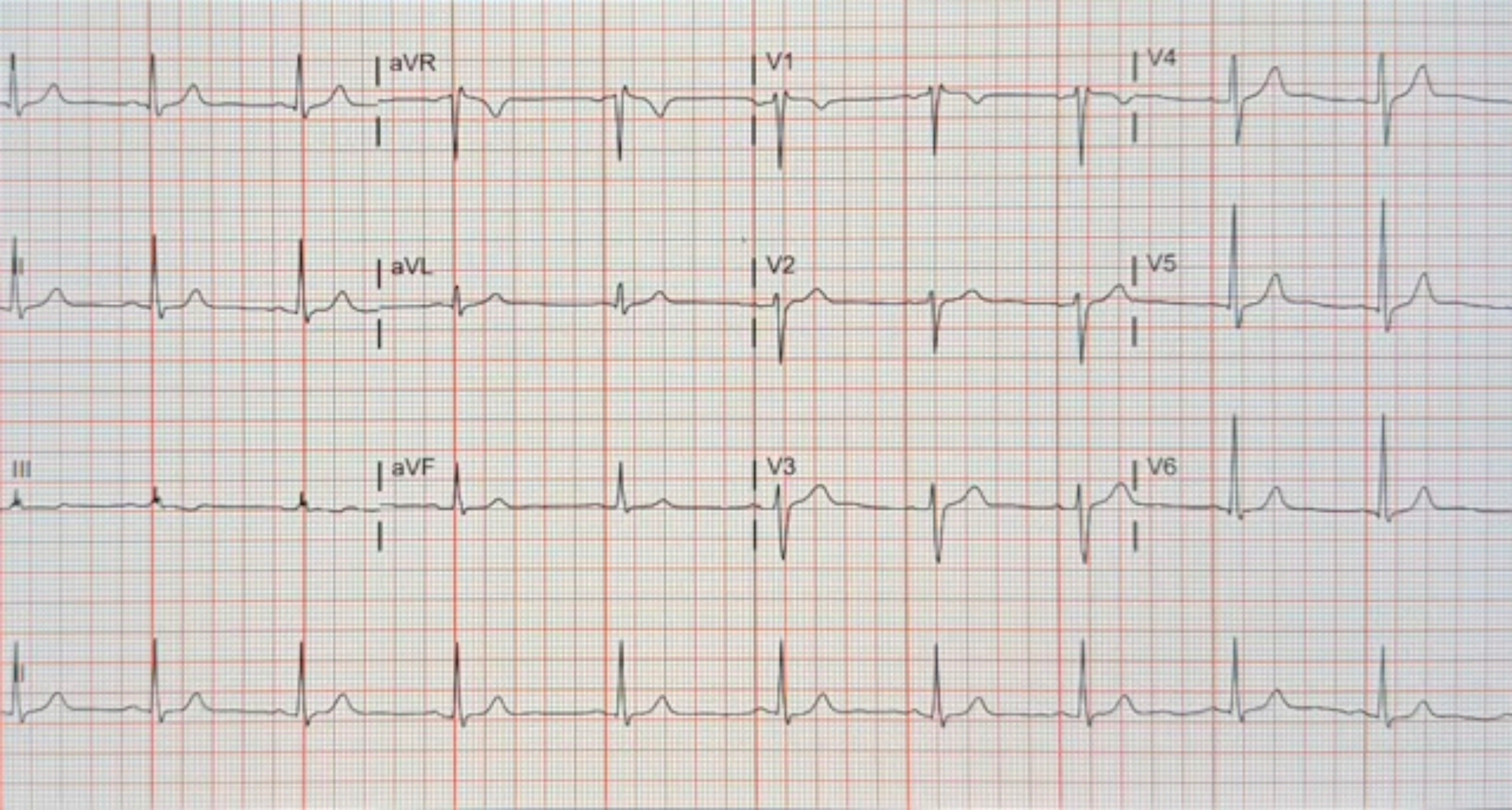

aVR
V1
V4
II
aVL
V2
V5
III
aVF
V3
V6
II

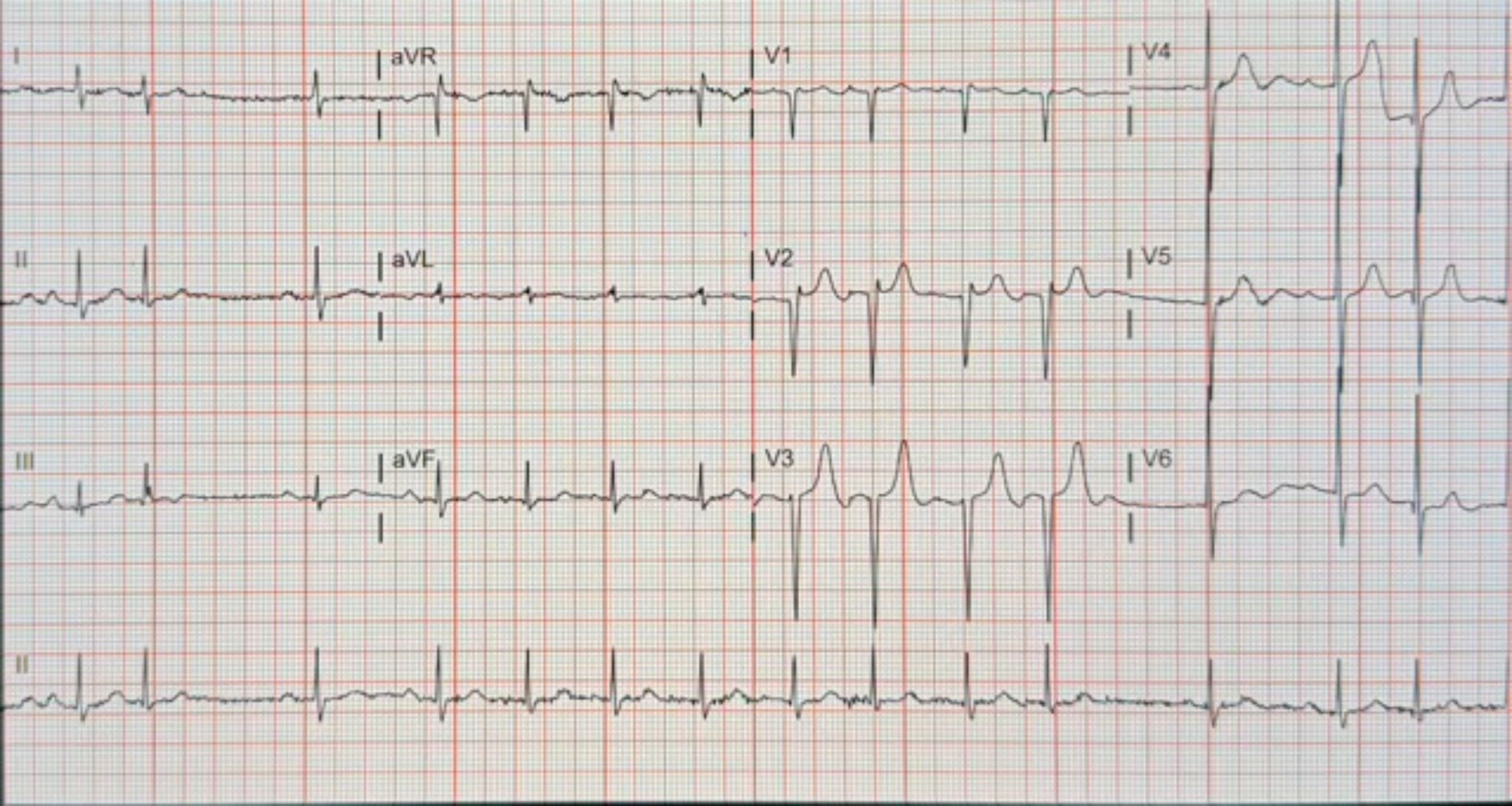
I
aVR
V1
V4
II
aVL
V2
V5
III
aVF
V3
V6
II

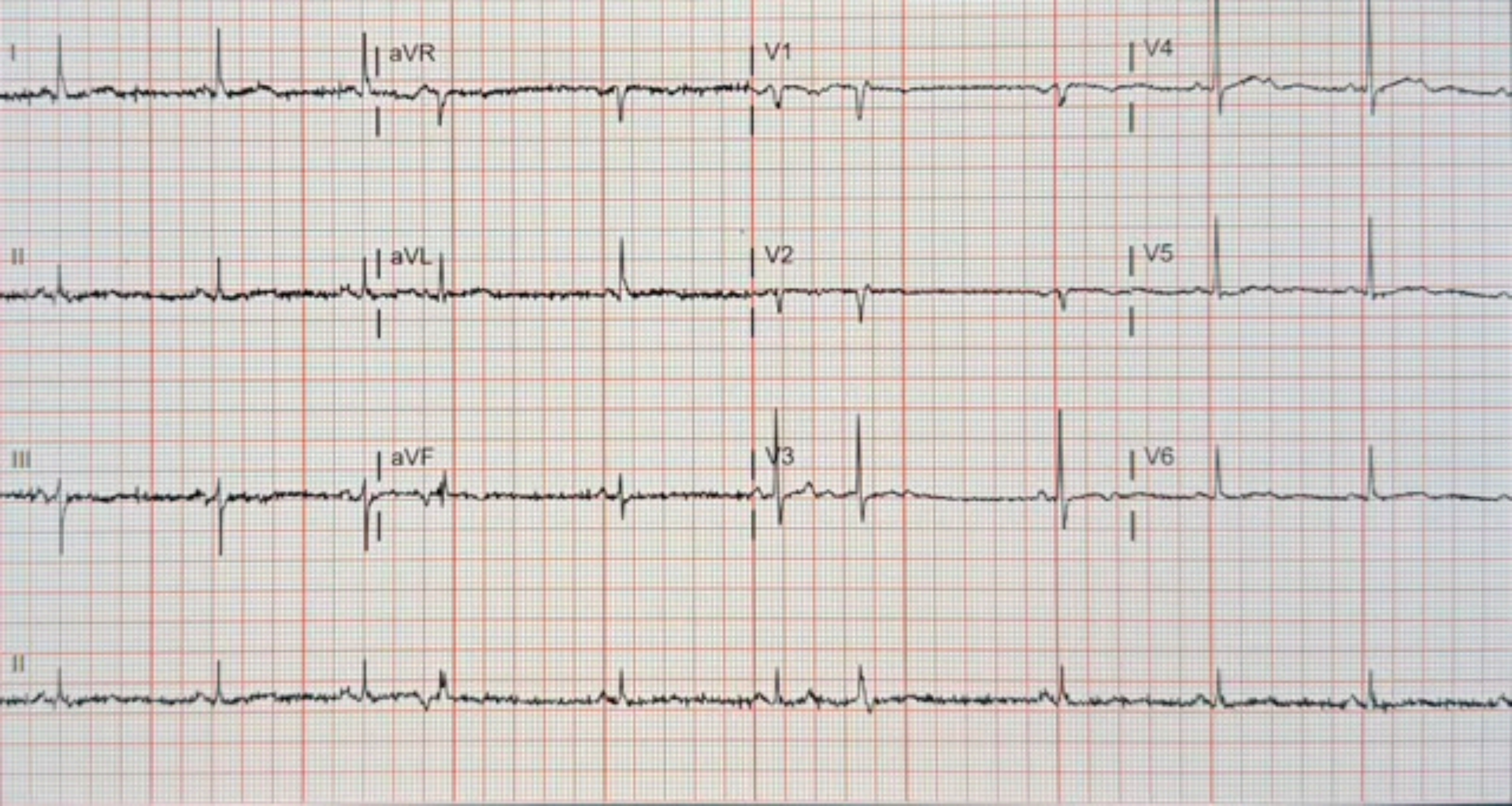
I
aVR
V1
V4
II
aVL
V2
V5
III
aVF
V3
V6
II

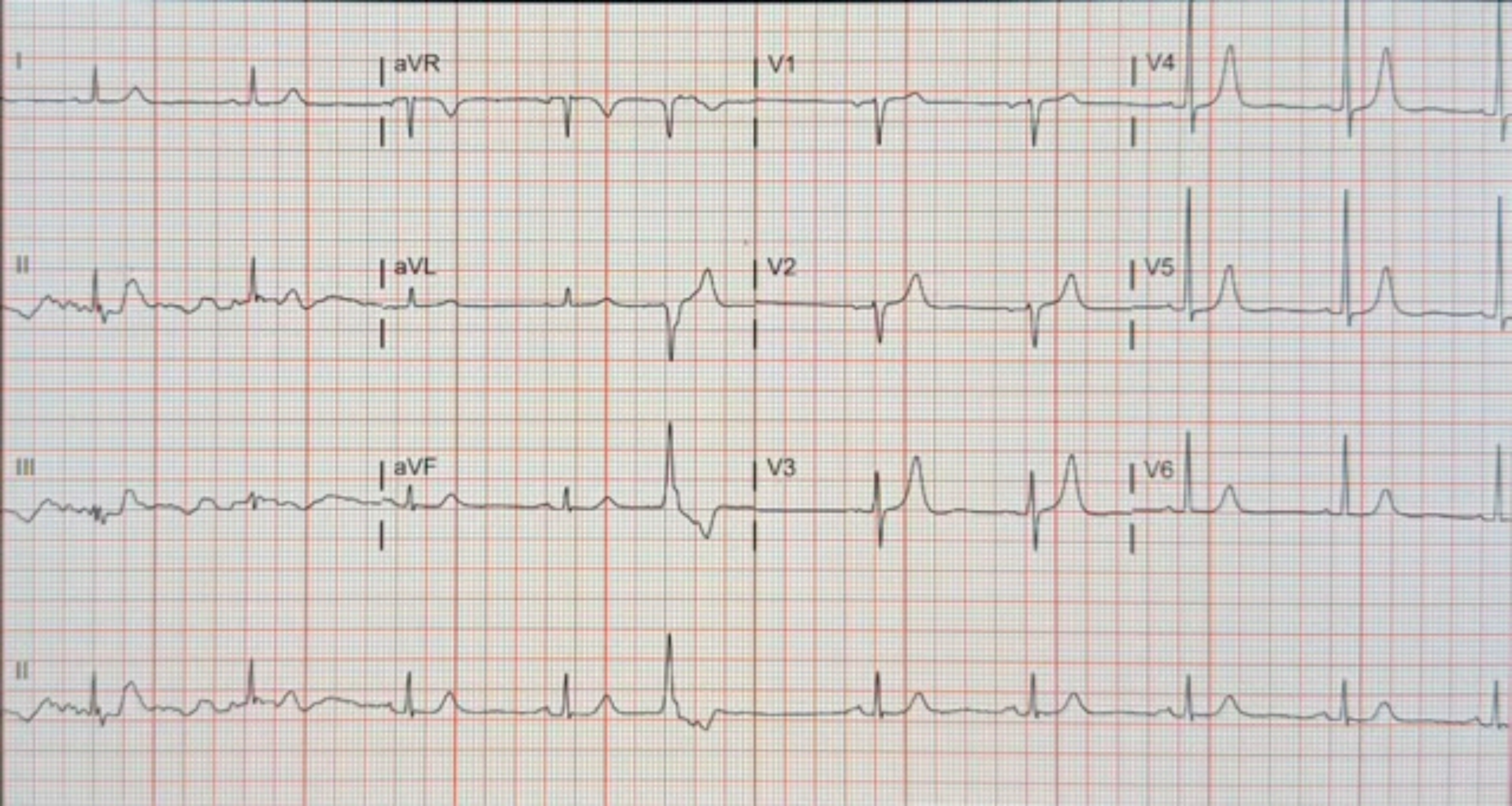
I
aVR
V1
V4
II
aVL
V2
V5
III
aVF
V3
V6
II

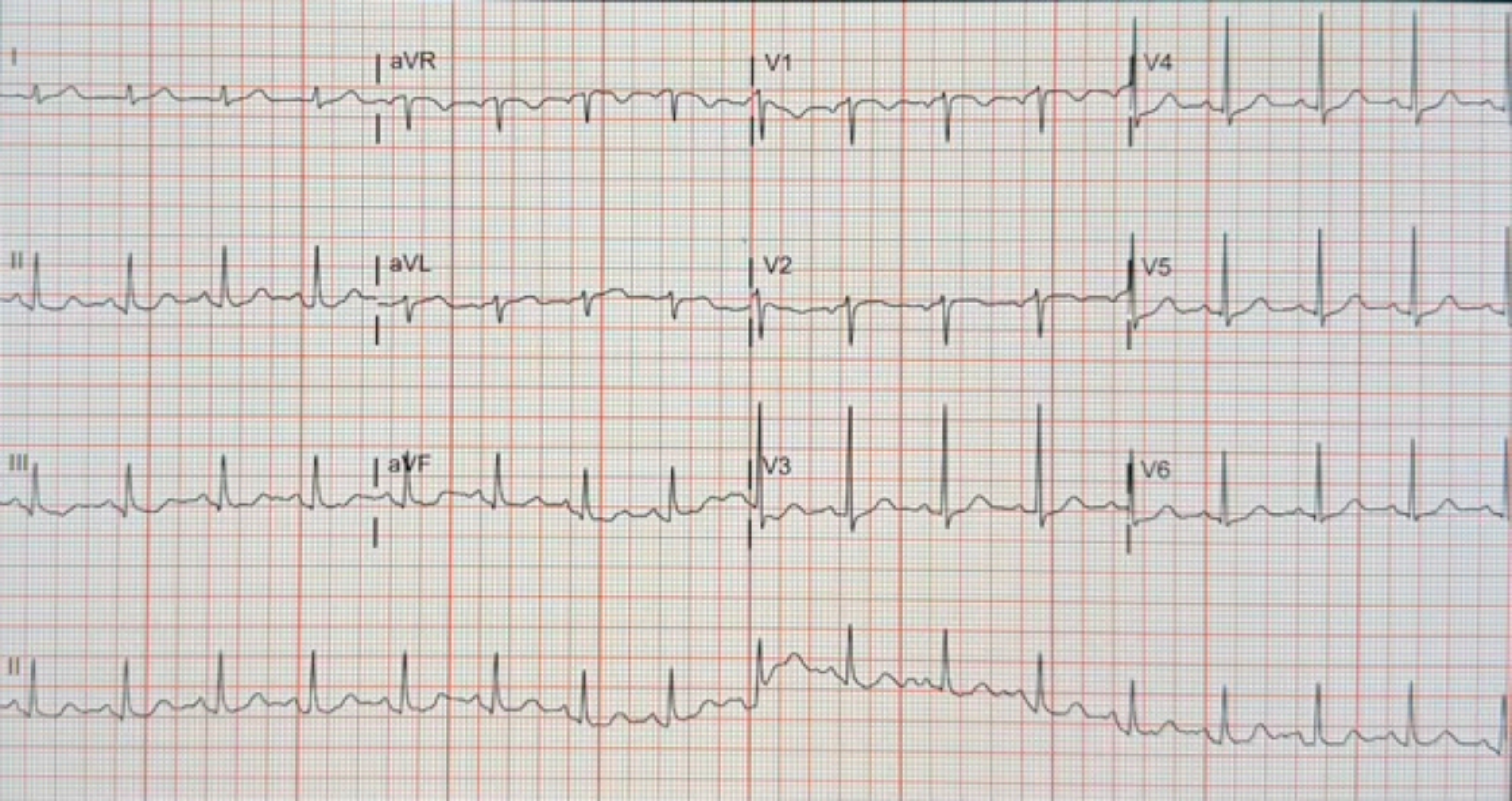
I
aVR
V1
V4
II
aVL
V2
V5
III
aVF
V3
V6
II

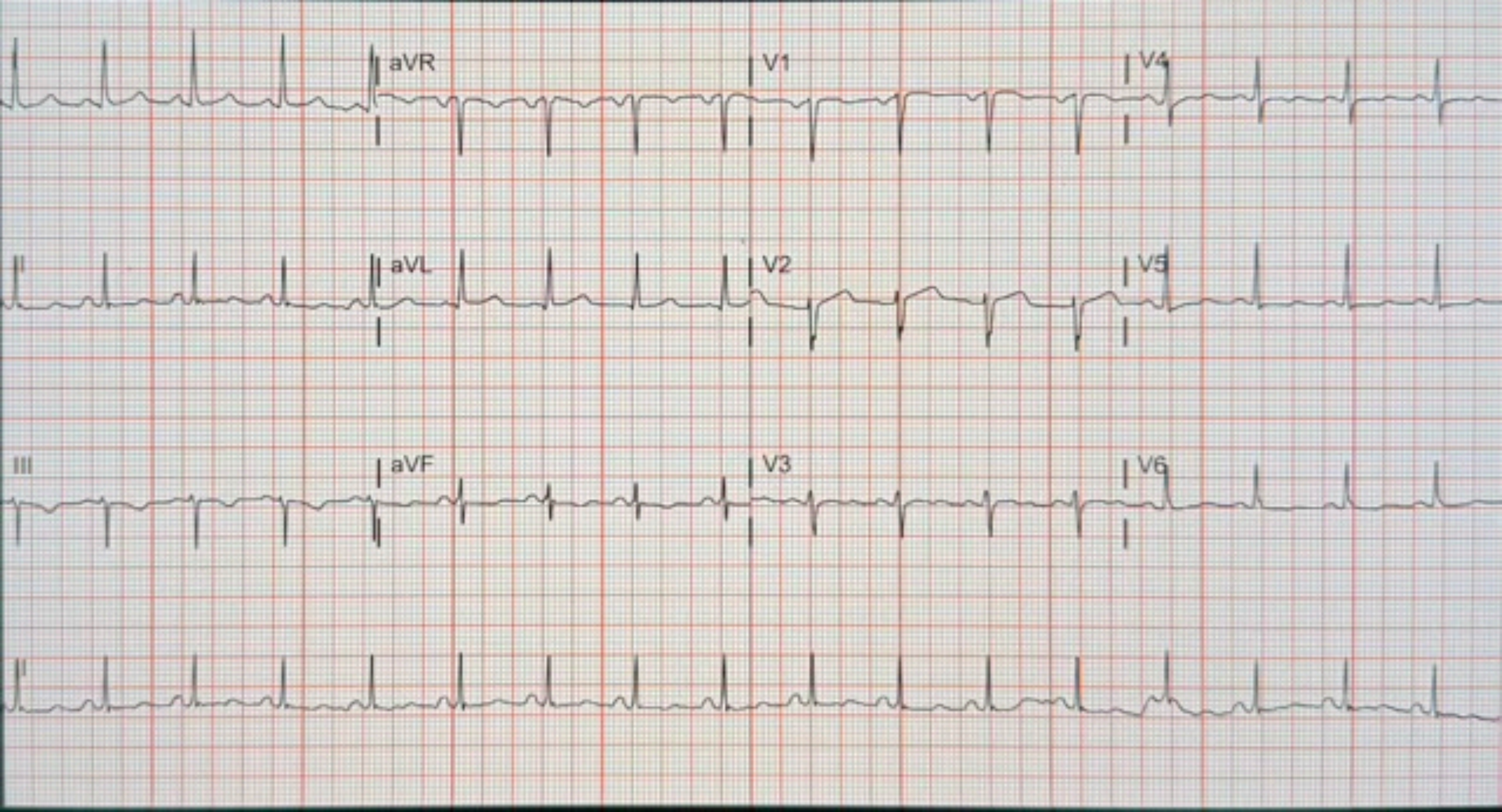
aVR
V1
V4
aVL
V2
V5
III
aVF
V3
V6

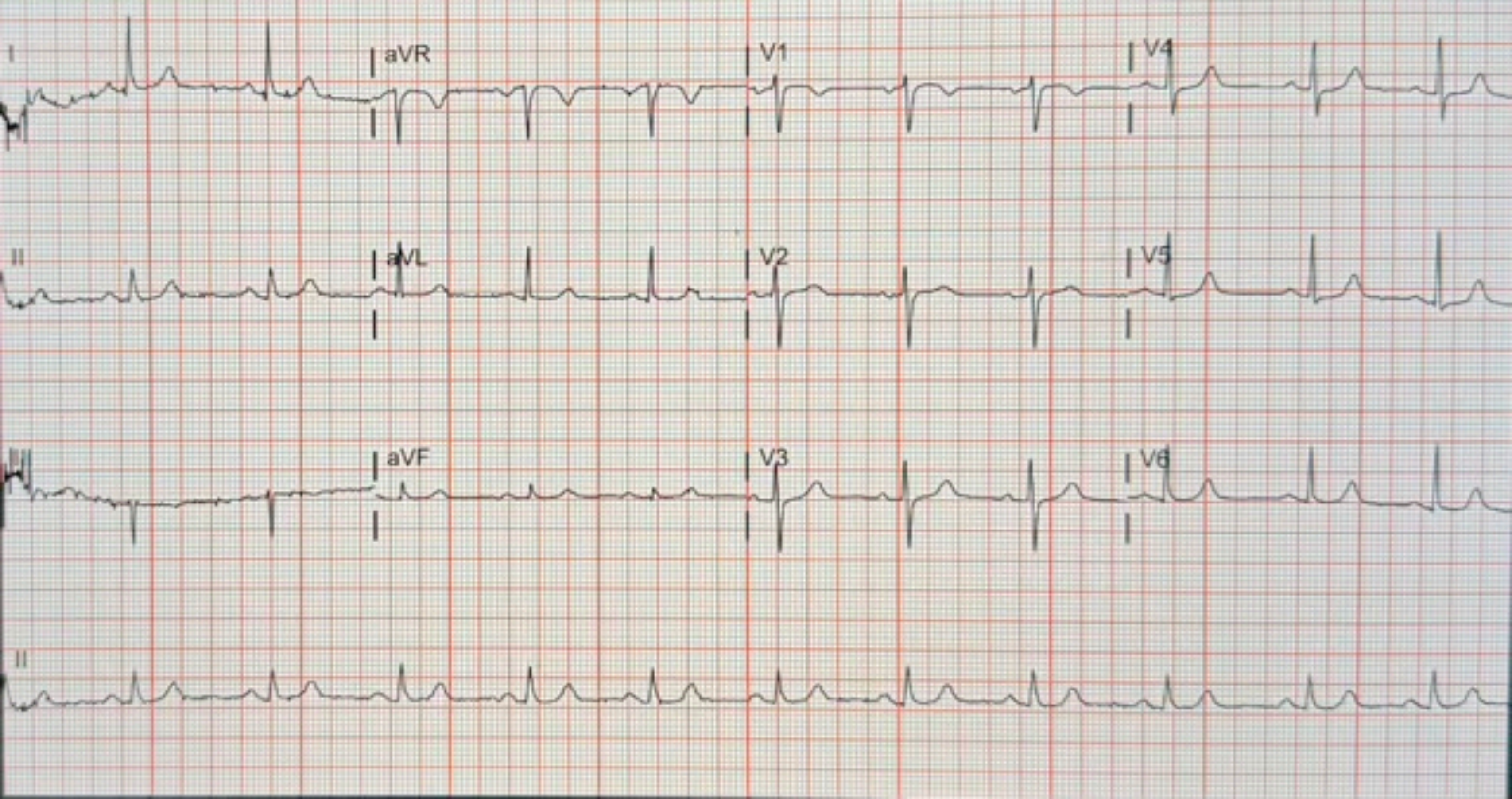
I
aVR
V1
V4
II
aVL
V2
V5
III
aVF
V3
V6
II

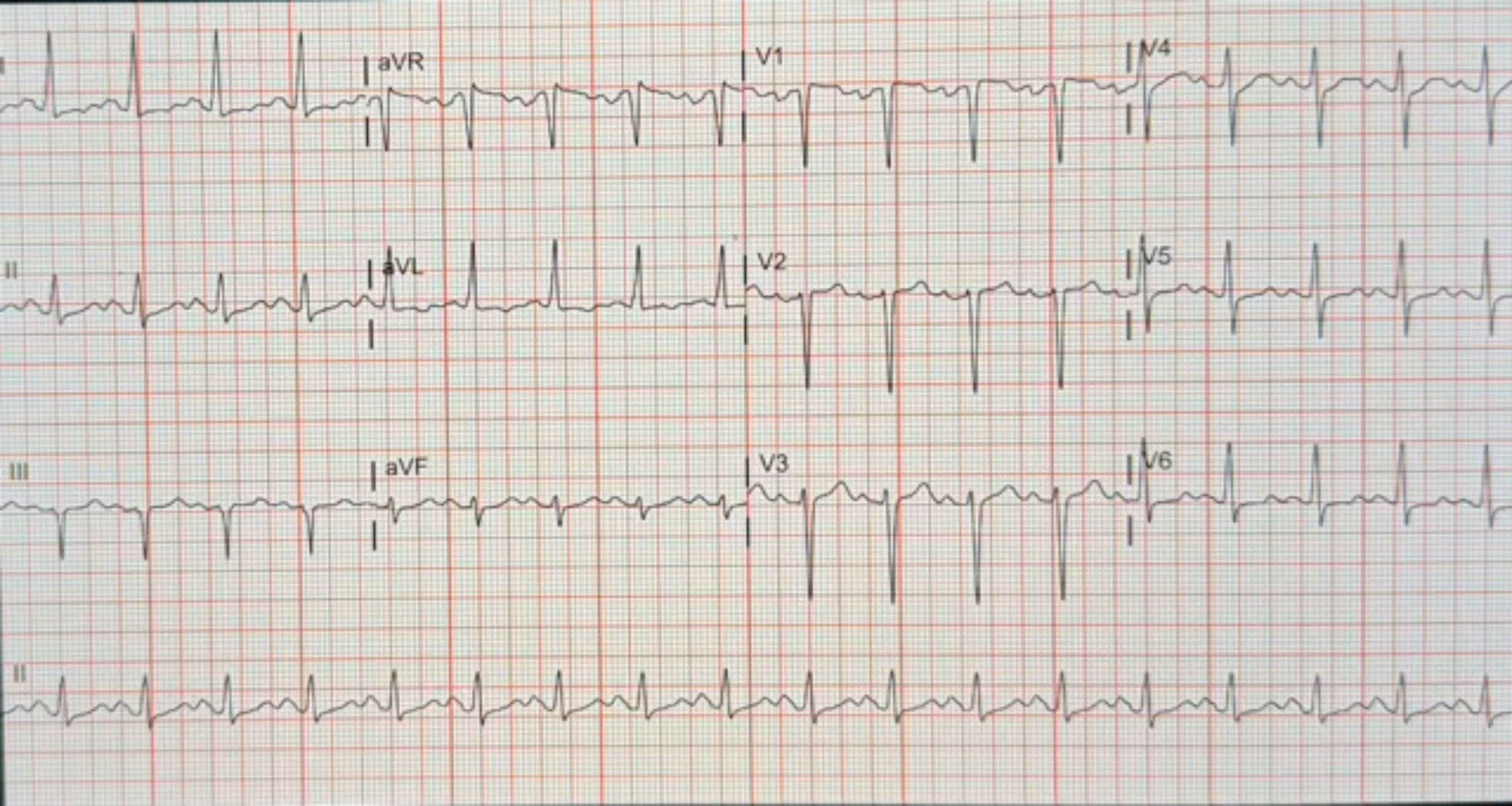
I
aVR
V1
V4
II
aVL
V2
V5
III
aVF
V3
V6
II

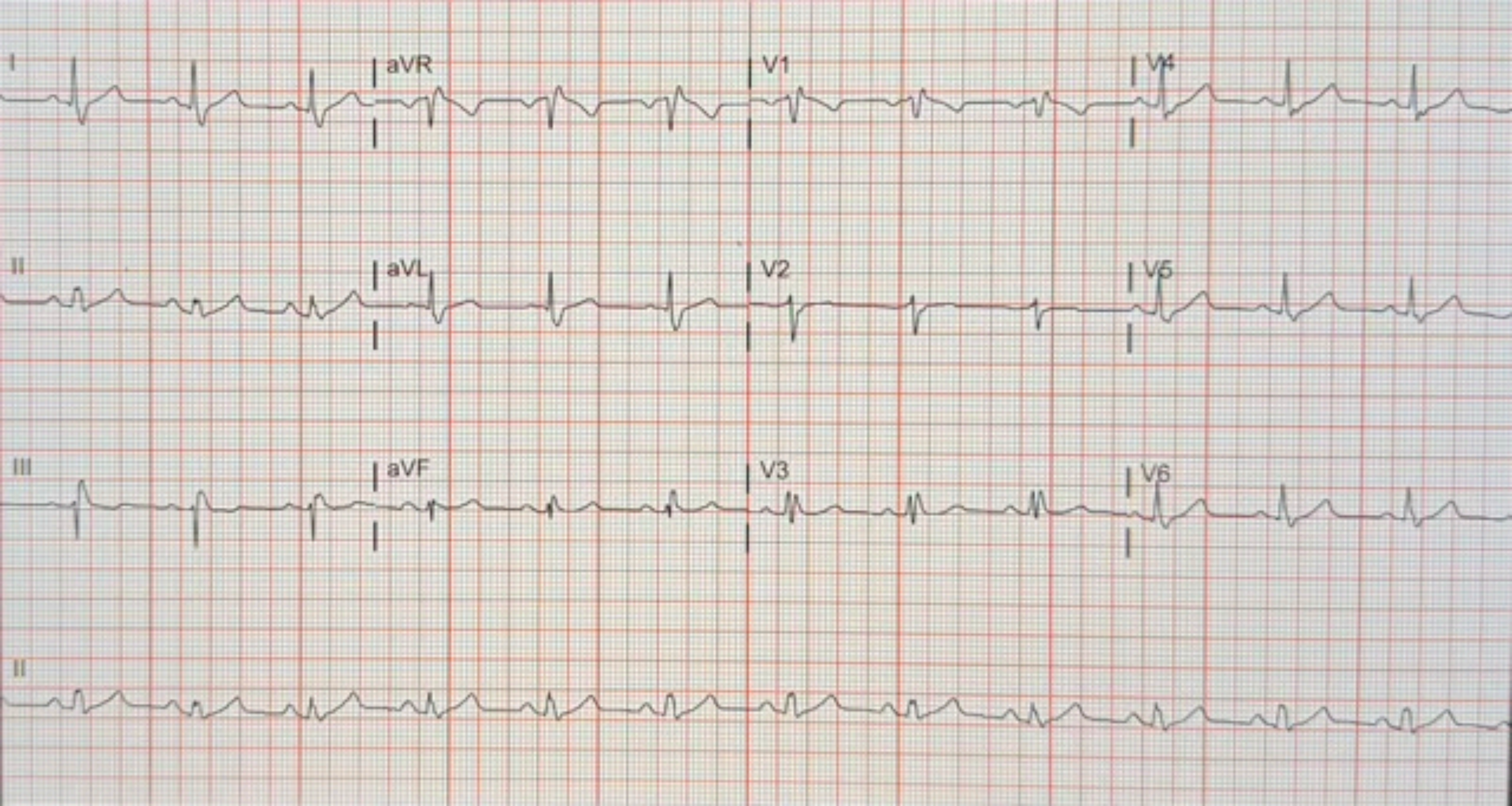

I
aVR
V1
V4
II
aVL
V2
V5
III
aVF
V3
V6
II

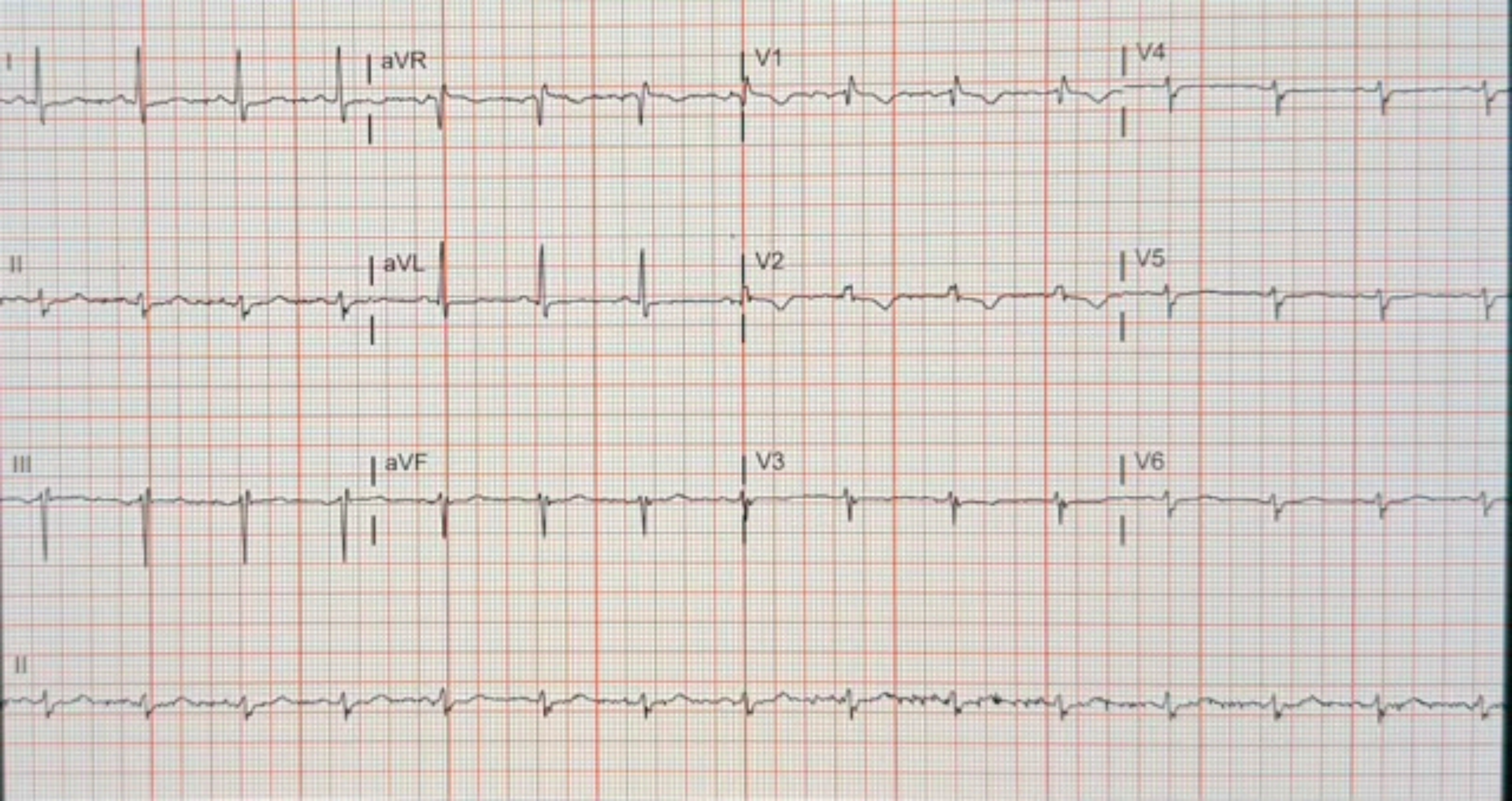

I
aVR
V1
V4
II
aVL
V2
V5
III
aVF
V3
V6
II

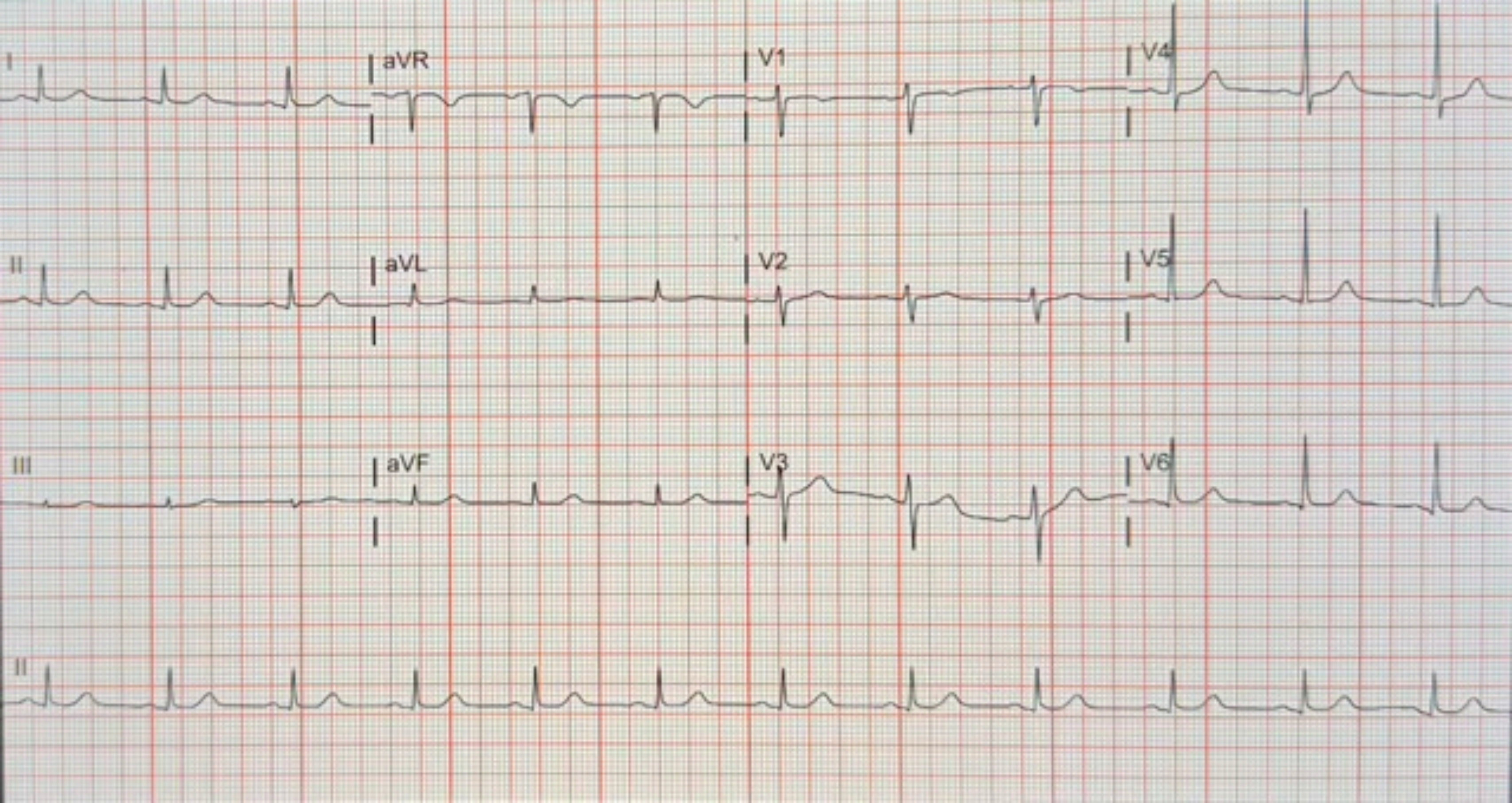
I
aVR
V1
V4
II
aVL
V2
V5
III
aVF
V3
V6
II

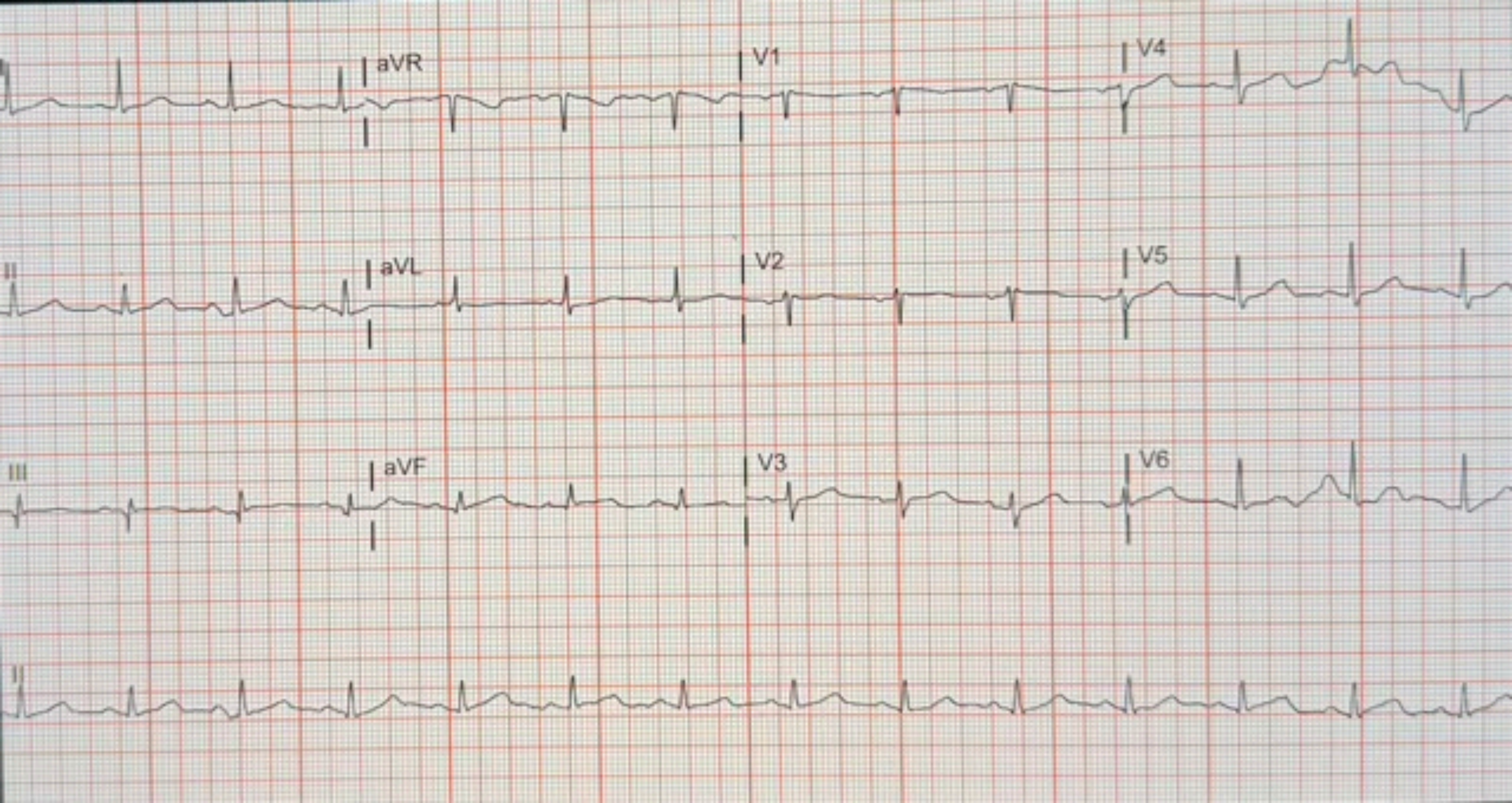
aVR
V1
V4
II
aVL
V2
V5
III
aVF
V3
V6
II

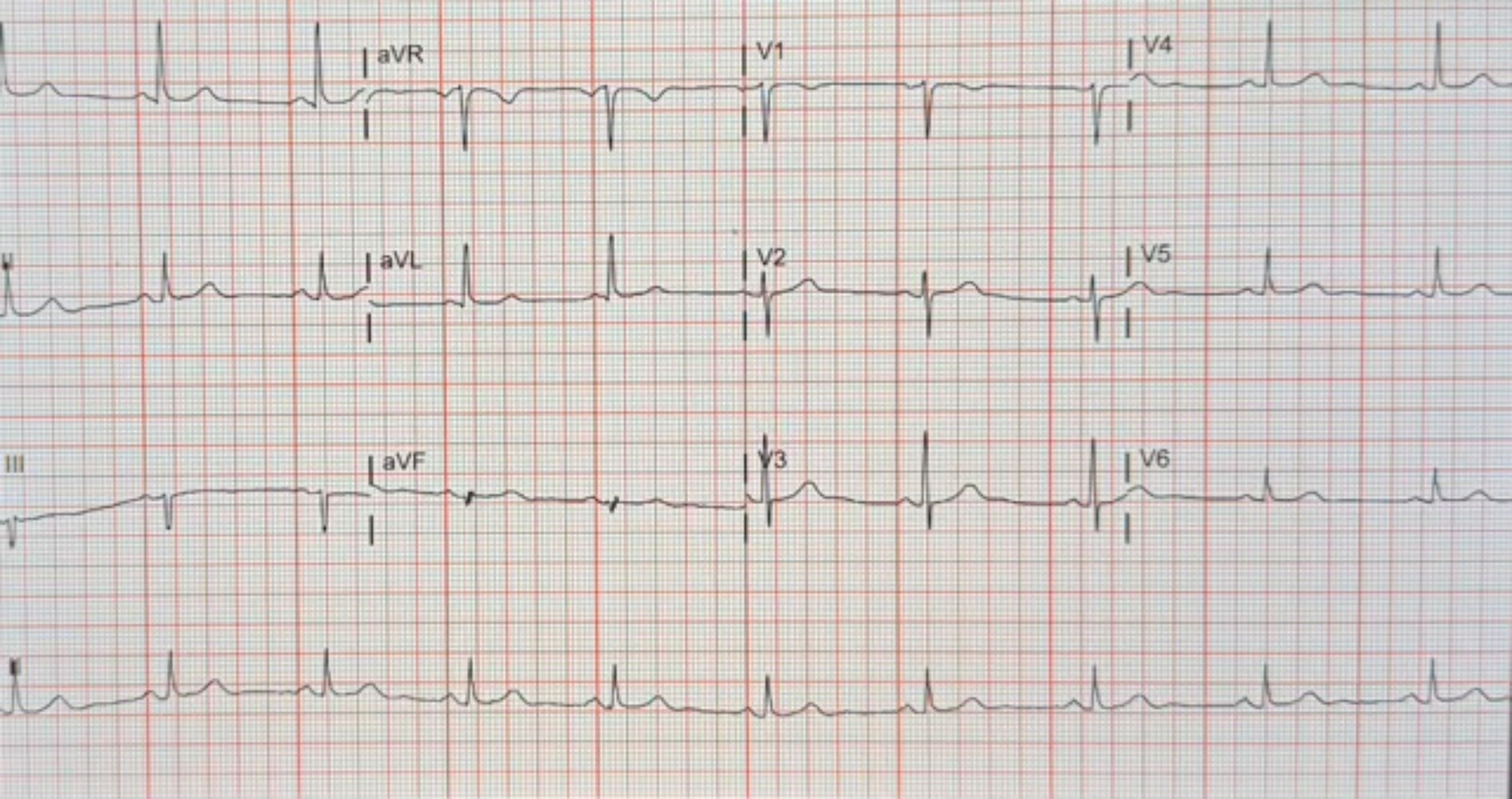

aVR
V1
V4
aVL
V2
V5
III
aVF
V3
V6

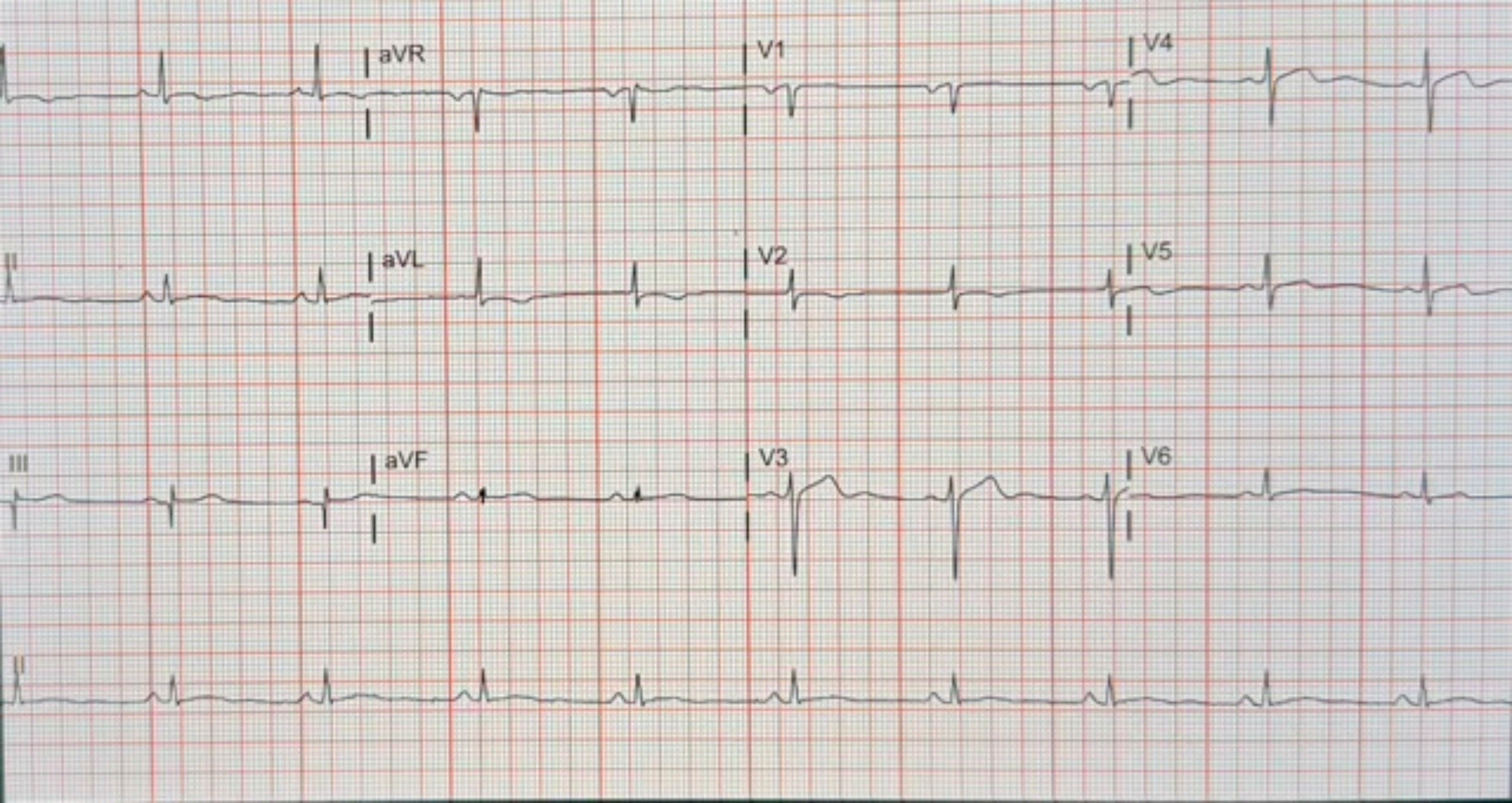
aVR
V1
V4
II
aVL
V2
V5
III
aVF
V3
V6
II

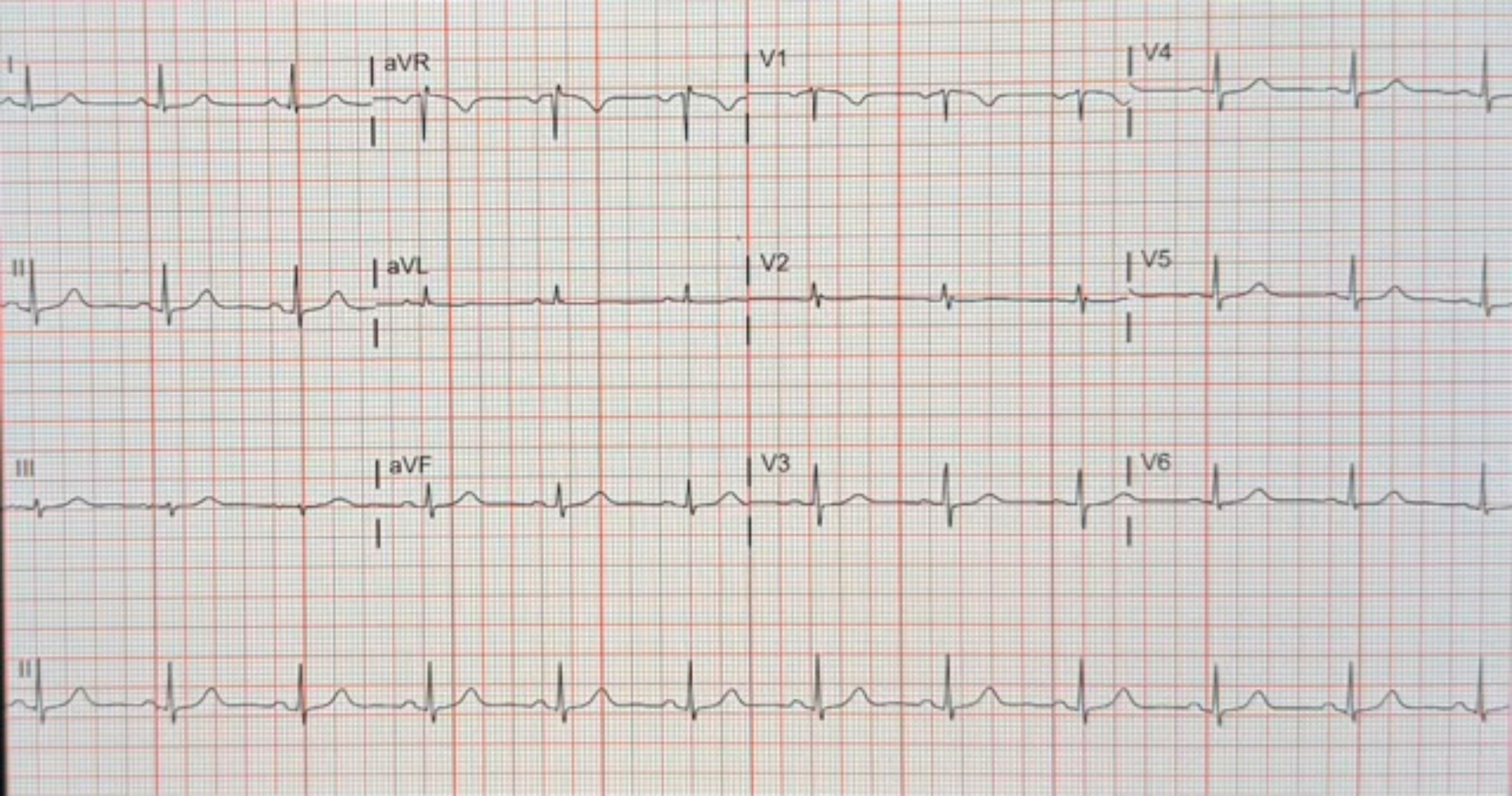
I
aVR
V1
V4
II
aVL
V2
V5
III
aVF
V3
V6
II

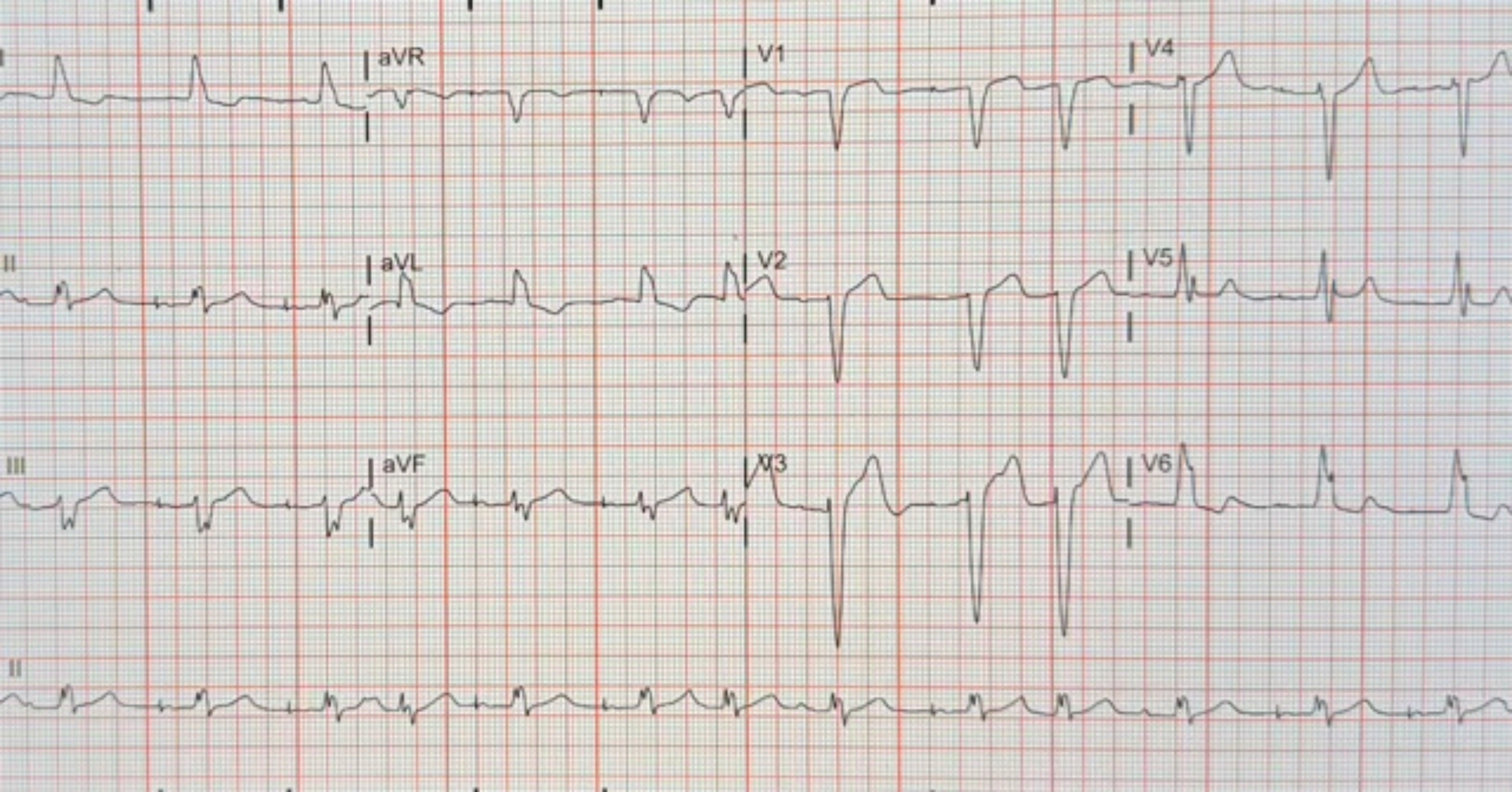
I
aVR
V1
V4
II
aVL
V2
V5
III
aVF
V3
V6
II

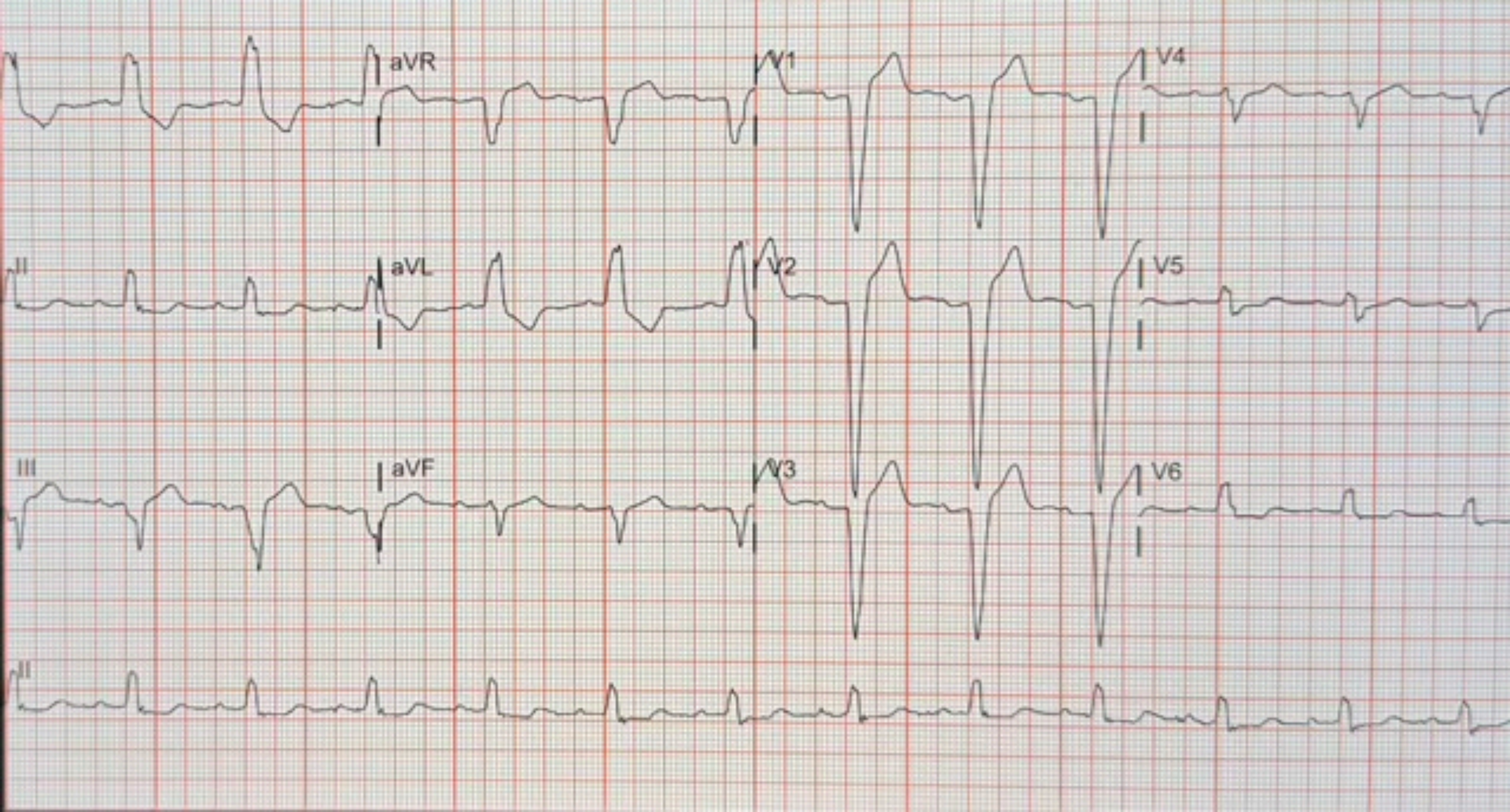

aVR
V1
V4
II
aVL
V2
V5
III
aVF
V3
V6
II

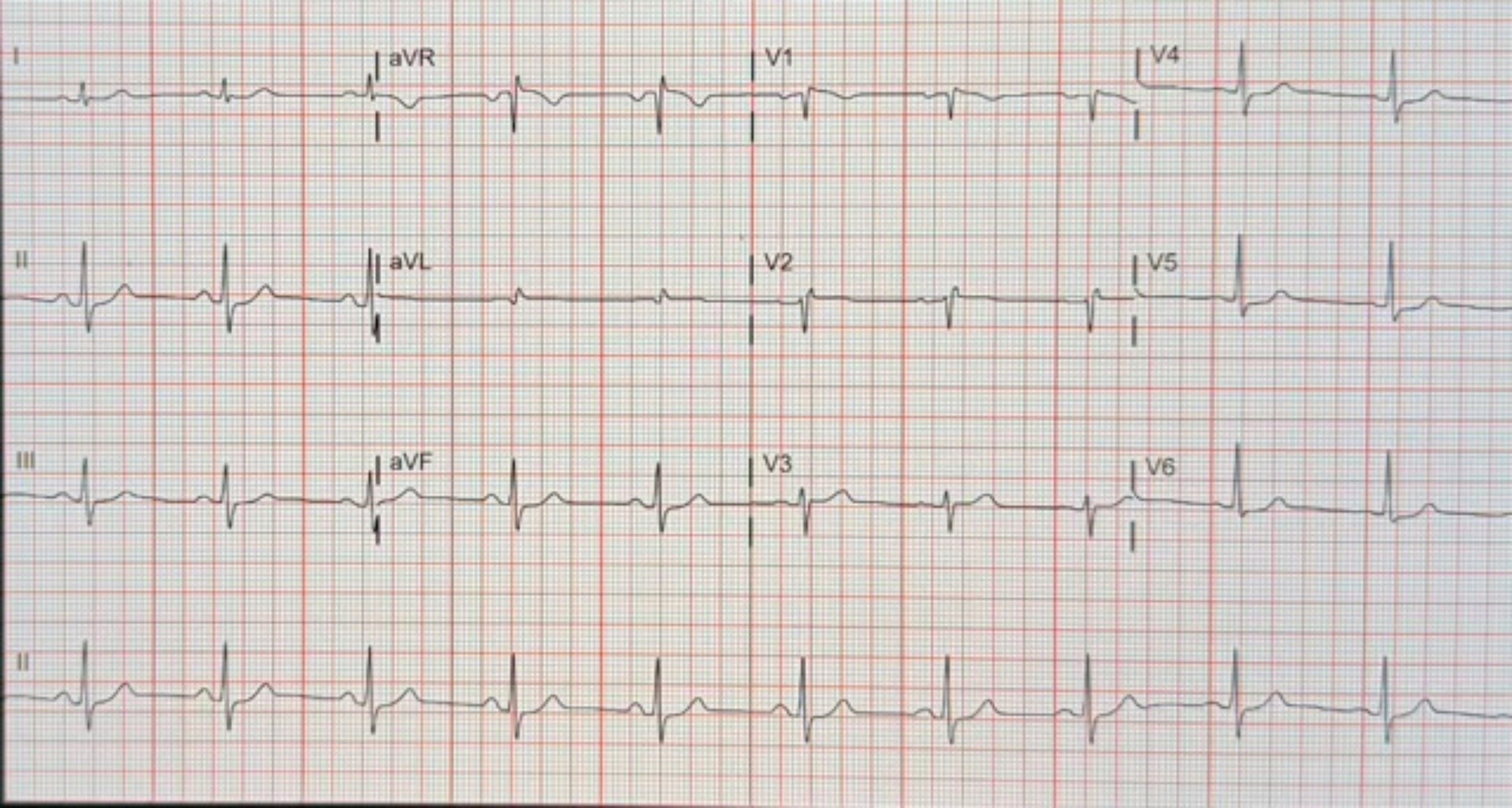
I
aVR
V1
V4
II
aVL
V2
V5
III
aVF
V3
V6
II

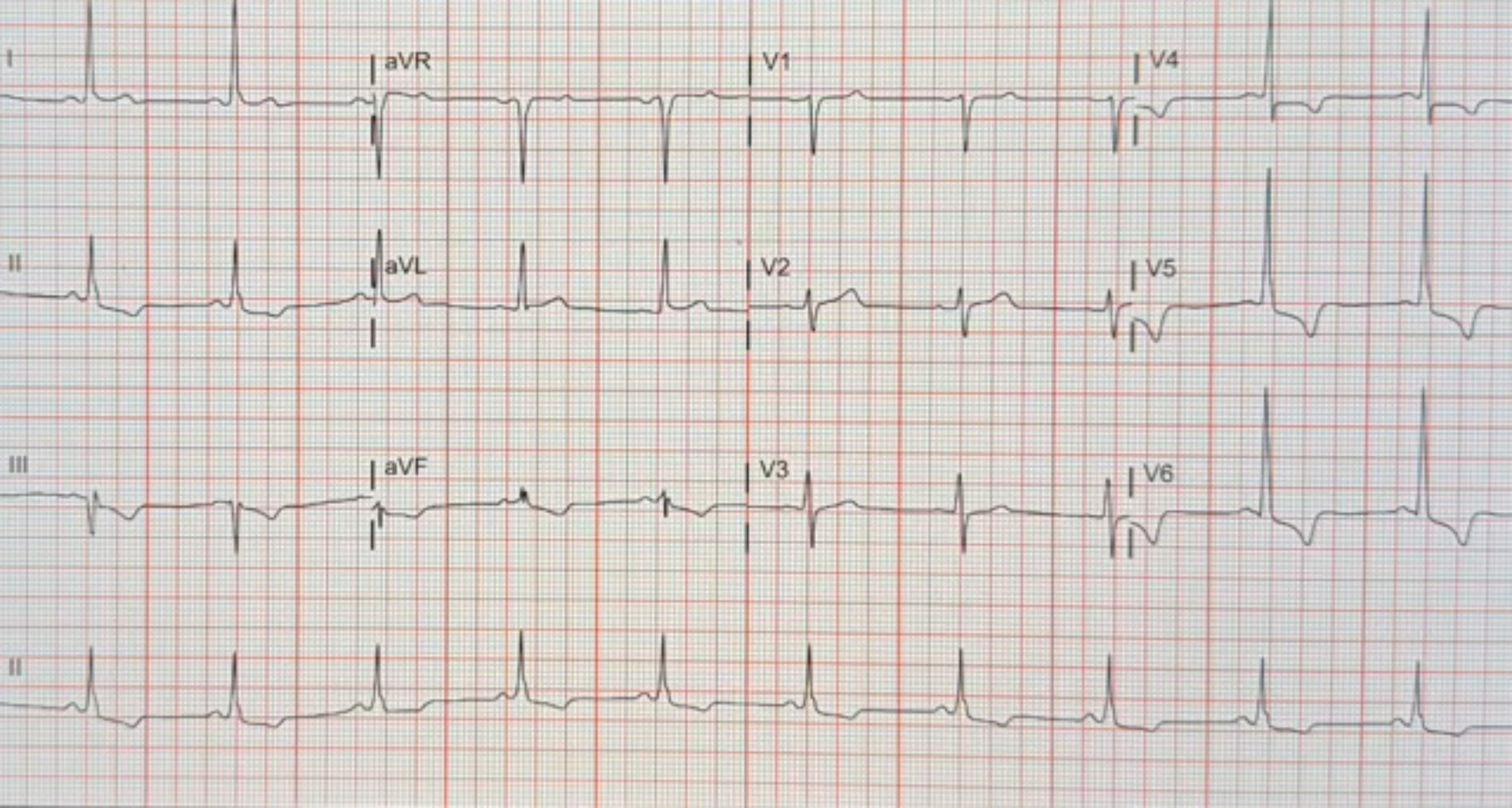

I
aVR
V1
V4
II
aVL
V2
V5
III
aVF
V3
V6
II

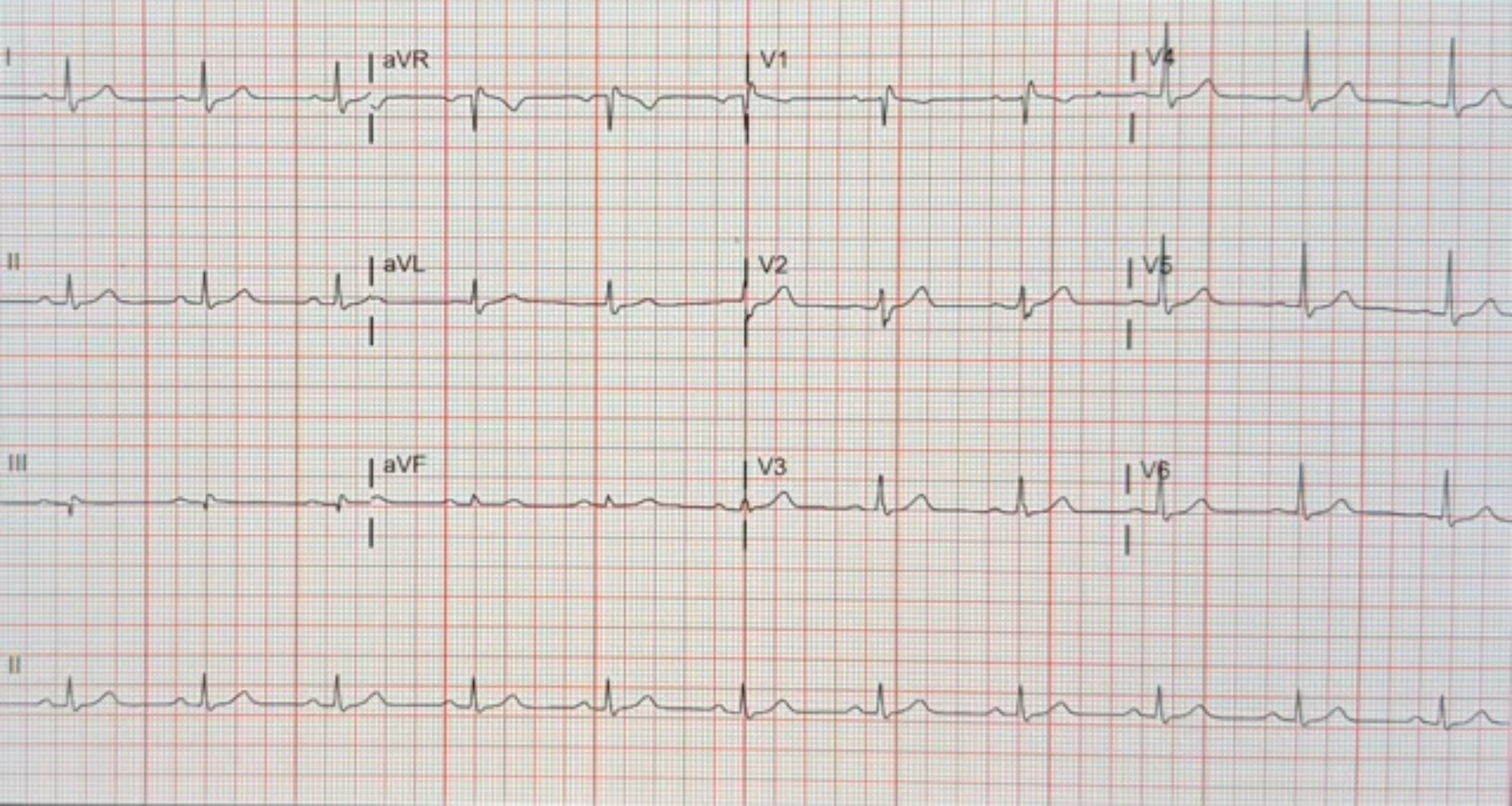
I
aVR
V1
V4
II
aVL
V2
V5
III
aVF
V3
V6
II

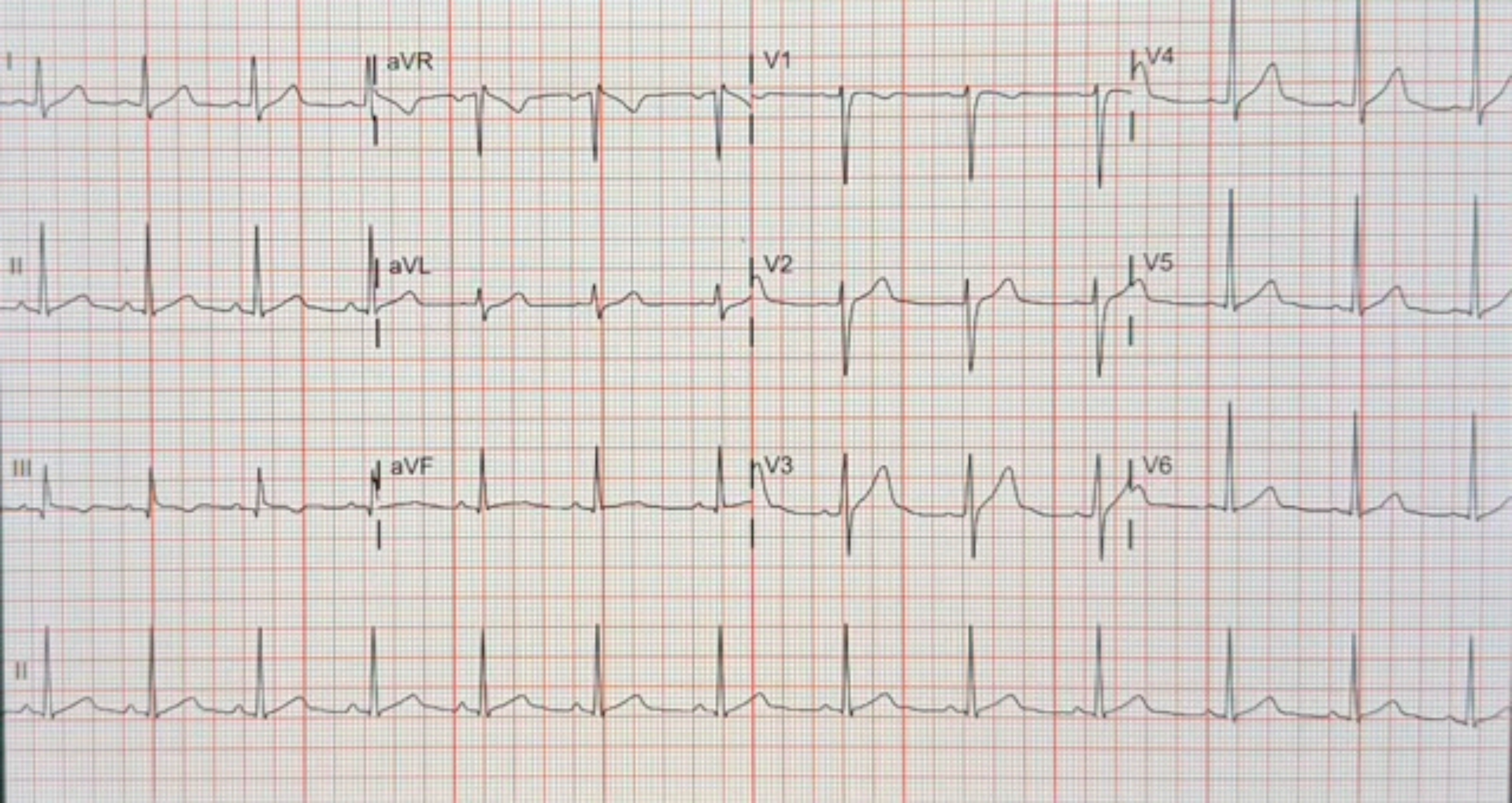
I
aVR
V1
V4
II
aVL
V2
V5
III
aVF
V3
V6
II

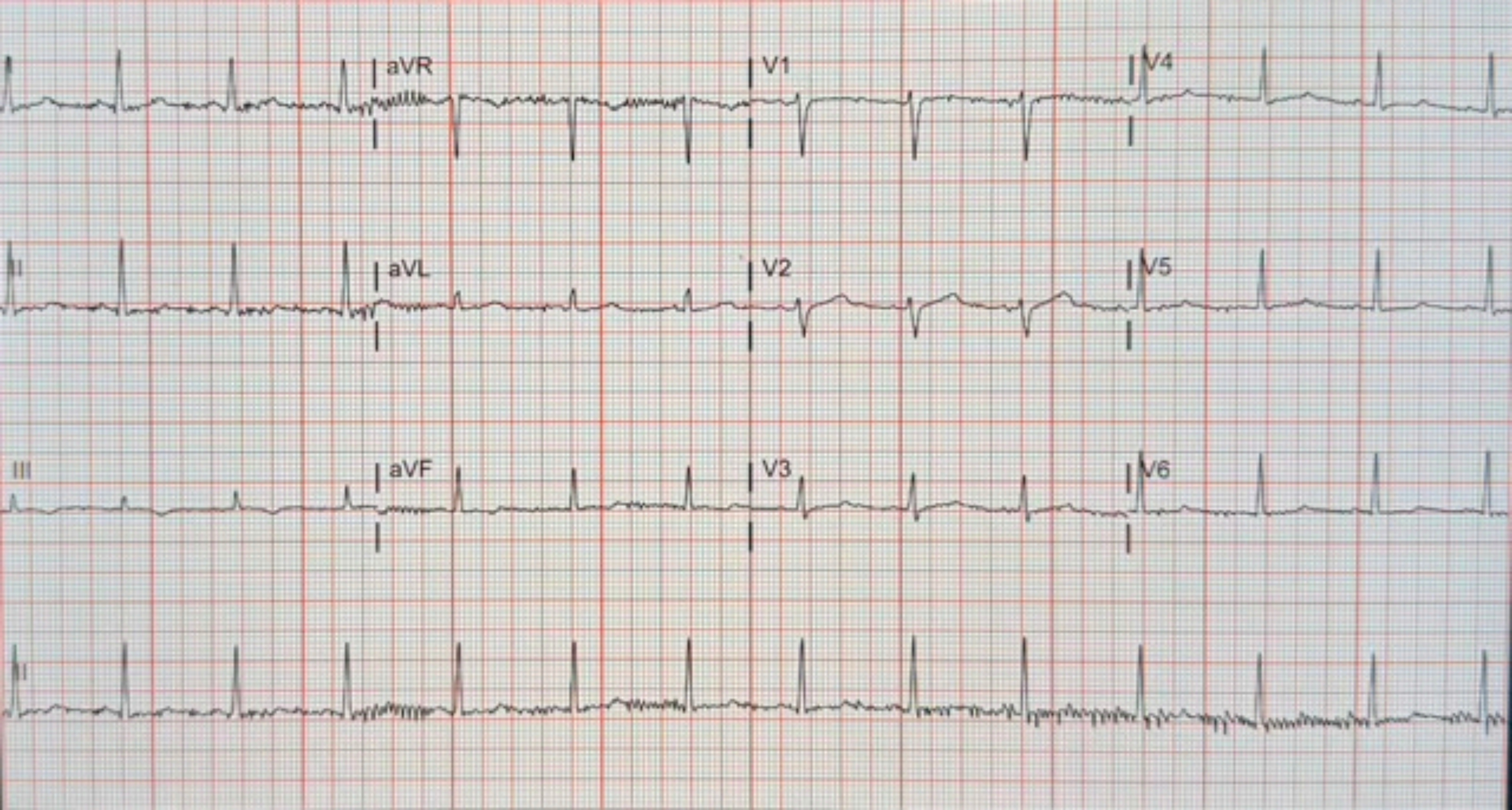
aVR
V1
V4
II
aVL
V2
V5
III
aVF
V3
V6
II

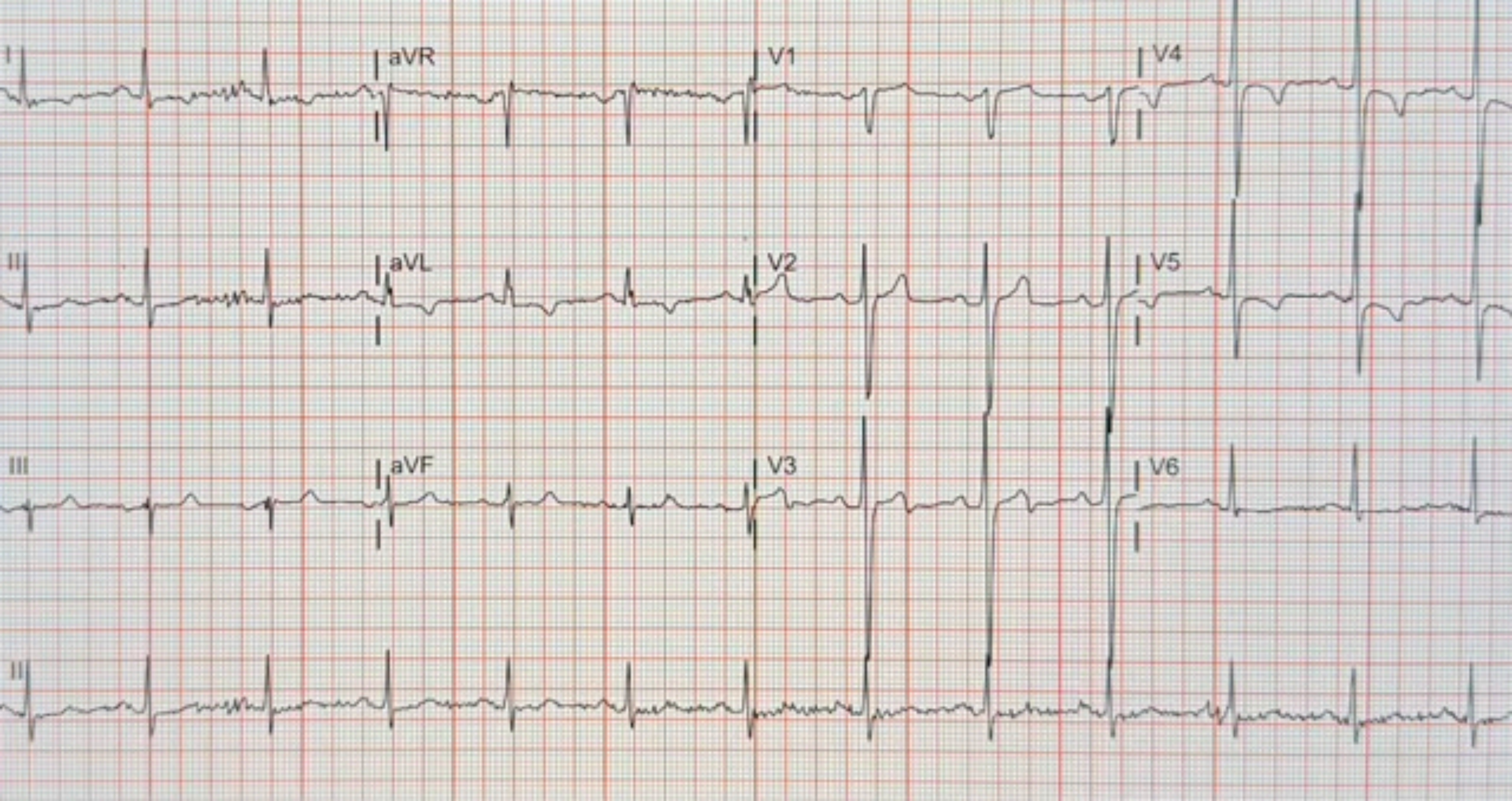

I
aVR
V1
V4
II
aVL
V2
V5
III
aVF
V3
V6
II

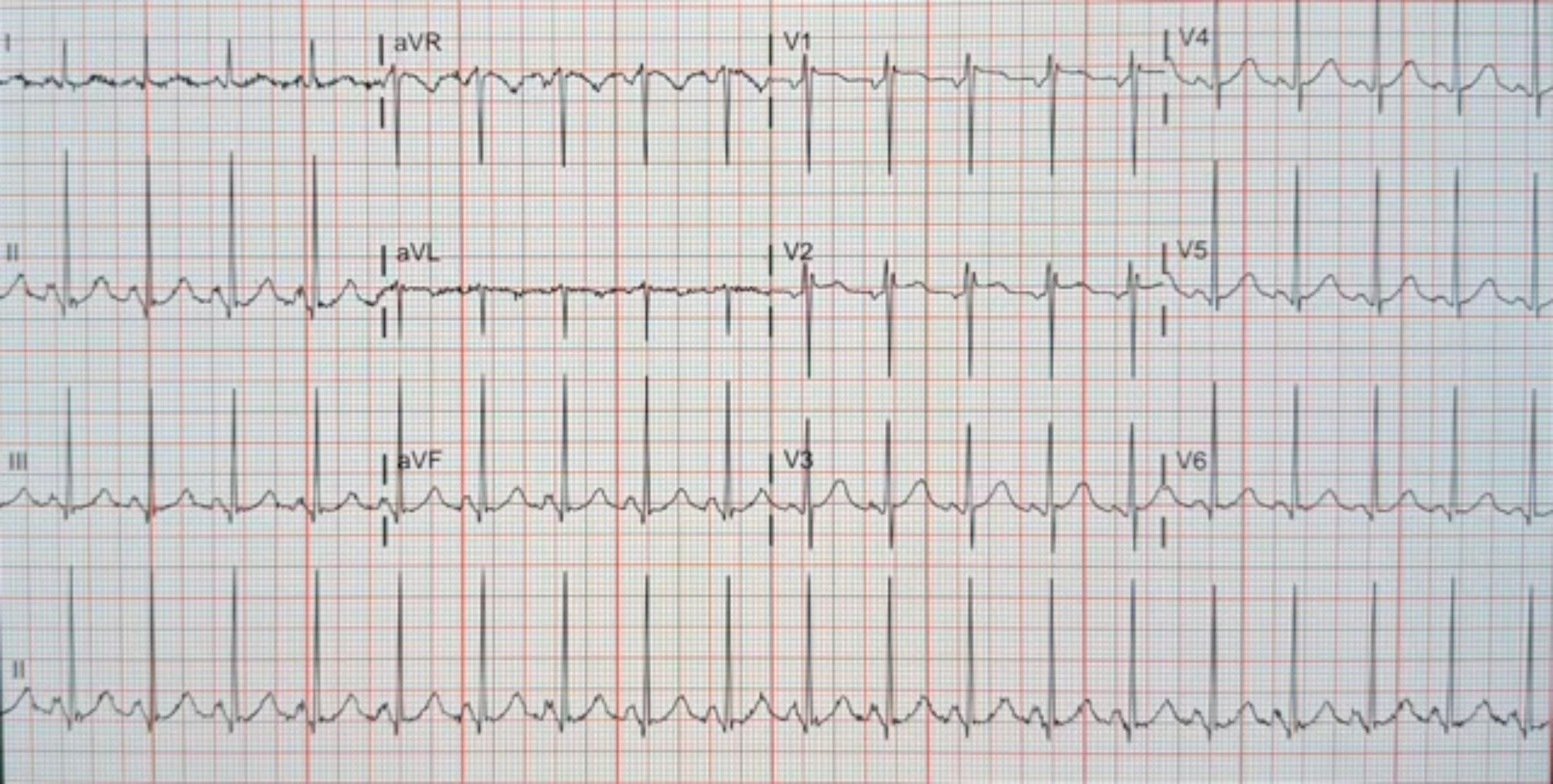
I
aVR
V1
V4
II
aVL
V2
V5
III
aVF
V3
V6
II

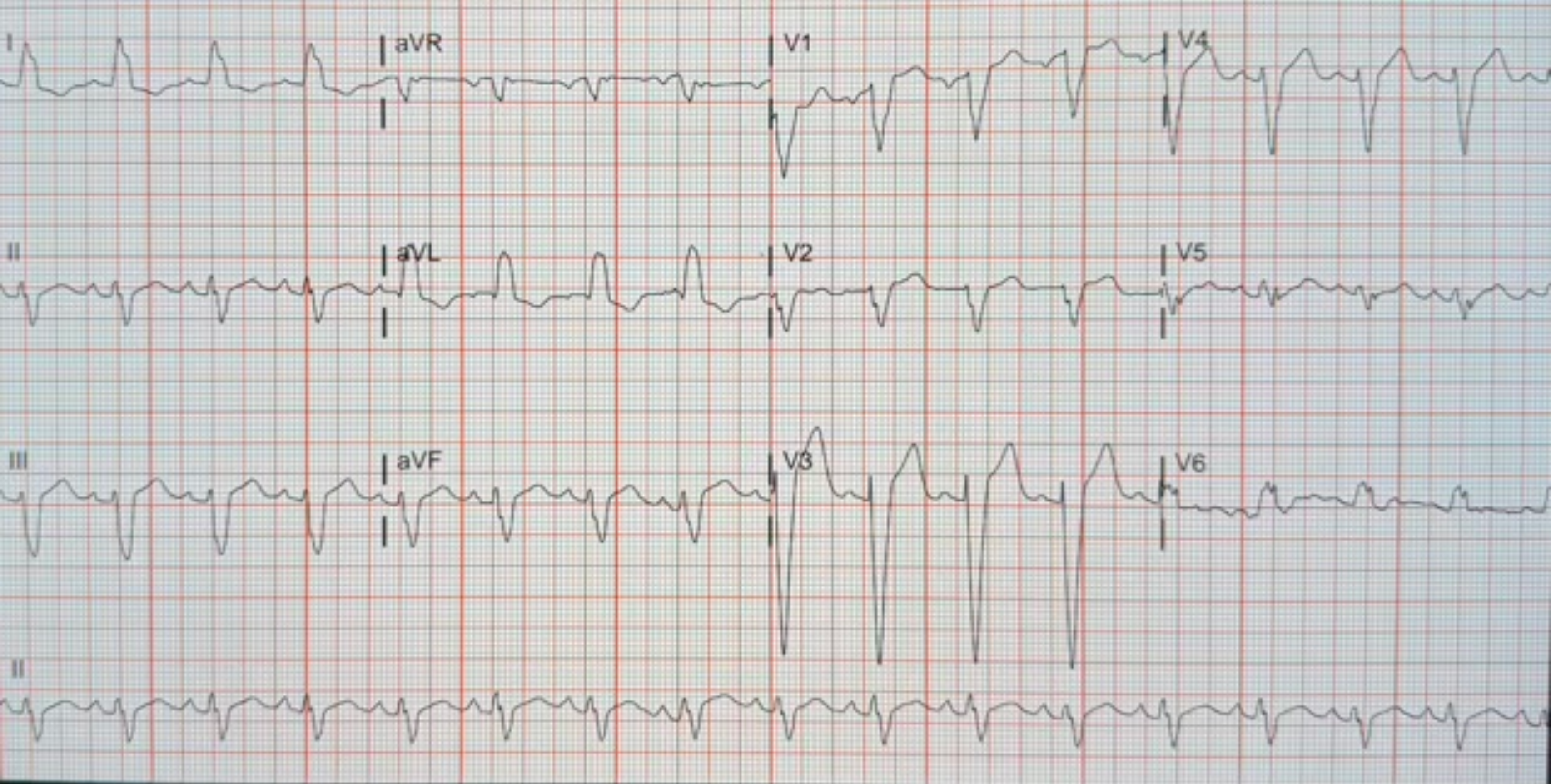
I
aVR
V1
V4
II
aVL
V2
V5
III
aVF
V3
V6
II

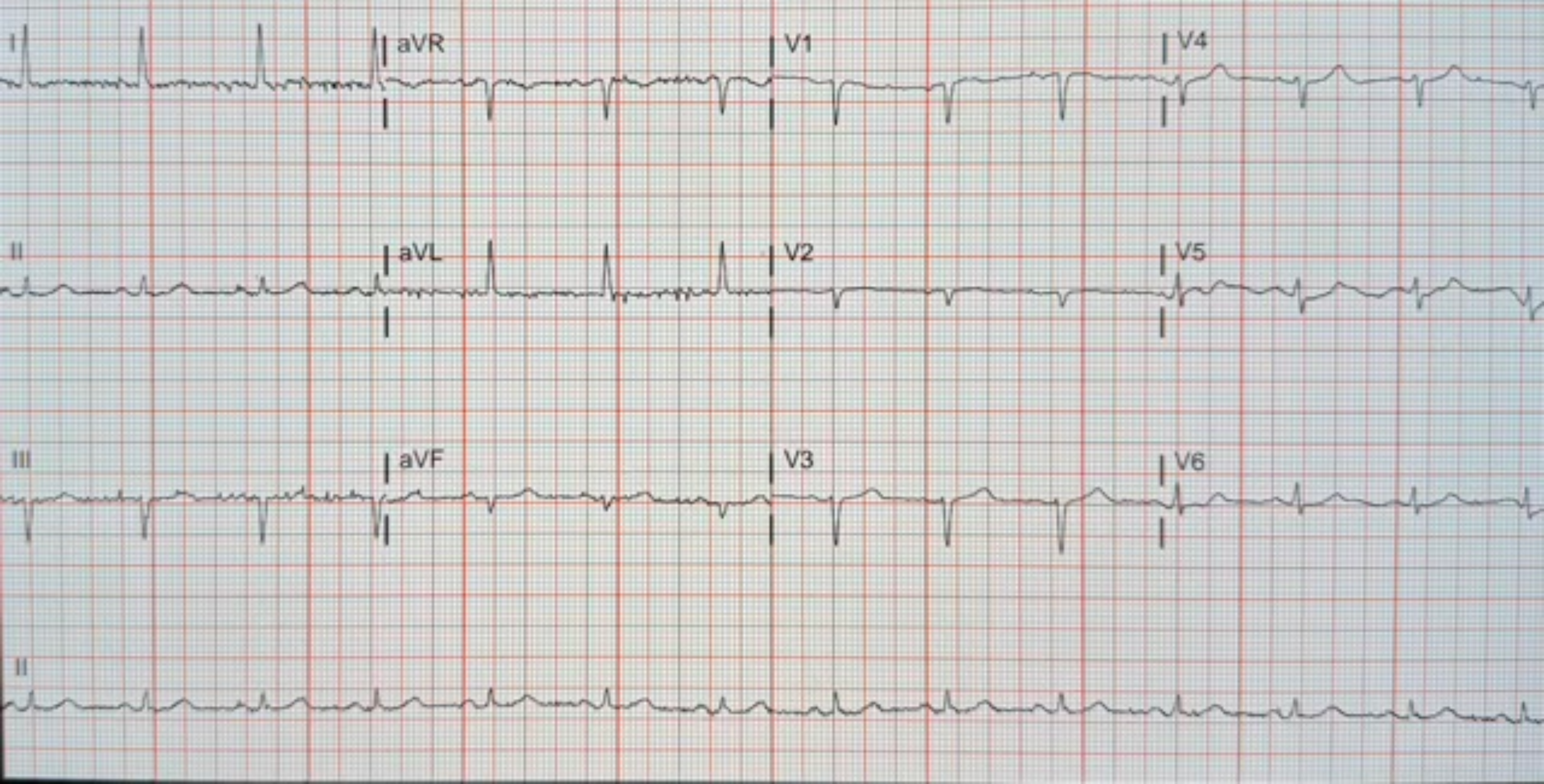
I
aVR
V1
V4
II
aVL
V2
V5
III
aVF
V3
V6
II

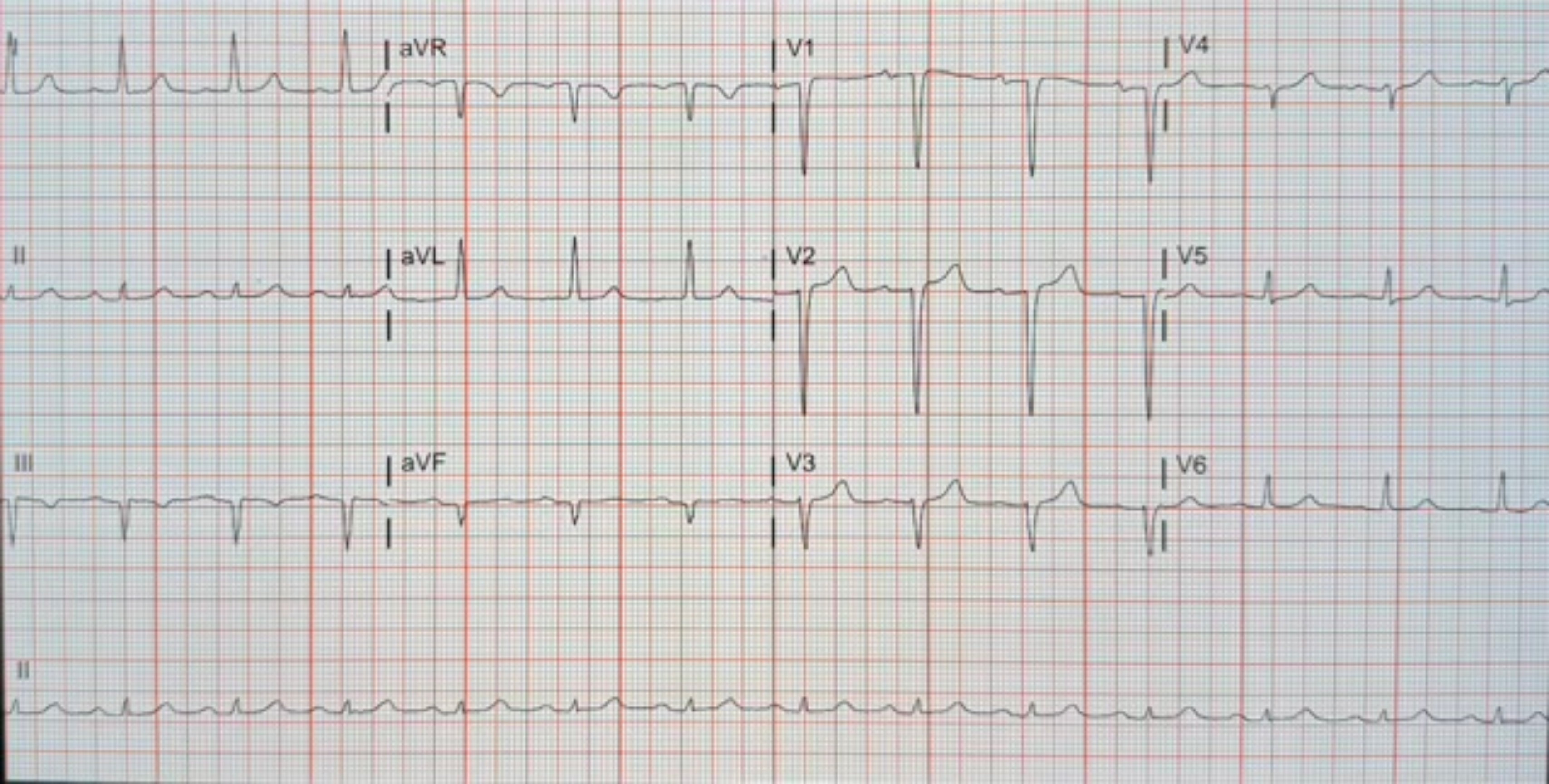
aVR
V1
V4
II
aVL
V2
V5
III
aVF
V3
V6
II

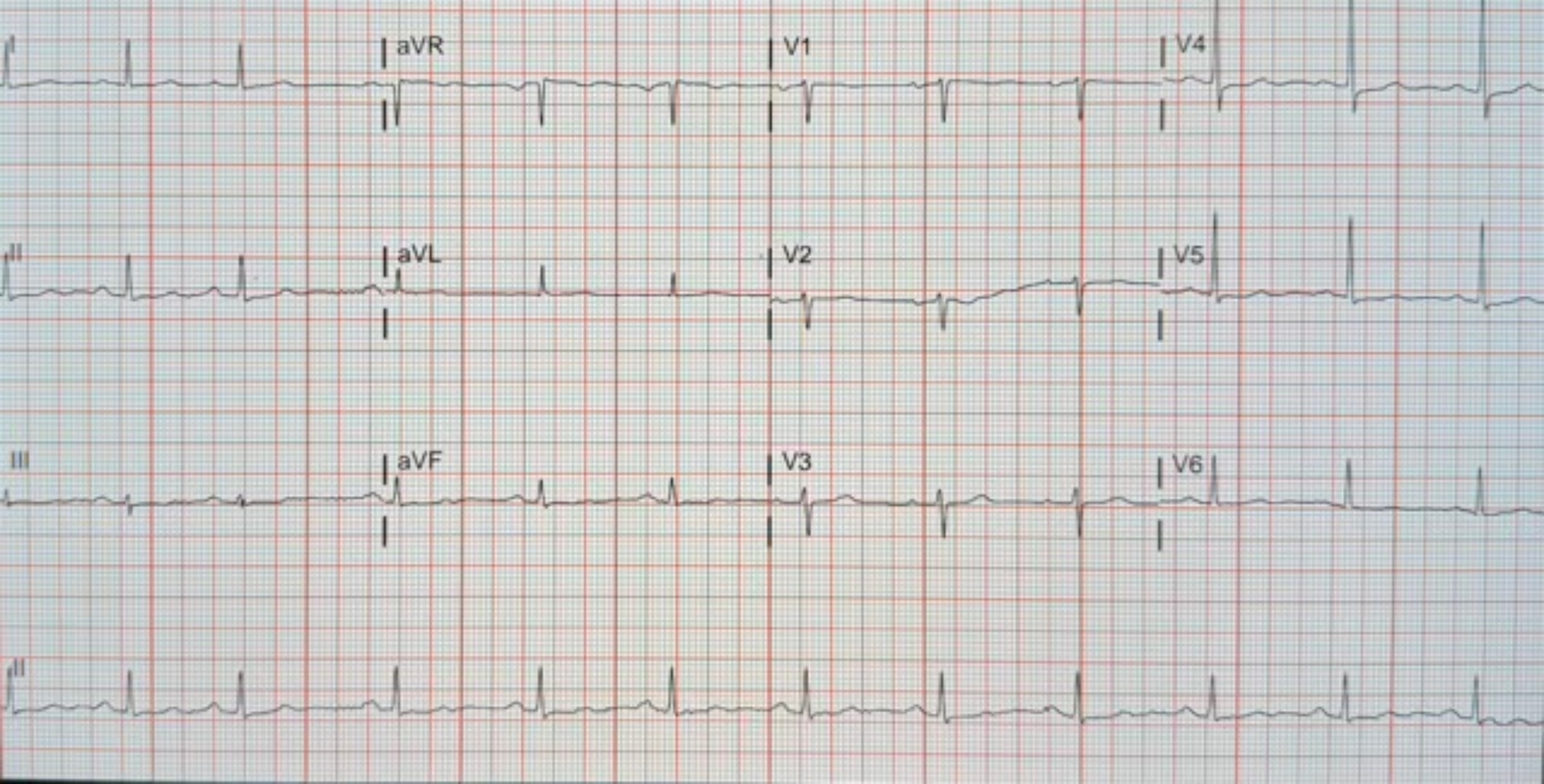
I
aVR
V1
V4
II
aVL
V2
V5
III
aVF
V3
V6
II

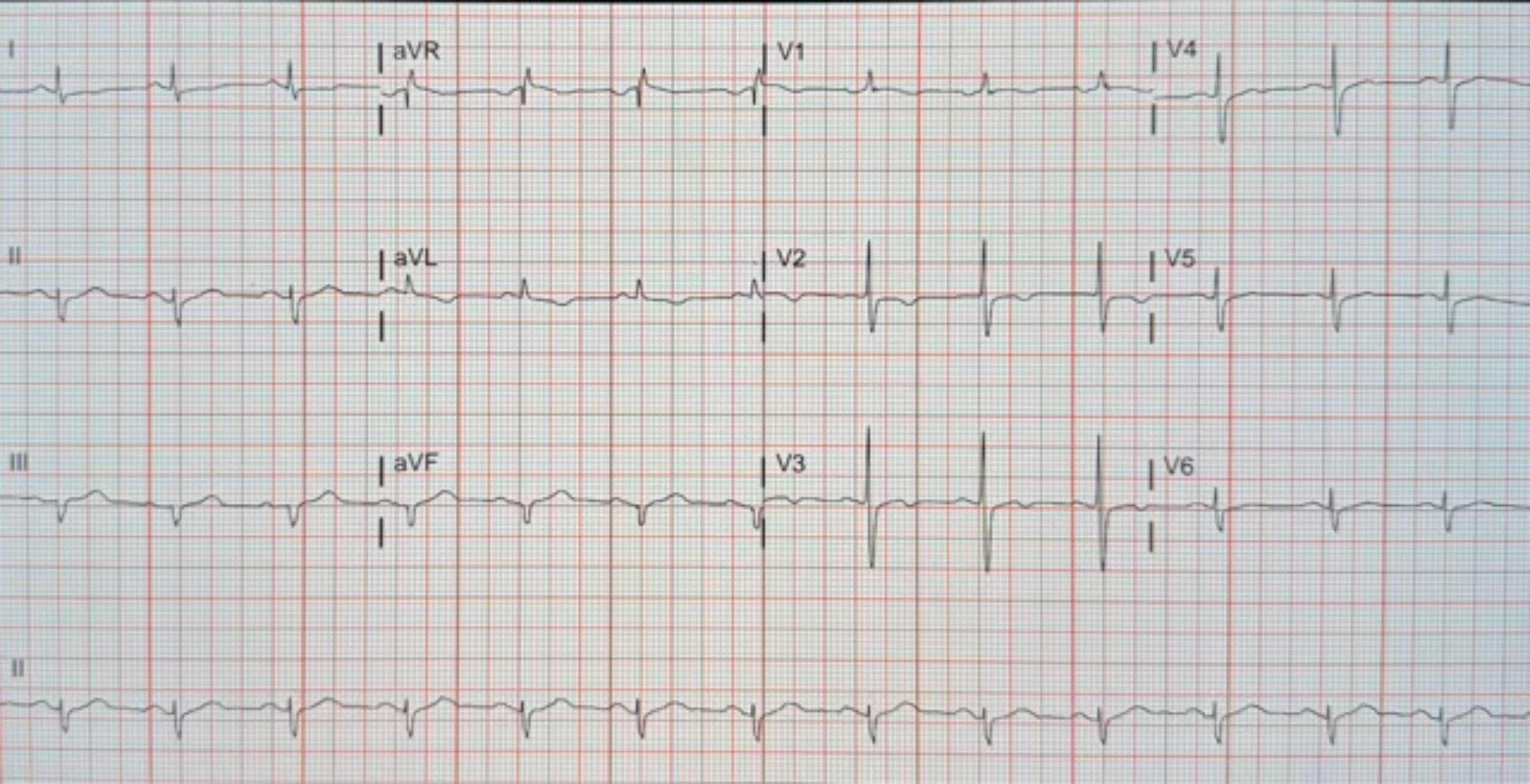
I
aVR
V1
V4
II
aVL
V2
V5
III
aVF
V3
V6
II

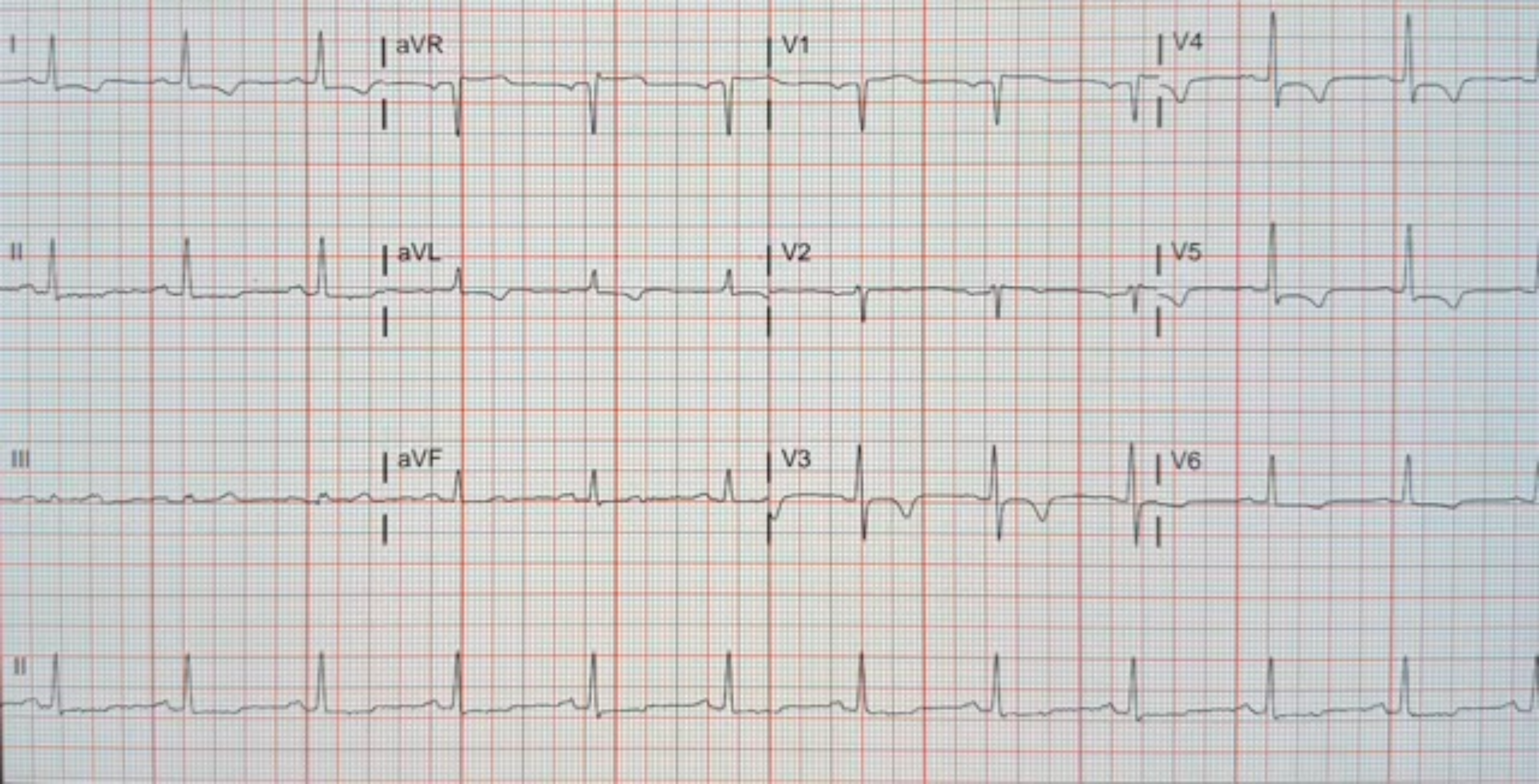
I
aVR
V1
V4
II
aVL
V2
V5
III
aVF
V3
V6
II

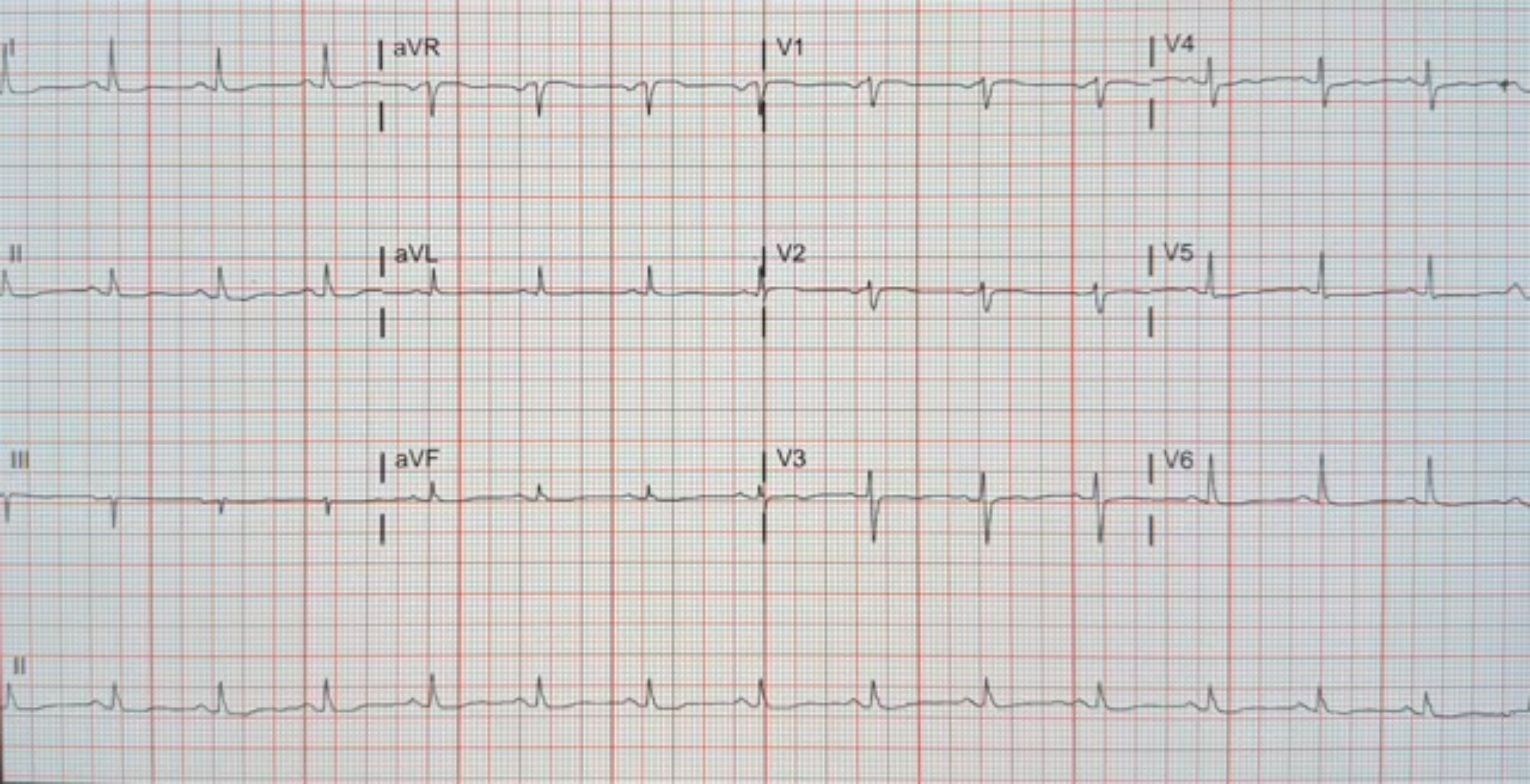
I
aVR
V1
V4
II
aVL
V2
V5
III
aVF
V3
V6
II

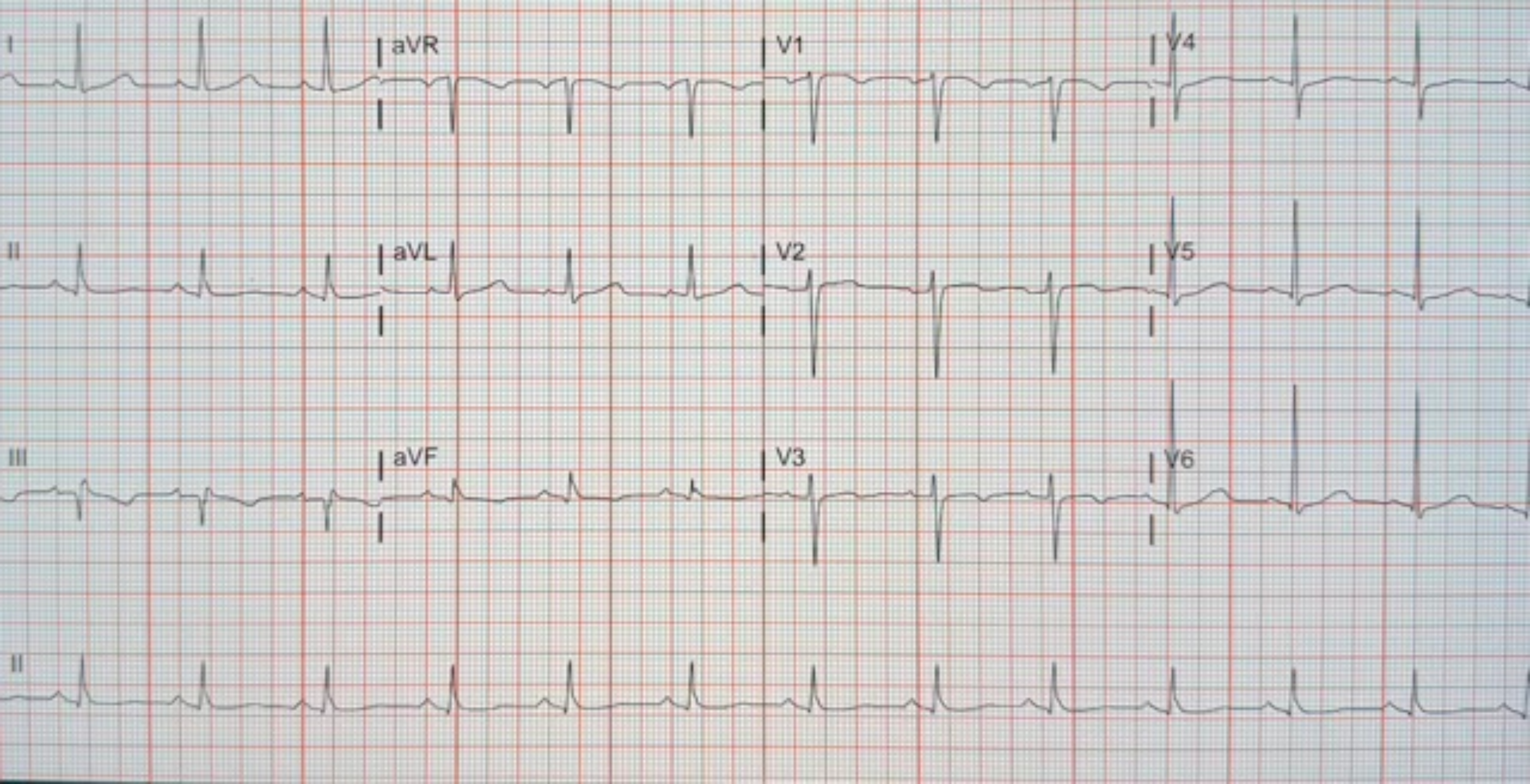
I
aVR
V1
V4
II
aVL
V2
V5
III
aVF
V3
V6
II

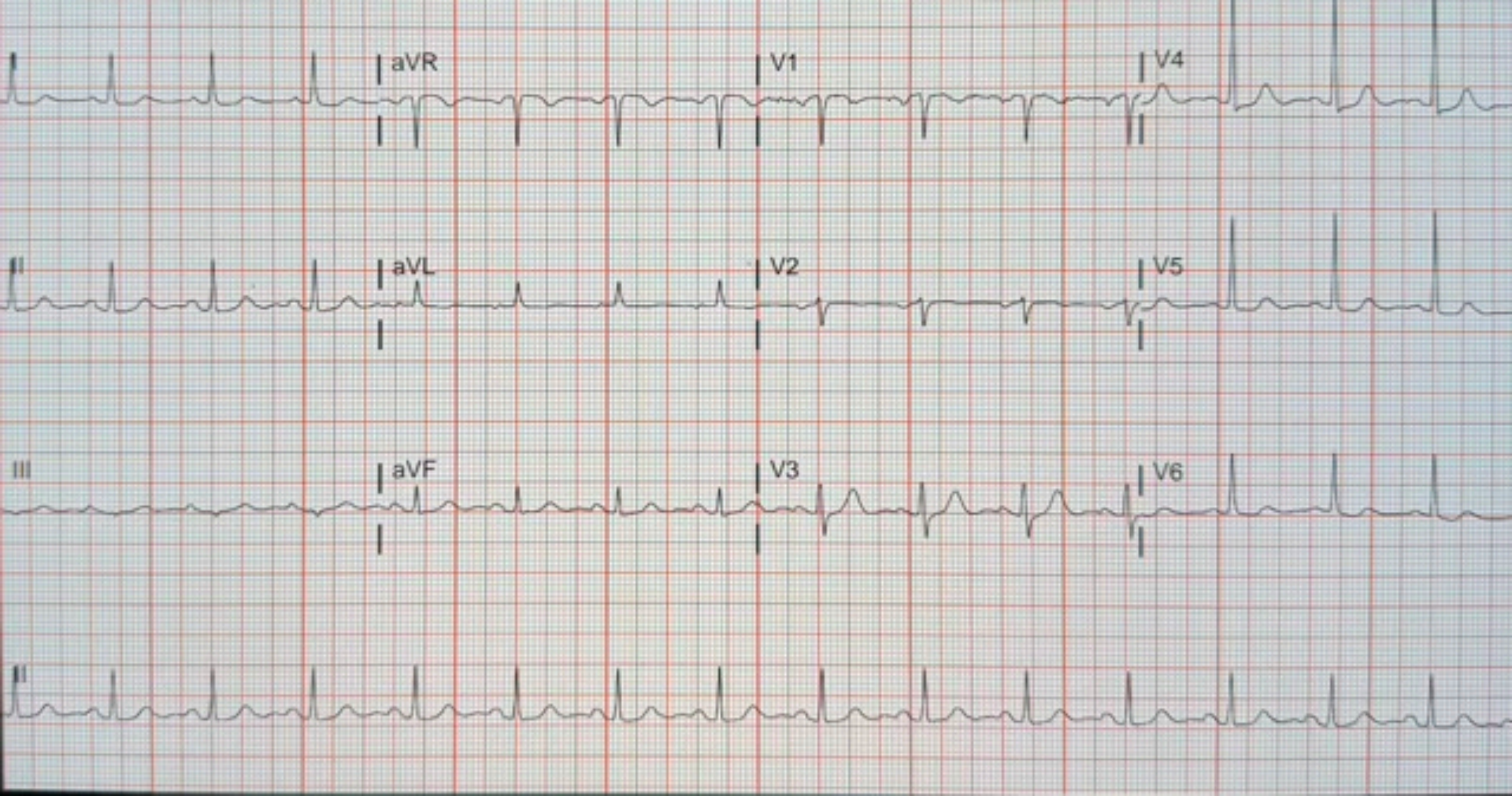
I
aVR
V1
V4
II
aVL
V2
V5
III
aVF
V3
V6
II

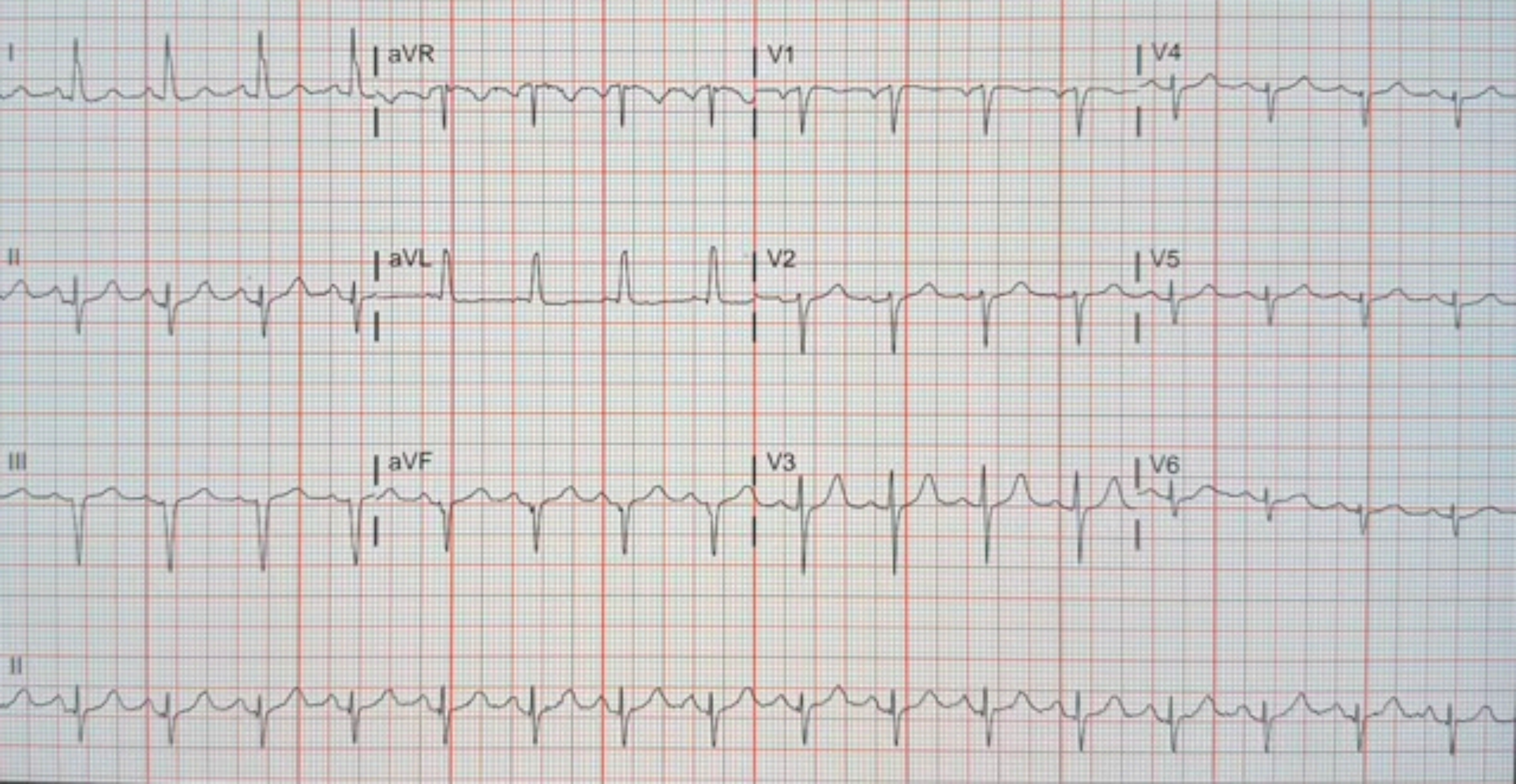
I
aVR
V1
V4
II
aVL
V2
V5
III
aVF
V3
V6
II

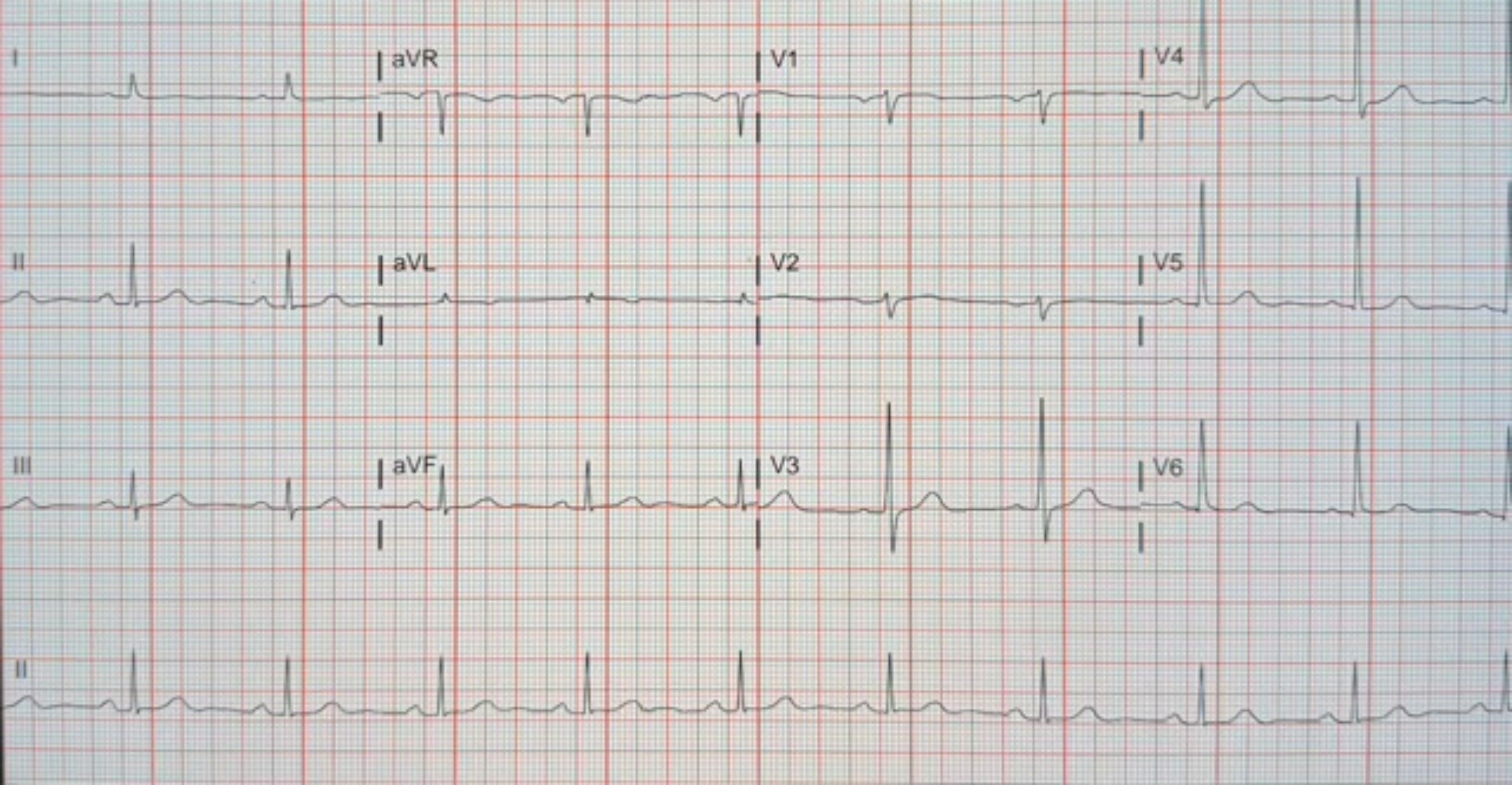
I
aVR
V1
V4
II
aVL
V2
V5
III
aVF
V3
V6
II

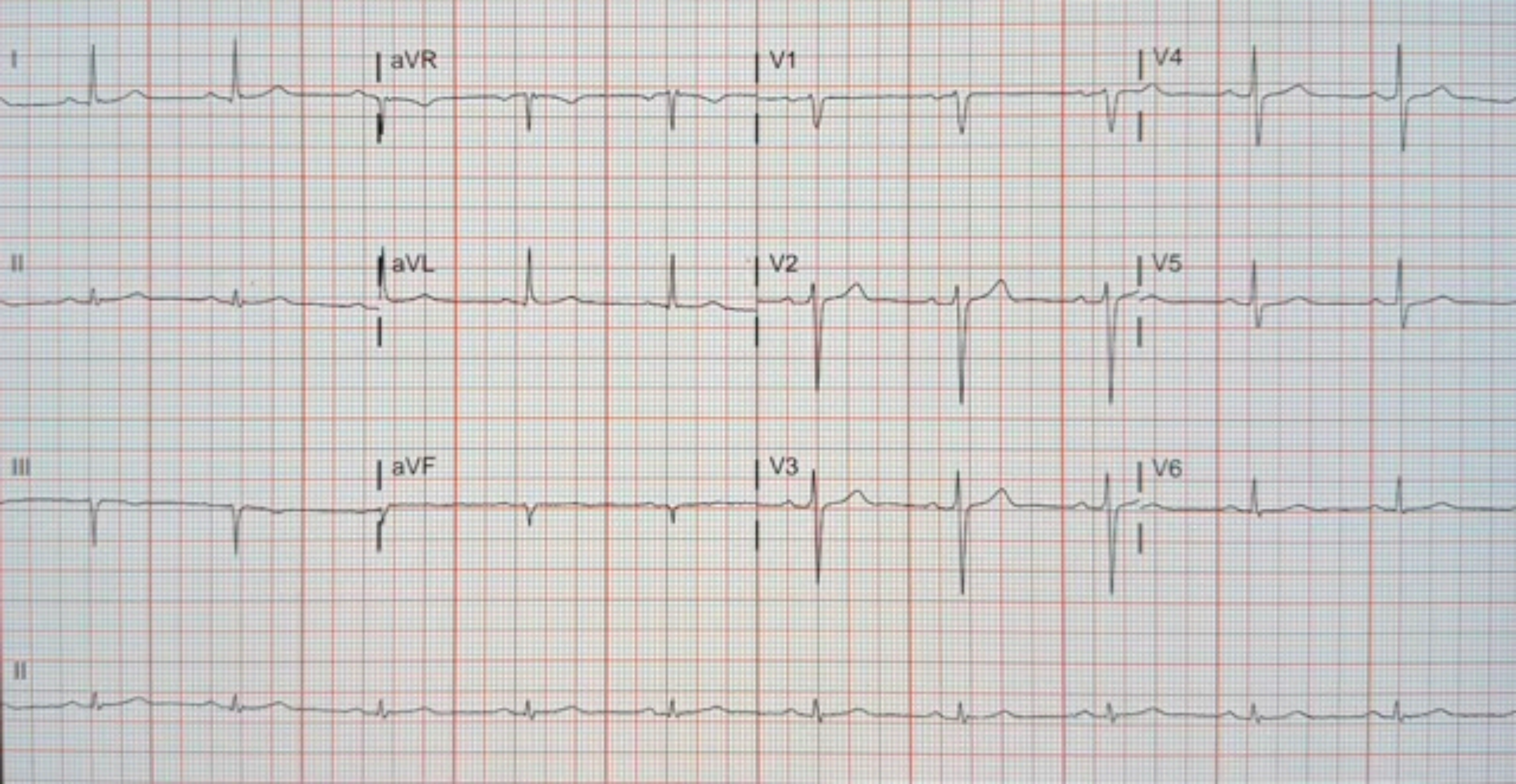
I
aVR
V1
V4
II
aVL
V2
V5
III
aVF
V3
V6
II

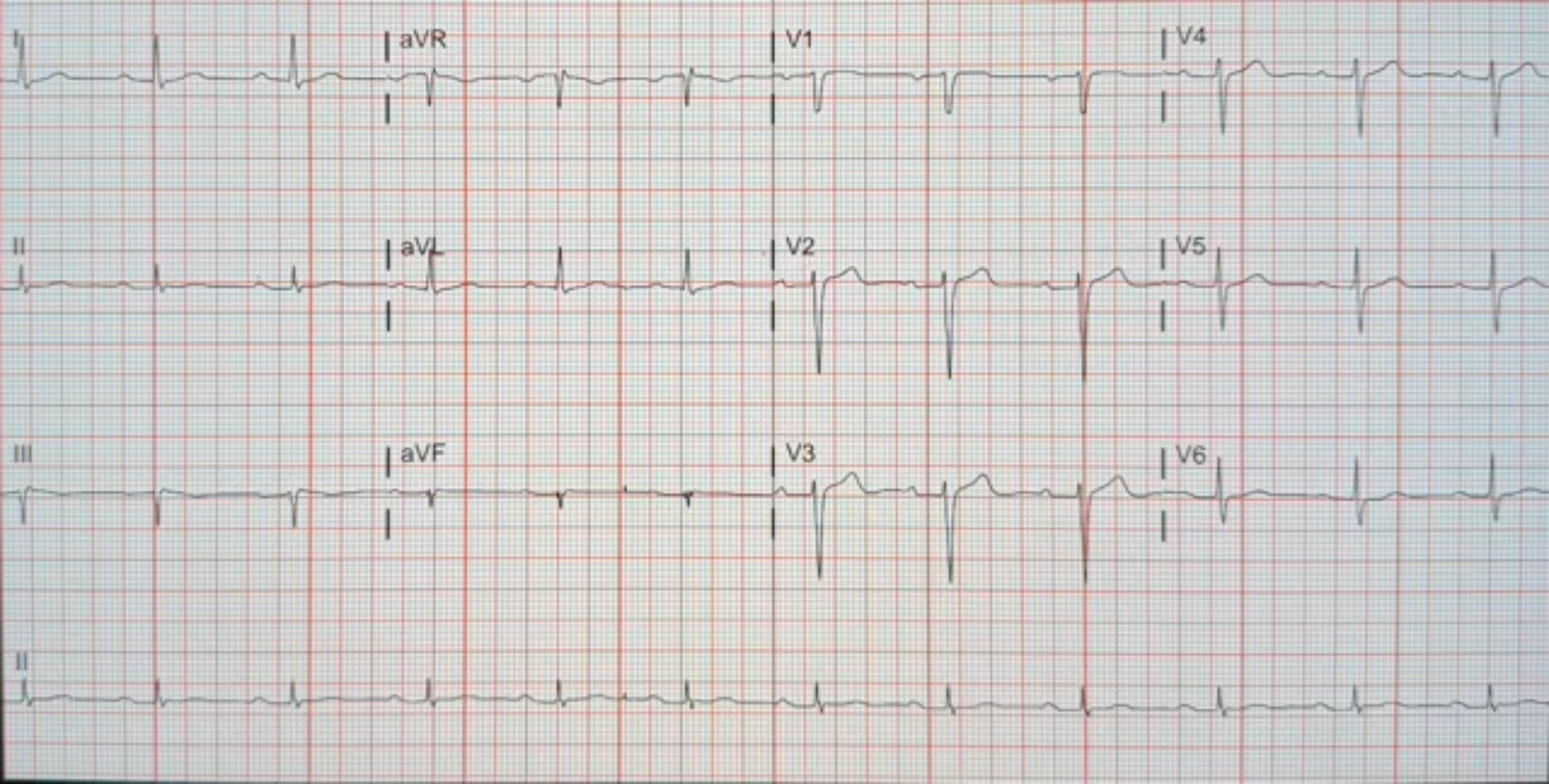
I
aVR
V1
V4
II
aVL
V2
V5
III
aVF
V3
V6
II

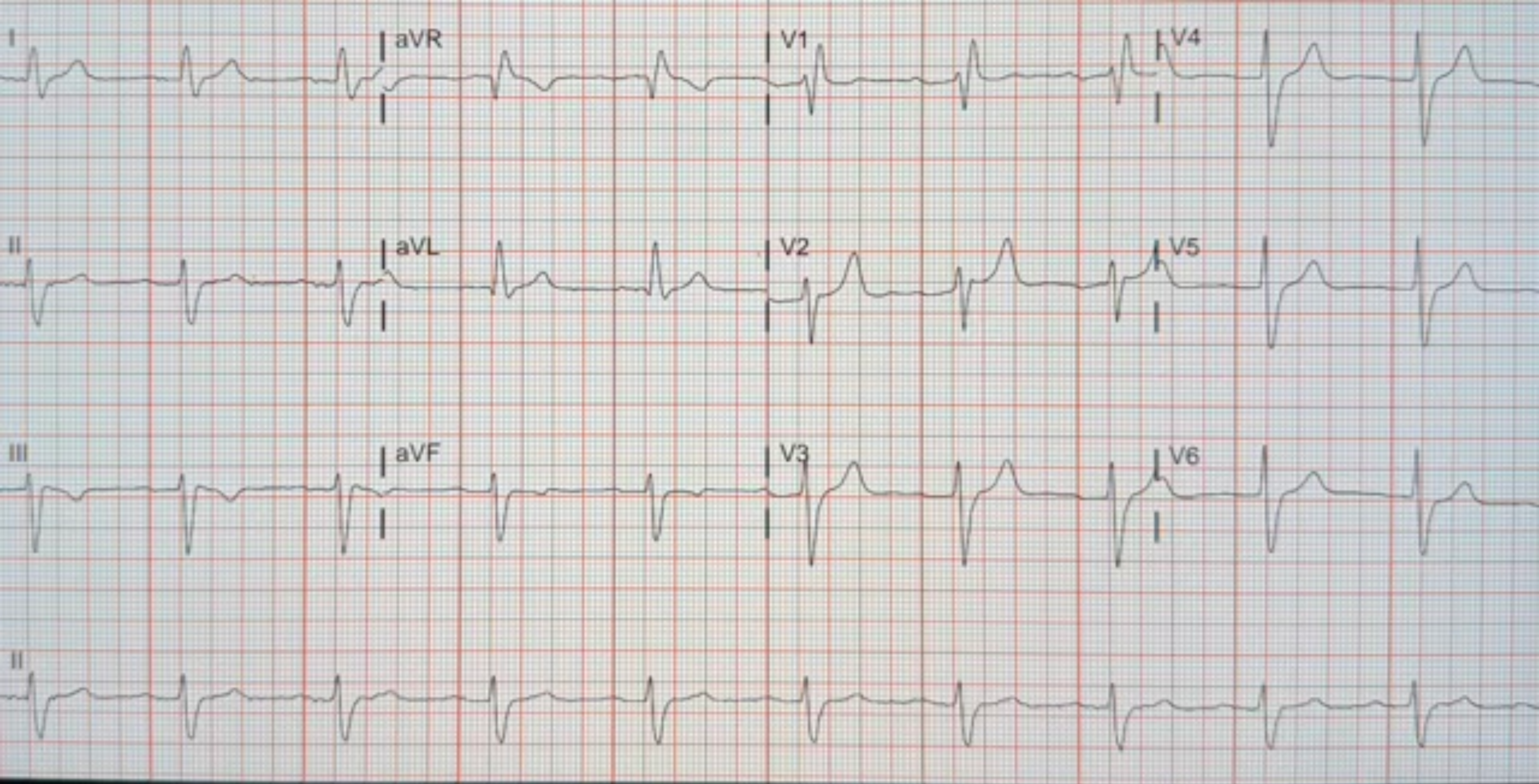
I
aVR
V1
V4
II
aVL
V2
V5
III
aVF
V3
V6
II

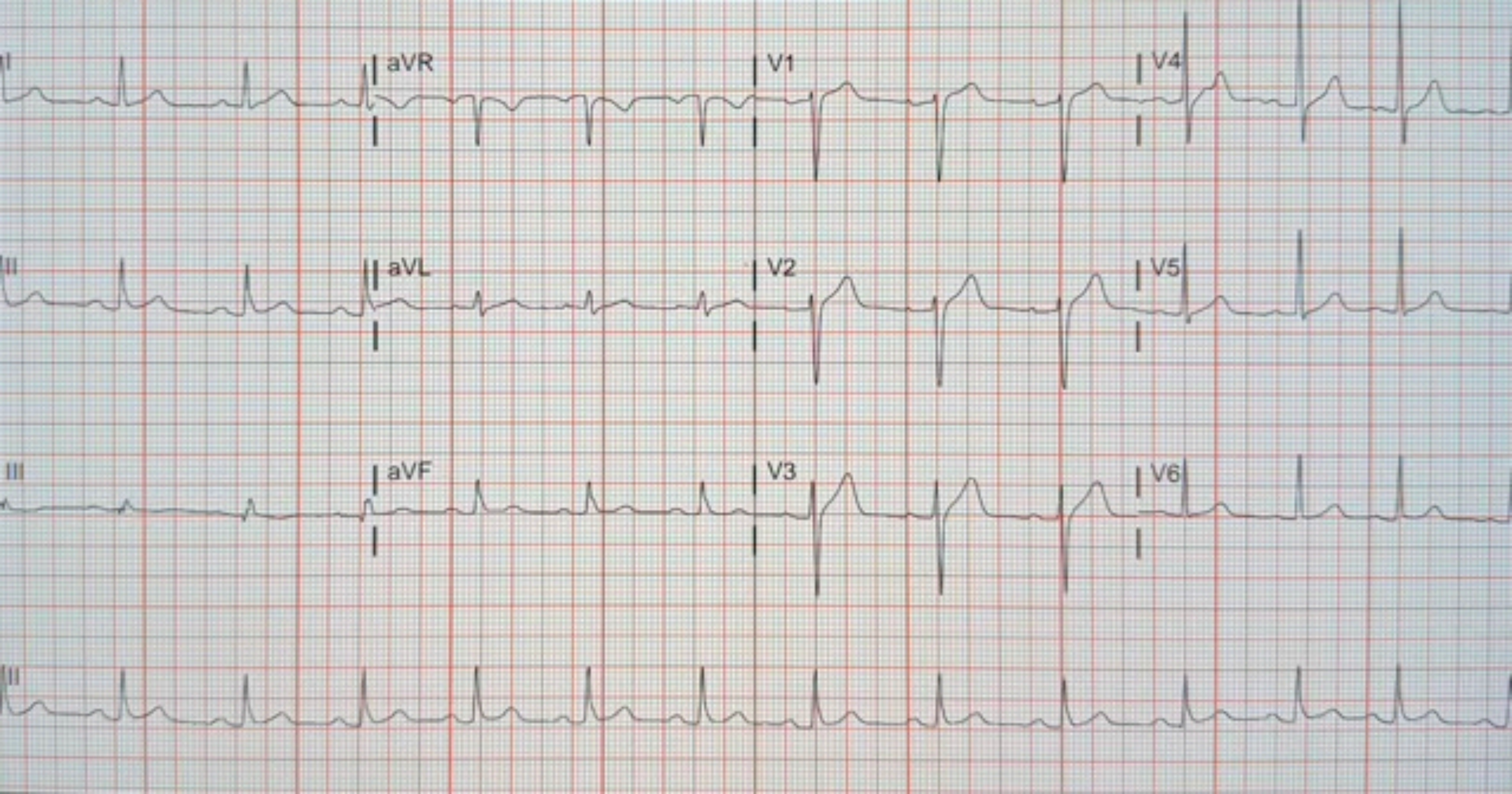

I
aVR
V1
V4
II
aVL
V2
V5
III
aVF
V3
V6
II

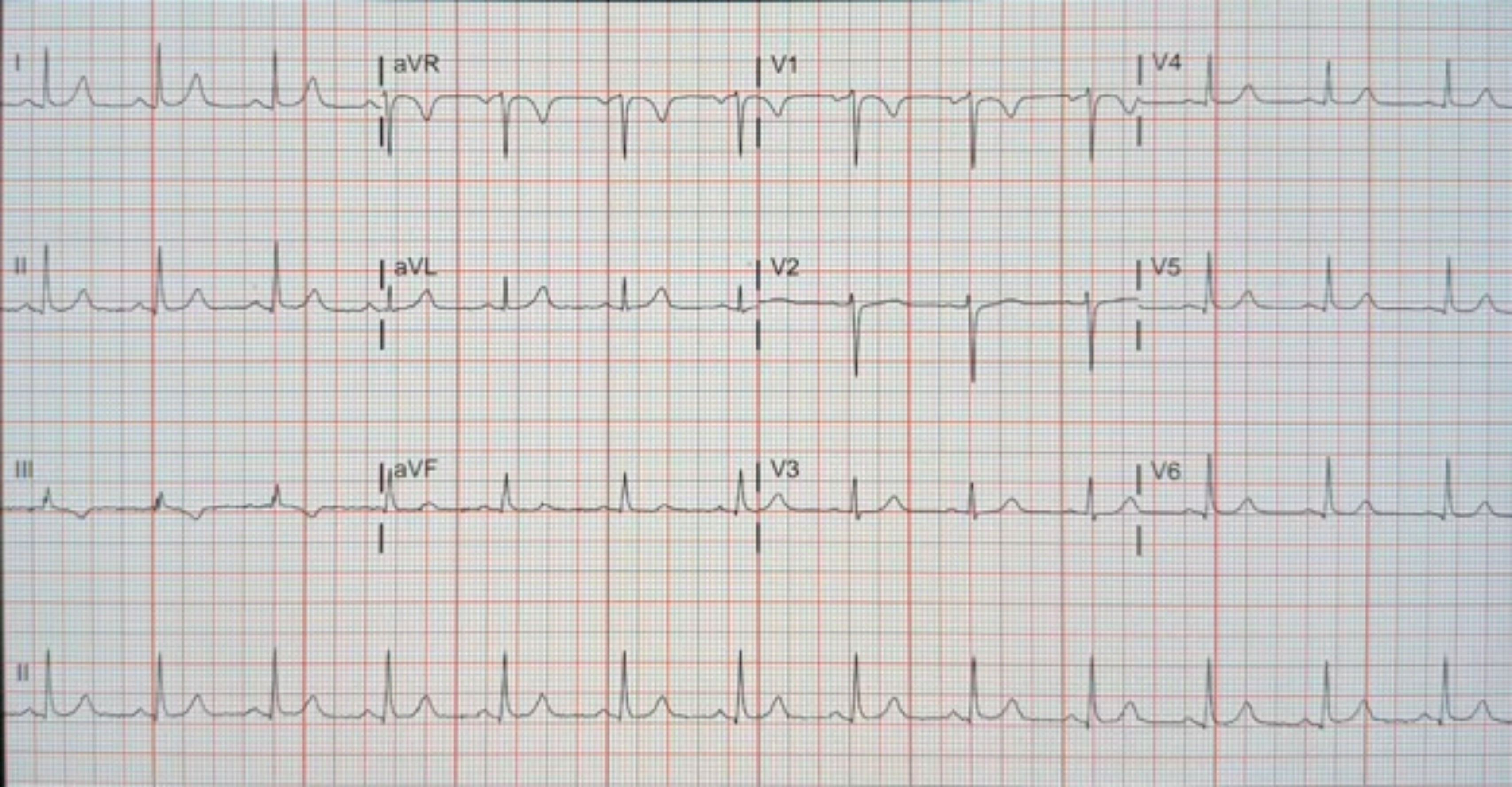
I
aVR
V1
V4
II
aVL
V2
V5
III
aVF
V3
V6
II

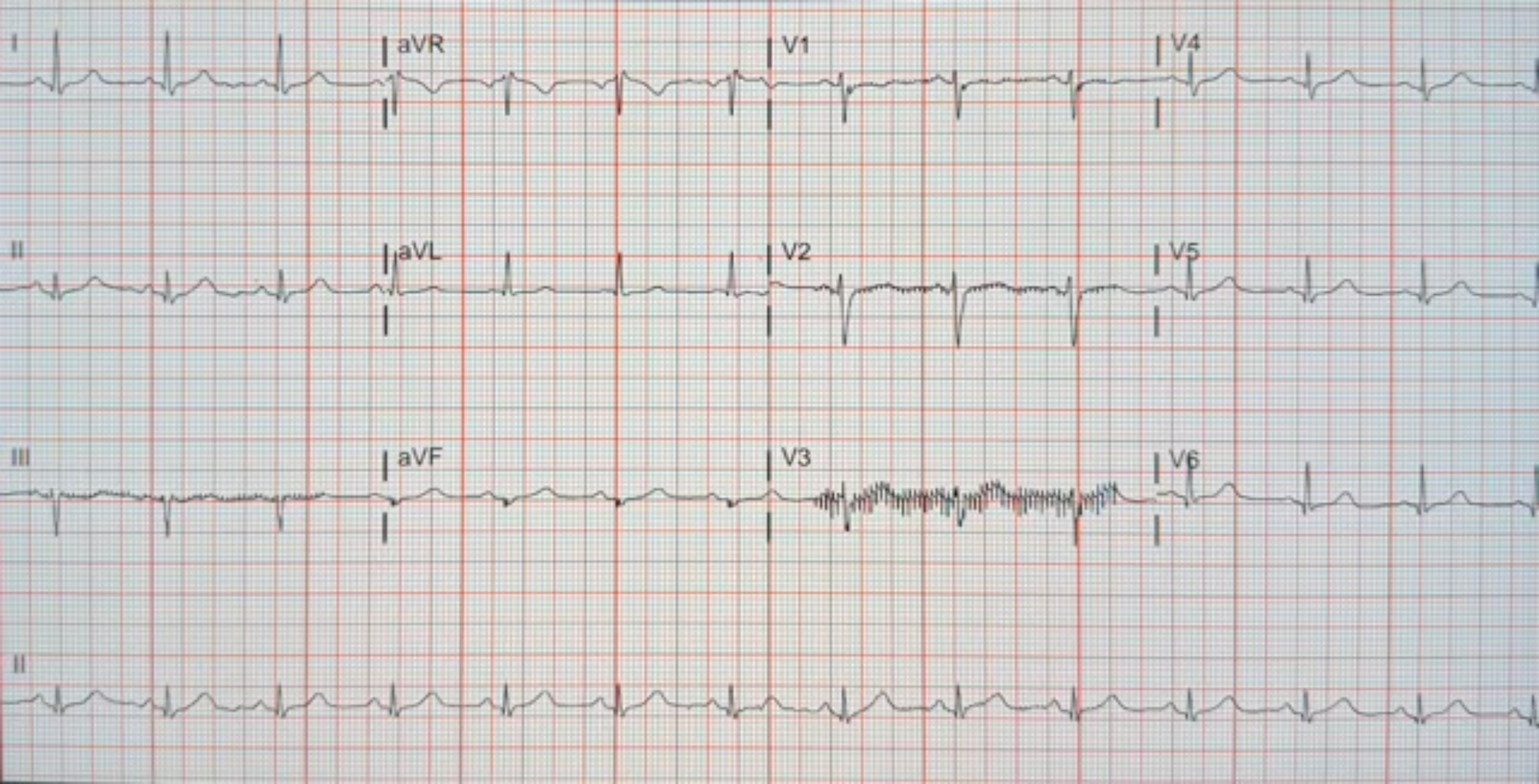
I
aVR
V1
V4
II
aVL
V2
V5
III
aVF
V3
V6
II

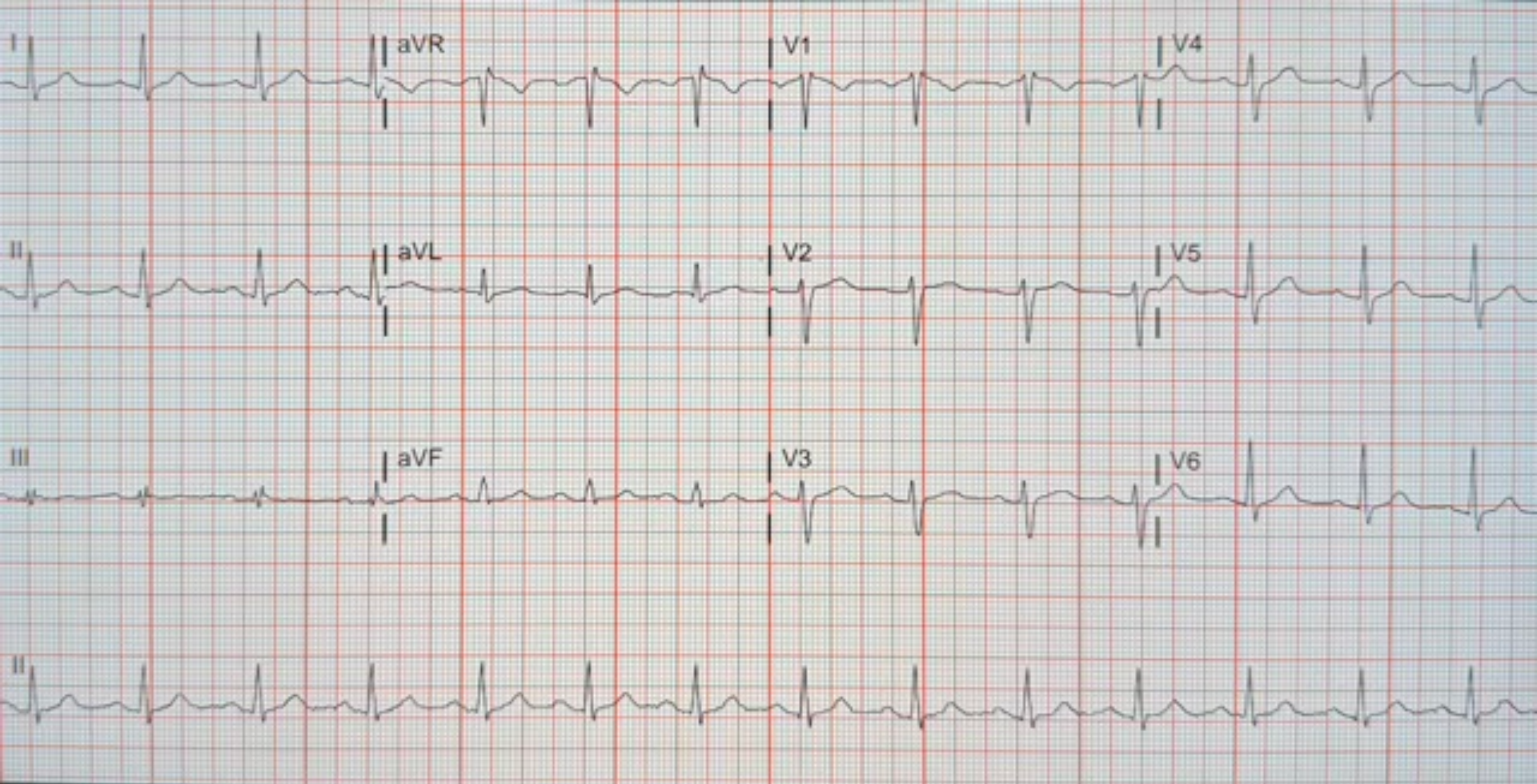

I
aVR
V1
V4
II
aVL
V2
V5
III
aVF
V3
V6
II

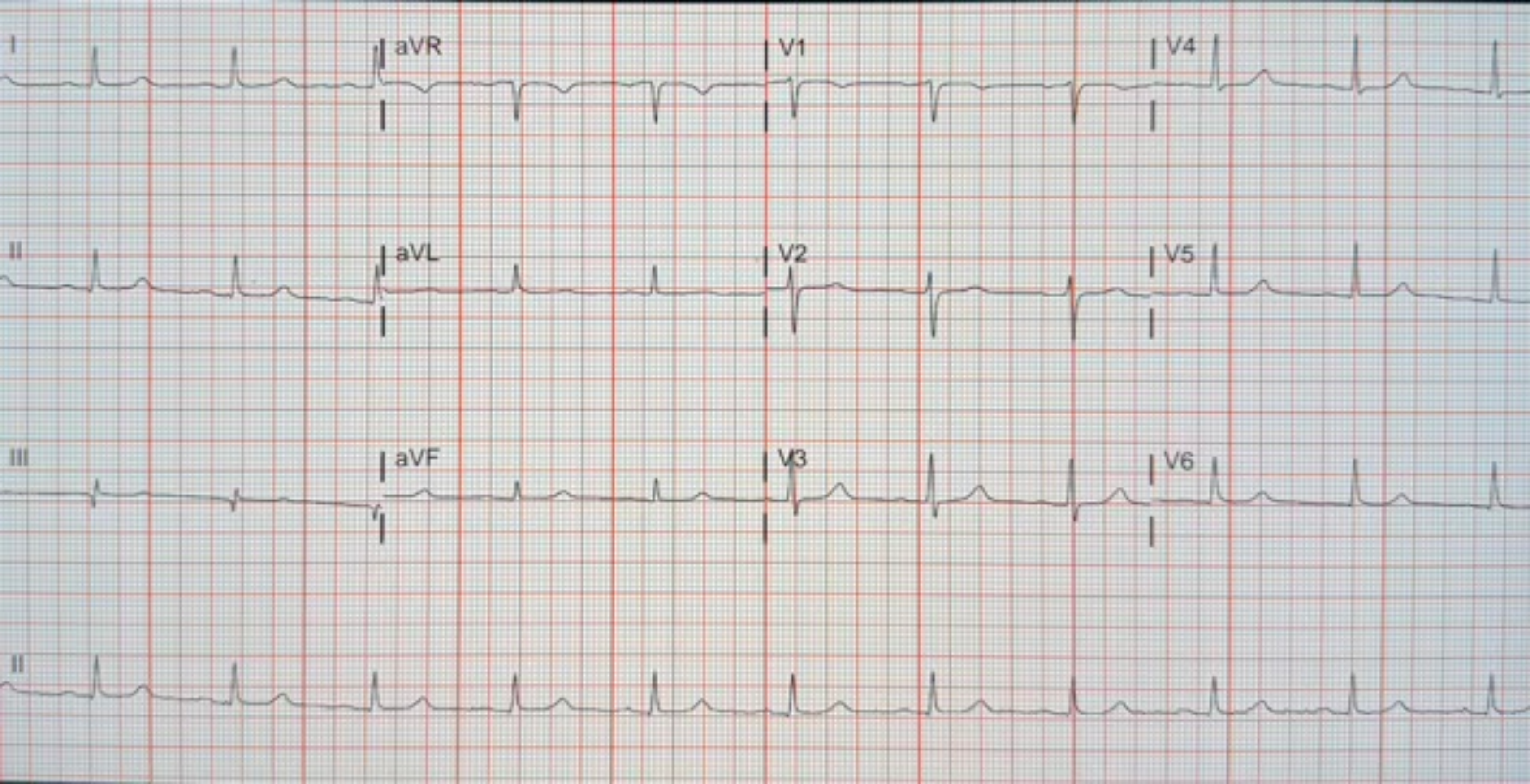
I
aVR
V1
V4
II
aVL
V2
V5
III
aVF
V3
V6
II

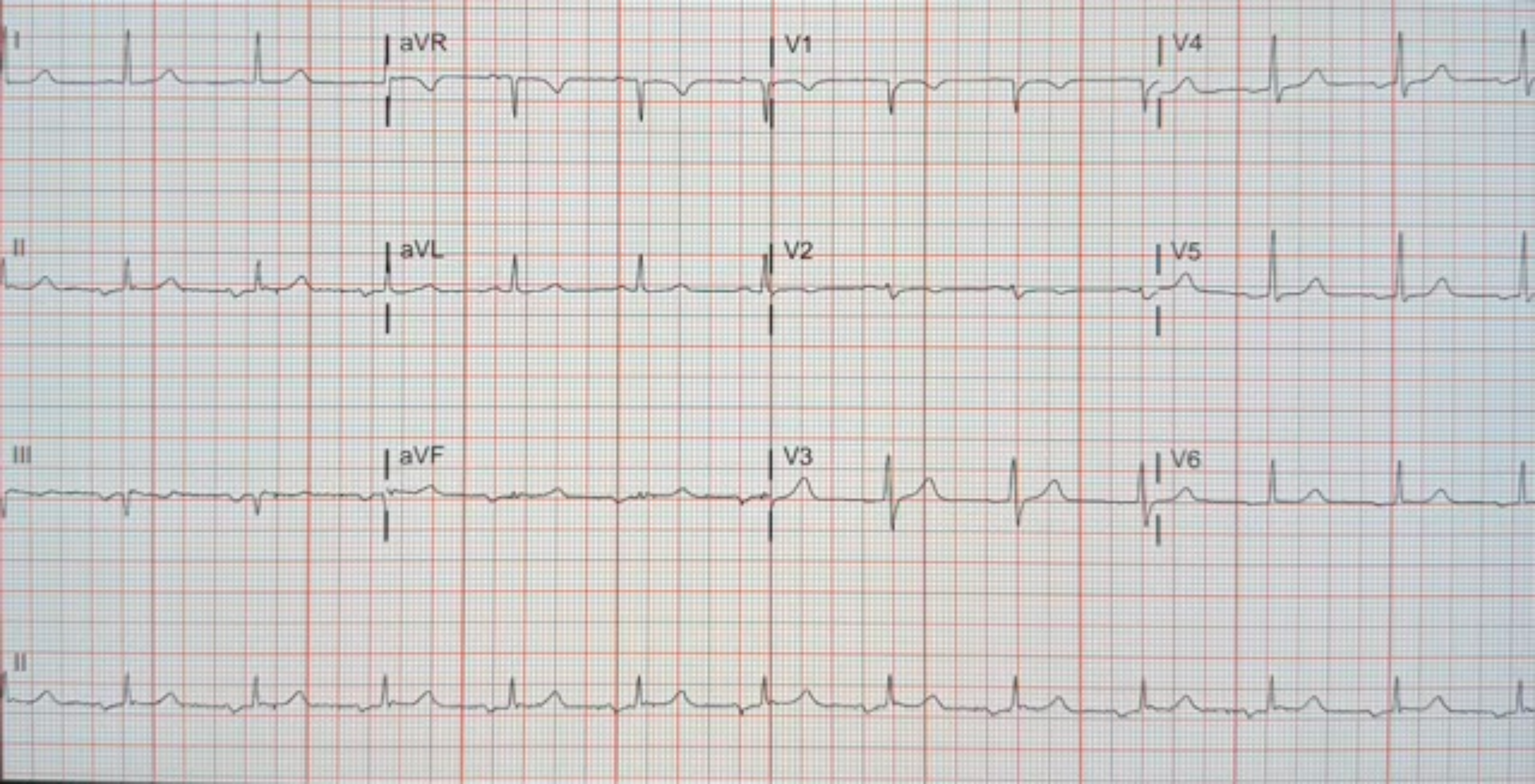
I
aVR
V1
V4
II
aVL
V2
V5
III
aVF
V3
V6
II

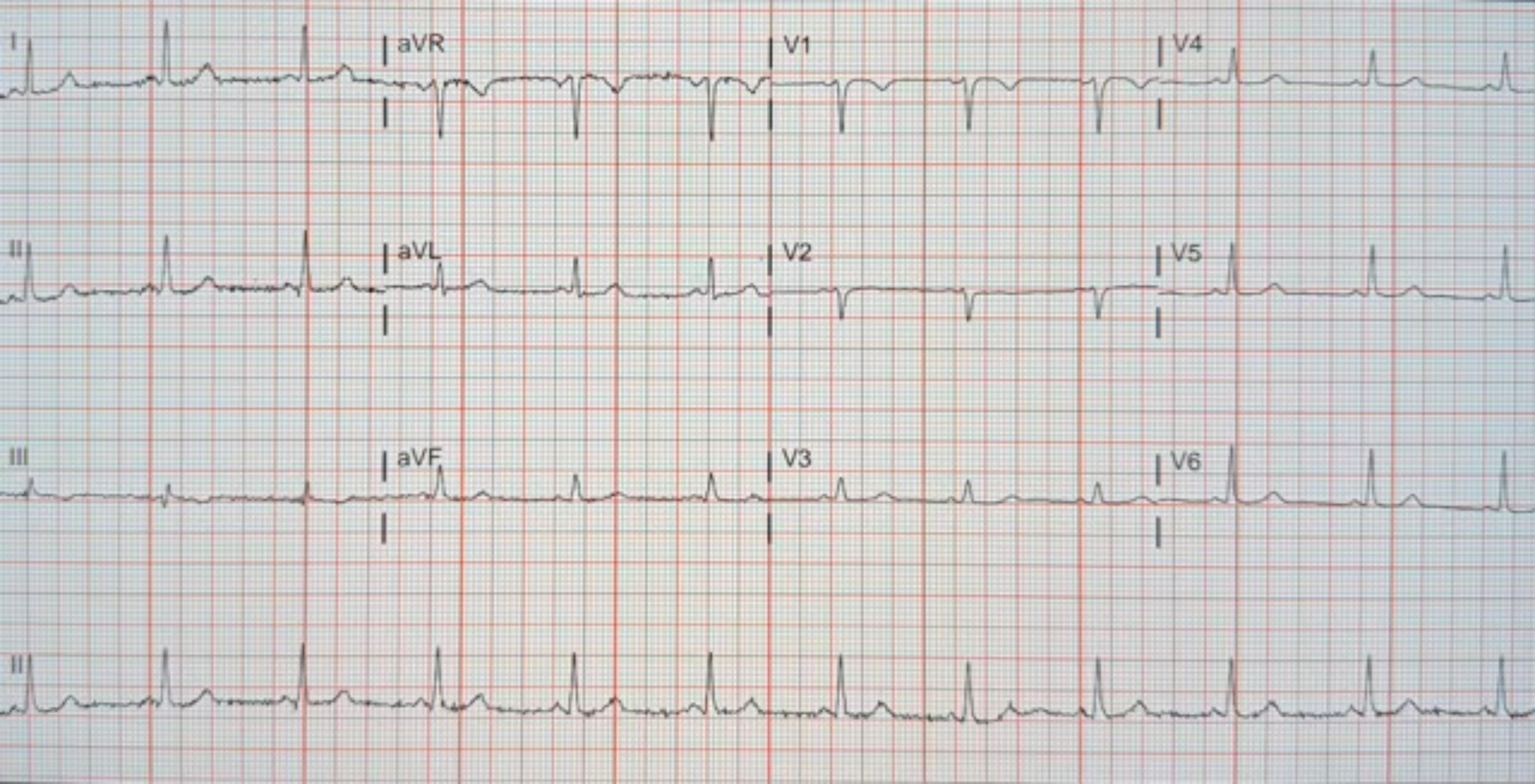
I
aVR
V1
V4
II
aVL
V2
V5
III
aVF
V3
V6
II

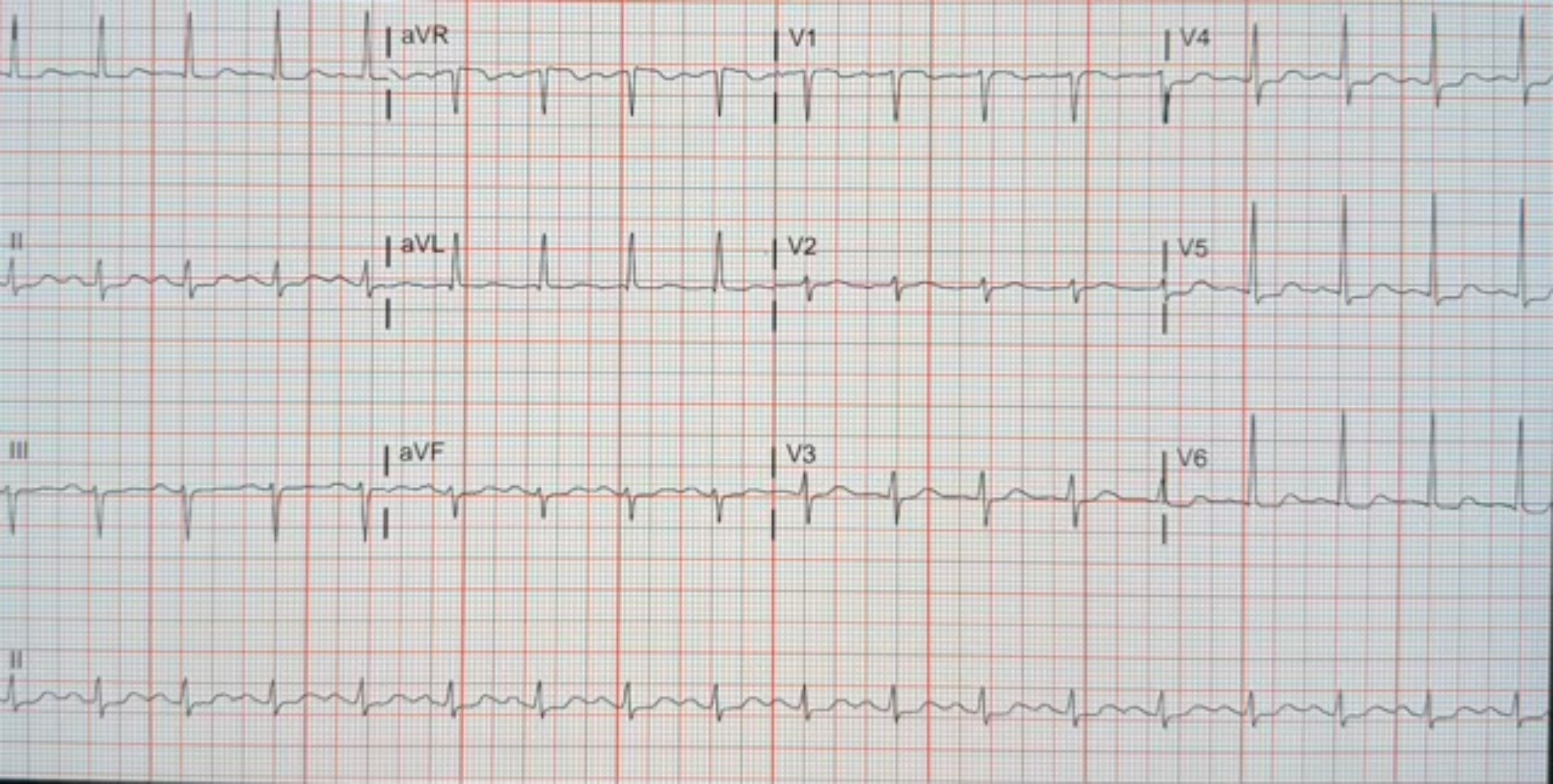
aVR
V1
V4
II
aVL
V2
V5
III
aVF
V3
V6
II

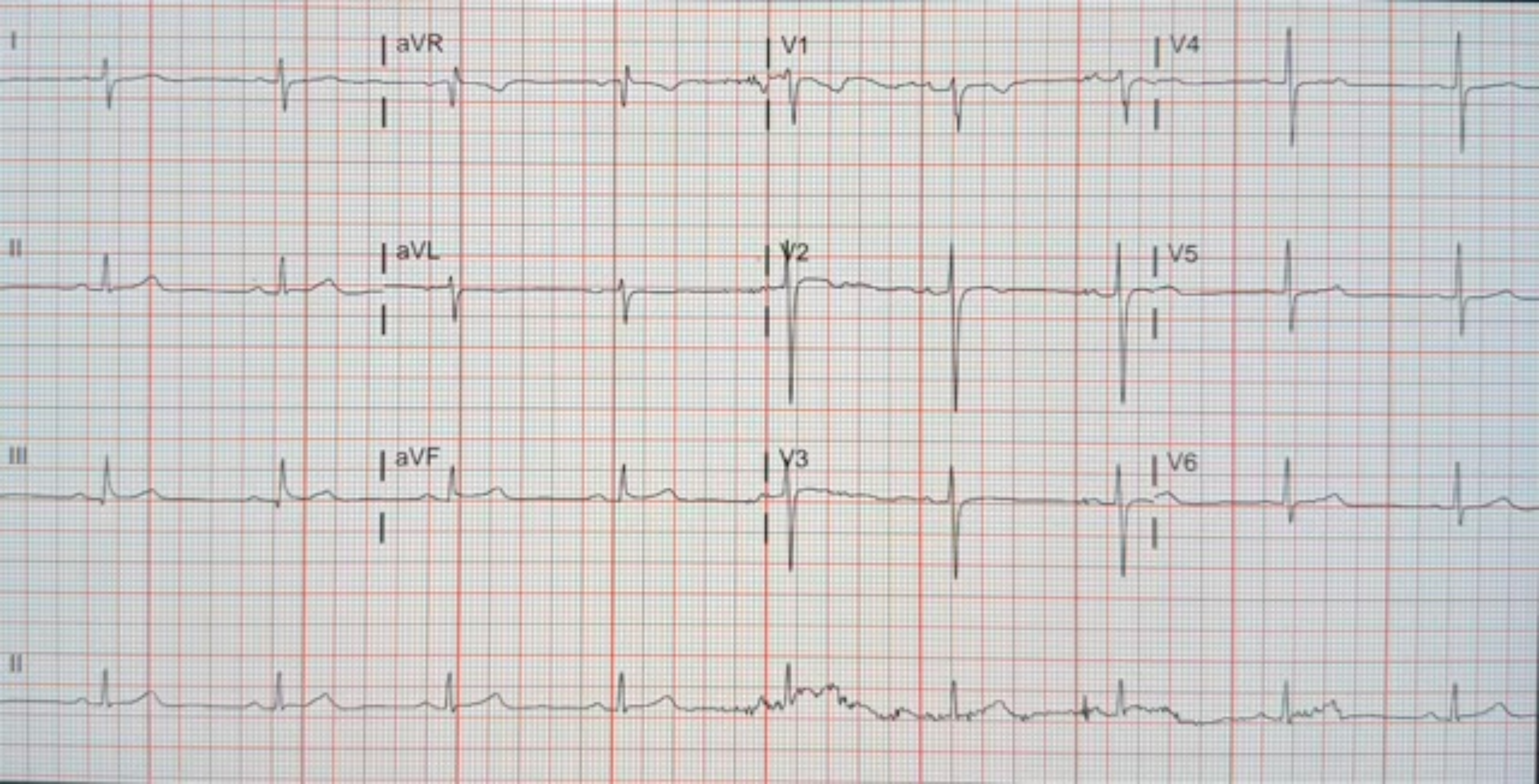
I
aVR
V1
V4
II
aVL
V2
V5
III
aVF
V3
V6
II

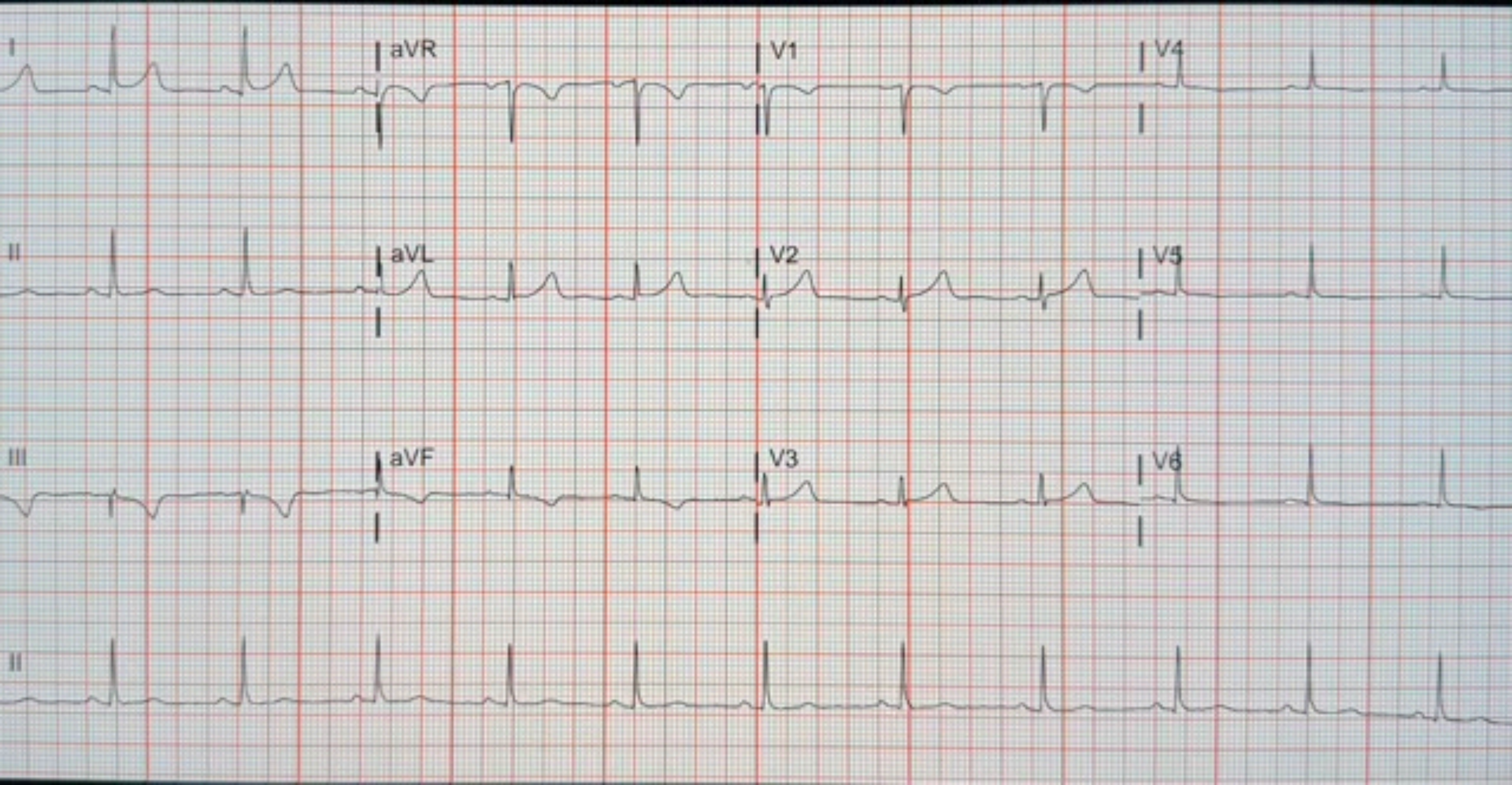
I
aVR
V1
V4
II
aVL
V2
V5
III
aVF
V3
V6
II

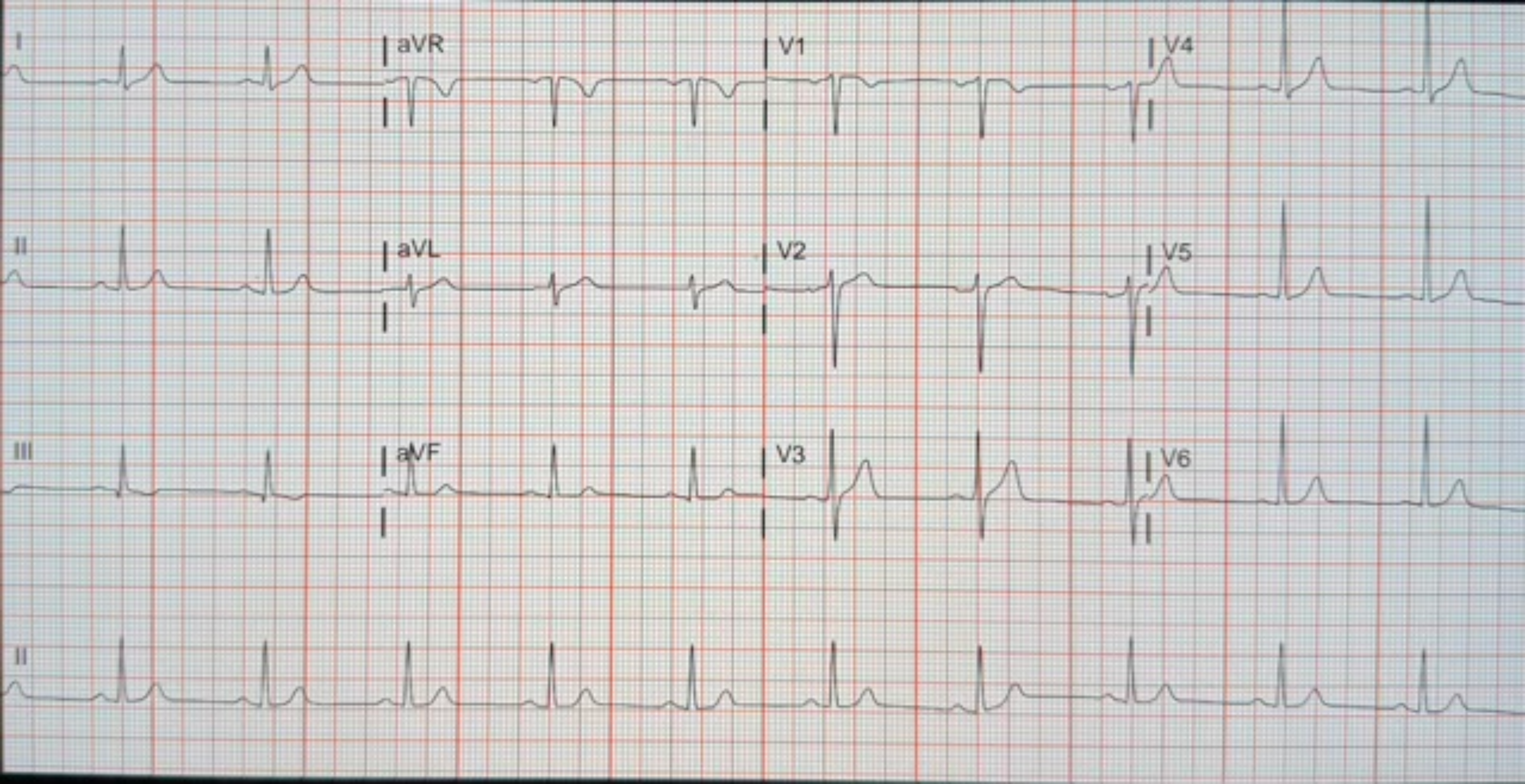
I
aVR
V1
V4
II
aVL
V2
V5
III
aVF
V3
V6
II

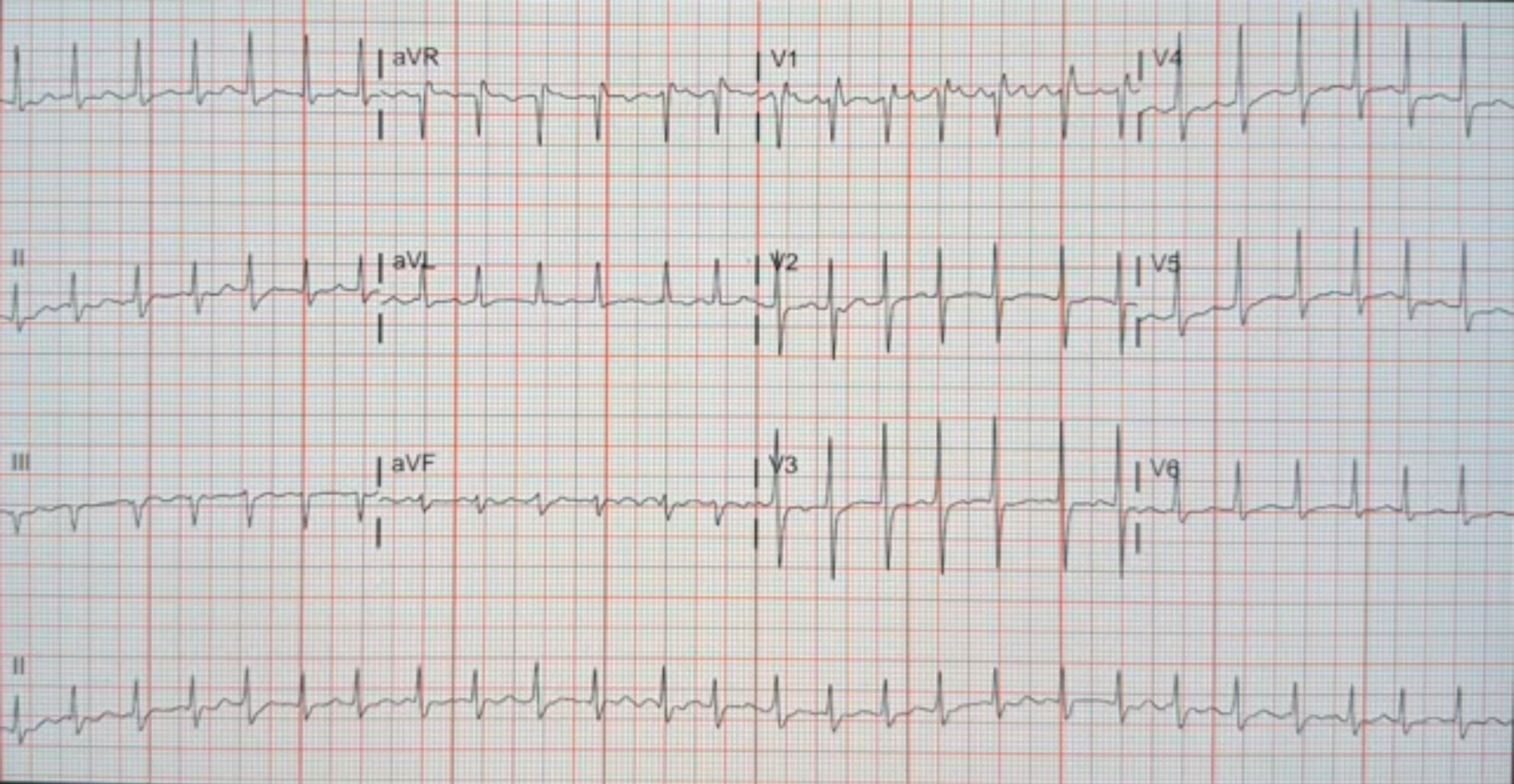
aVR
V1
V4
II
aVL
V2
V5
III
aVF
V3
V6
II

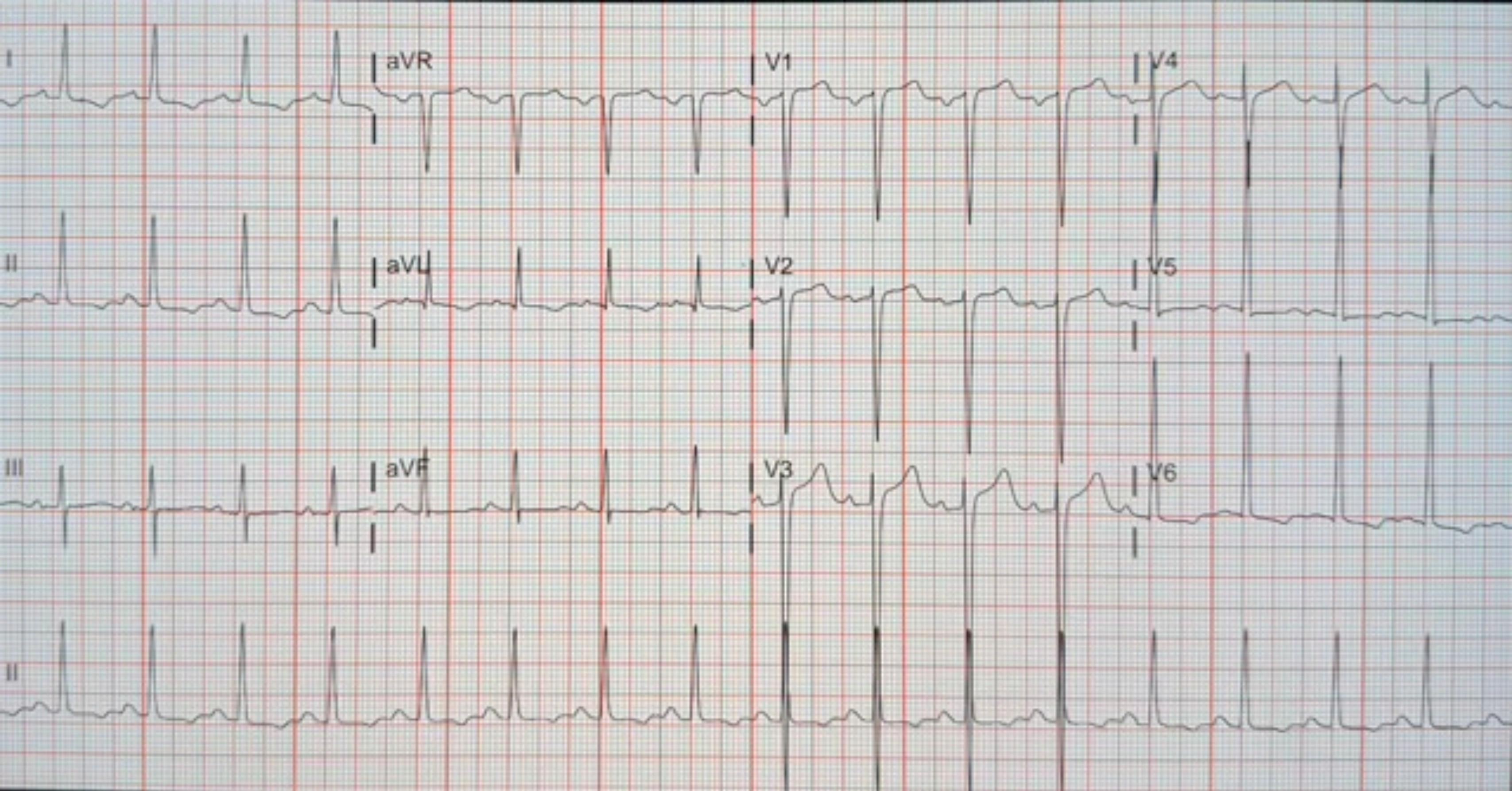
I
aVR
V1
V4
II
aVL
V2
V5
III
aVF
V3
V6
II

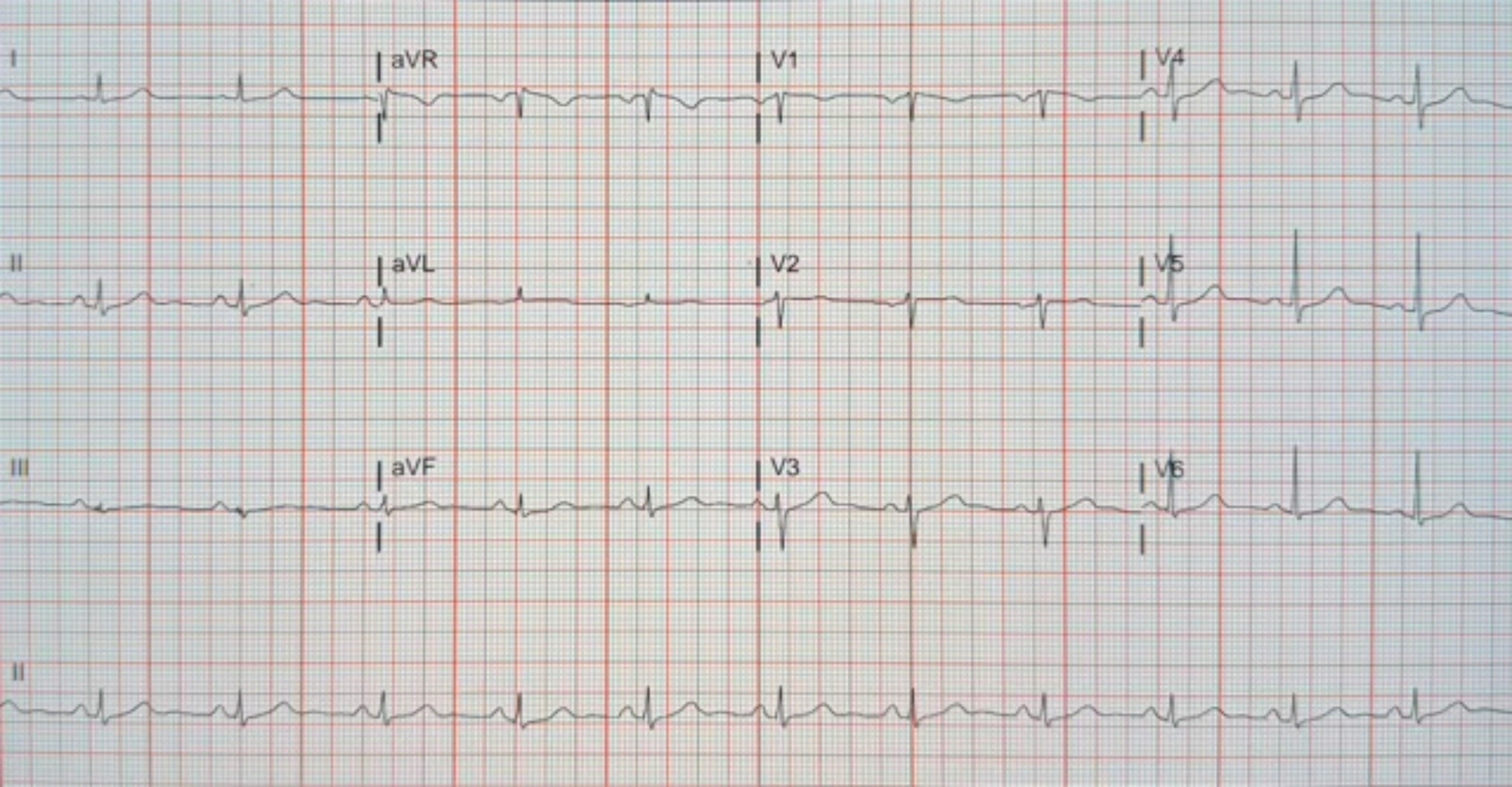
I
aVR
V1
V4
II
aVL
V2
V5
III
aVF
V3
V6
II

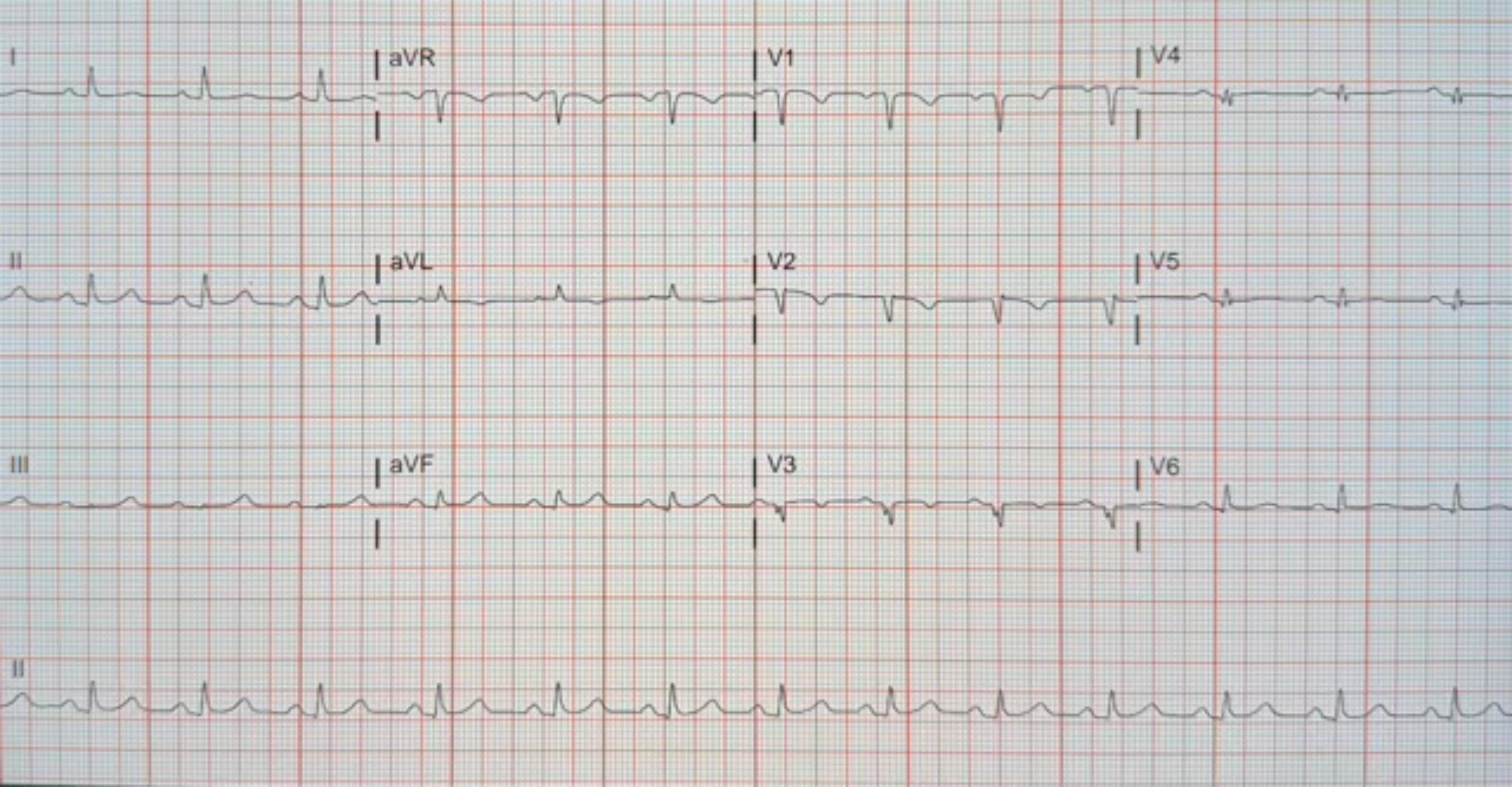
I
aVR
V1
V4
II
aVL
V2
V5
III
aVF
V3
V6
II

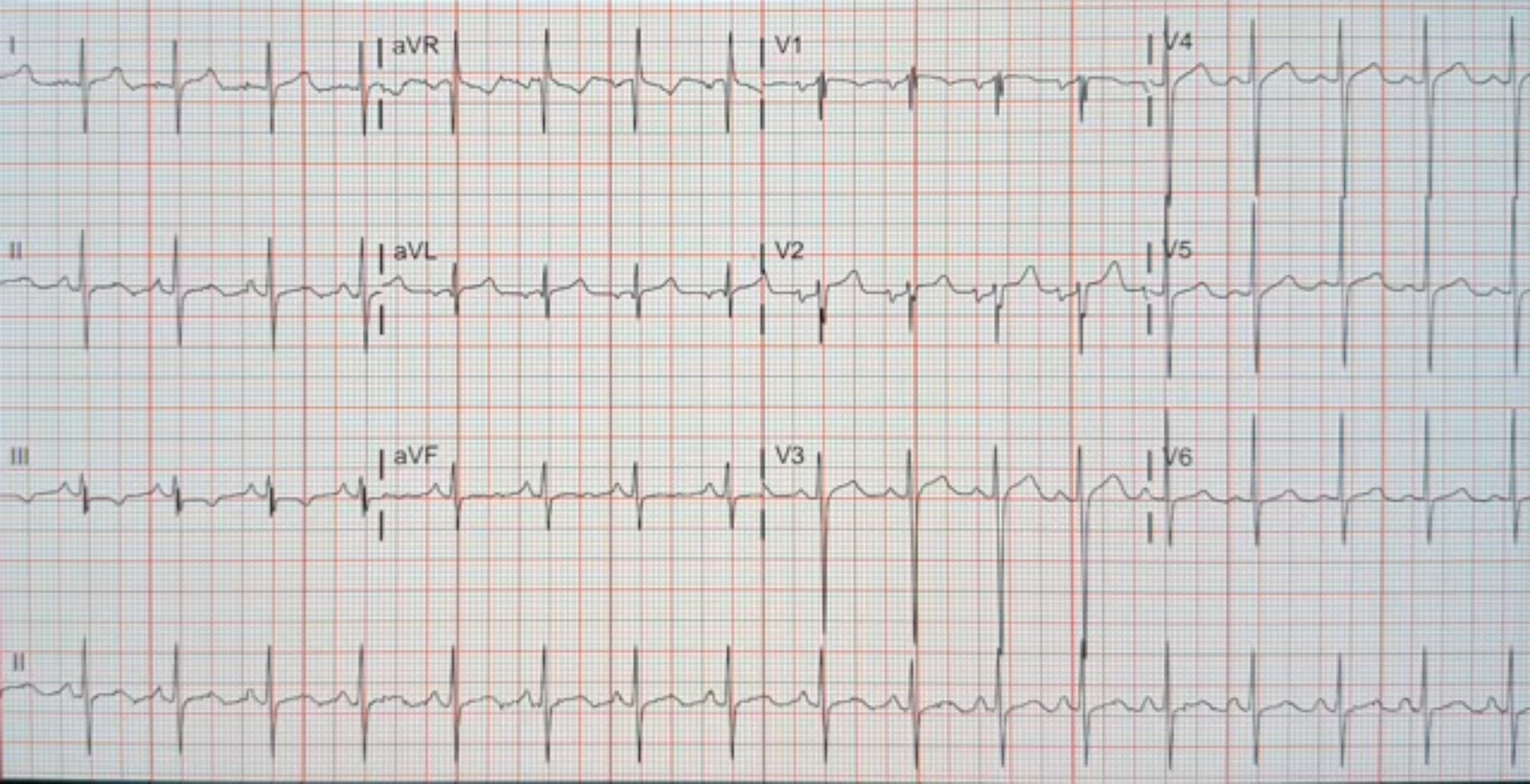
I
aVR
V1
V4
II
aVL
V2
V5
III
aVF
V3
V6
II

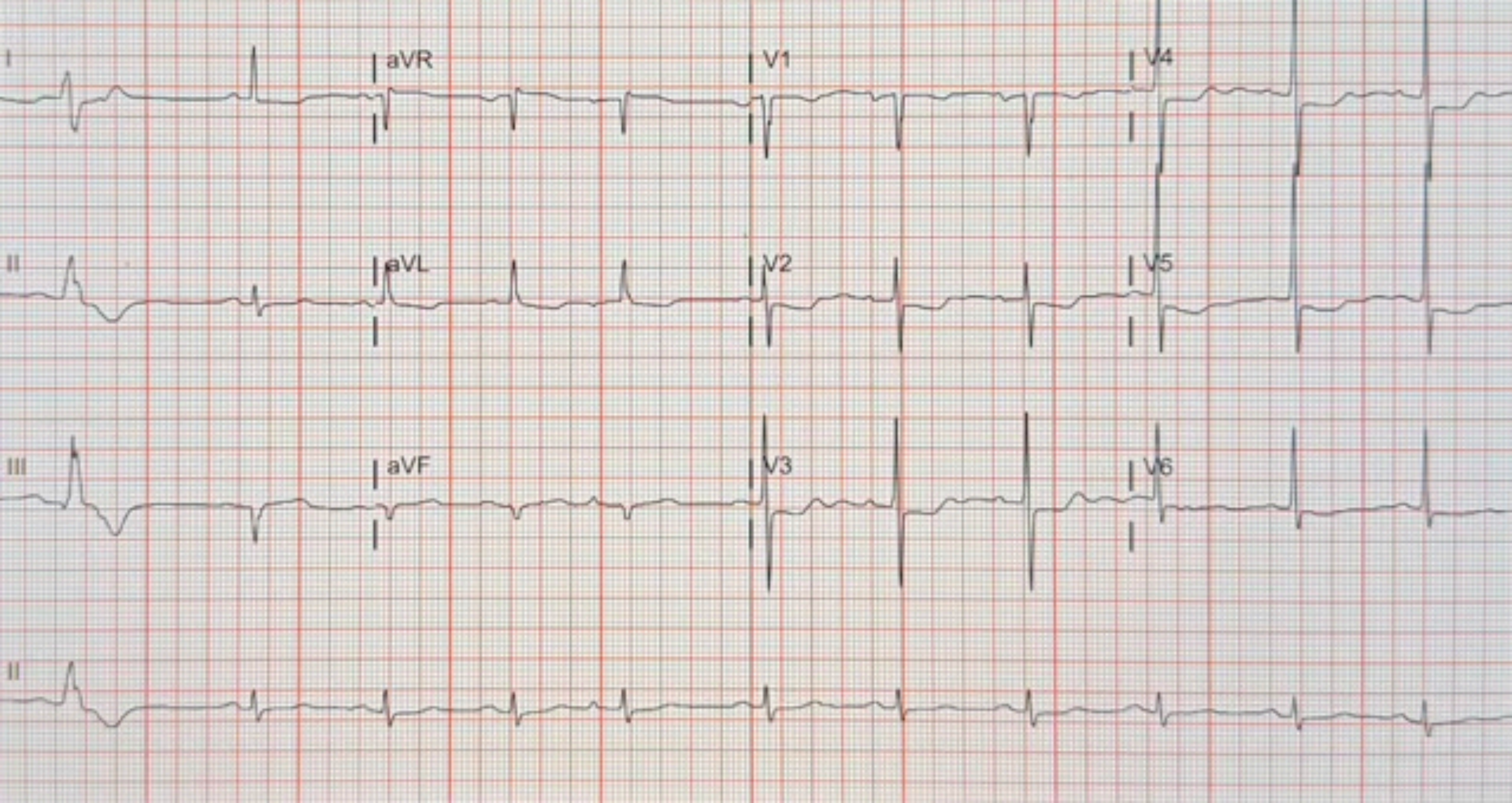
I
aVR
V1
V4
II
aVL
V2
V5
III
aVF
V3
V6
II

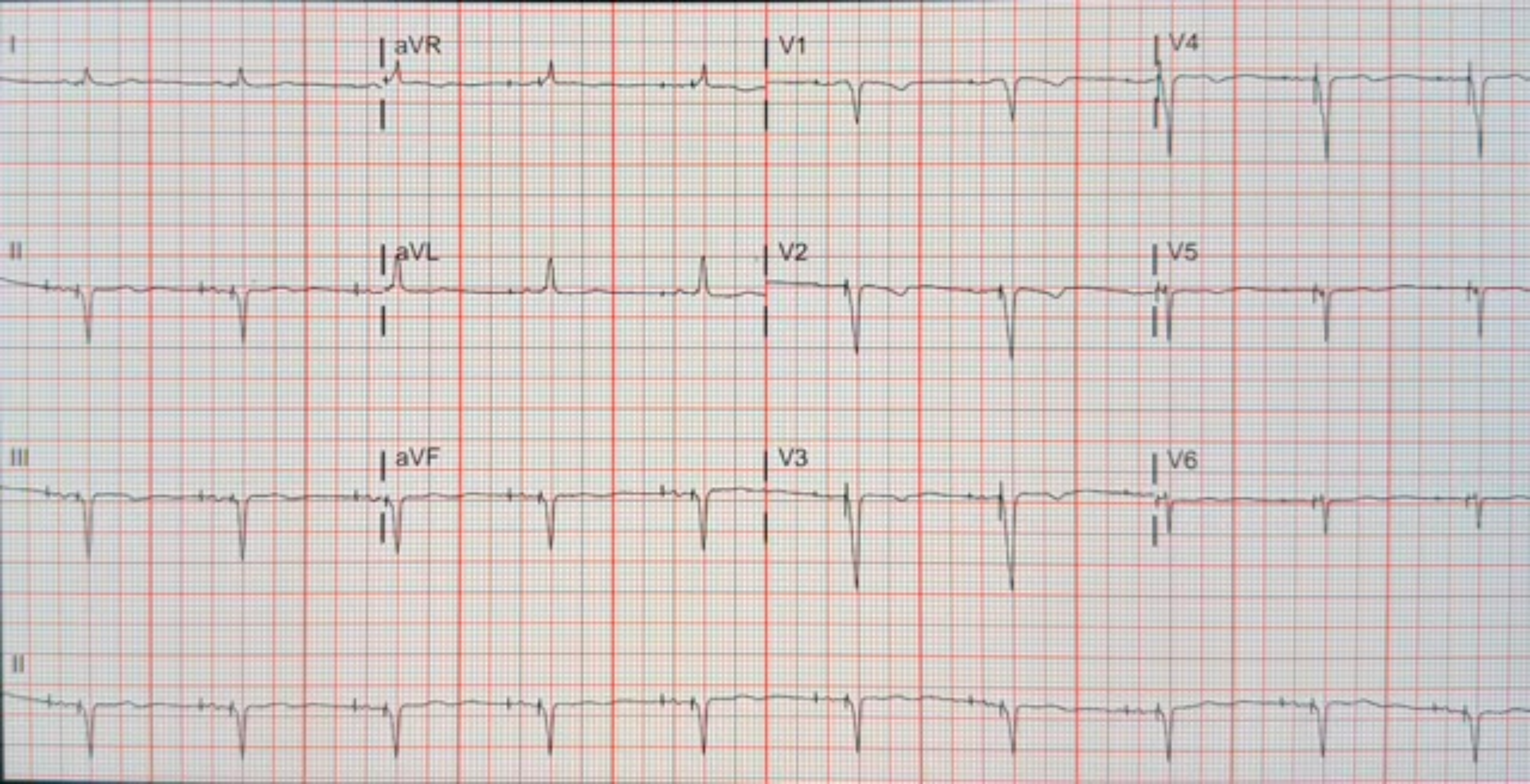
I
aVR
V1
V4
II
aVL
V2
V5
III
aVF
V3
V6
II

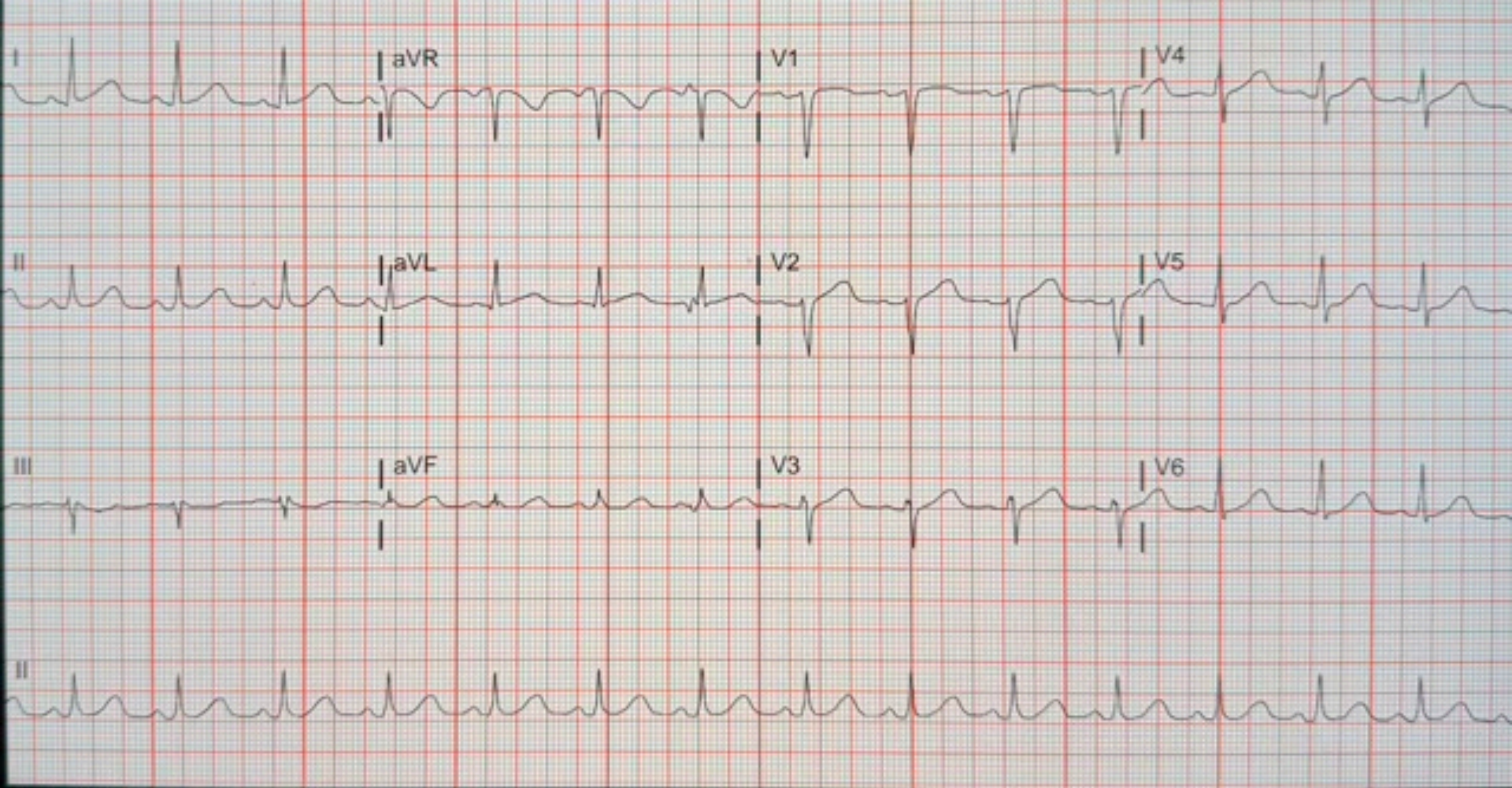
I
aVR
V1
V4
II
aVL
V2
V5
III
aVF
V3
V6
II

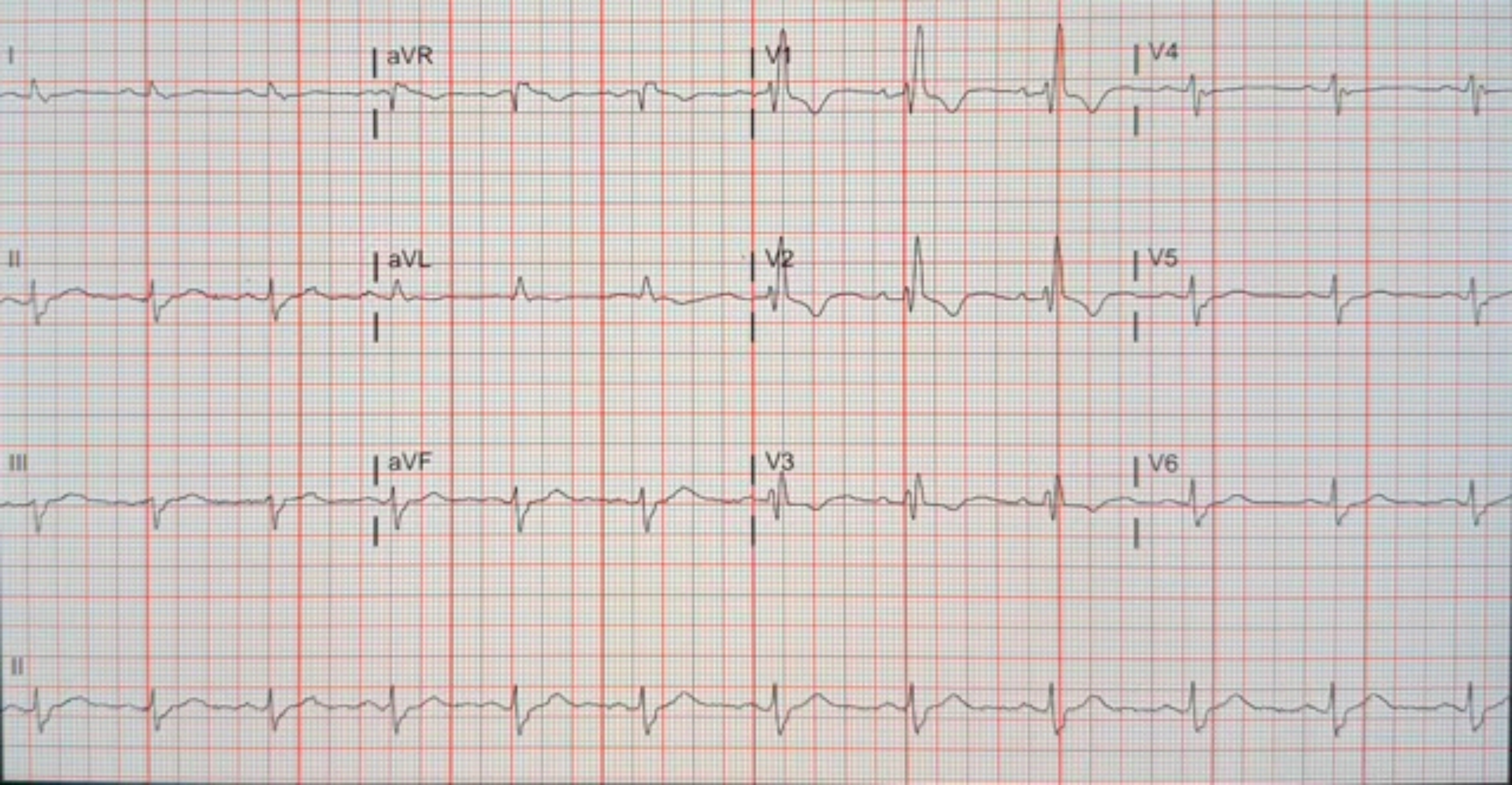
I
aVR
V1
V4
II
aVL
V2
V5
III
aVF
V3
V6
II

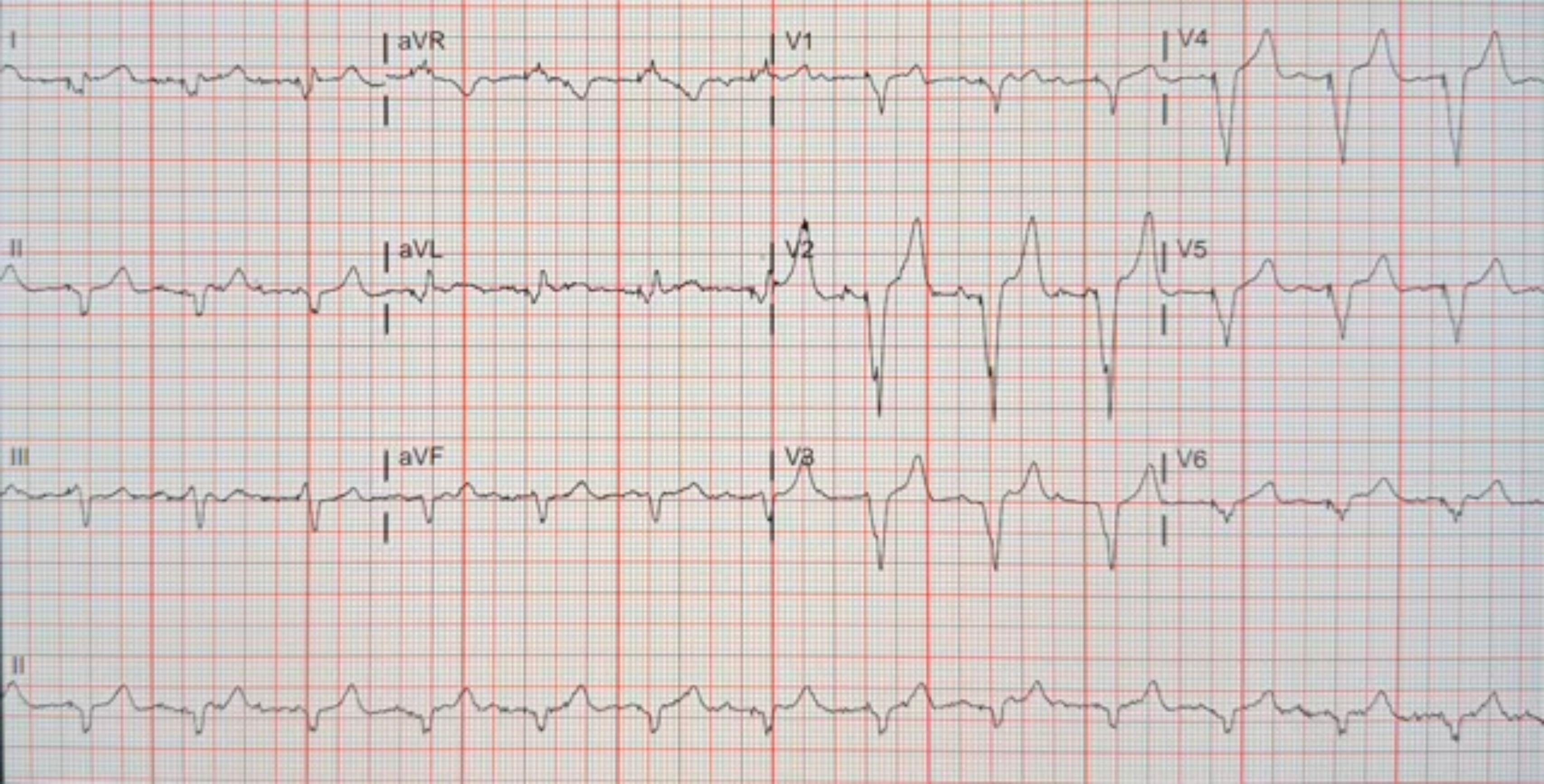
I
aVR
V1
V4
II
aVL
V2
V5
III
aVF
V3
V6
II

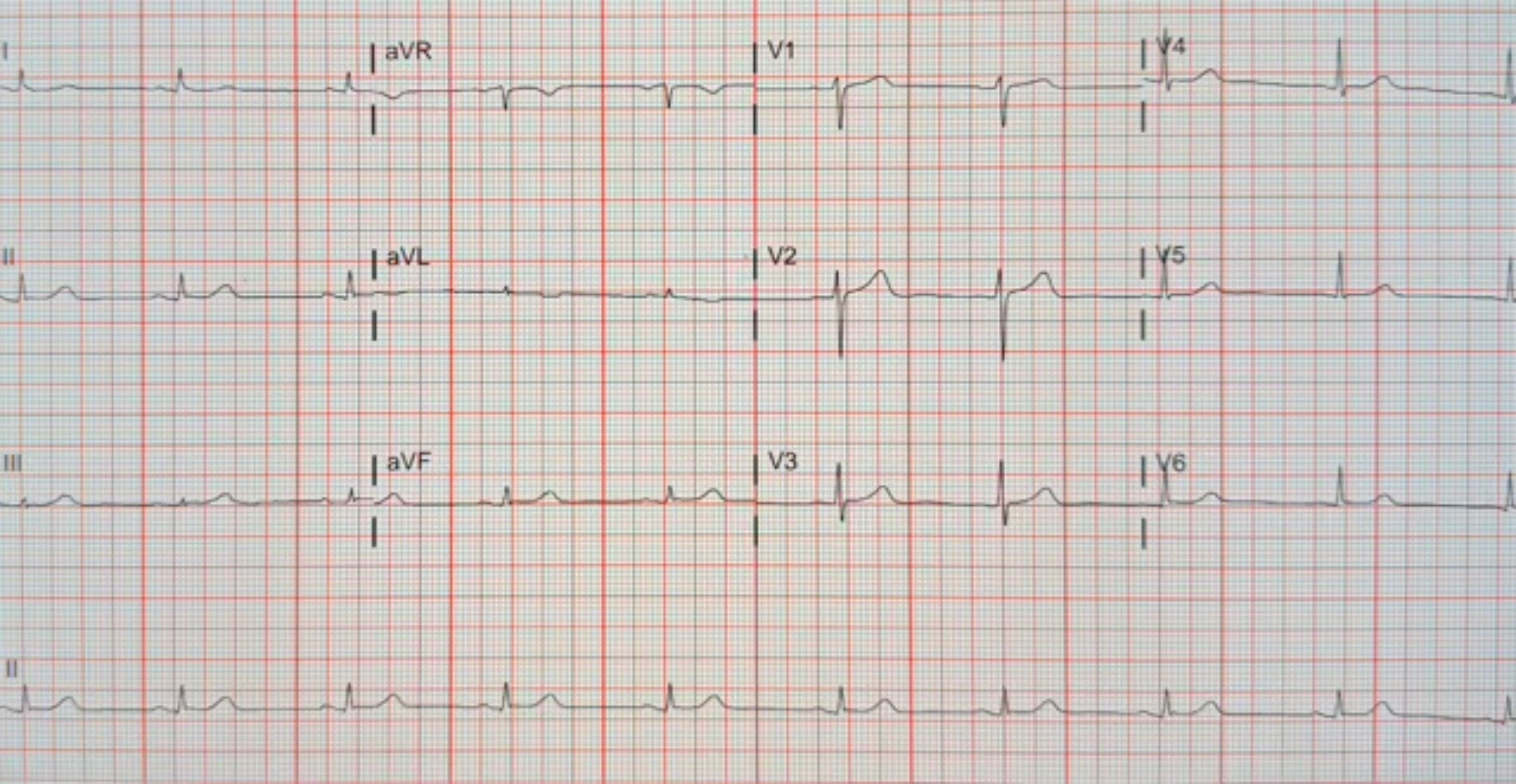
I
aVR
V1
V4
II
aVL
V2
V5
III
aVF
V3
V6
II

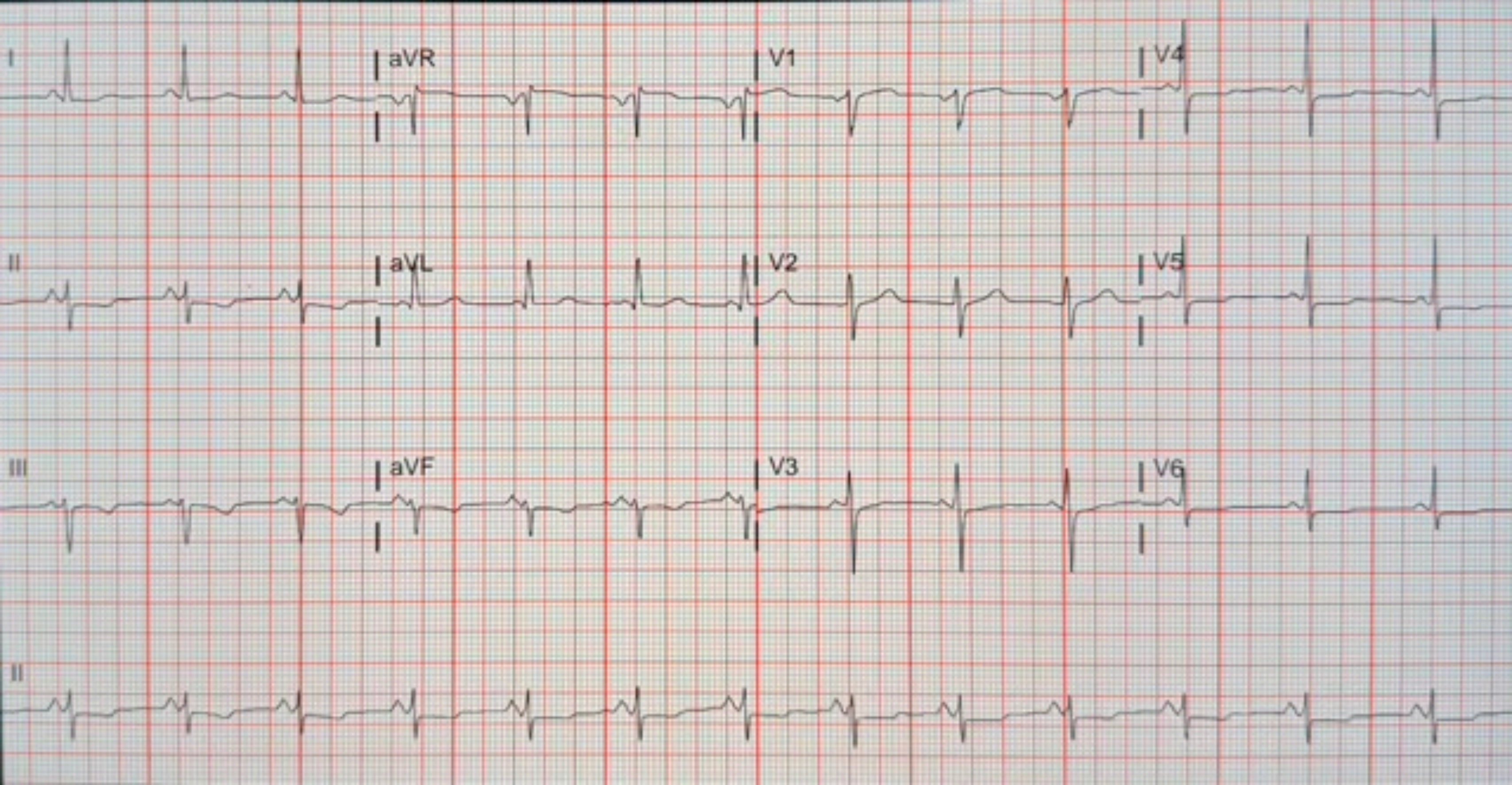
I
aVR
V1
V4
II
aVL
V2
V5
III
aVF
V3
V6
II

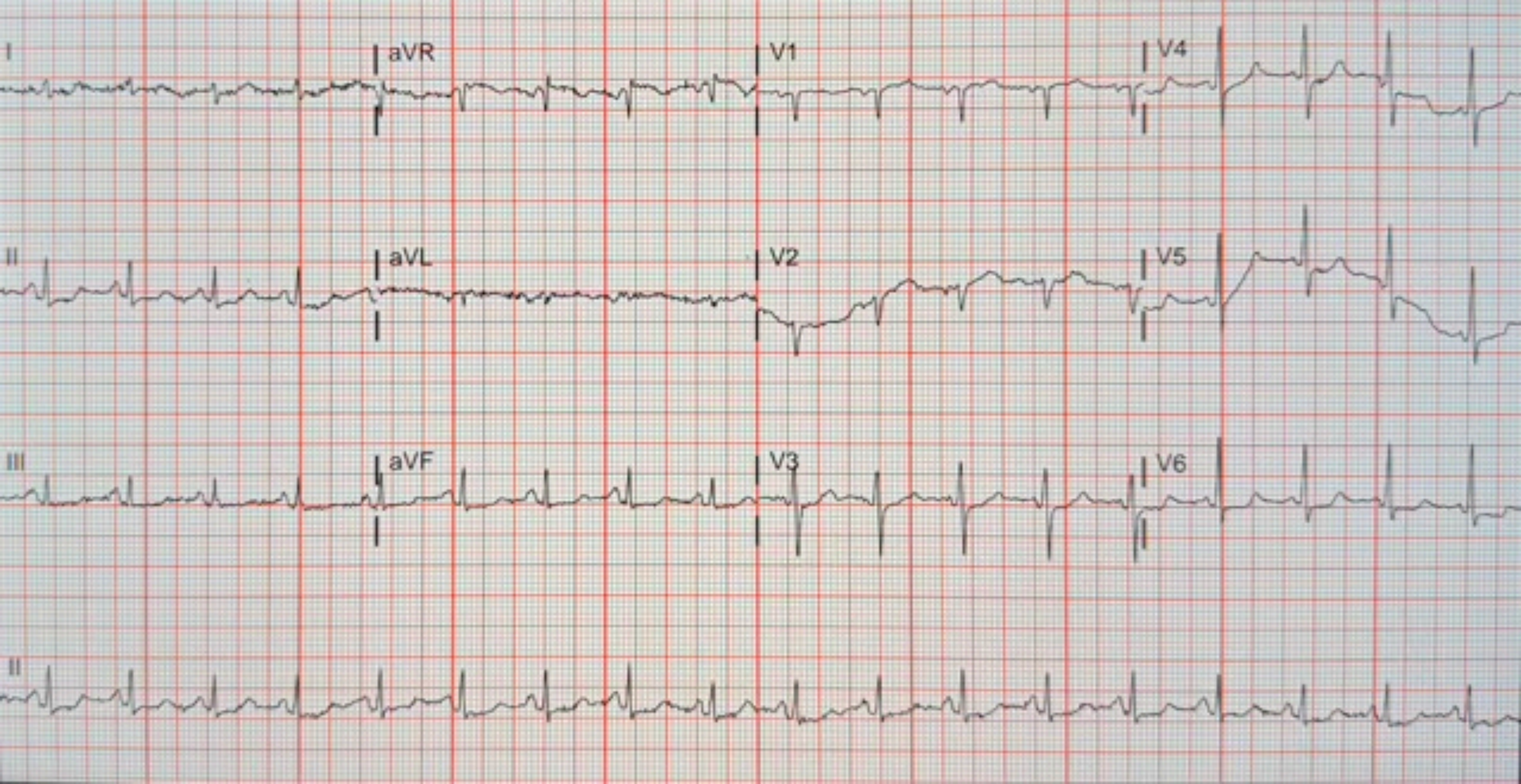
I
aVR
V1
V4
II
aVL
V2
V5
III
aVF
V3
V6
II

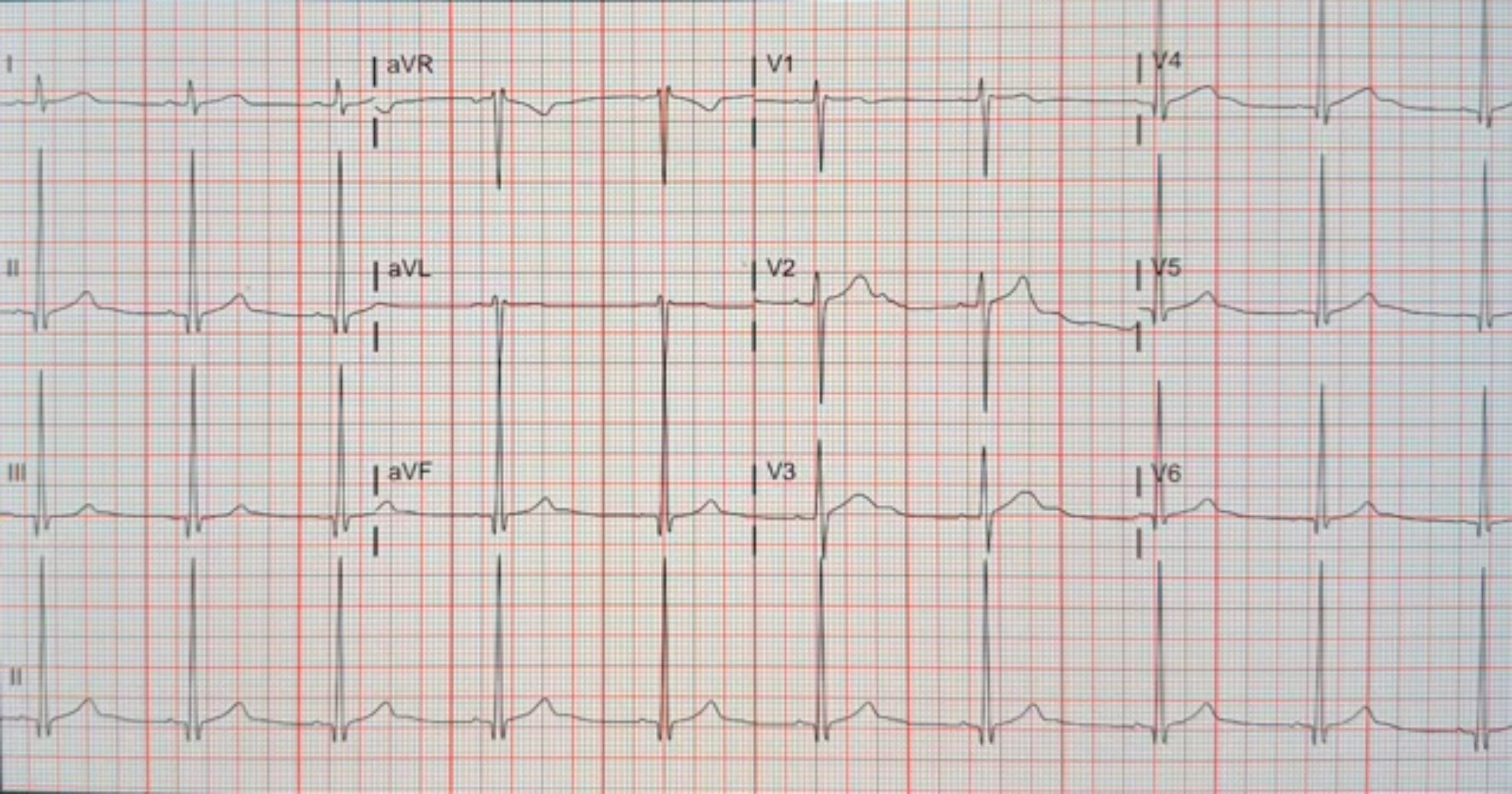
I
aVR
V1
V4
II
aVL
V2
V5
III
aVF
V3
V6
II

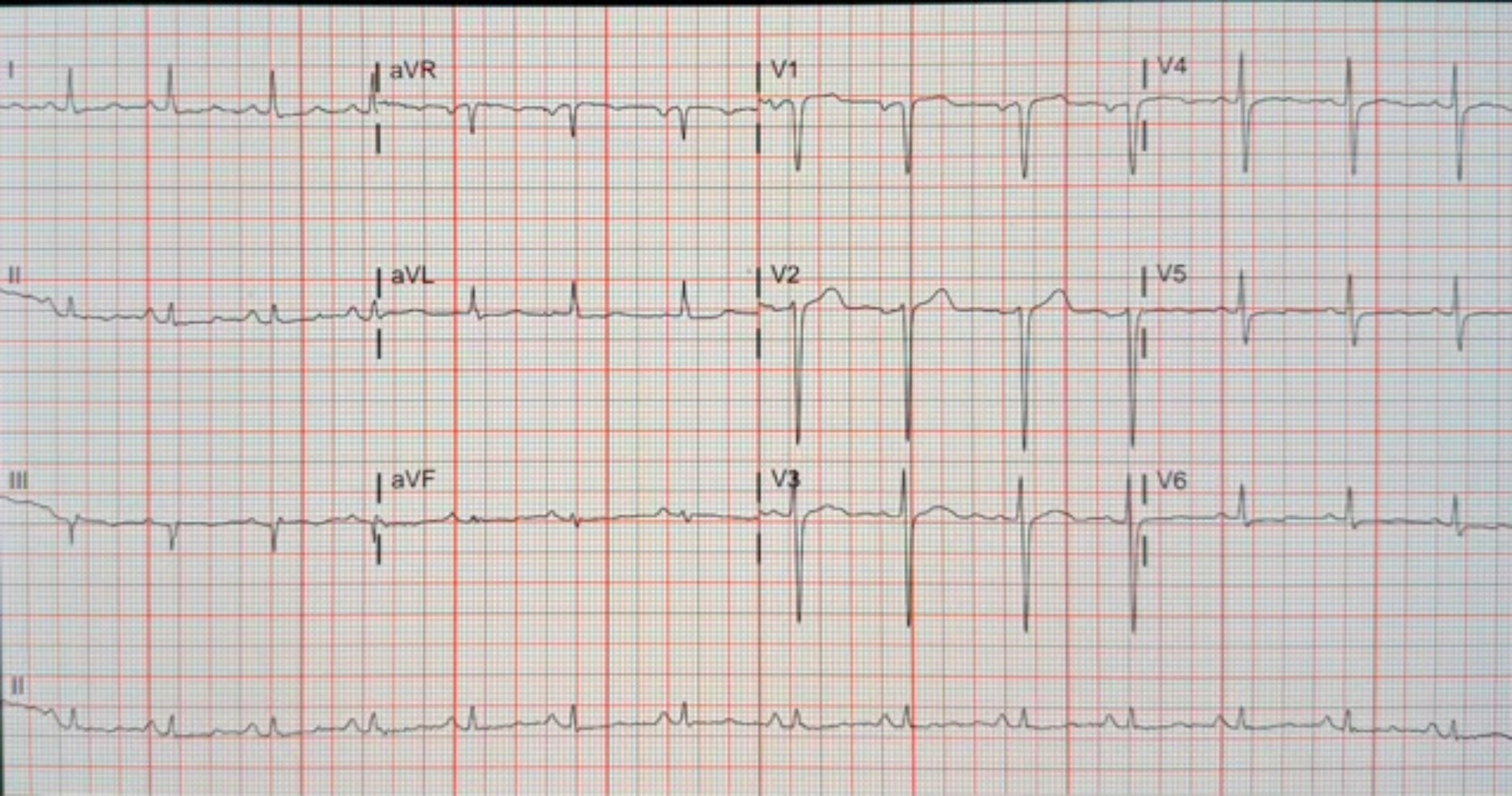
I
aVR
V1
V4
II
aVL
V2
V5
III
aVF
V3
V6
II

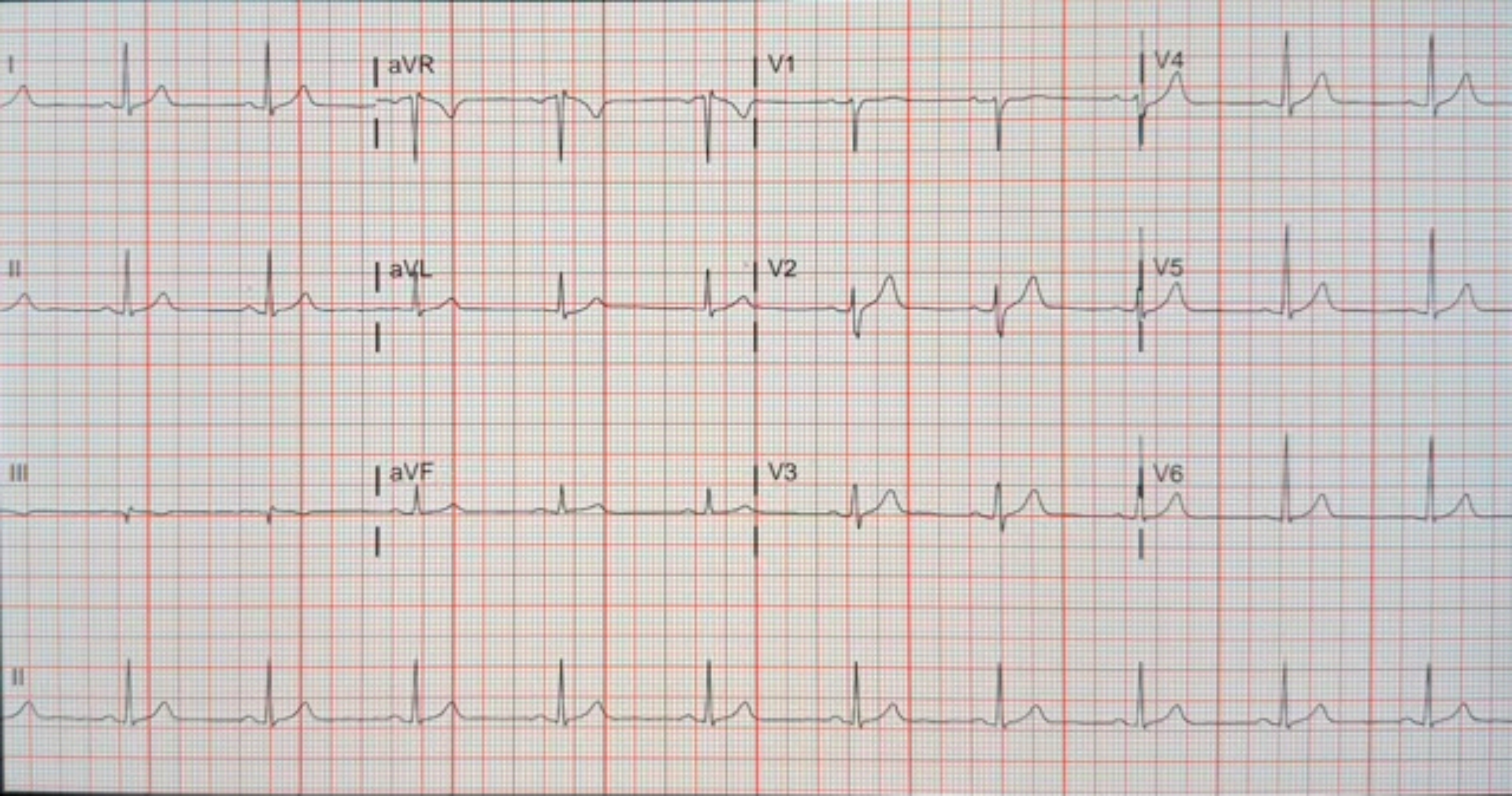
I
aVR
V1
V4
II
aVL
V2
V5
III
aVF
V3
V6
II

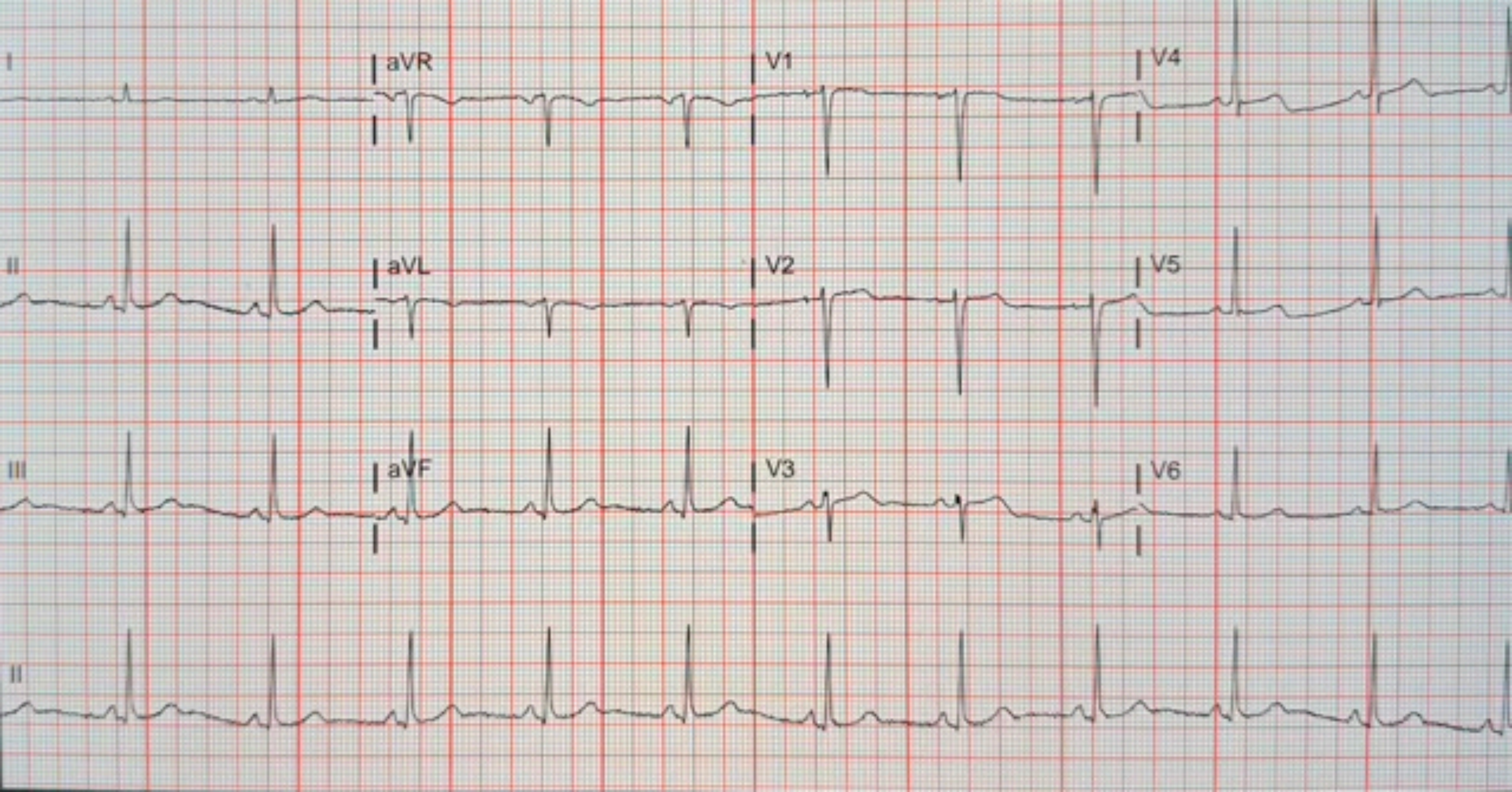
I
aVR
V1
V4
II
aVL
V2
V5
III
aVF
V3
V6
II

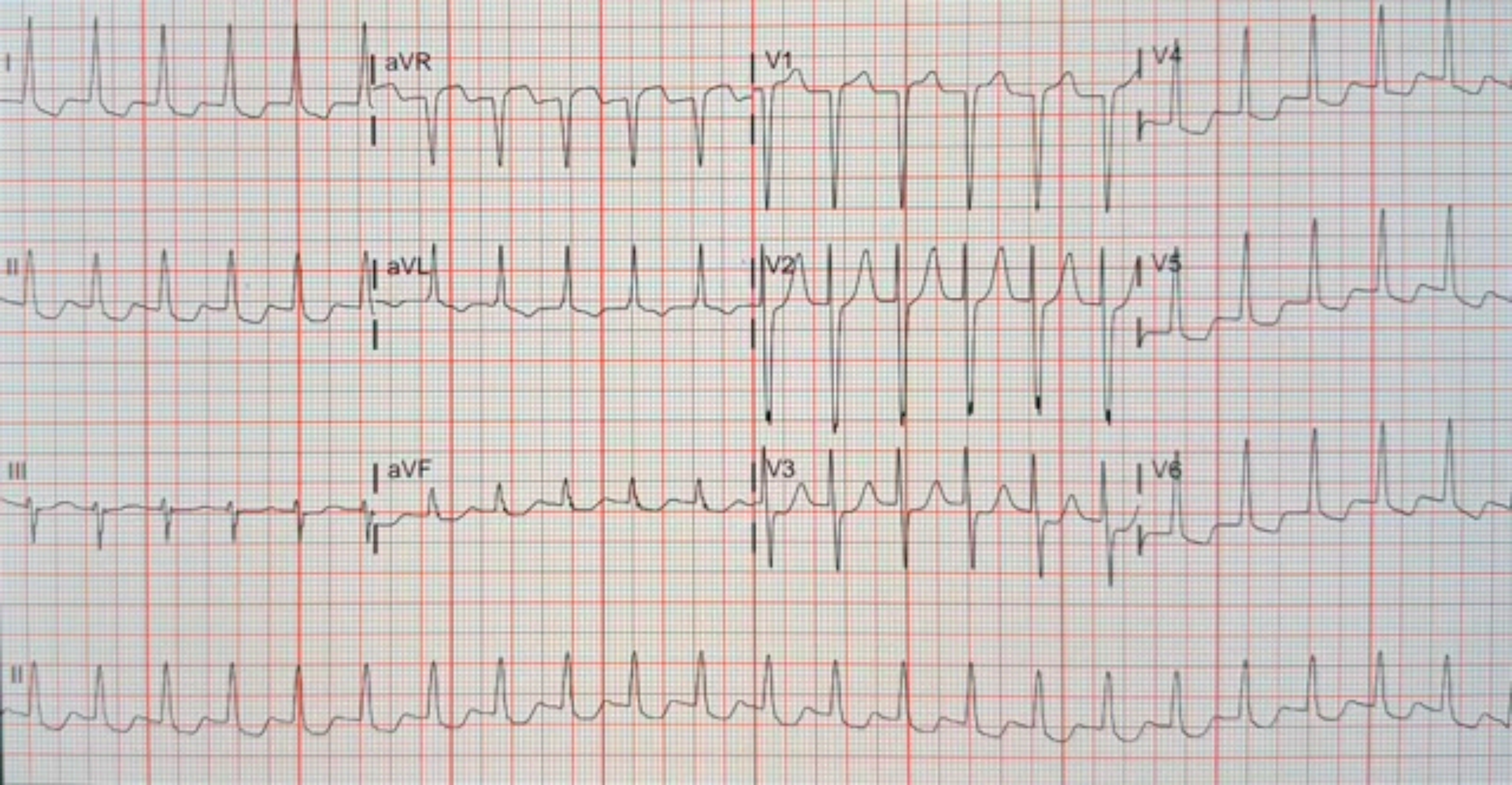
I
aVR
V1
V4
II
aVL
V2
V5
III
aVF
V3
V6
II

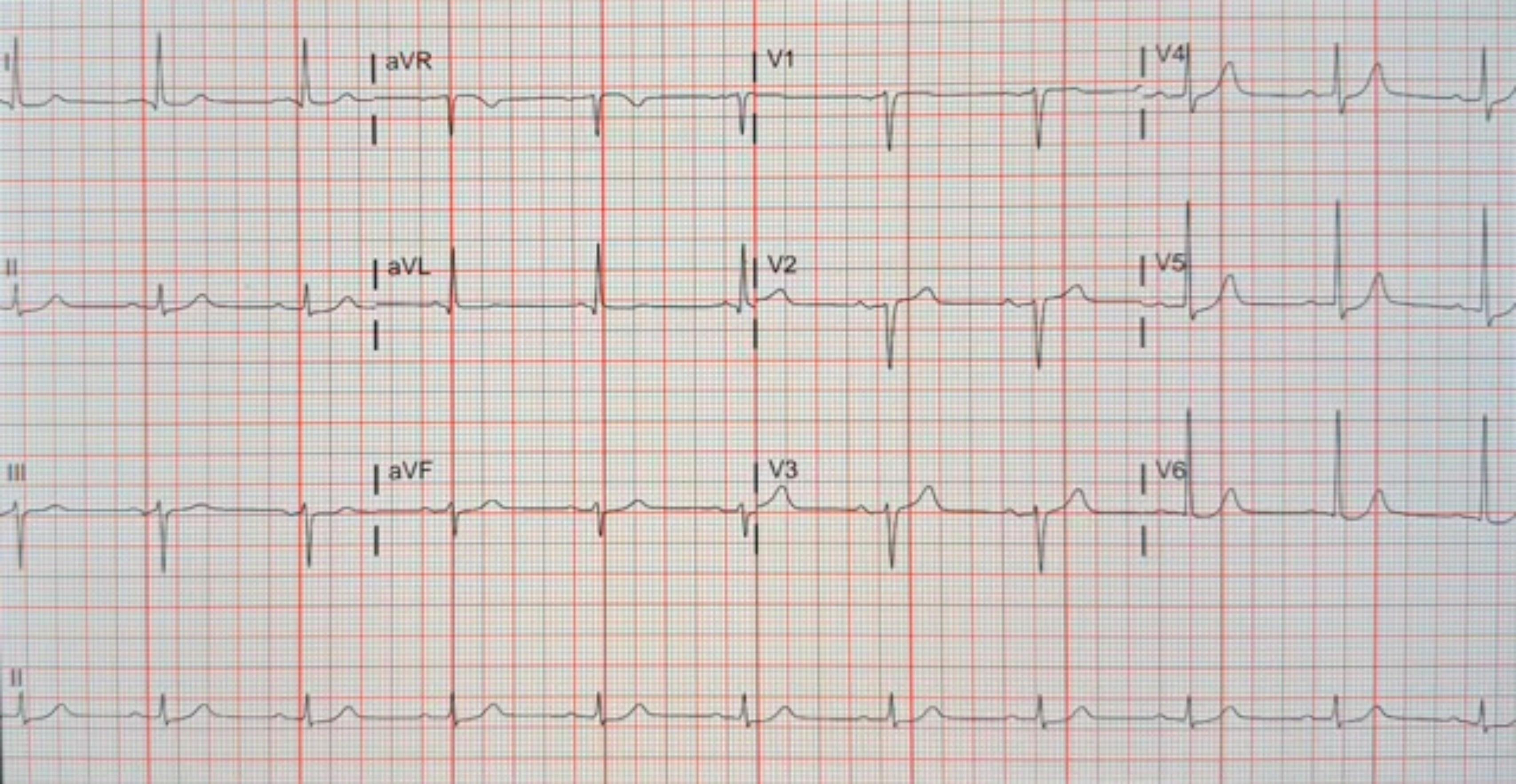
I
aVR
V1
V4
II
aVL
V2
V5
III
aVF
V3
V6
II

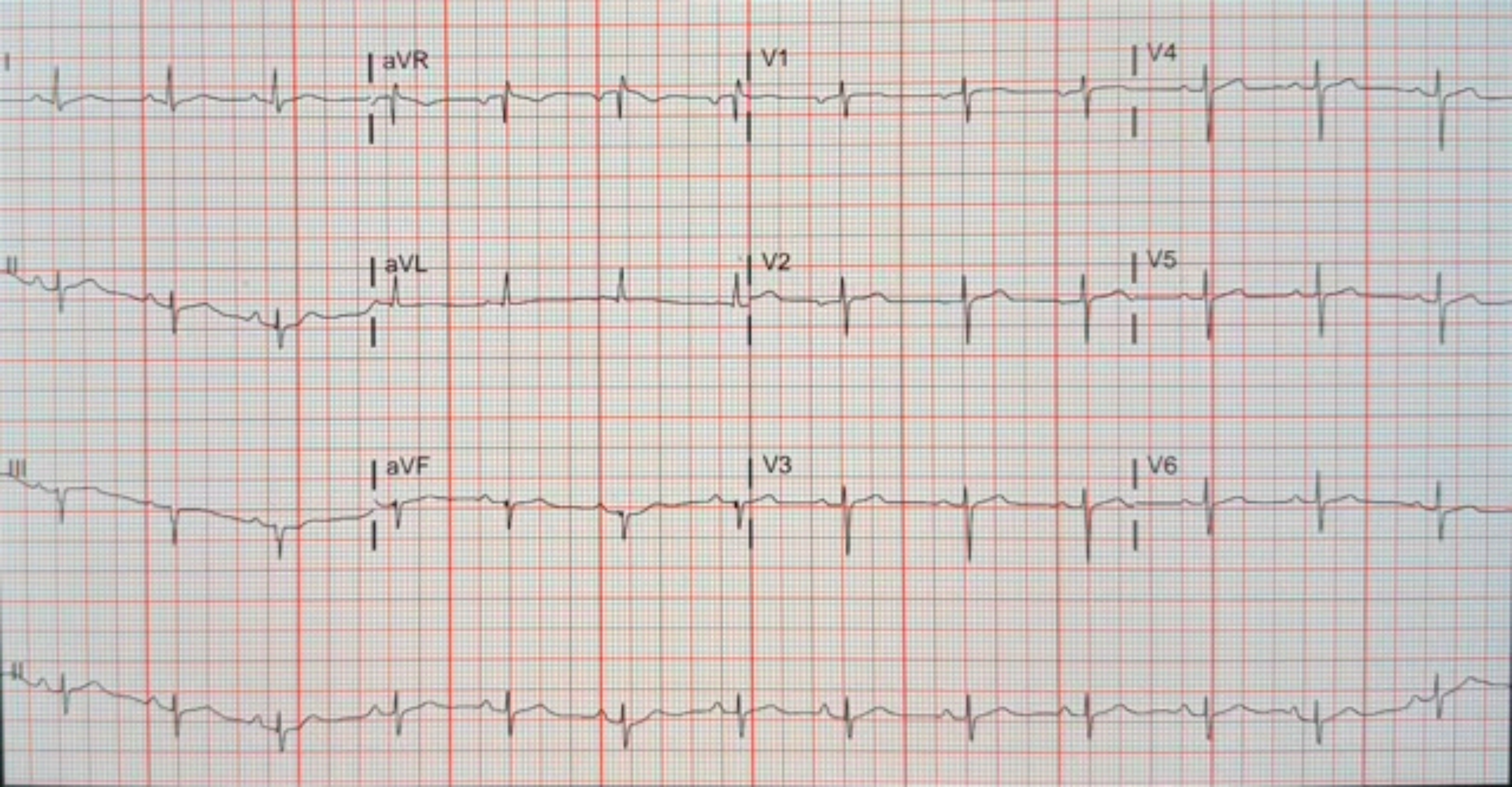
I
aVR
V1
V4
II
aVL
V2
V5
III
aVF
V3
V6
II

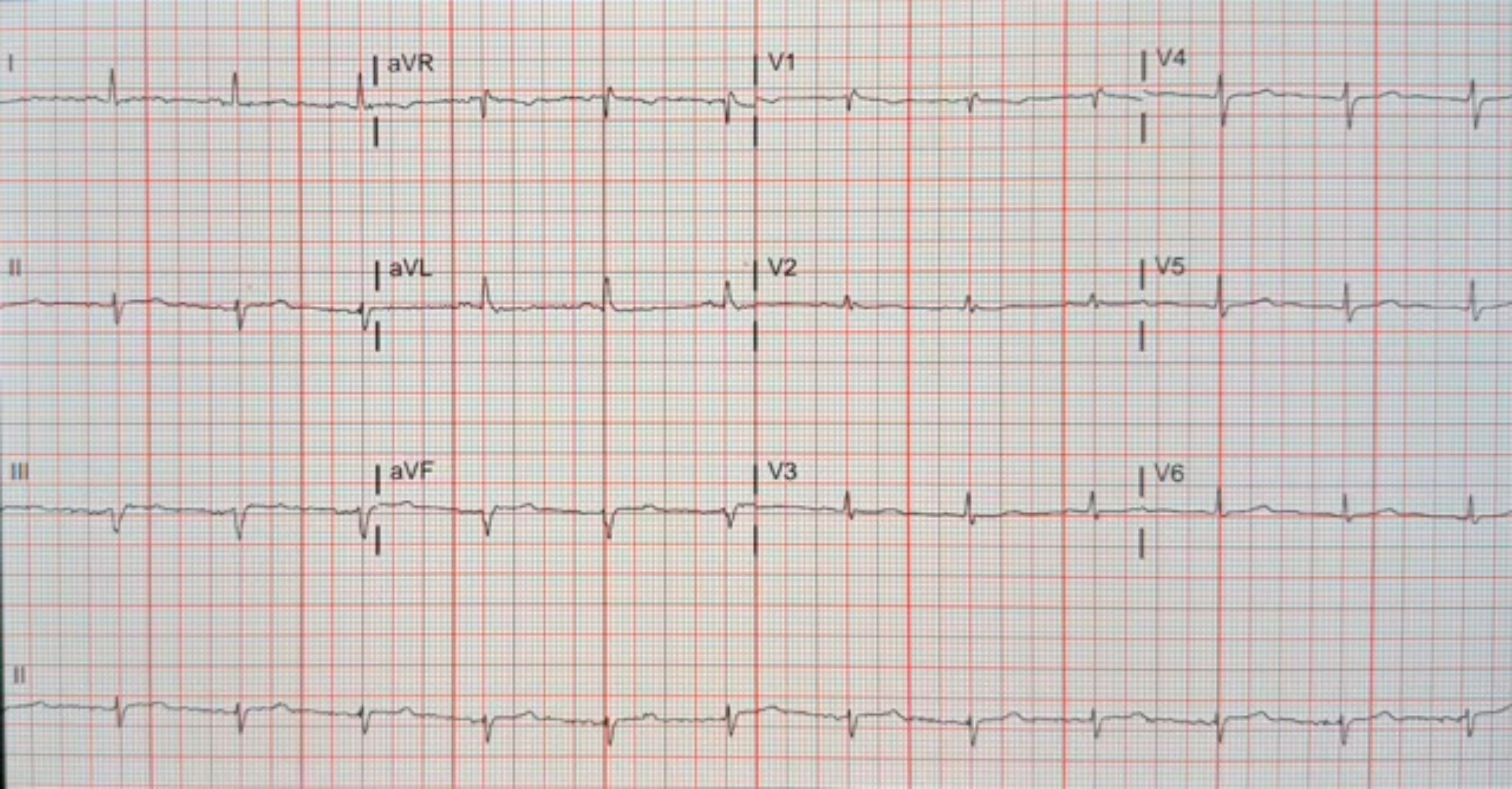

I
aVR
V1
V4
II
aVL
V2
V5
III
aVF
V3
V6
II

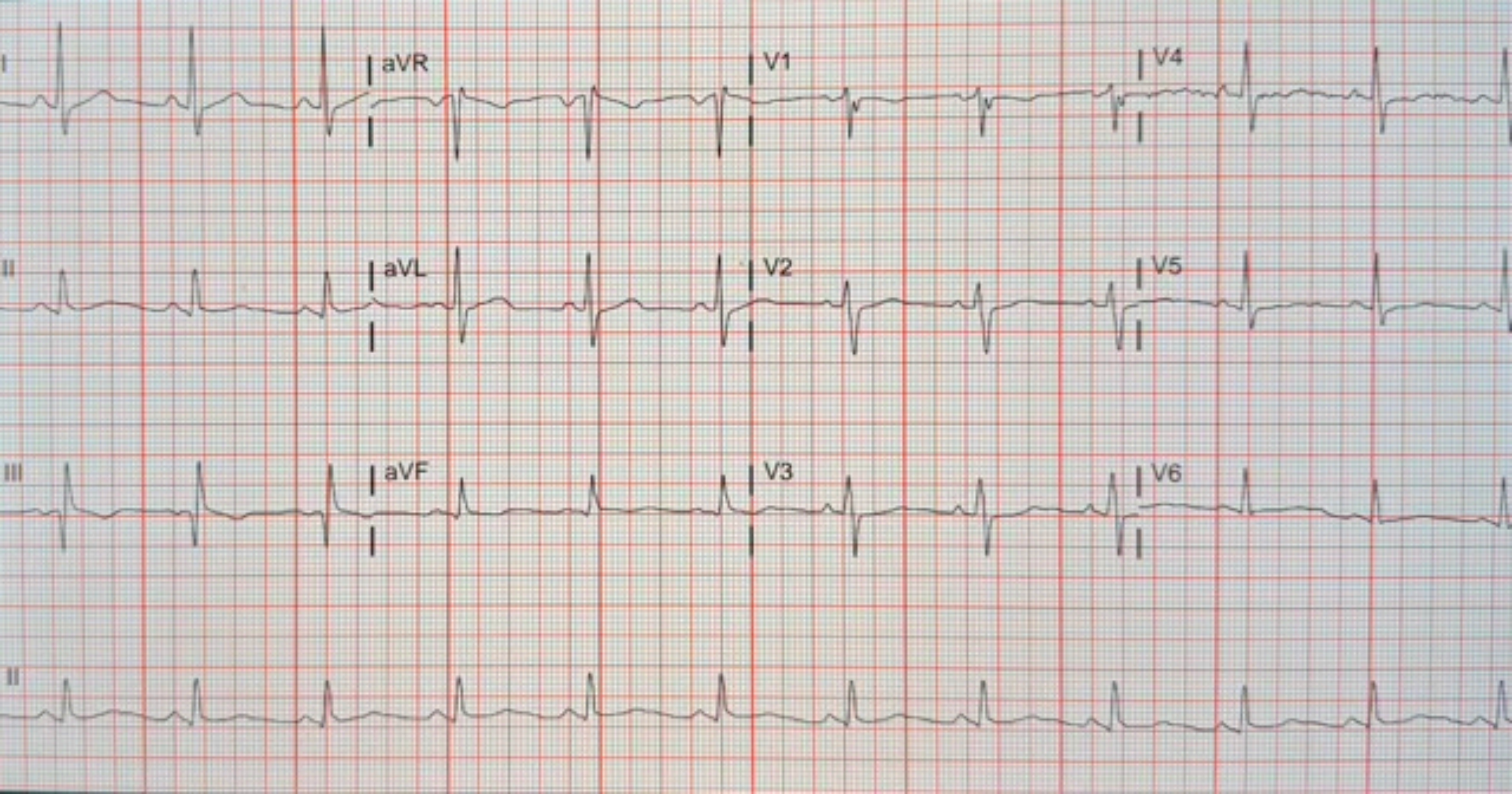
I
aVR
V1
V4
II
aVL
V2
V5
III
aVF
V3
V6
II

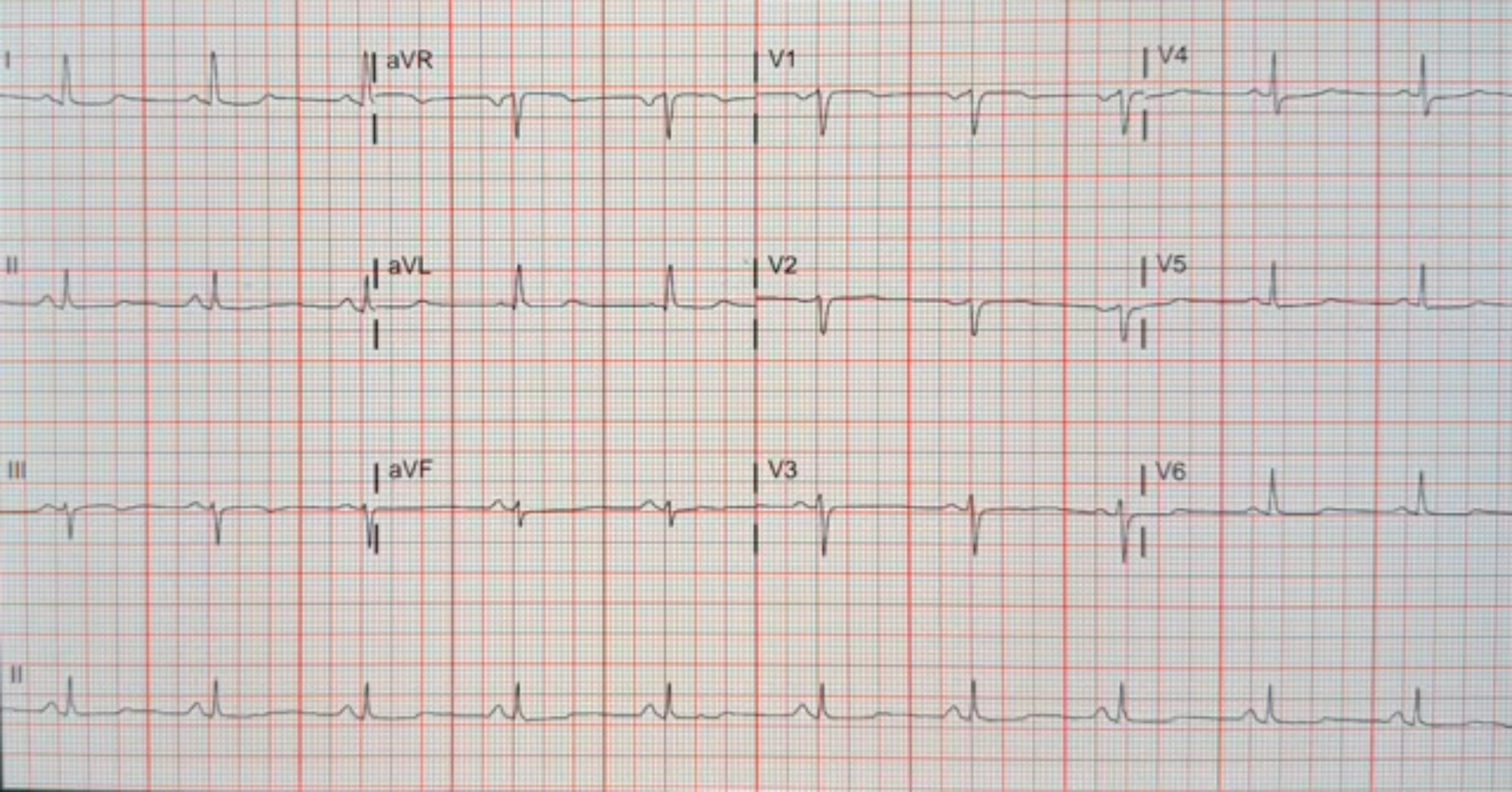
I
aVR
V1
V4
II
aVL
V2
V5
III
aVF
V3
V6
II

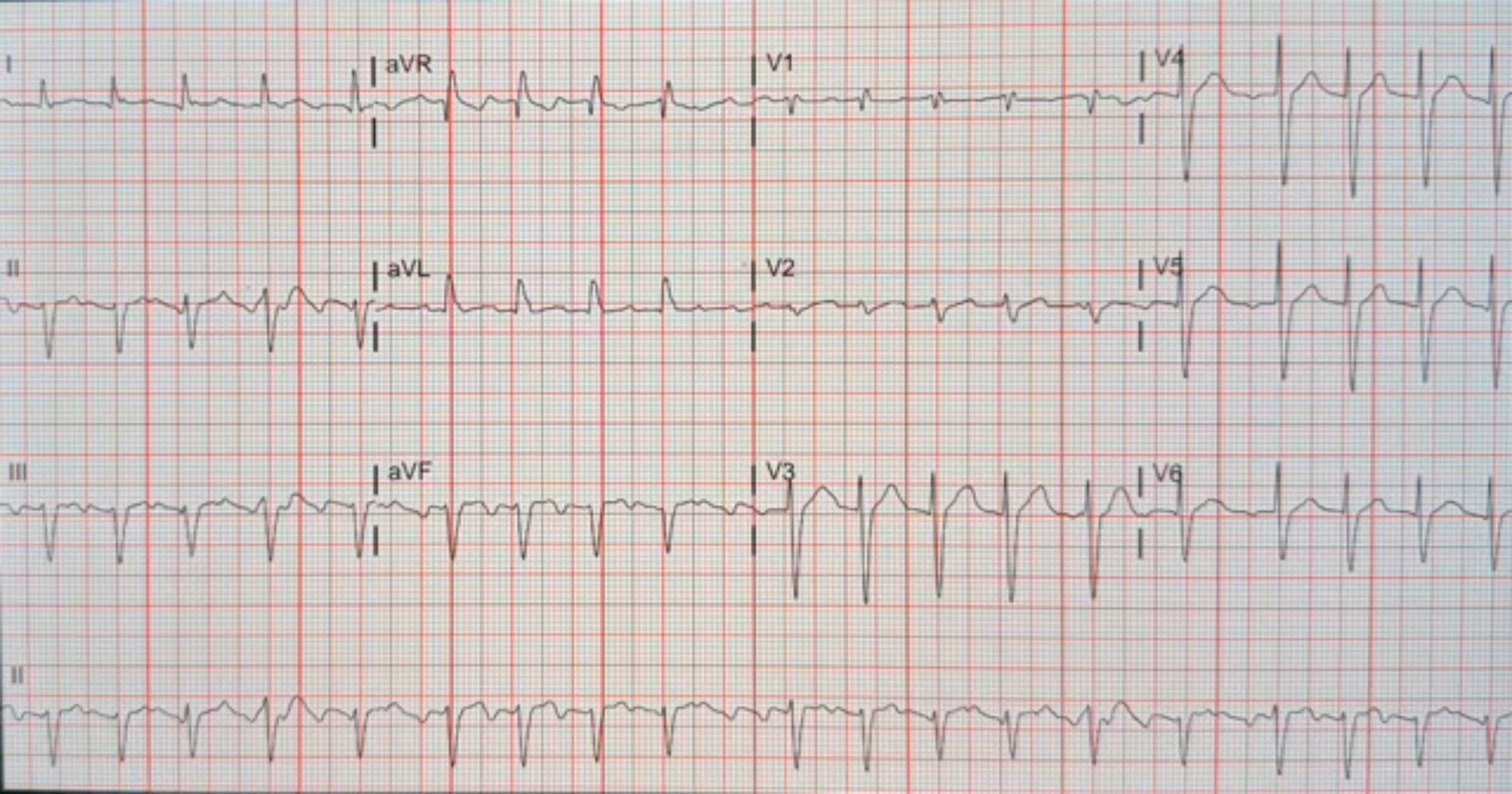
I
aVR
V1
V4
II
aVL
V2
V5
III
aVF
V3
V6
II

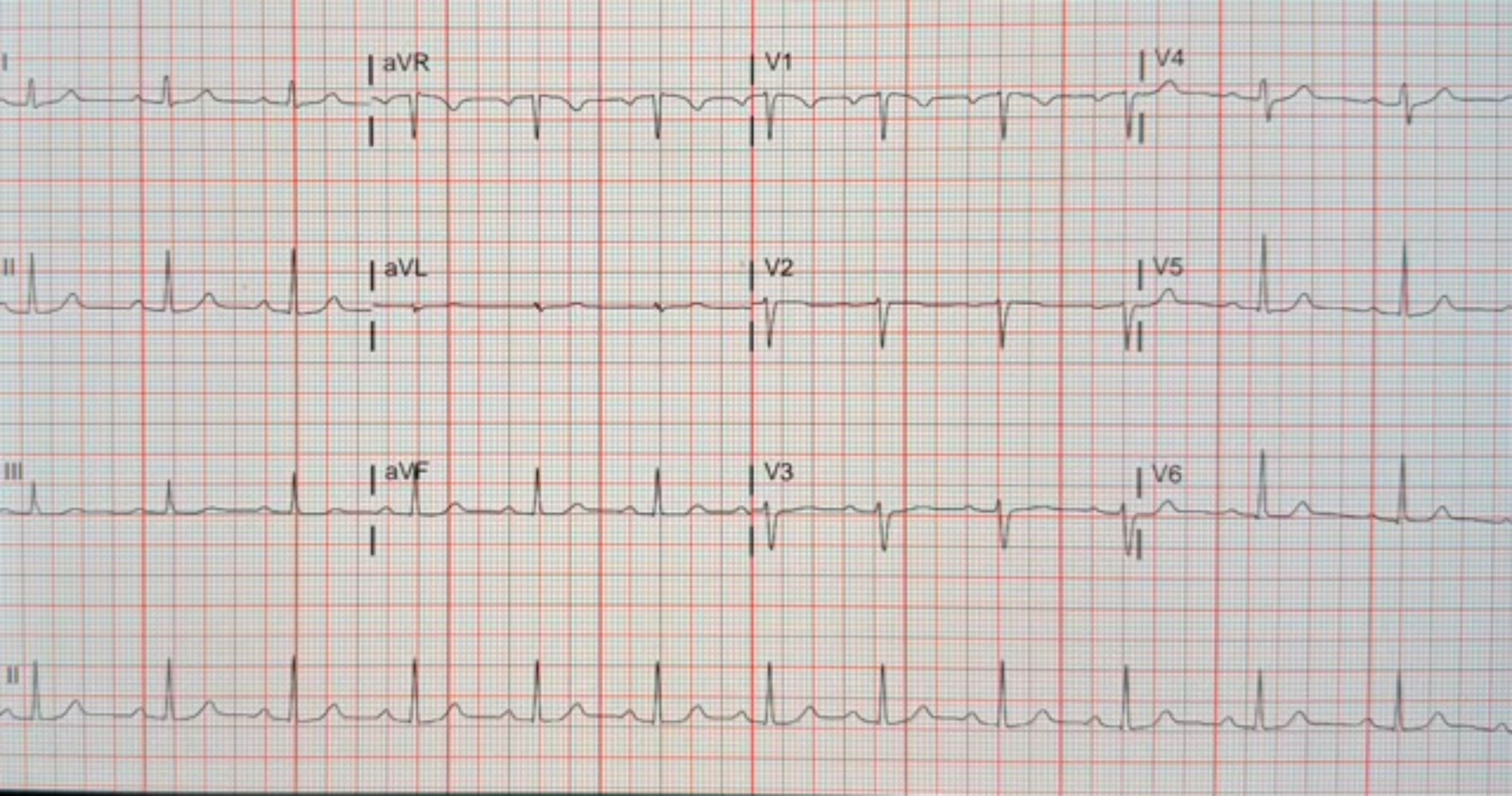

I
aVR
V1
V4
II
aVL
V2
V5
III
aVF
V3
V6
II

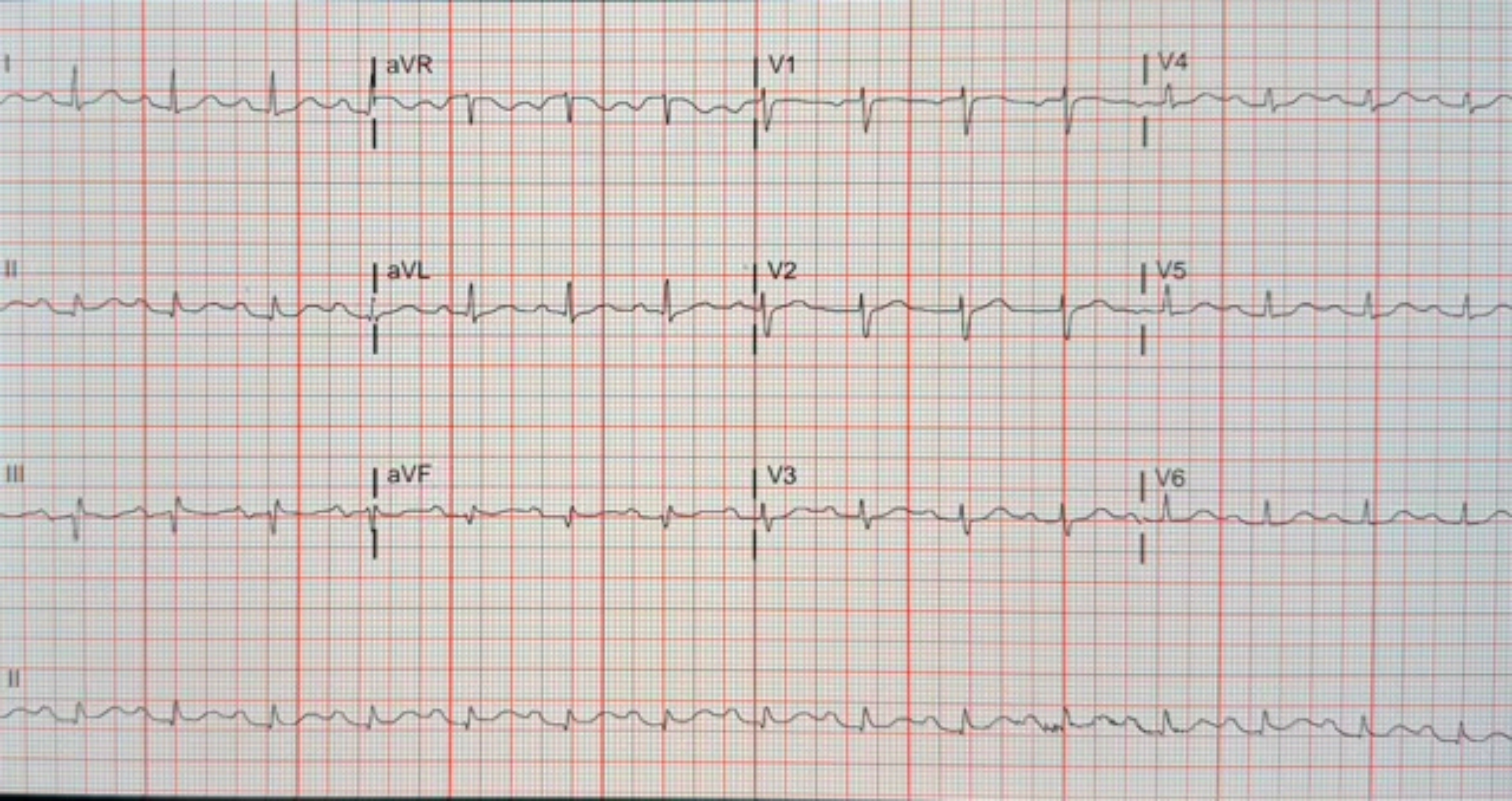
I
aVR
V1
V4
II
aVL
V2
V5
III
aVF
V3
V6
II

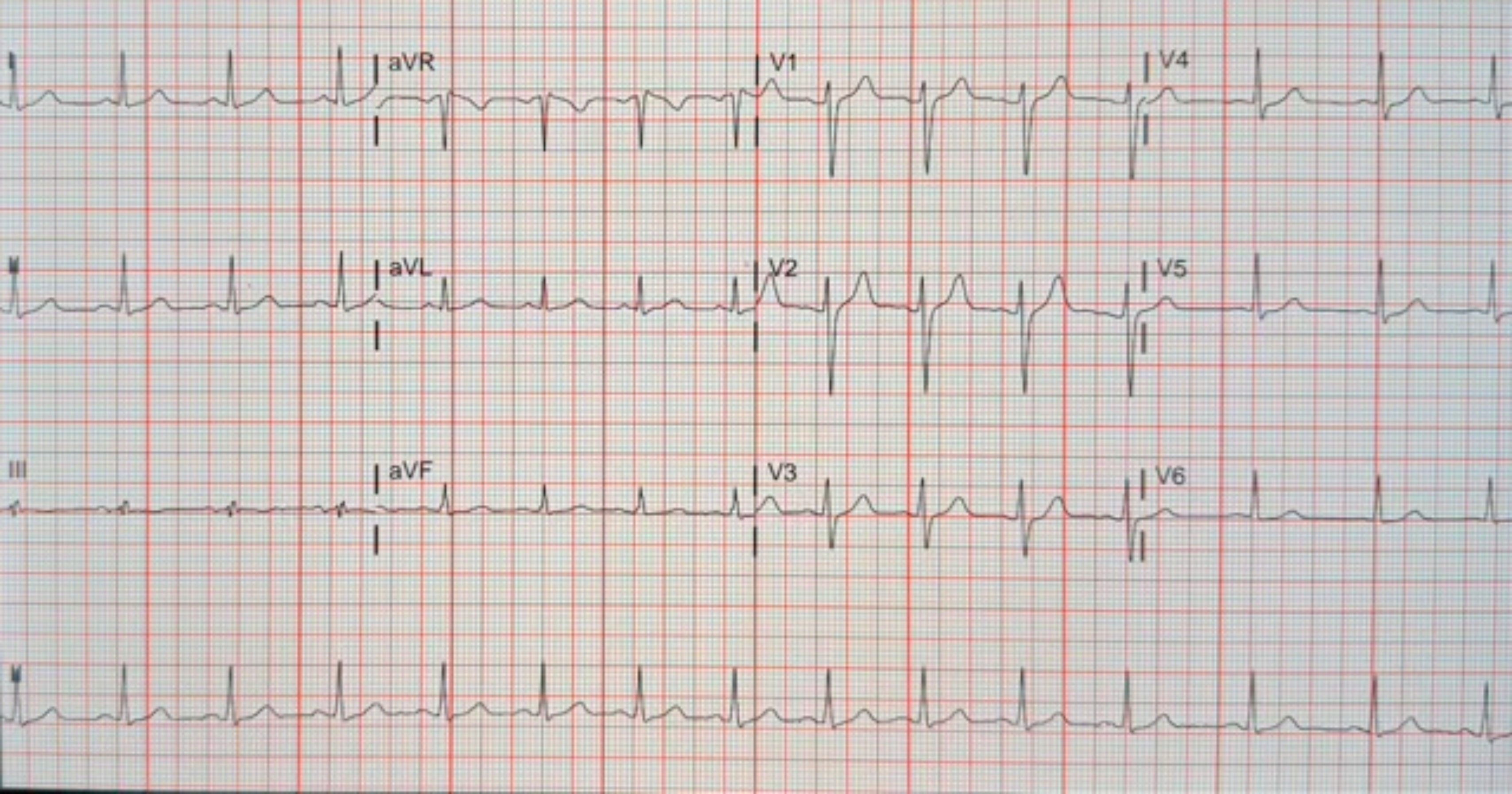
aVR
V1
V4
aVL
V2
V5
III
aVF
V3
V6

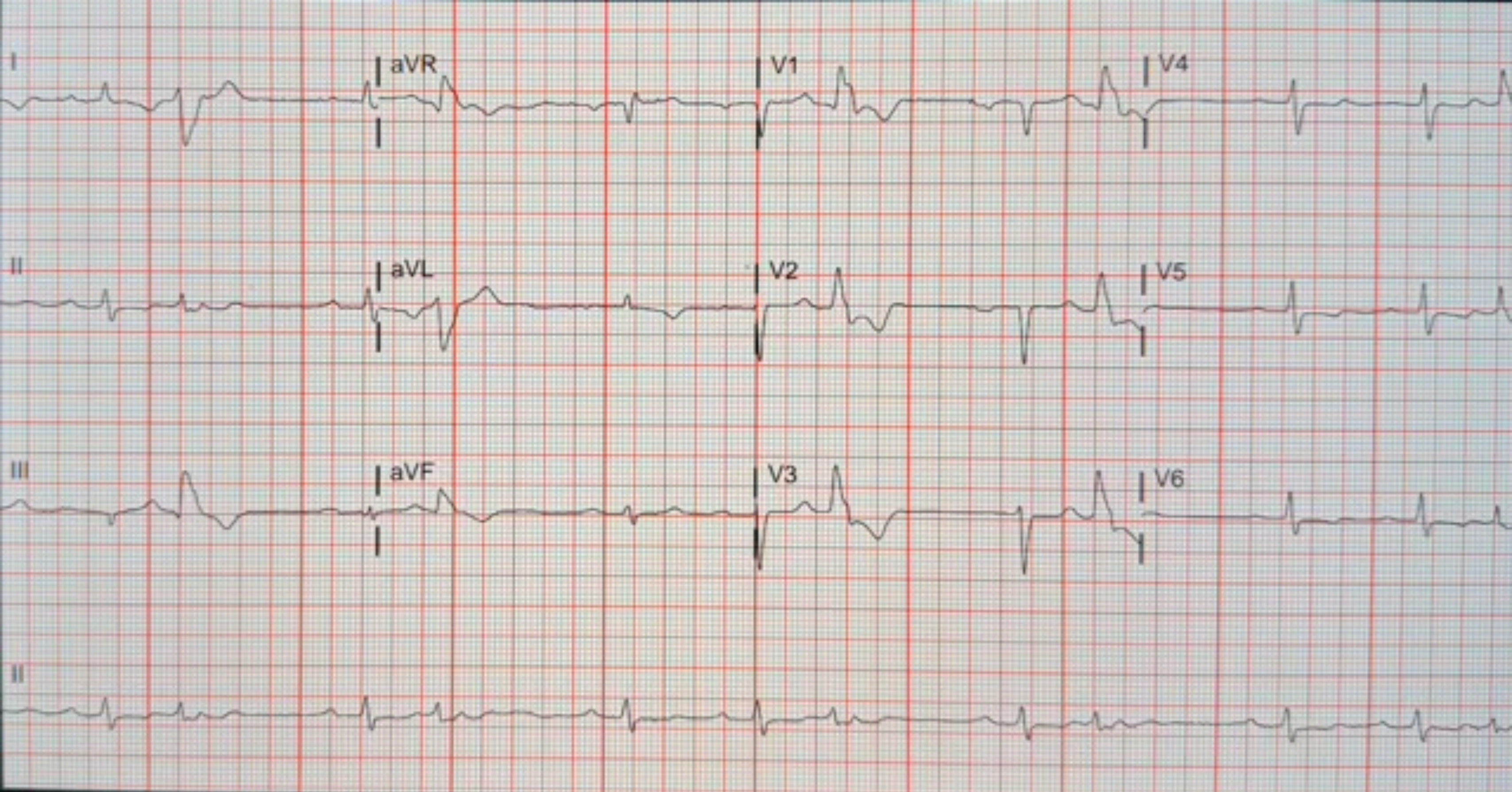
I
aVR
V1
V4
II
aVL
V2
V5
III
aVF
V3
V6
II

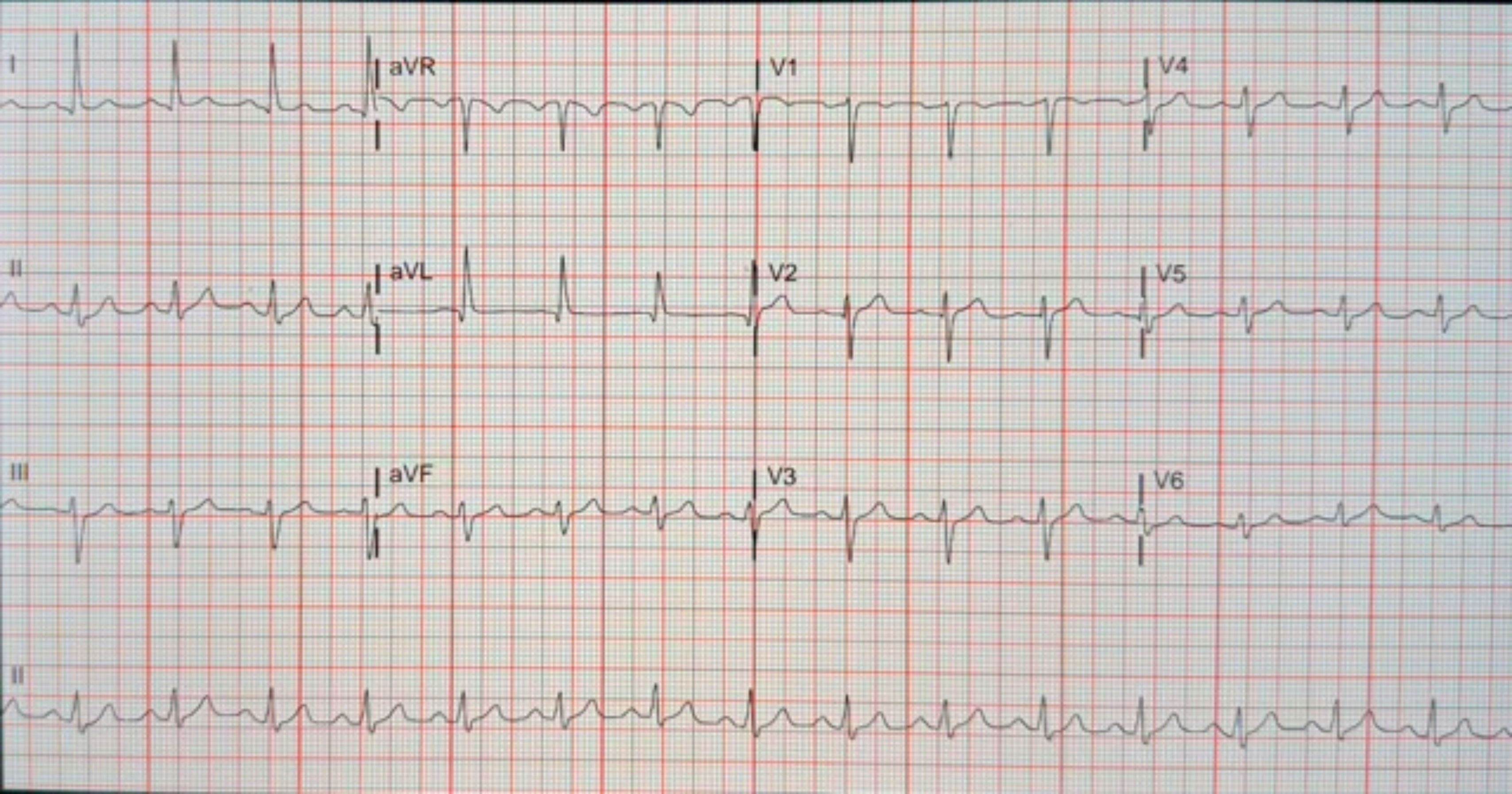
I
aVR
V1
V4
II
aVL
V2
V5
III
aVF
V3
V6
II

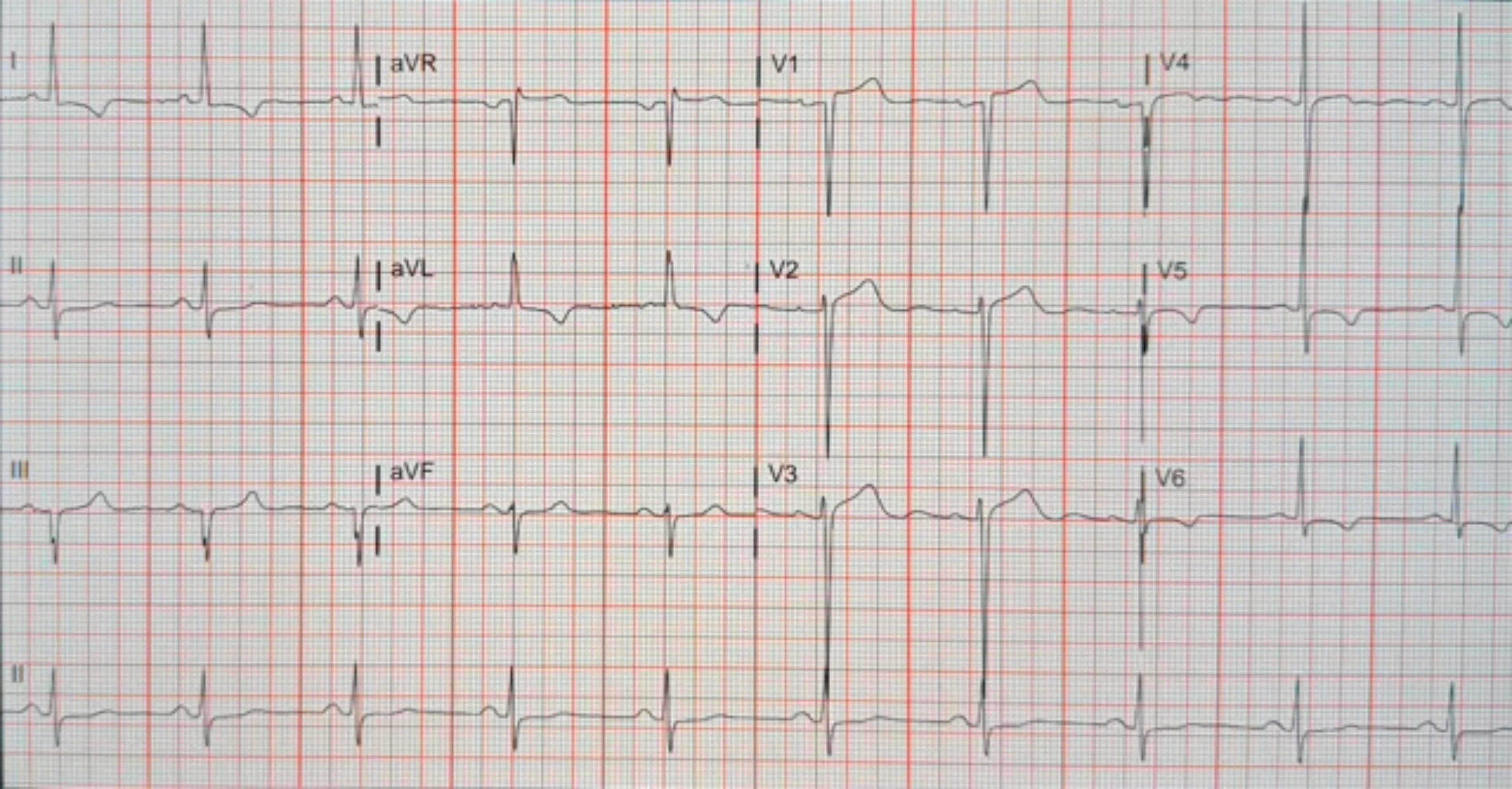
I
aVR
V1
V4
II
aVL
V2
V5
III
aVF
V3
V6
II

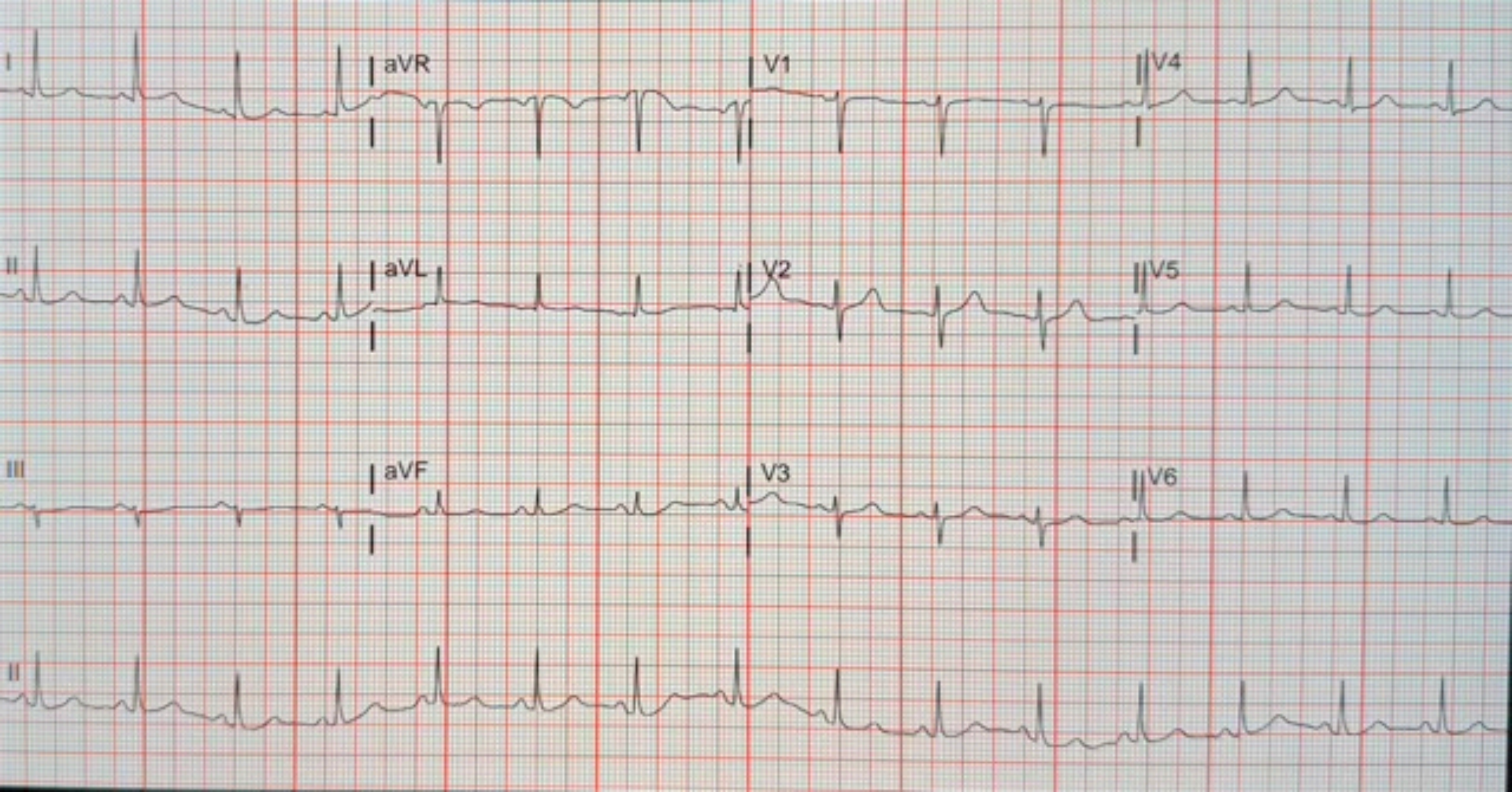
I
aVR
V1
V4
II
aVL
V2
V5
III
aVF
V3
V6
II

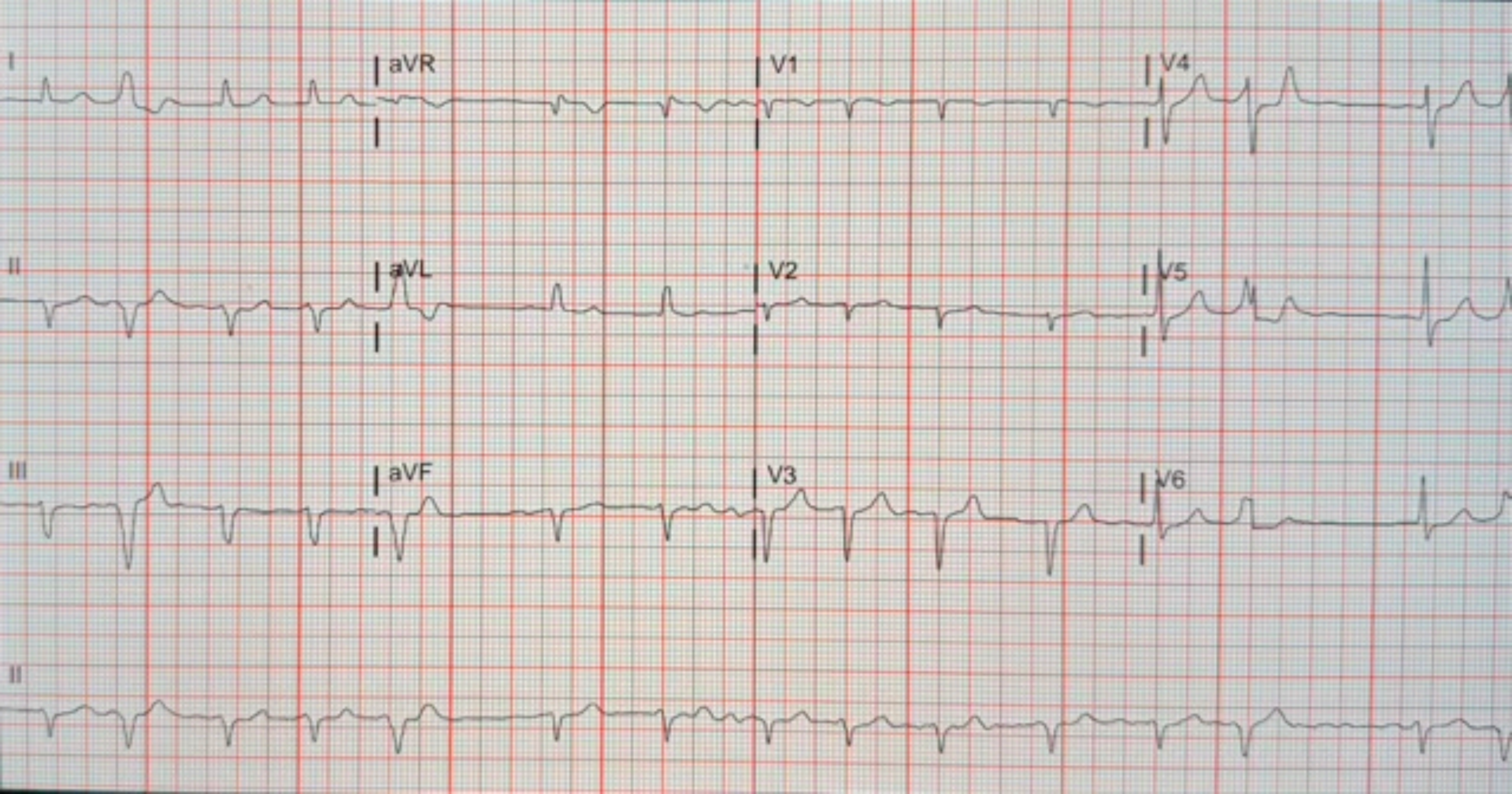
I
aVR
V1
V4
II
aVL
V2
V5
III
aVF
V3
V6
II

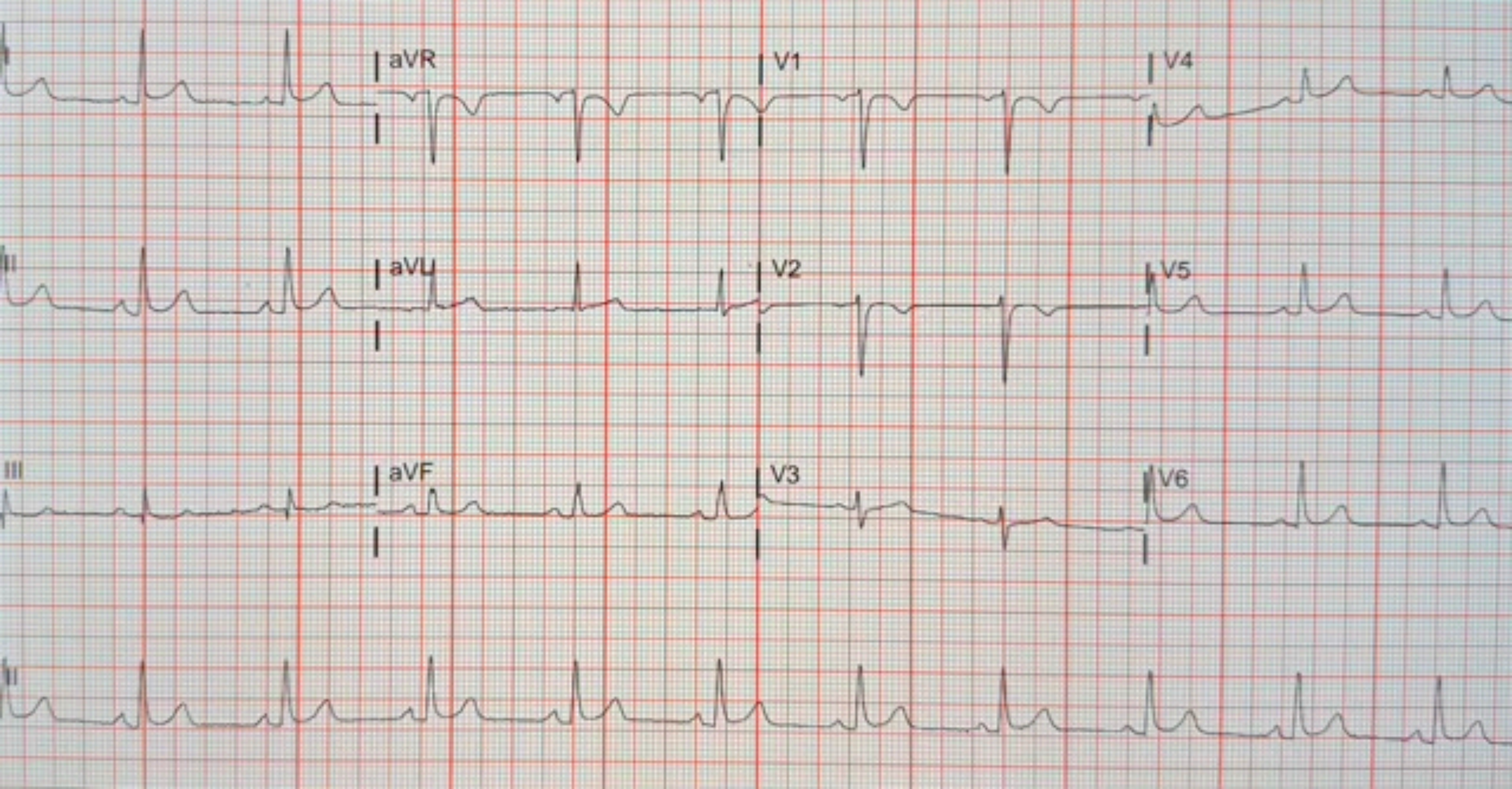
aVR
V1
V4
II
aVL
V2
V5
III
aVF
V3
V6
II

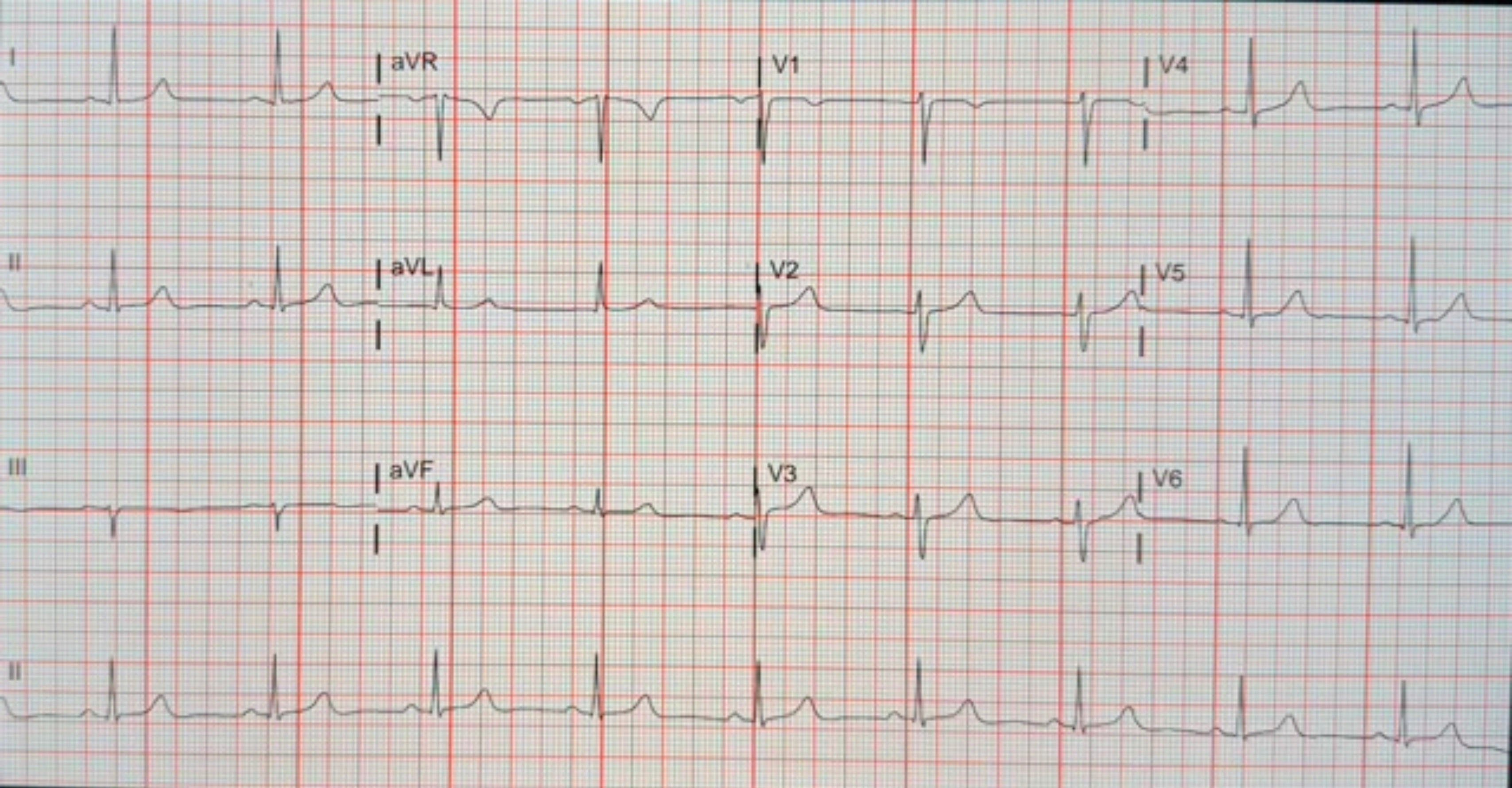
I
aVR
V1
V4
II
aVL
V2
V5
III
aVF
V3
V6
II

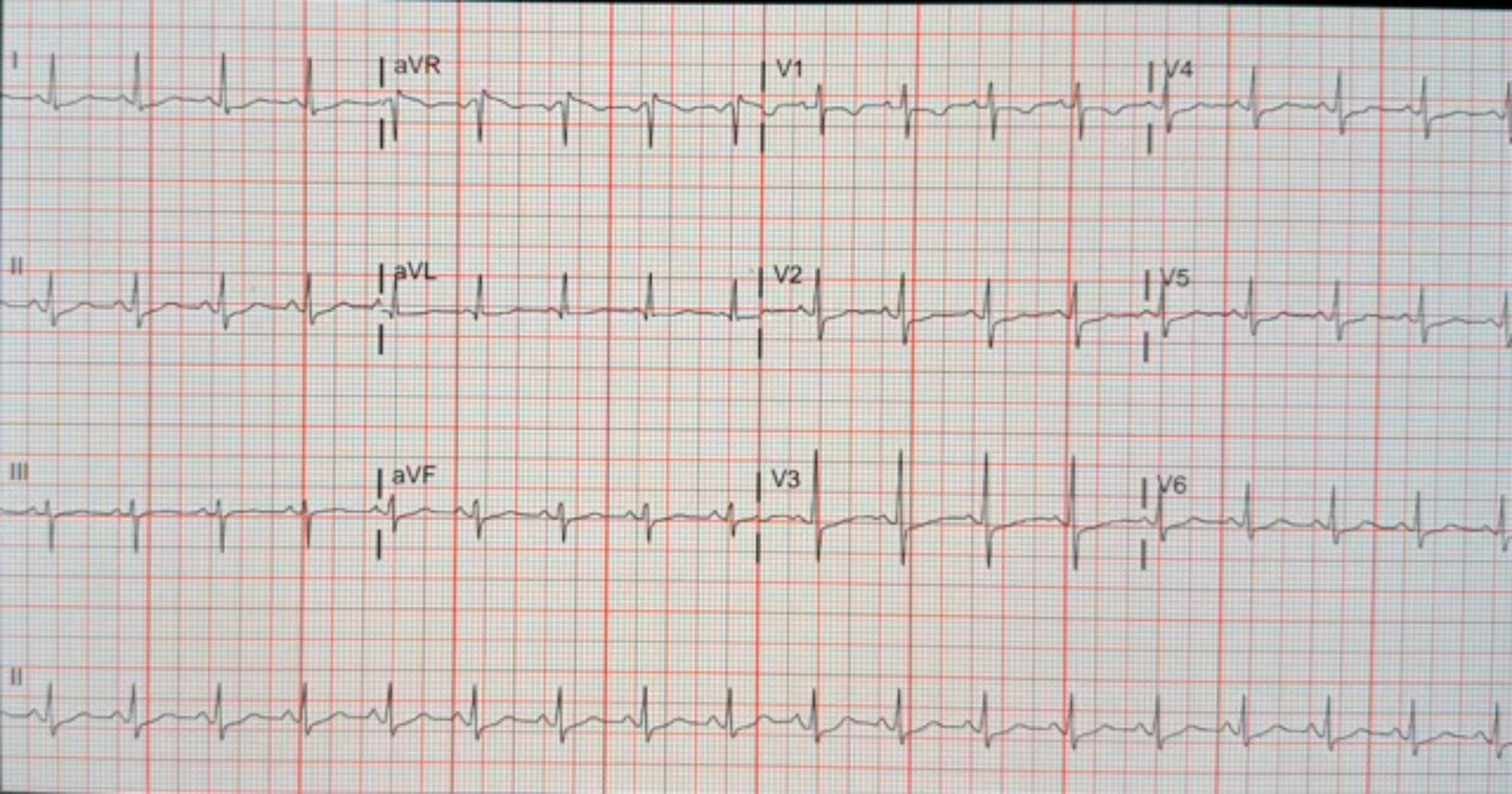
I
aVR
V1
V4
II
aVL
V2
V5
III
aVF
V3
V6
II

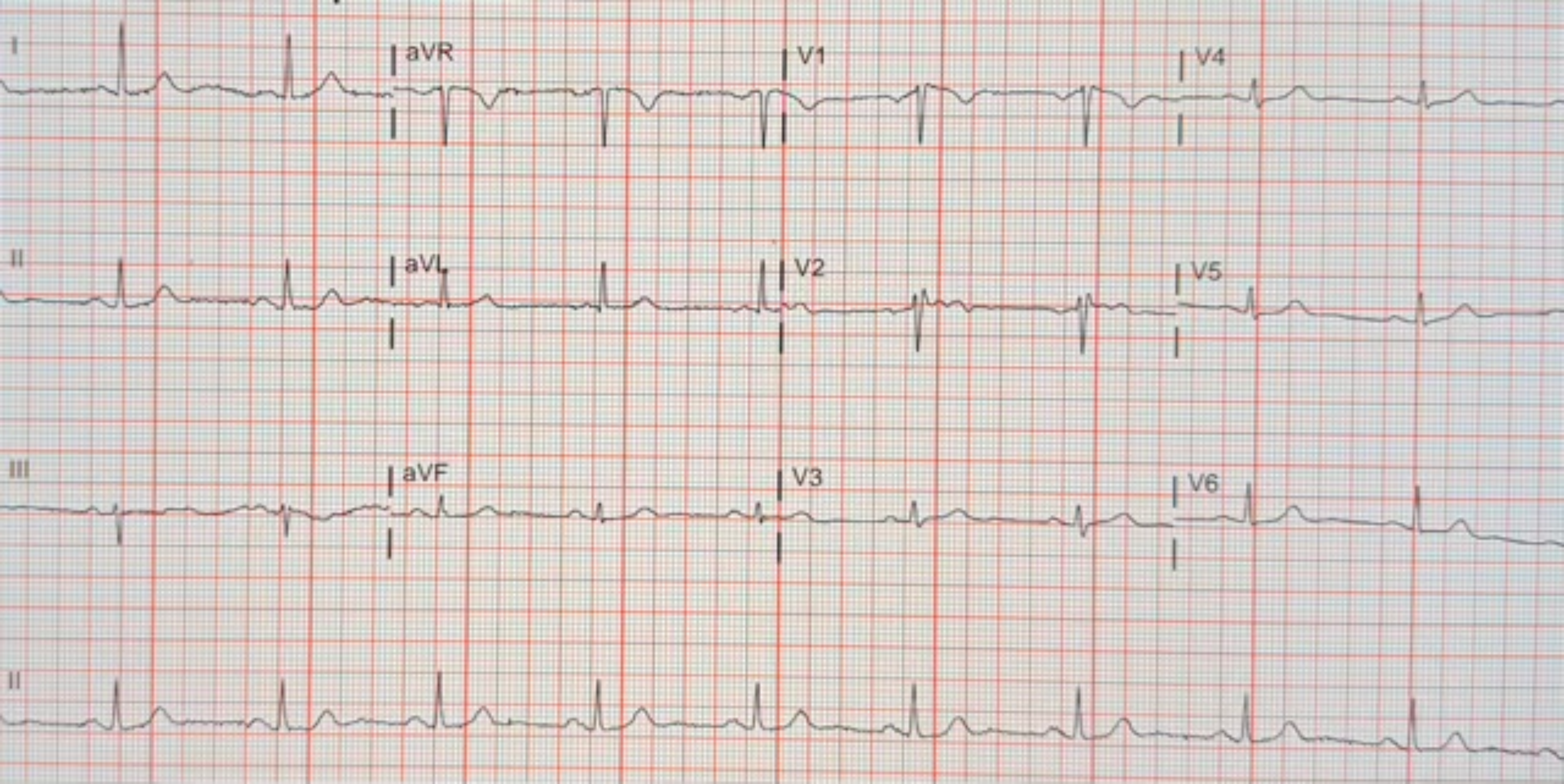
I
aVR
V1
V4
II
aVL
V2
V5
III
aVF
V3
V6
II

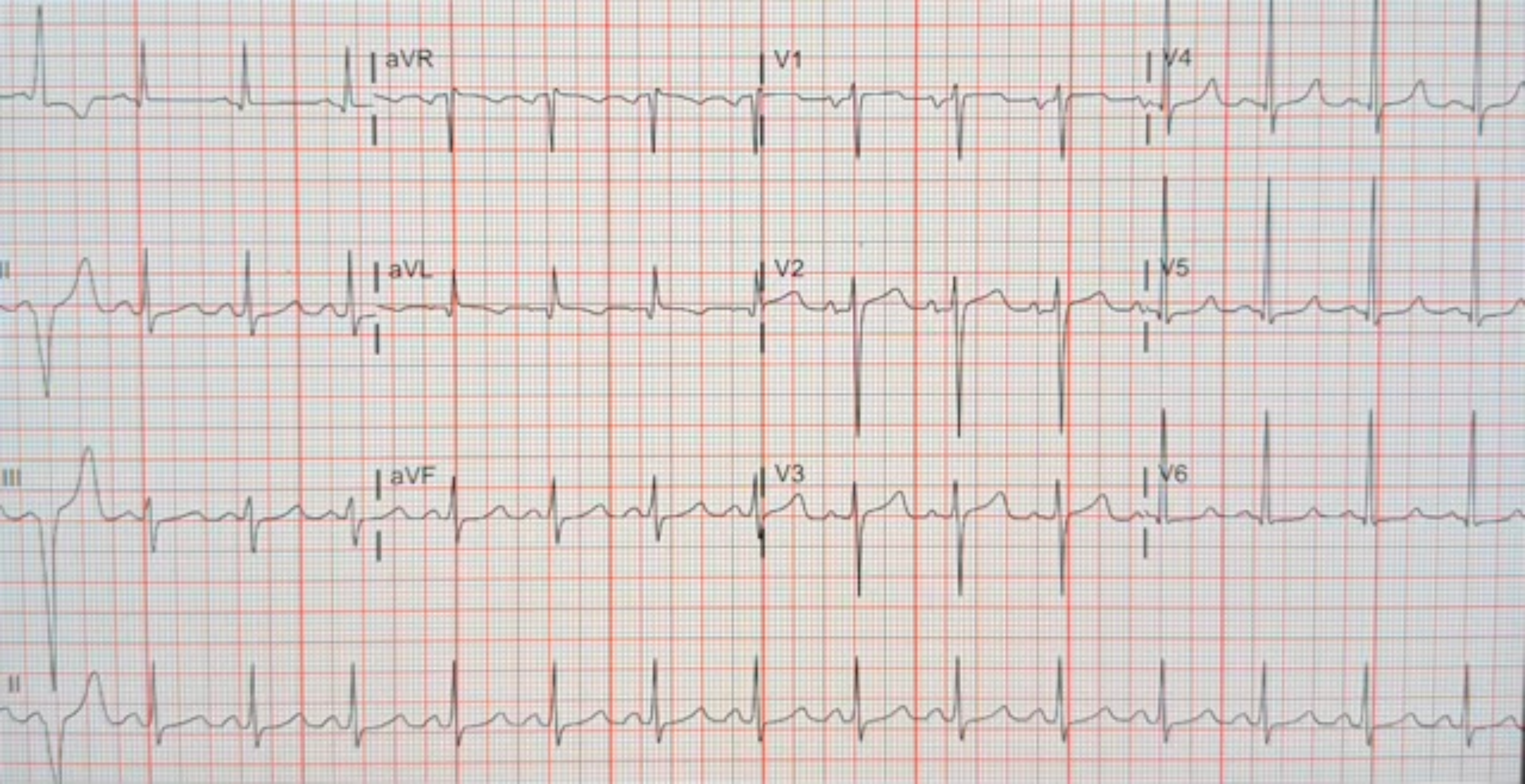
aVR
V1
V4
II
aVL
V2
V5
III
aVF
V3
V6
II

### Section 9. Smartphone photographs of 100 electrocardiogram printouts
### (30 Apple iPhone 15 Pro, 40 Apple iPhone 12, 30 Samsung Galaxy S21 5G; cropped)

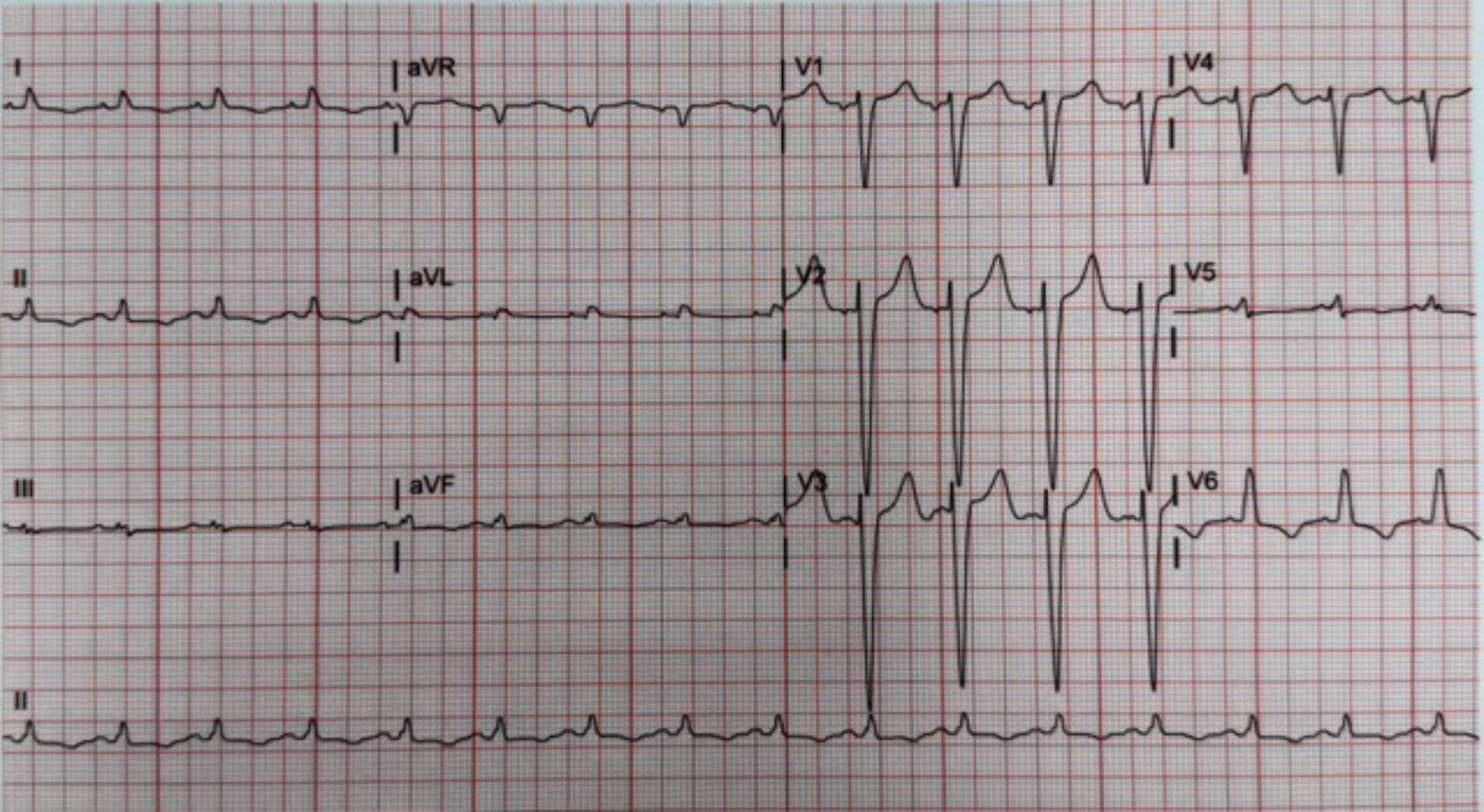
I
aVR
V1
V4
II
aVL
V2
V5
III
aVF
V3
V6
II

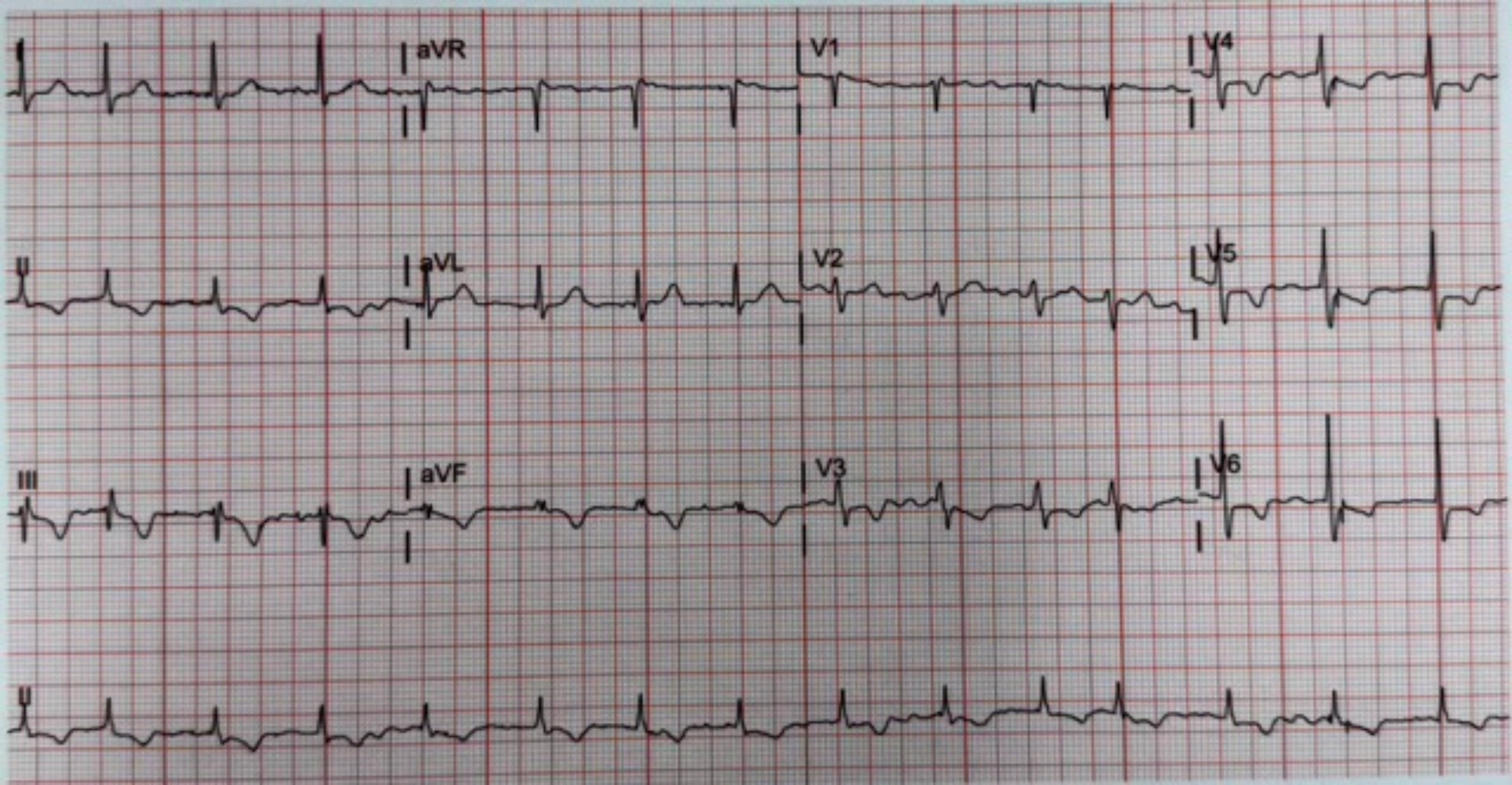

I
aVR
V1
V4
II
aVL
V2
V5
III
aVF
V3
V6
II

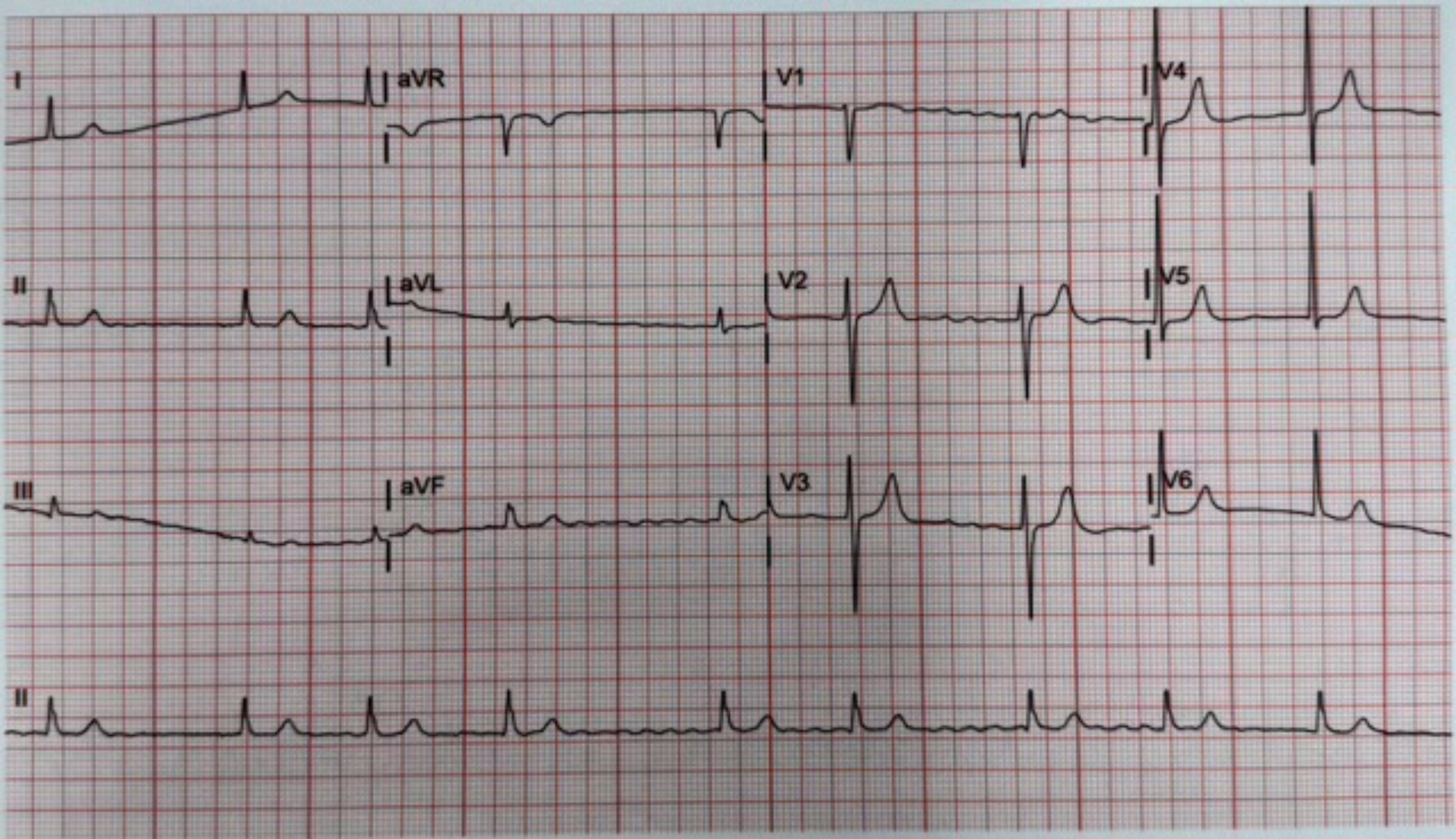
I
aVR
V1
V4
II
aVL
V2
V5
III
aVF
V3
V6
II

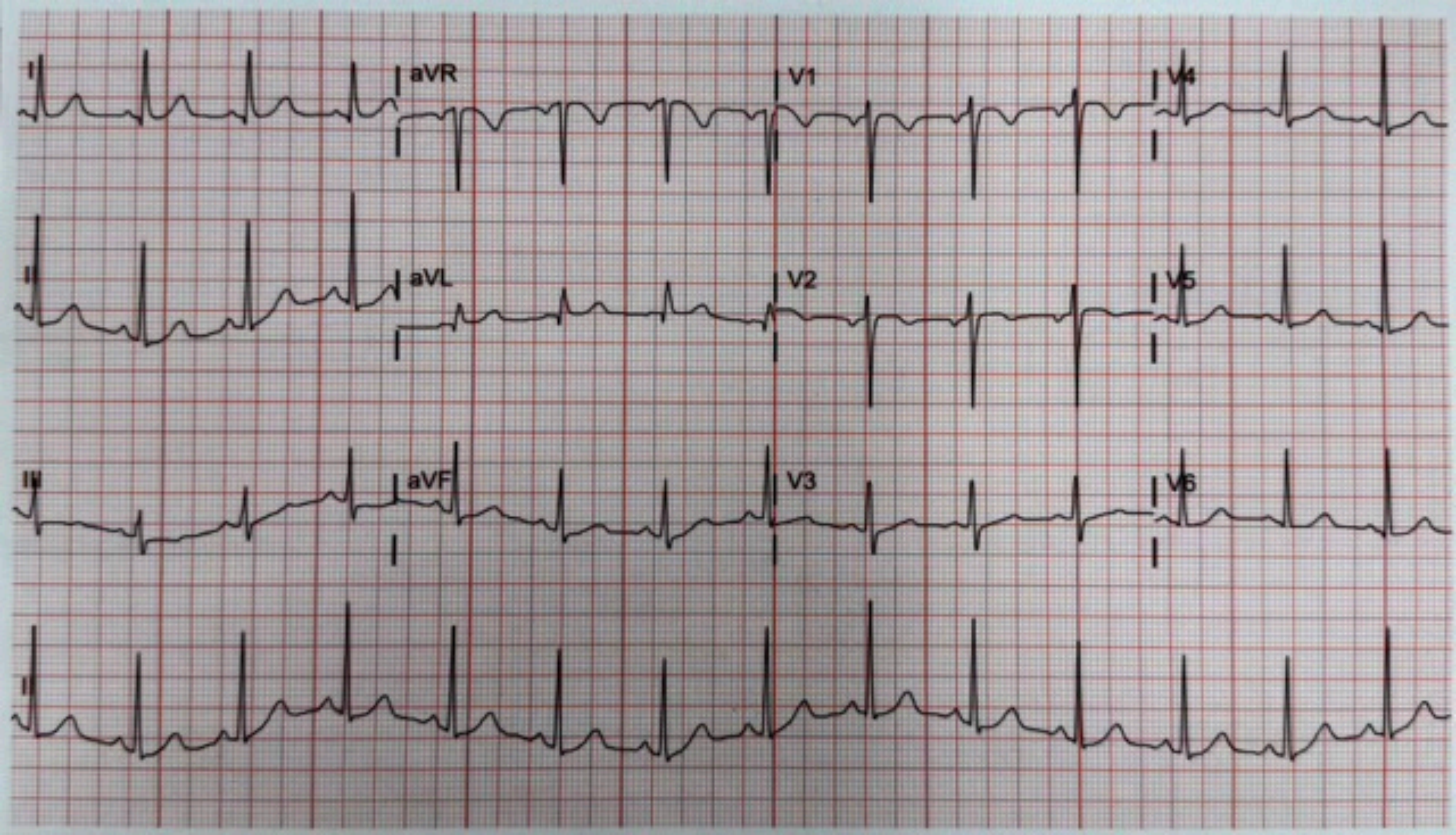
I
aVR
V1
V4
II
aVL
V2
V5
III
aVF
V3
V6
II

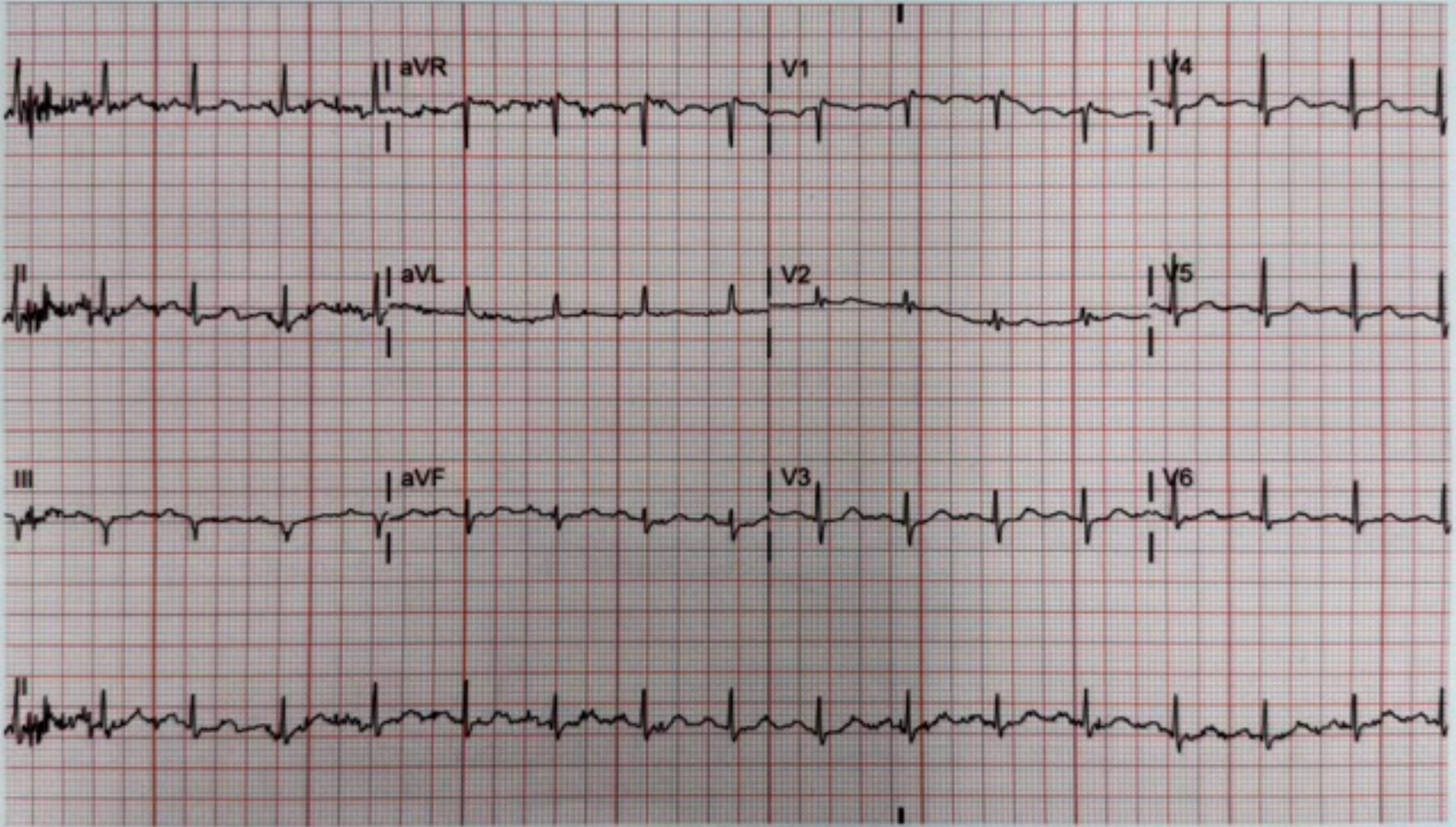

aVR
V1
V4
II
aVL
V2
V5
III
aVF
V3
V6
II

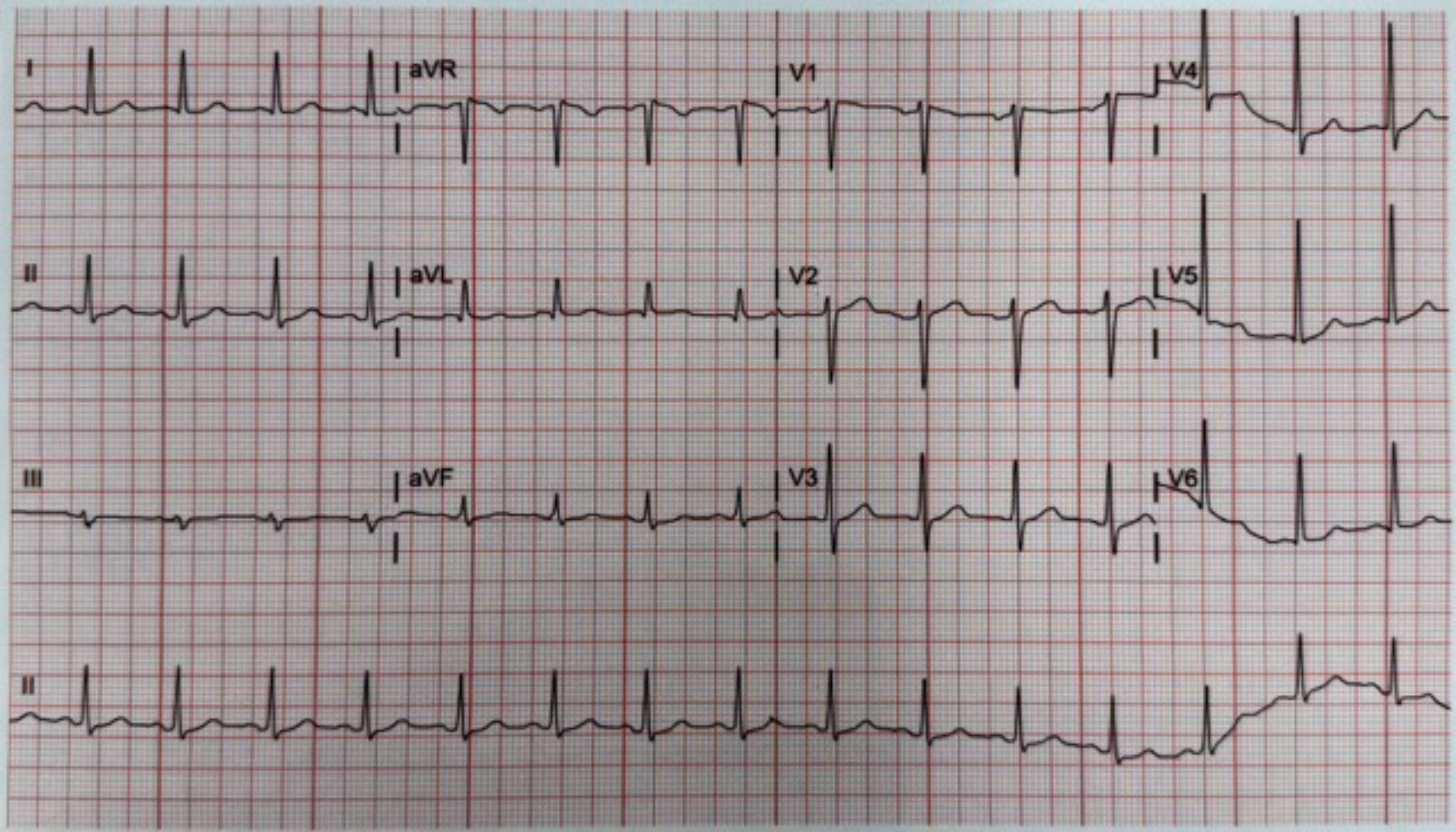
I
aVR
V1
V4
II
aVL
V2
V5
III
aVF
V3
V6
II

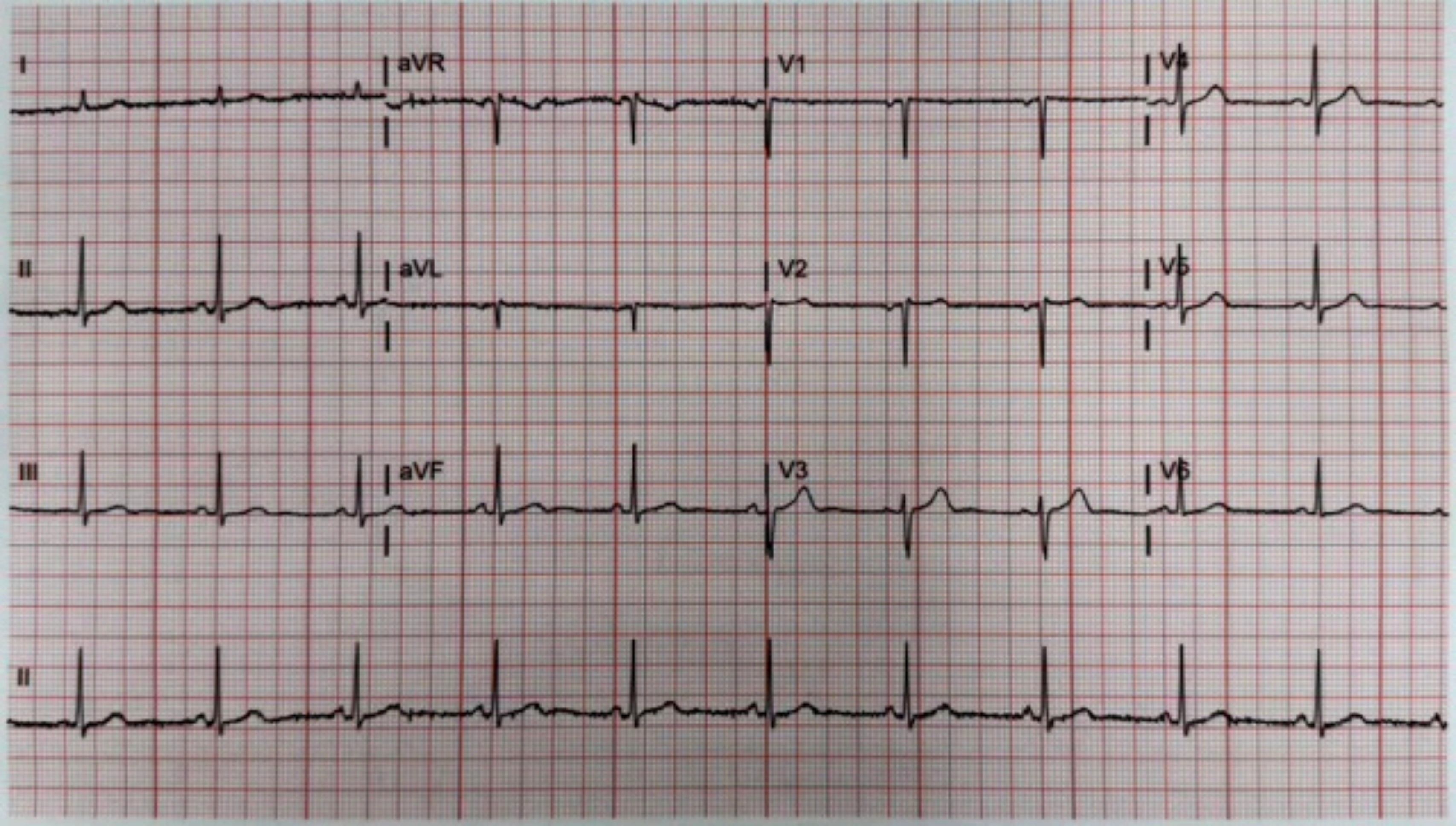
I
aVR
V1
II
aVL
V2
III
aVF
V3
II

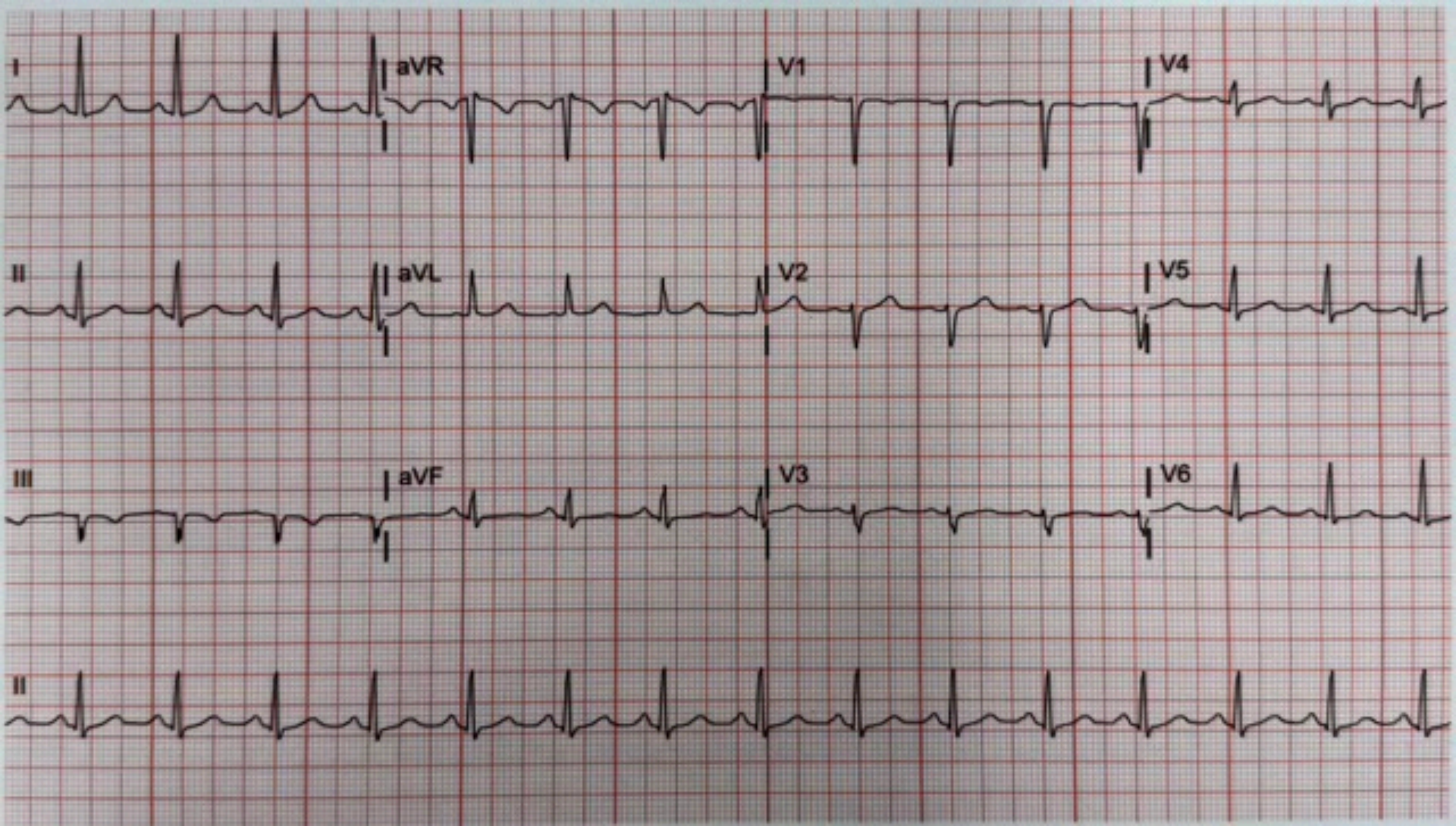
I
aVR
V1
V4
II
aVL
V2
V5
III
aVF
V3
V6
II

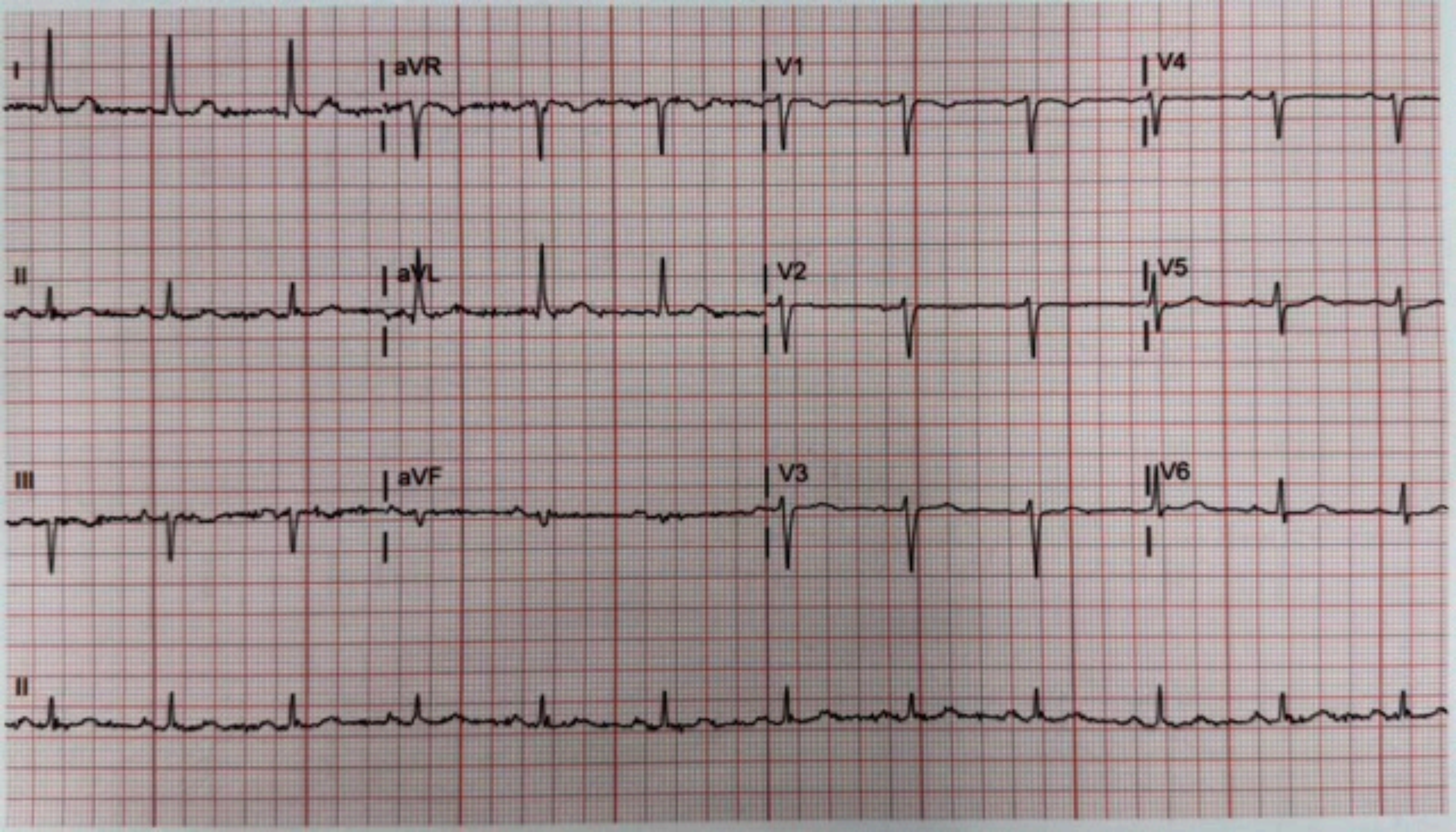
I
aVR
V1
V4
II
aVL
V2
V5
III
aVF
V3
V6
II

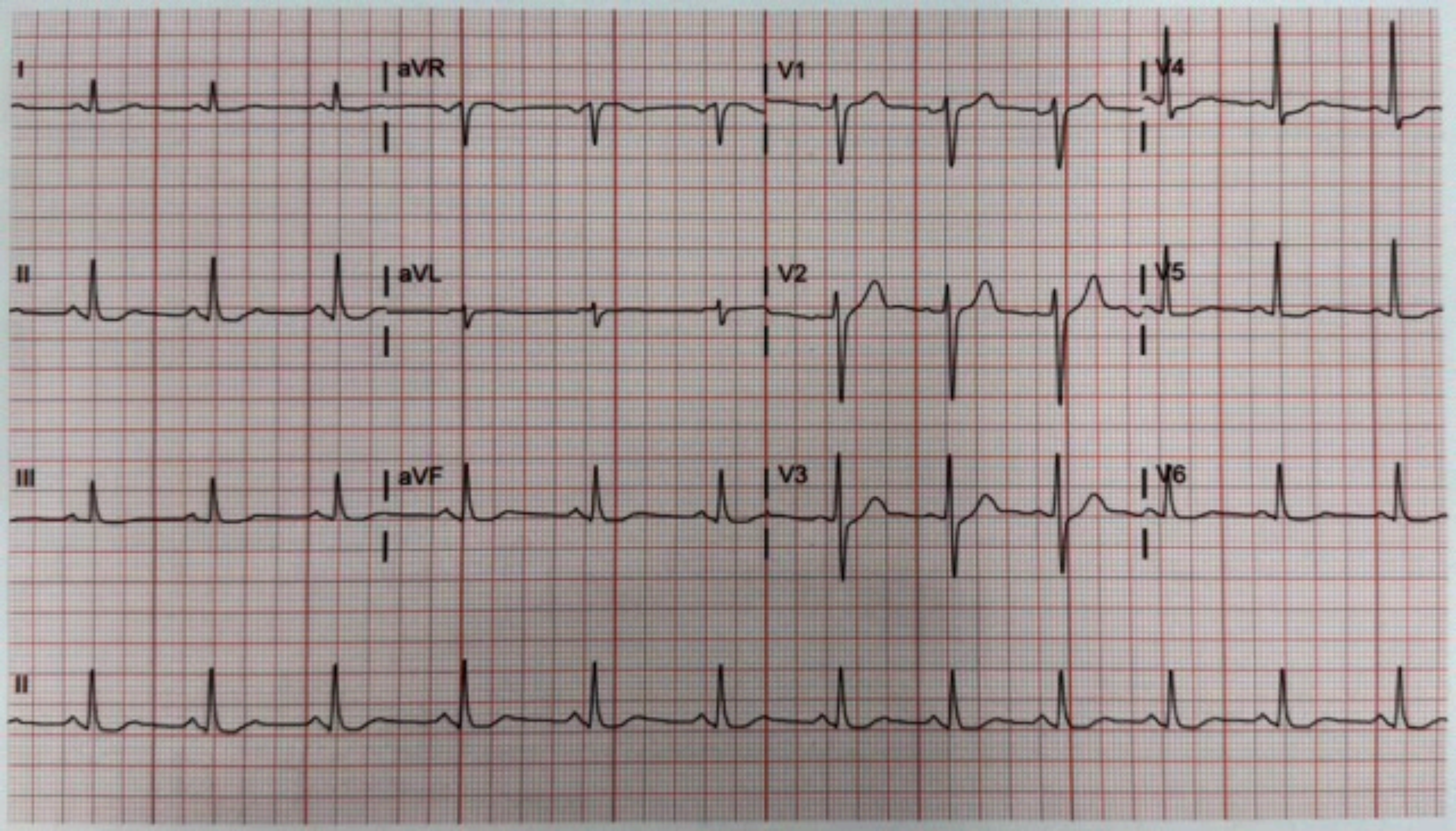
I
aVR
V1
V4
II
aVL
V2
V5
III
aVF
V3
V6
II

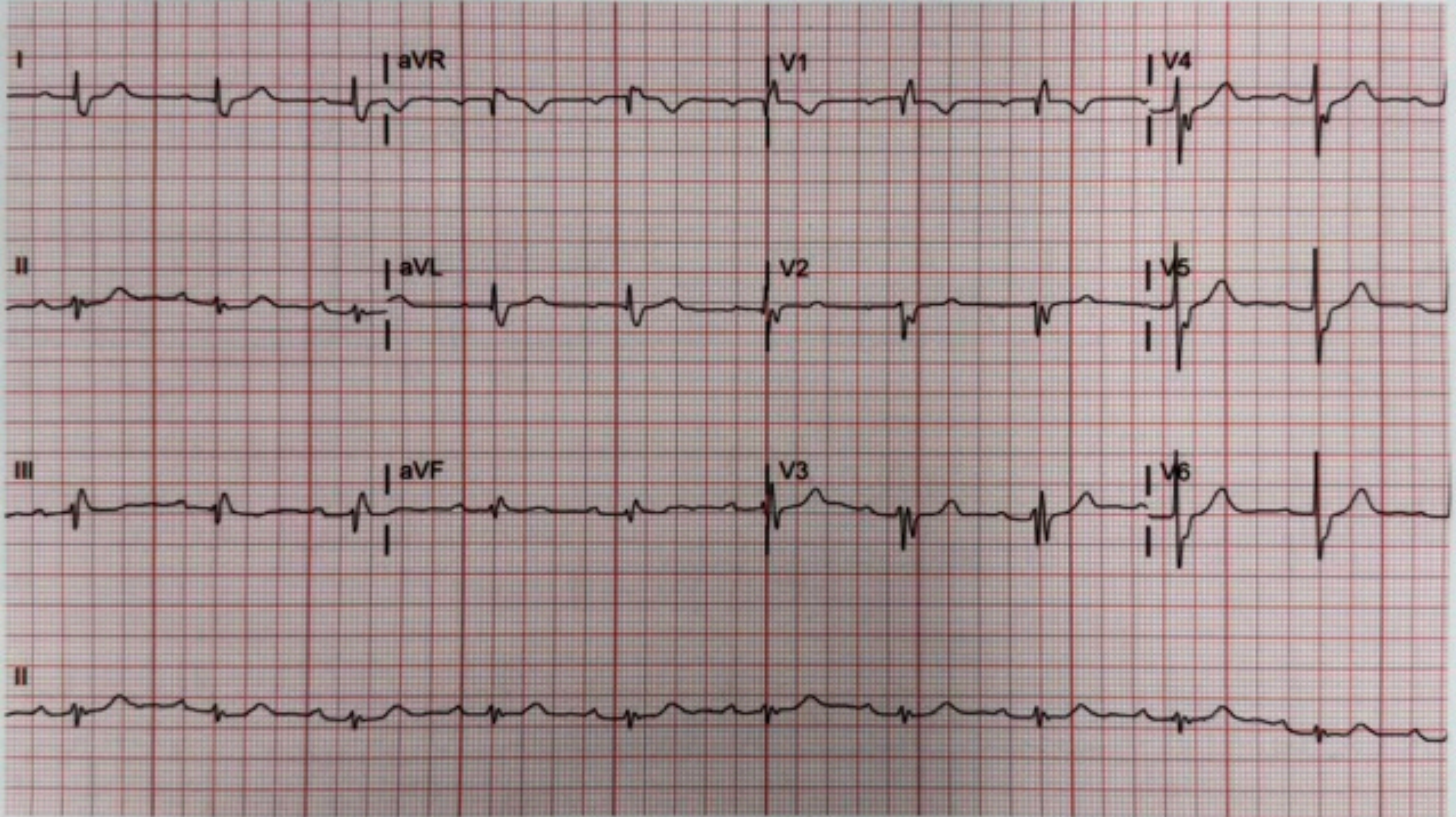
I
aVR
V1
V4
II
aVL
V2
V5
III
aVF
V3
V6
II

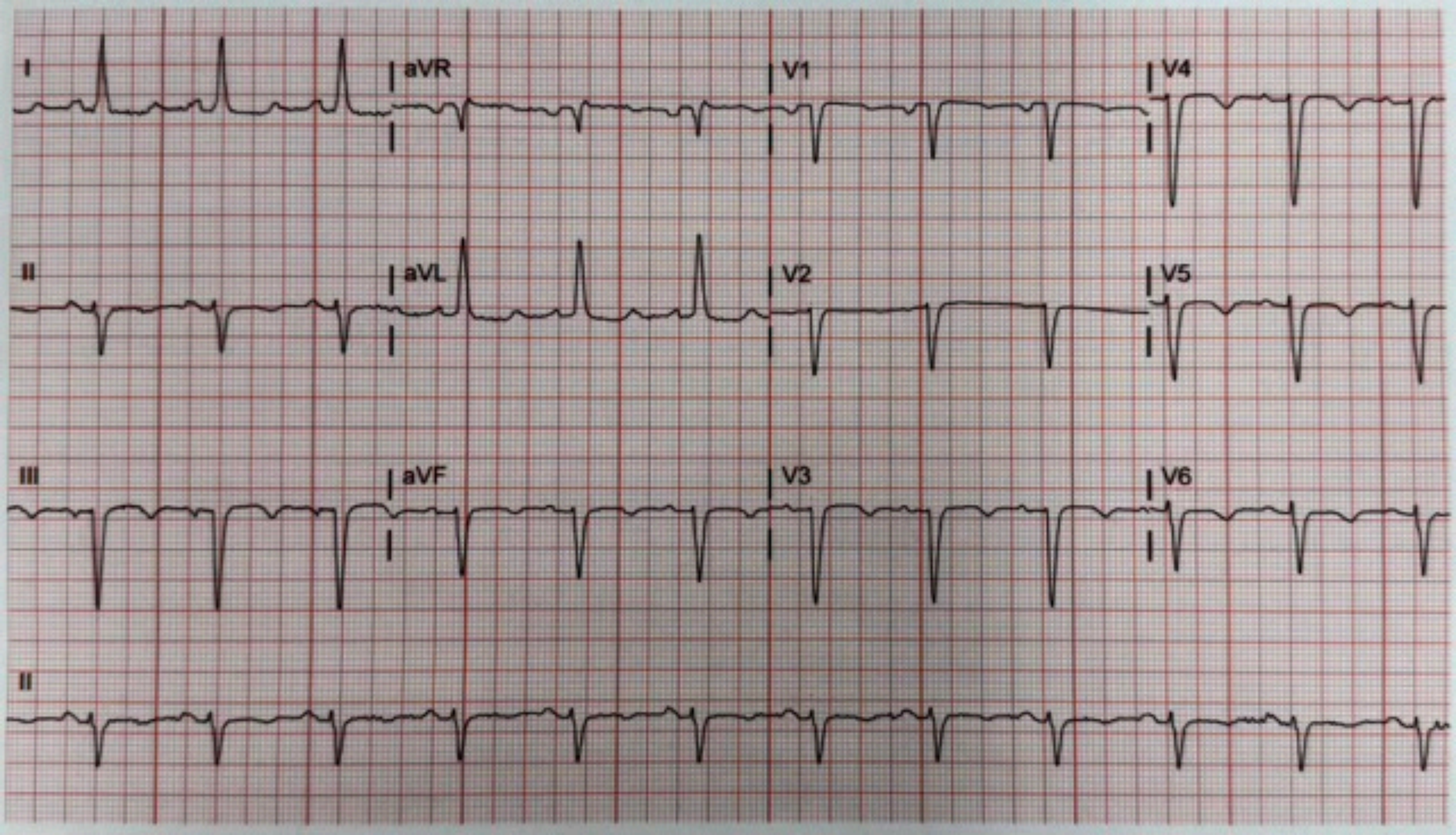
I
aVR
V1
V4
II
aVL
V2
V5
III
aVF
V3
V6
II

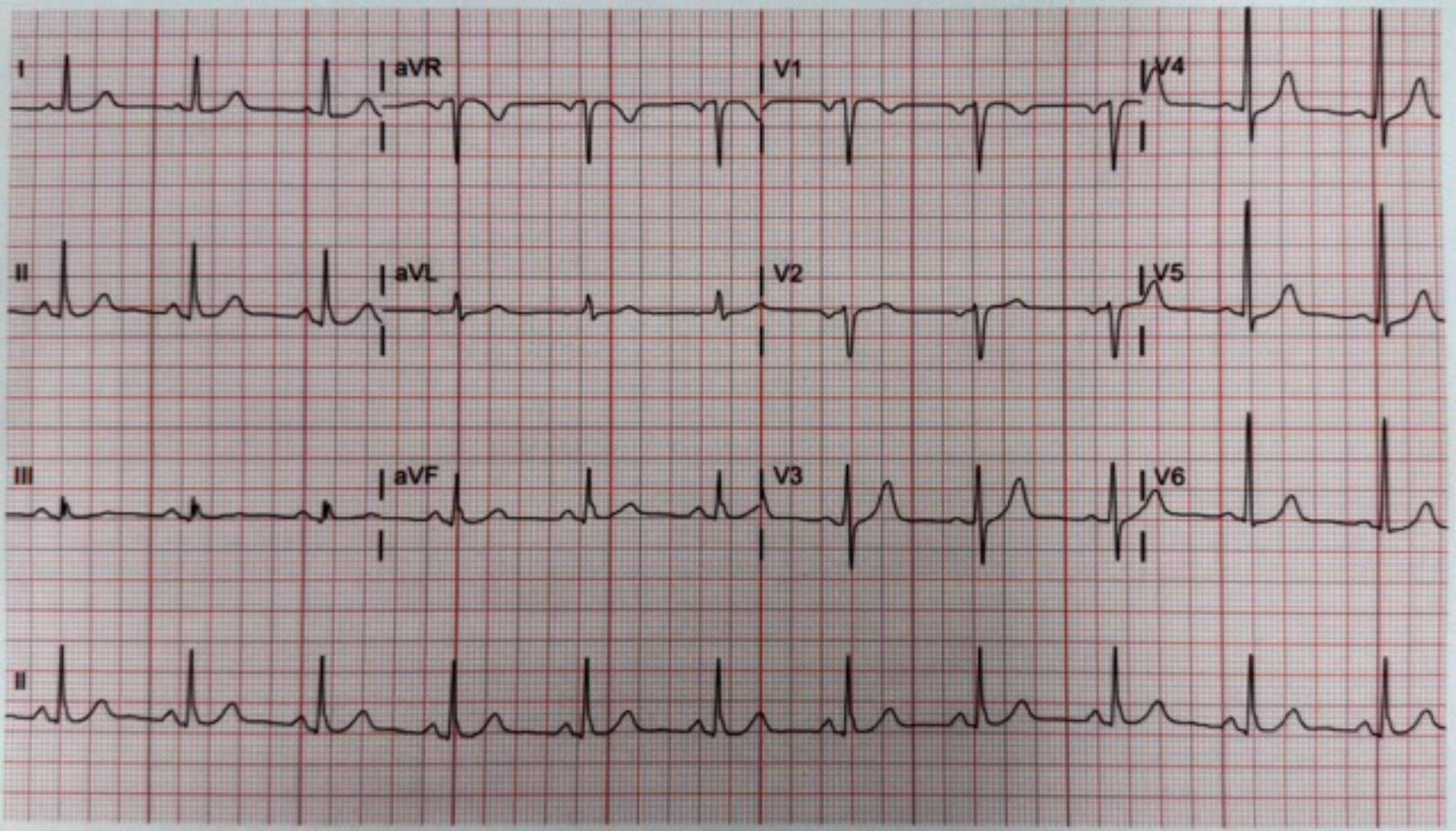
I
aVR
V1
V4
II
aVL
V2
V5
III
aVF
V3
V6
II

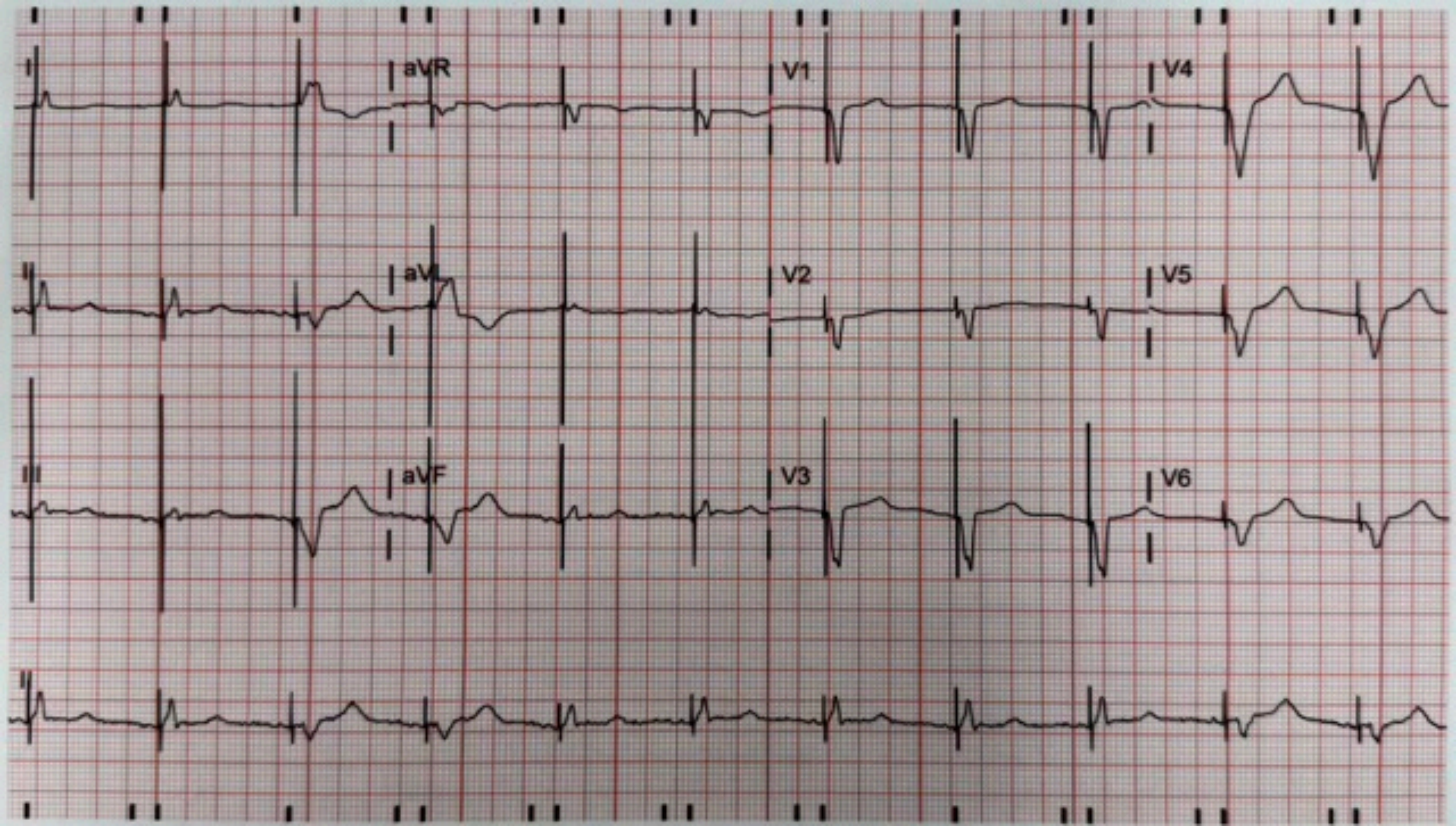
I
aVR
V1
V4
II
aVL
V2
V5
III
aVF
V3
V6
II

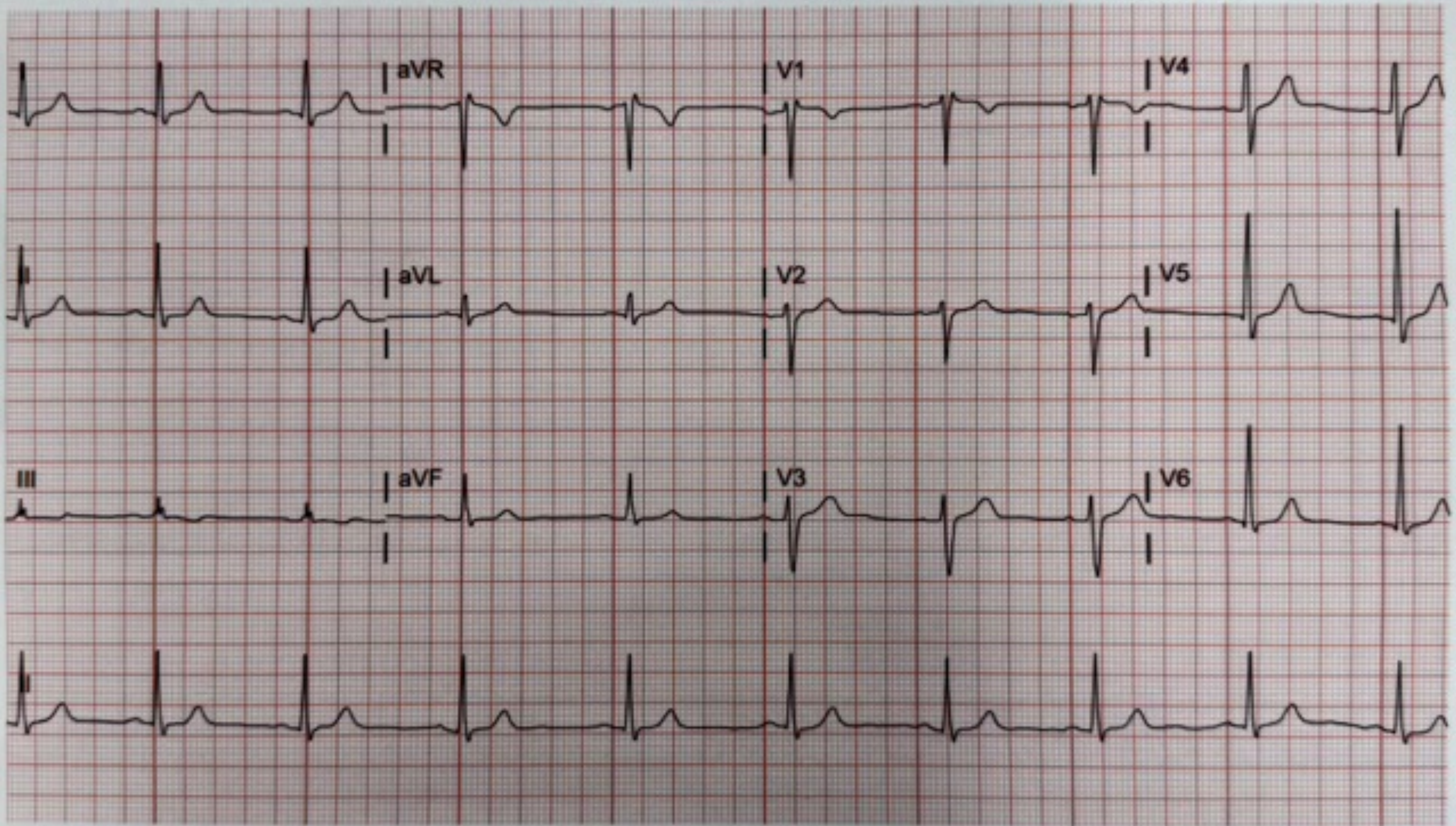

aVR
V1
V4
II
aVL
V2
V5
III
aVF
V3
V6
II

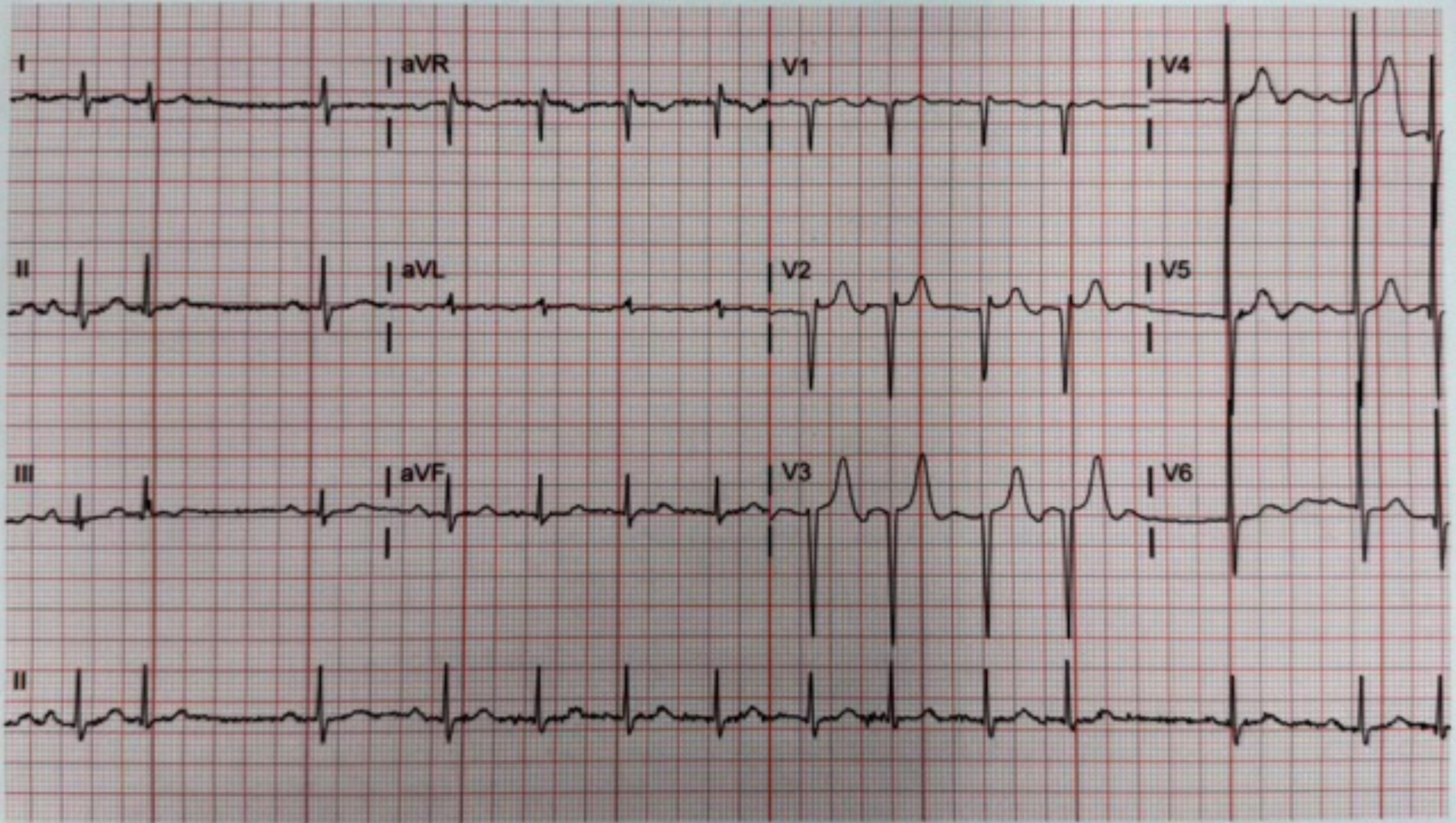
I
aVR
V1
V4
II
aVL
V2
V5
III
aVF
V3
V6
II

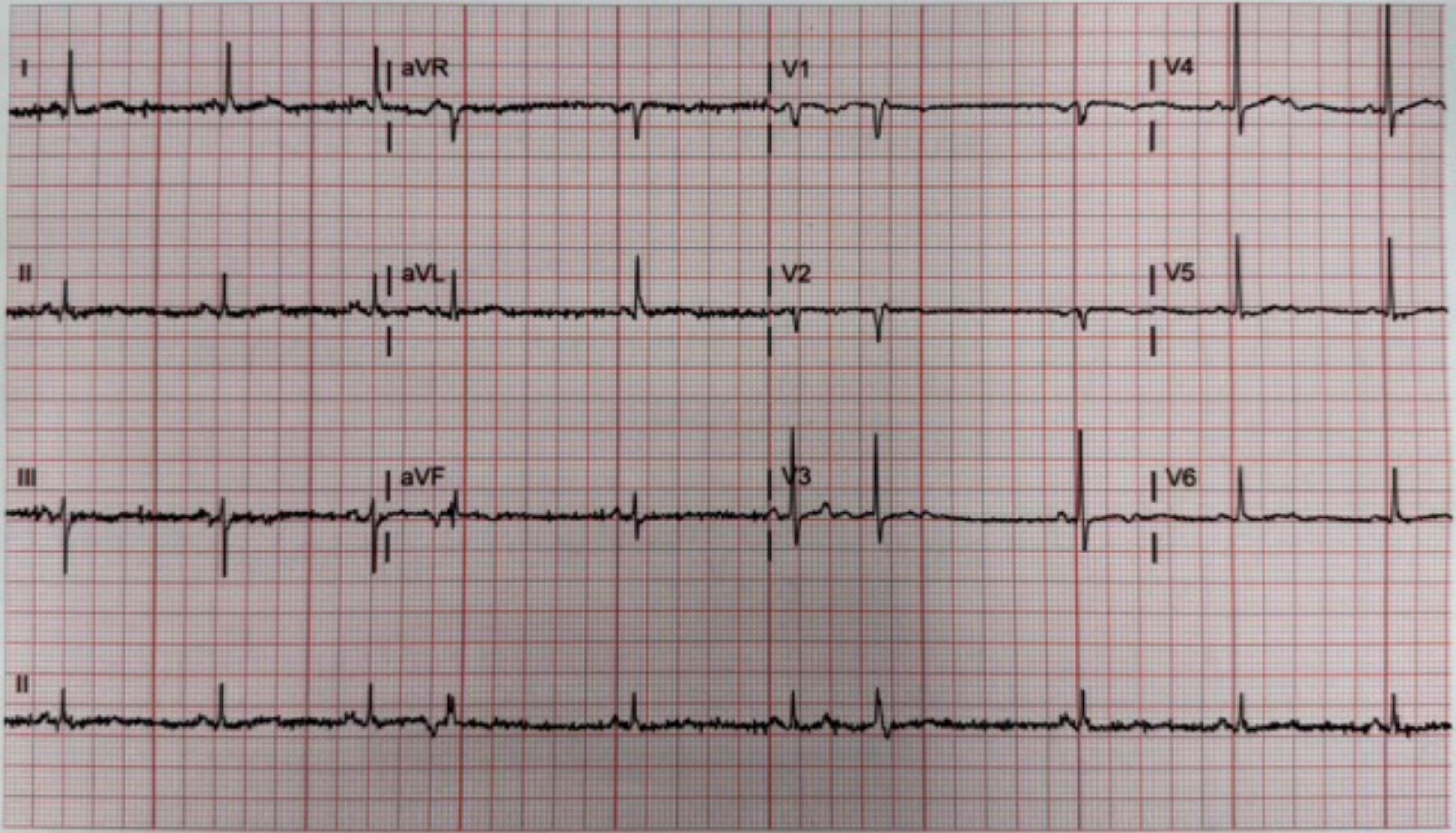
I
aVR
V1
V4
II
aVL
V2
V5
III
aVF
V3
V6
II

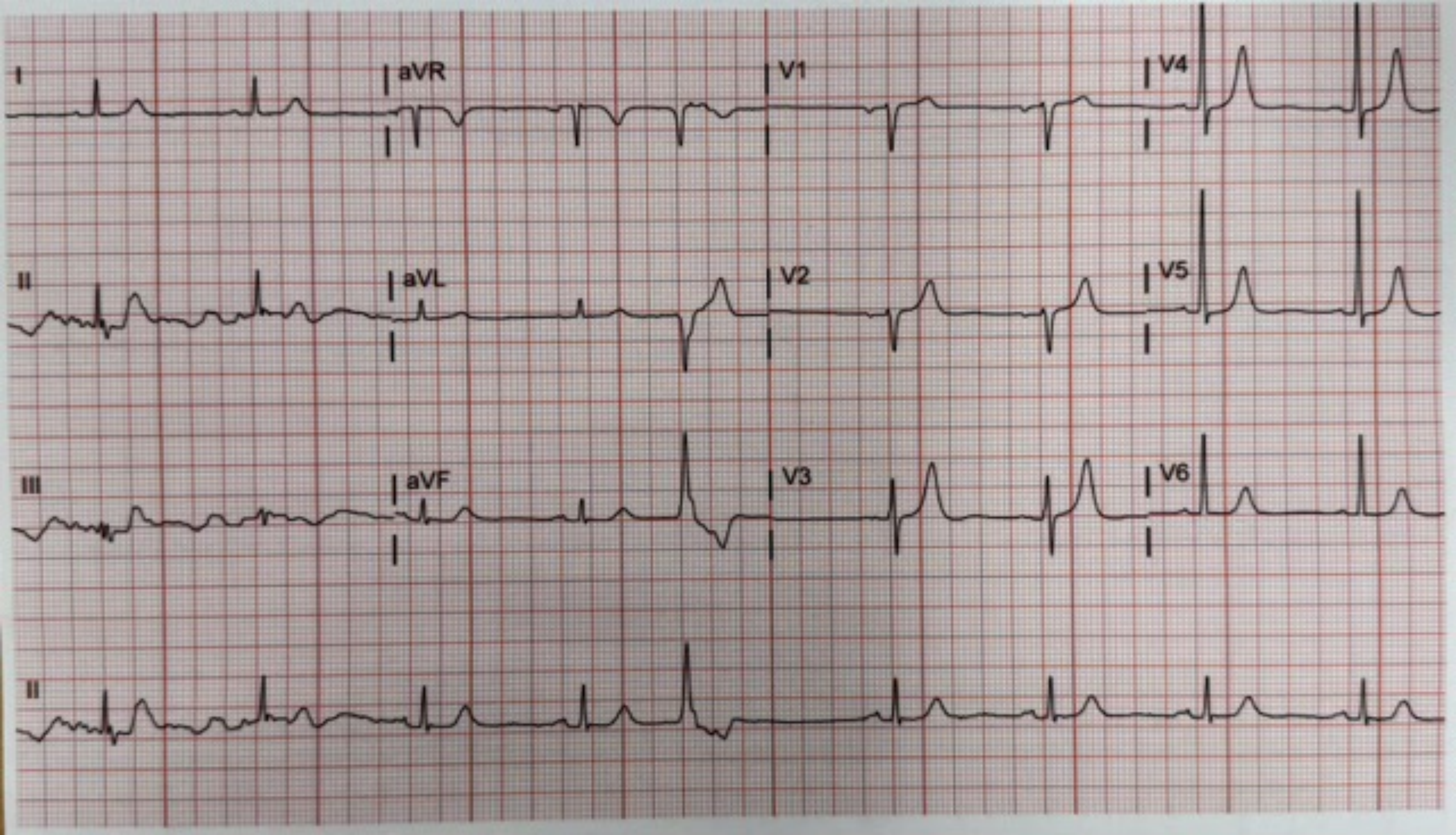

I
aVR
V1
V4
II
aVL
V2
V5
III
aVF
V3
V6
II

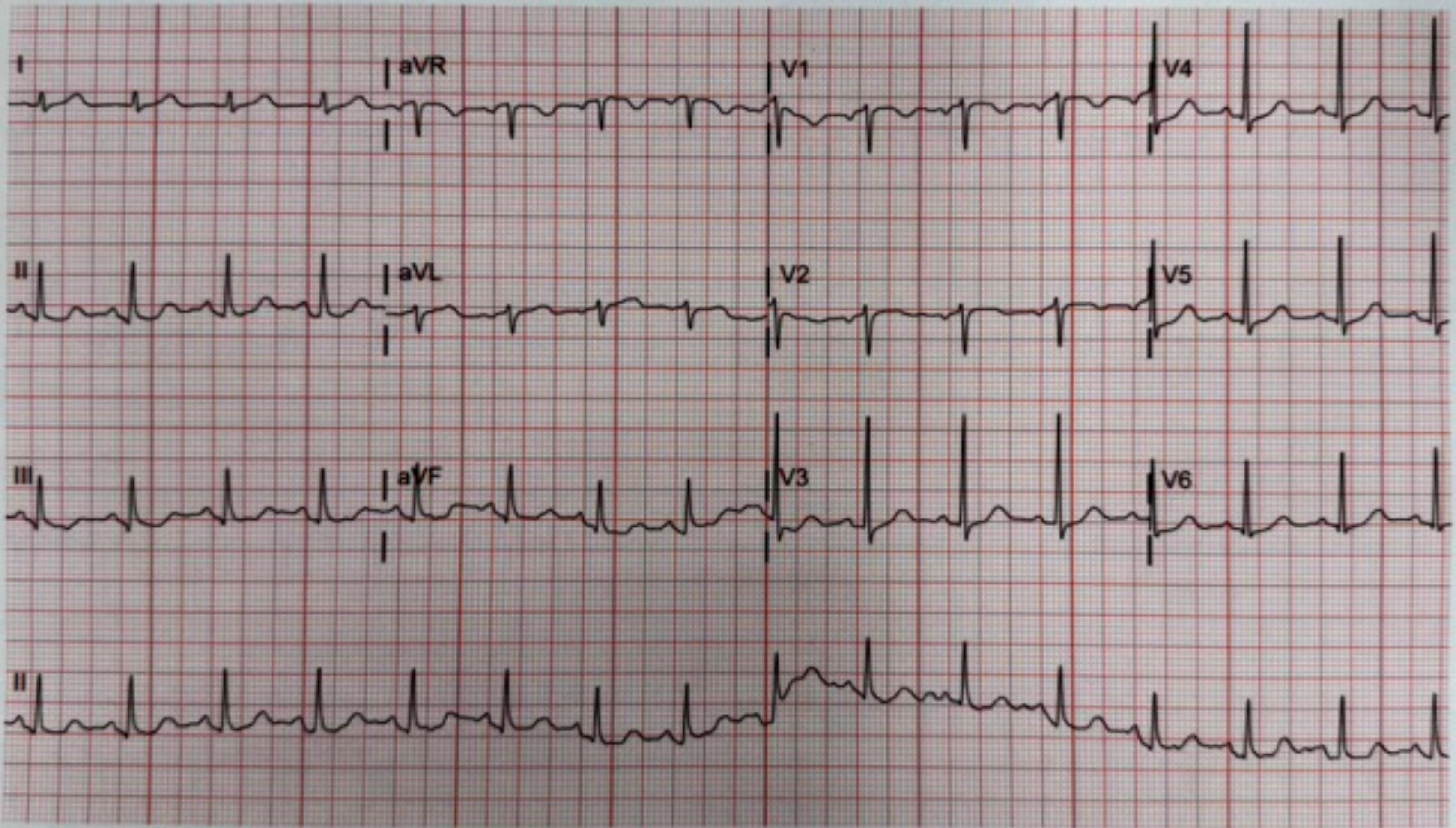
I
aVR
V1
V4
II
aVL
V2
V5
III
aVF
V3
V6
II

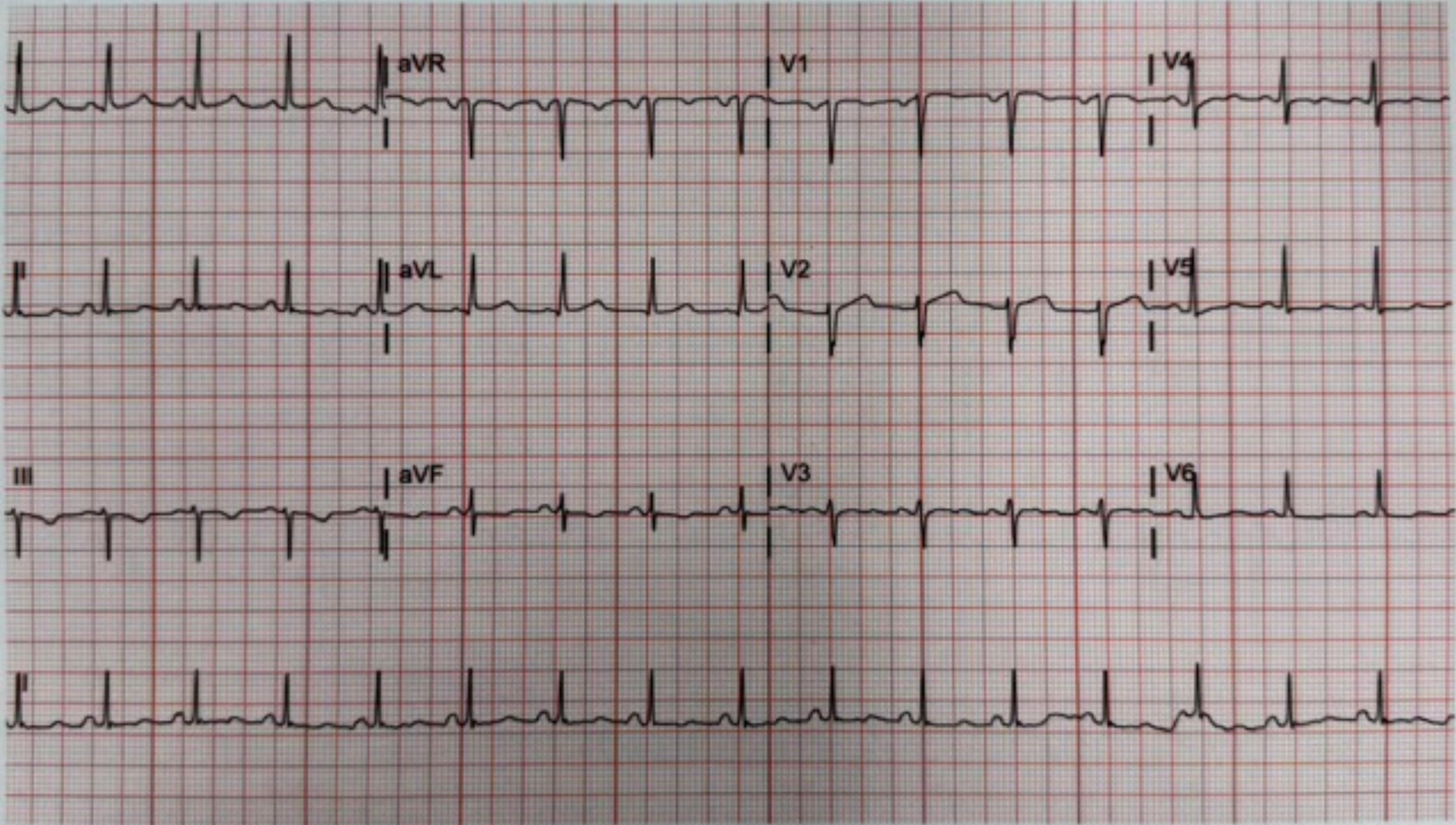
I
aVR
V1
V4
II
aVL
V2
V5
III
aVF
V3
V6
II

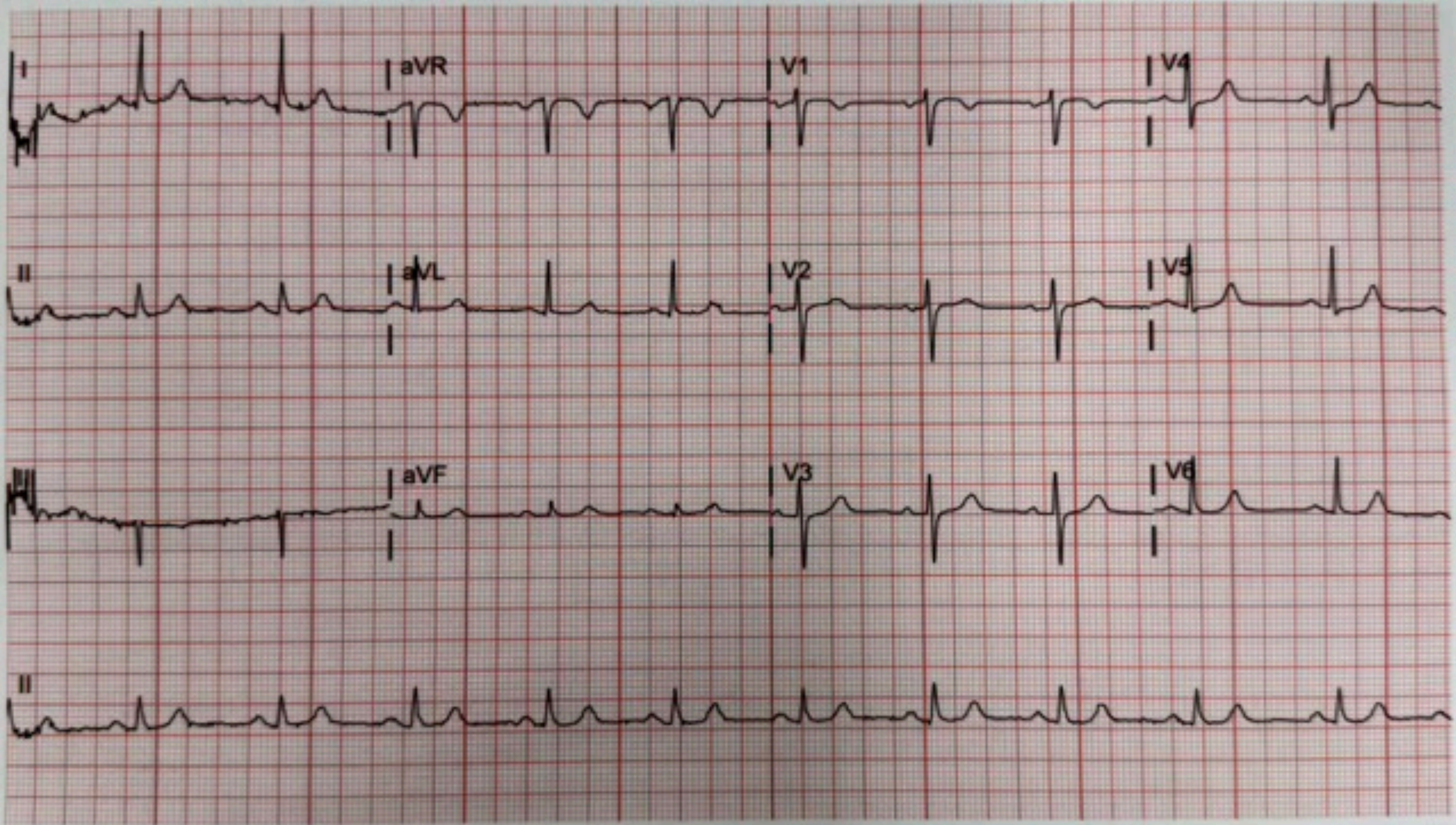
I
aVR
V1
V4
II
aVL
V2
V5
III
aVF
V3
V6
II

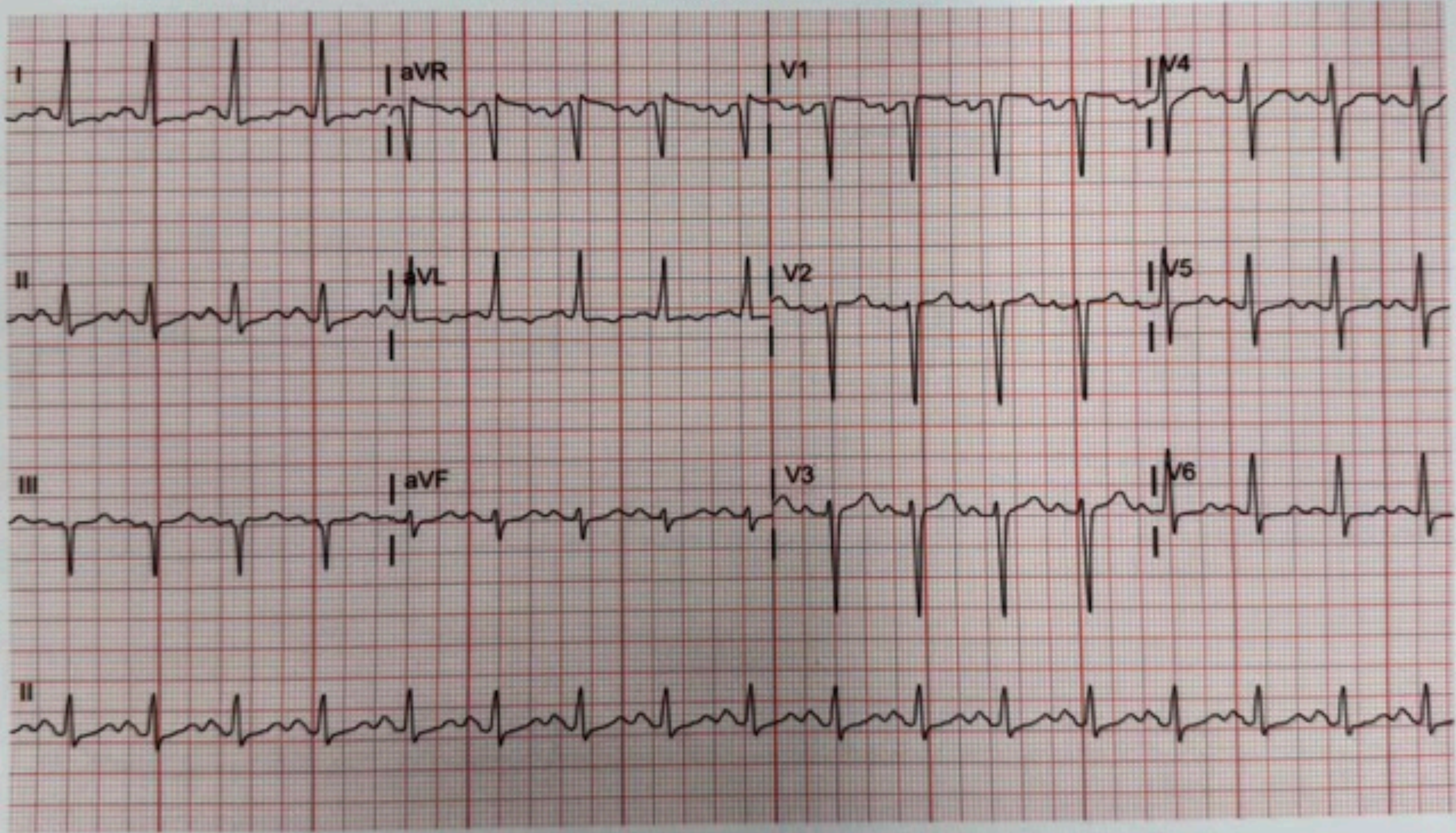
I
aVR
V1
V4
II
aVL
V2
V5
III
aVF
V3
V6
II

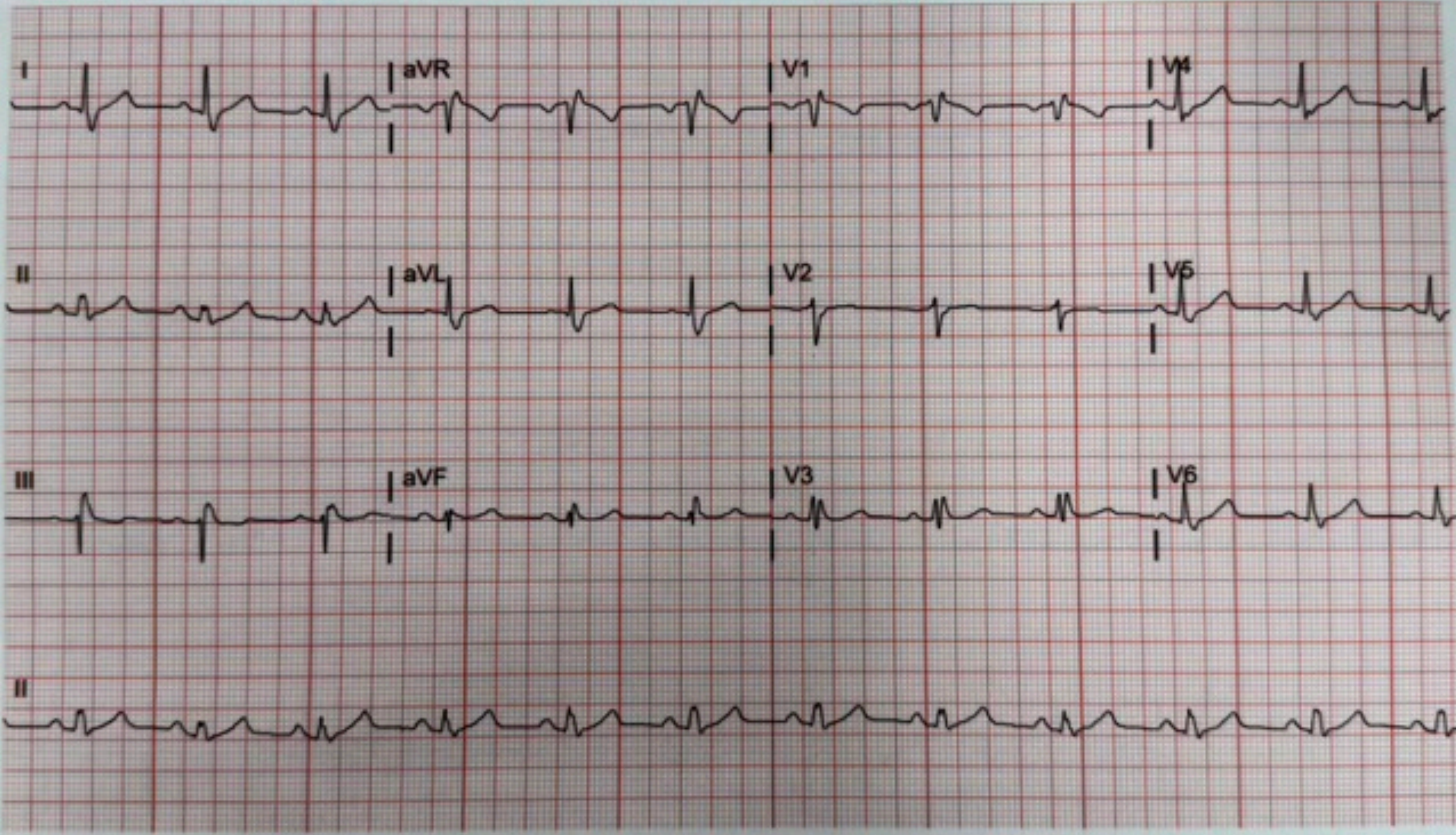
I
aVR
V1
V4
II
aVL
V2
V5
III
aVF
V3
V6
II

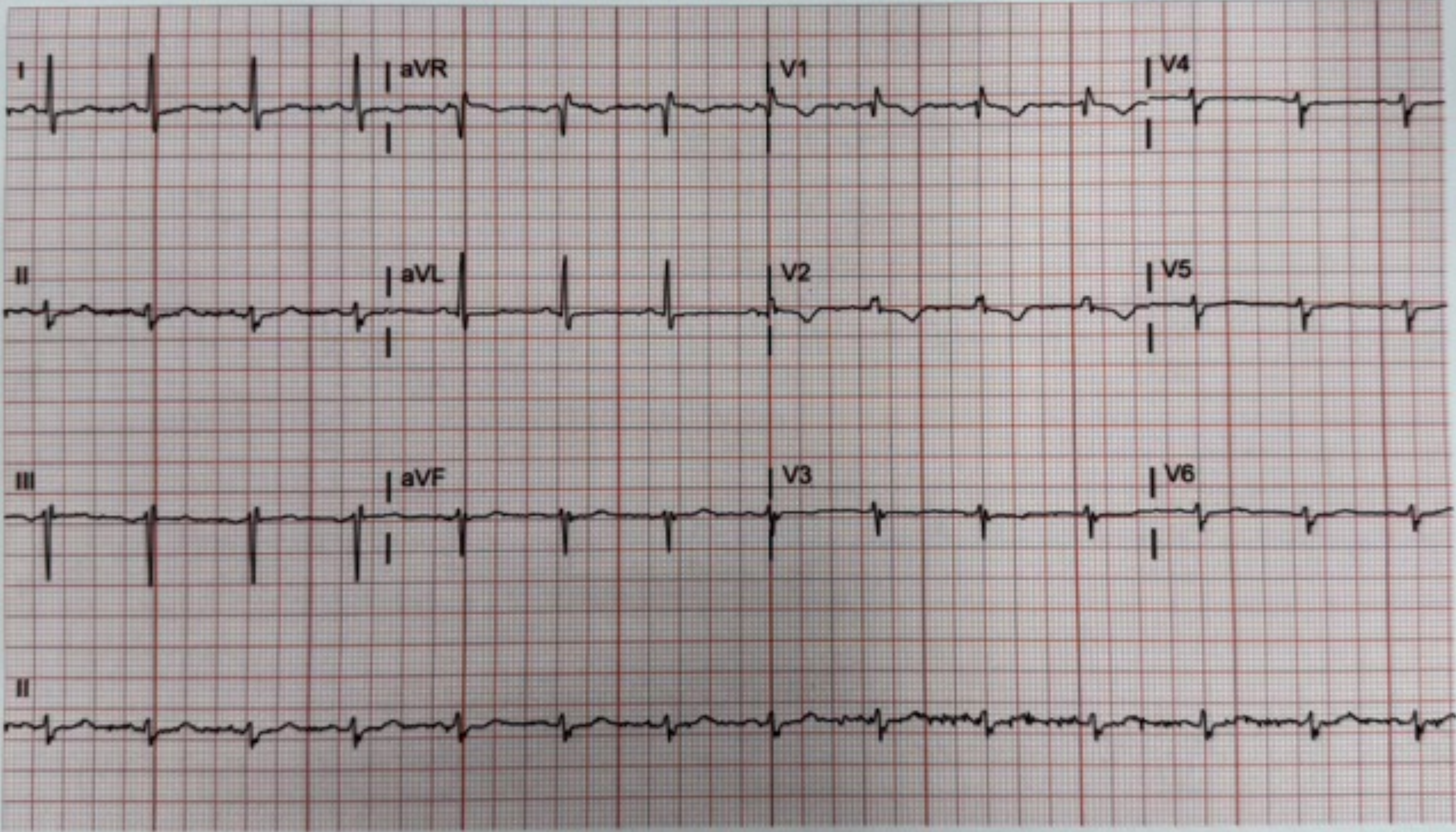
I
aVR
V1
V4
II
aVL
V2
V5
III
aVF
V3
V6
II

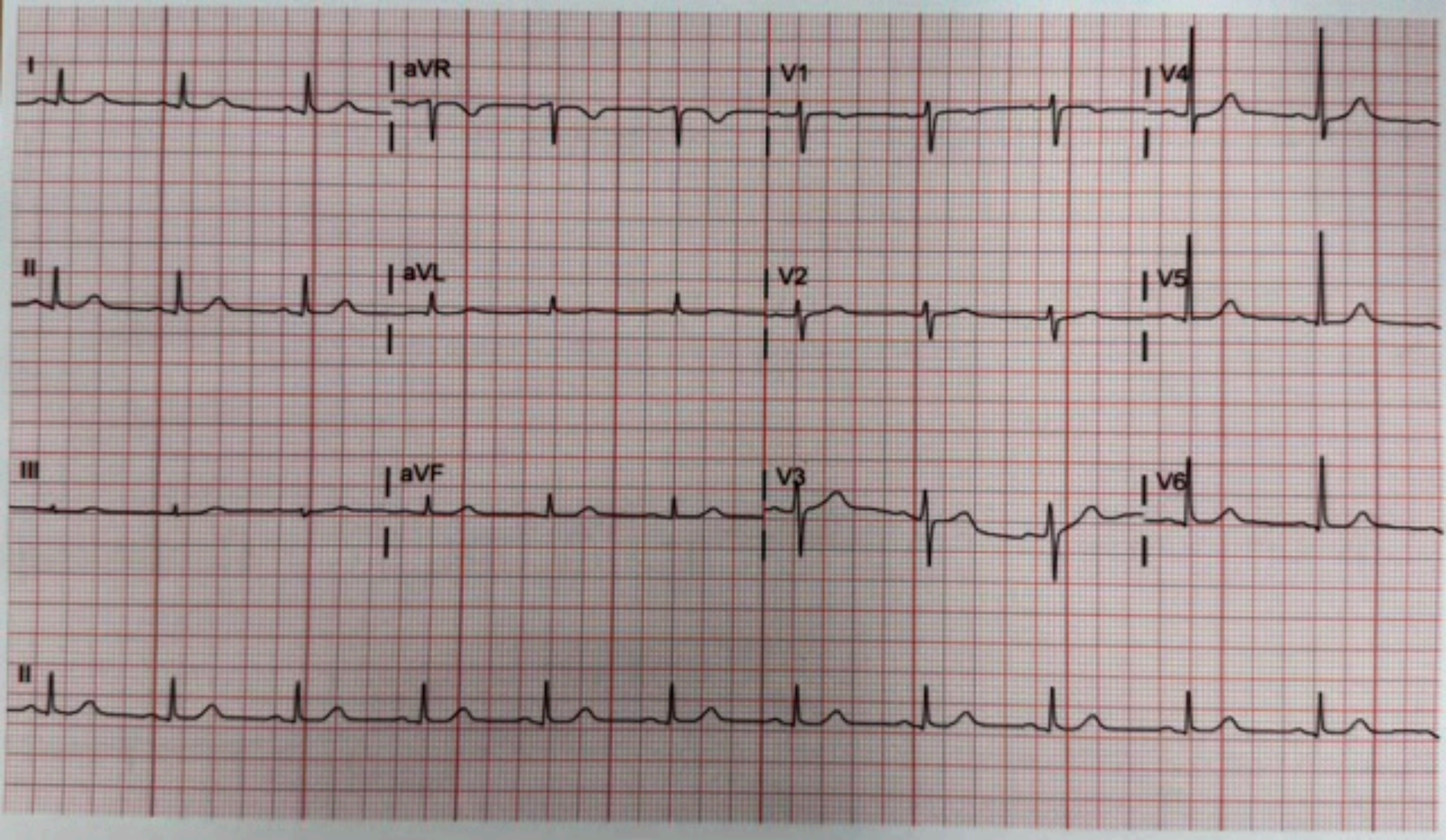
I
aVR
V1
V4
II
aVL
V2
V5
III
aVF
V3
V6
II

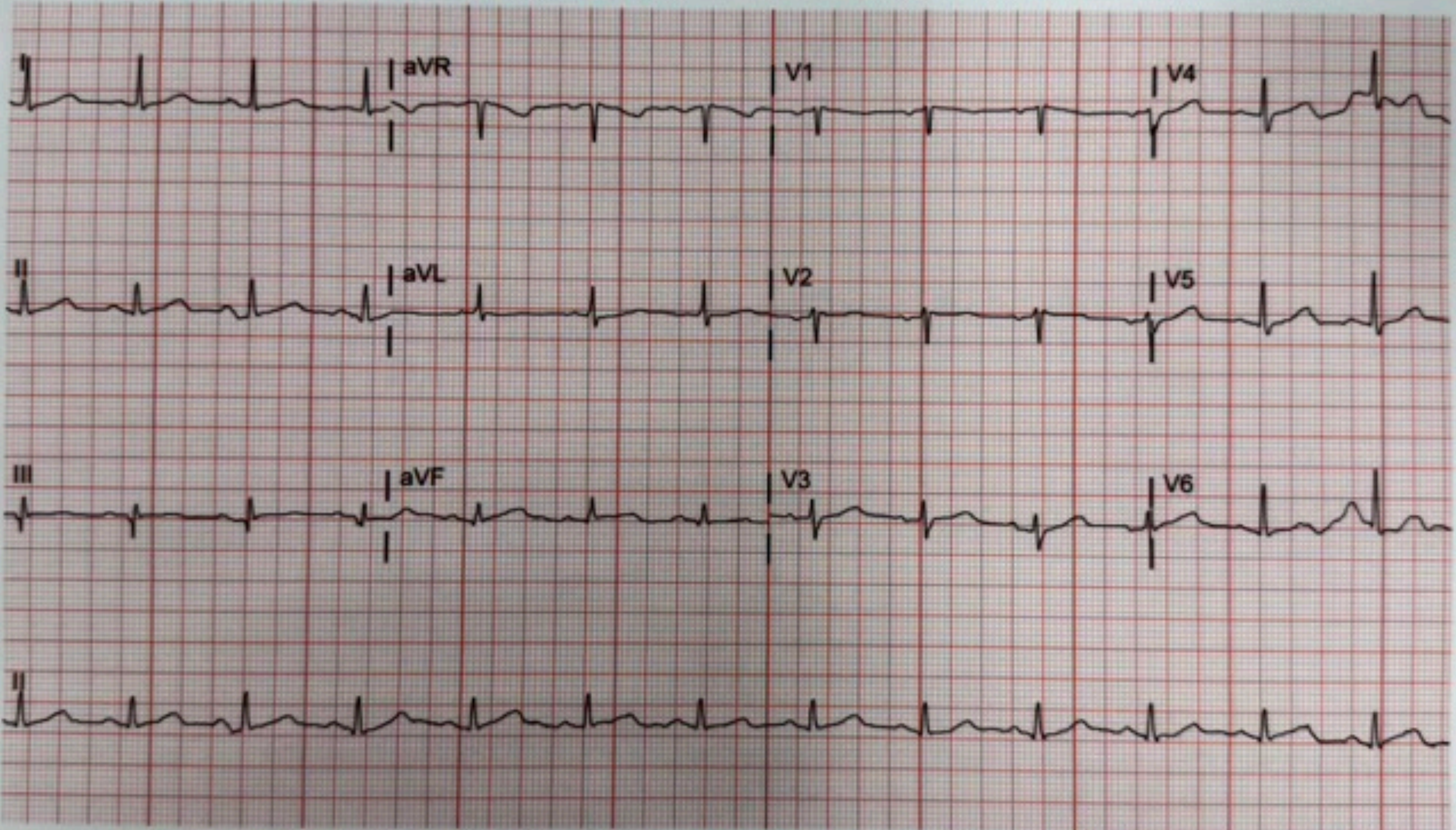
I
aVR
V1
V4
II
aVL
V2
V5
III
aVF
V3
V6
II

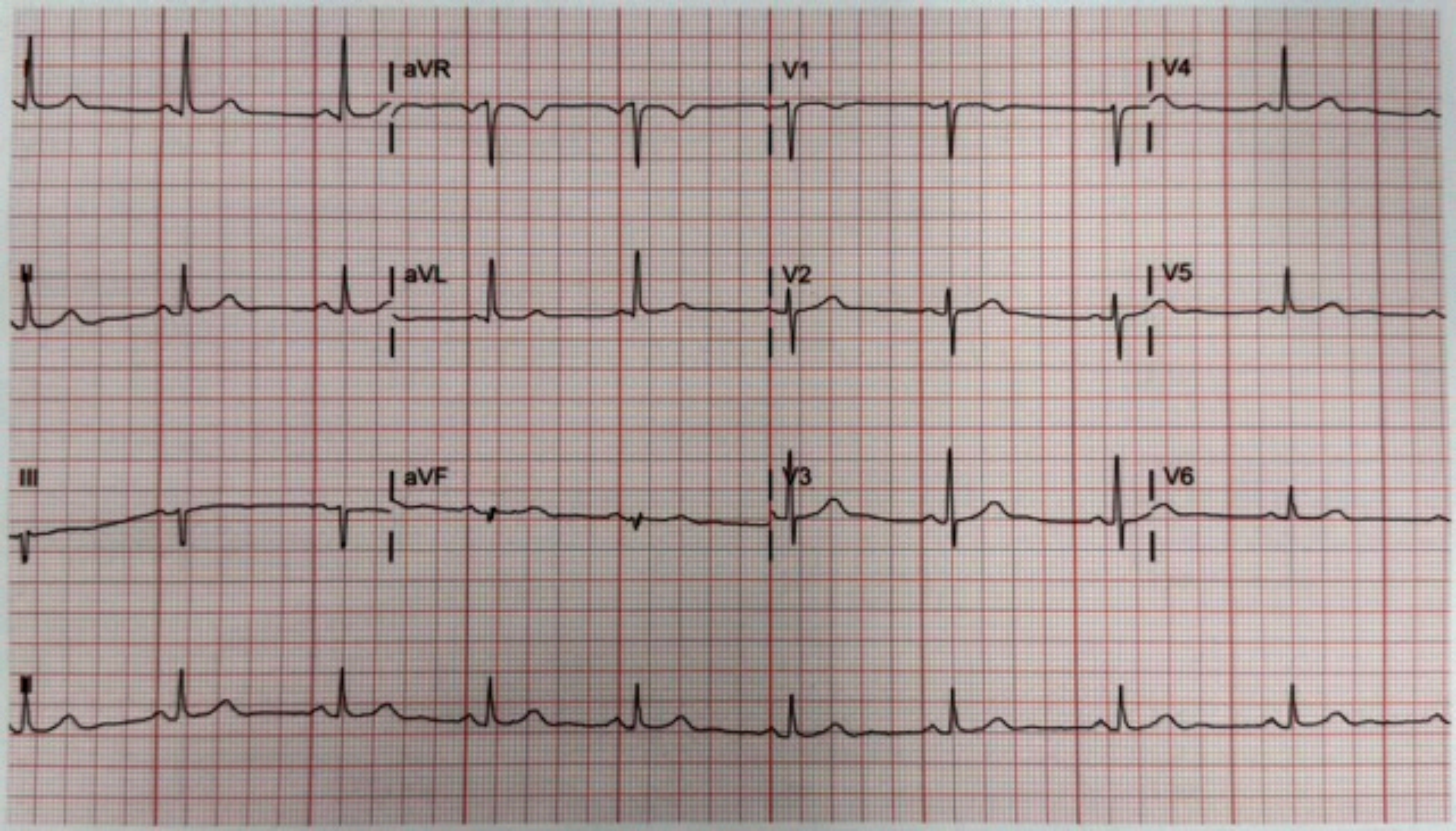
aVR
V1
V4
aVL
V2
V5
III
aVF
V3
V6

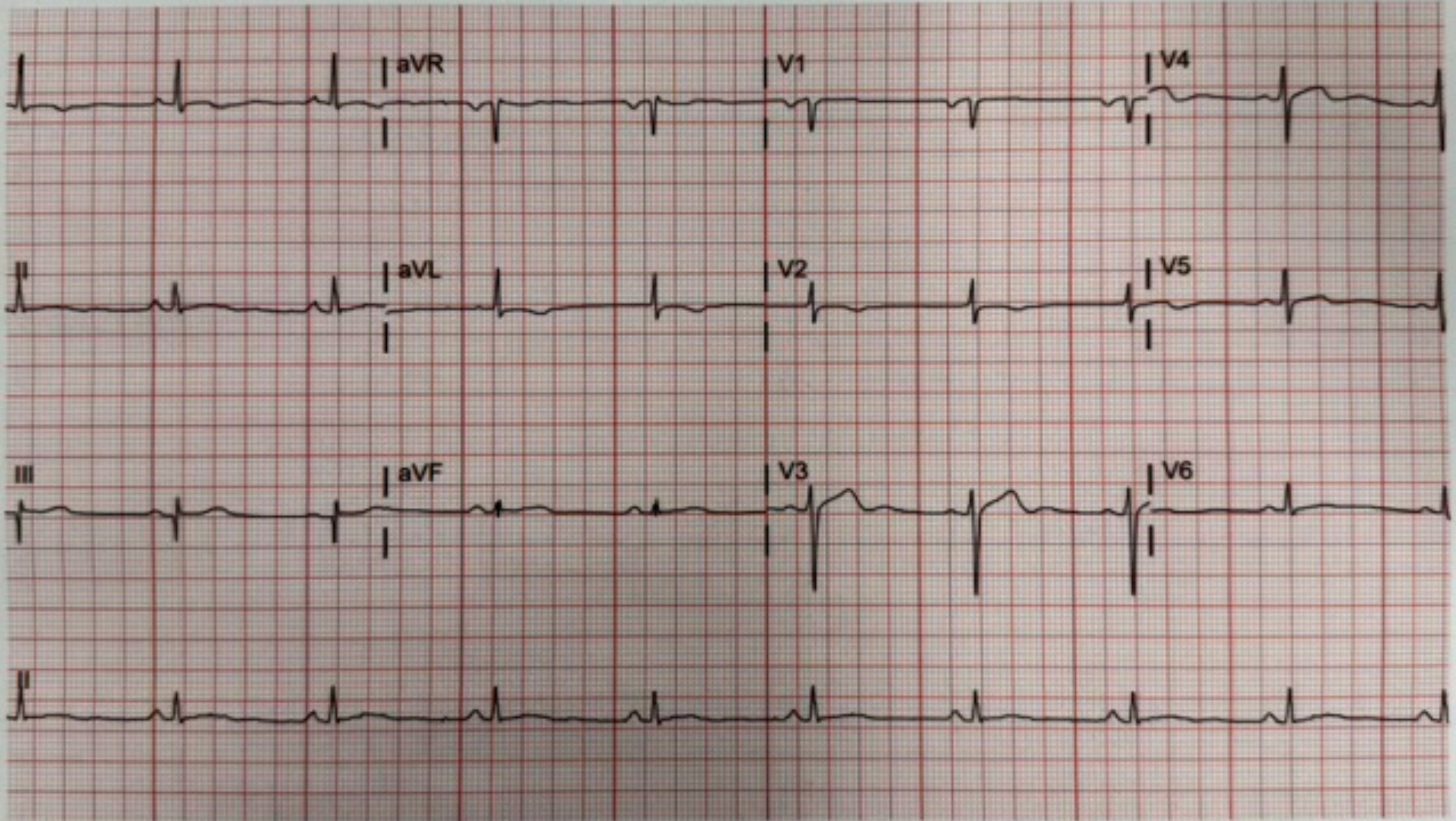
aVR
V1
V4
II
aVL
V2
V5
III
aVF
V3
V6
II

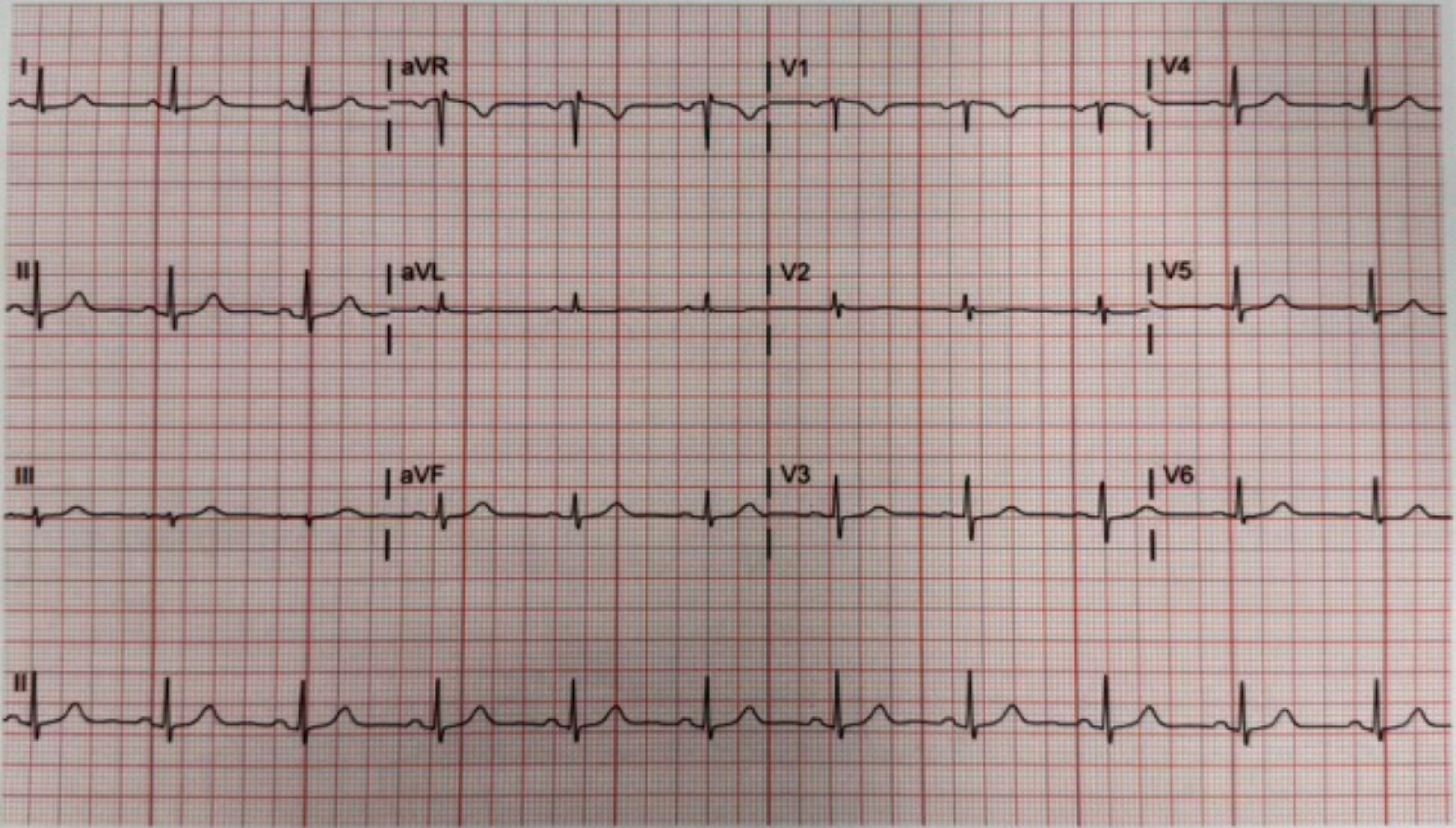
I
aVR
V1
V4
II
aVL
V2
V5
III
aVF
V3
V6
II

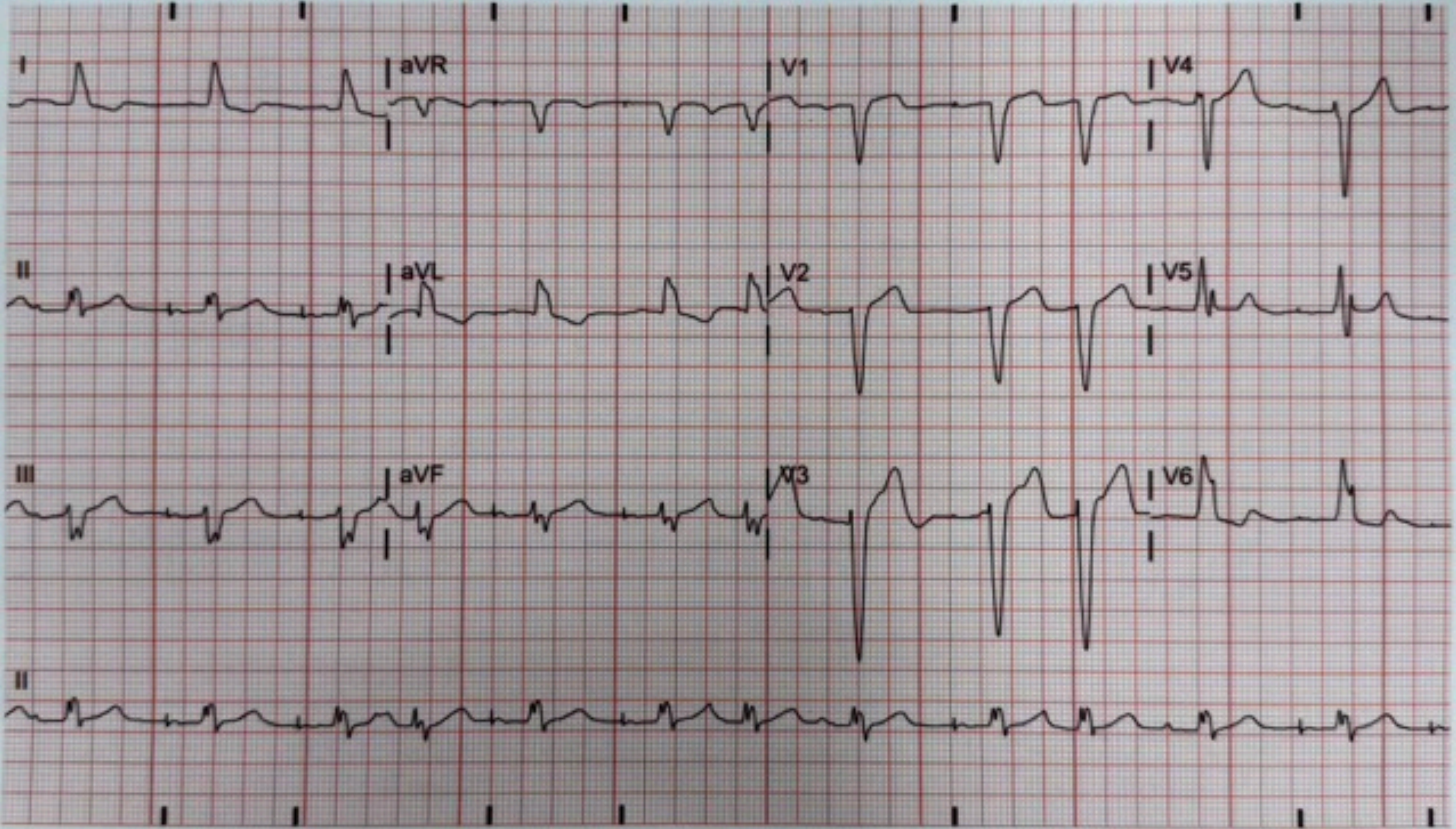
I
aVR
V1
V4
II
aVL
V2
V5
III
aVF
V3
V6
II

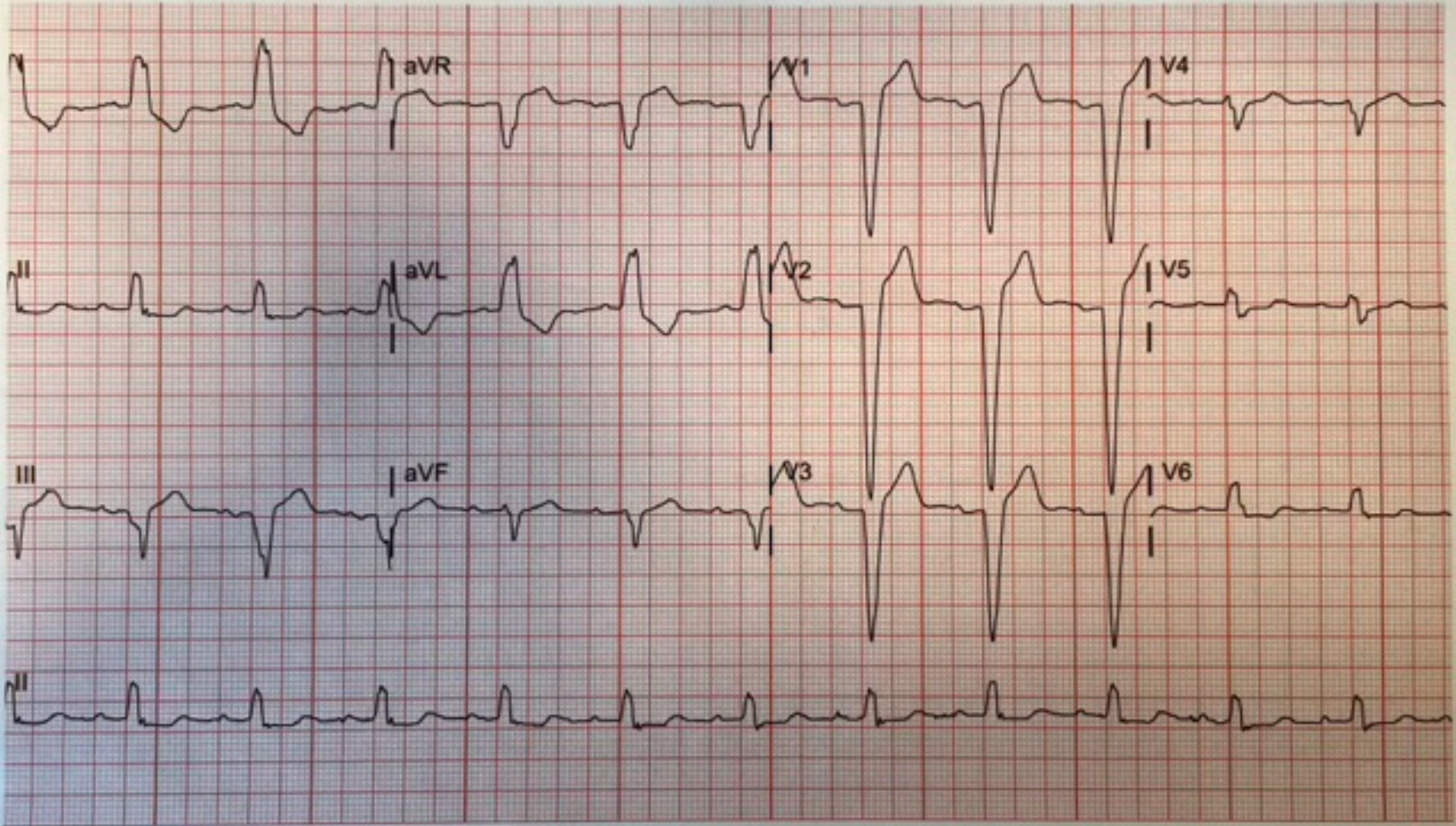
I
aVR
V1
V4
II
aVL
V2
V5
III
aVF
V3
V6
II

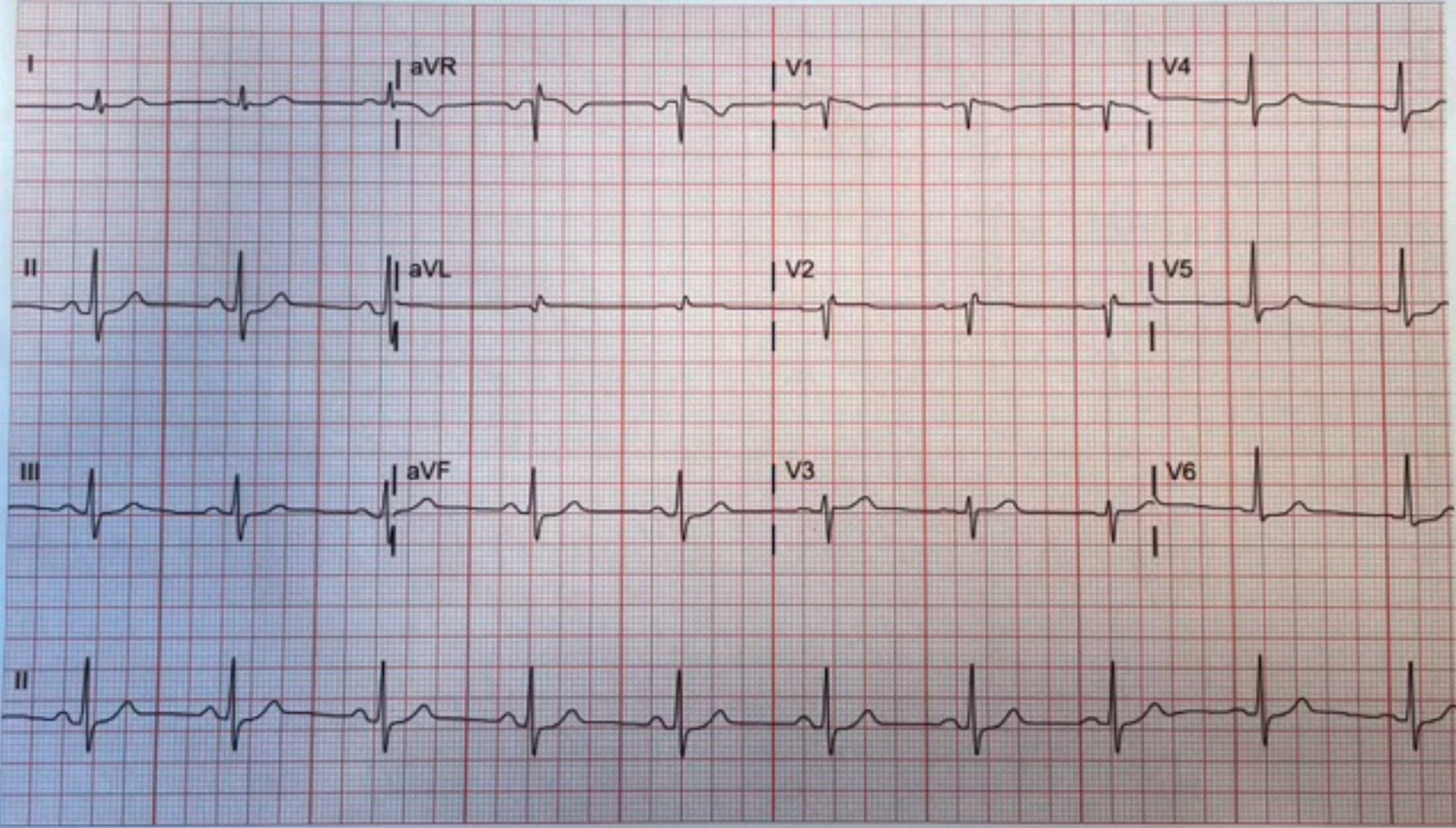
I
aVR
V1
V4
II
aVL
V2
V5
III
aVF
V3
V6
II

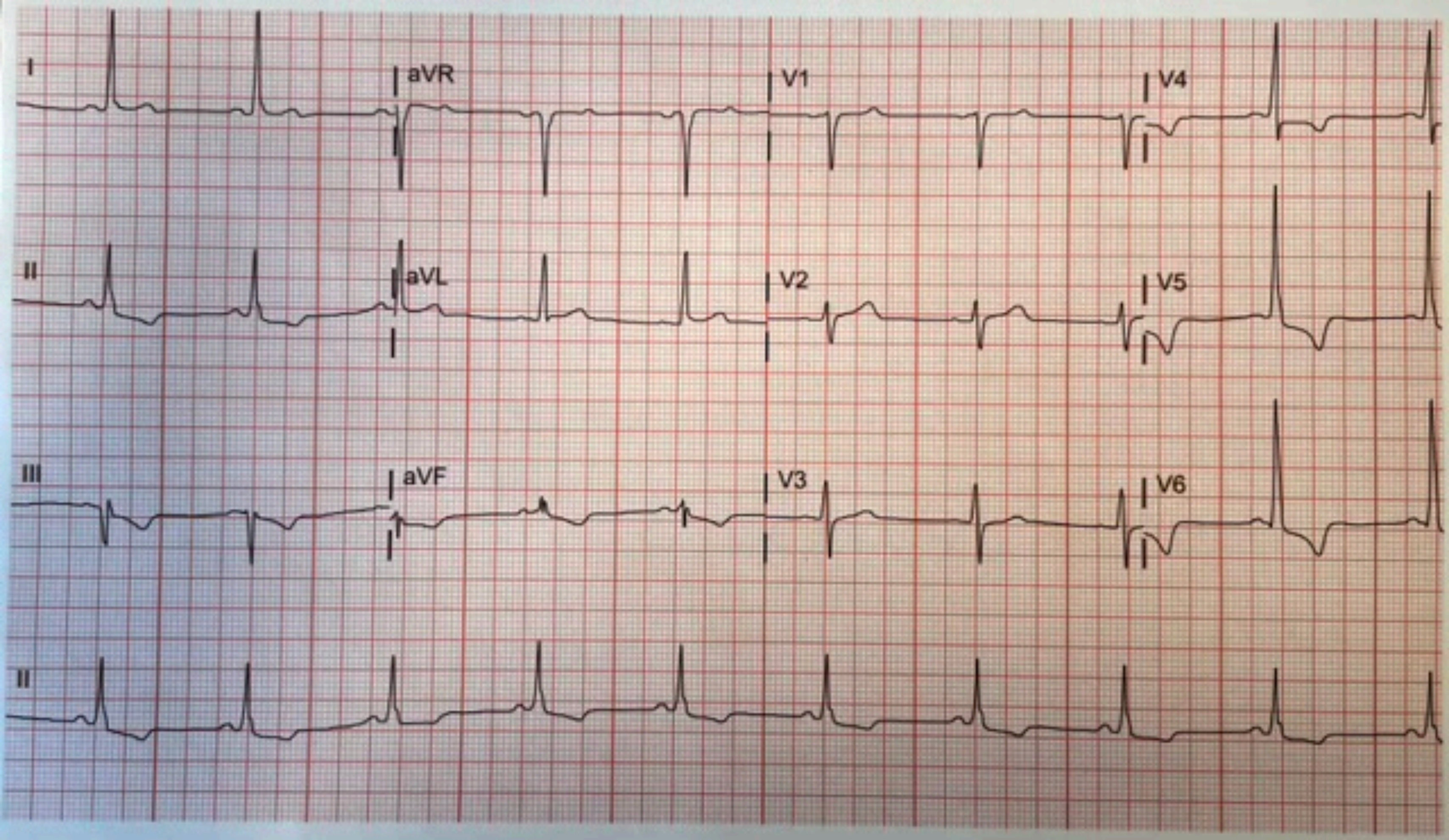
I
aVR
V1
V4
II
aVL
V2
V5
III
aVF
V3
V6
II

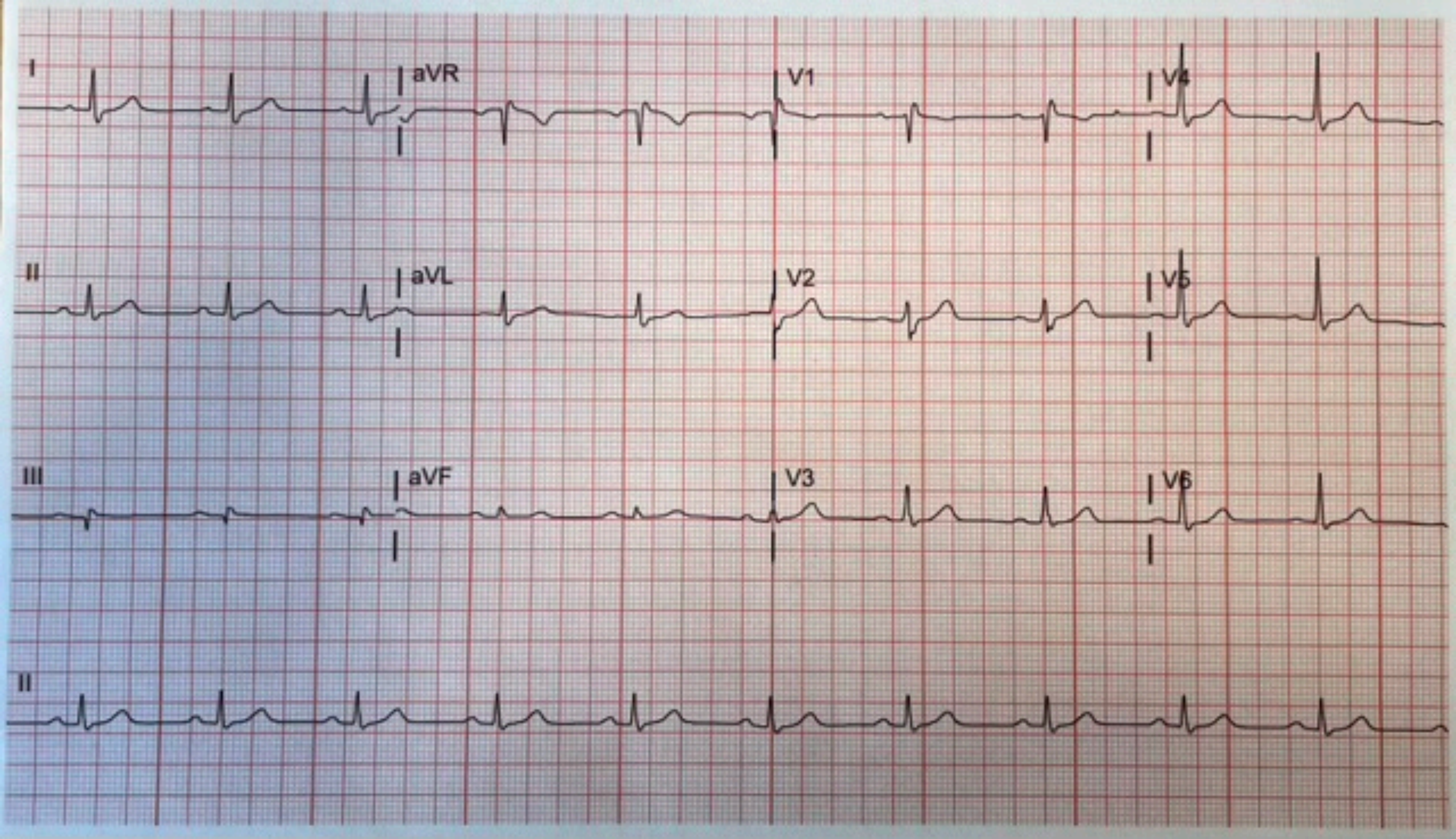
I
aVR
V1
V4
II
aVL
V2
V5
III
aVF
V3
V6
II

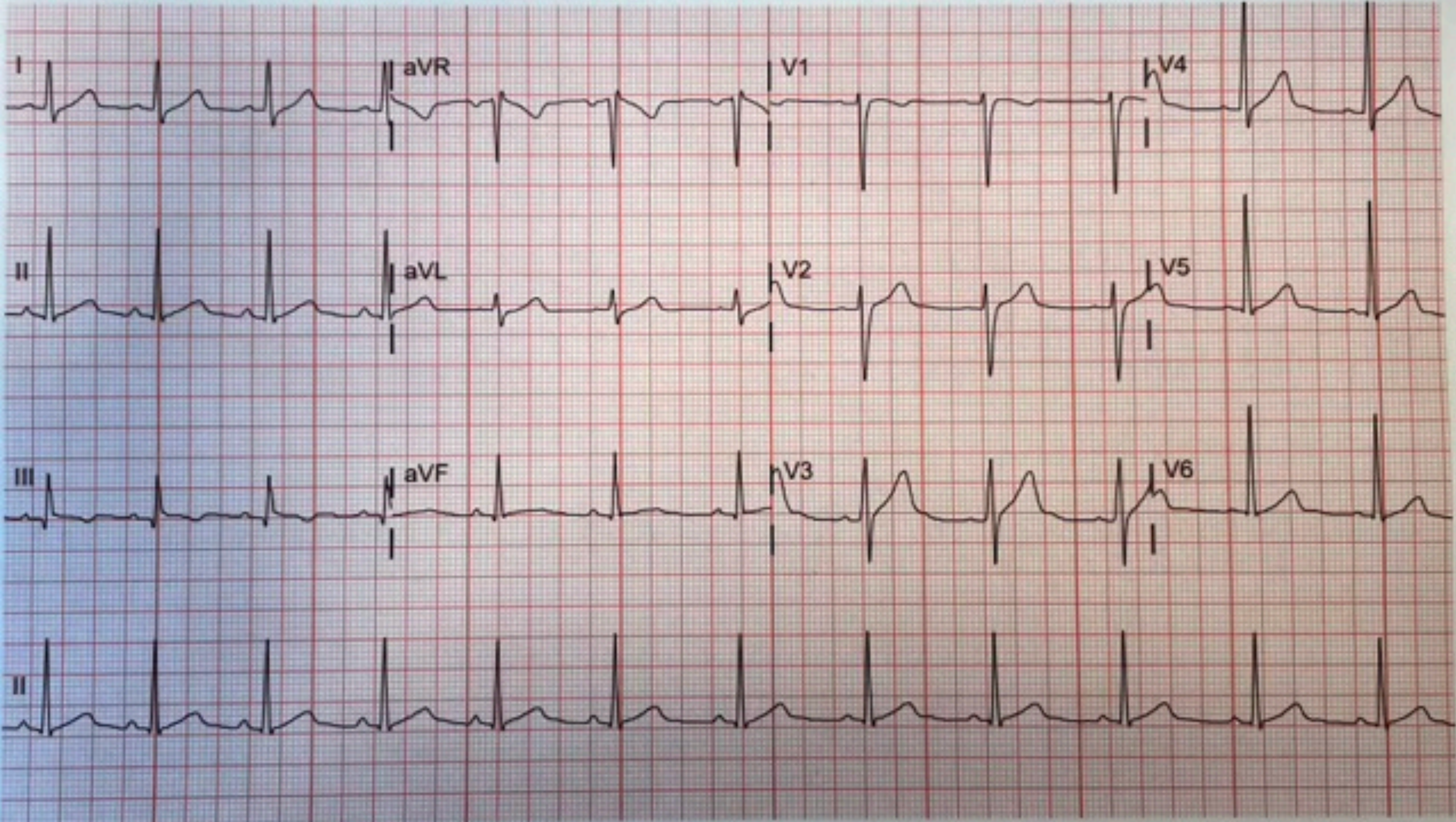
I
aVR
V1
V4
II
aVL
V2
V5
III
aVF
V3
V6
II

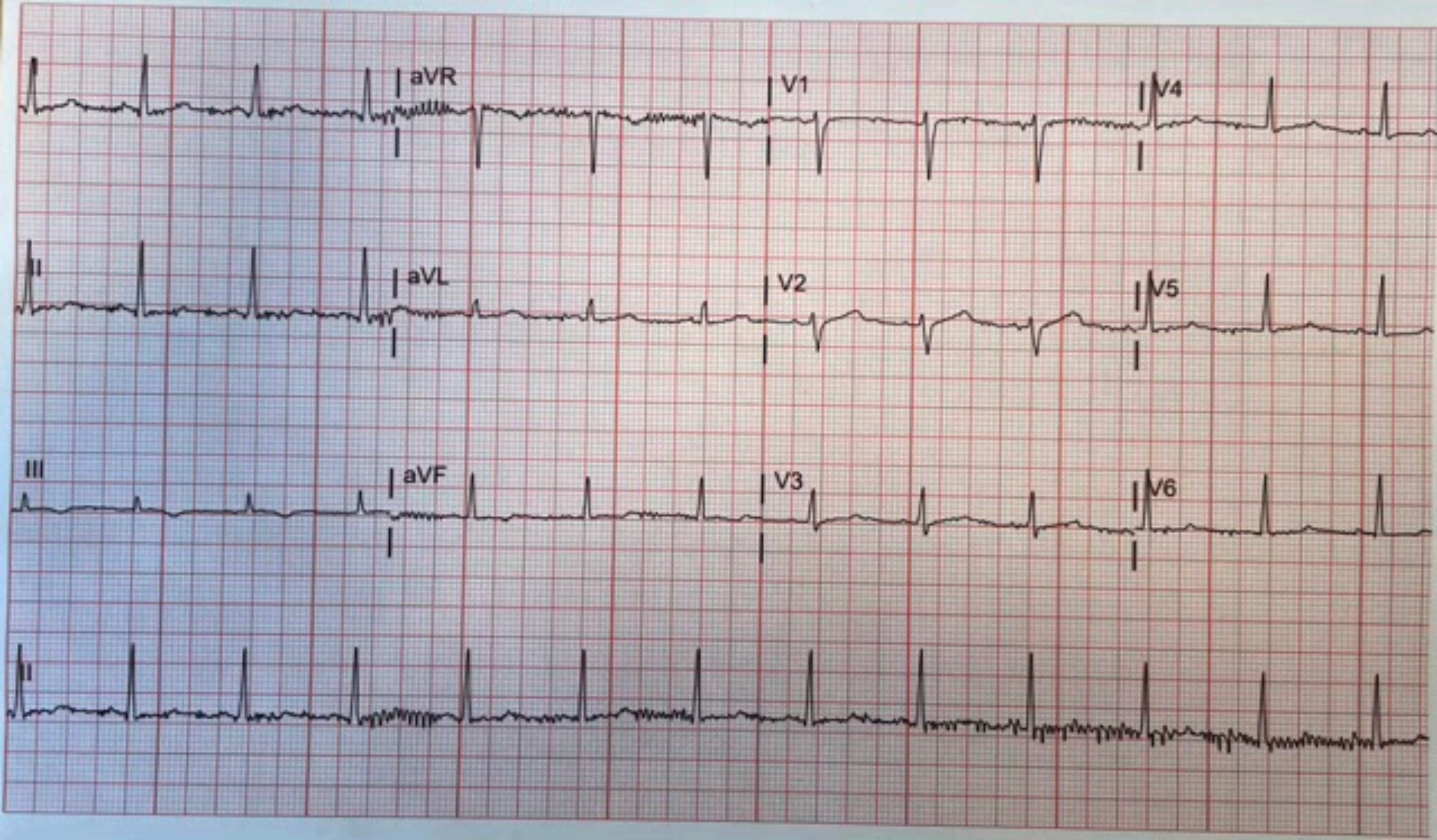
I
aVR
V1
V4
II
aVL
V2
V5
III
aVF
V3
V6
II

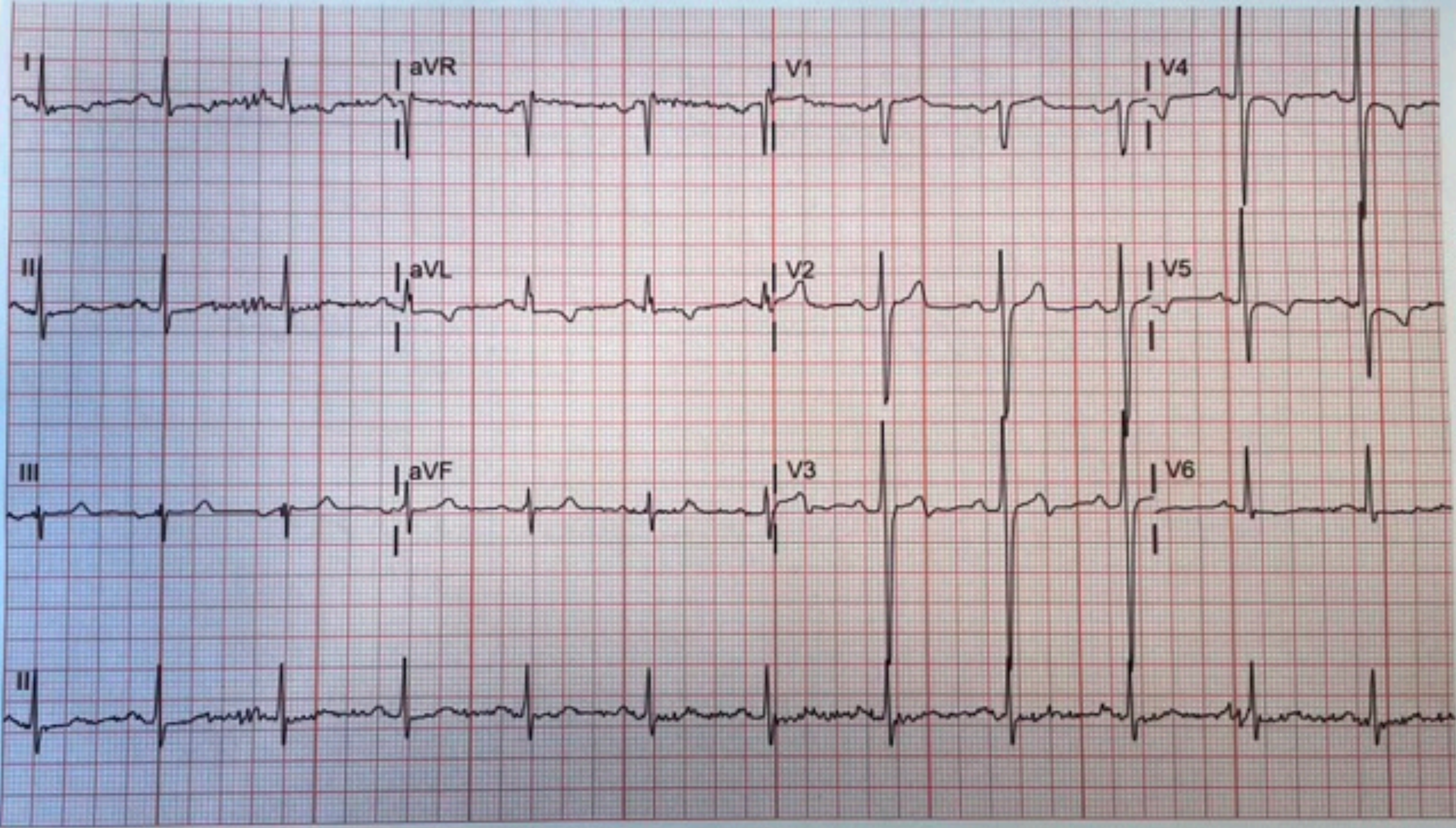

I
aVR
V1
V4
II
aVL
V2
V5
III
aVF
V3
V6
II

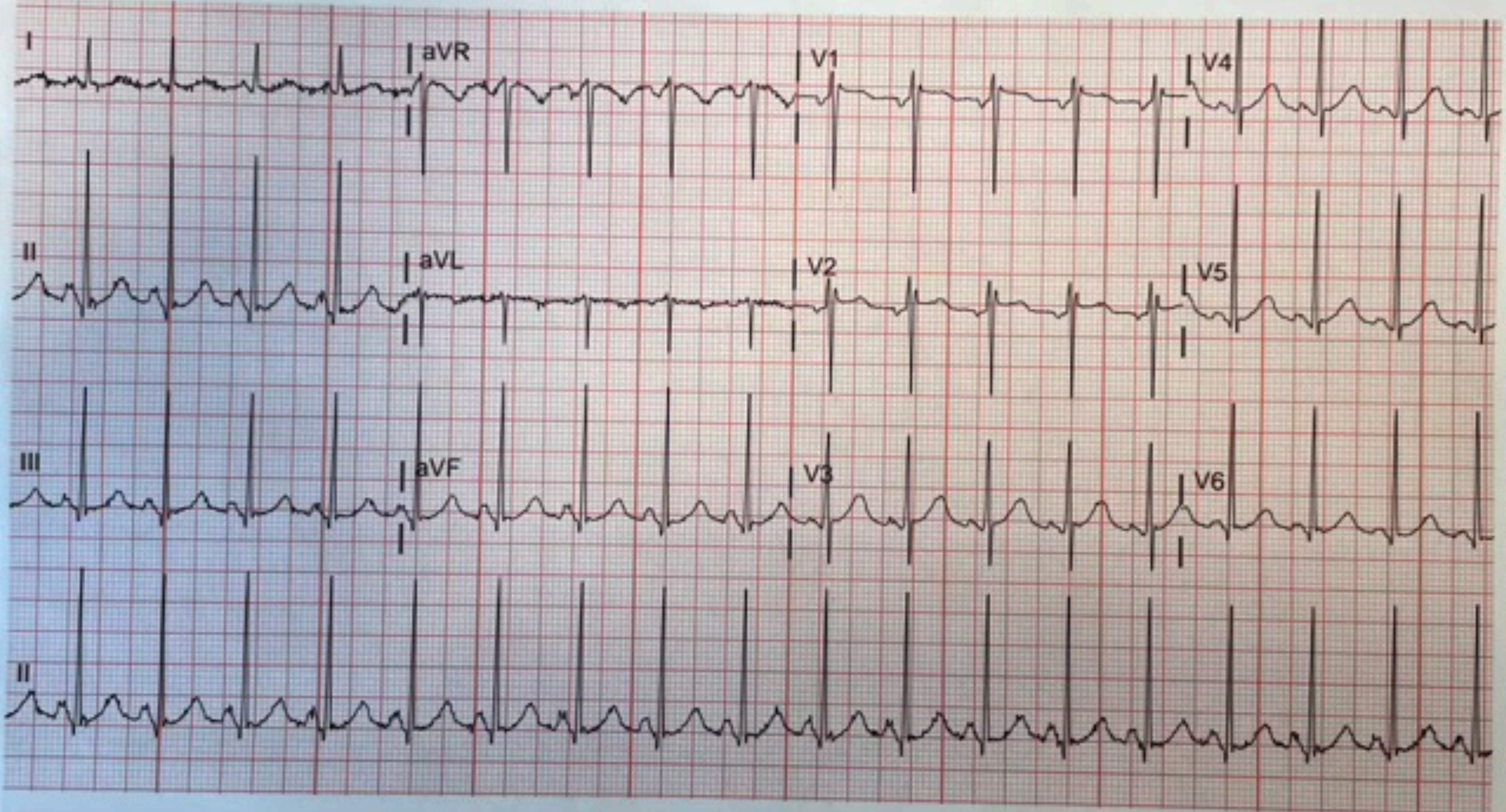
I
aVR
V1
V4
II
aVL
V2
V5
III
aVF
V3
V6
II

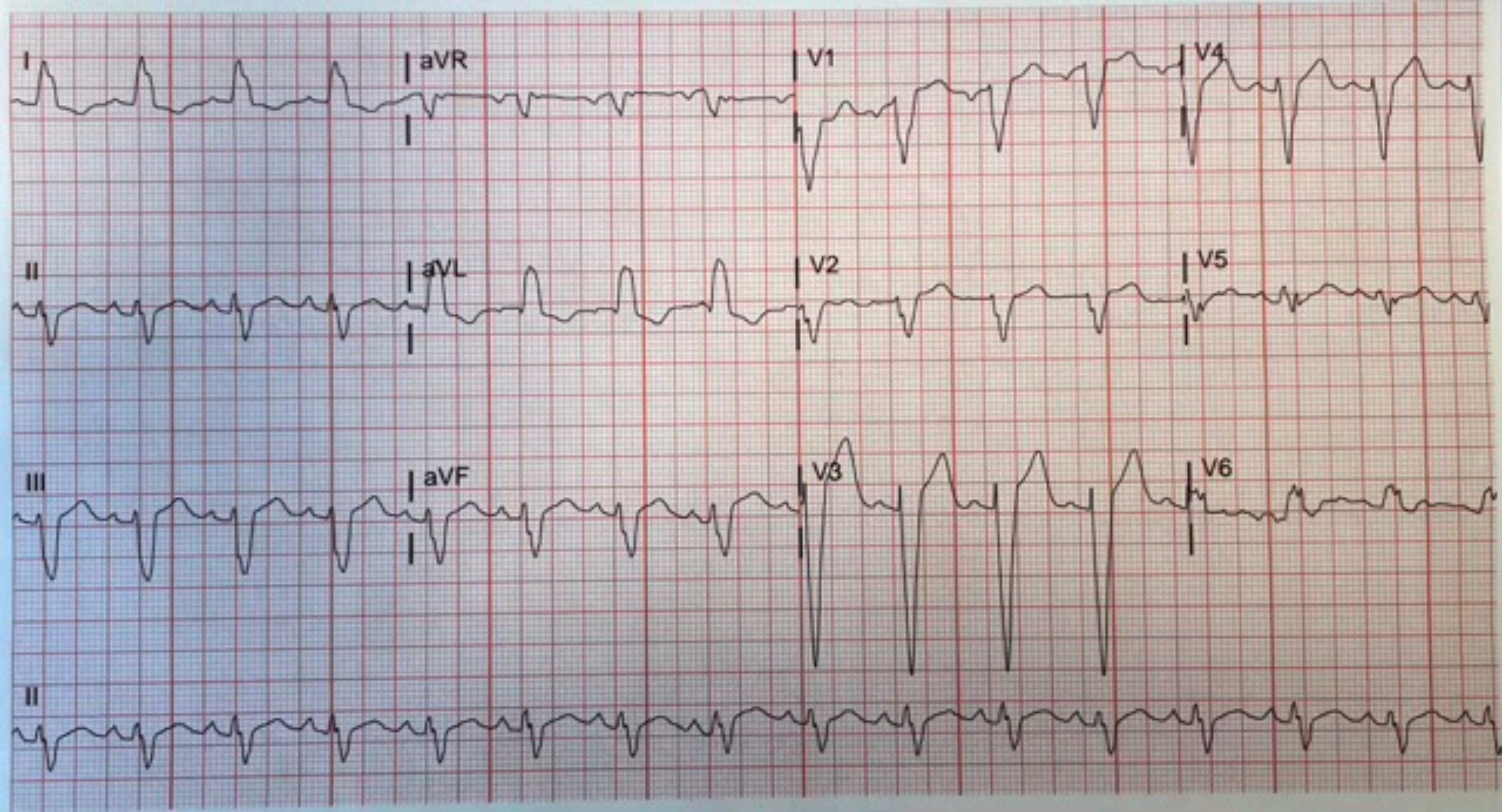
I
aVR
V1
V4
II
aVL
V2
V5
III
aVF
V3
V6
II

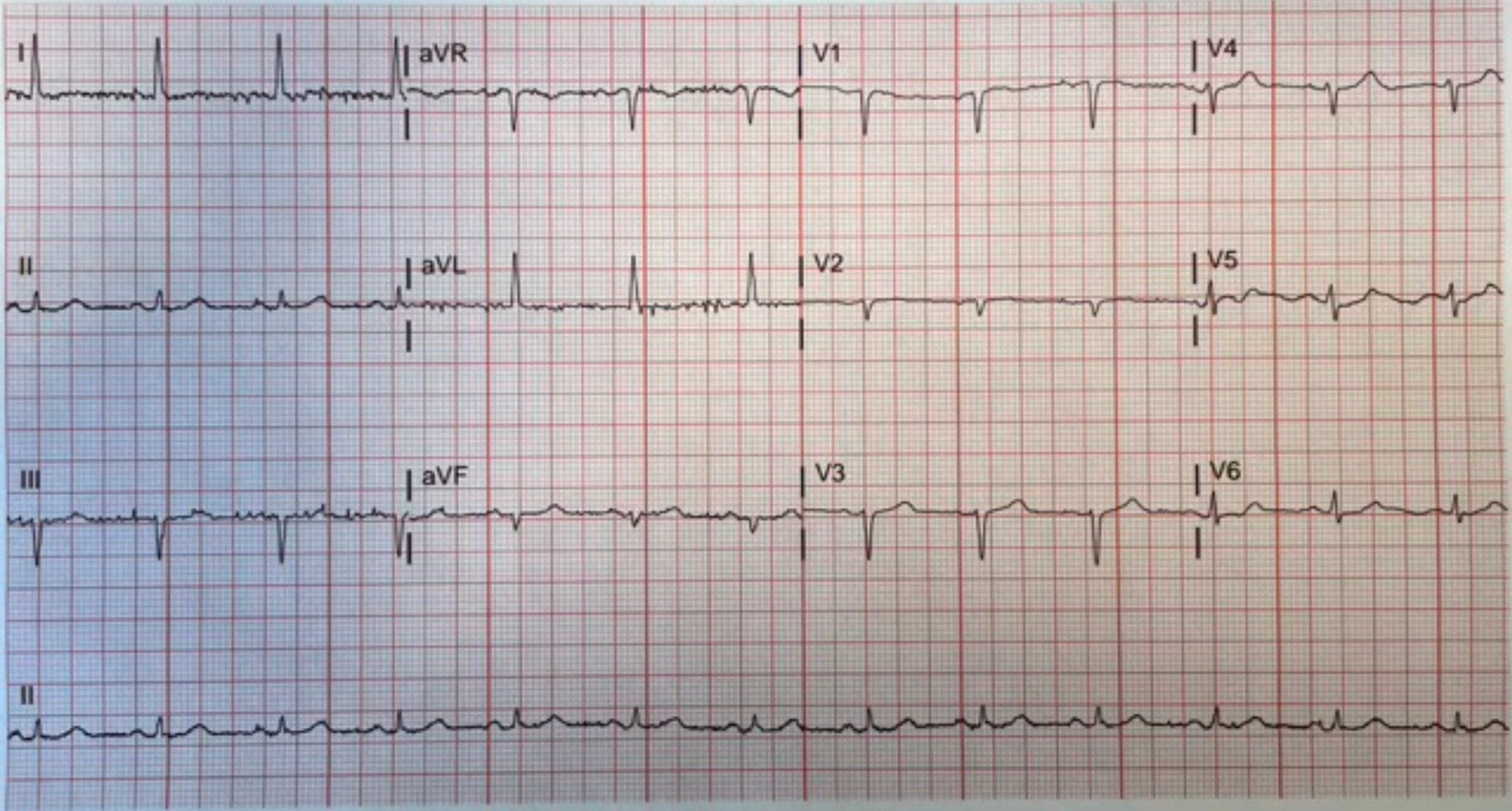
I
aVR
V1
V4
II
aVL
V2
V5
III
aVF
V3
V6
II

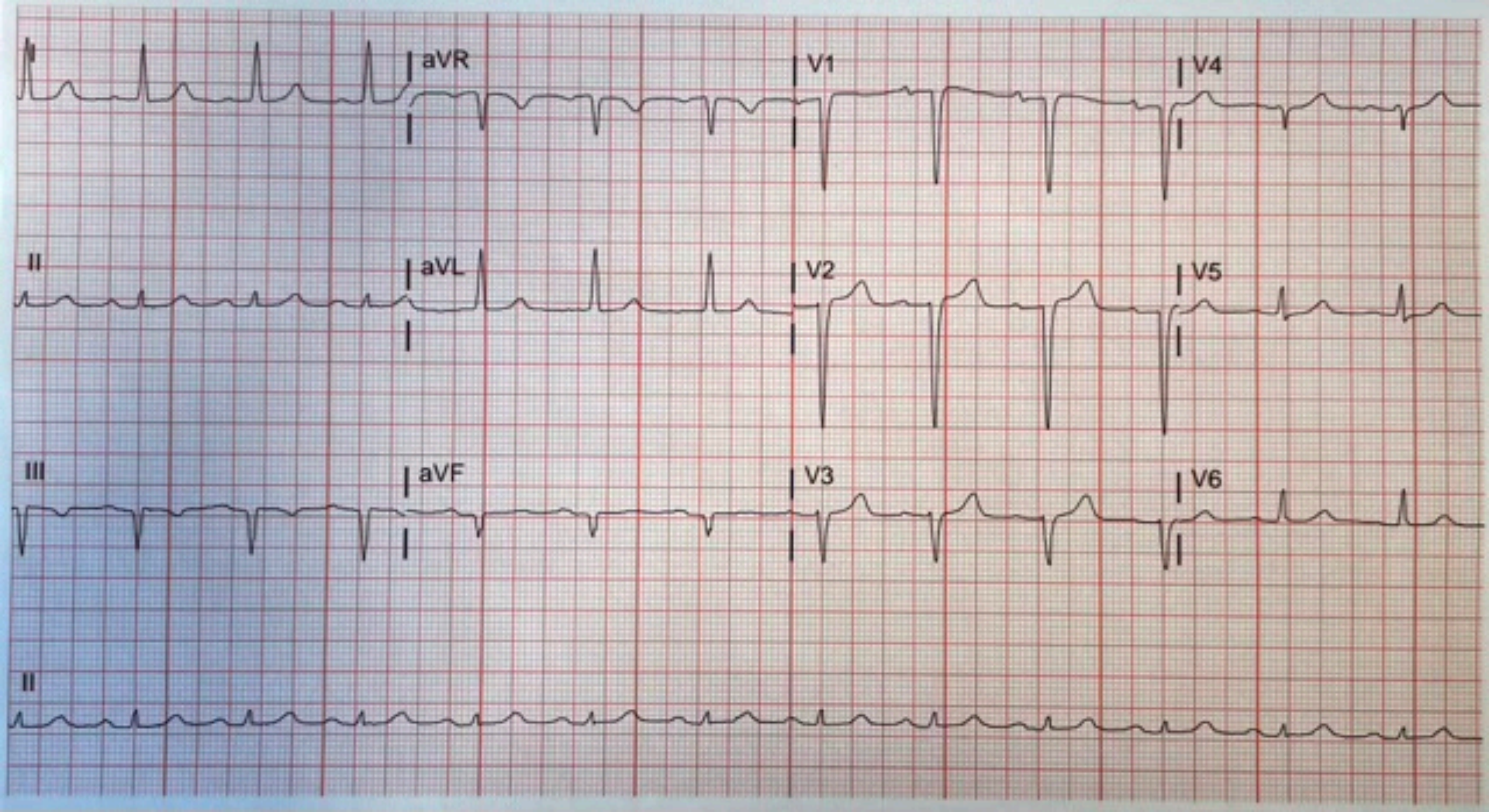
I
aVR
V1
V4
II
aVL
V2
V5
III
aVF
V3
V6
II

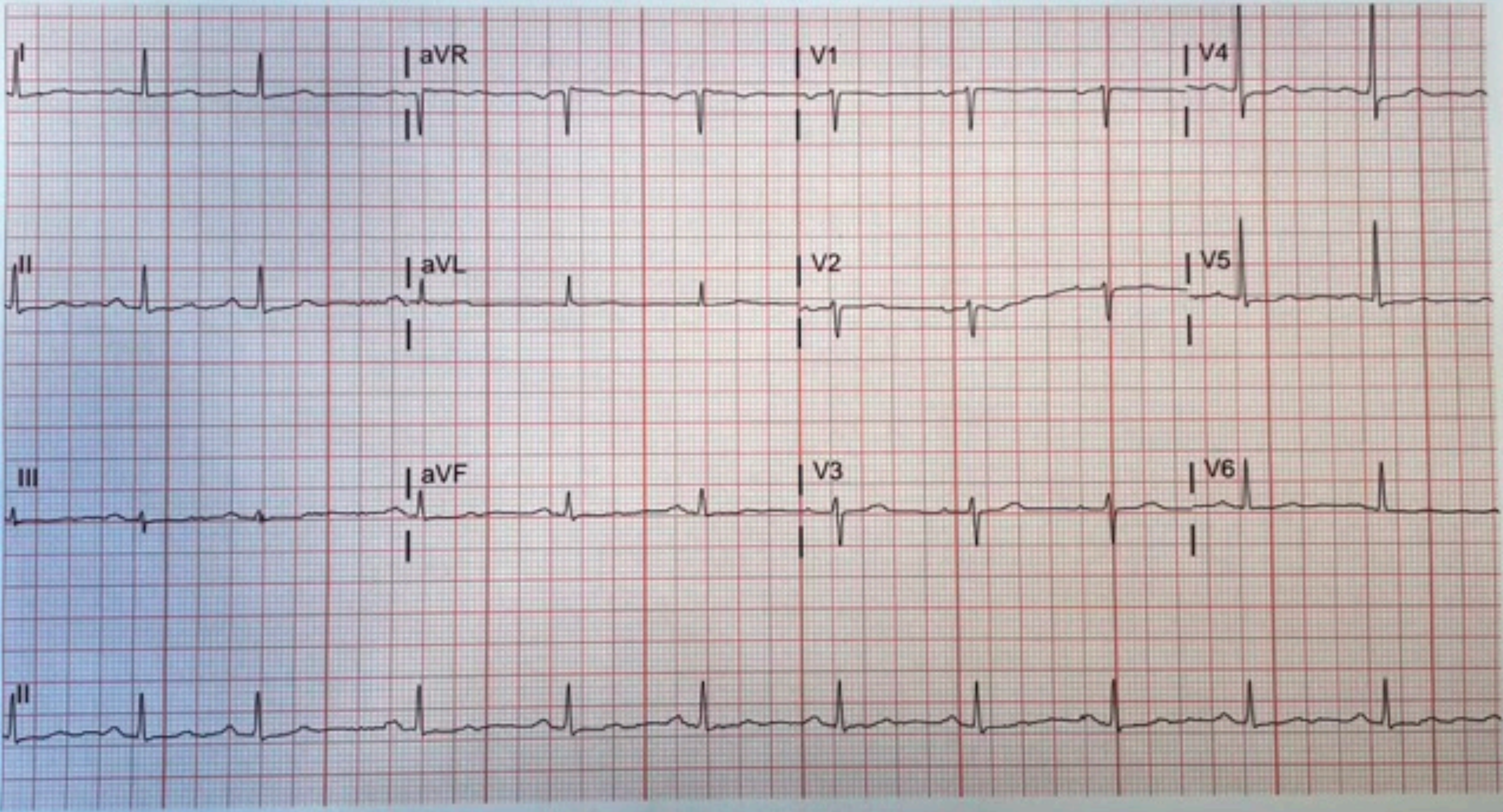
I
aVR
V1
V4
II
aVL
V2
V5
III
aVF
V3
V6
II

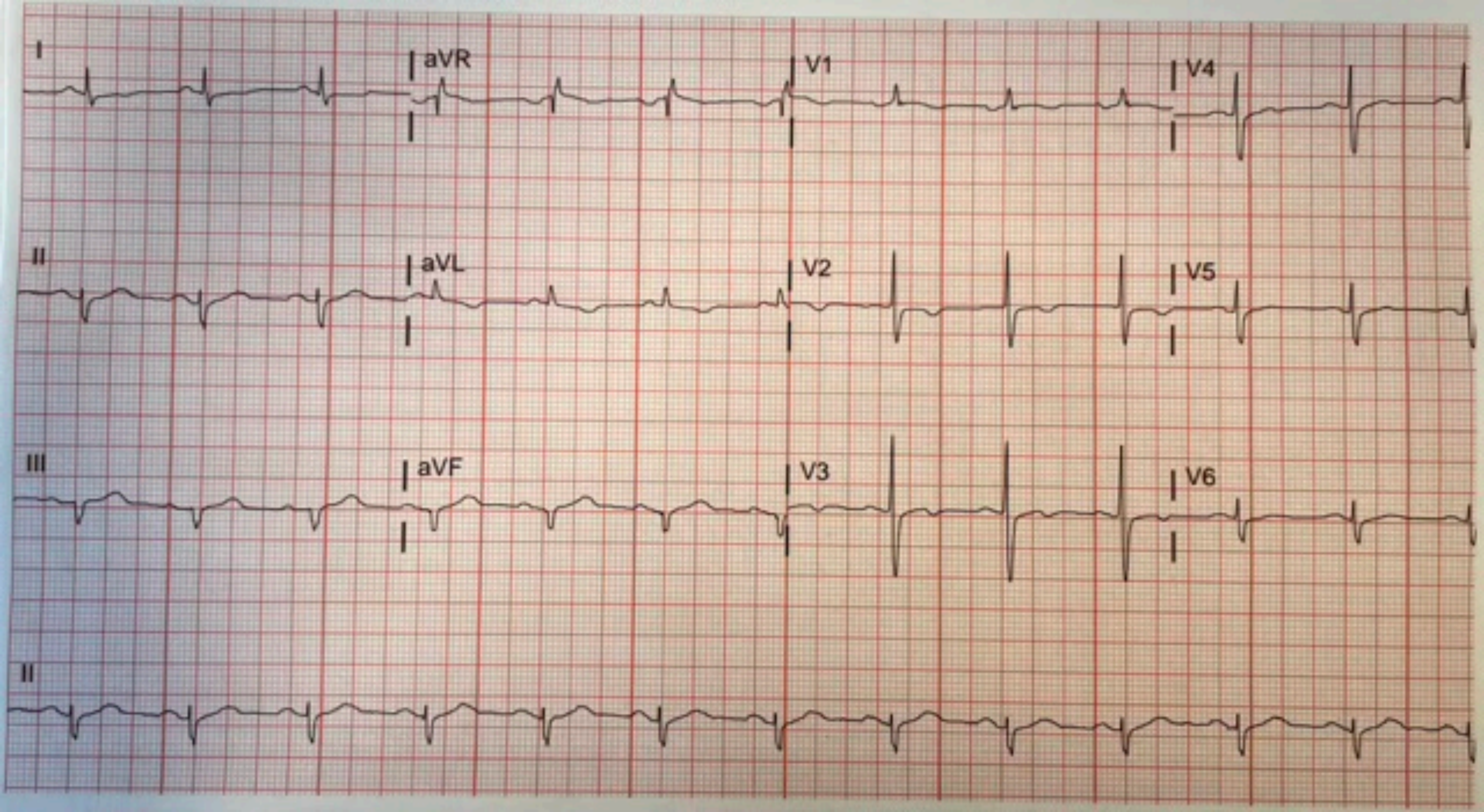
I
aVR
V1
V4
II
aVL
V2
V5
III
aVF
V3
V6
II

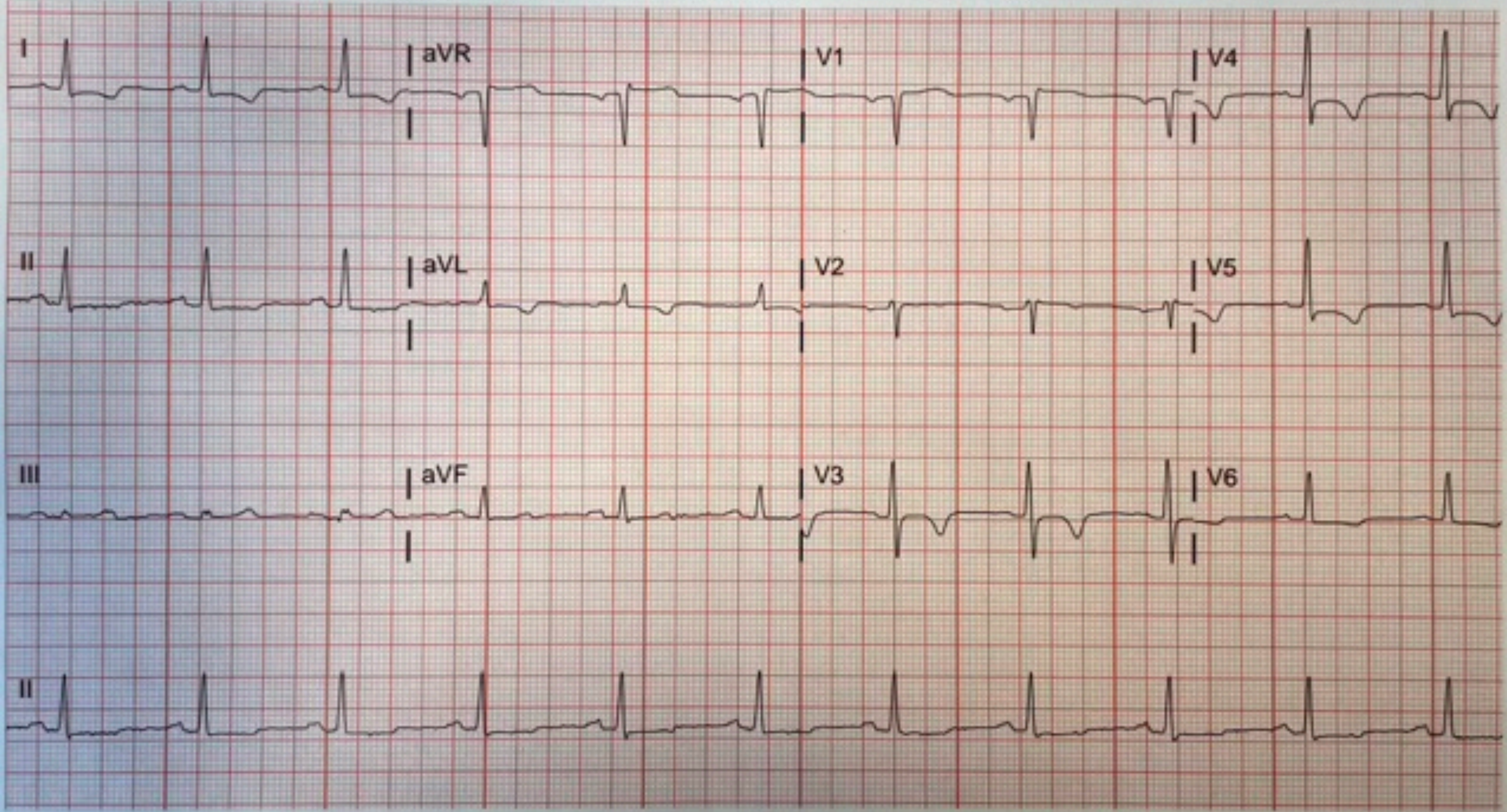
I
aVR
V1
V4
II
aVL
V2
V5
III
aVF
V3
V6
II

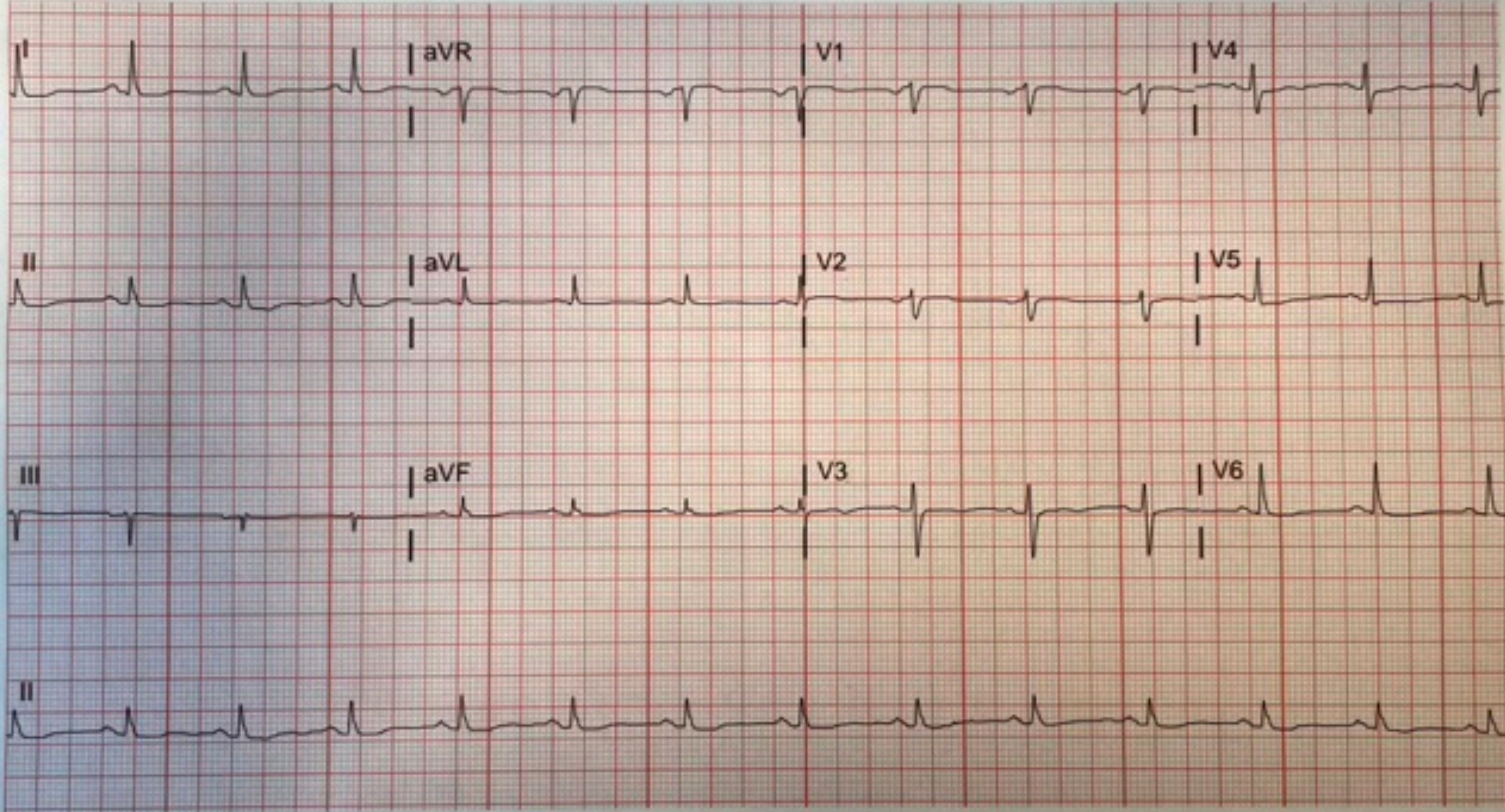
I
aVR
V1
V4
II
aVL
V2
V5
III
aVF
V3
V6
II

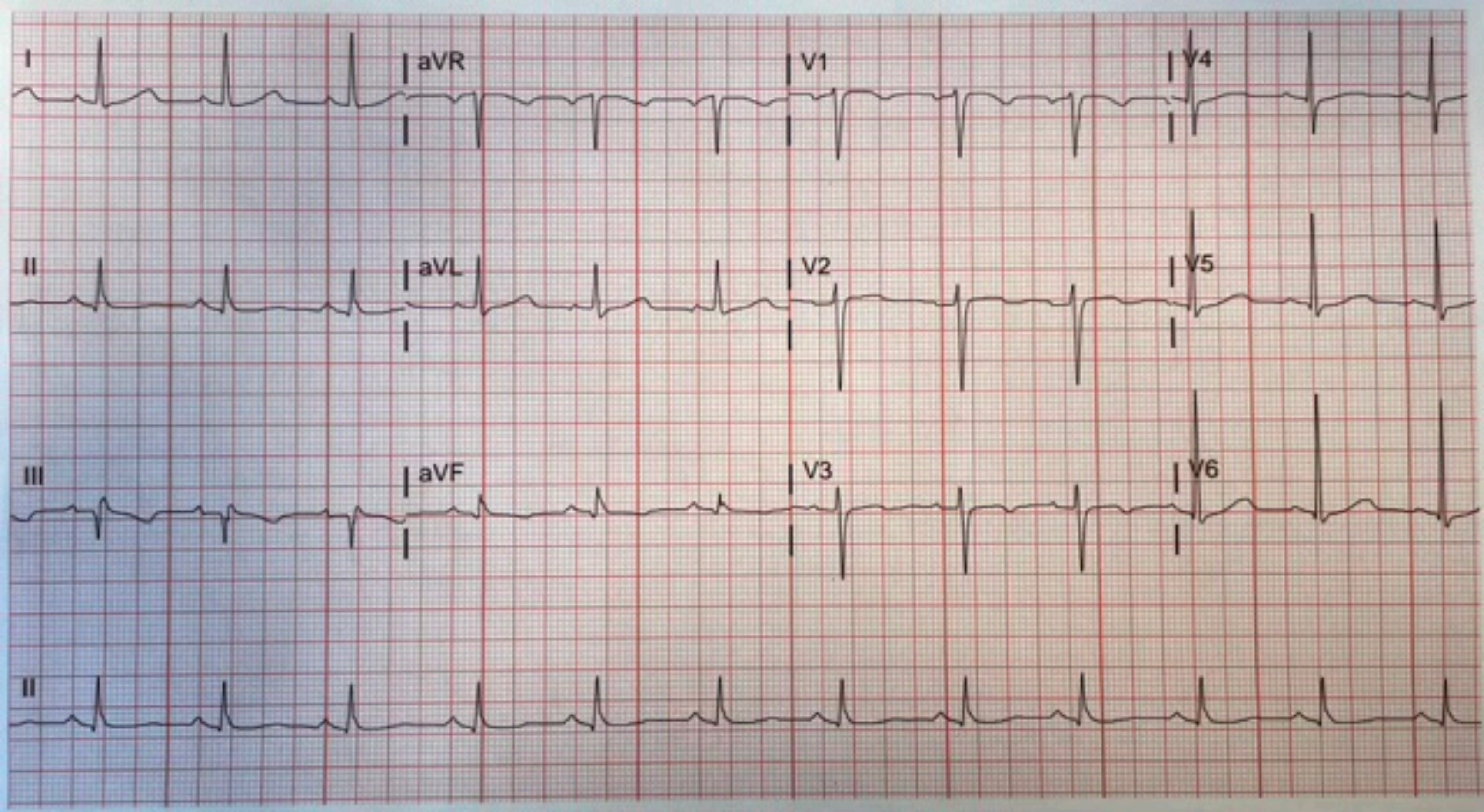
I
aVR
V1
V4
II
aVL
V2
V5
III
aVF
V3
V6
II

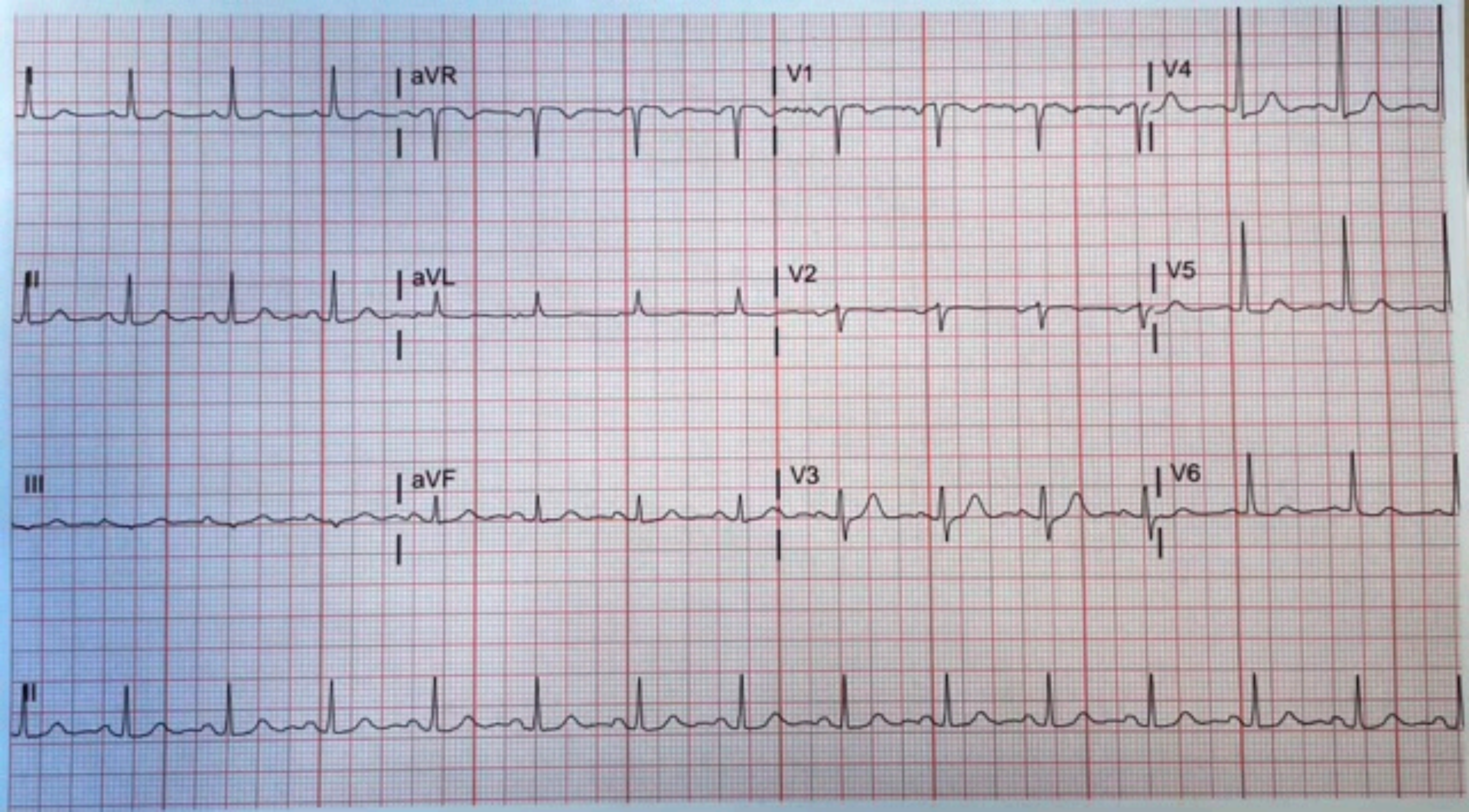
I
aVR
V1
V4
II
aVL
V2
V5
III
aVF
V3
V6
II

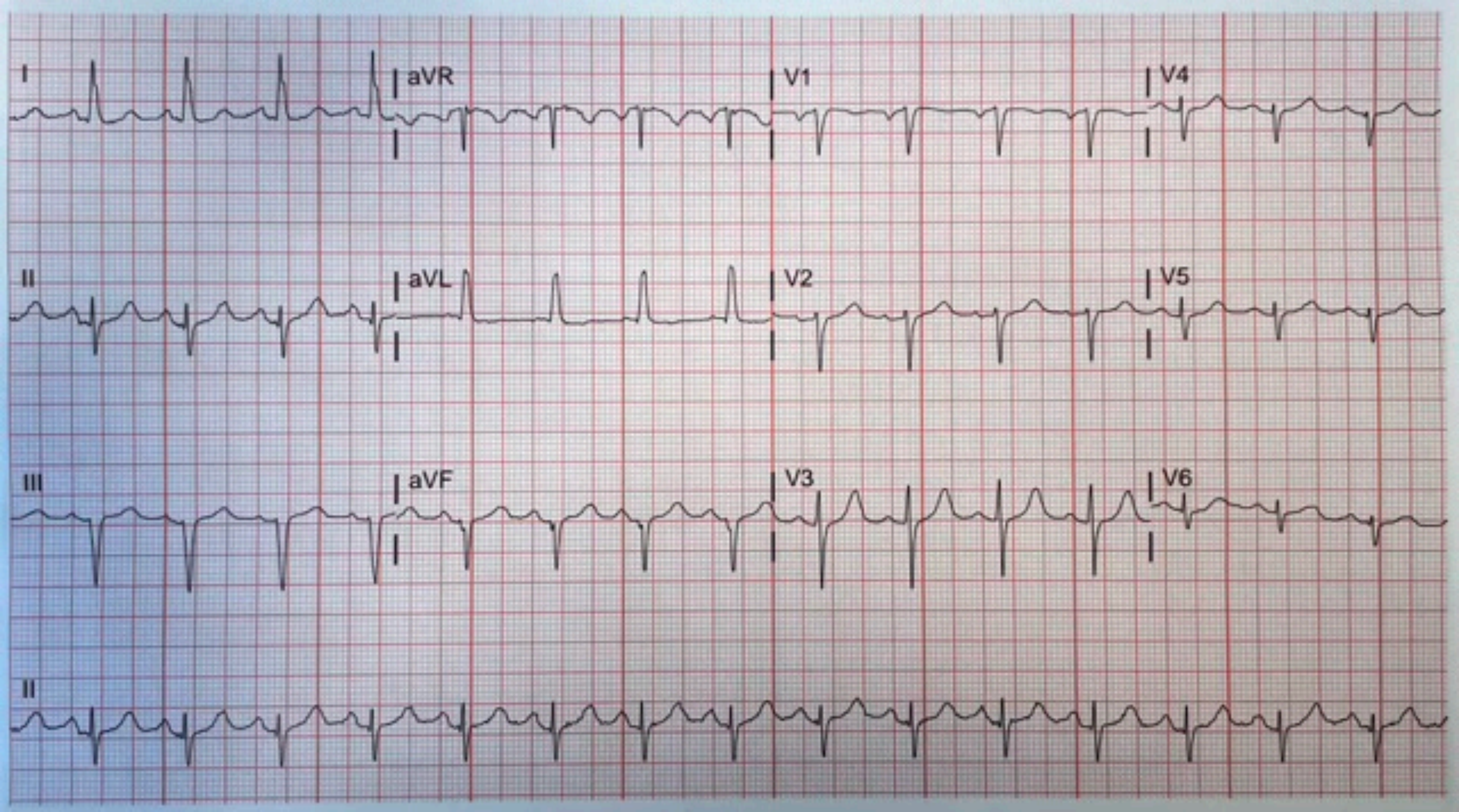
I
aVR
V1
V4
II
aVL
V2
V5
III
aVF
V3
V6
II

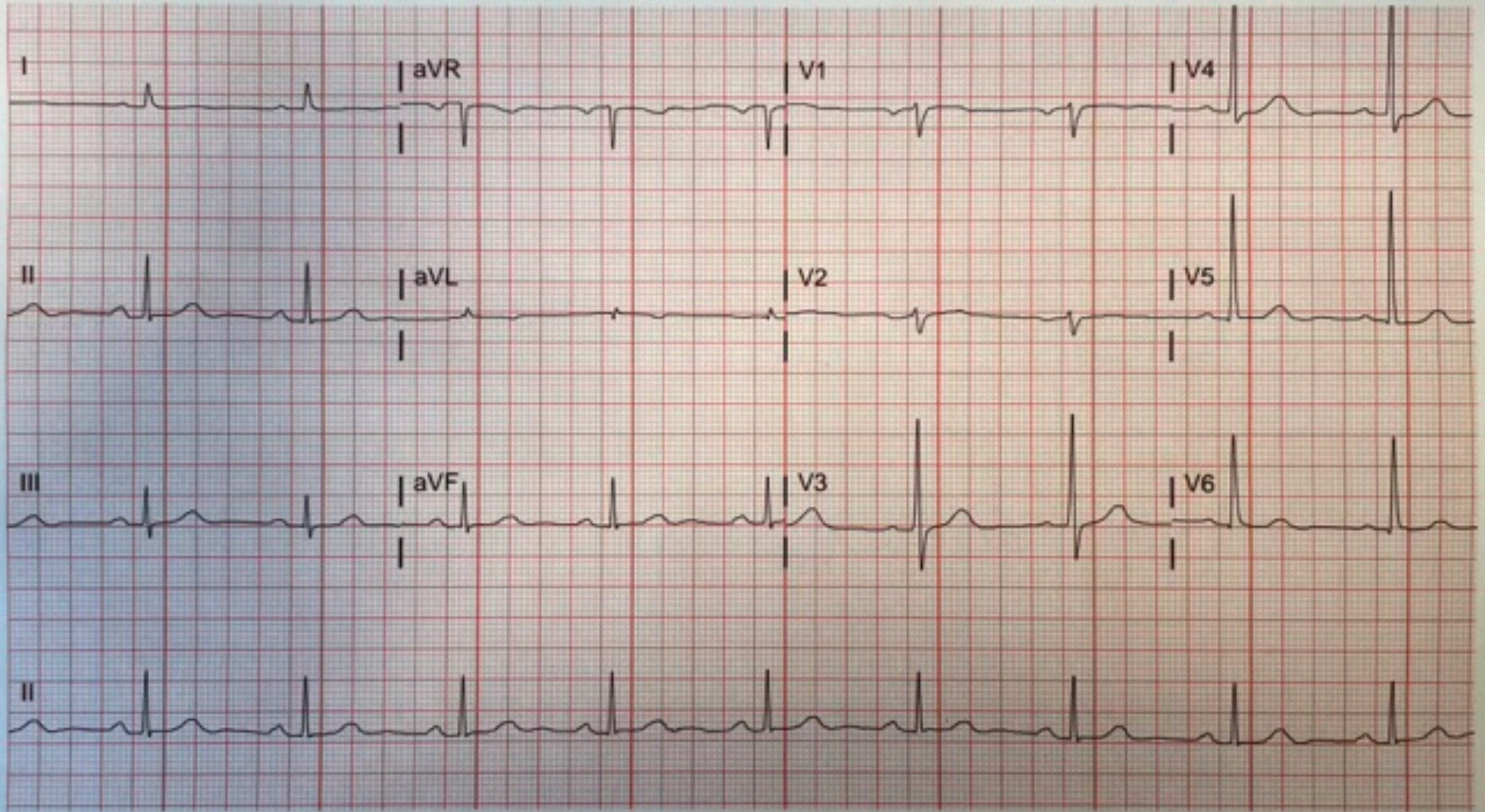
I
aVR
V1
V4
II
aVL
V2
V5
III
aVF
V3
V6
II

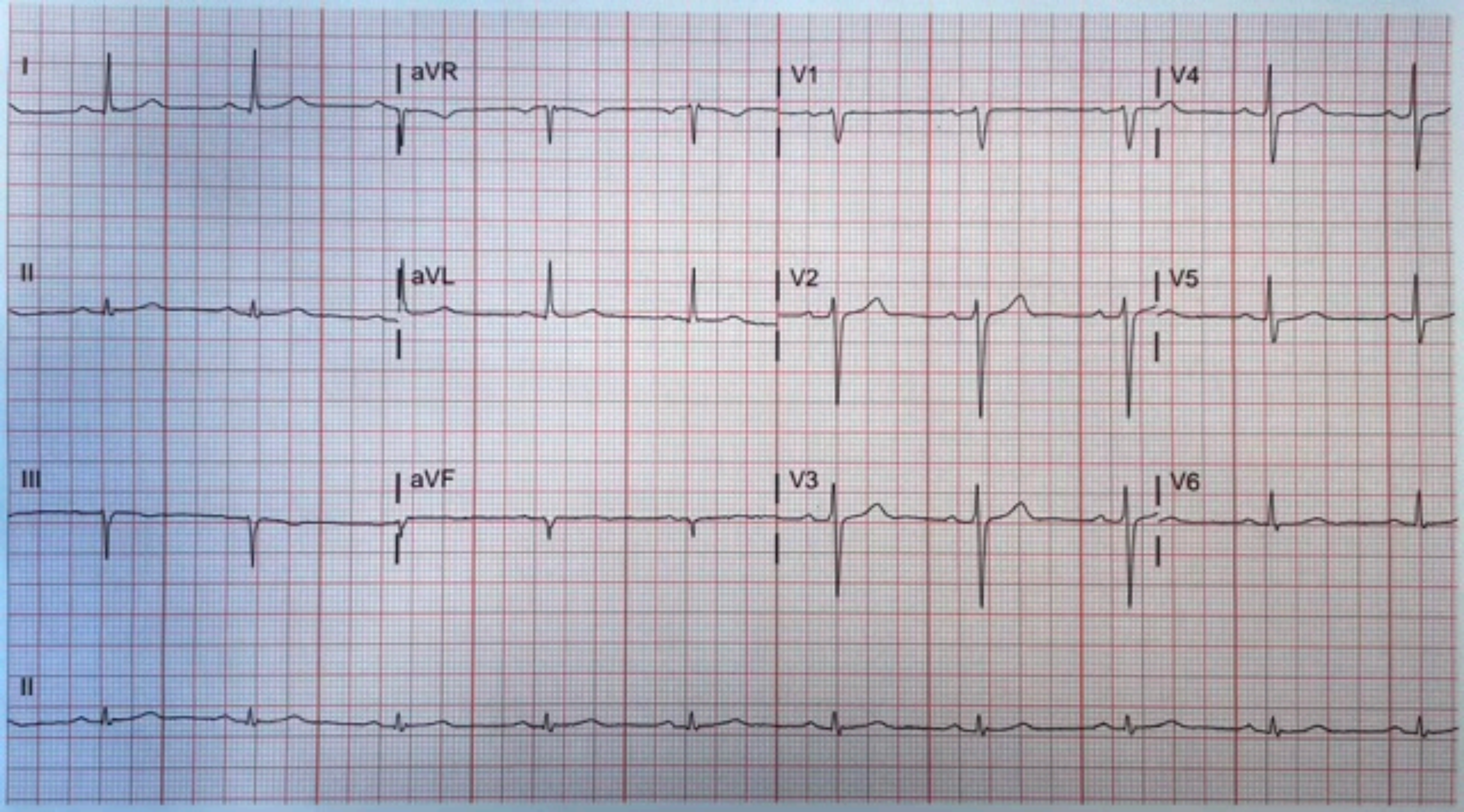
I
aVR
V1
V4
II
aVL
V2
V5
III
aVF
V3
V6
II

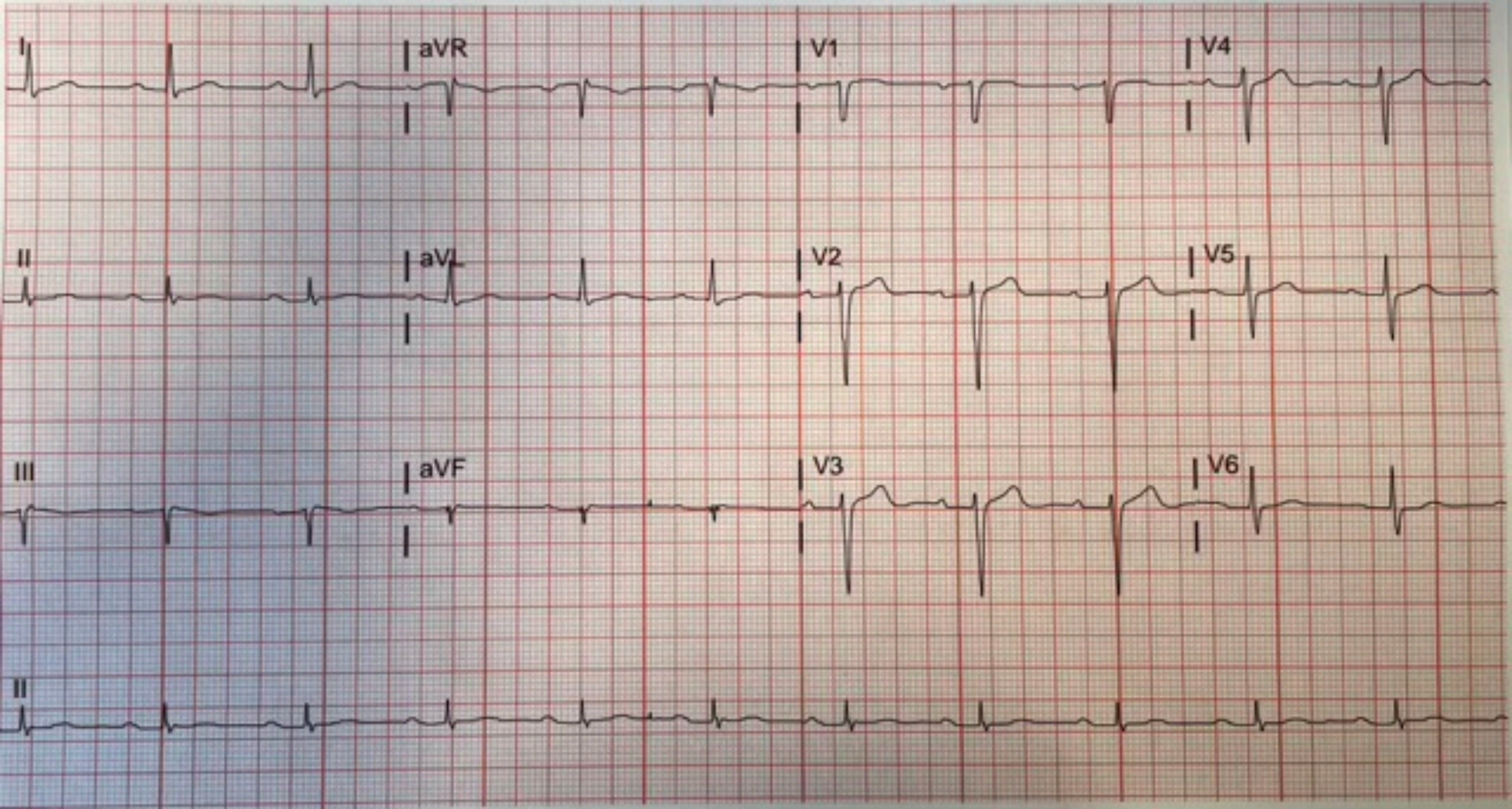
I
aVR
V1
V4
II
aVL
V2
V5
III
aVF
V3
V6
II

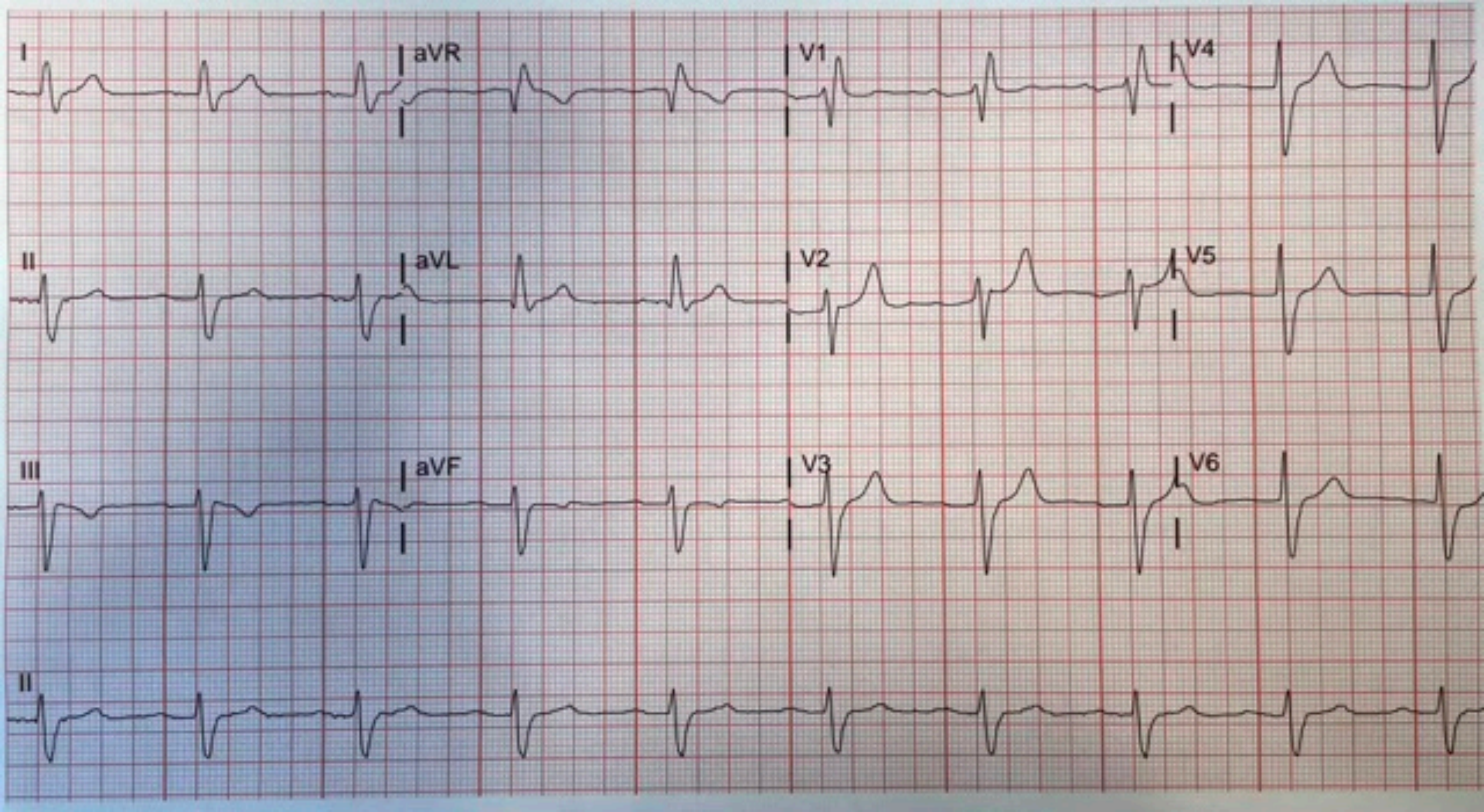
I
aVR
V1
V4
II
aVL
V2
V5
III
aVF
V3
V6
II

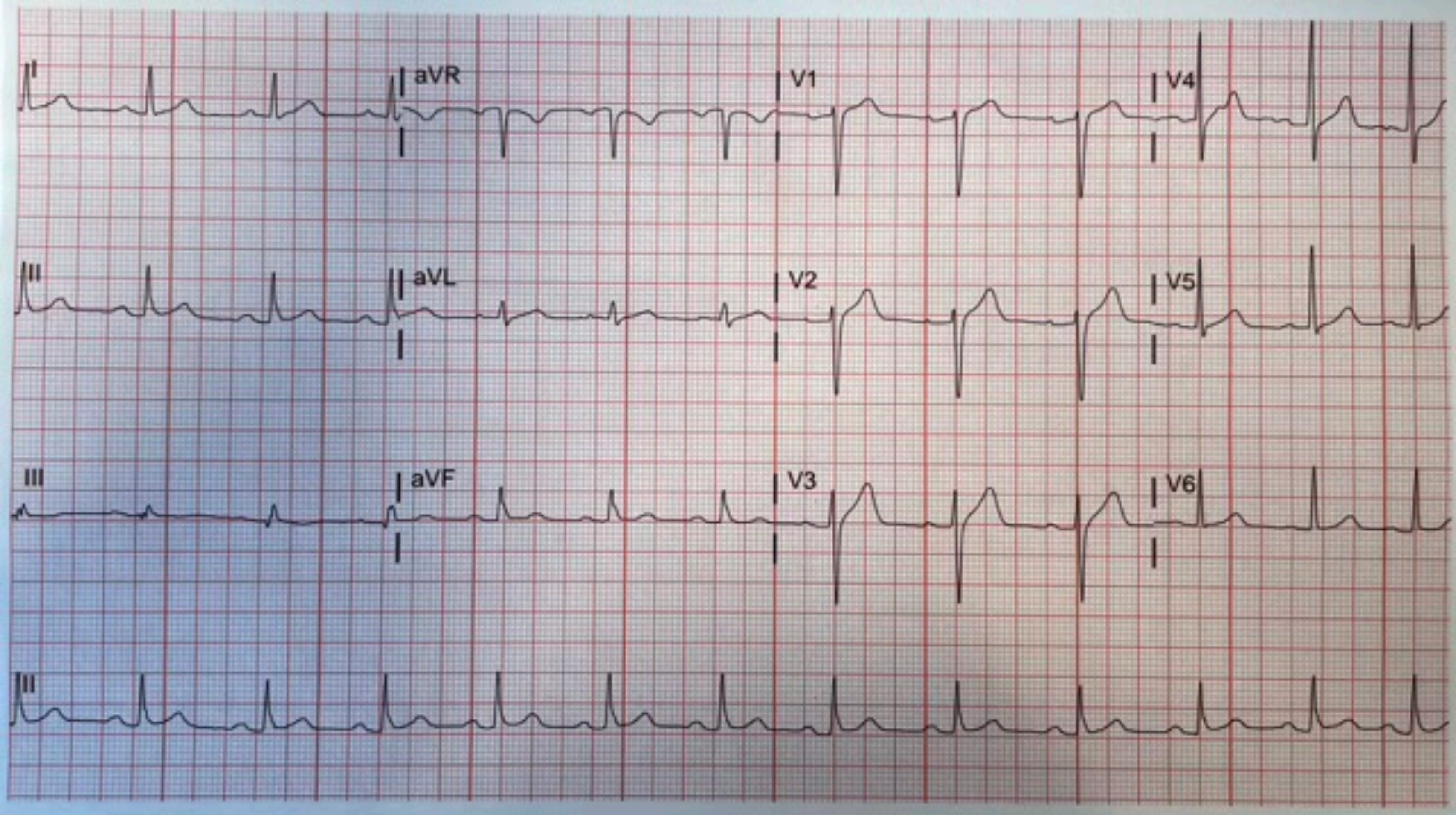
I
aVR
V1
V4
II
aVL
V2
V5
III
aVF
V3
V6
II

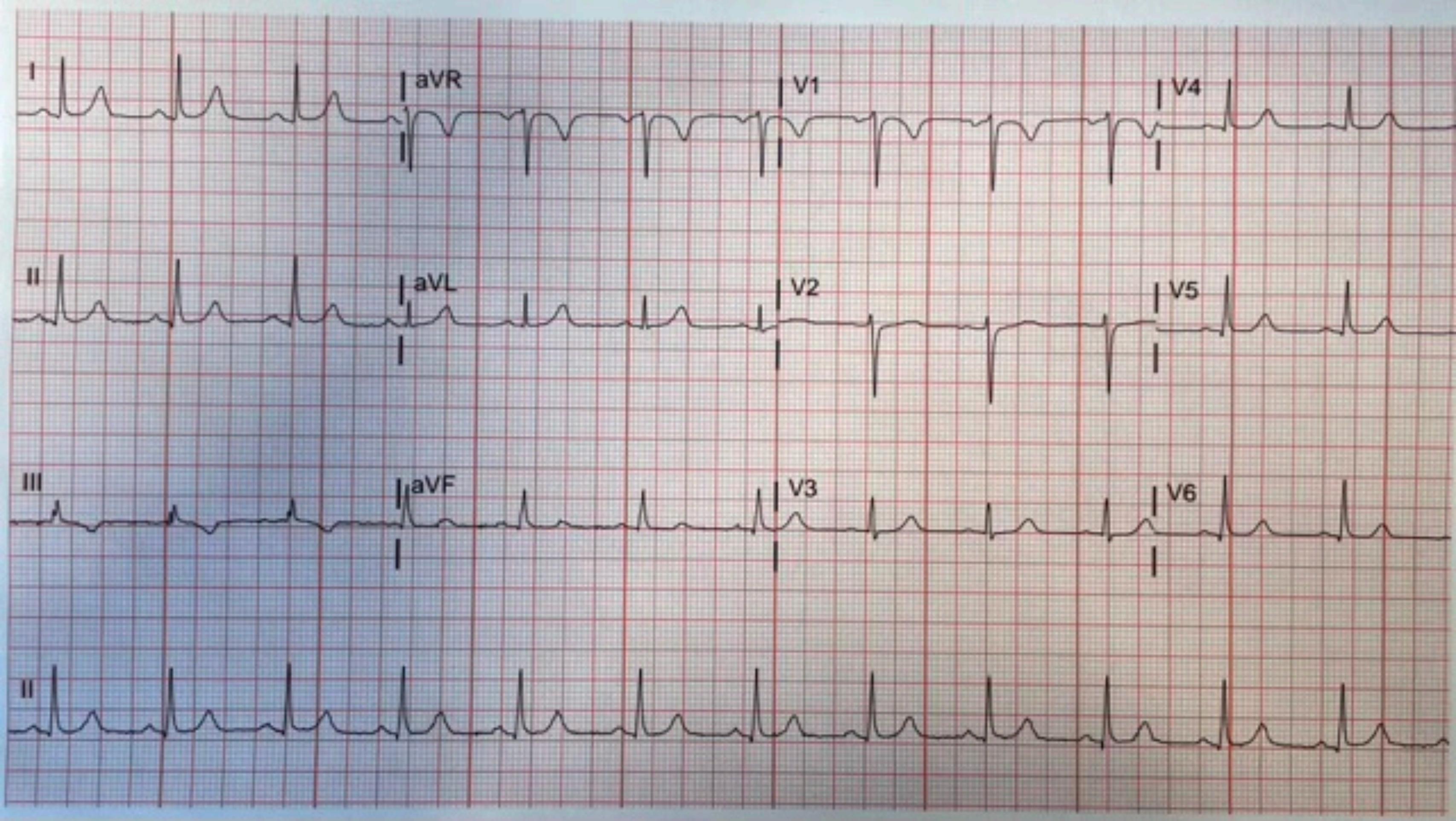
I
aVR
V1
V4
II
aVL
V2
V5
III
aVF
V3
V6
II

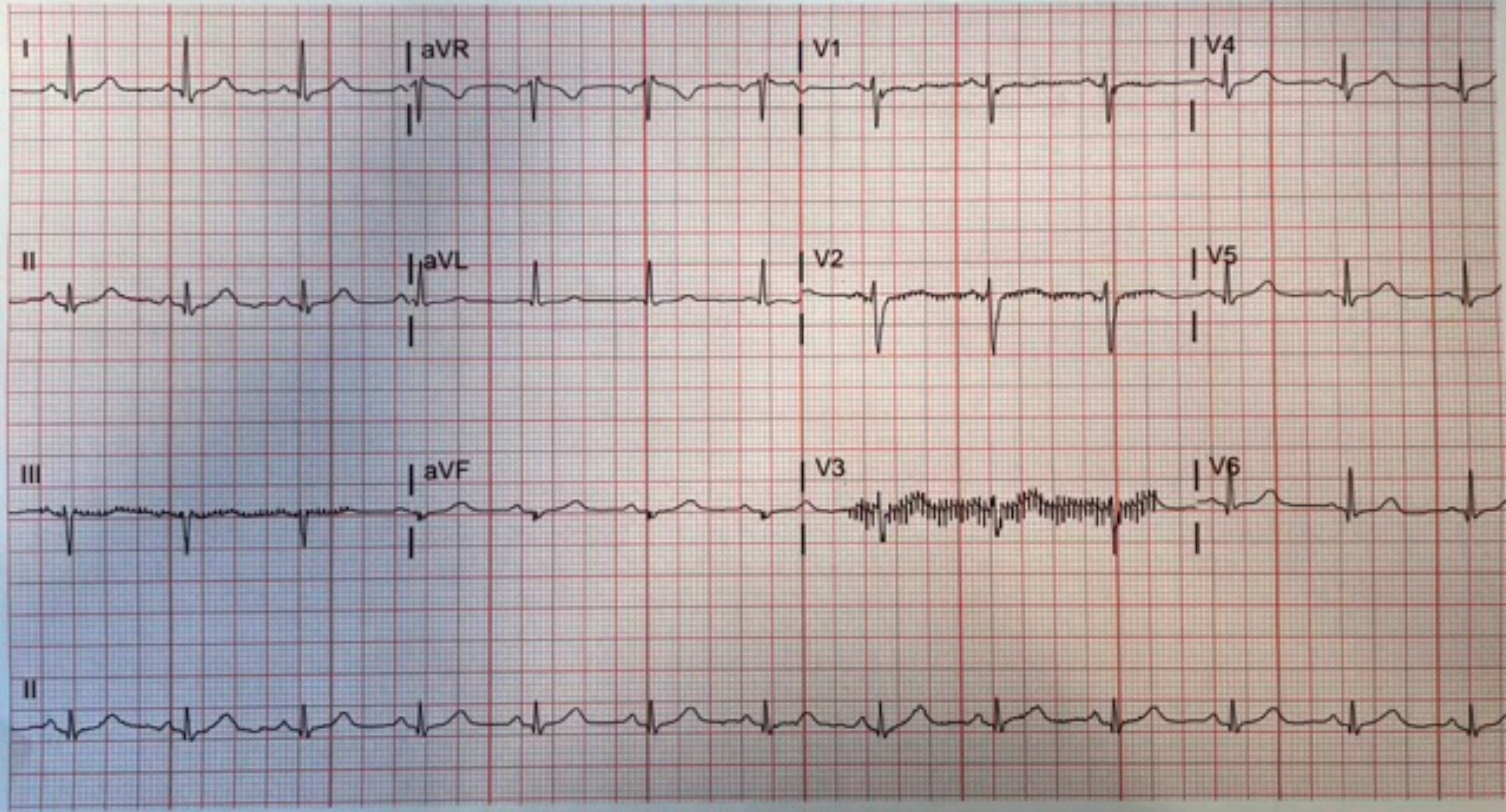
I
aVR
V1
V4
II
aVL
V2
V5
III
aVF
V3
V6
II

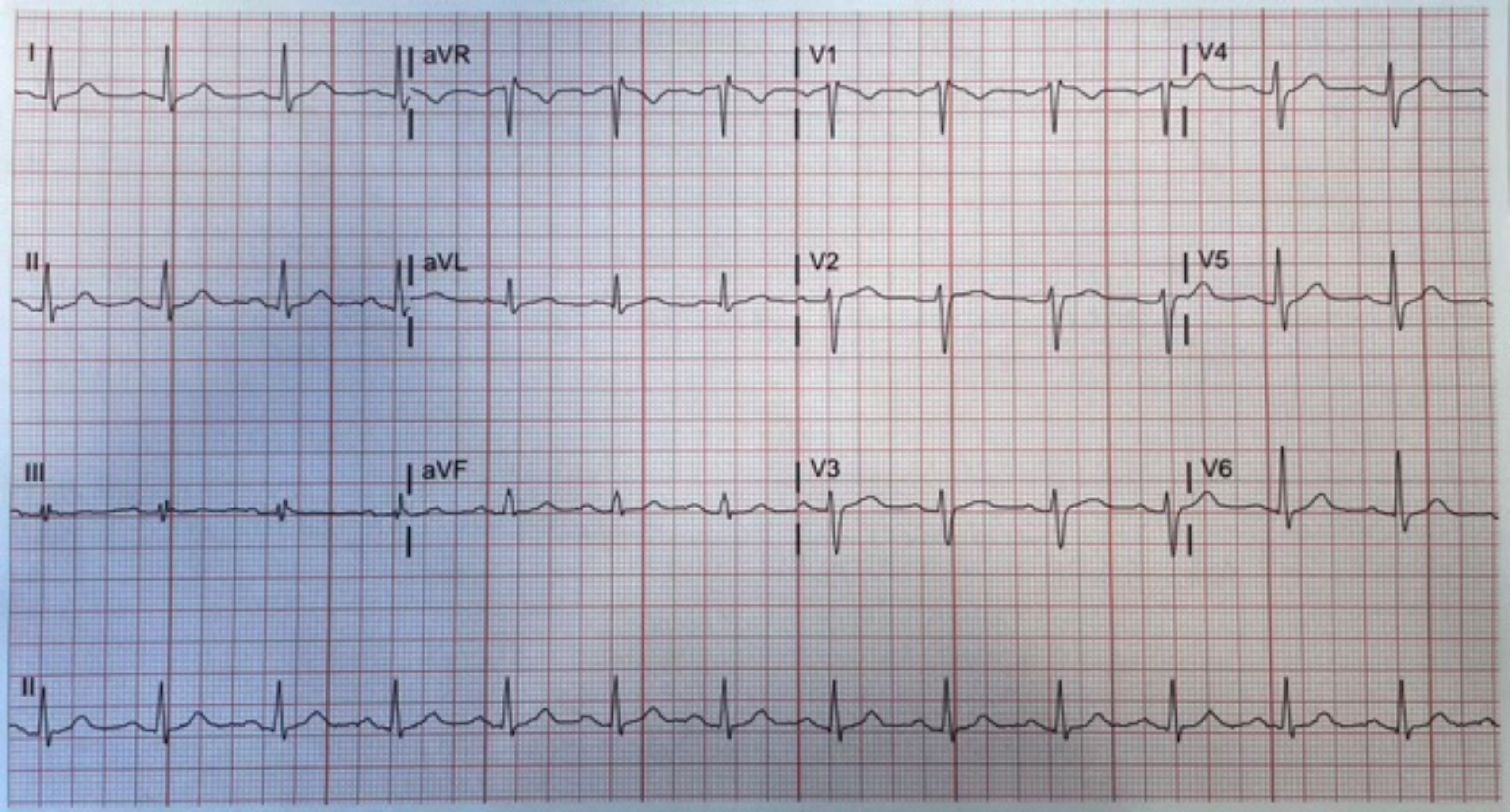
I
aVR
V1
V4
II
aVL
V2
V5
III
aVF
V3
V6
II

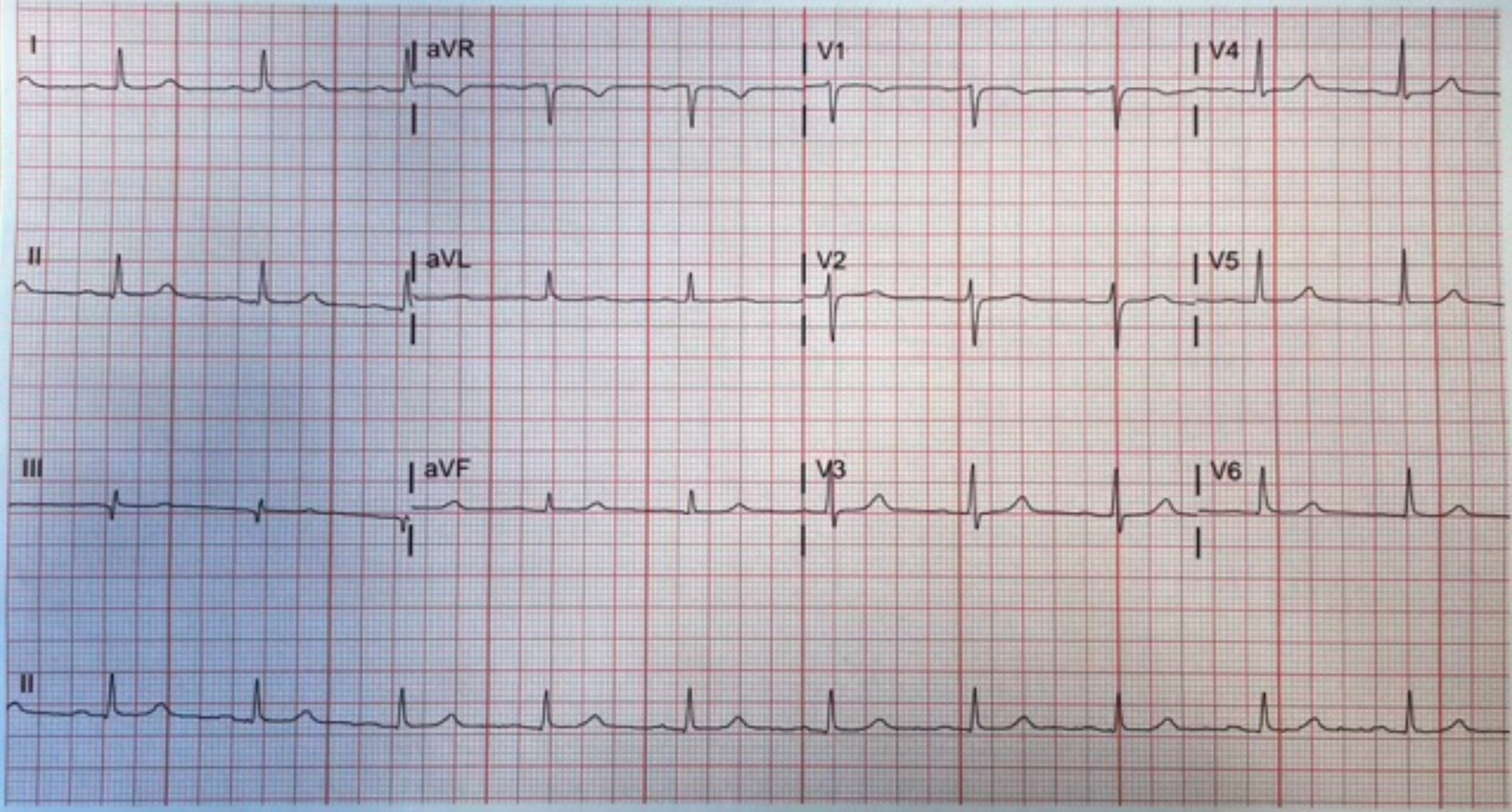
I
aVR
V1
V4
II
aVL
V2
V5
III
aVF
V3
V6
II

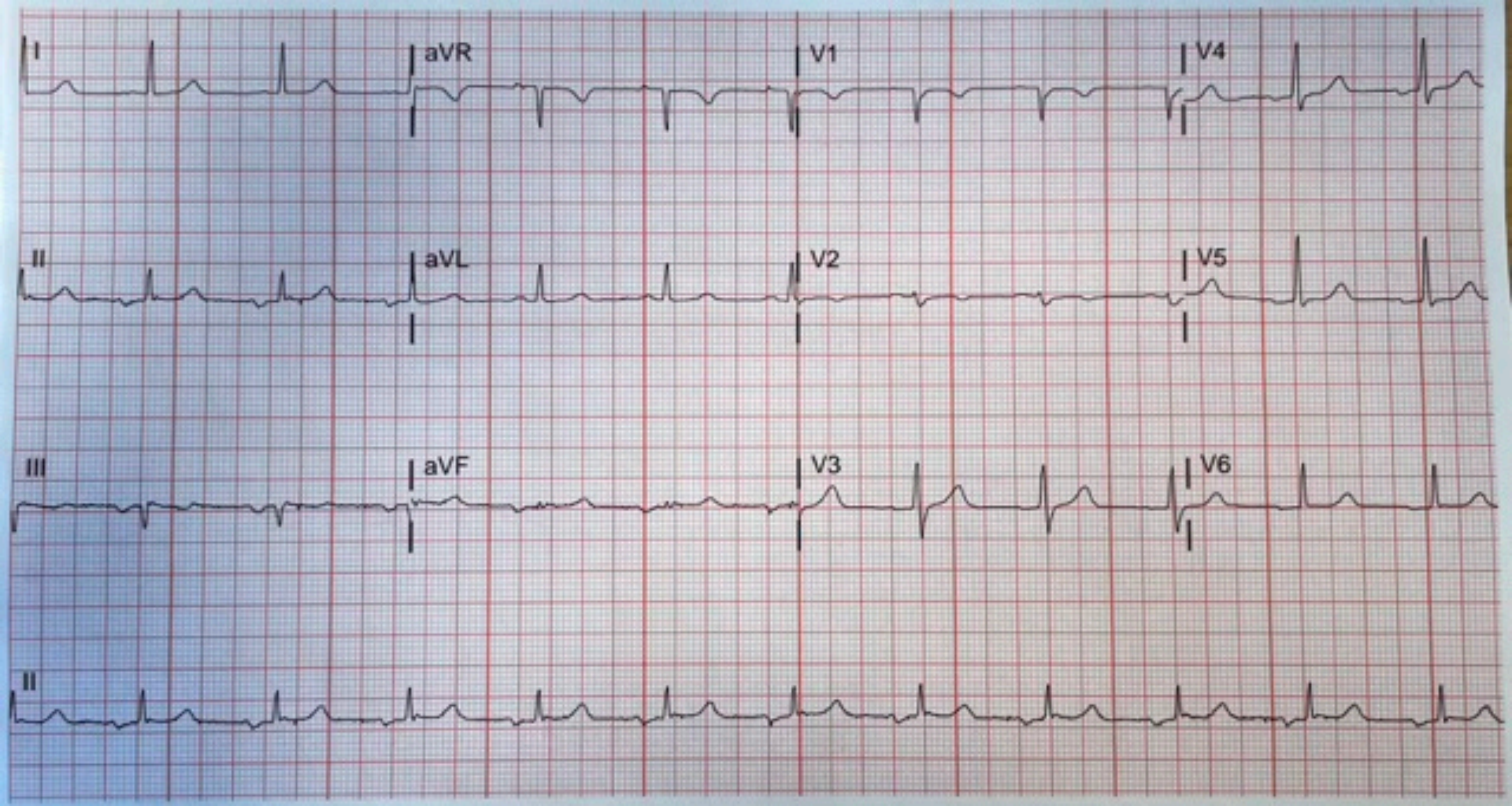
I
aVR
V1
V4
II
aVL
V2
V5
III
aVF
V3
V6
II

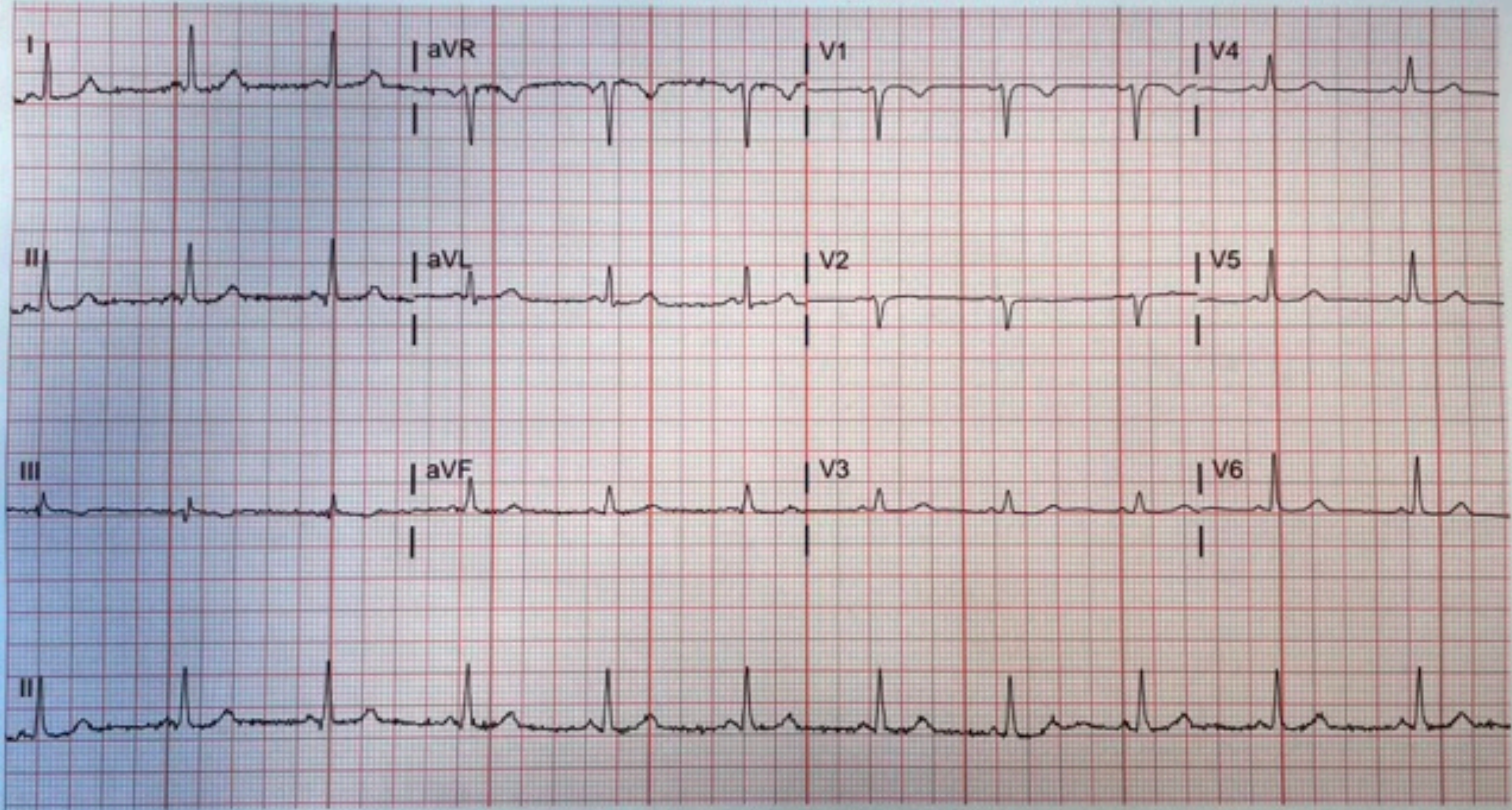
I
aVR
V1
V4
II
aVL
V2
V5
III
aVF
V3
V6
II

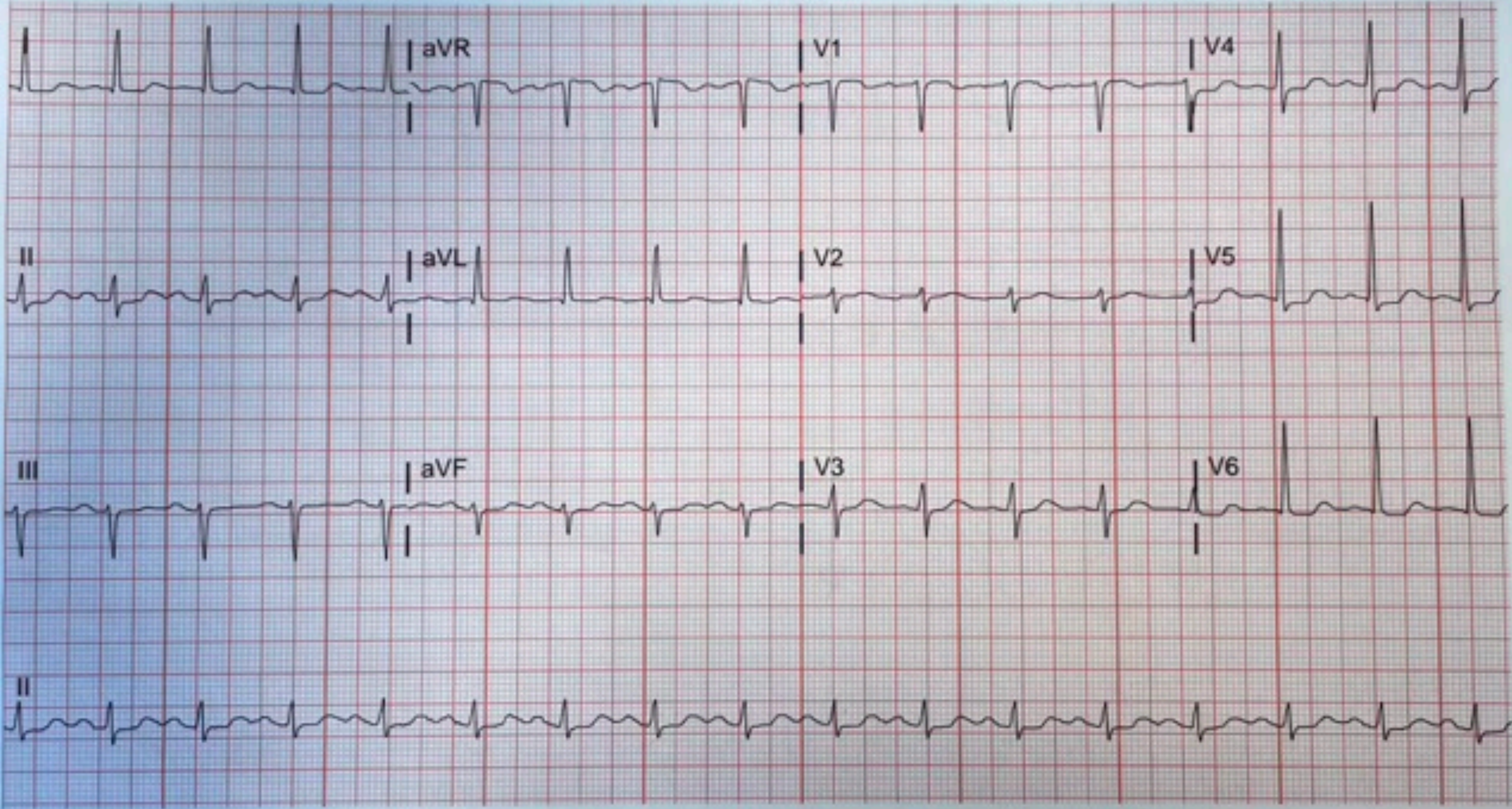
aVR
V1
V4
II
aVL
V2
V5
III
aVF
V3
V6
II

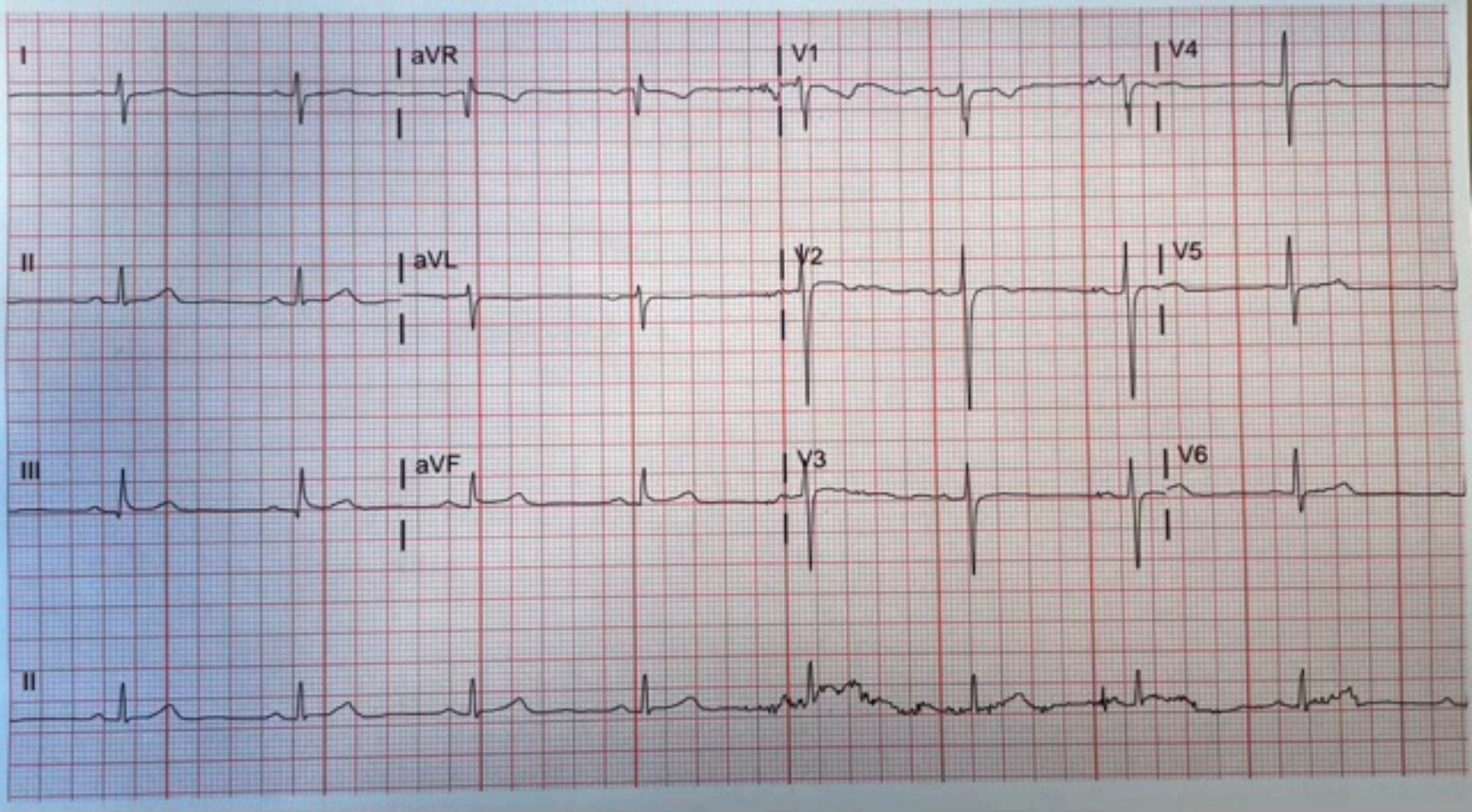
I
aVR
V1
V4
II
aVL
V2
V5
III
aVF
V3
V6
II

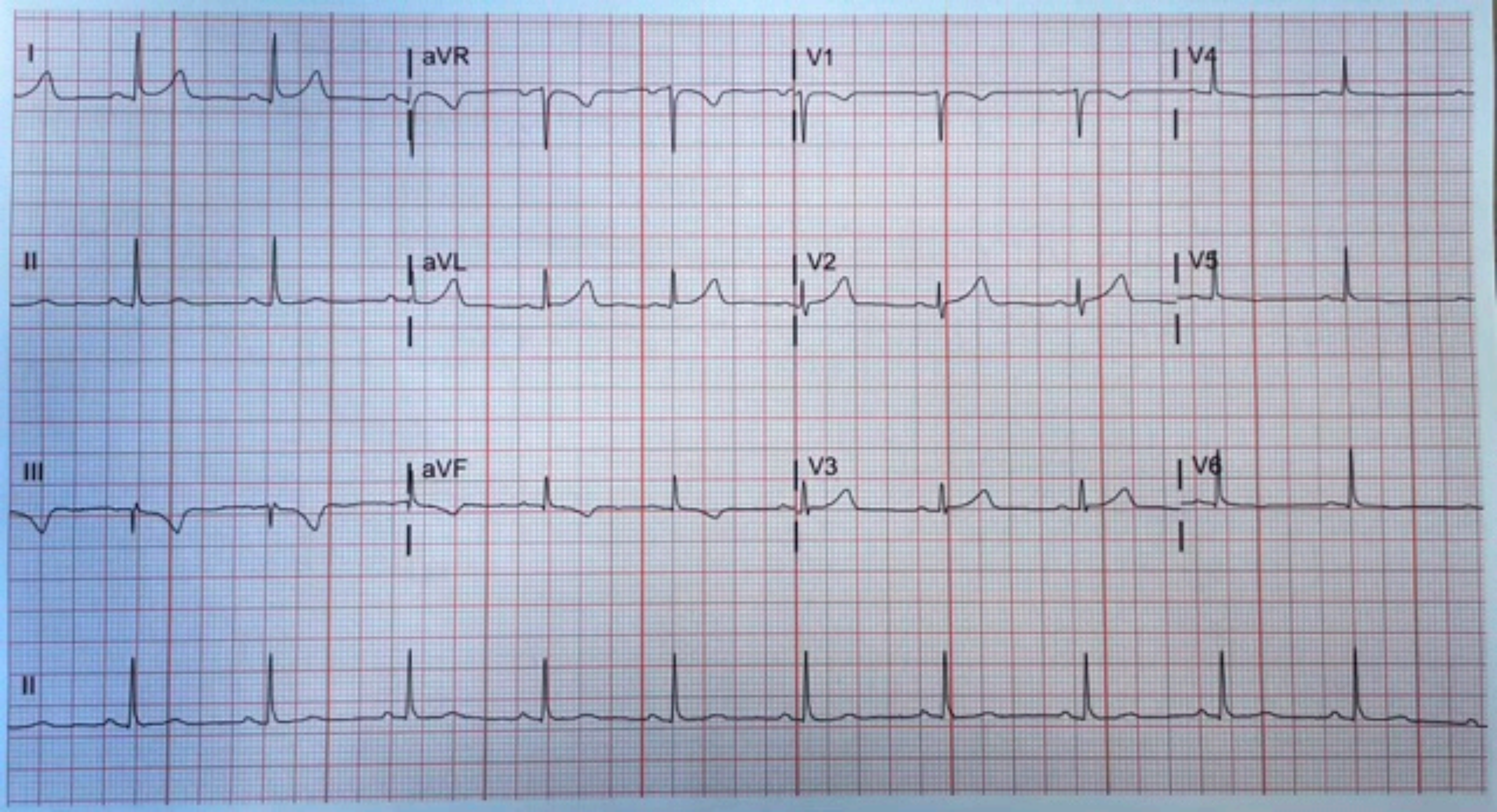
I
aVR
V1
V4
II
aVL
V2
V5
III
aVF
V3
V6
II

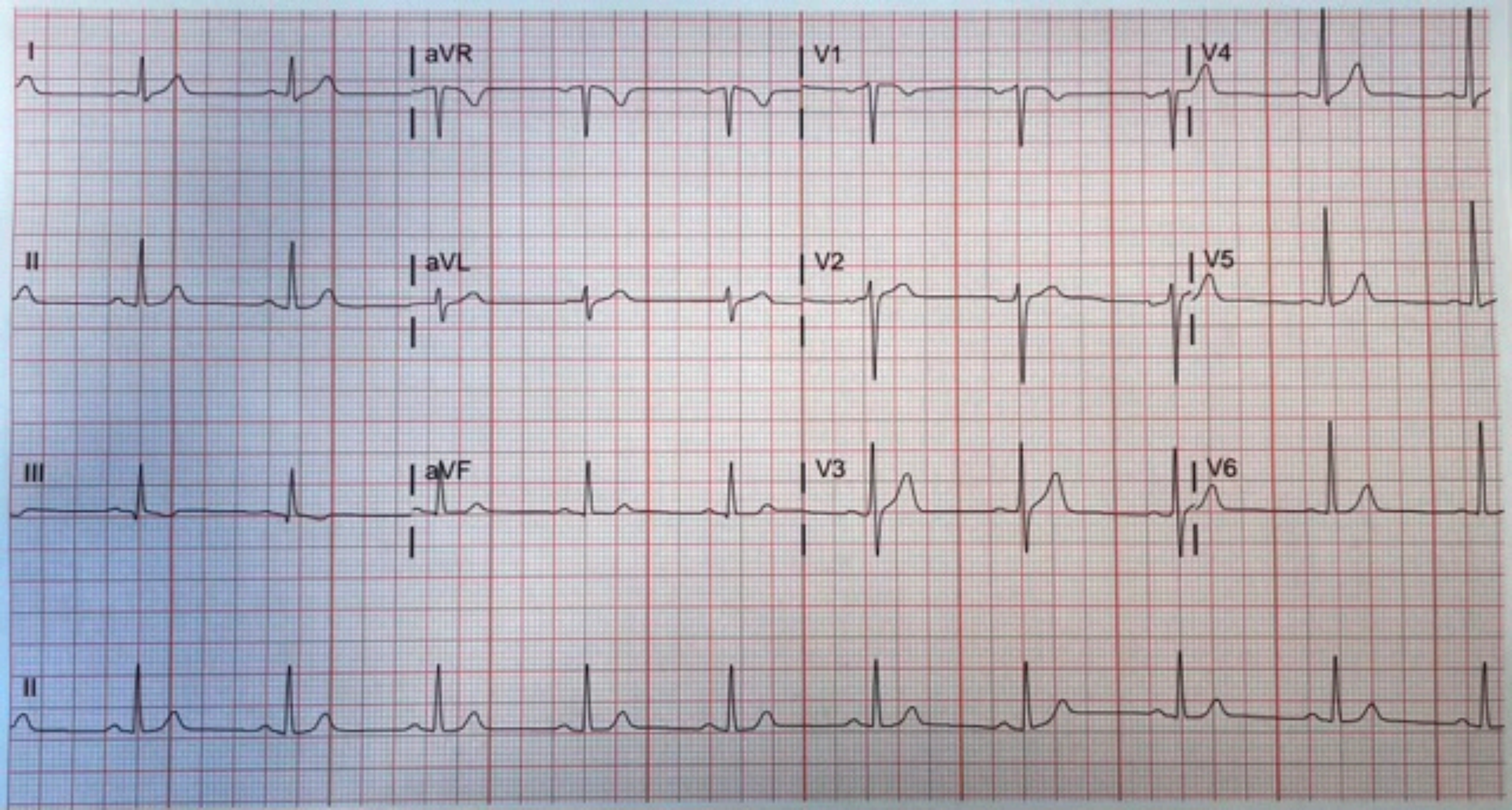
I
aVR
V1
V4
II
aVL
V2
V5
III
aVF
V3
V6
II

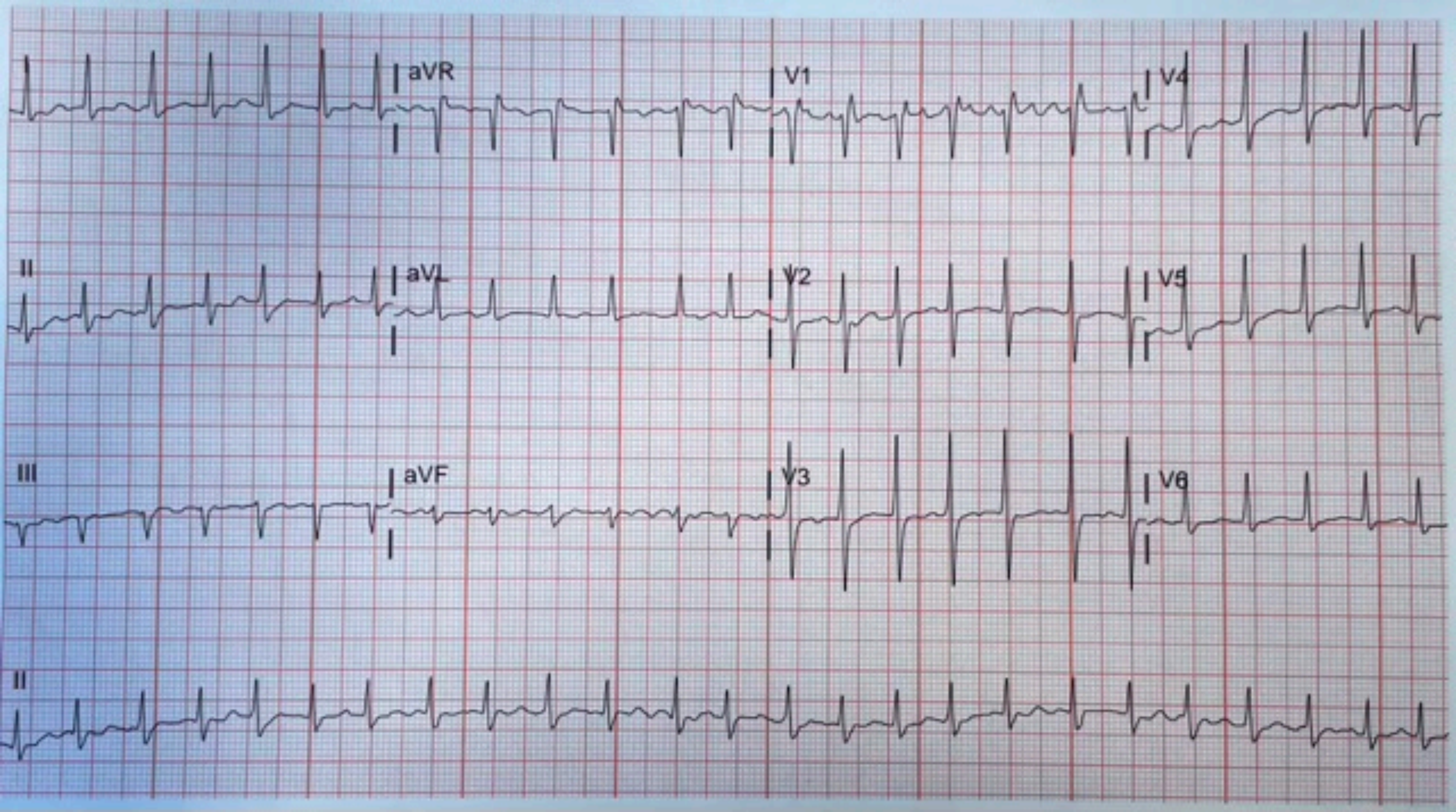
aVR
V1
V4
II
aVL
V2
V5
III
aVF
V3
V6
II

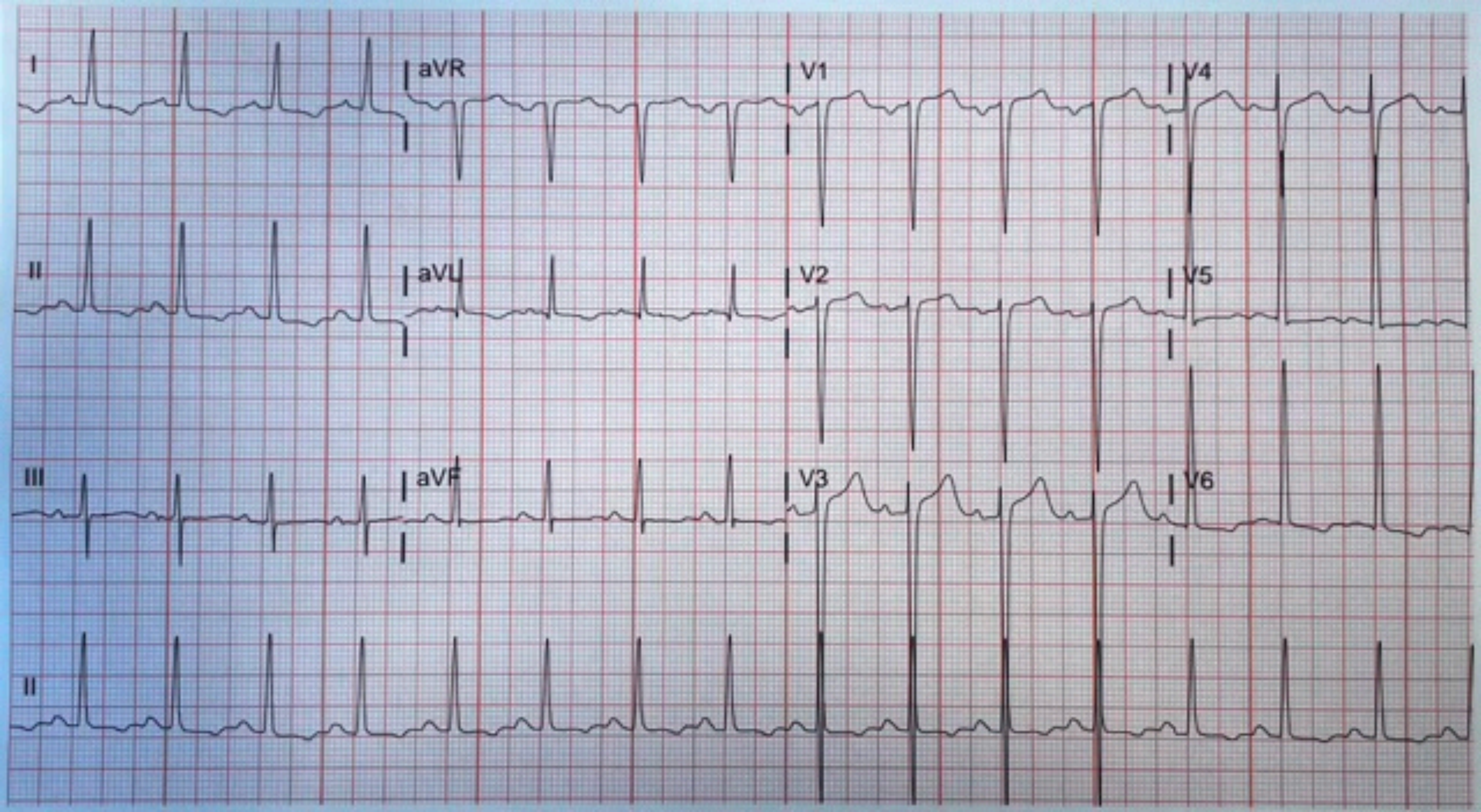
I
aVR
V1
V4
II
aVL
V2
V5
III
aVF
V3
V6
II

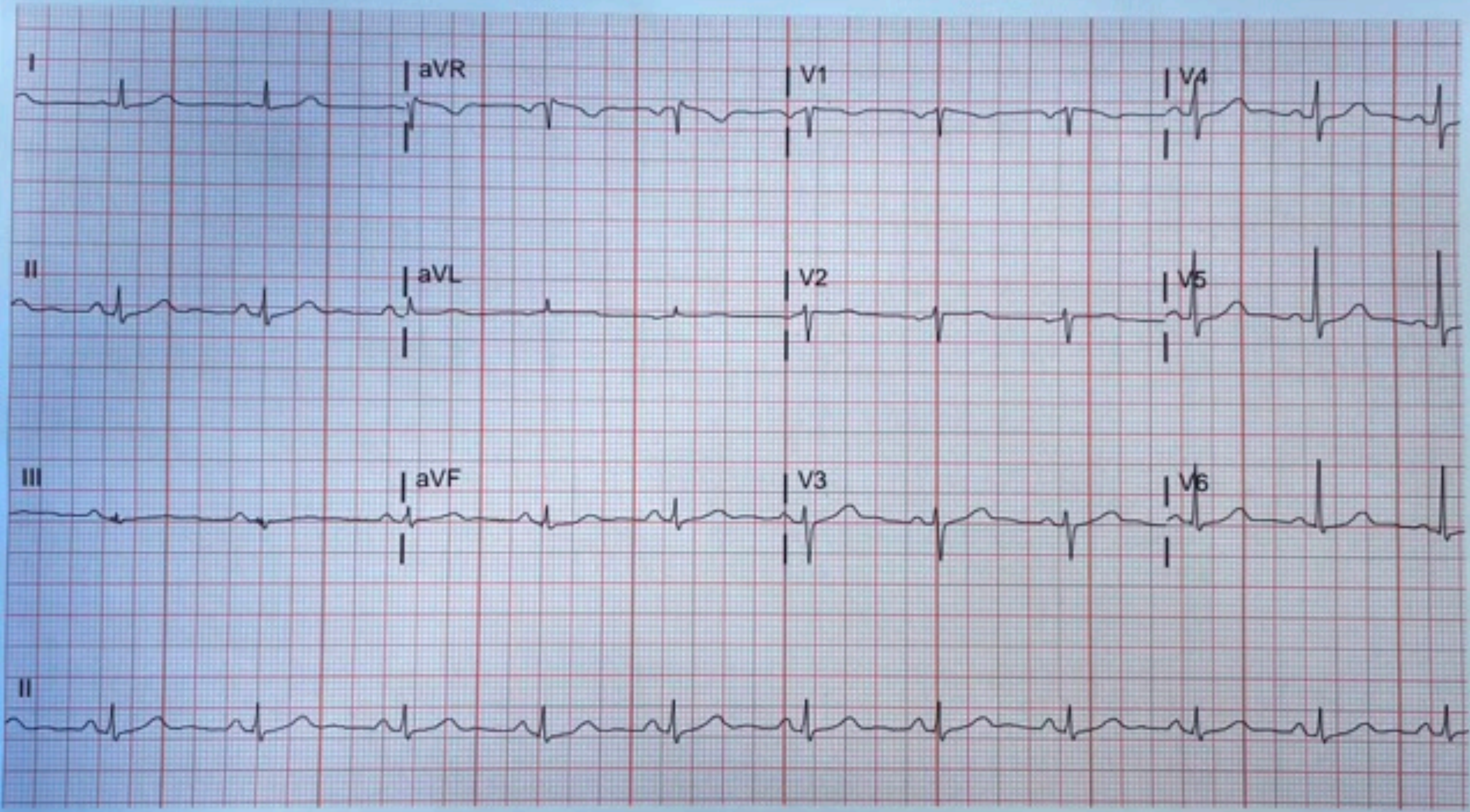
I
aVR
V1
V4
II
aVL
V2
V5
III
aVF
V3
V6
II

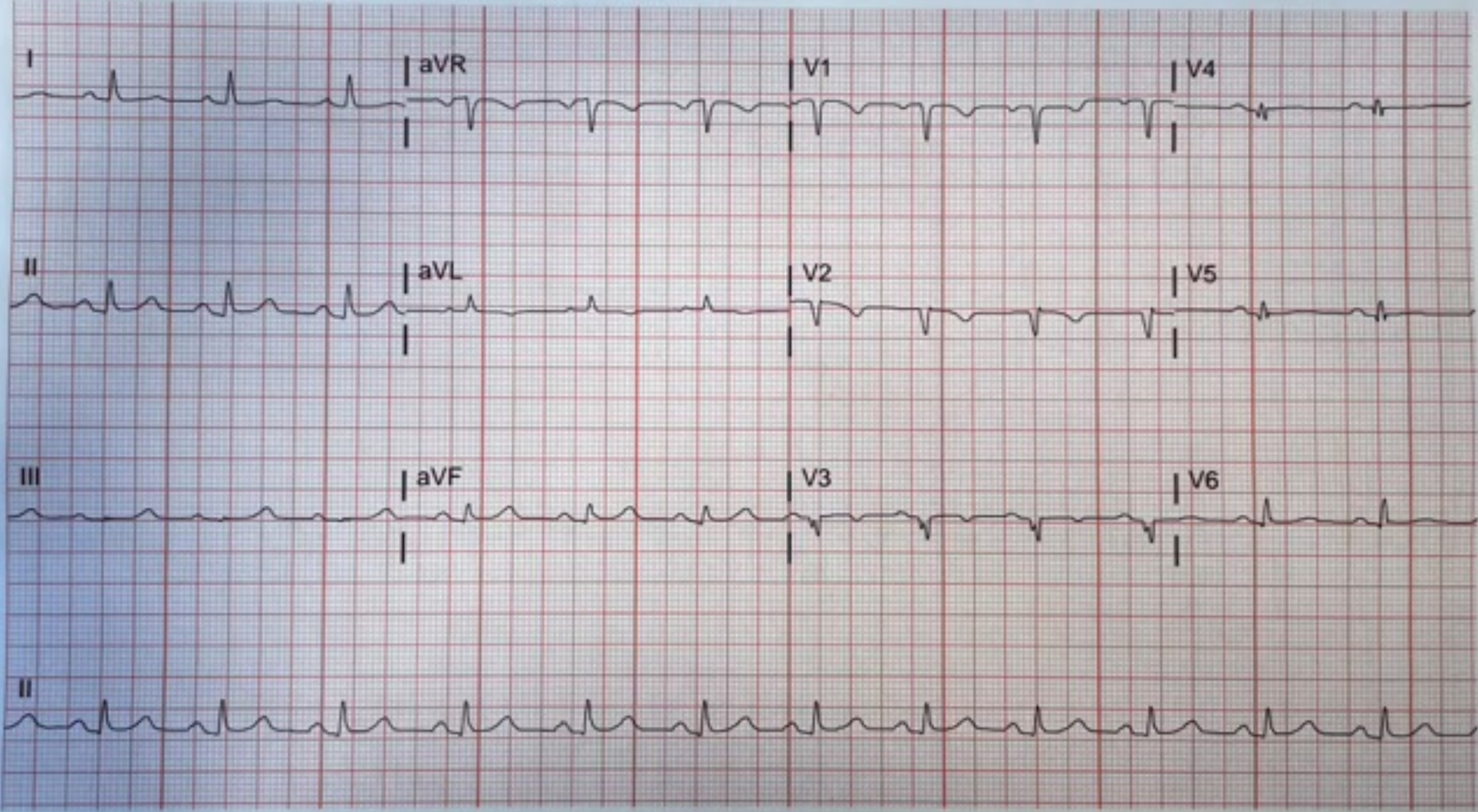
I
aVR
V1
V4
II
aVL
V2
V5
III
aVF
V3
V6
II

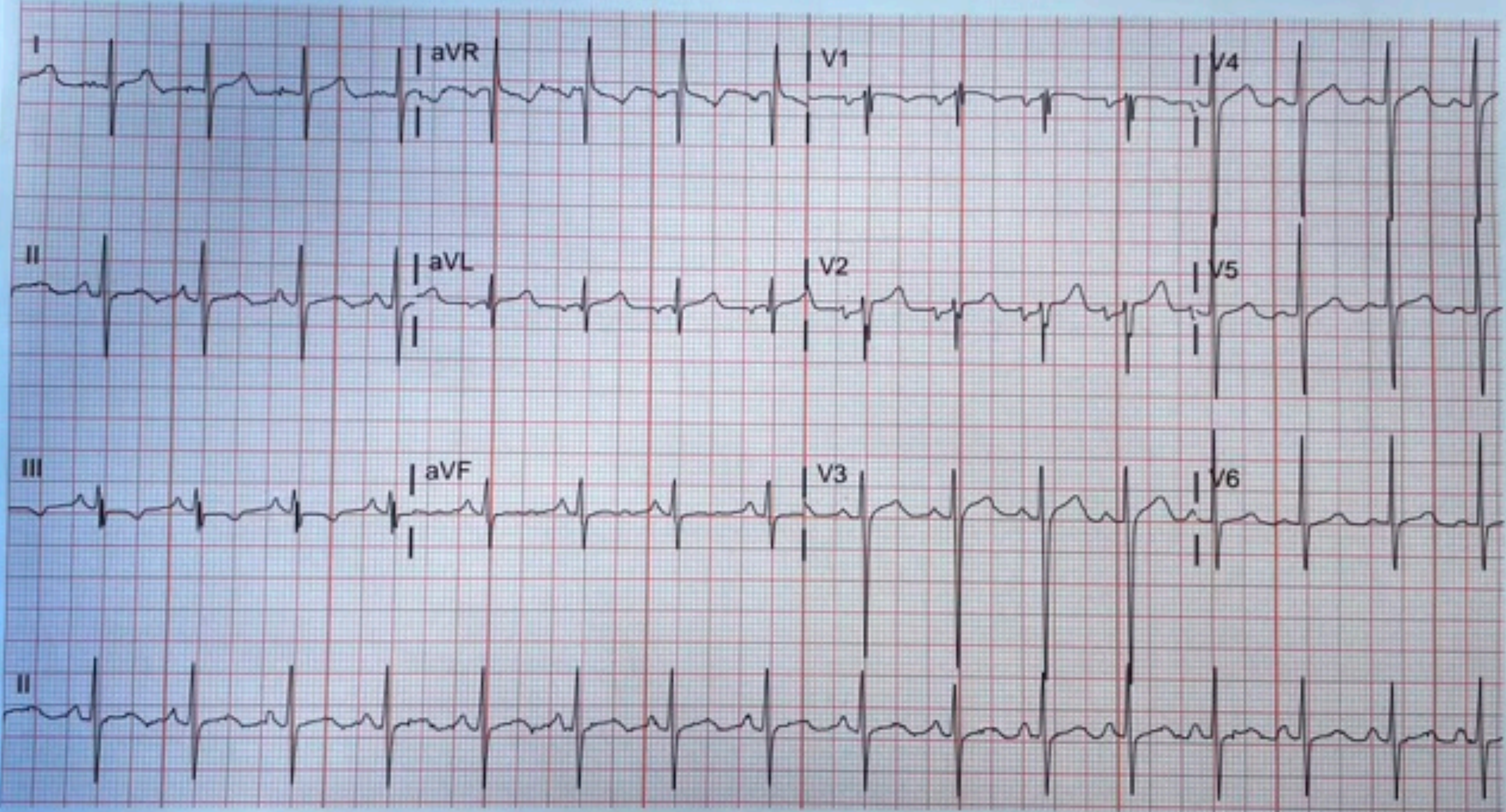
I
aVR
V1
V4
II
aVL
V2
V5
III
aVF
V3
V6
II

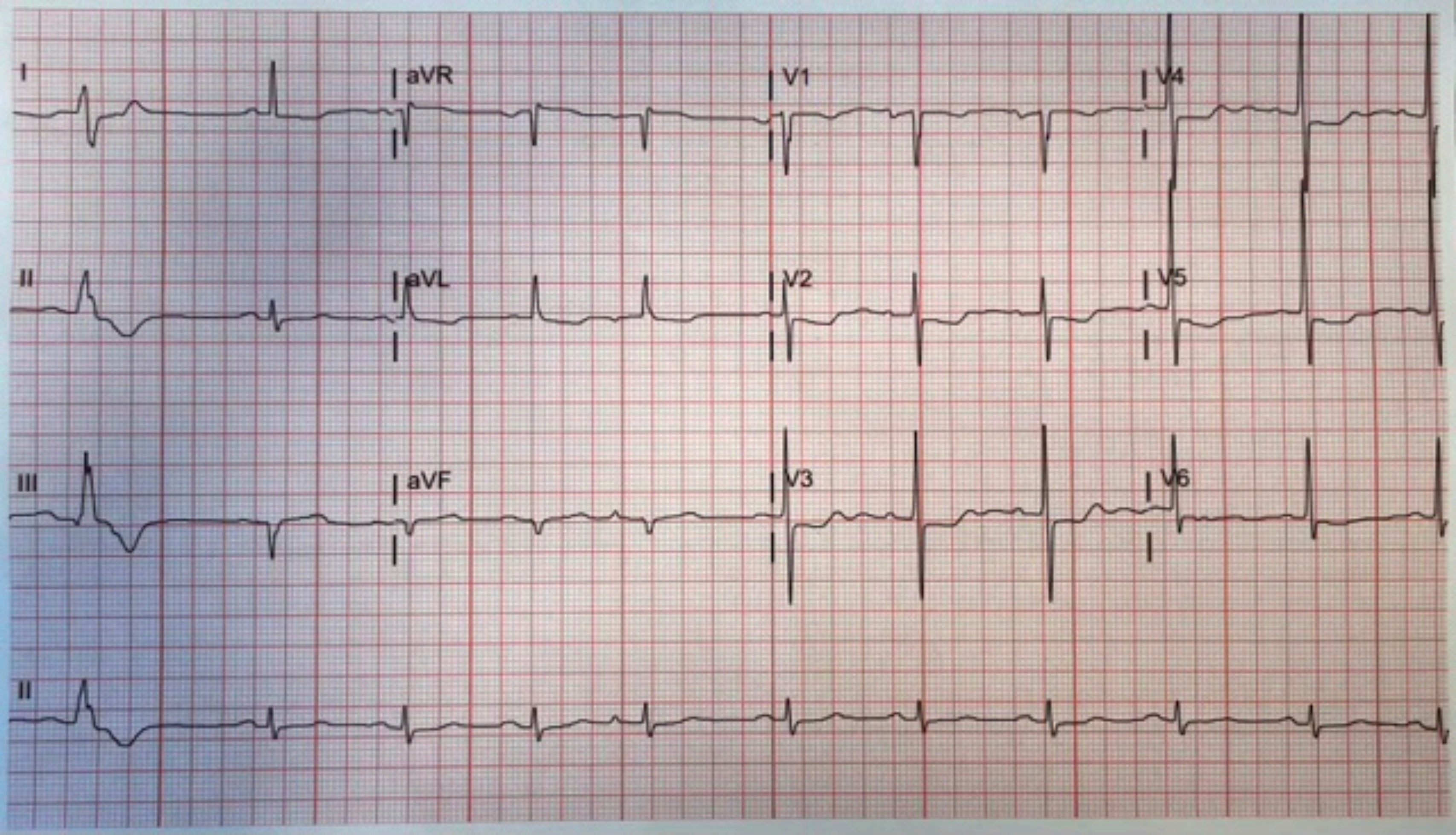
I
aVR
V1
V4
II
aVL
V2
V5
III
aVF
V3
V6
II

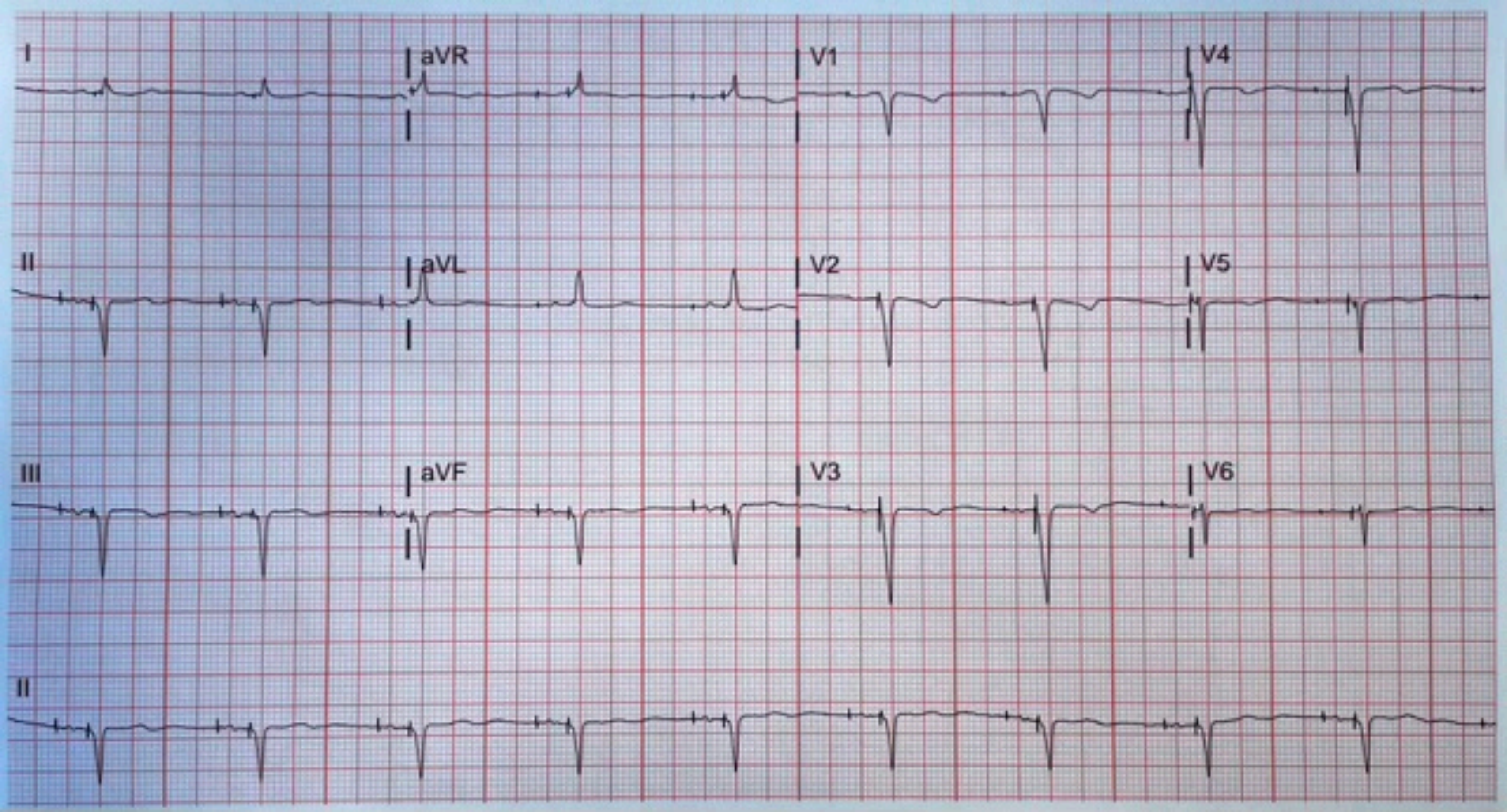
I
aVR
V1
V4
II
aVL
V2
V5
III
aVF
V3
V6
II

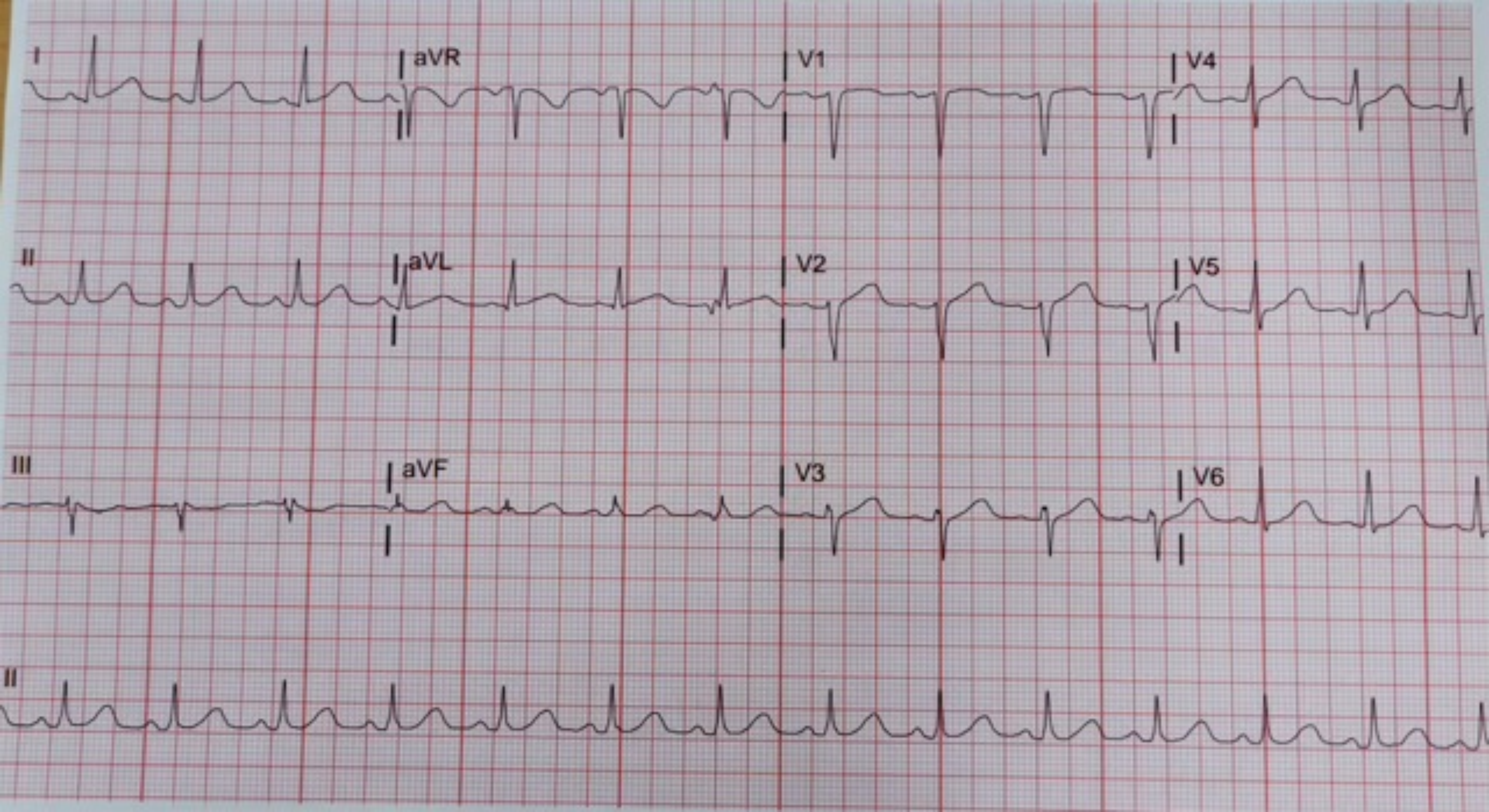
I
aVR
V1
V4
II
aVL
V2
V5
III
aVF
V3
V6
II

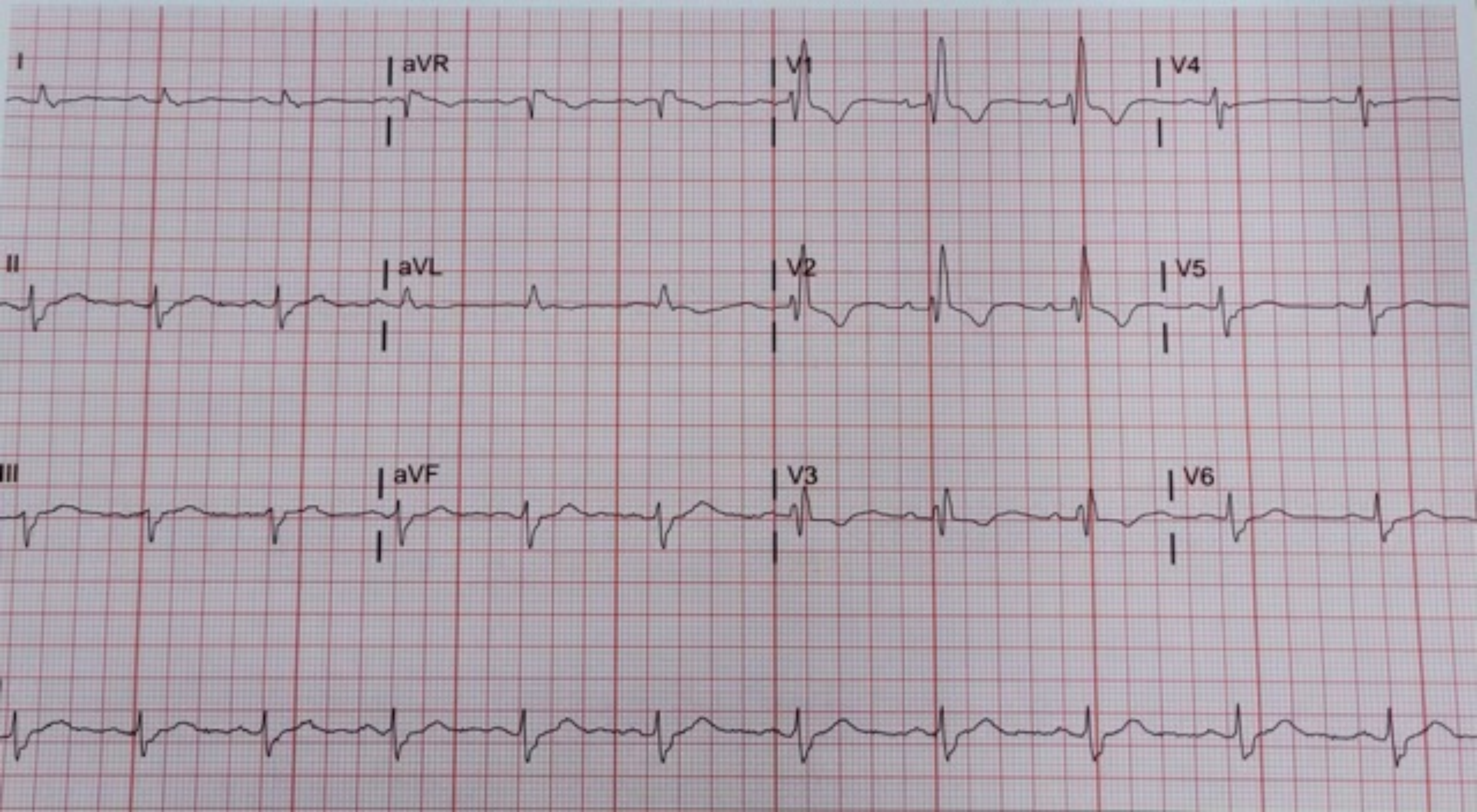

I
aVR
V1
V4
II
aVL
V2
V5
III
aVF
V3
V6

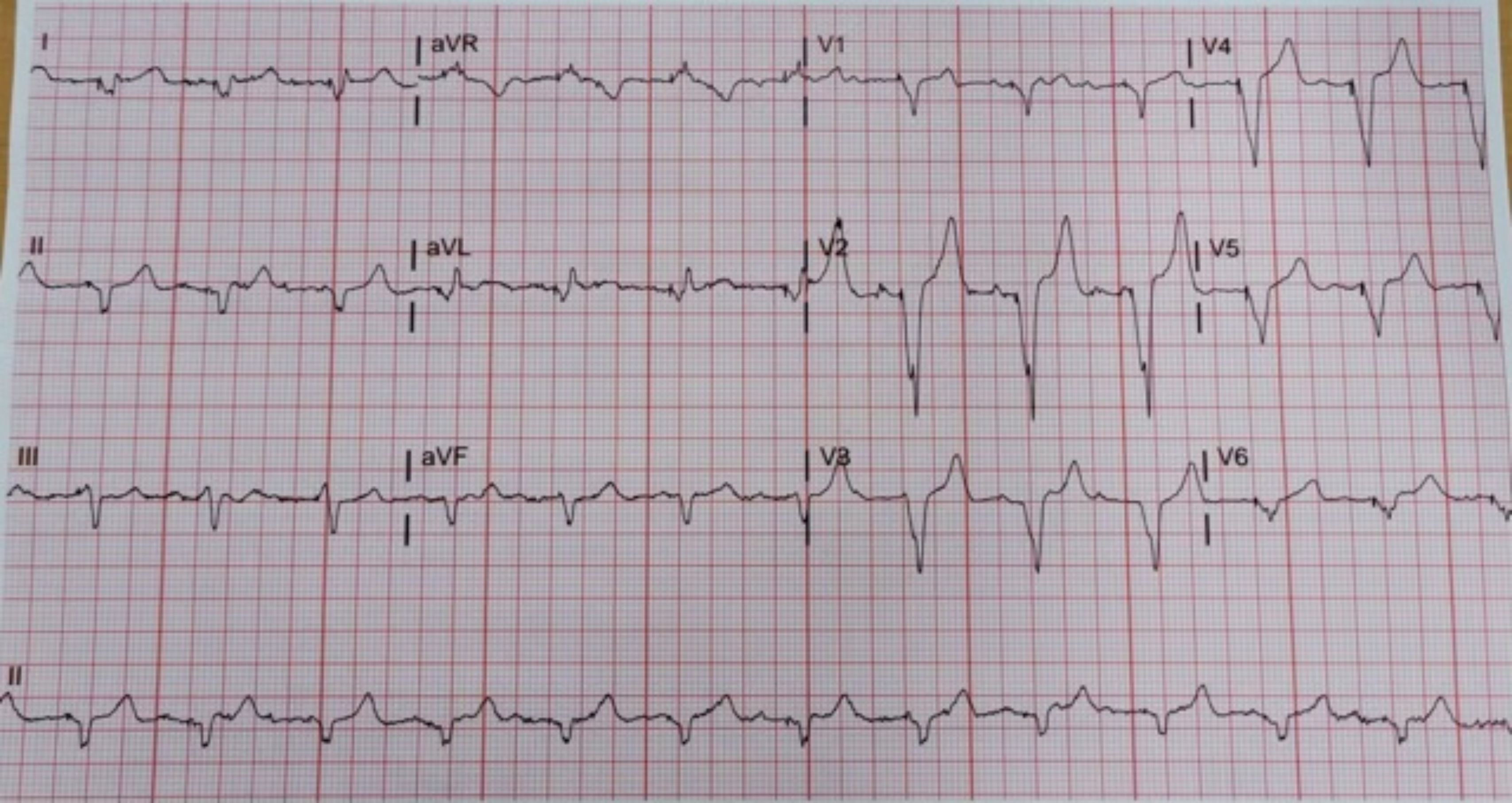
I
aVR
V1
V4
II
aVL
V2
V5
III
aVF
V3
V6
II

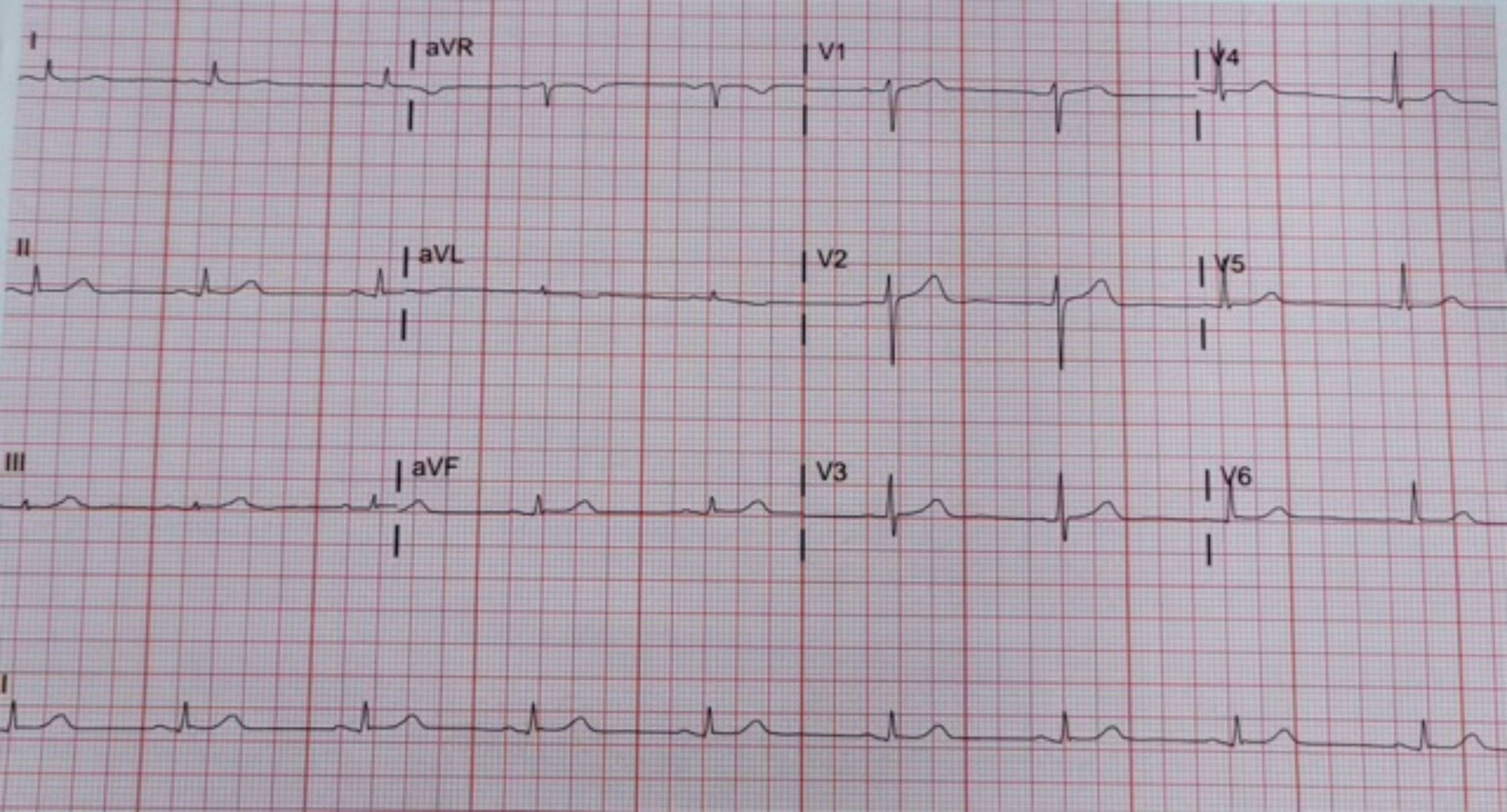

I
aVR
V1
V4
II
aVL
V2
V5
III
aVF
V3
V6
II

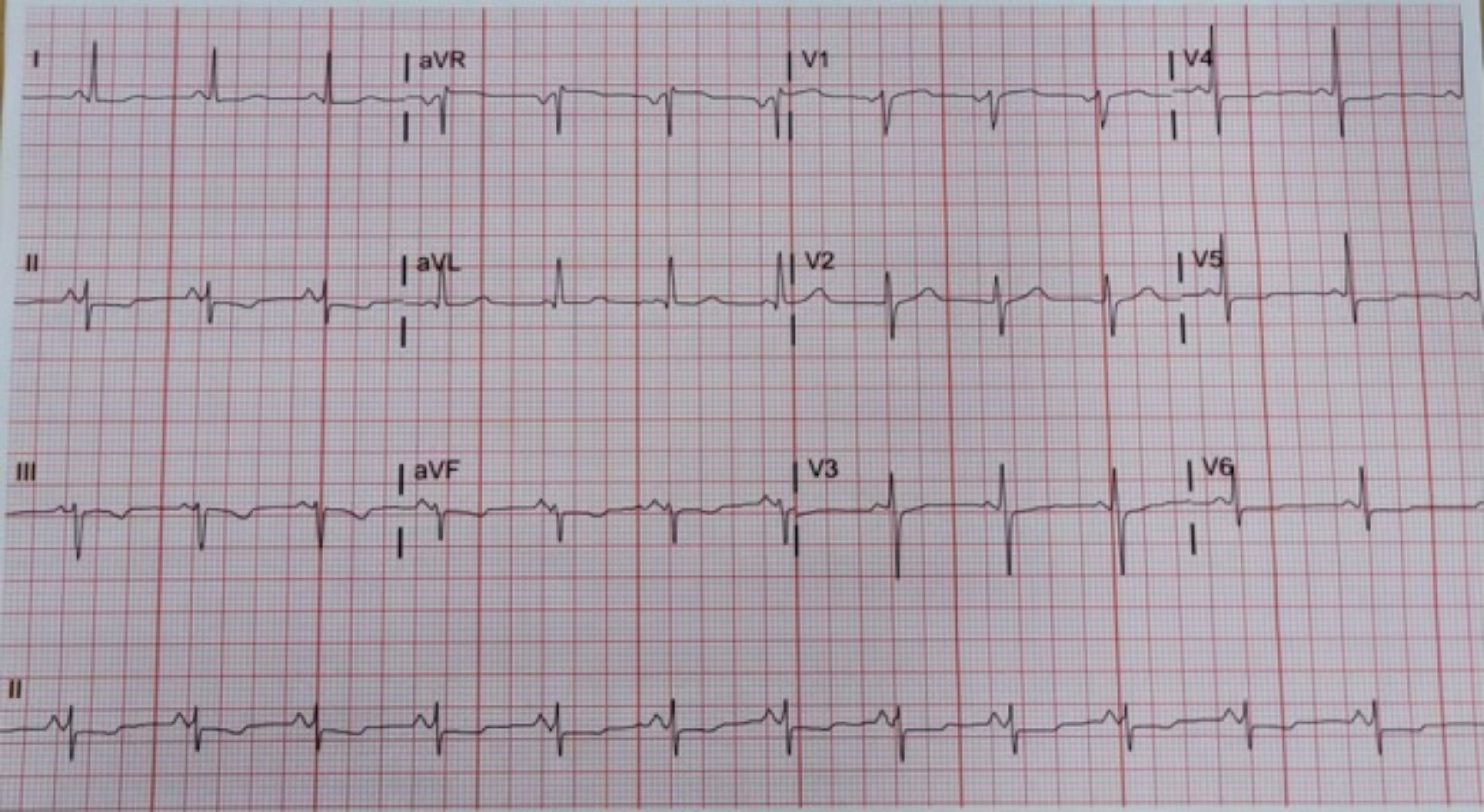
I
aVR
V1
V4
II
aVL
V2
V5
III
aVF
V3
V6
II

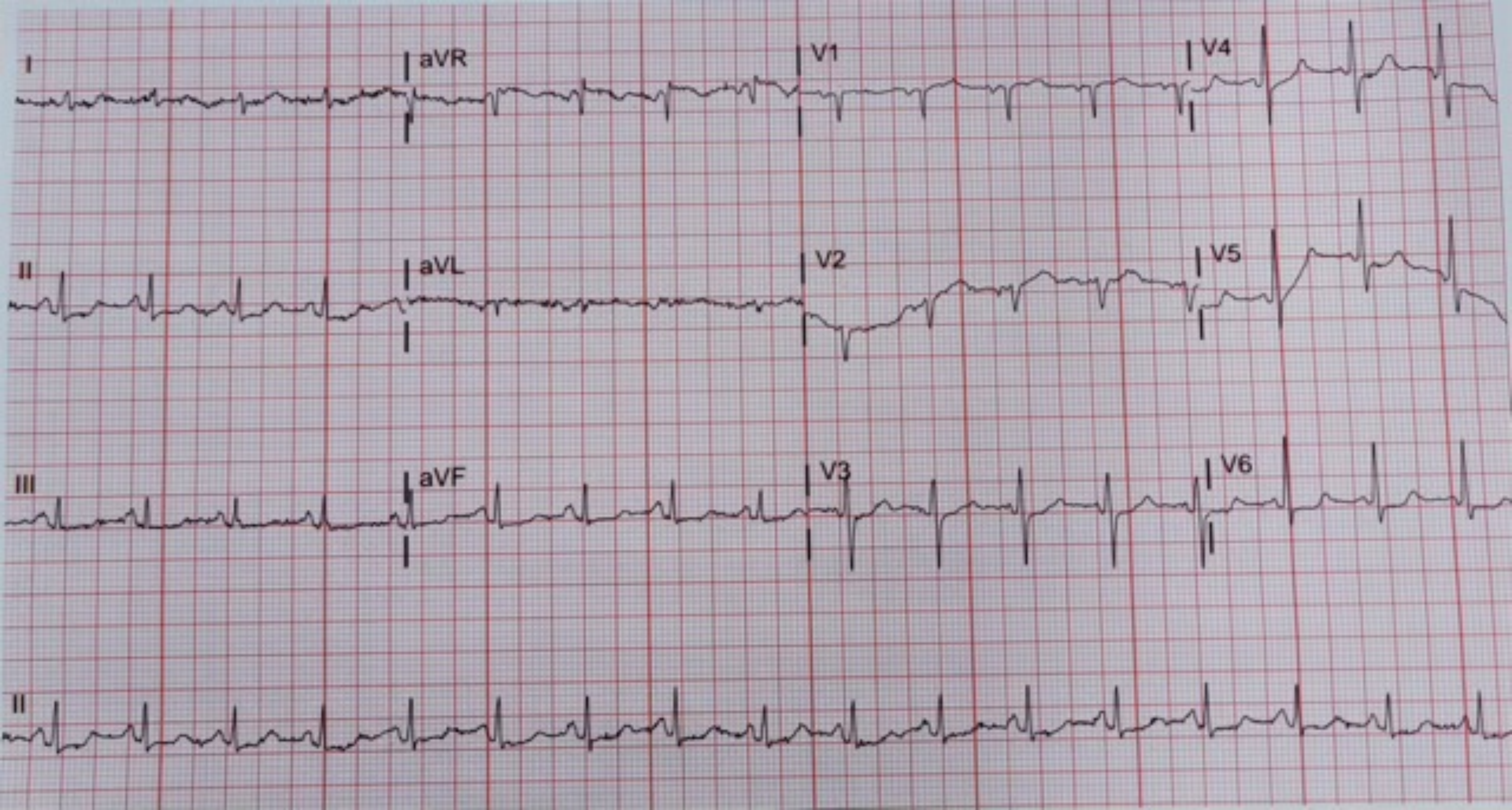
I
aVR
V1
V4
II
aVL
V2
V5
III
aVF
V3
V6
II

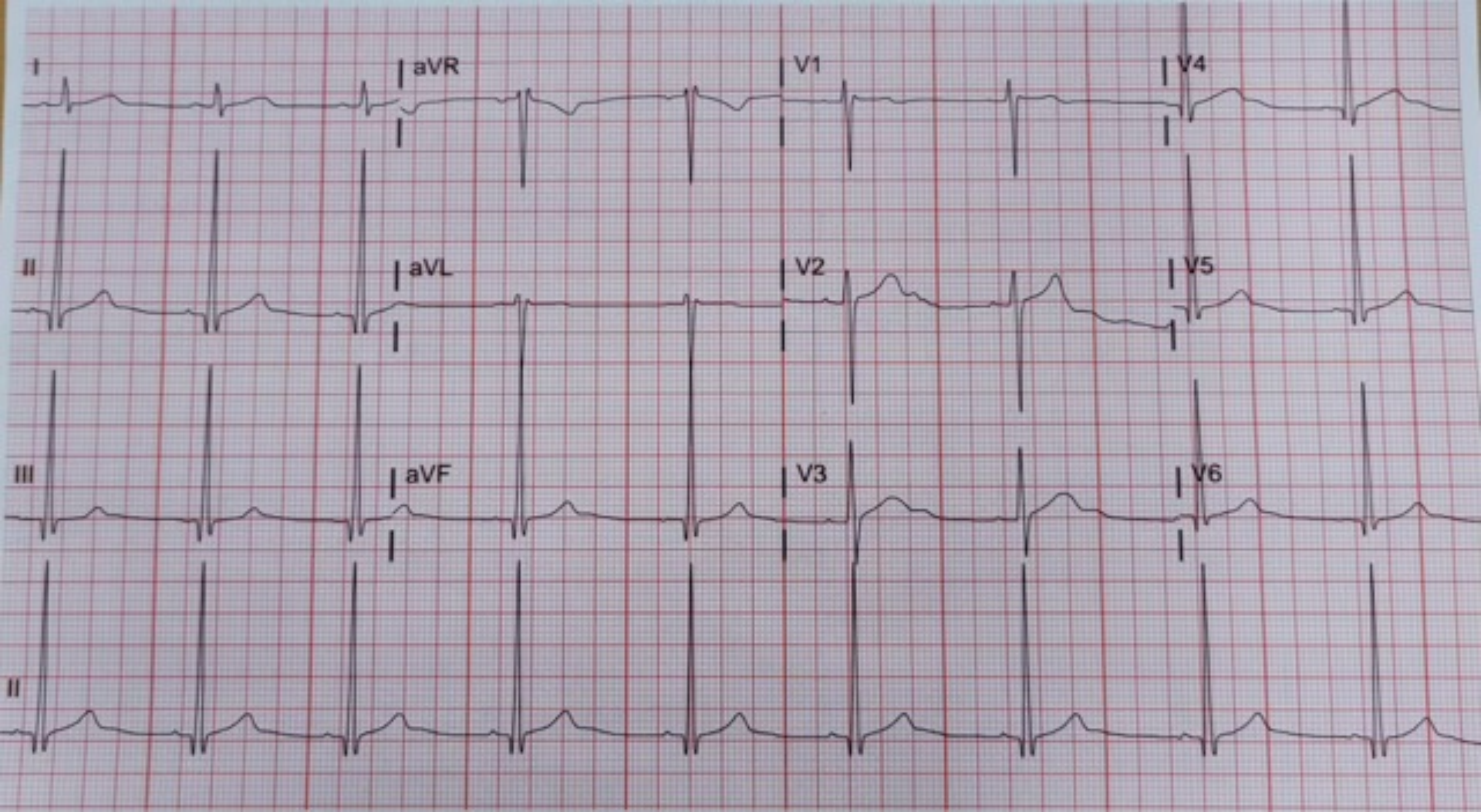
I
aVR
V1
V4
II
aVL
V2
V5
III
aVF
V3
V6
II

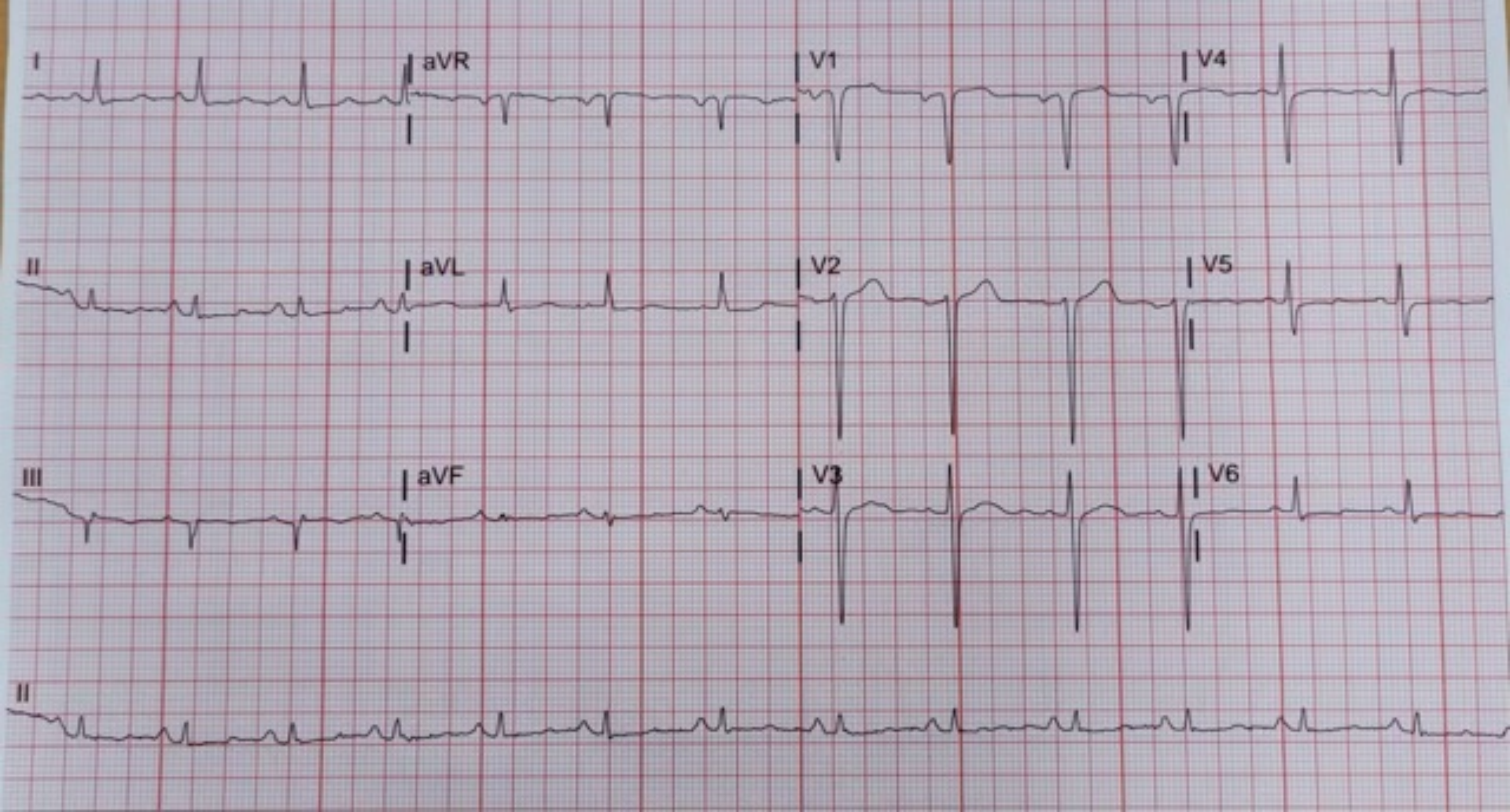
I
aVR
V1
V4
II
aVL
V2
V5
III
aVF
V3
V6
II

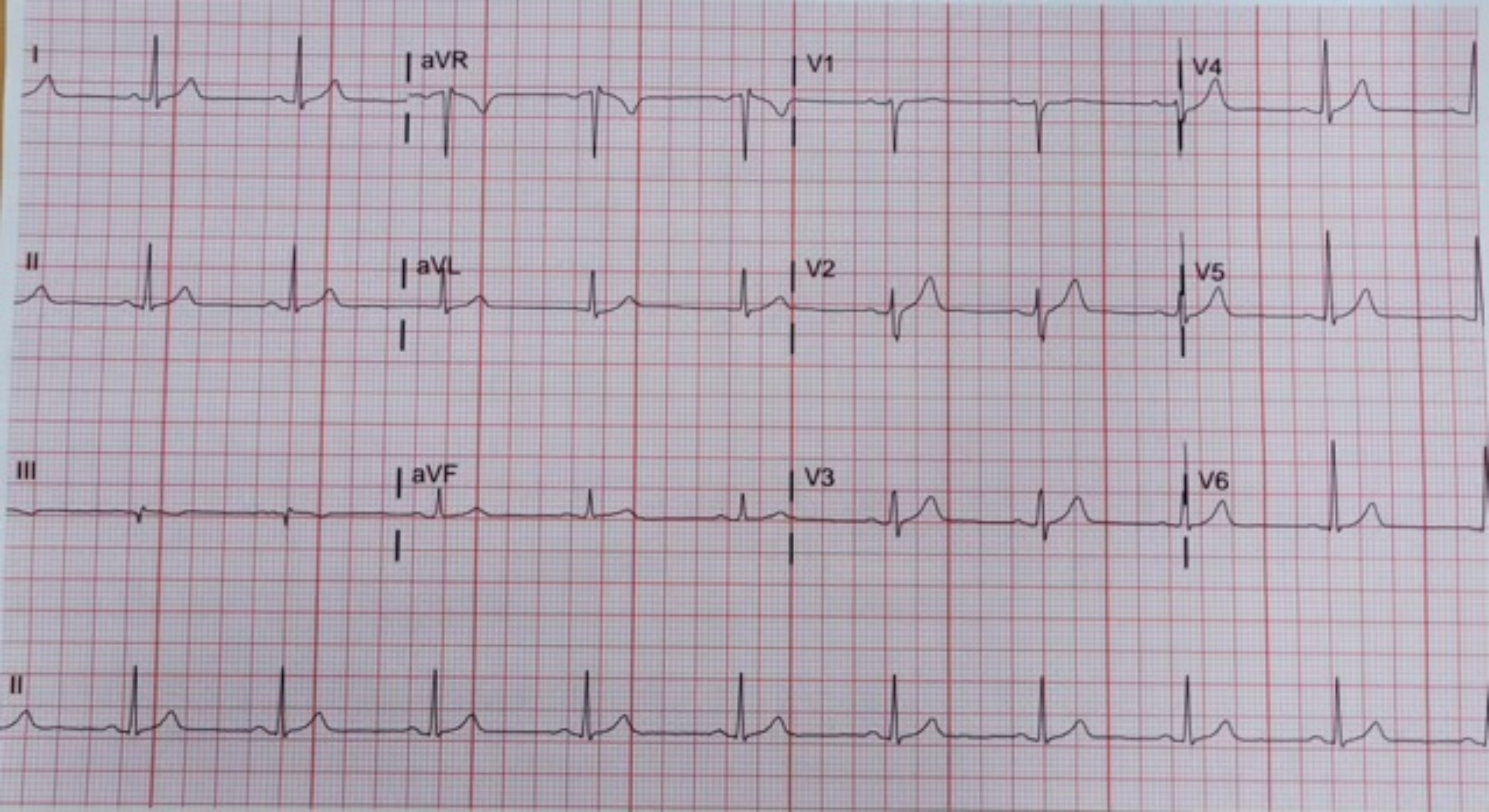
I
aVR
V1
V4
II
aVL
V2
V5
III
aVF
V3
V6
II

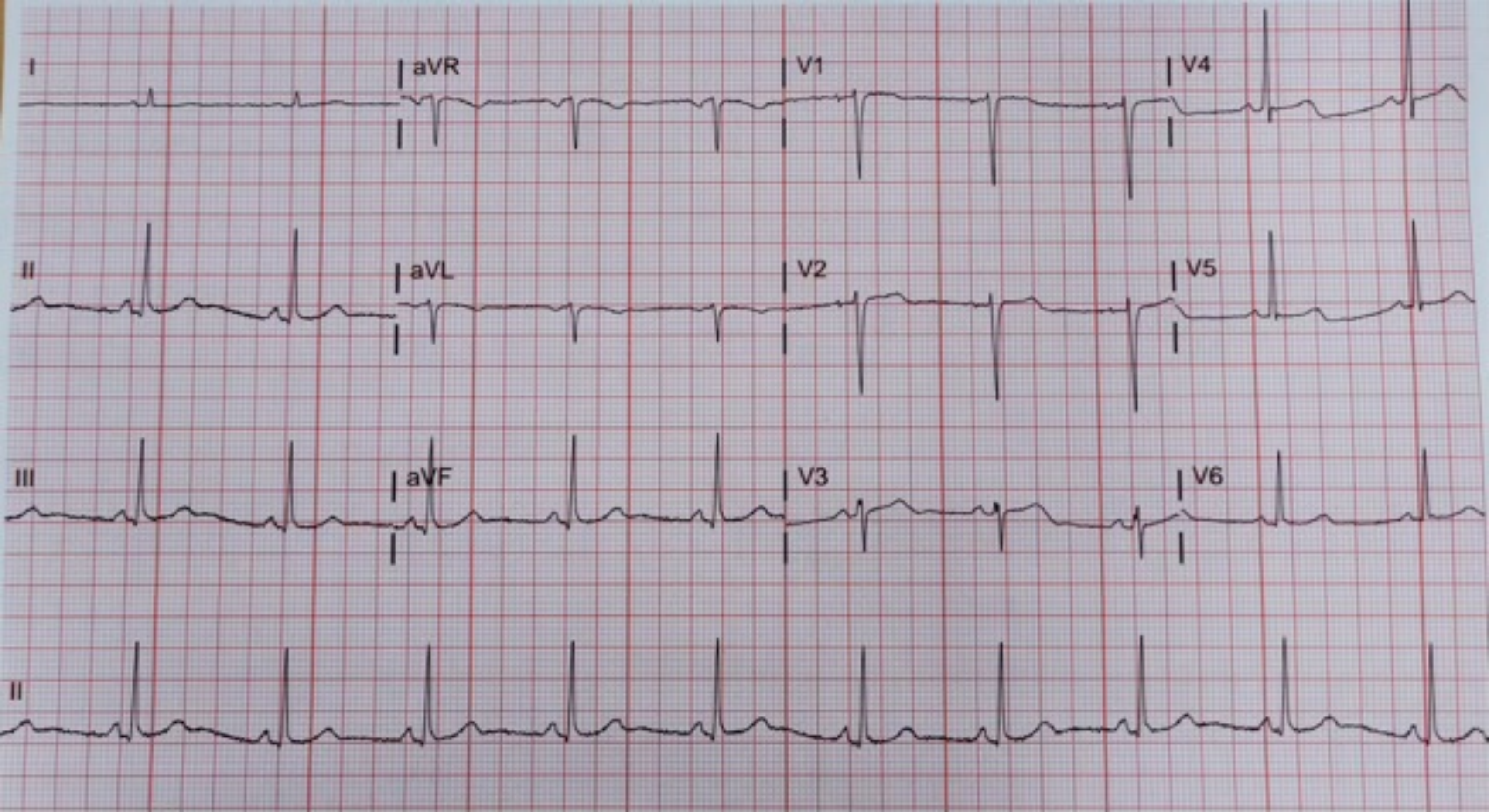
I
aVR
V1
V4
II
aVL
V2
V5
III
aVF
V3
V6
II

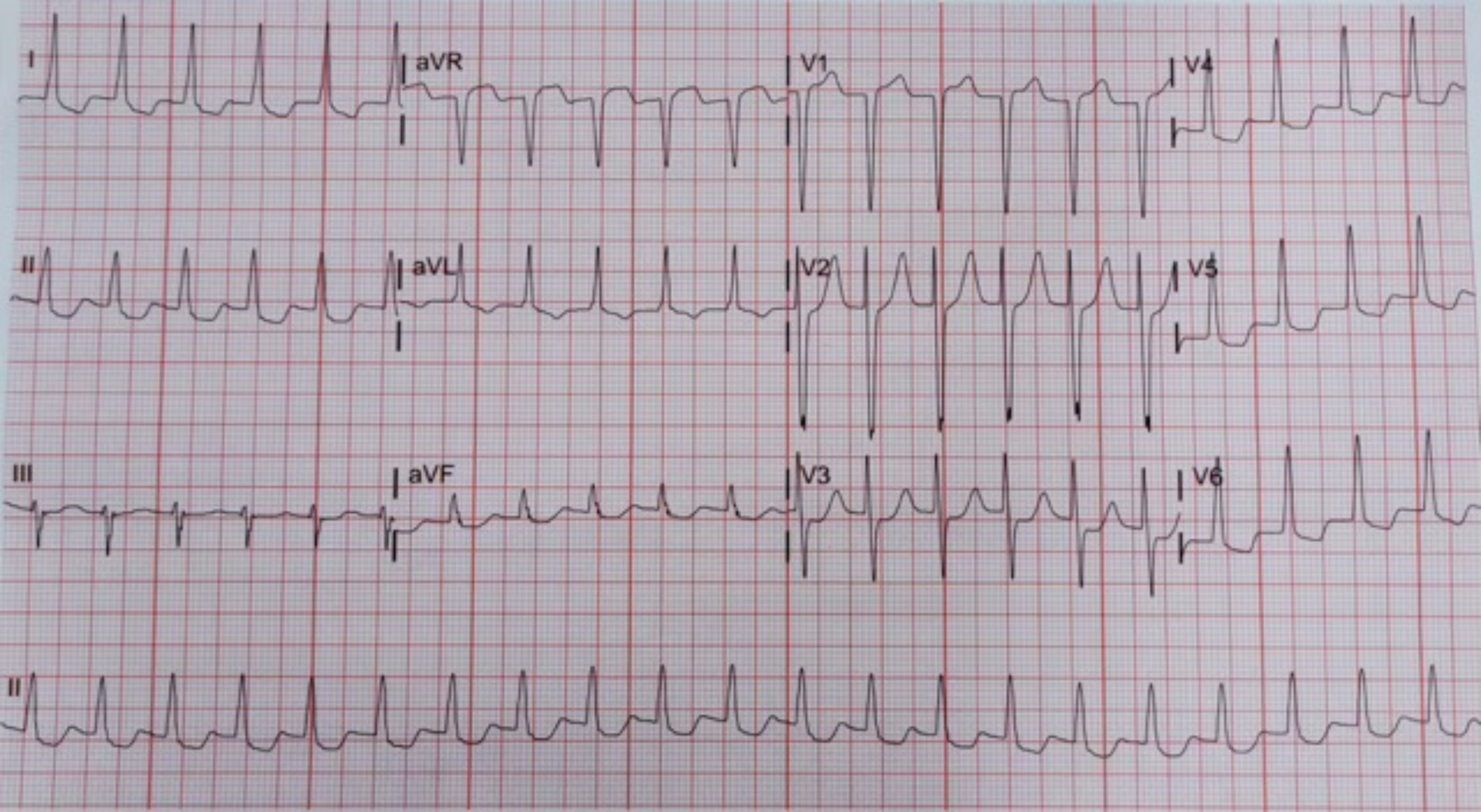
I
aVR
V1
V4
II
aVL
V2
V5
III
aVF
V3
V6
II

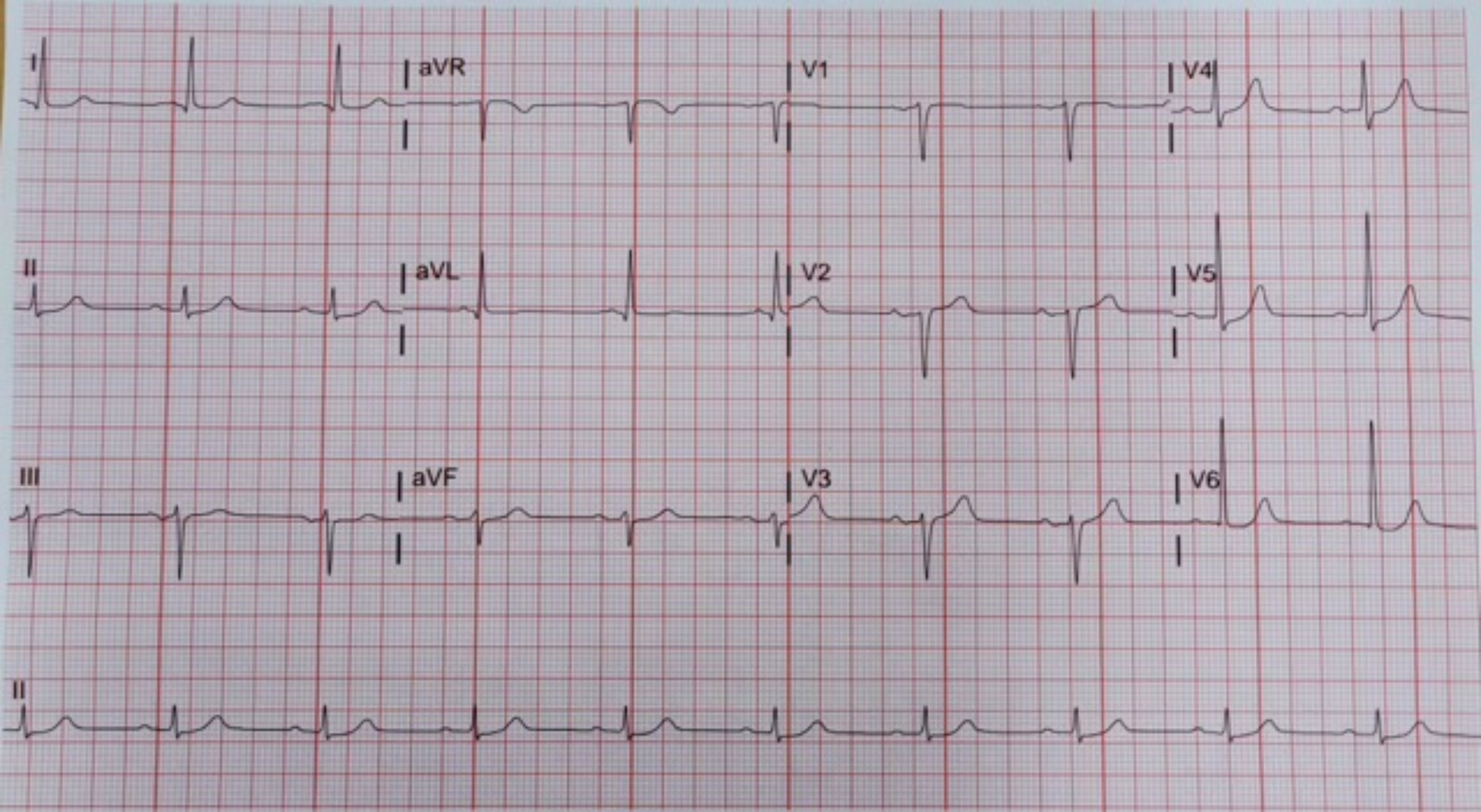
I
aVR
V1
V4
II
aVL
V2
V5
III
aVF
V3
V6
II

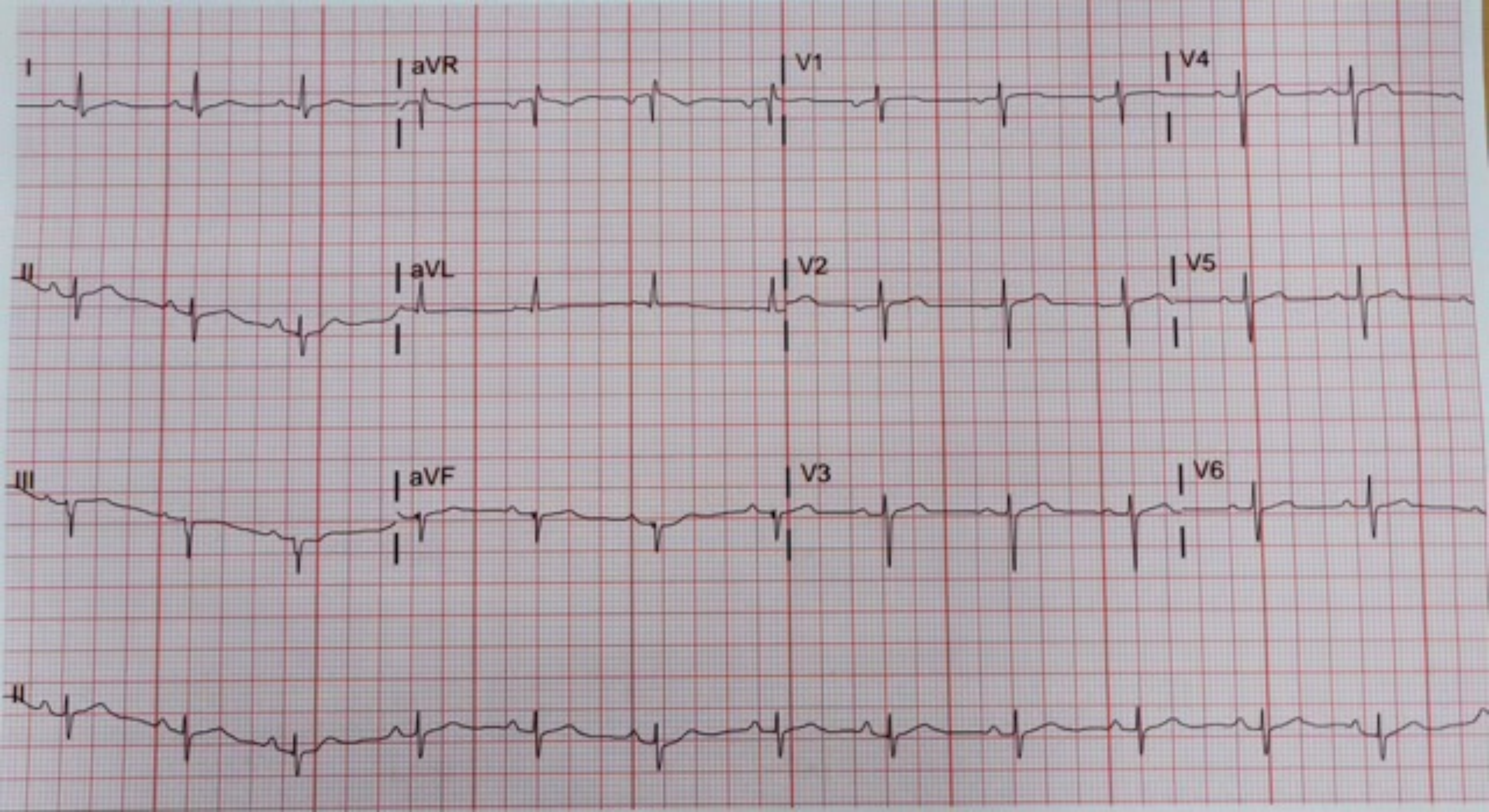
I
aVR
V1
V4
II
aVL
V2
V5
III
aVF
V3
V6
II

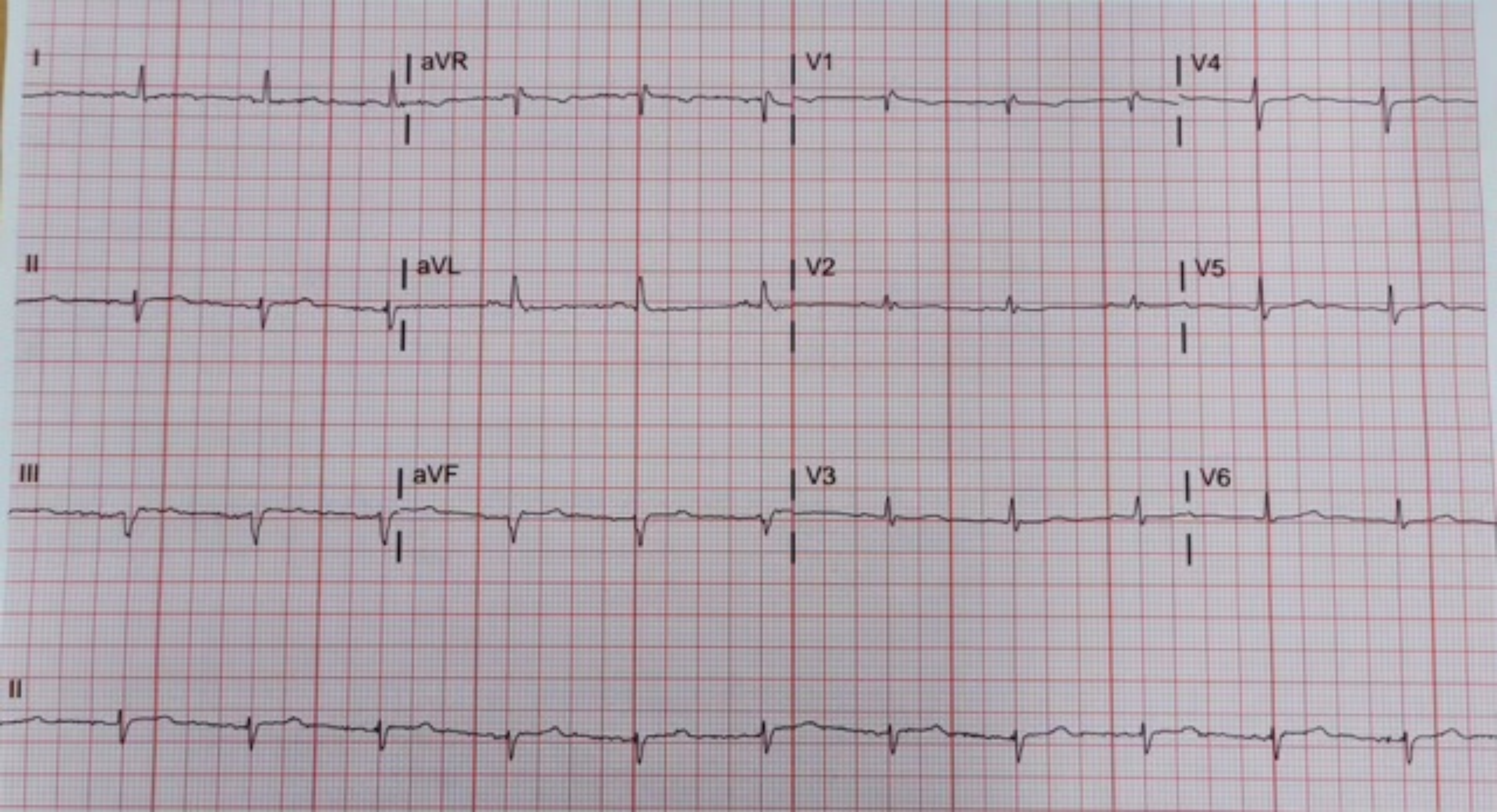
I
aVR
V1
V4
II
aVL
V2
V5
III
aVF
V3
V6
II

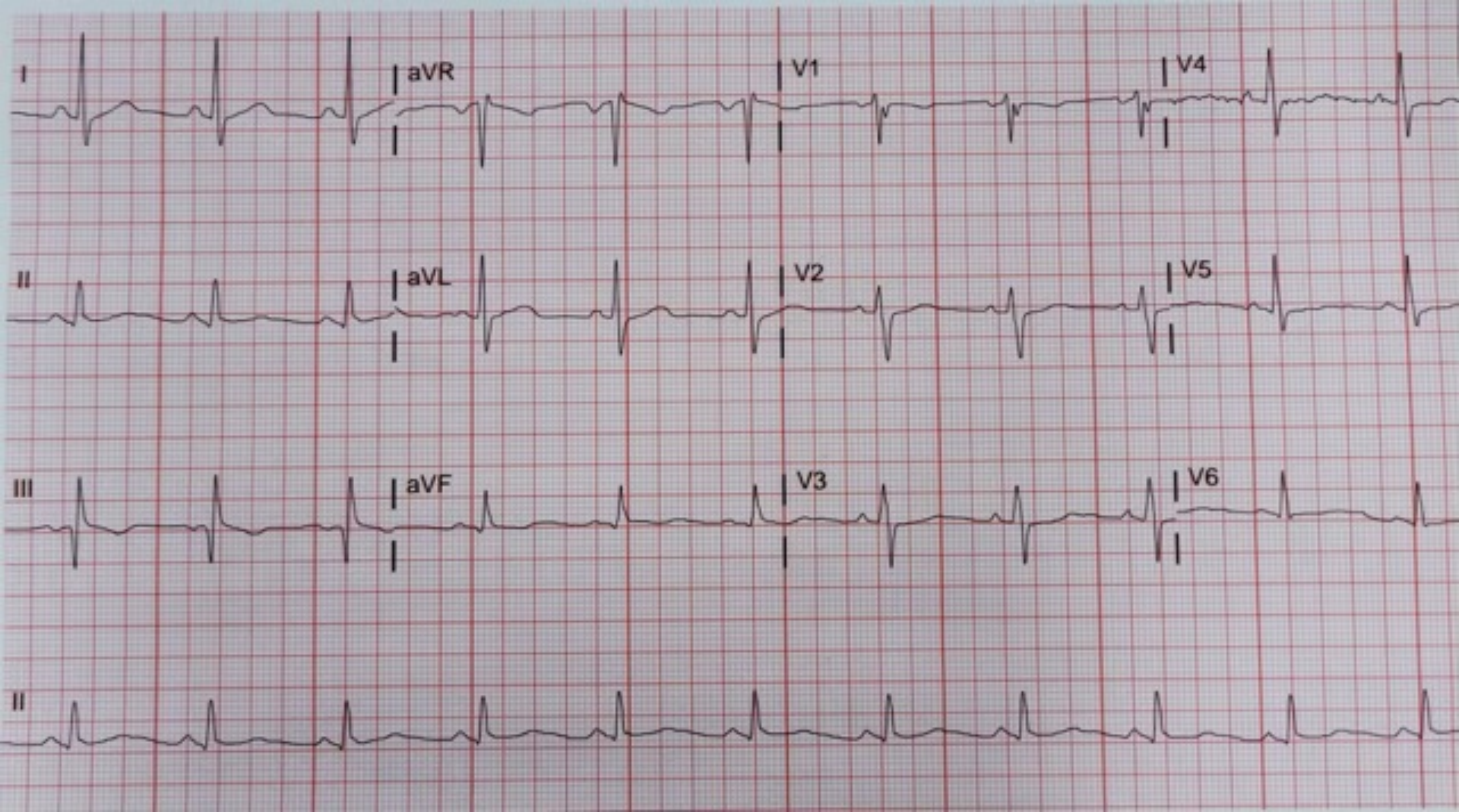
I
aVR
V1
V4
II
aVL
V2
V5
III
aVF
V3
V6
II

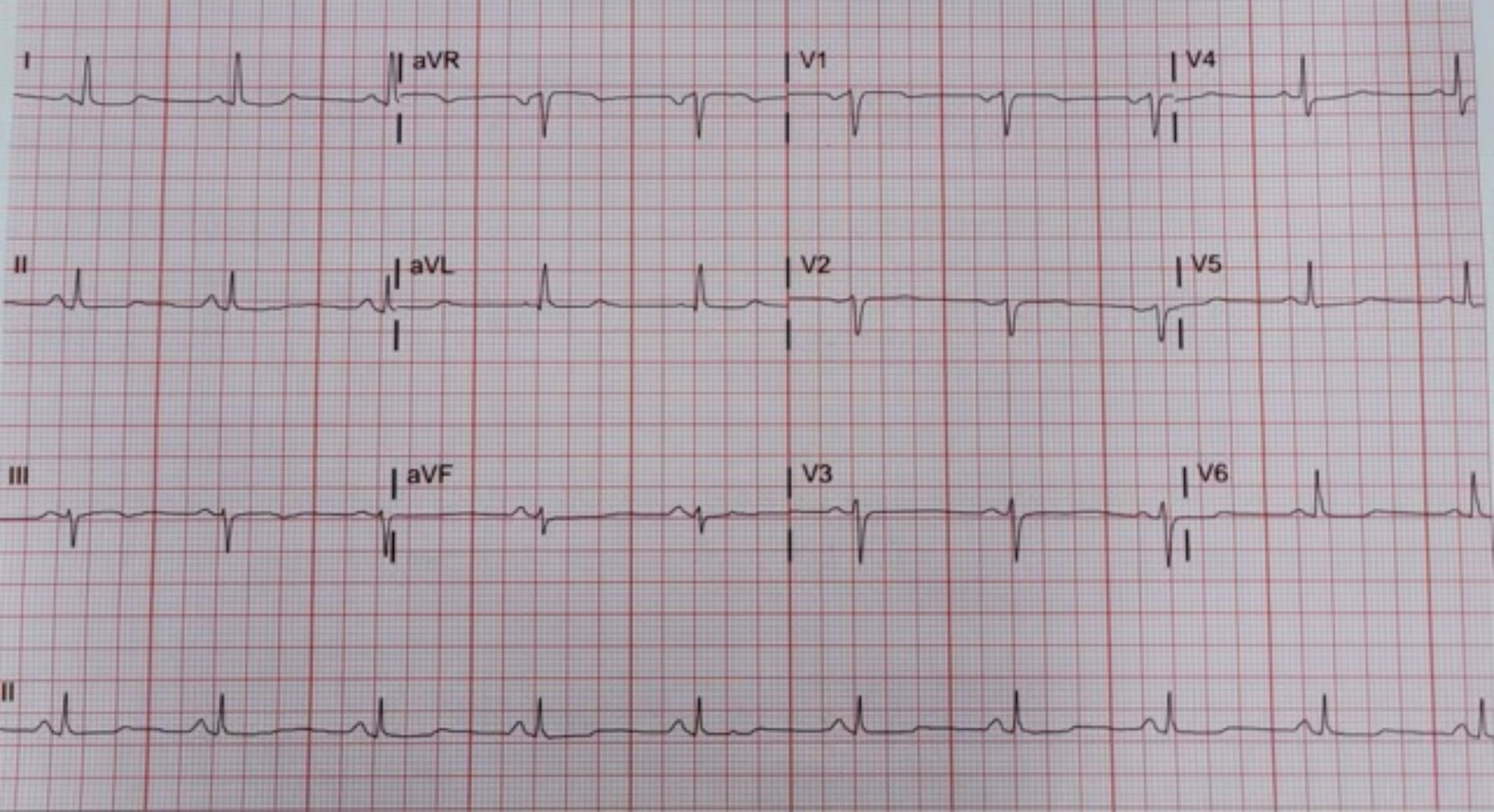
I
aVR
V1
V4
II
aVL
V2
V5
III
aVF
V3
V6
II

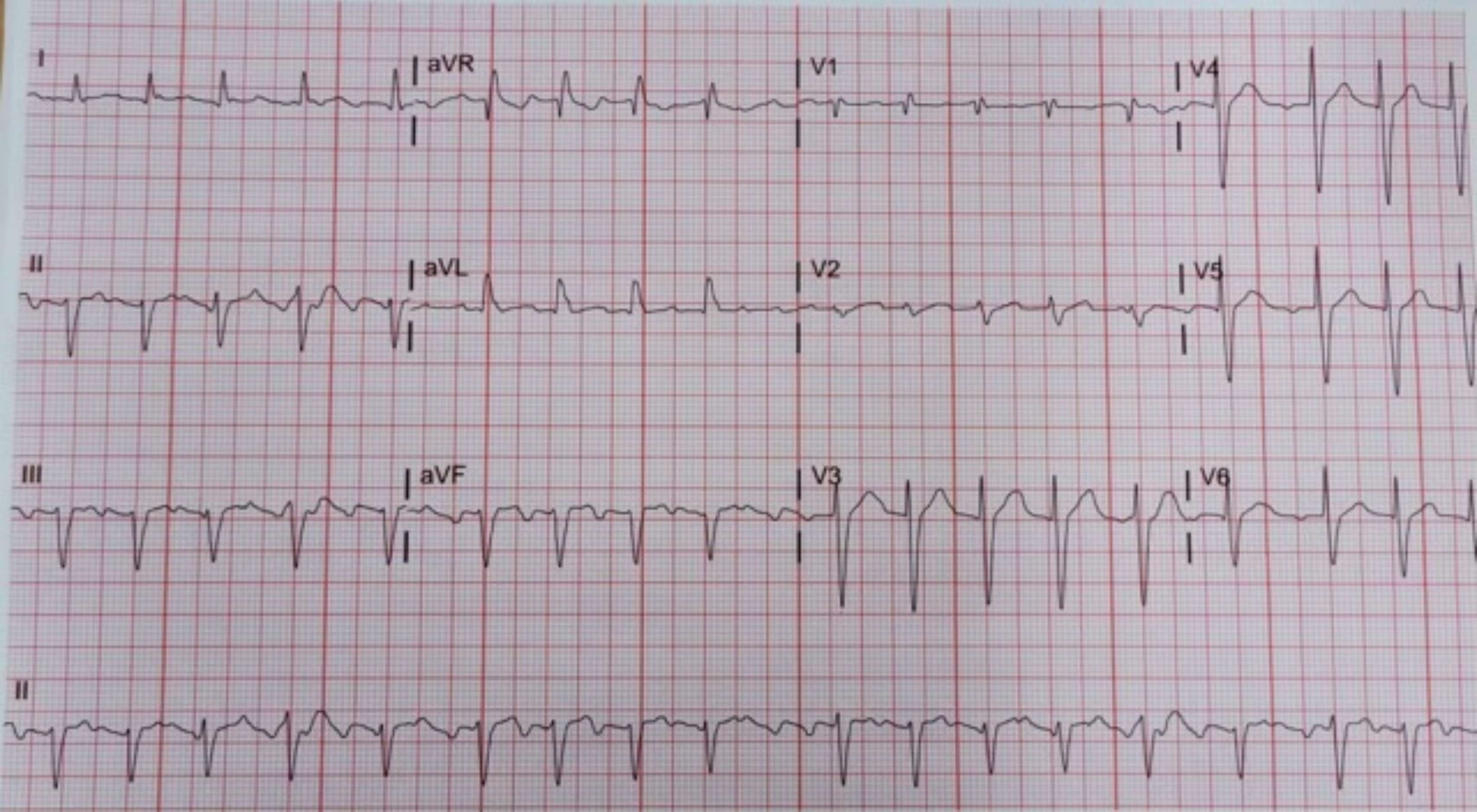

I
aVR
V1
V4
II
aVL
V2
V5
III
aVF
V3
V6
II

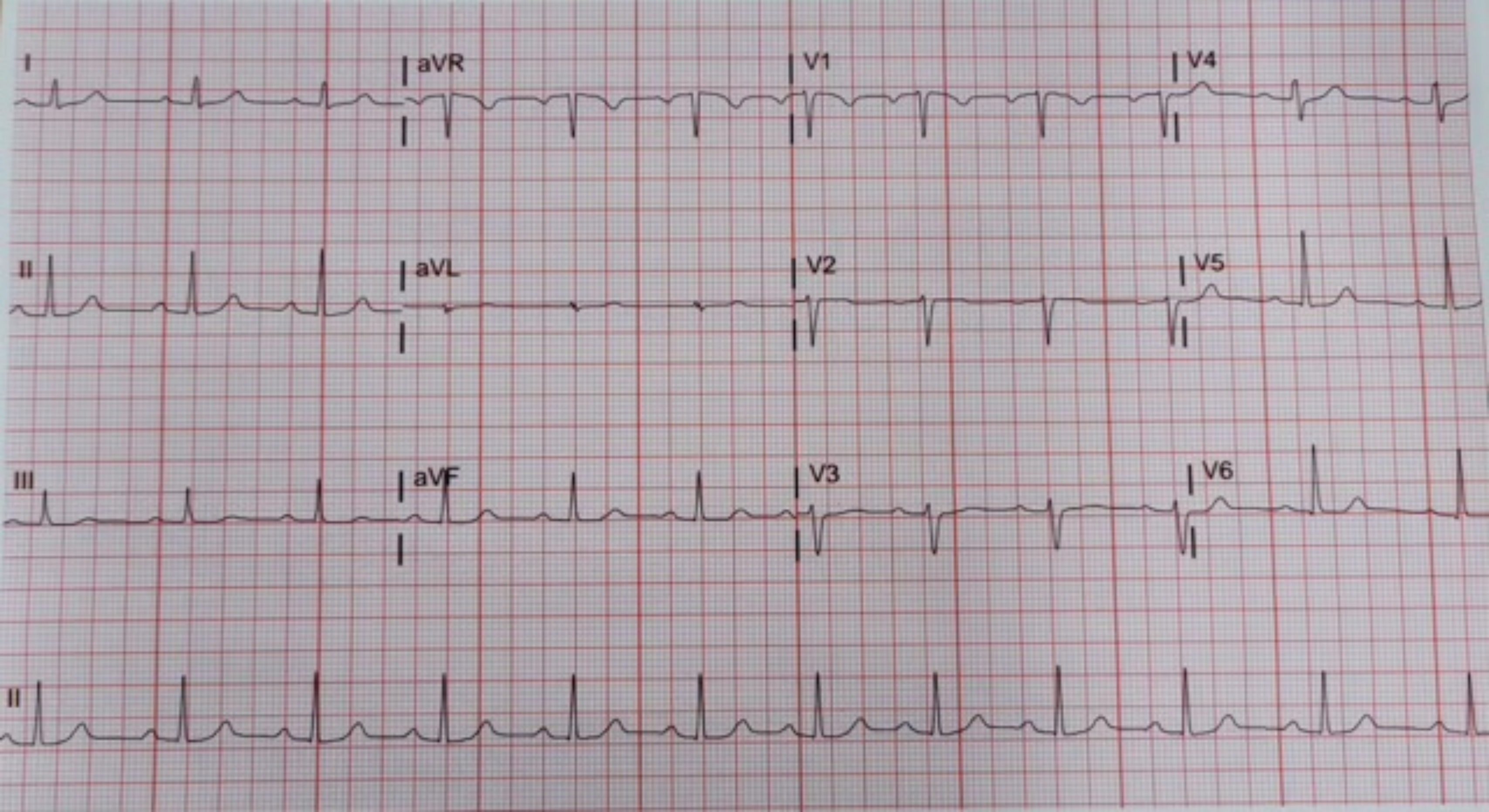
I
aVR
V1
V4
II
aVL
V2
V5
III
aVF
V3
V6
II

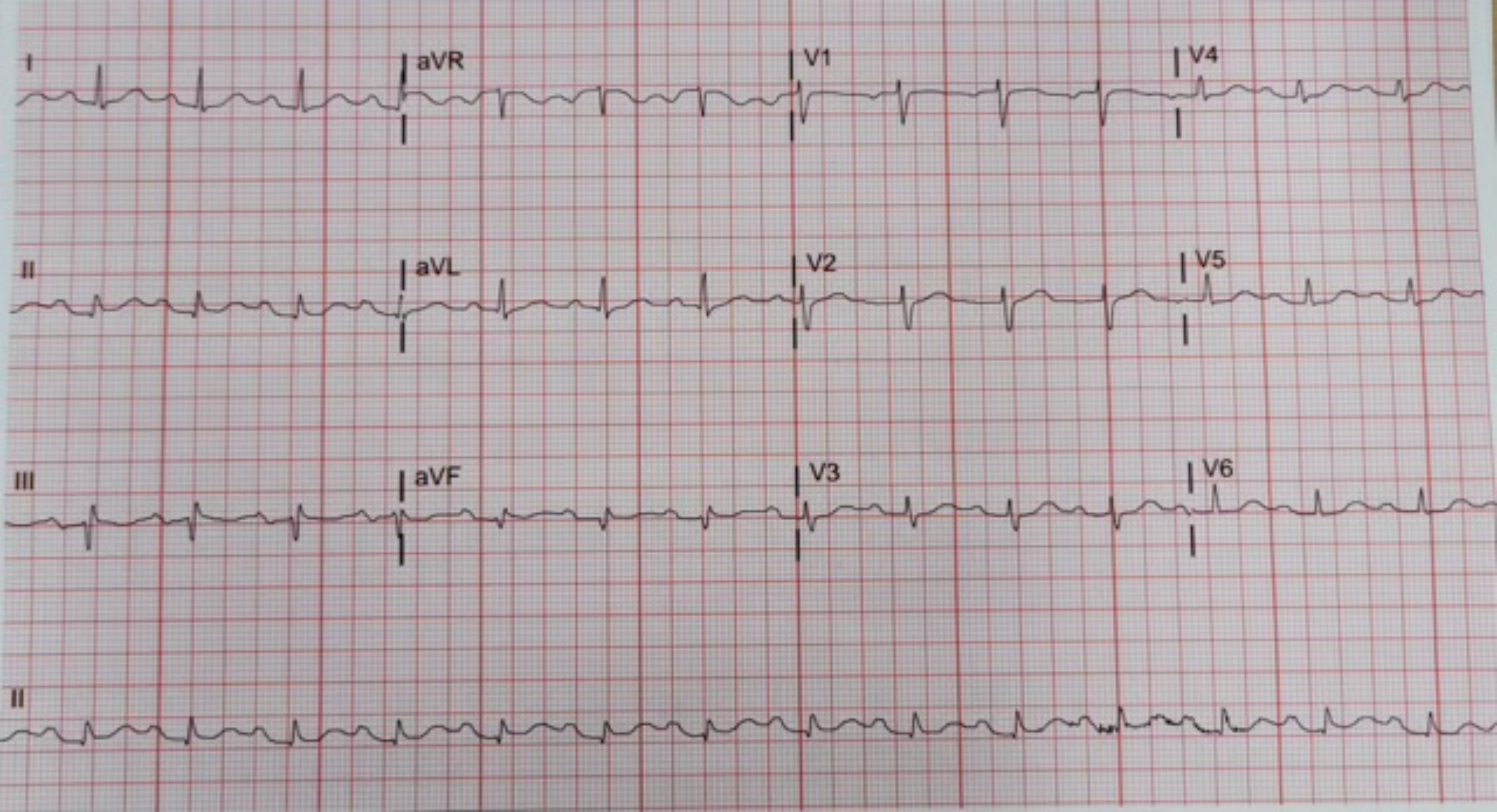
I
aVR
V1
V4
II
aVL
V2
V5
III
aVF
V3
V6
II

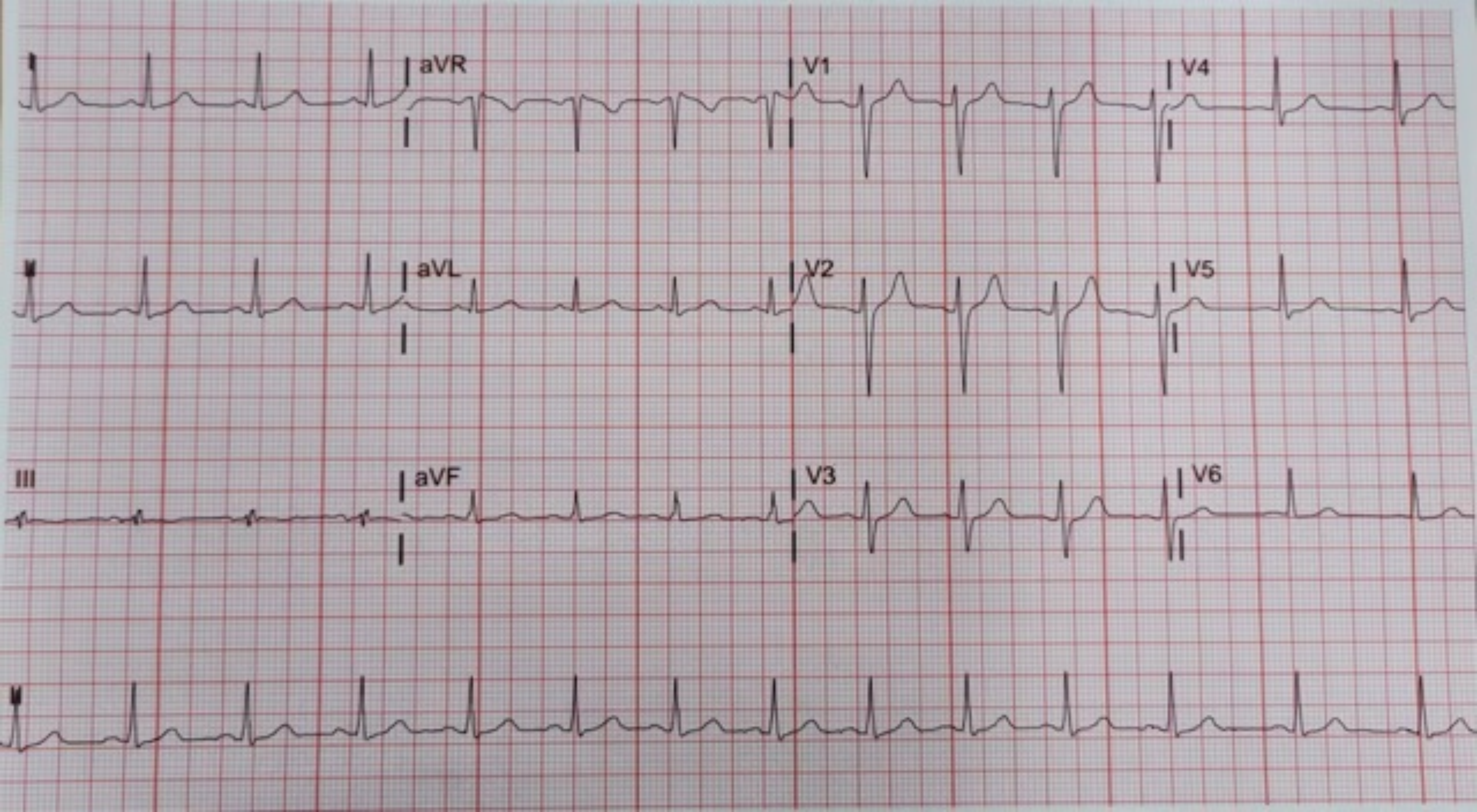
aVR
V1
V4
aVL
V2
V5
III
aVF
V3
V6

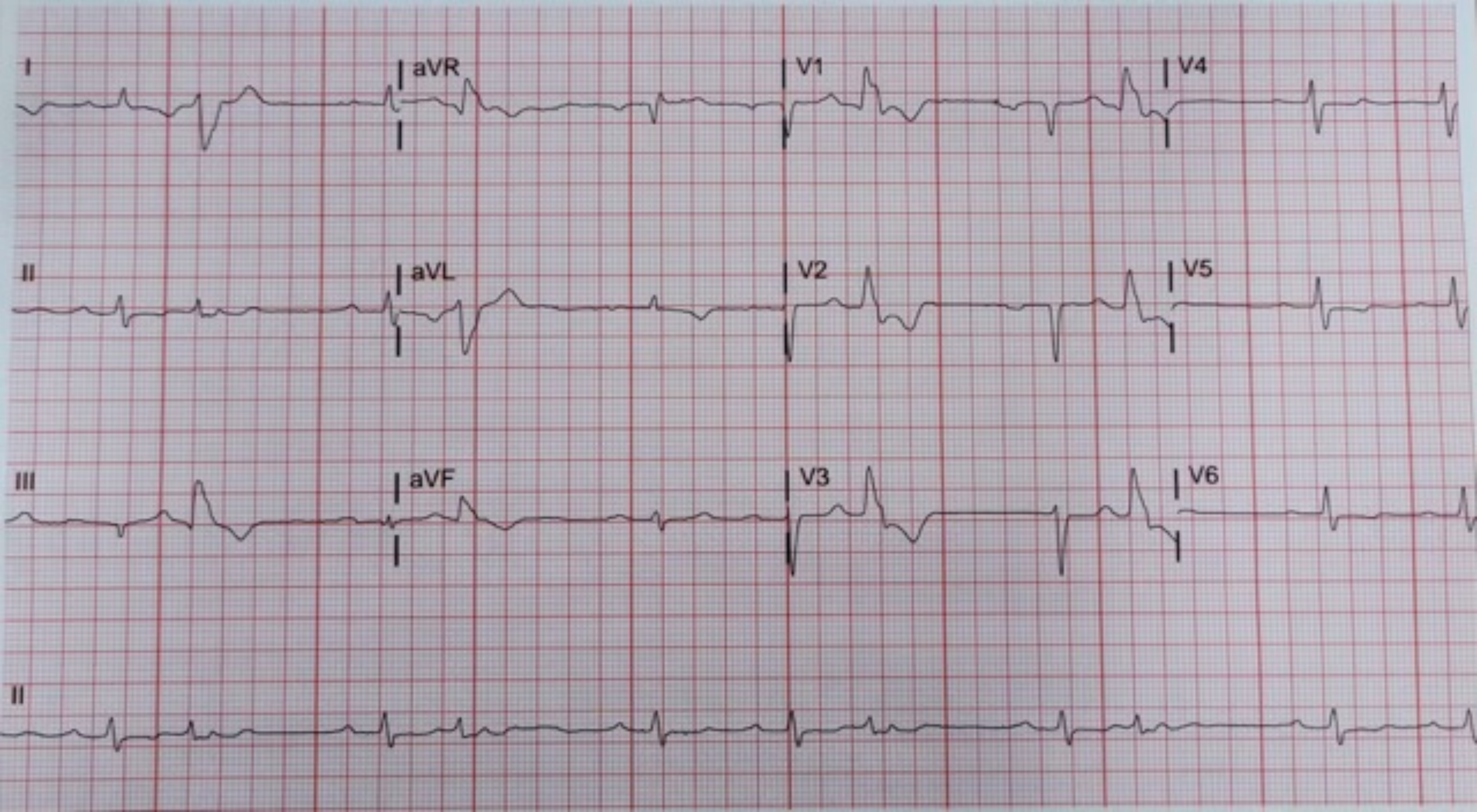
I
aVR
V1
V4
II
aVL
V2
V5
III
aVF
V3
V6
II

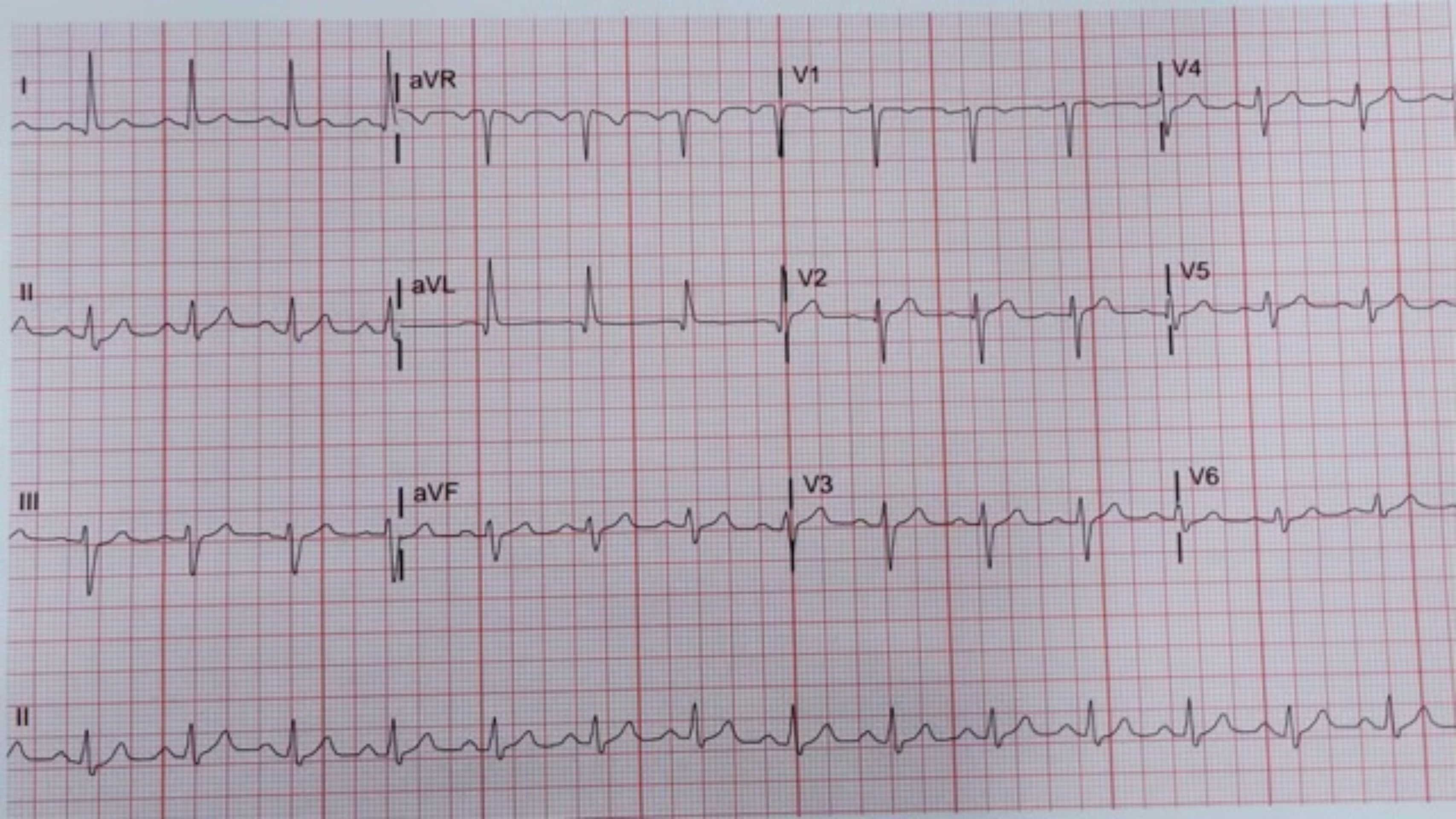
I
aVR
V1
V4
II
aVL
V2
V5
III
aVF
V3
V6
II

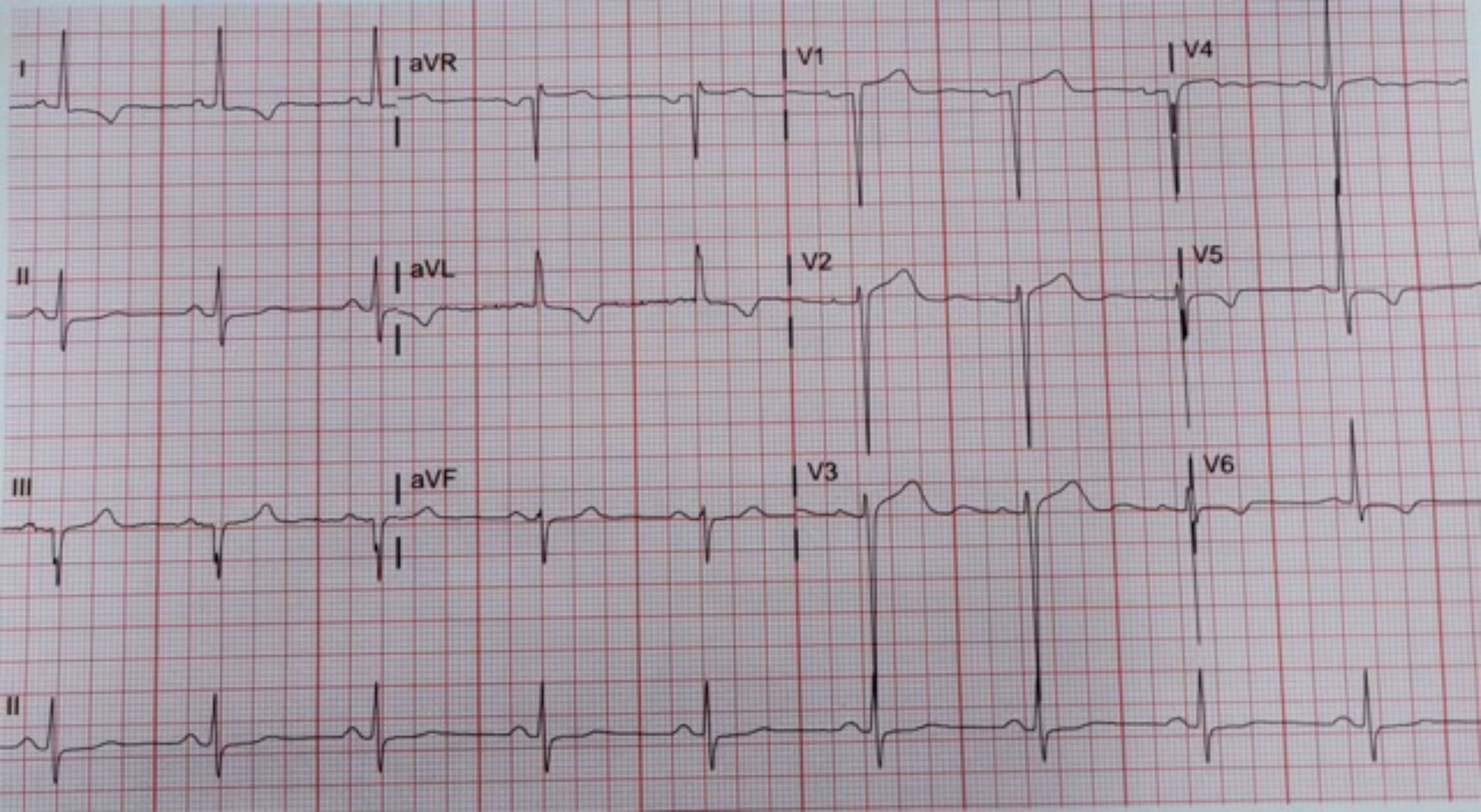
I
aVR
V1
V4
II
aVL
V2
V5
III
aVF
V3
V6
II

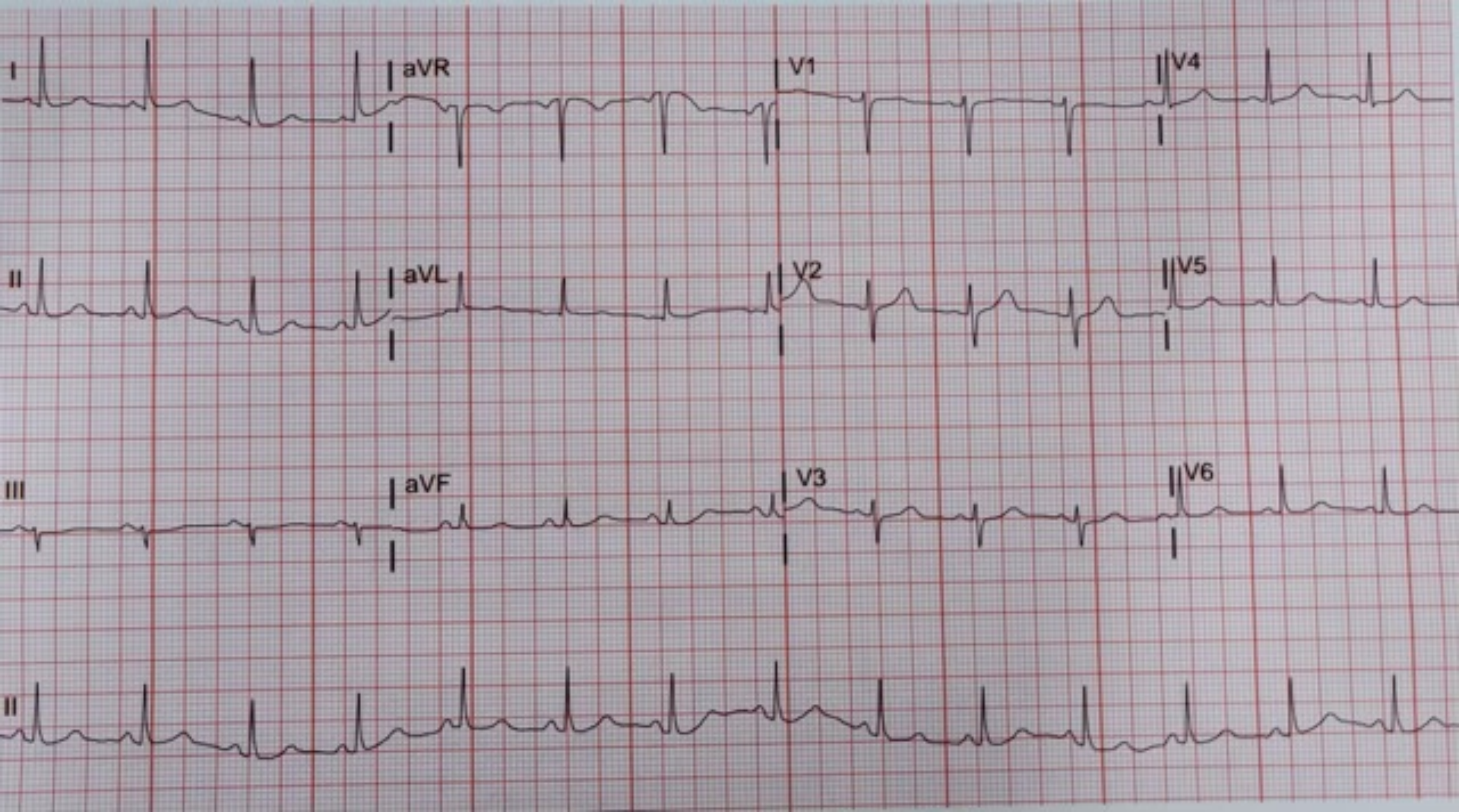
I
aVR
V1
V4
II
aVL
V2
V5
III
aVF
V3
V6
II

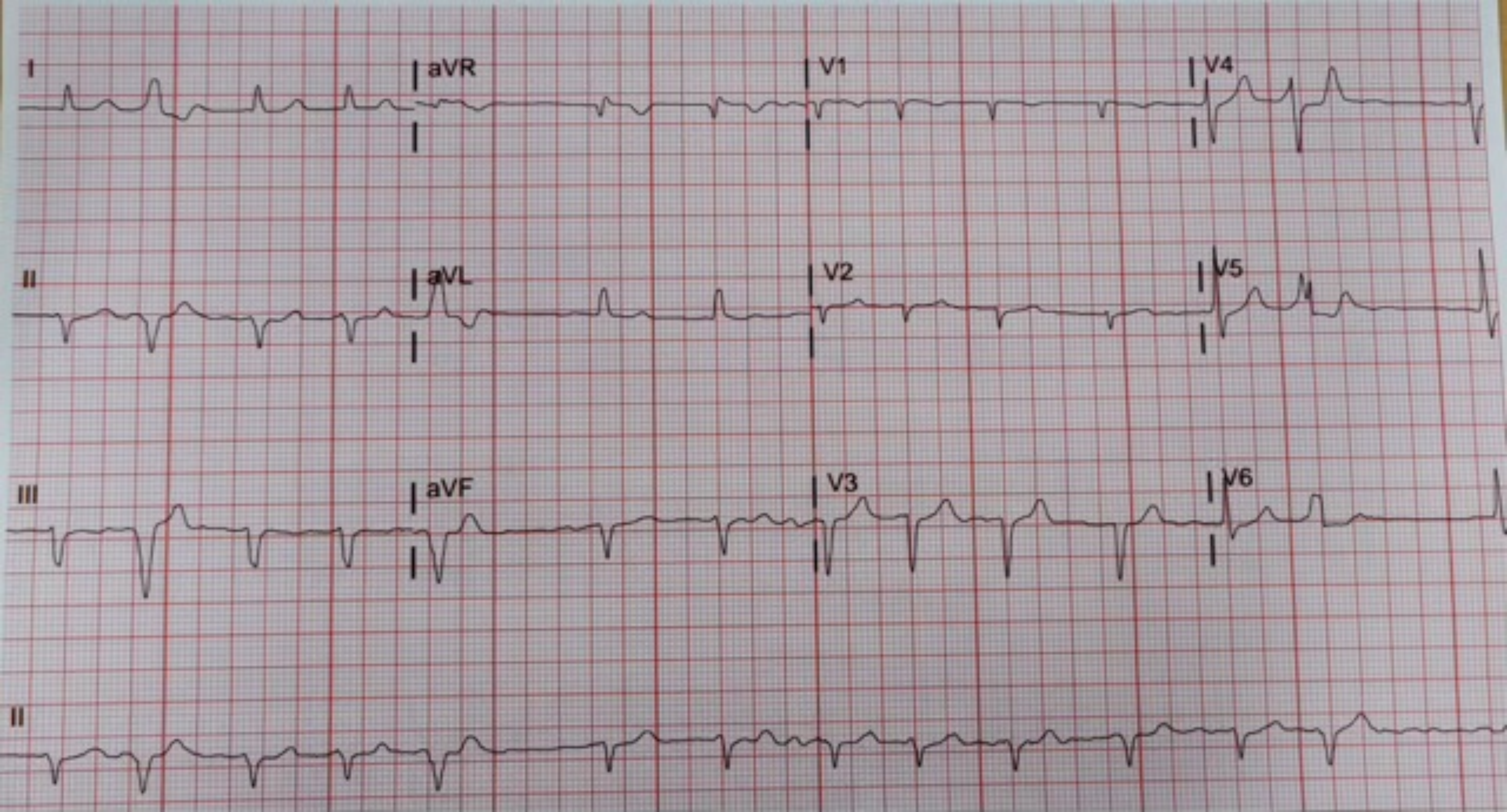
I
aVR
V1
V4
II
aVL
V2
V5
III
aVF
V3
V6
II

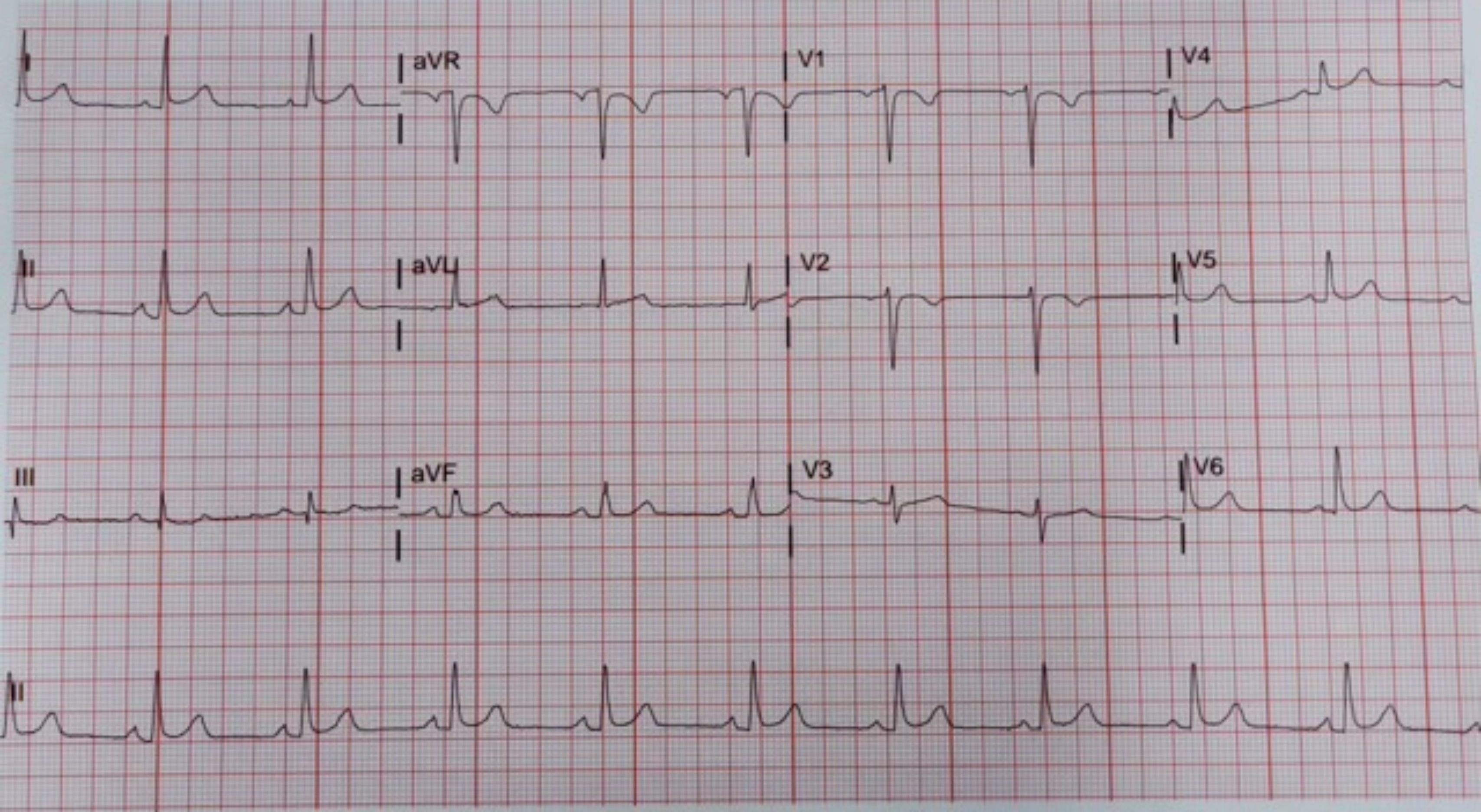
I
aVR
V1
V4
II
aVL
V2
V5
III
aVF
V3
V6
II

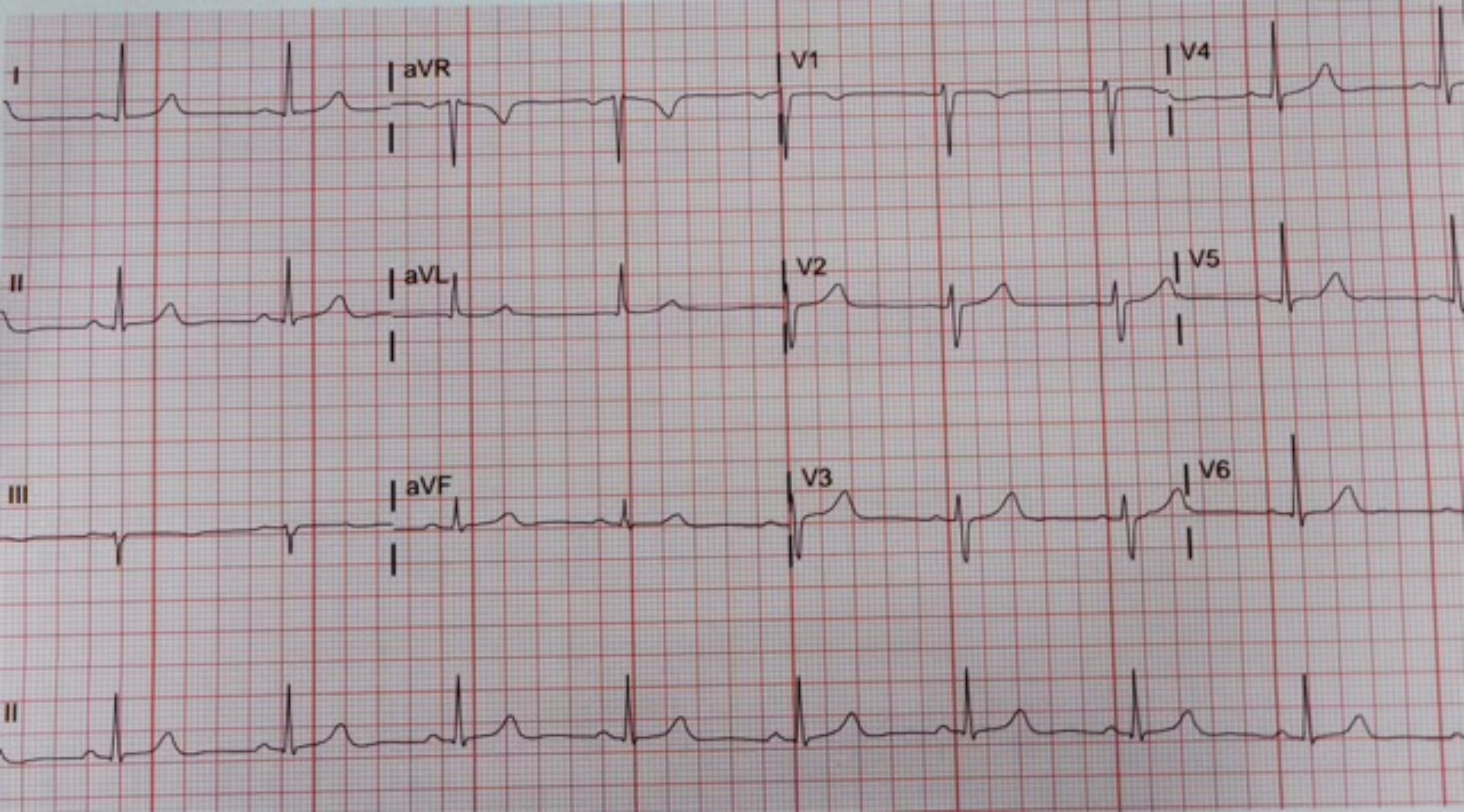
I
aVR
V1
V4
II
aVL
V2
V5
III
aVF
V3
V6
II

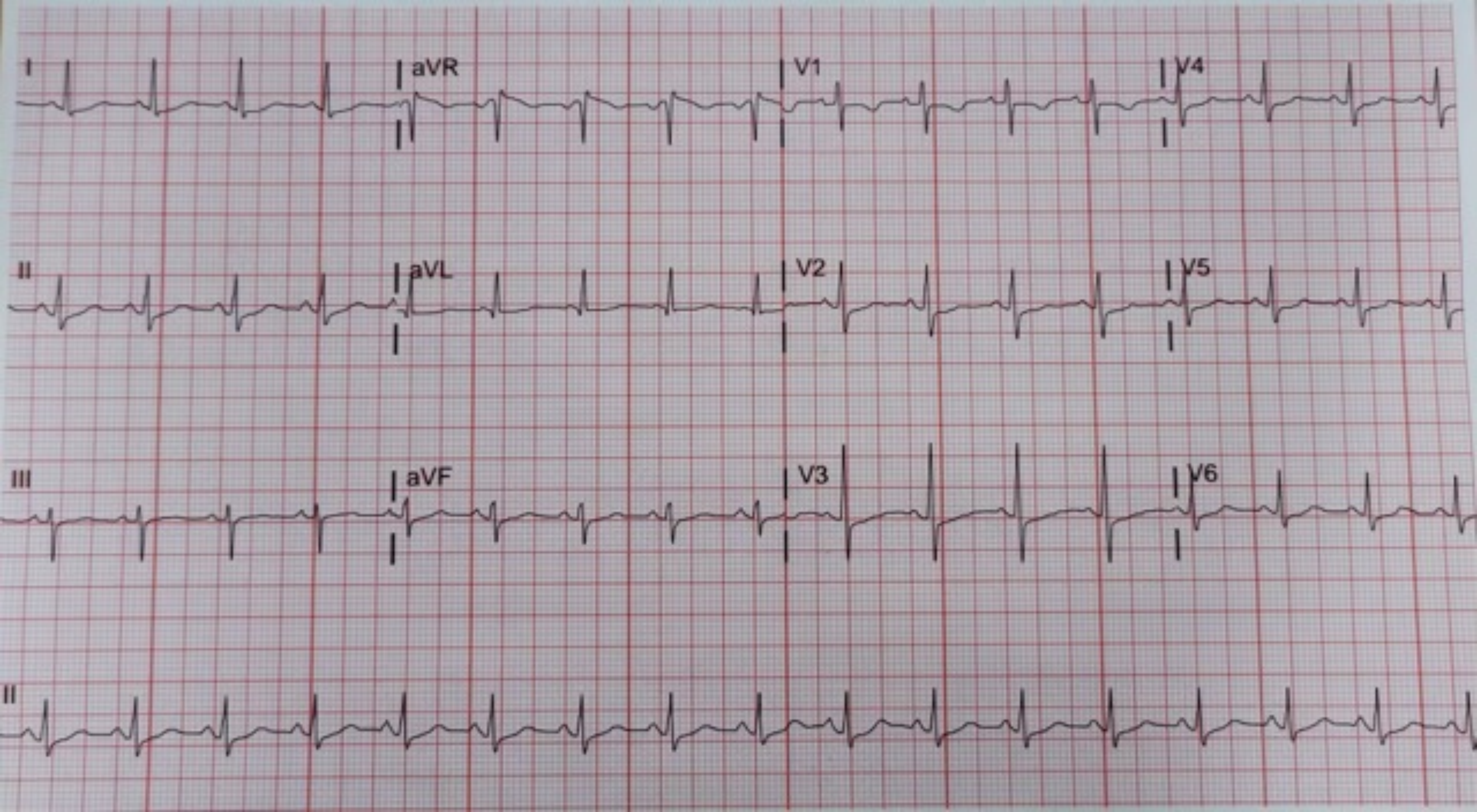
I
aVR
V1
V4
II
aVL
V2
V5
III
aVF
V3
V6
II

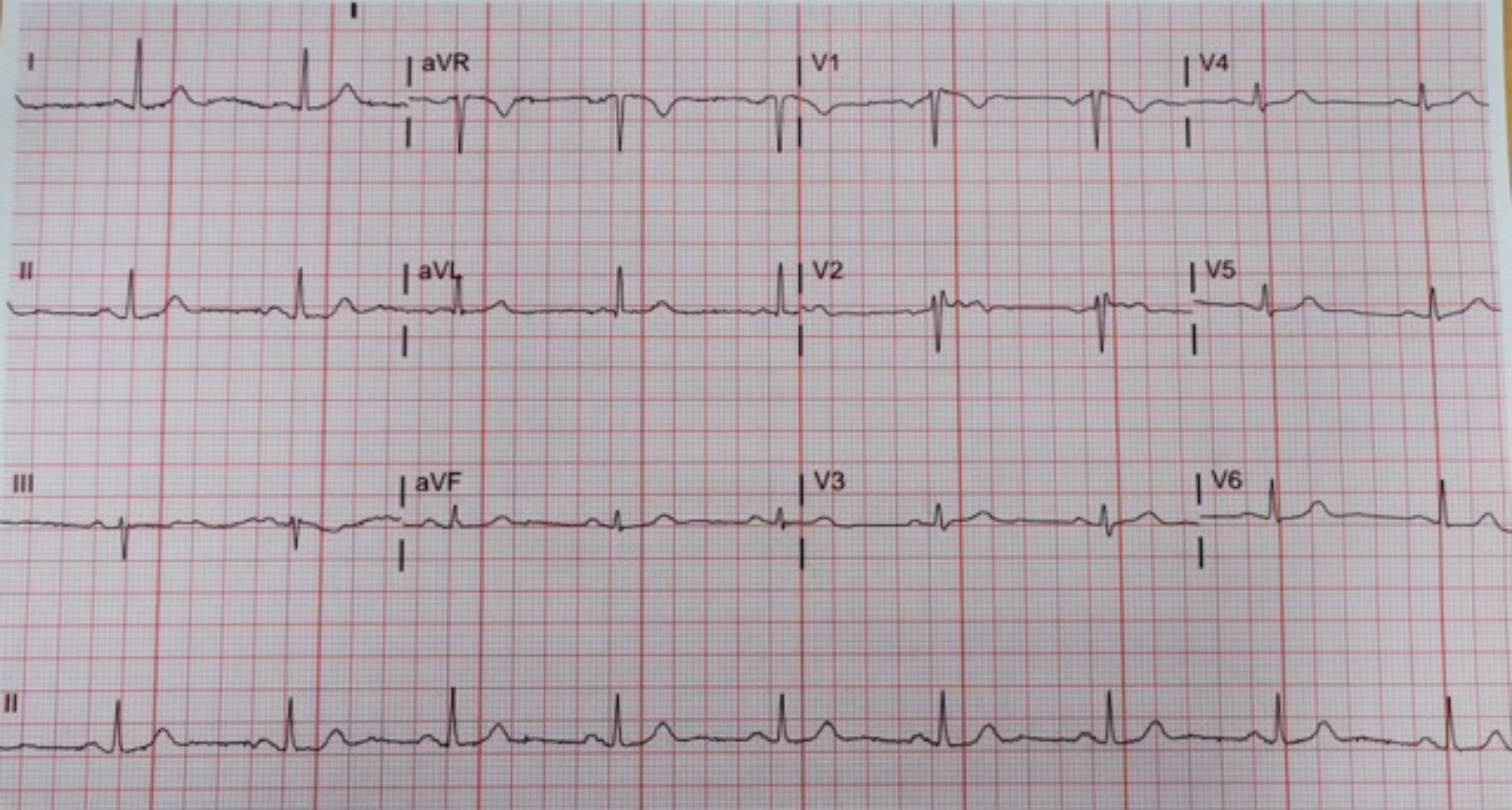
I
aVR
V1
V4
II
aVL
V2
V5
III
aVF
V3
V6
II

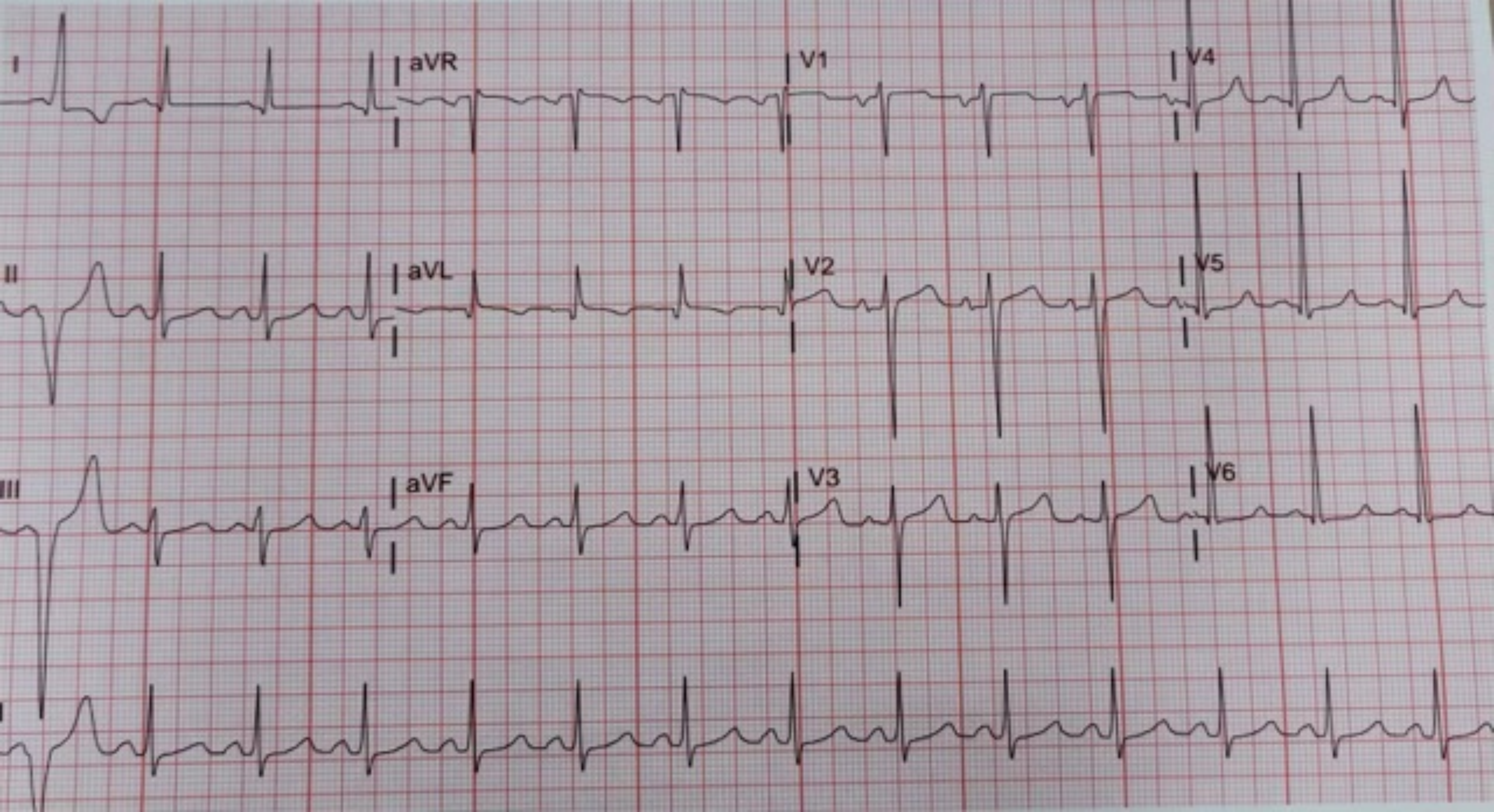
I
aVR
V1
V4
II
aVL
V2
V5
III
aVF
V3
V6
